# Genome-wide characterization of circulating metabolic biomarkers reveals substantial pleiotropy and novel disease pathways

**DOI:** 10.1101/2022.10.20.22281089

**Authors:** Minna K. Karjalainen, Savita Karthikeyan, Clare Oliver-Williams, Eeva Sliz, Elias Allara, Praveen Surendran, Weihua Zhang, Pekka Jousilahti, Kati Kristiansson, Veikko Salomaa, Matt Goodwin, David A. Hughes, Michael Boehnke, Lilian Fernandes Silva, Xianyong Yin, Anubha Mahajan, Matt J. Neville, Natalie R. van Zuydam, Renée de Mutsert, Ruifang Li-Gao, Dennis O. Mook-Kanamori, Ayse Demirkan, Jun Liu, Raymond Noordam, Stella Trompet, Zhengming Chen, Christiana Kartsonaki, Liming Li, Kuang Lin, Fiona A. Hagenbeek, Jouke Jan Hottenga, René Pool, M. Arfan Ikram, Joyce van Meurs, Toomas Haller, Yuri Milaneschi, Mika Kähönen, Pashupati P. Mishra, Peter K. Joshi, Erin Macdonald-Dunlop, Massimo Mangino, Jonas Zierer, Ilhan E. Acar, Carel B. Hoyng, Yara T.E. Lechanteur, Lude Franke, Alexander Kurilshikov, Alexandra Zhernakova, Marian Beekman, Erik B. van den Akker, Ivana Kolcic, Ozren Polasek, Igor Rudan, Christian Gieger, Melanie Waldenberger, Folkert W. Asselbergs, China Kadoorie Biobank Collaborative Group, Estonian Biobank Research Team, FinnGen Consortium, Caroline Hayward, Jingyuan Fu, Anneke I. den Hollander, Cristina Menni, Tim D. Spector, James F. Wilson, Terho Lehtimäki, Olli T. Raitakari, Brenda W.J.H. Penninx, Tonu Esko, Robin G. Walters, J. Wouter Jukema, Naveed Sattar, Mohsen Ghanbari, Ko Willems van Dijk, Fredrik Karpe, Mark I. McCarthy, Markku Laakso, Marjo-Riitta Järvelin, Nicholas J. Timpson, Markus Perola, Jaspal S. Kooner, John C. Chambers, Cornelia van Duijn, P. Eline Slagboom, Dorret I. Boomsma, John Danesh, Mika Ala-Korpela, Adam S. Butterworth, Johannes Kettunen

## Abstract

Genome-wide association analyses using high-throughput metabolomics platforms have led to novel insights into the biology of human metabolism^1–7^. This detailed knowledge of the genetic determinants of systemic metabolism has been pivotal for uncovering how genetic pathways influence biological mechanisms and complex diseases^8–11^. Here we present a genome-wide association study of 233 circulating metabolic traits quantified by nuclear magnetic resonance spectroscopy in up to 136,016 participants from 33 predominantly population-based cohorts. We discover over 400 independent loci and assign likely causal genes at two-thirds of these using detailed manual curation of highly plausible biological candidates. We highlight the importance of sample- and participant characteristics, such as fasting status and sample type, that can have significant impact on genetic associations, revealing direct and indirect associations on glucose and phenylalanine. We use detailed metabolic profiling of lipoprotein- and lipid-associated variants to better characterize how known lipid loci and novel loci affect lipoprotein metabolism at a granular level. We demonstrate the translational utility of comprehensively phenotyped molecular data, characterizing for the first time the metabolic associations of an understudied phenotype, intrahepatic cholestasis of pregnancy. Finally, we observe substantial genetic pleiotropy for multiple metabolic pathways and illustrate the importance of careful instrument selection in Mendelian randomization analysis, revealing a putative causal relationship between acetoacetate and hypertension. Our publicly available results provide a foundational resource for the community to examine the role of metabolism across diverse diseases.

## MAIN TEXT

Large genome-wide association studies (GWASs) coupled with metabolic profiling platforms have successfully identified many loci associated with circulating metabolic traits^1–7,12–16^. For example, studies combining genomics with detailed metabolic profiling from a high-throughput metabolomics platform based on nuclear magnetic resonance (NMR) spectroscopy^17^ have allowed the identification of dozens of loci for circulating lipid, lipoprotein and fatty acid traits and small molecules such as amino acids^2,4,5,9,18,19^. These studies have provided novel insights into the biology of human metabolism and have guided large-scale epidemiological studies, such as Mendelian randomization analyses to infer causal relationships^17^. Here, using the same NMR metabolomics platform from Nightingale Health with an updated quantification version, we considerably extend our previous GWAS^4^ of 123 circulating metabolic traits in up to ∼25,000 participants to study of 233 traits in more than 135,000 participants.

### Genetic discovery

GWAS was performed under the additive model separately in each of 33 cohorts (Supplementary Table S1). Subsequent meta-analysis involved 233 metabolic traits (Supplementary Table S2), including 213 lipid and lipoprotein parameters or fatty acids, and 20 non-lipid traits (amino acids, ketone bodies, and glycolysis/gluconeogenesis, fluid balance and inflammation-related metabolites). After variant filtering and quality control, up to 13,389,637 imputed autosomal single-nucleotide polymorphisms (SNPs) were included in the meta-analysis in up to 136,016 participants.

In the meta-analysis, we detected genome-wide significant associations for all 233 metabolic traits (Supplementary Figures S1-S3, Supplementary Tables S4 and S5) with extensive pleiotropy and polygenicity. We detected 276 broad regions (defined as a +/-500 Kb region around the set of genome-wide significant SNPs) associated with at least one metabolic trait (Figure 1A, Supplementary Table S4). Eighty-six of these regions were associated with just a single metabolic trait, whereas most regions harbored associations with multiple traits (Figure 1B and 1C), up to a maximum of 214 associated traits at the well-characterized lipid-associated *APOE* region. The lipid and lipoprotein traits were mostly demonstrably polygenic, with some traits having associations at >60 loci, whereas most non-lipid traits had substantially fewer associated loci (<20), including three glucose-metabolism related traits (lactate, pyruvate, glycerol) having fewer than five associated loci (Supplementary Table S5). The non-lipid traits accounted for most of the regions with a single associated trait (*n*=67; 79%), and the majority (*n*=163; 57%) of the regions with non-lipid trait associations had fewer than five associated metabolic traits in total. By contrast, the lipid, lipoprotein and fatty acid trait-associated regions (*n*=186) were generally more pleiotropic with 75% (*n*=140) of the regions being associated with five or more traits. Within the 276 regions, we found 8,795 lead SNP – lead trait associations corresponding to 1,447 unique lead SNPs (Supplementary Table S5). After resolving independent signals based on pairwise linkage disequilibrium, we concluded that the 276 broad regions involved at least 443 independent loci.

**Figure 1.**
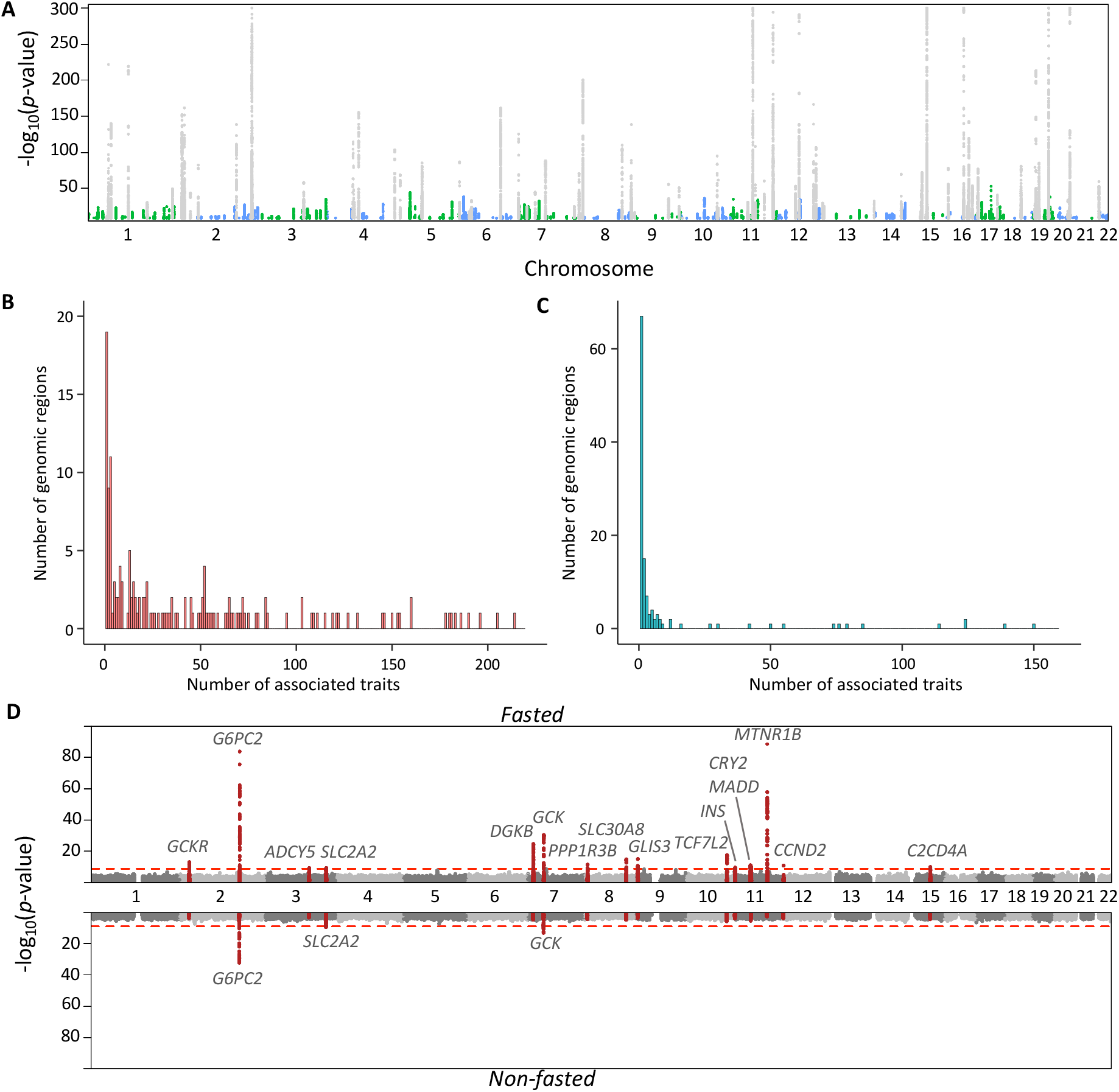
Results of the GWAS meta-analysis of 233 metabolic traits. The metabolic trait associations are summarized in a Manhattan plot (panel A). Loci that do not overlap with those identified in the previous large-scale NMR metabolomics GWAS^4,5^ are shown in blue and green. Only genome-wide significant SNPs (p < 1.8 × 10^−9^) are shown and -log_10_(p-values) were capped at 300. Numbers of associated metabolic traits at the 276 associated genomic regions are shown separately for genomic regions in which the lead trait was a lipid, lipoprotein or fatty acid trait (155 loci; median 24 traits per locus; panel B) and for those in which the lead trait was a non-lipid trait (121 loci; median one trait per locus; panel C). Results of the genome-wide association study of glucose are shown in panel D separately for the fasted (top) cohorts (total n=68,559) and non-fasted (total n=58,112) cohorts. The red line indicates the threshold for genome-wide significance. 500-kb regions around lead SNPs in the fasted cohorts are highlighted in both top and bottom panels.

### Associations in UK Biobank and the effects of fasting and sample type

The availability of NMR data from the UK Biobank resource^20^ (March 2021 release) allowed us to check for associations of the lead variants in an independent population and to assess the effects of participant characteristics and sample-related factors on our associations. Of the 8,502 lead SNP – metabolic trait pairs that could be tested in up to 115,078 UK Biobank European-ancestry participants, 5,443 (64.0%) associated at *p* <5 × 10^−8^, while a further 777 (9.1%; 332 unique SNPs) associated at *p* <1 × 10^−5^ (Supplementary Table S6). In addition to subtle differences in population ancestry between the studies, we identified sample type and fasting status as major drivers of non-replication. The UK Biobank NMR measurements were performed on EDTA plasma samples, whereas the current meta-analysis involved predominantly serum samples. For example, several of non-replicating associations with phenylalanine were in coagulation-related loci (e.g., *KLKB1, F12, KNG1, FGB*) but these signals were absent in UK Biobank (Supplementary Table S6; Supplementary Figure S4), suggesting that the removal of clotting factors in the preparation of serum can reveal associations with phenylalanine via coagulation. Similarly, we found associations with glucose that did not replicate in the UK Biobank, including a well-known association at *MTNR1B* (melatonin receptor 1B)^21^, a key regulator in glucose metabolism (rs10830963; meta-analysis *p*-value=1.5 × 10^−60^; UK Biobank *p*-value=0.60). The UK Biobank predominantly includes non-fasted samples, but the current meta-analysis mainly consists of cohorts (27 cohorts) with fasted samples (Supplementary Table S1). We therefore conducted a fasting-stratified meta-analysis, which suggested that some of these associations were driven by cohorts with predominantly fasted samples (Figure 1D, Supplementary Figure S5) and hence are absent in UK Biobank. In addition to *MTNR1B* rs10830963 (*p*-values 2.9 × 10^−89^ and 0.57 in meta-analysis of fasted and non-fasted cohorts, respectively), the association of which was also previously shown to be absent in non-fasting samples^22^, *GLIS3* (GLIS family zinc finger 3, a known diabetes risk gene^23^ with a role in pancreatic β-cell biology) rs10974438 represents another example of an association that was not robustly replicated in UK Biobank (meta-analysis *p*-value=4.0 × 10^−14^; UK Biobank *p*-value=0.001) and was characterized by the absence of signals in the non-fasted cohorts (*p*-values 1.1 × 10^−15^ and 0.14 in meta-analysis of fasted and non-fasted cohorts; Supplementary Figure S5). We note that the effects of the sample type and fasting status require careful consideration when interpreting the results of GWAS of metabolic traits and conducting downstream analyses, such as Mendelian randomization studies using trait-associated variants as instruments.

### Novel loci and candidate genes

We conducted extensive manual curation to prioritize 231 likely causal genes with clear biological relevance to the associated trait(s) at 297 (67.0%) of the 443 loci (Methods). As some regions were extremely complex and pleiotropic due to overlapping genetic associations of up to 11 independent lead variants with heterogeneous associations across the metabolic traits, we characterized these loci in detail to pinpoint potential multiple likely causal genes within each locus (Supplementary Table S5). For example, in a 7.6-Mb region on chromosome 16 with 139 associated metabolic traits, we identified six distinct biologically relevant potential causal genes: *LCAT* (lecithin-cholesterol acyltransferase, associated with multiple lipoprotein subclass measures), *SLC7A6* (solute carrier family 7 member 6, associated with acetate and creatinine), *PDPR* (pyruvate dehydrogenase phosphatase regulatory subunit, associated with pyruvate), *AARS* (alanyl-tRNA synthetase 1, associated with amino acids), *TAT* (tyrosine aminotransferase, associated with tyrosine) and *HP* (haptoglobin, associated with a range of lipoprotein subclass measures, fatty acids, cholesterol, apolipoprotein B and glycoprotein acetylation). This locus exemplifies the complexity of the metabolic trait-associated loci. For additional loci without an obvious biological candidate, we assigned a further 39 likely causal genes based on SNP function or the presence of likely functional (missense, stop gained or splice region) variants in strong LD (*r*^*2*^≥0.8) with the lead variant (Supplementary Table S5).

We performed an extensive comparison of the discovered associations to previously reported genetic associations of metabolic traits and traditional clinical lipids (HDL-C, LDL-C, triglycerides, total cholesterol; Supplementary Table S5). In comparison to previous large-scale NMR metabolomics GWASs^4,5^, we identified 212 additional associated genomic regions (Supplementary Table S4). These included 138 novel genomic regions for the lipoprotein, lipid, and fatty acid traits, and 113 novel regions associated with the non-lipid traits. New associations for several lipoprotein subclass measures were detected in loci previously associated with clinical lipids, such as the locus containing *LDLRAP1* encoding low density lipoprotein receptor adapter protein 1, which is involved in cholesterol metabolism. This locus was previously known to be associated with LDL-C and total cholesterol^24,25^, and we found associations at this locus with several lipoprotein subclass measures, lipids and fatty acids (Supplementary Table S5). Our analyses also identified genetic associations with detailed lipoprotein subclass measures in loci not reported to be associated with traditional clinical lipids: *ACOX1* (encoding Peroxisomal acyl-coenzyme A oxidase 1 with a function in fatty acid oxidation), *SOAT2* (encoding sterol O-acyltransferase 2 with a function in cholesterol metabolism), and *ST3GAL6* (encoding Type 2 lactosamine alpha-2,3-sialyltransferase with a function in glycolipid metabolism) represent examples of biologically plausible genes associated with a range of lipoprotein subclass measures, lipids, and apolipoprotein B.

Novel loci were also detected for the small molecules, such as phenylalanine and glutamine. For phenylalanine, we detected associations at 13 loci. Novel phenylalanine-associated loci include both a well-known metabolic trait-associated locus (*FADS1/FADS2*) and two novel, biologically plausible loci (*GSTA2, SLC2A4RG*). For example, *SLC2A4RG* encodes SLC2A4 regulator, a transcription factor involved in the activation of SLC2A4 (GLUT4), a key regulator of glucose transport. For glutamine, we detected associations at 26 loci. Interestingly, seven of the loci were only associated with glutamine (*GLS, PLCL1, SFXN1, KCNK16, MED23, SLC25A29, PCK1*). Thus, these associations likely represent biology local to glutamine, most of the loci having biologically plausible candidate genes with roles in glutamine metabolism (*GLS*), amino acid transport (*SFXN1, SLC25A29*) or glucose and gluconeogenesis-related pathways (*PCK1, KCNK16*). *KCNK16*, a known type 2 diabetes susceptibility gene encoding potassium two pore domain channel subfamily K member 16, is a pancreatic potassium channel, and represents an example of a novel glutamine-associated locus with a role in glucose biology^26,27^.

### Characterizing metabolic effects of apolipoprotein B-associated variants

To provide insights into the distinct ways in which lipid loci can affect the continuum of lipoprotein metabolism, we characterized clusters of genes with similar metabolic association profiles. The effect estimates were scaled by dividing all effect estimates of a given SNP using the strongest association effect estimate across all metabolic associations in each locus. This way, the scaled effect estimates for all SNPs were between -1 and 1, and the statistical strength of an association affects the clustering less while more emphasis is given to the association landscape in guiding the clustering. We concentrated on 134 loci with nominal evidence (*p* < 0.05) of an association with apolipoprotein B (apoB), as recent studies have highlighted the predominant role of apoB in coronary artery disease etiology^28–30^. Despite the strong correlation structure within the lipid and apolipoprotein traits, we identified several loci with association patterns that do not follow the between-trait correlation structure (Figure 2A, Supplementary Figures S6 and S7). For example, some loci (*APOC1, TIMD4*) are strongly associated with all the apoB-containing particles (VLDL, IDL, and LDL), while other loci are predominantly associated with IDL and LDL particles (*PCSK9, HMGCR, TRIM5*), with VLDL and the largest HDL particles (*IRS1, CD300LG*), or with medium and small HDL particles (*APOA2, CERS2*). Several SNPs also exhibit discordant associations within highly correlated metabolic traits (e.g., *LPA* and *APOH* within apoB-containing particles and *FADS1-2-3* within both apoB-containing and HDL particles; Figure 2A). Genes known to play a role in LDL-related metabolism (*PCSK9, APOB, HMGCR, LDLR*) clustered closely together, demonstrating that our clustering approach is reflecting at least some known biological similarity (Figure S7).

**Figure 2.**
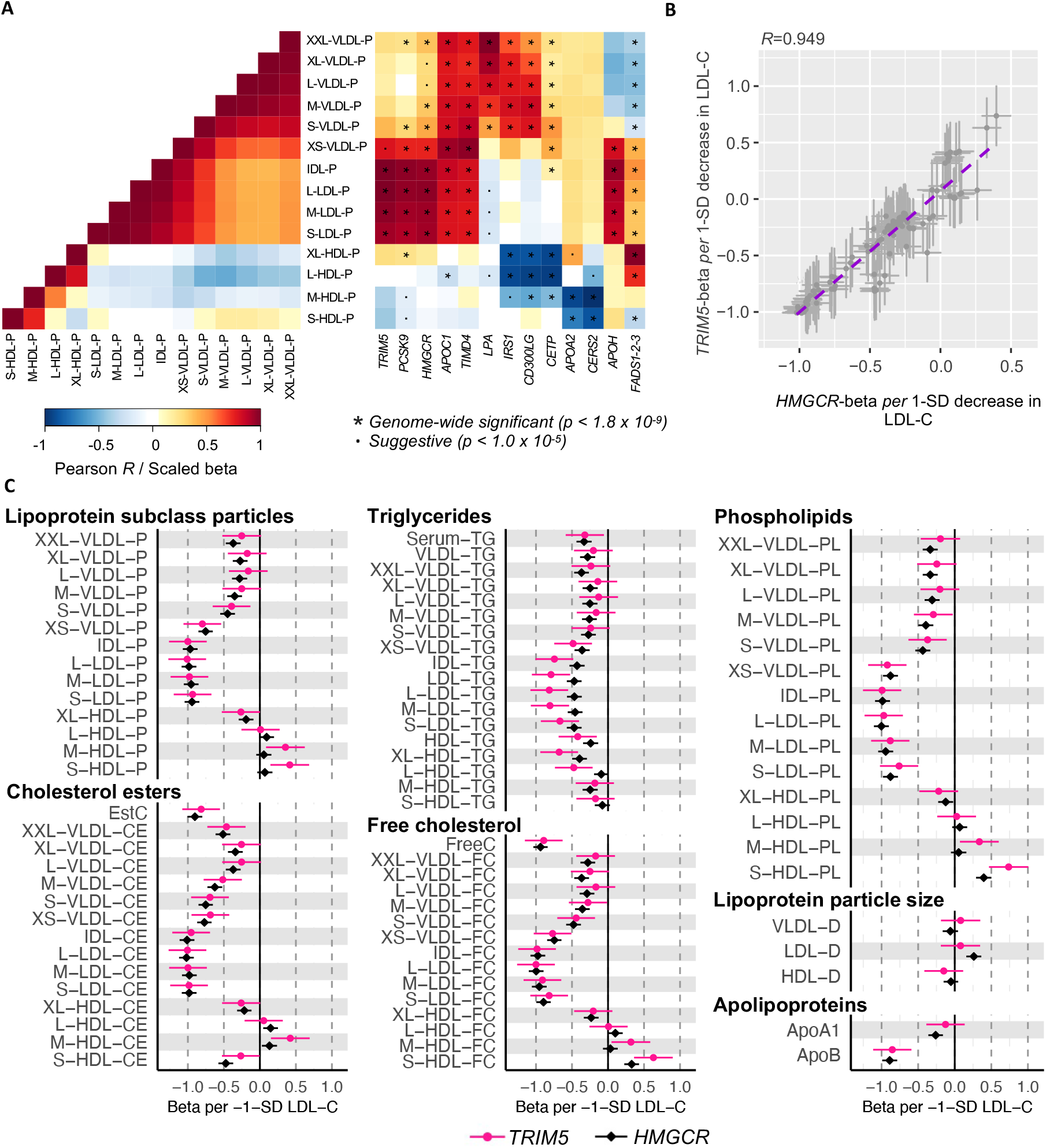
Effects of SNPs across the lipoprotein and lipid traits. (A) Heatmaps of the correlation structure of lipoprotein subclass particle concentrations (left) and the association landscapes of exemplar SNPs (right). In the heat maps, pairwise correlations of lipoprotein subclass particle concentrations (calculated in FINRISK1997; left) and effect estimates for the SNP-metabolic trait associations (right) are visualized by a color range. The SNP effect sizes were scaled relative to the absolute maximum effect size in each locus. Each column represents a single SNP, and each row corresponds to a single metabolic measure. A scatterplot (B) and forest plots (C) of the effect estimates for TRIM5 and HMGCR lead SNPs (rs11601507 and rs12916, respectively) across the lipoprotein and lipid traits. A best fit regression line is illustrated (purple dashed line) in panel B along with an estimate of Pearson’s correlation coefficient R in the title. The effect estimates (SD units) were scaled relative to a 1-SD decrease in LDL cholesterol.

Metabolic profiles of 84 novel loci that were not identified in the previous NMR GWASs^2,4,5^ were characterized here using the clustering approach (Supplementary Figures S6 and S7). As the approach we have taken uses scaled effect estimates, our results are not directly comparable to previous studies which have used unscaled effect estimates^9^ or numbers of associations *per* lipoprotein type^5^ in clustering. Even though most loci, such as the master regulator genes *PCSK9* and *LDLR*, clustered similarly as reported previously^5,9^, the new genetically calibrated approach applied here can specifically add to the understanding of the detailed metabolic effects of less well-known lipid-associated loci as their metabolic association patterns have not been previously characterized. *TRIM5* (encoding tripartite motif-containing protein 5) is an example of a poorly characterized locus associated with 42 lipoprotein and lipid traits (Supplementary Table S5). *TRIM5* is best known for its role in antiviral host defense^31^, but variants near *TRIM5* have also been associated with risk of liver fibrosis in HIV/HCV co-infected patients and altered levels of liver enzymes^32^ and were recently reported to associate with risk of coronary artery disease^33^. Interestingly, the metabolic effects on the lipoprotein and lipid traits of the lead *TRIM5* variant (rs11601507, p.Val112Ile) appear similar to those of the *HMGCR* variant rs12916 (Figures 2B and 2C), the metabolic effects of which are concordant with those of statin therapy^34–36^. The mechanism by which *TRIM5* affects lipid and lipoprotein levels and predisposes to coronary artery disease is unclear and it has been speculated to be related to innate immunity^37^. However, our data suggest that TRIM5 may be affecting the hepatic cholesterol synthesis pathway, raising the possibility that inhibition of TRIM5 could provide an alternative therapeutic pathway for reducing the risk of cardiovascular disease via lowering the concentrations of circulating atherosclerotic apoB-containing lipoprotein particles.

### Characterizing the roles of metabolic trait-associated variants in diseases

To investigate the roles of the metabolic trait-associated variants in disease, we scanned all the disease and trait associations of the 1,447 lead SNPs in the (1) FinnGen study (Data Freeze 7, up to 309,154 participants, 3,095 phenotypes), a dataset linking genomic information from Finnish participants to digital health care data^38^, and in (2) PhenoScanner, a curated database of genotype-phenotype associations from published GWAS^39,40^ (Supplementary Table S5).

Most (*n*=1,189) of the 1,447 lead SNPs had previously reported associations (*p* < 5 × 10^−8^) with traits or diseases, including directly relevant outcomes such as use of statin medication and hypercholesterolemia (Supplementary Table S5). Seven metabolic trait-associated loci (*GCKR, ABCG8, ABCB11, ABCB1, CYP7A1, SERPINA1, HNF4A*) were associated (*p* < 5 × 10^−8^) with risk of intrahepatic cholestasis of pregnancy (ICP) in FinnGen (Figure 3A, Supplementary Table S7), of which all but *ABCG8* had robust evidence of colocalization or shared regional associations with the metabolic trait associations (Supplementary Table S8). ICP is a cholestatic disorder with onset in the second or third trimester of pregnancy, characterized by pruritus and elevated concentrations of serum aminotransferases and bile acids. ICP increases the risk of meconium staining of amniotic fluid, preterm delivery, fetal bradycardia, fetal distress, and fetal loss^41^. The genetic background of ICP is poorly characterized with few published GWASs^7,42^ and the metabolic impact of the ICP-loci has not been characterized. Compared to results of a recent ICP GWAS that included data from meta-analysis of an earlier FinnGen release (Data Freeze 4) and two other cohorts^42^, associations at nine loci (*GCKR, ABCG8, ABCB11, ABCB1, CYP7A1, SERPINA1, GAPDHS*/*TMEM147, SULT2A1, HNF4A*) were replicated here and three novel loci (*UGT8, NUP153, HKDC1*) were additionally identified. A pathway analysis of the ICP-associated loci showed that biological processes related to bile acid, glucose, and lipid metabolism were enriched for ICP (Supplementary Table S9), consistent with the metabolic trait associations. For some loci (*CYP7A1, ABCB1, SERPINA1*), the most profound associations were detected for IDL and LDL particles, while two loci (*HNF4A* and *GCKR*) were more pleiotropic with effects across both apoB-containing and HDL particles (Figure 3B). At three of the loci (*CYP7A1, ABCB1, SERPINA1*) the ICP-predisposing alleles were associated with higher concentrations of IDL and LDL subclass measures, while the directions were opposite for others (*GCKR, ABCB11* and *HNF4A*). This information may be useful when considering these genes for therapeutic targets, as targets that adversely influence atherosclerotic lipids in pregnant women may be undesirable, despite the relatively short treatment period. By characterizing the associations of ICP-associated loci with metabolic traits in detail, we exemplify the value of combining the metabolic association information with disease associations to elucidate the metabolic underpinnings of poorly understood conditions.

**Figure 3.**
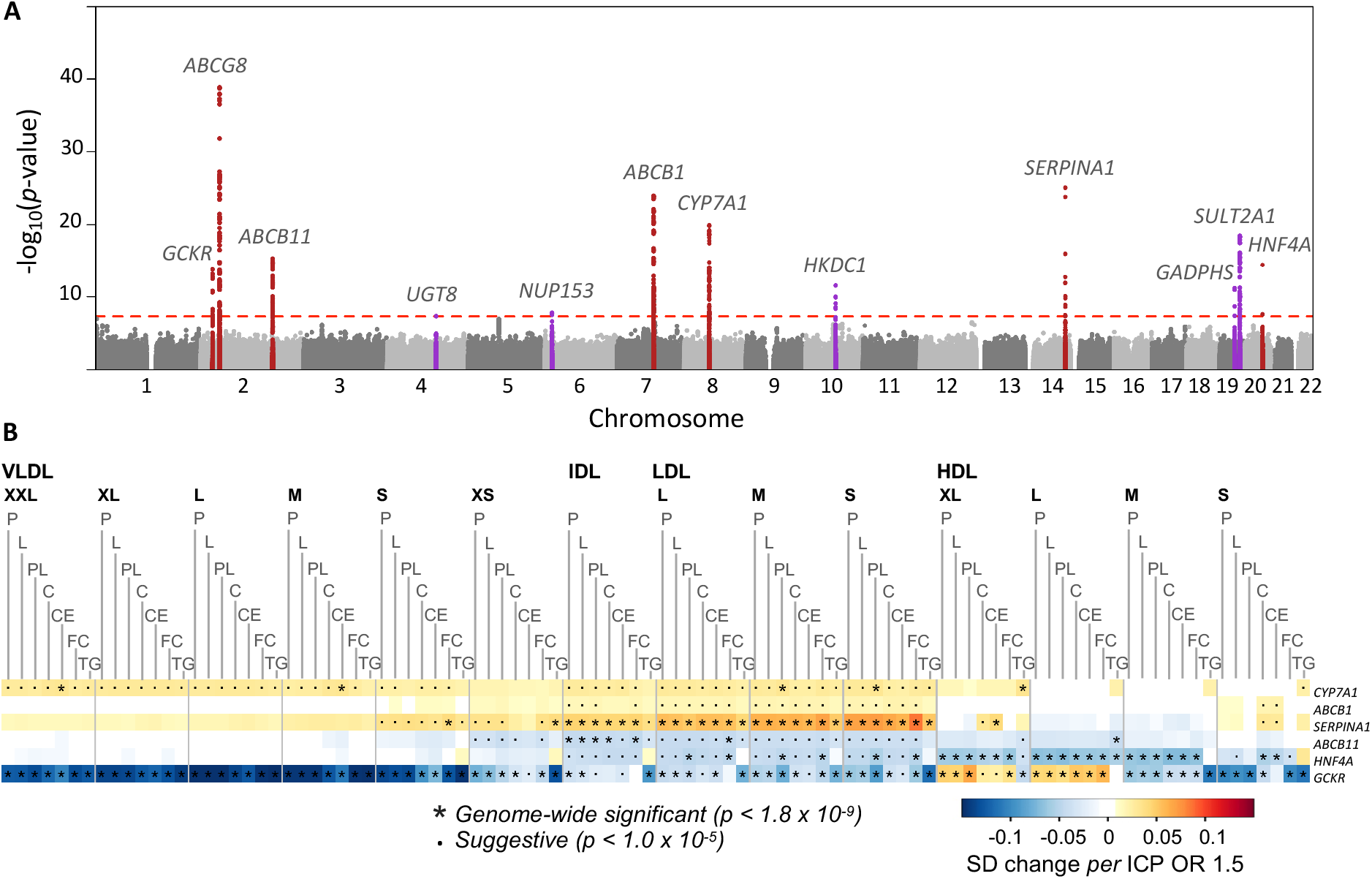
Metabolic trait associated variants are associated with intrahepatic cholestasis of pregnancy. A Manhattan plot of genome-wide association study of intrahepatic cholestasis of pregnancy (ICP) (A) and a heat map of loci associated with metabolic traits and ICP (B). Twelve loci were associated with ICP in the FinnGen study (1,460 cases, 172,286 controls). In the Manhattan plot (A), 500 Kb regions flanking the lead SNPs are highlighted, and the nearest gene is indicated for each signal. Loci that overlap with the loci identified in the NMR meta-analysis are indicated in red. In the heat map (B), loci that likely had shared causal variants with the metabolic traits were included. The heat map illustrates the resemblances of the association landscapes. Each row represents a single SNP, each column corresponds to a single metabolic measure, and the scaled effect estimates for the SNP-metabolite associations are visualized with a color range. The associations were scaled with respect to their associations with ICP (SD change per ICP OR 1.5).

### Mendelian randomization identifies a causal relationship between acetoacetate and hypertension

Finally, we exploited the absence of UK Biobank from our GWAS meta-analysis to perform a two-sample Mendelian randomization (MR) analysis to investigate associations of genetically predicted levels of the twenty non-lipid traits with 460 Phecodes and 52 quantitative traits from the UK Biobank. Initial MR analyses using all lead variants for each trait as genetic instruments identified 503 significant associations (*p* < 4.88 × 10^−6^) under the inverse variance weighted model, including positive associations between glucose and diabetes, creatinine and renal failure, and amino acids with diabetes (Supplementary Table S10), all of which represent well-known causal relationships. Restricting the analyses to less pleiotropic variants (associated with fewer than five NMR traits), the association estimates were on average considerably weaker with less between-variant heterogeneity (median absolute beta=0.058 *vs*. 0.152; Q-statistic 34.2 *vs*. 385.6, Supplementary Figure S8), suggesting that pleiotropy was driving many of the initial MR associations. This clearly emphasizes that pleiotropy should be carefully considered when selecting instrument SNPs for MR to avoid false interpretations about potential causal relationships.

As an example, the MR results for acetoacetate were substantially affected by the inclusion of more pleiotropic SNPs in the instrument (Figure 4). Acetoacetate is a ketone body that is produced primarily in the liver during fasting and which has been associated with several cardiometabolic conditions including heart failure^43^ and diabetes^44^ in biochemical and epidemiological studies. In the GWAS, we identified associations for acetoacetate at ten loci (only one associated locus, *APOA5*, was identified in the previous NMR GWAS meta-analysis^4^), and MR yielded twenty robust associations (Figure 4A). These included associations with triglycerides, HDL-cholesterol and remnant-cholesterol, likely reflecting the inclusion of well-known lipid loci (*LPL, APOA5, TRIB1, APOC1, GALNT2, PPP1R3B*) in the instrument. The less pleiotropic instrument for acetoacetate included only four loci: *HMGCS2* (3-hydroxy-3-methylglutaryl-CoA synthase 2), *OXTC1* (3-oxoacid CoA-transferase 1), *CYP2E1* (cytochrome P450 family 2 subfamily E member 1) and *SLC2A4* (solute carrier family 2 member 4), all of which have direct roles in ketone body or glycemic-related pathways. Using these four variants only the positive association with hypertension (OR *per* 1-SD higher genetically predicted acetoacetate level = 1.41, *p* = 6.9 × 10^−7^) was robust (Figures 4A and 4B) and was also replicated in FinnGen (OR 1.45, *p* = 4.5 × 10^−5^) (Figure 4C). Consistent with these results, acetoacetate has recently been suggested as a biomarker for hypertension^45^. The discovery regarding this potential causal relationship between acetoacetate and hypertension is noteworthy since the data on the role of ketogenic diets in hypertension are suggestive but inconclusive^46^ and ketone bodies have also emerged as potential therapeutics for coronary disease^47^. A recent study in the UK Biobank demonstrated that some loci and pathways associated with the non-lipid NMR traits are highly pleiotropic, with the less pleiotropic variants often reflecting biology more proximal to the traits^48^. This is also in line with our findings as demonstrated by the identification of several pleiotropic triglyceride-related genes that are associated with acetoacetate levels, as well as four less pleiotropic acetoacetate-associated loci with direct links to pathways related to ketone biology. These results accentuate that genetic pleiotropy can be common for metabolic measures, even for some non-lipid traits, and that careful selection of variants for MR is crucial to avoid bias due to pervasive pleiotropy.

**Figure 4.**
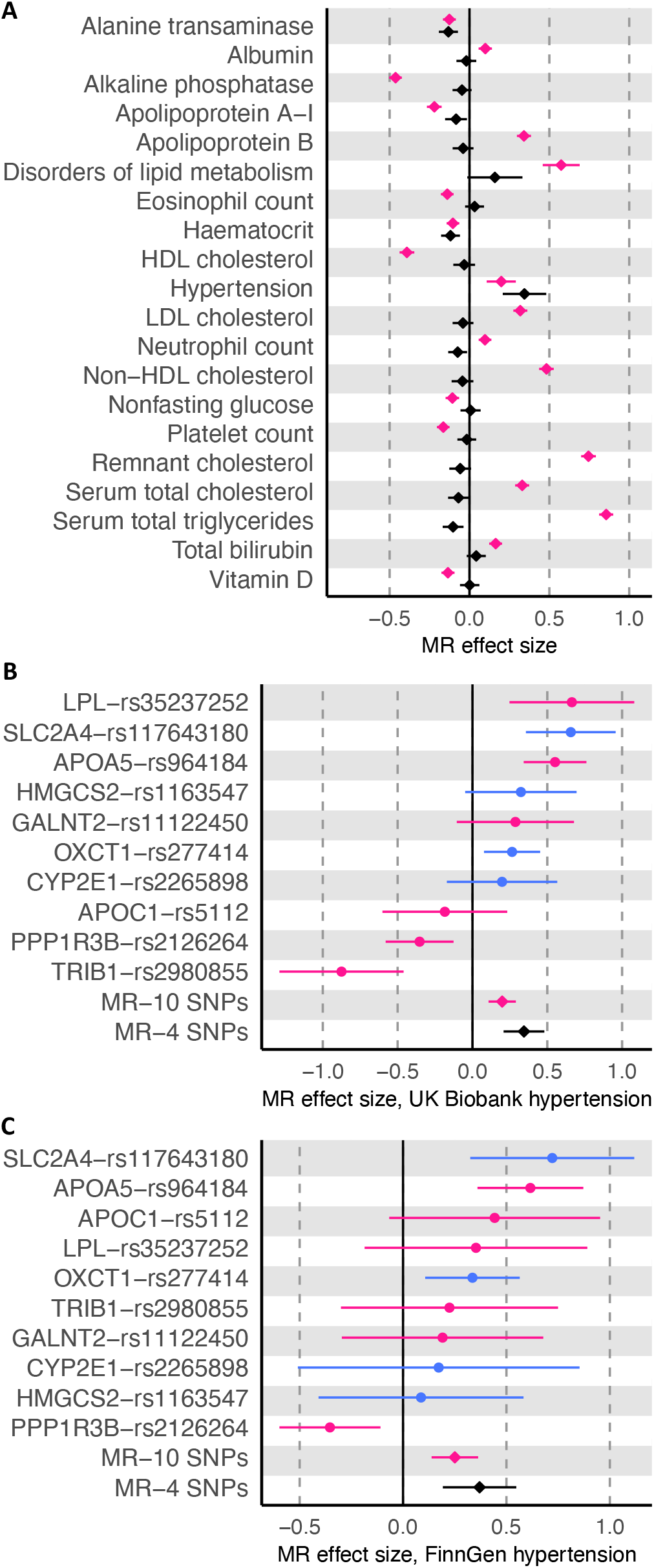
Mendelian randomization suggests a causal association between acetoacetate and hypertension. In panel A, effect estimates (betas per 1-SD increase in acetoacetate) are shown for the UK Biobank outcomes that were significant (p < 4.88 × 10^−6^) with the full (pleiotropic, n = 10, pink) or strict (non-pleiotropic, n = 4, black) set of instruments. Panels B and C show the effect estimates in Mendelian randomization (MR) analysis with hypertension in the UK Biobank (panel B) and FinnGen (panel C) as the outcomes. Single-SNP MR effect estimates and 95% confidence intervals are shown, with the SNPs in the strict instrument colored blue and the other SNPs colored pink. Mendelian randomization effect estimates are shown with pink and black diamonds for the full instrument (all ten SNPs) and strict instrument (four non-pleiotropic SNPs).

## CONCLUSION

Through this large-scale, genome-wide meta-analysis including more than 136,000 participants, we discovered over 8,000 genetic associations of circulating metabolic biomarkers involving over 400 loci. The five-fold increase in sample size and doubling of the number of metabolic traits compared to our previous GWAS meta-analysis of NMR metabolic traits led to a dramatic increase in the number of significant associations (62 associated loci previously^4^), leading to a substantial improvement in understanding of genetic regulation of systemic metabolism. Key features of our meta-analysis are the inclusion of participants from 33 cohorts, enabling the discovery of many new robust associations with evidence from independent datasets. Through internal comparisons across these datasets and external comparison with UK Biobank, we have highlighted the important role that sample and participant characteristics, such as sample type and fasting status, can play in revealing or masking genetic associations, with significant consequences for biological interpretation and downstream analyses. Our extensive manual curation to identify highly likely causal genes at nearly 300 associated loci provides a useful resource to further biological understanding of the associations and allows high-confidence identification of causal genes for disease associations that colocalize. For the remaining loci, our results provide a starting point for identification of genes that have not been known to be involved in metabolic regulation thus far. Our comparison of the fine-grained metabolic associations across the lipoprotein measures allows for the identification of clusters of genes with similar metabolic profiles, suggesting TRIM5 as a potential therapeutic target for lowering pro-atherogenic lipid levels, and therefore cardiovascular diseases, due to its similarity to HMGCR, the target for statins. By making the summary statistics publicly available, we provide a valuable resource for Mendelian randomization studies and have illustrated the potential pitfalls of using pleiotropic variants as genetic instrumental variables. Finally, we have illustrated the potential to use these findings to shed light on inadequately characterized diseases by examining the metabolic effects of genetic variants associated with intrahepatic cholestasis of pregnancy, a disease with a largely unknown genetic background.

## METHODS

### NMR metabolomics

In this work, we expand our previous genome-wide association study of 123 human metabolic traits in ∼25,000 individuals^4^ to include additional cohorts and a more comprehensive panel of metabolic traits. Up to 233 serum metabolic traits were quantified in 33 cohorts (total sample size up to 136,016) using an updated quantification version of the same NMR metabolomics platform^17^ as in the previous study. The NMR metabolomics platform provides data of lipoprotein subclasses and their lipid concentrations and compositions, apolipoprotein A1 (apo-AI) and apoB, cholesterol and triglyceride measures, albumin, various fatty acids and low-molecular-weight metabolites, e.g., amino acids, glycolysis-related measures and ketone bodies. In this work, the metabolic traits were quantified in the following cohorts (described in detail in Supplementary Notes and Supplementary Table S1): Avon Longitudinal Study of Parents and Children (ALSPAC), China Kadoorie Biobank (CKB), Estonian Genome Center of University of Tartu Cohort (EGCUT), The Erasmus Rucphen Family study (ERF), European Genetic Database (EUGENDA), FINRISK 1997 (FR97), FINRISK 2007 (FR07, i.e. DILGOM), The INTERVAL Bioresource (INTERVAL), CROATIA-Korcula Study (KORCULA), LifeLines-DEEP (LLD), Leiden Longevity Study (LLS), eight subcohorts from the London Life Sciences Prospective Population Study (LOLIPOP), The Metabolic Syndrome in Men study (METSIM), The Netherlands Epidemiology of Obesity Study (NEO), The Netherlands Study of Depression and Anxiety (NESDA), Northern Finland Birth Cohort 1966 (NFBC1966), NFBC1986, The Netherlands Twin Register (NTR), Oxford Biobank (OBB), Orkney Complex Disease Study (ORCADES), PROspective Study of Pravastatin in the Elderly at Risk (PROSPER), three subcohorts from the Rotterdam Study (RS), TwinsUK (TUK), and The Cardiovascular Risk in Young Finns Study (YFS). Most of the cohorts consisted of individuals of European origin (six Finnish and 21 non-Finnish), and six cohorts had individuals of Asian origin (one Han Chinese and five South Asian). All participants gave informed consent and all studies were approved by the ethical committees of the participating centers.

### Genome-wide association study

A GWAS was performed for 233 metabolic traits (Supplementary Table S2) in each of 33 cohorts (Supplementary Table S1), leading to inclusion of up to 136,016 individuals with both NMR metabolic trait measurements and genome-wide SNP data available. Pregnant individuals or those under lipid-lowering medication were excluded from the study. SNPs were imputed using the Haplotype Reference Consortium release 1.1 or the 1000 Genomes Project Phase 3 release, and GWAS was performed under the additive model separately in each cohort (details in Supplementary Table S3). Before analyses, the metabolic trait distributions were adjusted for age, sex, principal components and relevant study-specific covariates (See Supplementary Table S3), and inverse rank normal transformation of trait residuals was performed. The cohorts were combined in fixed-effect meta-analysis with METAL^49^, and the SNPs were filtered to those present in at least seven cohorts. The NMR metabolic traits are highly correlated and therefore using the Bonferroni correction to account for multiple testing would result in an overconservative threshold for genome-wide significance. We therefore used the number of PCs (28) explaining >95% variation in the metabolic traits defined in the largest cohort, INTERVAL, to correct for multiple-testing, and our genome-wide significance threshold was set to *p* < 1.8 × 10^−9^ (standard genome-wide significance level, *p* < 5 × 10^−8^, divided by 28). After the primary GWAS, a fasting-stratified analysis was performed; in this analysis 27 of the cohorts were classified as fasted (total *n*=68,559) and five cohorts were classified as non-fasted (total *n*=58,112; see Supplementary Table S1). To define associated loci across the metabolic traits, we defined a 500-kb window flanking each SNP meeting the significance threshold, pooled together these windows from all metabolic traits for each chromosome, and iteratively merged the windows. As this approach can lead to inclusion of multiple independent signals within these loci, we further defined potential independent signals that reside within the defined loci based on pairwise linkage disequilibrium (LD; *r*^*2*^ cut-off of 0.3, defined in INTERVAL and FINRISK97) of all the lead SNPs within each locus. We assigned the associated lead SNPs to most likely causal genes based on two criteria: 1) we prioritized genes with clear biological relevance to the associated metabolic traits; and 2) if no biologically plausible causal gene was detected and the lead SNP was a functional variant (missense, splice region or stop gained) or in high LD (*r*^*2*^>0.8 in INTERVAL) with such variant, the gene with the functional variant was assigned as the most likely candidate gene. If criteria 1 and 2 were not fulfilled, the nearest gene was indicated as the candidate gene.

### Replication using publicly available resources

UK Biobank SNP – metabolic trait summary statistics were downloaded (https://gwas.mrcieu.ac.uk/datasets/?gwas_id_icontains=met-d) from the IEU Open GWAS Project^50^. These summary statistics were derived from the publicly available March 2021 release of the UK Biobank data in which the metabolic traits were measured with a similar NMR technology (newer version of the Nightingale Health platform) as in our study. The data was used to compare the association of our lead SNP – metabolic trait pairs within the 276 associated regions. Two thresholds were used to define an association in the UK Biobank data: the standard genome-wide significance level (*p* < 5 × 10^−8^) and the suggestive level of significance (*p* <1 × 10^−5^).

### Comparing to previous associations

We performed an extensive comparison of our metabolic trait associations to previous genome-wide association studies of metabolic traits. Our comparisons were divided into three groups: 1) comparison to results of previously published large GWAS of circulating NMR traits^4,5^; 2) comparison with loci associated with clinical lipids (including those from the UK Biobank September 2019 version 3 release)^20,24,25^; and 3) comparison with an extensive list of associations from previous metabolite and metabolomic studies^11,13,51–64^. The comparisons were performed by indicating: 1) co-located known variants; 2) any known associations within a 500 kb flank of a lead SNP; or 3) known associations in LD (*r*^*2*^>0.3, defined in INTERVAL) with a lead SNP. Since our comparisons included studies with available summary statistics, comparing our associations to those from a recent study on sixteen non-lipid NMR traits^48^ was not possible.

In addition to comparing to previous metabolic trait associations, we screened previous disease and trait associations (*p* value cut-off 5 × 10^−8^) of the lead SNPs using PhenoScanner, v2^39,40^. In addition, we screened the FinnGen^38^ Data Freeze 7 summary statistics of 3,095 disease endpoints for overlapping associations (*p* value cut-off 5 × 10^−8^).

### Characterizing metabolic effects of lipoprotein and lipid associated loci

To compare the metabolic effects of lipoprotein lipid and apolipoprotein associated variants, the effect estimates were visualized as color-coded heat maps. To allow comparison of SNP effects, the estimates were scaled relative to the highest absolute value of the estimate for each SNP. In this analysis, we included lead SNPs at the 276 initially defined regions that were associated with any of the lipoprotein lipids or apolipoproteins at genome-wide significance and nominally associated (*p* < 0.05) with apolipoprotein B. We used these criteria to restrict the analysis to SNPs associated with apolipoprotein B, because apolipoprotein B is known to be a causal part of lipoprotein metabolism for cardiovascular disease ^28–30^. To exclude signals with similar effects across the metabolic traits due to the same causal gene, we included only a single SNP from the initially defined genomic regions that had multiple independent signals if the patterns of metabolic traits associations were similar (*R*>0.5). In the heat maps each line represents a single SNP, each column corresponds to a single metabolic measure, and the scaled effect estimates for the SNP-metabolite associations are visualized with a color range. Directions of effects are shown in relation to the allele associated with increased apolipoprotein B. To group SNPs with similar effects together, dendrograms were constructed based on hierarchical clustering of the scaled SNP effects. Heat maps were constructed using the heatmap.2 function of the gplots v. 3.0.3 R package.

### Characterizing metabolic associations of intrahepatic cholestasis of pregnancy

We assessed overlap of our metabolic trait associations with intrahepatic cholestasis of pregnancy (ICP) using summary statistics from the FinnGen study ^38^ Data Freeze 7 (O15_ICP; 1,460 cases 172,286 controls). ICP cases were defined through hospital discharge registry, ICD10 code O26.6 and ICD9 codes 6467A and 6467X. Using the candidate gene assignments of each associated locus, we performed gene ontology (GO) enrichment analysis to search for enriched biological process and molecular function GO terms^65,66^. We assessed colocalizations of association signals using Hypothesis Prioritisation for multi-trait Colocalization (HyPrColoc) R library in which an efficient deterministic Bayesian algorithm is used to detect colocalization across vast numbers of traits simultaneously^67^. We searched for colocalization at single causal variants and shared regional associations. To visualize SNP effects across lipid and lipoprotein traits, heat maps were constructed using the heatmap.2 function of the gplots v. 3.0.3 R package. The following SNPs were included in the heatmaps: *GCKR*-rs1260326, *ABCB11*-rs10184673, *ABCB1*-rs17209837, *CYP7A1*-rs9297994, *SERPINA1*-rs28929474 and *HNF4A*-rs1800961. Effects of the metabolic trait-associated SNPs were scaled relative to an odds ratio of 1.5 for ICP.

### Mendelian randomization

Two-sample Mendelian randomization was performed using twenty NMR non-lipid metabolic traits [including amino acids (alanine, glutamine, glycine, histidine, isoleucine, leucine, valine, phenylalanine, tyrosine), ketone bodies (acetate, acetotoacetate, 3-hydroxybutyrate), and glycolysis/gluconeogenesis (glucose, lactate, pyruvate, glycerol, citrate), fluid balance (albumine, creatinine) or inflammation-related (glycoprotein acetylation) metabolic traits] as exposures and 460 Phecodes and 52 quantitative traits from the UK Biobank^20^ as outcomes. We defined two sets of instruments for the analyses that are referred to as full and strict instruments. As initial instruments we used the 334 lead variants (a single instrument SNP *per* each defined associated locus) associated with these traits (‘full instruments’). To avoid potential pleiotropy, we also selected a subset of 193 variants (‘strict instruments’) that had fewer than 5 associations across all 233 metabolic traits. We defined disease outcomes in UK Biobank using a curated list of major Phecodes available in the PheWAS R package^68,69^. To restrict our analysis to major disease outcomes, we discarded any sub-categories (i.e., Phecodes with four or more characters) and removed outcomes with fewer than 100 events across up to 367,542 unrelated European-ancestry UK Biobank participants. The resulting 460 diseases were grouped into 15 broad domains: circulatory system, dermatologic, digestive, endocrine/metabolic, genitourinary, haematopoietic, infectious diseases, mental disorders, musculoskeletal, neoplasms, neurological, pregnancy complications, respiratory, sense organs, symptoms. We also analyzed 52 quantitative traits available in UK Biobank, including blood pressure, lung function measures, blood cell traits and clinical chemistry biomarkers. In our replication analysis (acetoacetate as the exposure and hypertension as the outcome), we used essential hypertension from the FinnGen study^38^ Data Freeze 7 as the outcome (Hypertension essential, I9_HYPTENSESS; 70,651 cases, 223,663 controls). Cases were defined through hospital discharge registry, ICD10 code I10, ICD9 codes 4019X and 4039A, ICD8 codes 40199, 40299, 40399, 40499, 40209, 40100, 40291, 40191 and 40290.

We performed univariable Mendelian randomization using the inverse-variance weighted method for each instrument^70^. We also performed sensitivity analyses using MR-Egger regression to account for unmeasured pleiotropy^71^ and weighted median regression to assess robustness to invalid genetic instruments^72^. Our primary analyses were based on fixed-effect models, but as sensitivity analyses we used random-effect models to account for between-variant heterogeneity, which we quantified using the I-squared statistic. The MR analyses were performed using the MendelianRandomization package v. 0.5.1^73^ or the TwoSampleMR package v. 0.5.3^74^. Single-SNP MR estimates were based on the Wald ratio. We considered the fixed-effects inverse-variance weighted method as the main MR model but report the results of all models in Supplementary Table S10. To account for multiple-testing, associations with *p* < 4.88 × 10^−6^ were considered significant (Bonferroni correction to account for testing of 20 metabolic traits with 512 outcomes).

### FinnGen study

In the present study, we used GWAS summary statistics of 3,095 disease endpoints from FinnGen Data Freeze 7. Full description of the FinnGen study^38^ and data analysis steps is provided in Supplementary Notes.

## Data Availability

All data produced in the present study are available upon reasonable request to the authors. The data will be made publicly available at the time of final publication.

## DATA AVAILABILITY

Full summary statistics of this study will be made publicly available upon publication.

## ACKNOWLEDGEMENTS

Please see Supplementary Notes for acknowledgements and funding.

## SUPPLEMENTARY MATERIALS

**Supplementary Notes**. This file contains study descriptions, acknowledgements, and funding information.

**Supplementary Figures**. Supplementary Figures are included in four files: Figures S1-S3 in separate files, Figures S4-S8 in a combined file. Figure contents are described below.

**Fig. S1**. Manhattan plots showing the meta-analysis results of 233 metabolic traits.

**Fig. S2**. Regional associations plots for the most significantly associated metabolic traits in each genomic region.

**Fig. S3**. Forest plots showing the associations of the lead SNPs in each cohort.

**Fig. S4**. Mirrored manhattan plot showing the results of genome-wide association study of phenylalanine in the NMR meta-analysis and UK Biobank.

**Fig. S5**. Examples of glucose associations for fasted and non-fasted cohorts.

**Fig. S6**. Heat map of lipoprotein and lipid associations.

**Fig. S7**. A zoomed heat map of lipoprotein and lipid associations.

**Fig. S8**. Influence of pleiotropy on Mendelian randomization estimates.

**Supplementary Tables**. This file contains Supplementary Tables S1-S11 (described below).

**Table S1**. Description of studies.

**Table S2**. List of NMR metabolic traits and their abbreviations.

**Table S3**. Details of genotyping and GWAS analyses.

**Table S4**. Genomic regions associated with metabolic traits. The genomic regions associated with the 233 NMR metabolic traits are indicated, along with candidate gene assignments and GWAS results for the lead SNP of the most significant metabolic trait within each region. Genomic positions refer to hg19. Effects are given for allele 1.

**Table S5**. Significant lead SNP – metabolic trait associations. GWAS results for all significant lead SNP – metabolic trait associations within the defined associated genomic regions are shown, along with comparisons to previous GWASs. Genomic positions refer to hg19. Effects are given for allele 1.

**Table S6**. Associations of lead SNPs with NMR metabolic traits in UK Biobank.

**Table S7**. Lead SNPs at loci associated with intrahepatic cholestasis of pregnancy in FinnGen Data Freeze 7.

**Table S8**. HyprColoc analysis for shared regional associations and colocalization with intrahepatic cholestasis of pregnancy in FinnGen R7.

**Table S9**. Pathway analysis of intrahepatic cholestasis of pregnancy.

**Table S10**. Mendelian randomization results. The results of Mendelian randomization (MR) analyses of twenty non-lipid traits with 460 Phecodes and 52 quantitative traits from the UK Biobank are shown. The MR estimates are shown separately for analyses performed with full and strict (non-pleiotropic) instruments.

**Table S11**. List of FinnGen contributors.

