## Supplementary Figure Legends and Supplementary Figures S4-S8 for "Genome-wide characterization of circulating metabolic biomarkers reveals substantial pleiotropy and novel disease pathways"

### Supplementary figures for the manuscript entitled *Genome-wide characterization of circulating metabolic biomarkers reveals substantial pleiotropy and novel disease pathways*

*Supplementary Figures are included in four files: Figures S1-S3 in separate files; Figures S4-S8 and legends of all figures in this combined file. Figure contents are listed below.*


**Figure S1.** Manhattan plots showing the meta-analysis results of 233 metabolic traits.

**Figure S2.** Regional associations plots for the most significantly associated metabolic traits in each genomic region.

**Figure S3.** Forest plots showing the associations of the lead SNPs in each cohort.

**Figure S4.** Mirrored Manhattan plot showing the results of genome-wide association study of phenylalanine in the NMR meta-analysis and UK Biobank.

**Figure S5.** Examples of glucose associations for fasted and non-fasted cohorts.

**Figure S6.** Heat map of lipoprotein and lipid associations.

**Figure S7.** A zoomed heat map of lipoprotein and lipid associations.

**Figure S8.** Influence of pleiotropy on Mendelian randomization estimates.

***Figure S1. Manhattan plots showing the NMR GWAS meta-analysis results of 233 metabolic traits.*** *The red line indicates the threshold for genome-wide significance ($p < 1.8 \times 10^{-9}$). -$\log_{10}$(p-values) were capped at 300. Metabolic trait abbreviations can be found in Supplementary Table S2.*

*See separate file (SupplFigureS1.pdf)*

***Figure S2. Regional associations plots for the most significantly associated metabolic traits in each genomic region.*** *Locus numbers (corresponding to those shown in Table S4) and lead metabolic traits in each region are indicated above the plots. 500-kb flanking regions around each lead SNP are shown. The linkage disequilibrium values ($r^2$) are based on the 1000Genomes European population.*

*See separate file (SupplFigureS2.pdf)*

***Figure S3. Forest plots showing the associations of the lead SNPs in each cohort.*** *Effect estimates are shown for the most significantly associated metabolic trait in each genomic region. Cohort acronyms can be found in Supplementary Table S1.*

*See separate file (SupplFigureS3.pdf)*

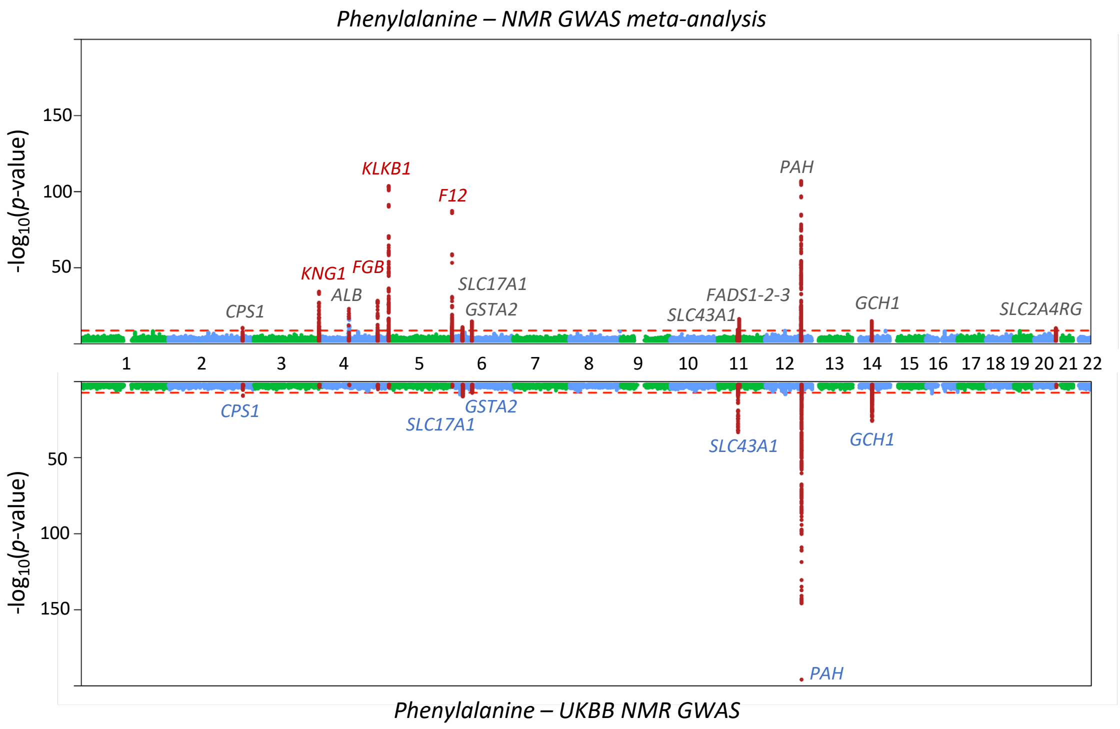


***Figure S4. Mirrored Manhattan plot showing the results of genome-wide association study of phenylalanine in the NMR GWAS meta-analysis and UK Biobank.*** *The top panel of the mirrored Manhattan plot shows the NMR GWAS meta-analysis results (n=136,016) and the bottom panel the UKBB results (n=115,025). The red lines indicate the thresholds for genome-wide significance (top panel* $p < 1.8 \times 10^{-9}$*; bottom panel* $p < 5 \times 10^{-8}$*). 500-kb regions around lead SNPs in the NMR GWAS are highlighted. Loci indicated in red have roles in coagulation-related pathways. Loci indicated in blue were genome-wide significant in both NMR GWAS meta-analysis and UK Biobank.*

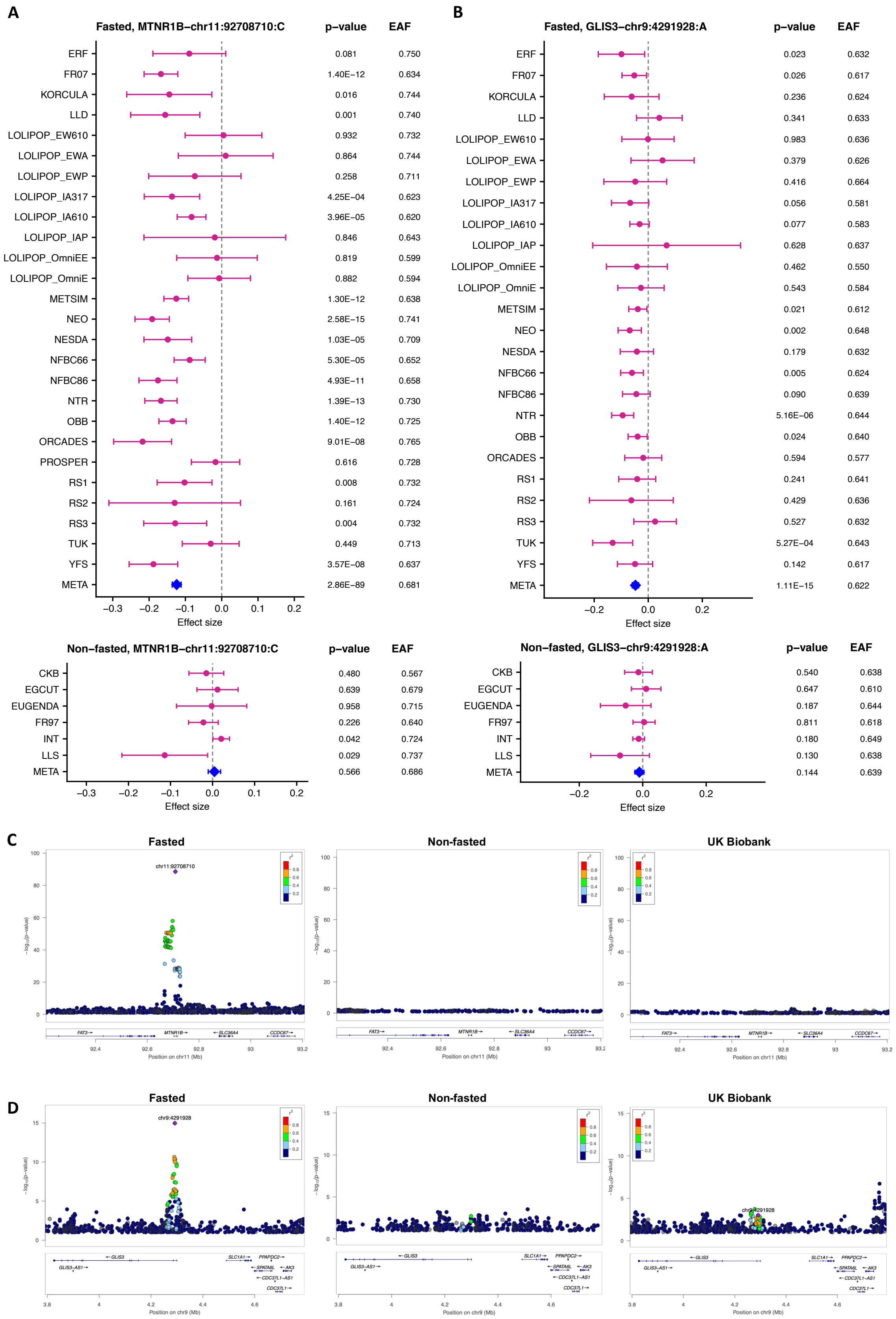


***Figure S5. Examples of glucose associations for fasted and non-fasted cohorts.*** *The forest plots in panels A and B show examples of two lead SNPs in which glucose associations were significant in the fasted cohorts (top) and non-significant in the non-fasted cohorts (bottom).*

*These associations were absent in the UK Biobank. Effect allele frequencies (EAF) and p-values are indicated for each cohort. Cohort acronyms can be found in Supplementary Table S1. Panels C and D show regional association plots of the MTNR1B (C) and GLIS3 (D) loci in the fasted (left) and non-fasted (center) cohorts and in UK Biobank (right). SNPs with p>0.1 are shown. 500-kb flanking regions around each lead SNP are shown. The linkage disequilibrium values ($r^2$) are based on the 1000Genomes European population.*

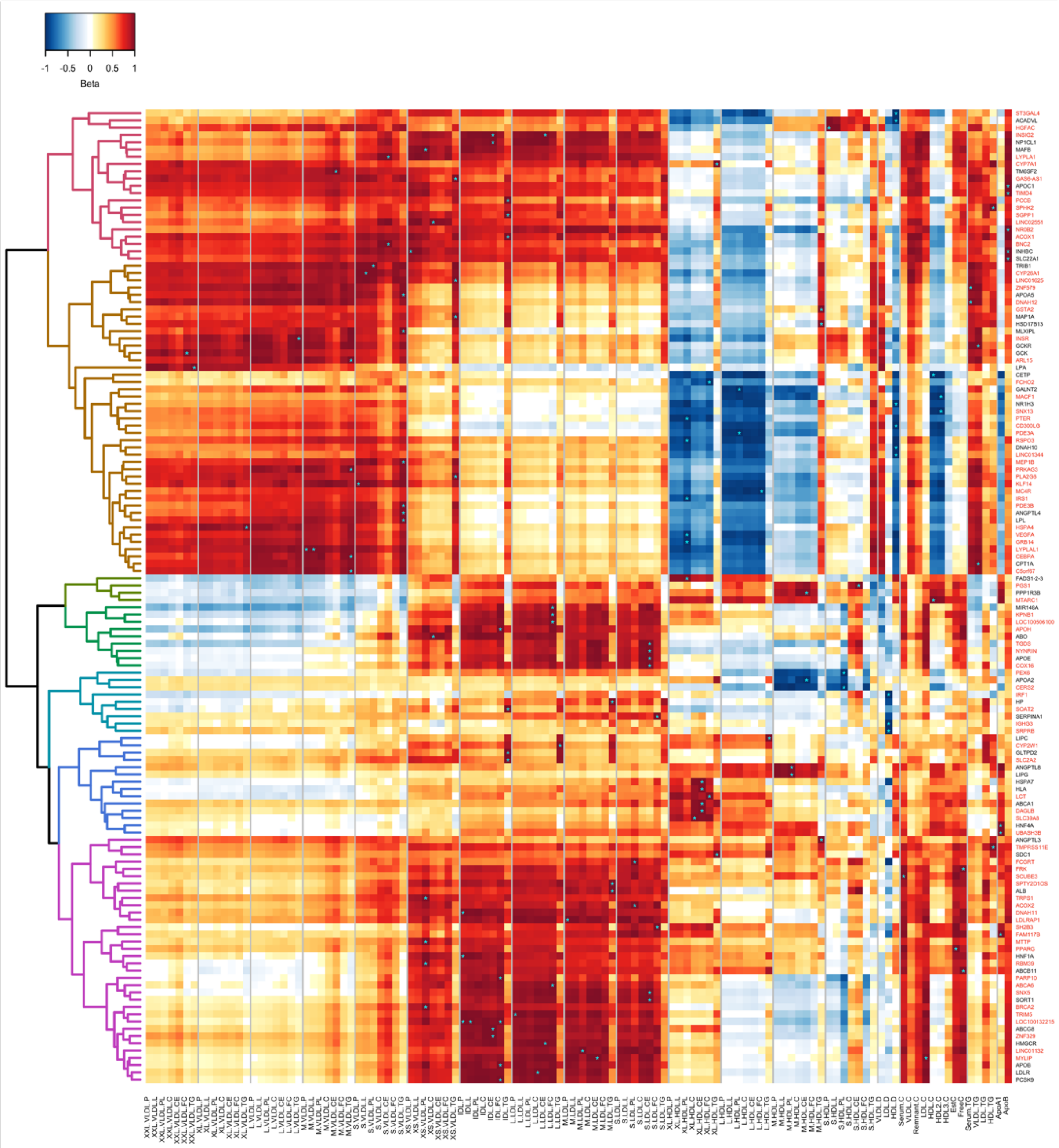


***Figure S6. Heat map of lipoprotein and lipid associations.*** *Lipoprotein and lipid associated loci with similar association patterns across the lipoprotein measures were grouped together in a dendrogram based on hierarchical clustering of the SNP effects (see Methods). The apolipoprotein B associated loci (n=134, p < 0.05) were included since apolipoprotein B represents a causal part of the lipoprotein metabolism for cardiovascular disease. The heat map illustrates the resemblances of the association landscapes; each row represents a single SNP, each column corresponds to a single metabolic measure, and the scaled effect estimates (see Methods) for the SNP-metabolite associations are visualized with a color range. Loci that were not identified in the previous large-scale NMR metabolomics GWAS are indicated by red. Traits with absolute maximum effects in each locus are indicated by asterisks. For clarity, two of the clusters (brown and purple) are highlighted in Supplementary Figure S7.*

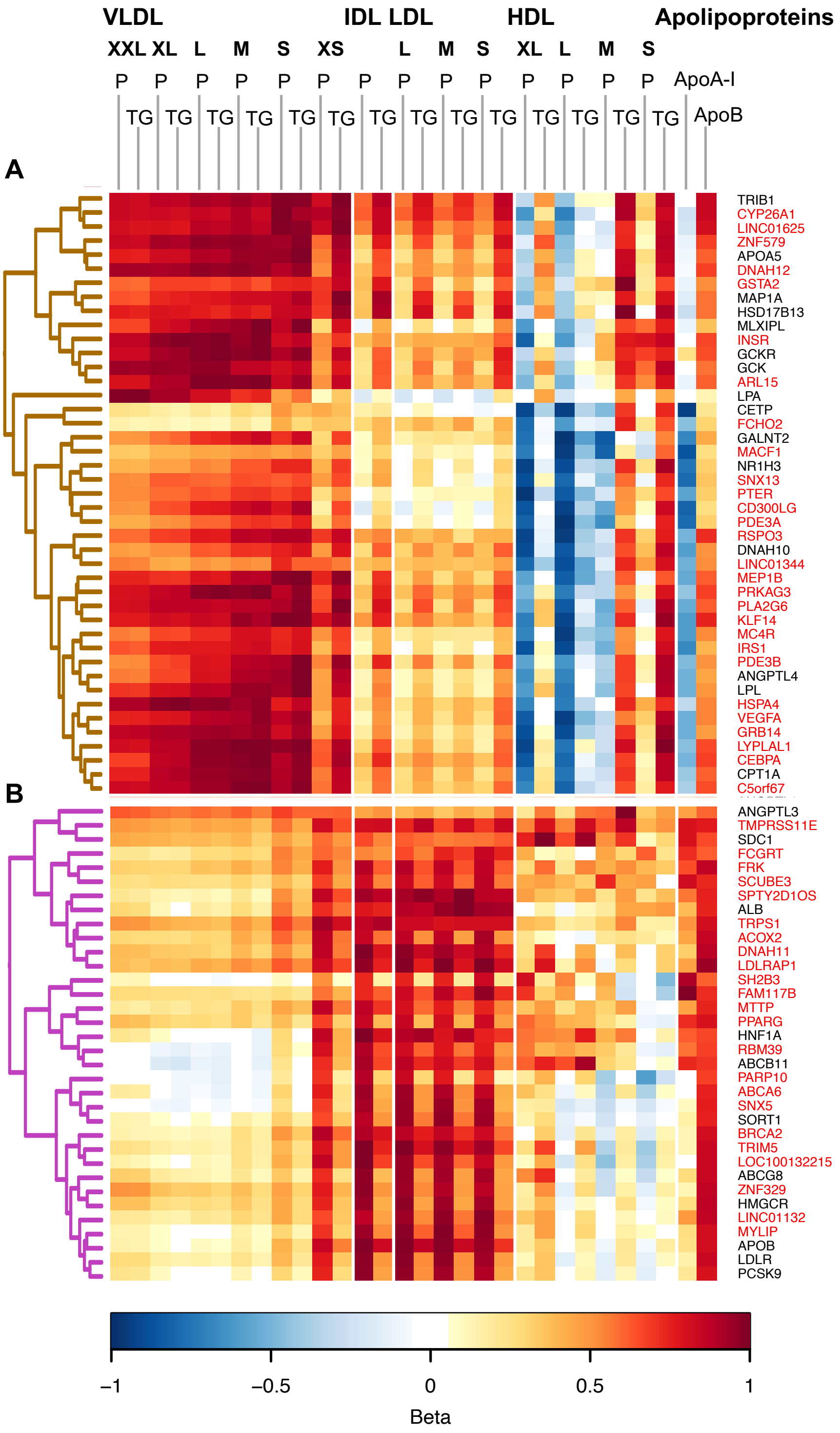


***Figure S7. A zoomed heat map of lipoprotein and lipid associations.*** *The full heat map including all the loci and a full set of lipoprotein traits is shown in Supplementary Figure S6. For clarity, two of the clusters are highlighted here. For details, see legend for Supplementary Figure S6. Panels A and B corresponding to the brown and purple branches of the dendrogram shown in the full-sized heat map, respectively. Effect sizes were scaled relative to the absolute maximum effect size (beta) in each locus. In the heat map, each row represents a single SNP, each column corresponds to a single metabolic measure, and the effect estimates for the SNP-metabolite associations are visualized with a color range. Loci highlighted in red were not identified in the previous NMR metabolomics GWAS.*

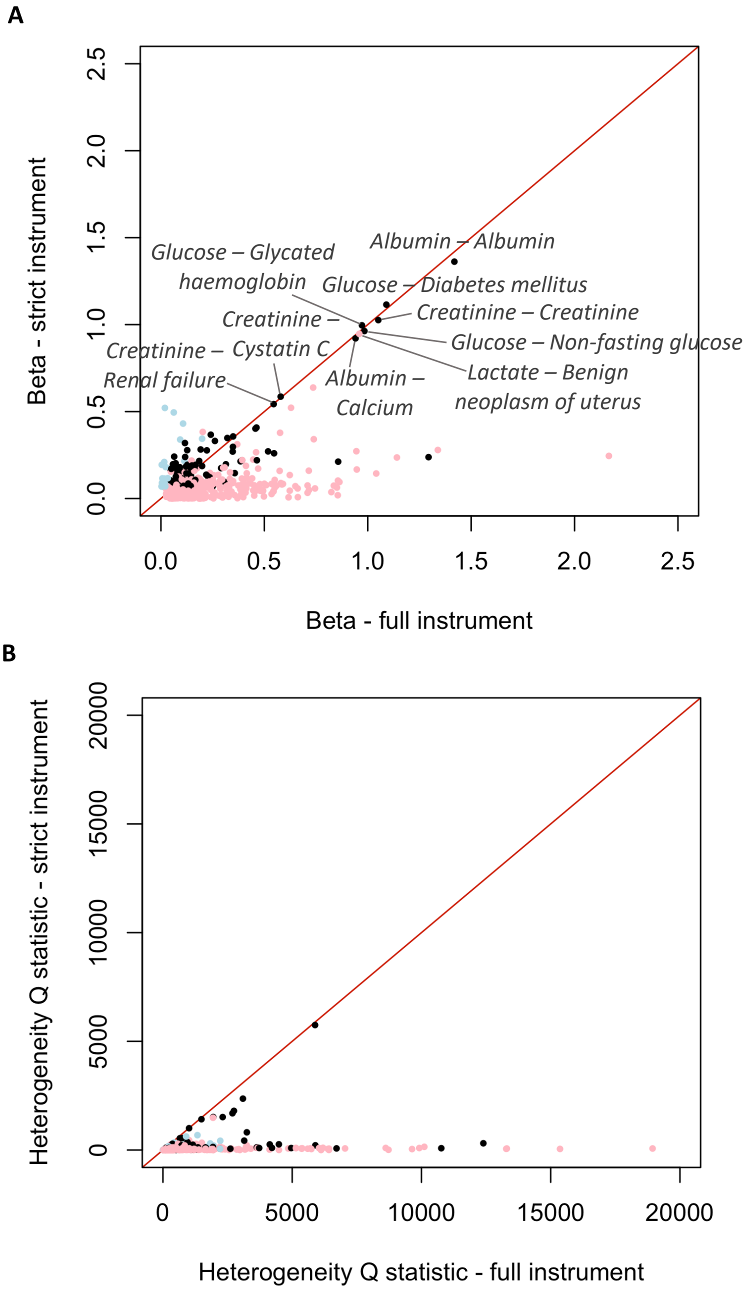


***Figure S8. Influence of pleiotropy on Mendelian randomization estimates.*** *The effect estimates (absolute betas) (panel A) and heterogeneity Q statistics (panel B) from the Mendelian randomization (MR) analyses using the full (pleiotropic) and strict (non-pleiotropic) MR instruments are shown. Associations that were significant ($p < 4.88 \times 10^{-6}$) using either the full or strict instrument or both were included; some of the significant exposure-outcome associations are indicated. Estimates indicated in light pink and light blue were not significant with the strict and full instruments, respectively. For clarity, very large beta (>5) and Q values (>60,000) were excluded from the plots.*
