## Supplementary Figure S1 for "Genome-wide characterization of circulating metabolic biomarkers reveals substantial pleiotropy and novel disease pathways"

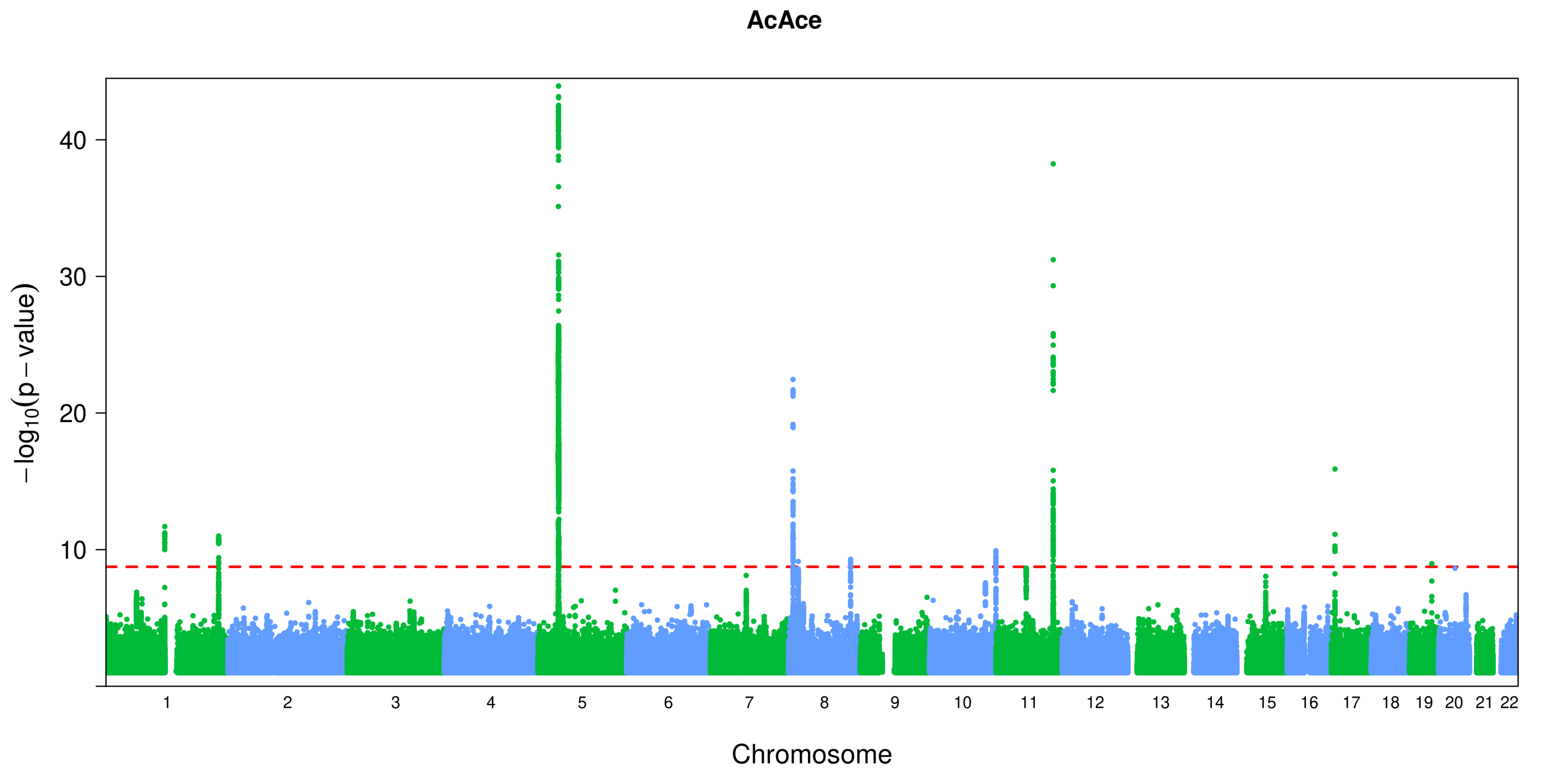
AcAce
40
30
20
10
$-\log_{10}(p-\text{value})$
Chromosome

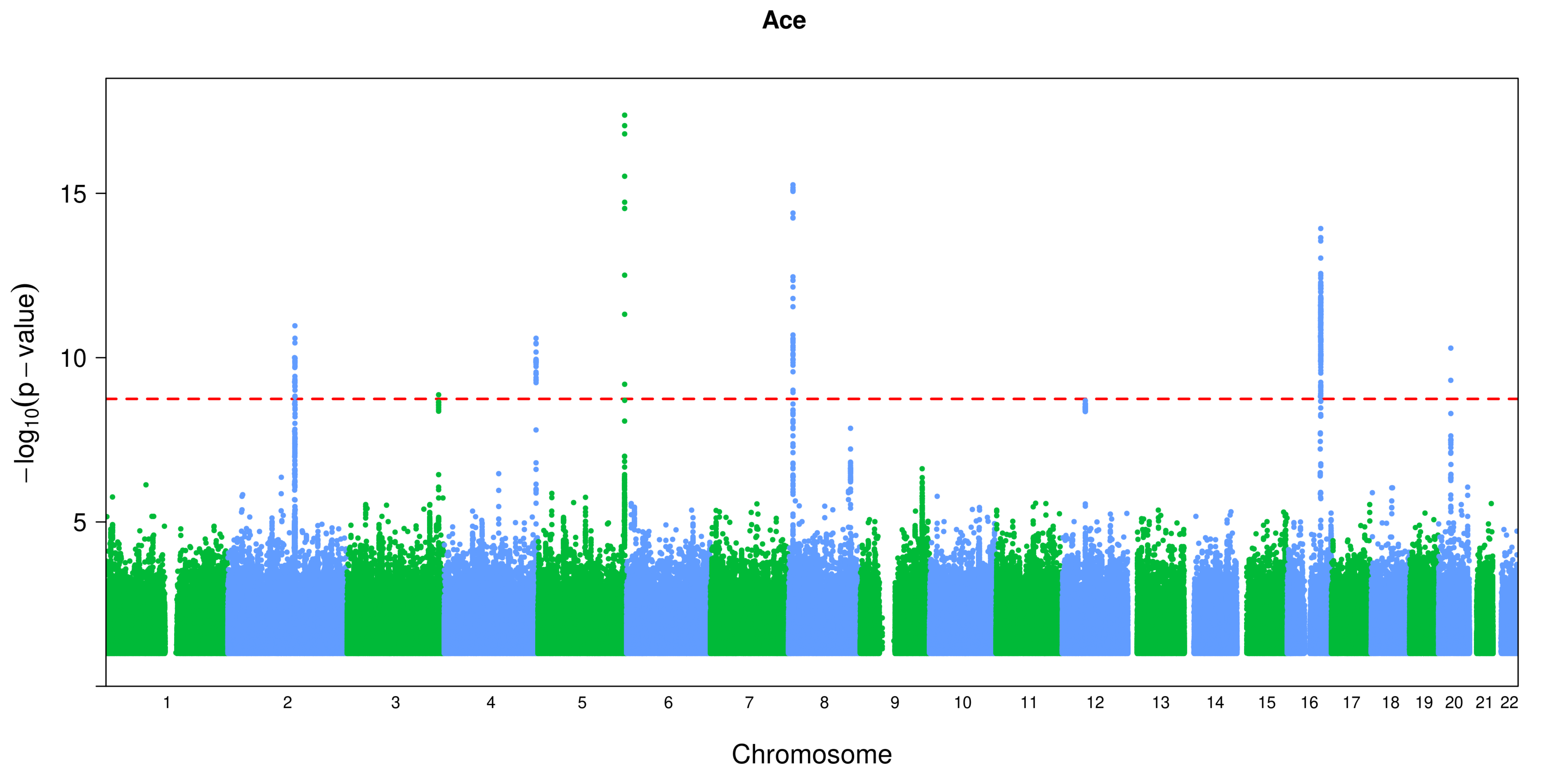

Ace
15
10
5
−log10(p − value)
Chromosome

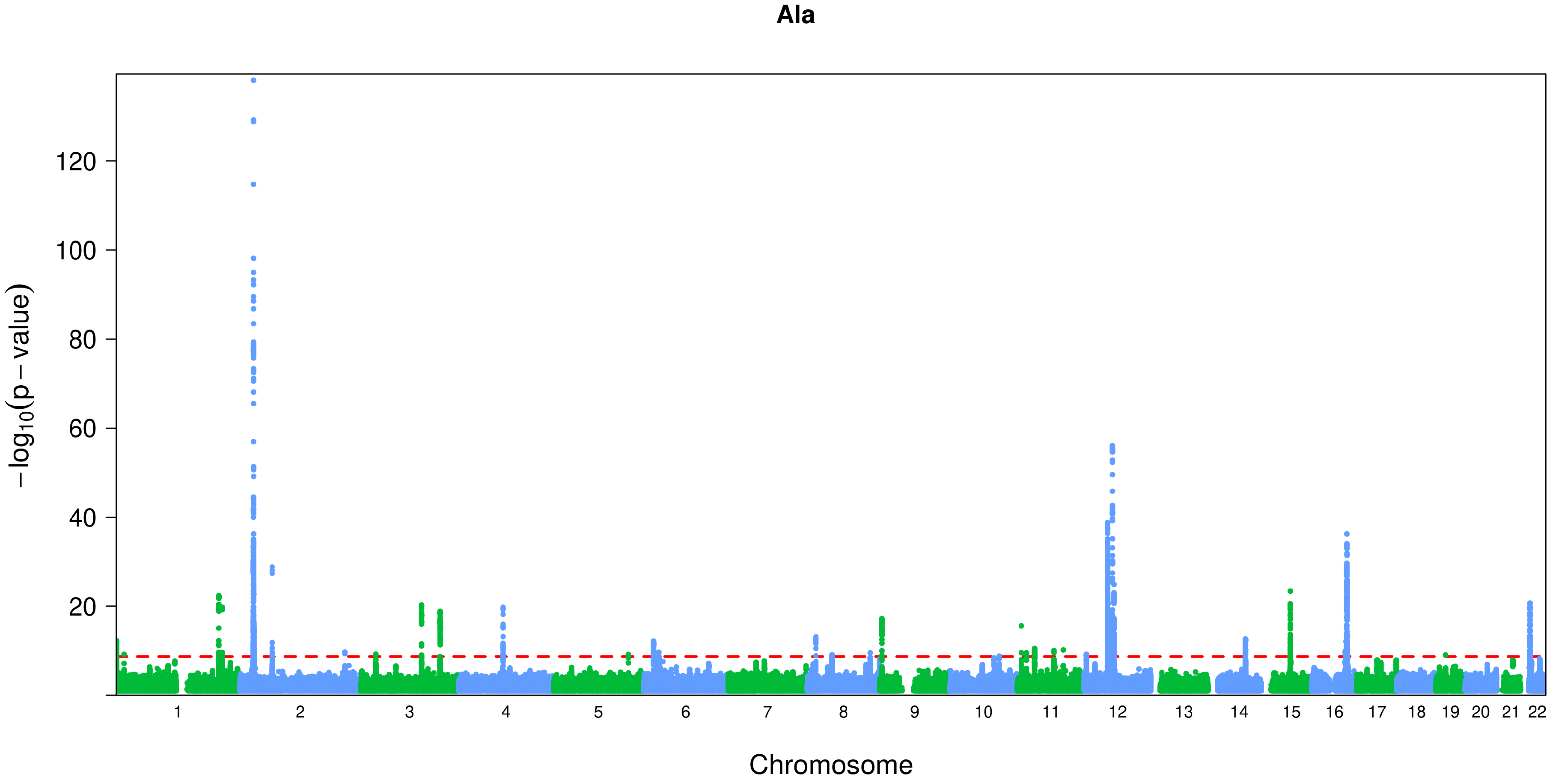
Ala
120
100
80
60
40
20
−log10(p − value)
Chromosome

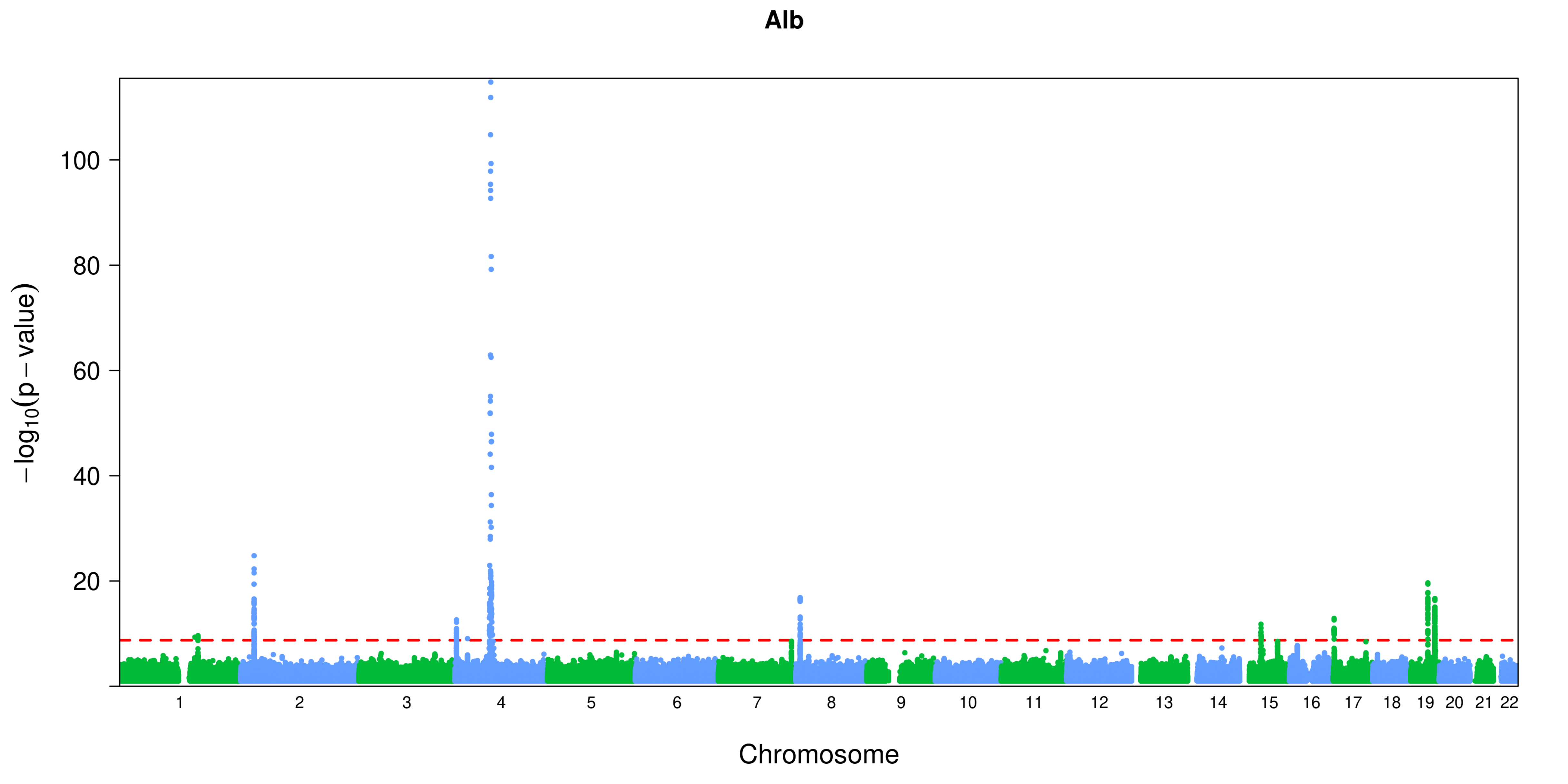
Alb
100
80
60
40
20
−log10(p − value)
Chromosome

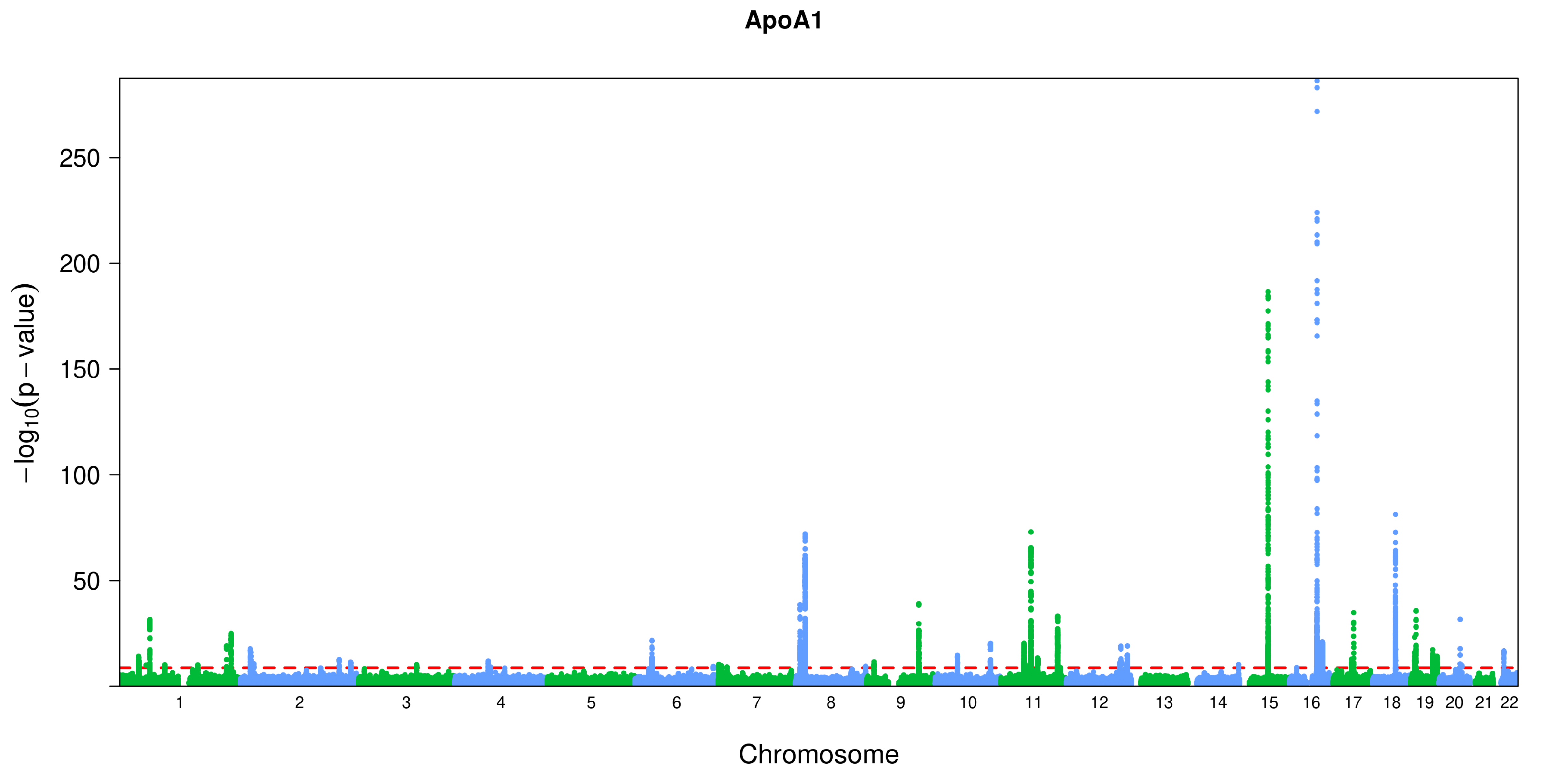
ApoA1
−log10(p − value)
250
200
150
100
50
Chromosome

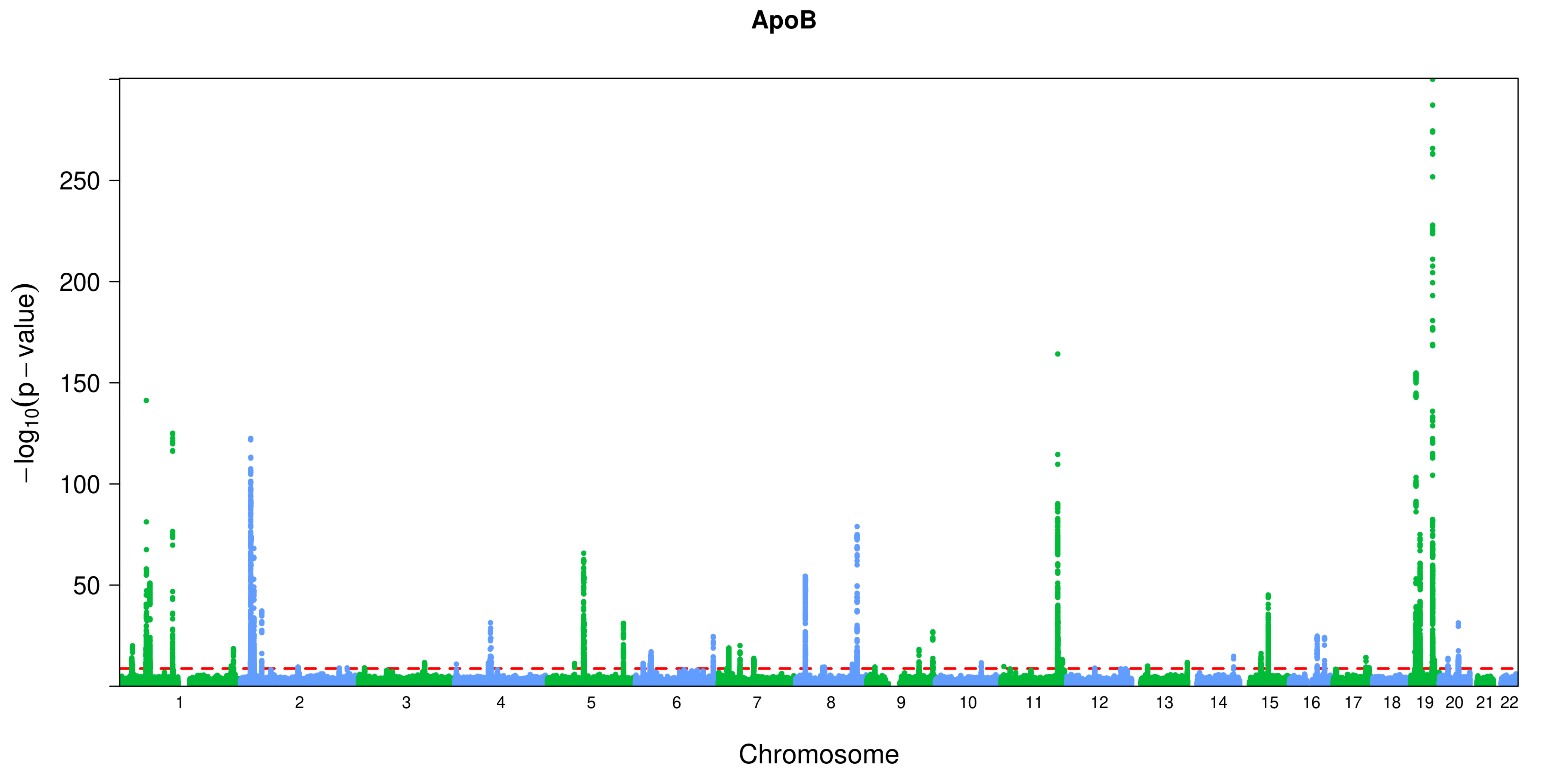

ApoB
250
200
150
100
50
−log10(p − value)
Chromosome

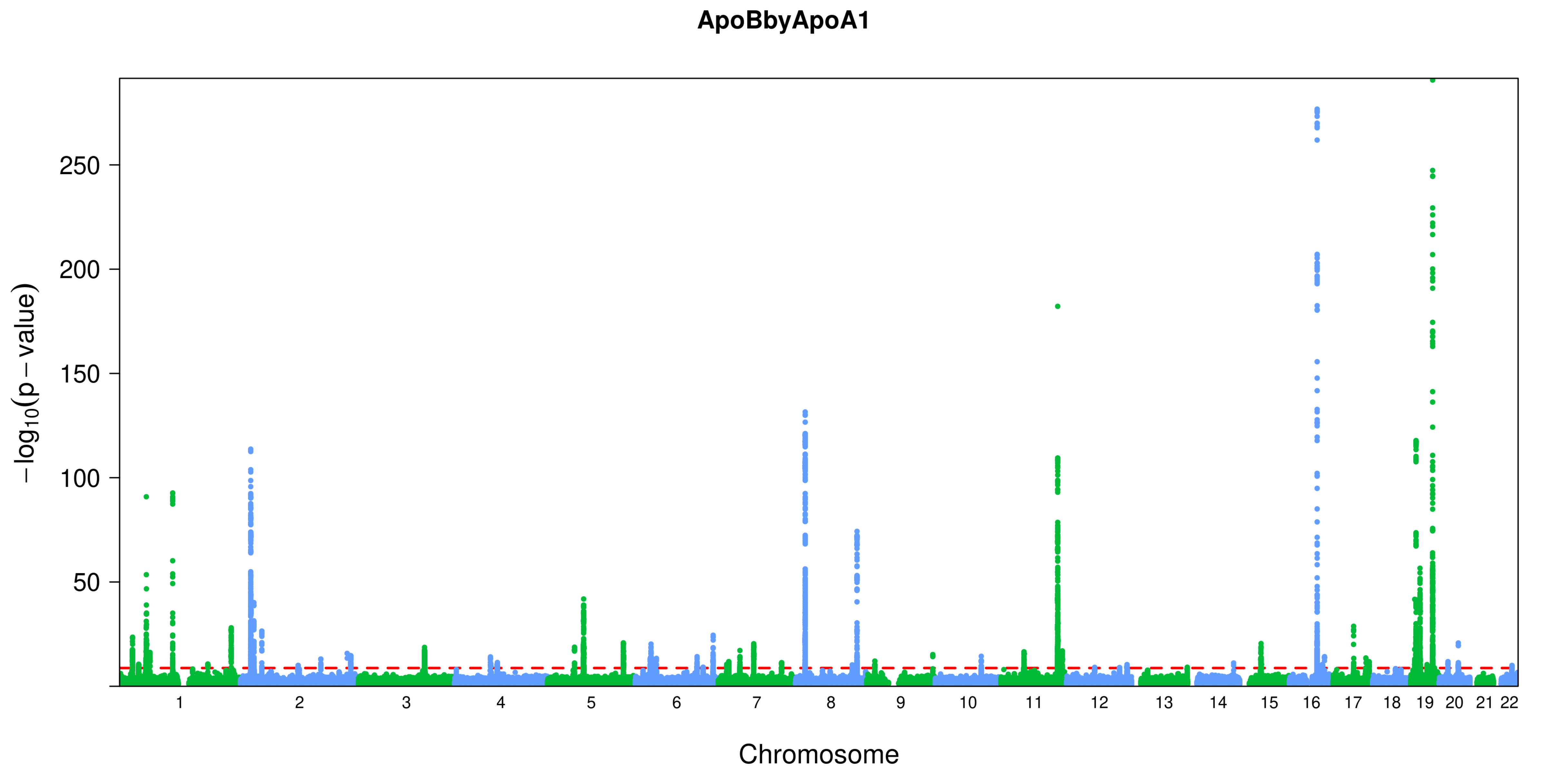

ApoBbyApoA1
−log10(p − value)
250
200
150
100
50
Chromosome

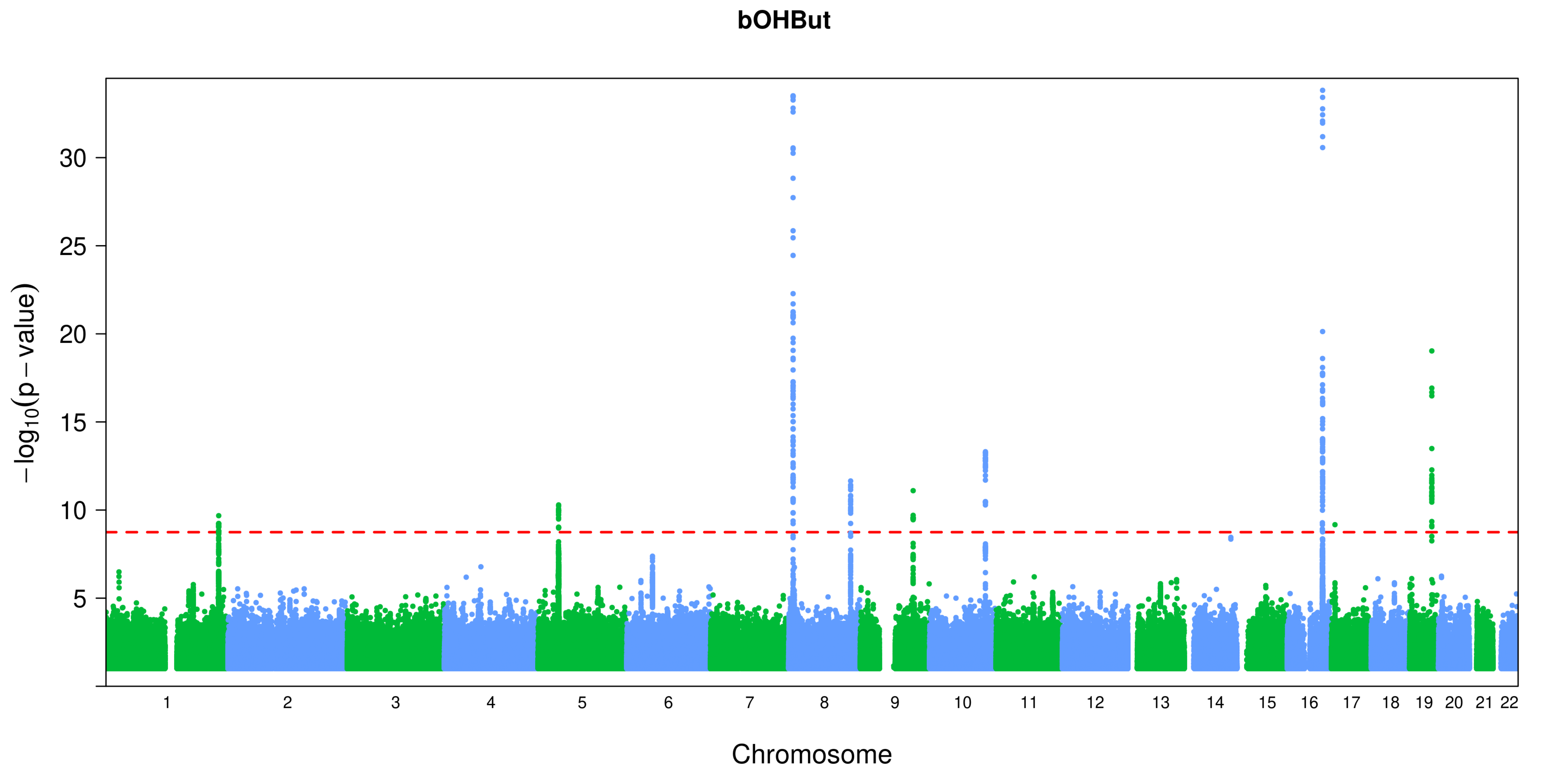
bOHBut
30
25
20
15
10
5
−log10(p − value)
Chromosome

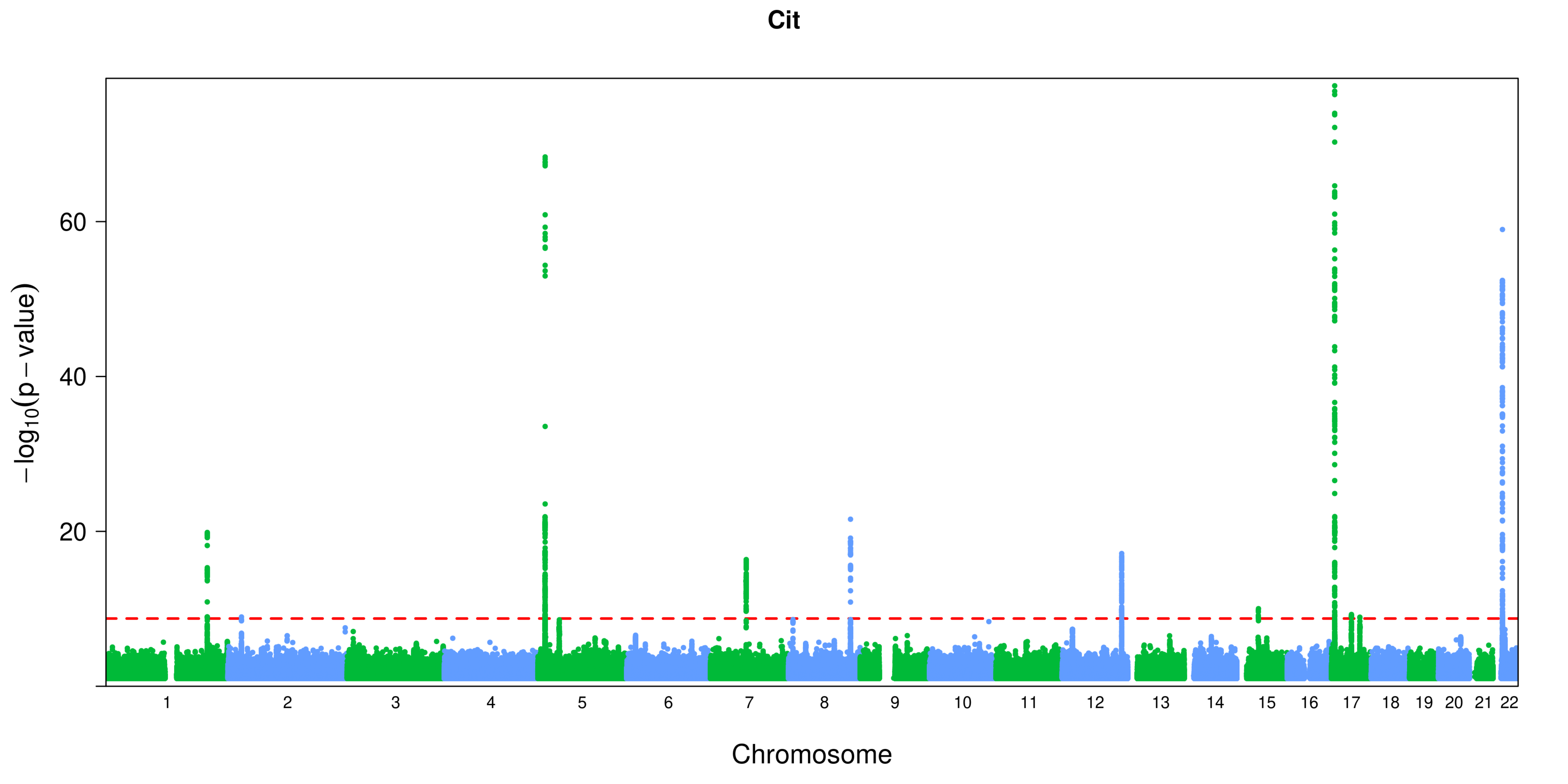
Cit
60
40
20
−log10(p − value)
Chromosome

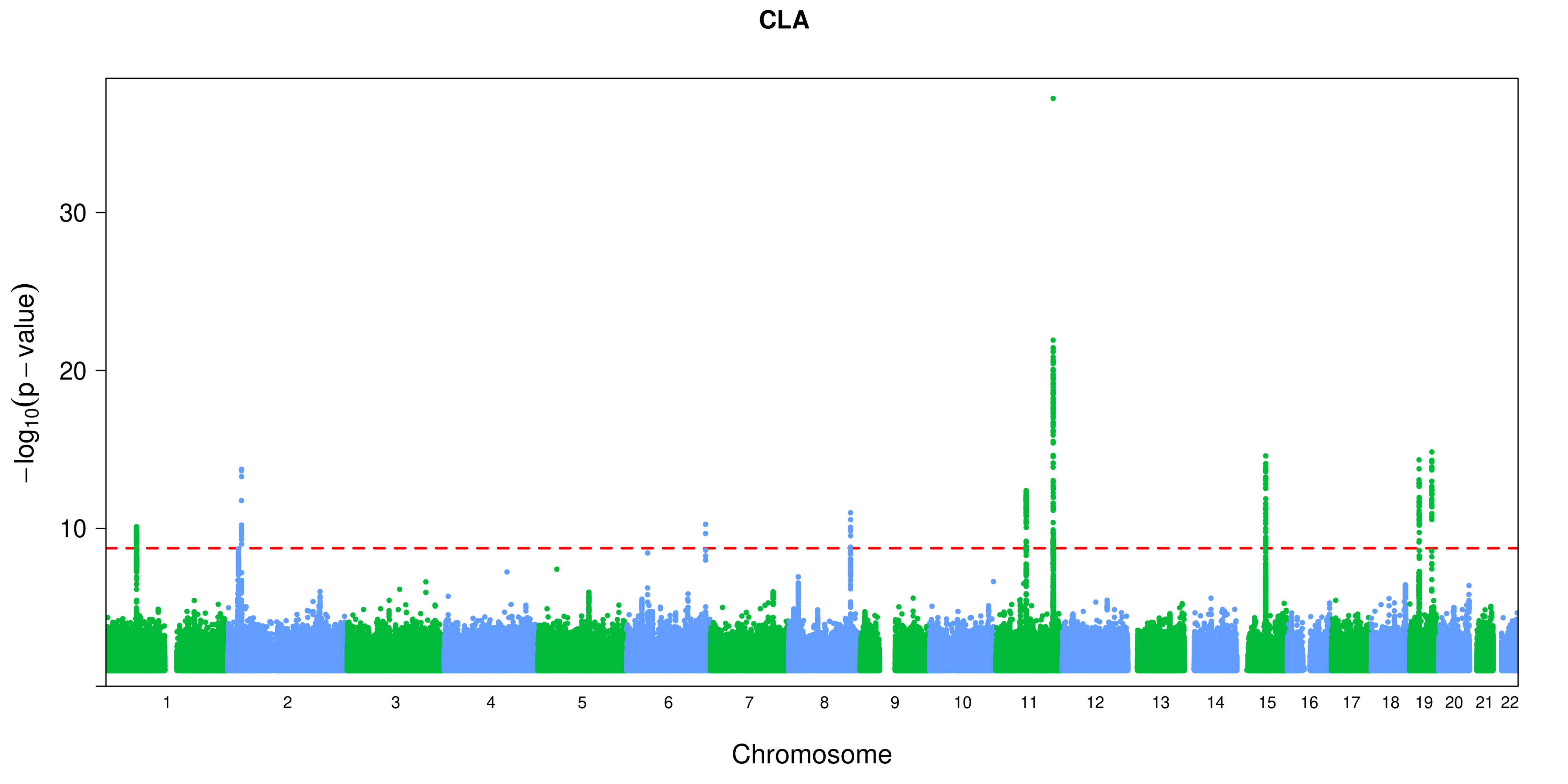
CLA
30
20
10
−log10(p − value)
Chromosome

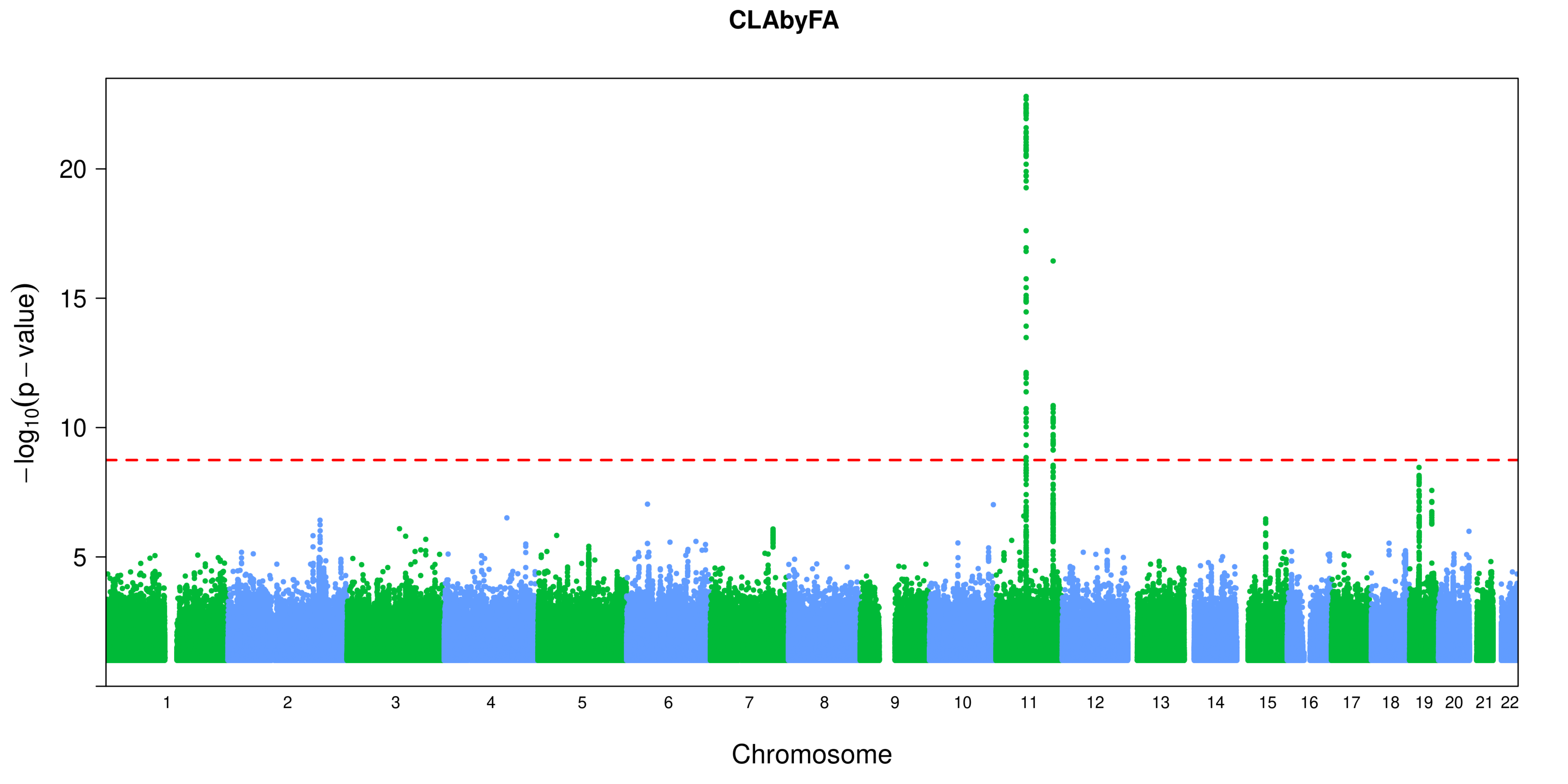
CLAbyFA
−log10(p − value)
20
15
10
5
Chromosome

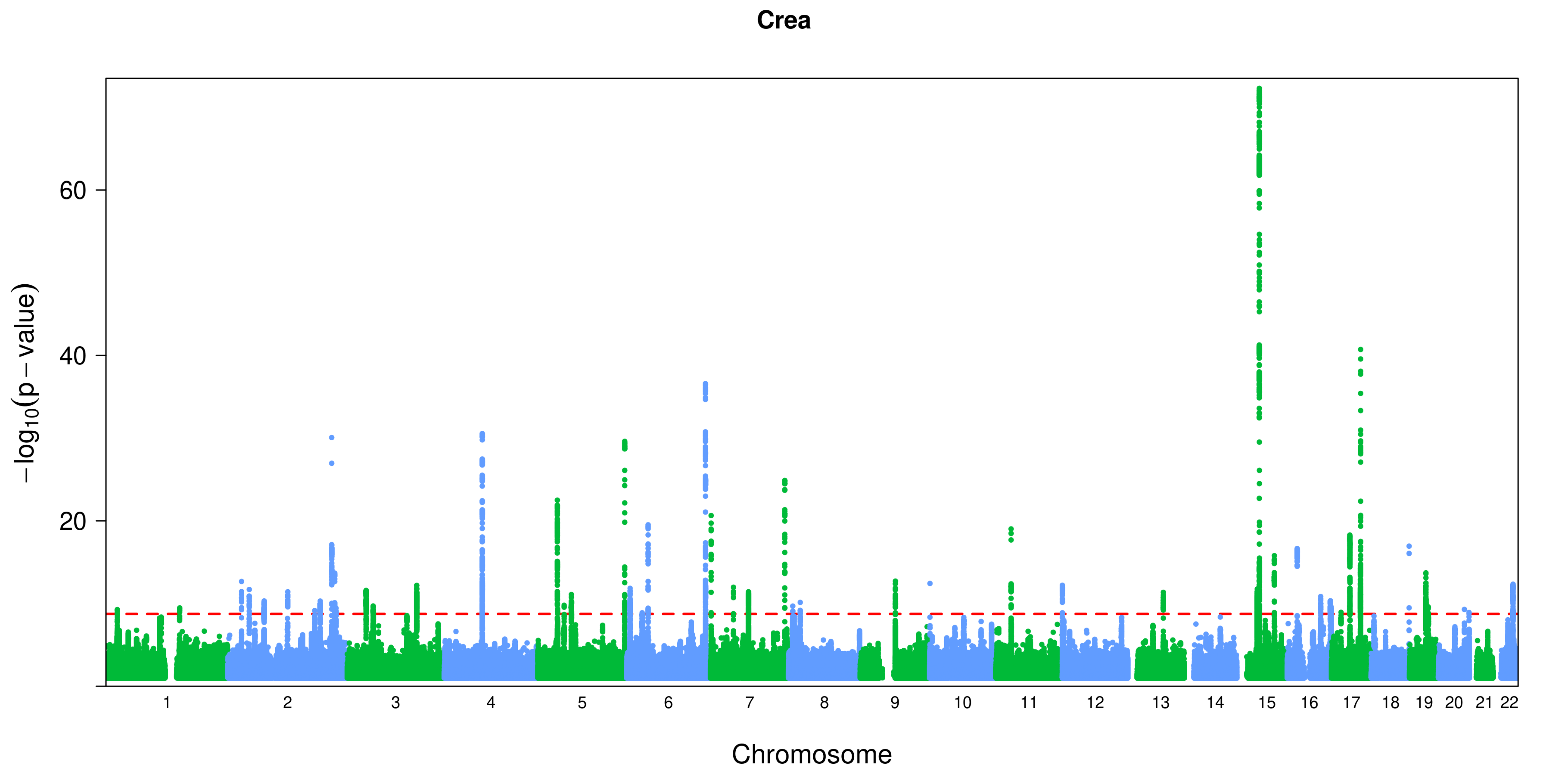
Crea
−log10(p − value)
60
40
20
Chromosome

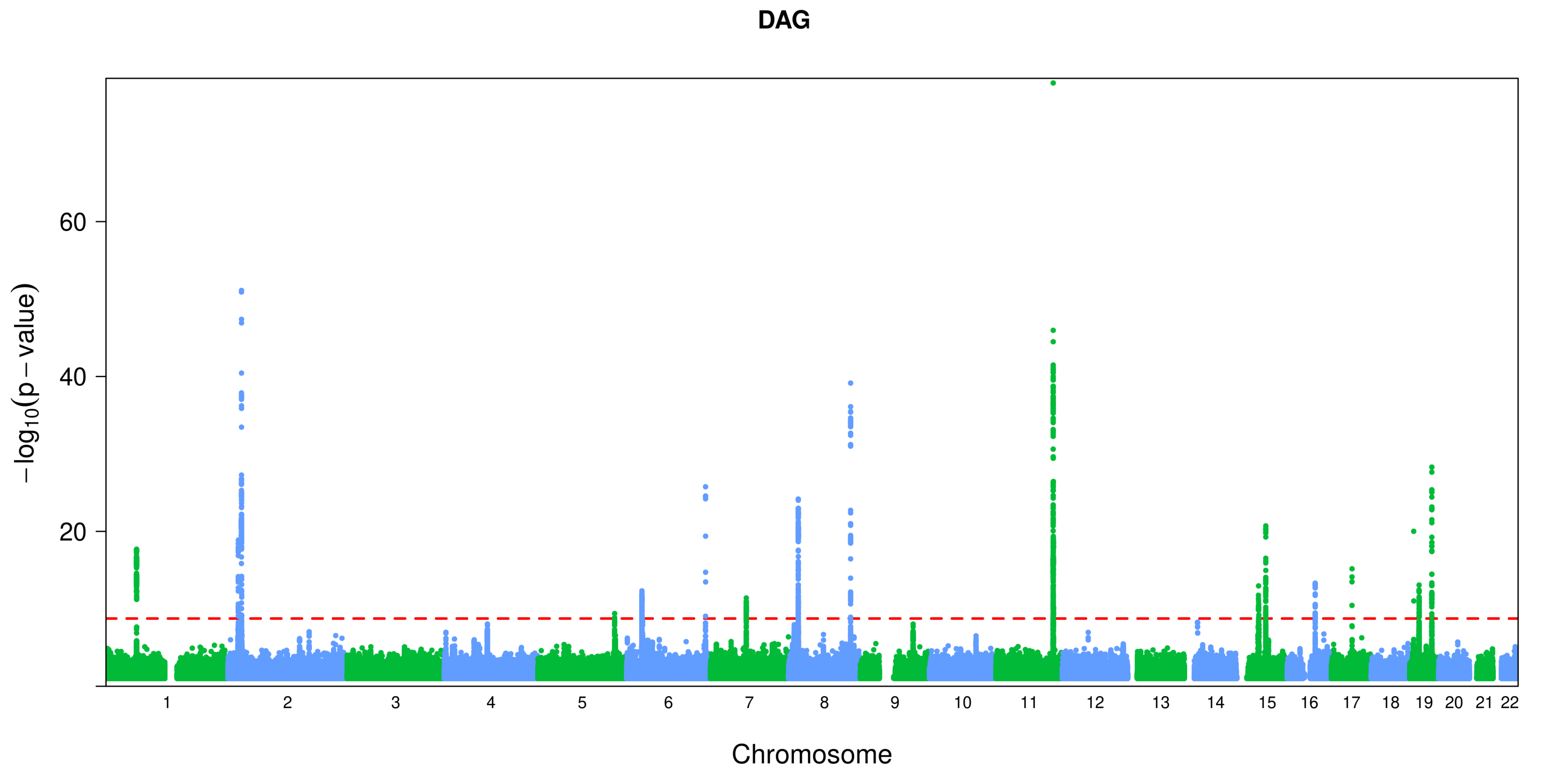
DAG
−log10(p − value)
60
40
20
Chromosome

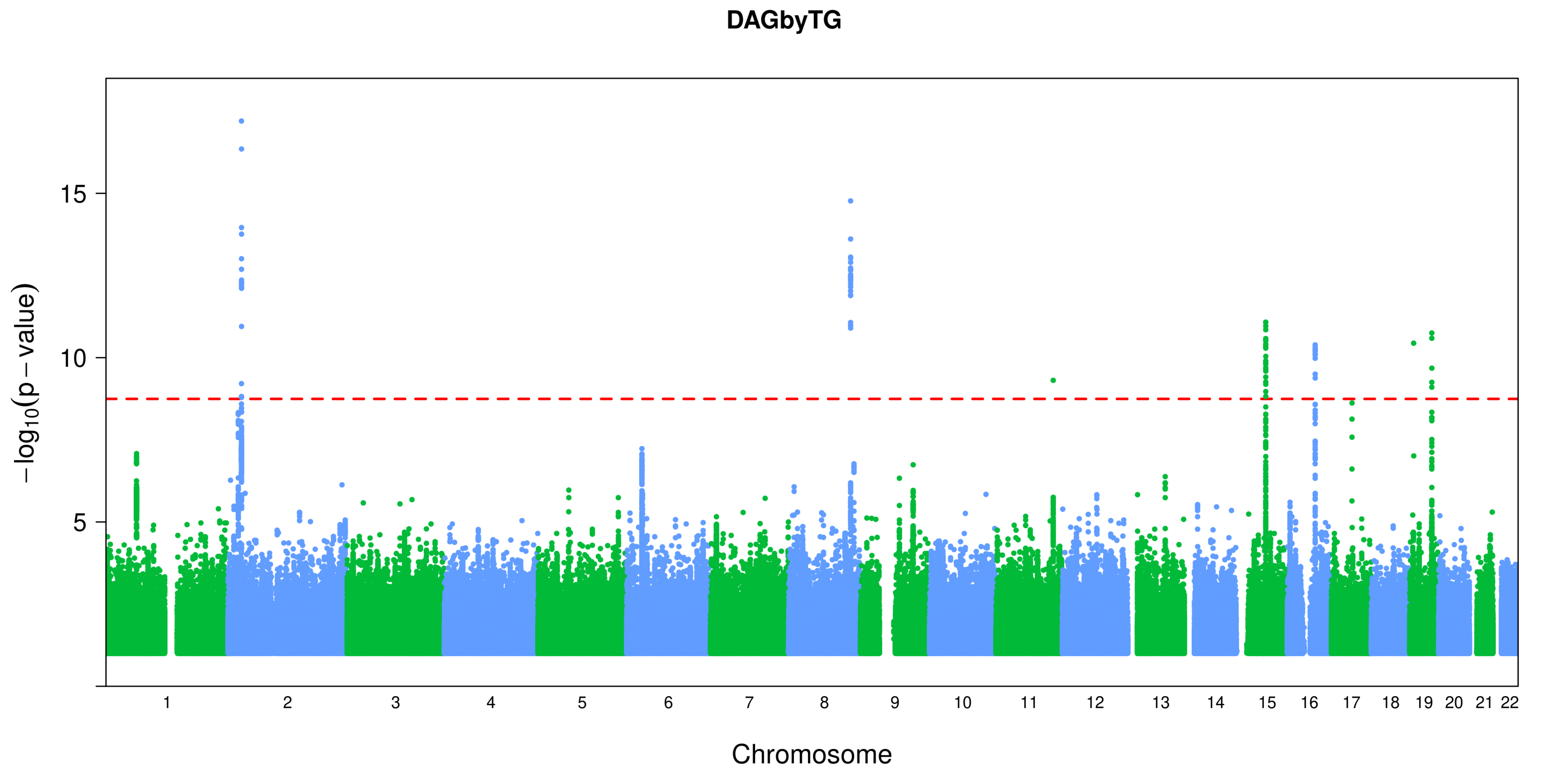
DAGbyTG
15
10
5
−log10(p − value)
Chromosome

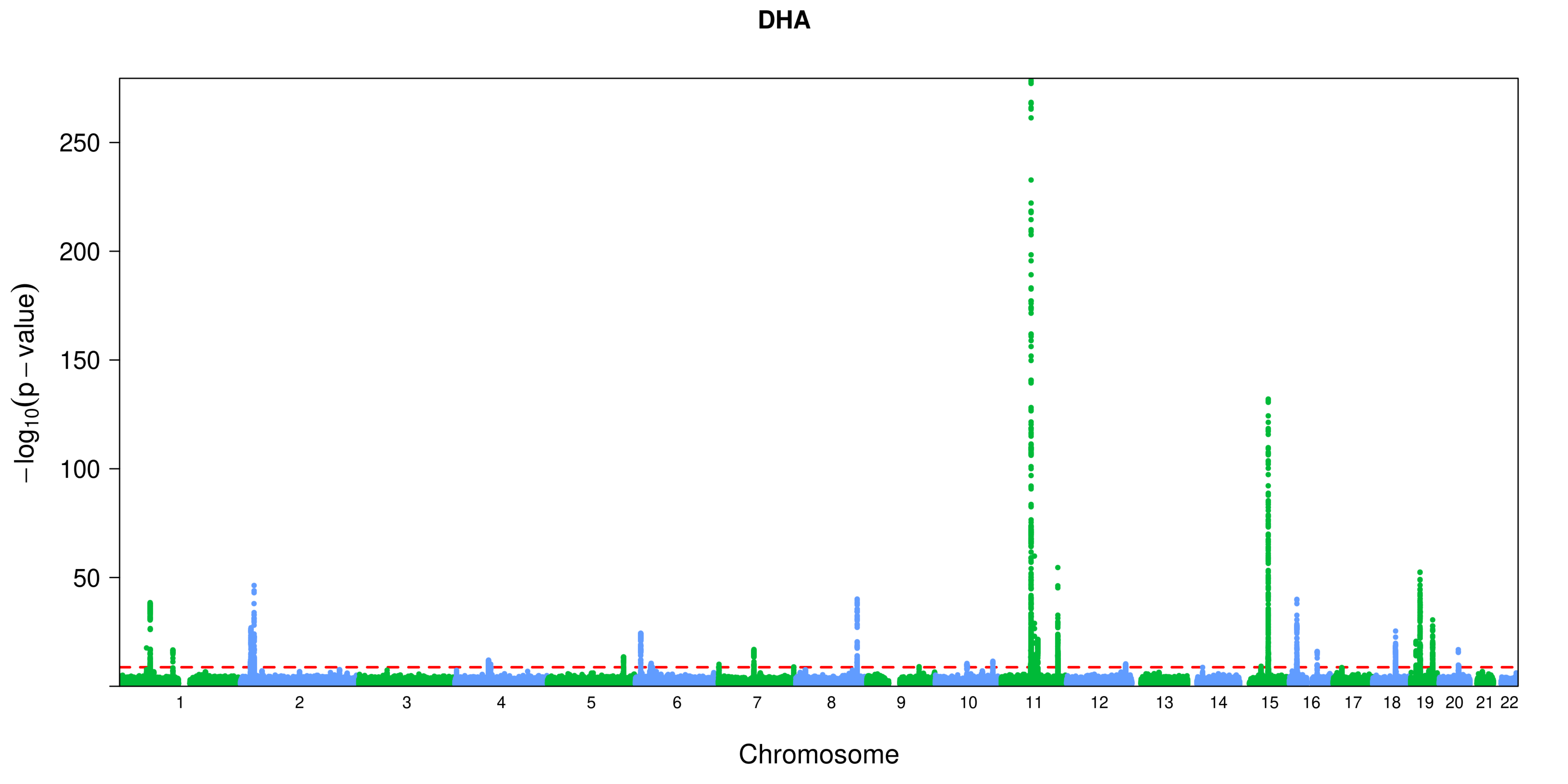
DHA
250
200
150
100
50
−log10(p − value)
Chromosome

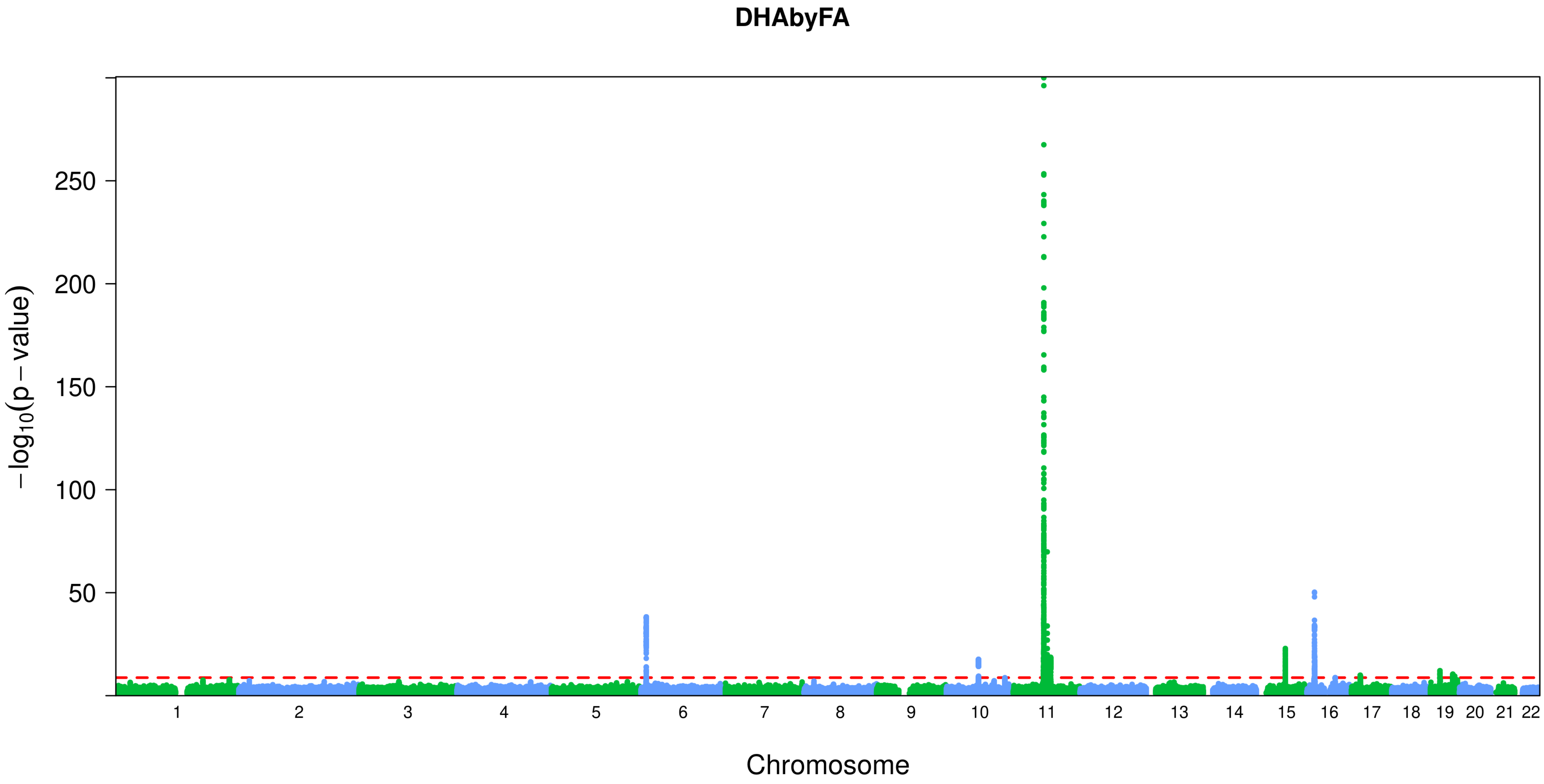

DHAbyFA
250
200
150
100
50
−log10(p−value)
Chromosome

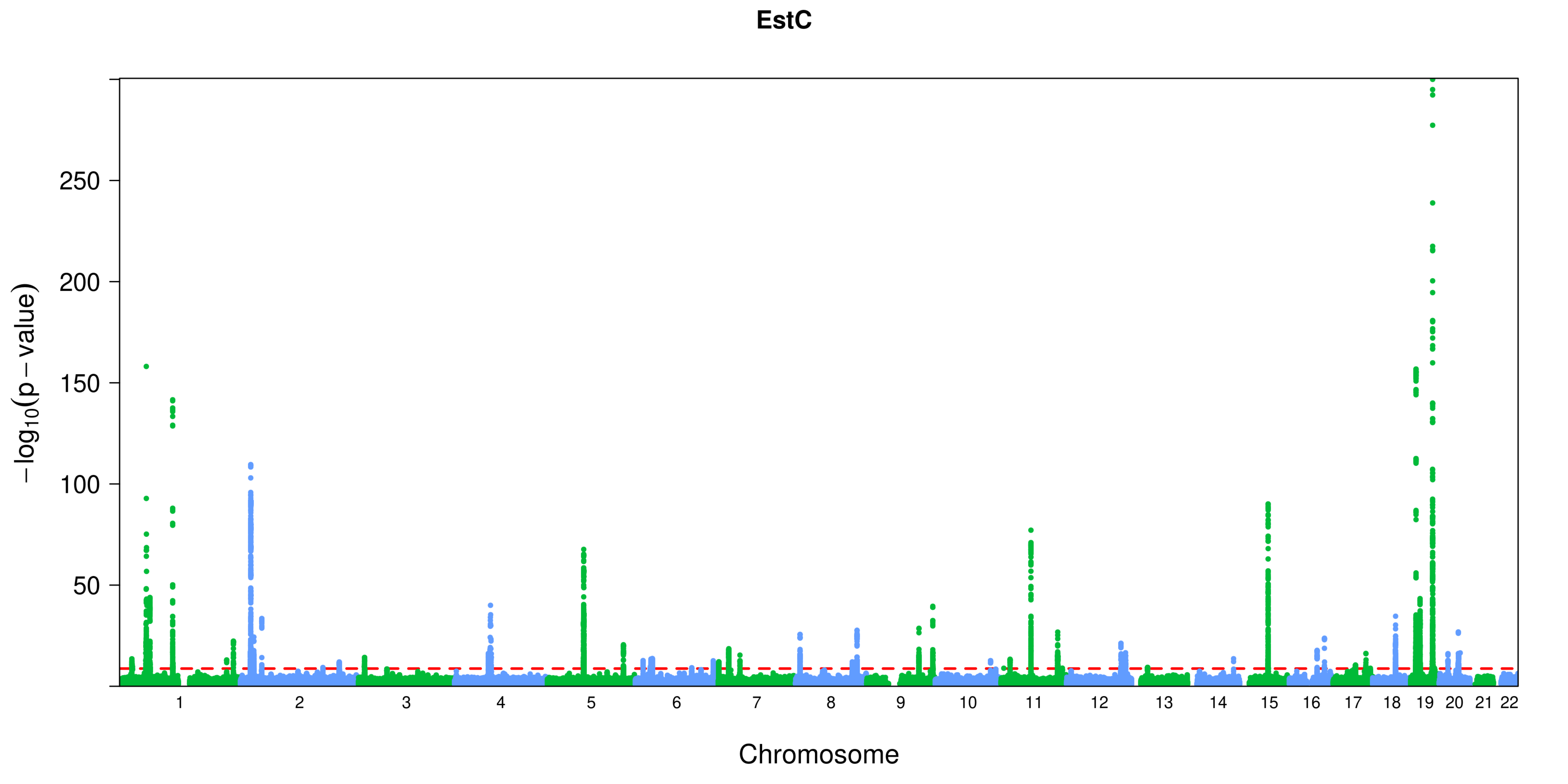
EstC
−log10(p − value)
250
200
150
100
50
Chromosome

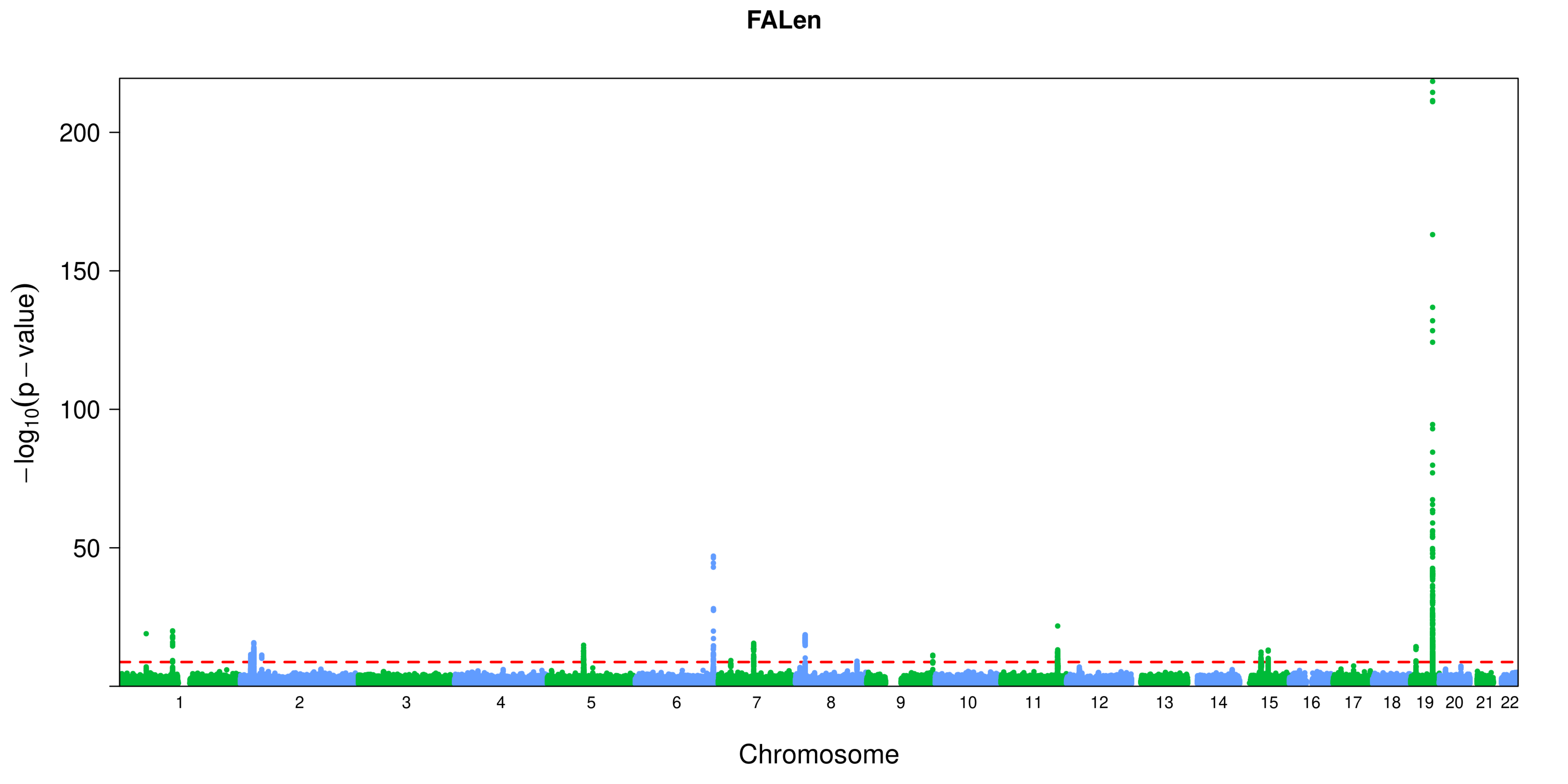
FALen
200
150
100
50
−log10(p − value)
Chromosome

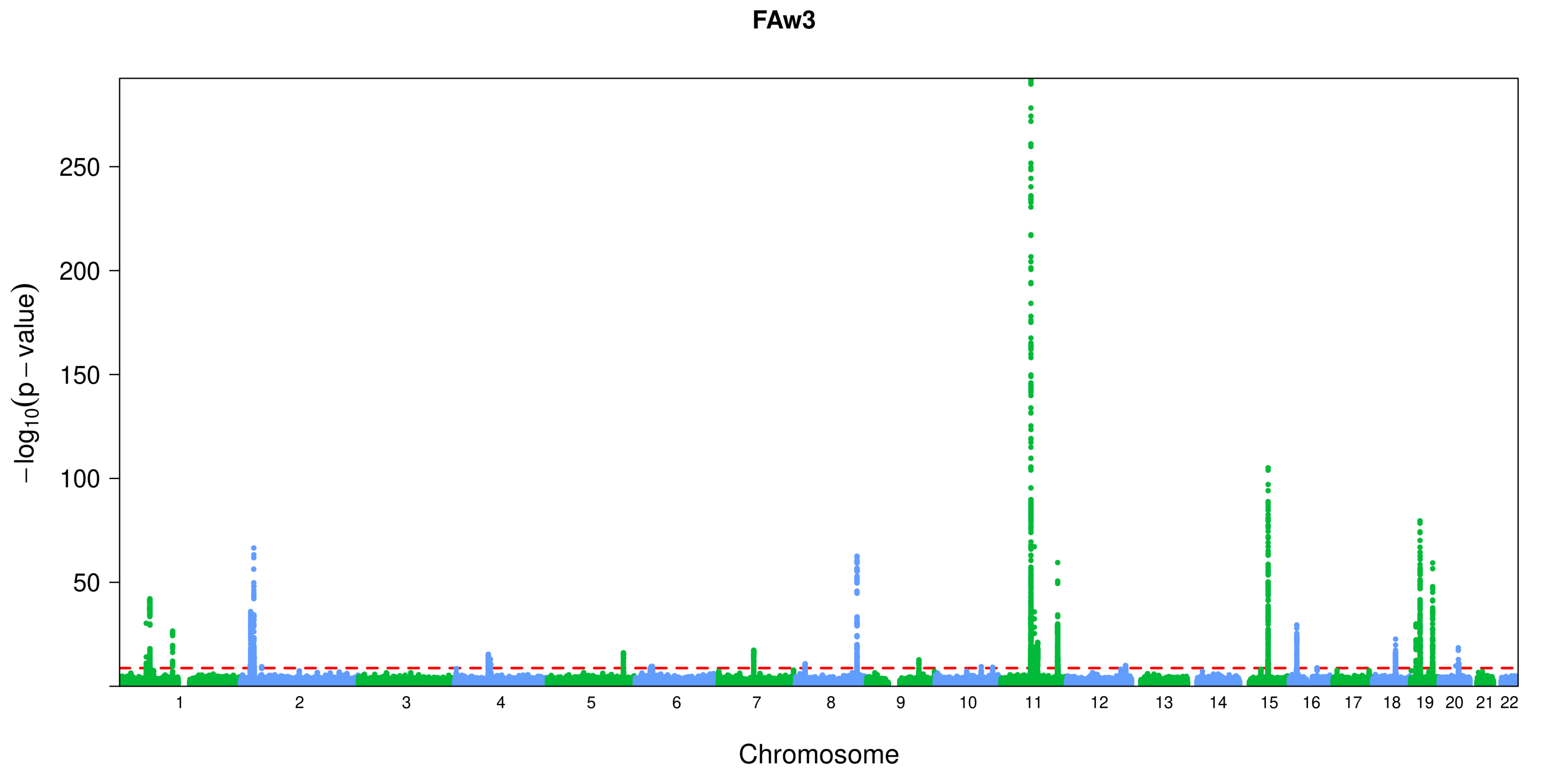

FAw3
250
200
150
100
50
$-\log_{10}(p-value)$
Chromosome

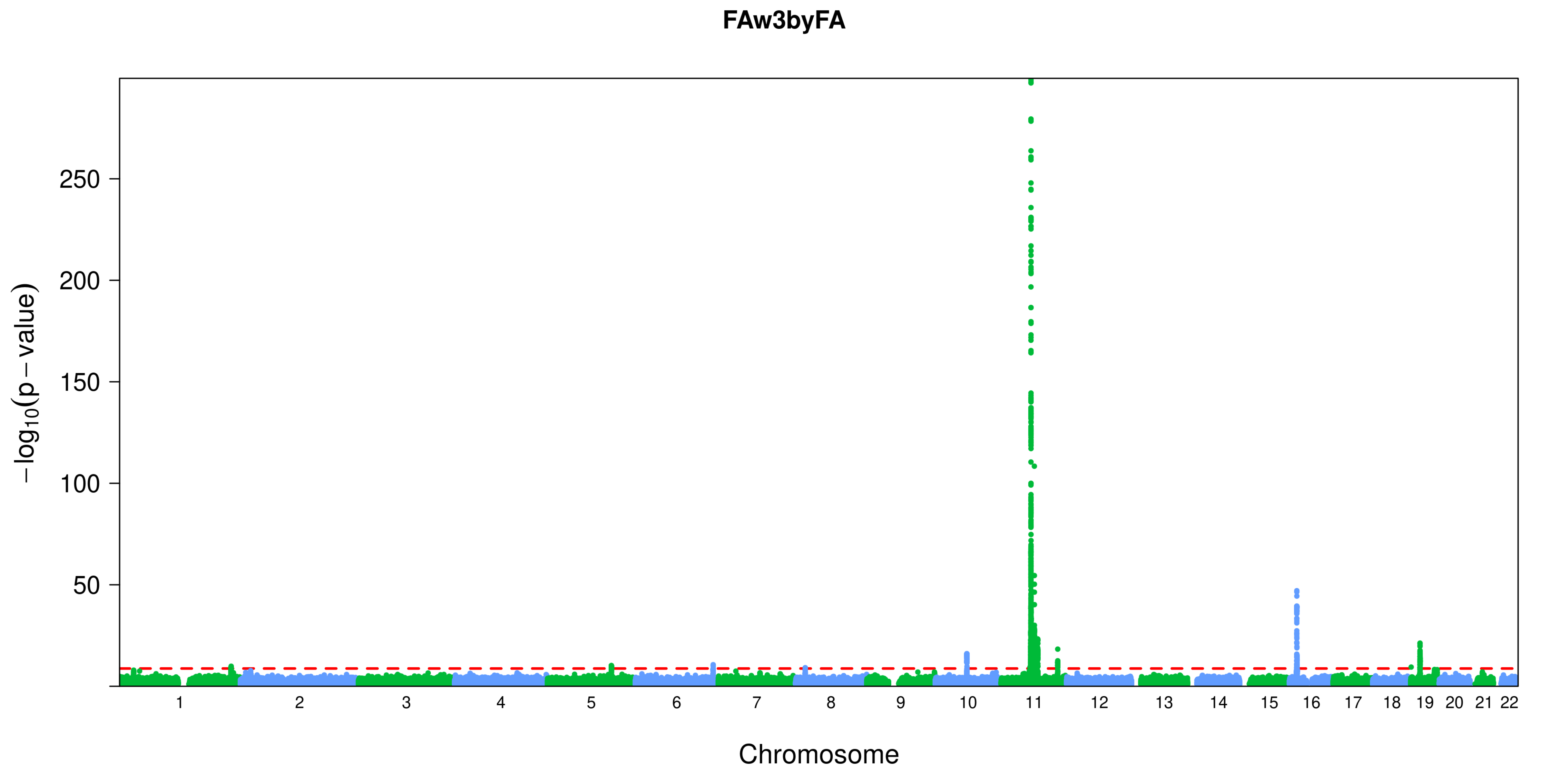

FAw3byFA
−log10(p − value)
250
200
150
100
50
Chromosome

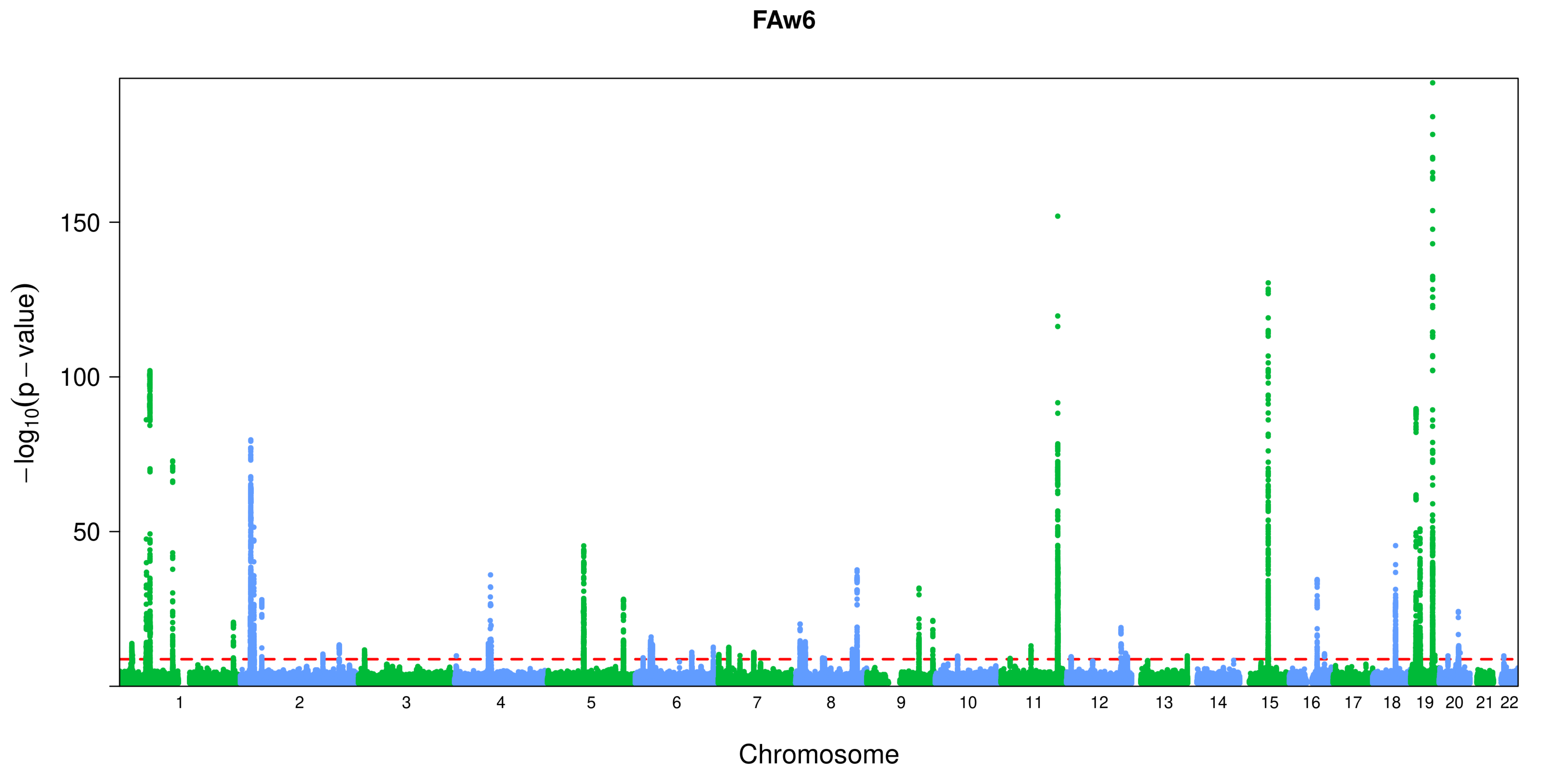
FAw6
150
100
50
−log10(p − value)
Chromosome

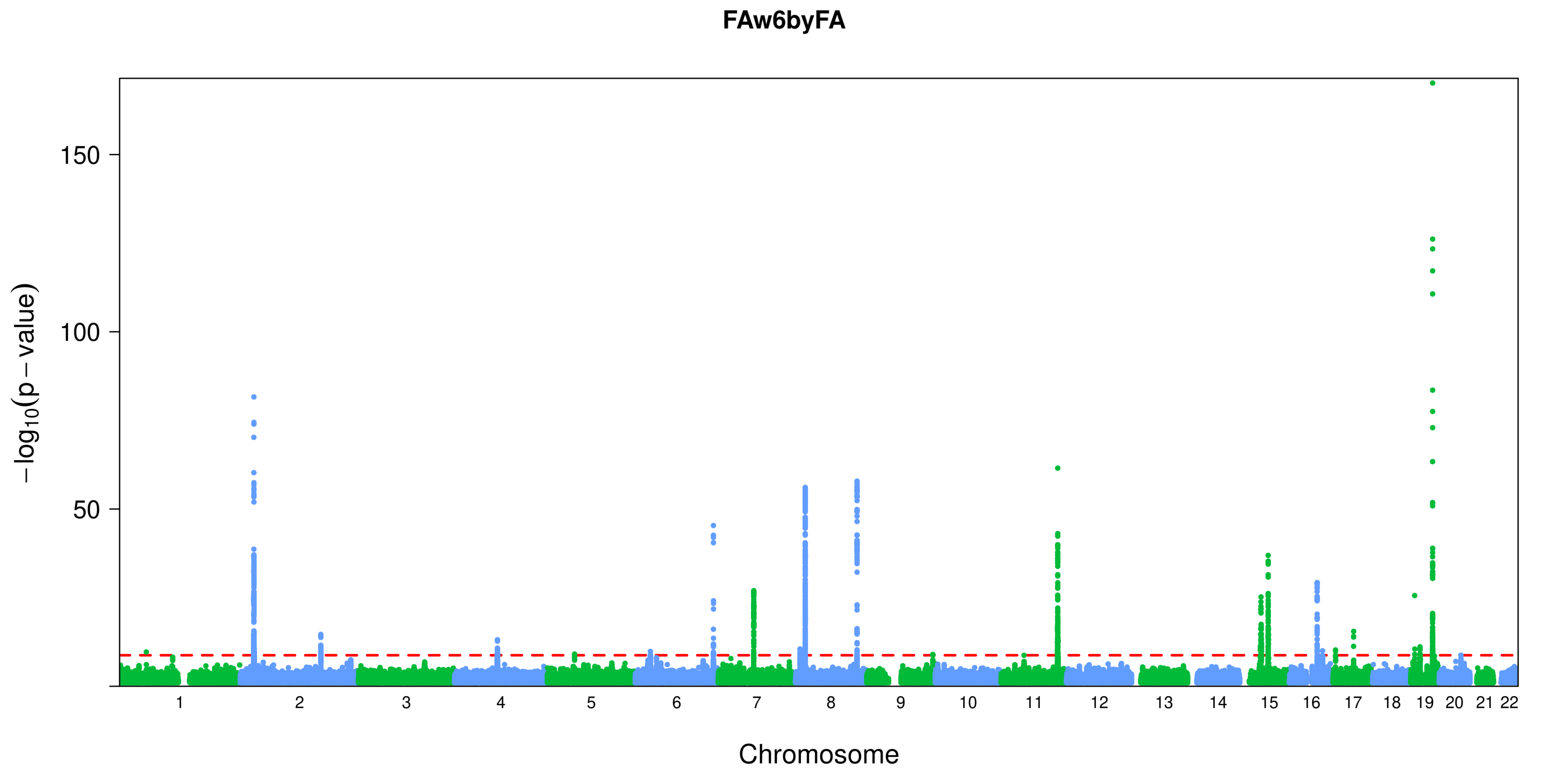
FAw6byFA
−log10(p − value)
150
100
50
1 2 3 4 5 6 7 8 9 10 11 12 13 14 15 16 17 18 19 20 21 22
Chromosome

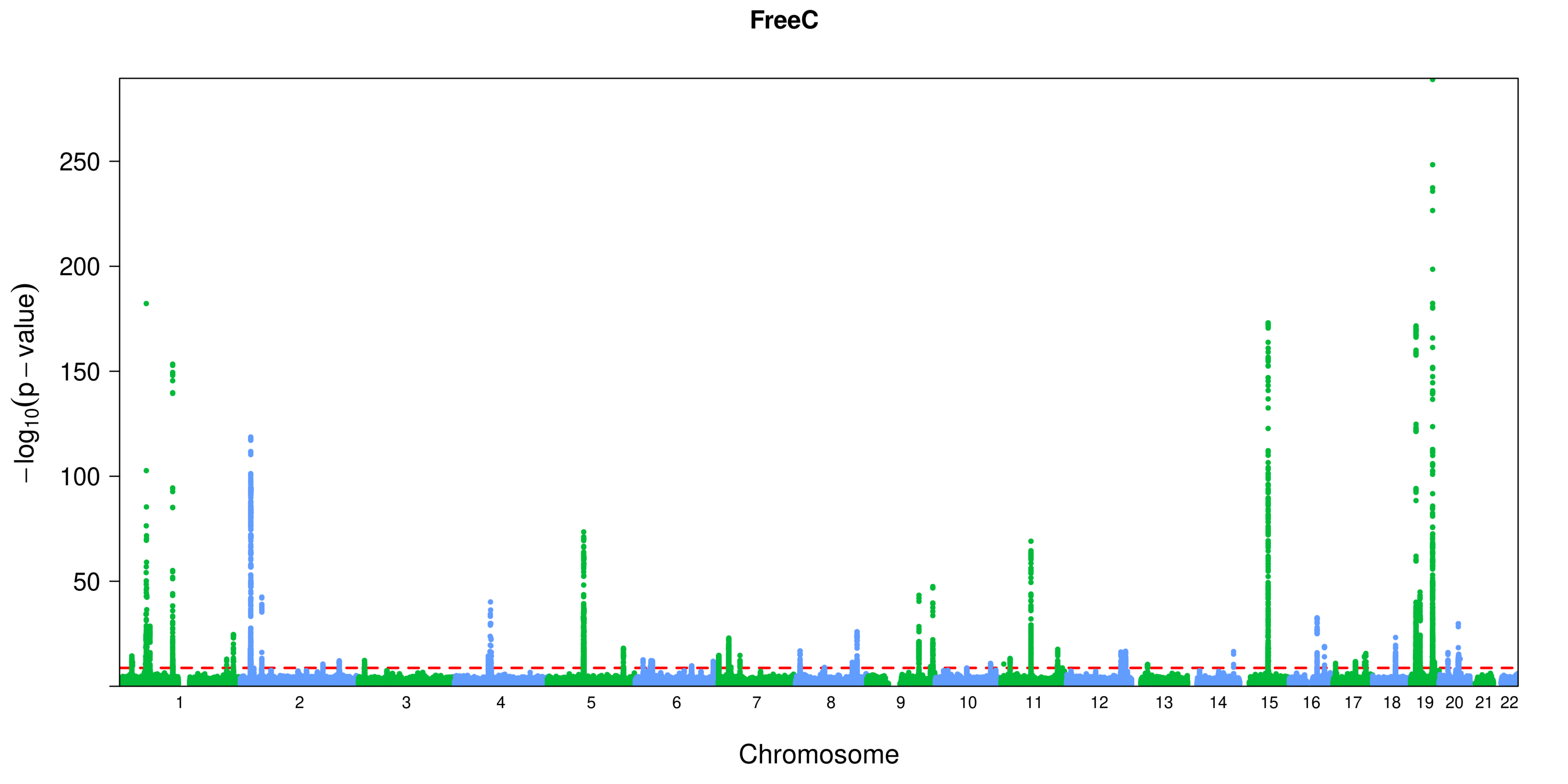
FreeC
$-\log_{10}(p-\text{value})$
250
200
150
100
50
Chromosome

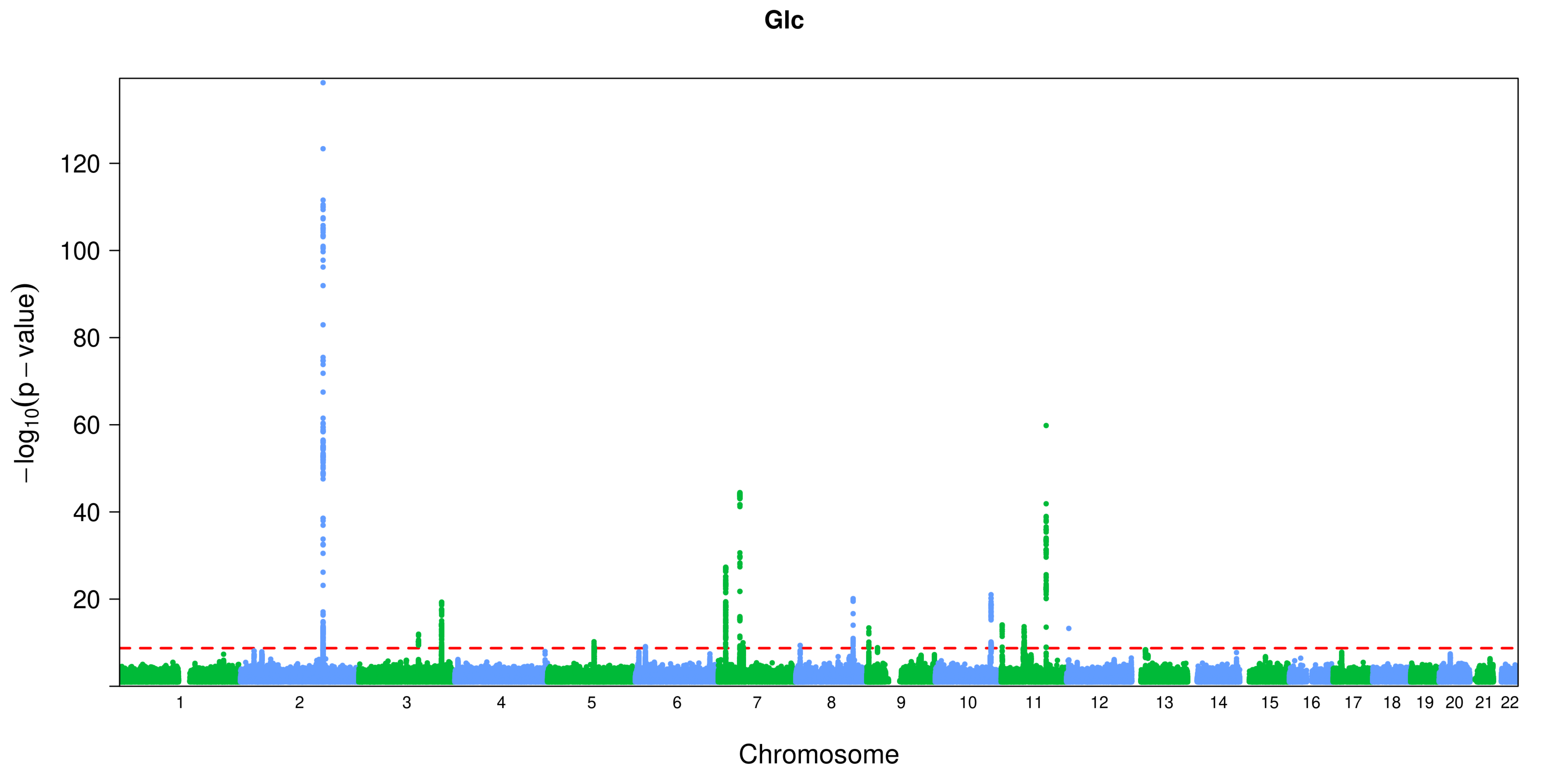
Glc
−log10(p − value)
120
100
80
60
40
20
Chromosome

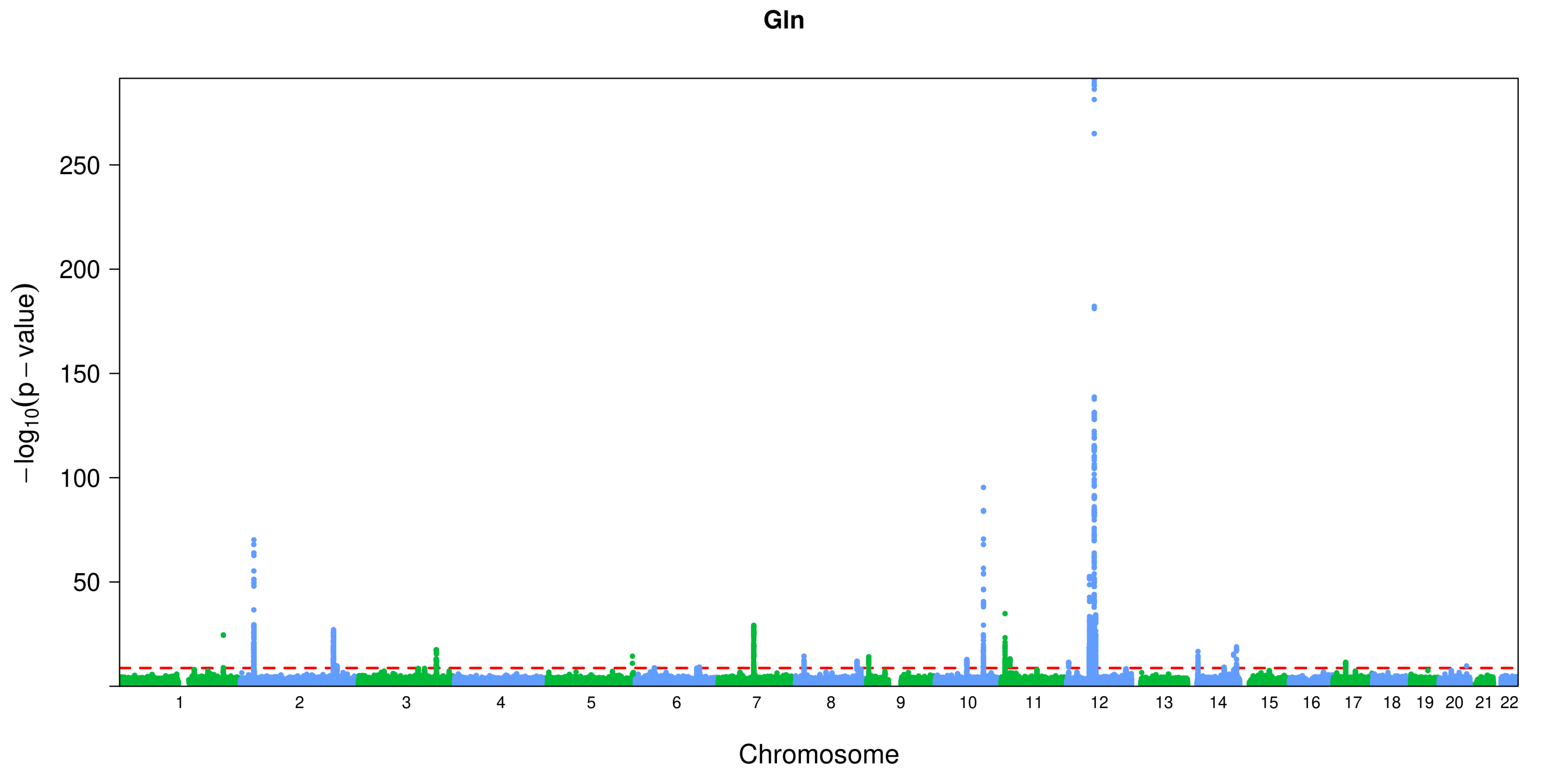

Gln
−log10(p − value)
250
200
150
100
50
Chromosome

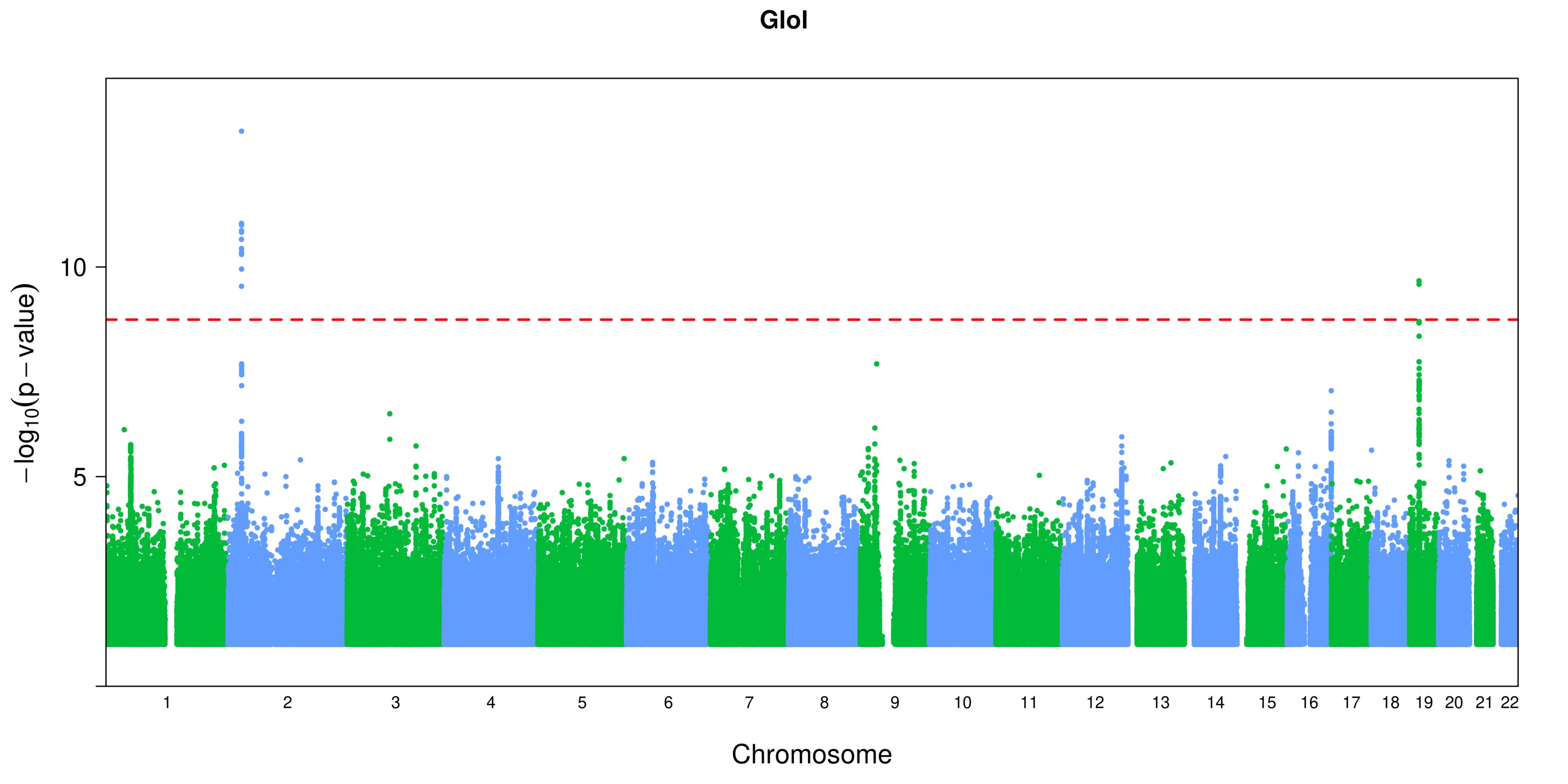
GloI
−log10(p − value)
10
5
Chromosome

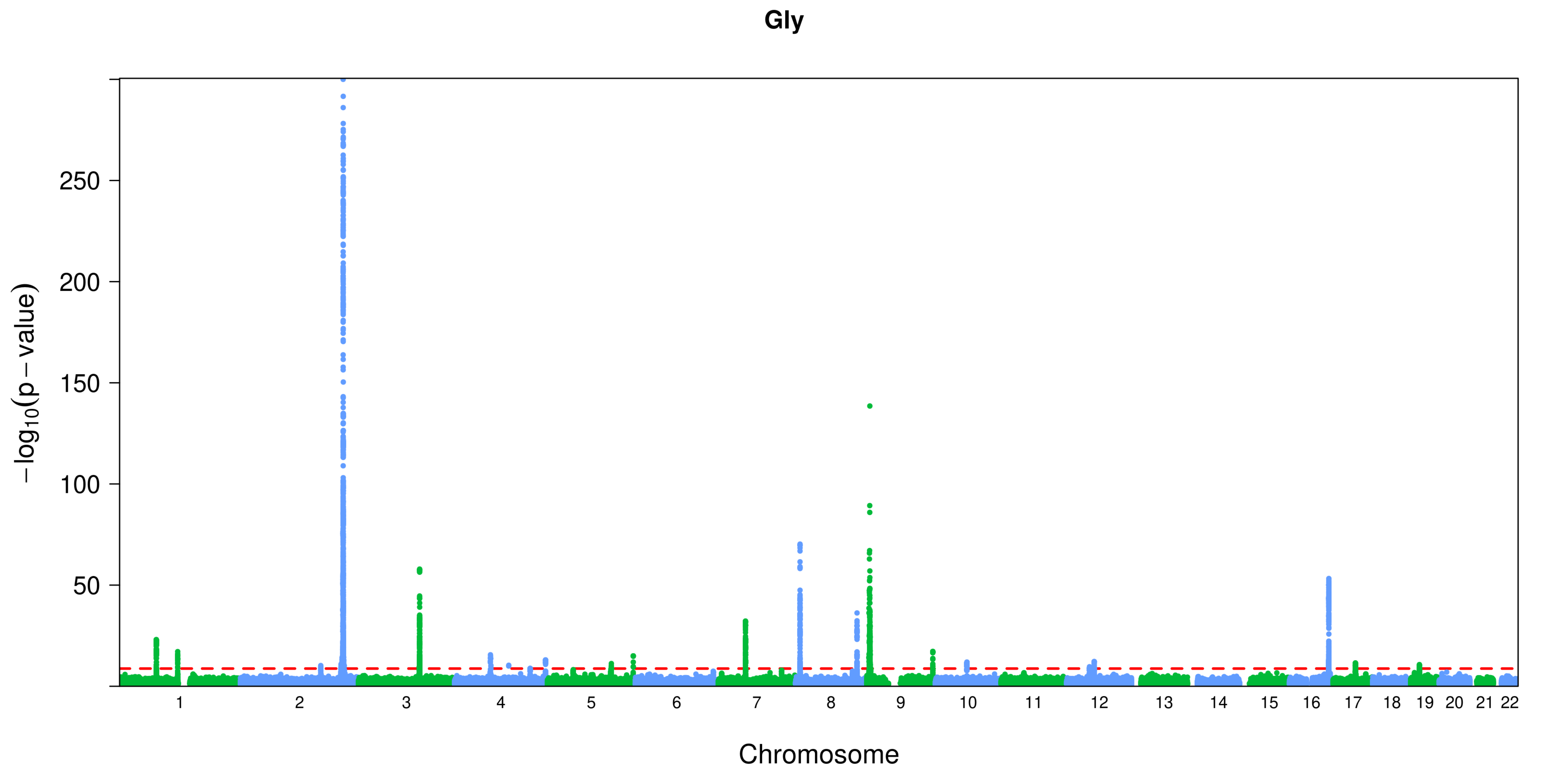
Gly
−log10(p − value)
250
200
150
100
50
Chromosome

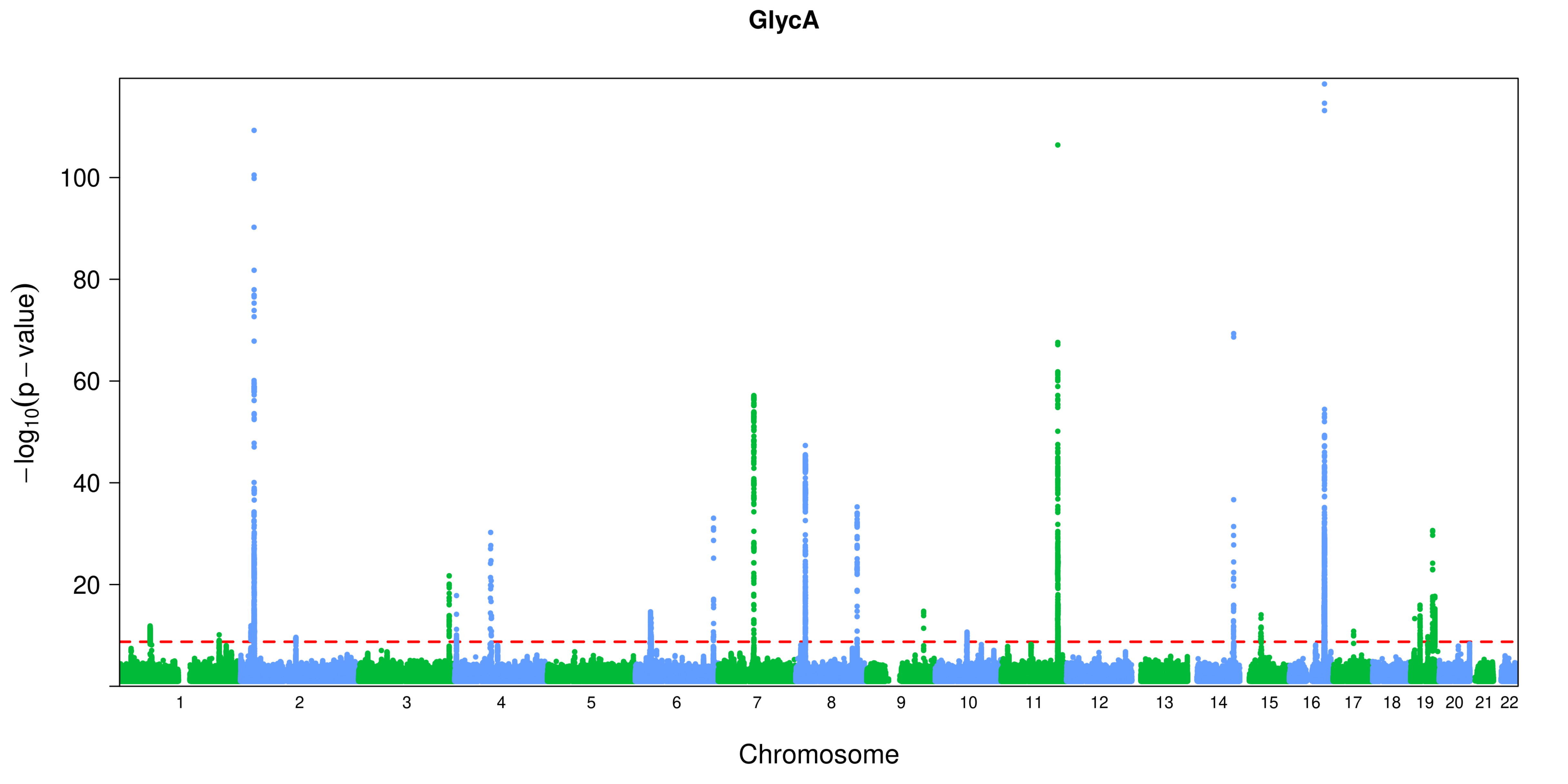
GlycA
−log10(p − value)
100
80
60
40
20
Chromosome

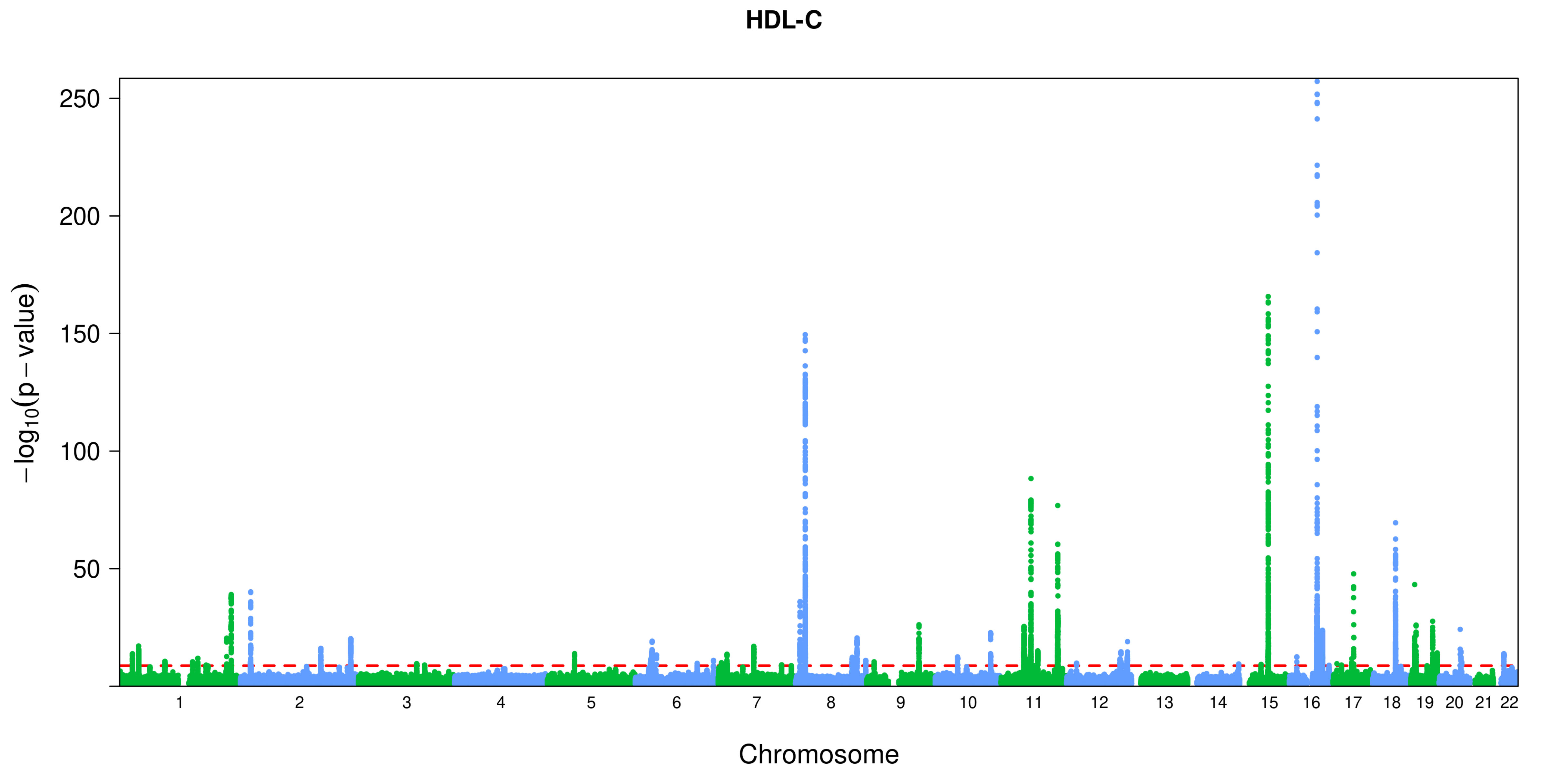

HDL-C
250
200
150
100
50
−log10(p − value)
Chromosome

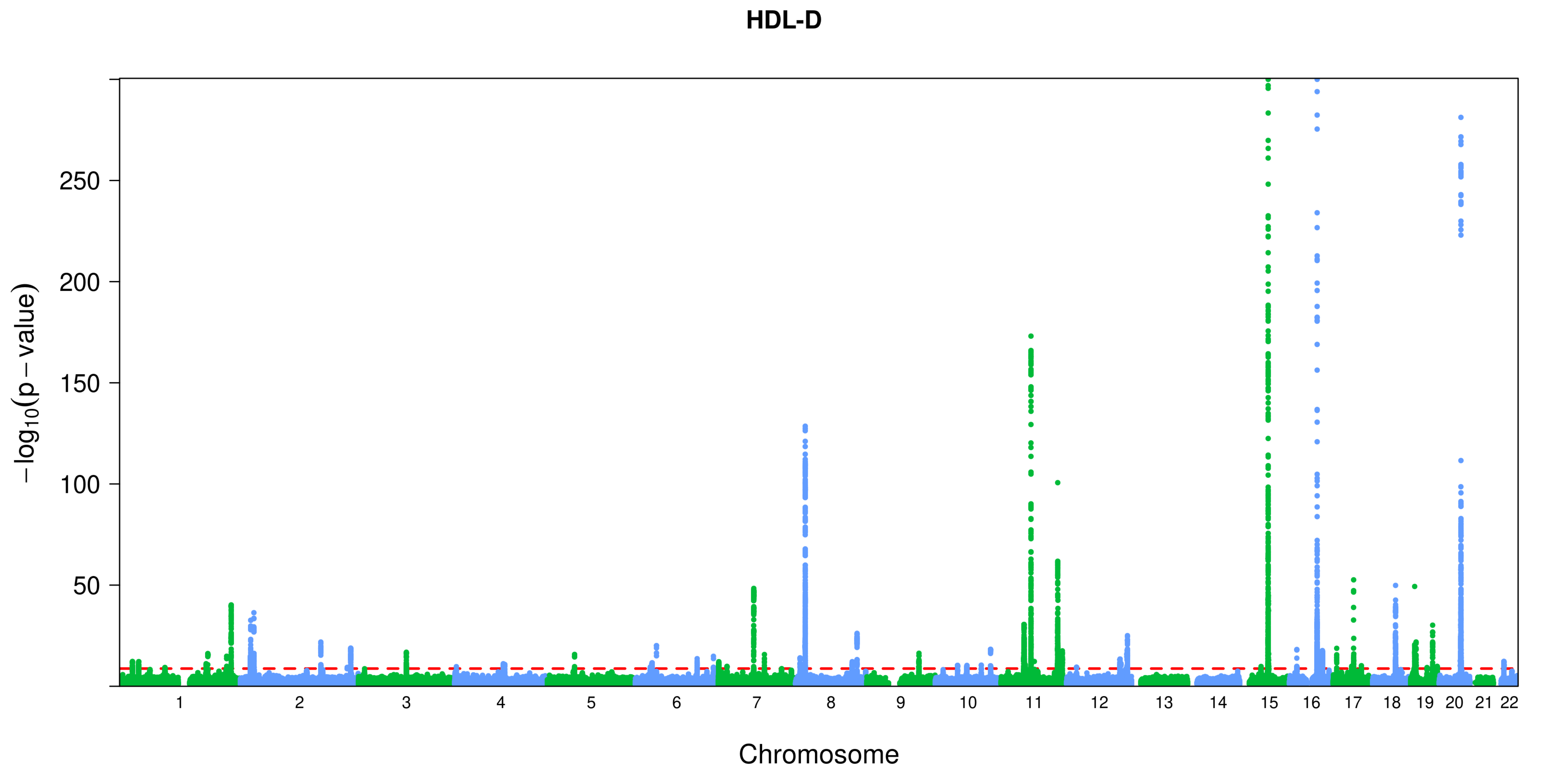
HDL-D
250
200
150
100
50
−log10(p − value)
Chromosome

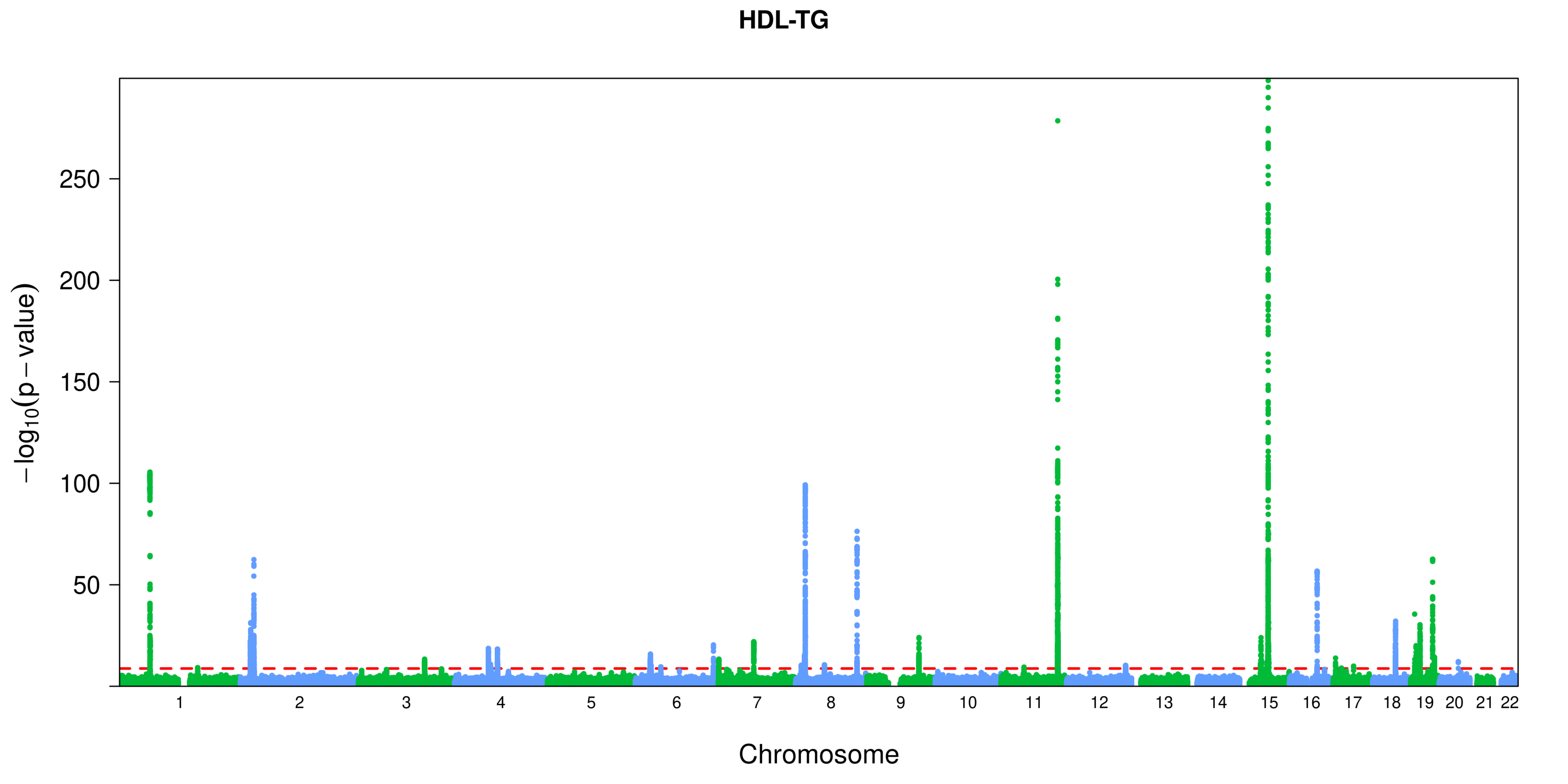
HDL-TG
250
200
150
100
50
−log10(p − value)
Chromosome

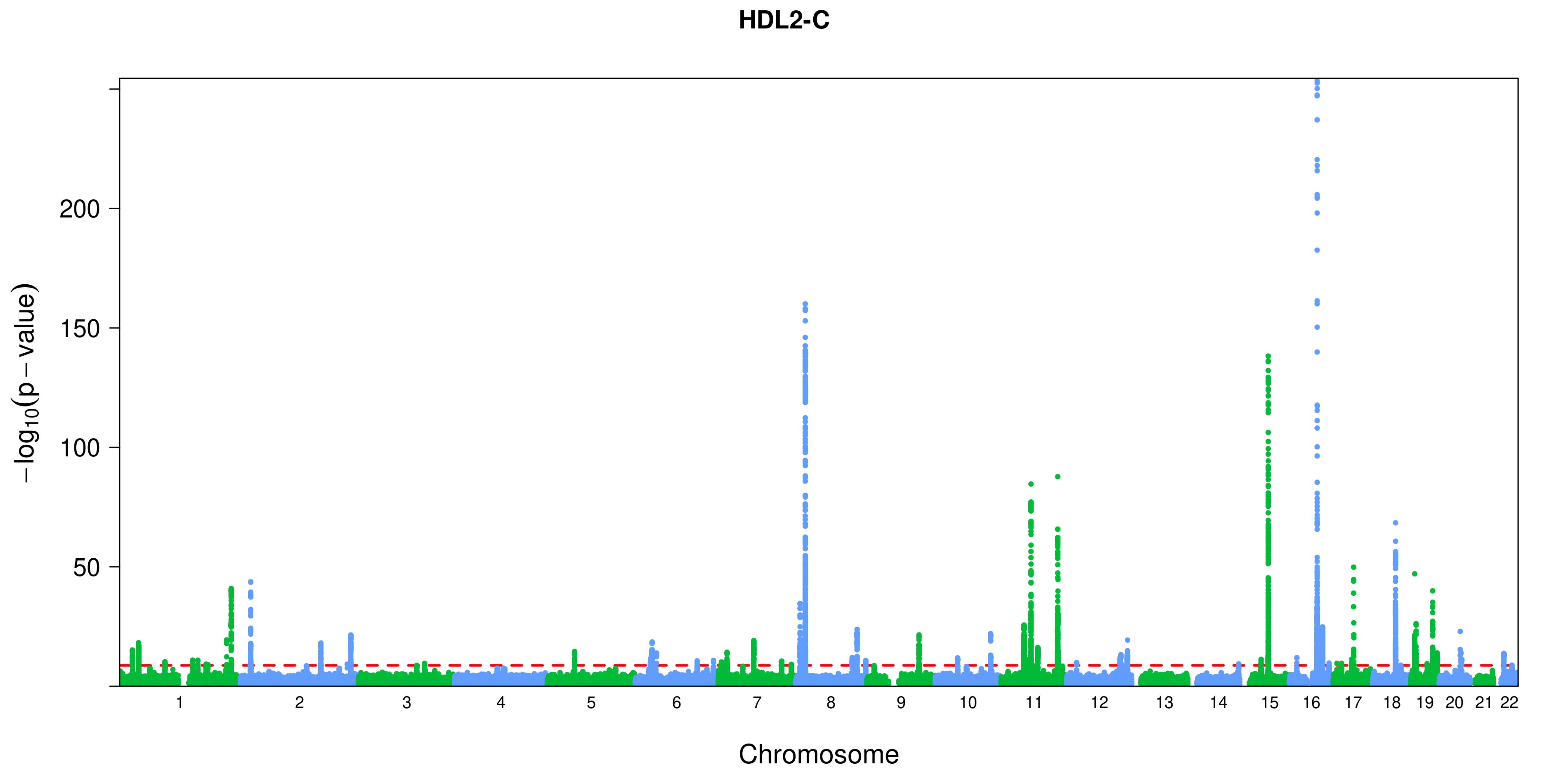

HDL2-C
−log10(p − value)
200
150
100
50
Chromosome

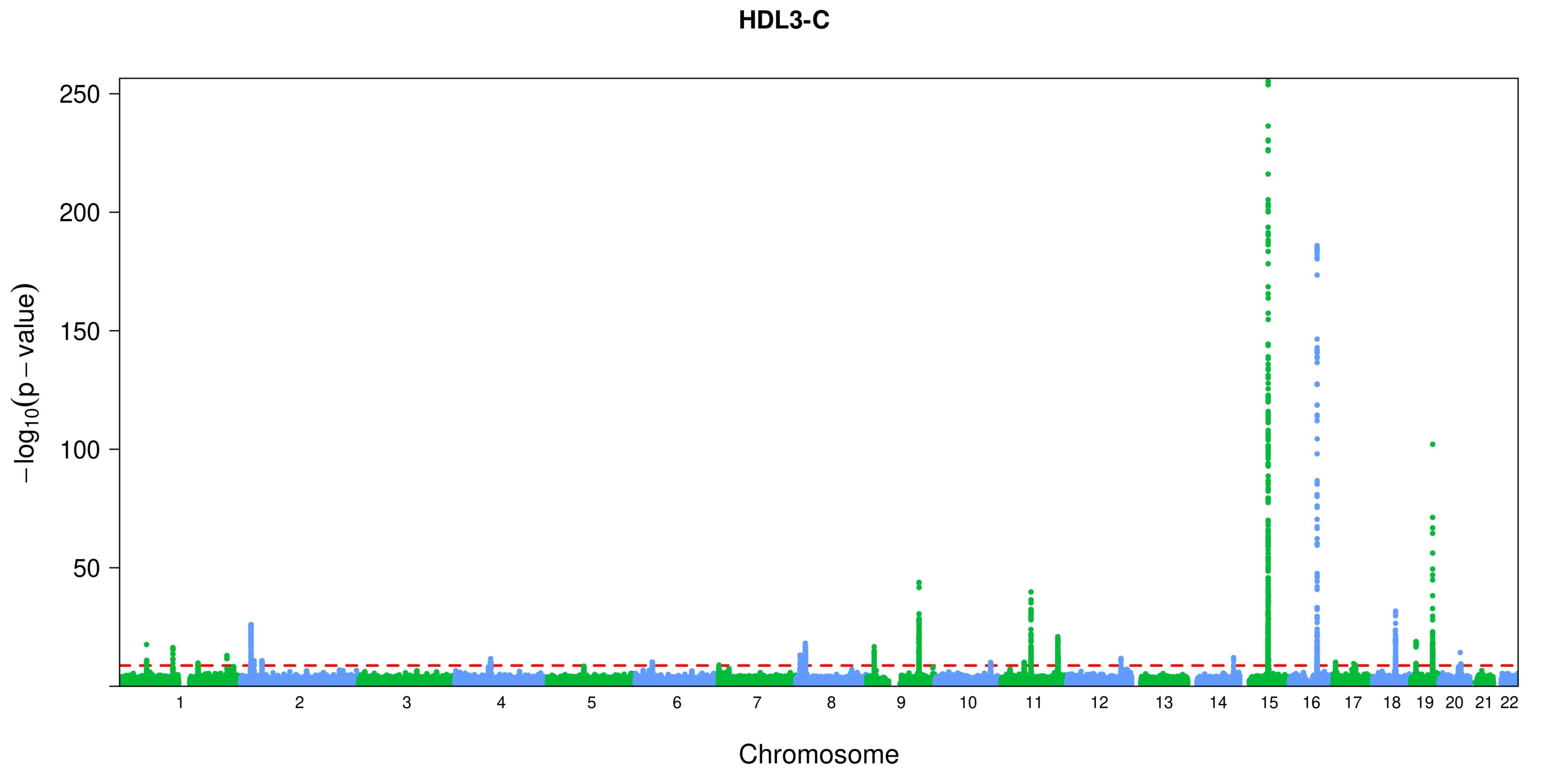
HDL3-C
250
200
150
100
50
$-\log_{10}(p-value)$
Chromosome

**His**

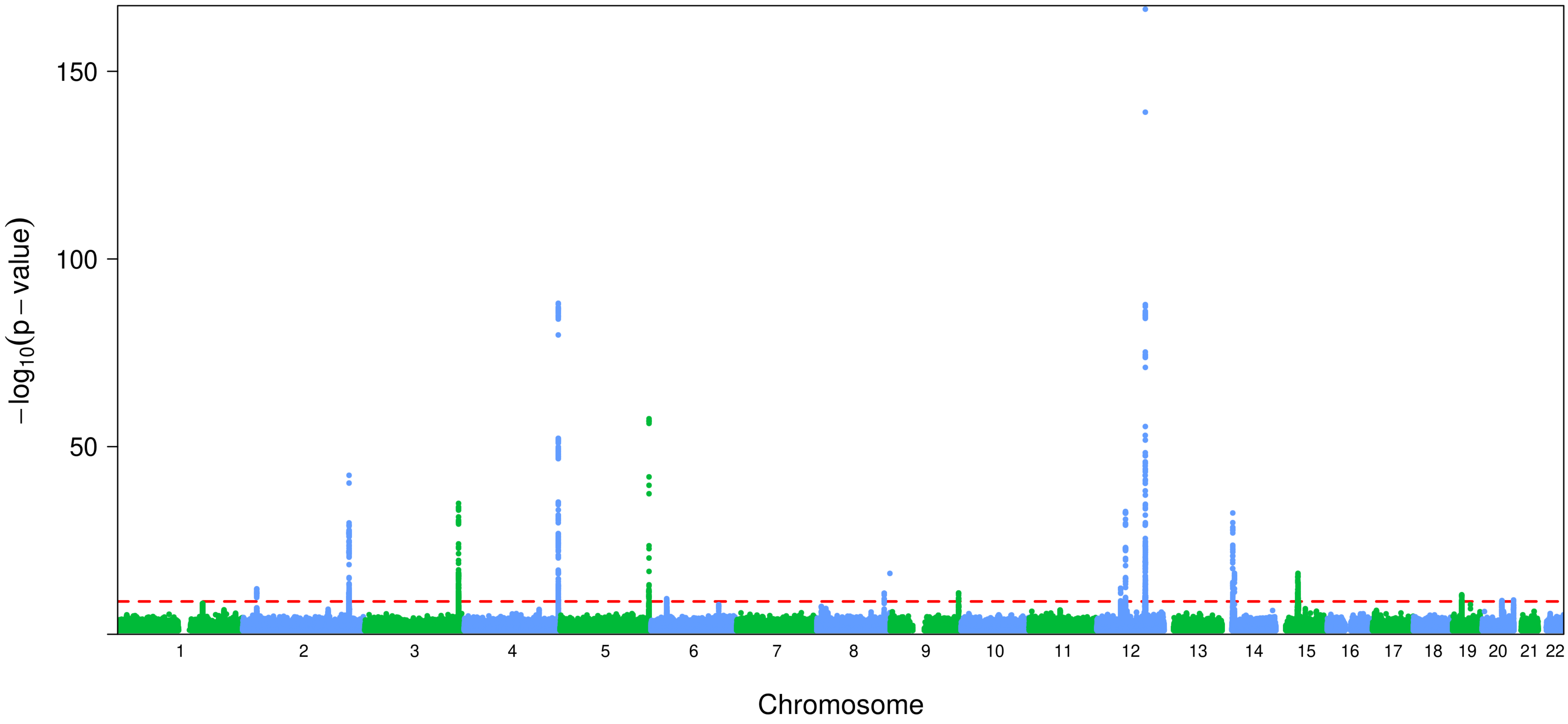
150
100
50
−log10(p − value)
Chromosome

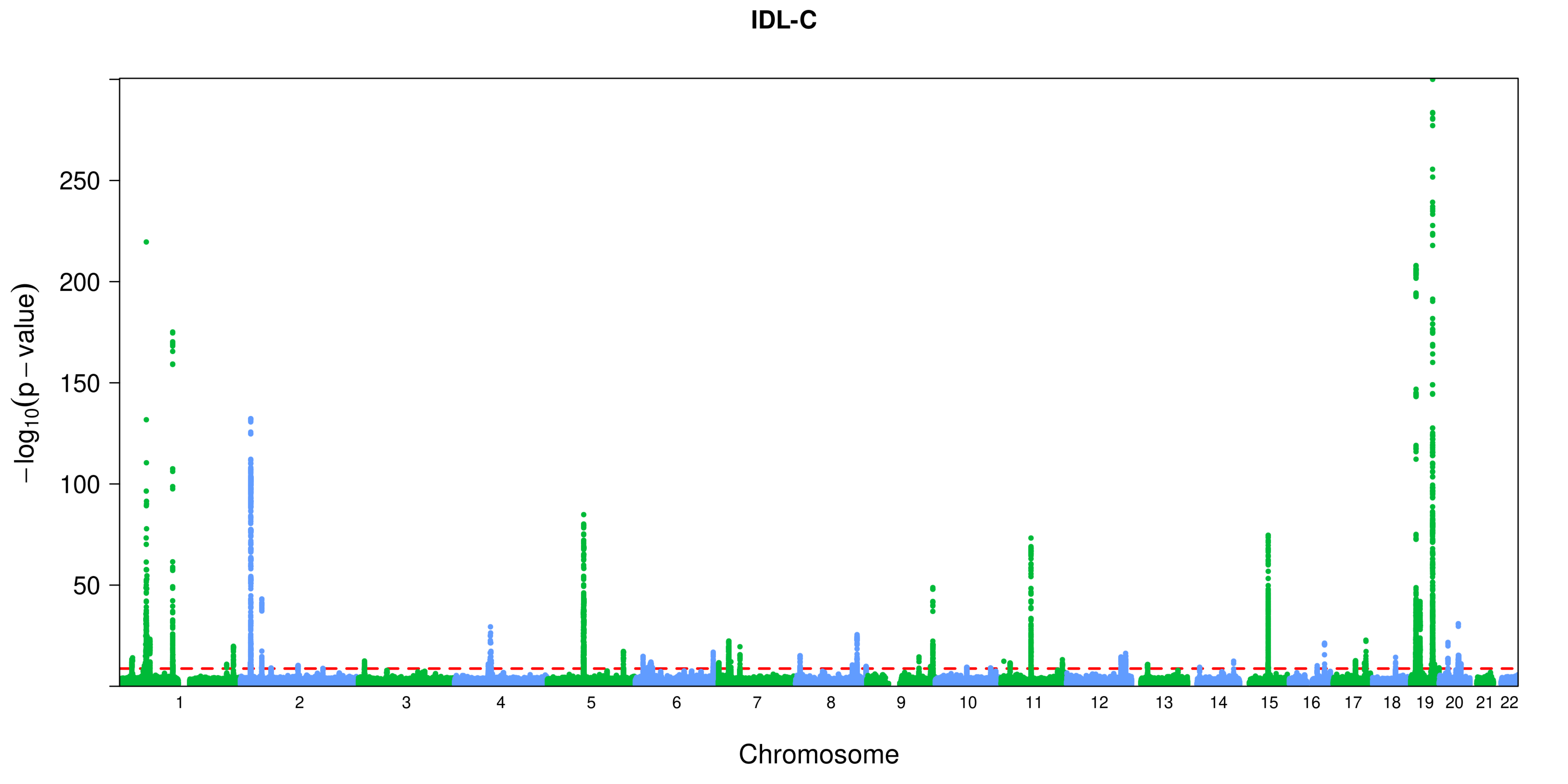

IDL-C
−log10(p − value)
250
200
150
100
50
Chromosome

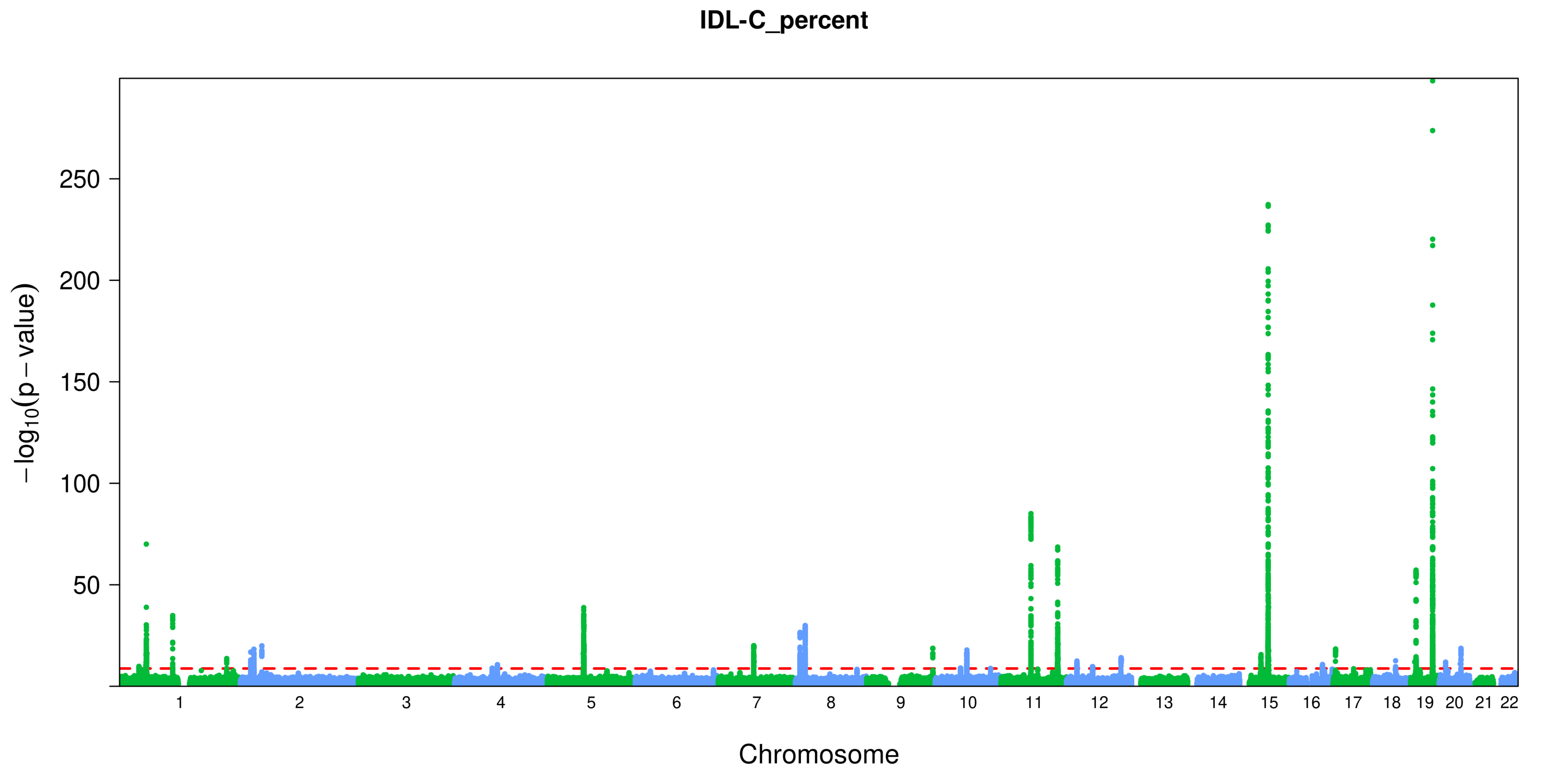
IDL-C_percent
250
200
150
100
50
−log10(p − value)
Chromosome

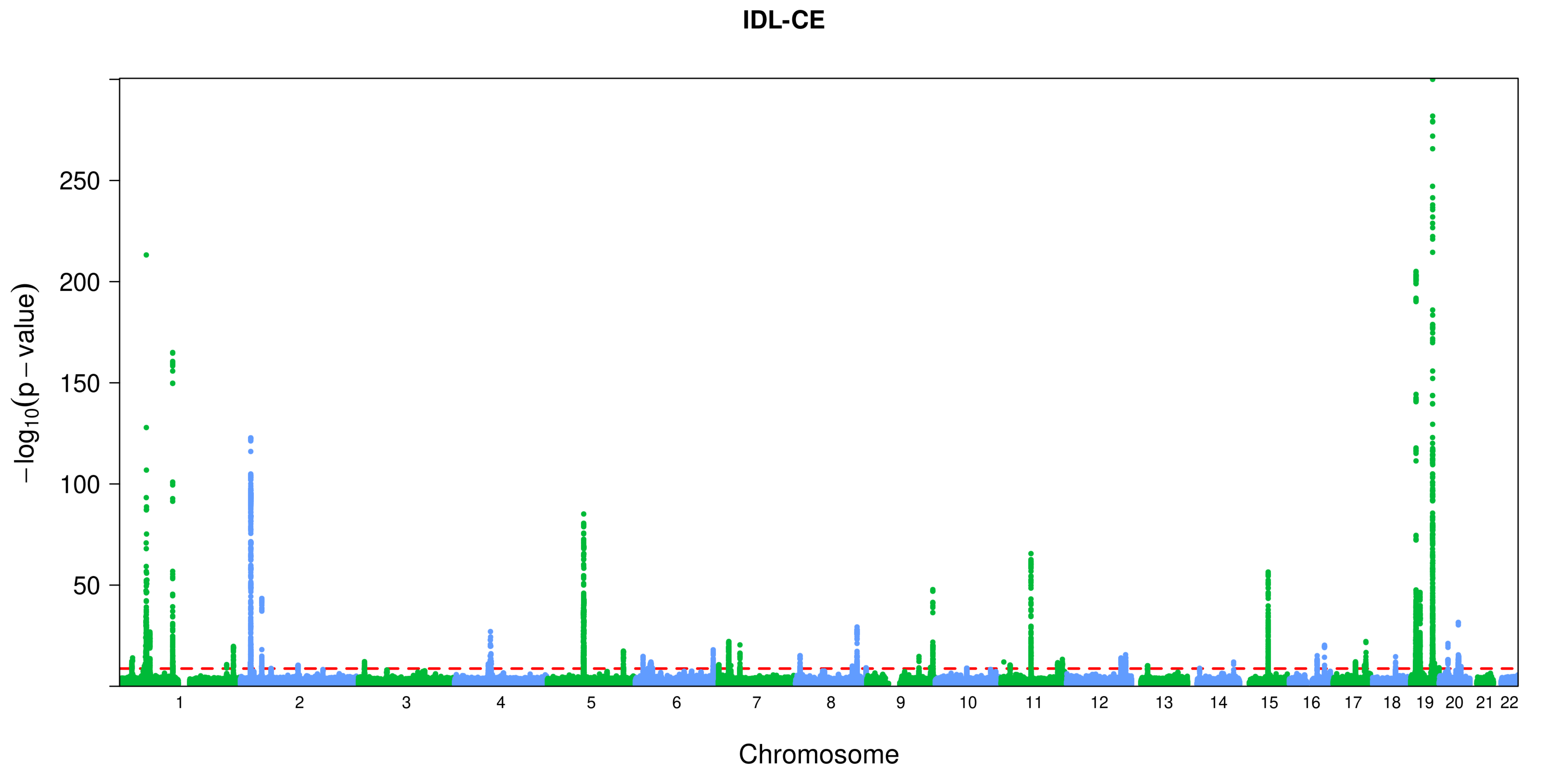
IDL-CE
−log10(p − value)
250
200
150
100
50
Chromosome

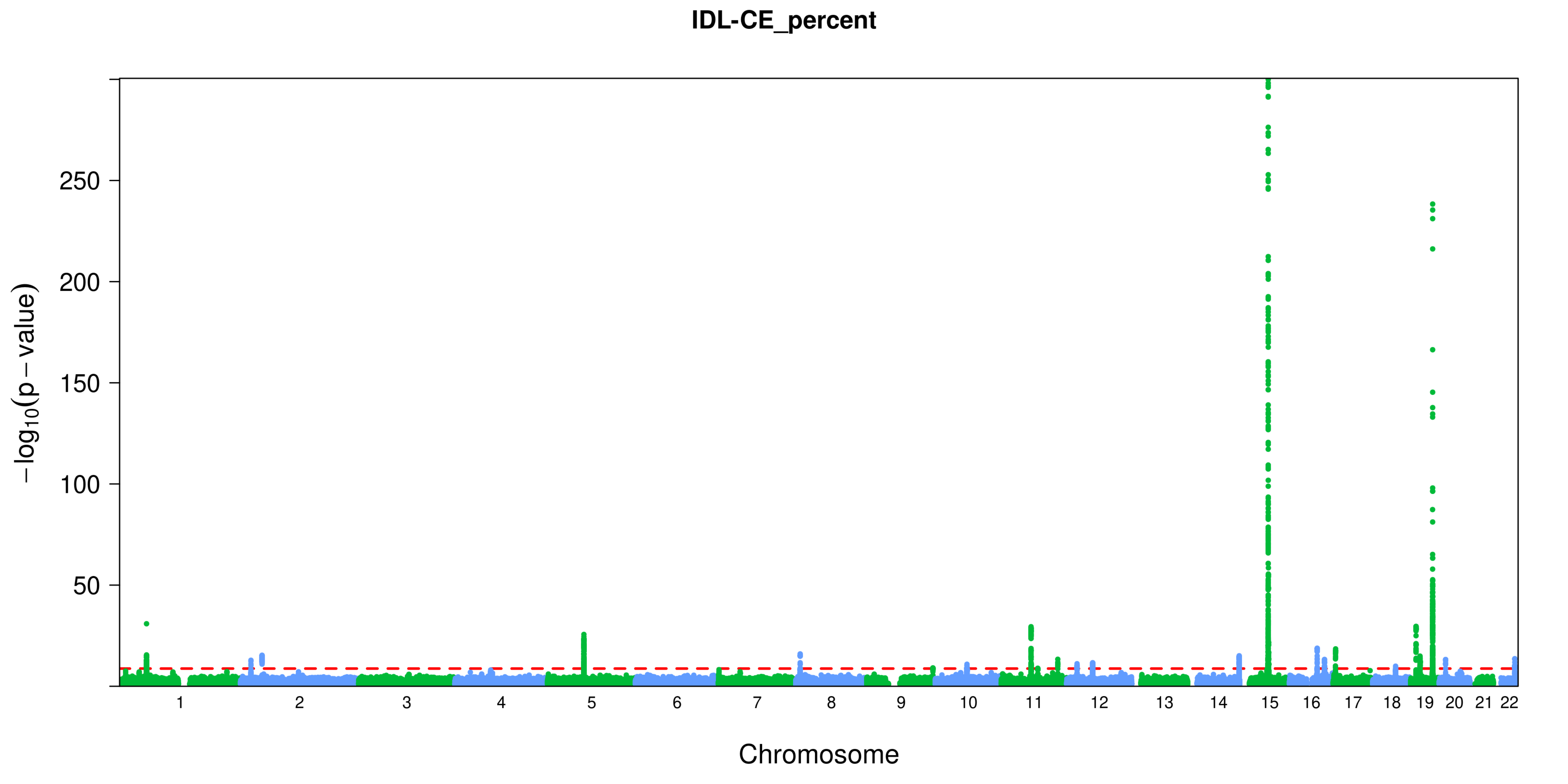
IDL-CE_percent
−log10(p − value)
250
200
150
100
50
Chromosome

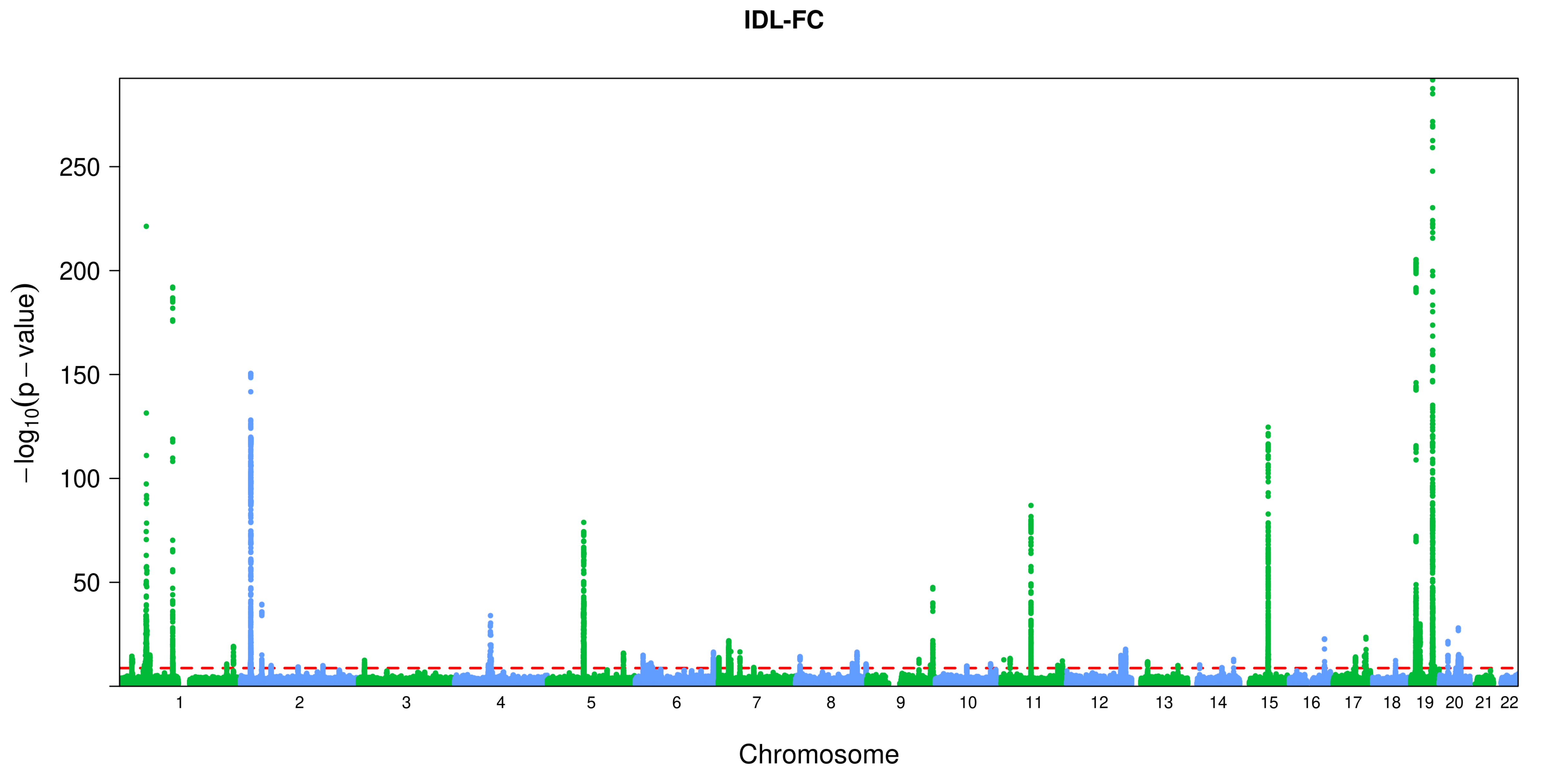
IDL-FC
250
200
150
100
50
−log10(p − value)
Chromosome

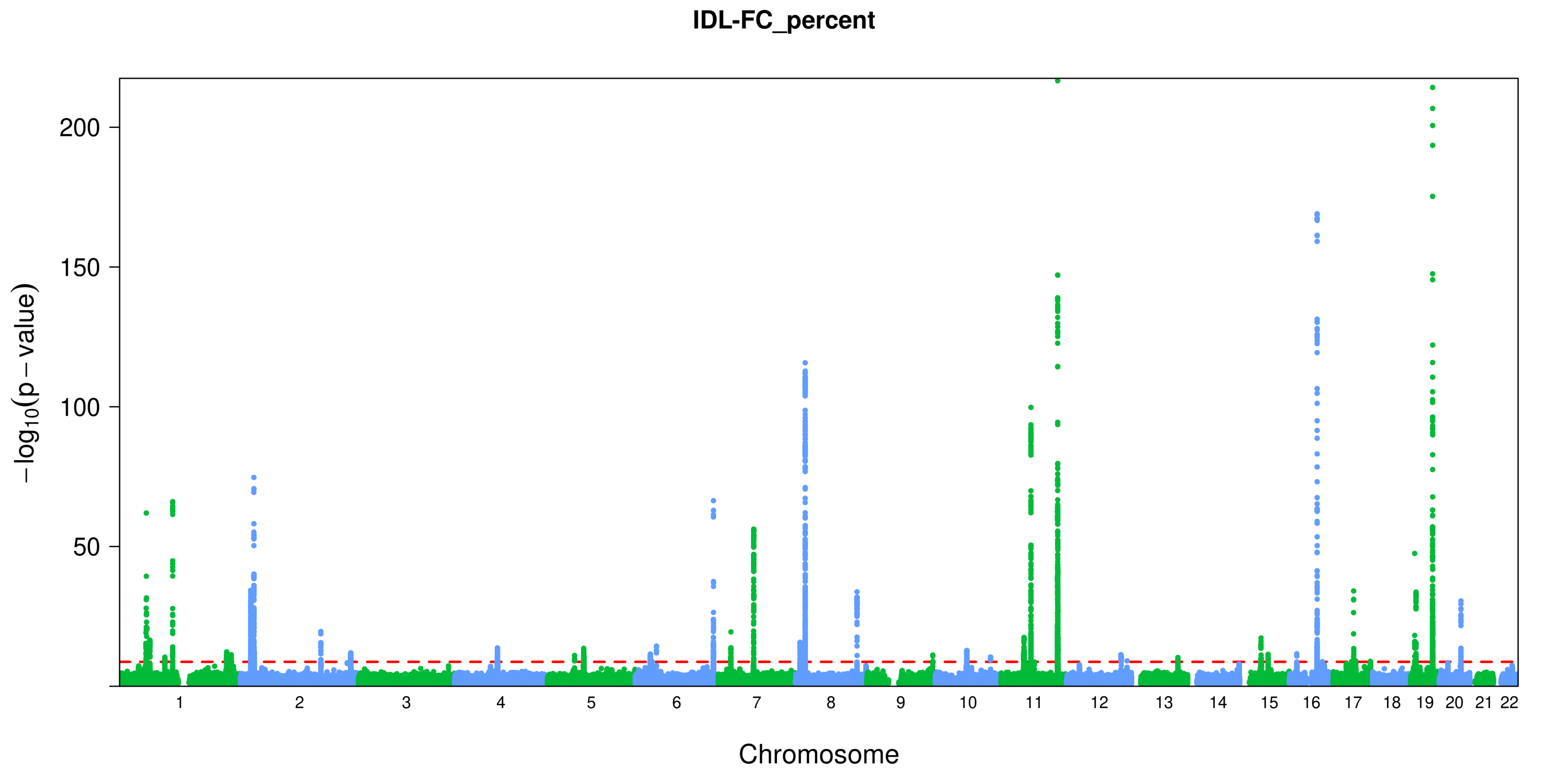
IDL-FC_percent
200
150
100
50
$-\log_{10}(p-\text{value})$
Chromosome

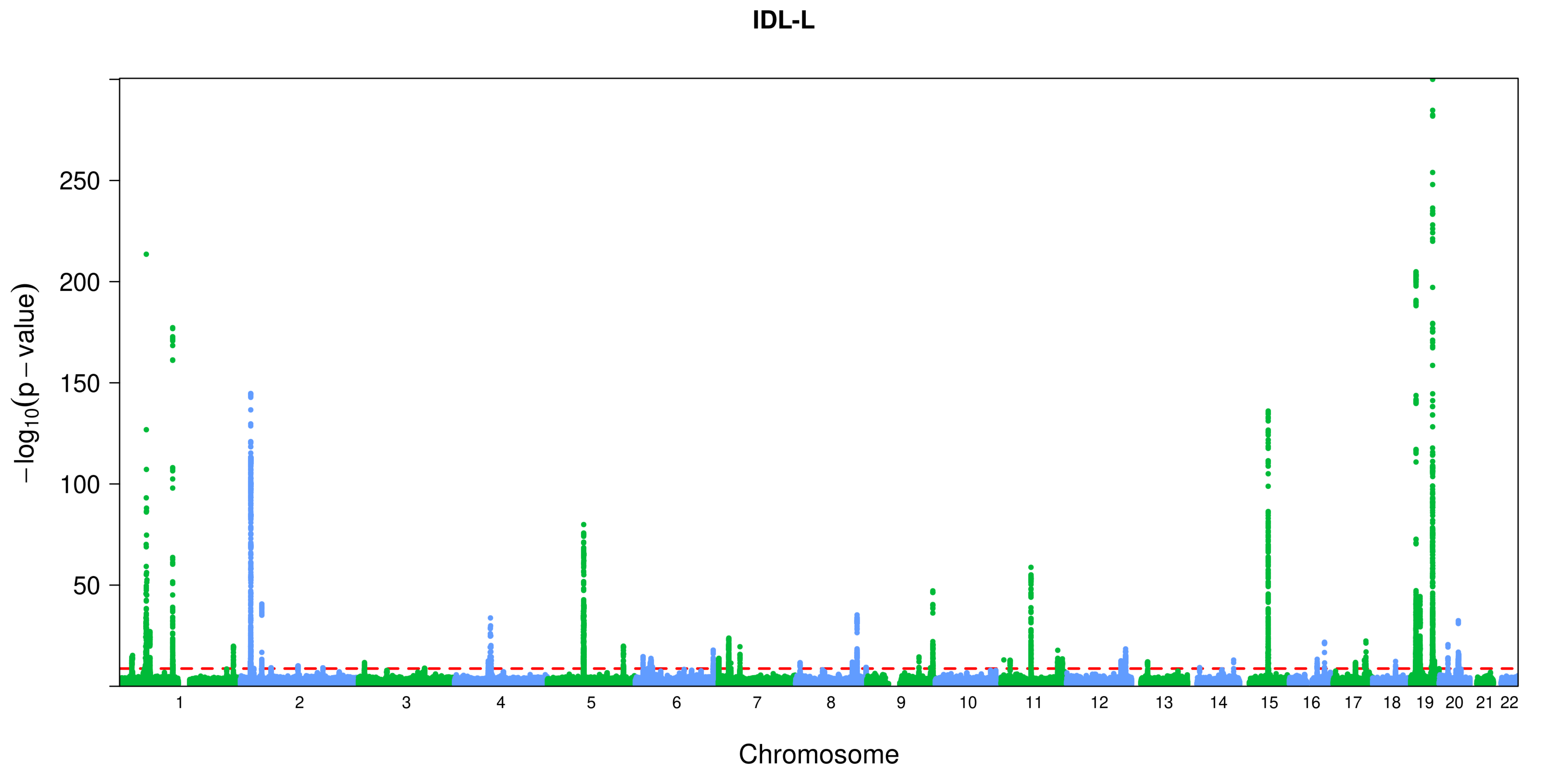
IDL-L
250
200
150
100
50
−log10(p − value)
Chromosome

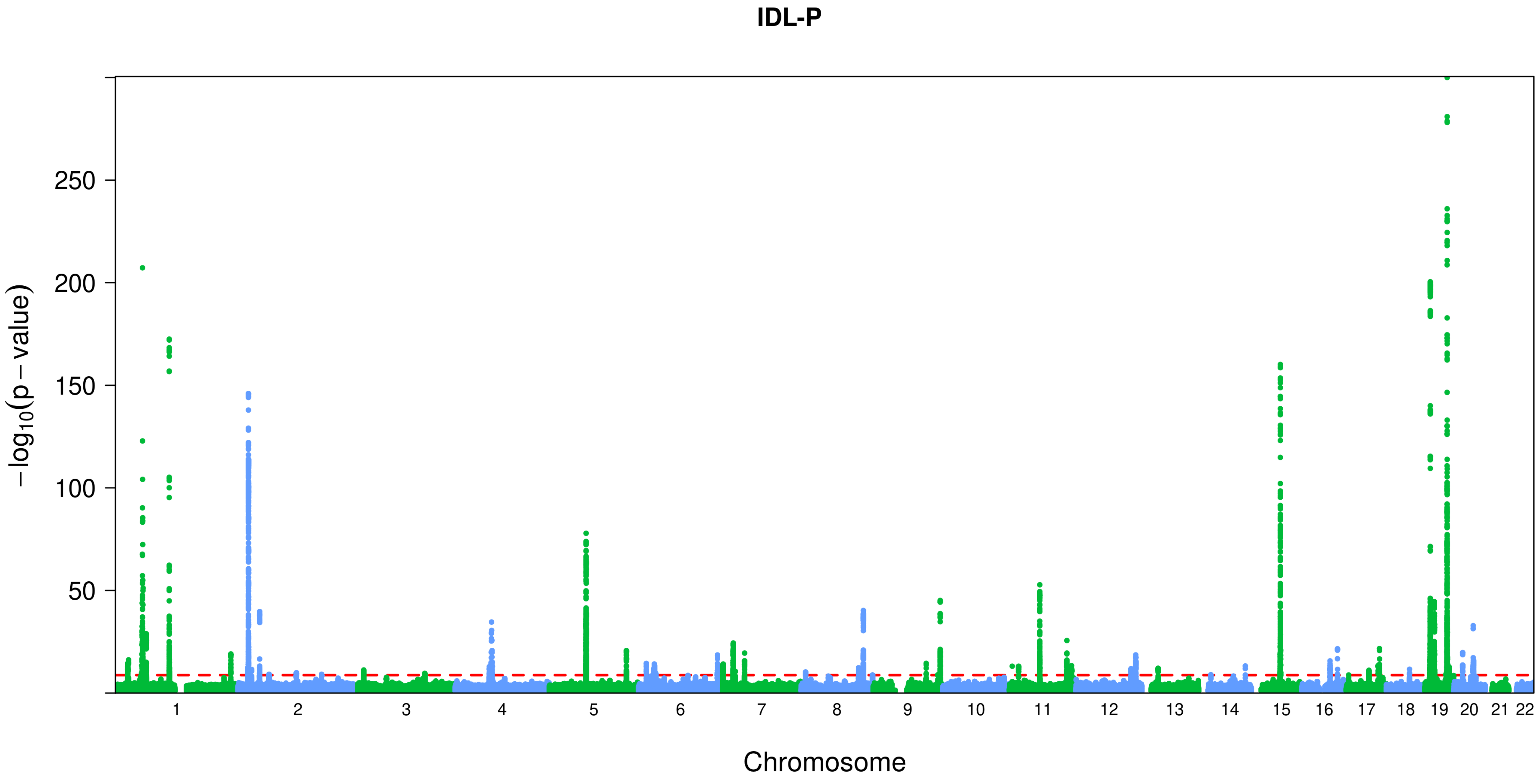

IDL-P
−log10(p − value)
250
200
150
100
50
Chromosome

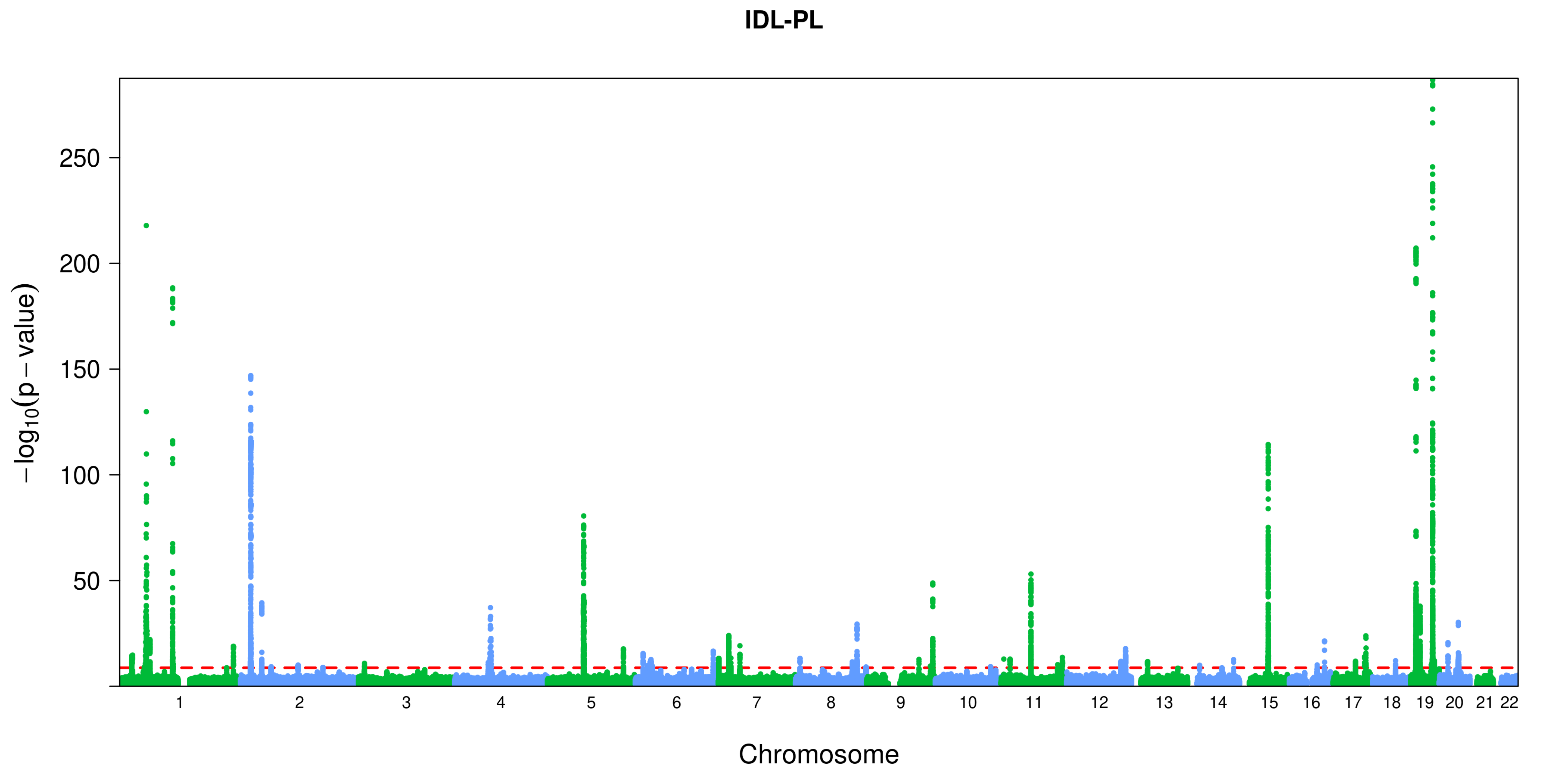
IDL-PL
−log10(p − value)
250
200
150
100
50
Chromosome

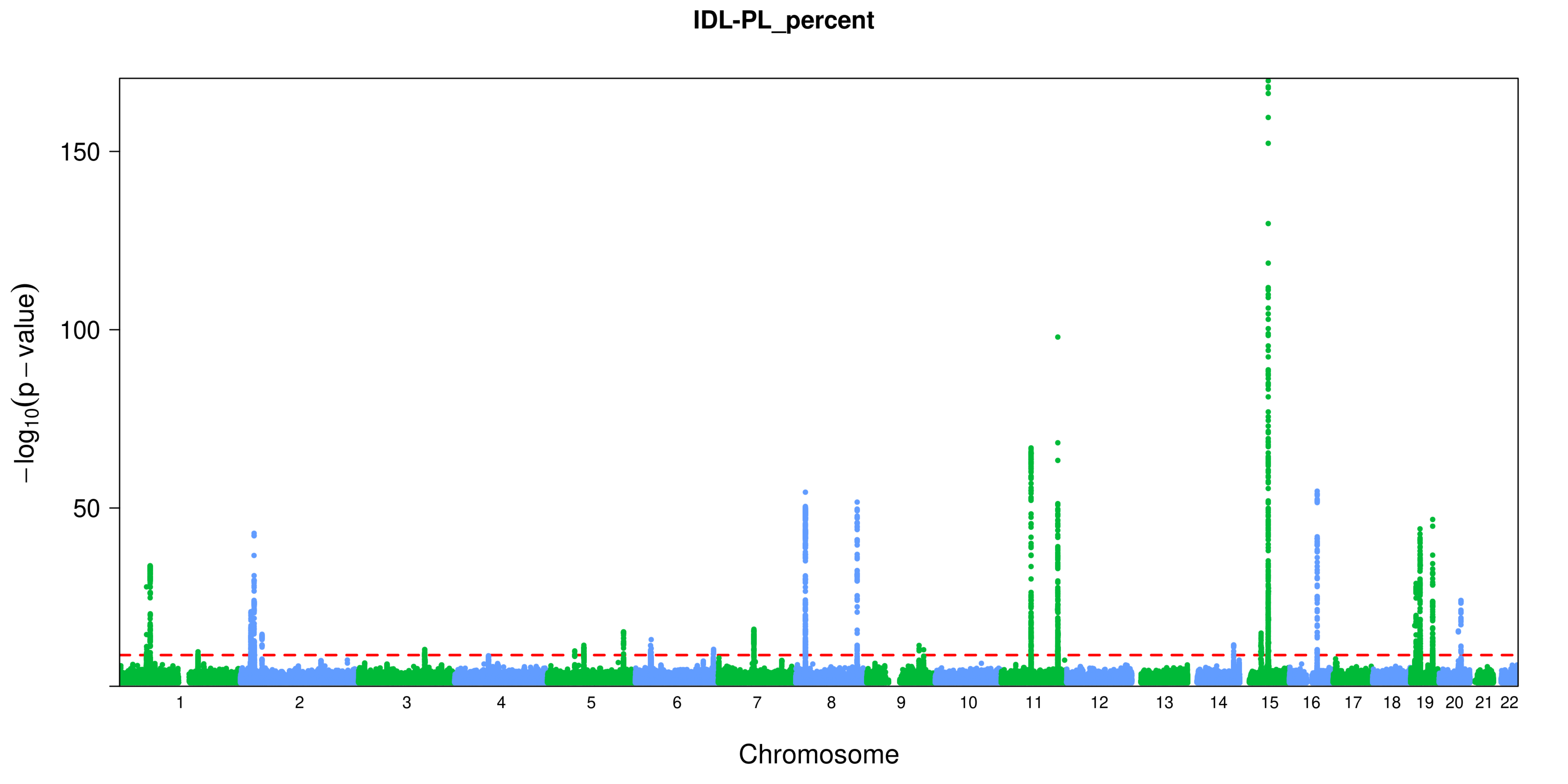

IDL-PL_percent
150
100
50
−log10(p − value)
Chromosome

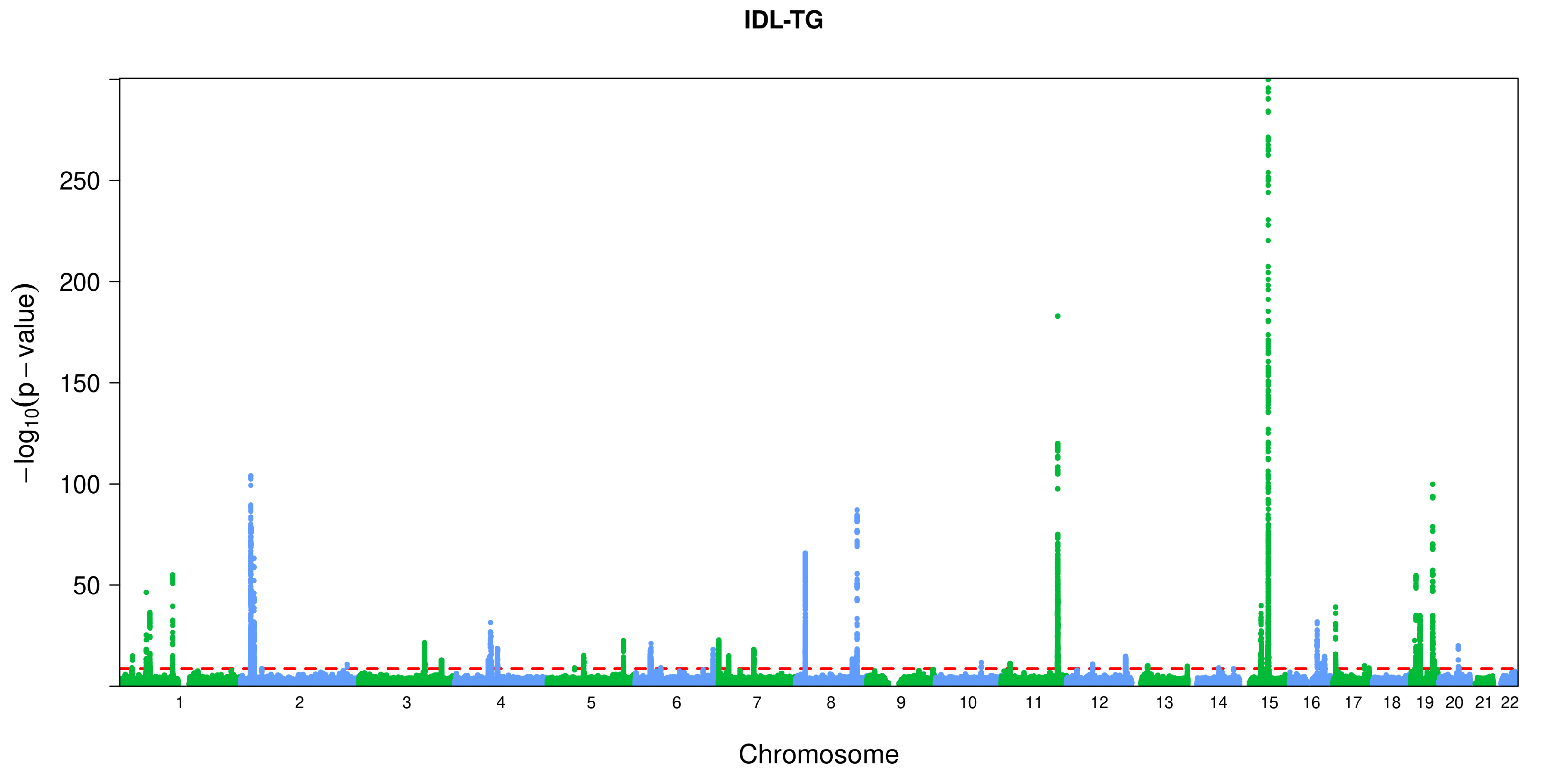
IDL-TG
−log10(p − value)
250
200
150
100
50
Chromosome

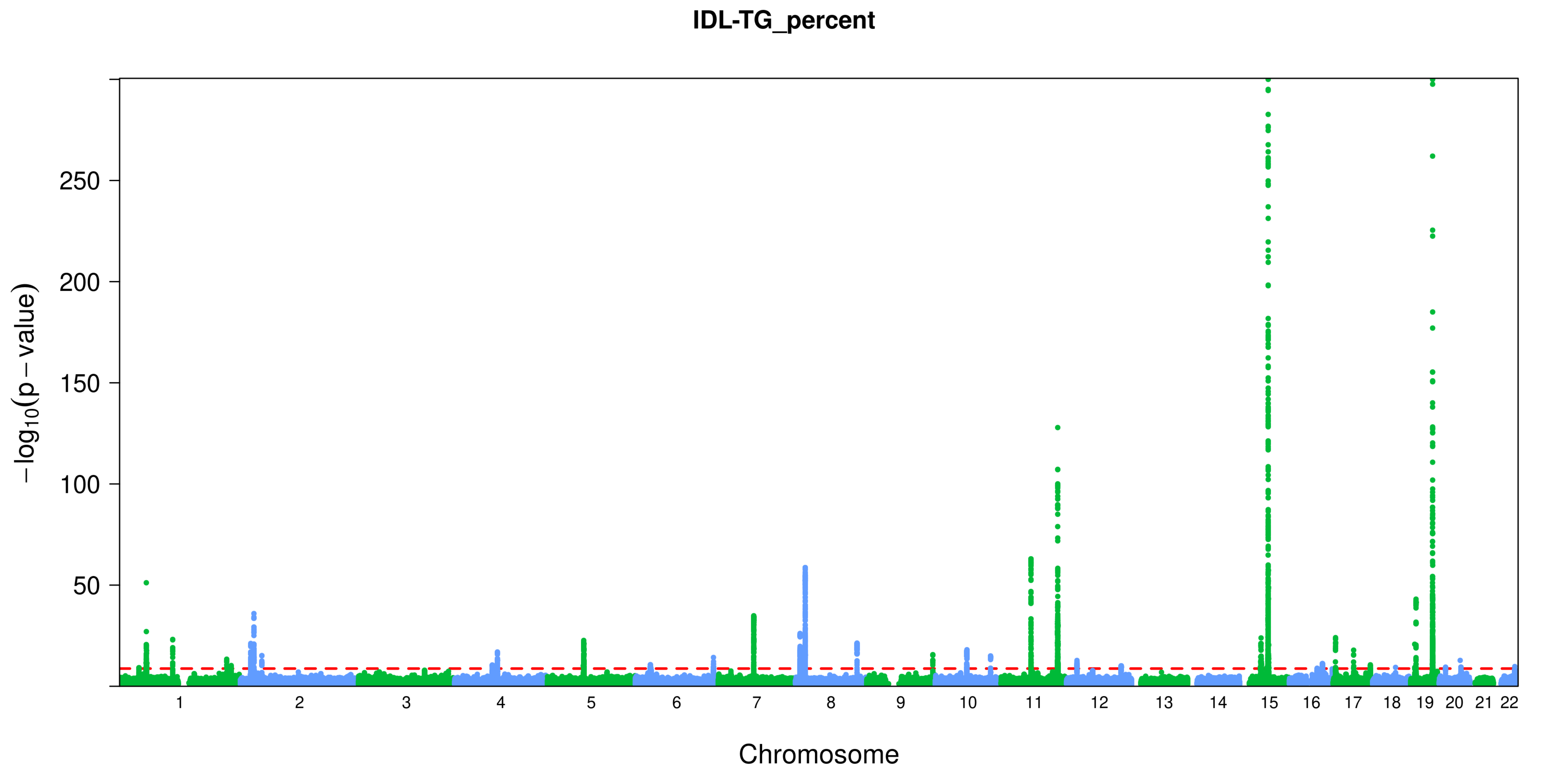
IDL-TG_percent
250
200
150
100
50
−log10(p − value)
Chromosome

### Ile

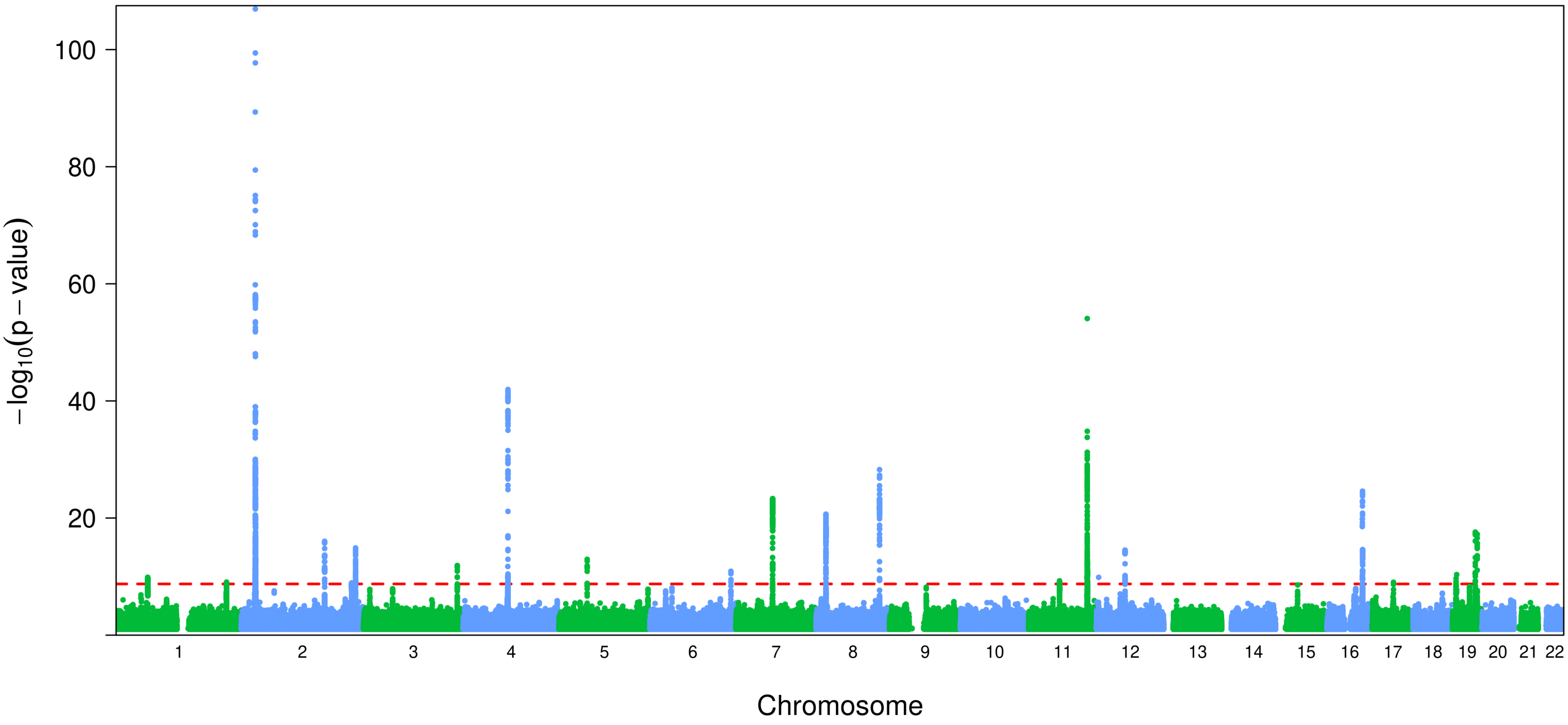

−log10(p − value)
100
80
60
40
20
Chromosome

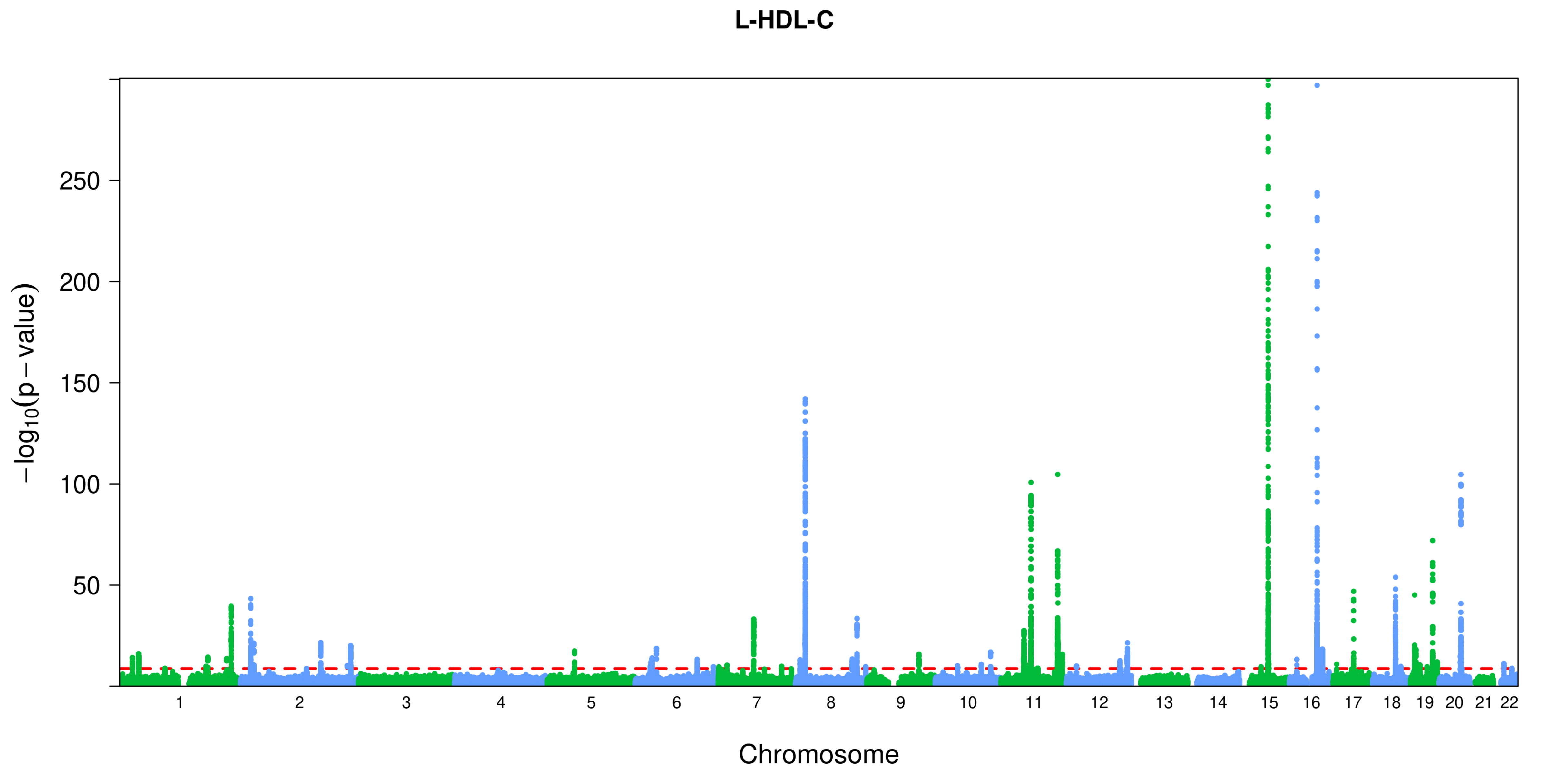

L-HDL-C
250
200
150
100
50
−log10(p − value)
Chromosome

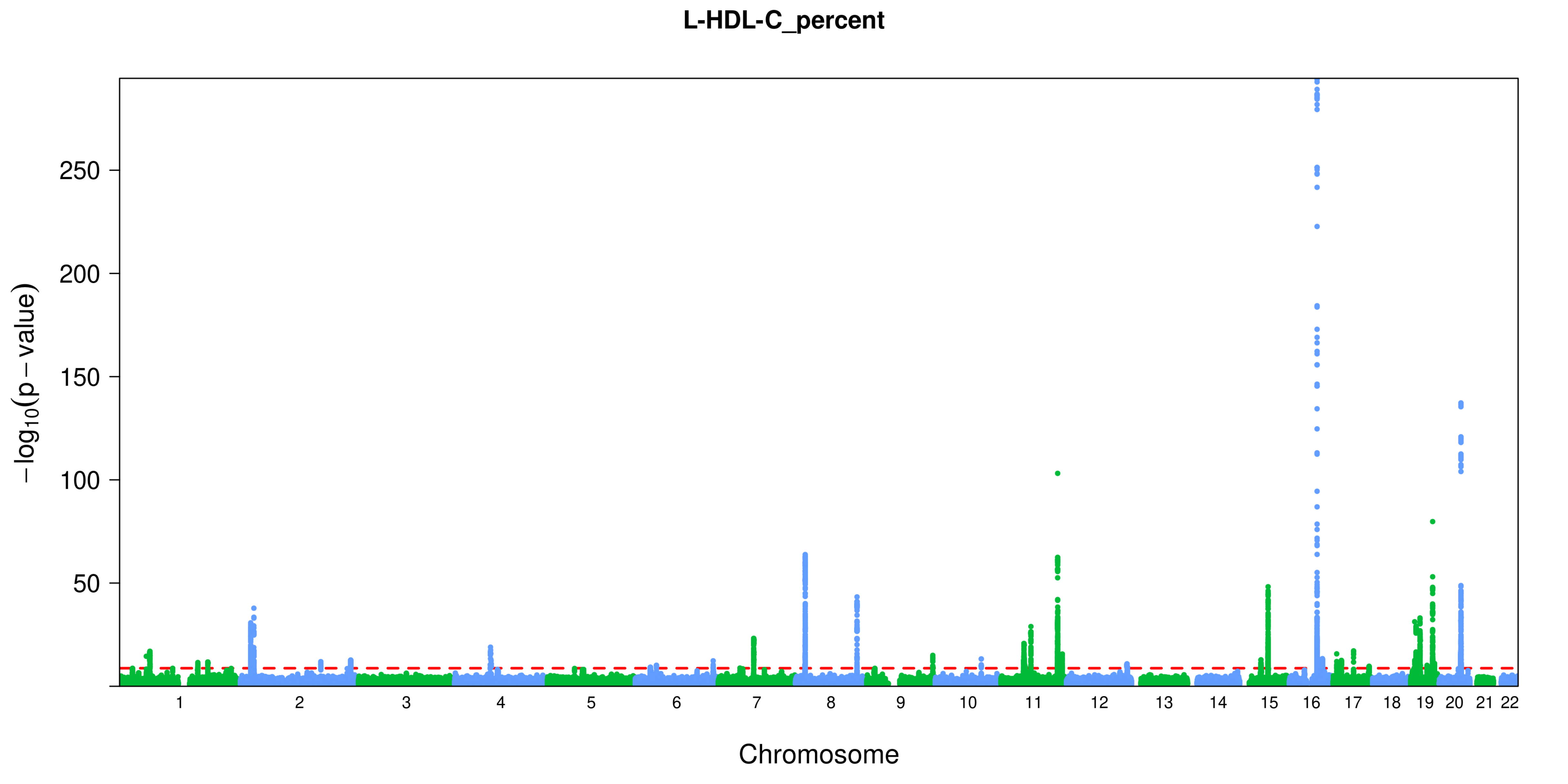

L-HDL-C_percent
−log10(p − value)
250
200
150
100
50
Chromosome

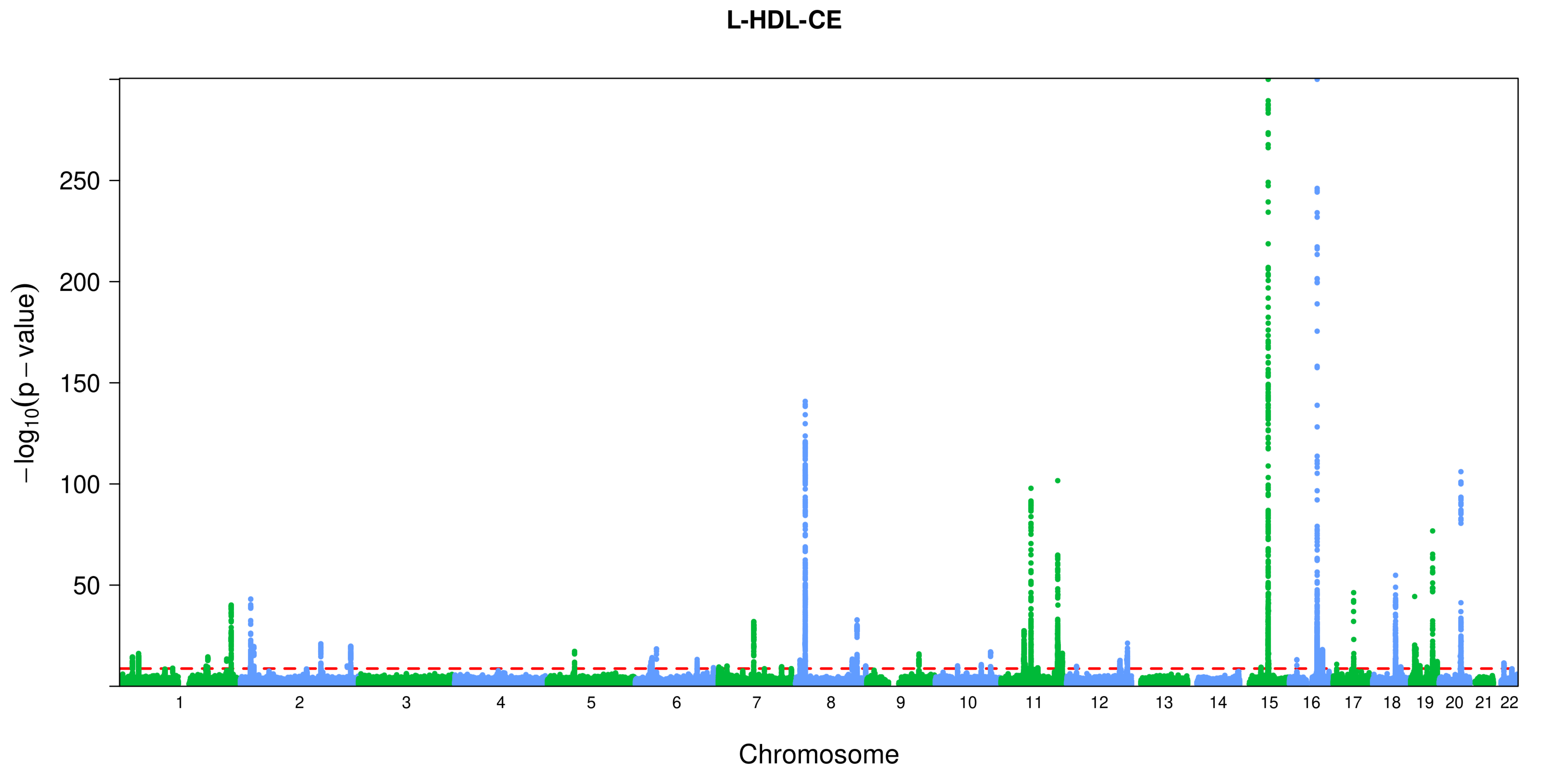

L-HDL-CE
250
200
150
100
50
−log10(p − value)
Chromosome

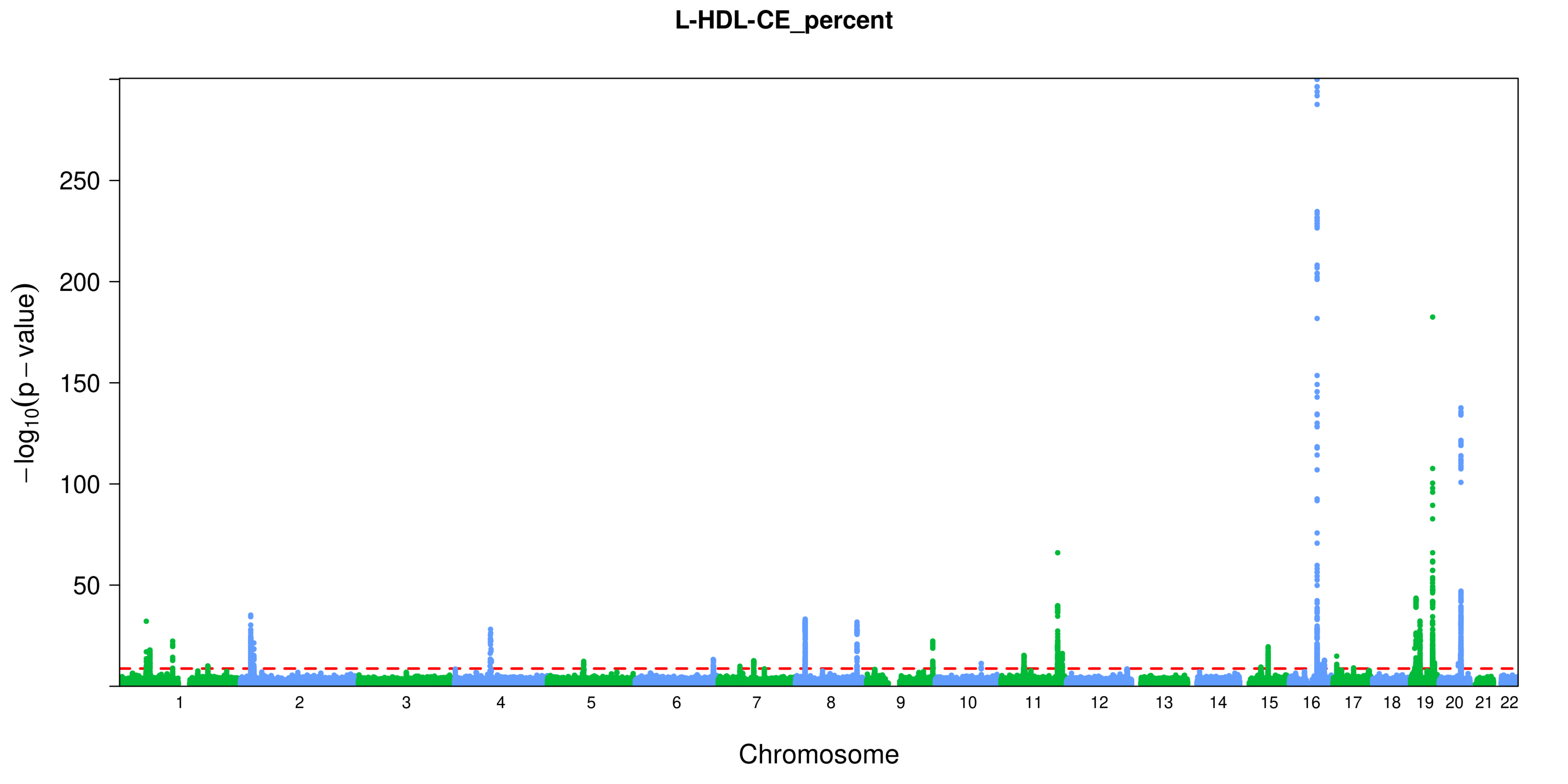

L-HDL-CE_percent
−log10(p − value)
250
200
150
100
50
Chromosome

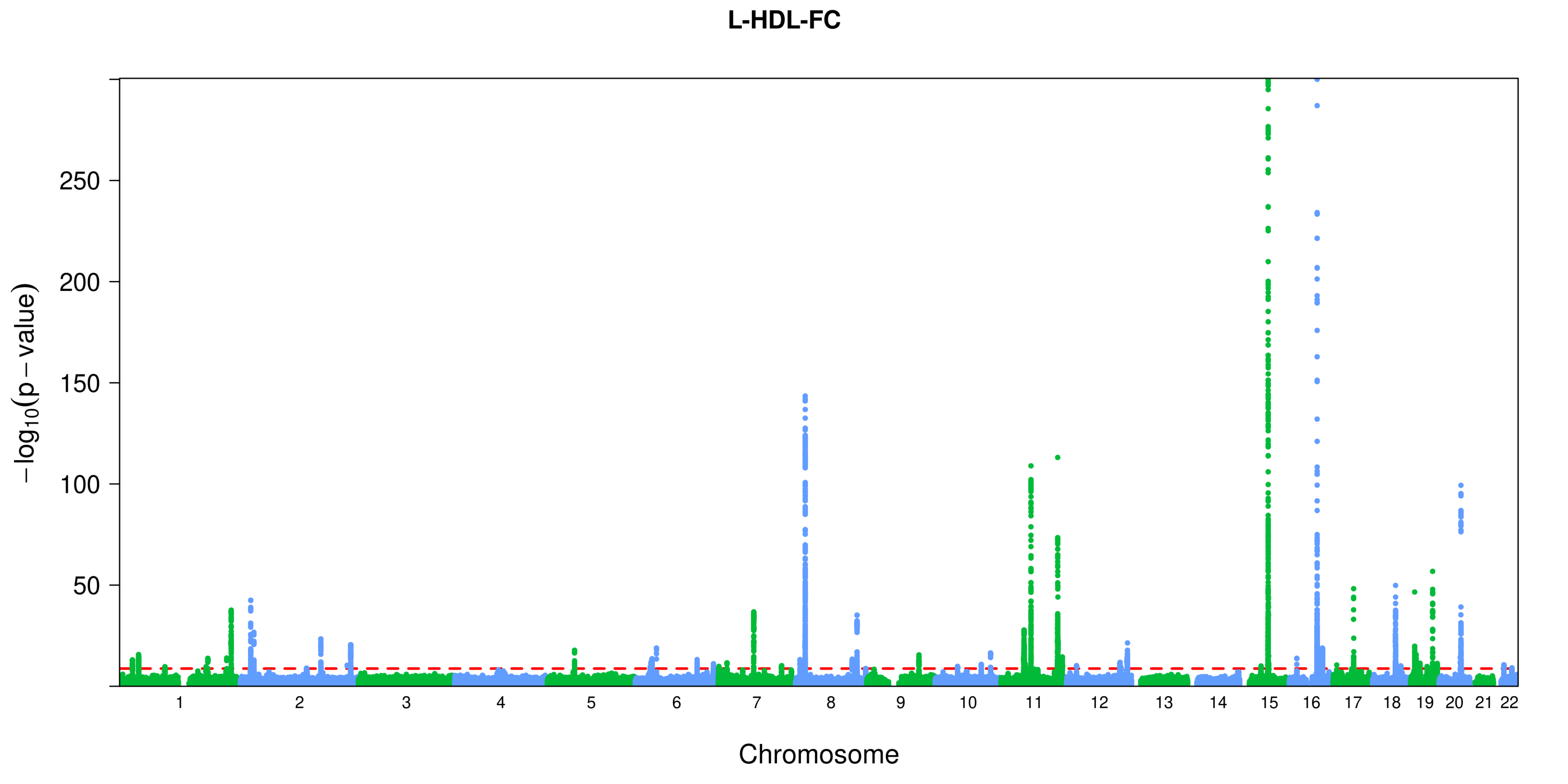
L-HDL-FC
250
200
150
100
50
$-\log_{10}(p-\text{value})$
Chromosome

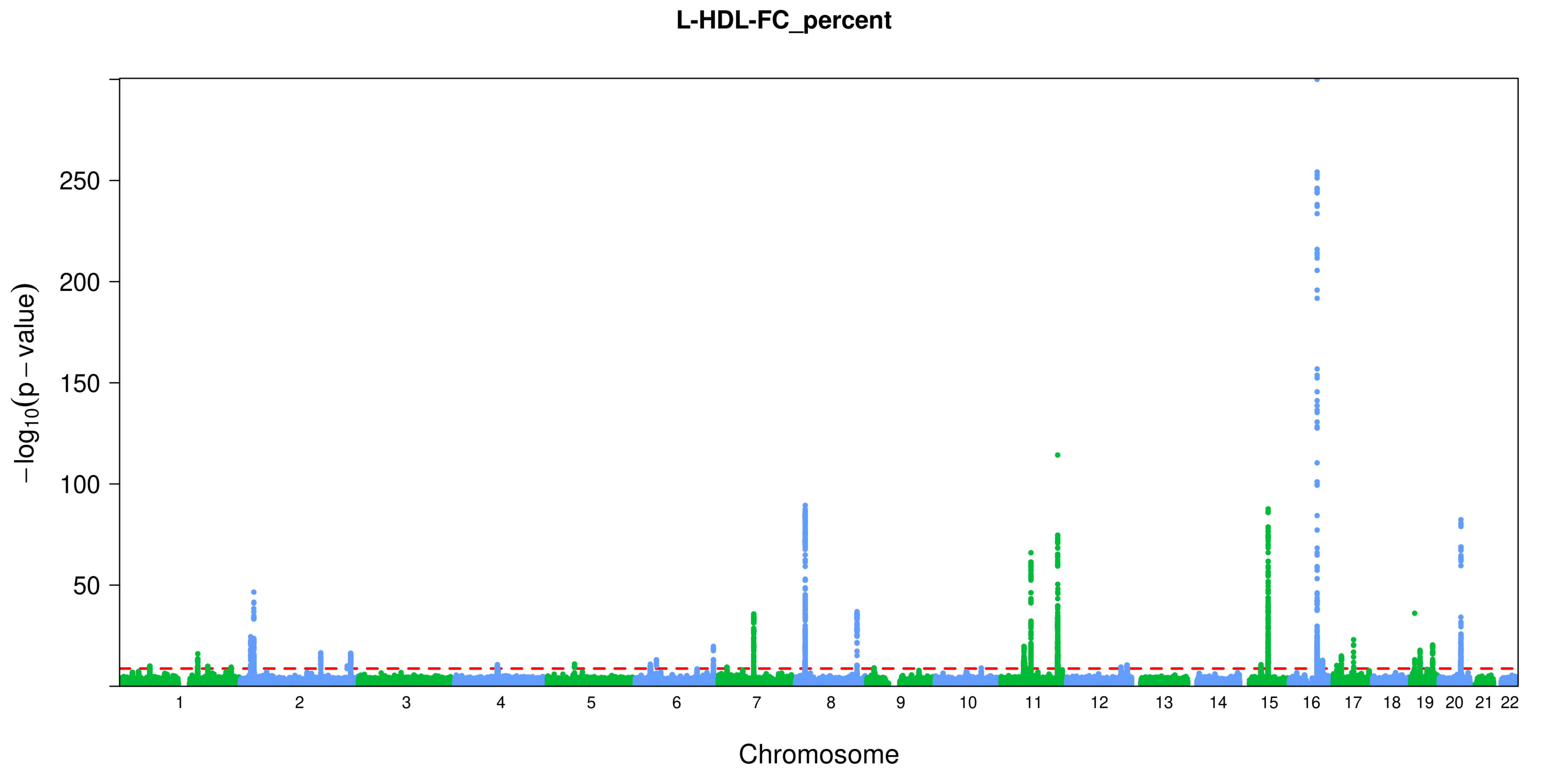
L-HDL-FC_percent
−log10(p − value)
250
200
150
100
50
Chromosome

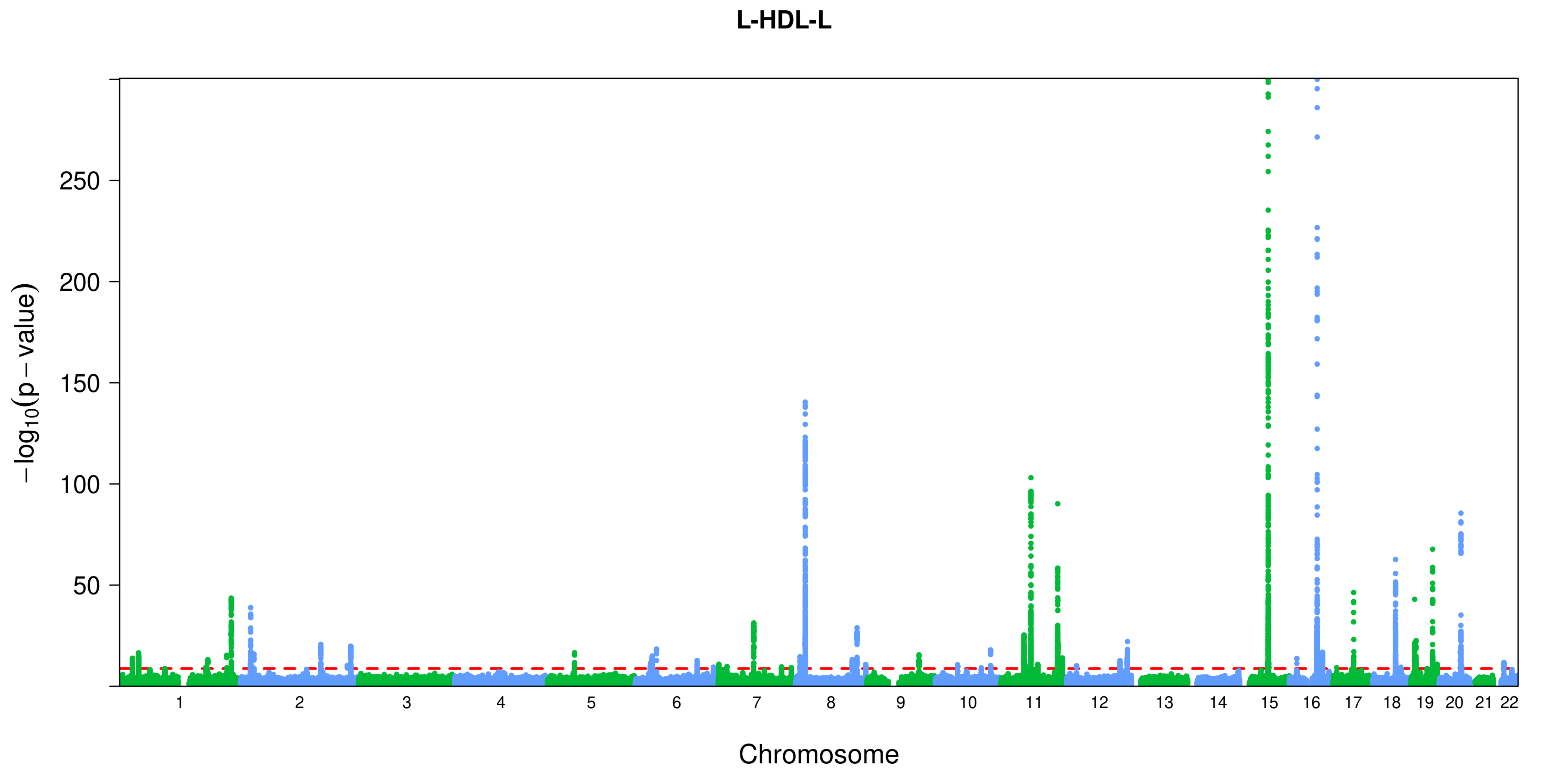
L-HDL-L
250
200
150
100
50
$-\log_{10}(p-\text{value})$
Chromosome

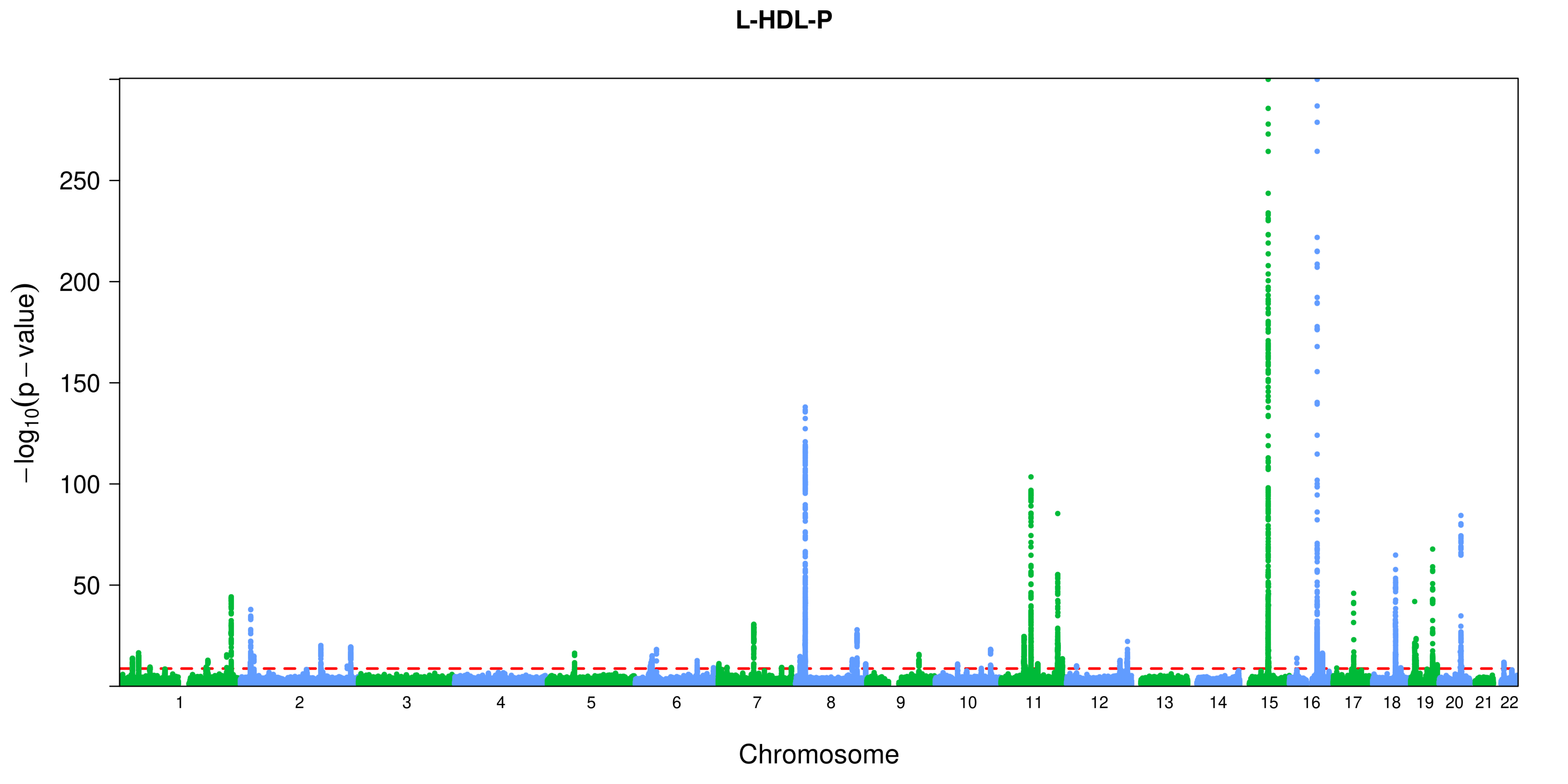
L-HDL-P
$-\log_{10}(p-\text{value})$
250
200
150
100
50
1 2 3 4 5 6 7 8 9 10 11 12 13 14 15 16 17 18 19 20 21 22
Chromosome

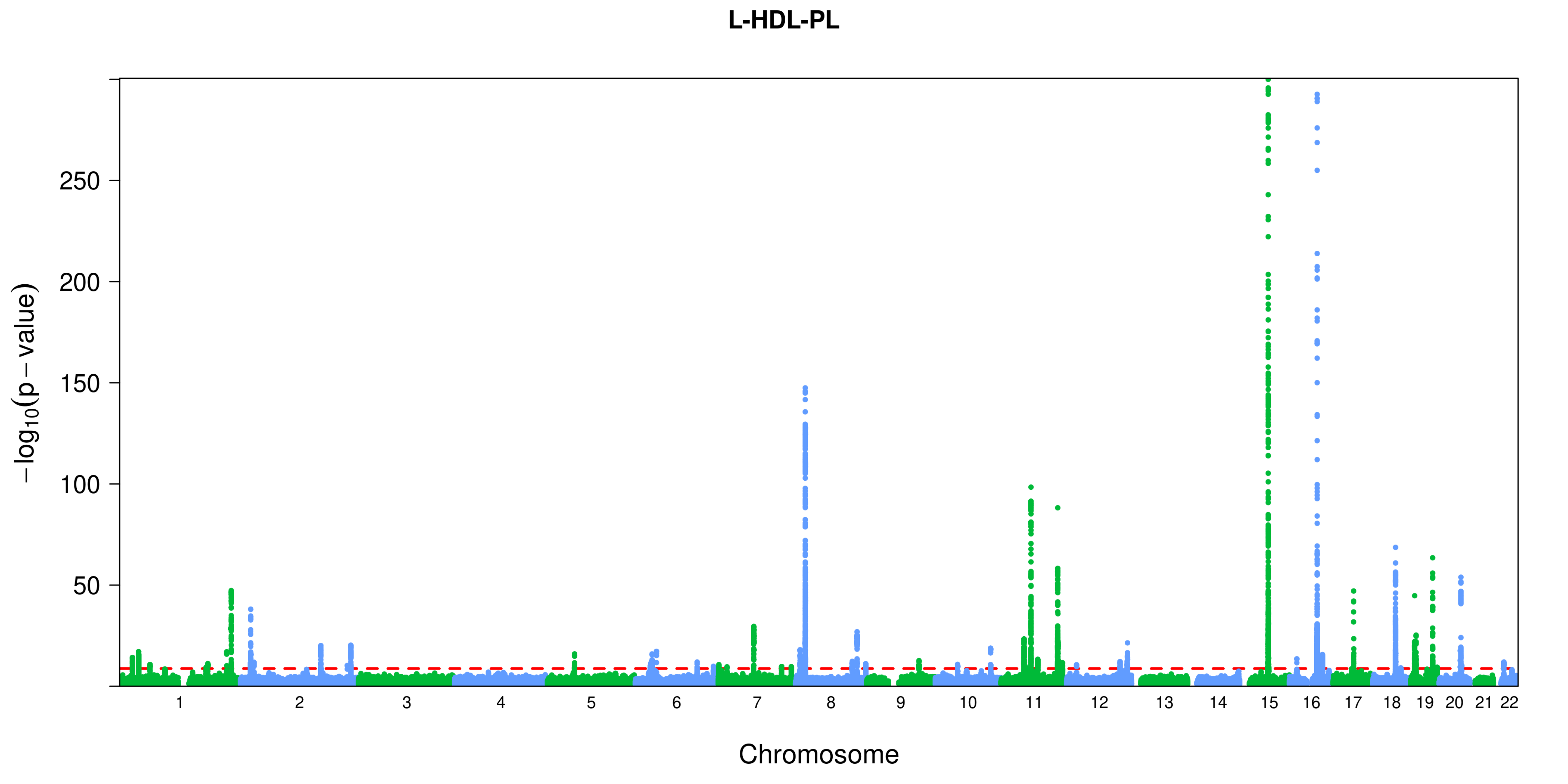

L-HDL-PL
250
200
150
100
50
$-\log_{10}(p-\text{value})$
Chromosome

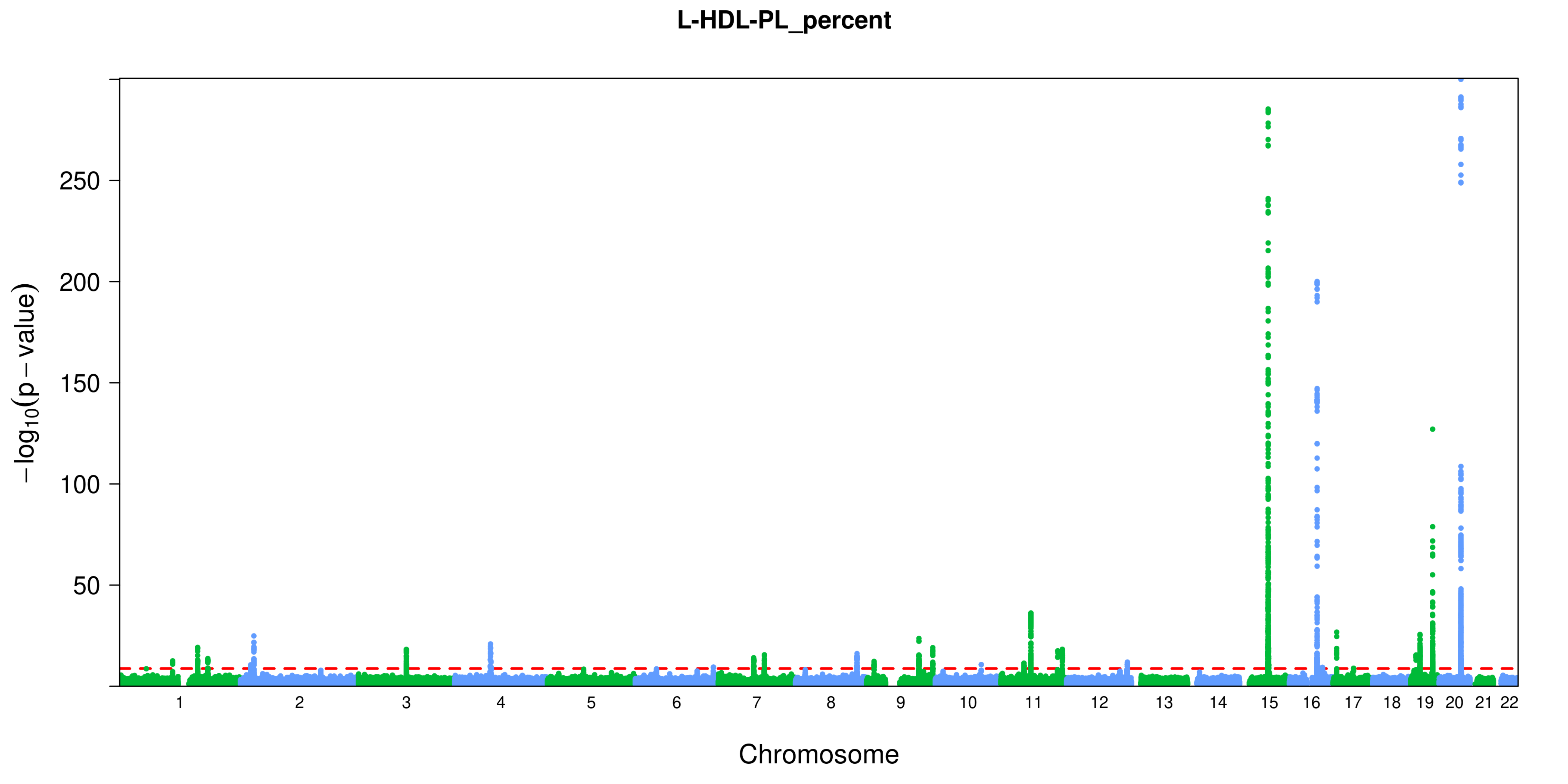

L-HDL-PL_percent
−log10(p − value)
250
200
150
100
50
Chromosome

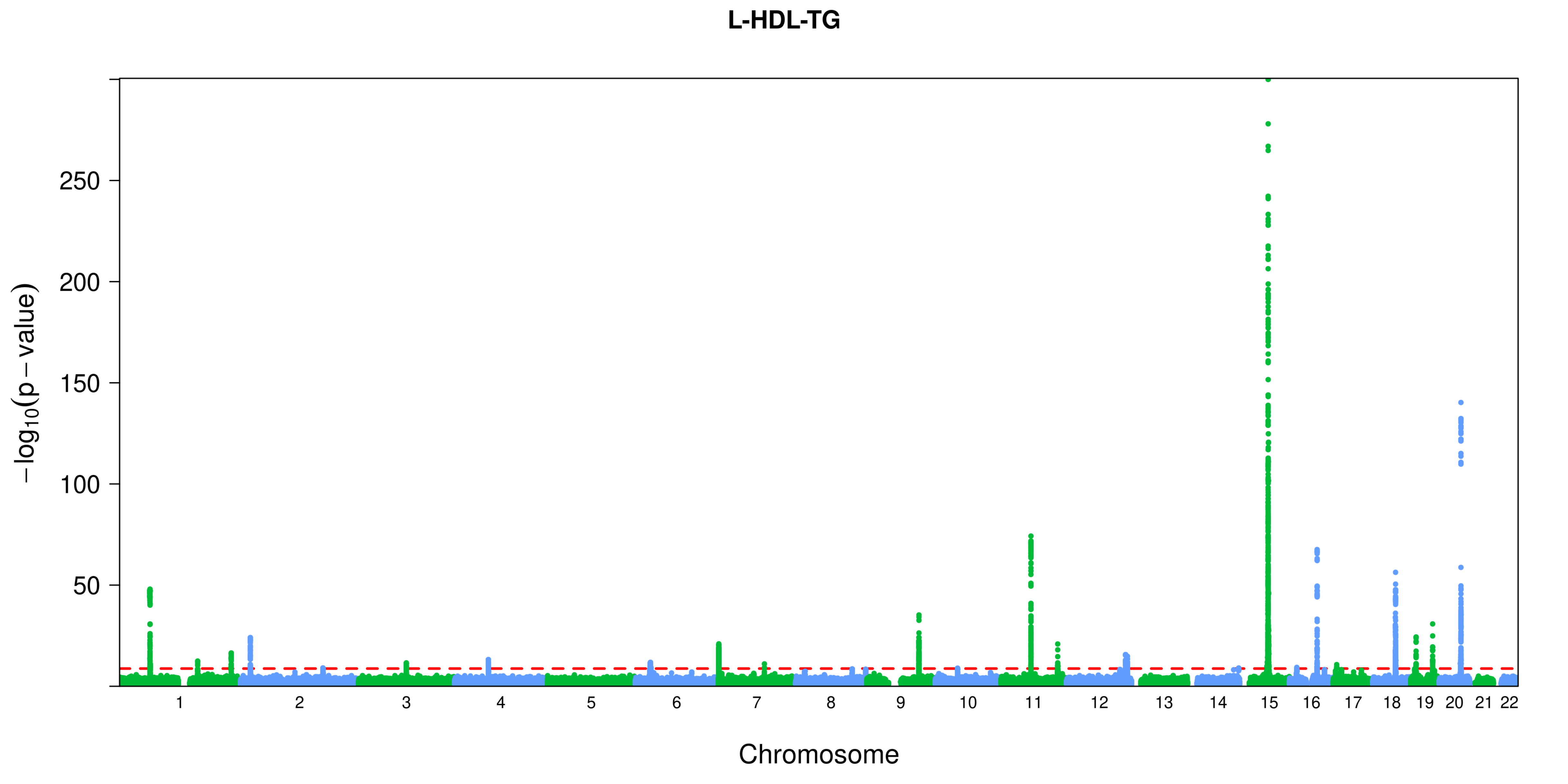

L-HDL-TG
250
200
150
100
50
$-\log_{10}(p-\text{value})$
Chromosome

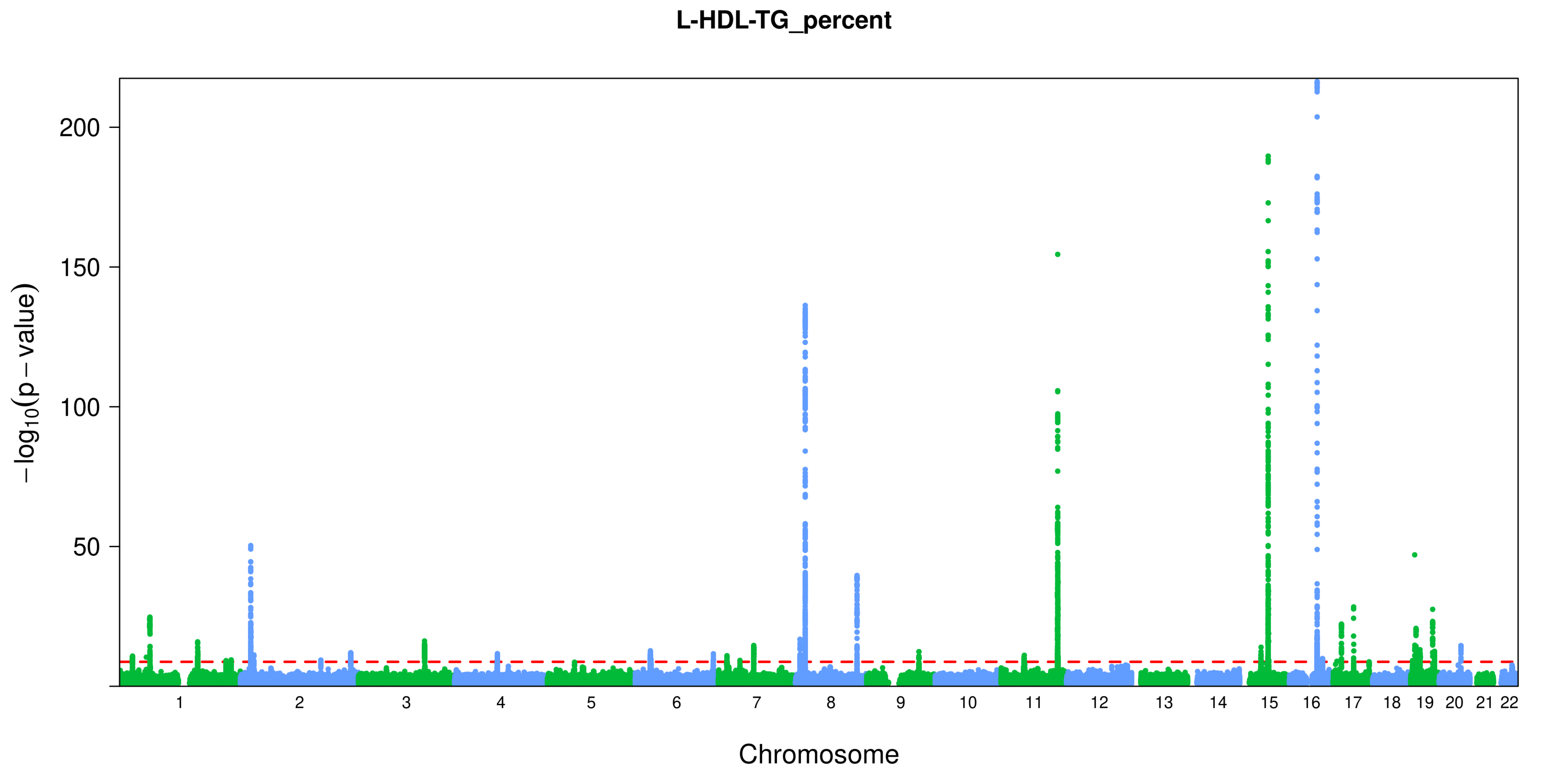
L-HDL-TG_percent
$-\log_{10}(p-\text{value})$
200
150
100
50
Chromosome

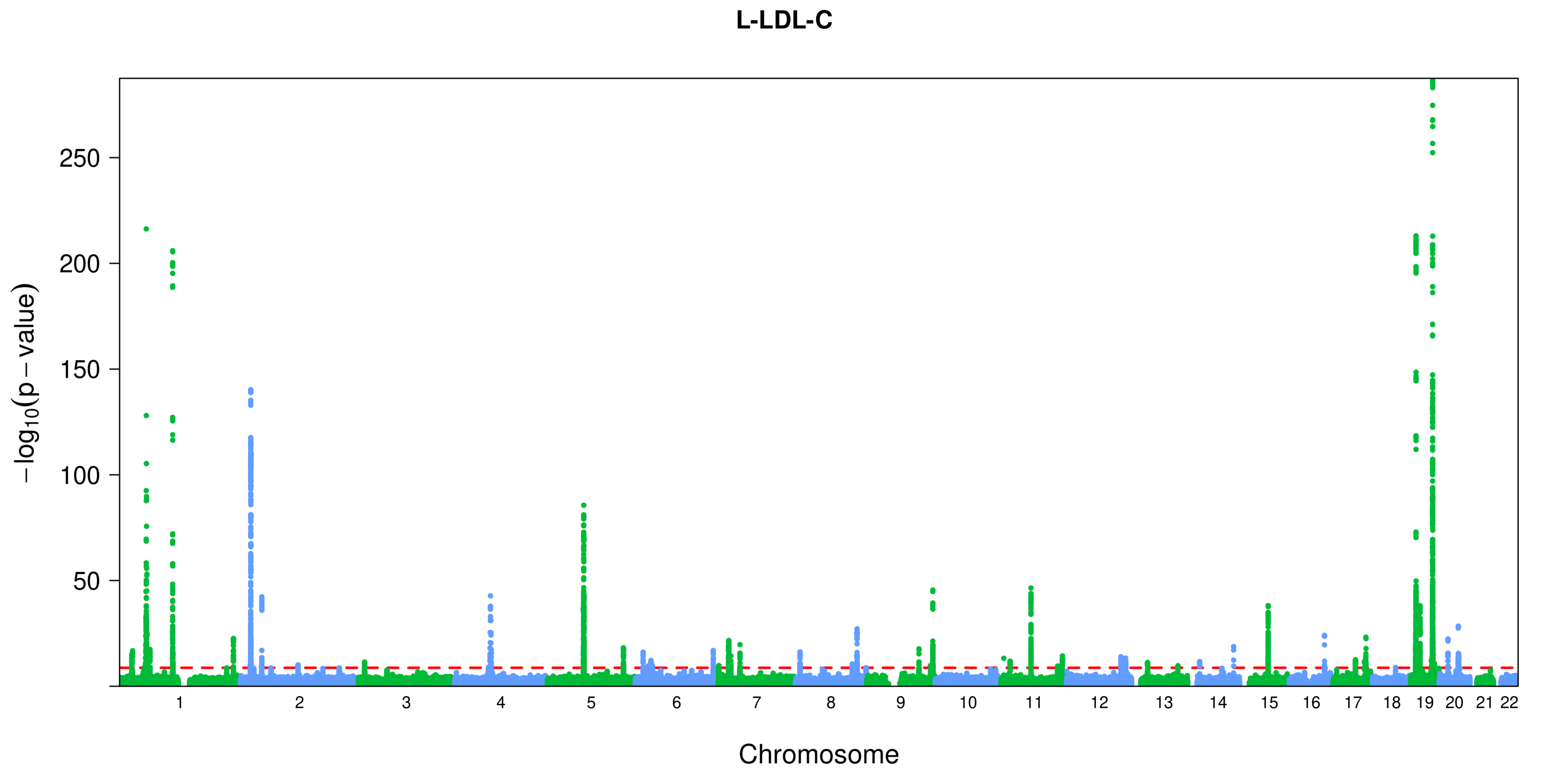

L-LDL-C
250
200
150
100
50
−log10(p − value)
Chromosome

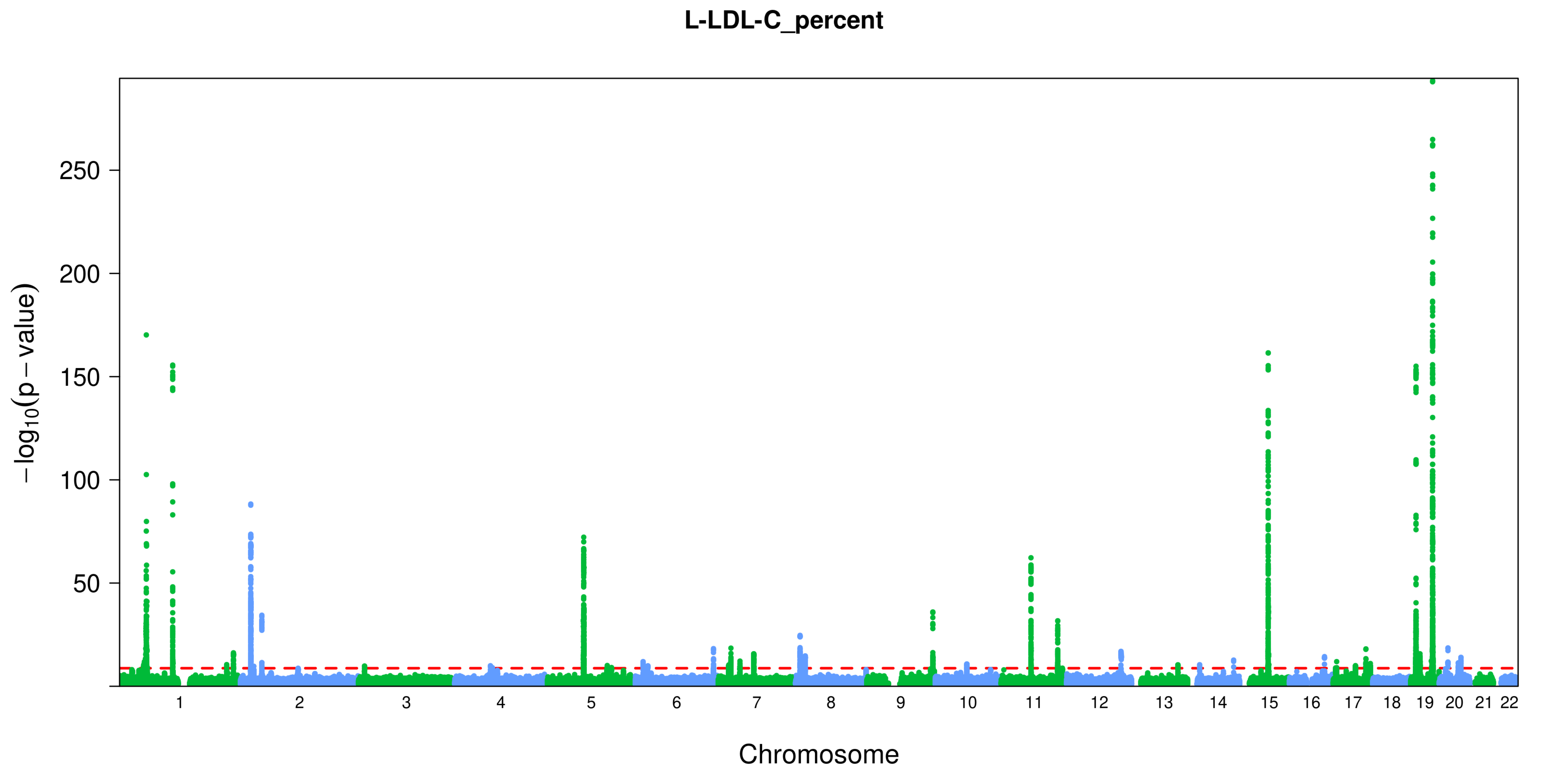

L-LDL-C_percent
$-\log_{10}(p-\text{value})$
250
200
150
100
50
Chromosome

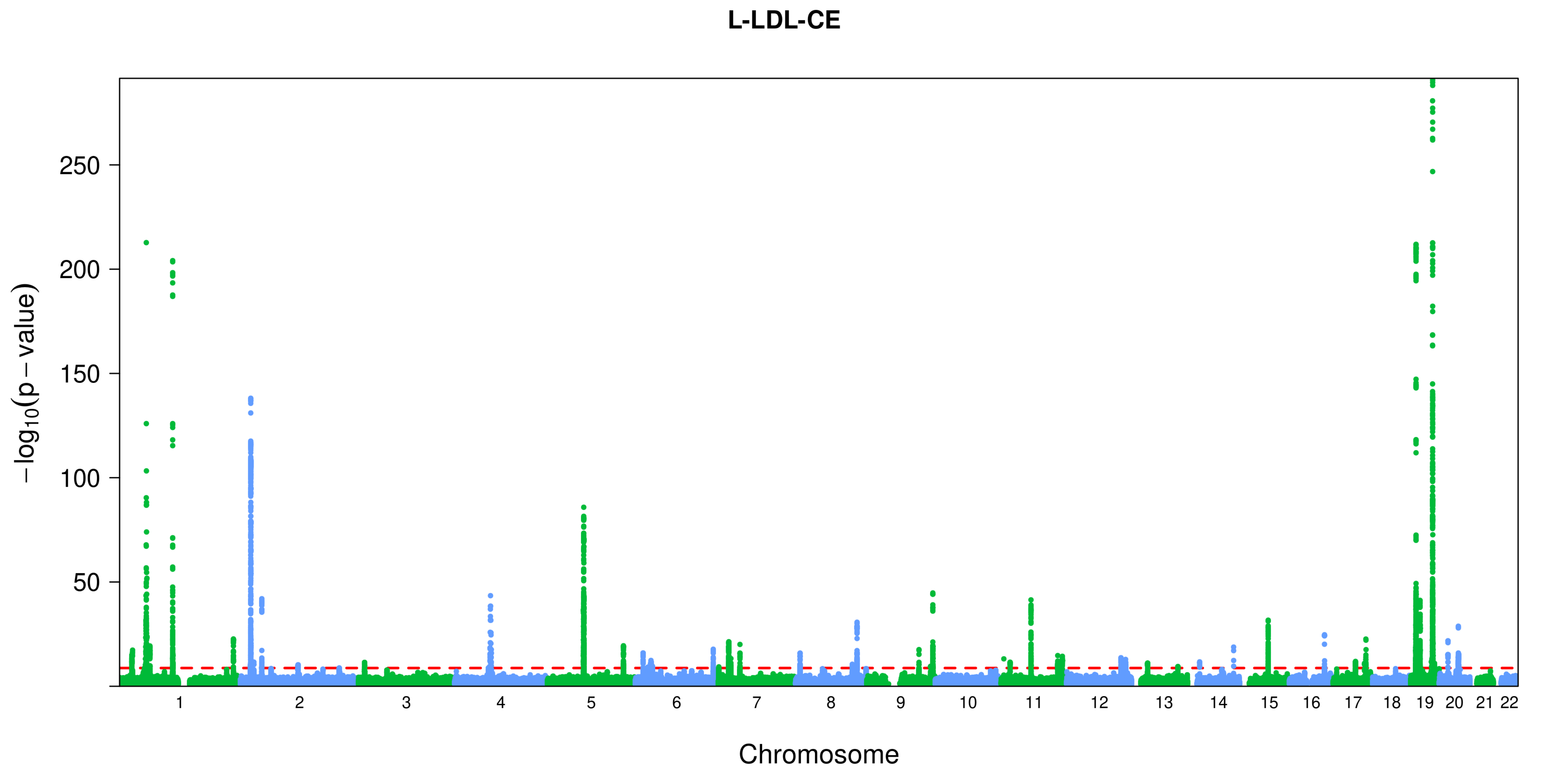
L-LDL-CE
−log10(p − value)
250
200
150
100
50
Chromosome

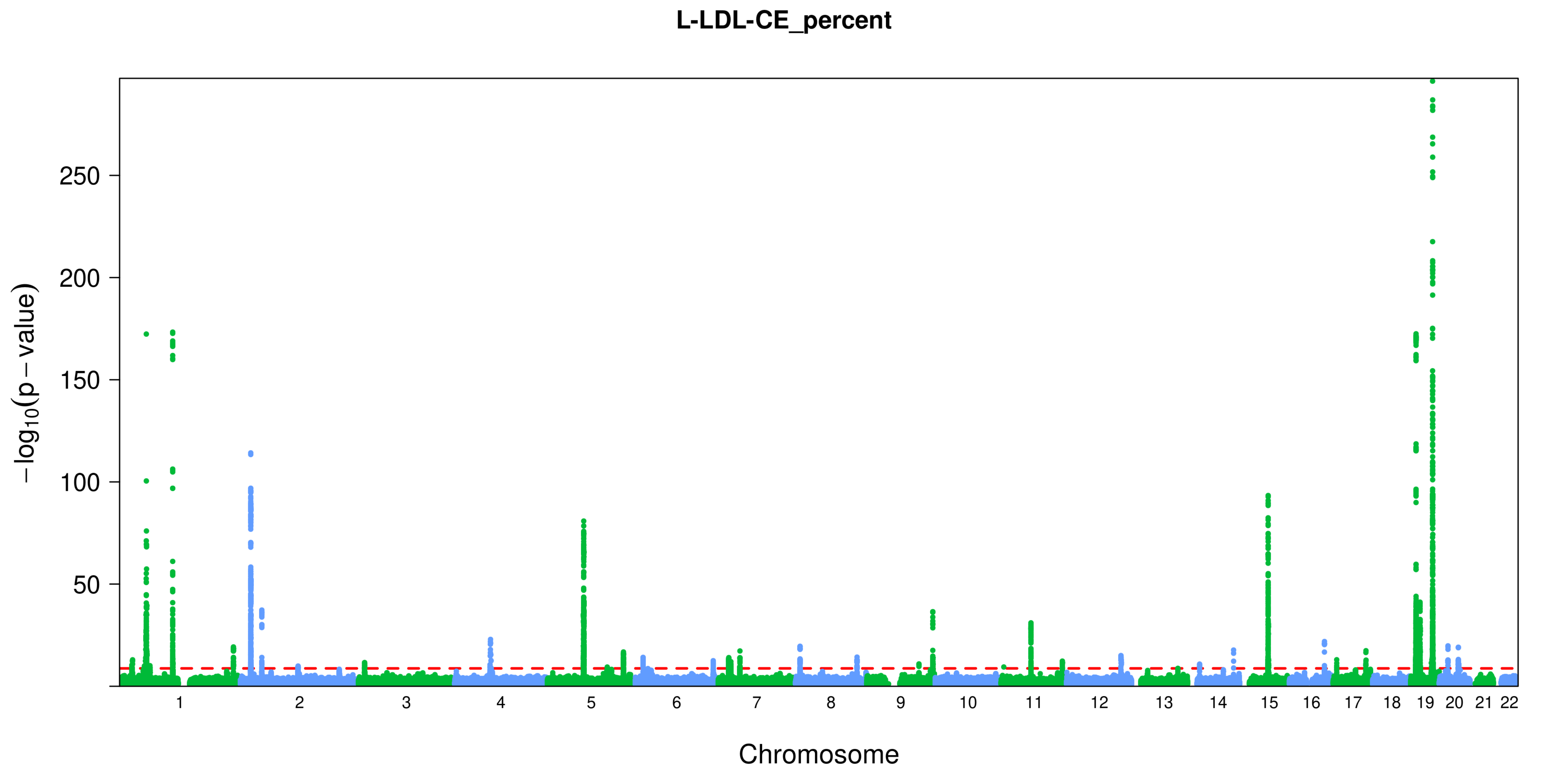
L-LDL-CE_percent
−log10(p − value)
250
200
150
100
50
Chromosome

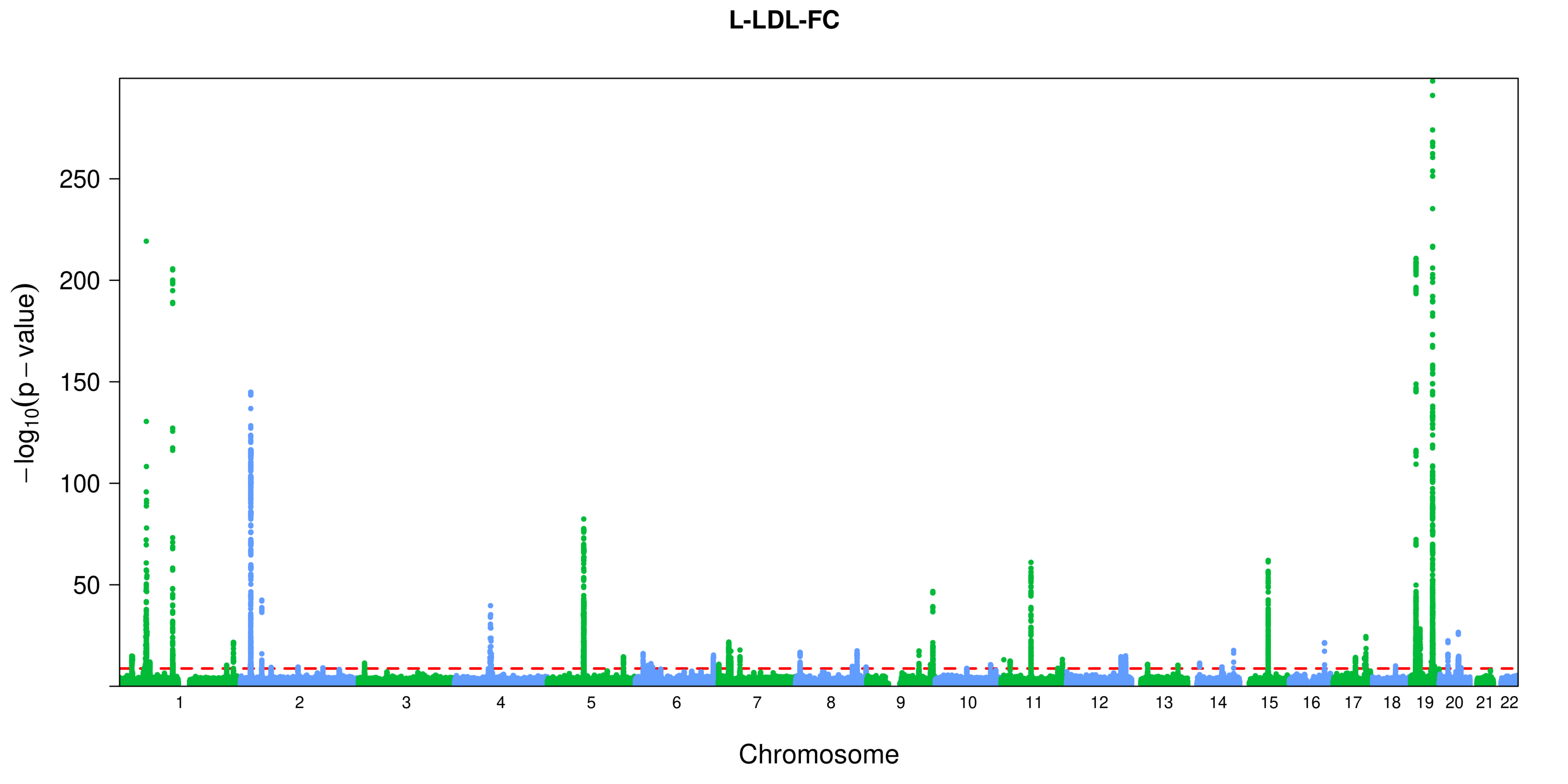
L-LDL-FC
−log10(p − value)
250
200
150
100
50
Chromosome

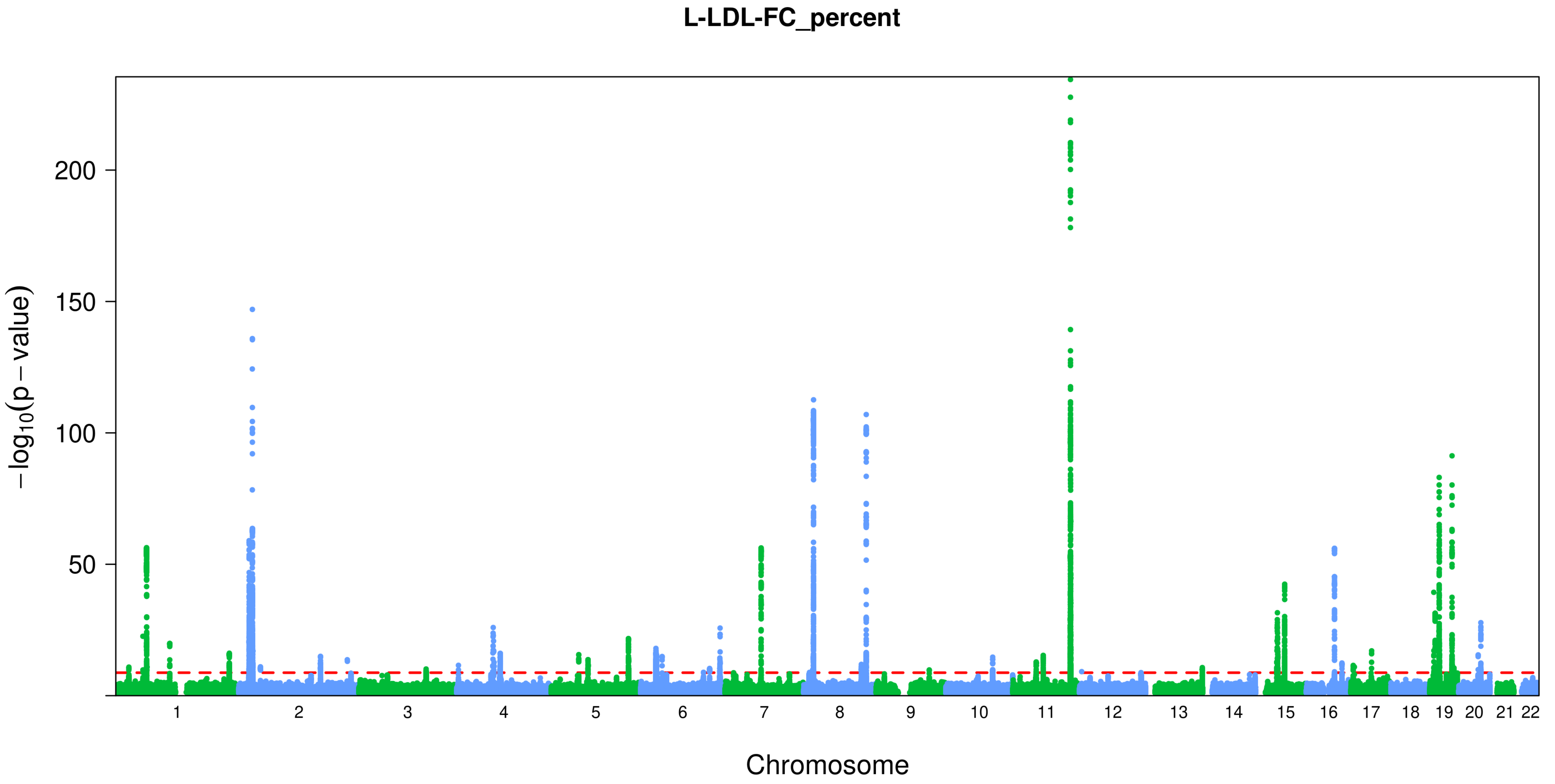

L-LDL-FC_percent
−log10(p − value)
200
150
100
50
Chromosome

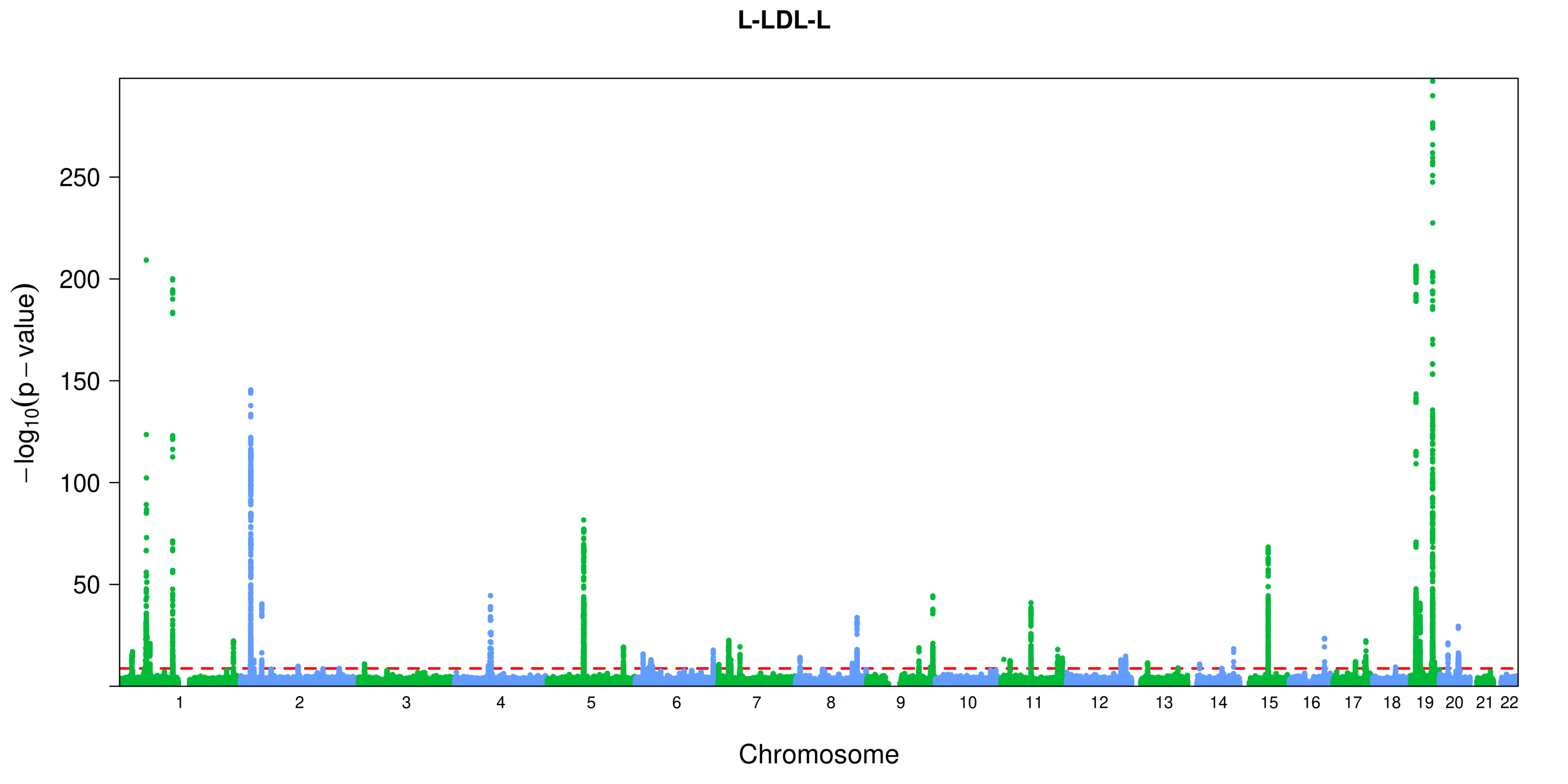

L-LDL-L
−log10(p − value)
250
200
150
100
50
Chromosome

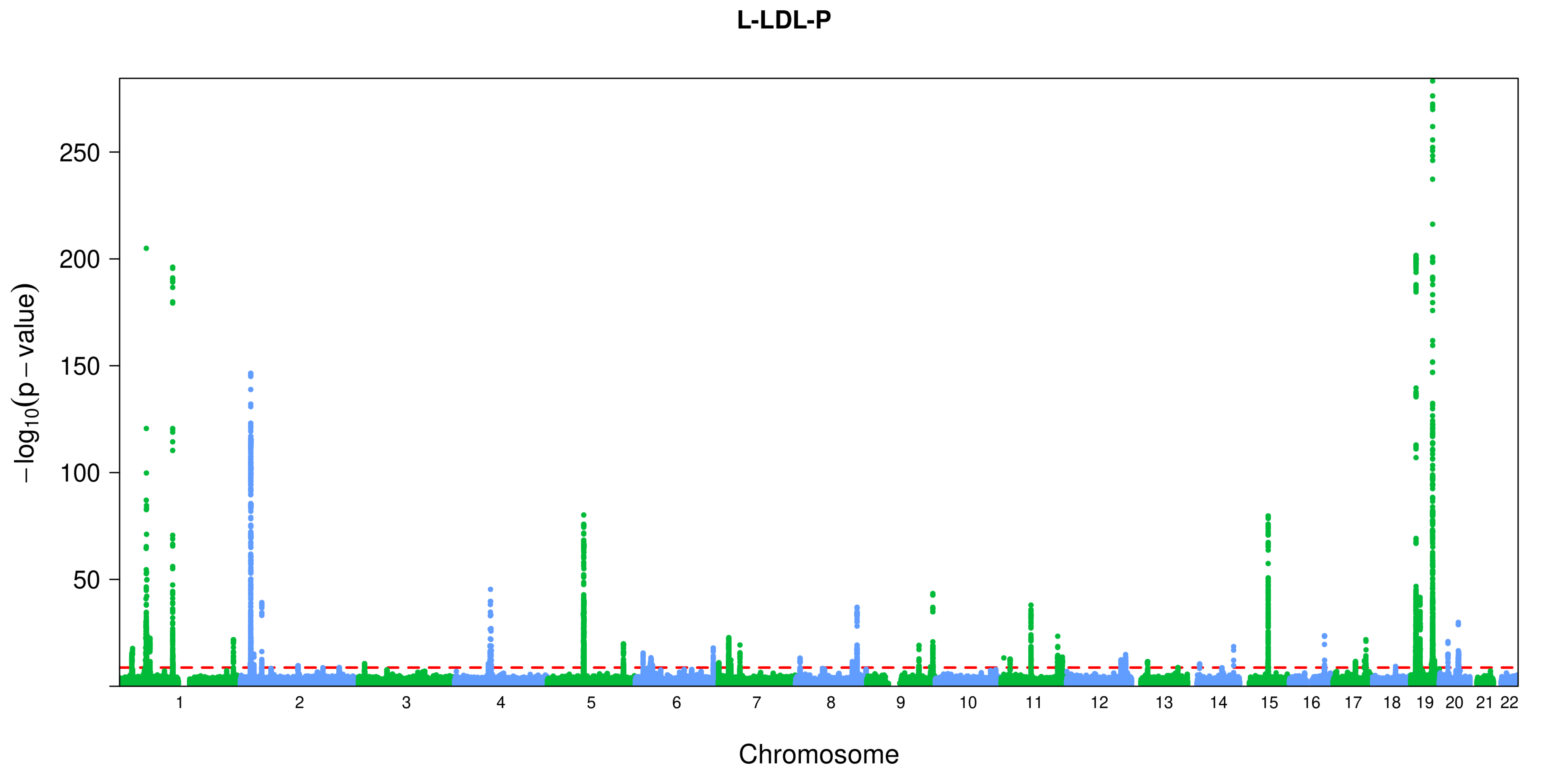

L-LDL-P
−log10(p − value)
250
200
150
100
50
Chromosome

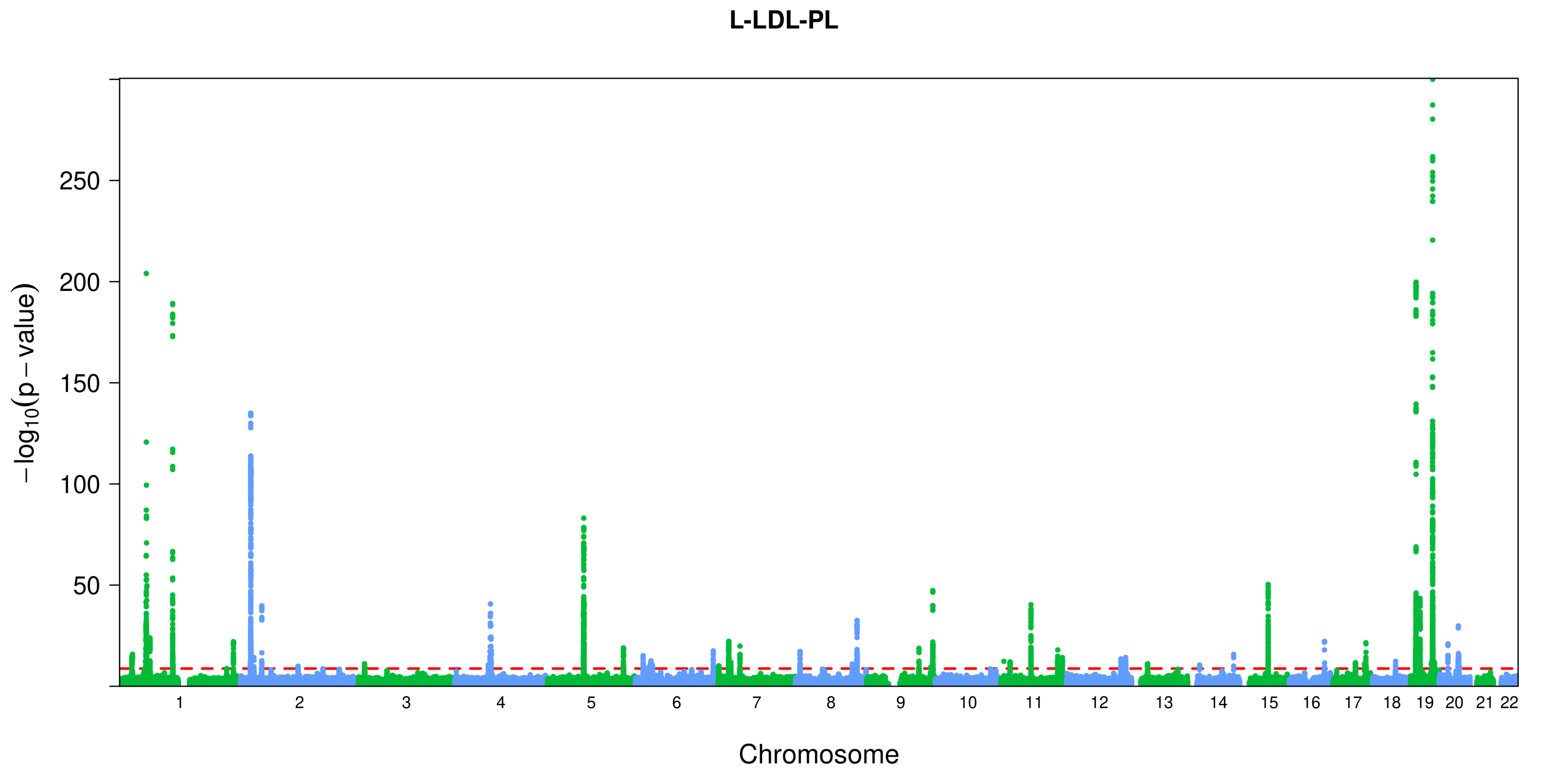
L-LDL-PL
−log10(p − value)
250
200
150
100
50
Chromosome

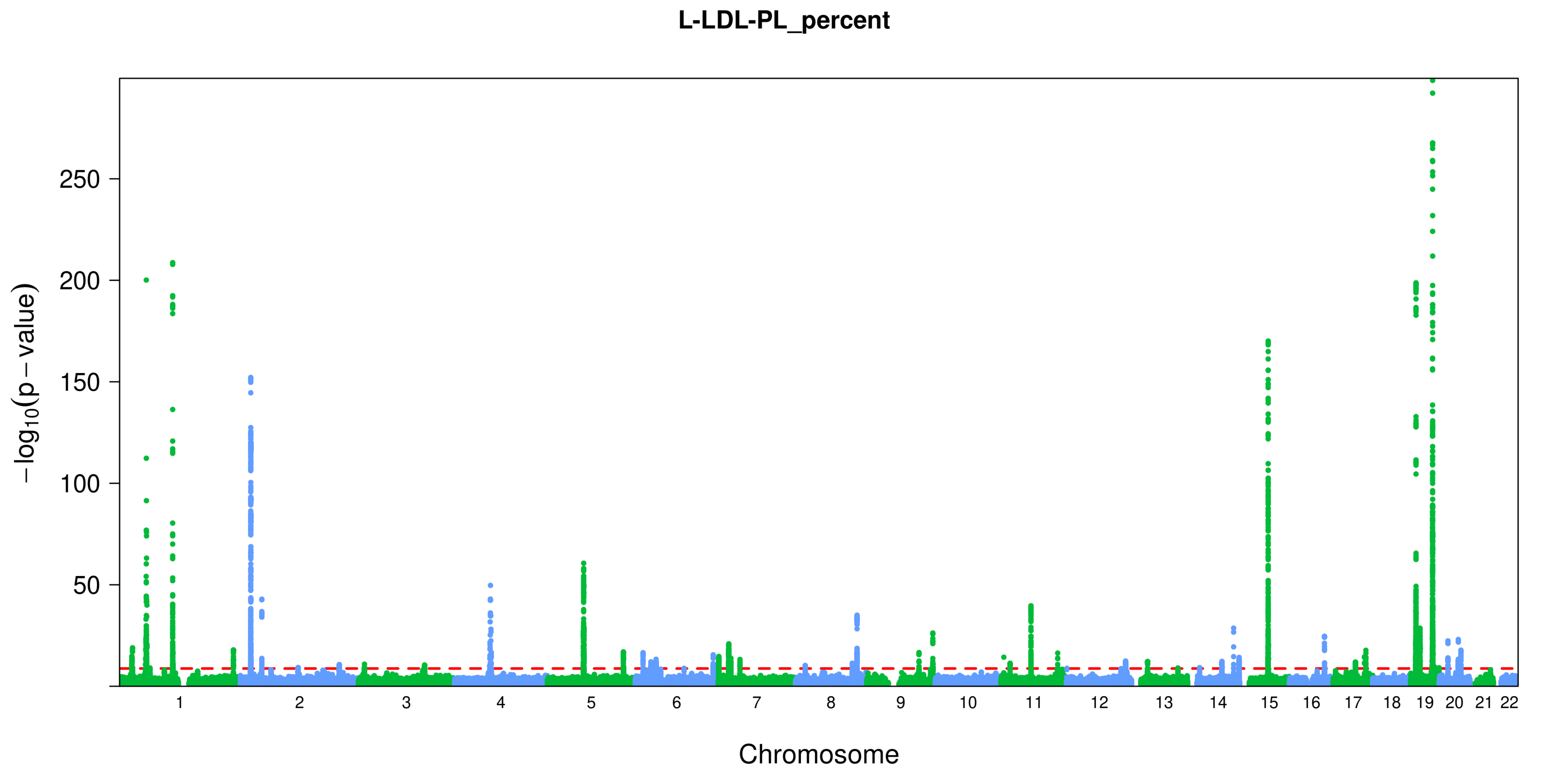

L-LDL-PL_percent
250
200
150
100
50
$-\log_{10}(p-\text{value})$
Chromosome

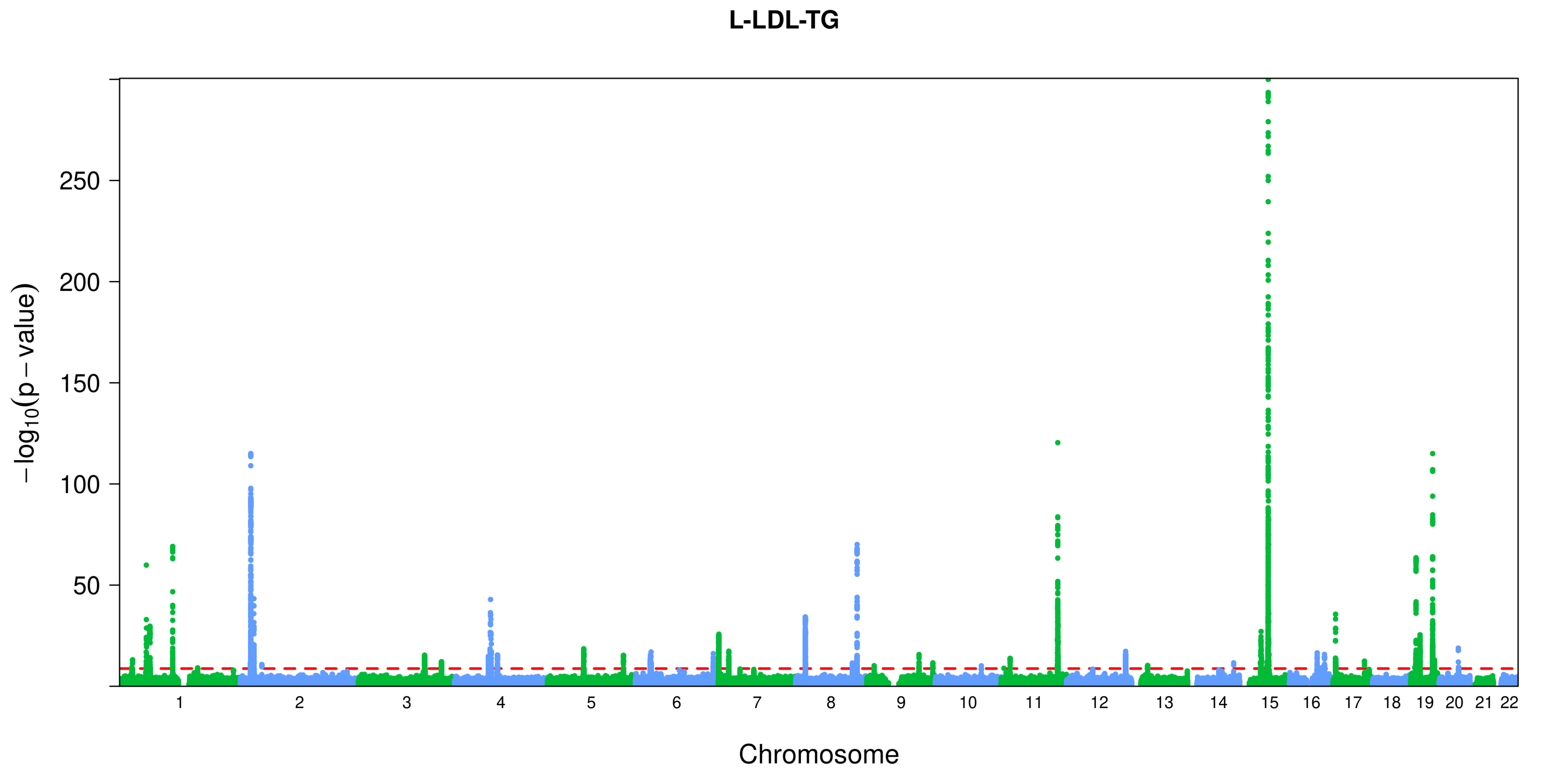
L-LDL-TG
−log10(p − value)
250
200
150
100
50
Chromosome

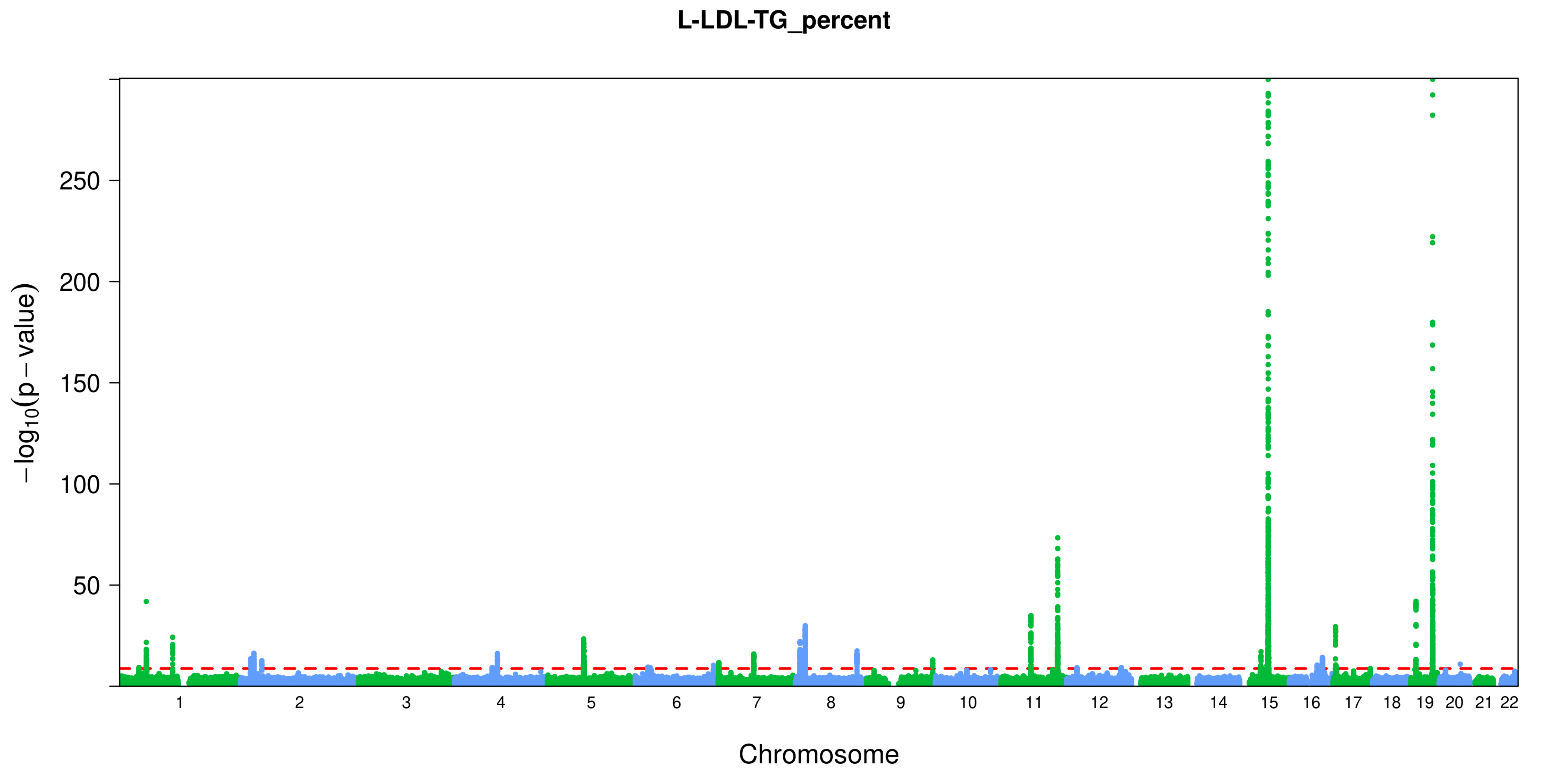
L-LDL-TG_percent
−log10(p − value)
250
200
150
100
50
Chromosome

L-VLDL-C

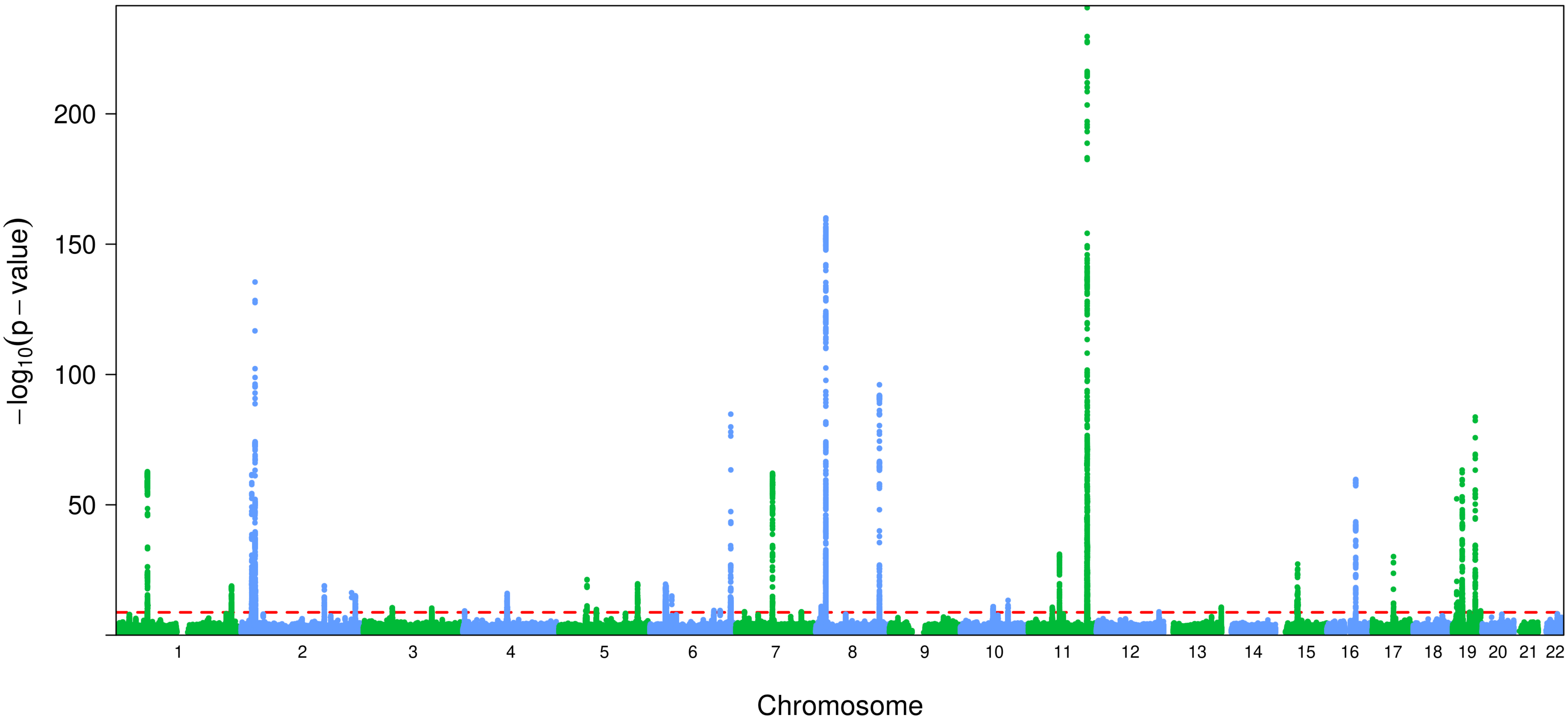

L-VLDL-C
200
150
100
50
−log10(p − value)
Chromosome

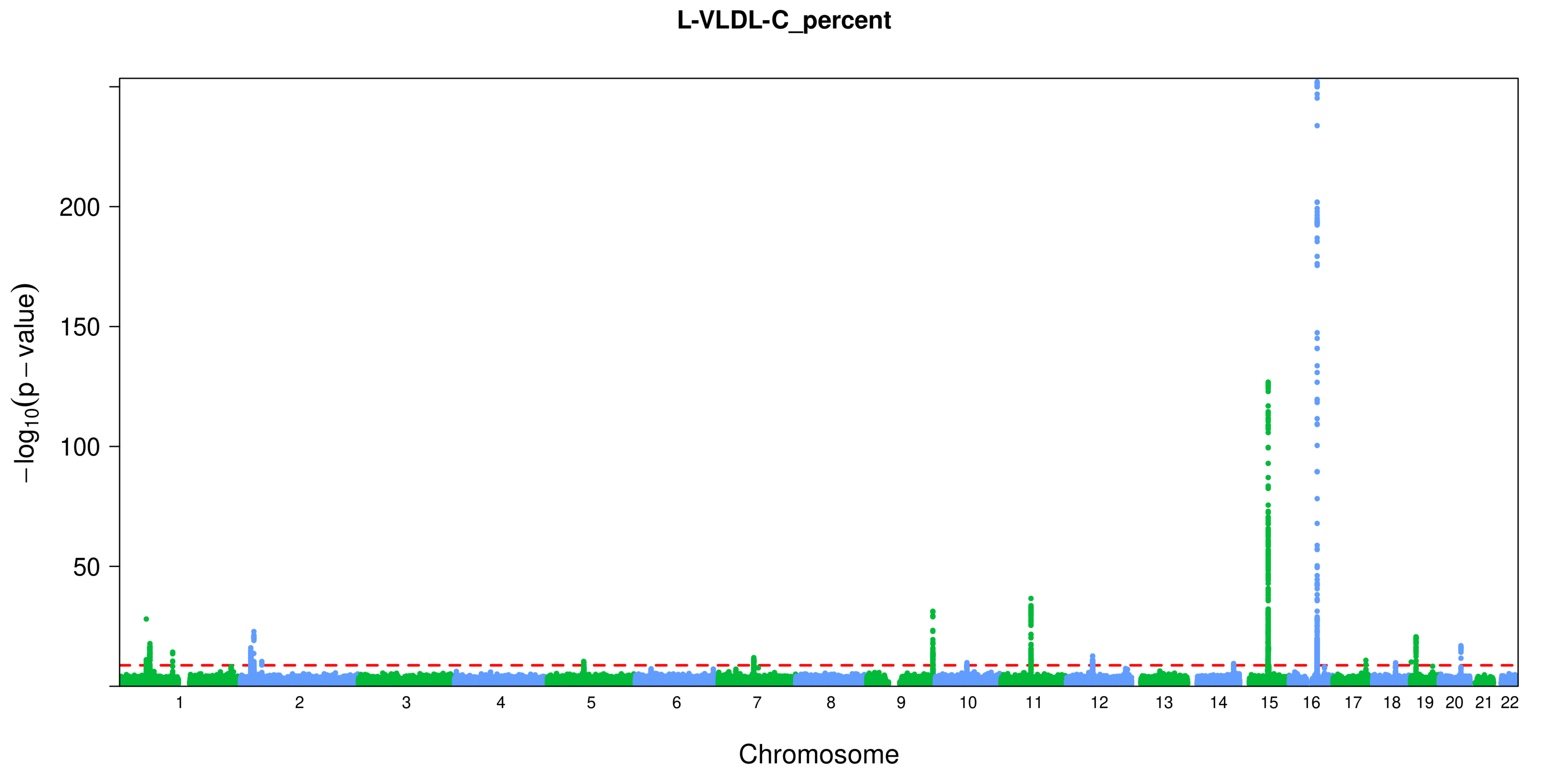

L-VLDL-C_percent
−log10(p − value)
200
150
100
50
Chromosome

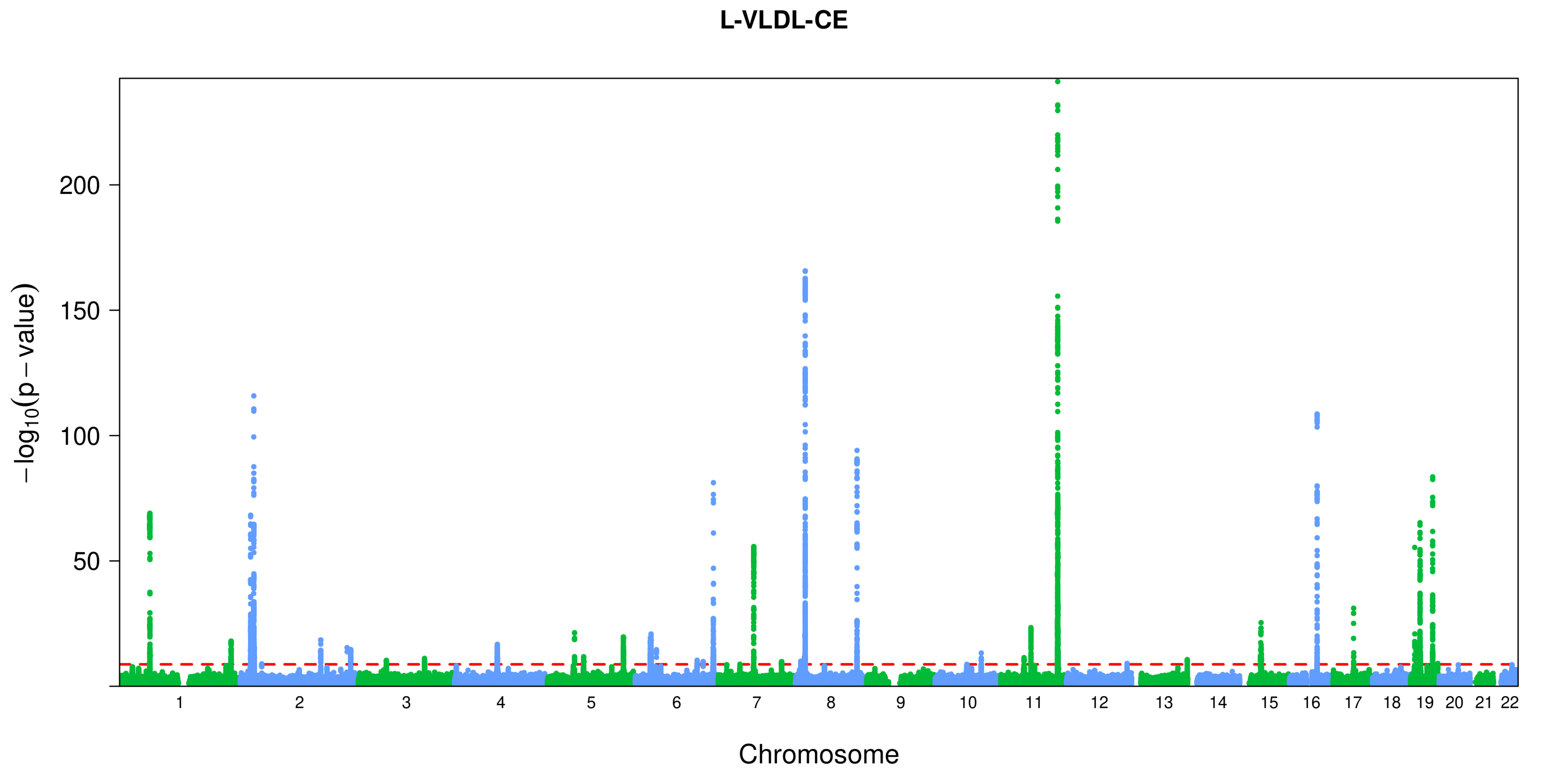
L-VLDL-CE
$-\log_{10}(p-\text{value})$
200
150
100
50
1 2 3 4 5 6 7 8 9 10 11 12 13 14 15 16 17 18 19 20 21 22
Chromosome

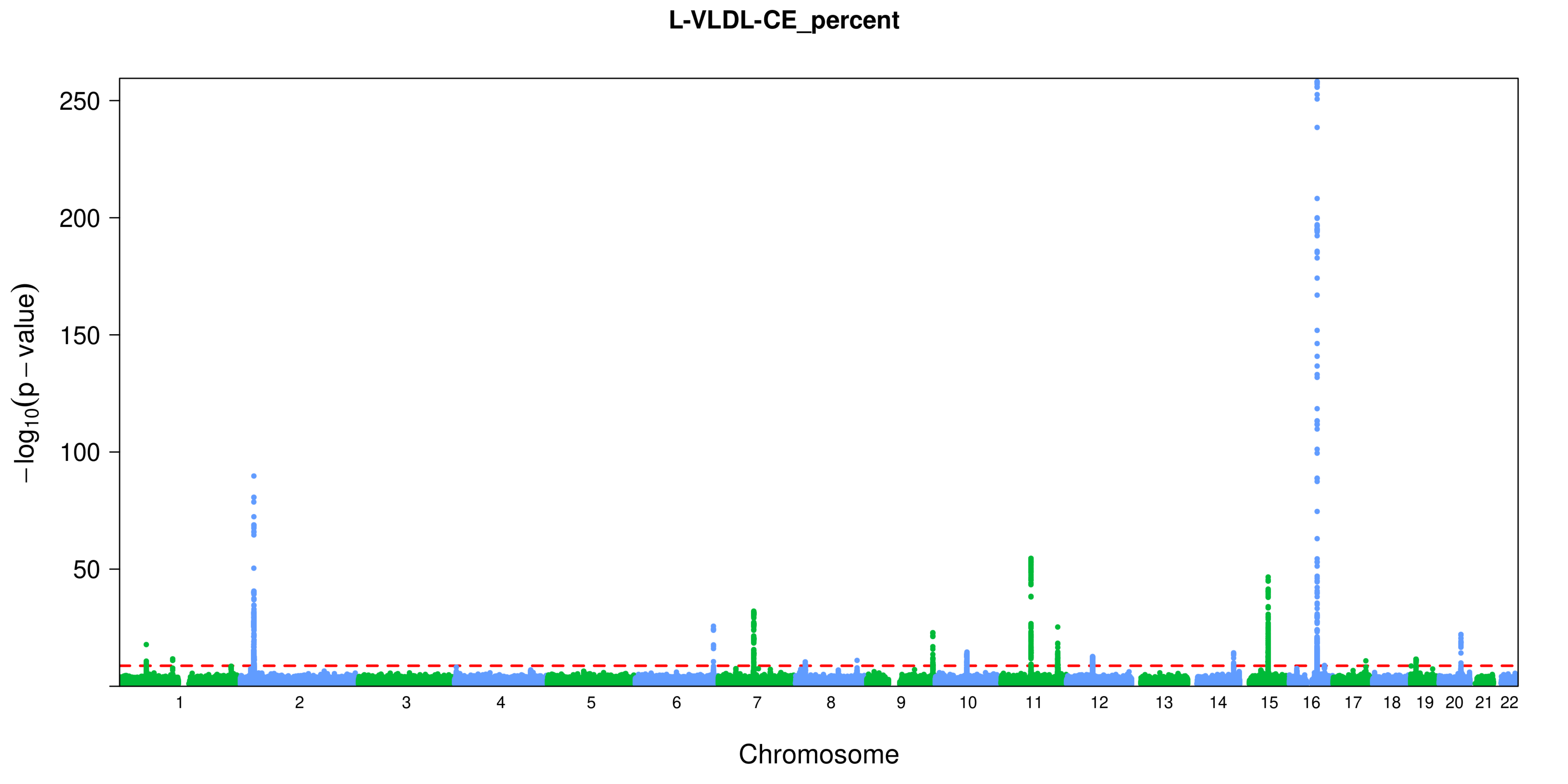

L-VLDL-CE_percent
−log10(p − value)
250
200
150
100
50
Chromosome

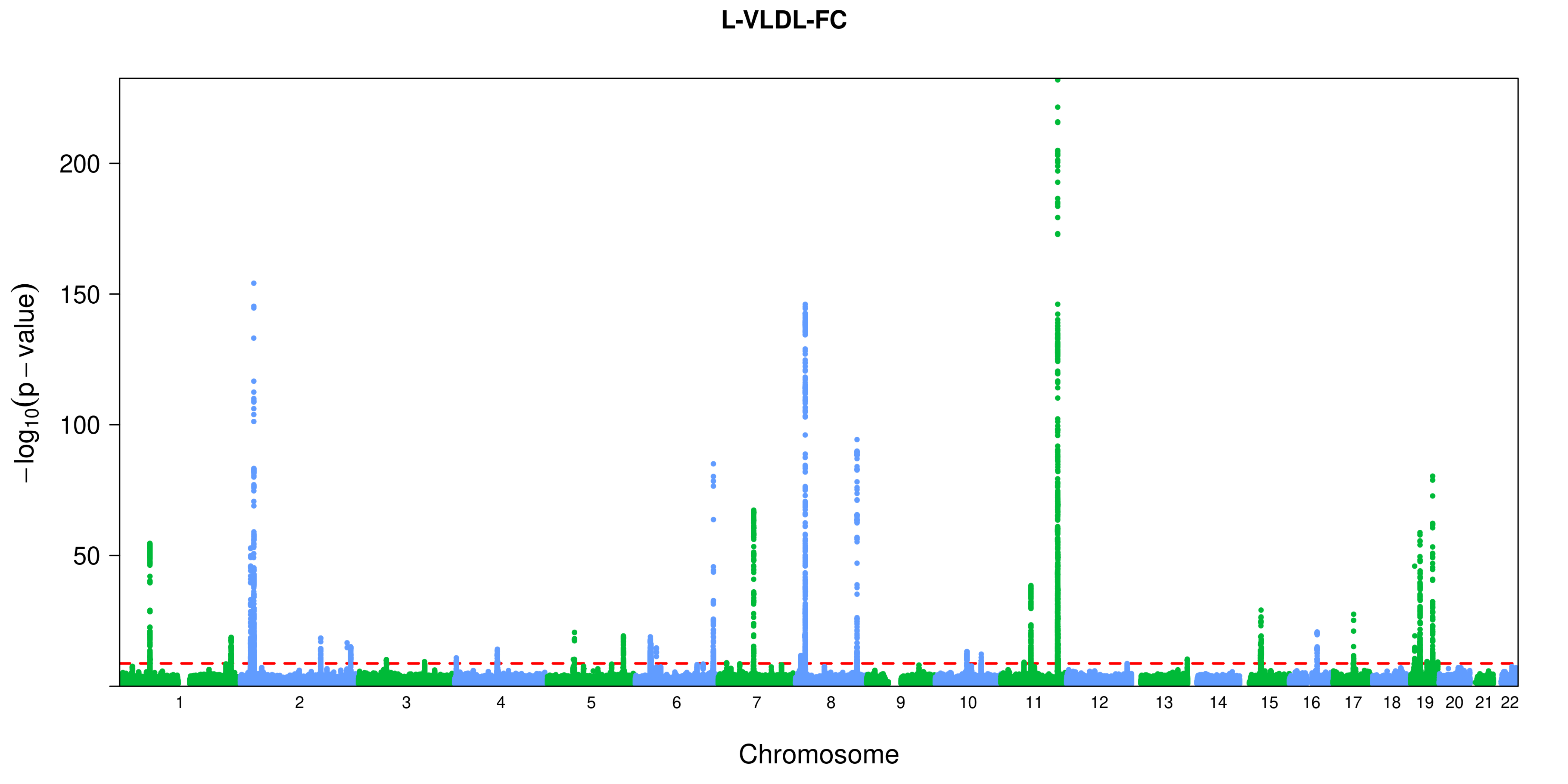
L-VLDL-FC
200
150
100
50
$-\log_{10}(p-\text{value})$
Chromosome

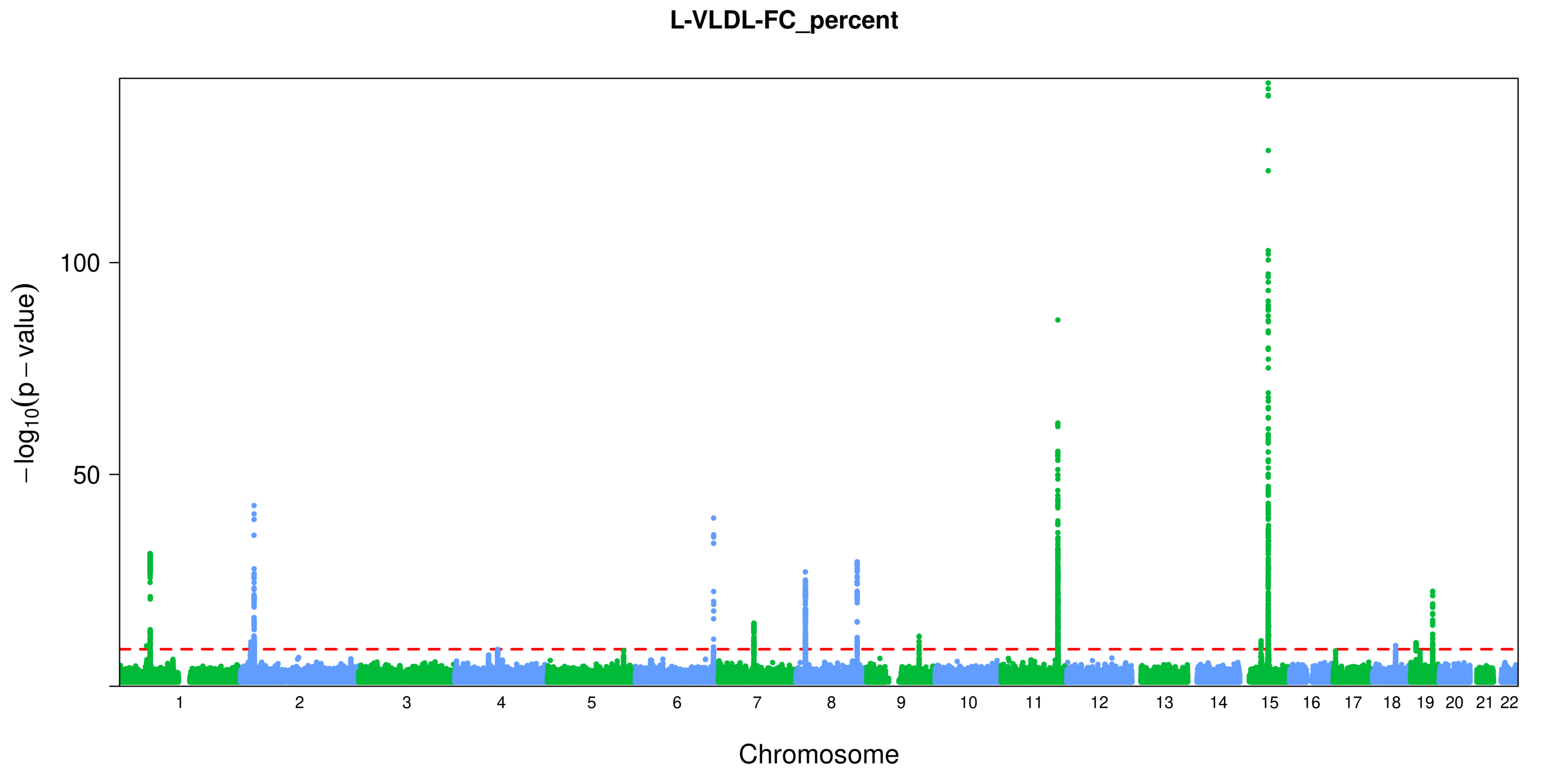

L-VLDL-FC_percent
100
50
−log10(p − value)
Chromosome

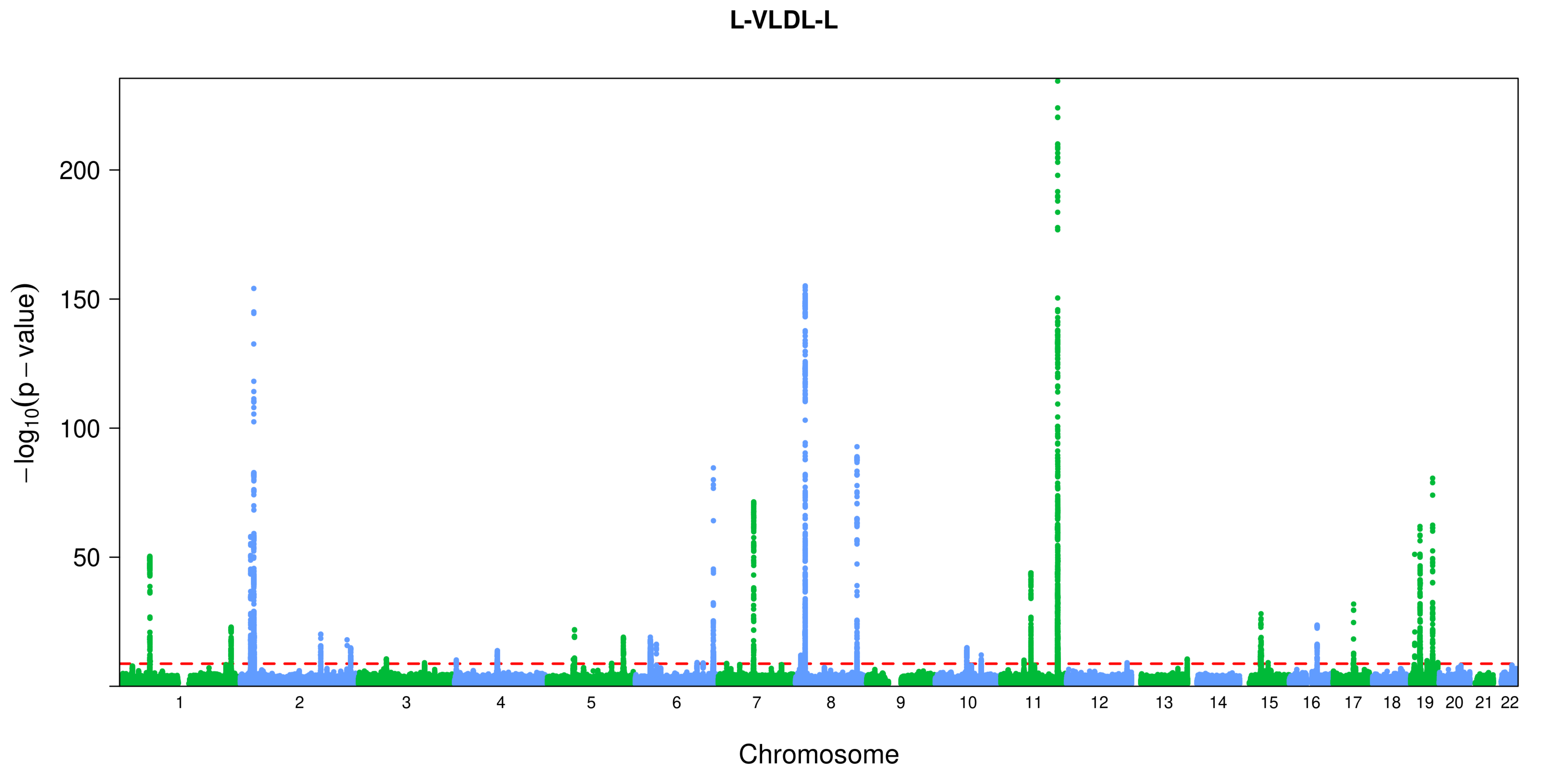
L-VLDL-L
200
150
100
50
$-\log_{10}(p - \text{value})$
1 2 3 4 5 6 7 8 9 10 11 12 13 14 15 16 17 18 19 20 21 22
Chromosome

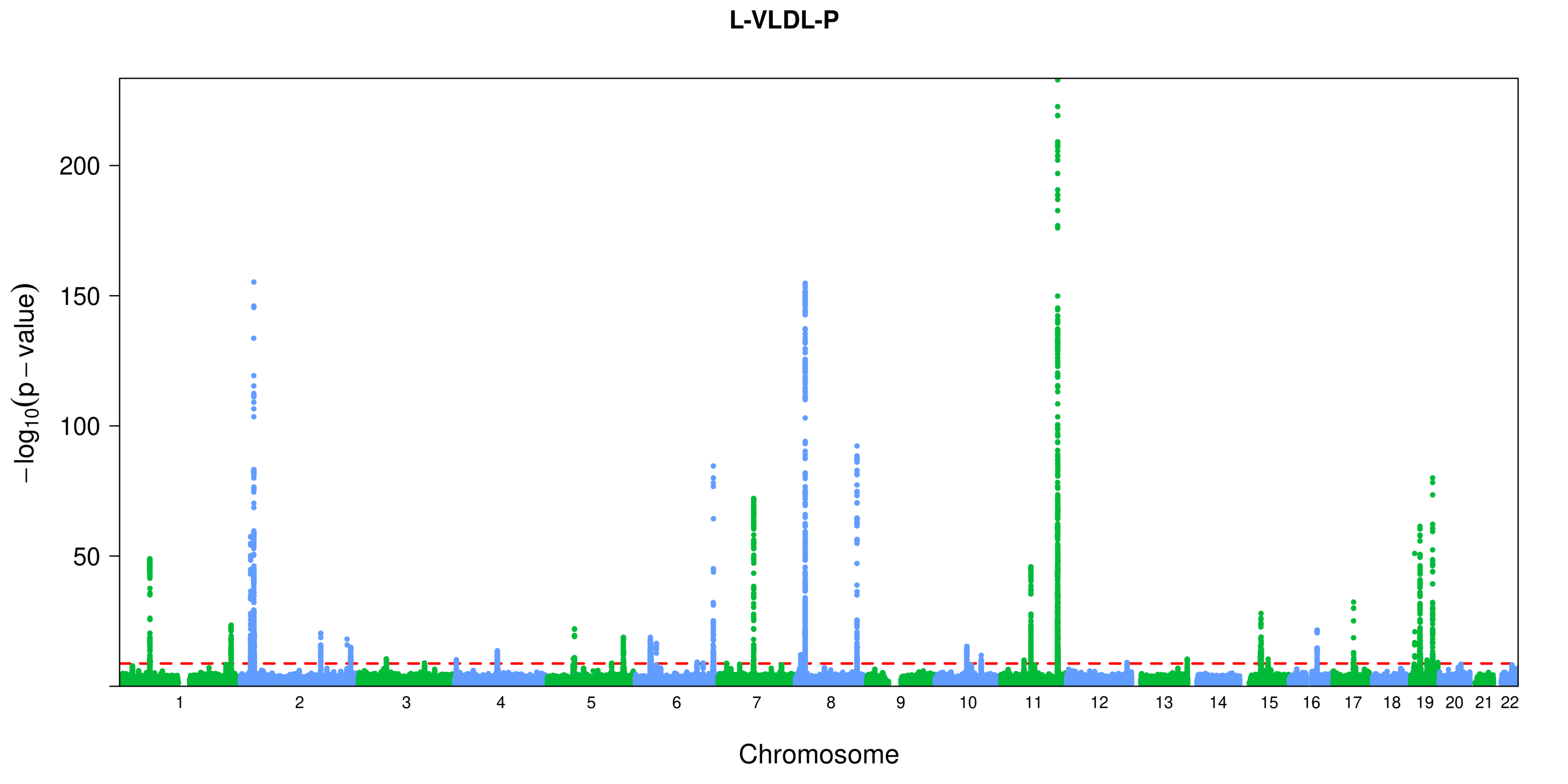

L-VLDL-P
200
150
100
50
$-\log_{10}(p-\text{value})$
Chromosome

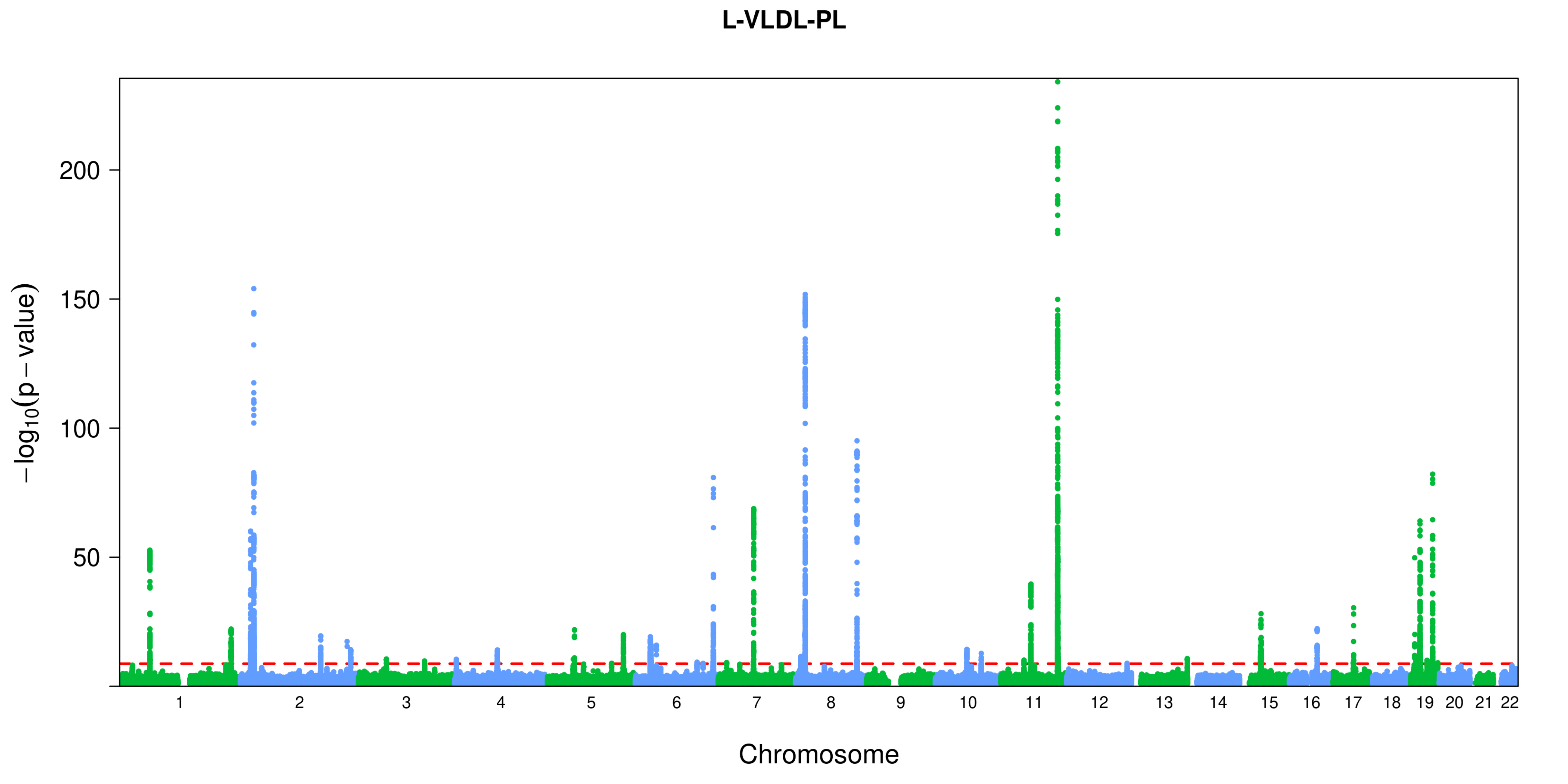
L-VLDL-PL
−log10(p − value)
200
150
100
50
Chromosome

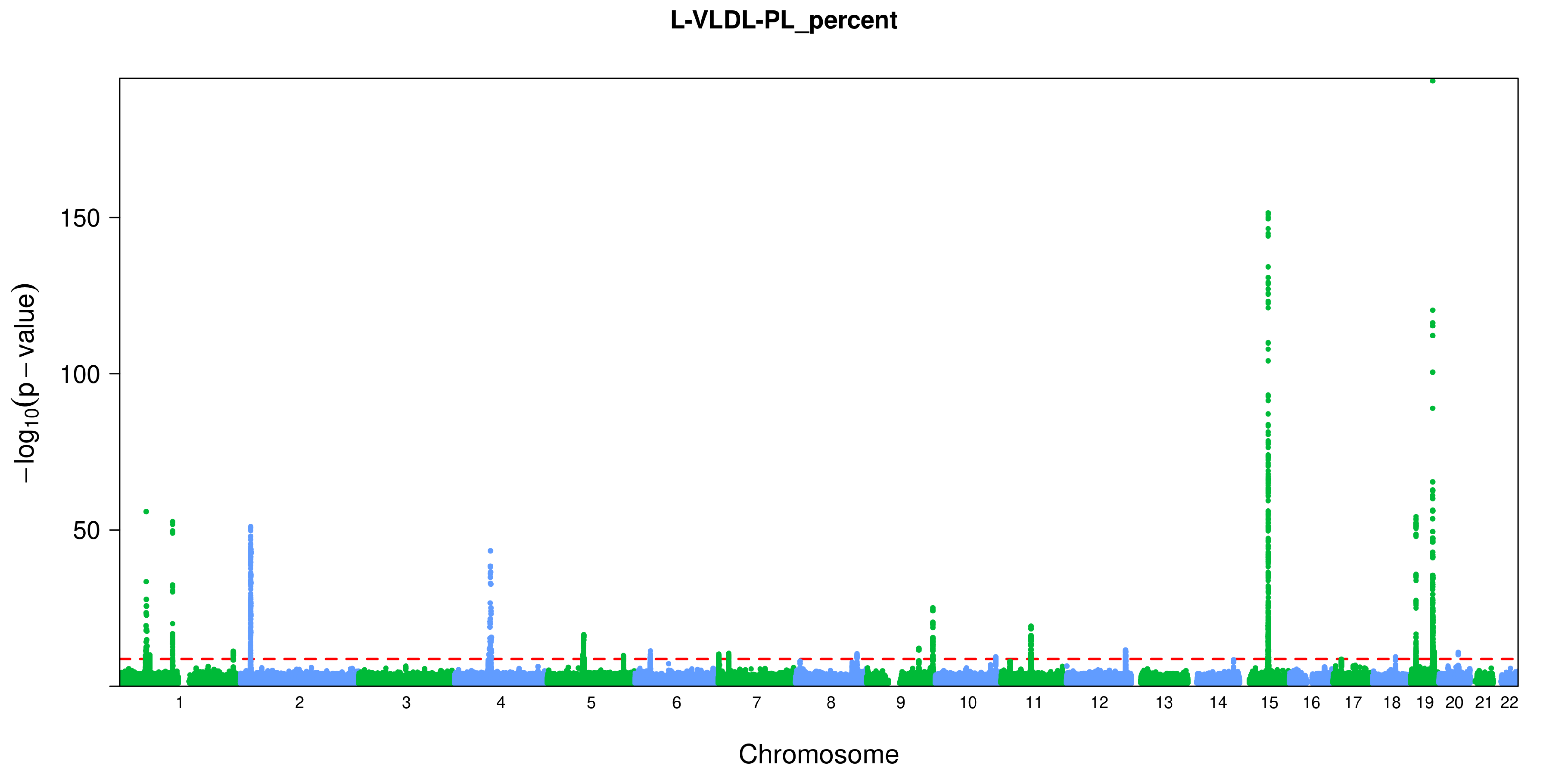

L-VLDL-PL_percent
150
100
50
$-\log_{10}(p-value)$
Chromosome

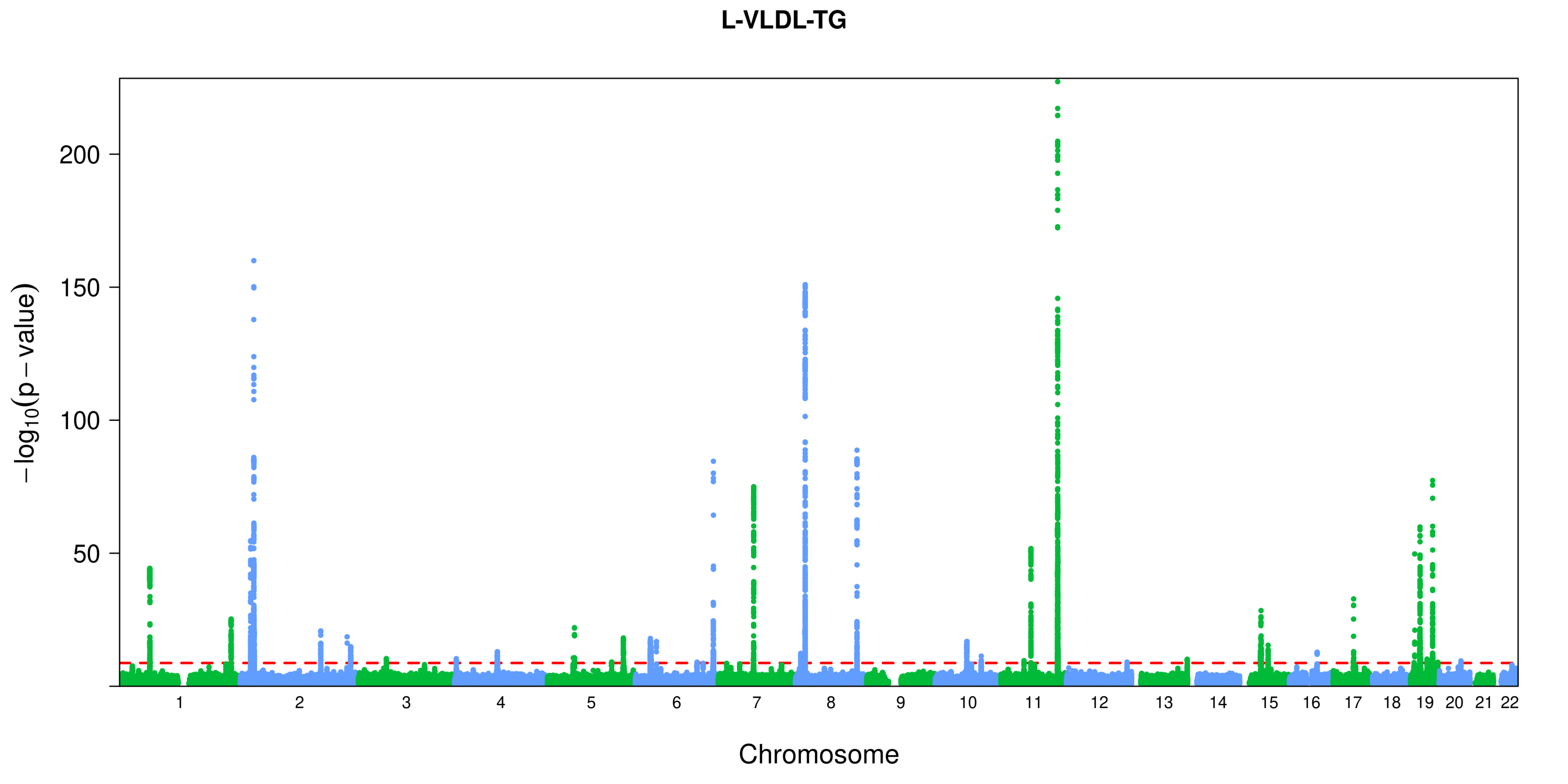
L-VLDL-TG
−log10(p − value)
200
150
100
50
Chromosome

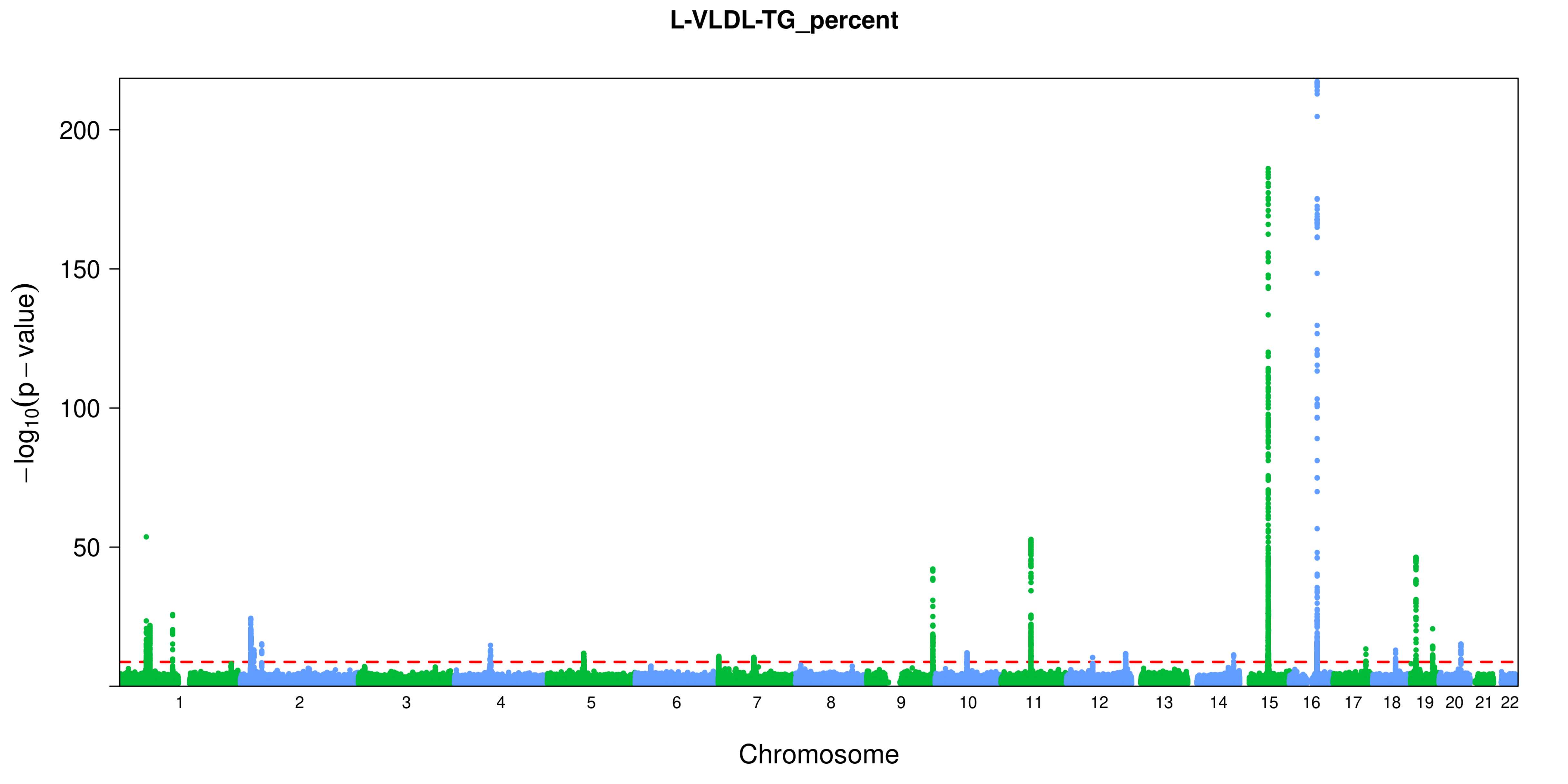
L-VLDL-TG_percent
−log10(p − value)
200
150
100
50
Chromosome

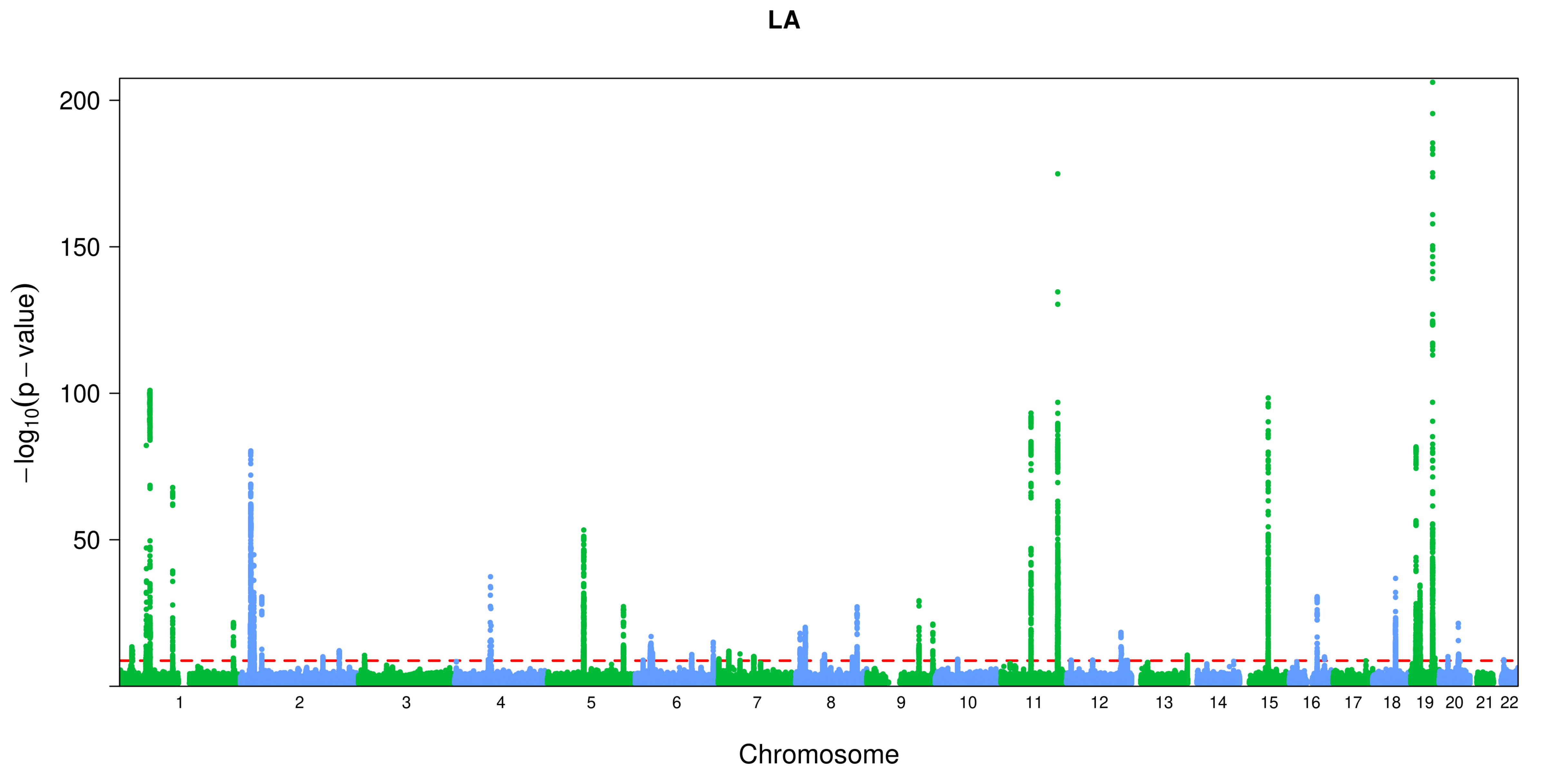

LA
200
150
100
50
$-\log_{10}(p-\text{value})$
Chromosome

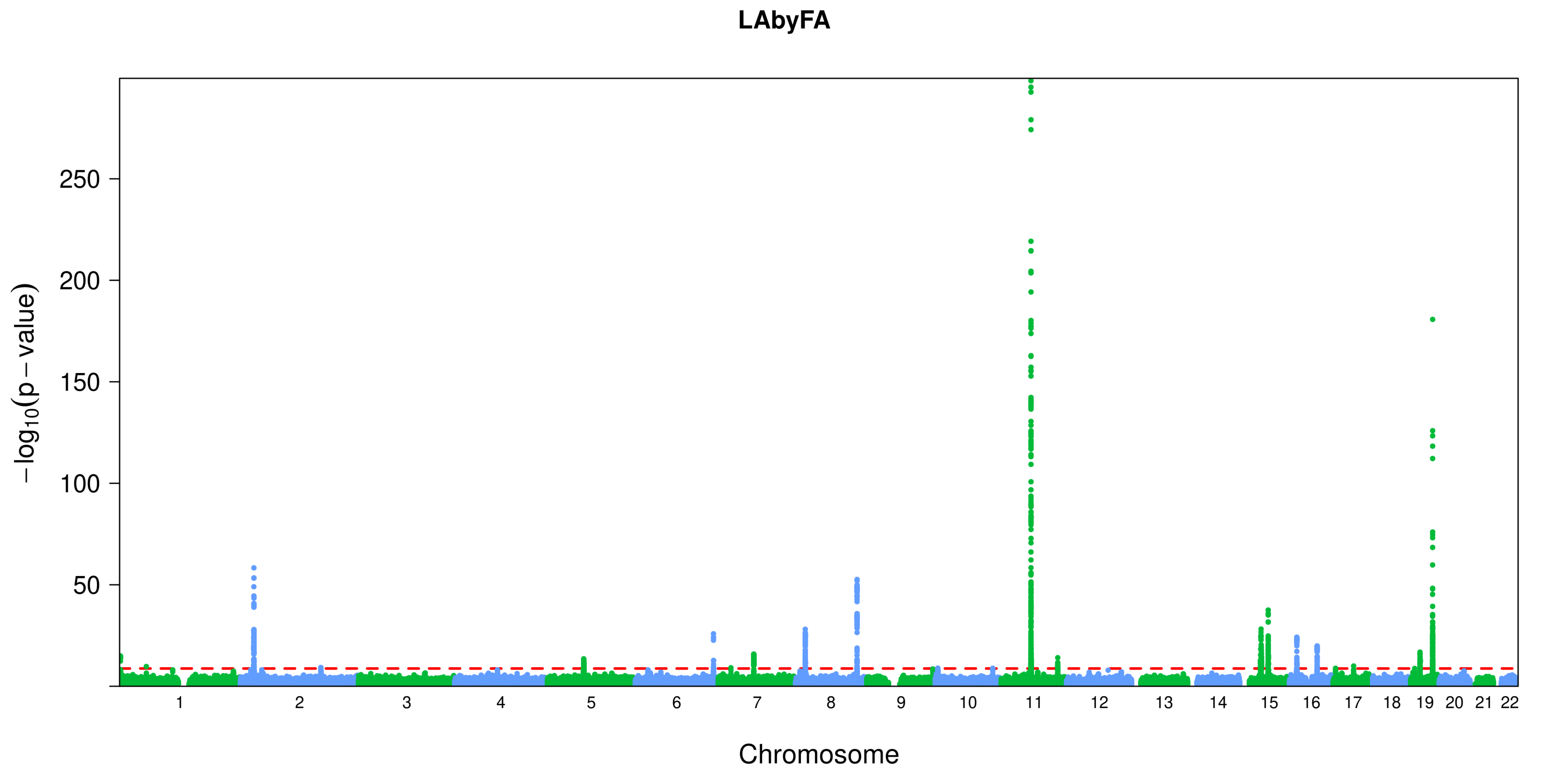
LAbyFA
250
200
150
100
50
−log10(p − value)
Chromosome

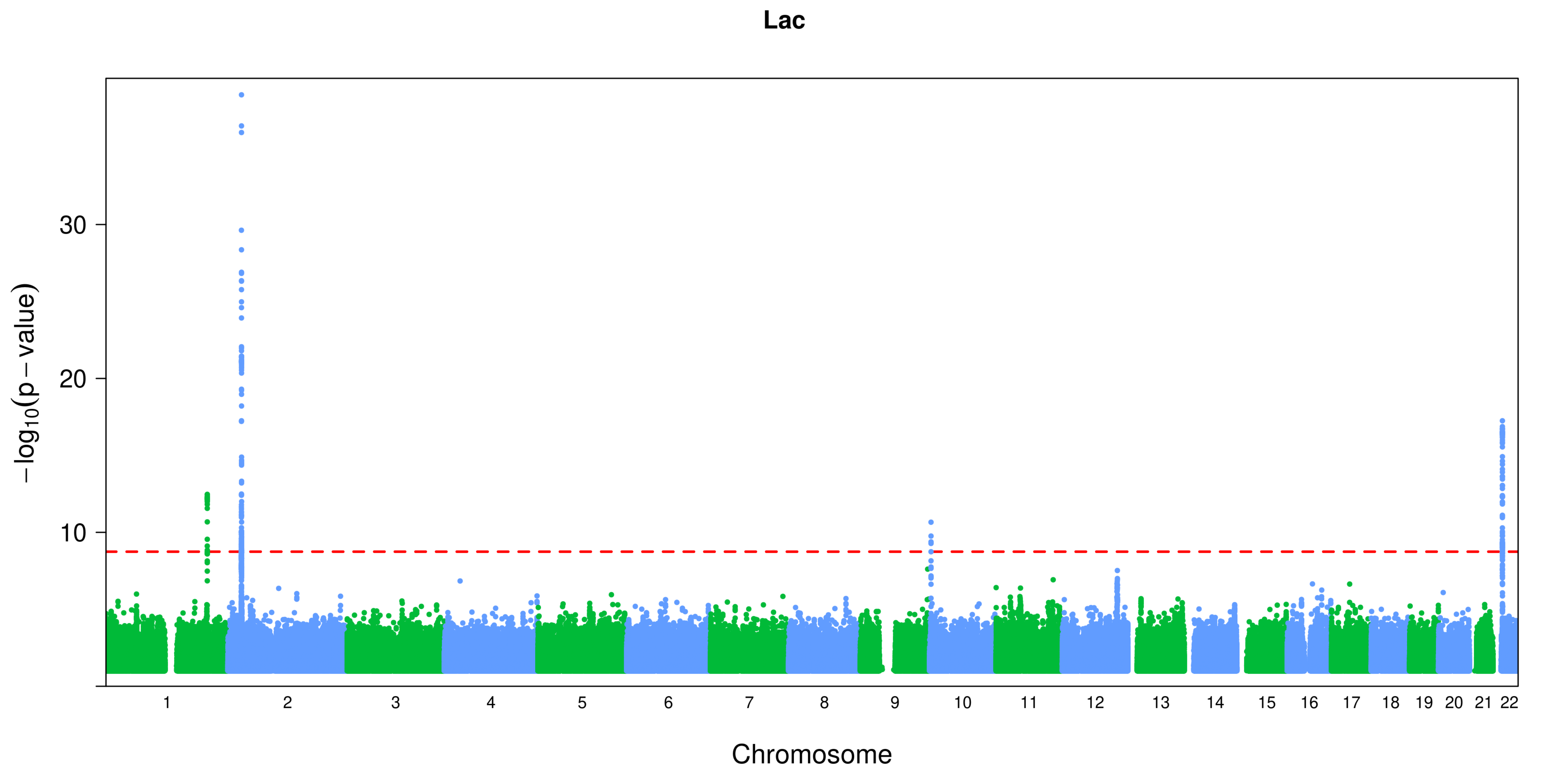

Lac
30
20
10
−log10(p − value)
Chromosome

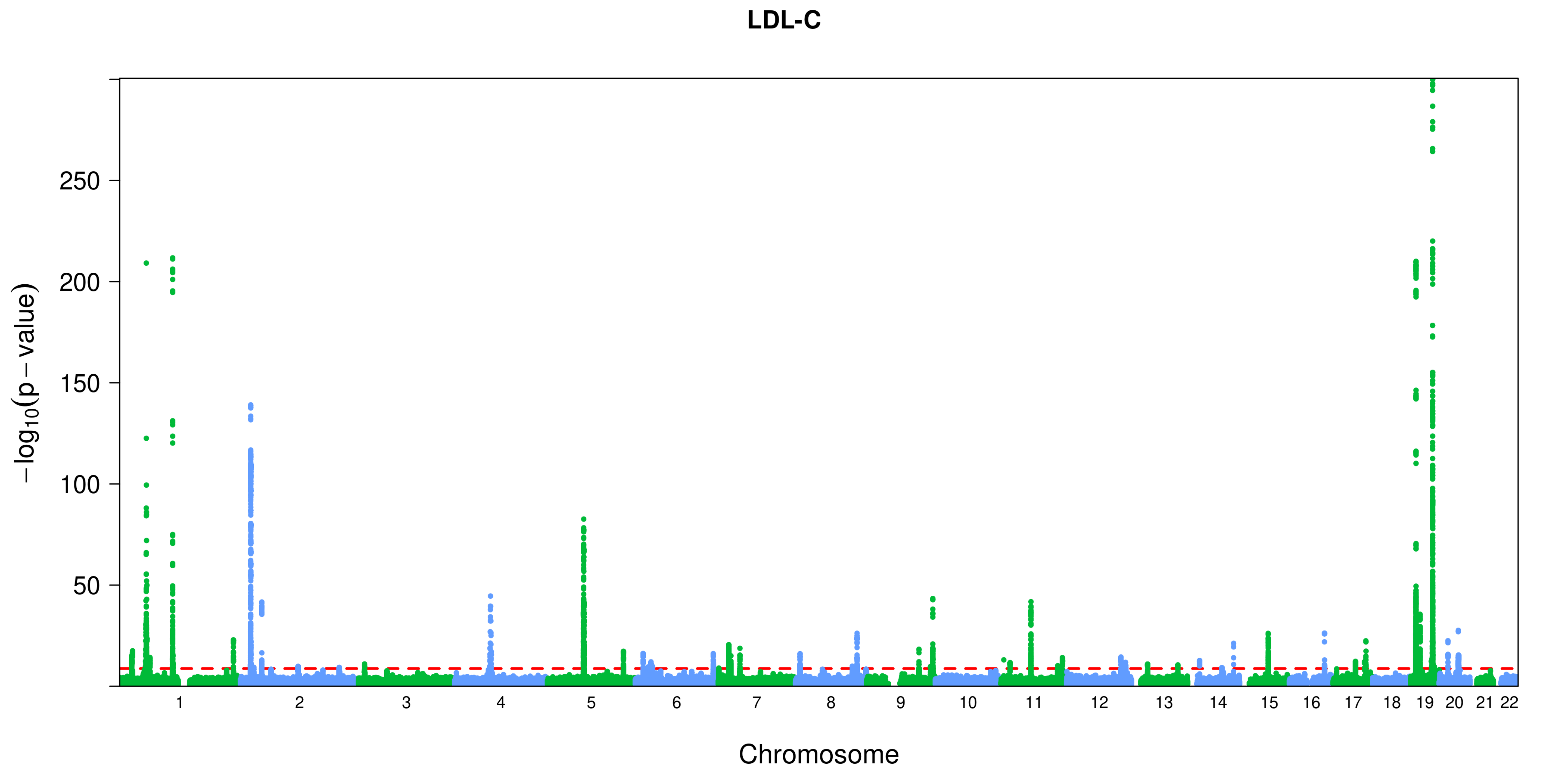

LDL-C
−log10(p − value)
250
200
150
100
50
Chromosome

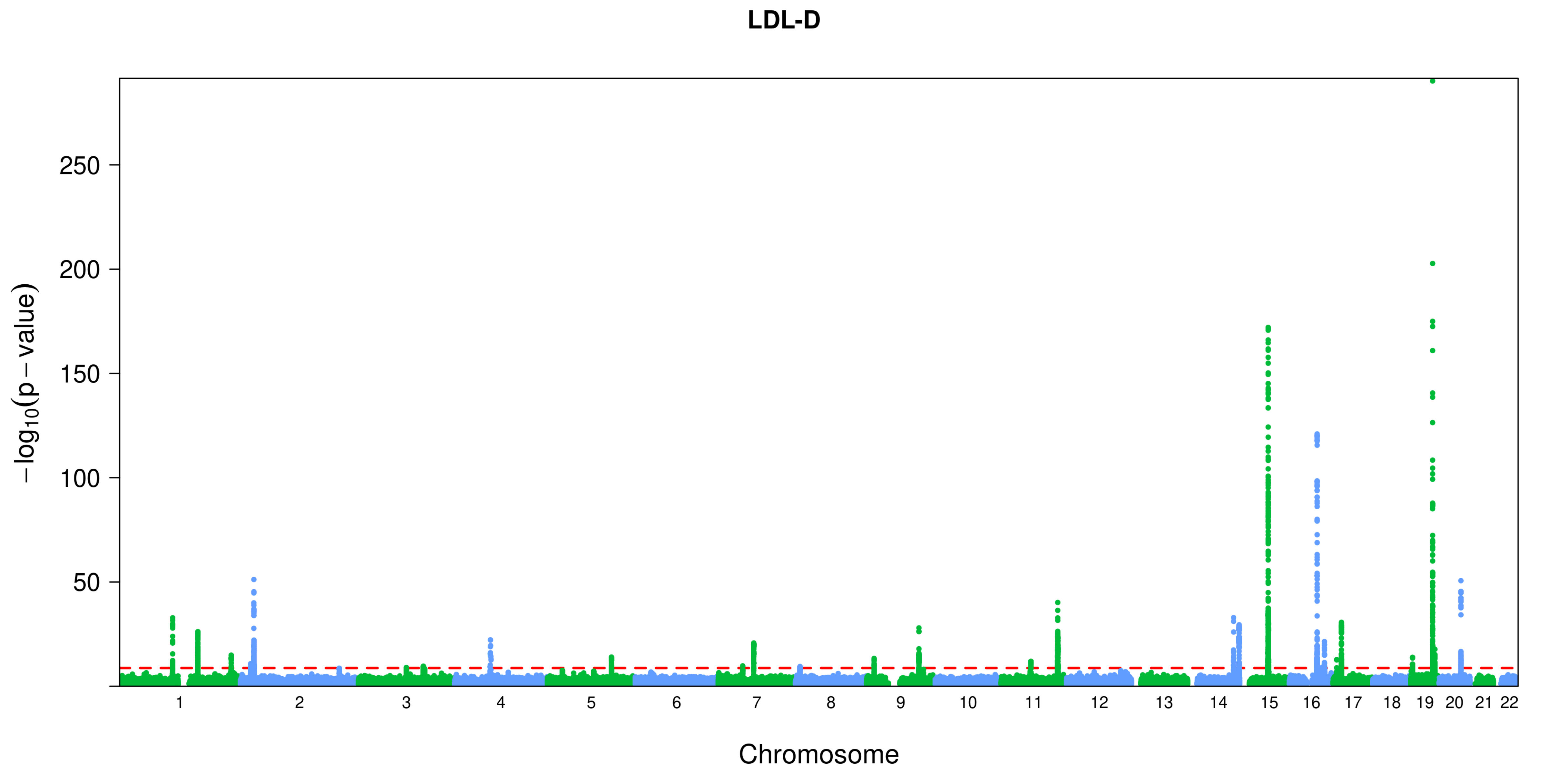

LDL-D
−log10(p − value)
250
200
150
100
50
Chromosome

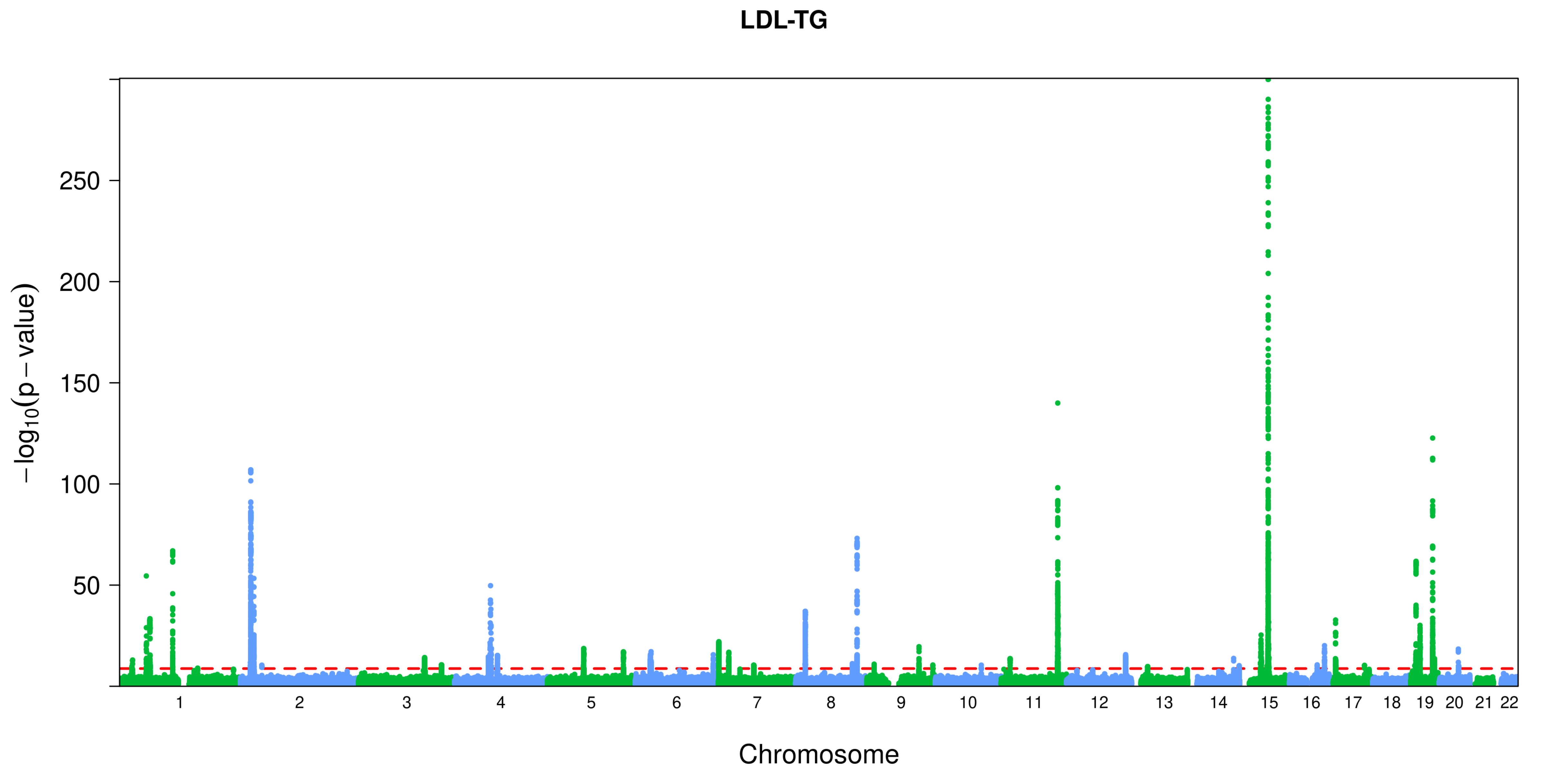

LDL-TG
−log10(p − value)
250
200
150
100
50
Chromosome

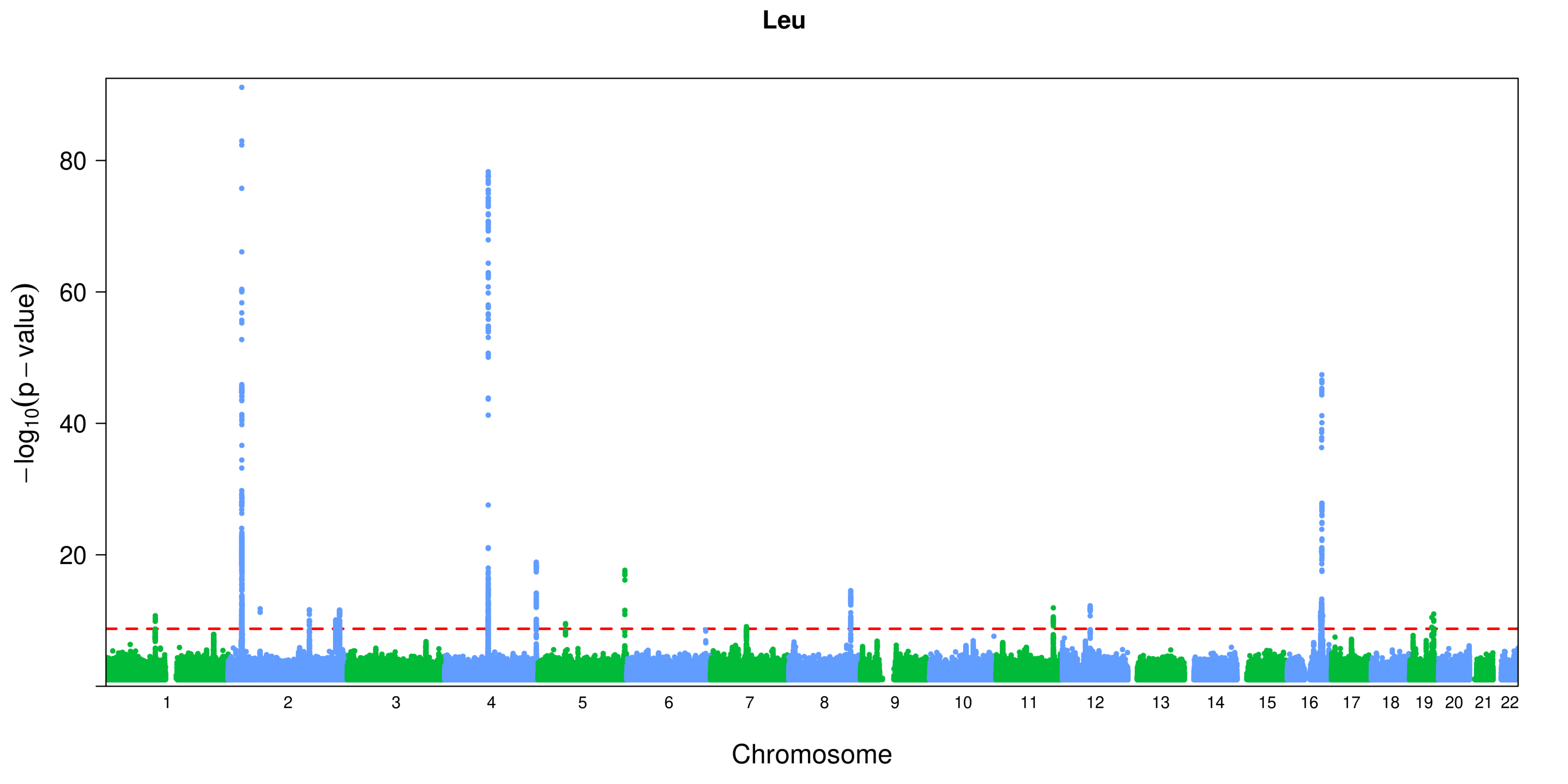
Leu
−log10(p − value)
80
60
40
20
Chromosome

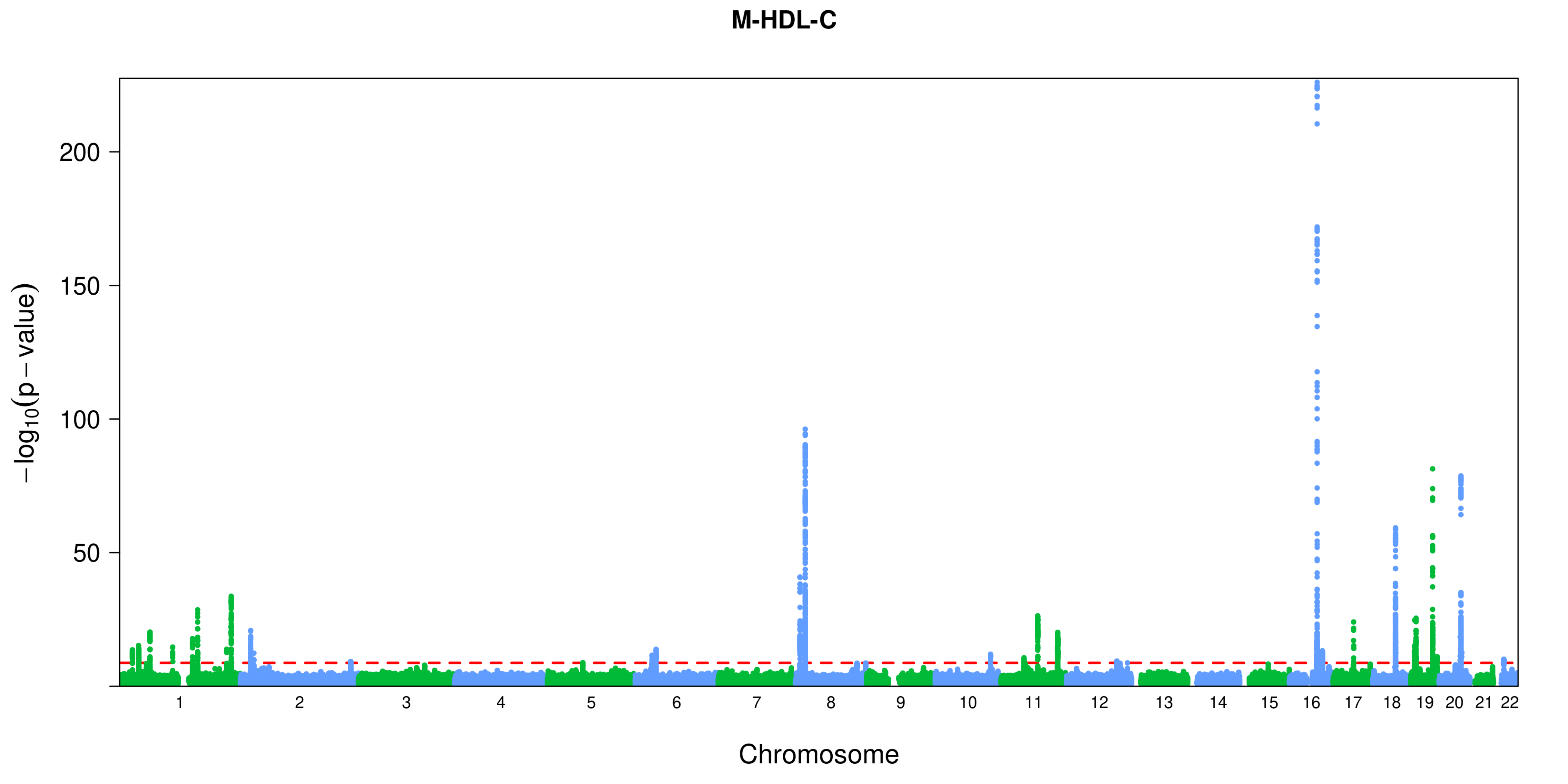

M-HDL-C
200
150
100
50
−log10(p − value)
Chromosome

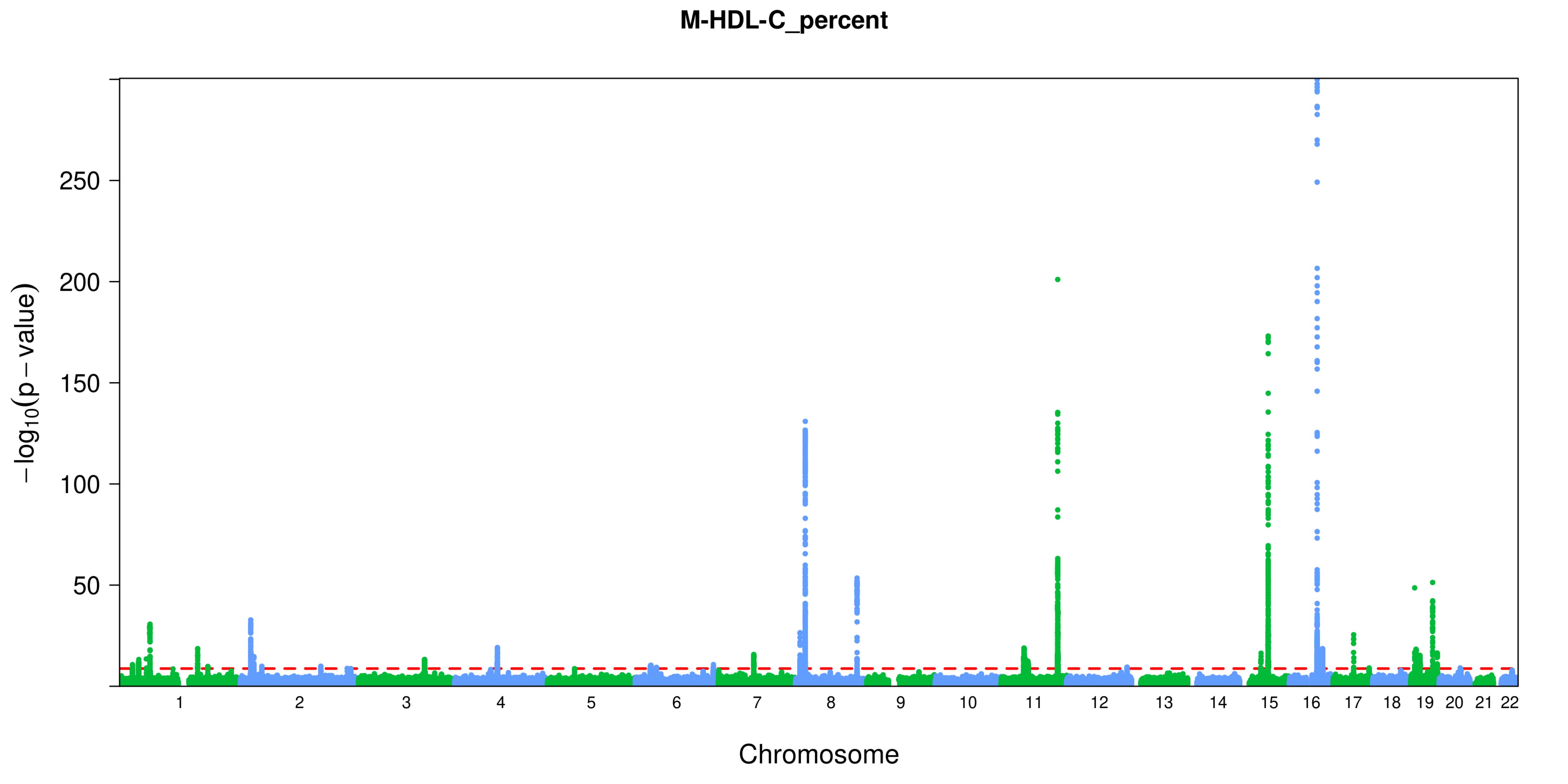
M-HDL-C_percent
−log10(p−value)
250
200
150
100
50
Chromosome

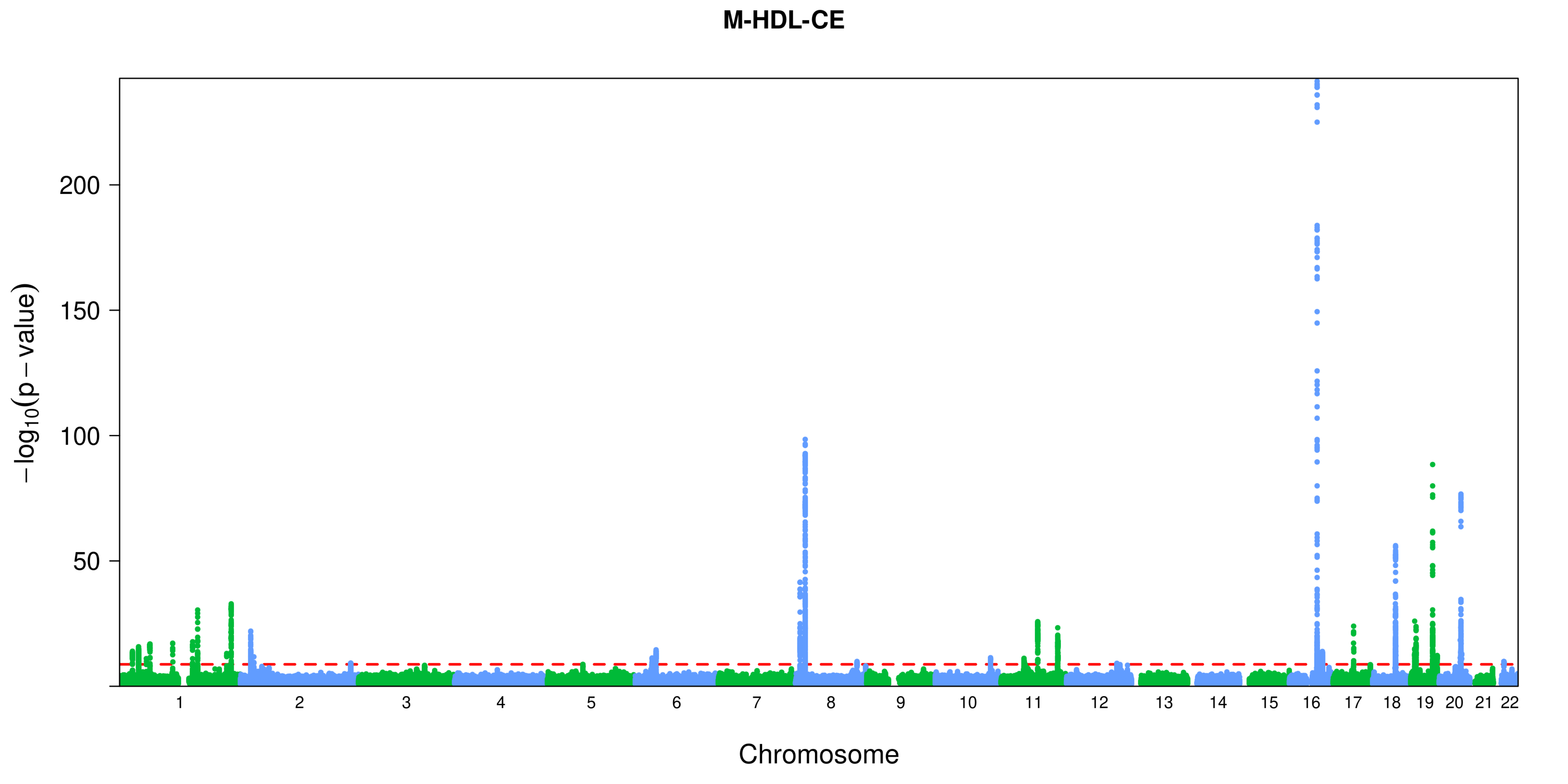
M-HDL-CE
−log10(p − value)
200
150
100
50
1 2 3 4 5 6 7 8 9 10 11 12 13 14 15 16 17 18 19 20 21 22
Chromosome

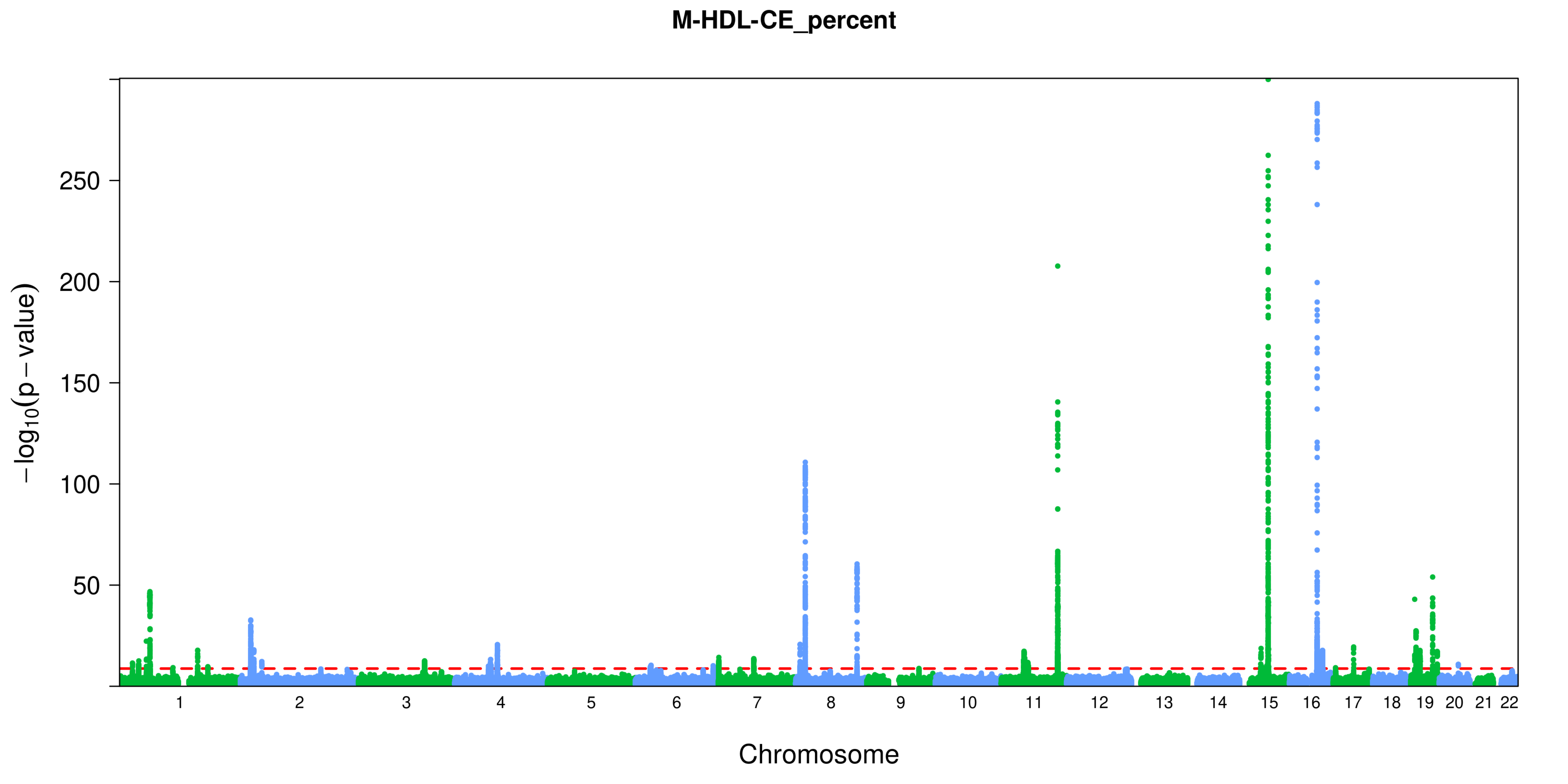
M-HDL-CE_percent
−log10(p − value)
250
200
150
100
50
Chromosome

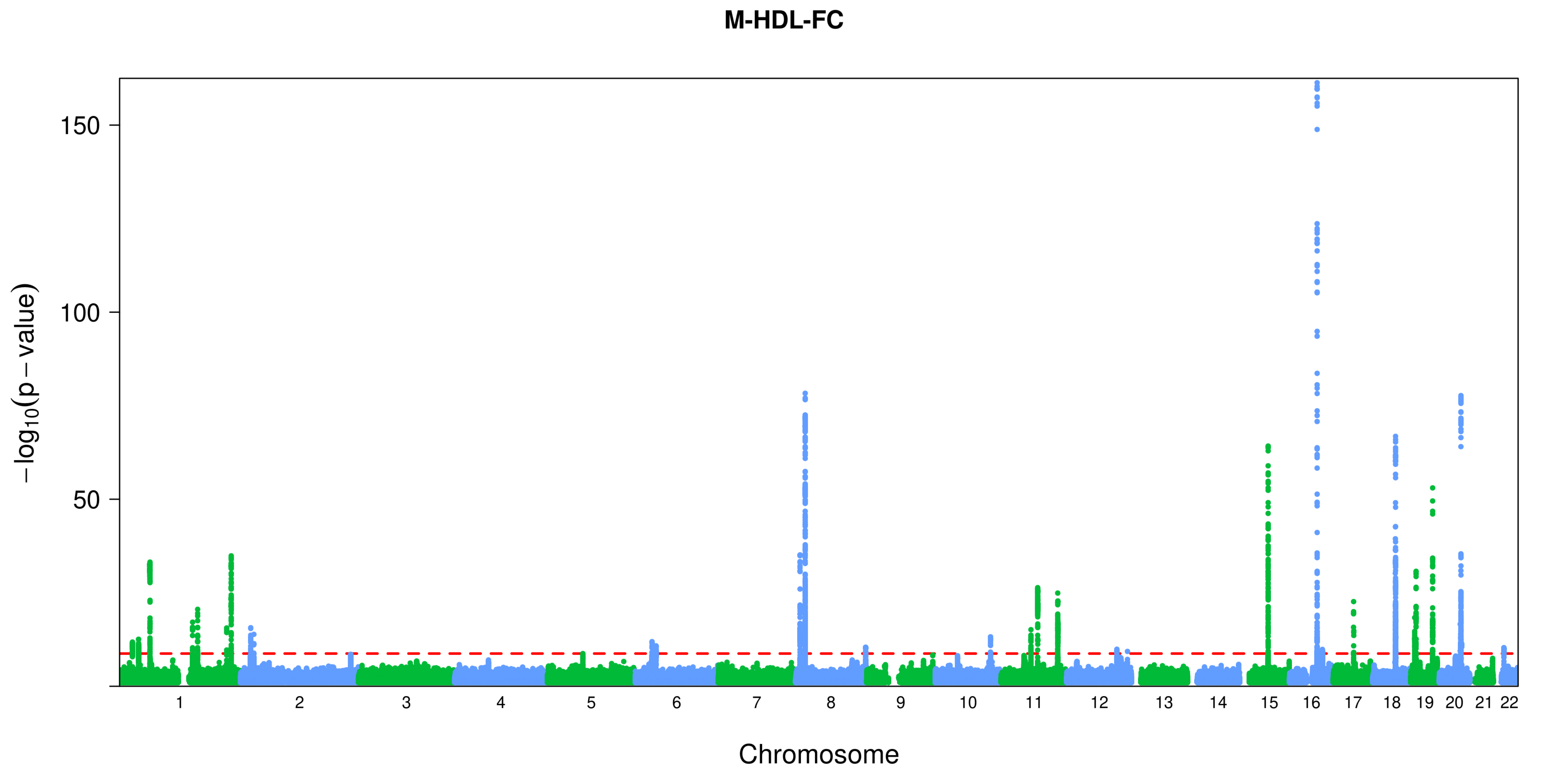

M-HDL-FC
150
100
50
−log10(p − value)
Chromosome

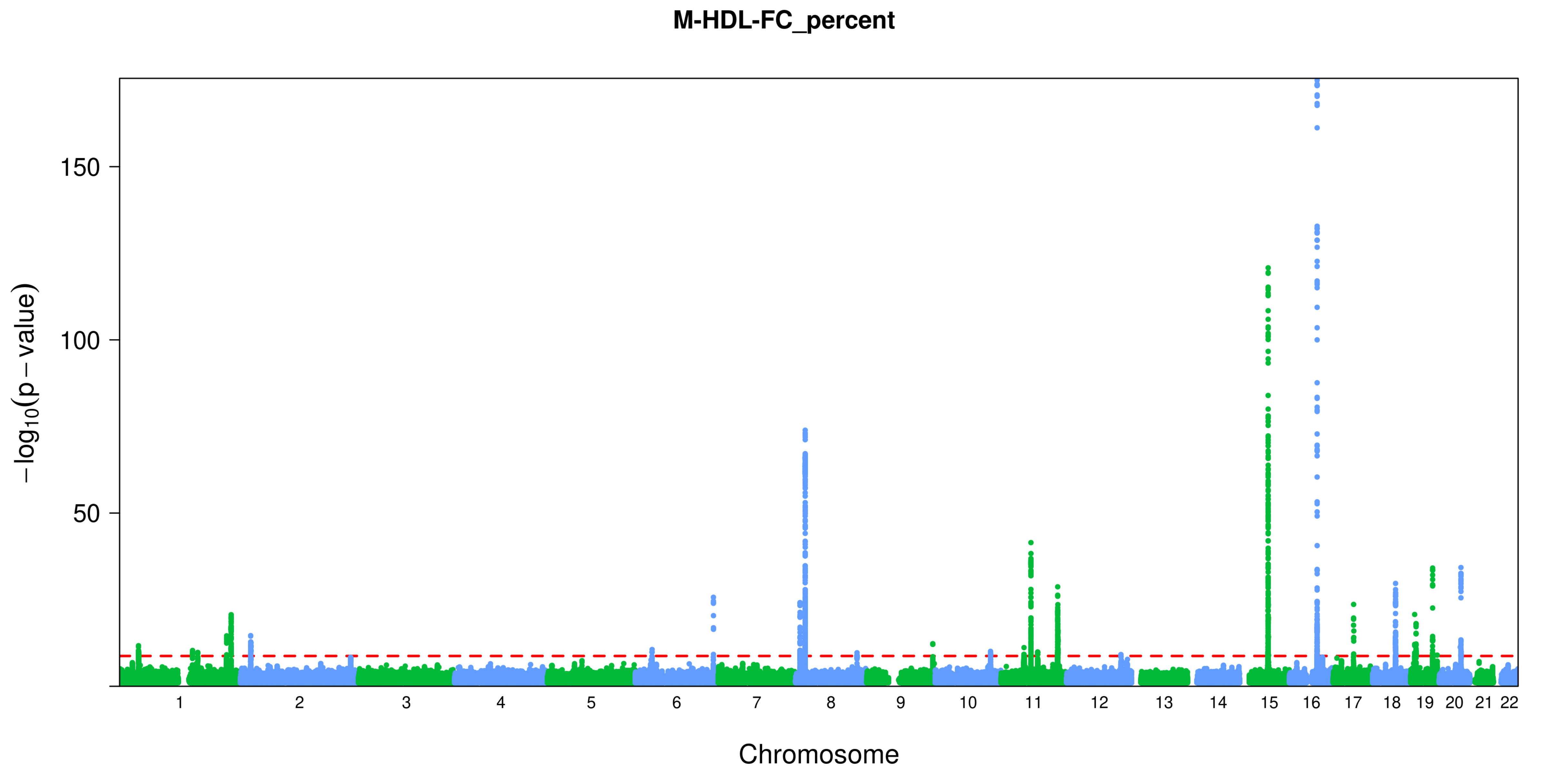
M-HDL-FC_percent
−log10(p − value)
150
100
50
Chromosome

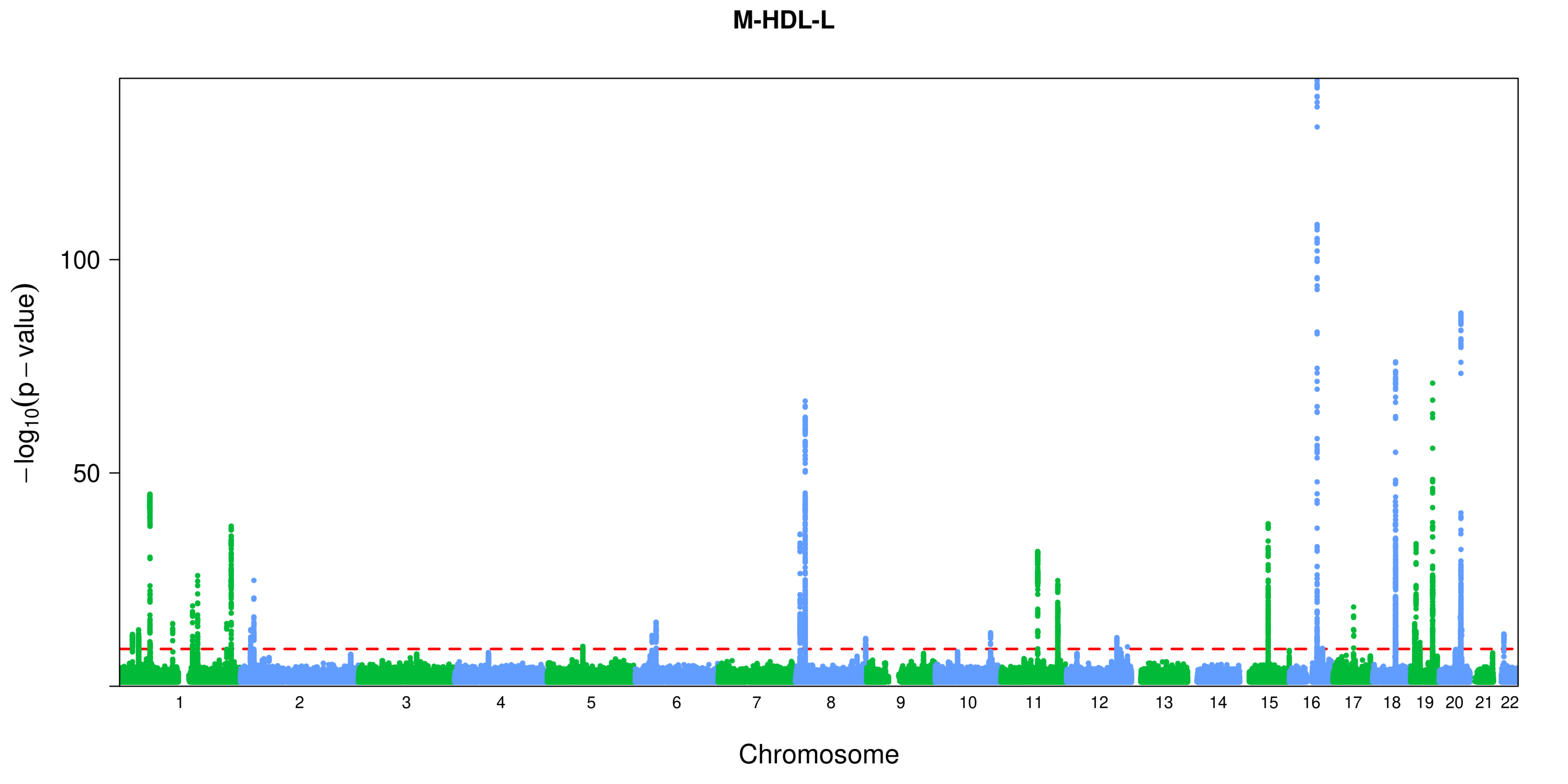
M-HDL-L
100
50
−log10(p − value)
Chromosome

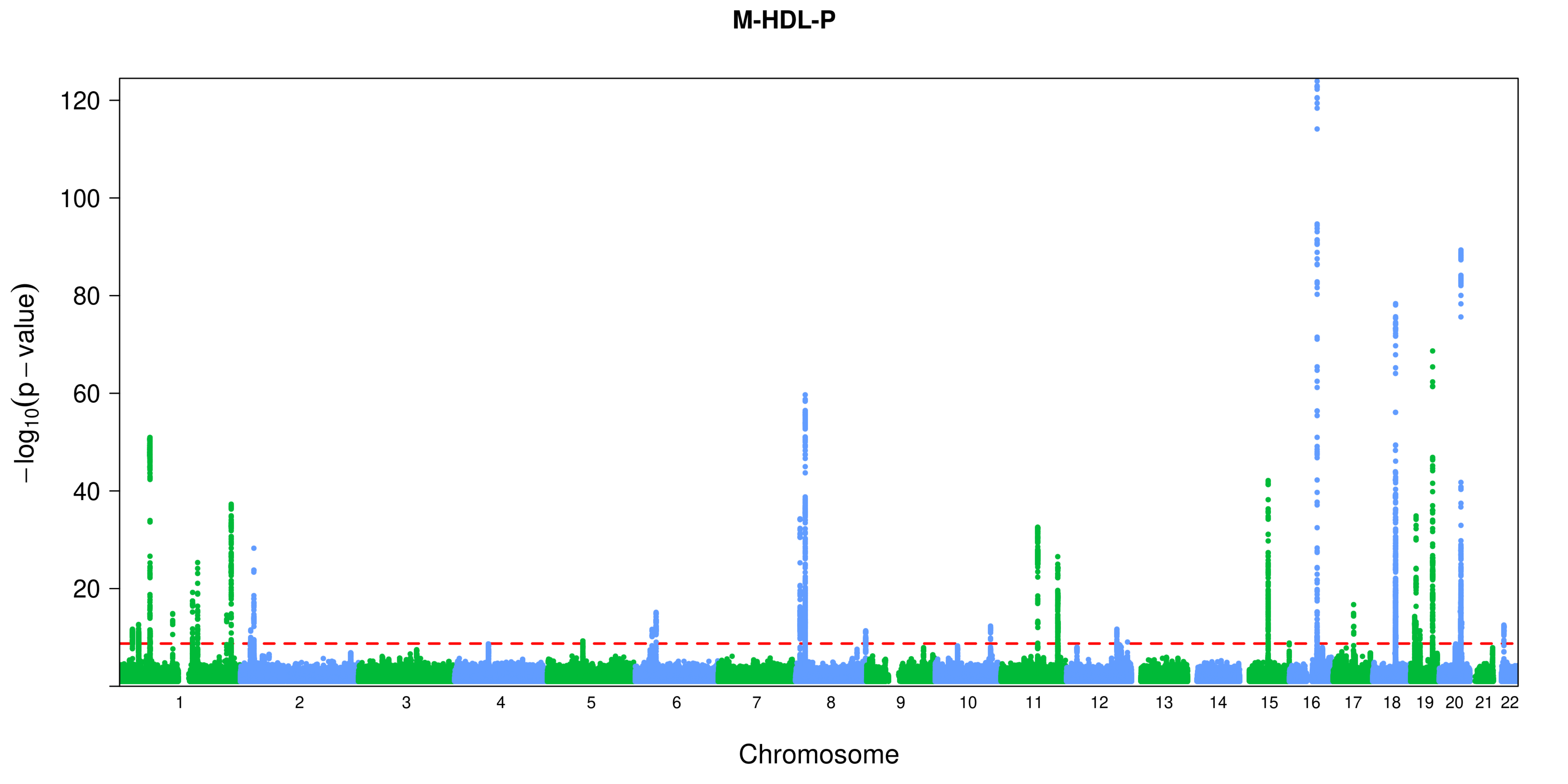
M-HDL-P
120
100
80
60
40
20
−log10(p − value)
Chromosome

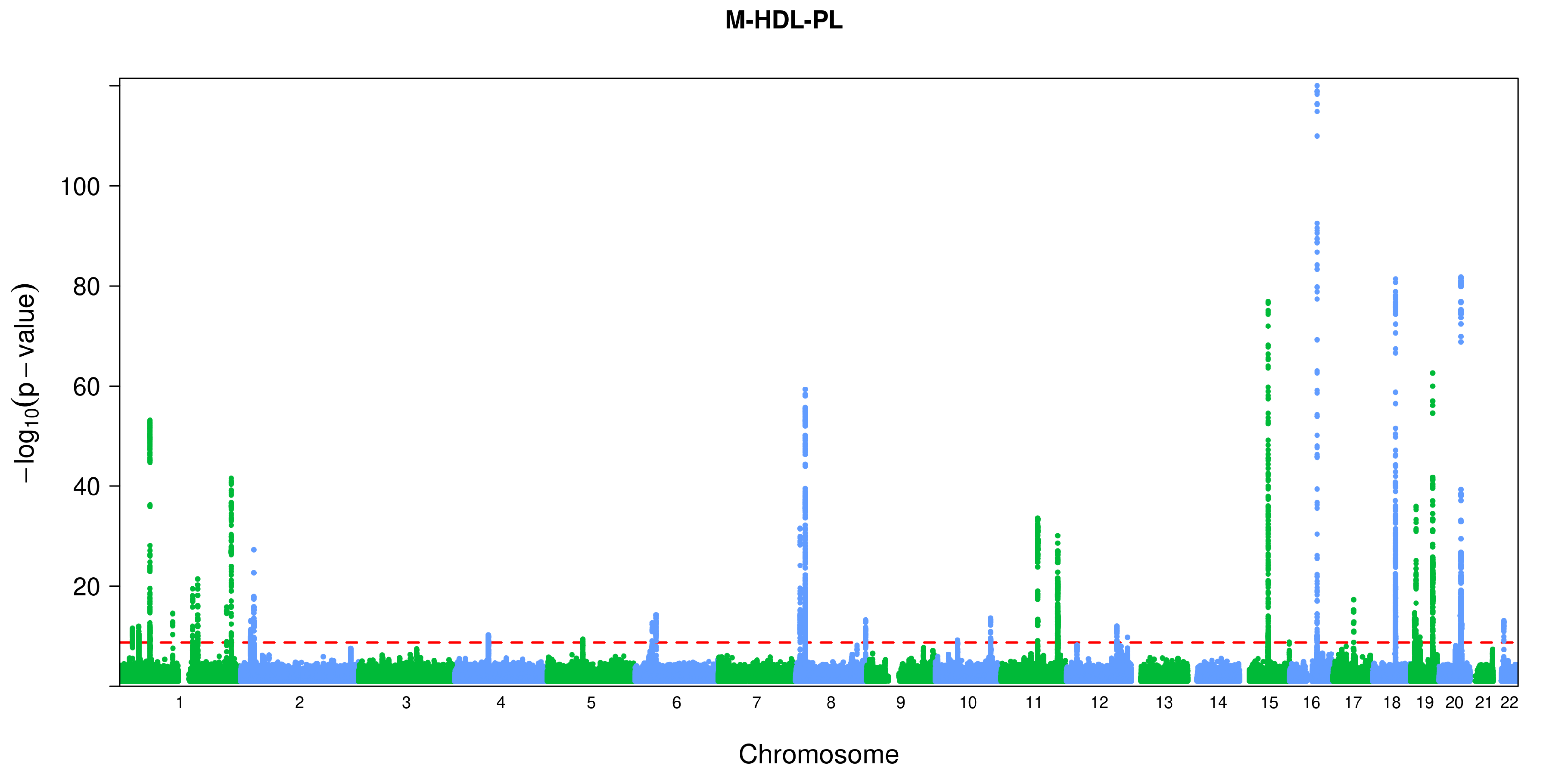
M-HDL-PL
−log10(p − value)
100
80
60
40
20
Chromosome

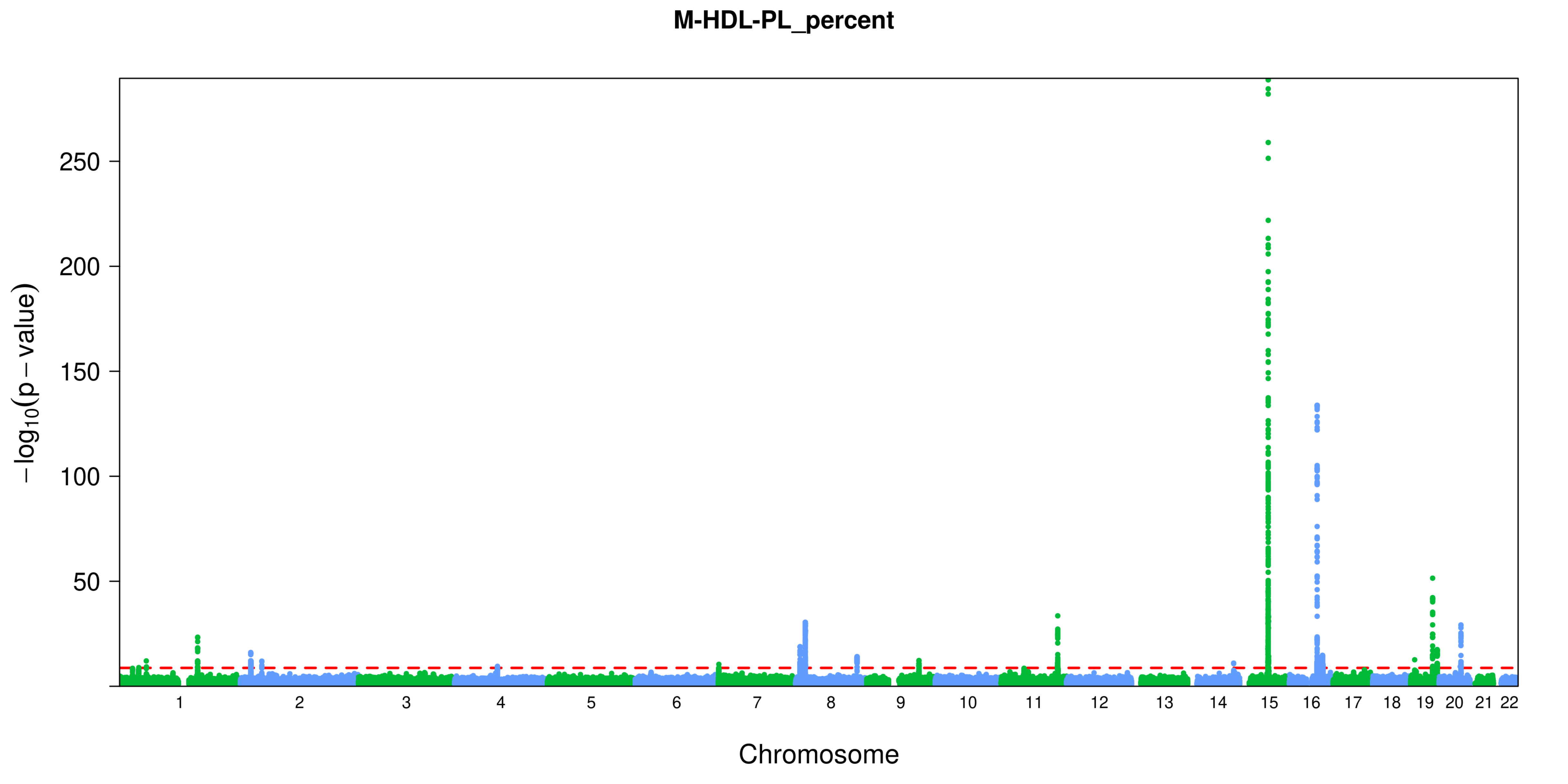
M-HDL-PL_percent
$-\log_{10}(p-\text{value})$
250
200
150
100
50
Chromosome

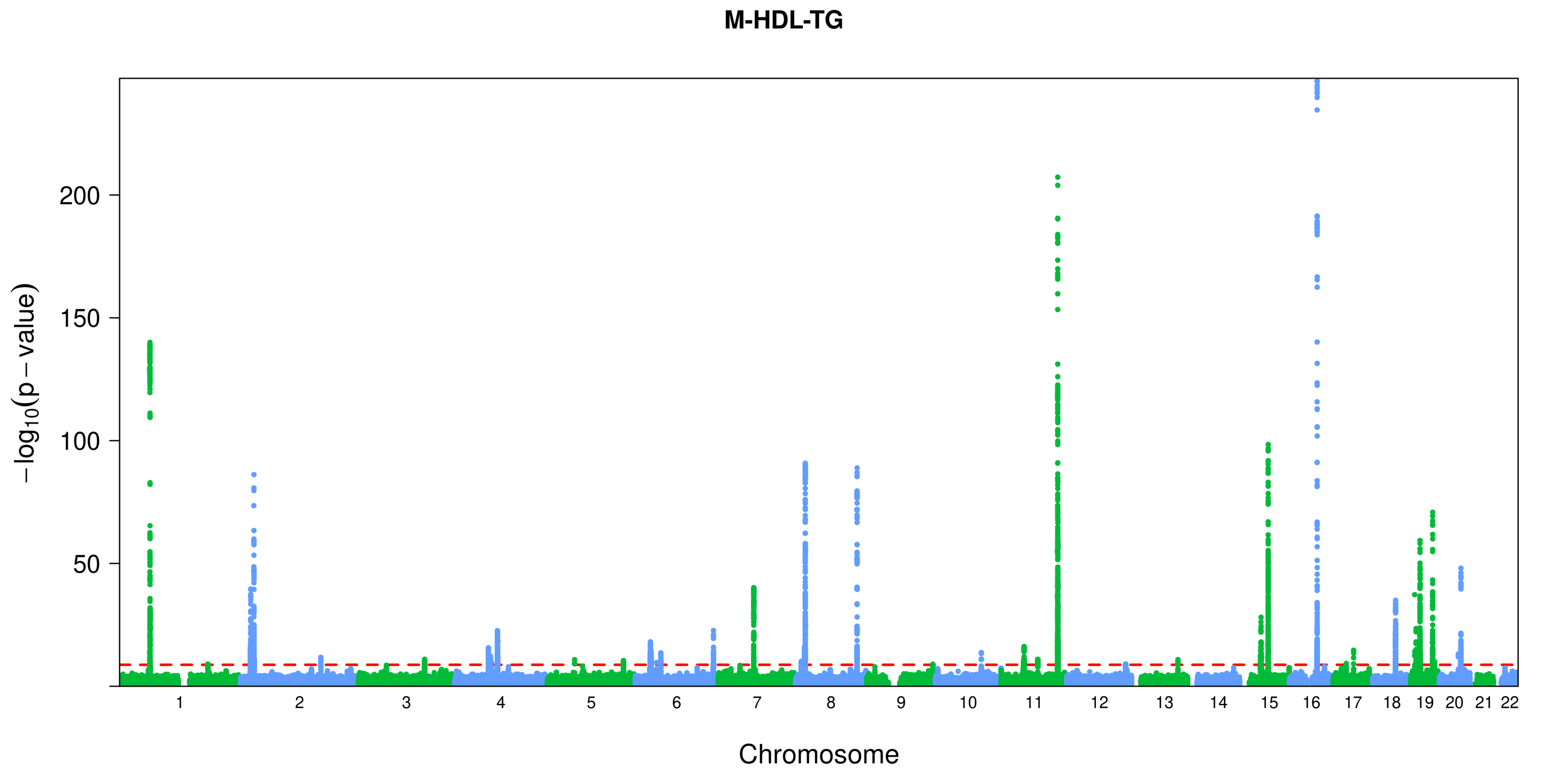
M-HDL-TG
−log10(p − value)
200
150
100
50
Chromosome

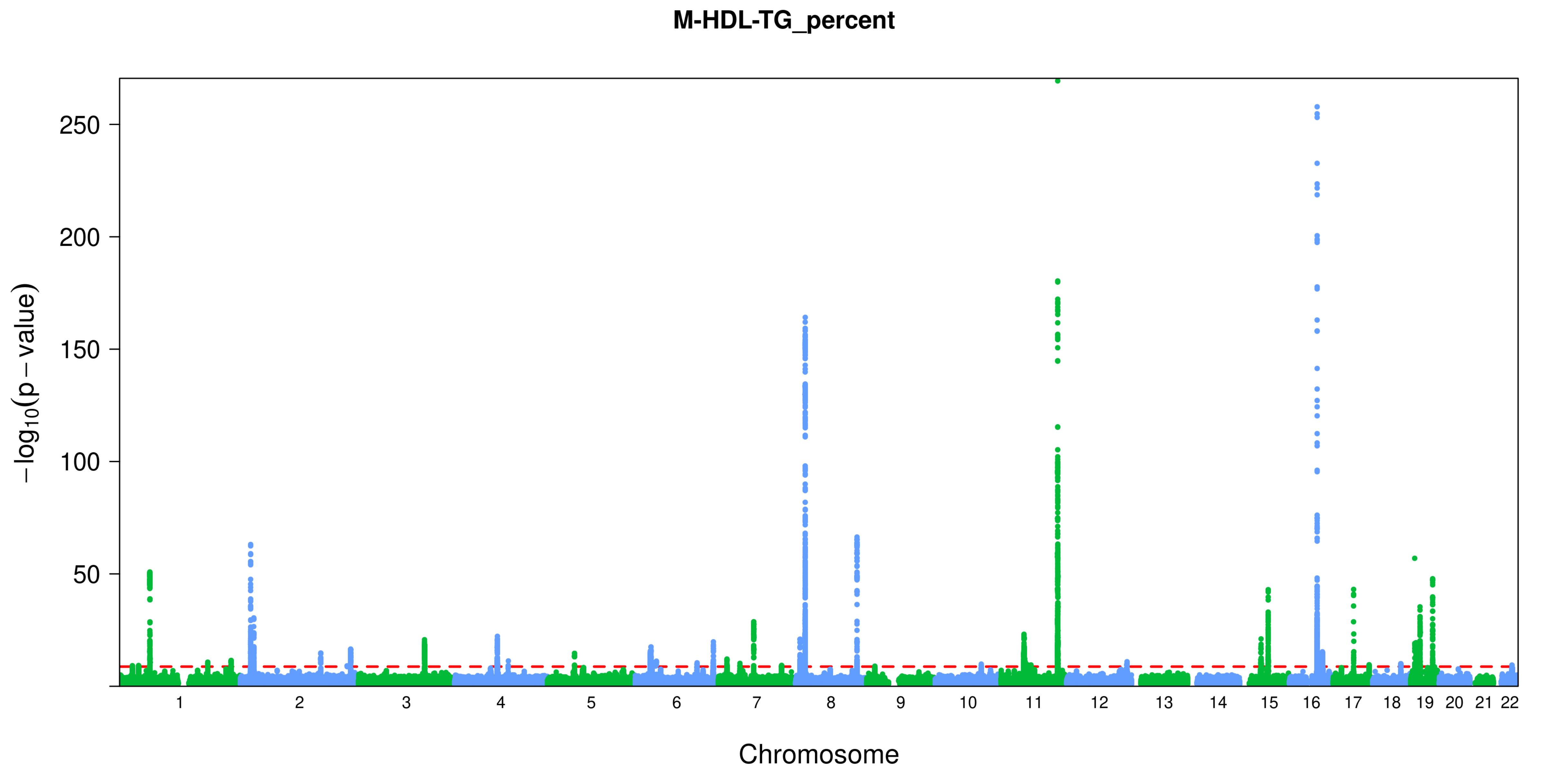

M-HDL-TG_percent
−log10(p − value)
250
200
150
100
50
Chromosome

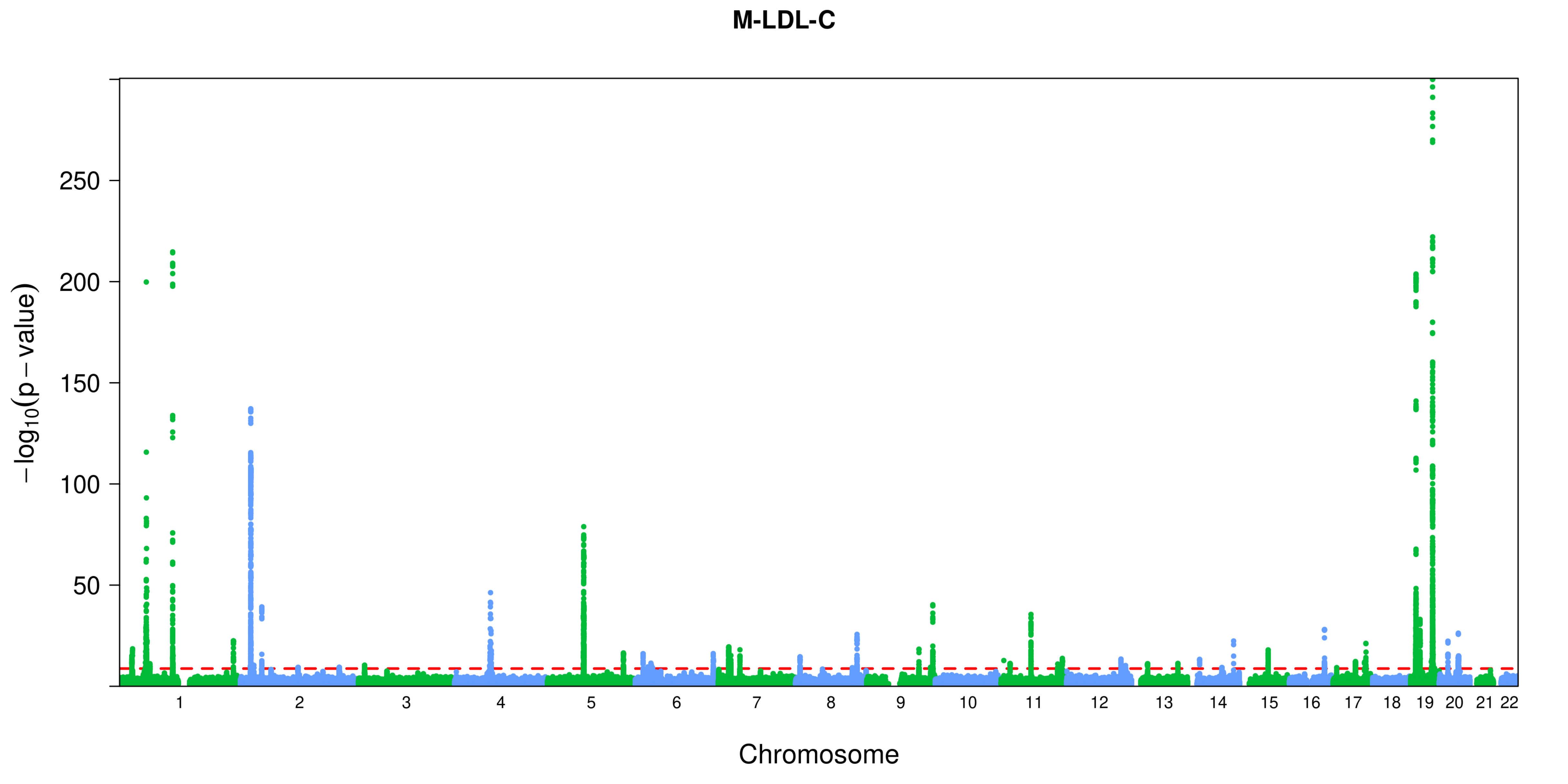
M-LDL-C
−log10(p − value)
250
200
150
100
50
Chromosome

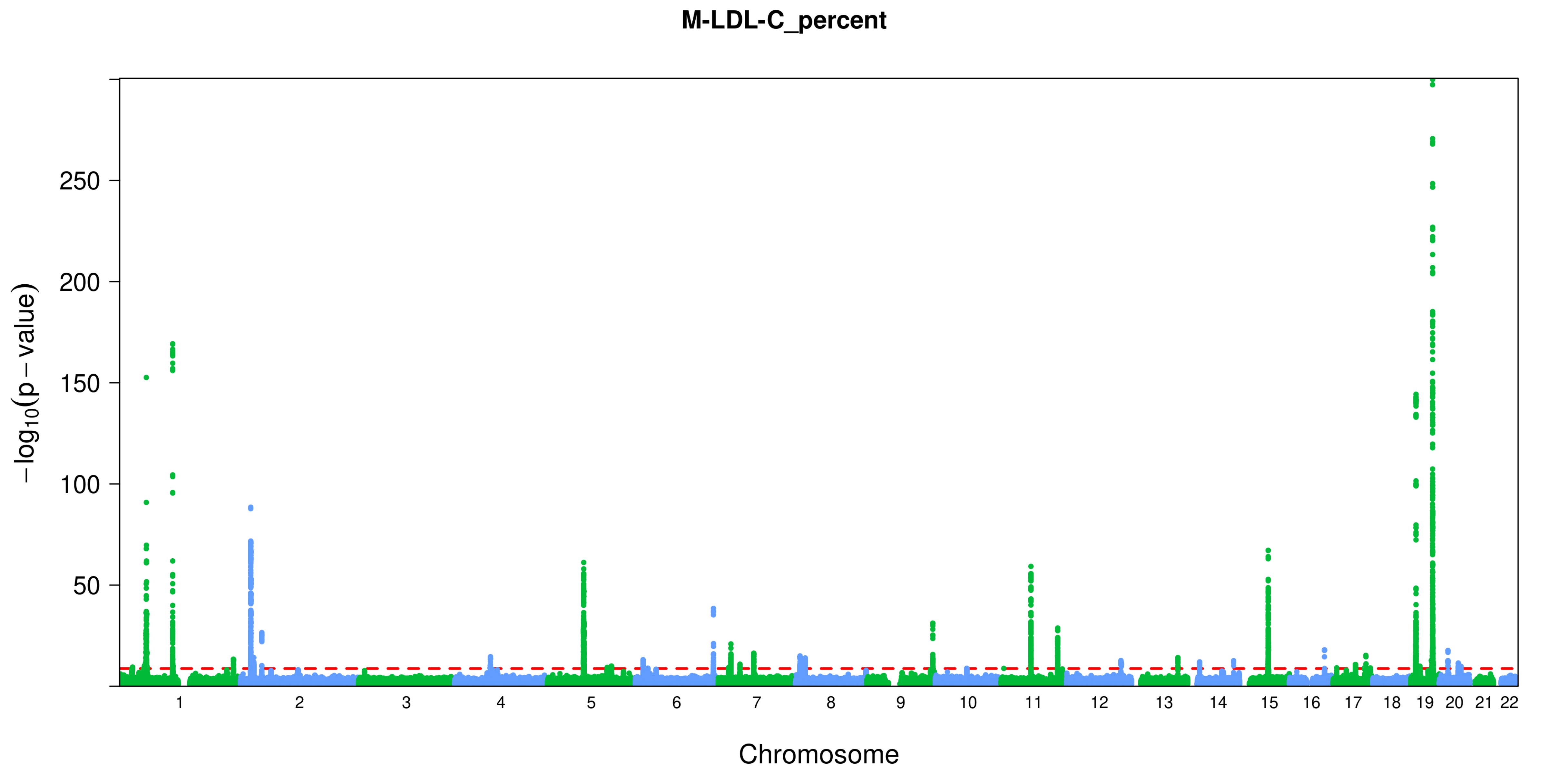

M-LDL-C_percent
250
200
150
100
50
−log10(p − value)
Chromosome

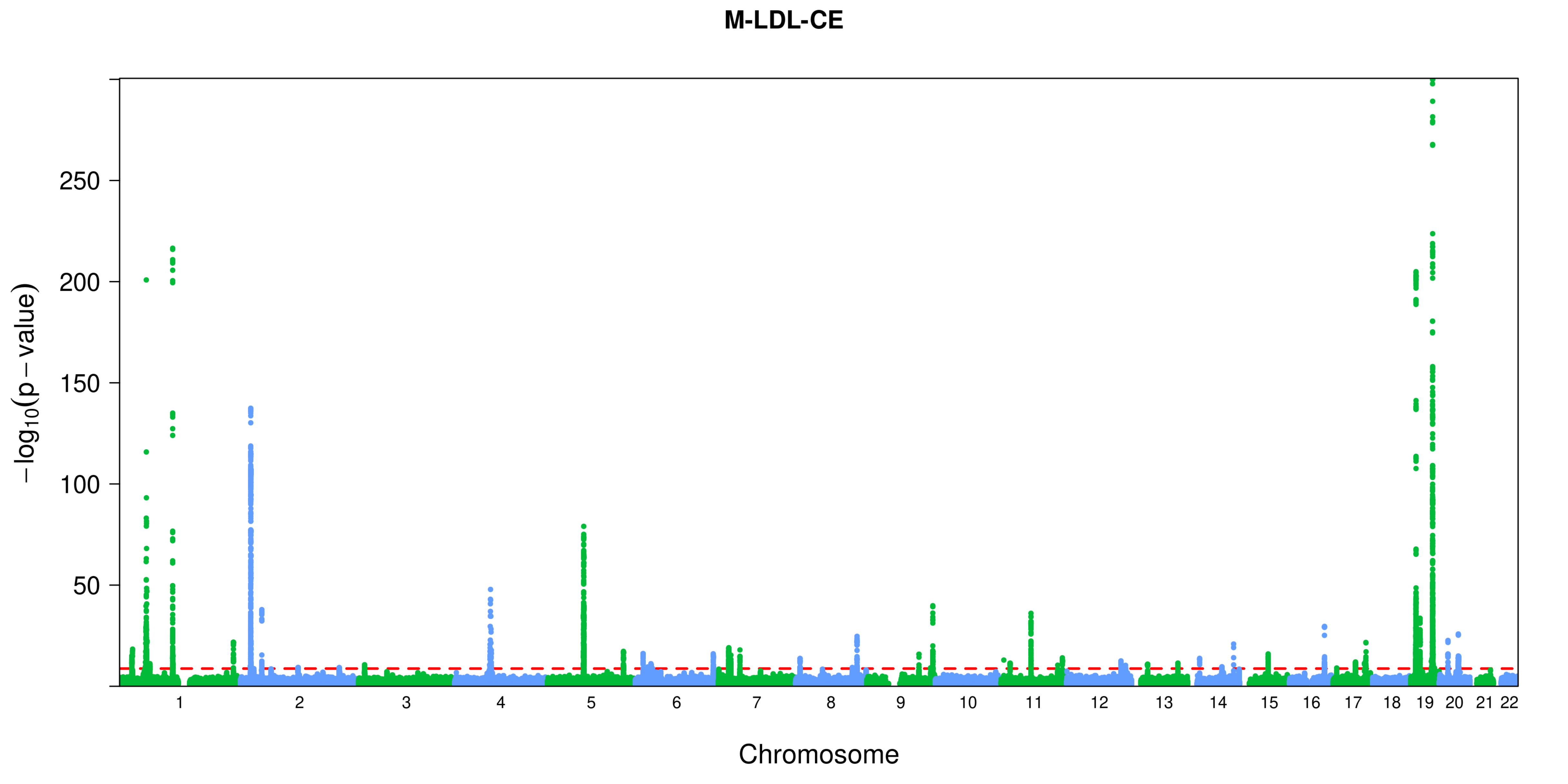
M-LDL-CE
250
200
150
100
50
−log10(p − value)
Chromosome

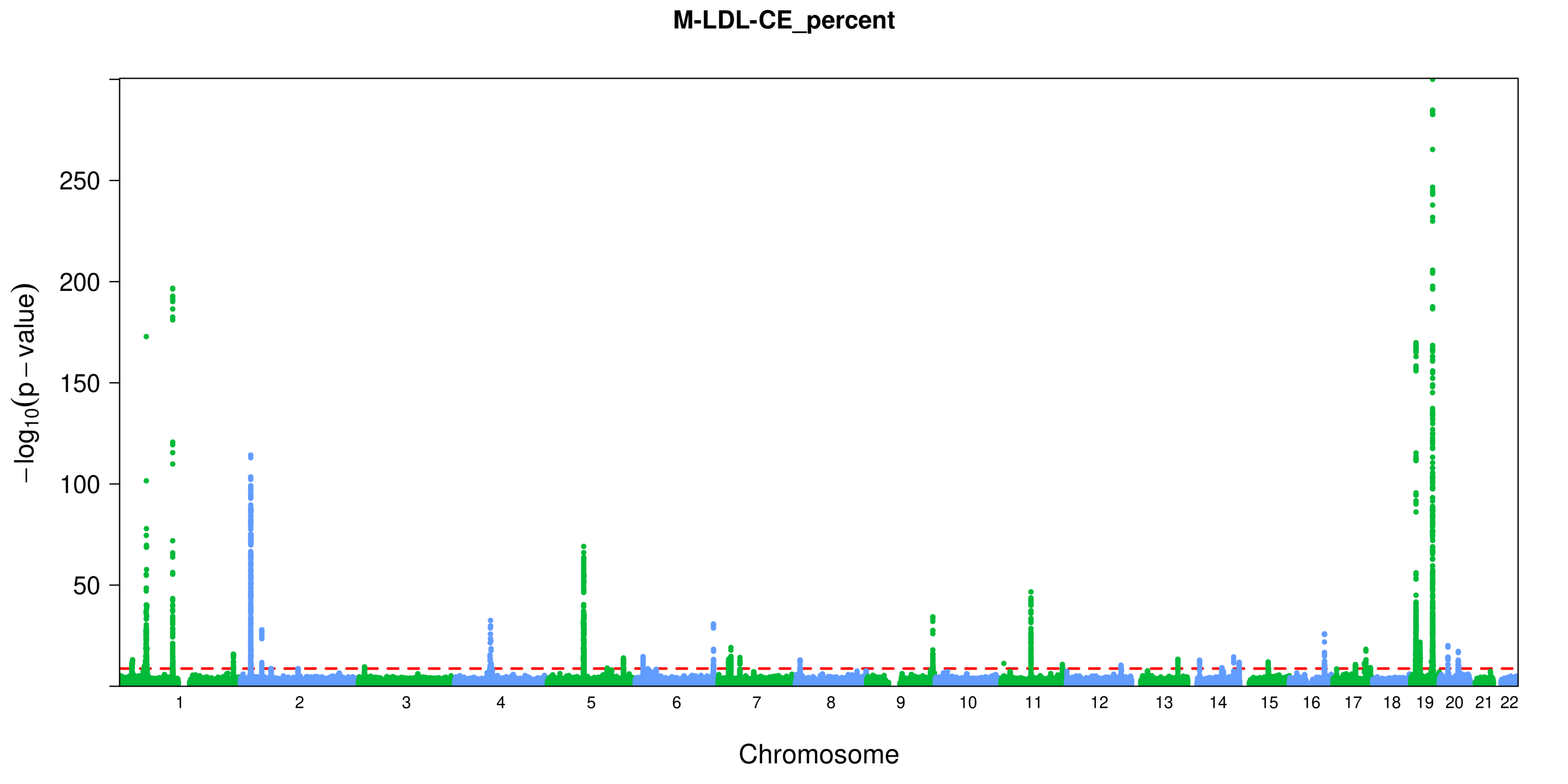
M-LDL-CE_percent
−log10(p − value)
250
200
150
100
50
Chromosome

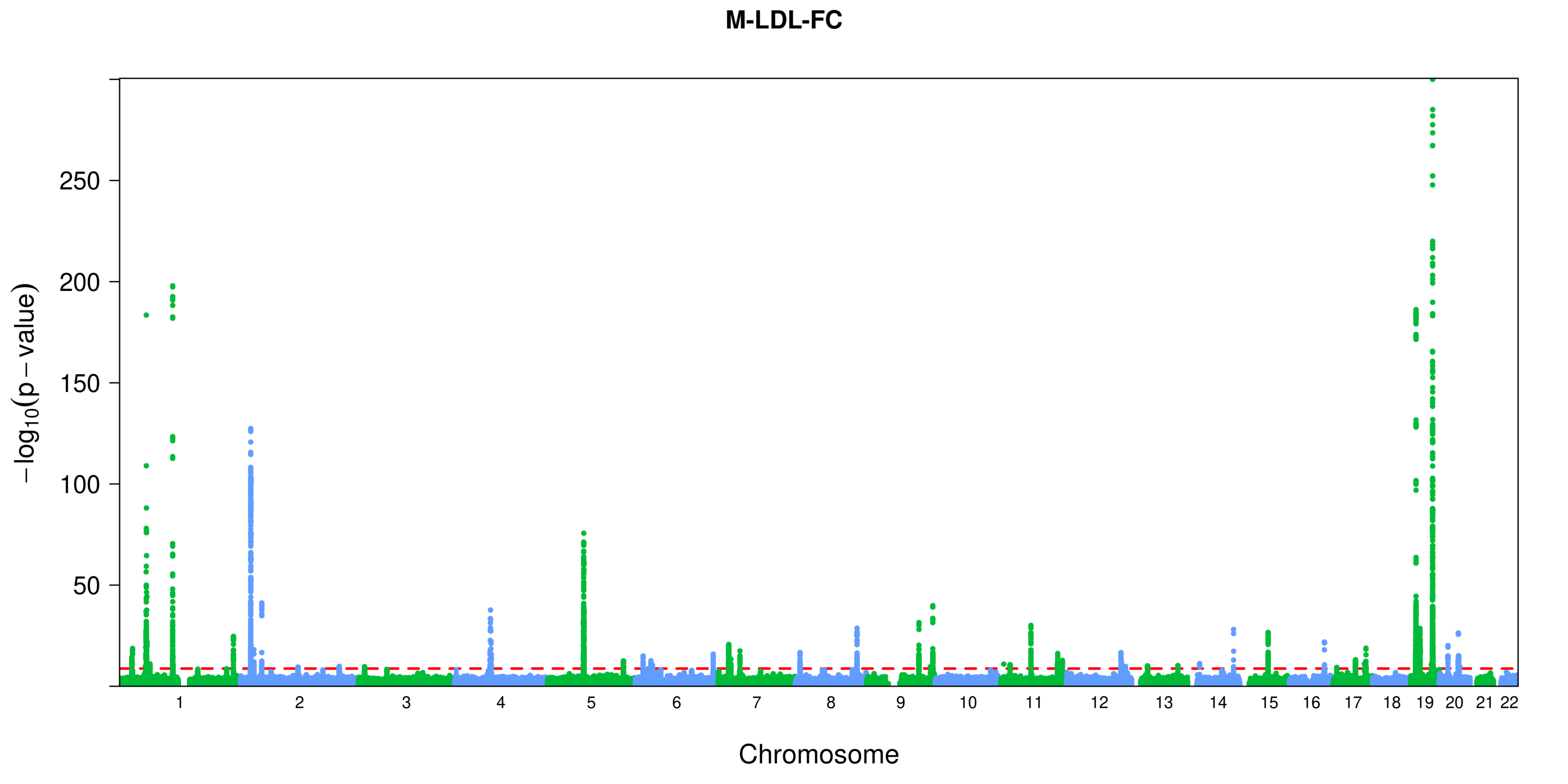

M-LDL-FC
250
200
150
100
50
−log10(p − value)
Chromosome

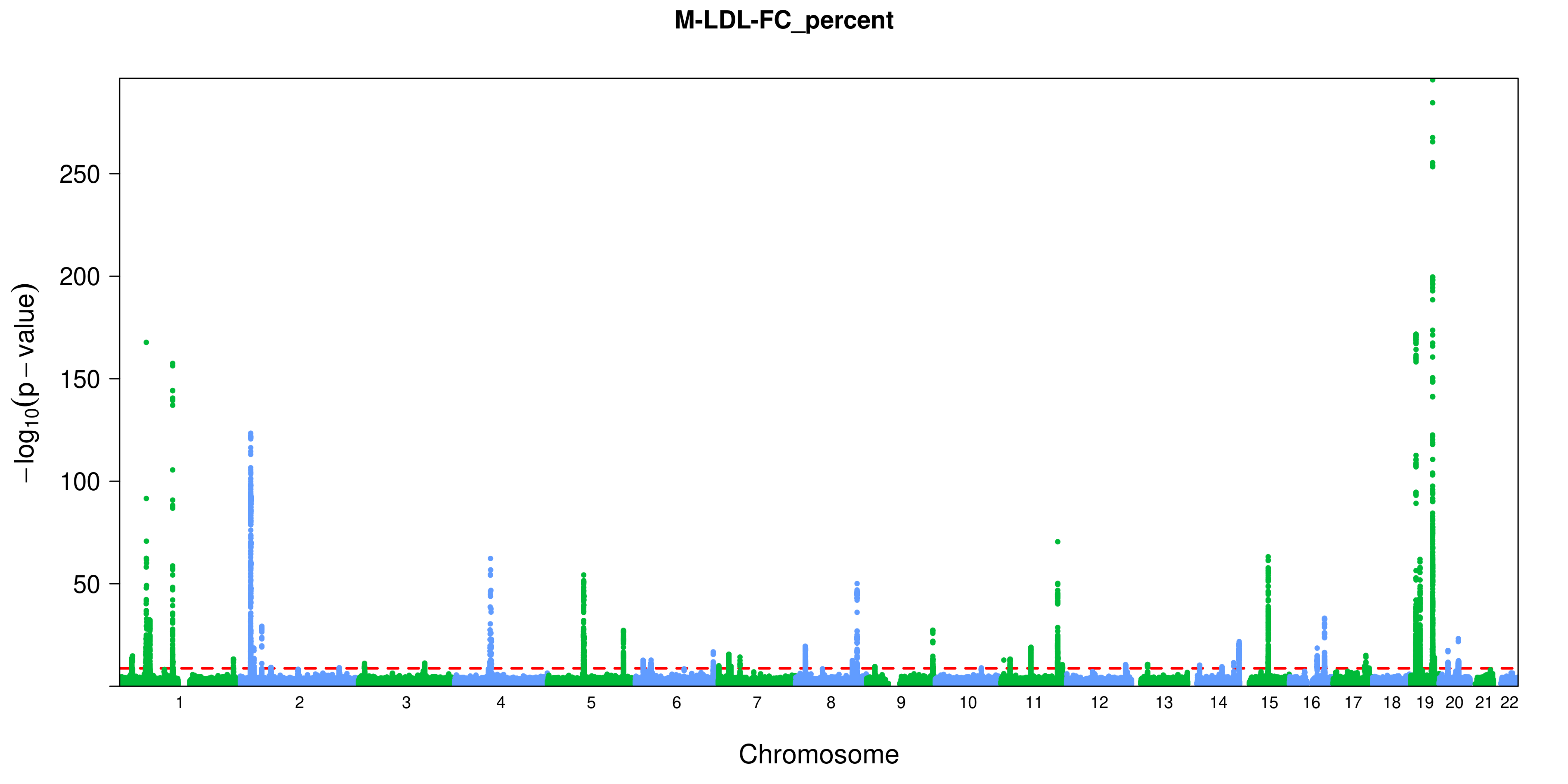
M-LDL-FC_percent
−log10(p − value)
250
200
150
100
50
Chromosome

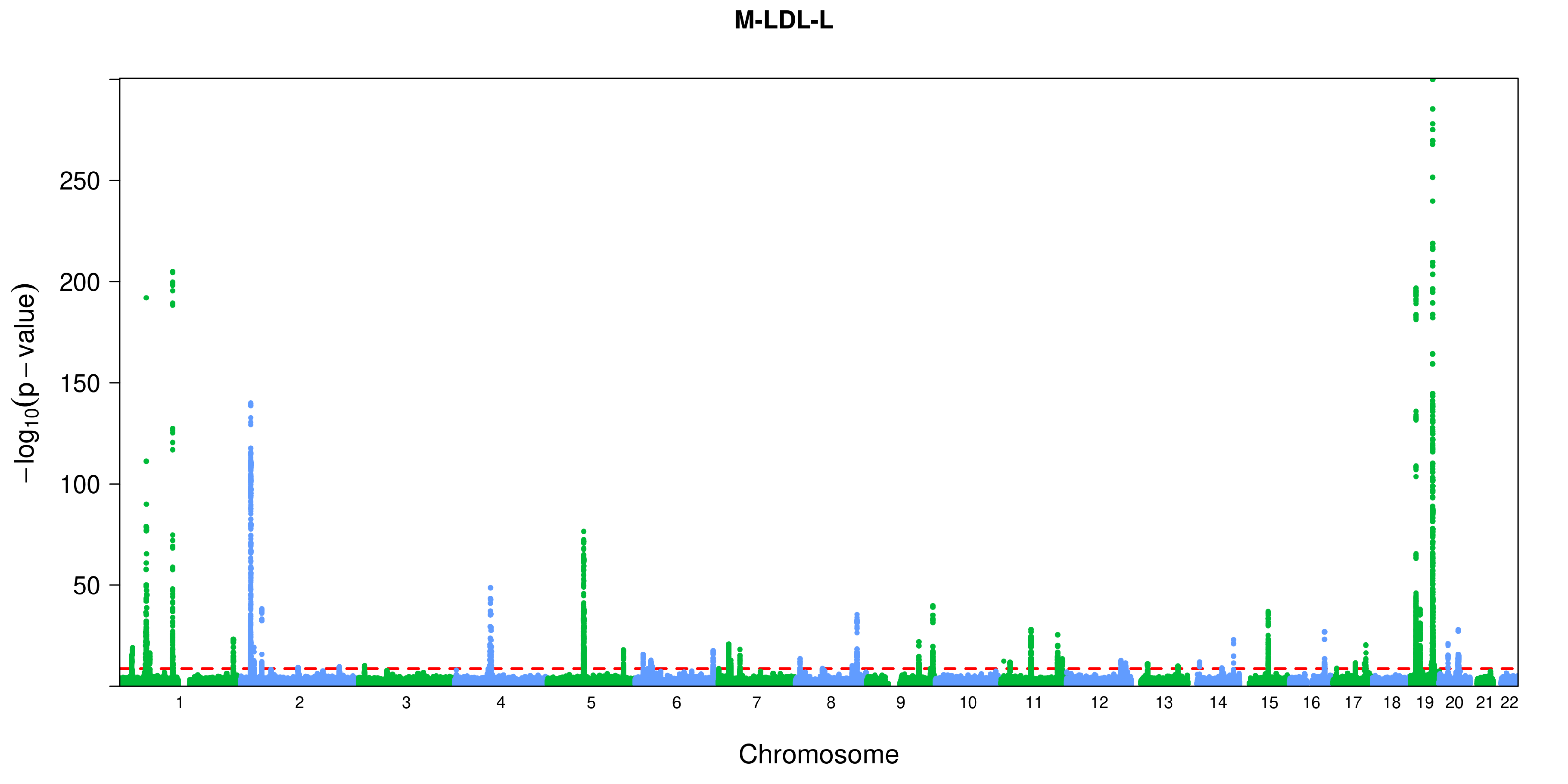

M-LDL-L
250
200
150
100
50
$-\log_{10}(p-\text{value})$
Chromosome

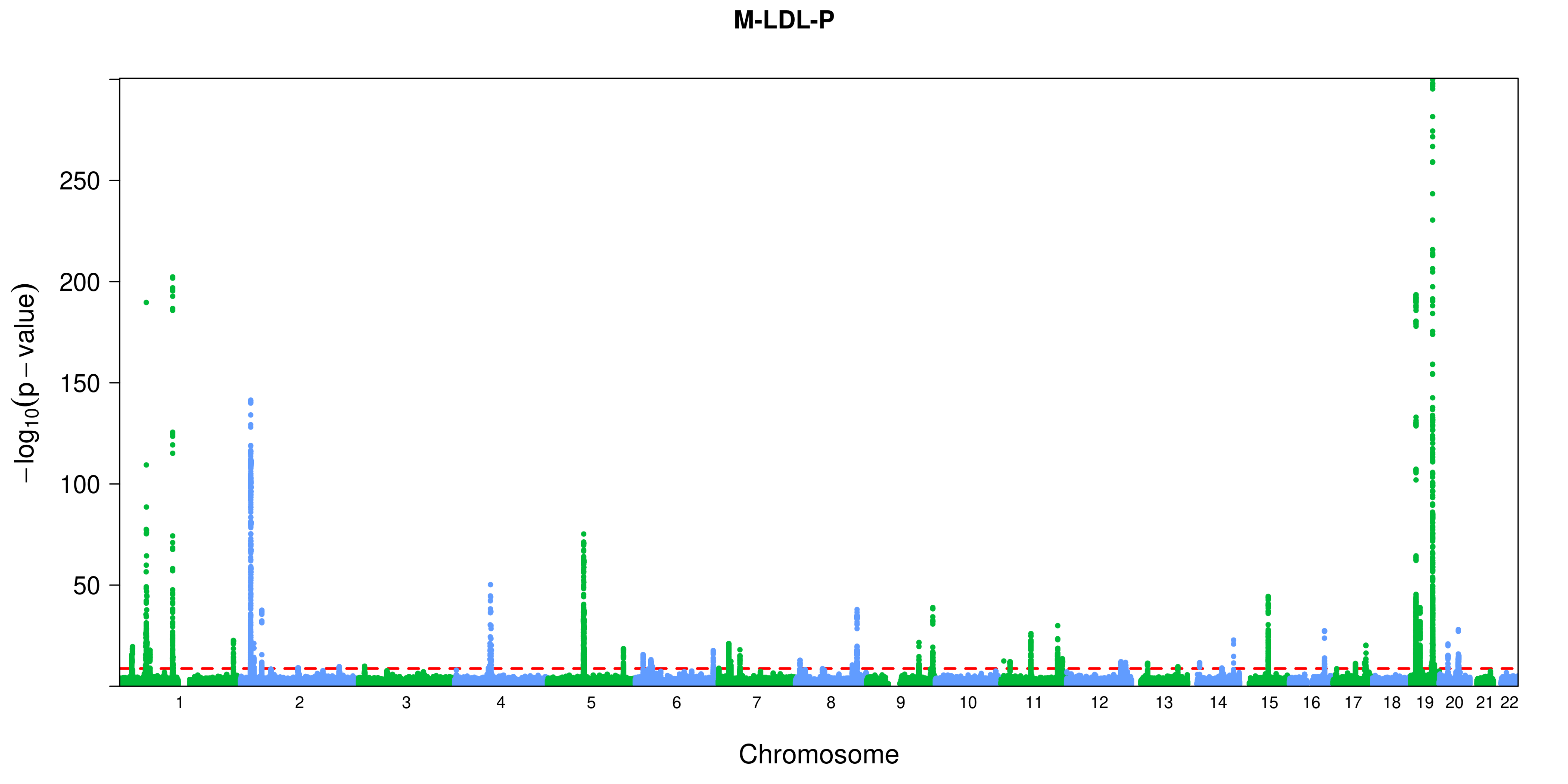
M-LDL-P
−log10(p − value)
250
200
150
100
50
Chromosome

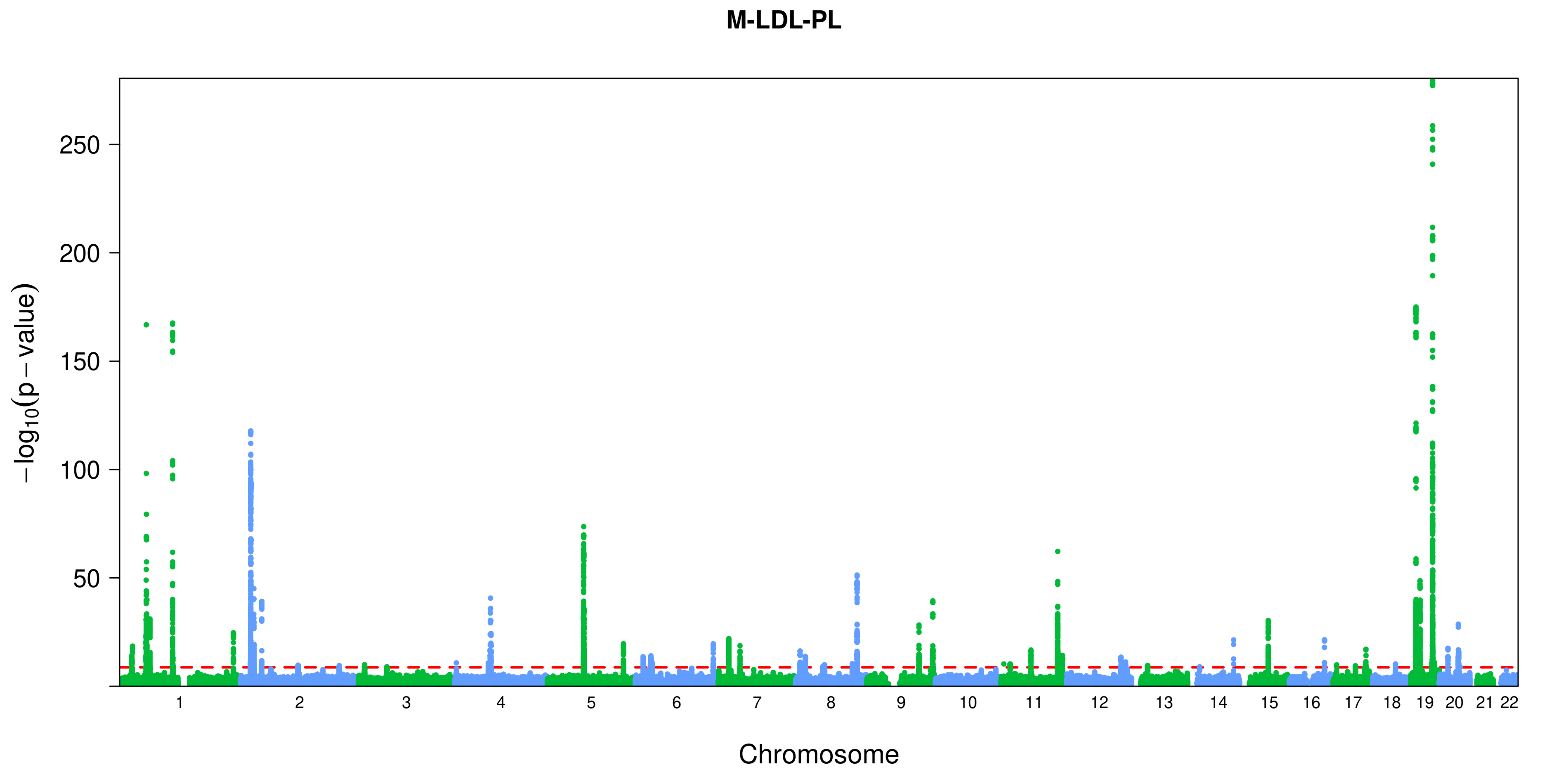
M-LDL-PL
250
200
150
100
50
−log10(p − value)
Chromosome

M-LDL-PL_percent

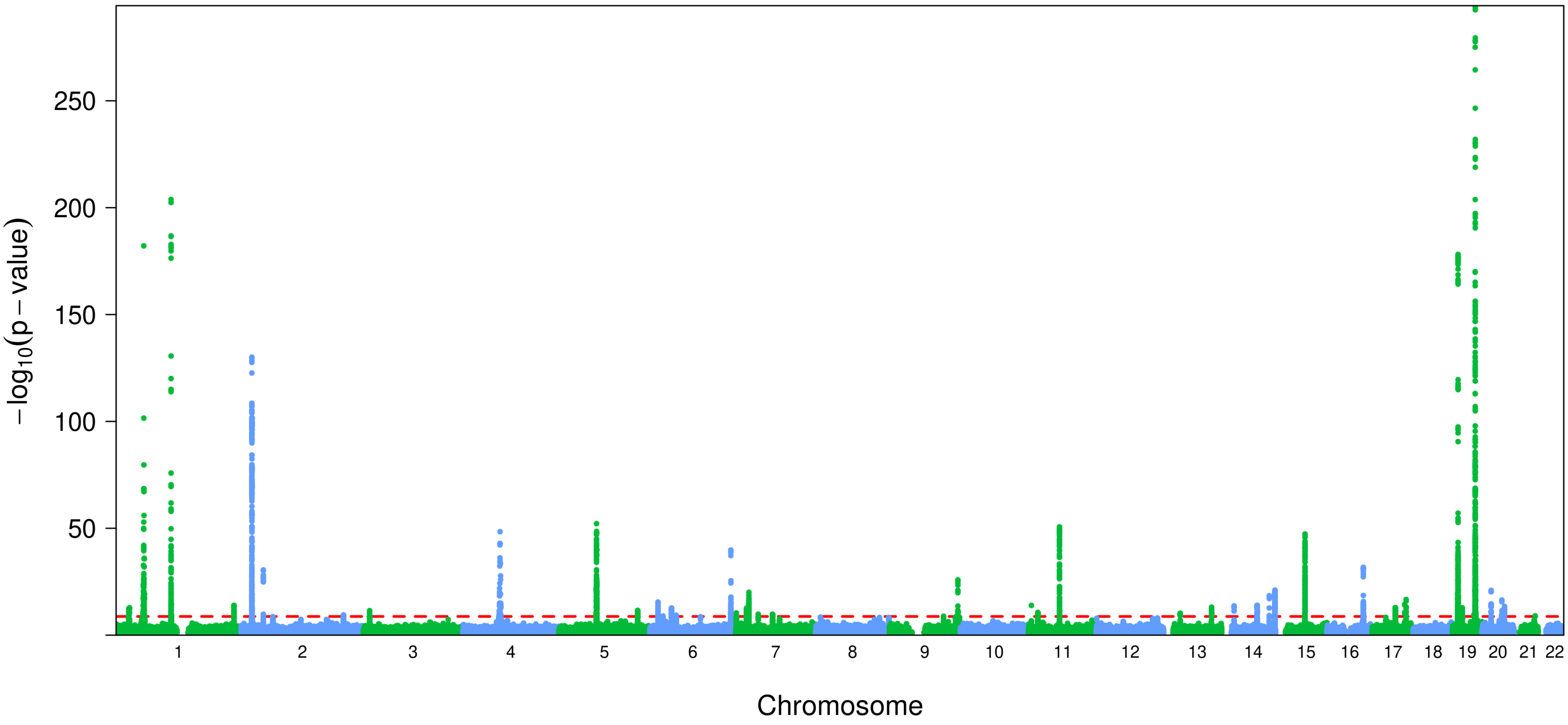

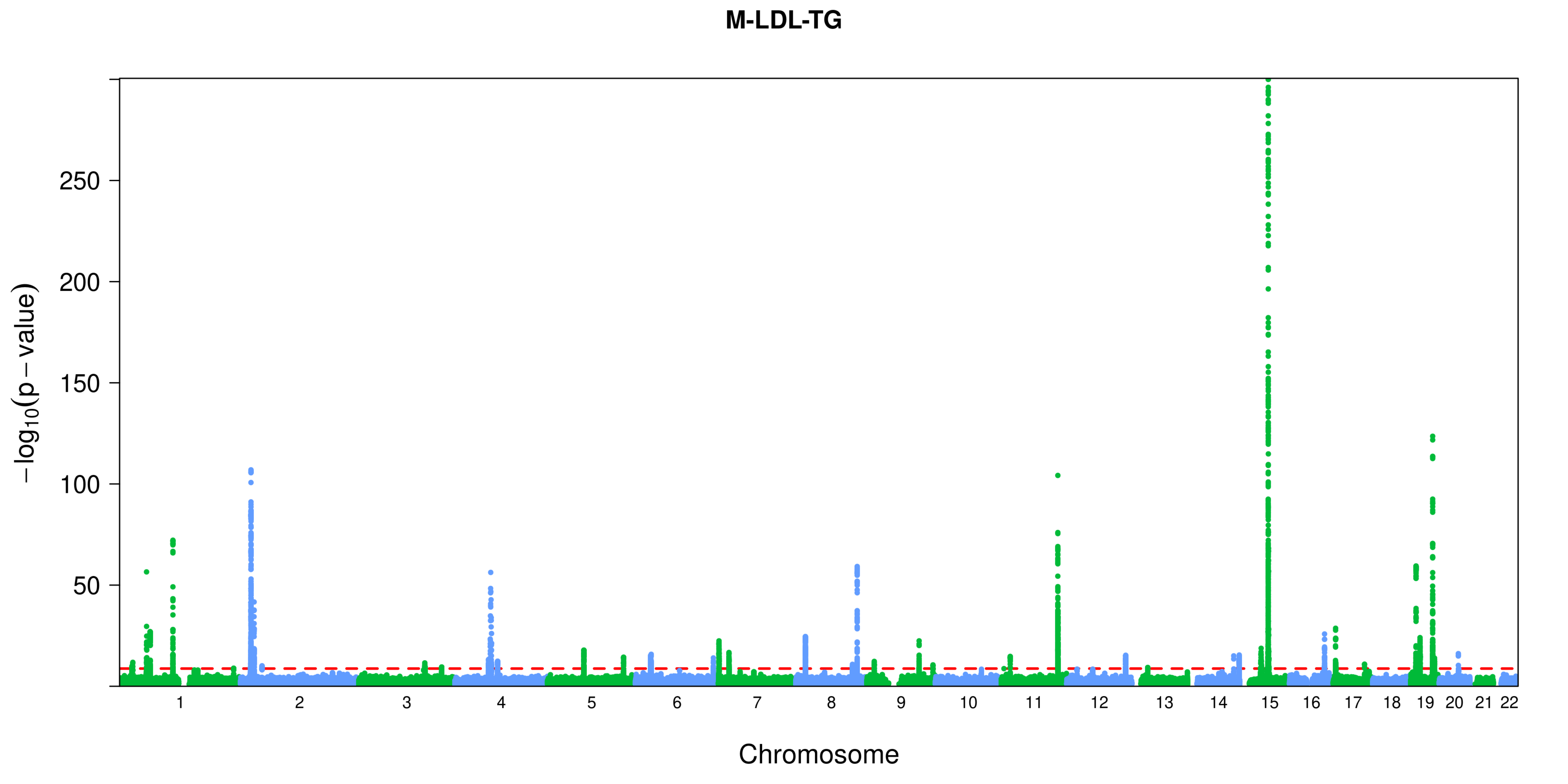
M-LDL-TG
−log10(p − value)
250
200
150
100
50
Chromosome

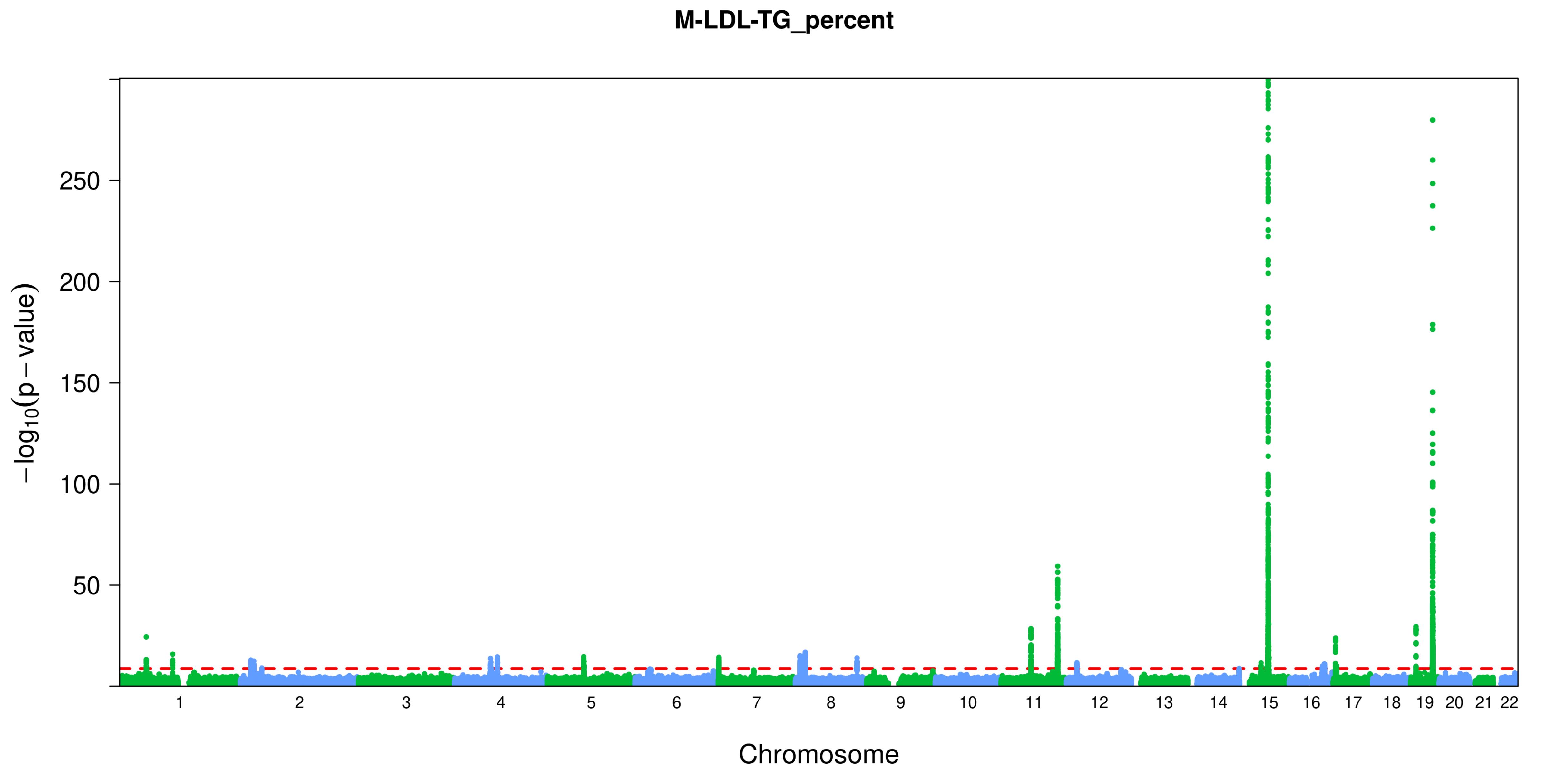

M-LDL-TG_percent
250
200
150
100
50
−log10(p − value)
Chromosome

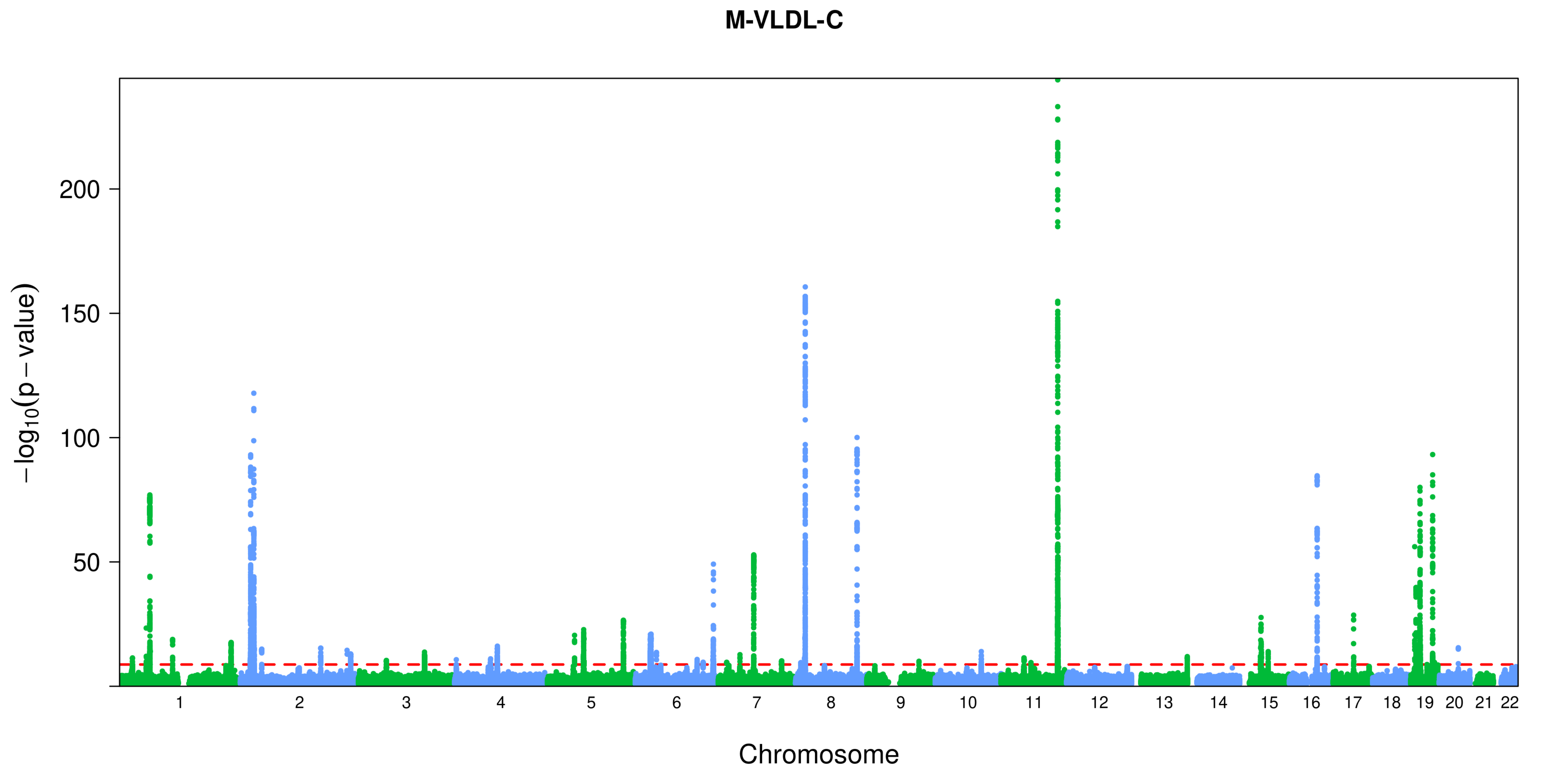
M-VLDL-C
$-\log_{10}(p-\text{value})$
200
150
100
50
Chromosome

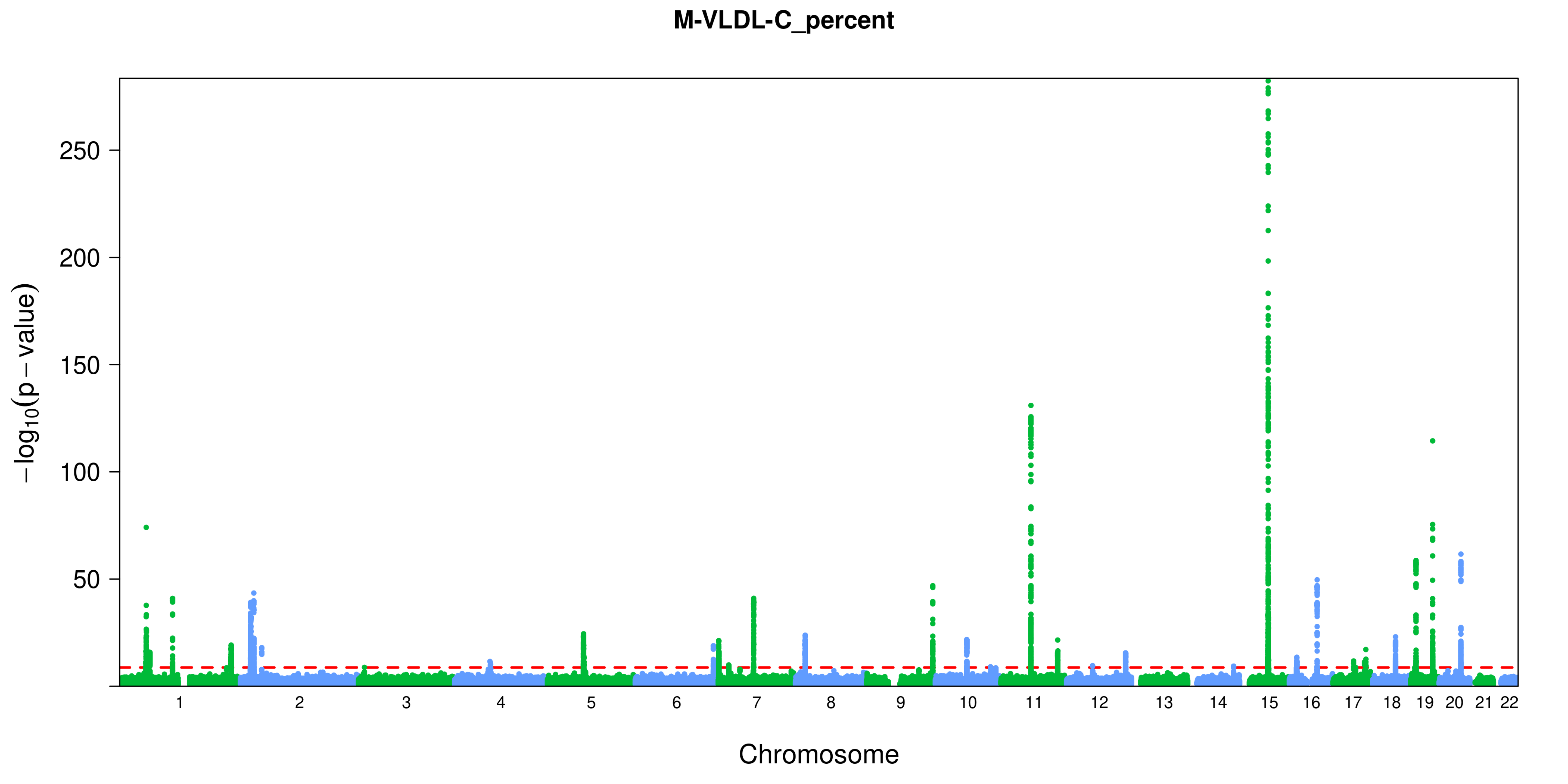
M-VLDL-C_percent
−log10(p − value)
250
200
150
100
50
Chromosome

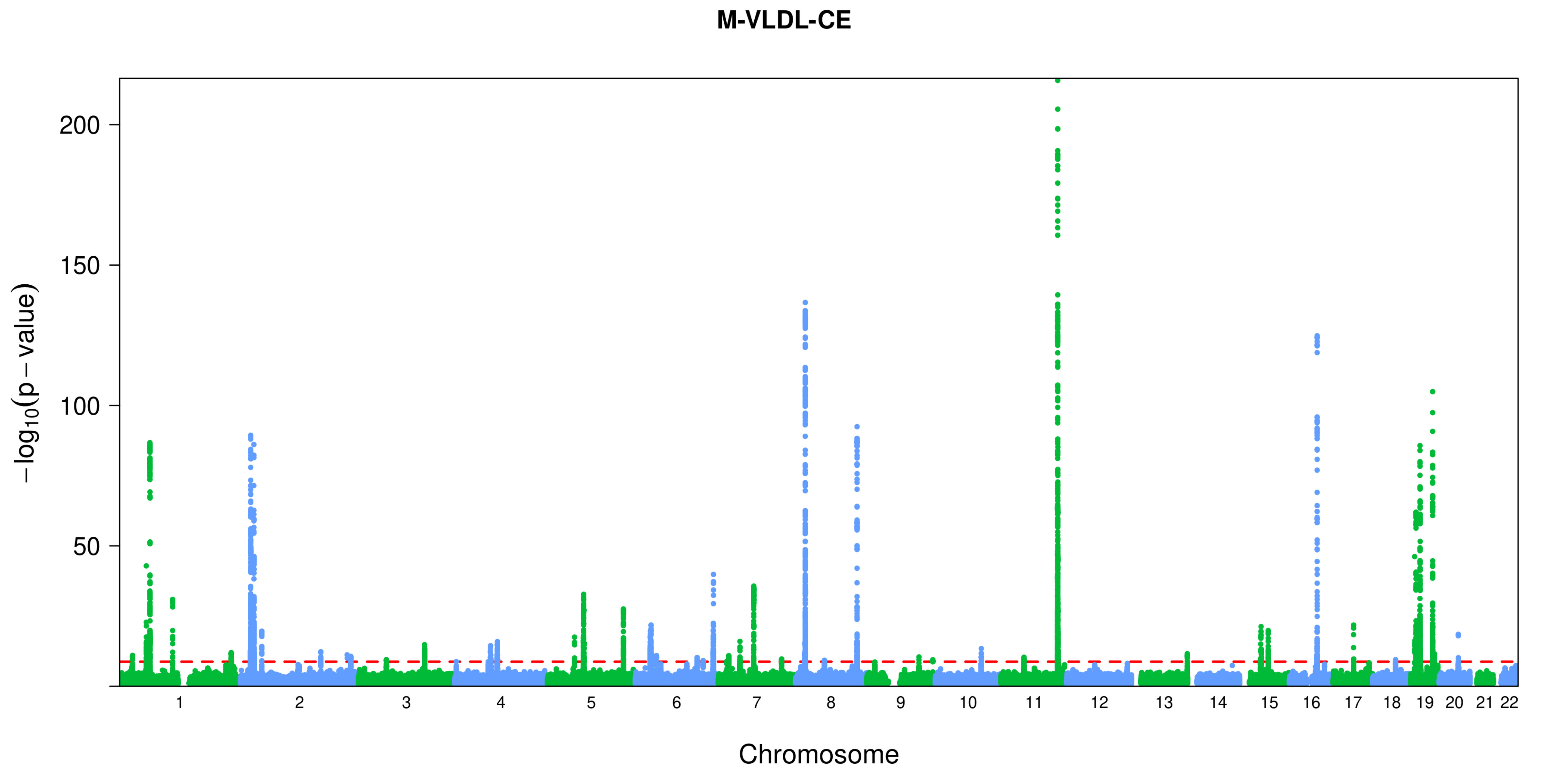
M-VLDL-CE
−log10(p − value)
200
150
100
50
Chromosome

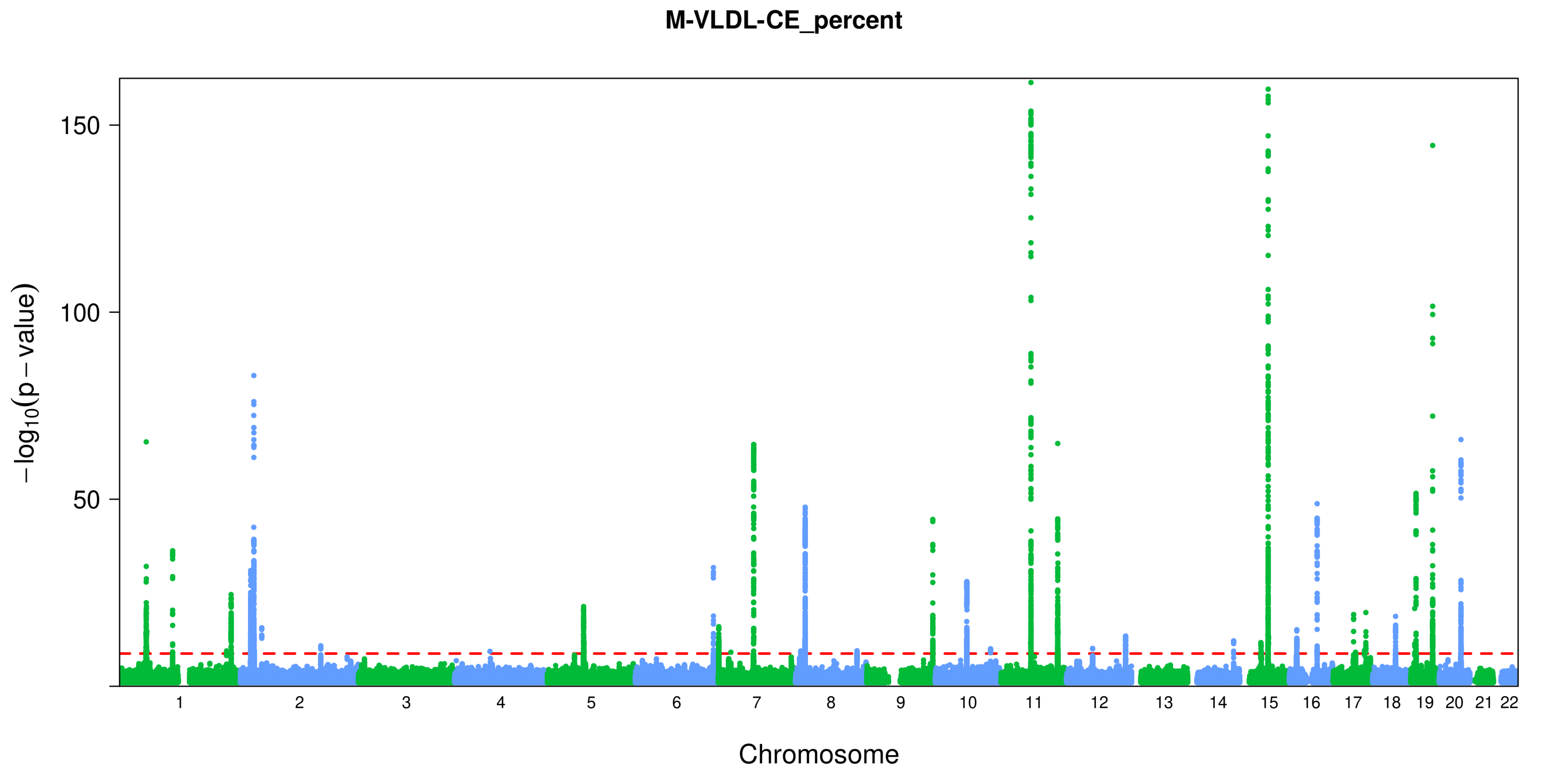
M-VLDL-CE_percent
150
100
50
−log10(p − value)
1 2 3 4 5 6 7 8 9 10 11 12 13 14 15 16 17 18 19 20 21 22
Chromosome

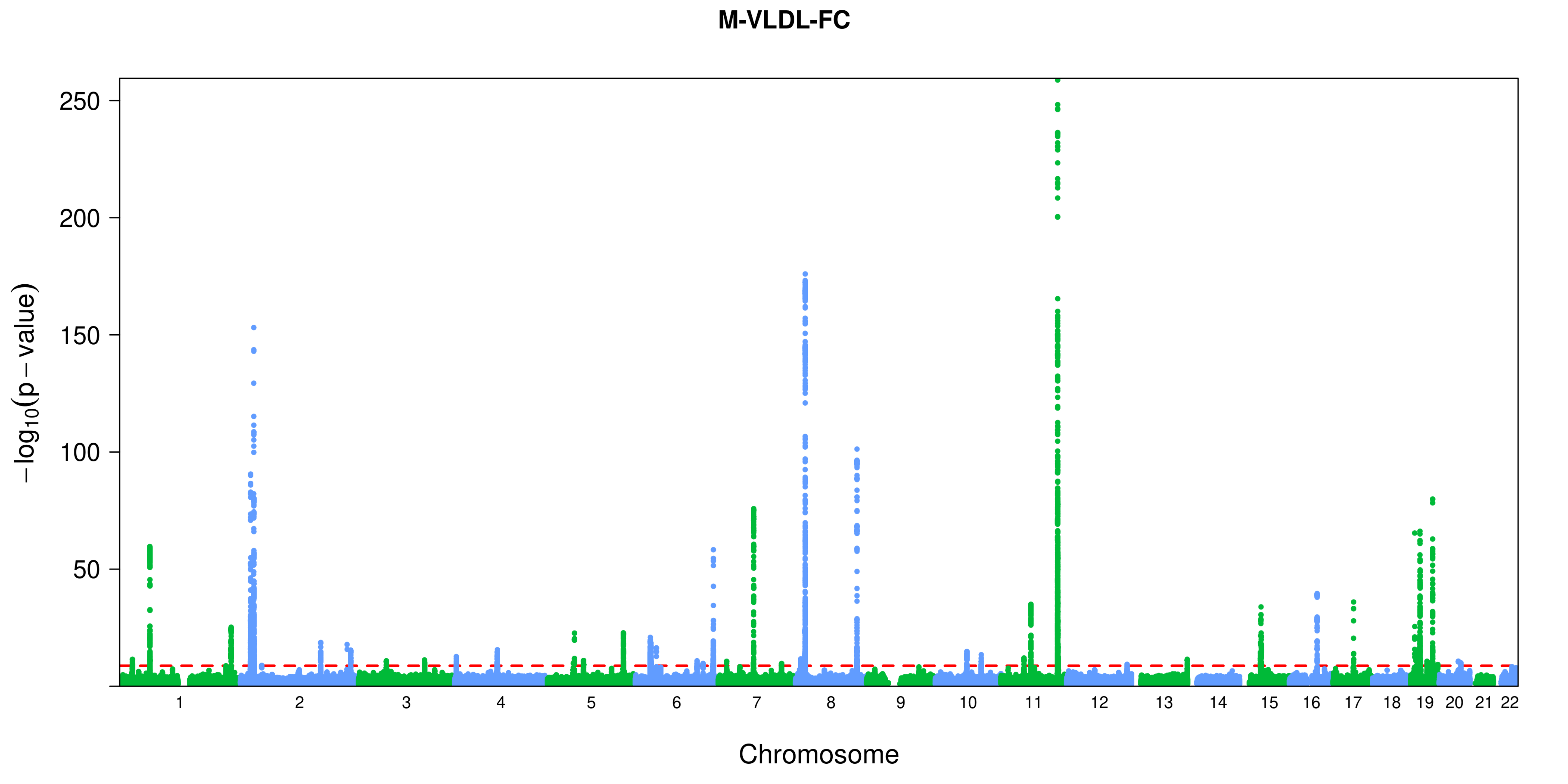

M-VLDL-FC
250
200
150
100
50
$-\log_{10}(p-\text{value})$
Chromosome

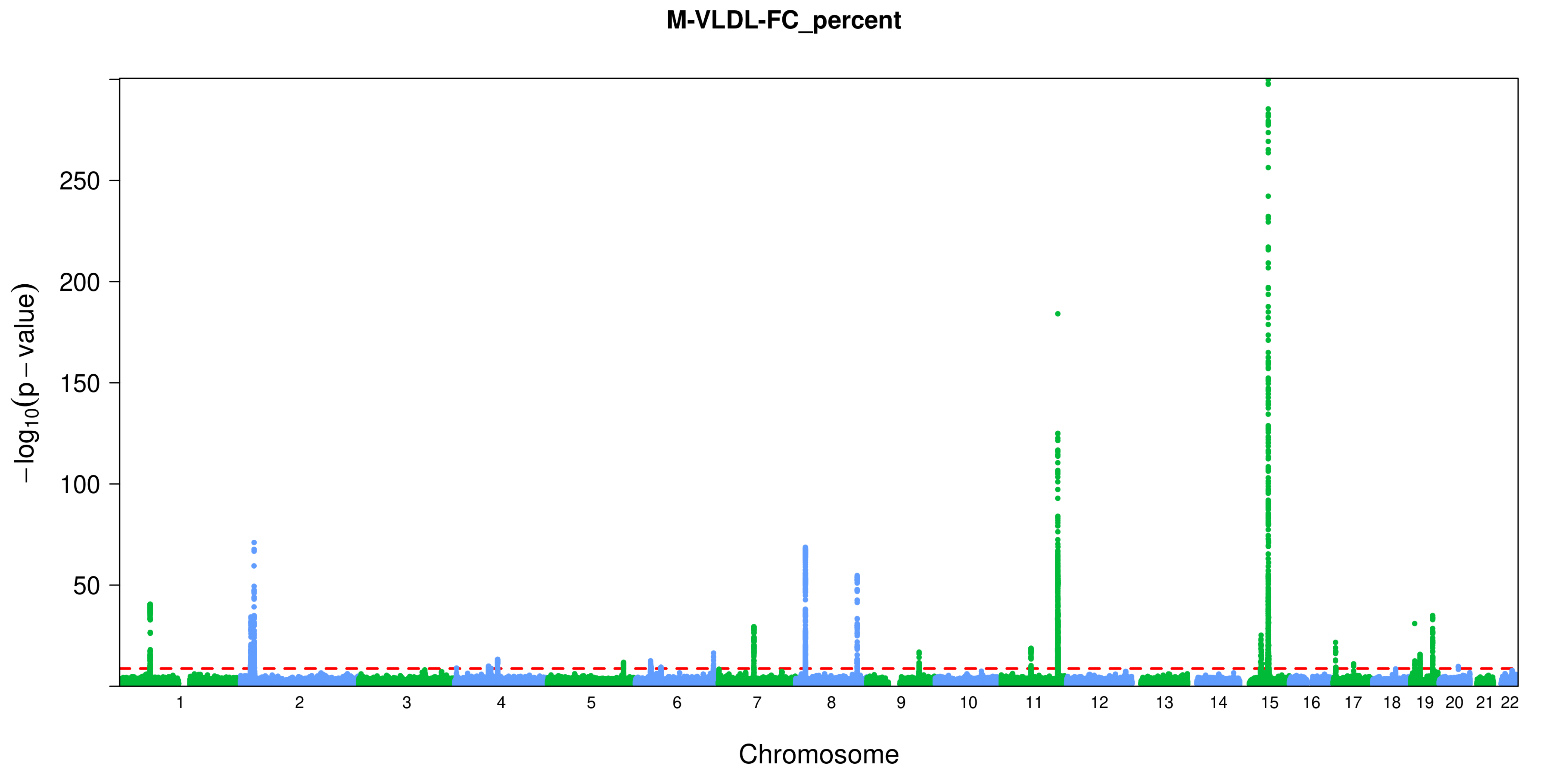
M-VLDL-FC_percent
−log10(p − value)
250
200
150
100
50
Chromosome

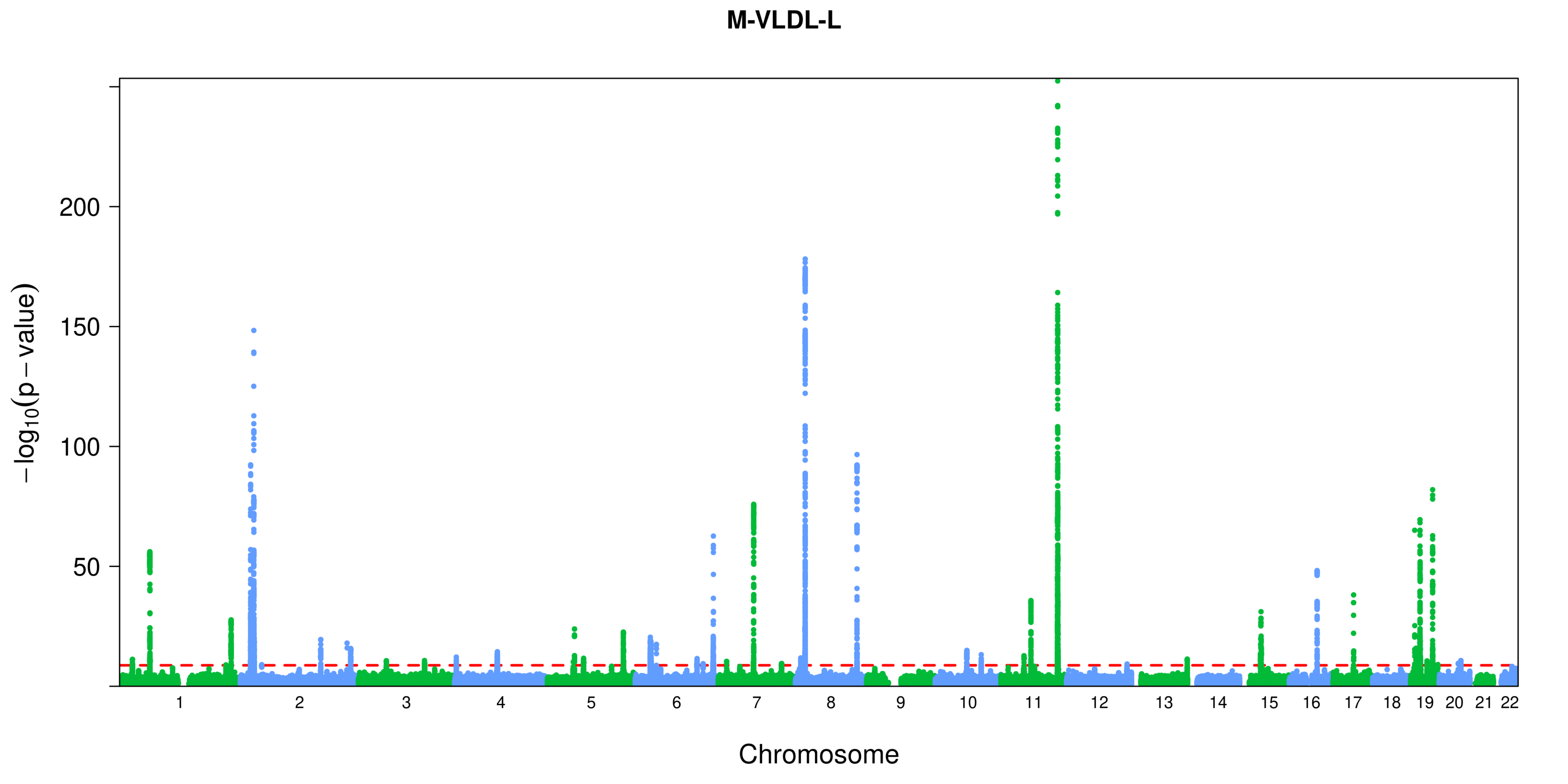

M-VLDL-L
−log10(p − value)
200
150
100
50
Chromosome

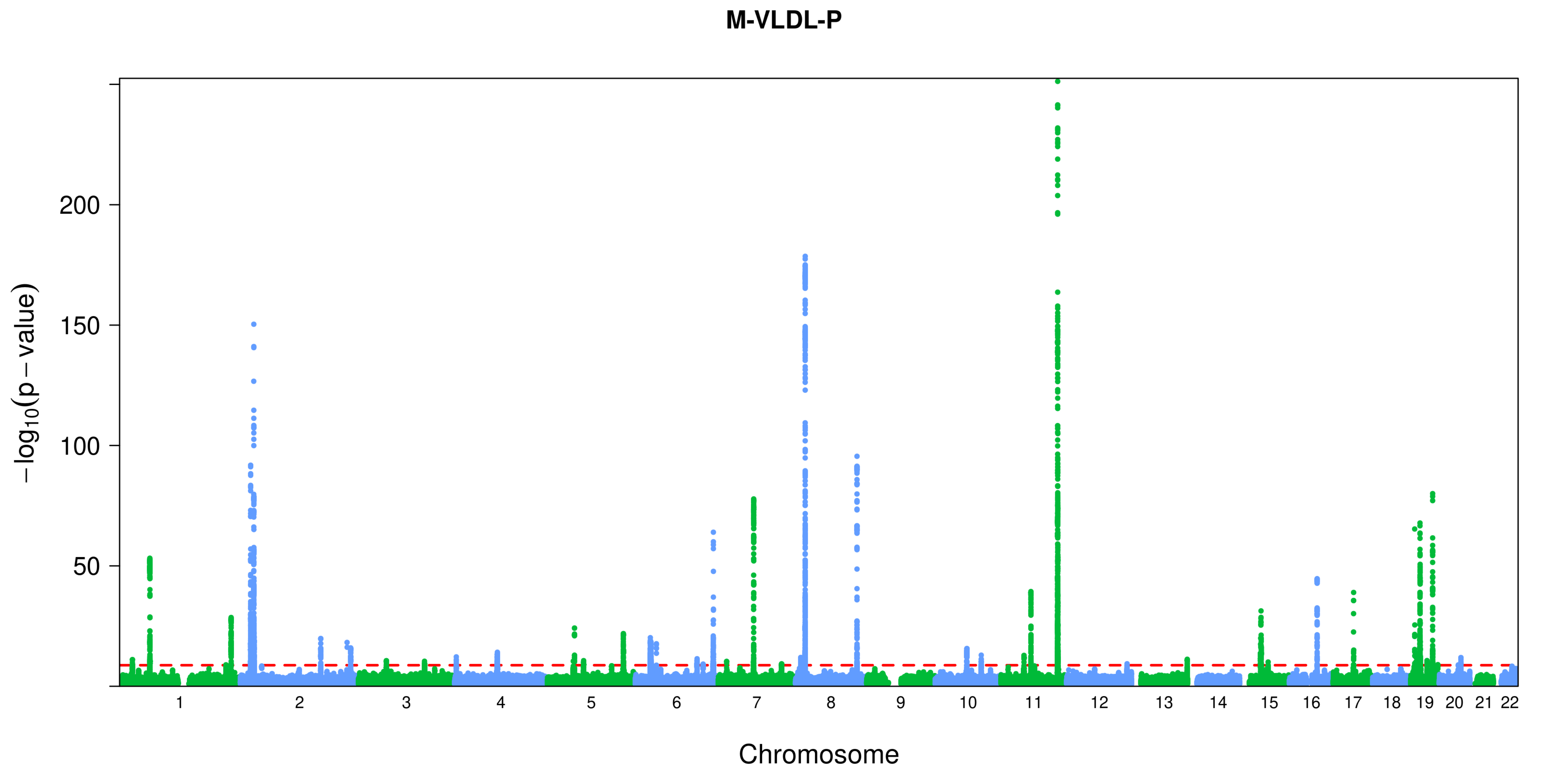
M-VLDL-P
200
150
100
50
$-\log_{10}(p-\text{value})$
Chromosome

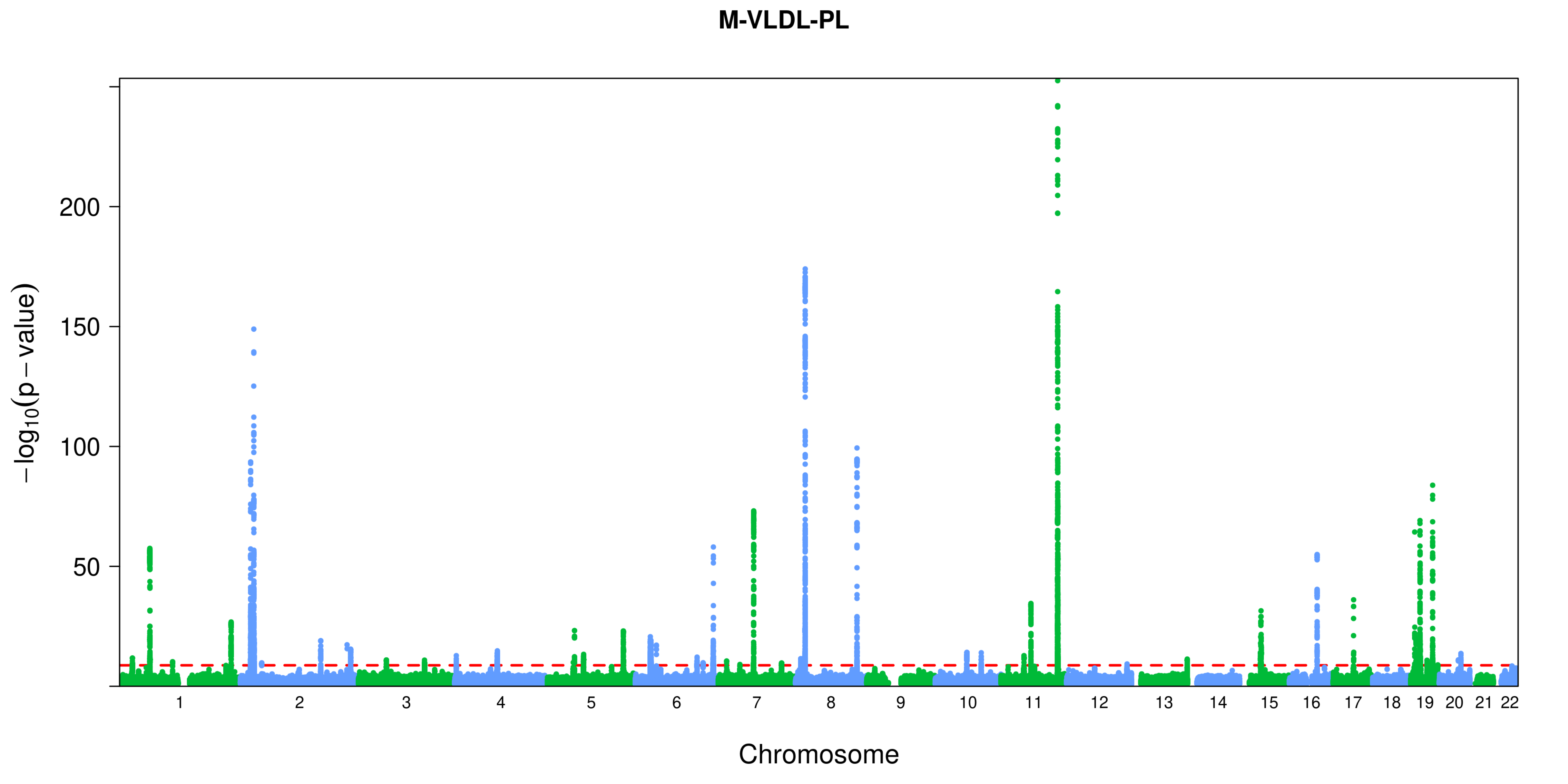
M-VLDL-PL
200
150
100
50
−log10(p − value)
Chromosome

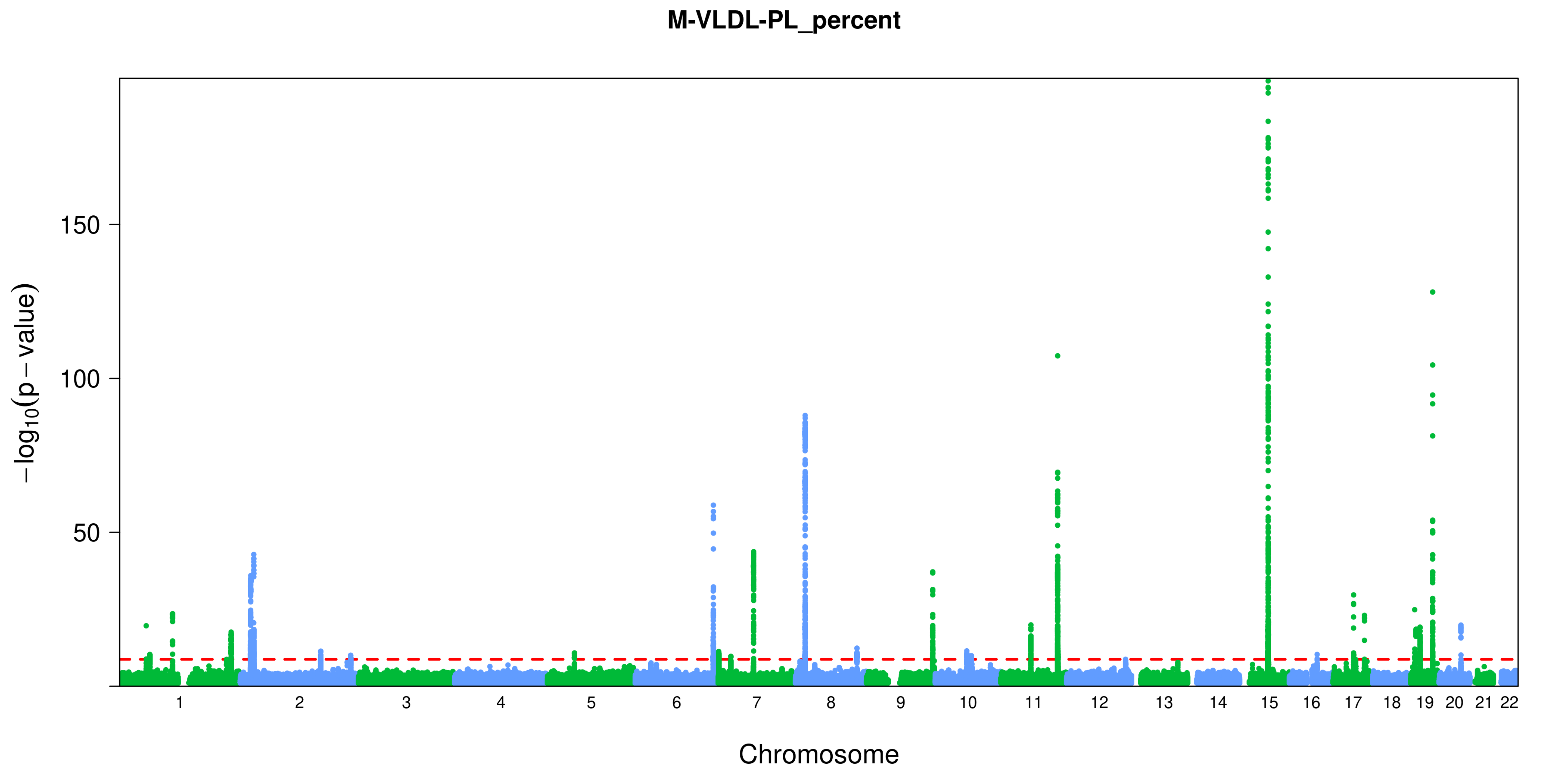
M-VLDL-PL_percent
150
100
50
$-\log_{10}(p-\text{value})$
Chromosome

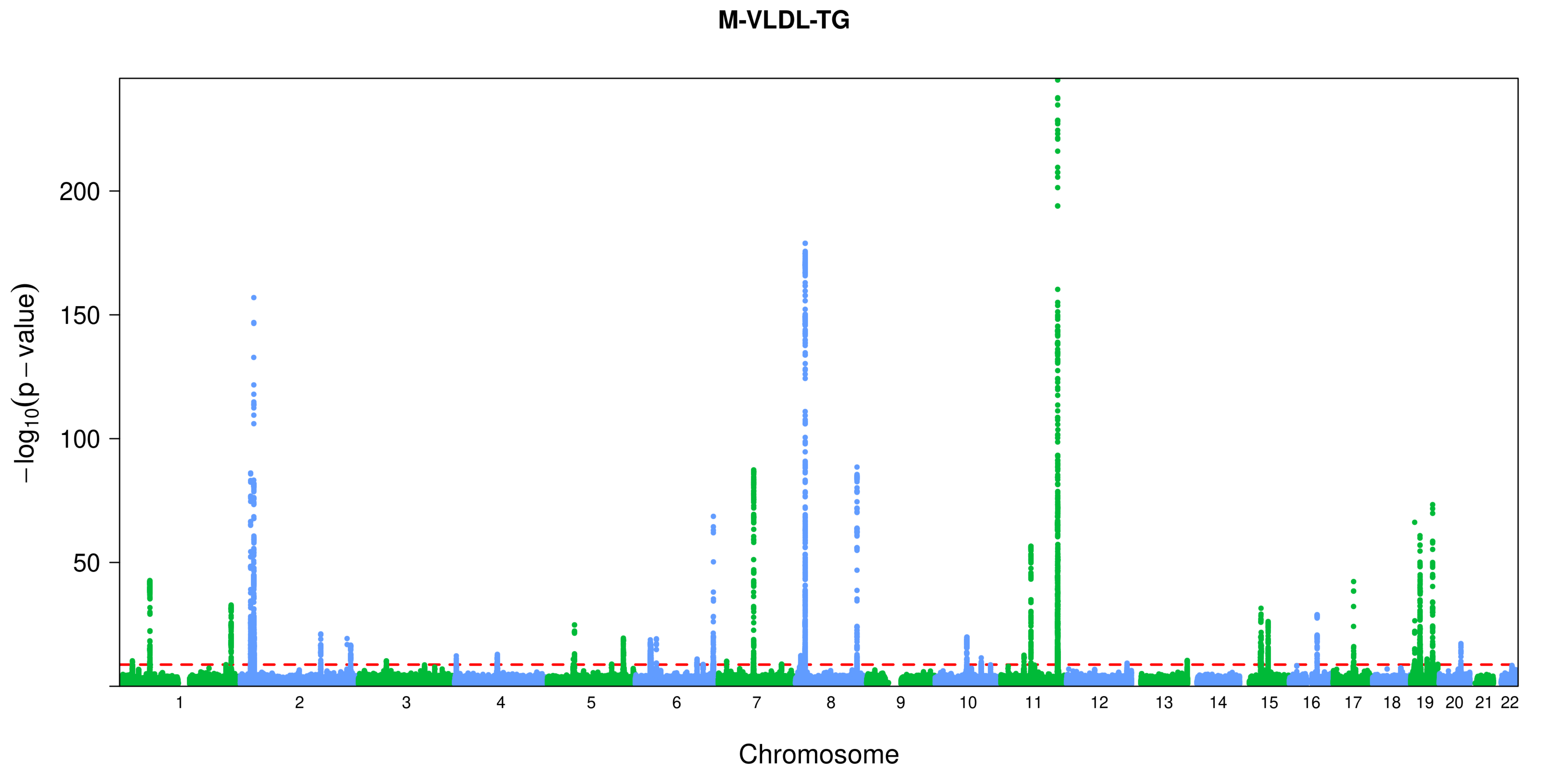

M-VLDL-TG
200
150
100
50
−log10(p − value)
Chromosome

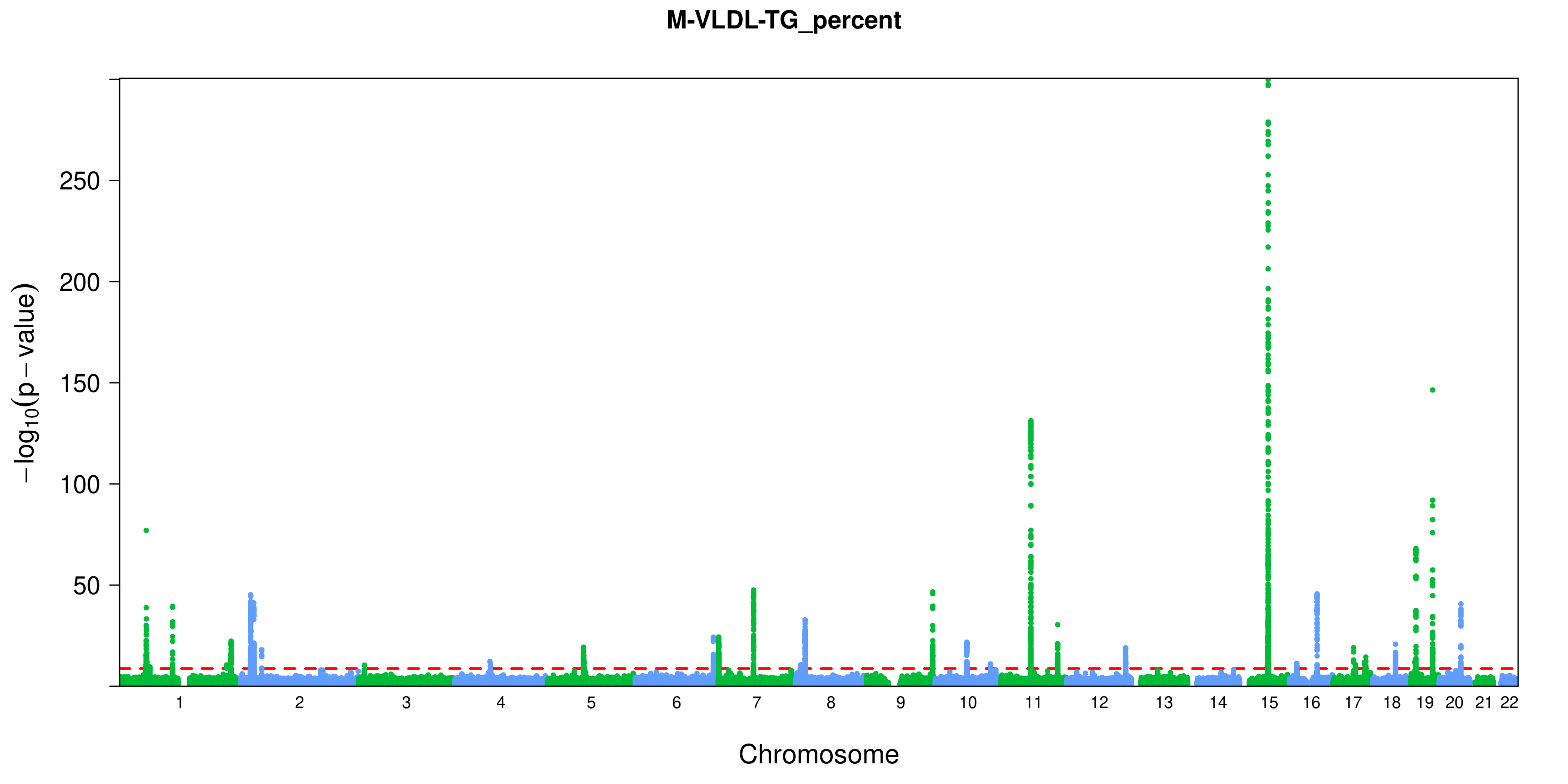
M-VLDL-TG_percent
$-\log_{10}(p-\text{value})$
250
200
150
100
50
Chromosome

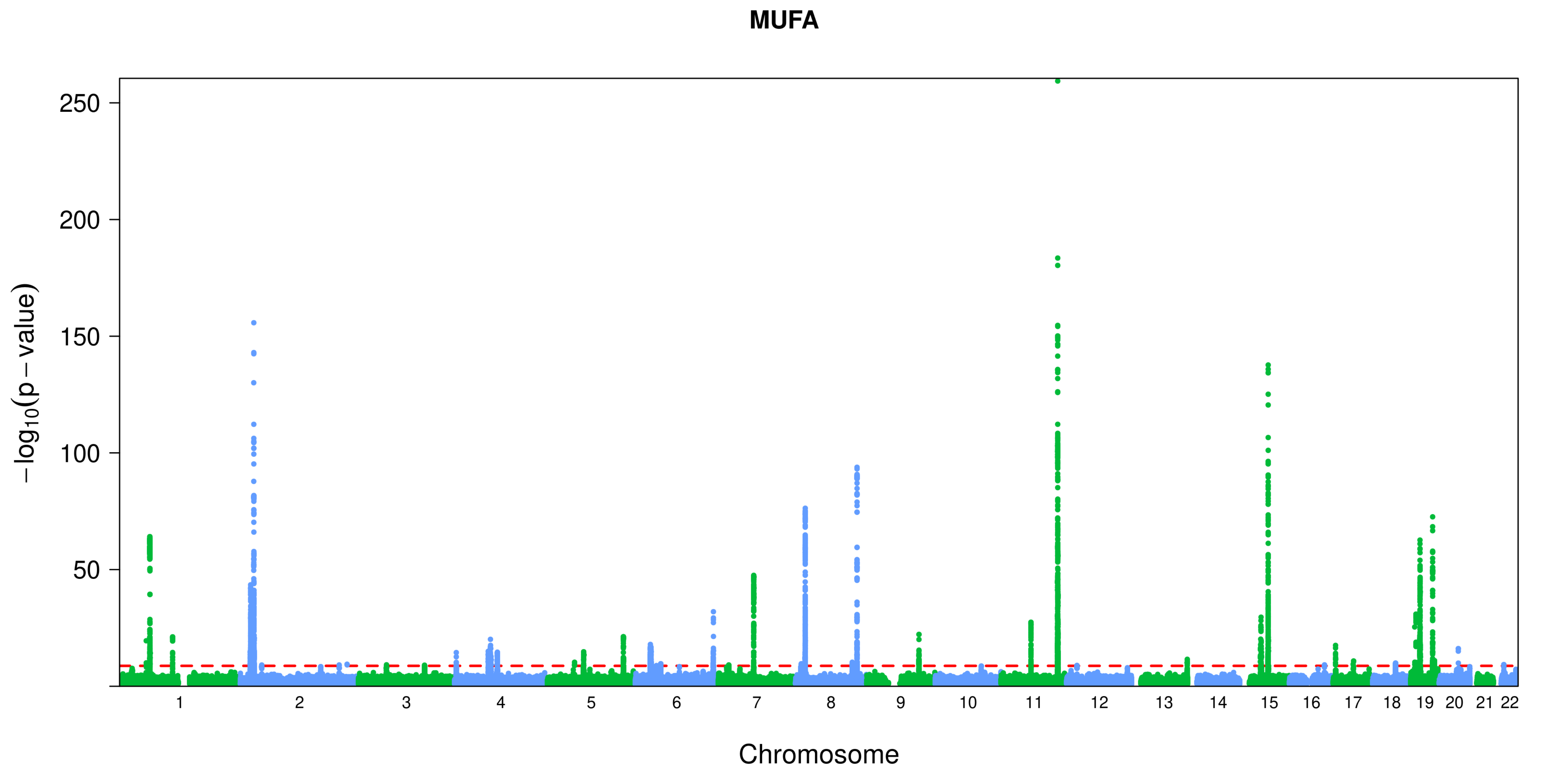
MUFA
250
200
150
100
50
−log10(p − value)
Chromosome

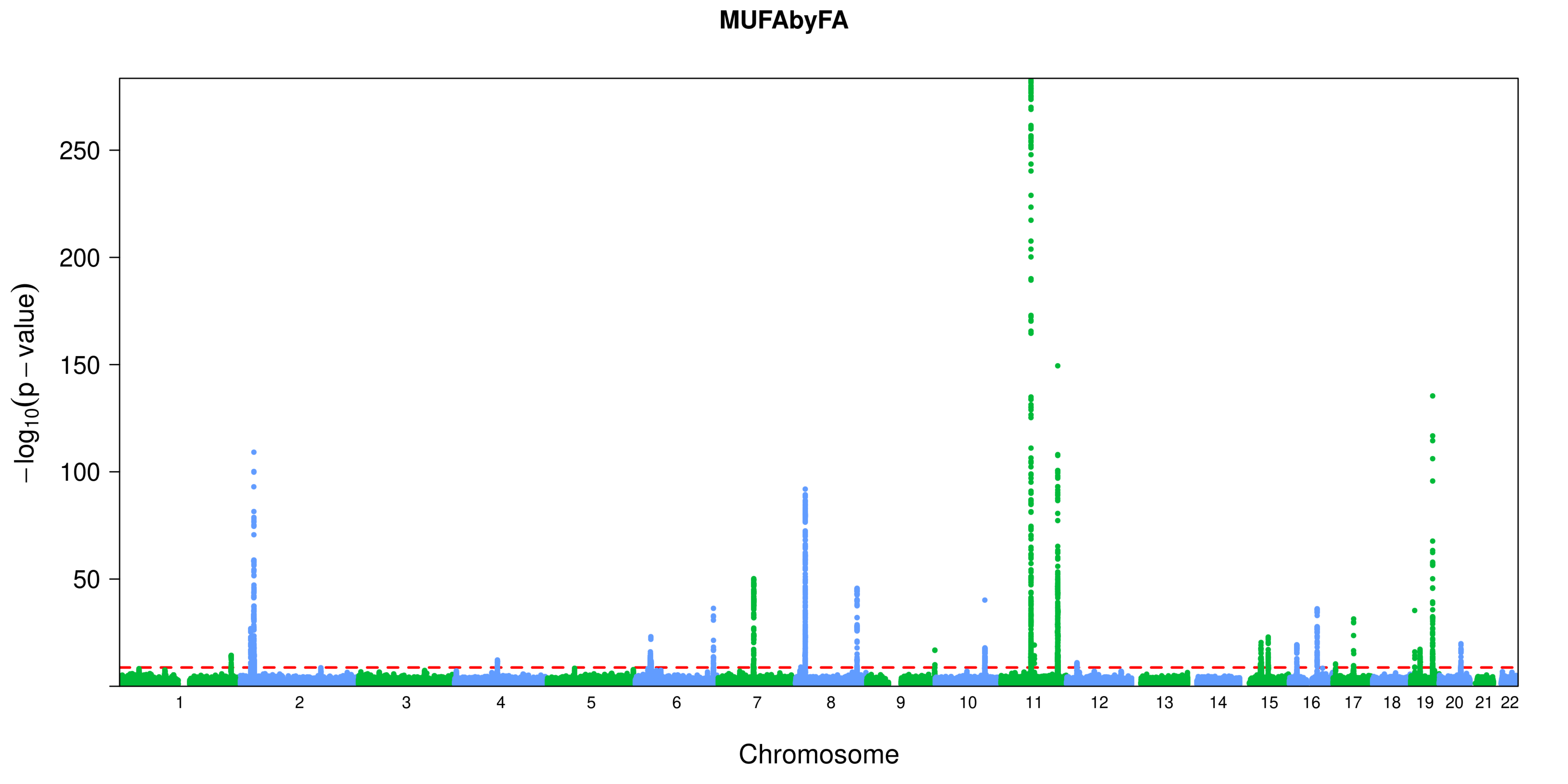
MUFAbyFA
−log10(p − value)
250
200
150
100
50
Chromosome

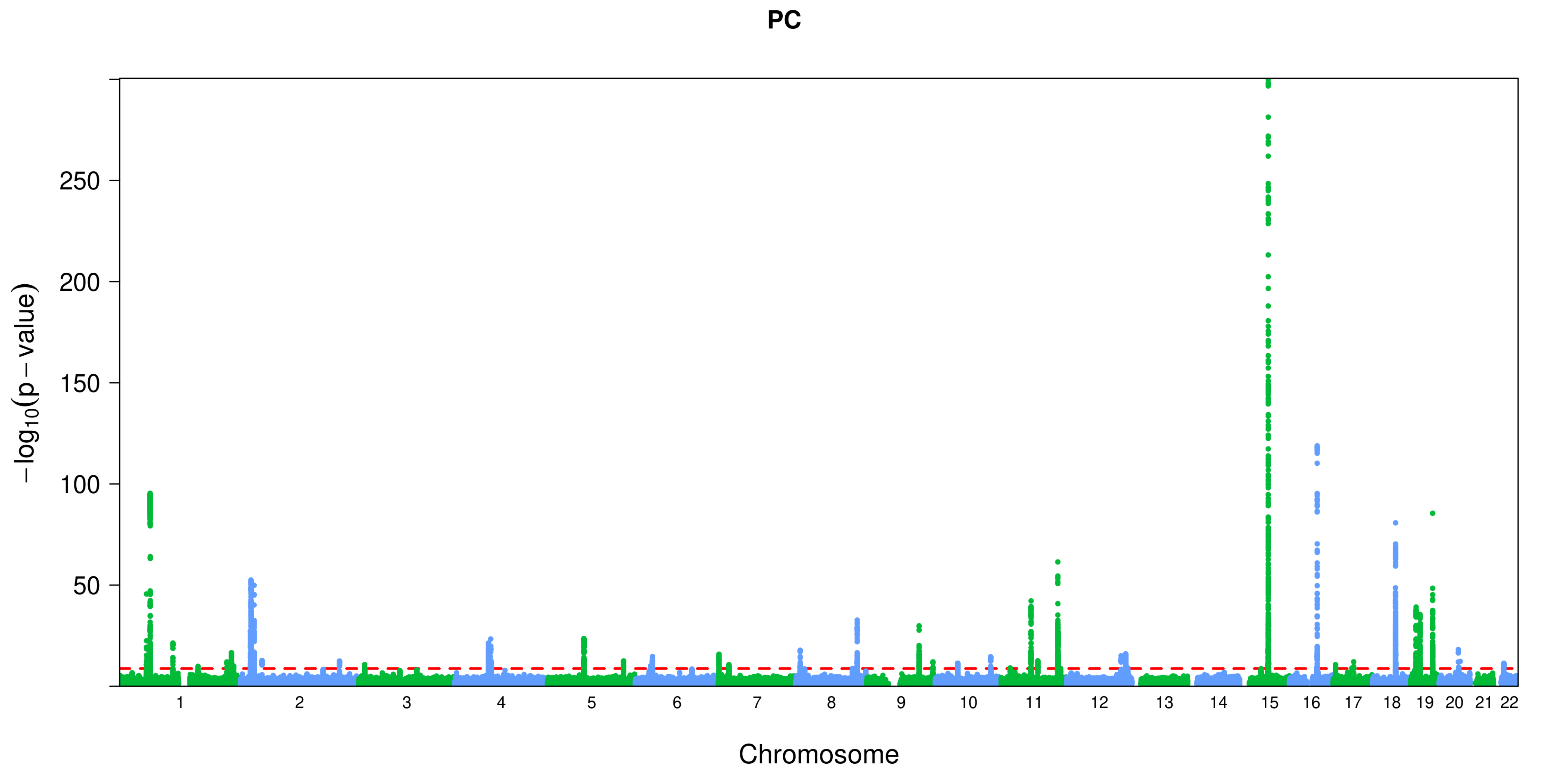

PC
250
200
150
100
50
−log10(p − value)
Chromosome

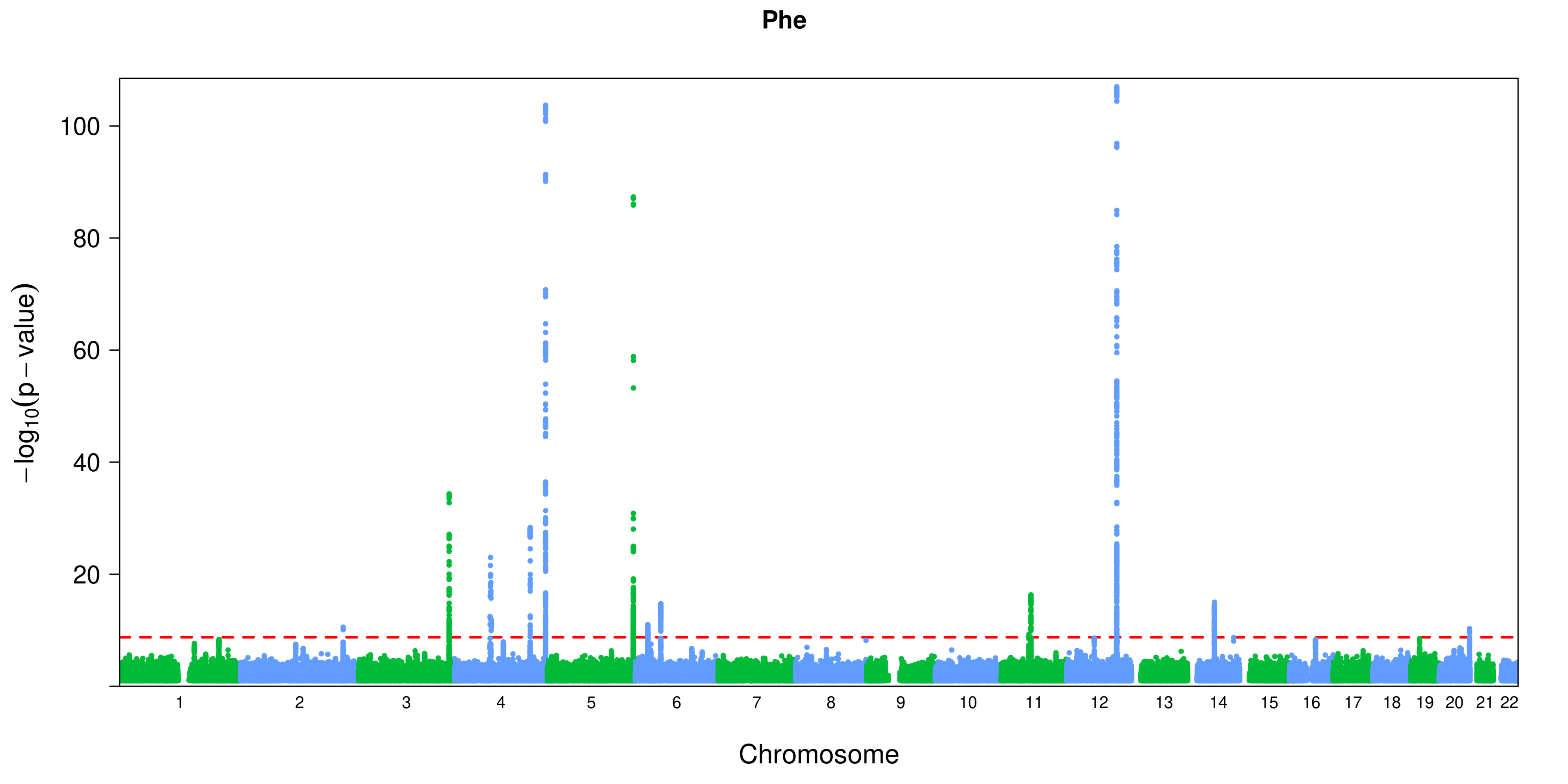
Phe
100
80
60
40
20
−log10(p − value)
Chromosome

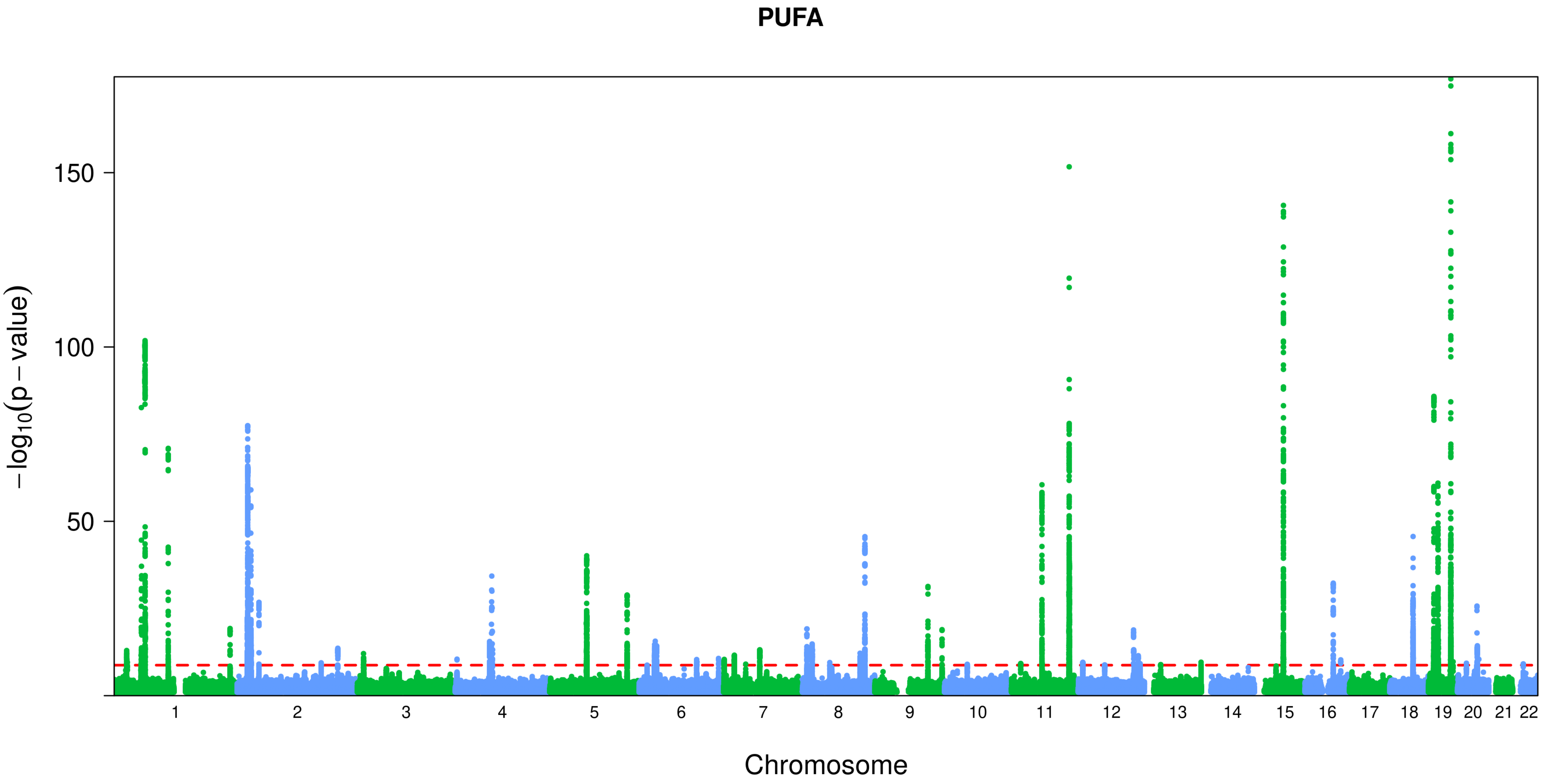
PUFA
−log10(p − value)
150
100
50
Chromosome

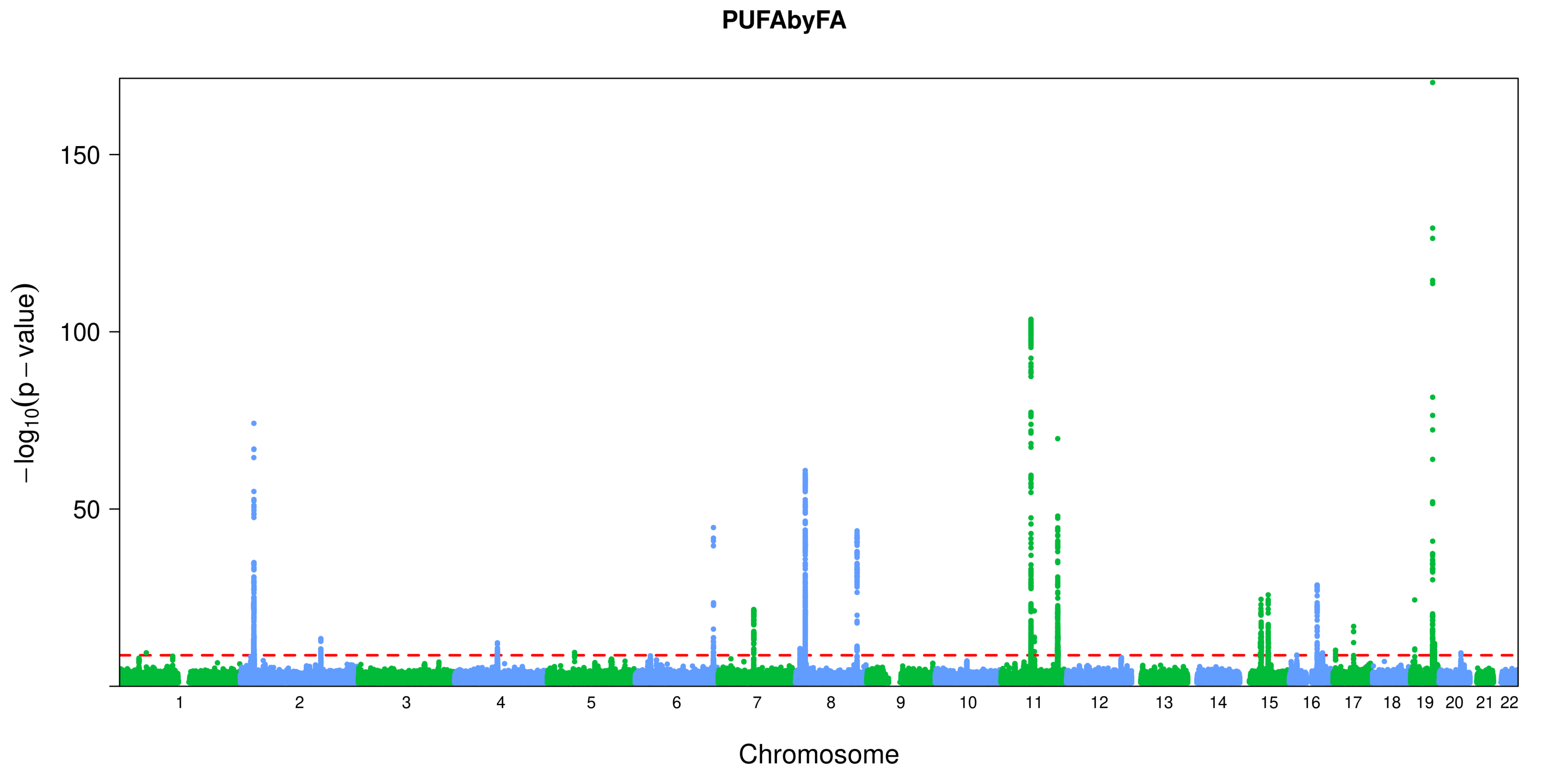
PUFAbyFA
−log10(p − value)
150
100
50
Chromosome

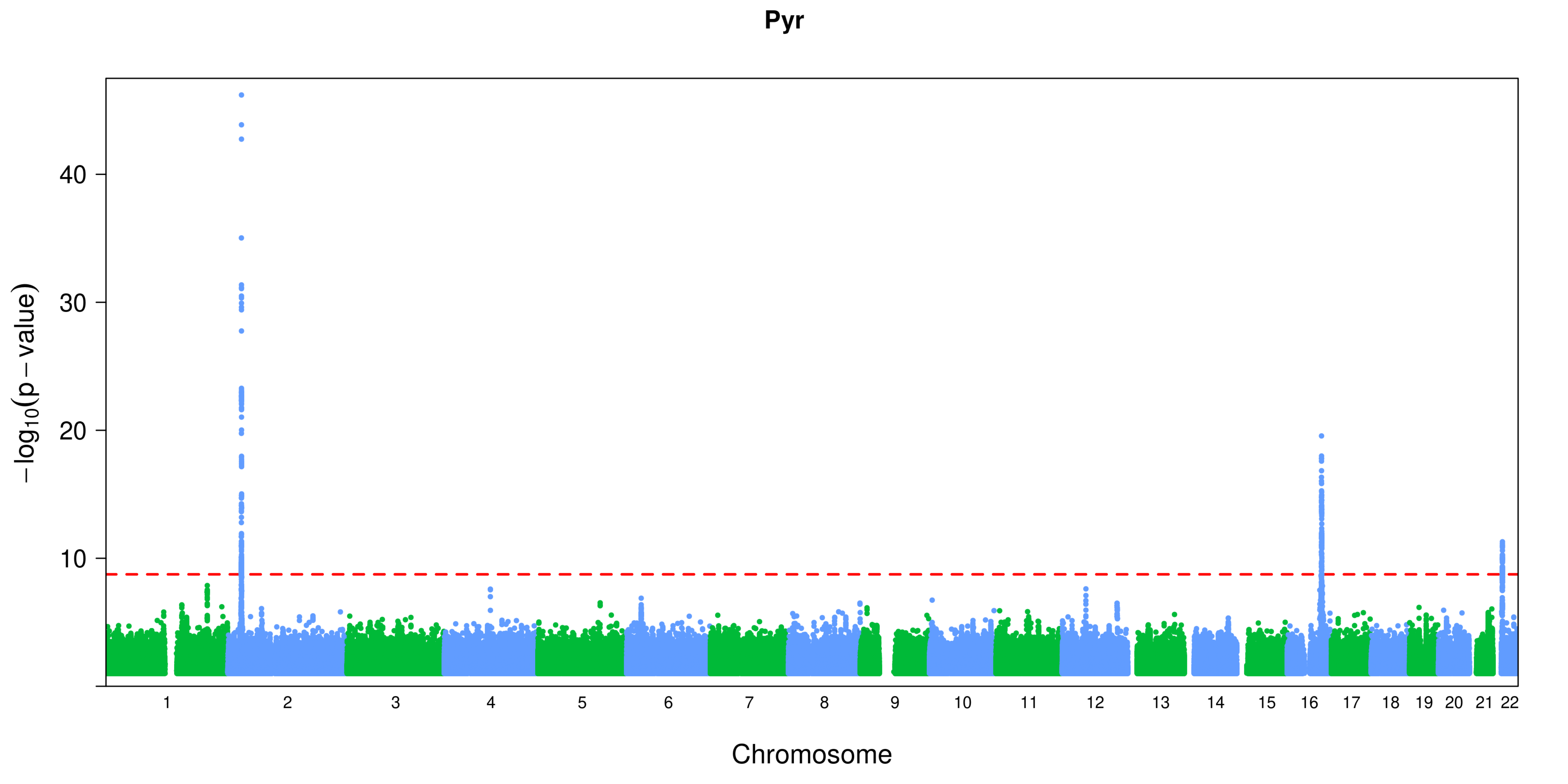

Pyr
40
30
20
10
−log10(p − value)
Chromosome

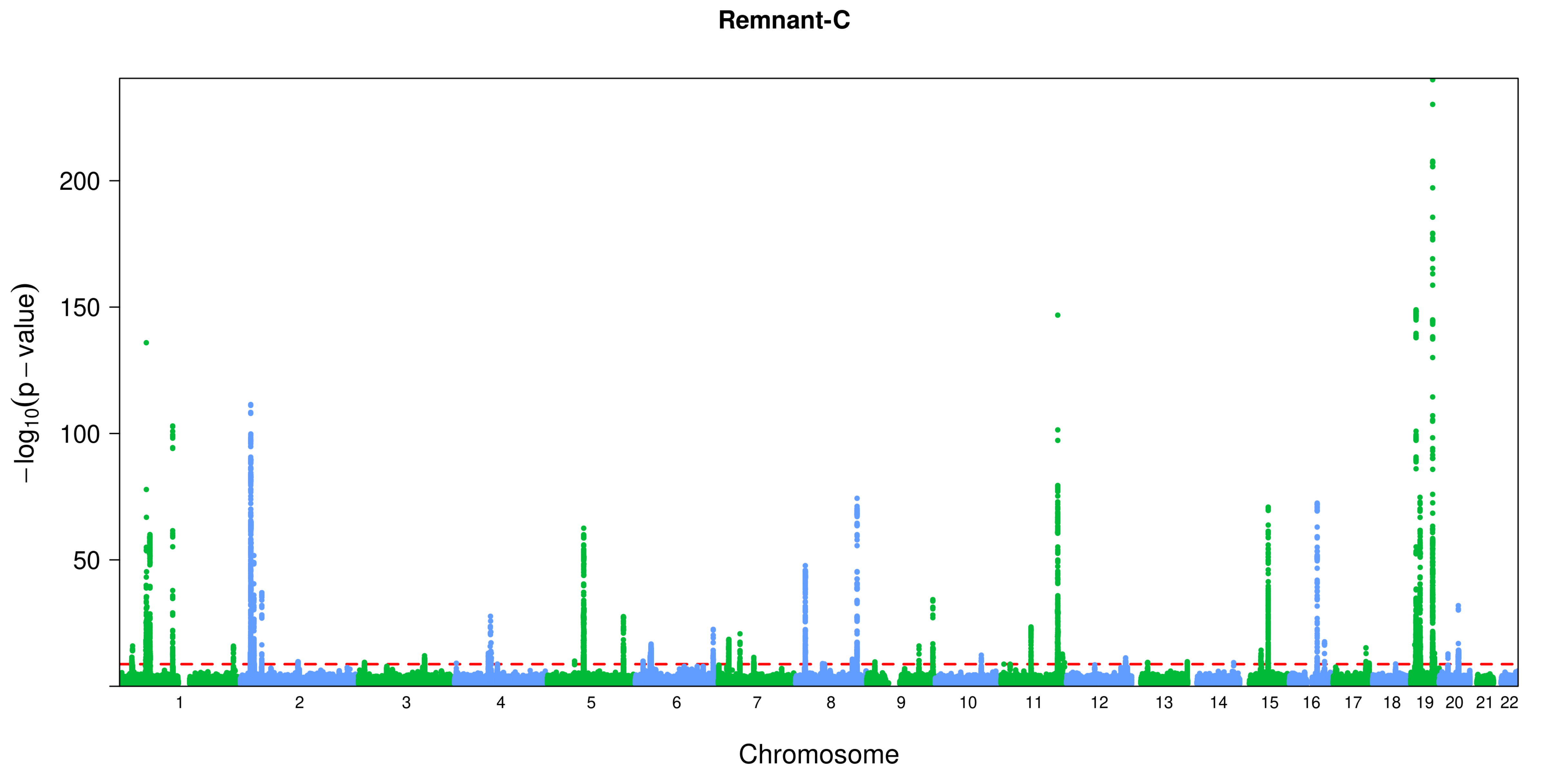
Remnant-C
200
150
100
50
$-\log_{10}(p-\text{value})$
Chromosome

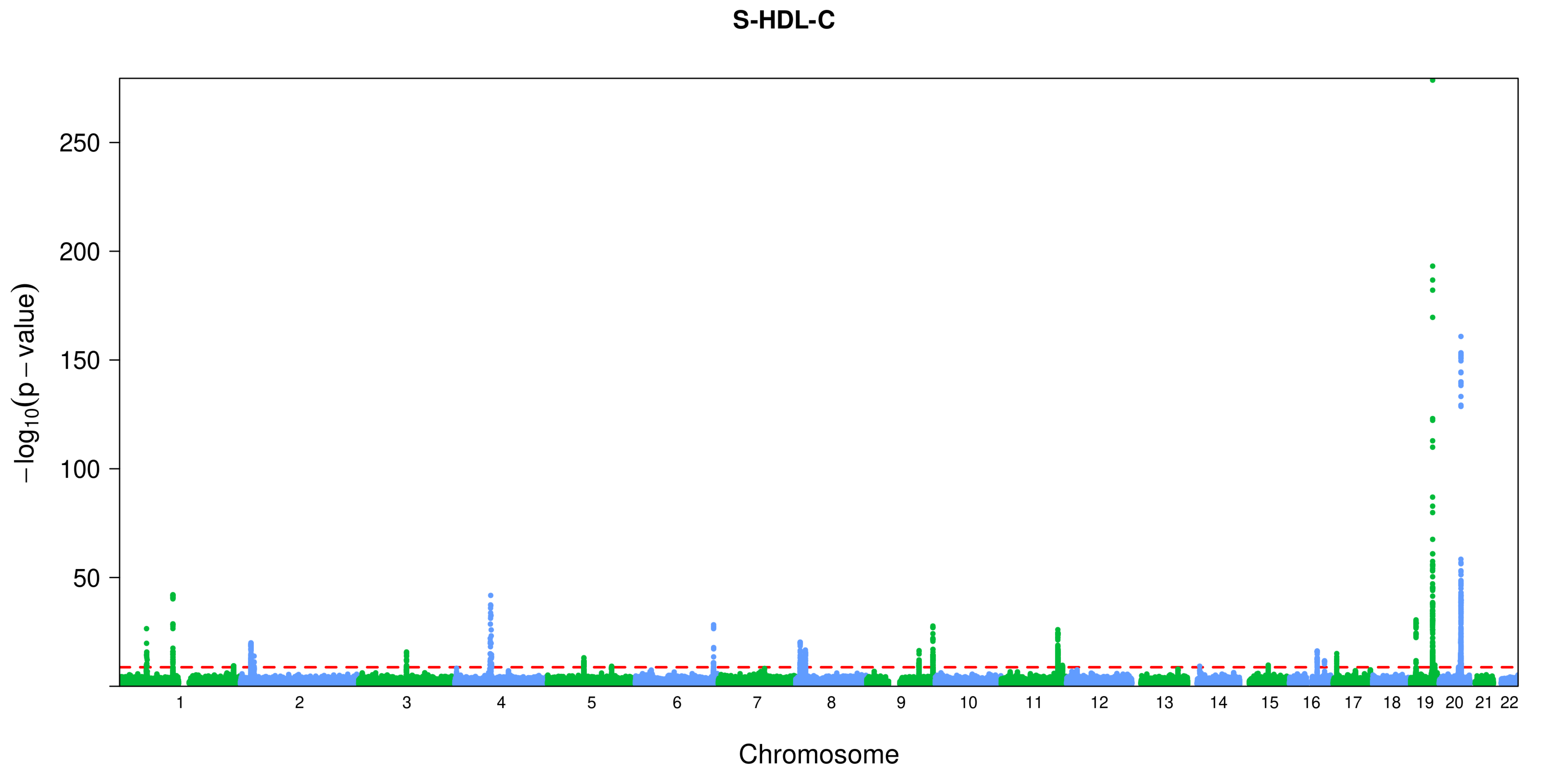
S-HDL-C
−log10(p − value)
250
200
150
100
50
Chromosome

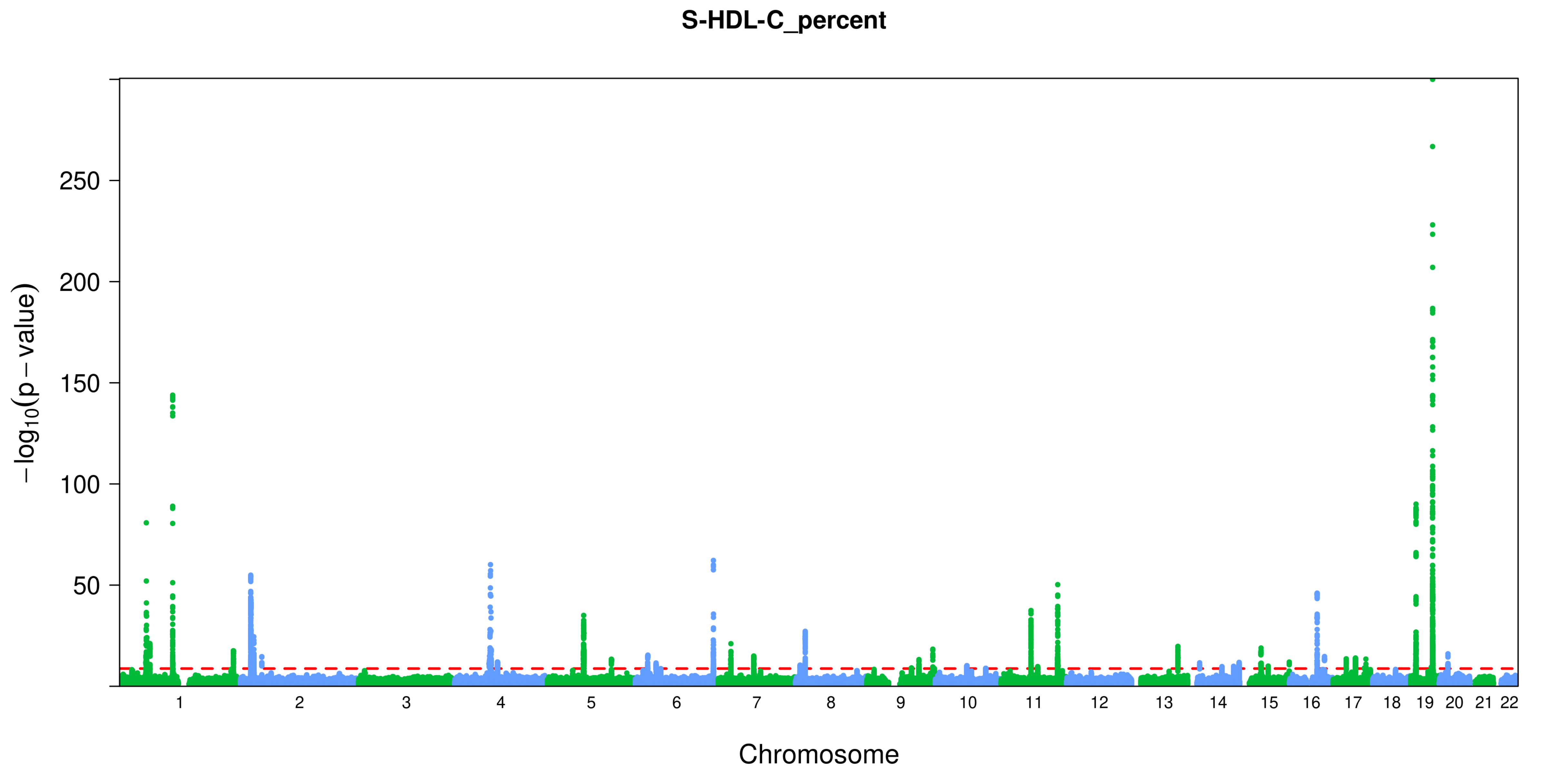
S-HDL-C_percent
−log10(p − value)
250
200
150
100
50
Chromosome

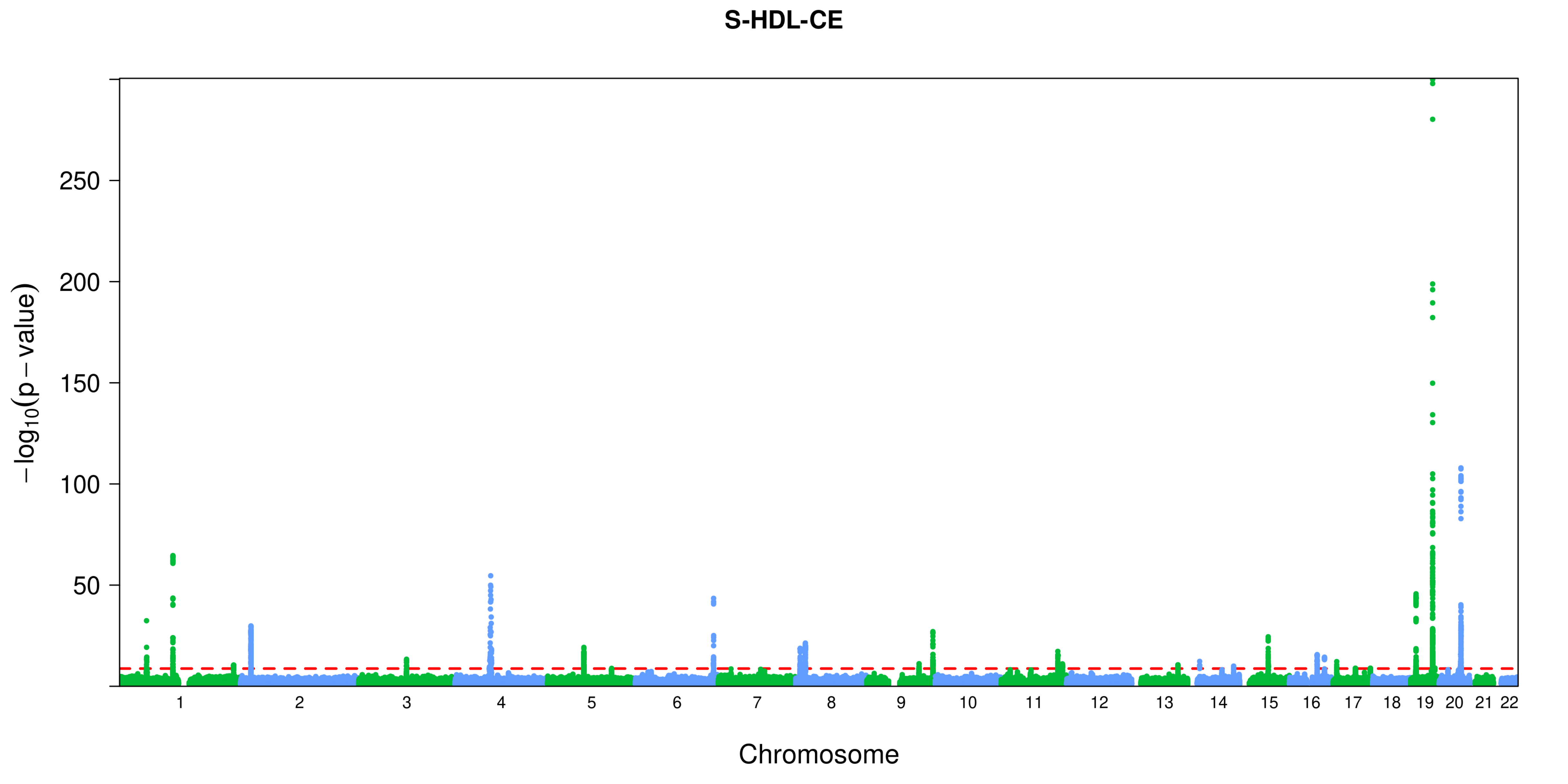
S-HDL-CE
−log10(p − value)
250
200
150
100
50
Chromosome

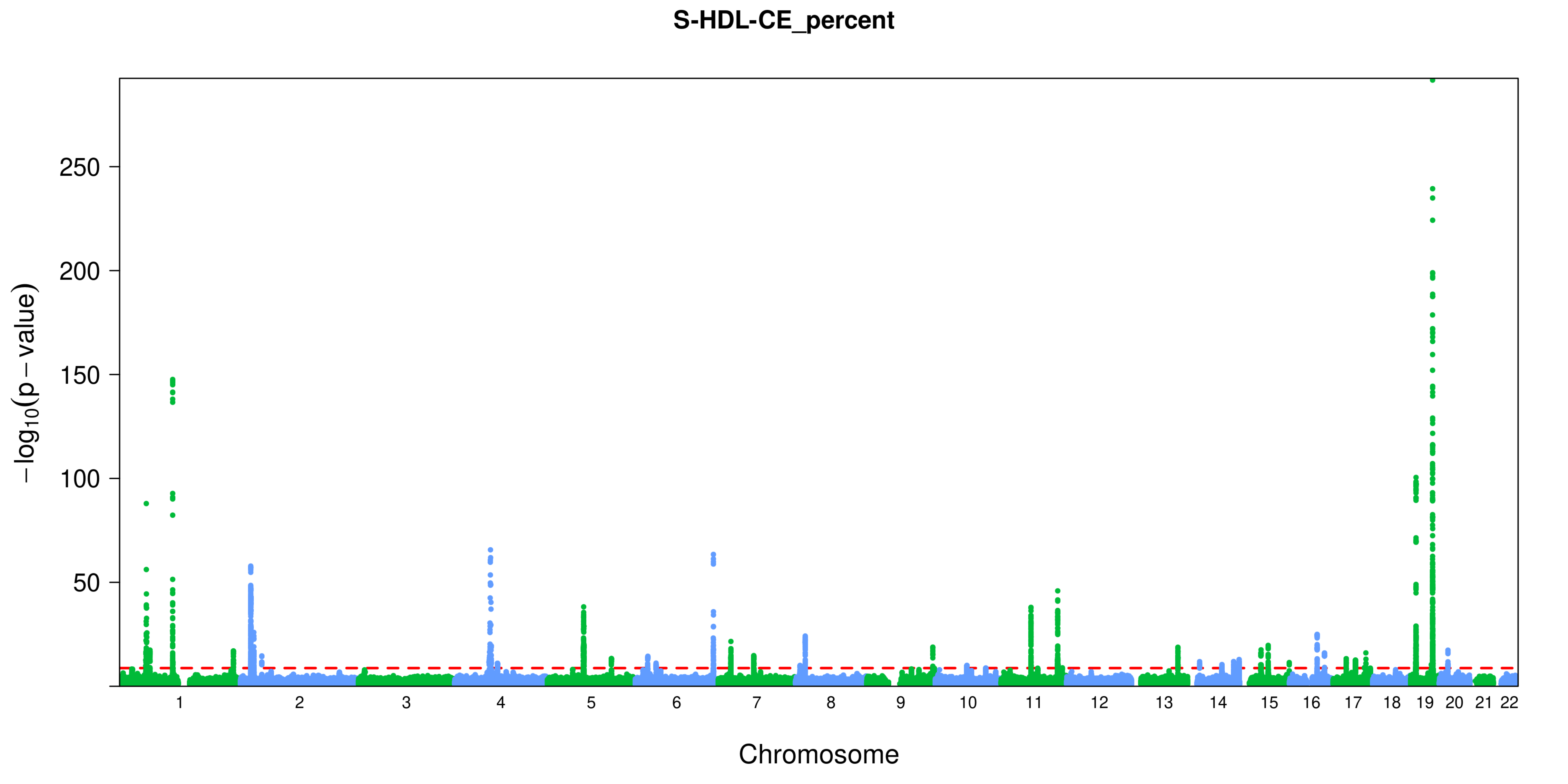

S-HDL-CE_percent
$-\log_{10}(p - value)$
250
200
150
100
50
Chromosome

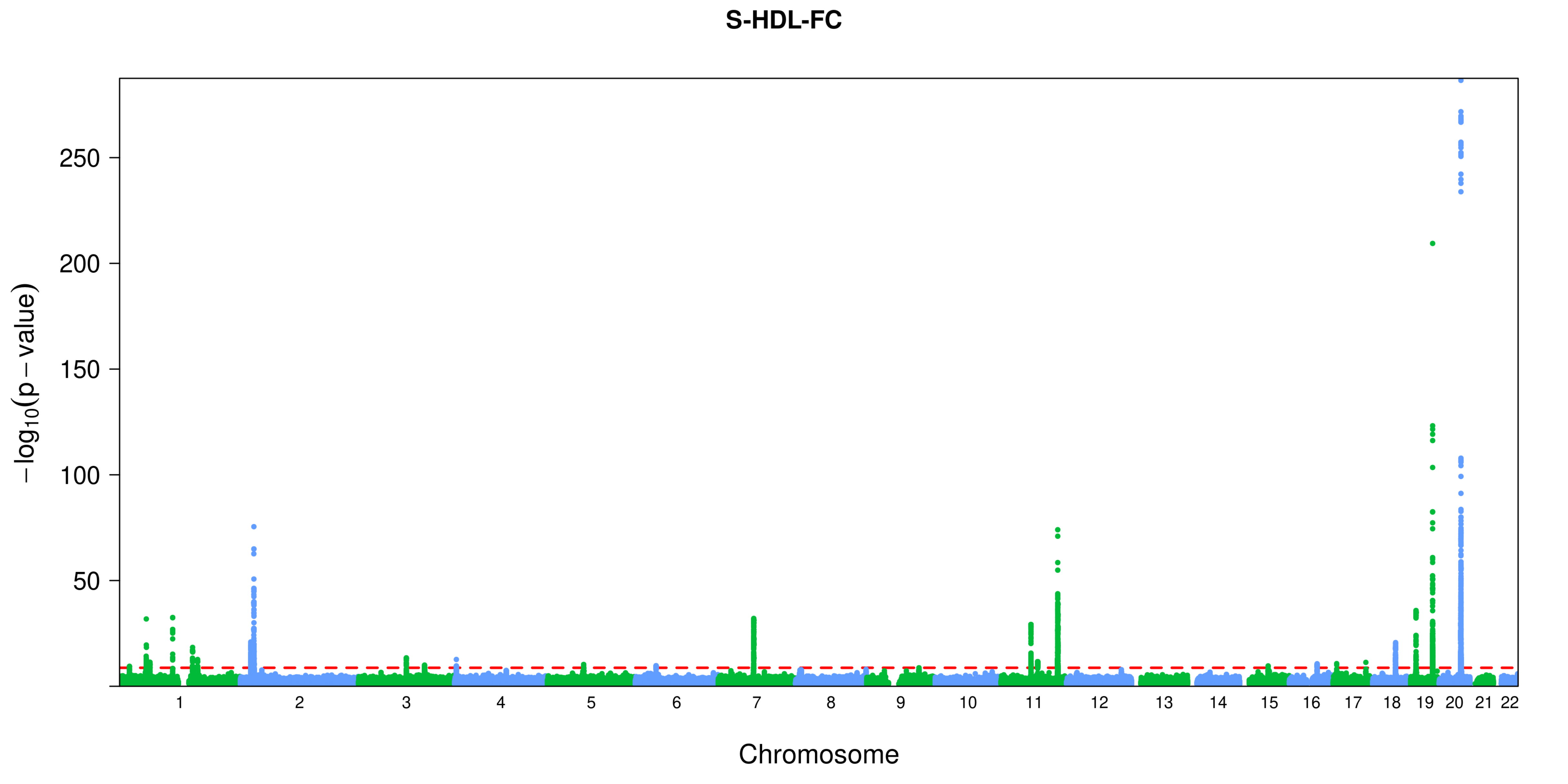

S-HDL-FC
−log10(p − value)
250
200
150
100
50
Chromosome

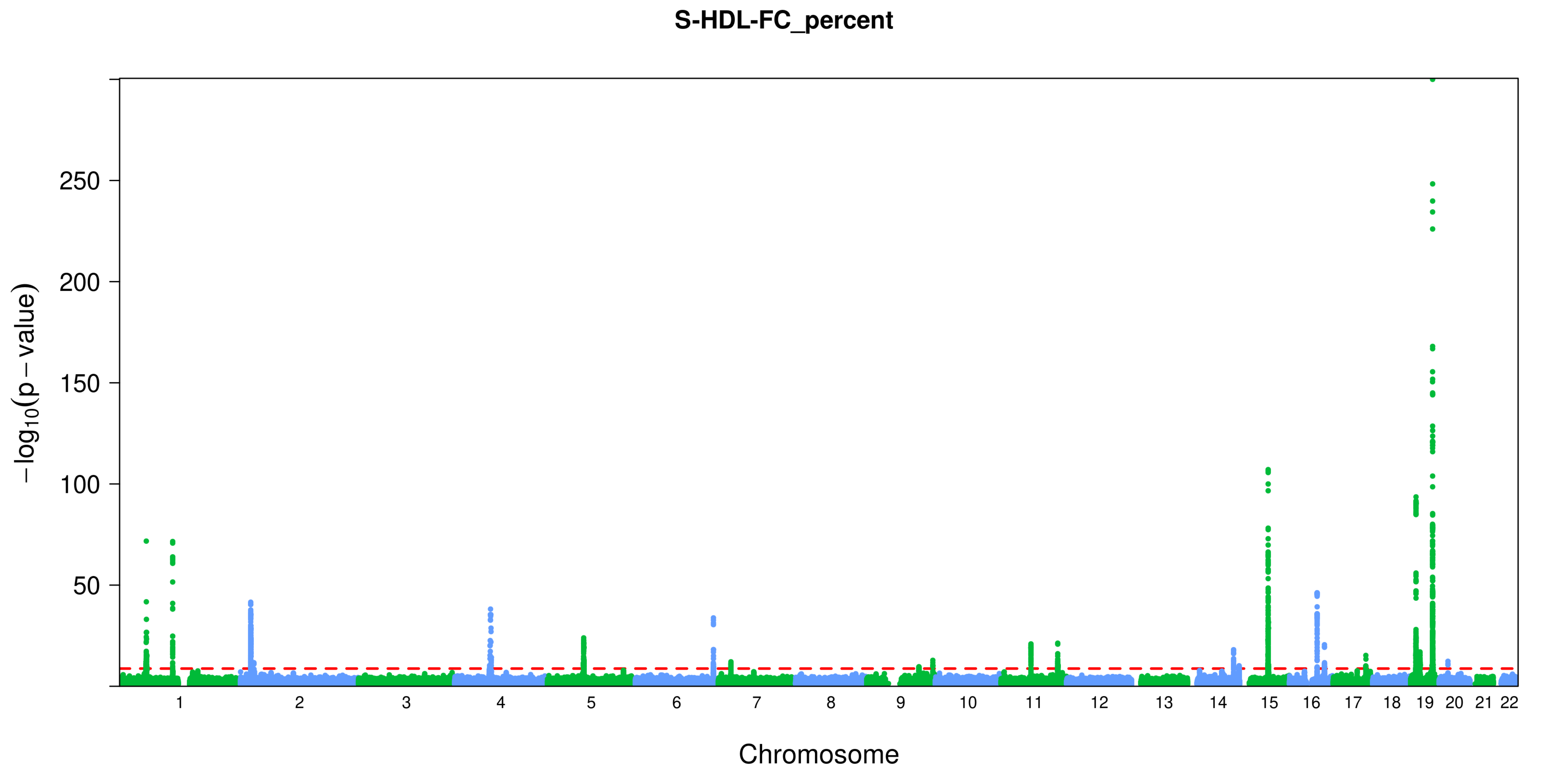
S-HDL-FC_percent
−log10(p − value)
250
200
150
100
50
Chromosome

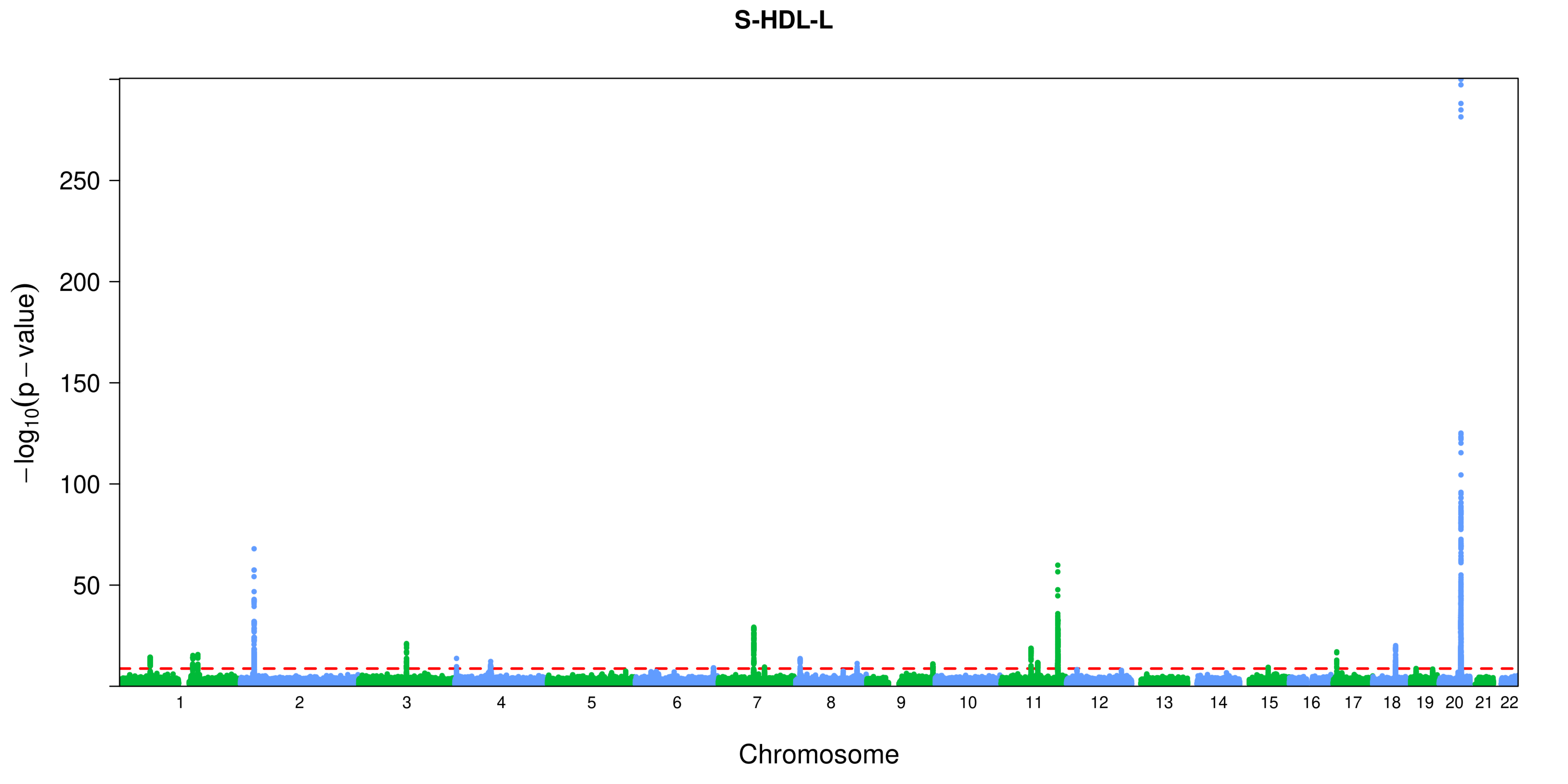
S-HDL-L
−log10(p − value)
250
200
150
100
50
Chromosome

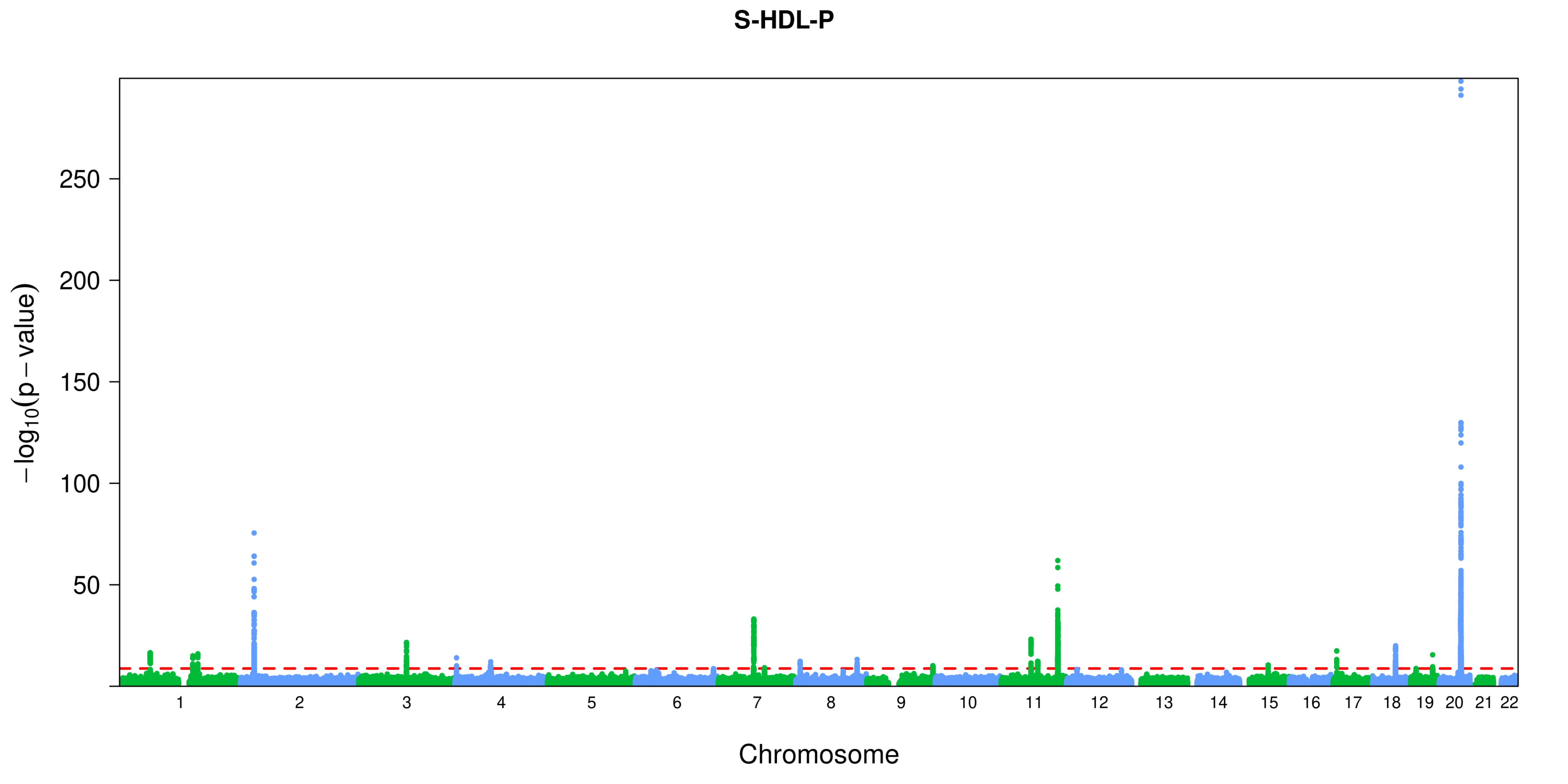

S-HDL-P
−log10(p − value)
250
200
150
100
50
Chromosome

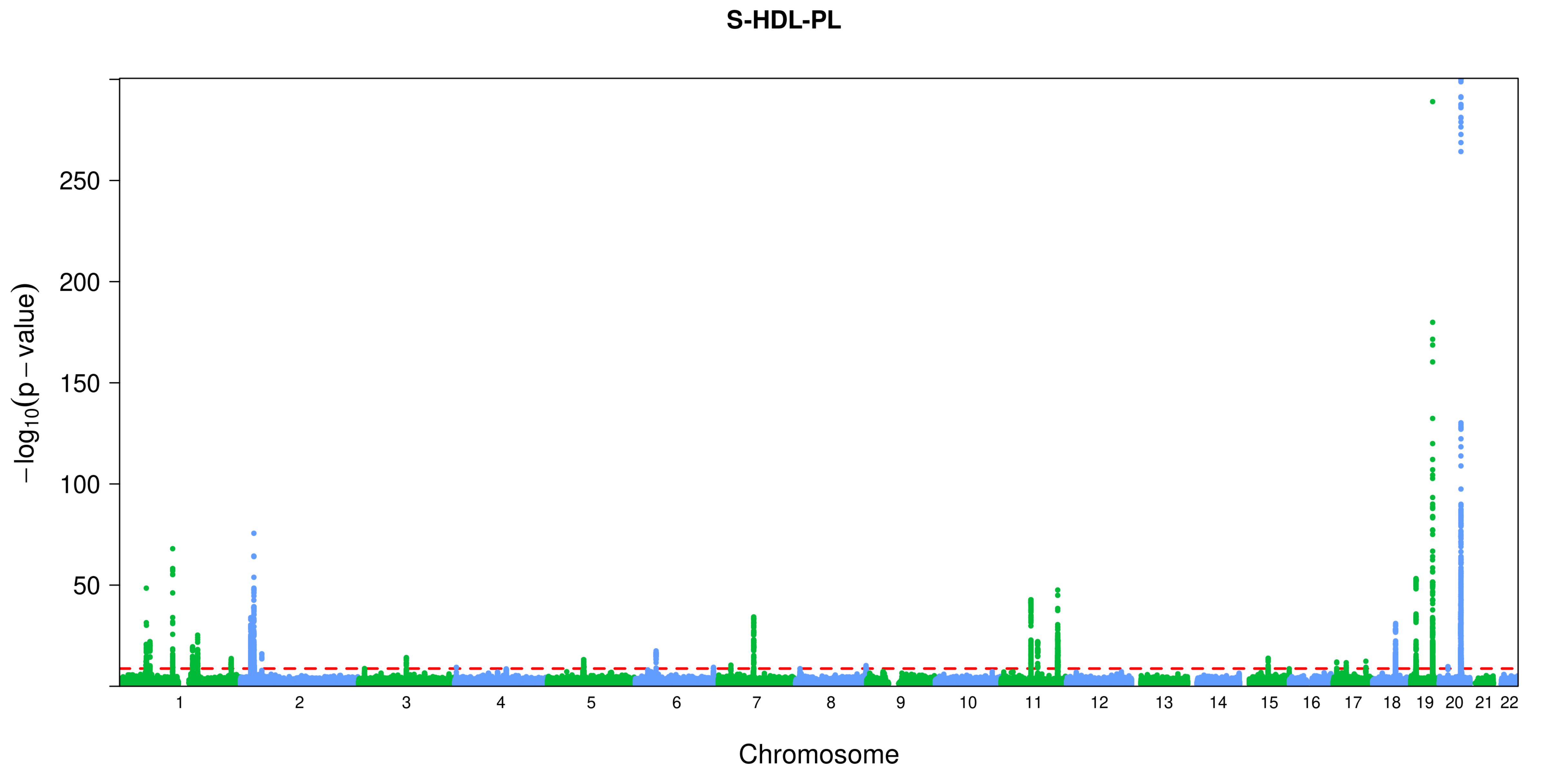
S-HDL-PL
250
200
150
100
50
−log10(p − value)
Chromosome

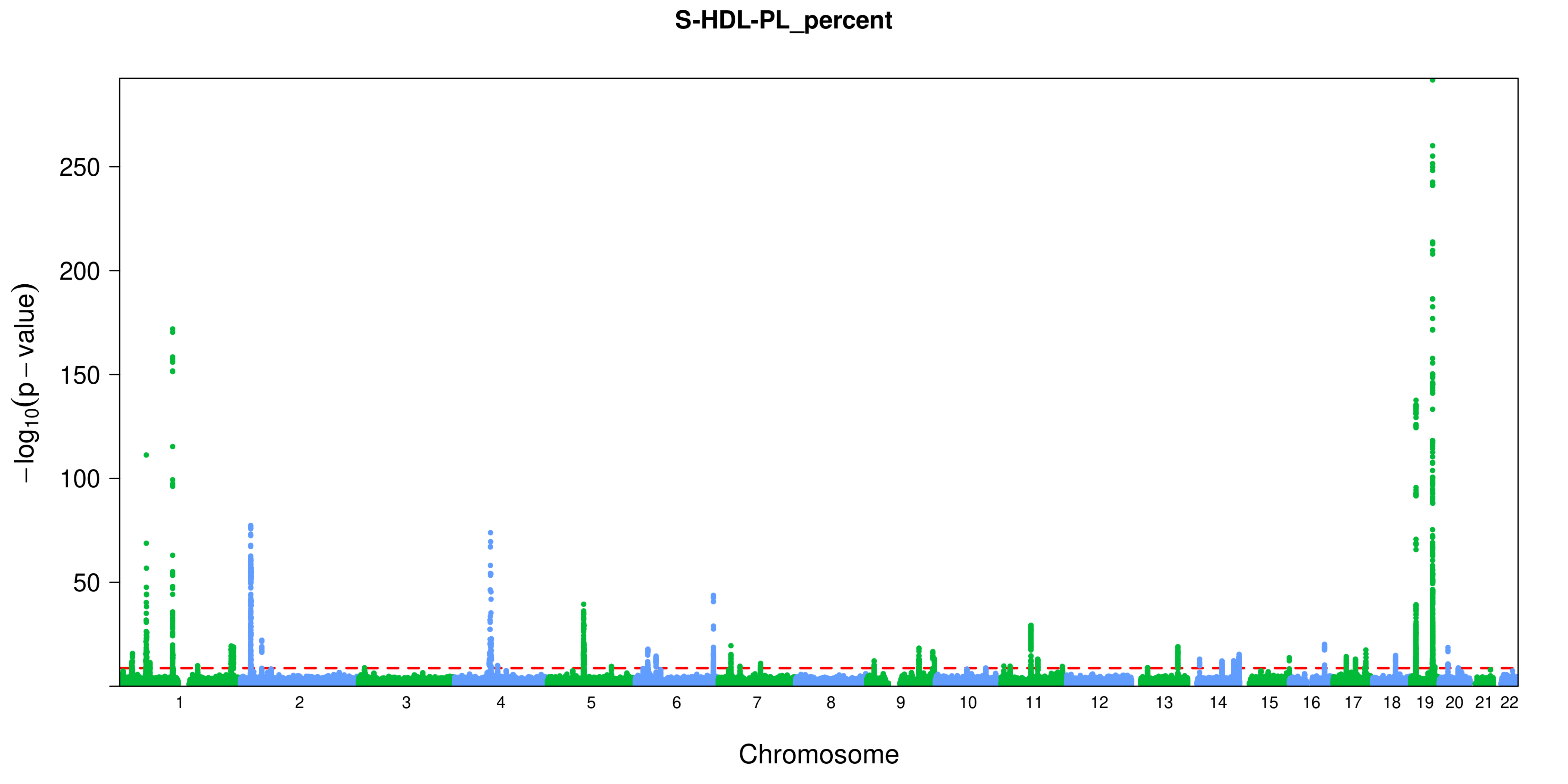
S-HDL-PL_percent
−log10(p − value)
250
200
150
100
50
Chromosome

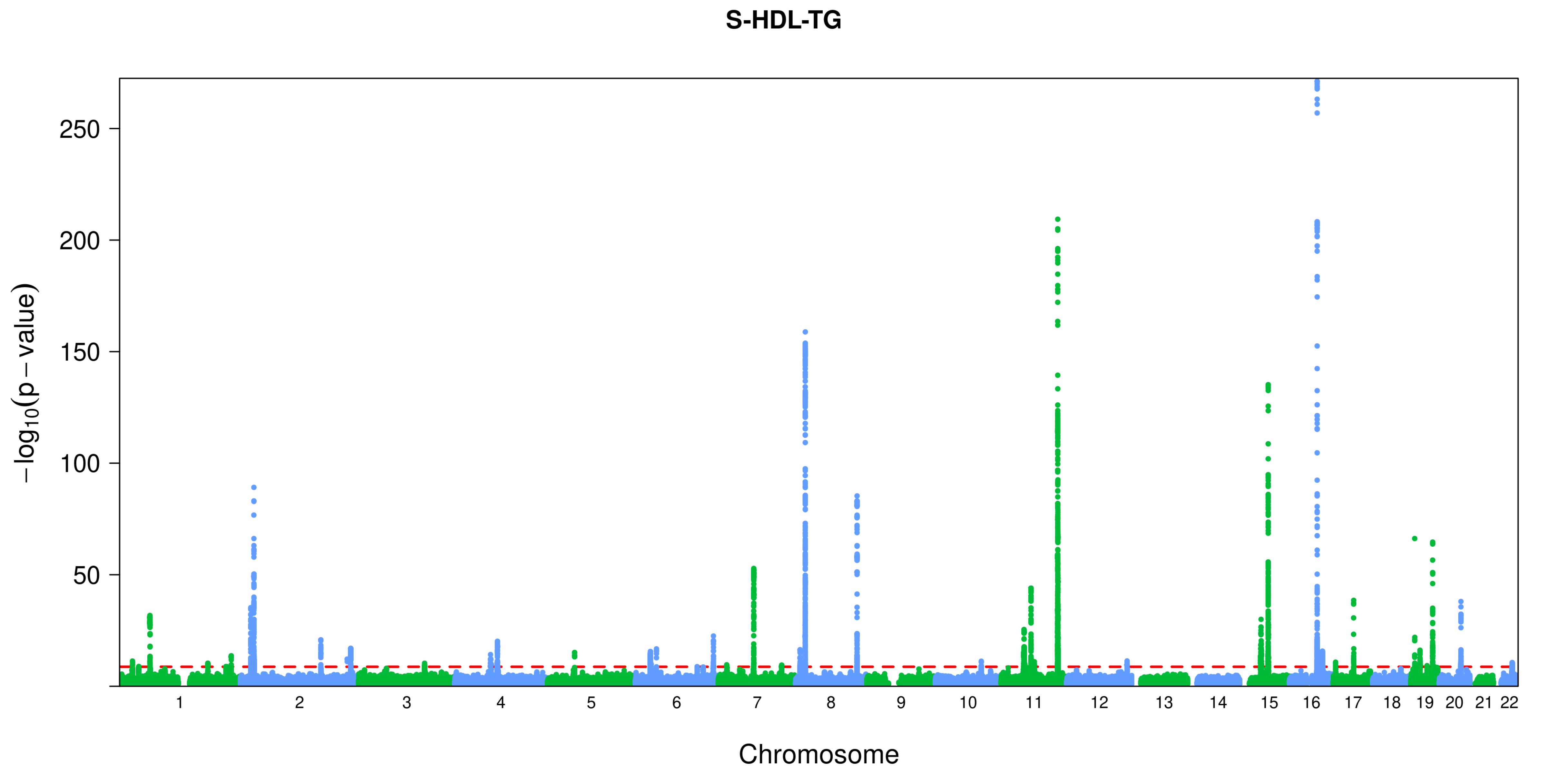

S-HDL-TG
−log10(p − value)
250
200
150
100
50
Chromosome

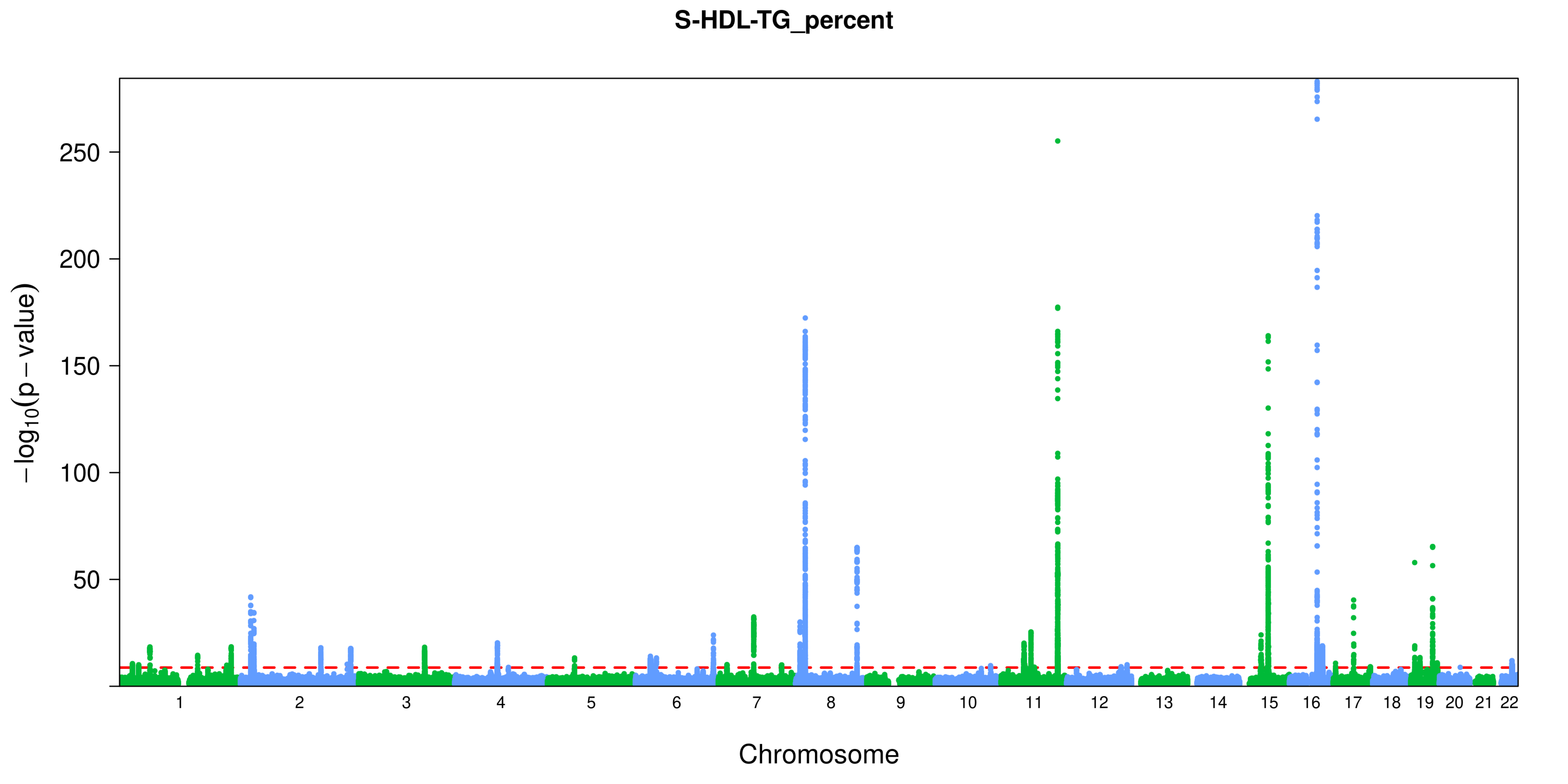
S-HDL-TG_percent
250
200
150
100
50
−log10(p − value)
1 2 3 4 5 6 7 8 9 10 11 12 13 14 15 16 17 18 19 20 21 22
Chromosome

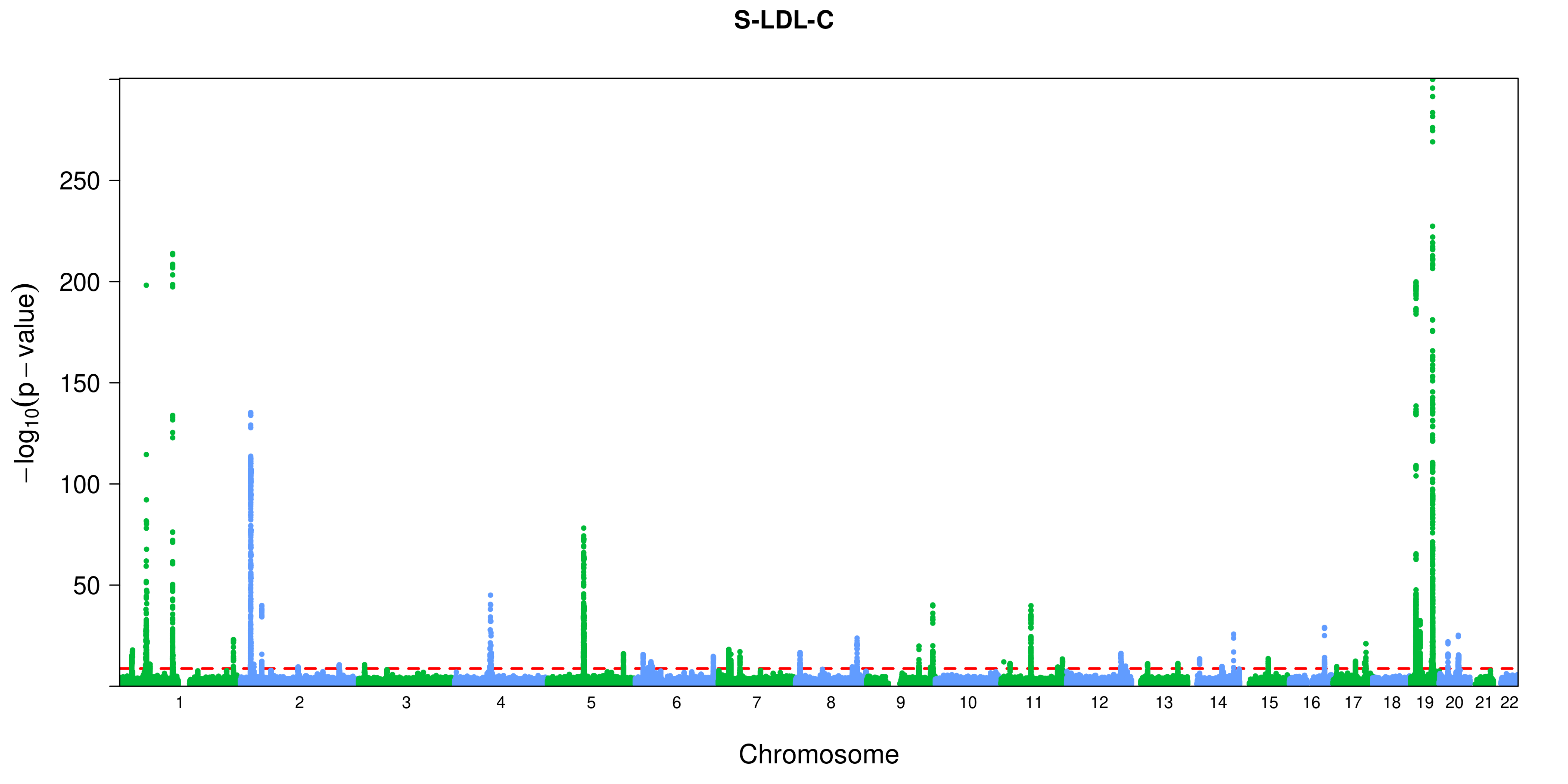
S-LDL-C
−log10(p − value)
250
200
150
100
50
Chromosome

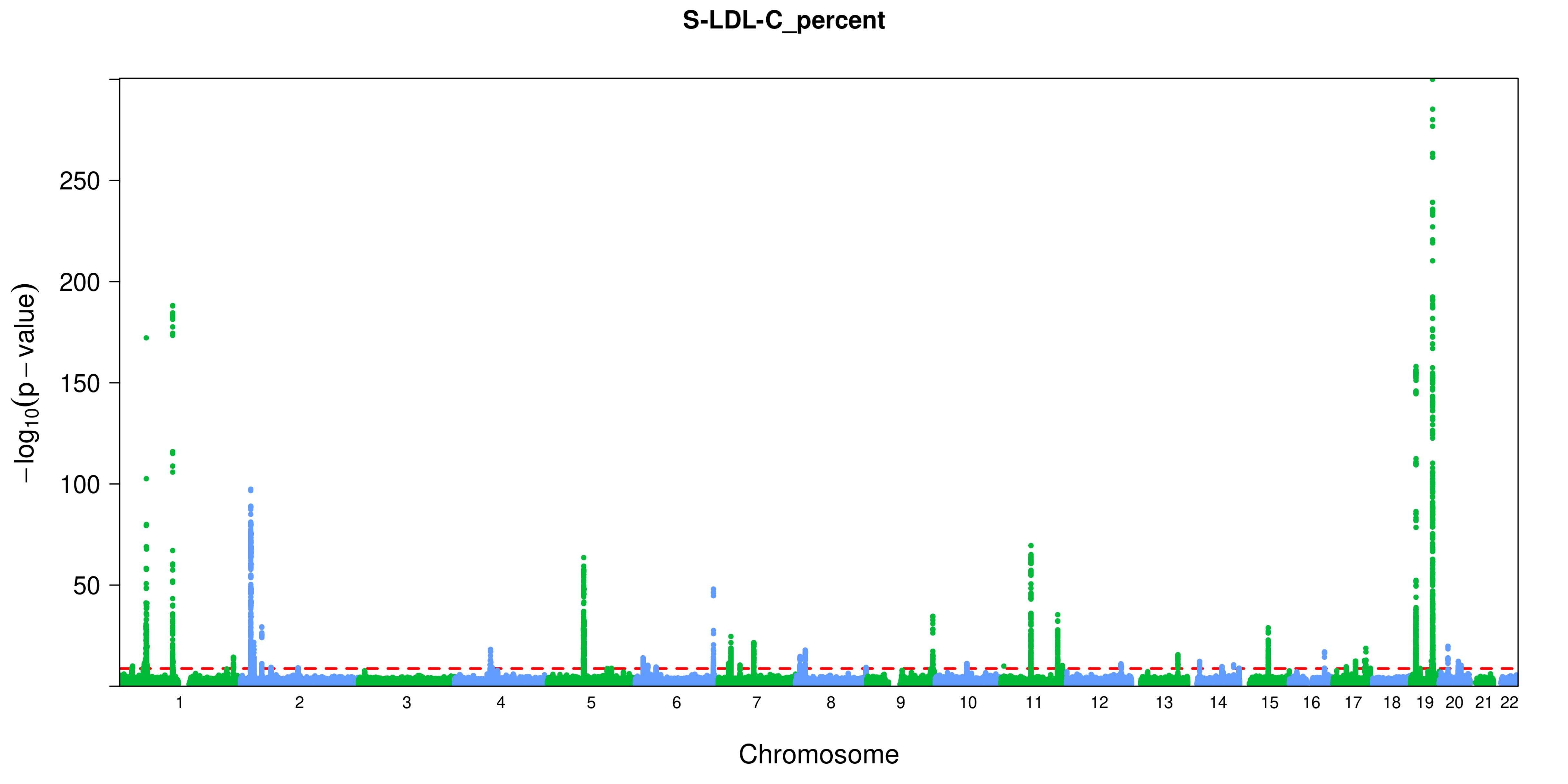
S-LDL-C_percent
−log10(p − value)
250
200
150
100
50
Chromosome

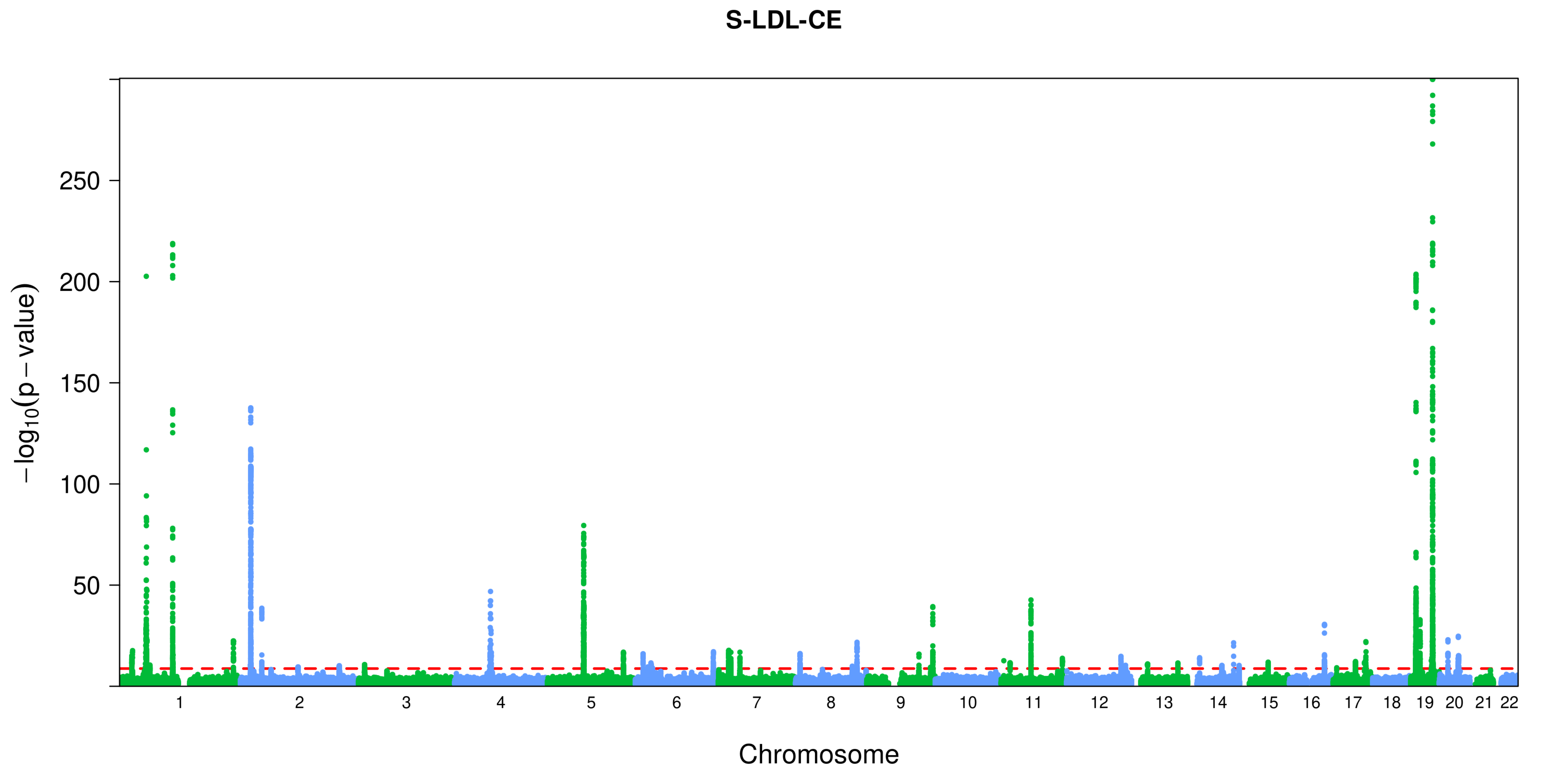
S-LDL-CE
250
200
150
100
50
$-\log_{10}(p-\text{value})$
1 2 3 4 5 6 7 8 9 10 11 12 13 14 15 16 17 18 19 20 21 22
Chromosome

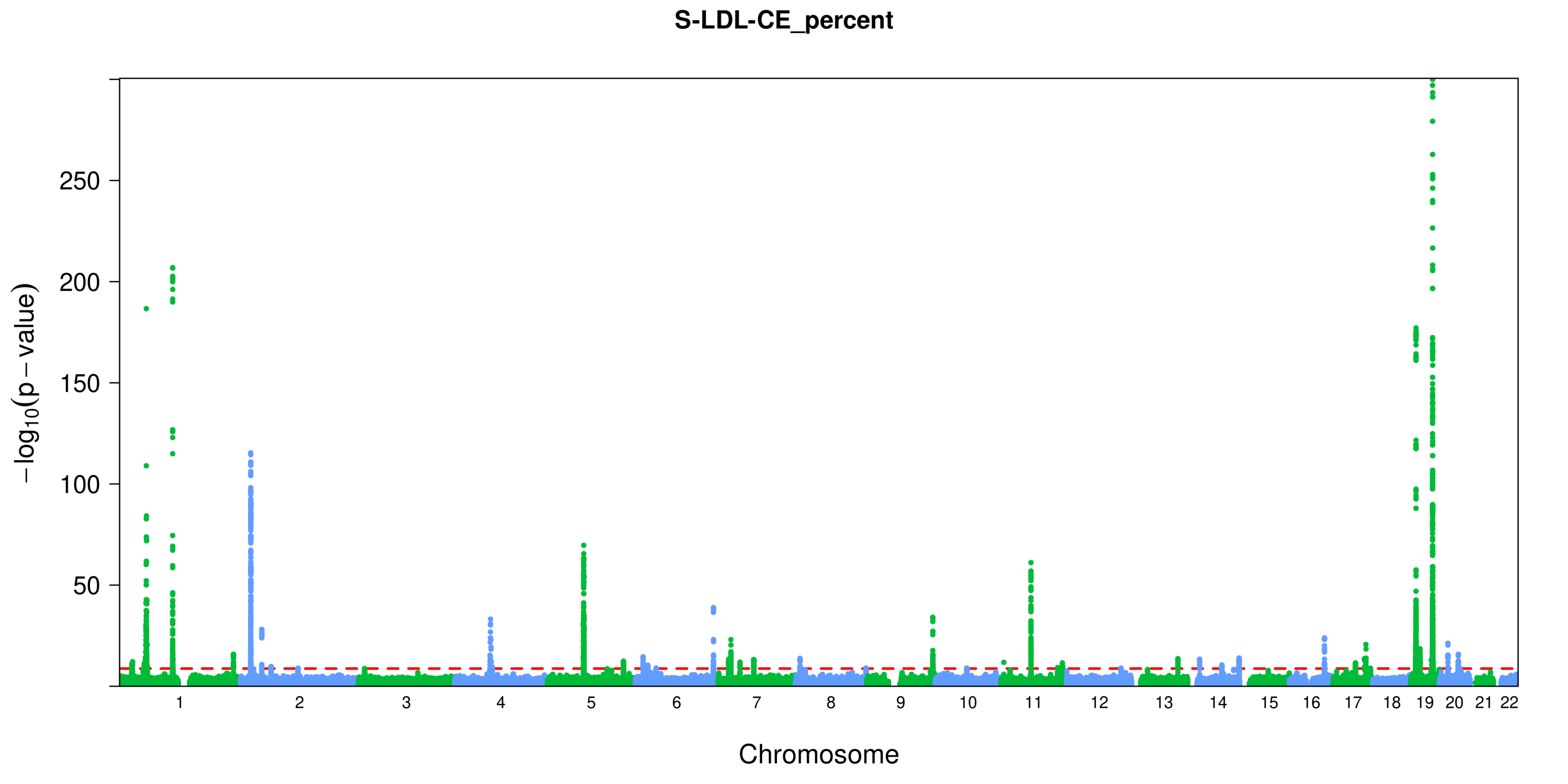

S-LDL-CE_percent
250
200
150
100
50
$-\log_{10}(p - value)$
Chromosome

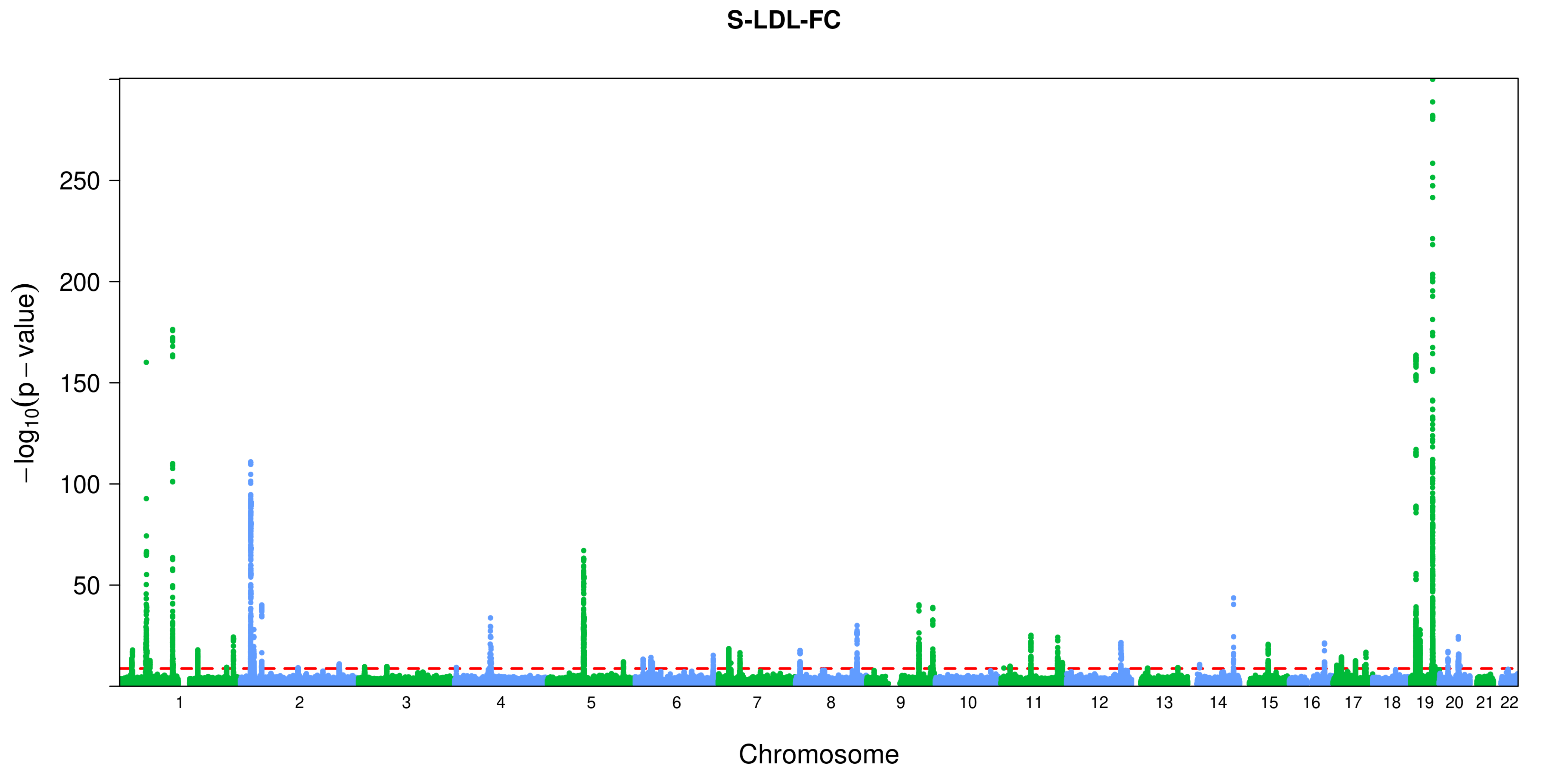

S-LDL-FC
$-\log_{10}(p-\text{value})$
250
200
150
100
50
Chromosome

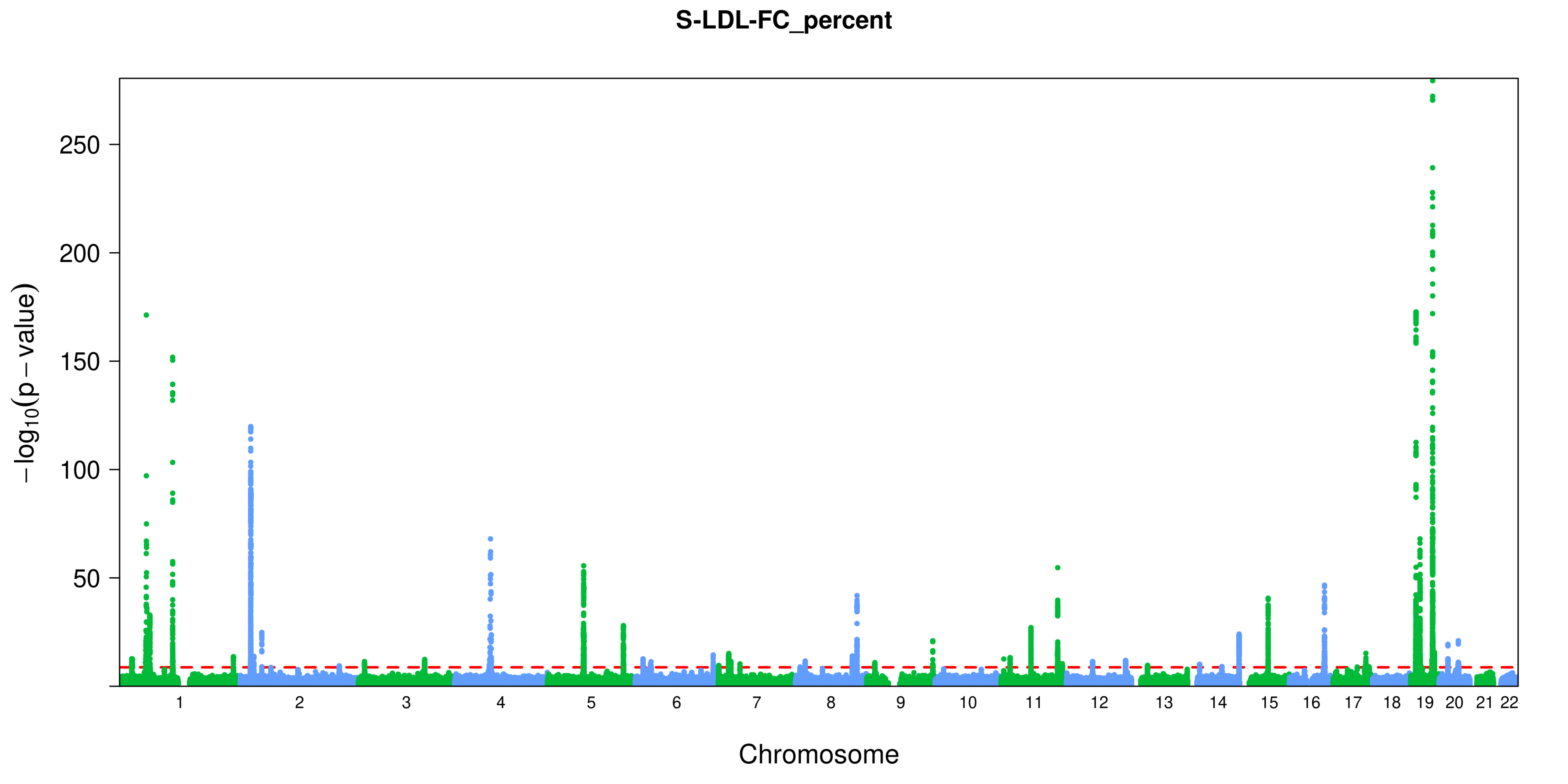
S-LDL-FC_percent
−log10(p − value)
250
200
150
100
50
Chromosome

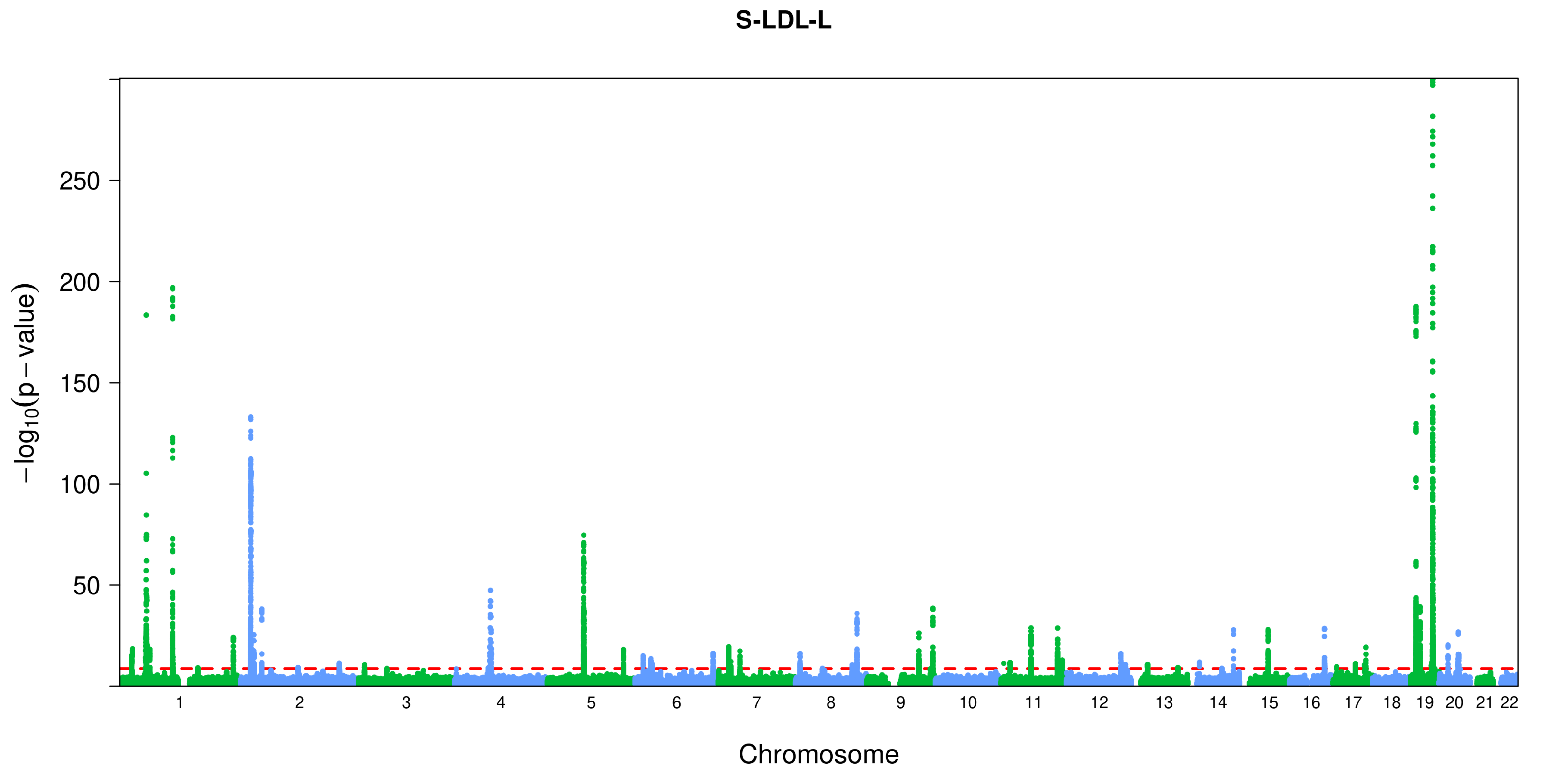
S-LDL-L
−log10(p − value)
250
200
150
100
50
Chromosome

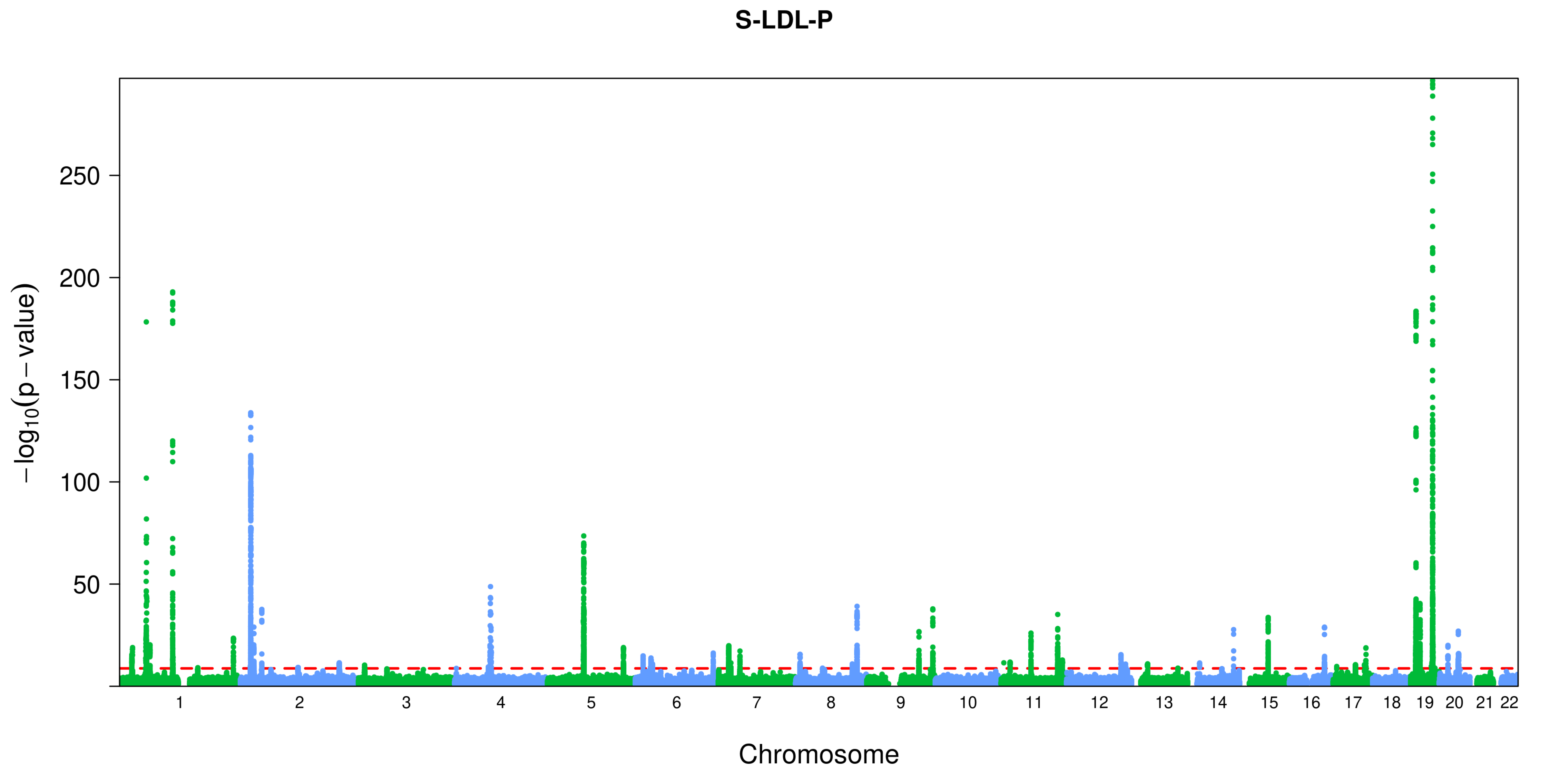
S-LDL-P
250
200
150
100
50
−log10(p − value)
Chromosome

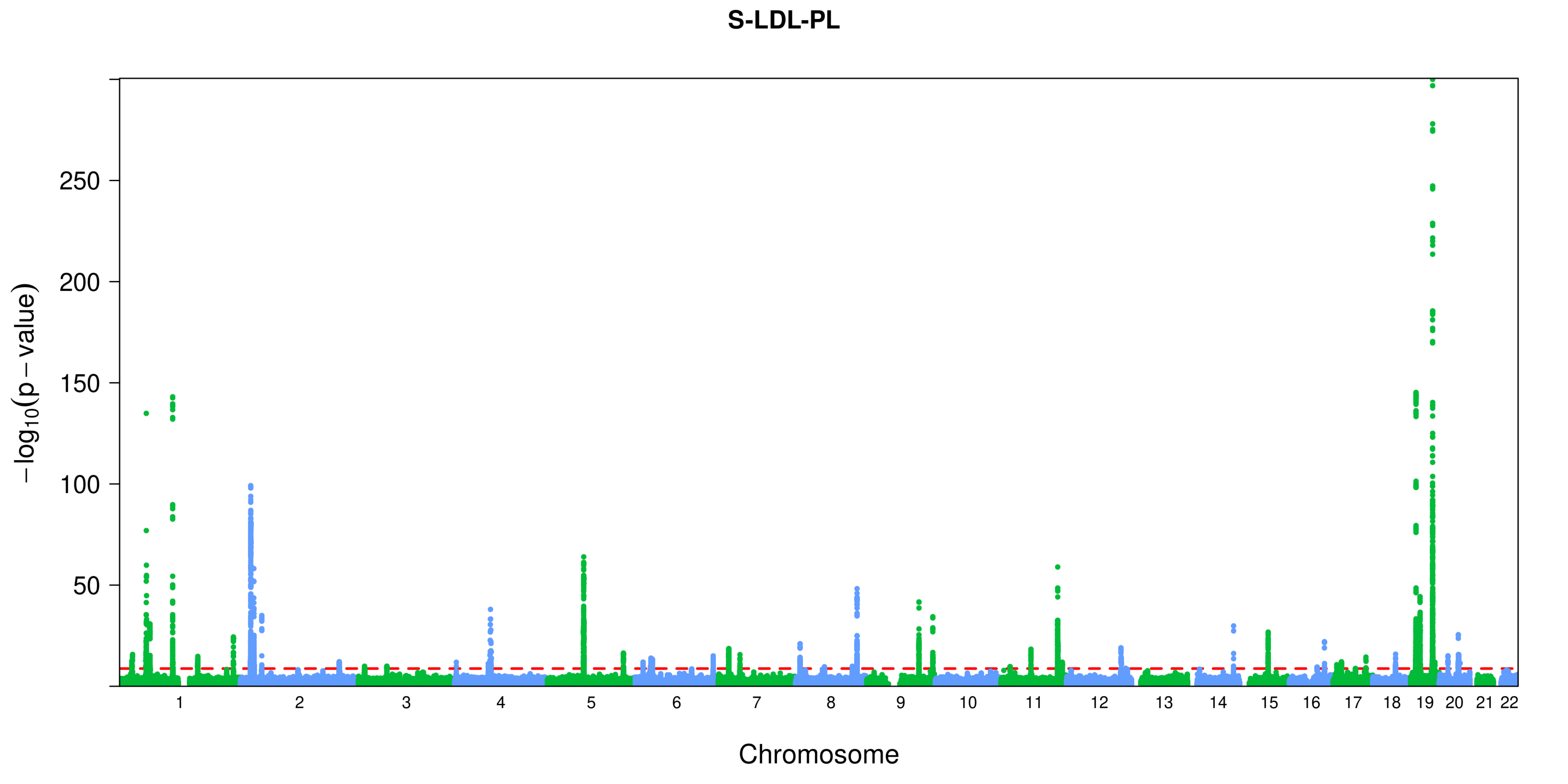
S-LDL-PL
250
200
150
100
50
−log10(p − value)
Chromosome

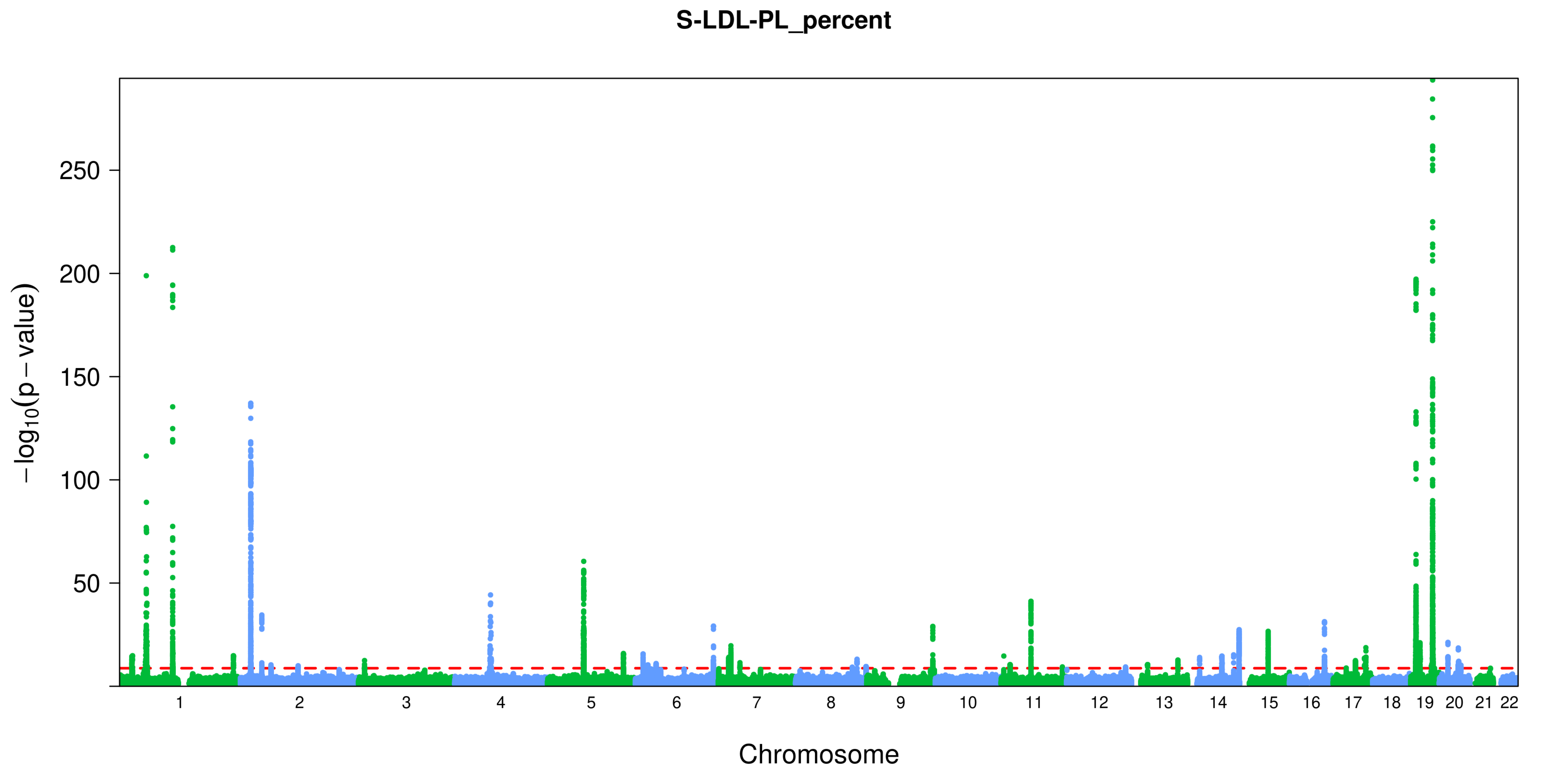
S-LDL-PL_percent
$-\log_{10}(p-\text{value})$
250
200
150
100
50
Chromosome

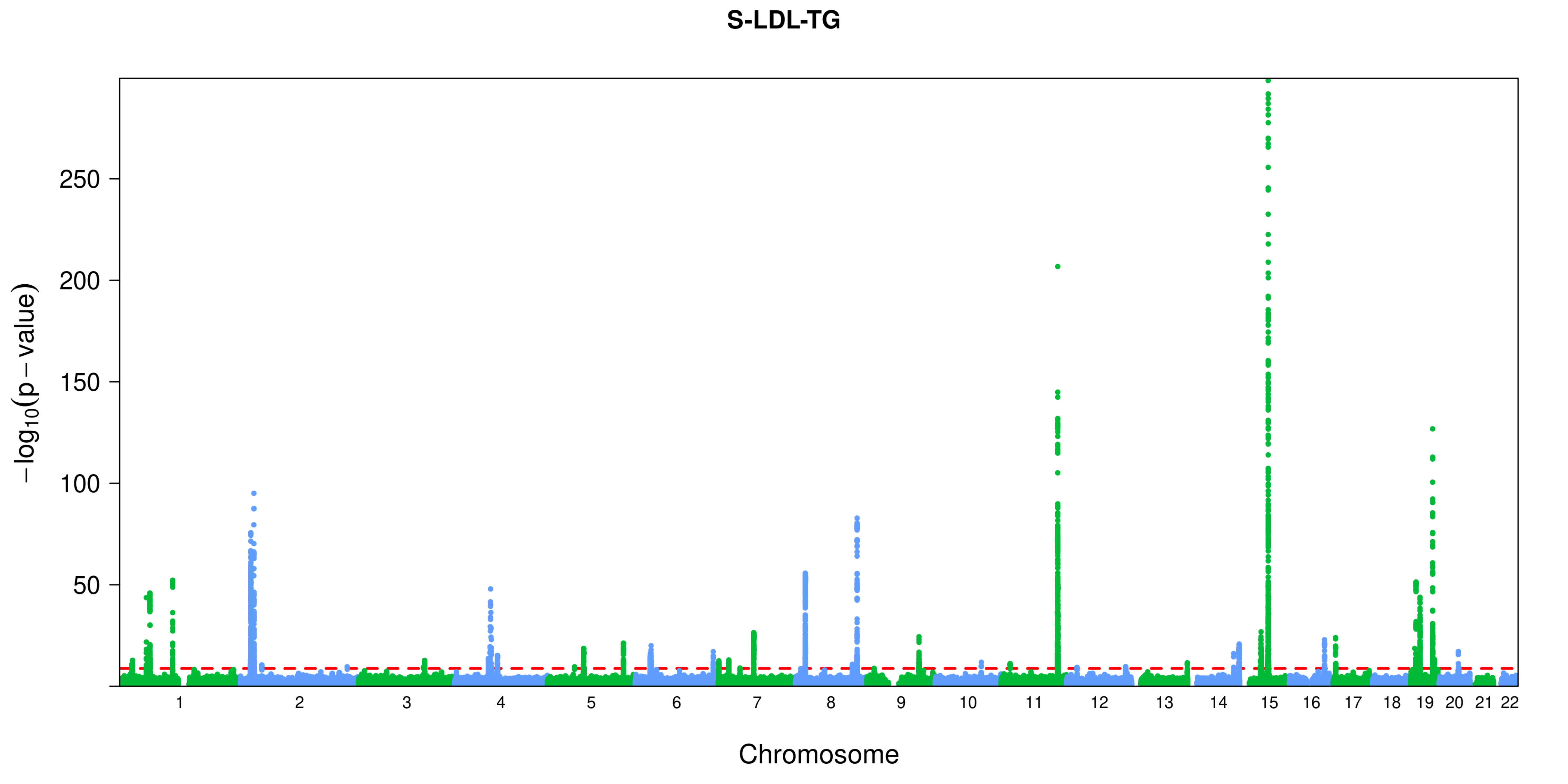

S-LDL-TG
−log10(p − value)
250
200
150
100
50
Chromosome

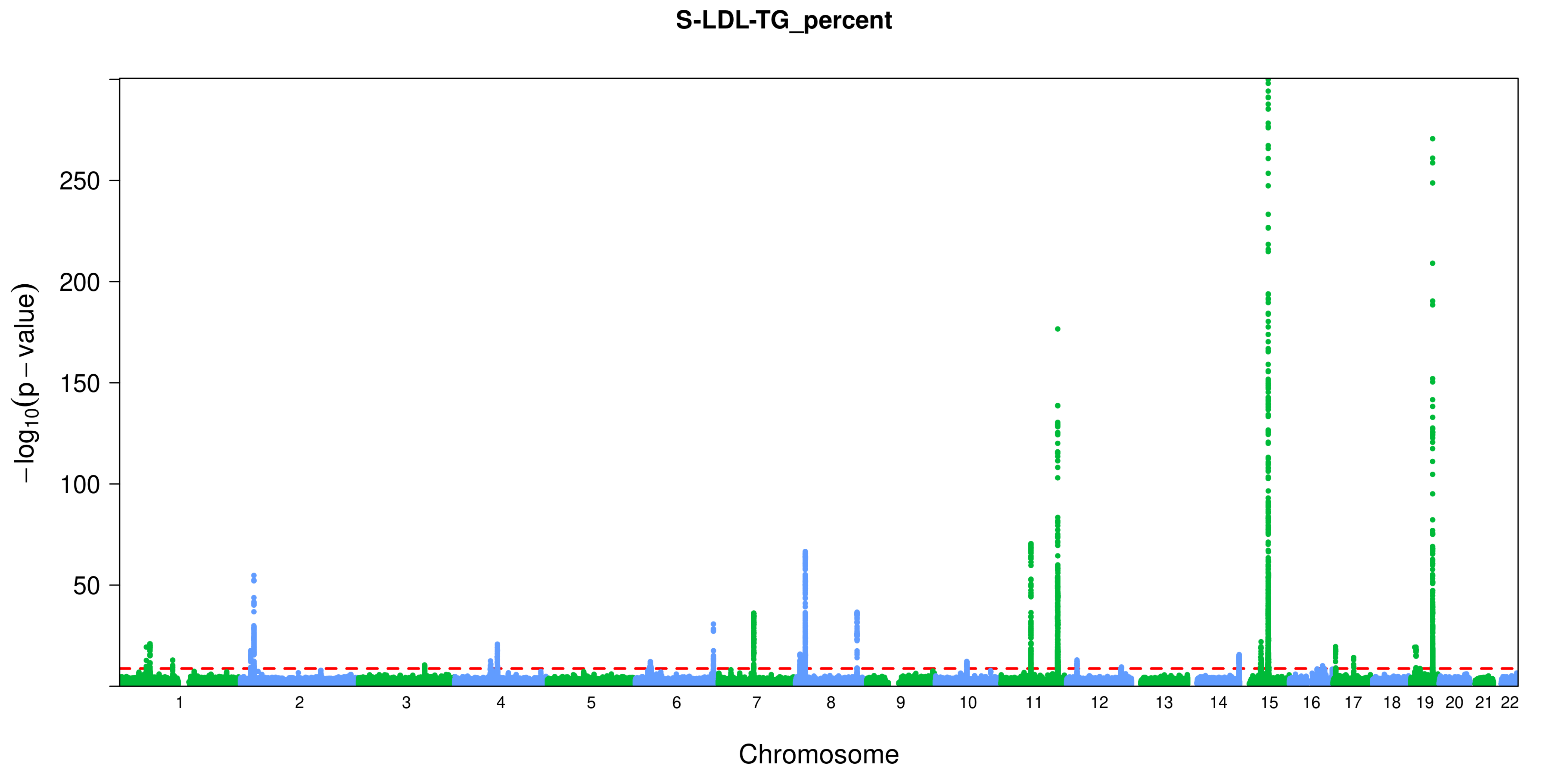
S-LDL-TG_percent
$-\log_{10}(p-\text{value})$
250
200
150
100
50
Chromosome

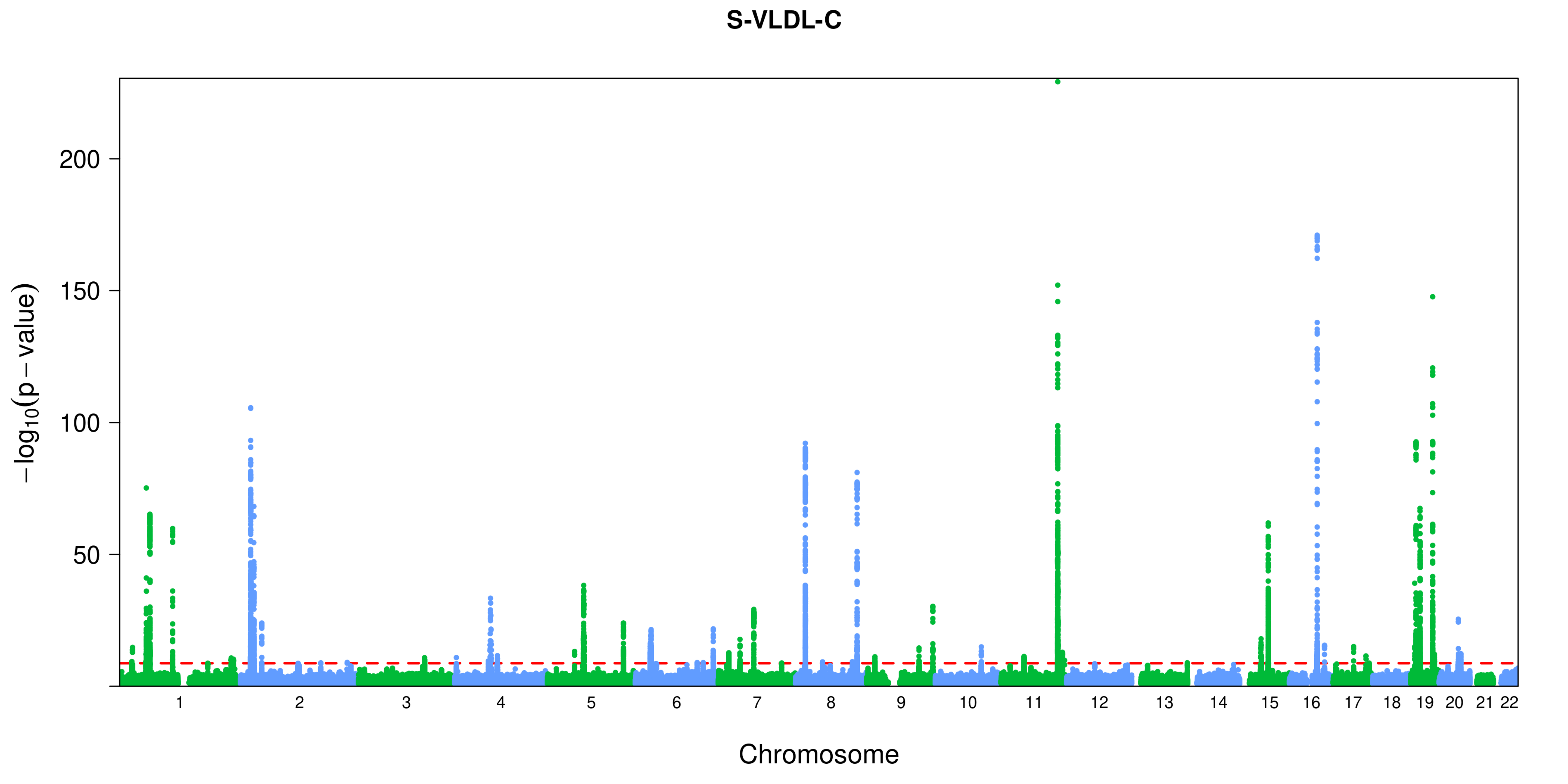
S-VLDL-C
−log10(p − value)
200
150
100
50
Chromosome

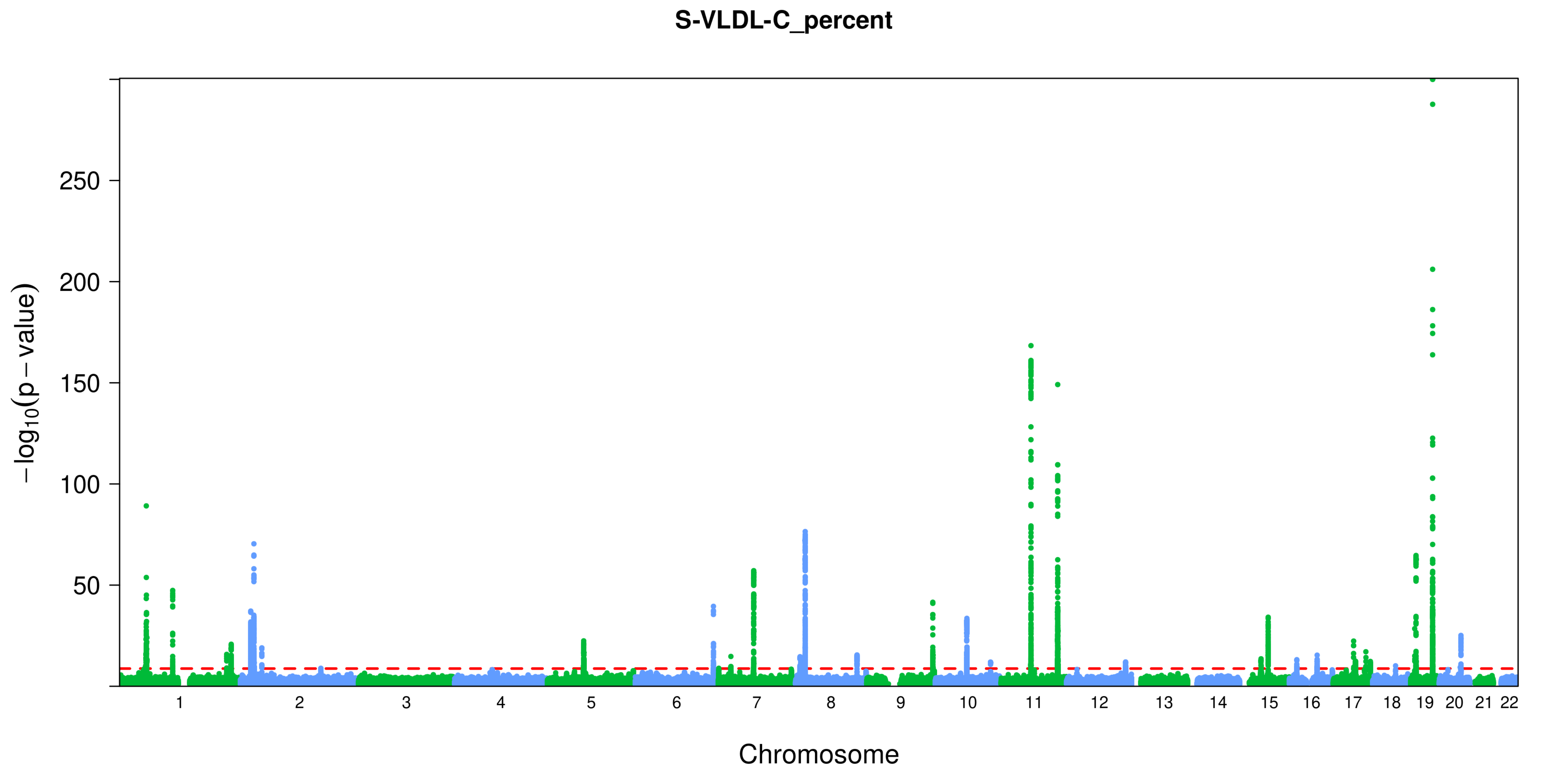

S-VLDL-C_percent
−log10(p − value)
250
200
150
100
50
1
2
3
4
5
6
7
8
9
10
11
12
13
15
16
17
18
19
20
21
22
Chromosome

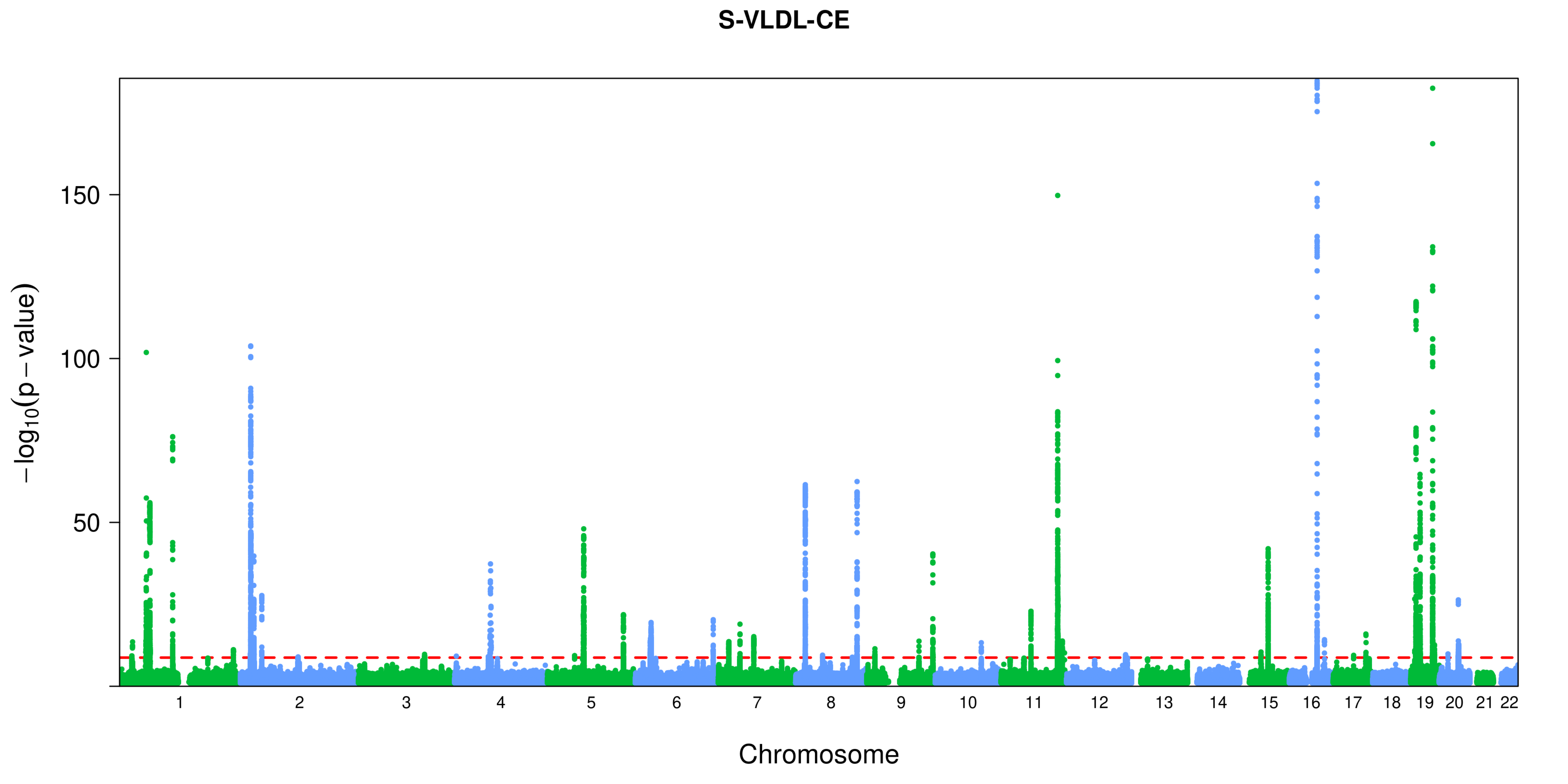
S-VLDL-CE
150
100
50
−log10(p − value)
Chromosome

**S-VLDL-CE_percent**

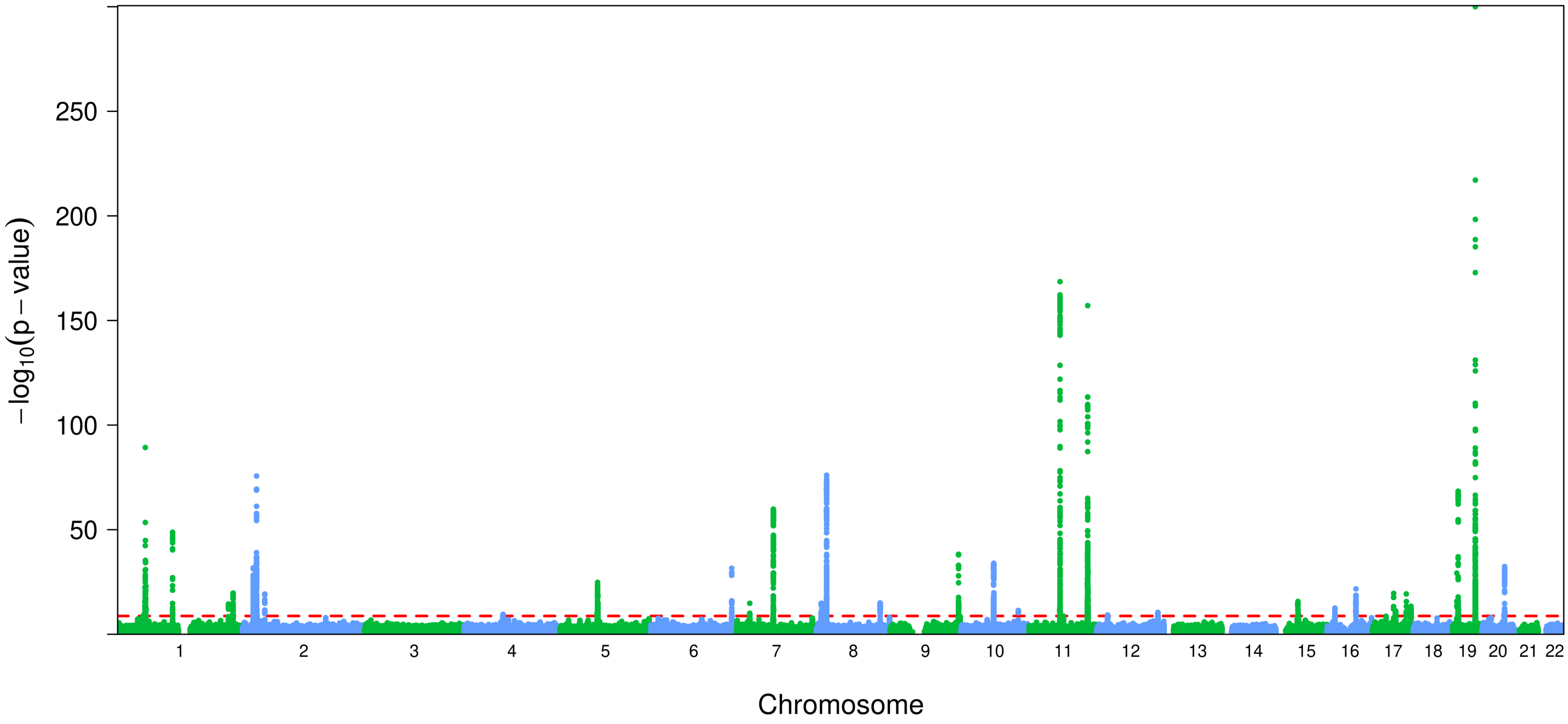

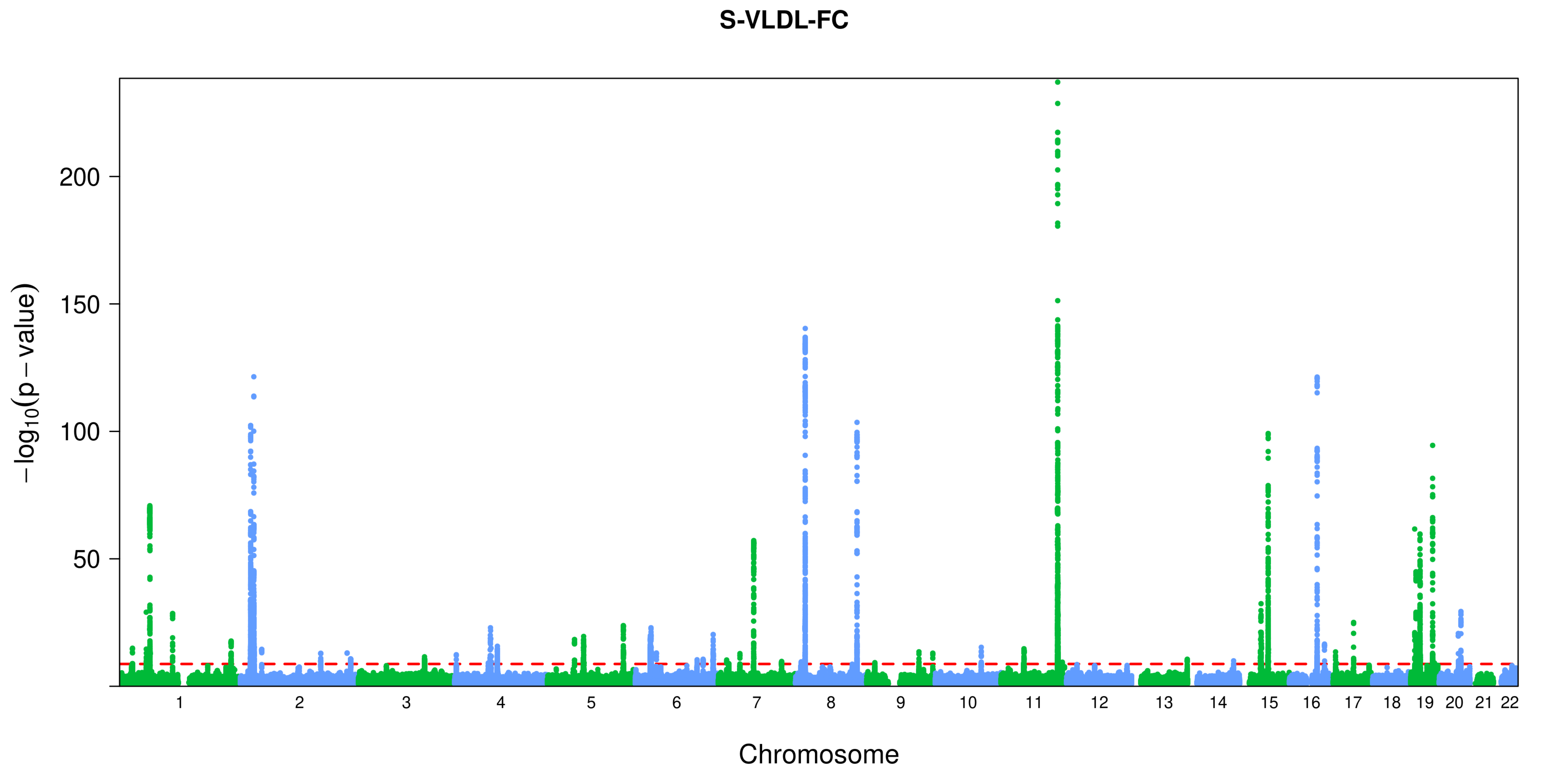

S-VLDL-FC
−log10(p − value)
200
150
100
50
Chromosome

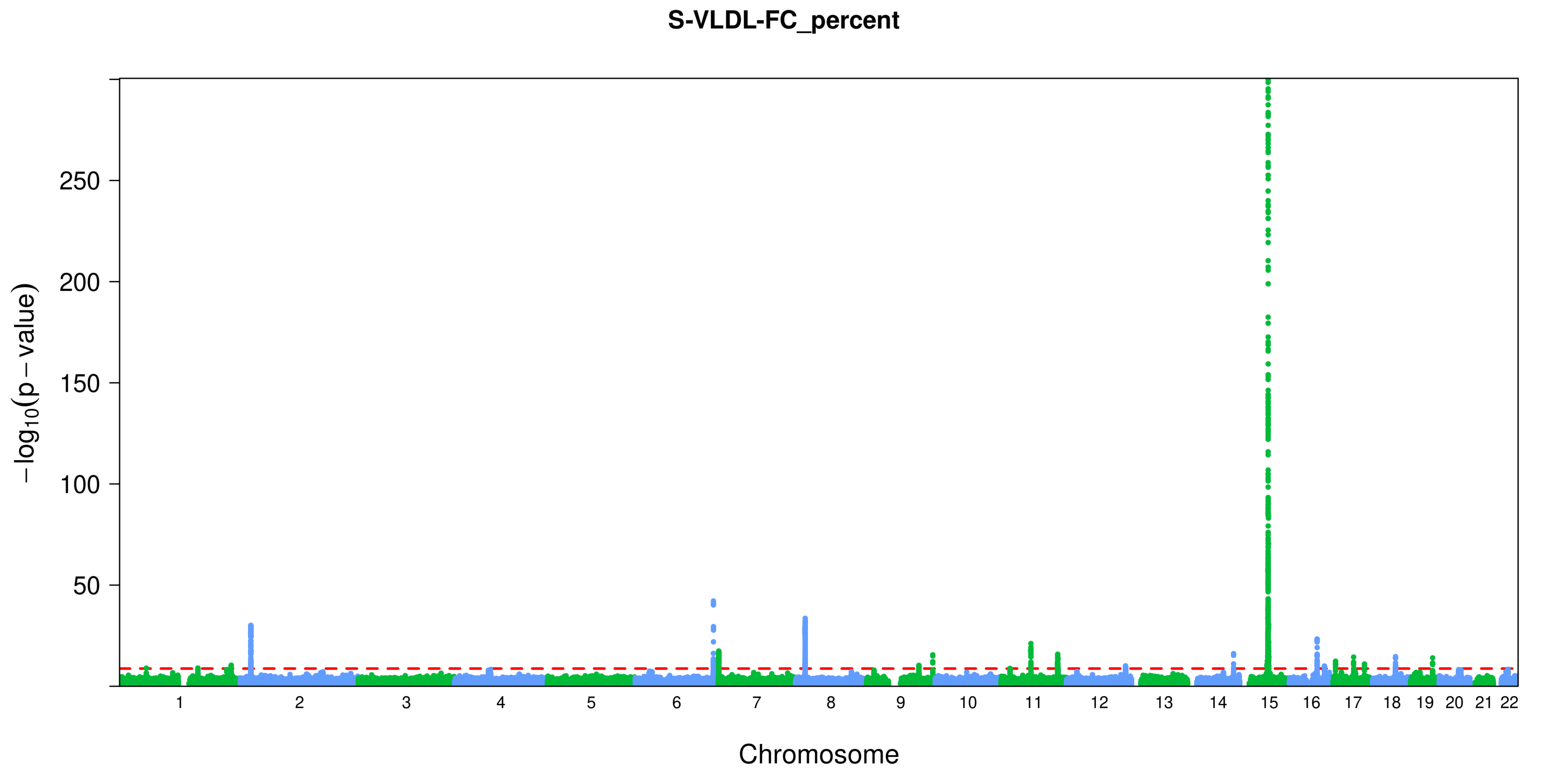
S-VLDL-FC_percent
−log10(p − value)
250
200
150
100
50
Chromosome

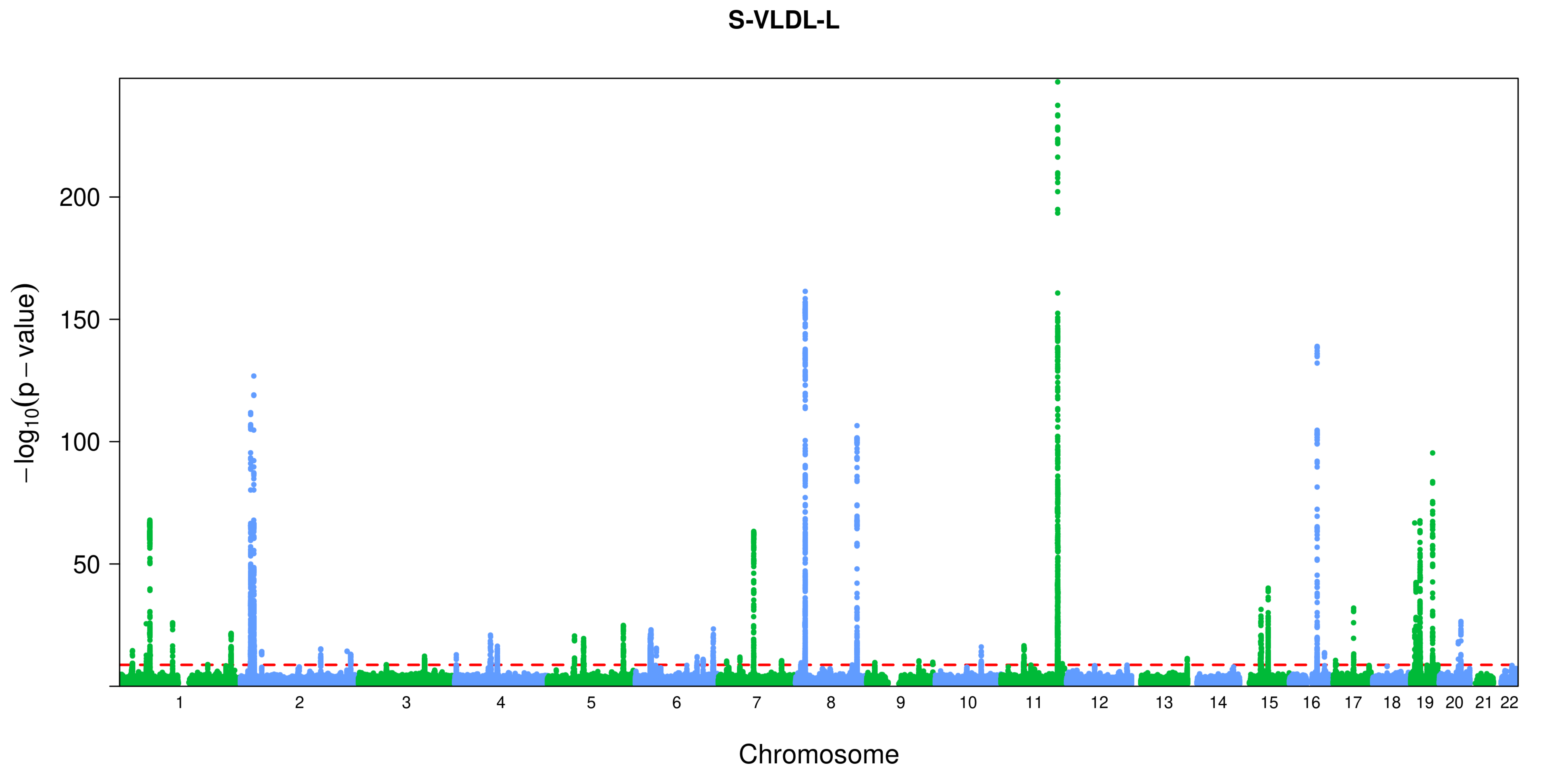
S-VLDL-L
200
150
100
50
$-\log_{10}(p-\text{value})$
Chromosome

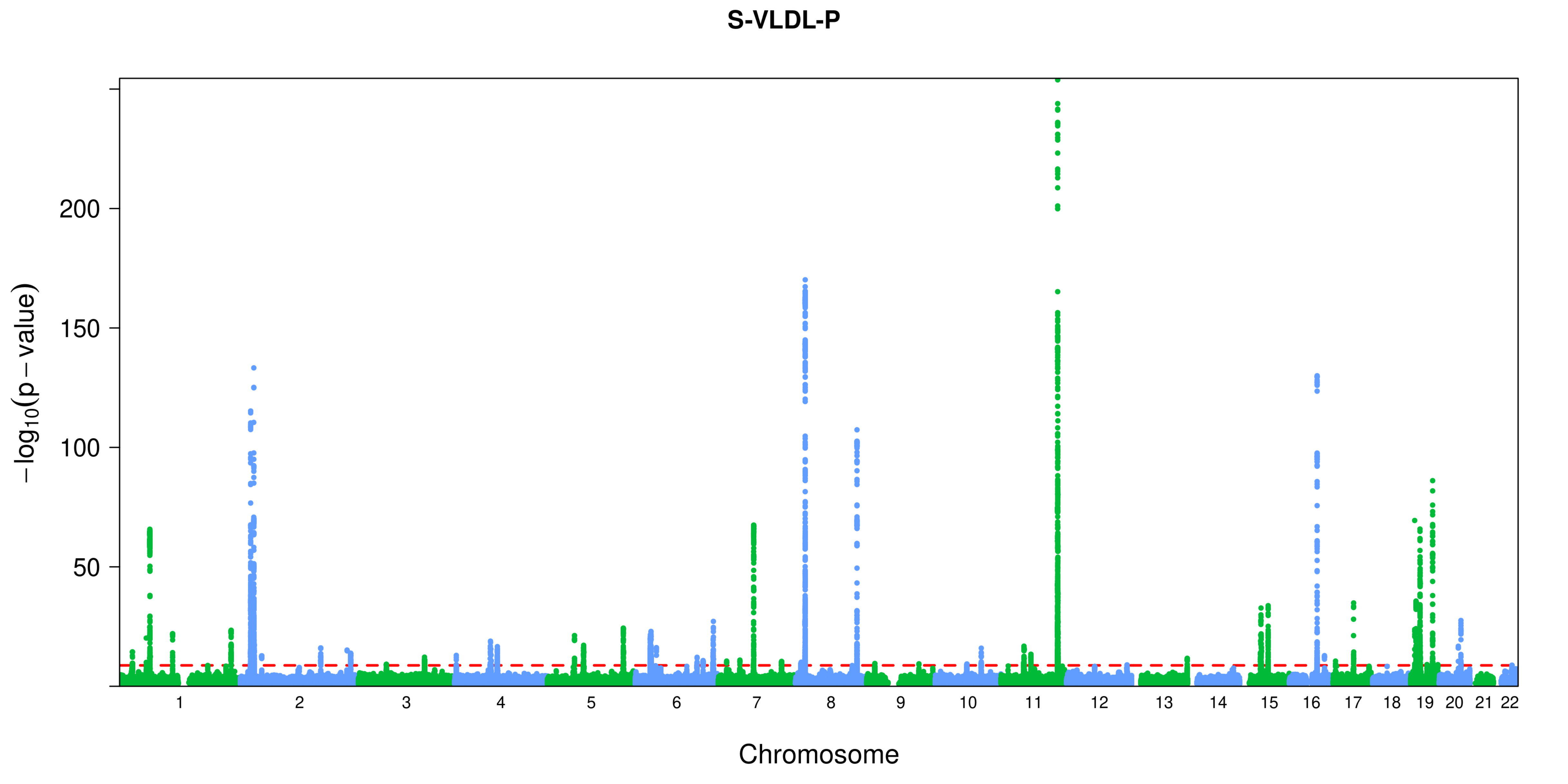
S-VLDL-P
−log10(p − value)
200
150
100
50
Chromosome

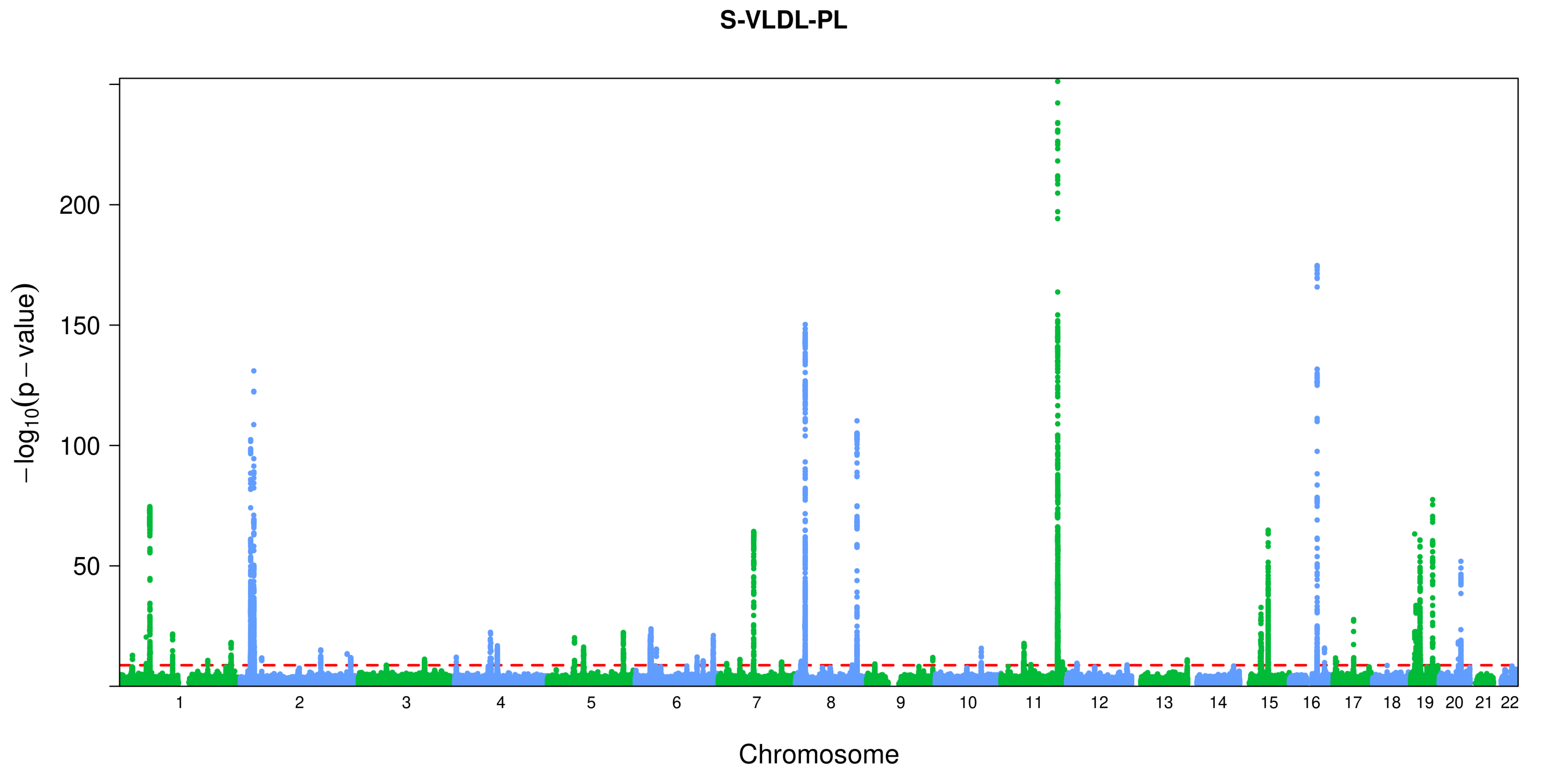
S-VLDL-PL
−log10(p − value)
200
150
100
50
Chromosome

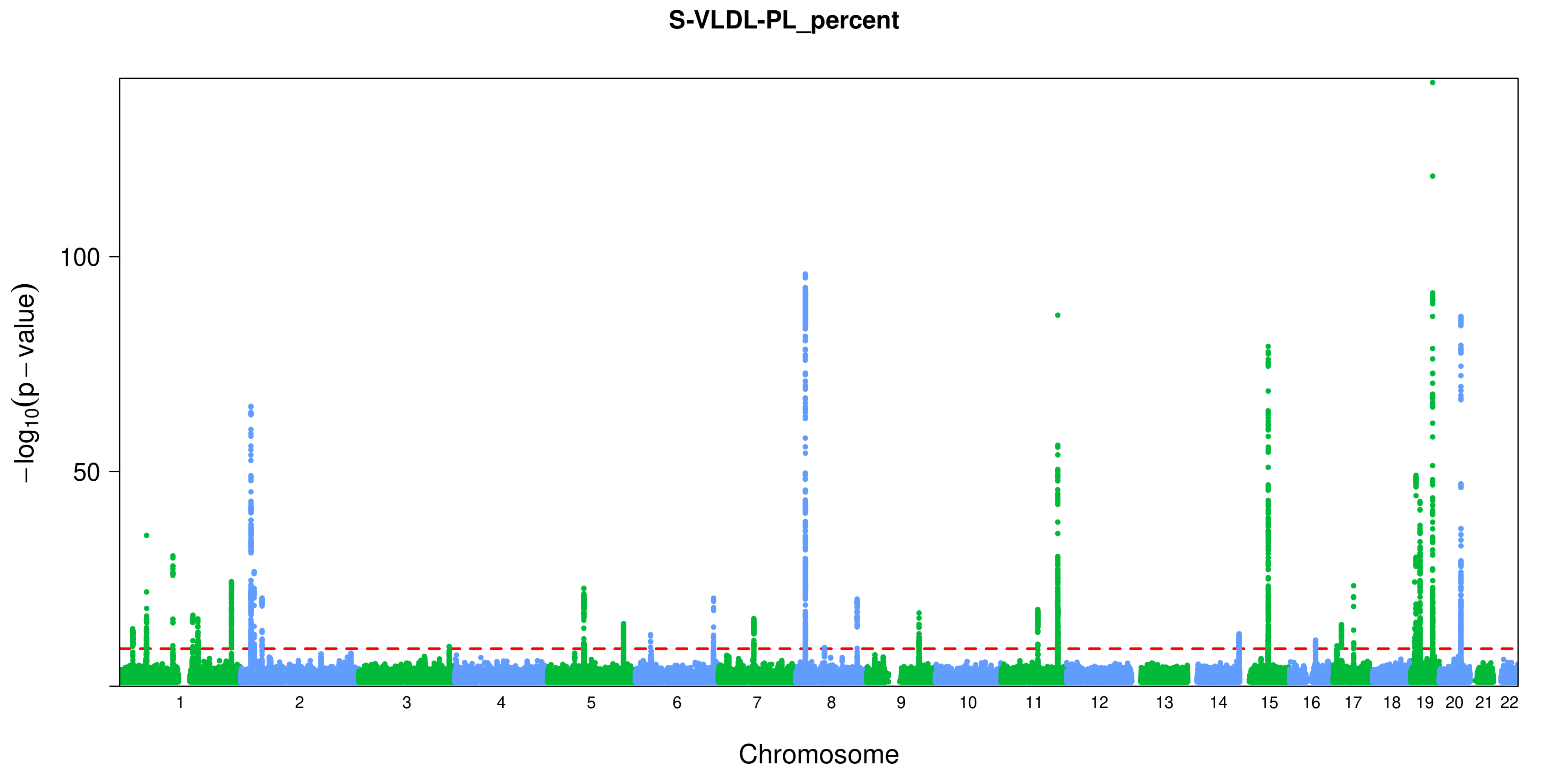
S-VLDL-PL_percent
100
50
−log10(p − value)
Chromosome

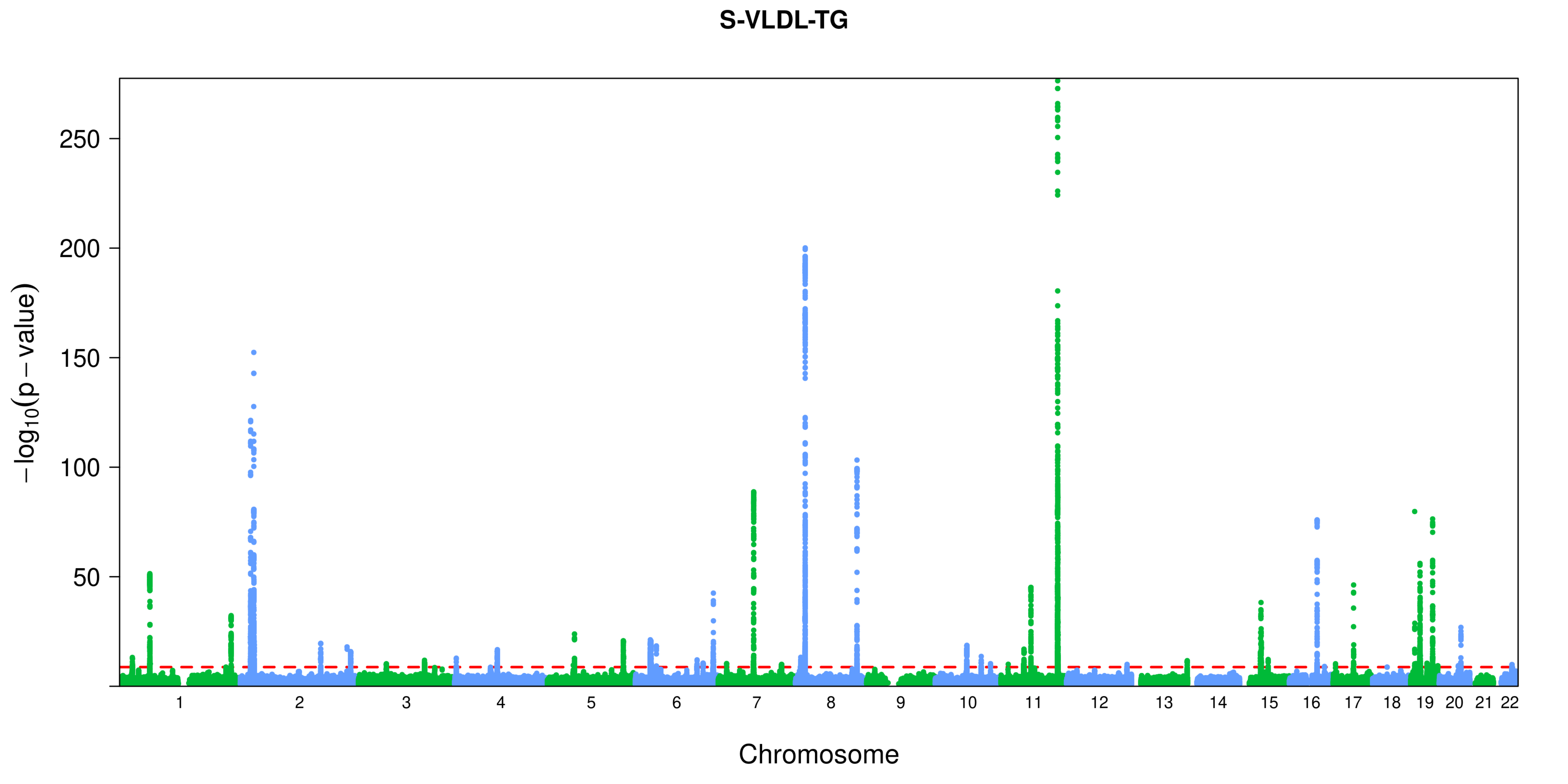
S-VLDL-TG
−log10(p − value)
250
200
150
100
50
Chromosome

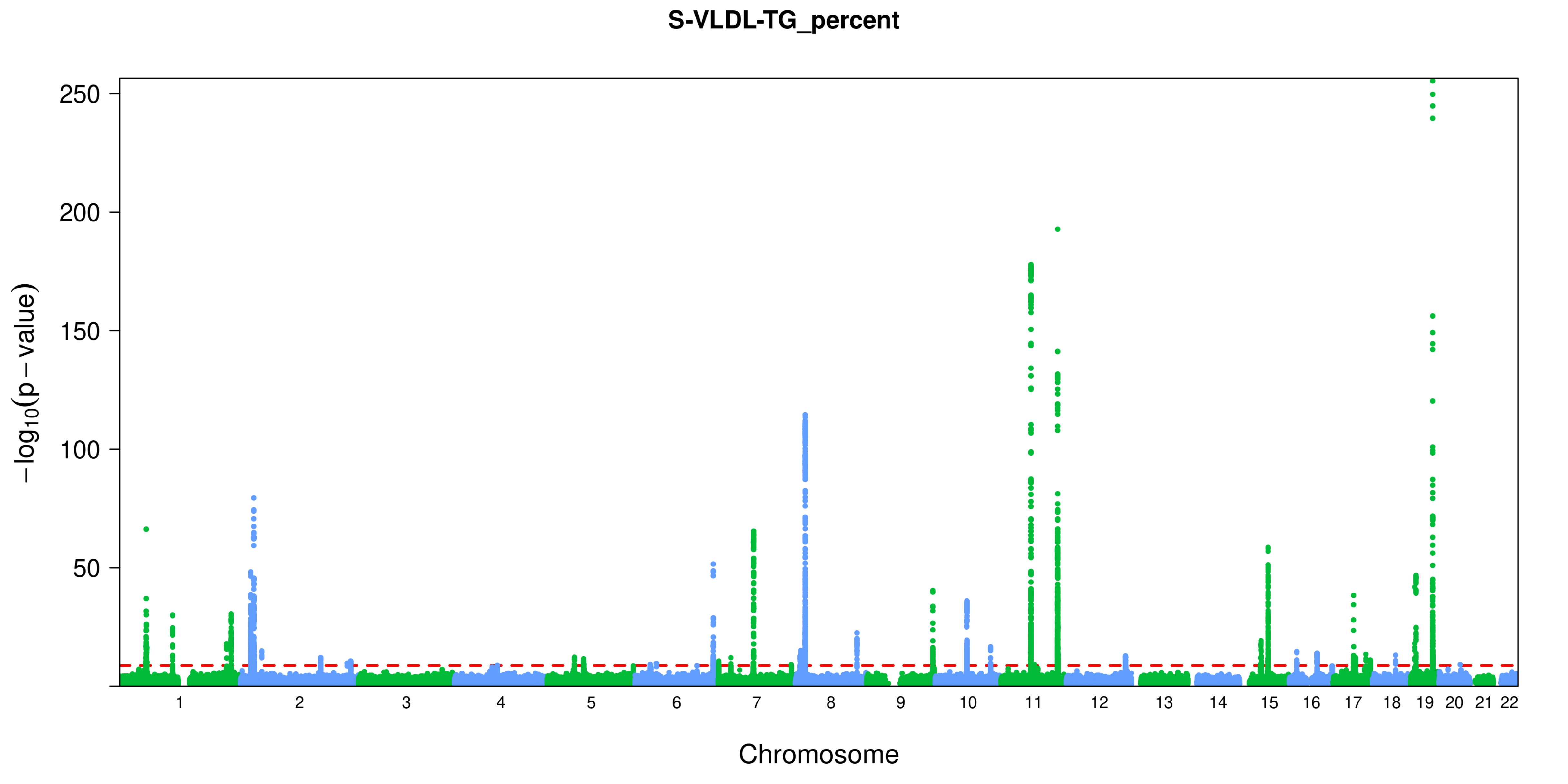

S-VLDL-TG_percent
250
200
150
100
50
−log10(p − value)
Chromosome

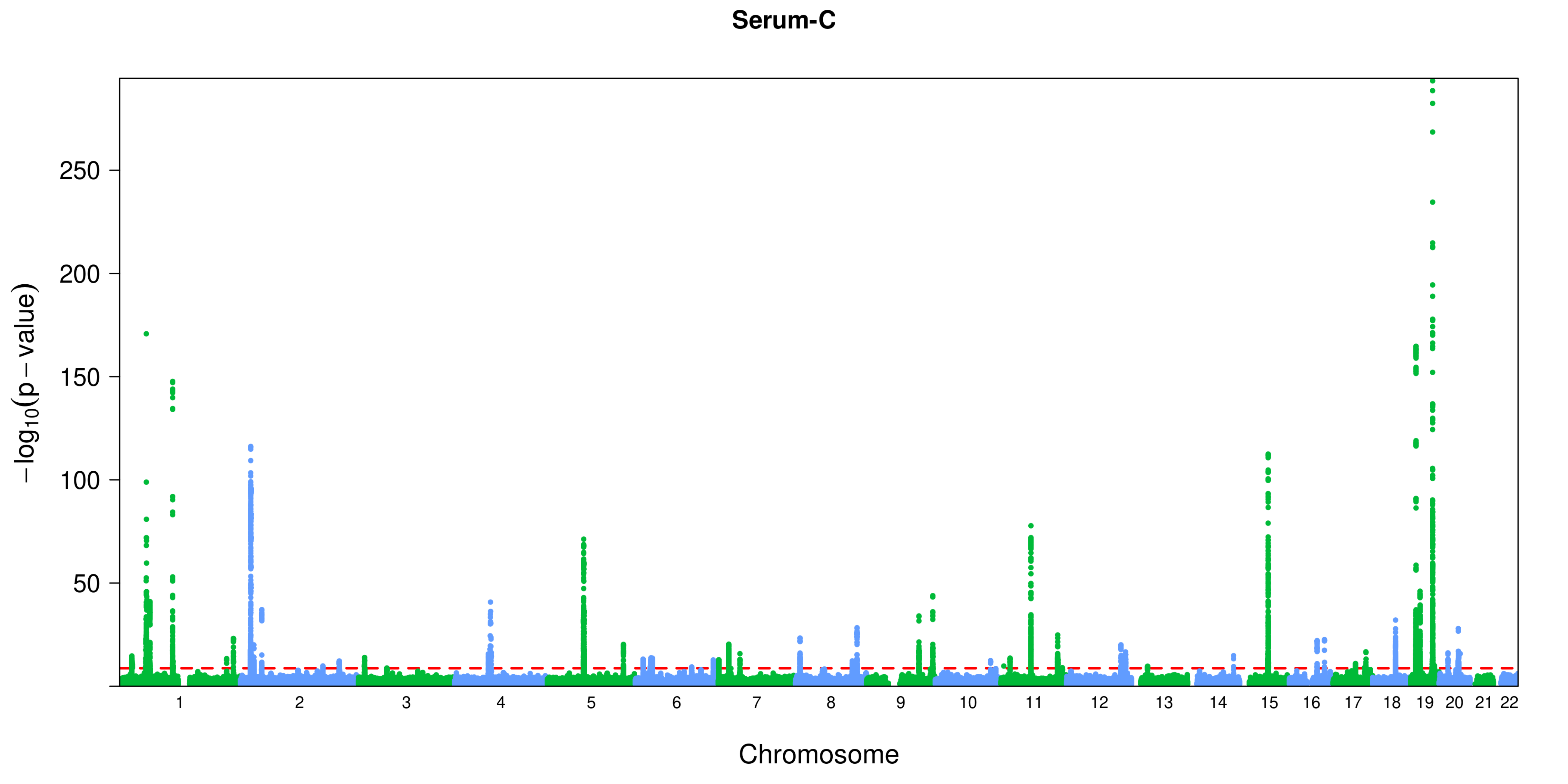

Serum-C
250
200
150
100
50
−log10(p − value)
Chromosome

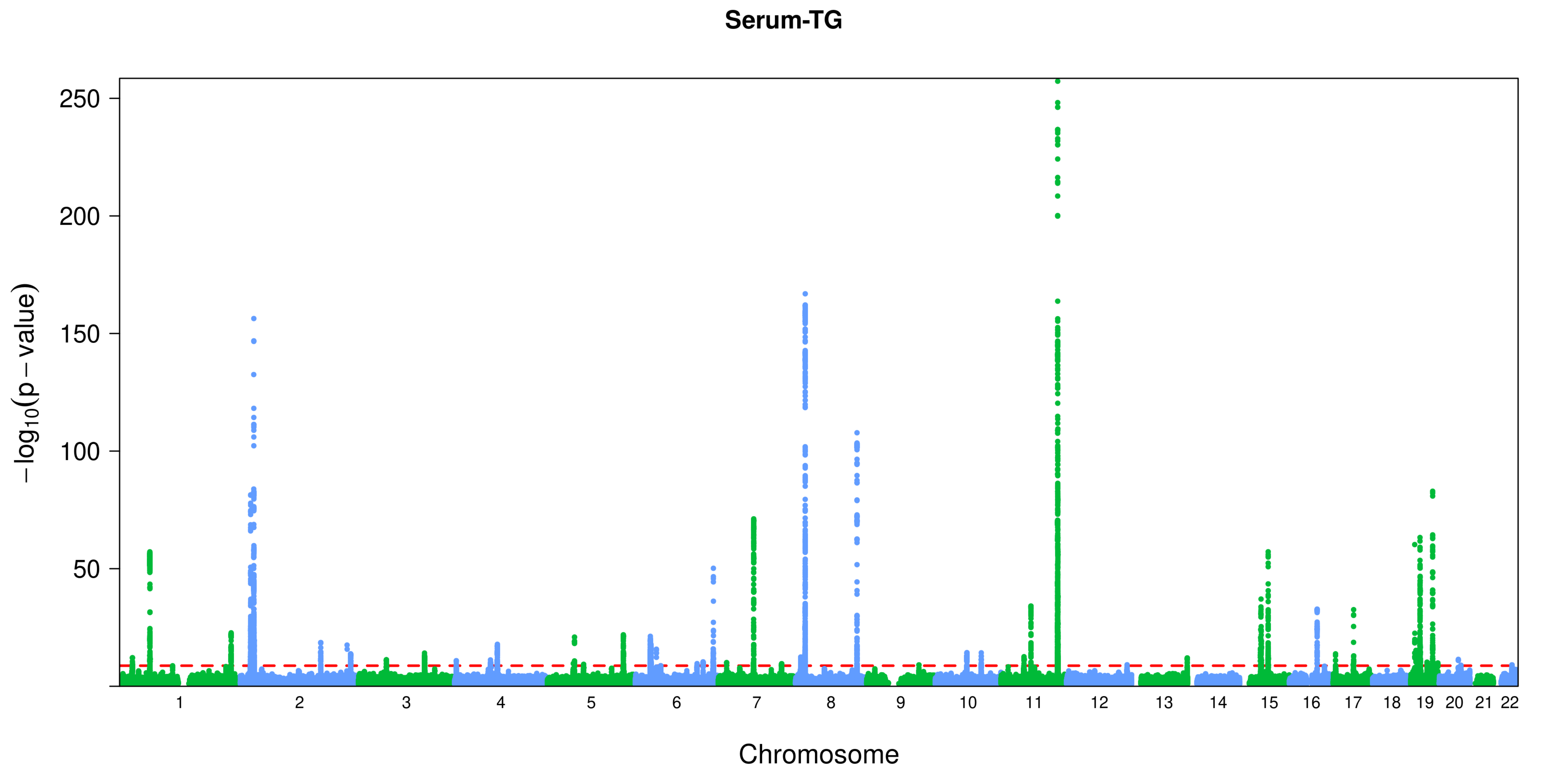

Serum-TG
250
200
150
100
50
−log10(p − value)
Chromosome

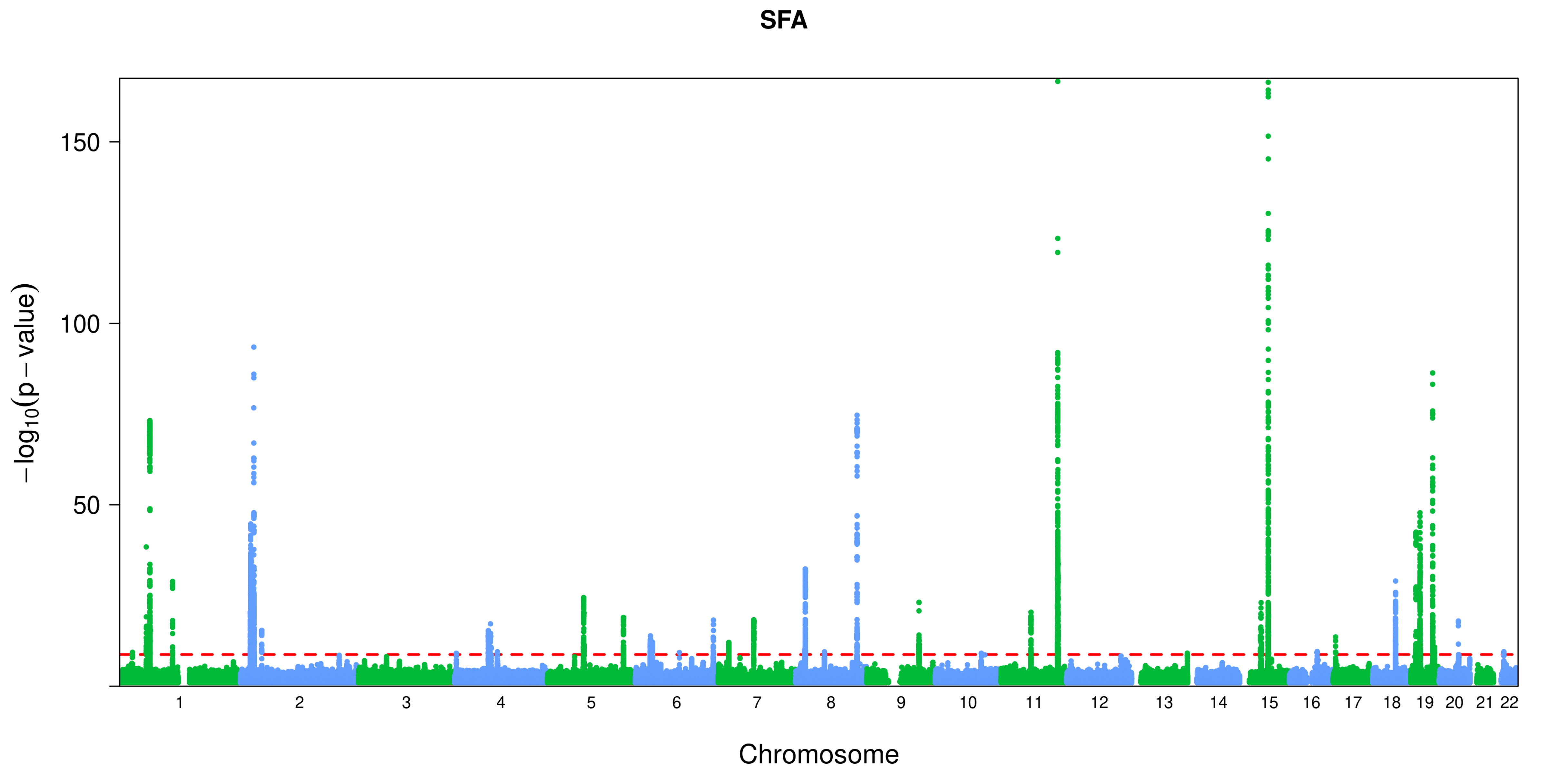
SFA
−log10(p−value)
150
100
50
Chromosome

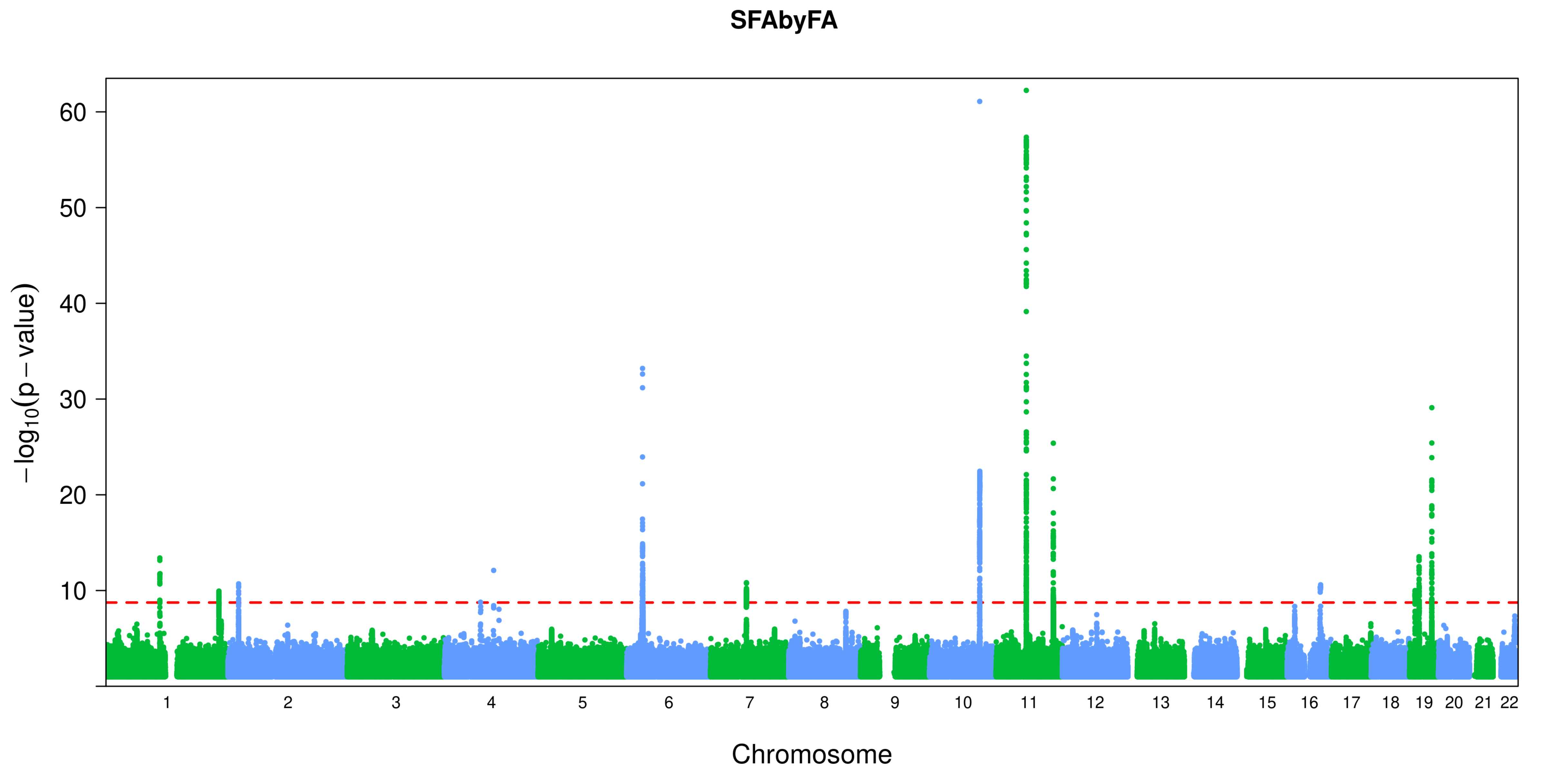

SFAbyFA
−log10(p − value)
60
50
40
30
20
10
Chromosome

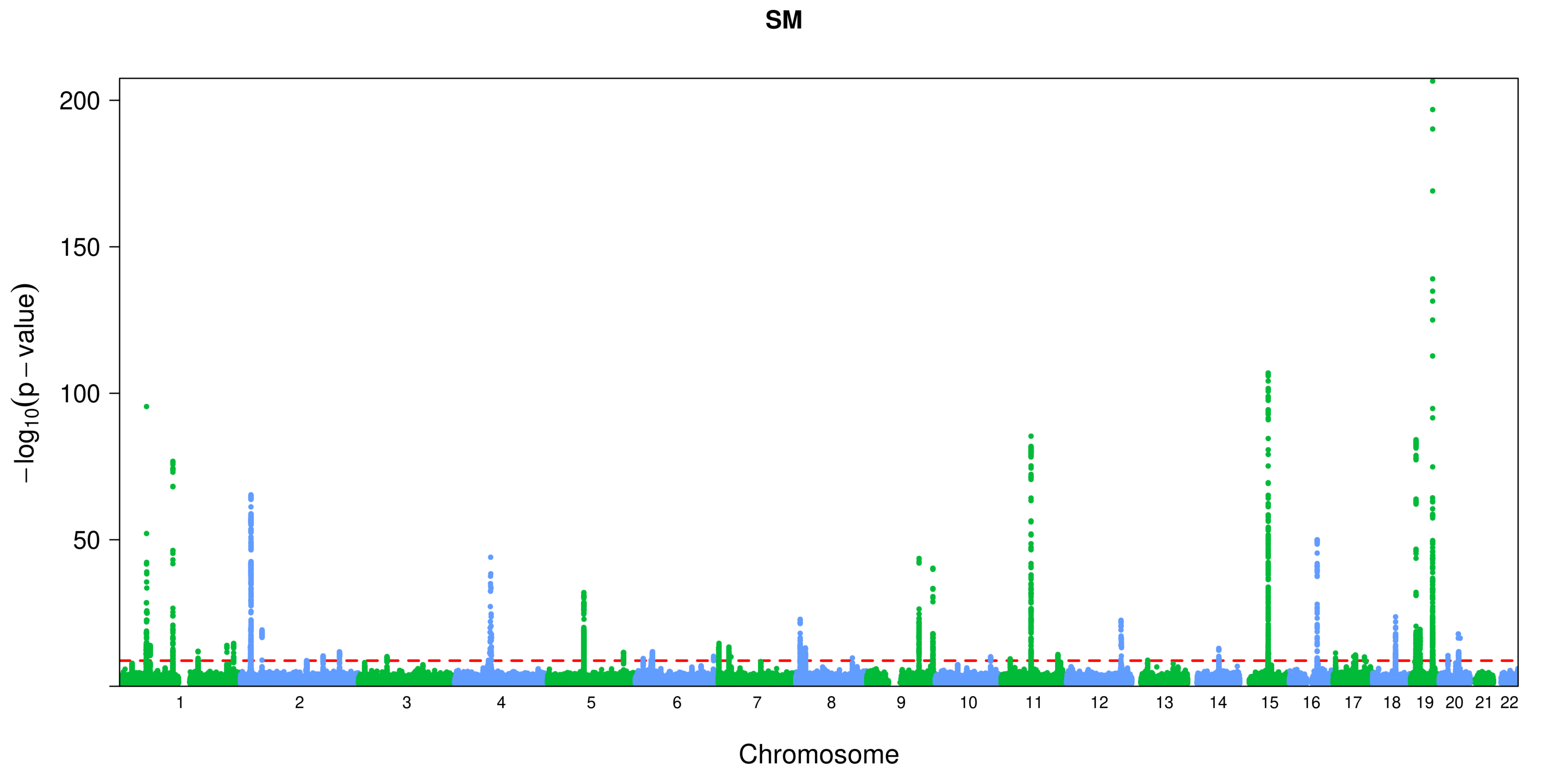
SM
200
150
100
50
−log10(p−value)
Chromosome

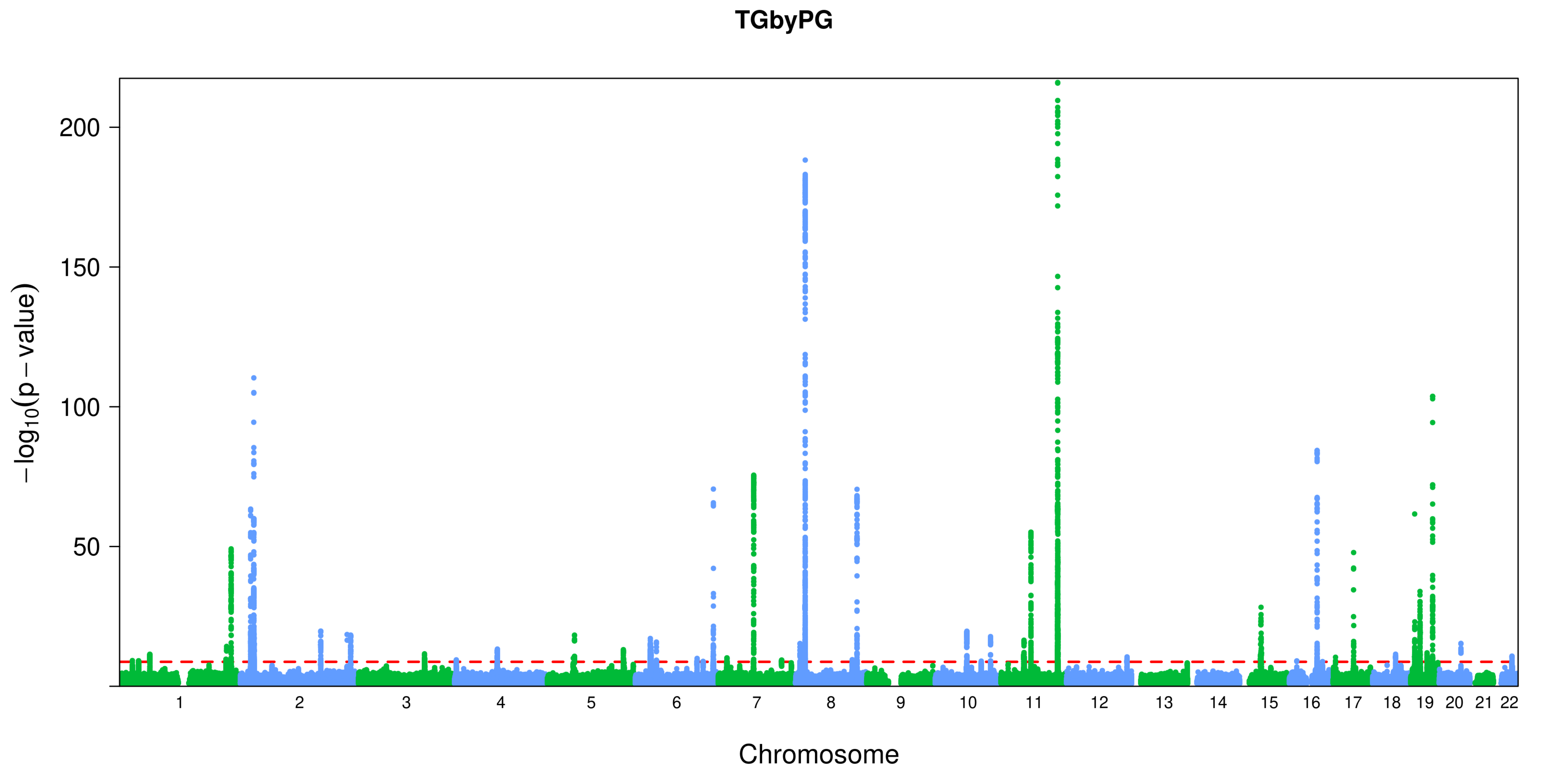
TGbyPG
200
150
100
50
$-\log_{10}(p-\text{value})$
Chromosome

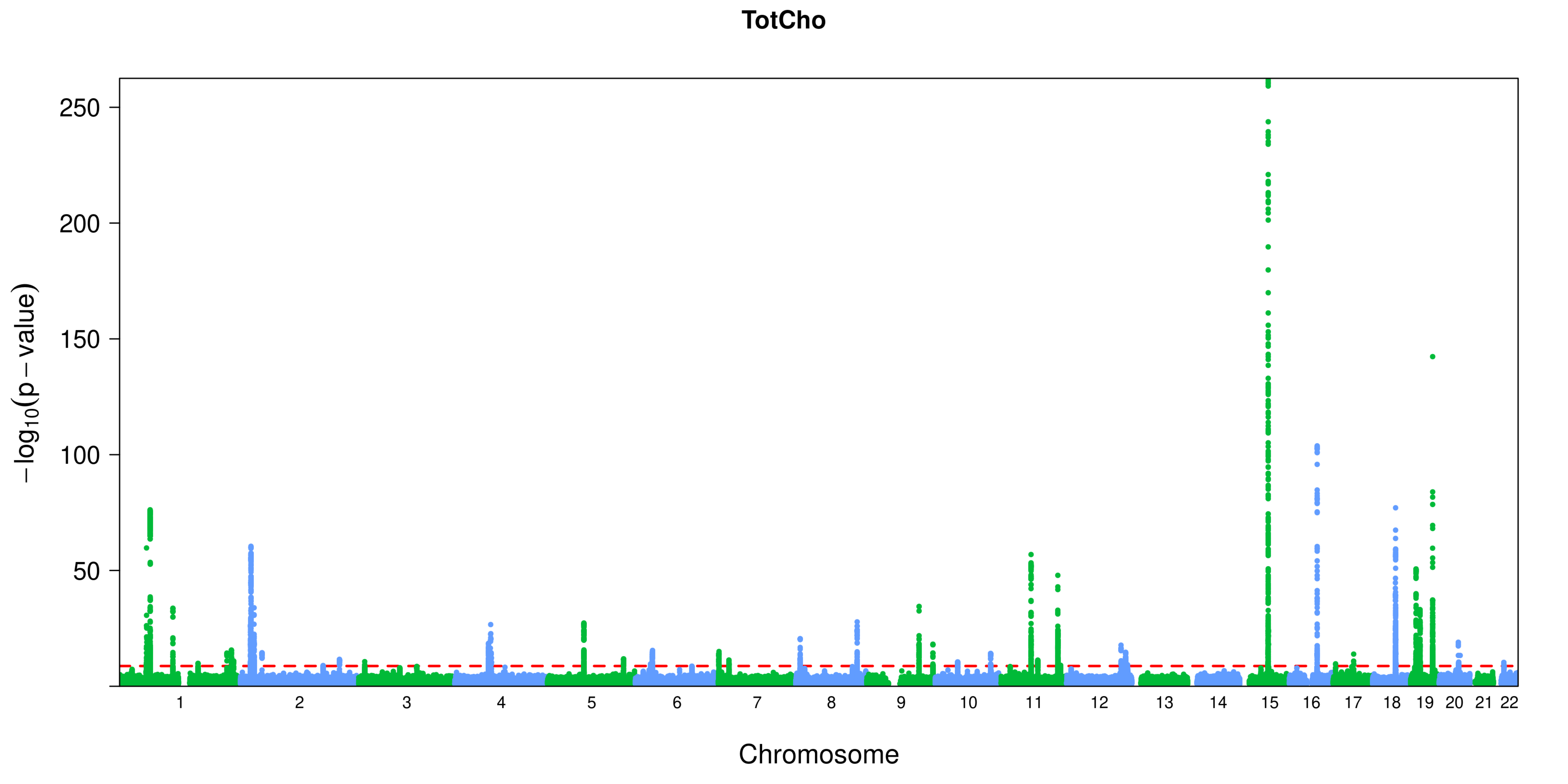
TotCho
250
200
150
100
50
−log10(p − value)
Chromosome

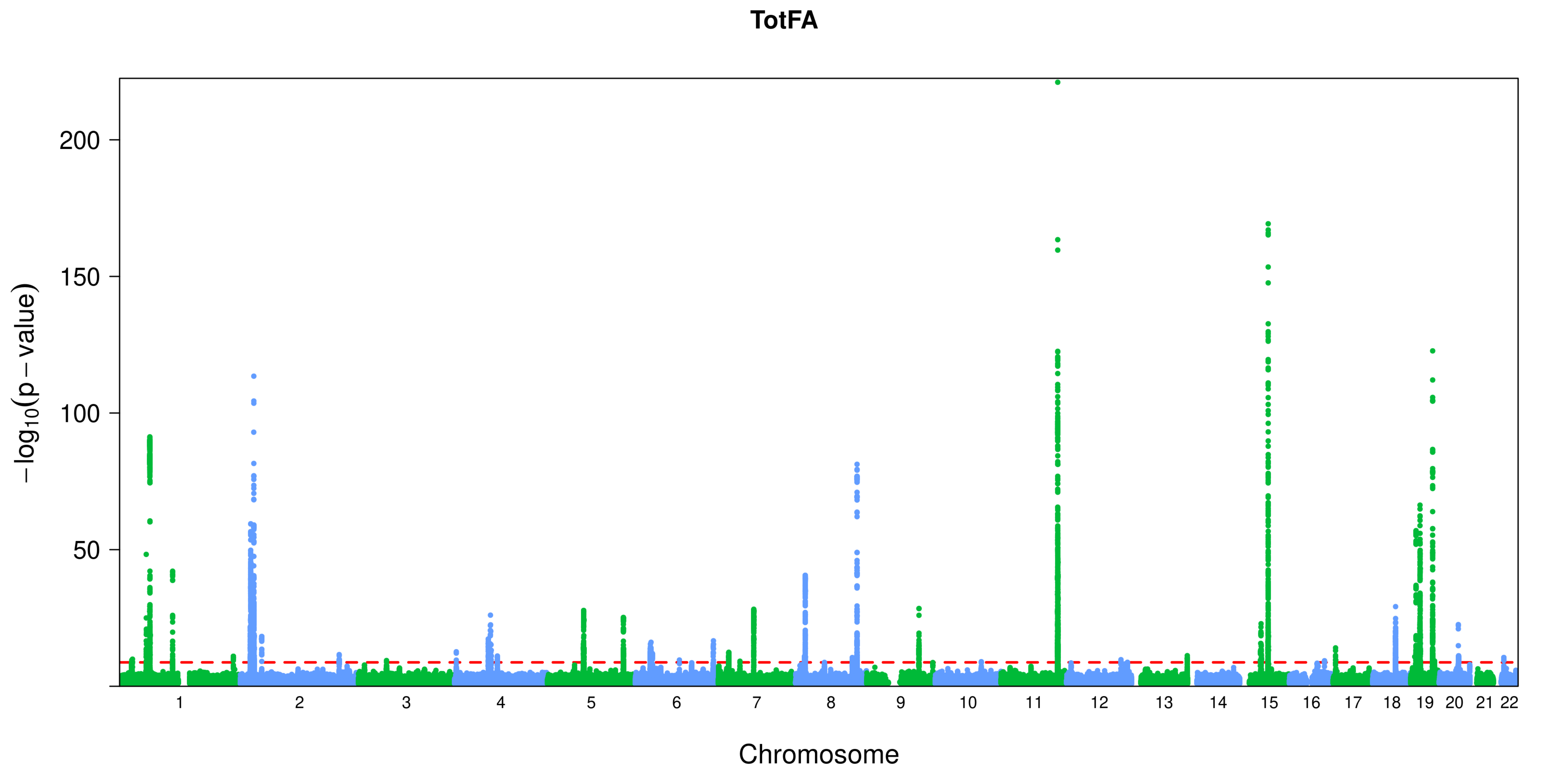

TotFA
200
150
100
50
$-\log_{10}(p-\text{value})$
Chromosome

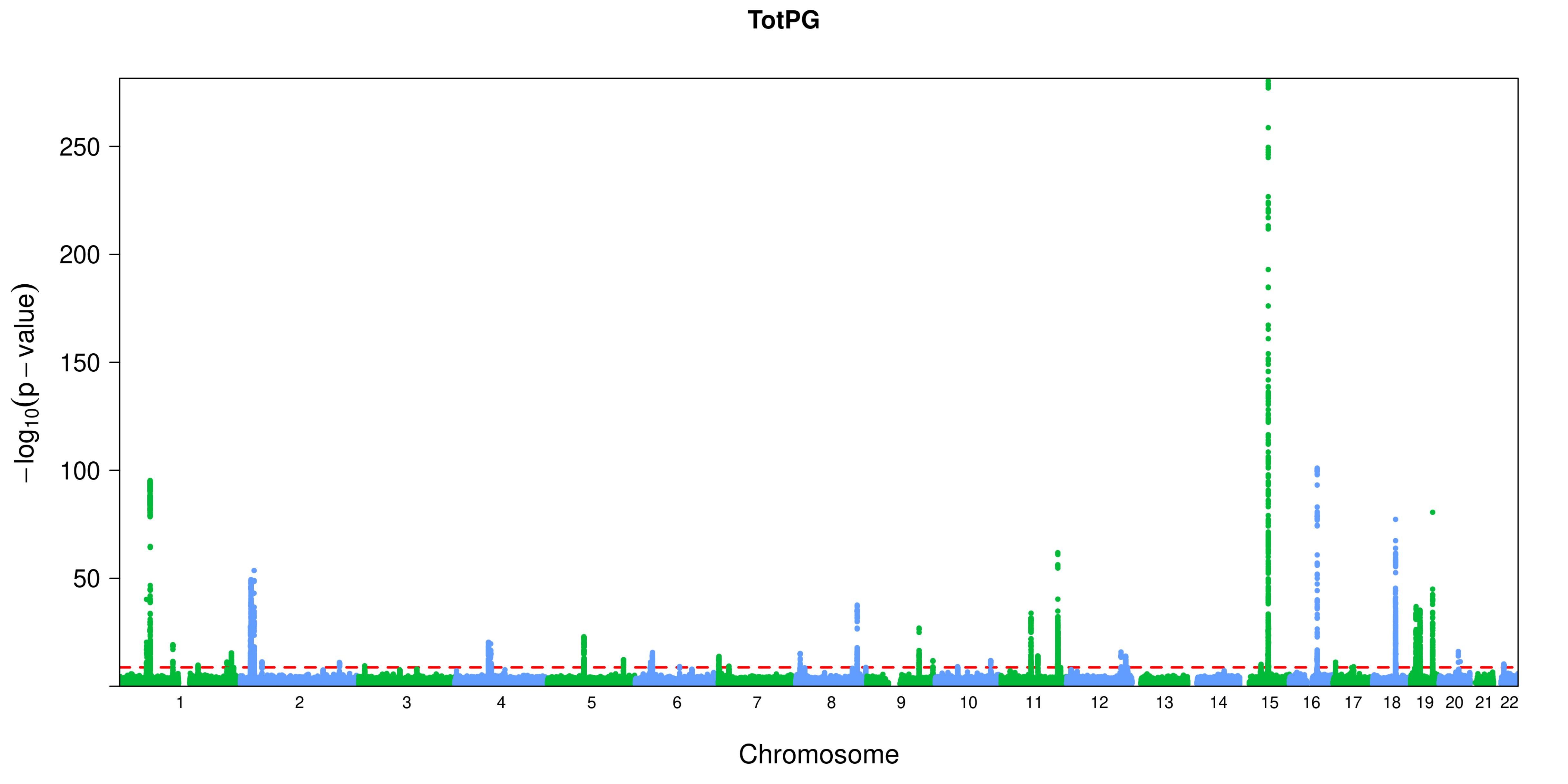

TotPG
250
200
150
100
50
−log10(p − value)
Chromosome

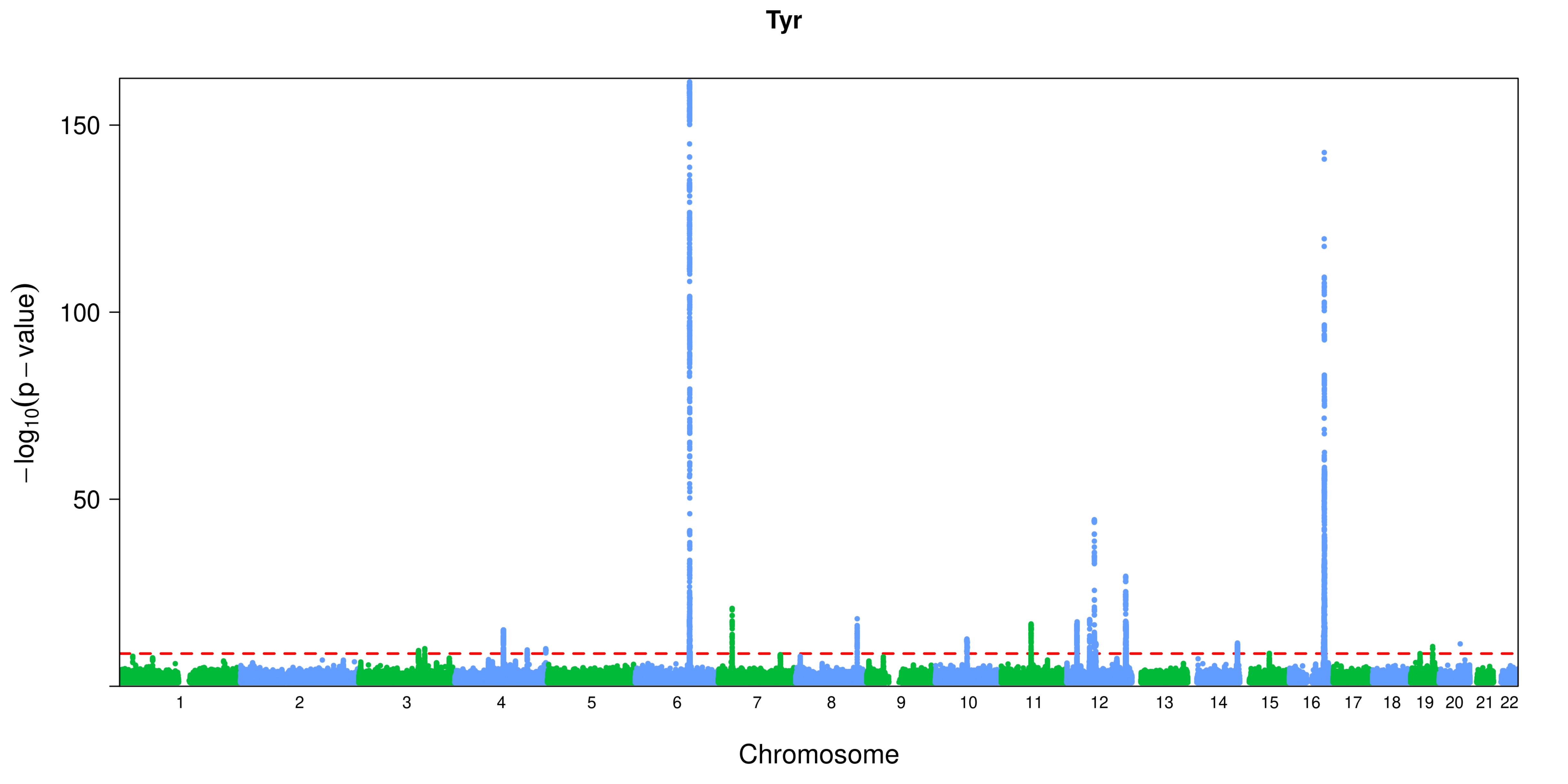
Tyr
150
100
50
−log10(p − value)
Chromosome

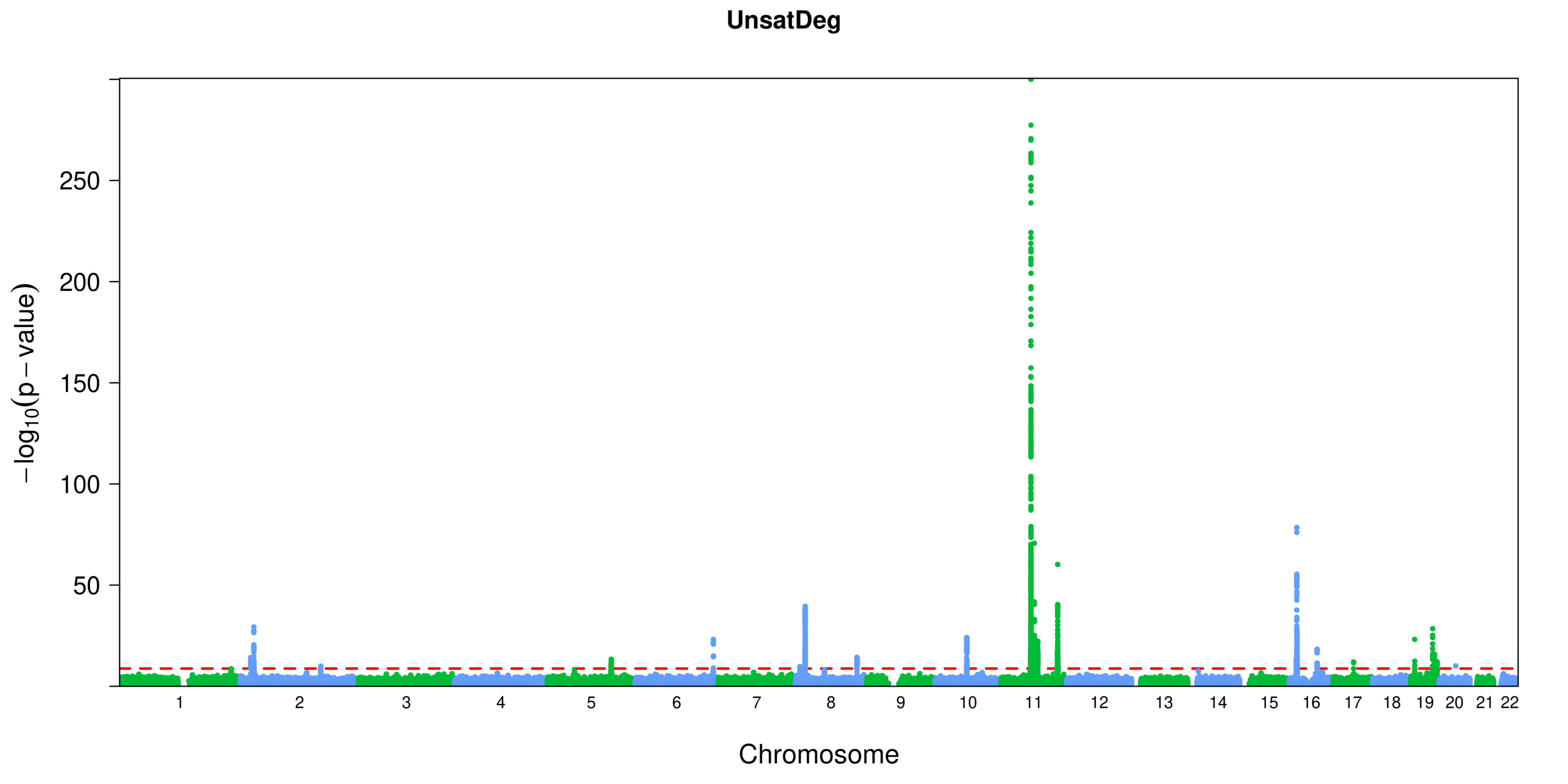

UnsatDeg
−log10(p − value)
250
200
150
100
50
Chromosome

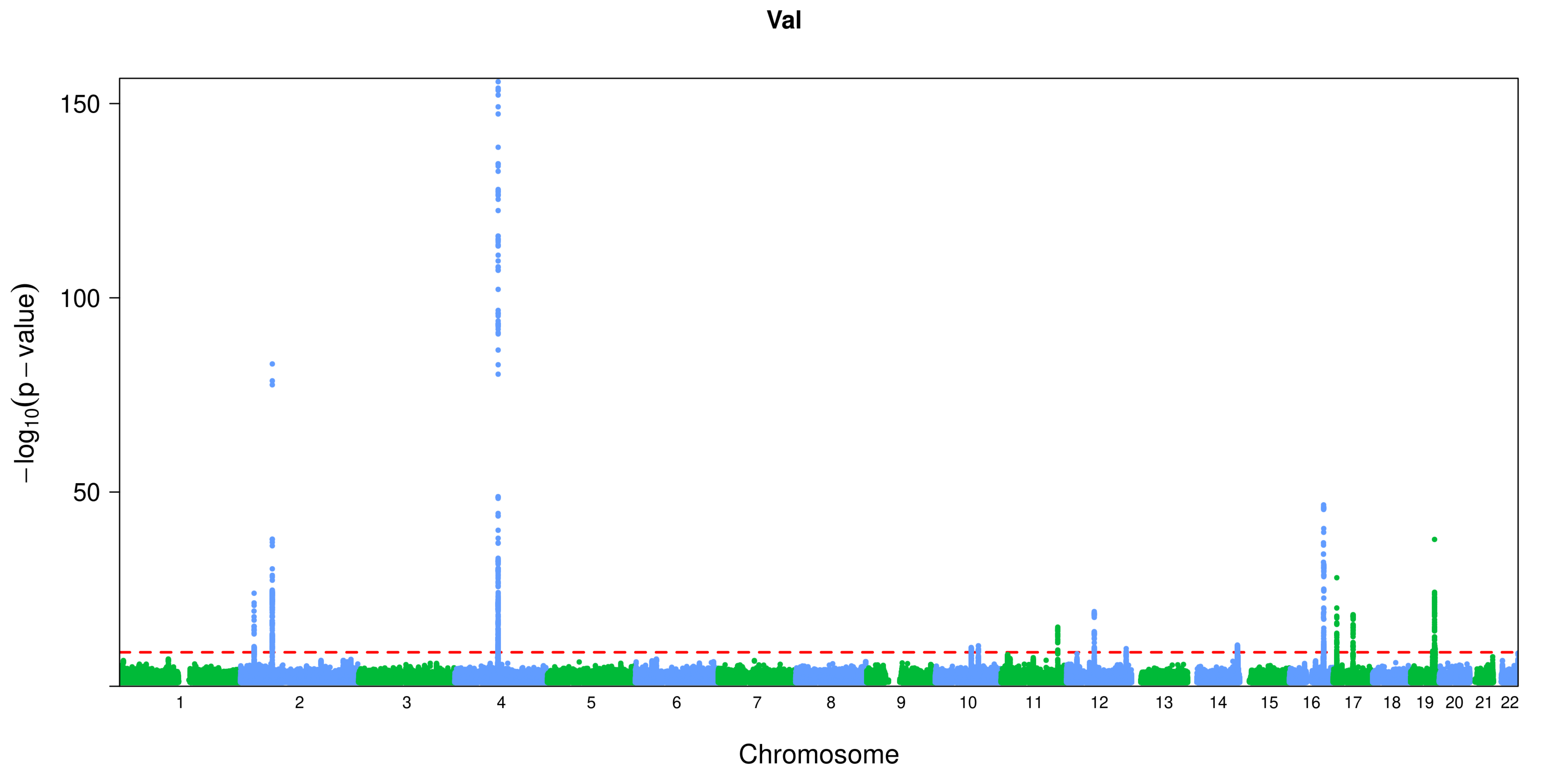

Val
150
100
50
−log10(p − value)
Chromosome

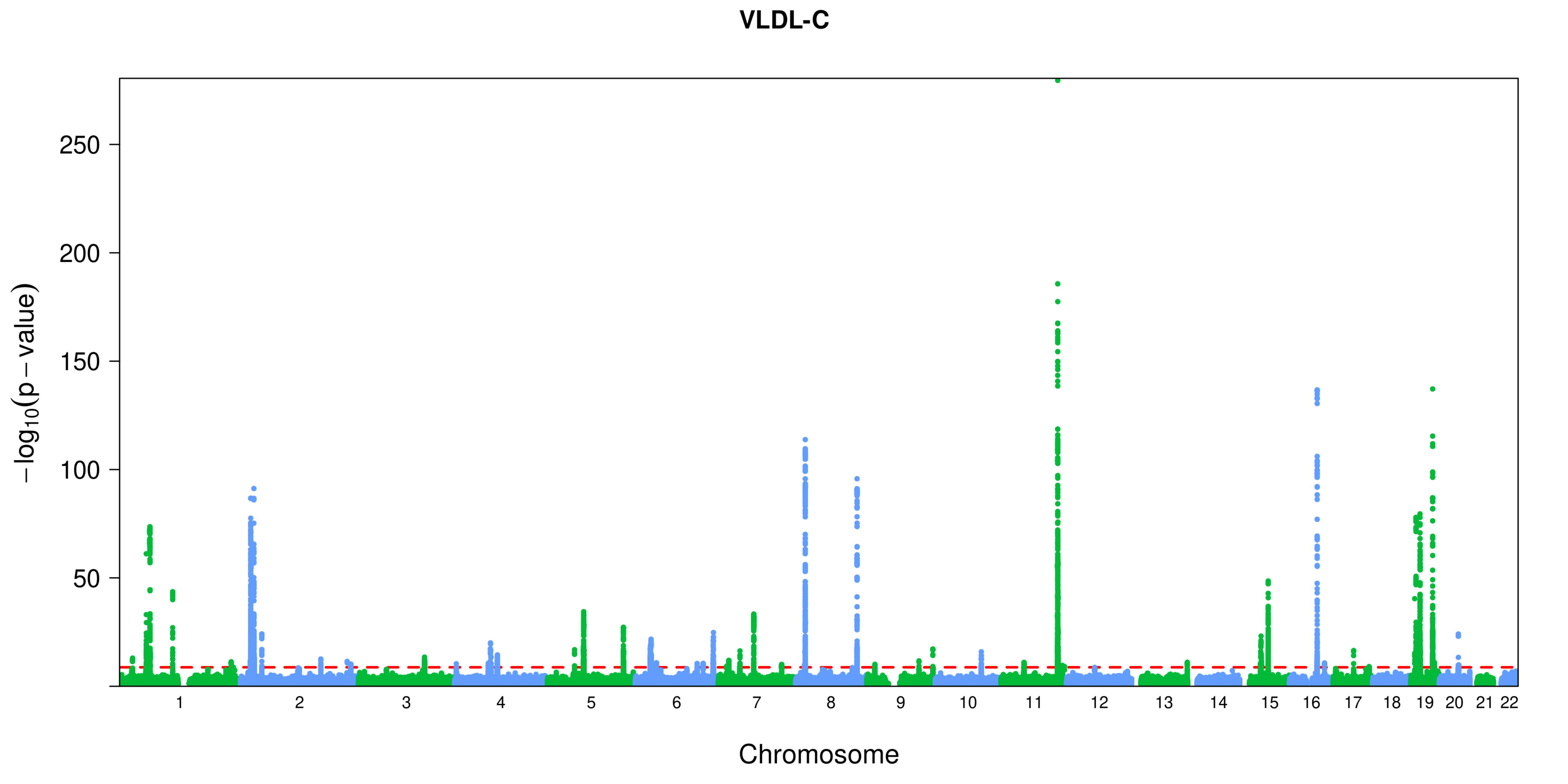
VLDL-C
−log10(p − value)
250
200
150
100
50
Chromosome

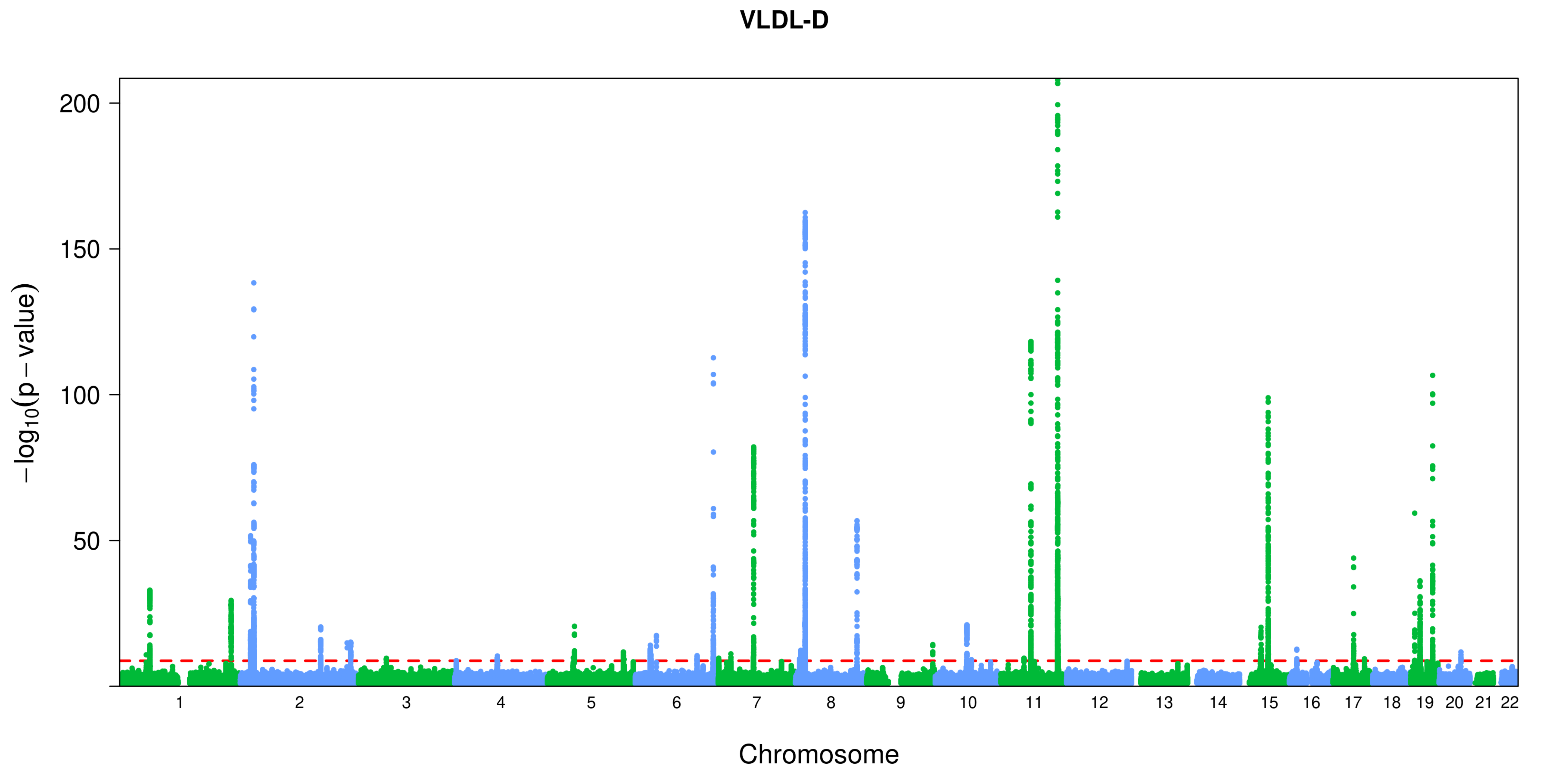
VLDL-D
200
150
100
50
−log10(p − value)
Chromosome

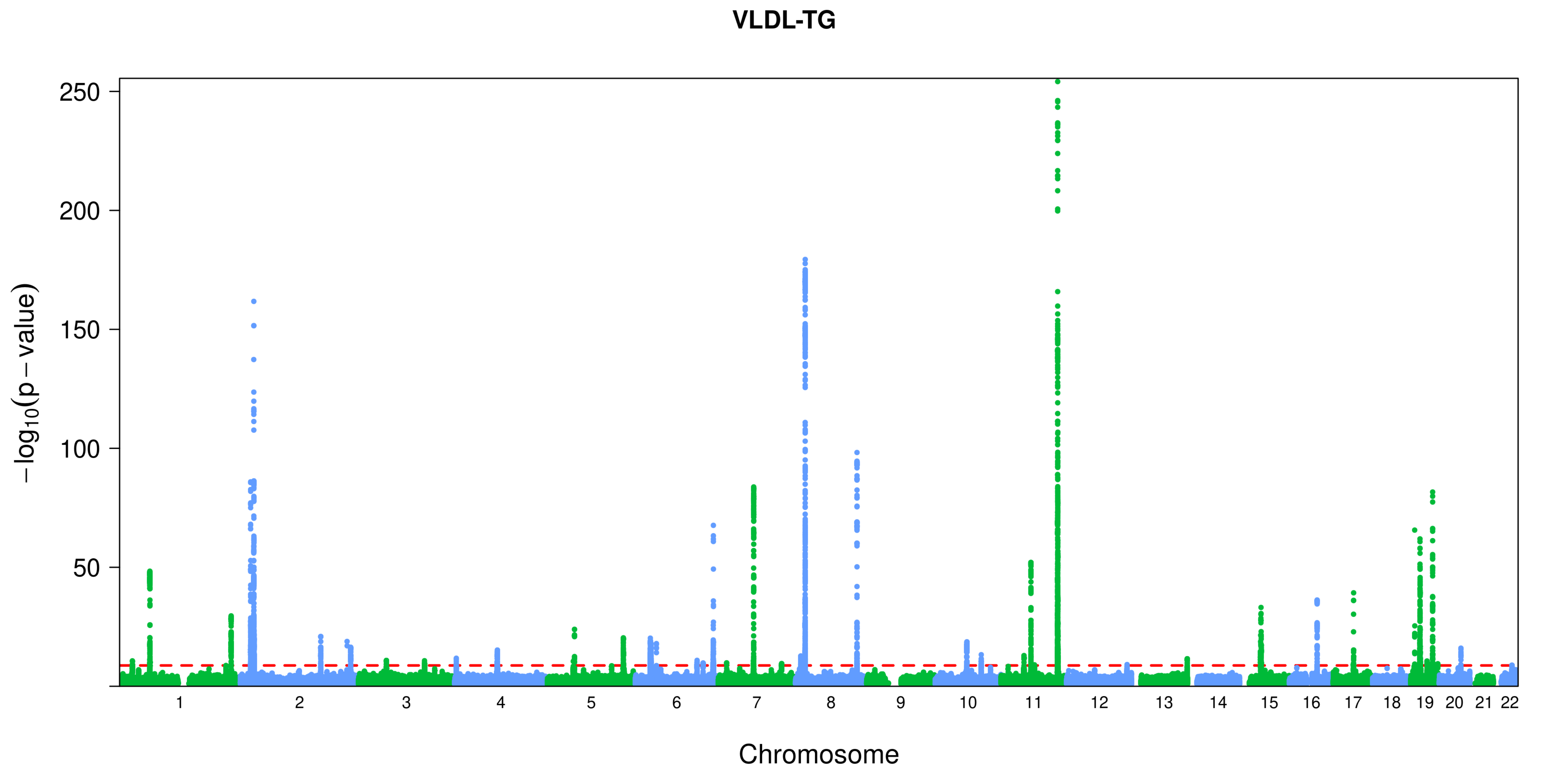
VLDL-TG
250
200
150
100
50
−log10(p − value)
Chromosome

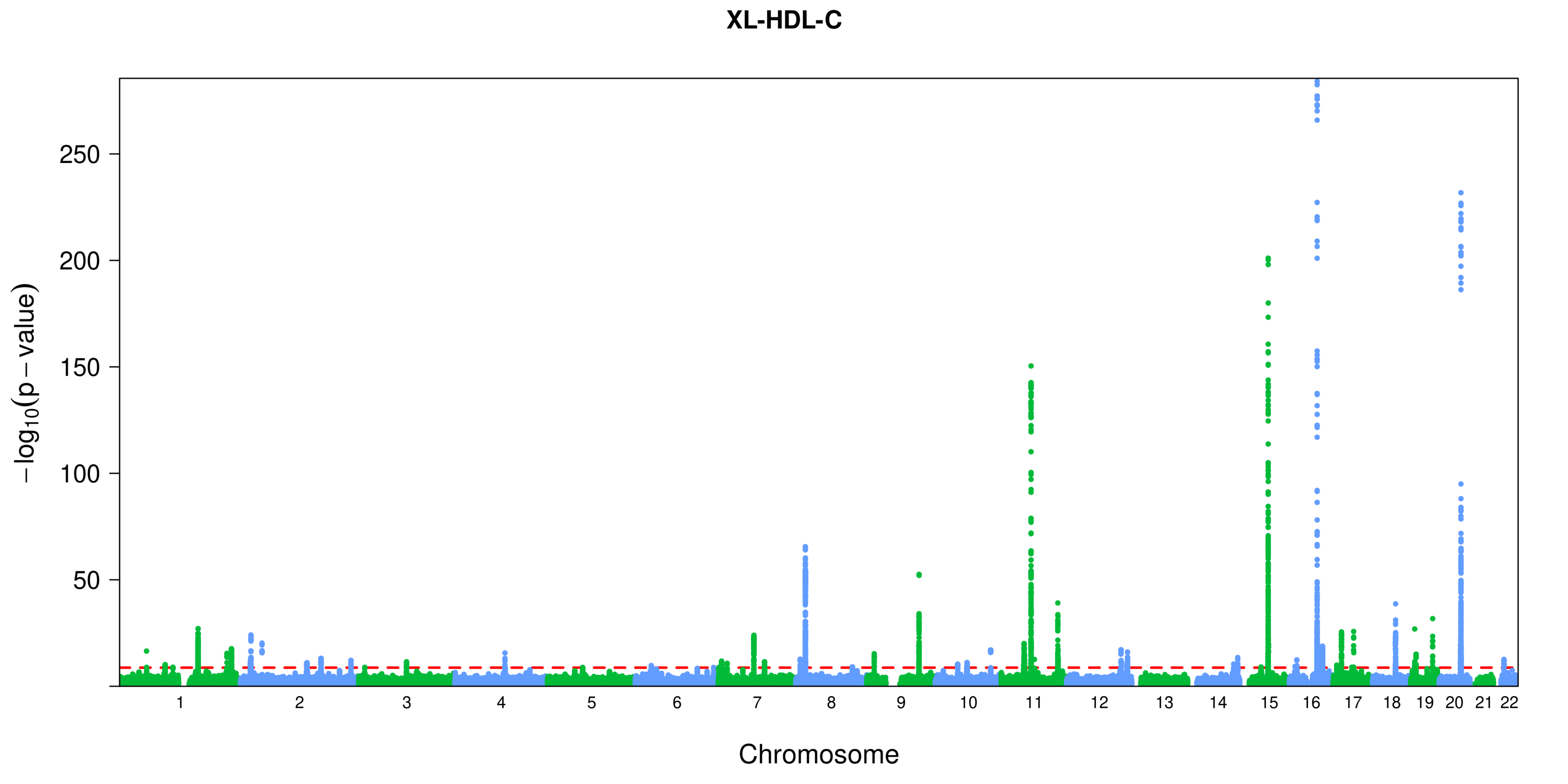
XL-HDL-C
250
200
150
100
50
−log10(p − value)
Chromosome

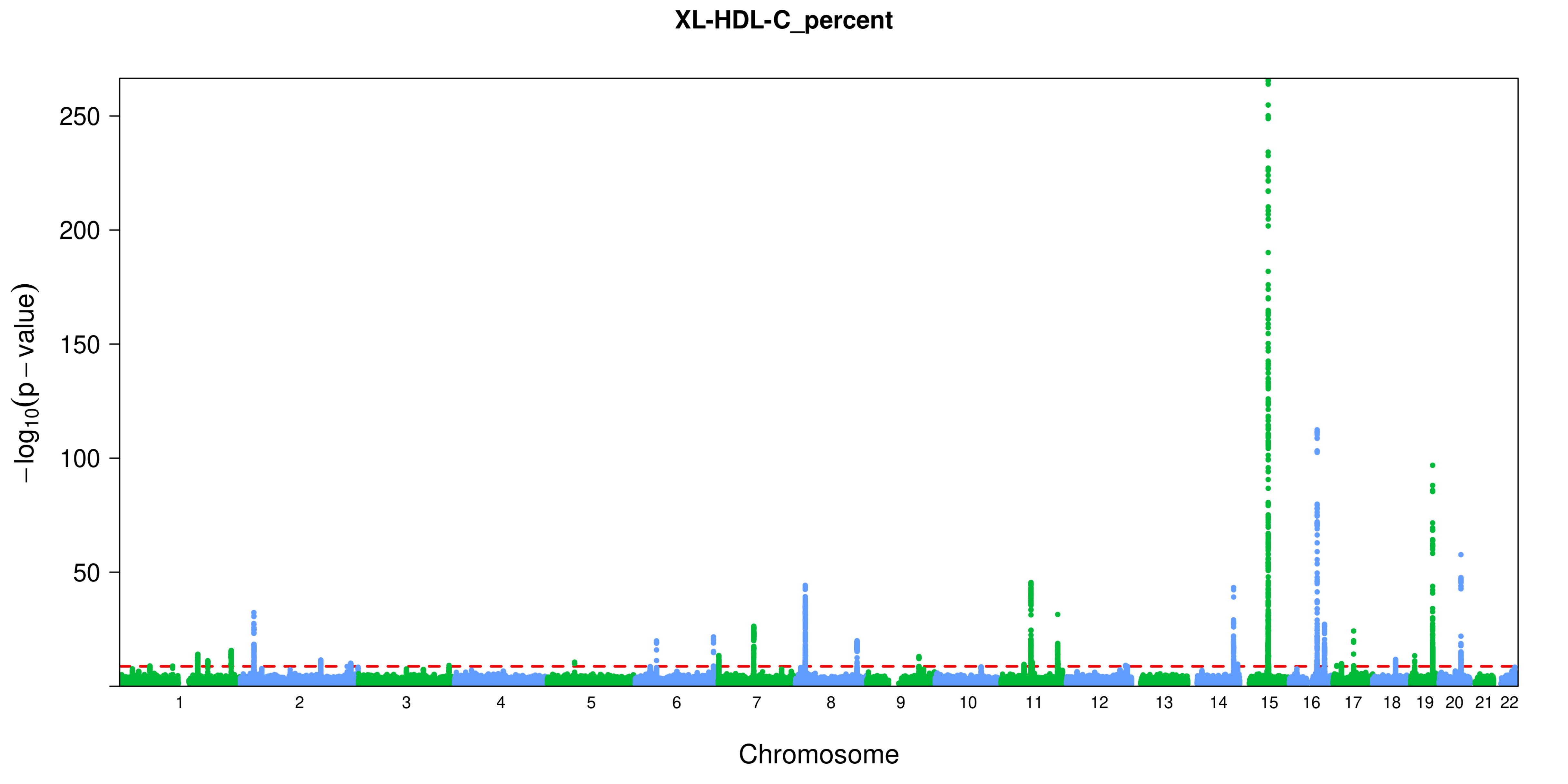

XL-HDL-C_percent
−log10(p − value)
250
200
150
100
50
1 2 3 4 5 6 7 8 9 10 11 12 13 14 15 16 17 18 19 20 21 22
Chromosome

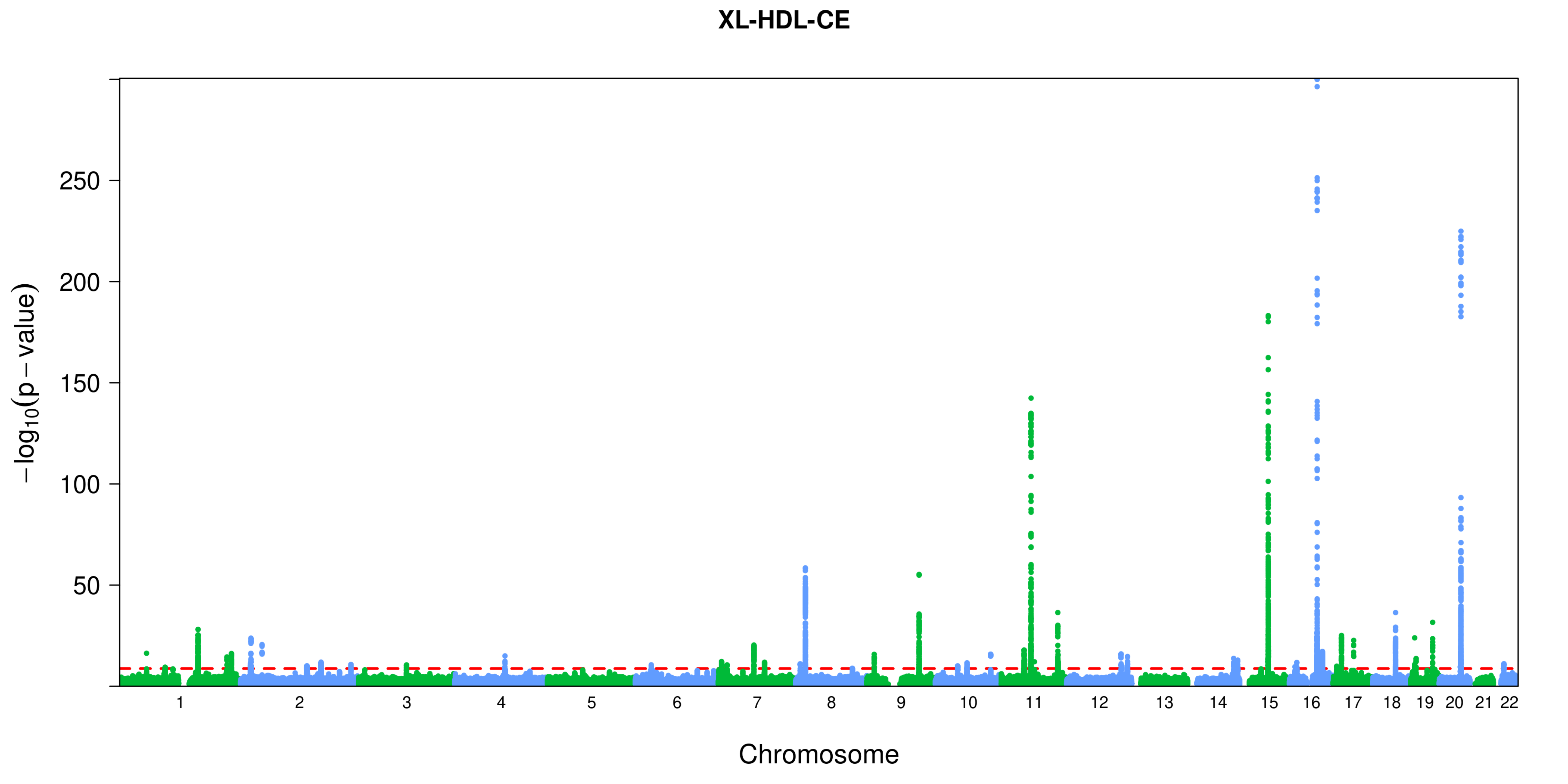

XL-HDL-CE
−log10(p − value)
250
200
150
100
50
Chromosome

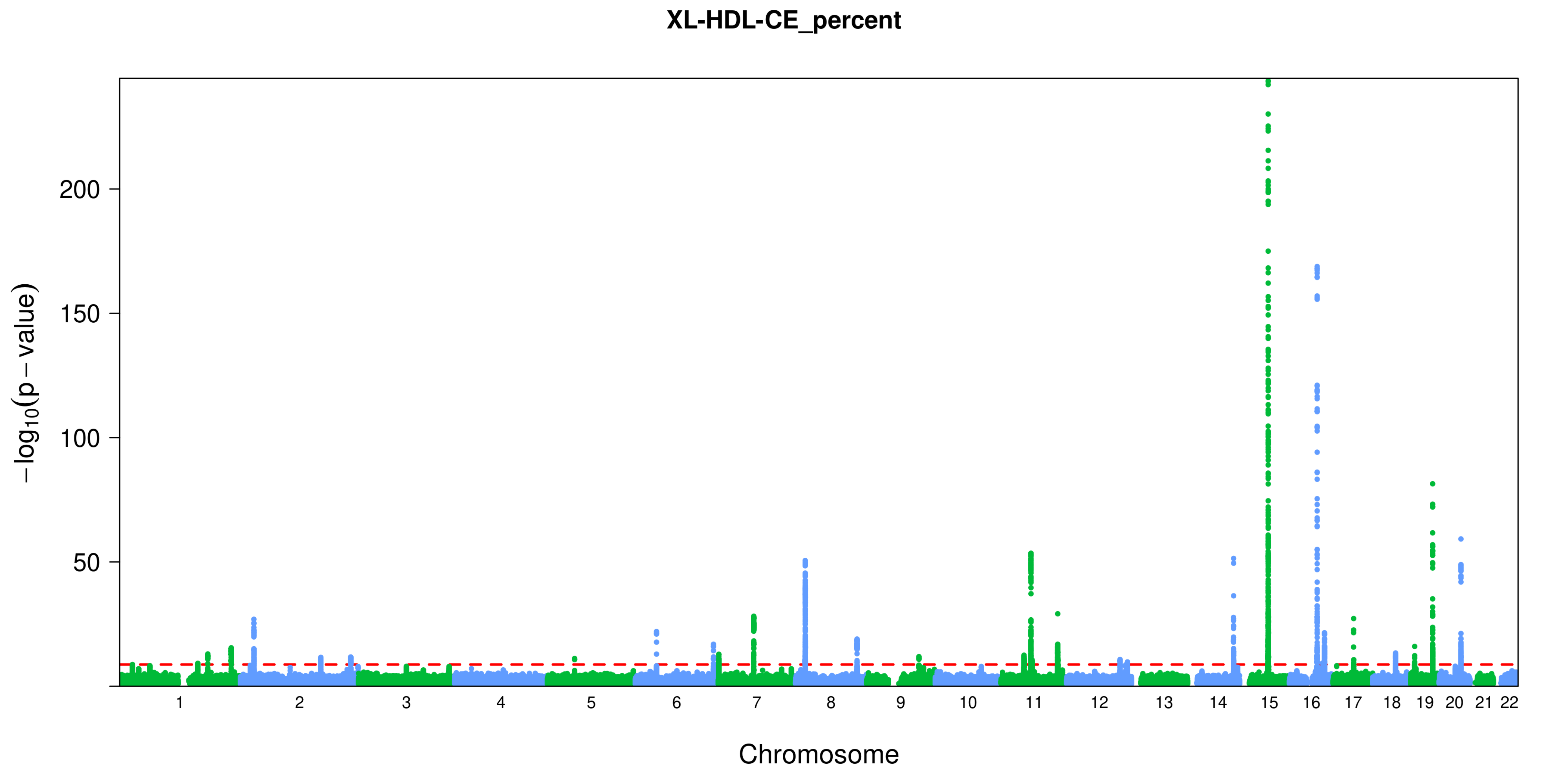

XL-HDL-CE_percent
−log10(p − value)
200
150
100
50
Chromosome

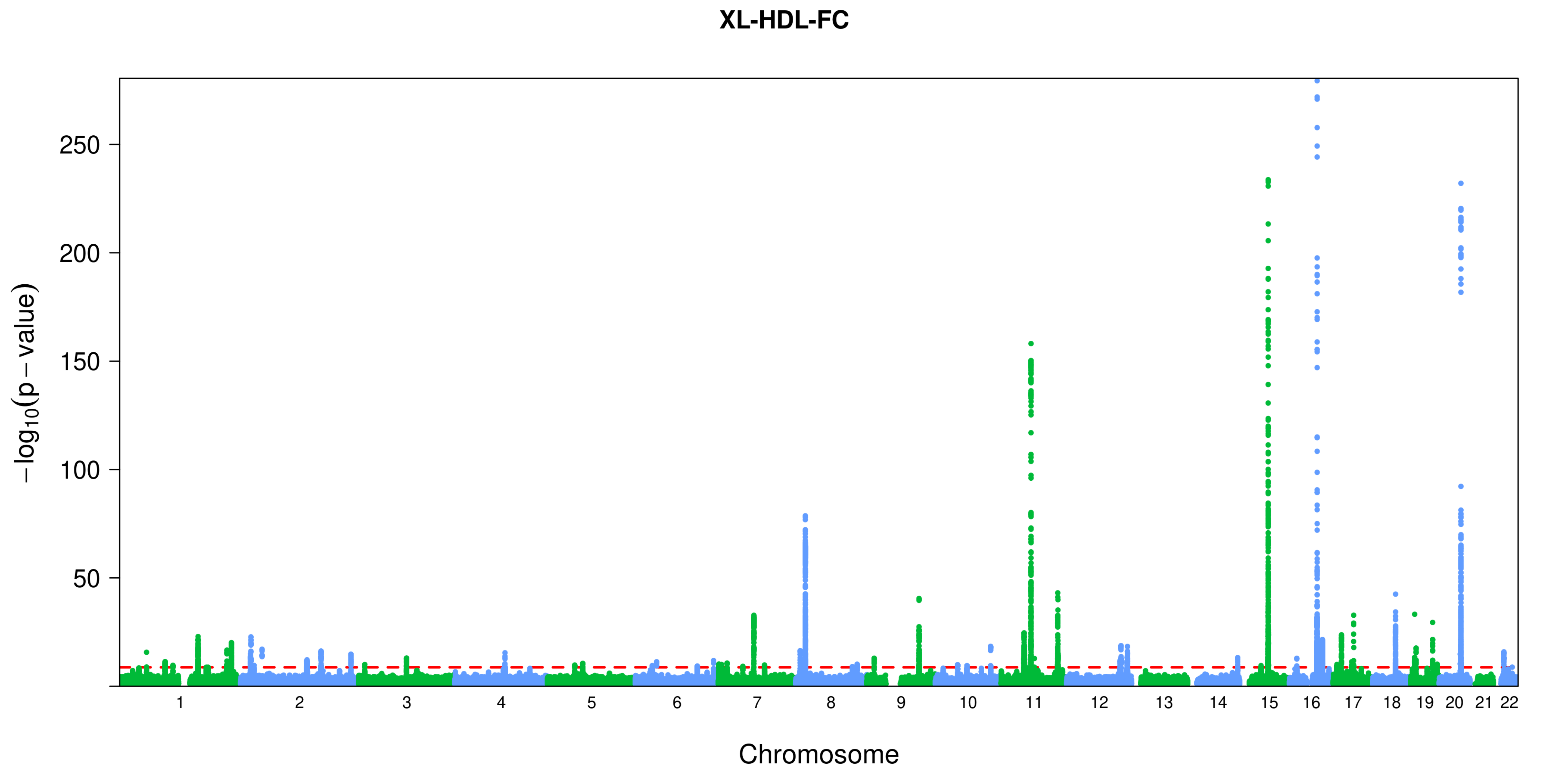
XL-HDL-FC
250
200
150
100
50
−log10(p − value)
Chromosome

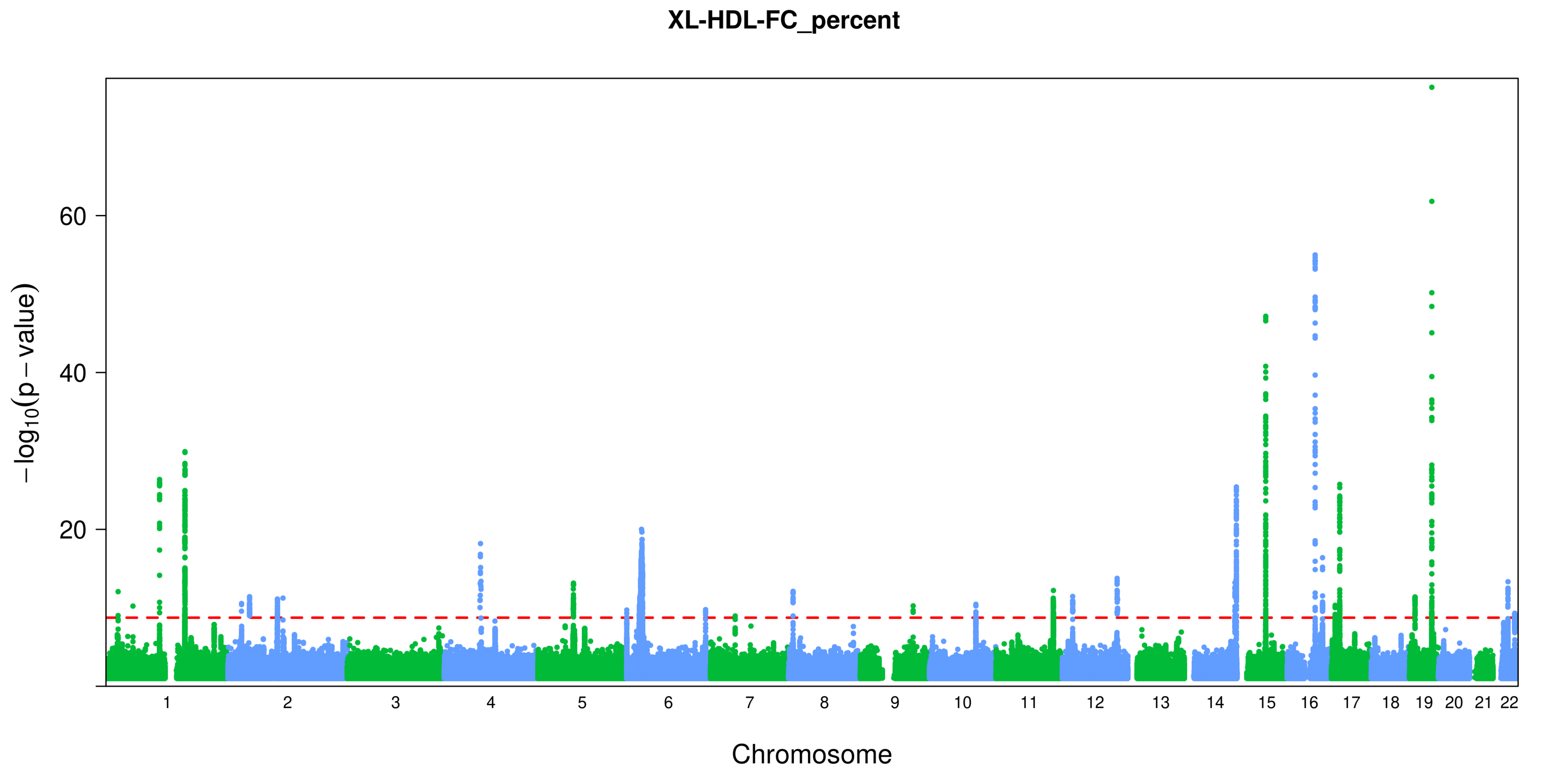
XL-HDL-FC_percent
−log10(p − value)
60
40
20
Chromosome

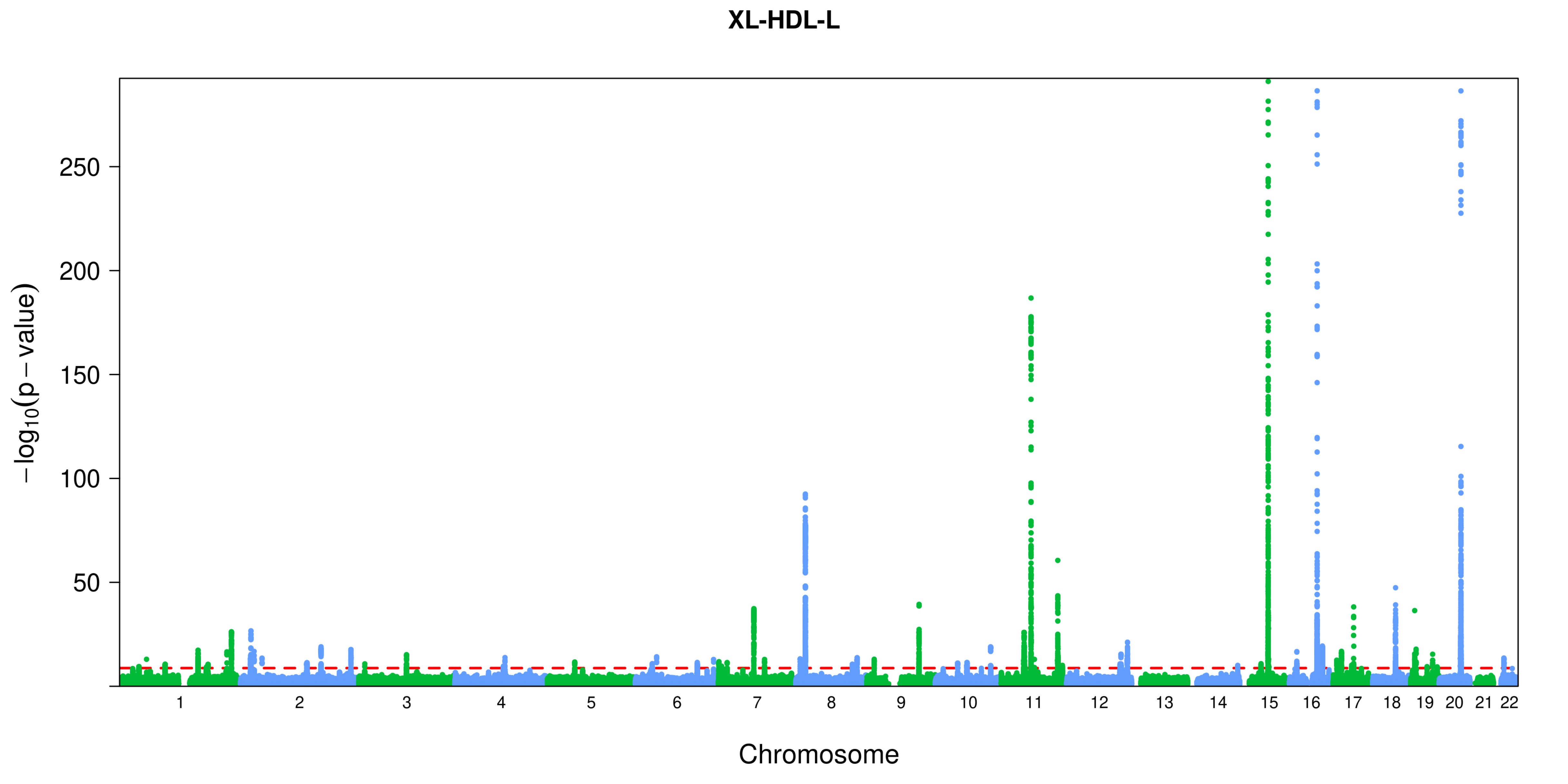

XL-HDL-L
250
200
150
100
50
$-\log_{10}(p-\text{value})$
Chromosome

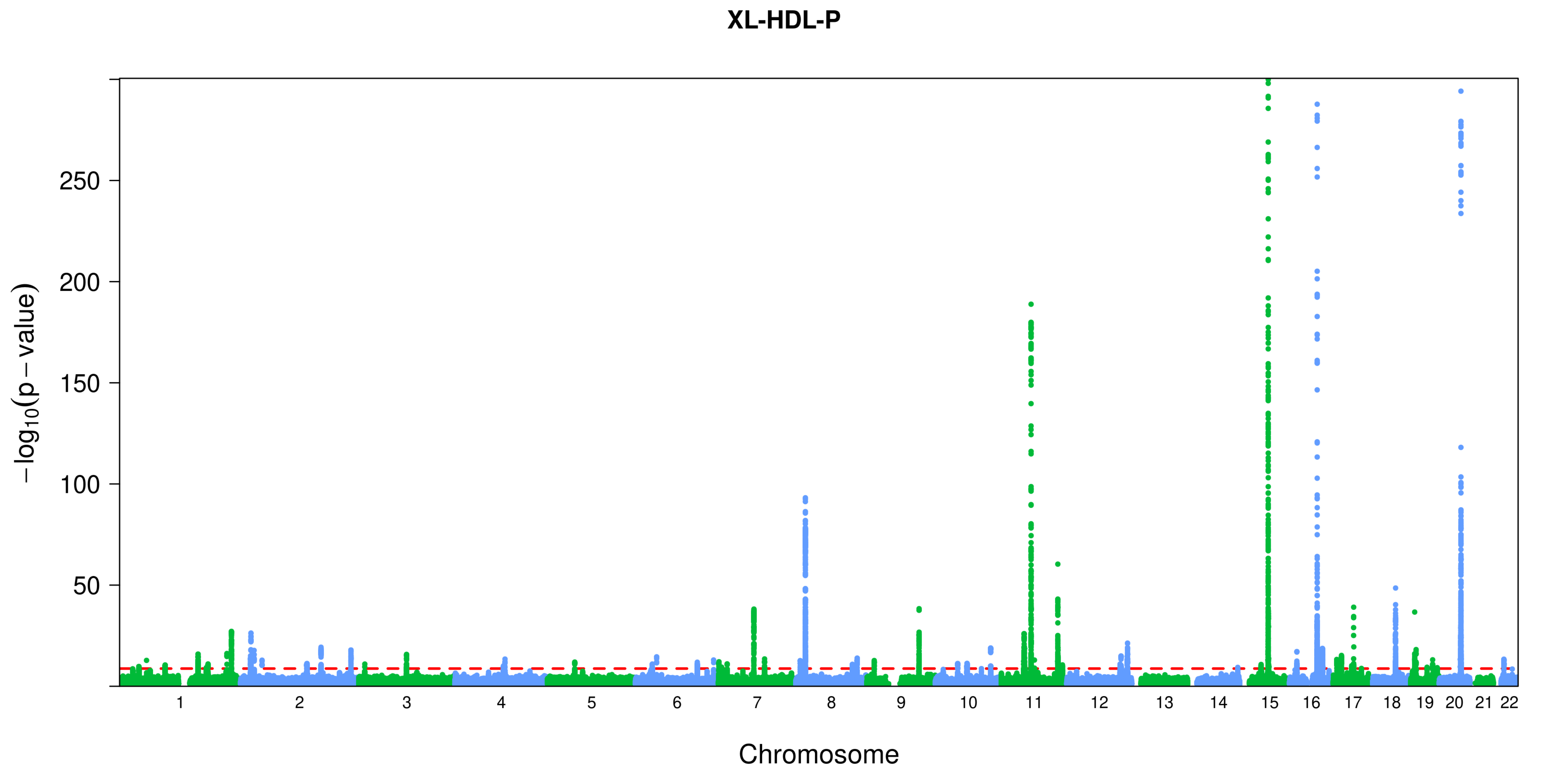
XL-HDL-P
250
200
150
100
50
−log10(p − value)
Chromosome

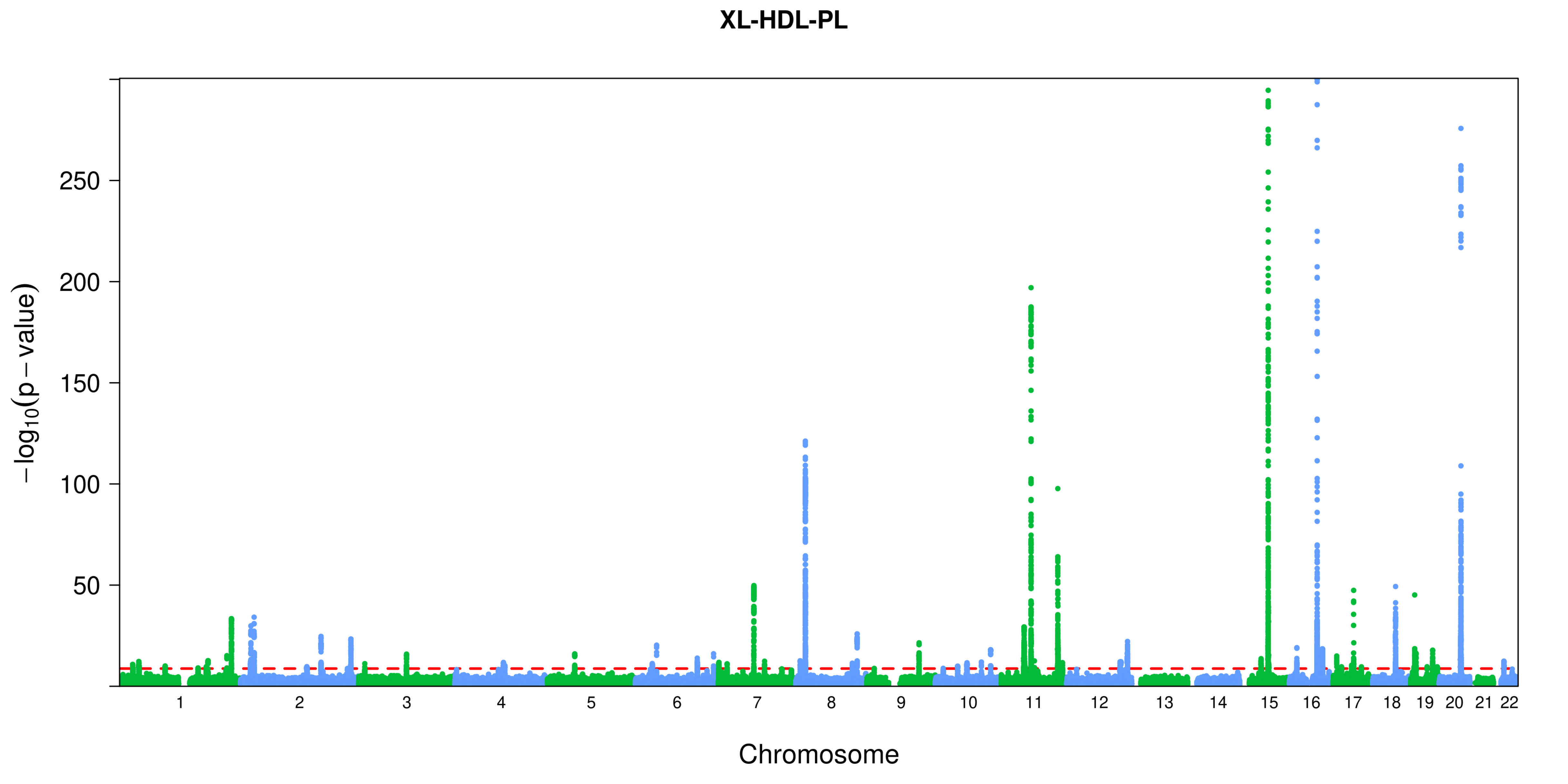

XL-HDL-PL
−log10(p − value)
250
200
150
100
50
Chromosome

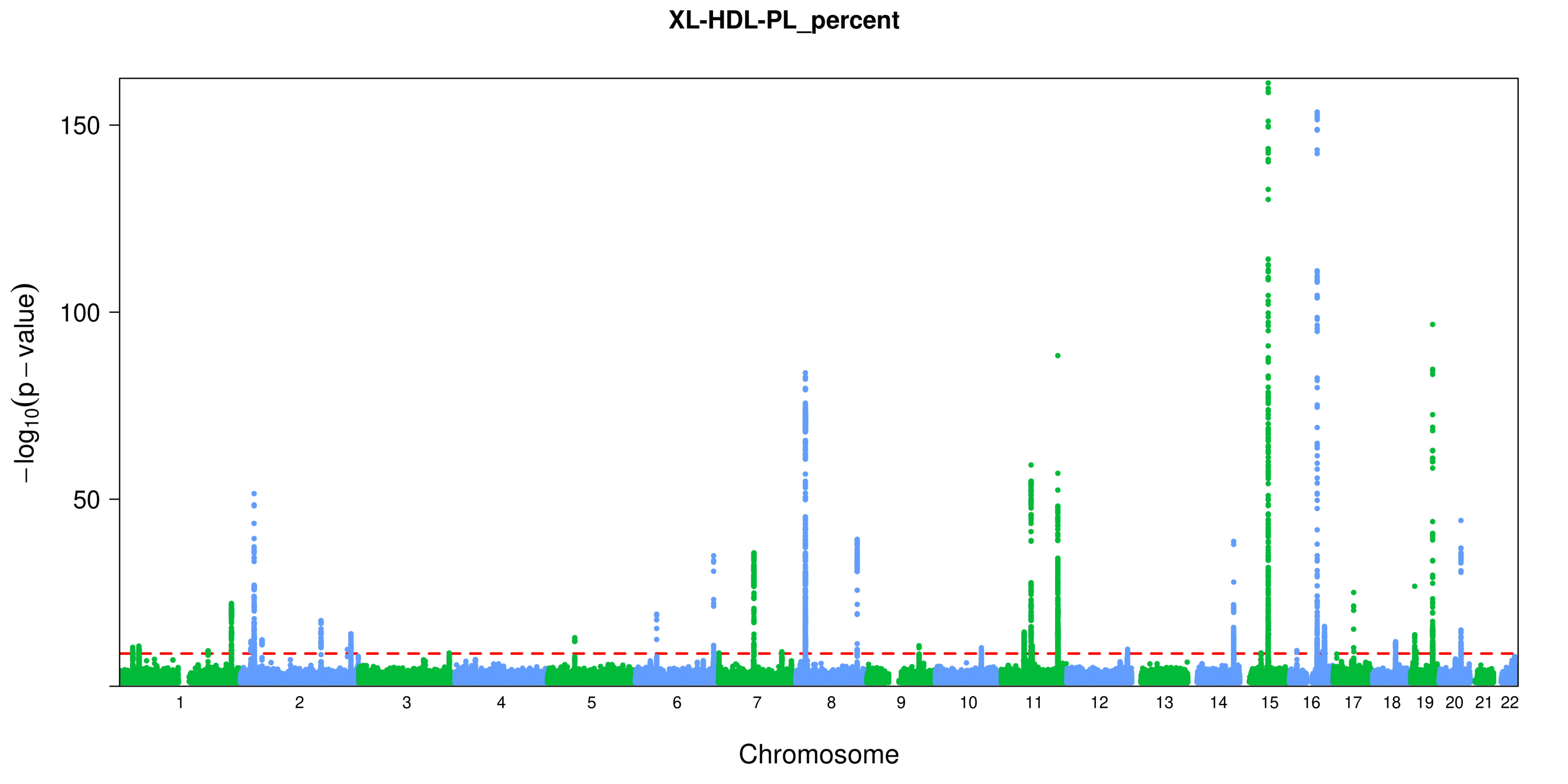

XL-HDL-PL_percent
150
100
50
−log10(p − value)
Chromosome

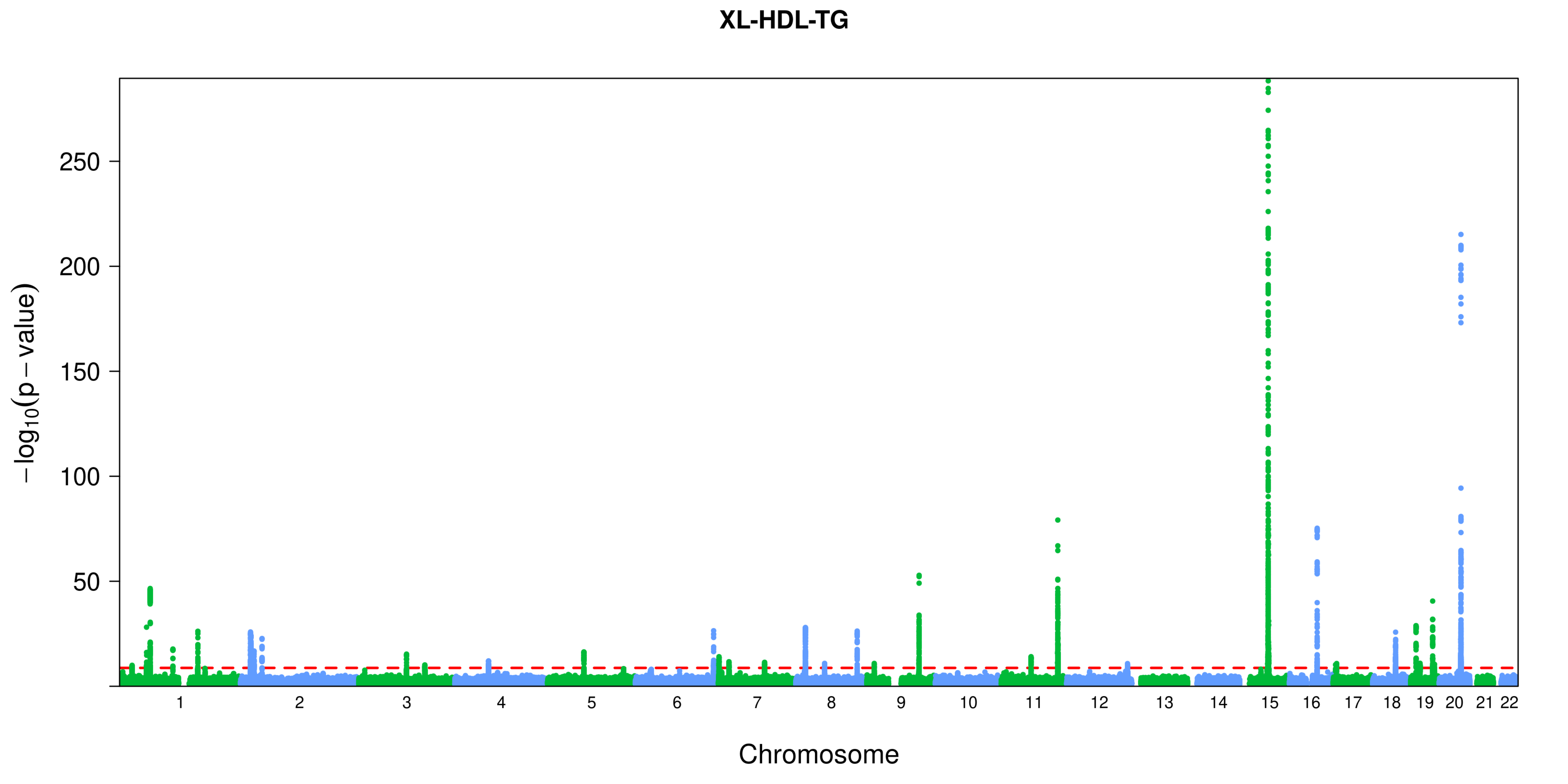
XL-HDL-TG
$-\log_{10}(p-\text{value})$
250
200
150
100
50
Chromosome

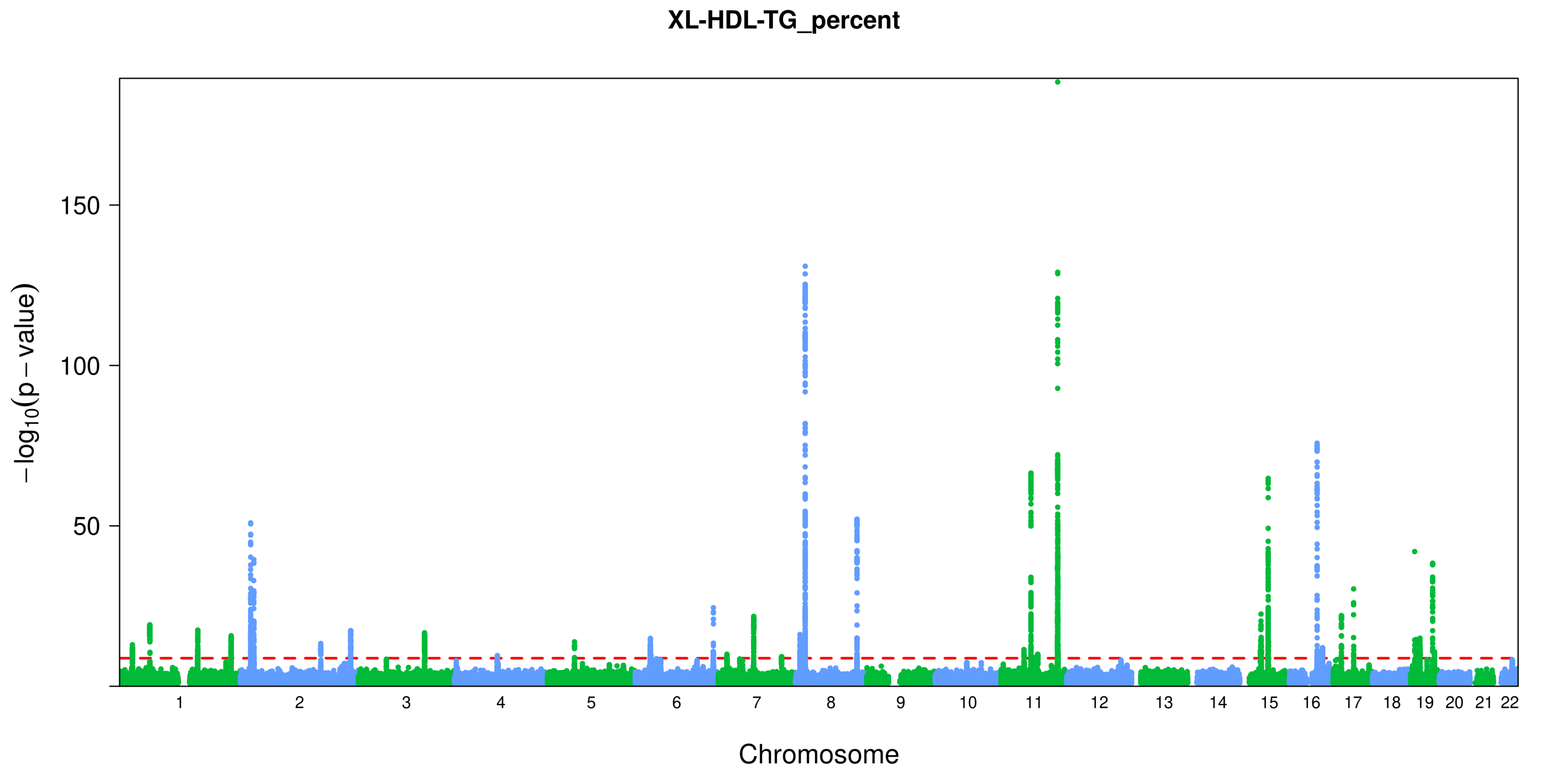

XL-HDL-TG_percent
150
100
50
−log10(p − value)
Chromosome

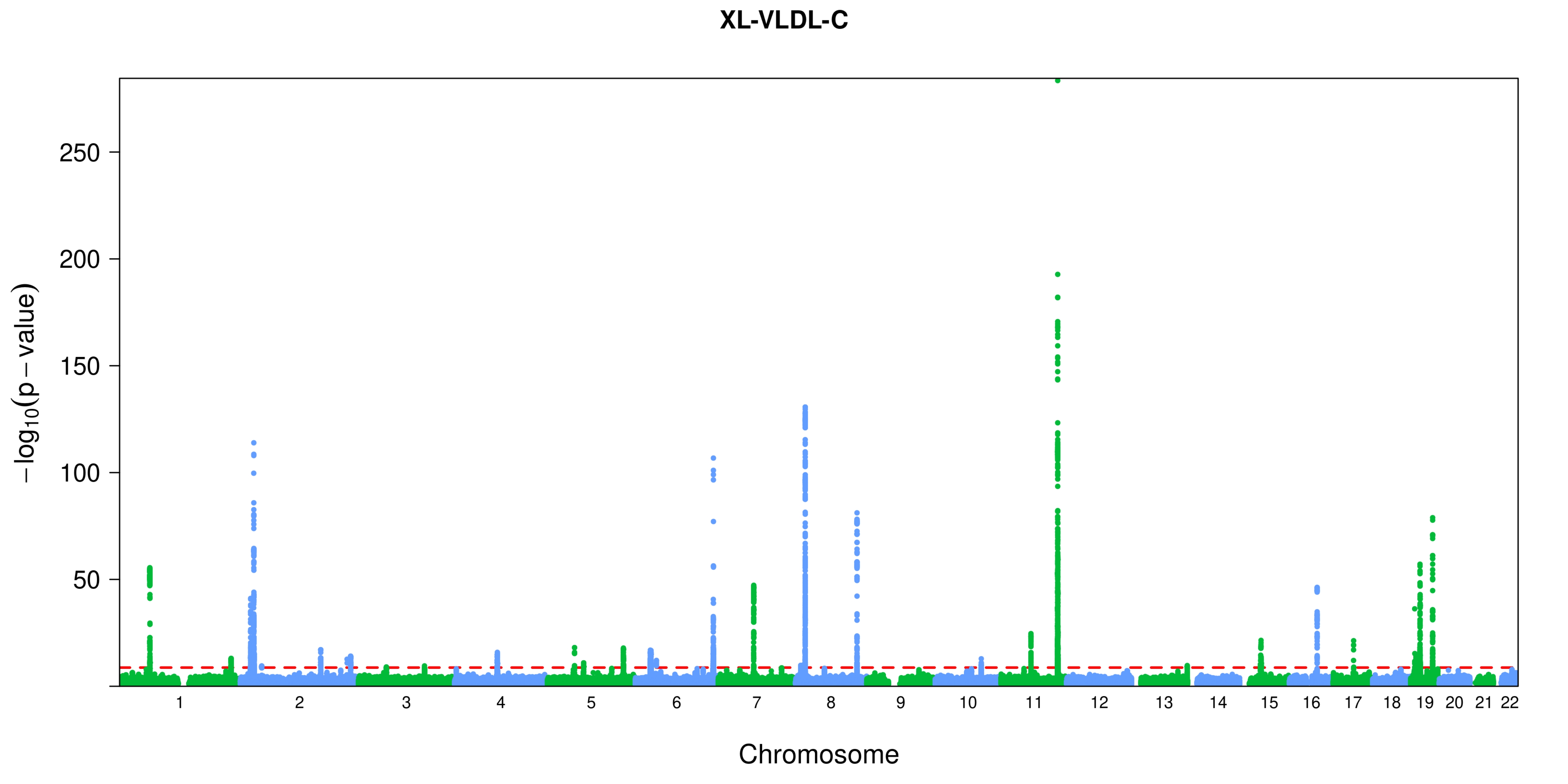
XL-VLDL-C
−log10(p − value)
250
200
150
100
50
Chromosome

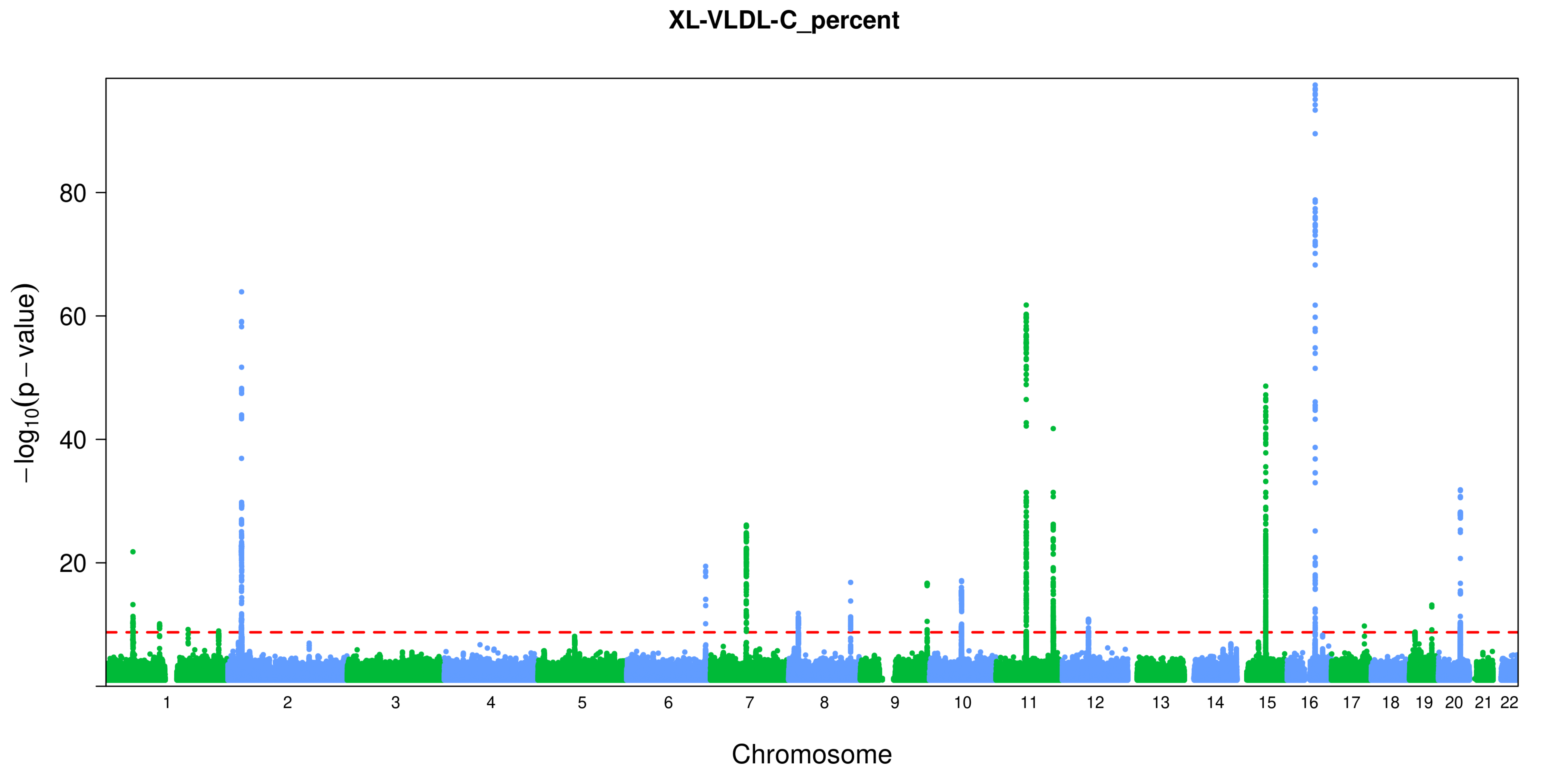
XL-VLDL-C_percent
−log10(p − value)
80
60
40
20
Chromosome

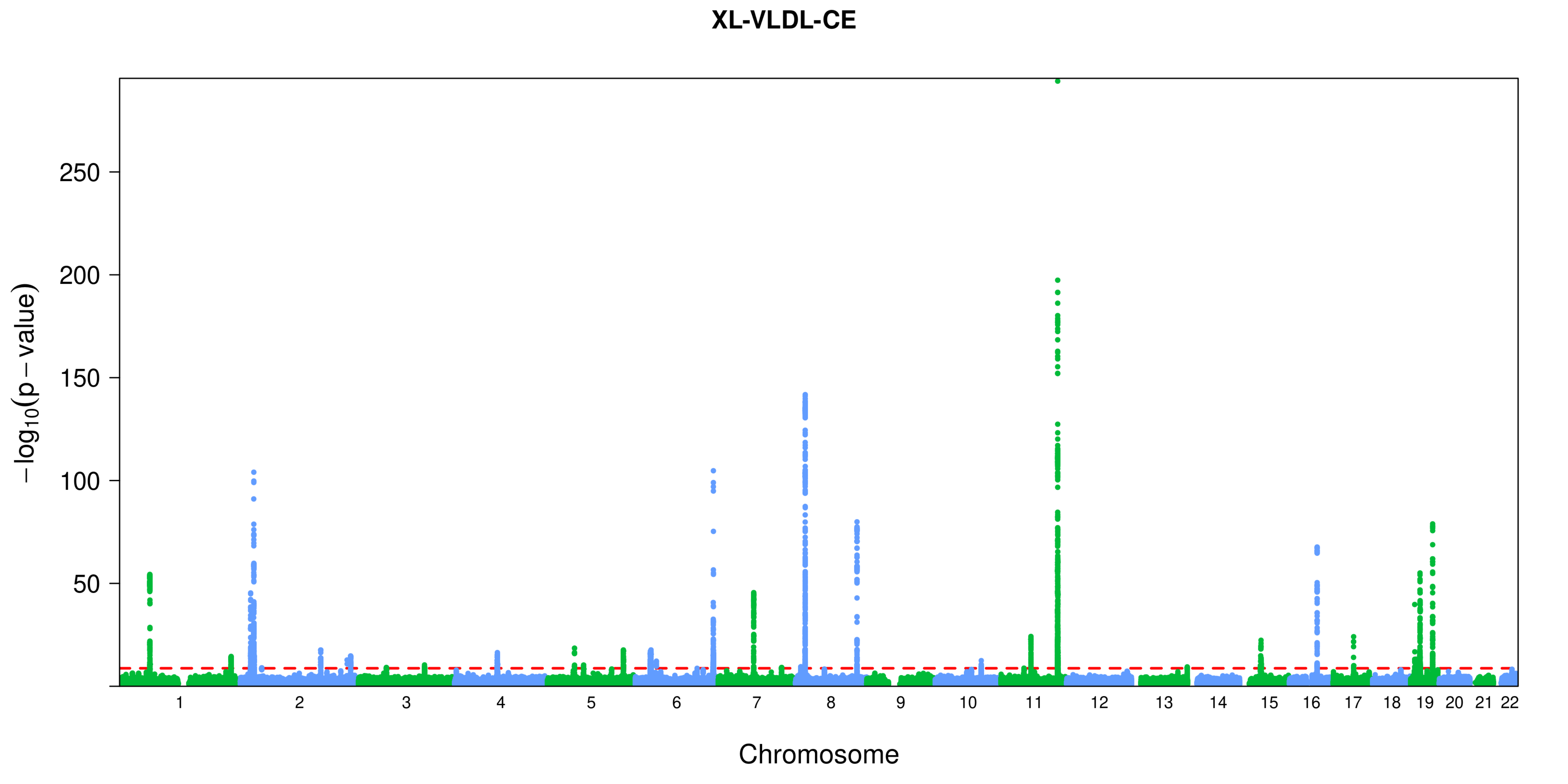

XL-VLDL-CE
−log10(p − value)
250
200
150
100
50
Chromosome

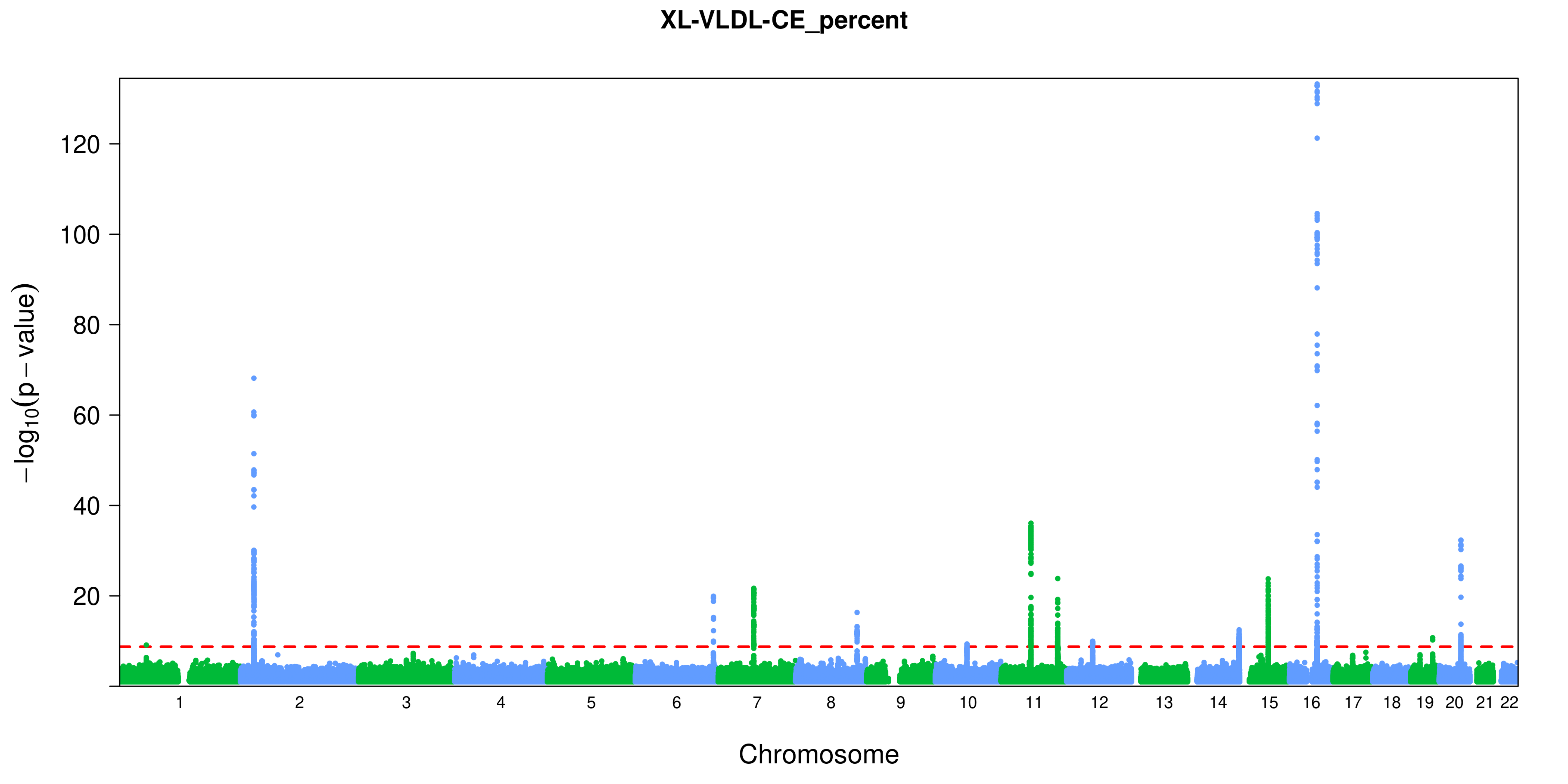
XL-VLDL-CE_percent
−log10(p − value)
0
20
40
60
80
100
120
Chromosome

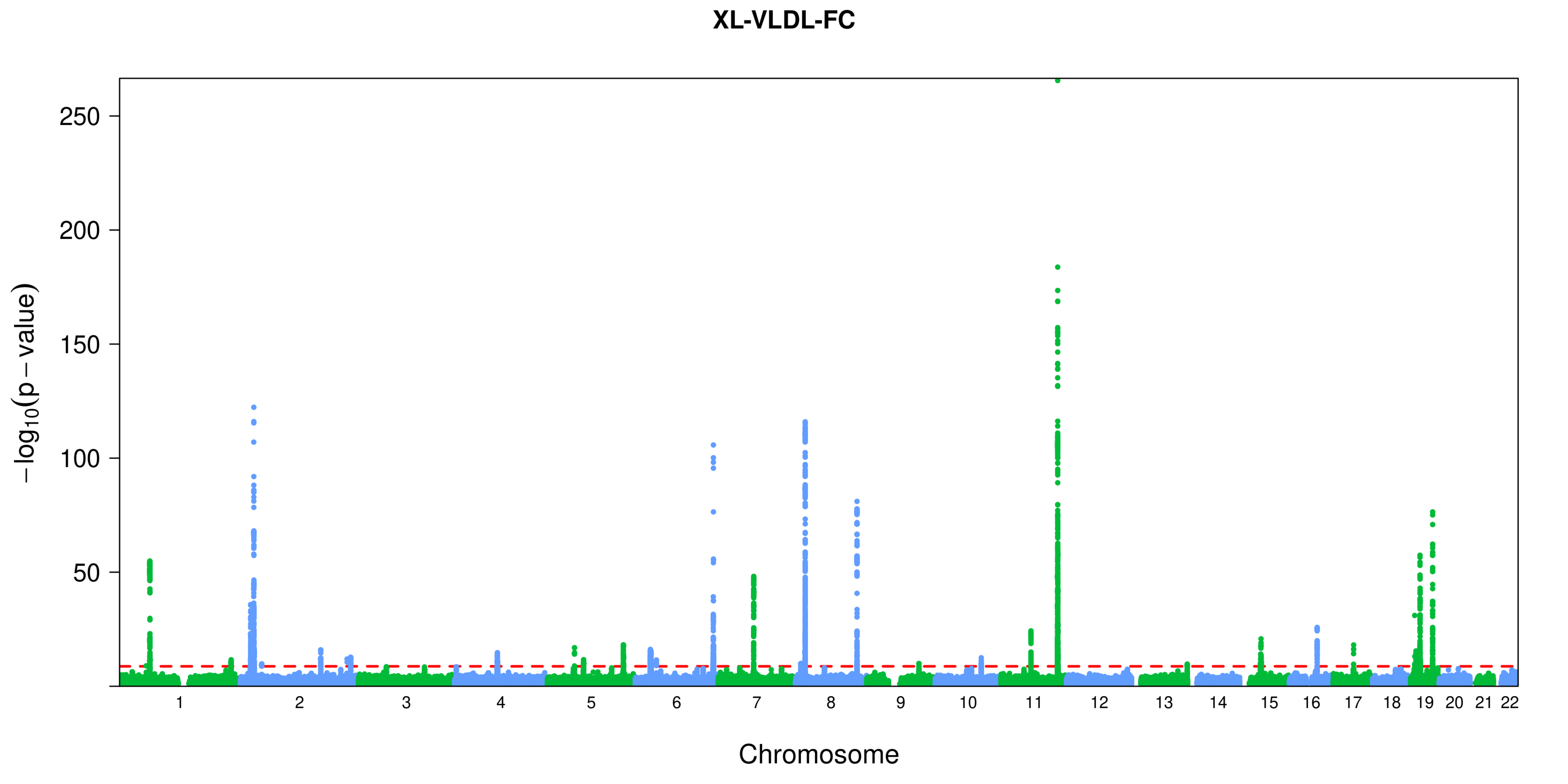
XL-VLDL-FC
250
200
150
100
50
−log10(p − value)
Chromosome

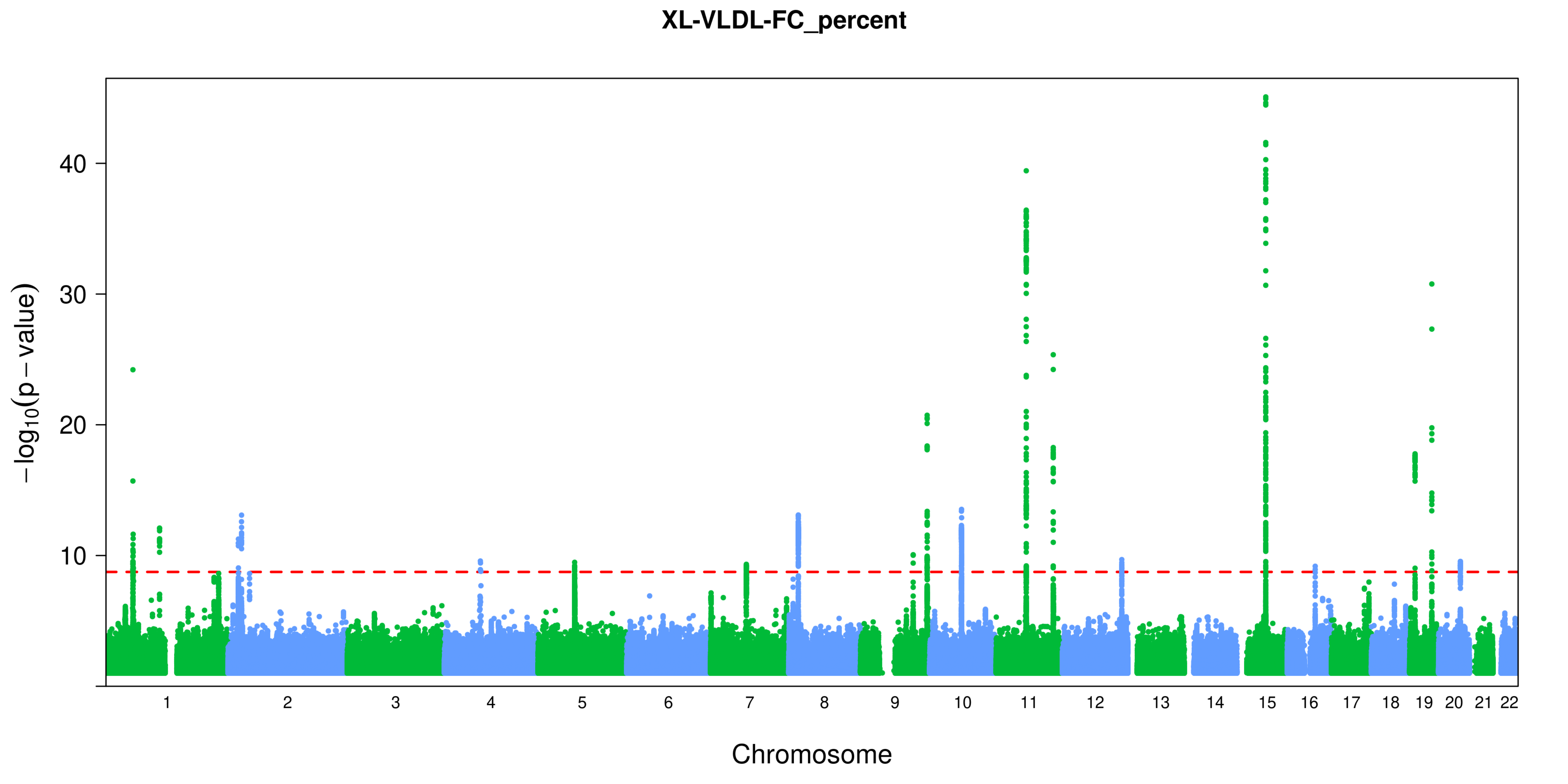

XL-VLDL-FC_percent
$-\log_{10}(p-\text{value})$
10
20
30
40
Chromosome

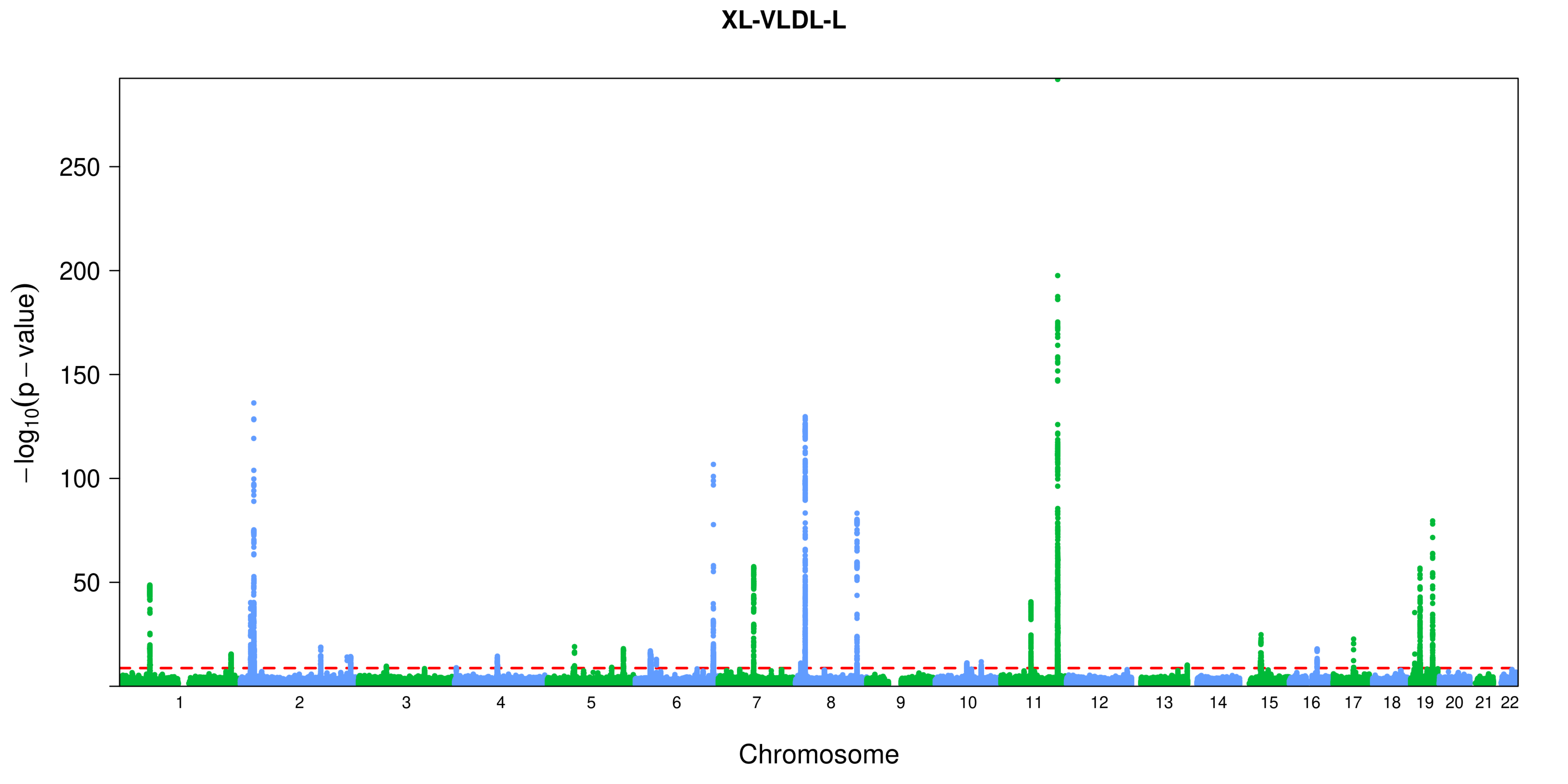

XL-VLDL-L
−log10(p − value)
250
200
150
100
50
Chromosome

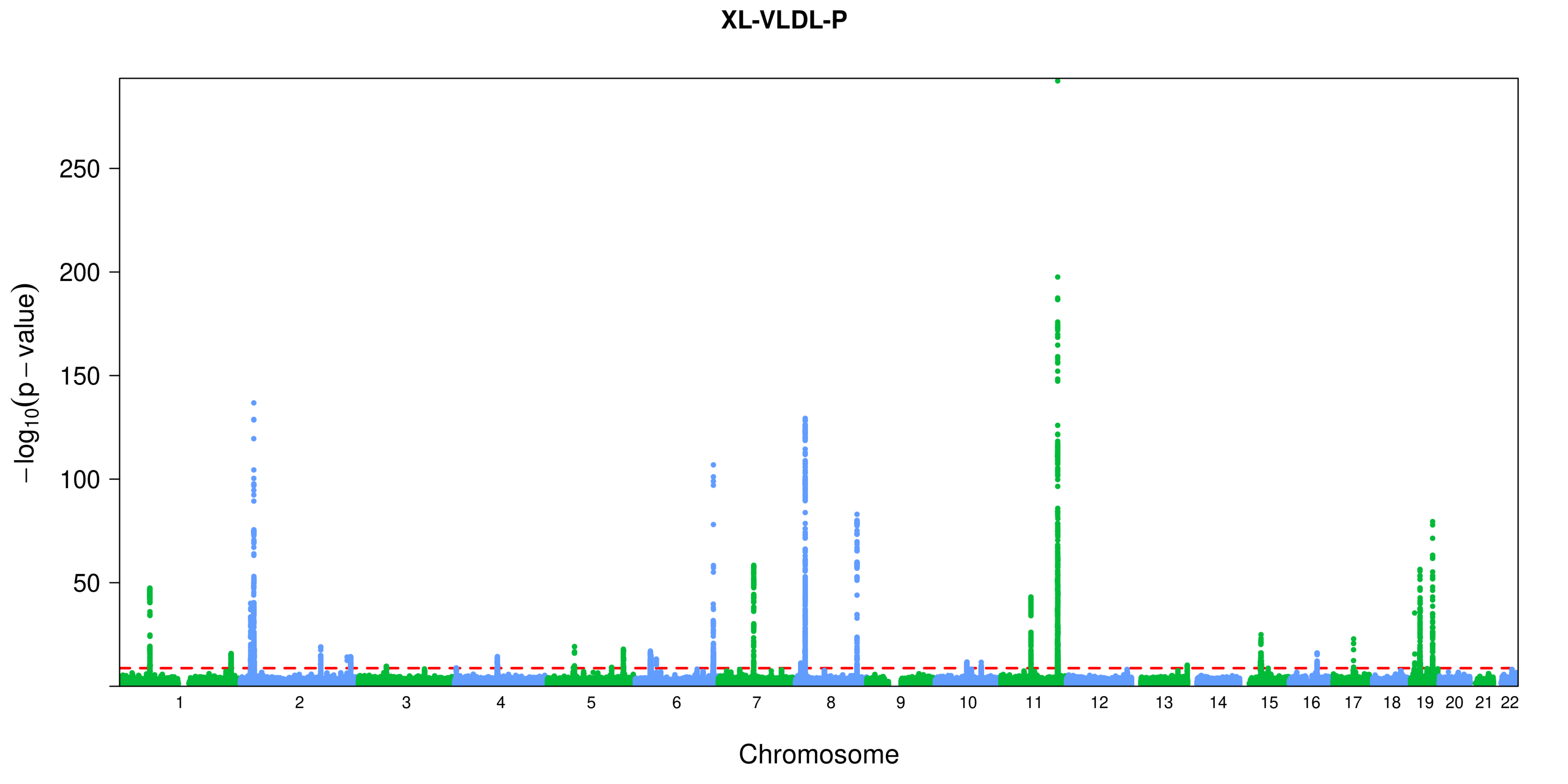

XL-VLDL-P
250
200
150
100
50
$-\log_{10}(p-\text{value})$
Chromosome

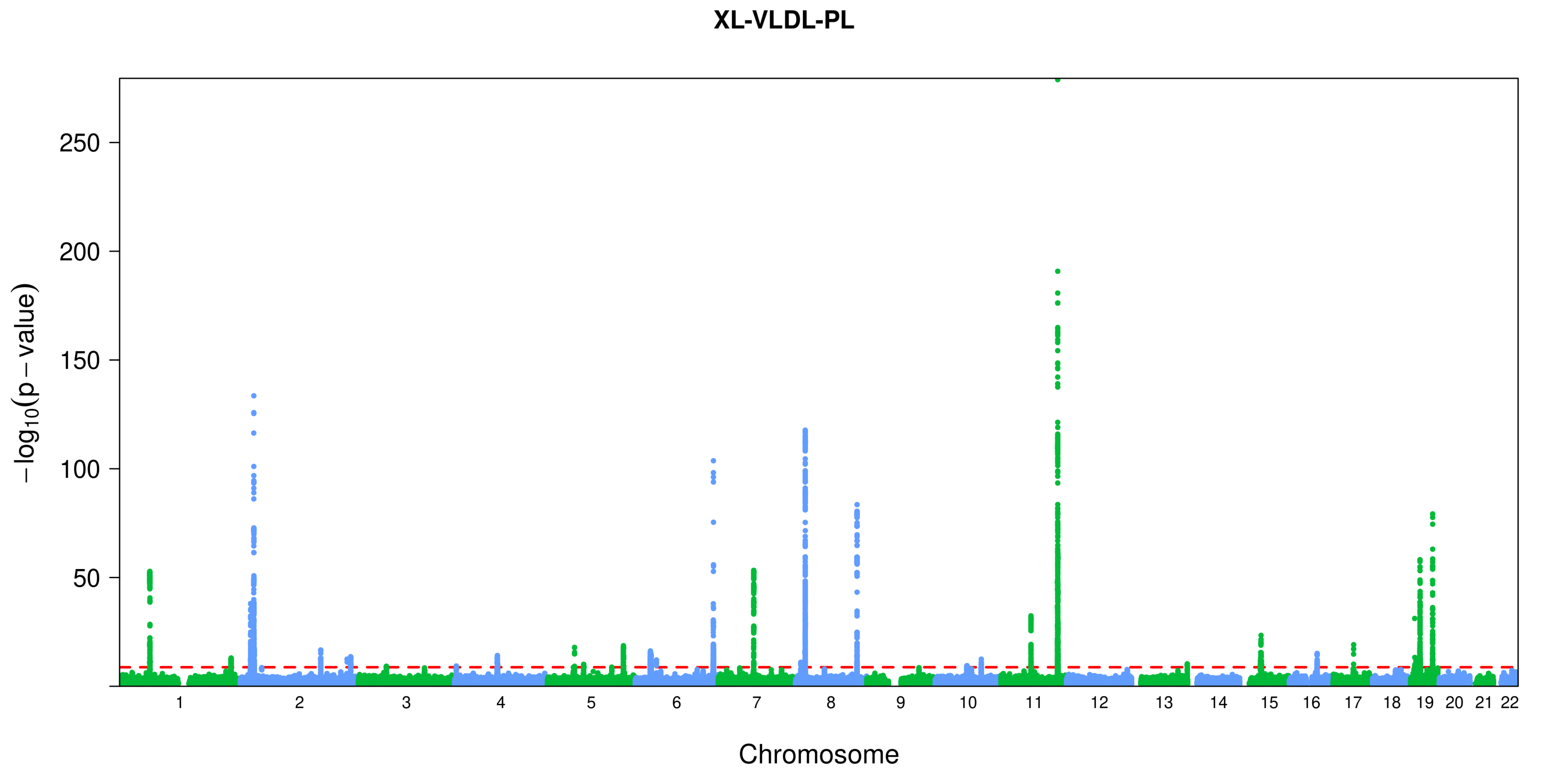
XL-VLDL-PL
$-\log_{10}(p-\text{value})$
250
200
150
100
50
Chromosome

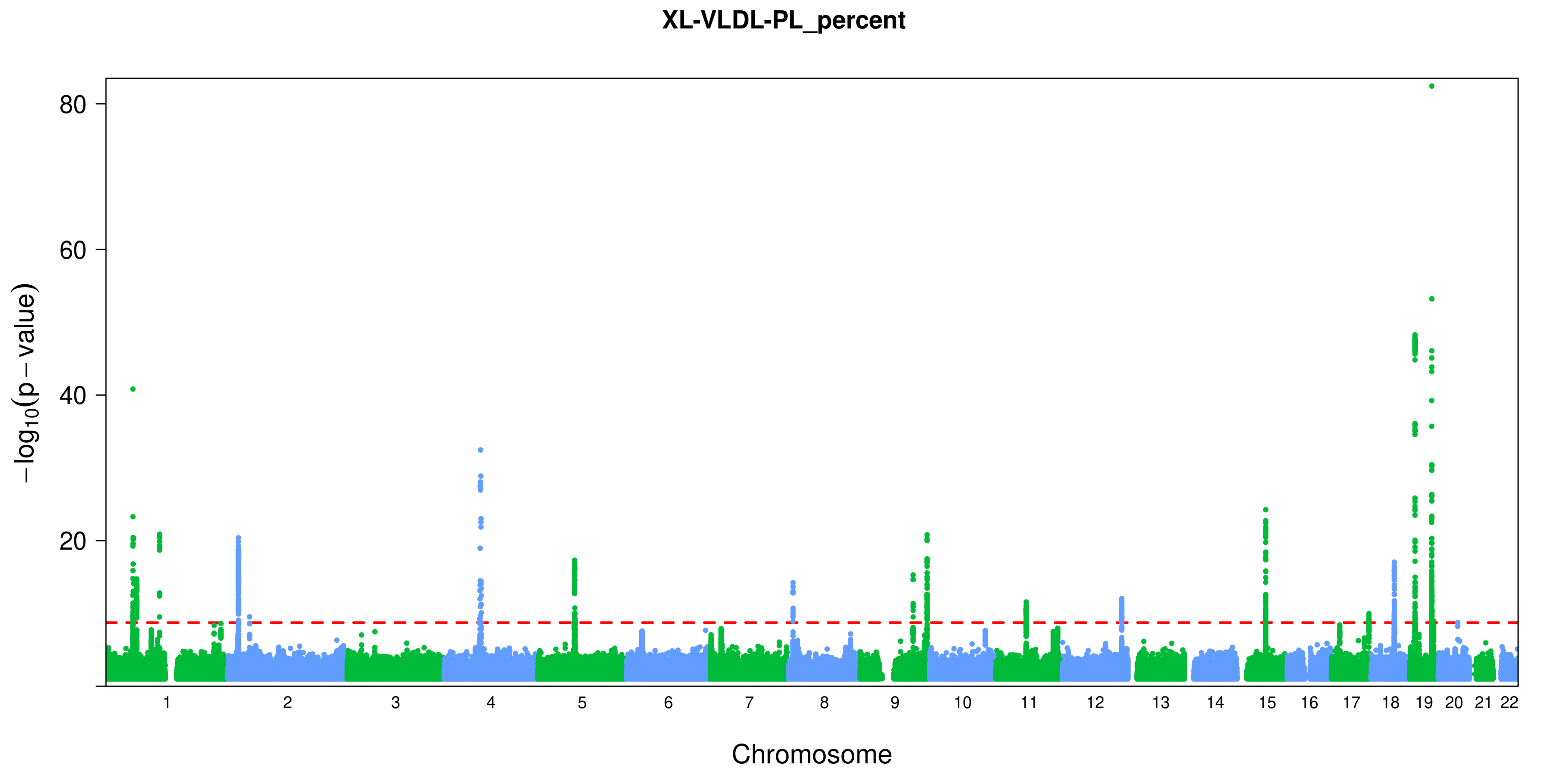
XL-VLDL-PL_percent
80
60
40
20
−log10(p − value)
Chromosome

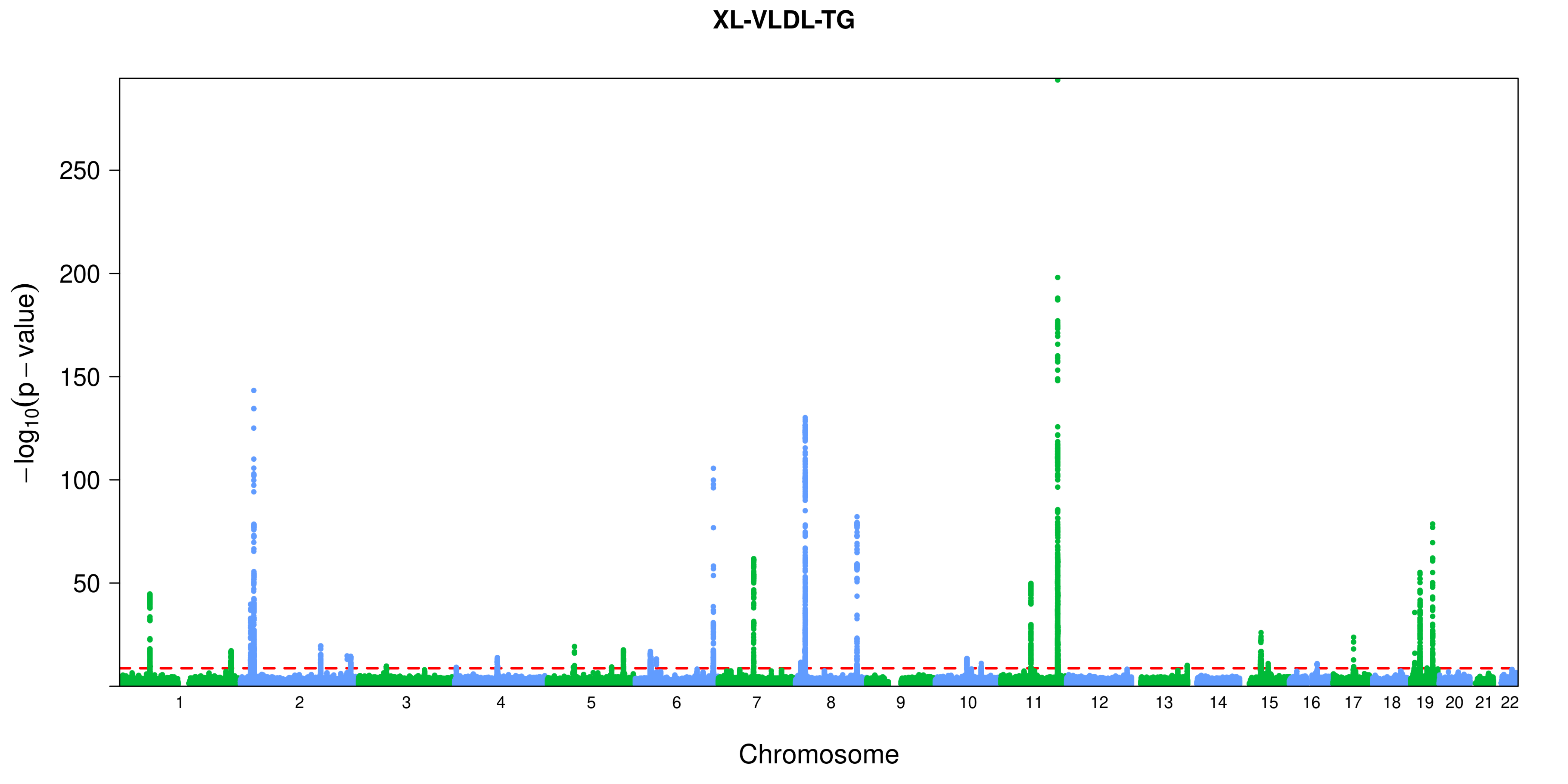

XL-VLDL-TG
−log10(p − value)
250
200
150
100
50
Chromosome

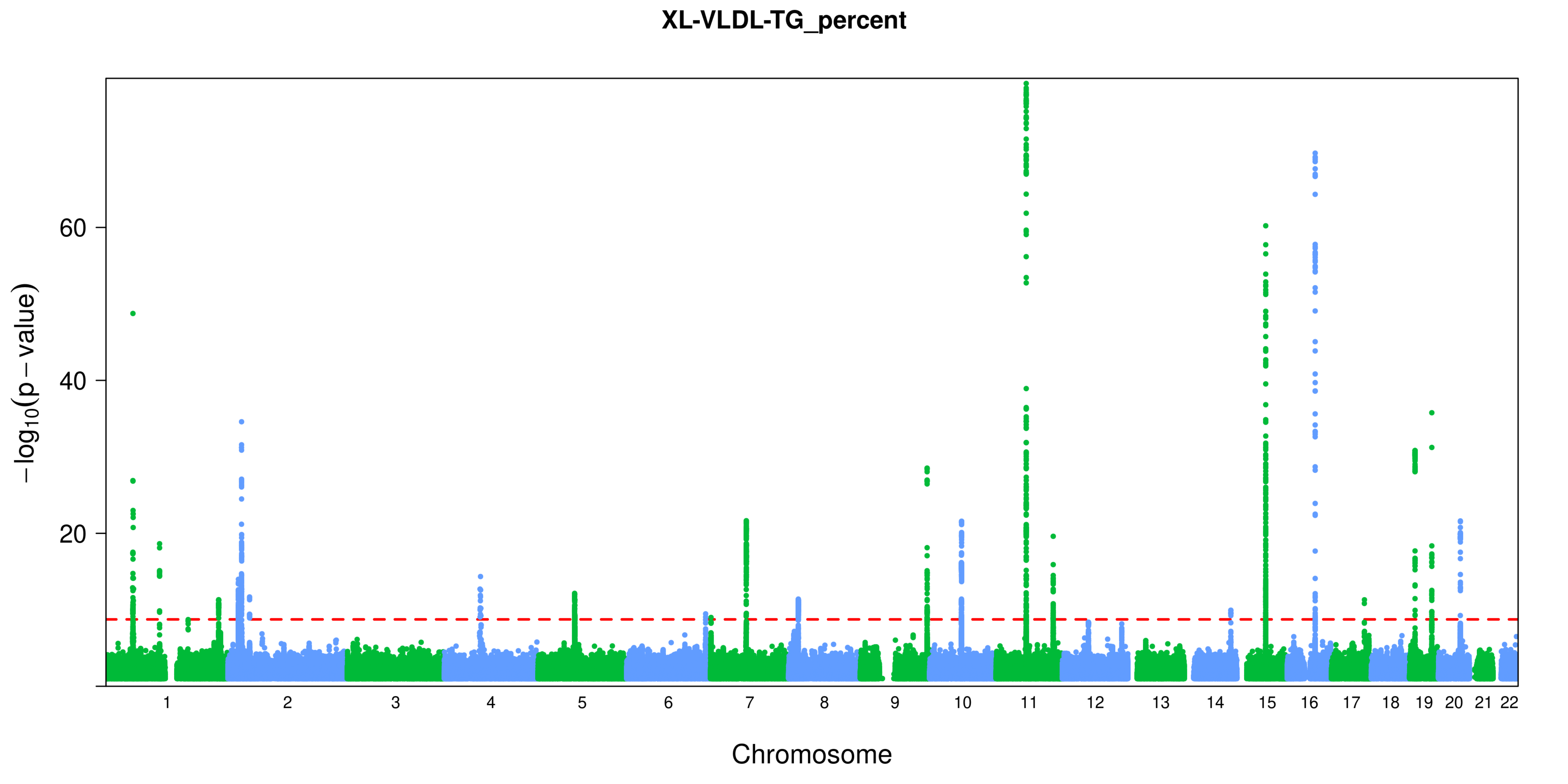
XL-VLDL-TG_percent
−log10(p − value)
60
40
20
Chromosome

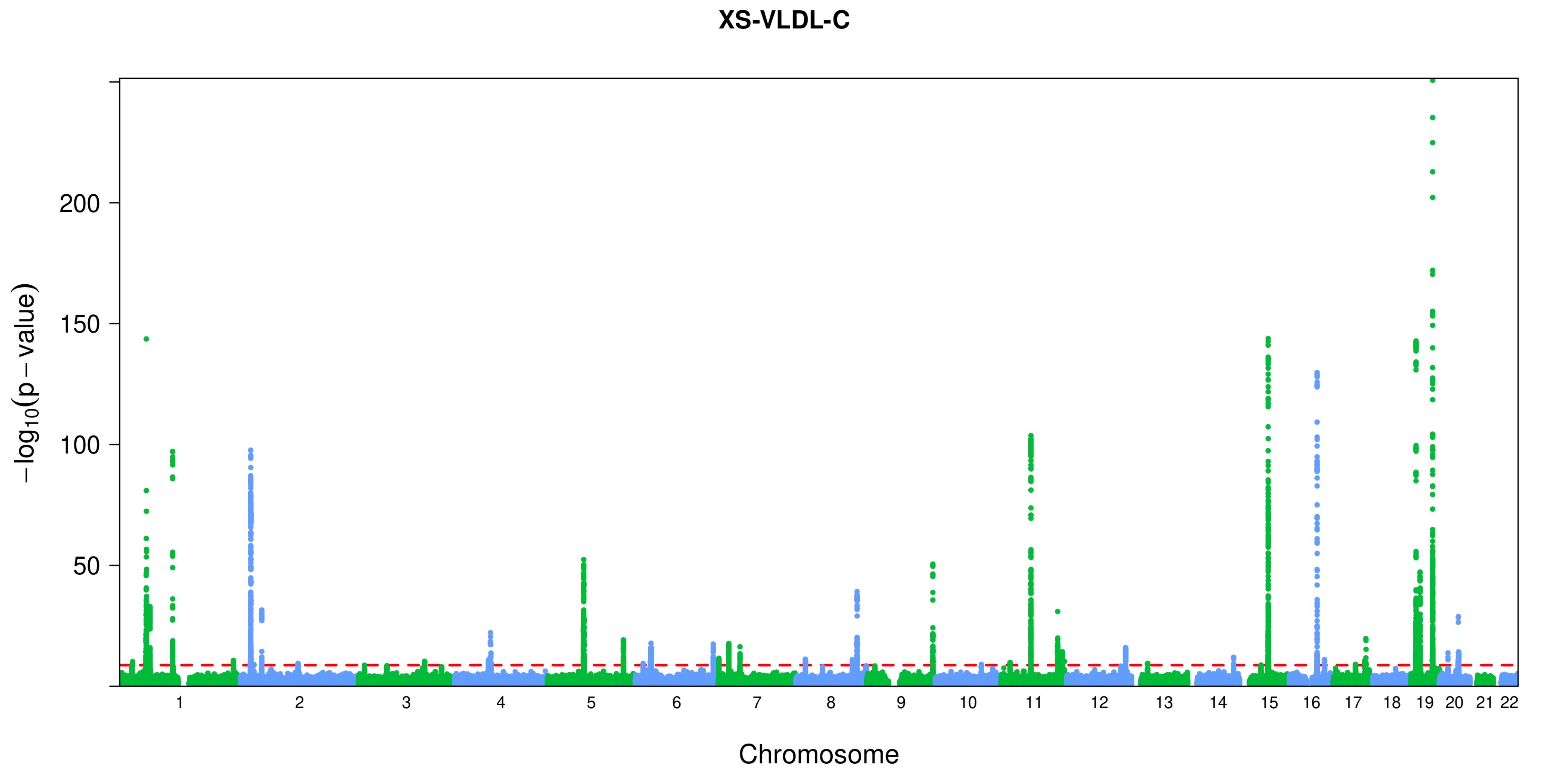
XS-VLDL-C
200
150
100
50
−log10(p − value)
Chromosome

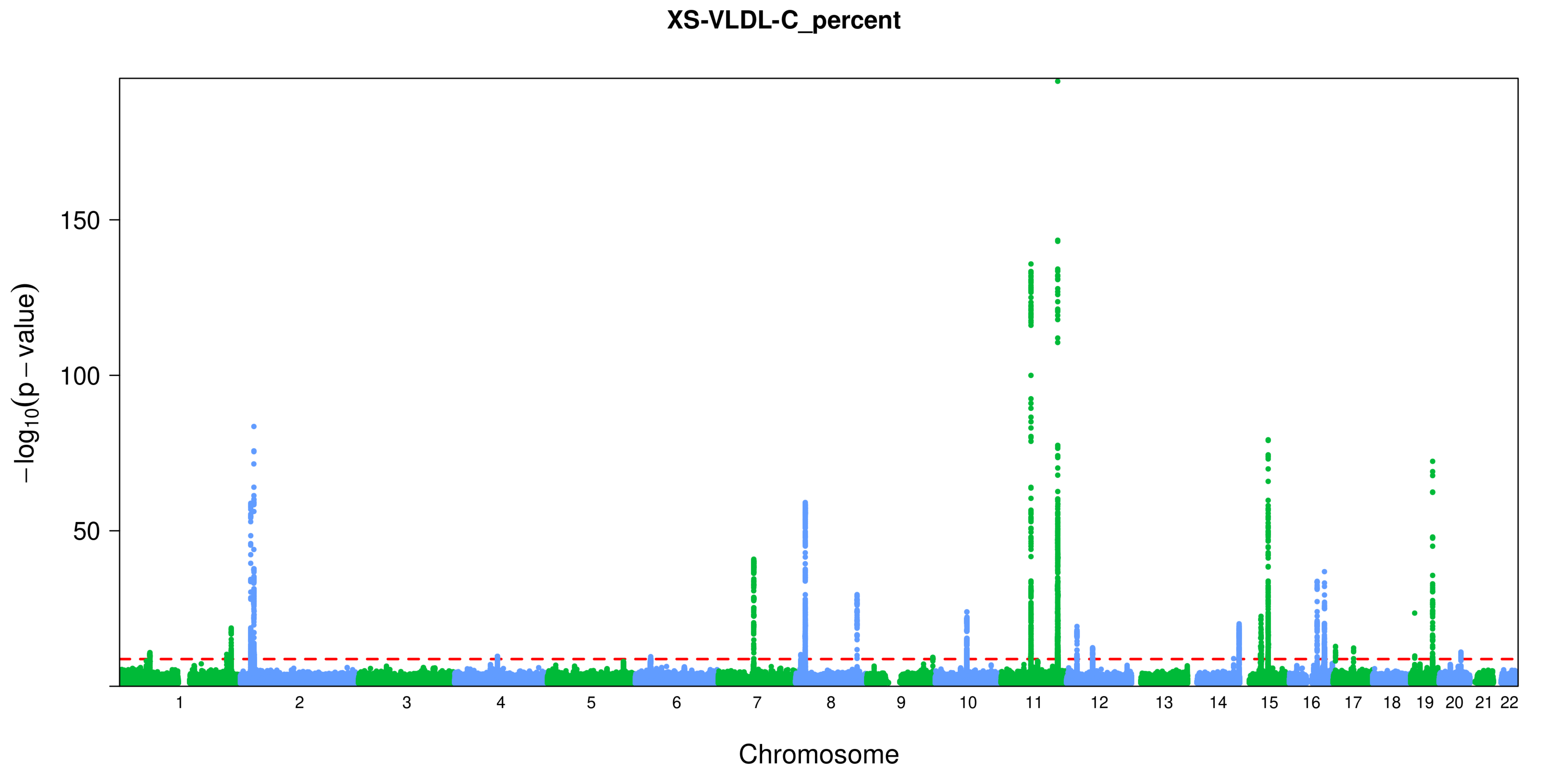
XS-VLDL-C_percent
150
100
50
$-\log_{10}(p-value)$
Chromosome

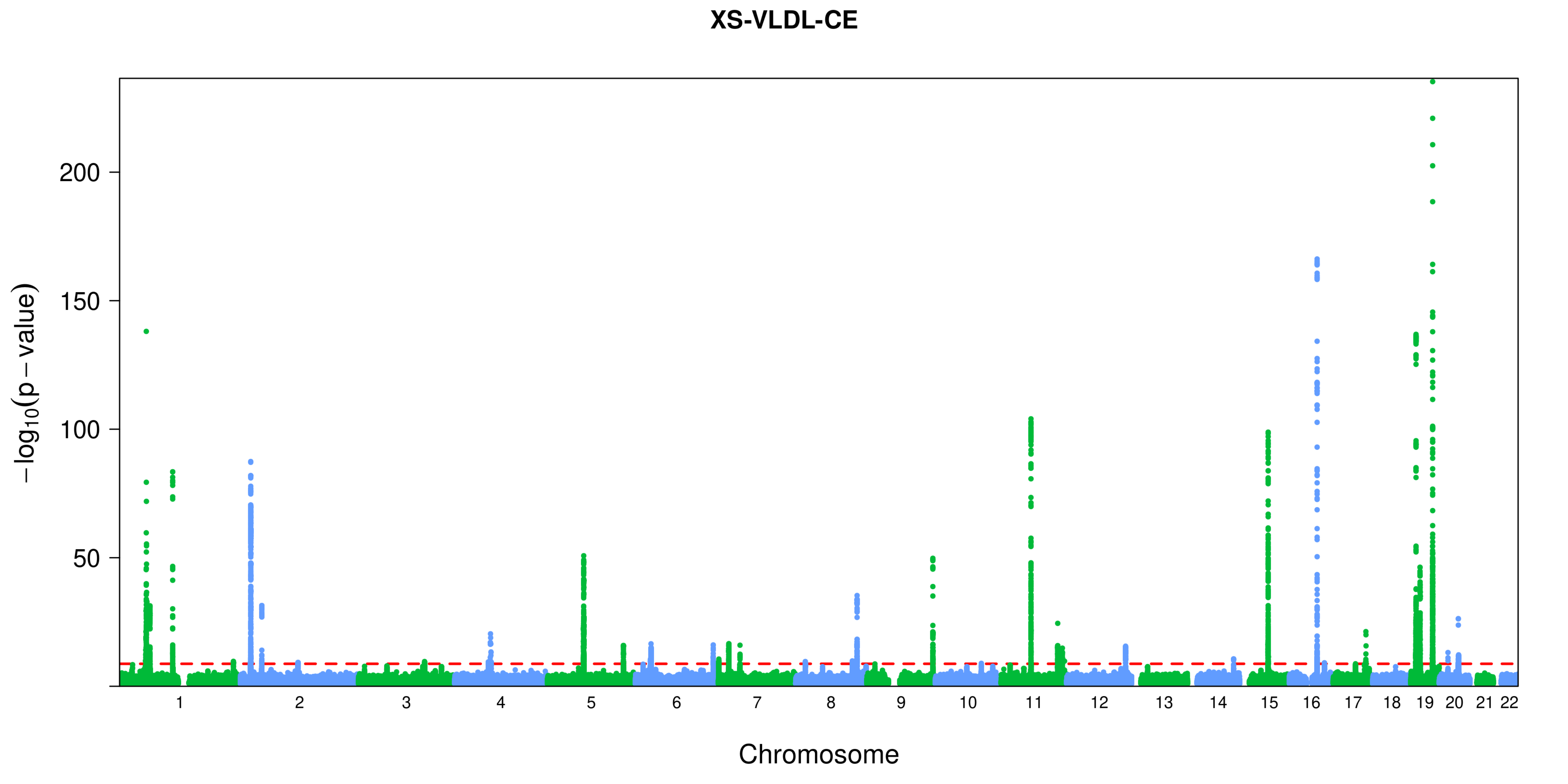
XS-VLDL-CE
$-\log_{10}(p-\text{value})$
200
150
100
50
Chromosome

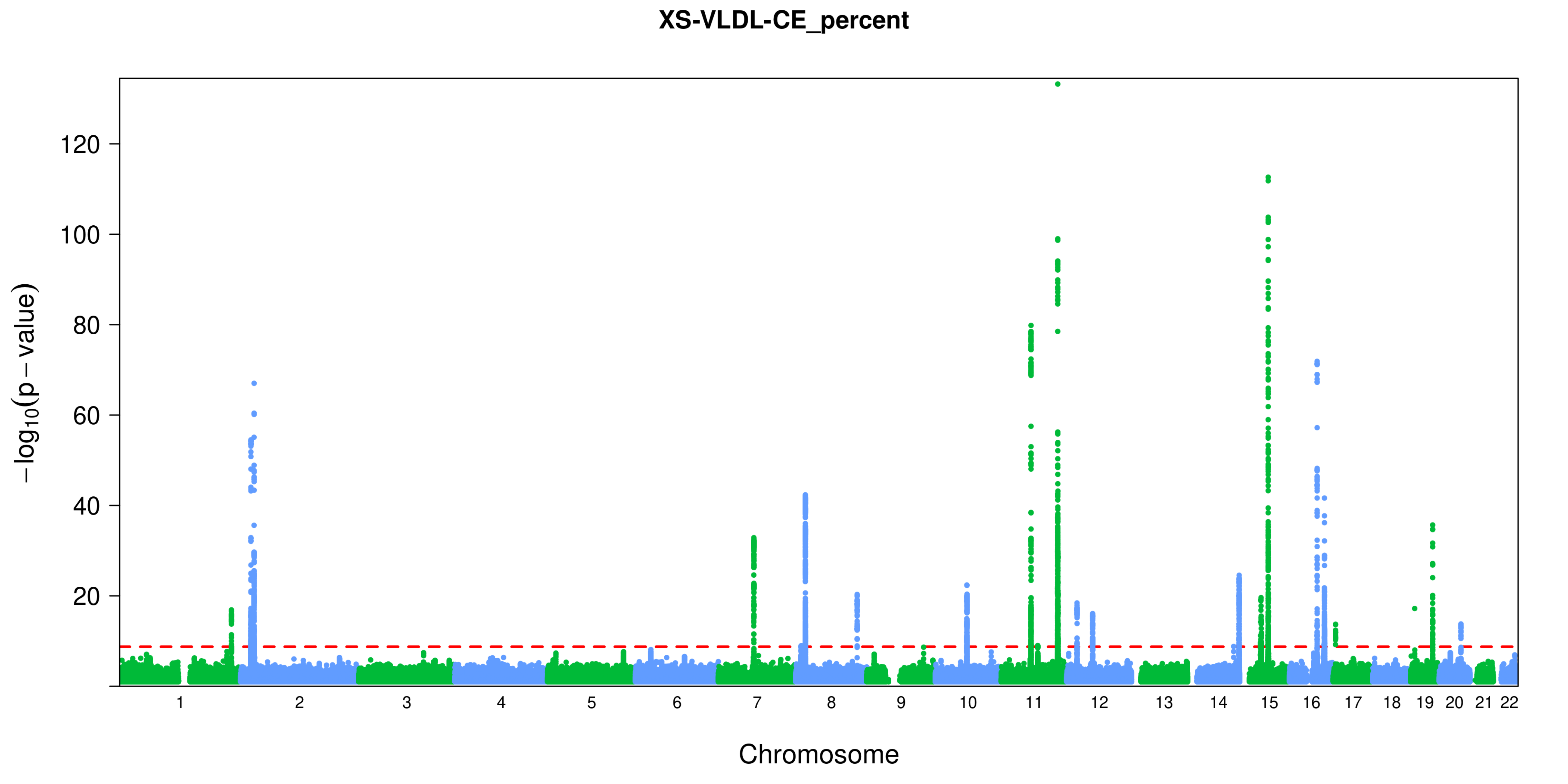

XS-VLDL-CE_percent
120
100
80
60
40
20
$-\log_{10}(p-\text{value})$
Chromosome

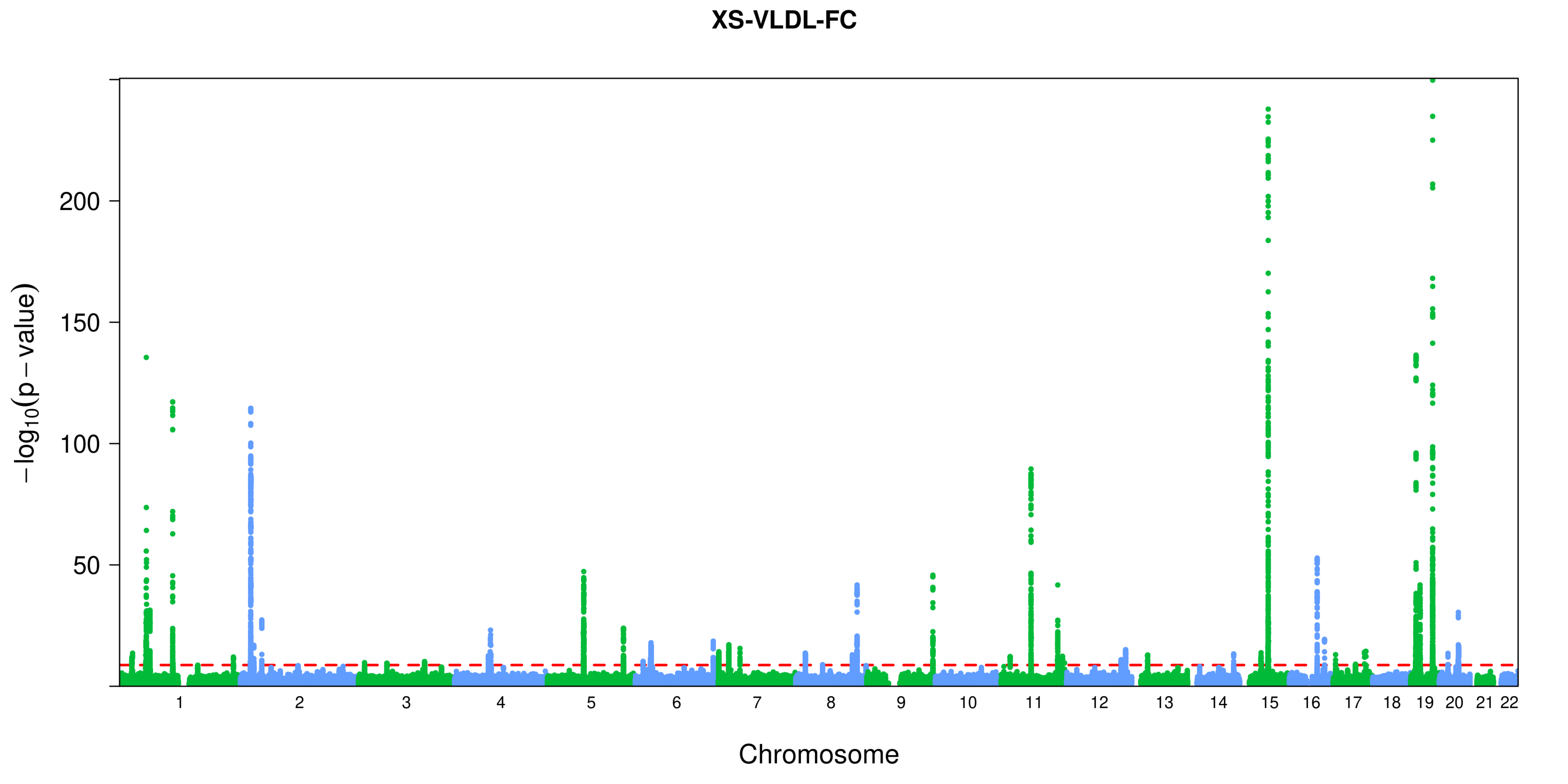
XS-VLDL-FC
$-\log_{10}(p-\text{value})$
200
150
100
50
Chromosome

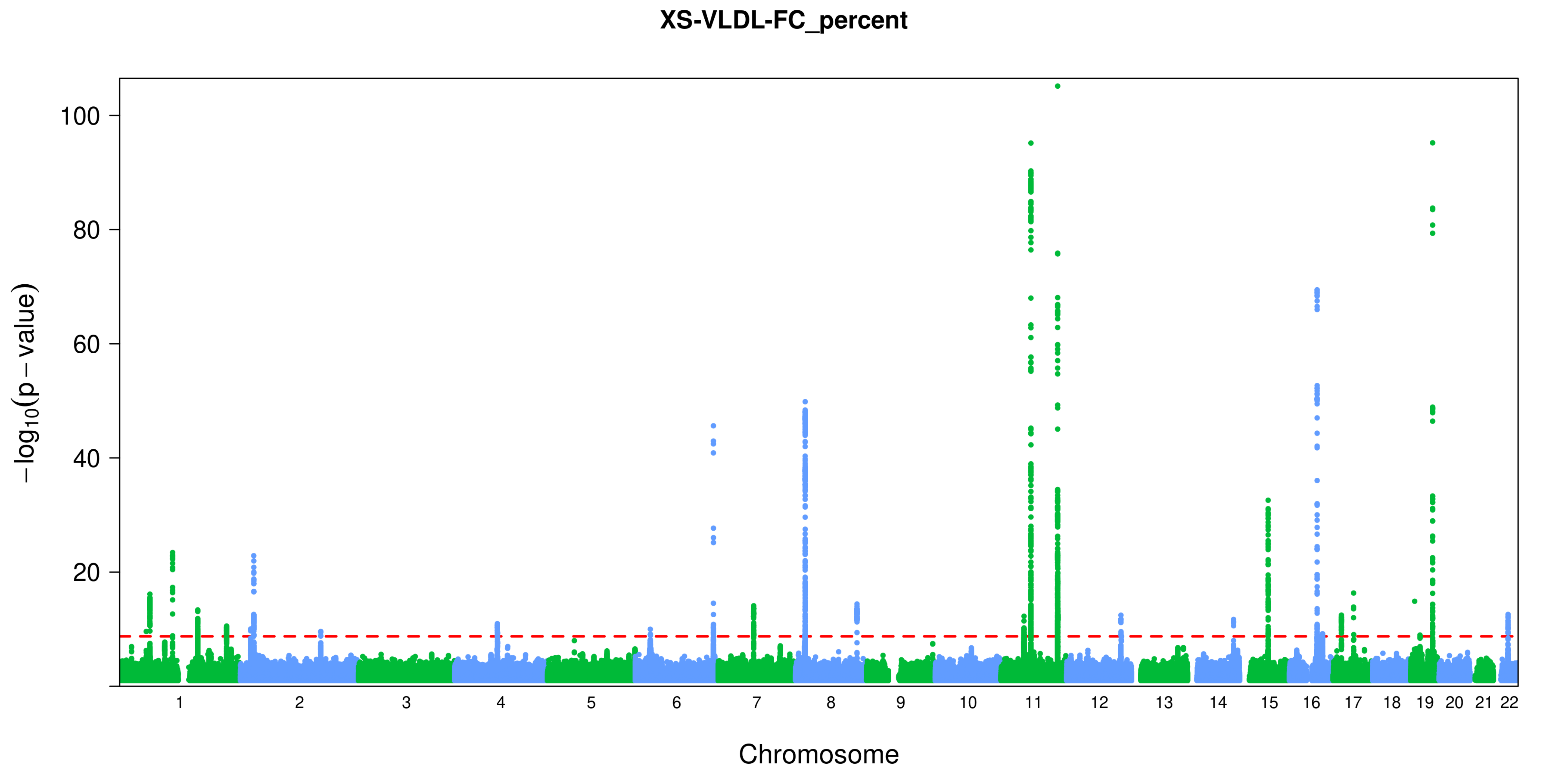

XS-VLDL-FC_percent
100
80
60
40
20
−log10(p − value)
Chromosome

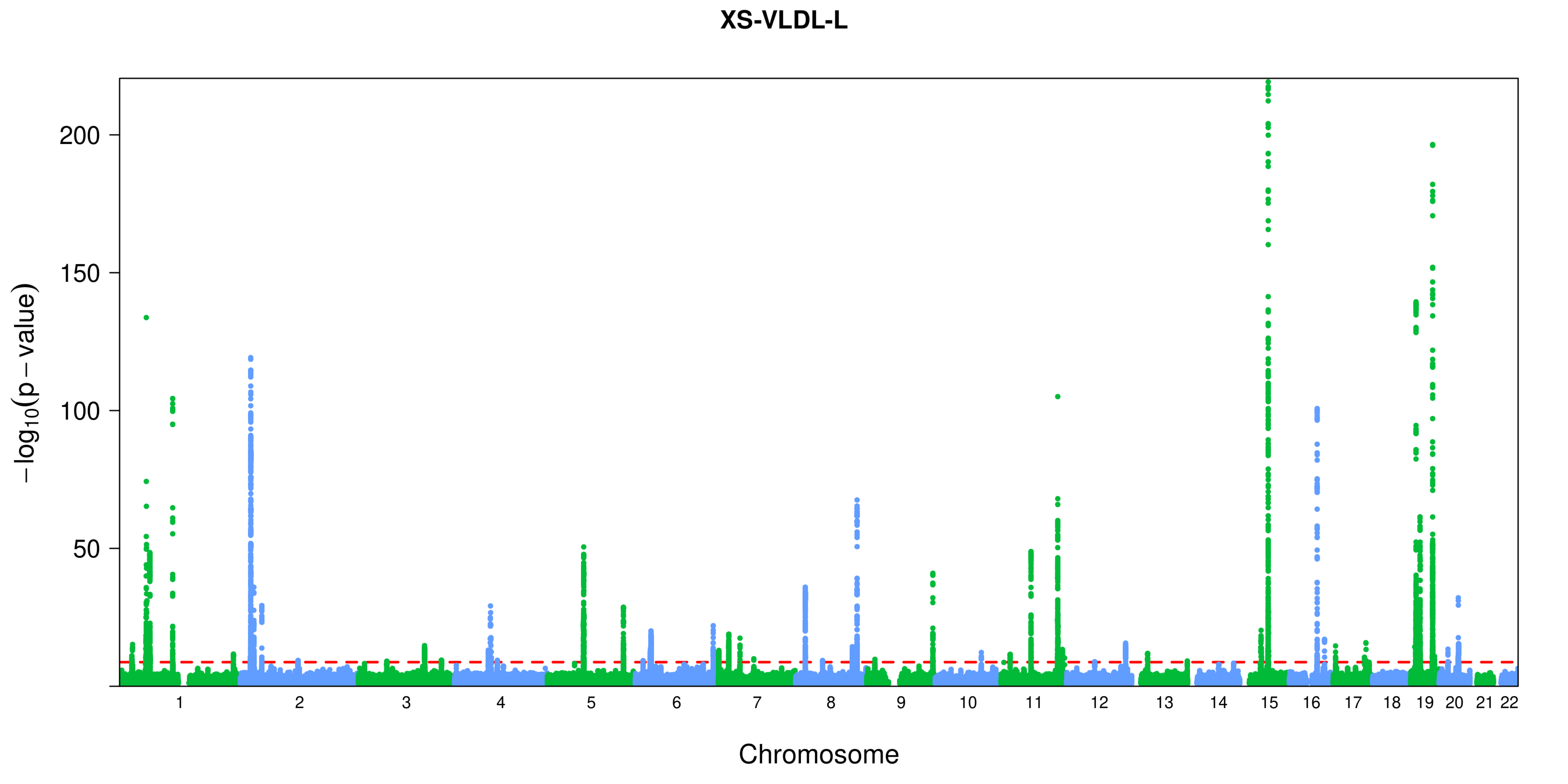

XS-VLDL-L
200
150
100
50
−log10(p − value)
Chromosome

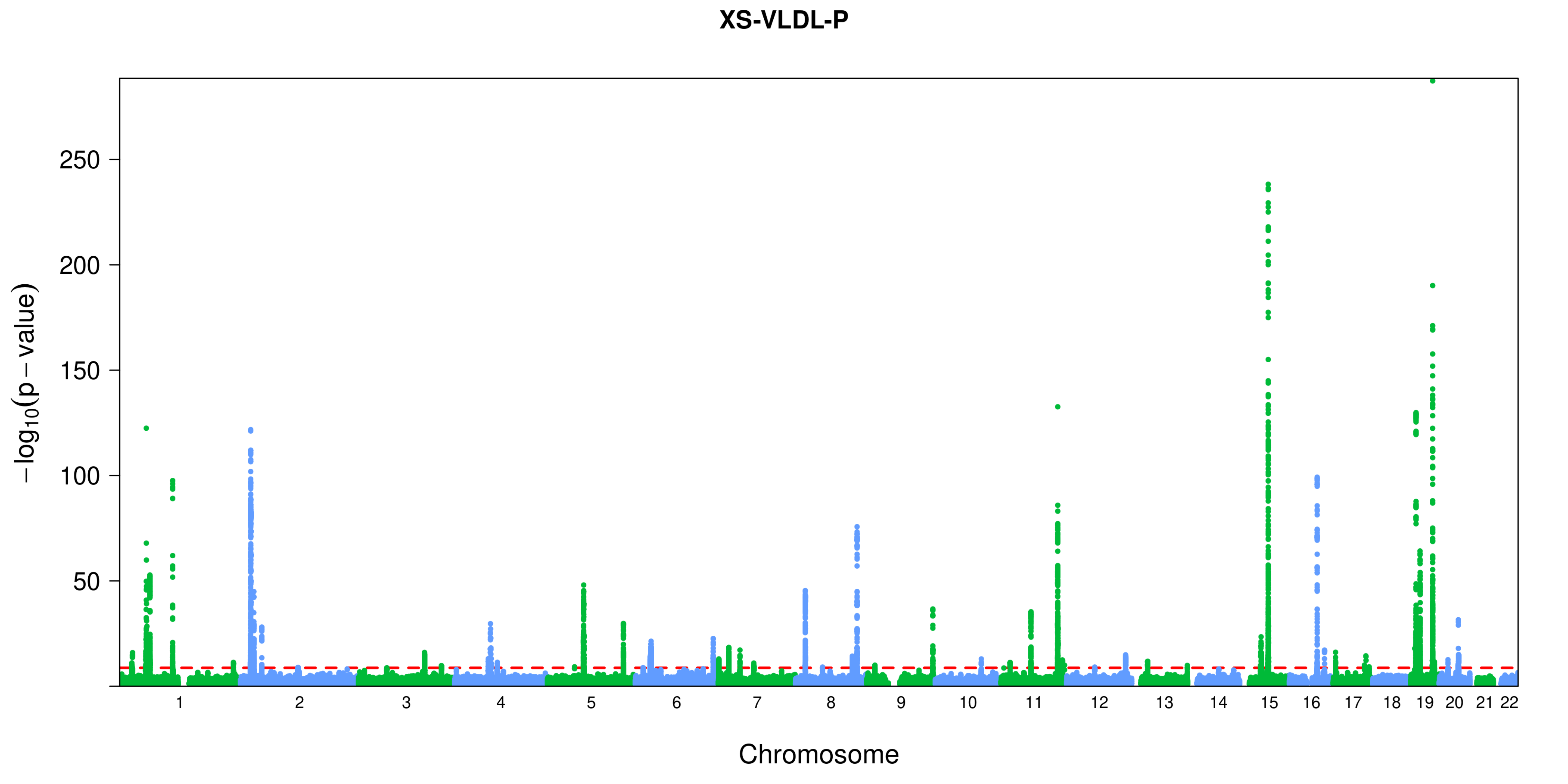

XS-VLDL-P
250
200
150
100
50
$-\log_{10}(p-\text{value})$
Chromosome

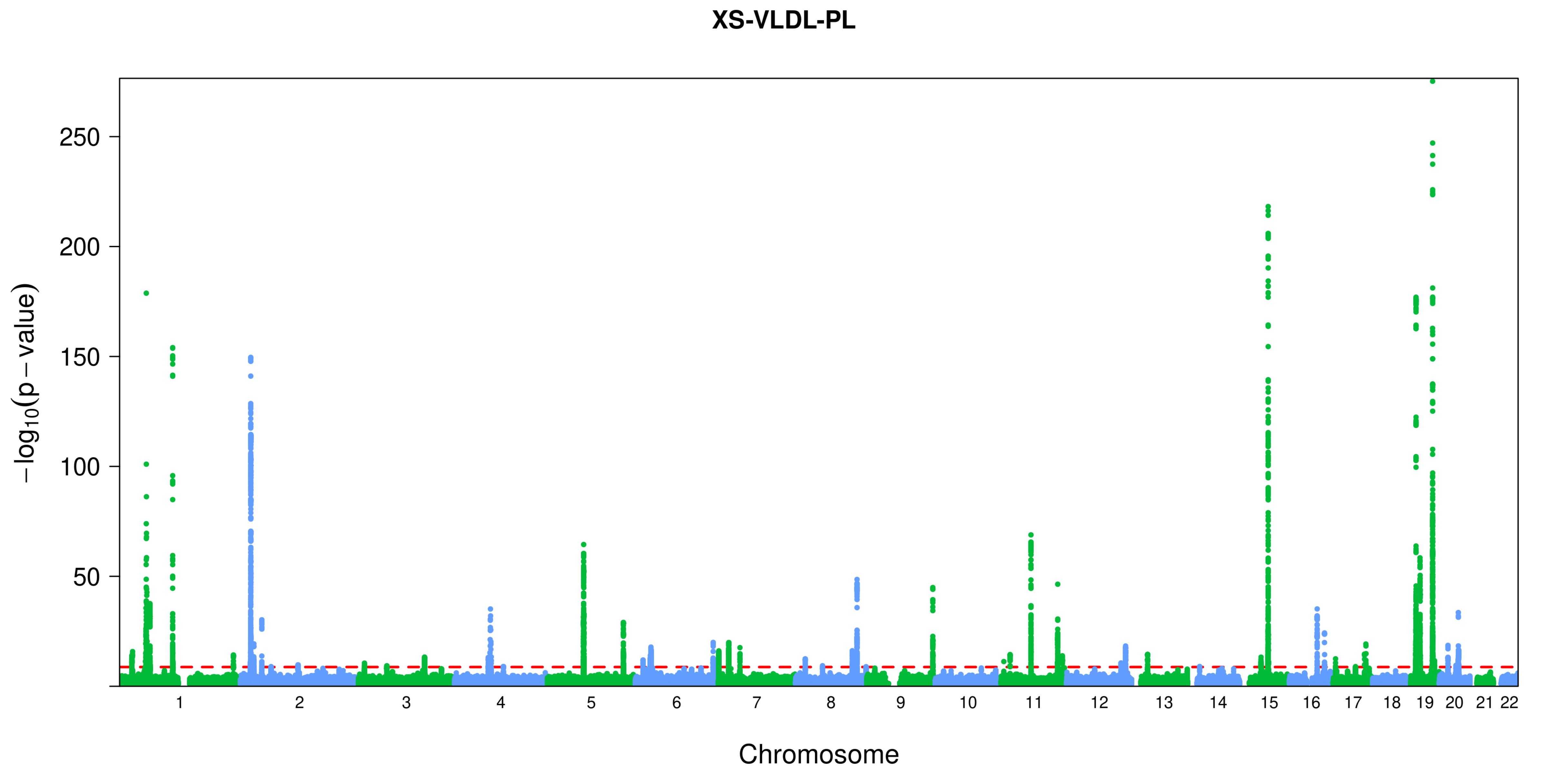
XS-VLDL-PL
−log10(p − value)
250
200
150
100
50
Chromosome

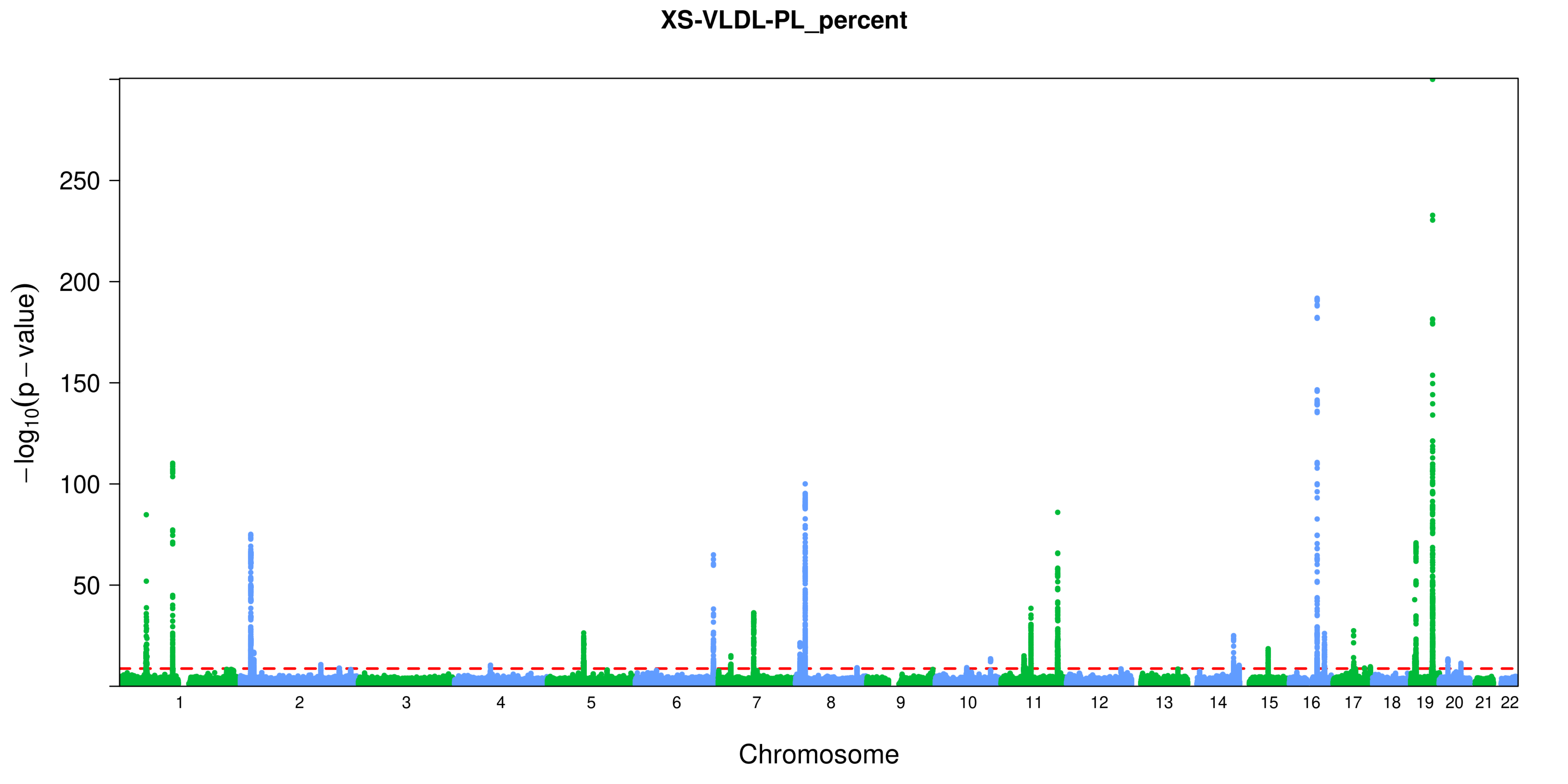

XS-VLDL-PL_percent
−log10(p − value)
250
200
150
100
50
Chromosome

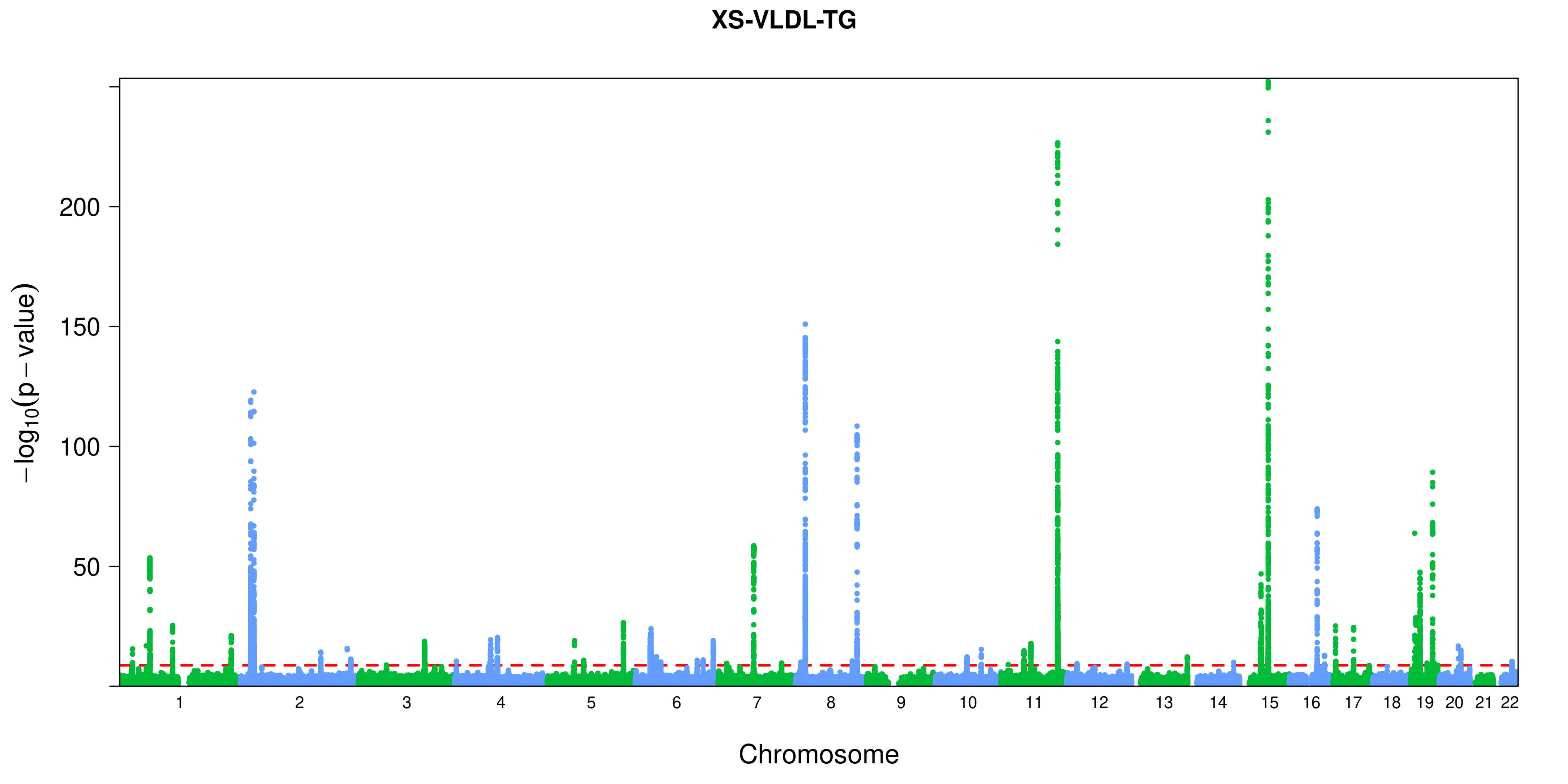
XS-VLDL-TG
−log10(p − value)
200
150
100
50
Chromosome

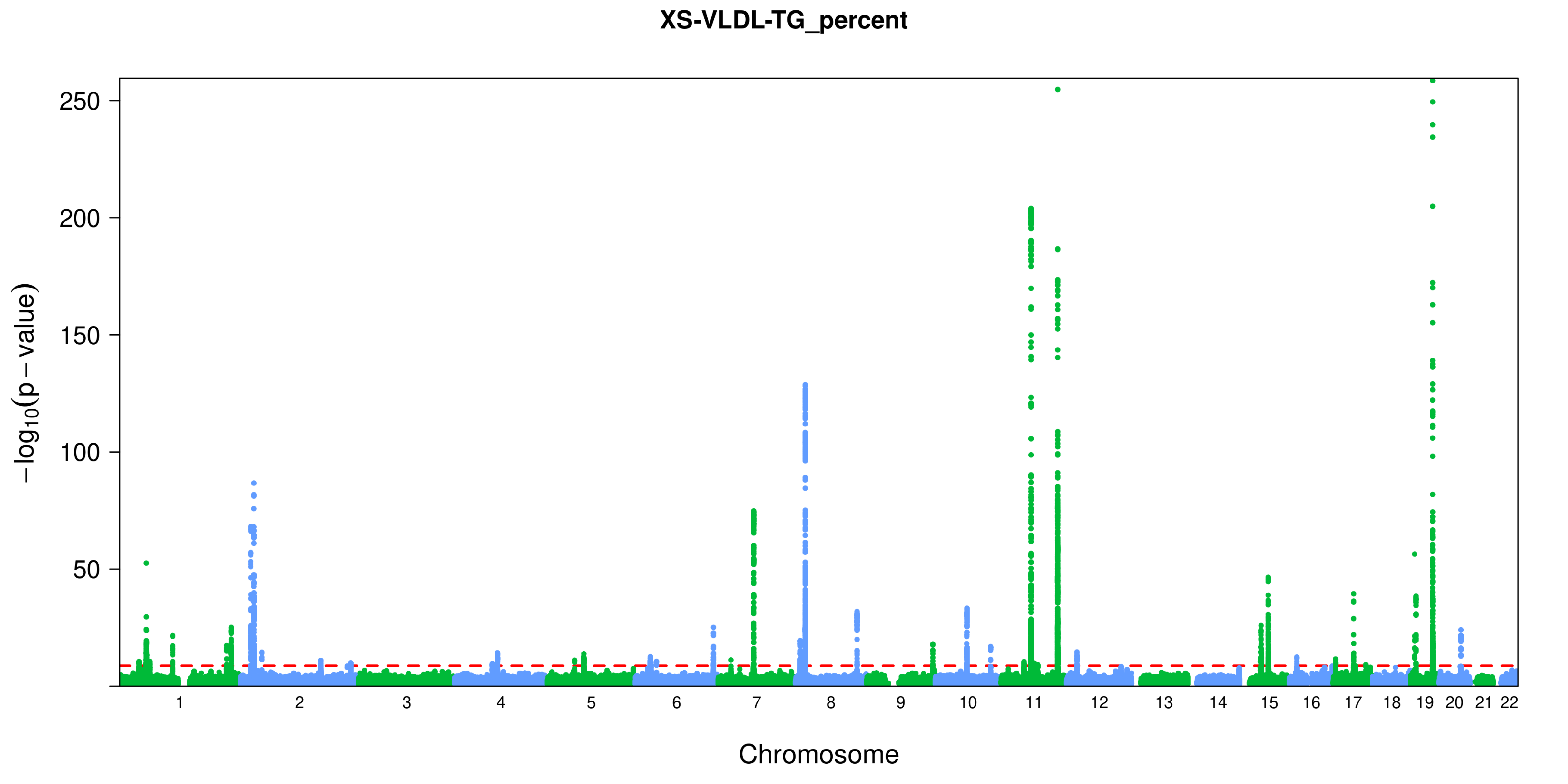

XS-VLDL-TG_percent
250
200
150
100
50
−log10(p − value)
Chromosome

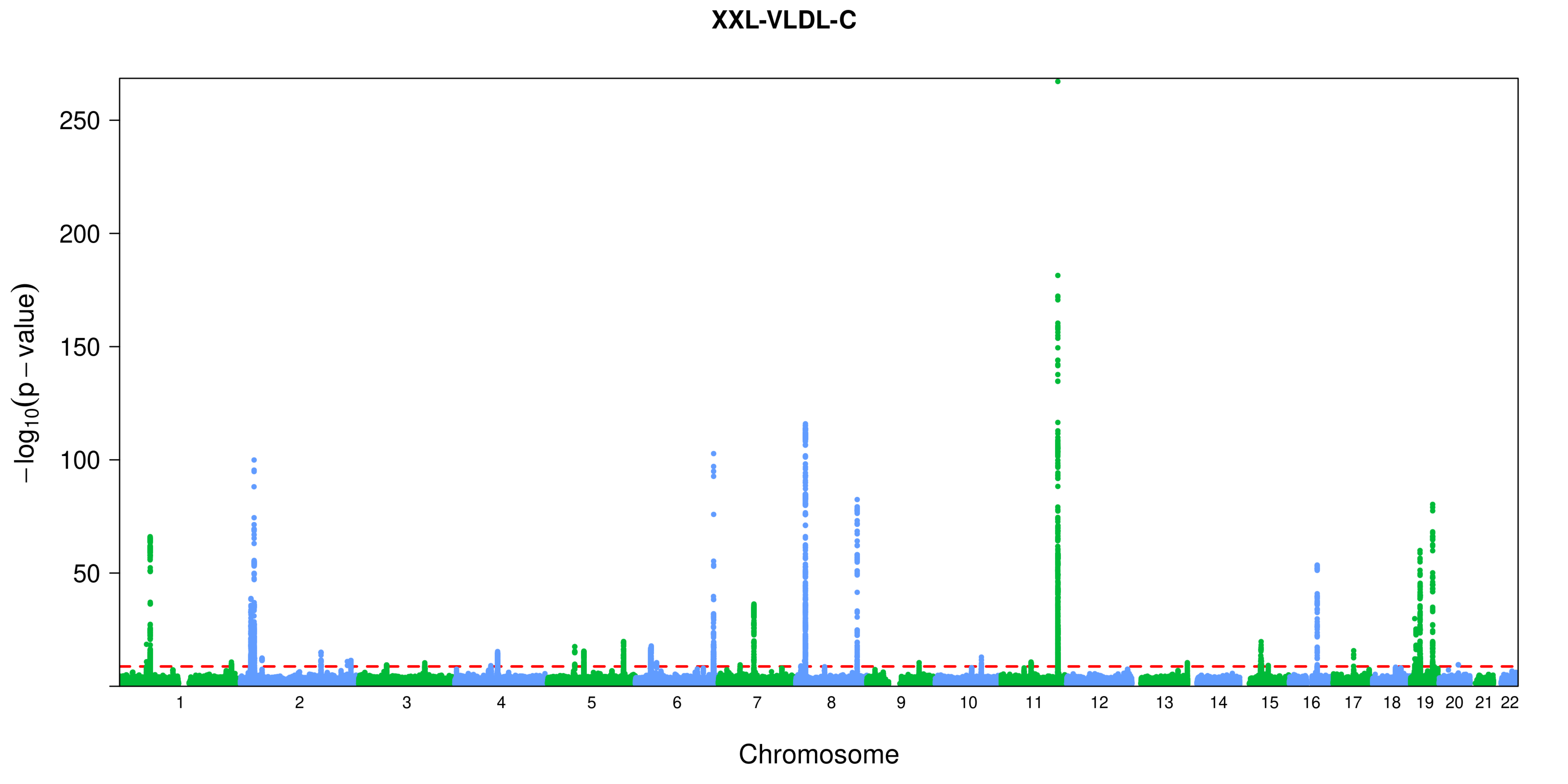
XXL-VLDL-C
250
200
150
100
50
−log10(p − value)
Chromosome

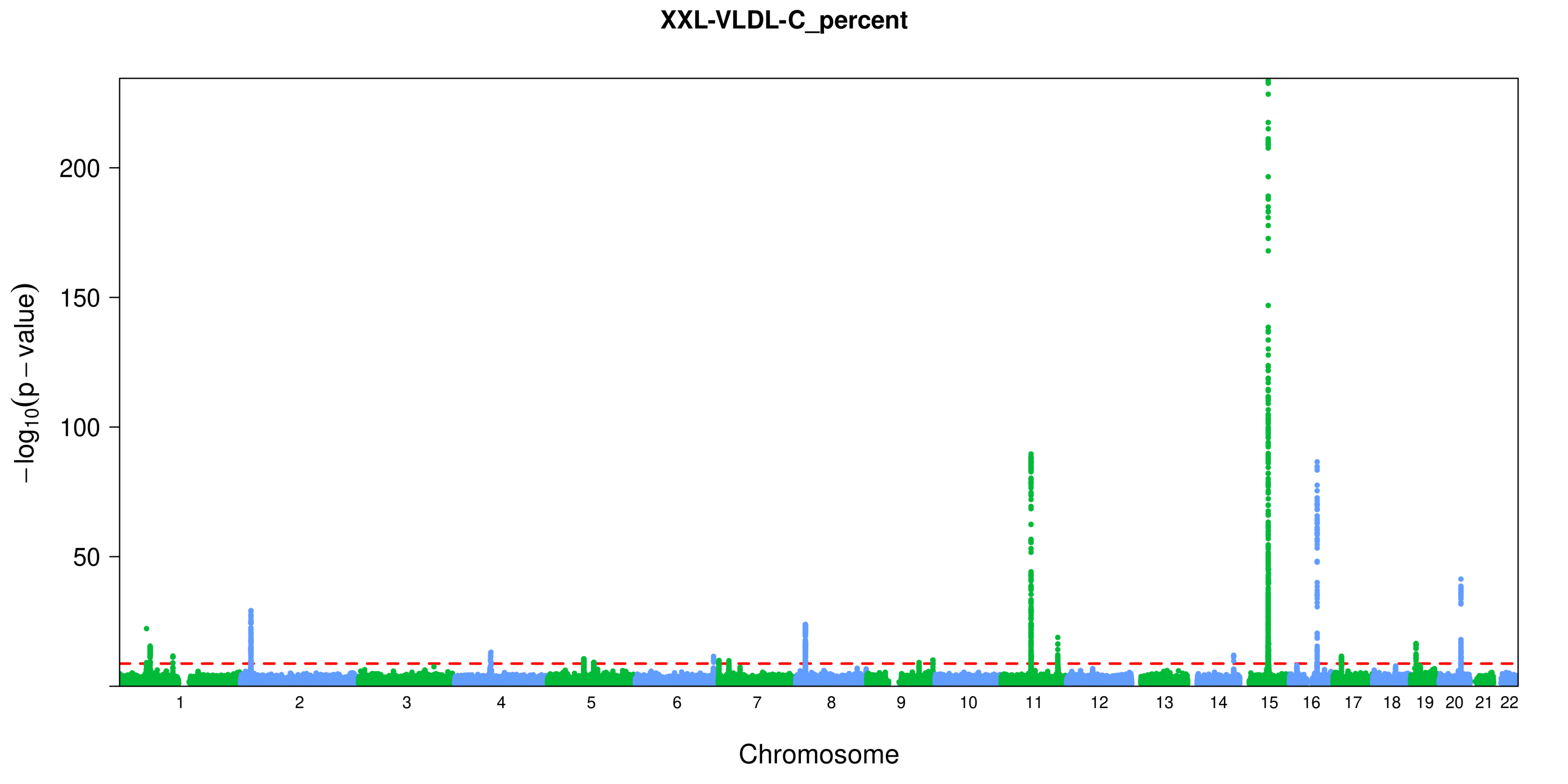
XXL-VLDL-C_percent
200
150
100
50
$-\log_{10}(p-\text{value})$
Chromosome

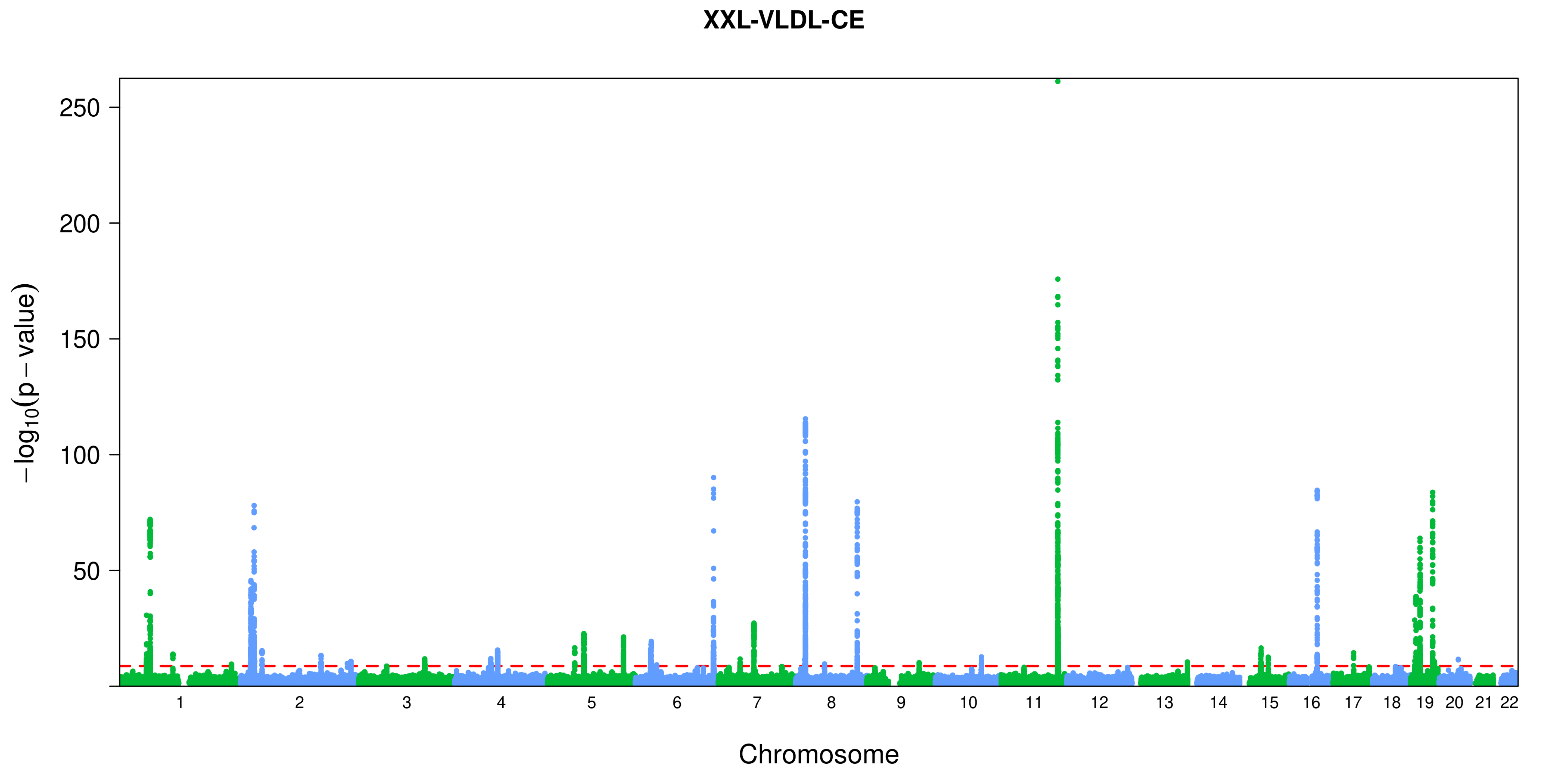

XXL-VLDL-CE
250
200
150
100
50
$-\log_{10}(p-\text{value})$
Chromosome

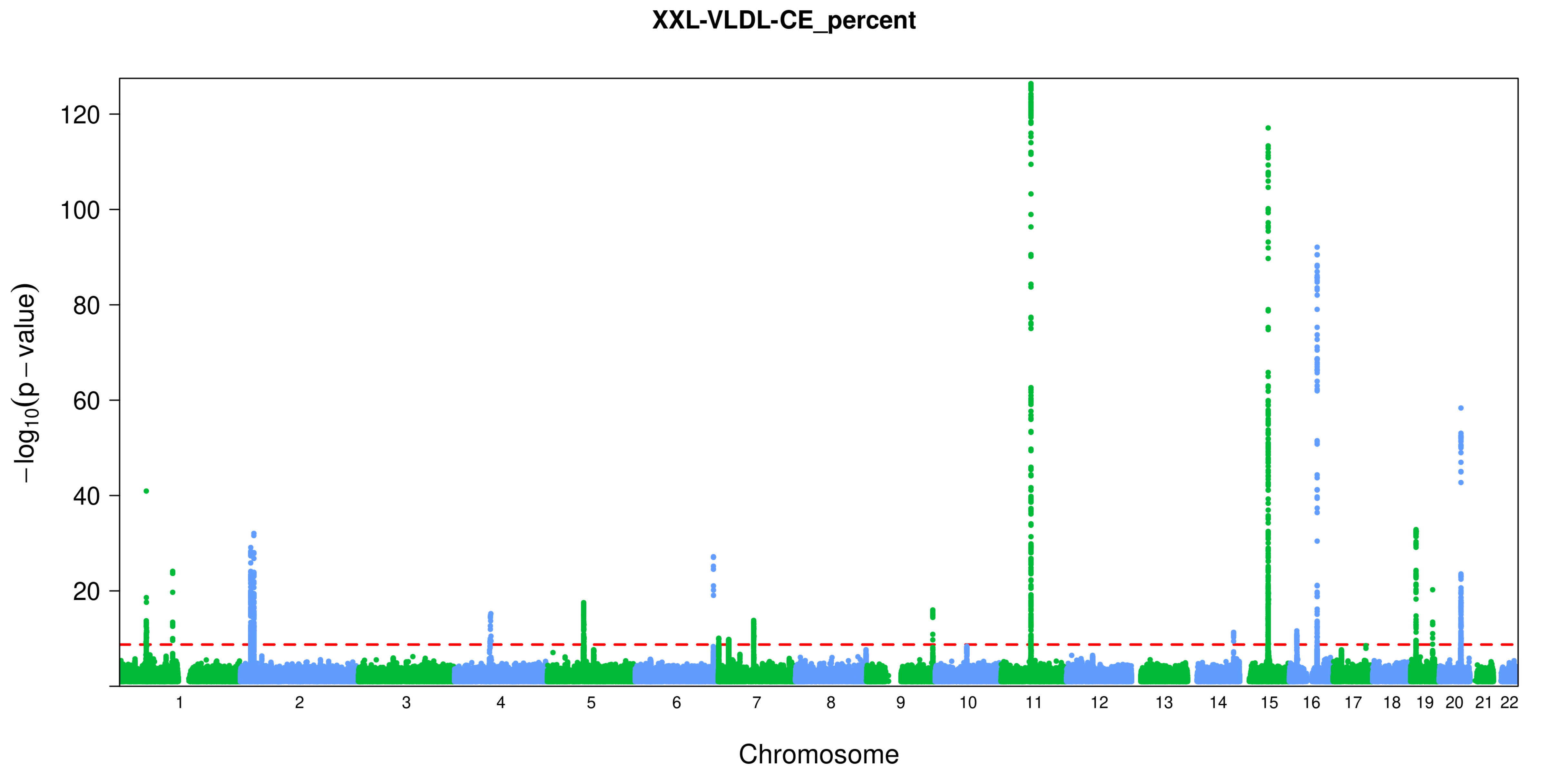
XXL-VLDL-CE_percent
−log10(p − value)
120
100
80
60
40
20
Chromosome

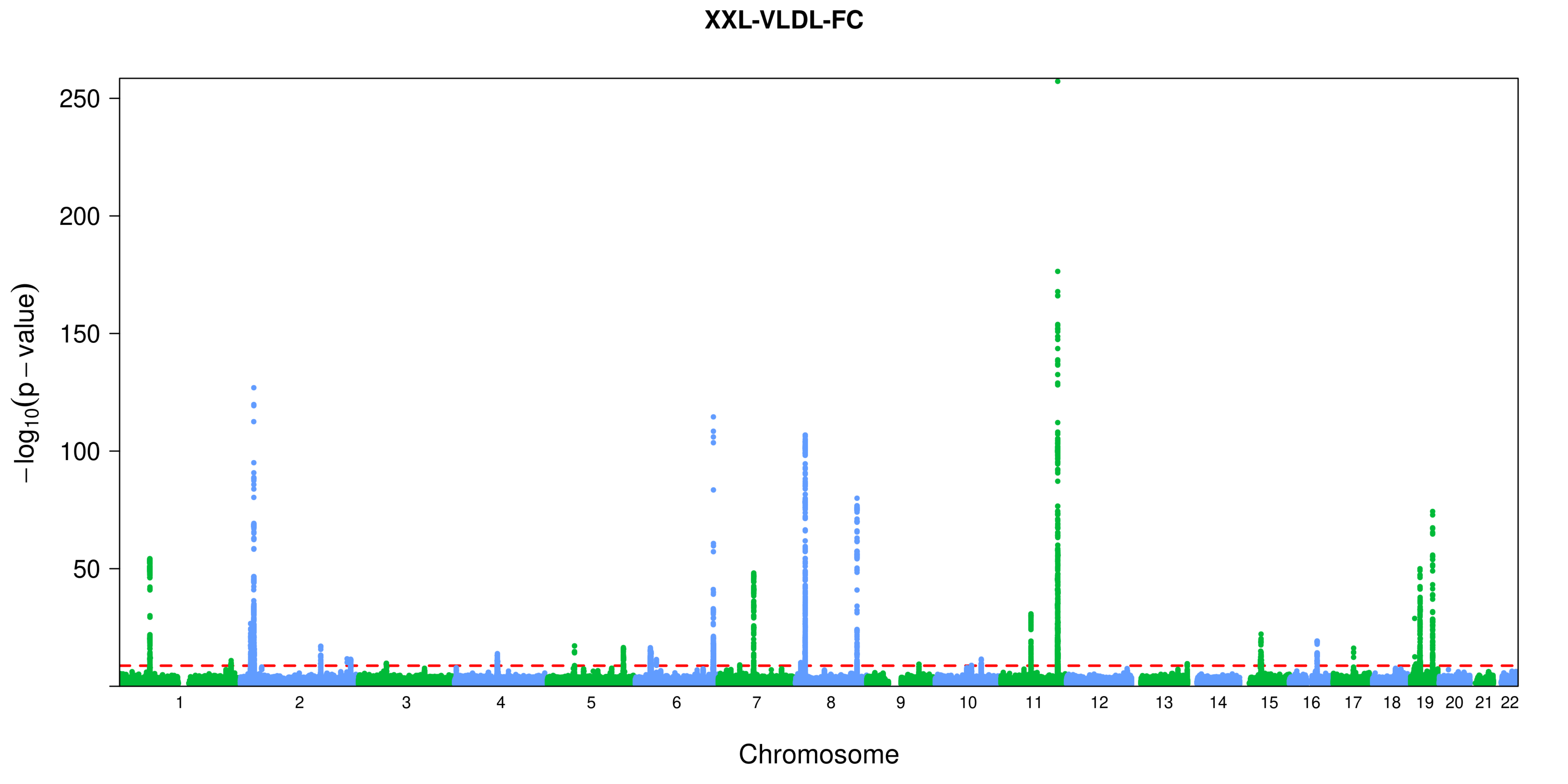

XXL-VLDL-FC
250
200
150
100
50
−log10(p − value)
Chromosome

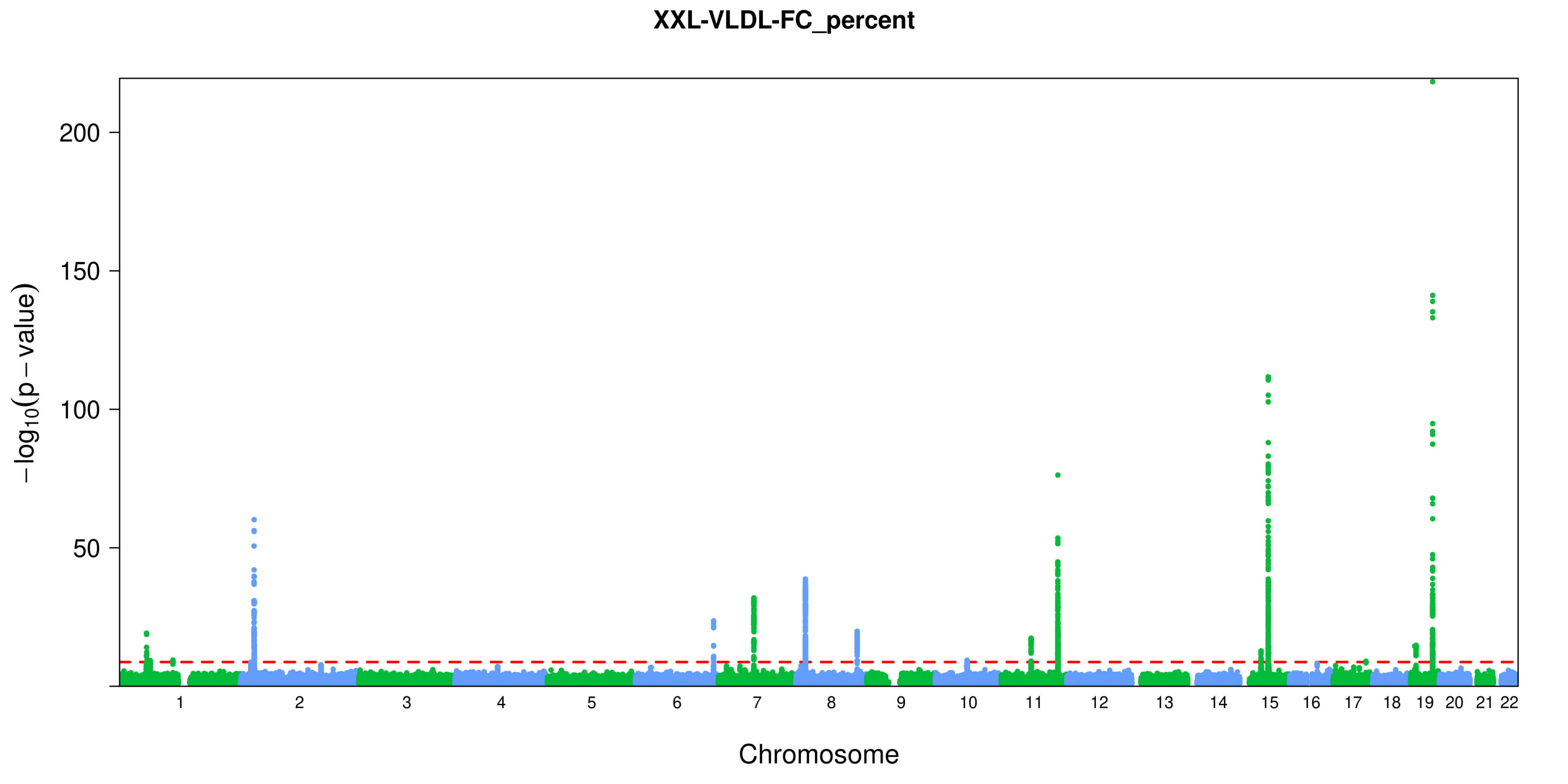

XXL-VLDL-FC_percent
200
150
100
50
$-\log_{10}(p-\text{value})$
1 2 3 4 5 6 7 8 9 10 11 12 13 14 15 16 17 18 19 20 21 22
Chromosome

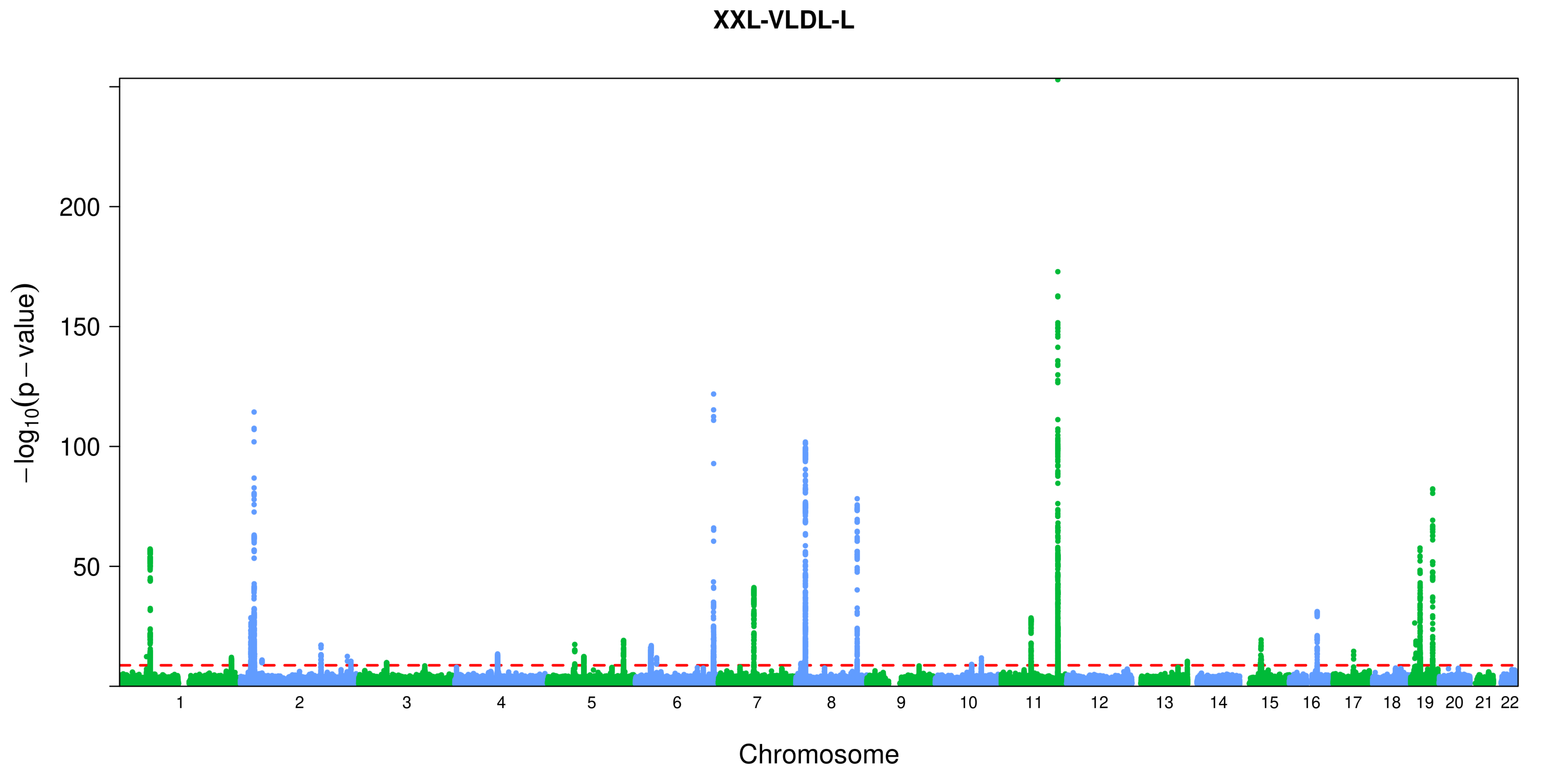
XXL-VLDL-L
−log10(p − value)
200
150
100
50
Chromosome

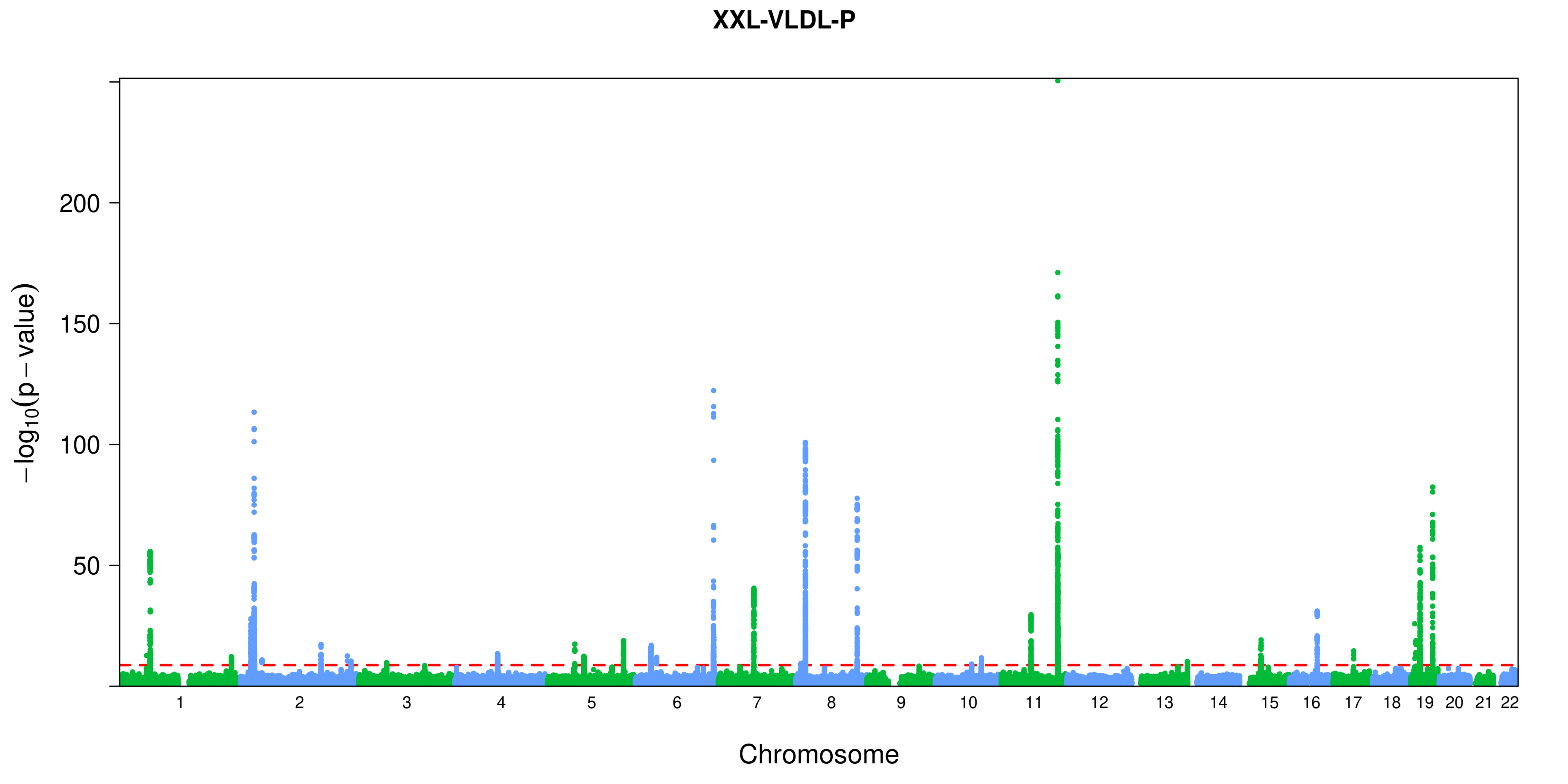
XXL-VLDL-P
$-\log_{10}(p-\text{value})$
200
150
100
50
Chromosome

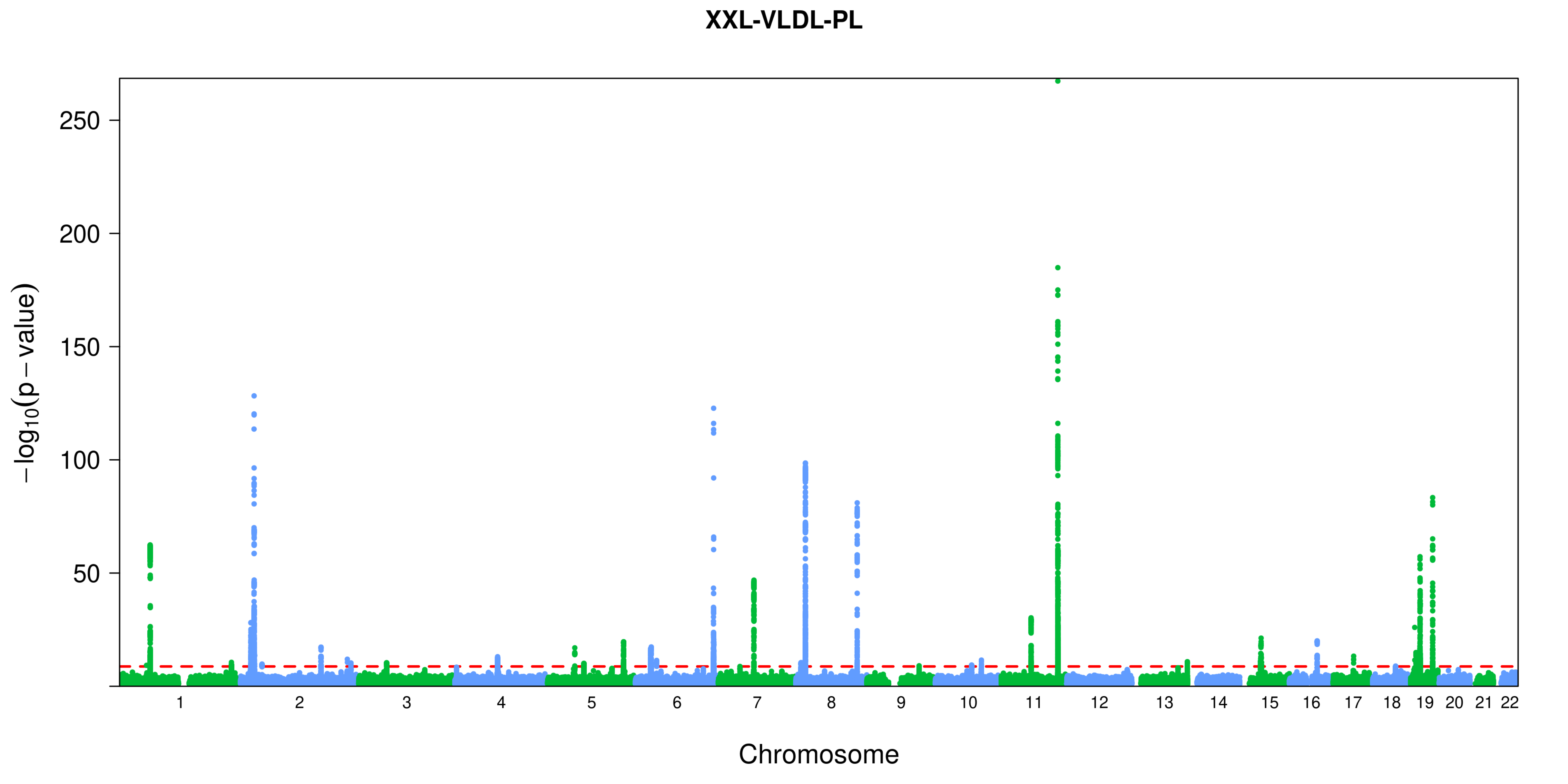
XXL-VLDL-PL
250
200
150
100
50
−log10(p − value)
Chromosome

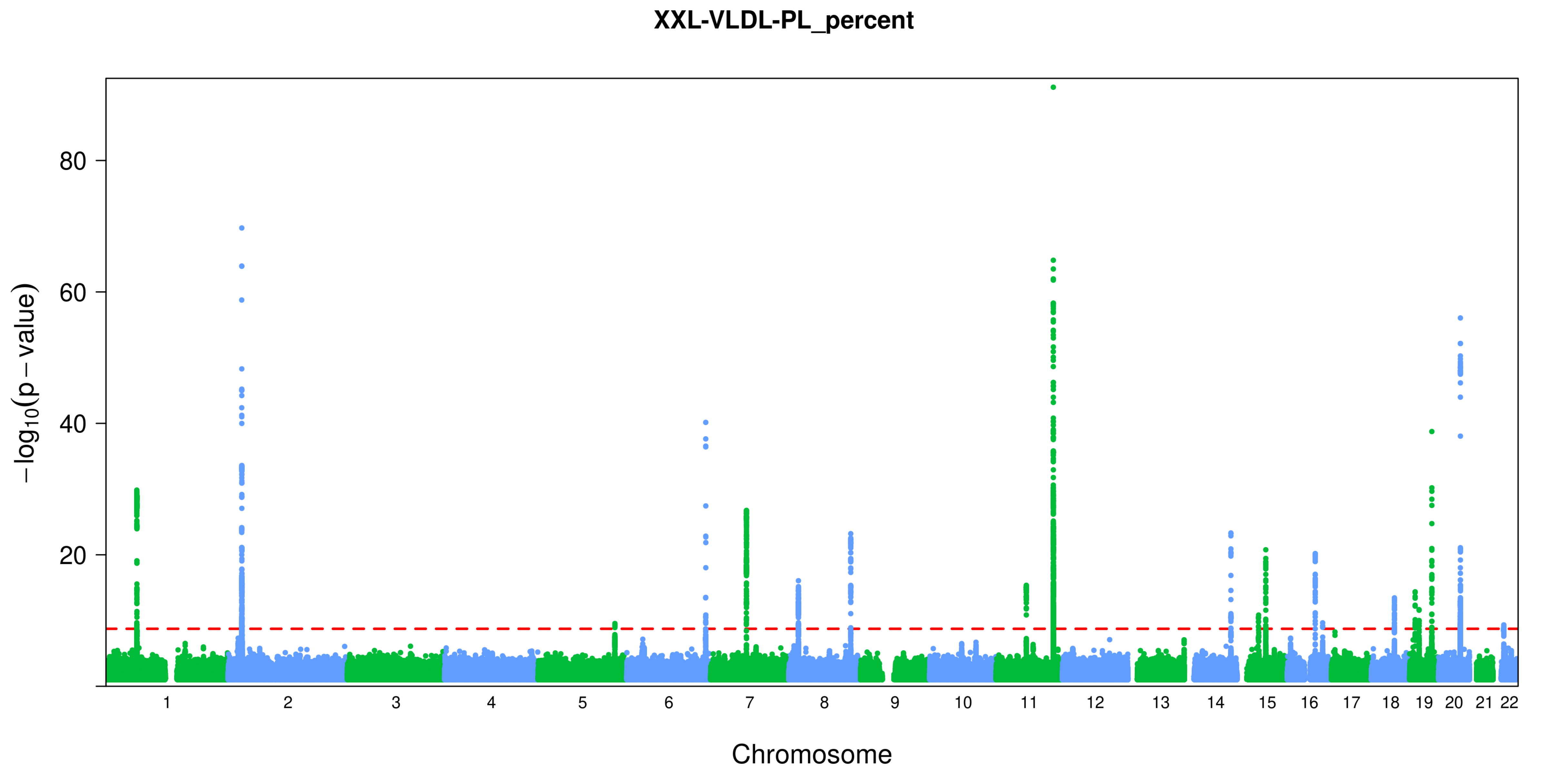
XXL-VLDL-PL_percent
80
60
40
20
−log10(p − value)
Chromosome

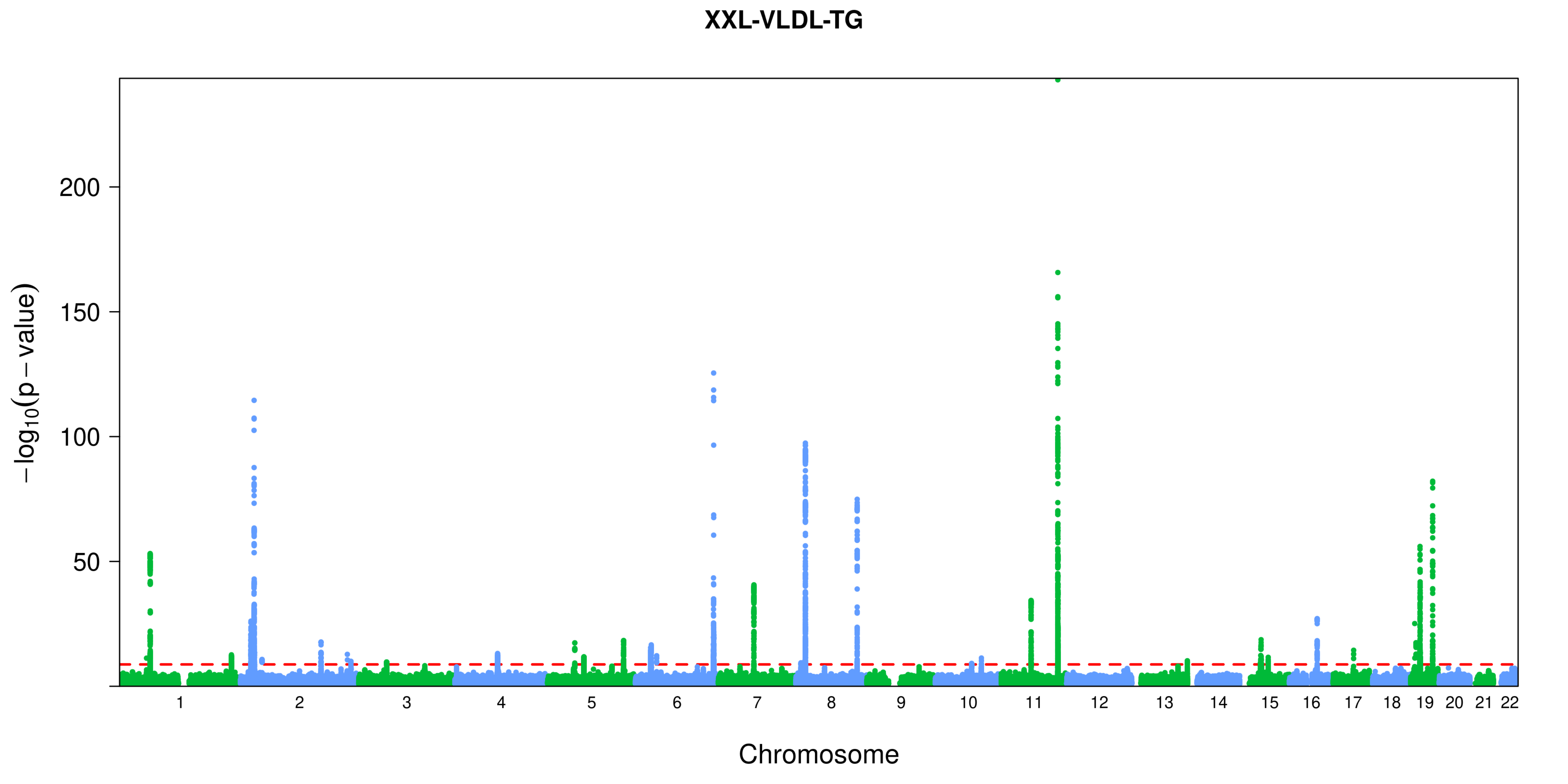
XXL-VLDL-TG
200
150
100
50
$-\log_{10}(p-\text{value})$
Chromosome

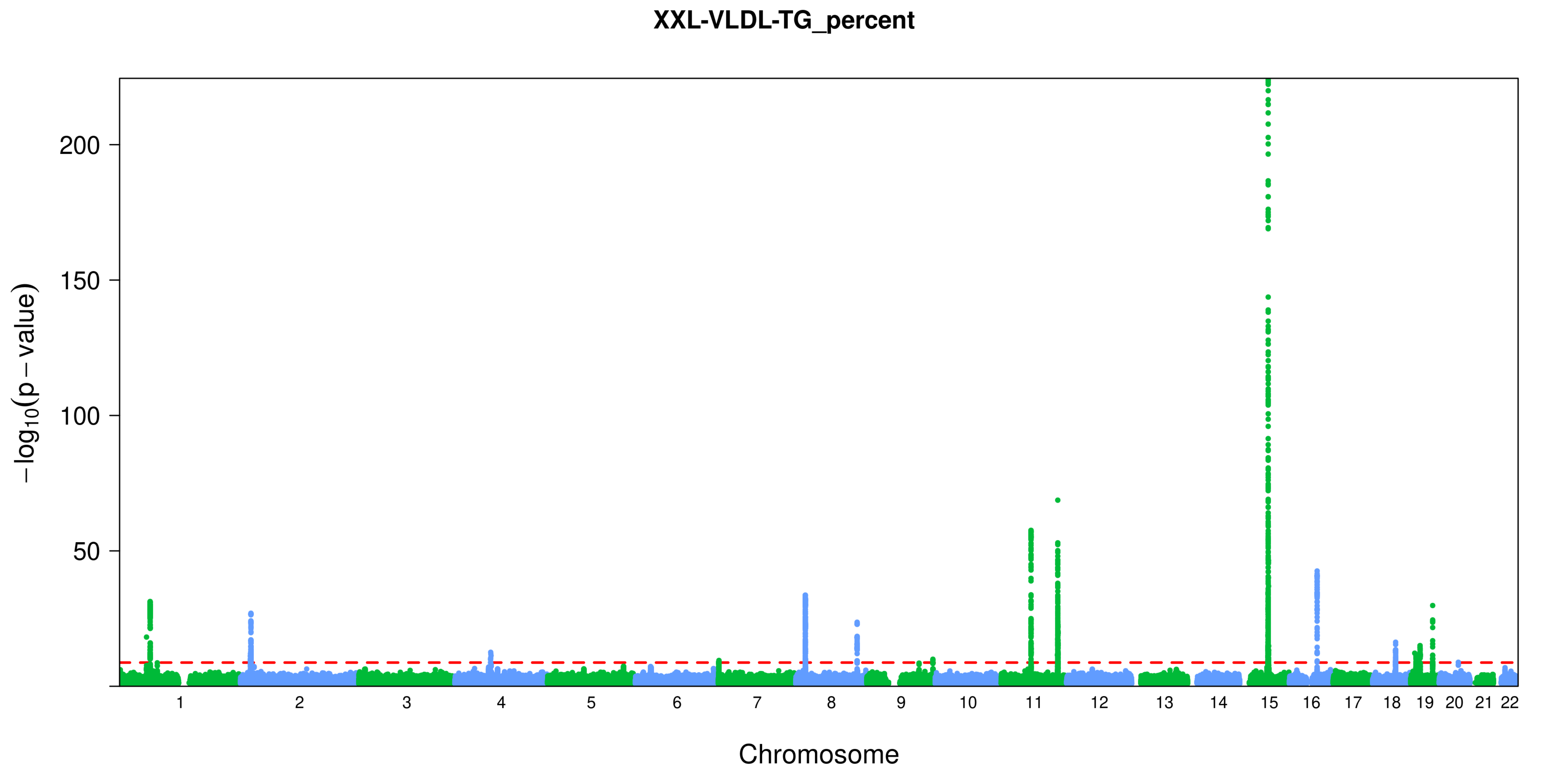
XXL-VLDL-TG_percent
200
150
100
50
−log10(p − value)
Chromosome
