## Supplementary Figure S2 for "Genome-wide characterization of circulating metabolic biomarkers reveals substantial pleiotropy and novel disease pathways"

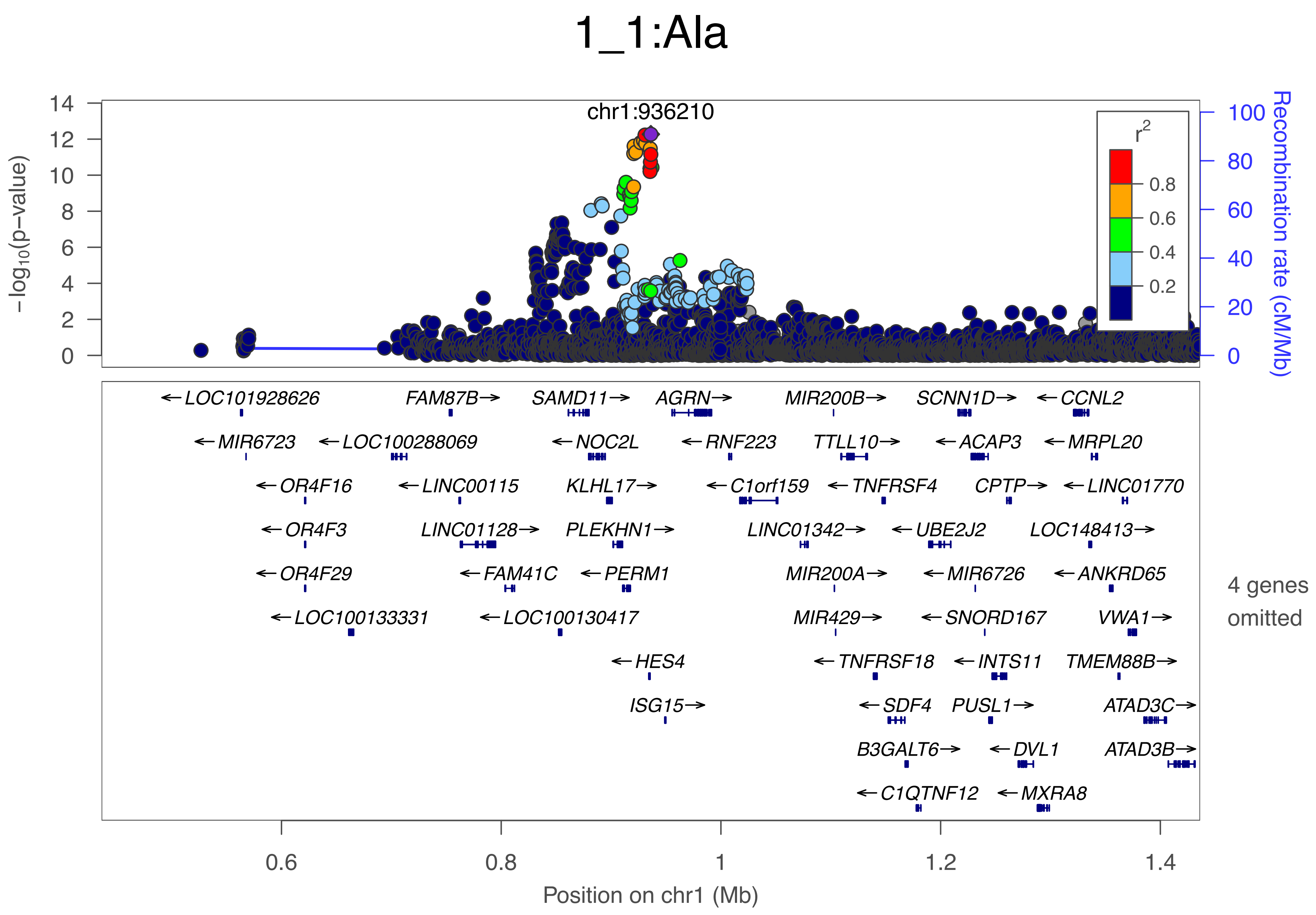
1_1:Ala
chr1:936210
−log10(p−value)
Recombination rate (cM/Mb)
r2
0.8
0.6
0.4
0.2
←LOC101928626
FAM87B→
SAMD11→
AGRN→
MIR200B→
SCNN1D→
←CCNL2
←MIR6723
←LOC100288069
←NOC2L
←RNF223
TTLL10→
←ACAP3
←MRPL20
←OR4F16
←LINC00115
KLHL17→
←C1orf159
←TNFRSF4
CPTP→
←LINC01770
←OR4F3
LINC01128→
PLEKHN1→
LINC01342→
←UBE2J2
LOC148413→
←OR4F29
←FAM41C
←PERM1
MIR200A→
←MIR6726
←ANKRD65
←LOC100133331
←LOC100130417
MIR429→
←SNORD167
VWA1→
←HES4
←TNFRSF18
←INTS11
TMEM88B→
ISG15→
←SDF4
PUSL1→
ATAD3C→
B3GALT6→
←DVL1
ATAD3B→
←C1QTNF12
←MXRA8
4 genes omitted
Position on chr1 (Mb)

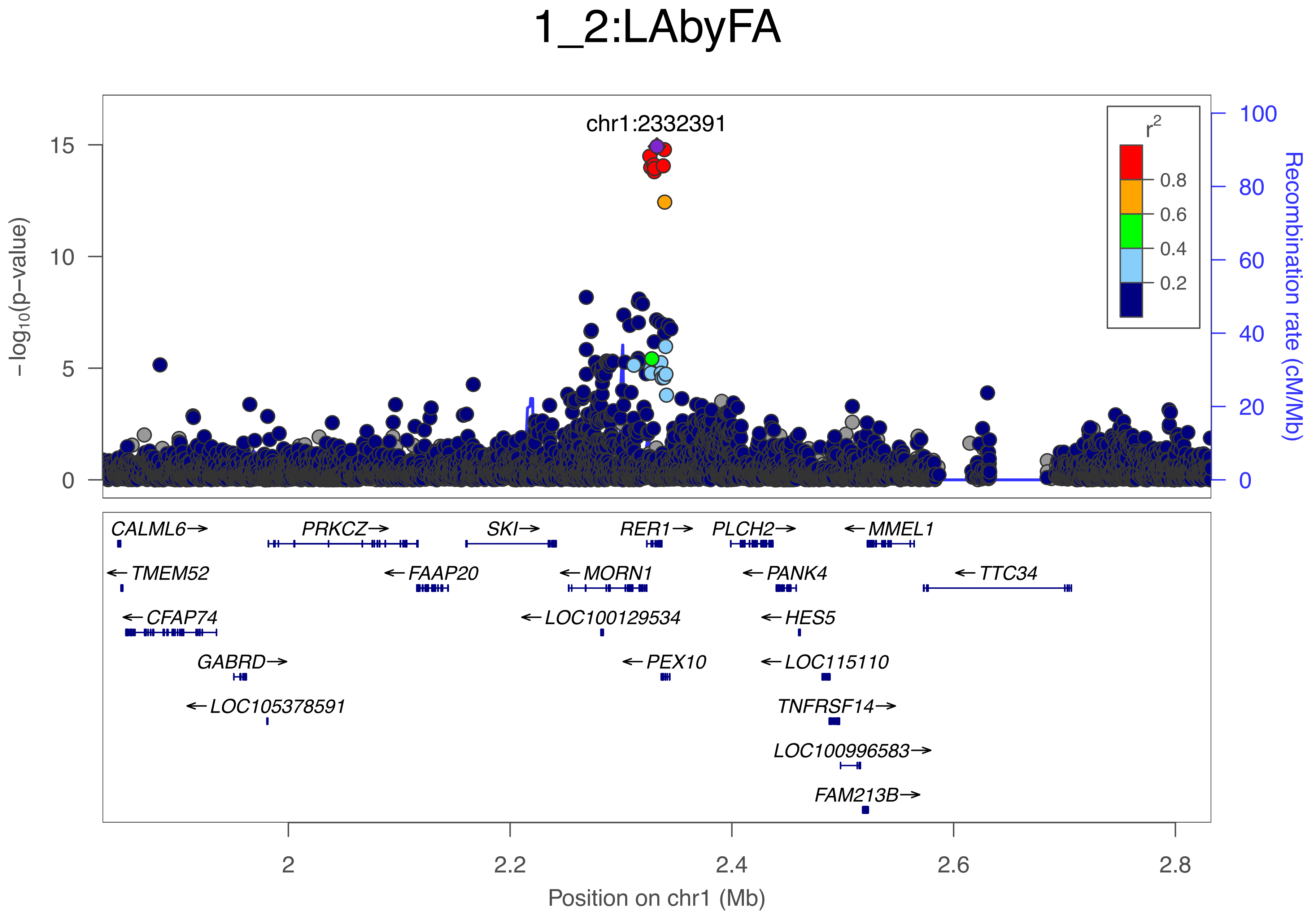
1_2:LAbyFA
chr1:2332391
r2
0.8
0.6
0.4
0.2
−log10(p−value)
Recombination rate (cM/Mb)
15
10
5
0
100
80
60
40
20
0
CALML6→
PRKCZ→
SKI→
RER1→
PLCH2→
←MMEL1
←TMEM52
←FAAP20
←MORN1
←PANK4
←TTC34
←CFAP74
←LOC100129534
←HES5
GABRD→
←PEX10
←LOC115110
←LOC105378591
TNFRSF14→
LOC100996583→
FAM213B→
2
2.2
2.4
2.6
2.8
Position on chr1 (Mb)

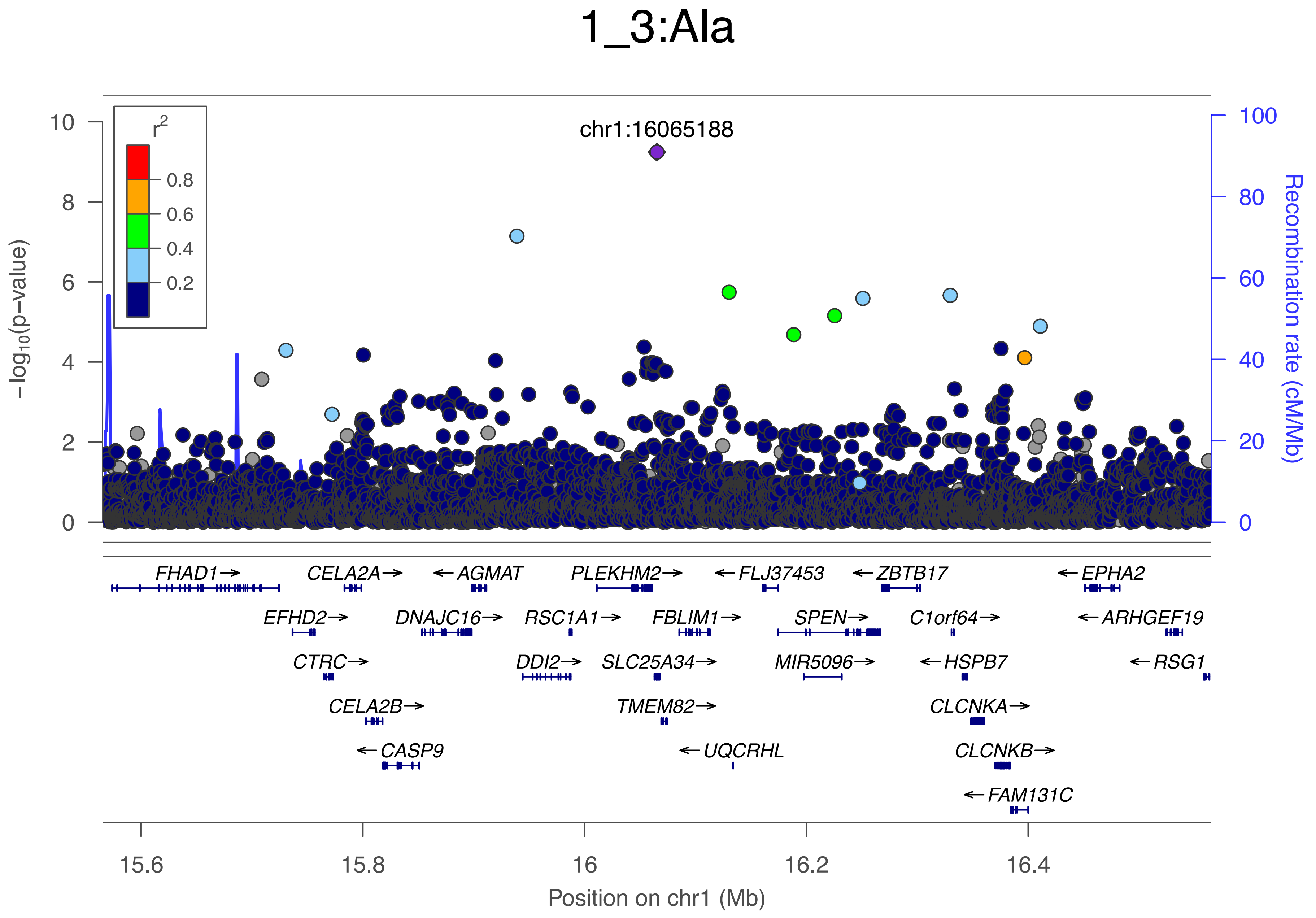
1_3:Ala
chr1:16065188
r2
0.8
0.6
0.4
0.2
-log10(p-value)
Recombination rate (cM/Mb)
FHAD1→
CELA2A→
←AGMAT
PLEKHM2→
←FLJ37453
←ZBTB17
←EPHA2
EFHD2→
DNAJC16→
RSC1A1→
FBLIM1→
SPEN→
C1orf64→
←ARHGEF19
CTRC→
DDI2→
SLC25A34→
MIR5096→
←HSPB7
←RSG1
CELA2B→
TMEM82→
CLCNKA→
←CASP9
←UQCRHL
CLCNKB→
←FAM131C
15.6
15.8
16
16.2
16.4
Position on chr1 (Mb)

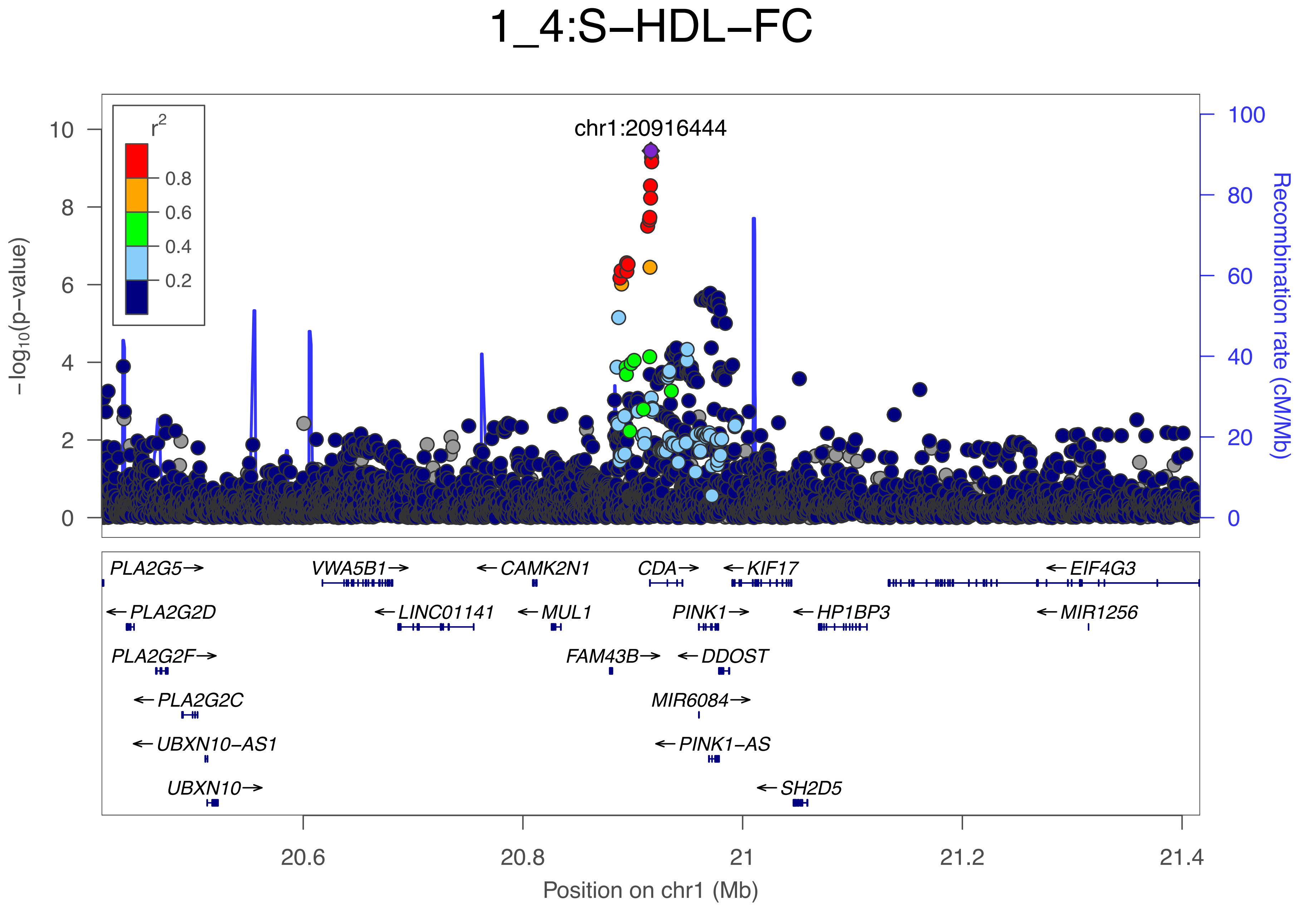

1_4:S−HDL−FC
chr1:20916444
r2
0.8
0.6
0.4
0.2
−log10(p−value)
Recombination rate (cM/Mb)
PLA2G5→
VWA5B1→
←CAMK2N1
CDA→
←KIF17
←EIF4G3
←PLA2G2D
←LINC01141
←MUL1
PINK1→
←HP1BP3
←MIR1256
PLA2G2F→
FAM43B→
←DDOST
←PLA2G2C
MIR6084→
←UBXN10−AS1
←PINK1−AS
UBXN10→
←SH2D5
Position on chr1 (Mb)

### 1_5:Crea

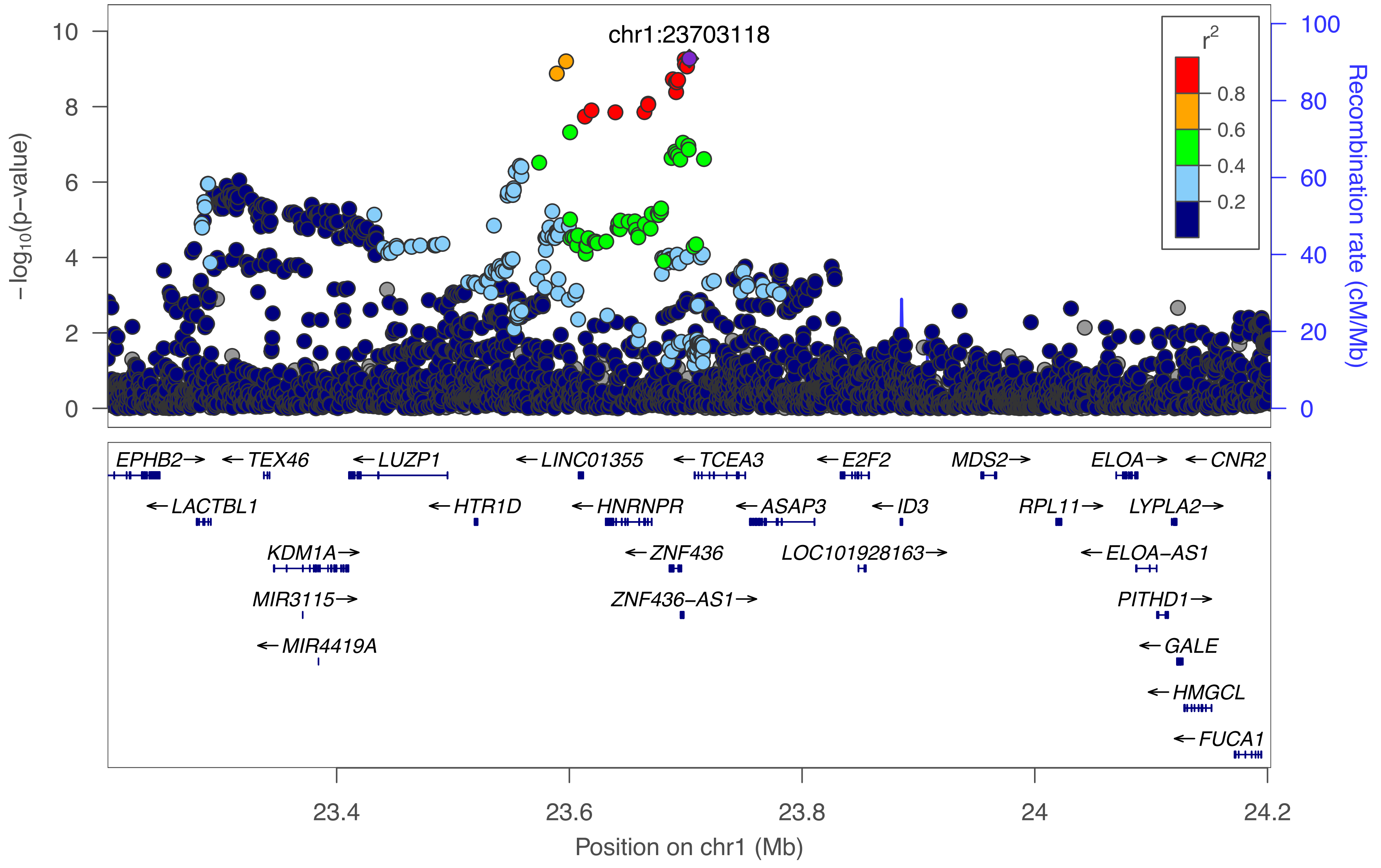
chr1:23703118
r2
0.8
0.6
0.4
0.2
−log10(p−value)
Recombination rate (cM/Mb)
10
8
6
4
2
0
100
80
60
40
20
0
EPHB2→
←TEX46
←LUZP1
←LINC01355
←TCEA3
←E2F2
MDS2→
ELOA→
←CNR2
←LACTBL1
←HTR1D
←HNRNPR
←ASAP3
←ID3
RPL11→
LYPLA2→
KDM1A→
←ZNF436
LOC101928163→
←ELOA−AS1
MIR3115→
ZNF436−AS1→
PITHD1→
←MIR4419A
←GALE
←HMGCL
←FUCA1
23.4
23.6
23.8
24
24.2
Position on chr1 (Mb)

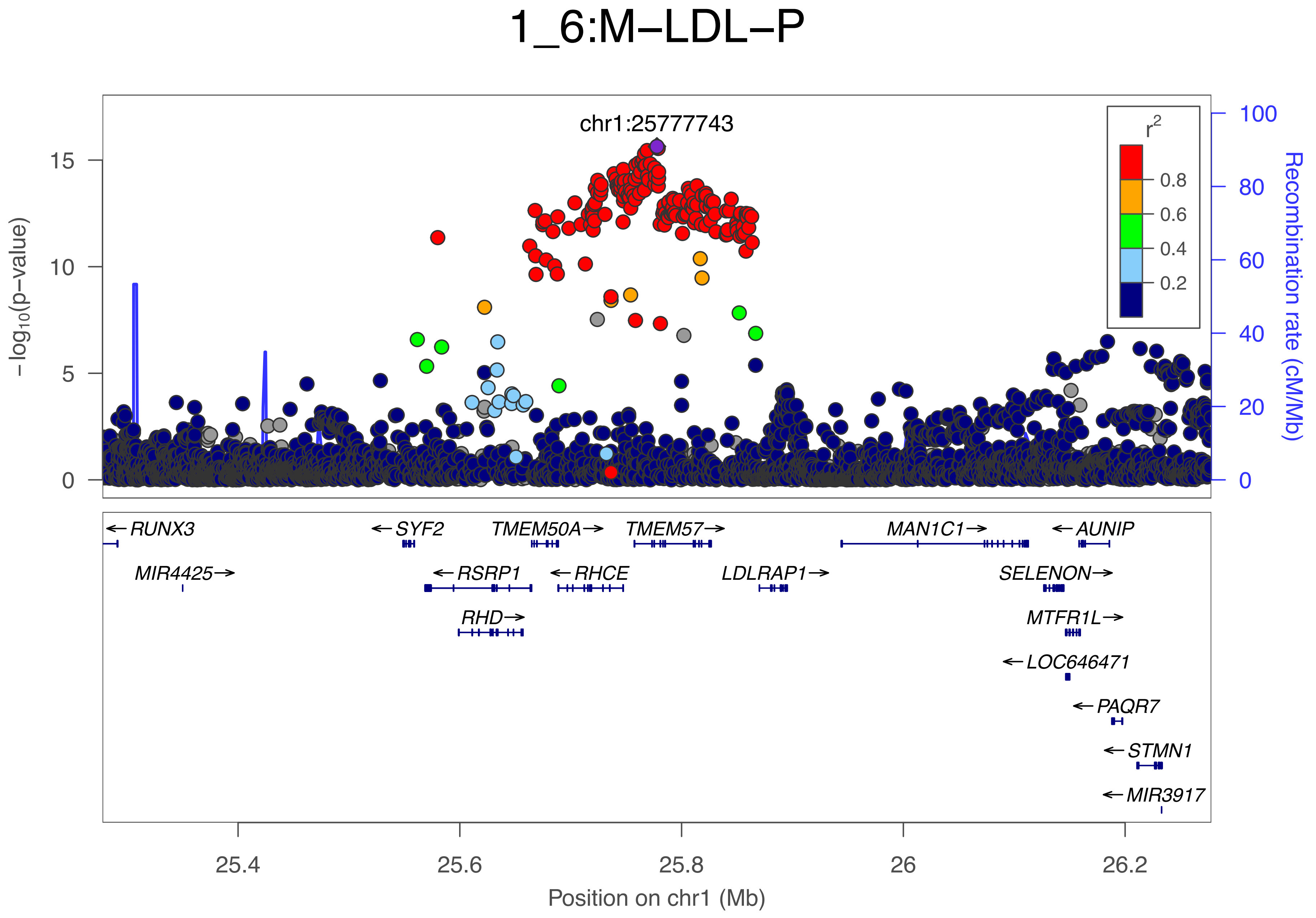
1_6:M–LDL–P
chr1:25777743
−log10(p−value)
Recombination rate (cM/Mb)
r2
0.8
0.6
0.4
0.2
15
10
5
0
100
80
60
40
20
0
RUNX3
SYF2
TMEM50A
TMEM57
MAN1C1
AUNIP
MIR4425
RSRP1
RHCE
LDLRAP1
SELENON
RHD
MTFR1L
LOC646471
PAQR7
STMN1
MIR3917
25.4
25.6
25.8
26
26.2
Position on chr1 (Mb)

### 1_7:ApoBbyApoA1

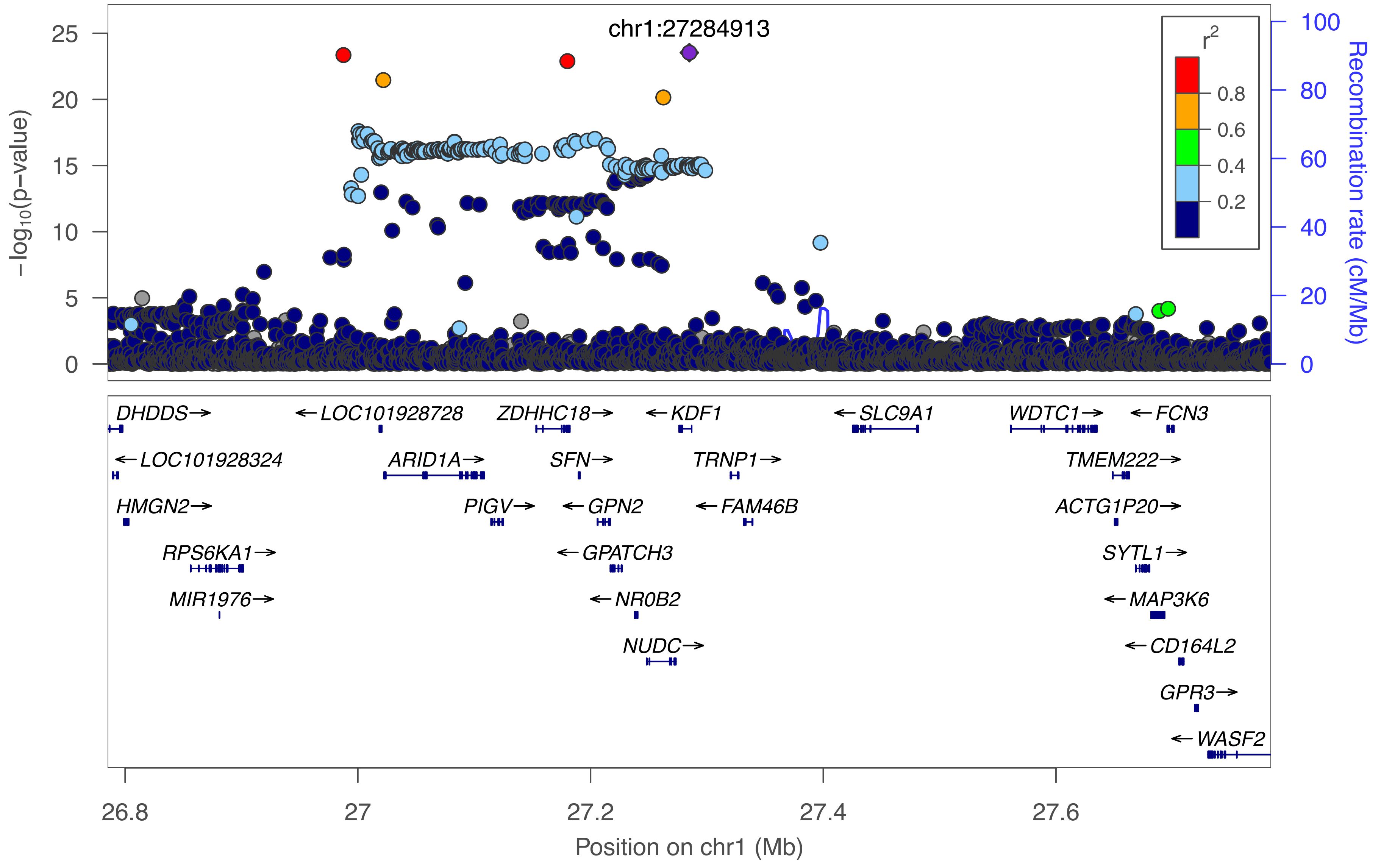

### 1_8:HDL2−C

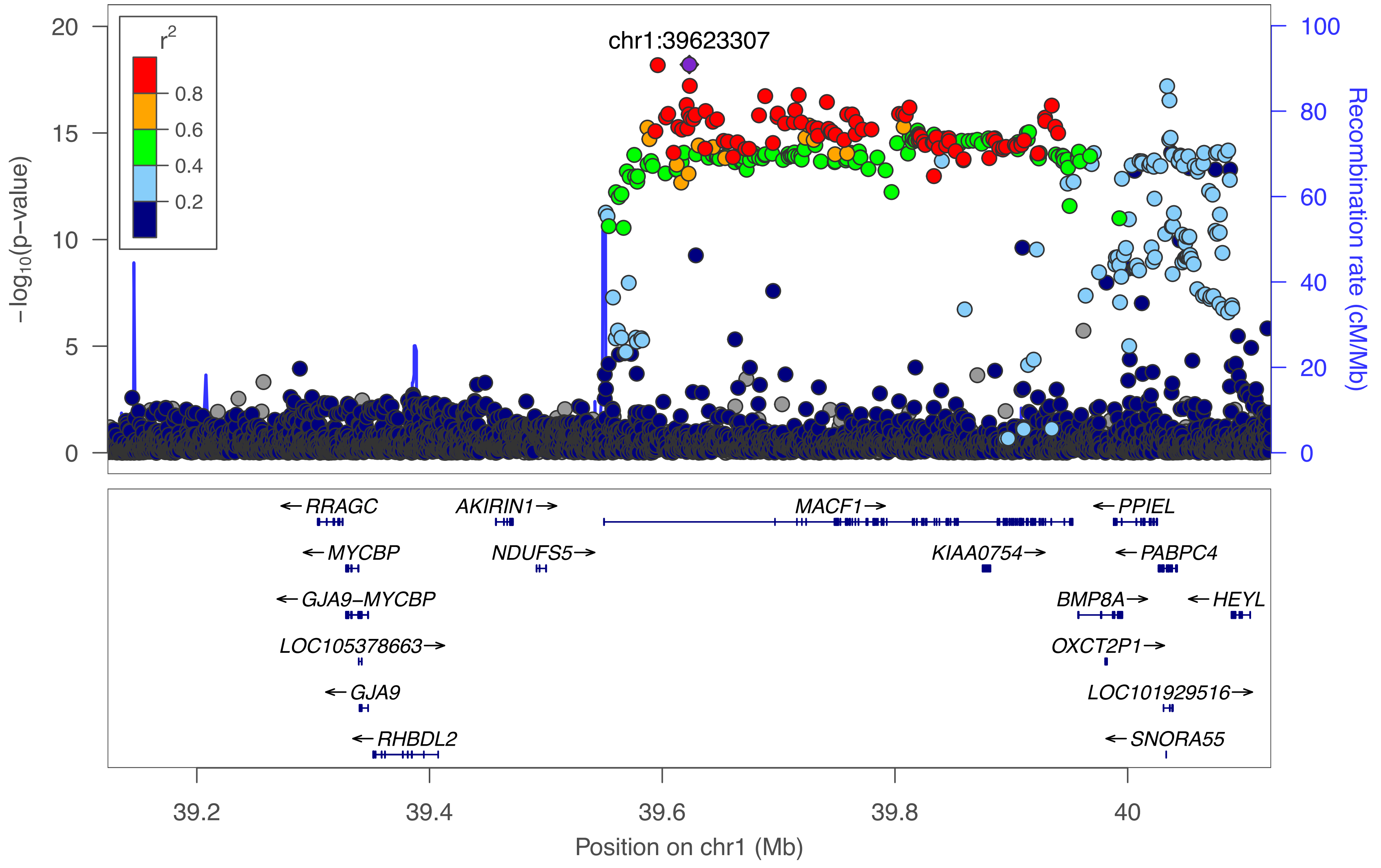

chr1:39623307
r2
0.8
0.6
0.4
0.2
−log10(p−value)
Recombination rate (cM/Mb)
RRAGC
AKIRIN1
MACF1
PPIEL
MYCBP
NDUFS5
KIAA0754
PABPC4
GJA9−MYCBP
BMP8A
HEYL
LOC105378663
OXCT2P1
GJA9
LOC101929516
RHBDL2
SNORA55
39.2
39.4
39.6
39.8
40
Position on chr1 (Mb)

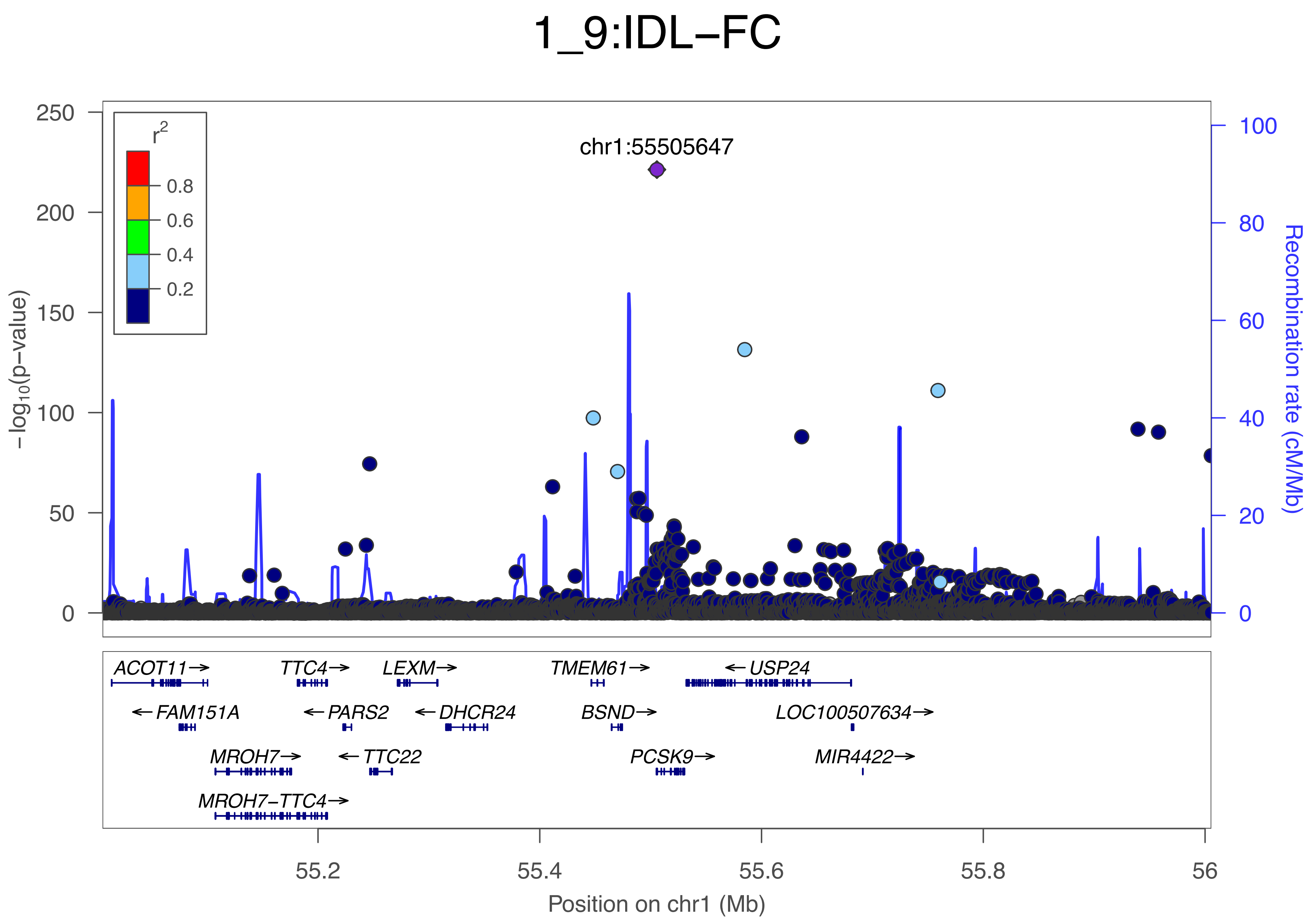
1_9:IDL−FC
chr1:55505647
r2
0.8
0.6
0.4
0.2
−log10(p−value)
0
50
100
150
200
250
Recombination rate (cM/Mb)
0
20
40
60
80
100
ACOT11→
TTC4→
LEXM→
TMEM61→
←USP24
←FAM151A
←PARS2
←DHCR24
BSND→
LOC100507634→
MROH7→
←TTC22
PCSK9→
MIR4422→
MROH7−TTC4→
55.2
55.4
55.6
55.8
56
Position on chr1 (Mb)

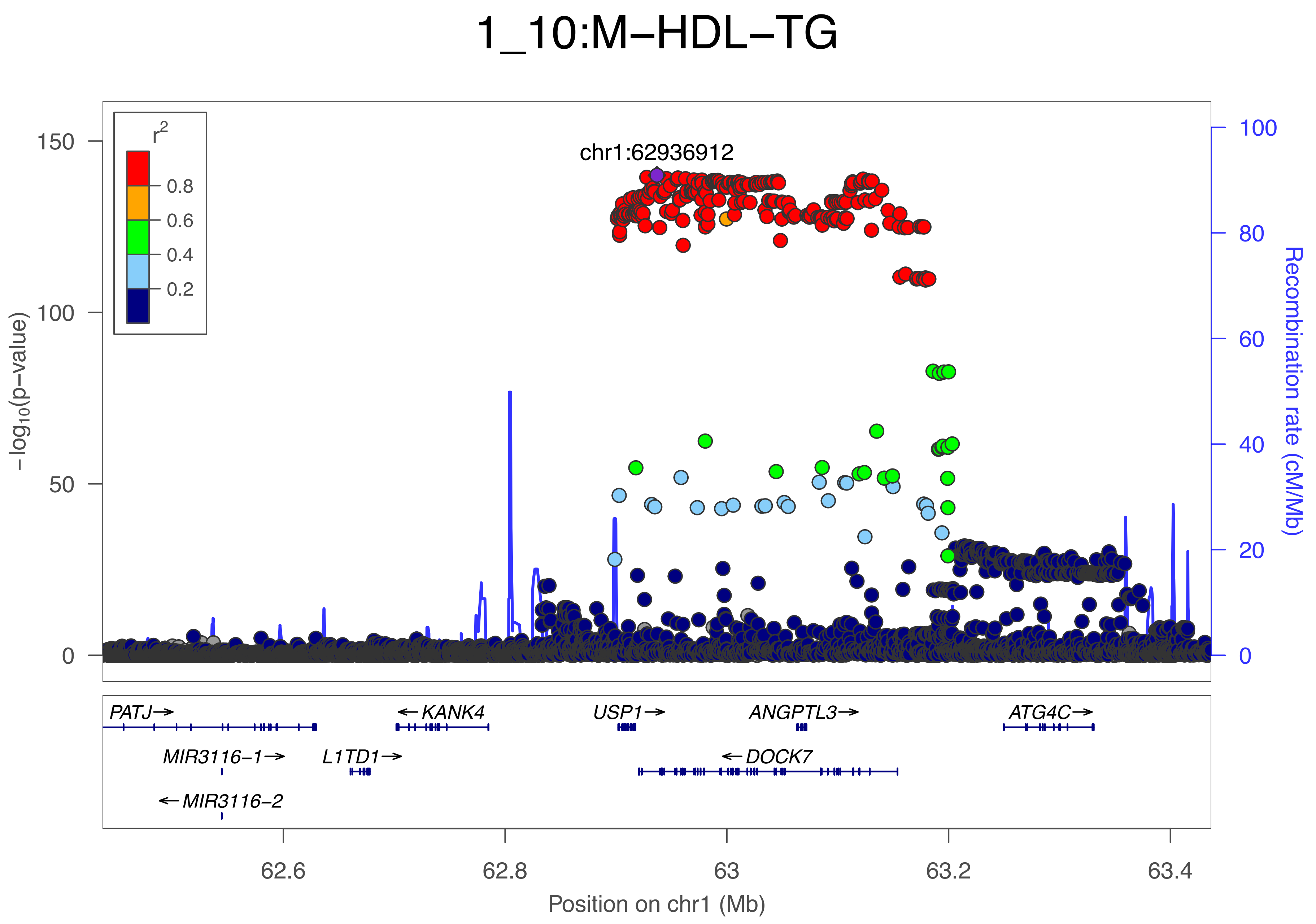

1_10:M−HDL−TG
chr1:62936912
r2
0.8
0.6
0.4
0.2
150
100
50
0
−log10(p−value)
100
80
60
40
20
0
Recombination rate (cM/Mb)
PATJ→
←KANK4
USP1→
ANGPTL3→
ATG4C→
MIR3116−1→
L1TD1→
←DOCK7
←MIR3116−2
62.6
62.8
63
63.2
63.4
Position on chr1 (Mb)

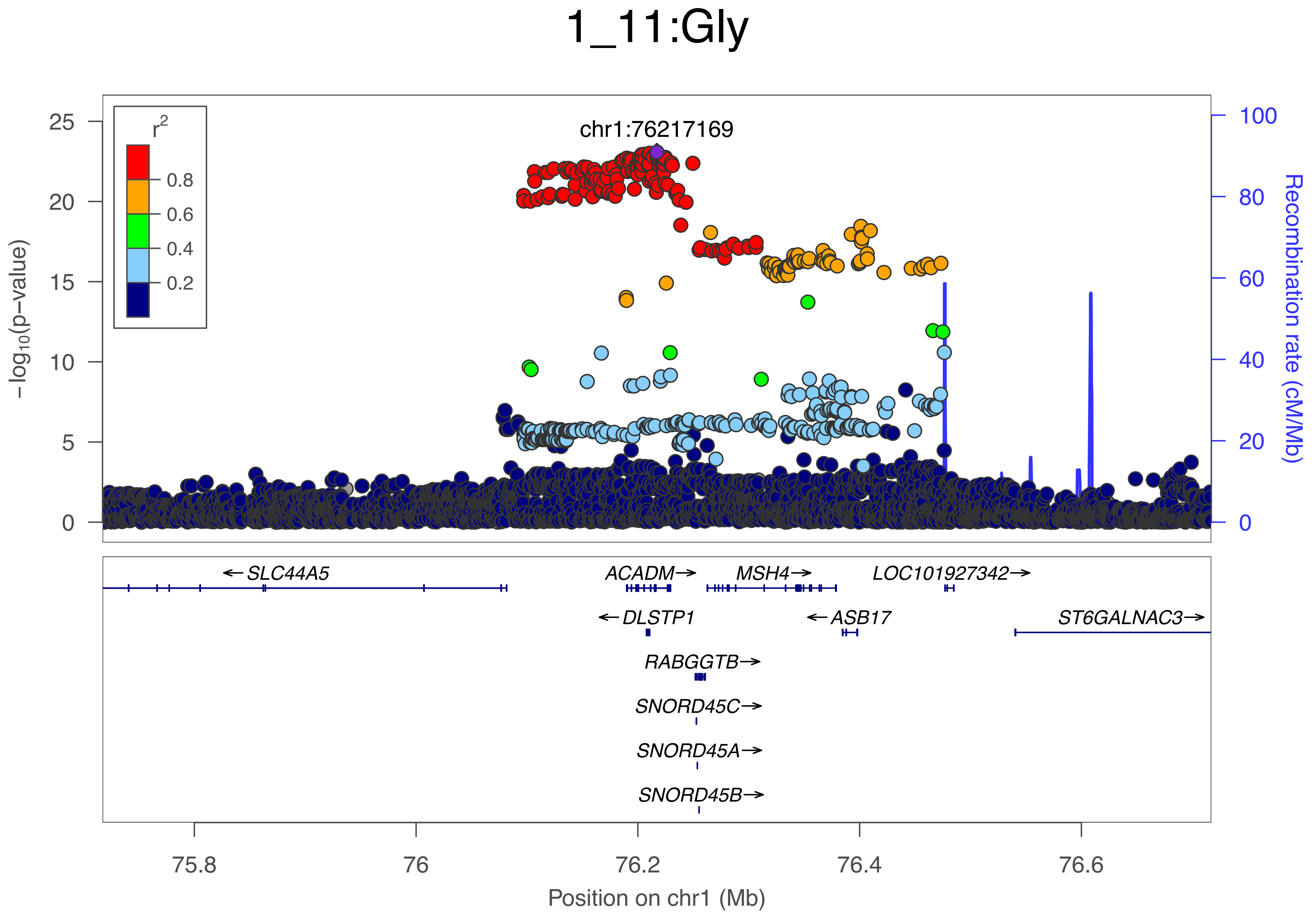
1_11:Gly
chr1:76217169
r2
0.8
0.6
0.4
0.2
−log10(p−value)
Recombination rate (cM/Mb)
SLC44A5
ACADM
MSH4
LOC101927342
DLSTP1
ASB17
ST6GALNAC3
RABGGTB
SNORD45C
SNORD45A
SNORD45B
75.8
76
76.2
76.4
76.6
Position on chr1 (Mb)

### 1_12:XL−HDL−FC

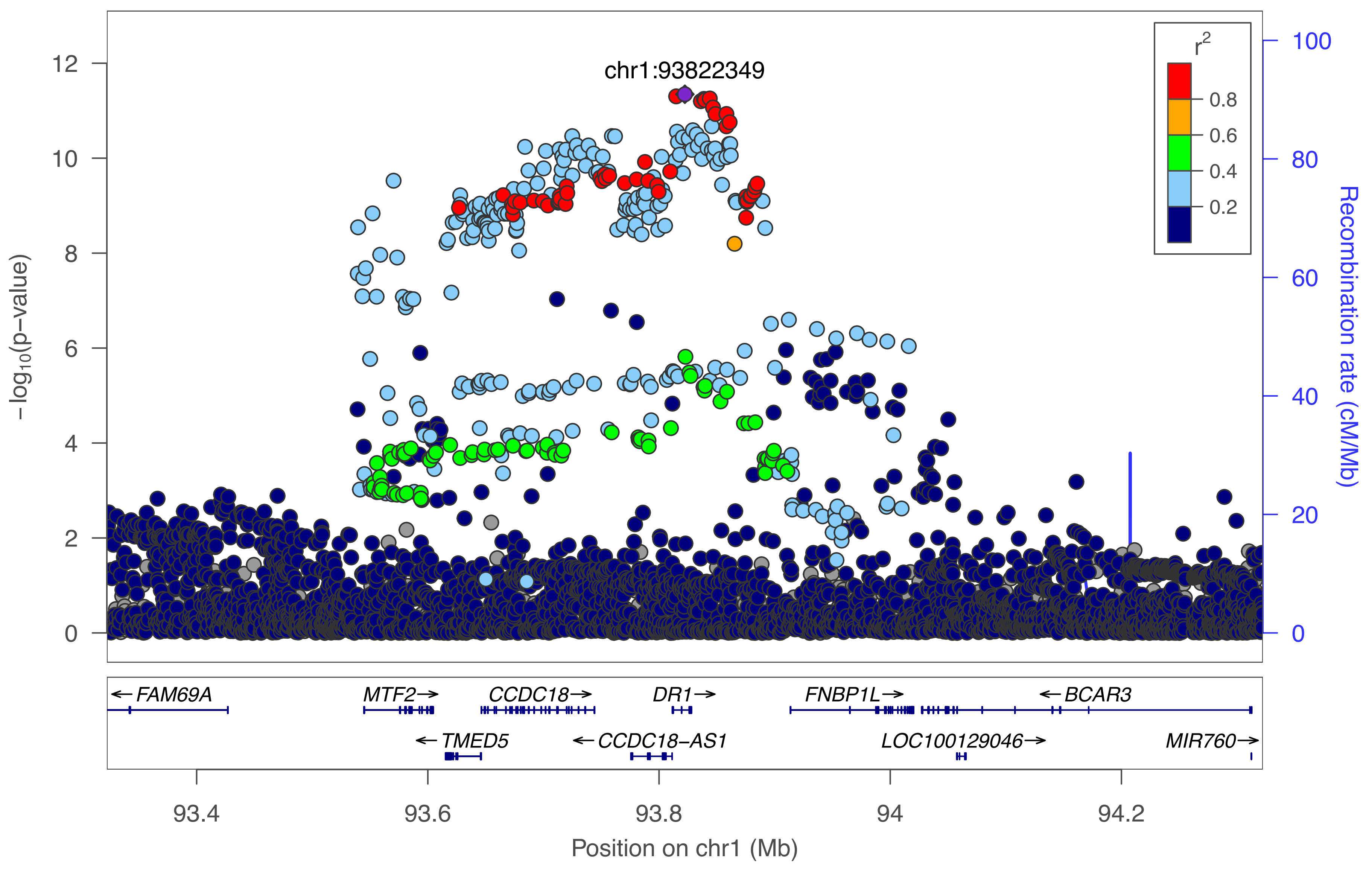

chr1:93822349
r²
0.8
0.6
0.4
0.2
−log10(p−value)
Recombination rate (cM/Mb)
←FAM69A
MTF2→
CCDC18→
DR1→
FNBP1L→
←BCAR3
←TMED5
←CCDC18−AS1
LOC100129046→
MIR760→
93.4
93.6
93.8
94
94.2
Position on chr1 (Mb)

### 1_13:Leu

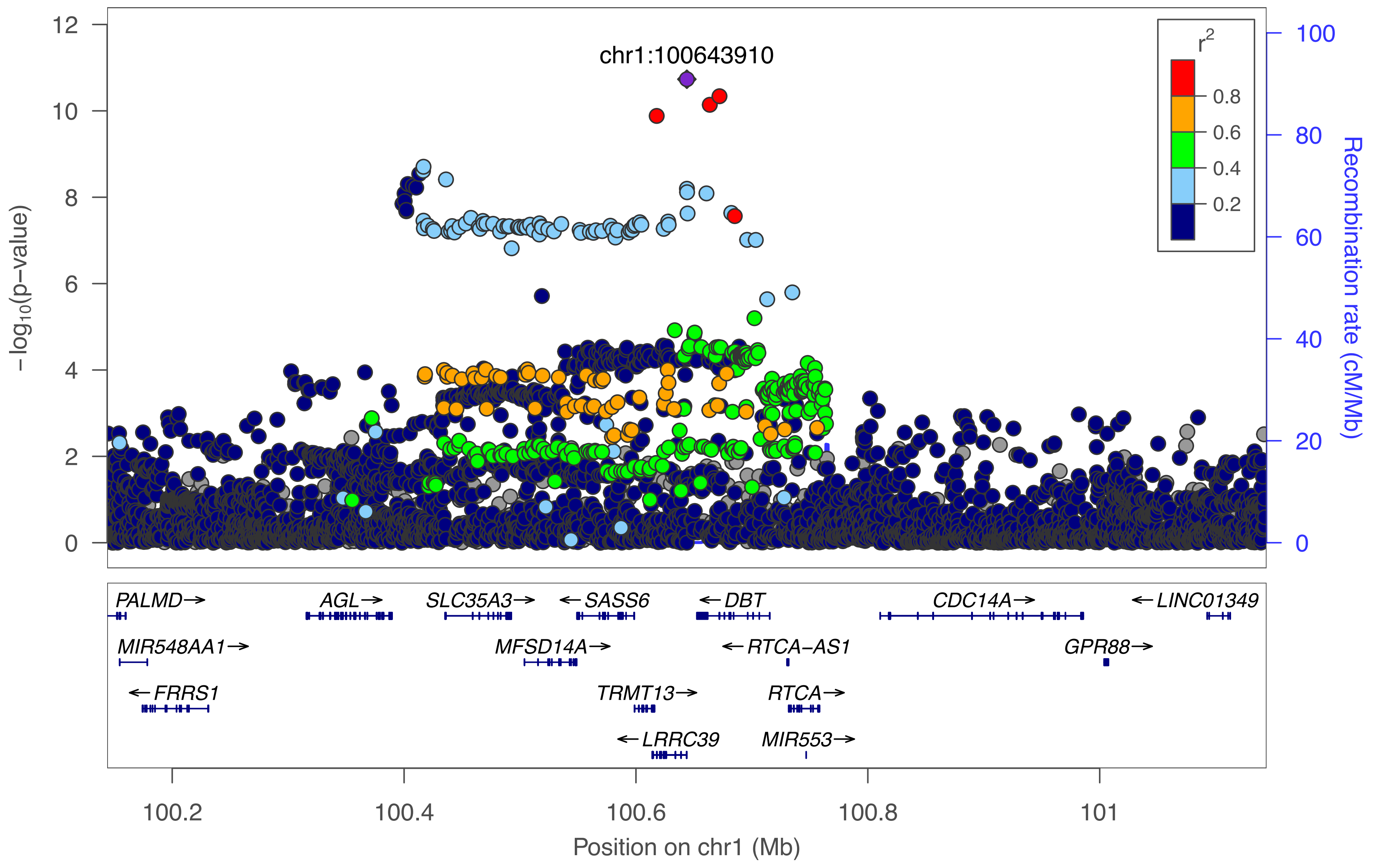

chr1:100643910
r²
0.8
0.6
0.4
0.2
−log10(p−value)
Recombination rate (cM/Mb)
PALMD→
AGL→
SLC35A3→
←SASS6
←DBT
CDC14A→
←LINC01349
MIR548AA1→
MFSD14A→
←RTCA−AS1
GPR88→
←FRRS1
TRMT13→
RTCA→
←LRRC39
MIR553→
100.2
100.4
100.6
100.8
101
Position on chr1 (Mb)

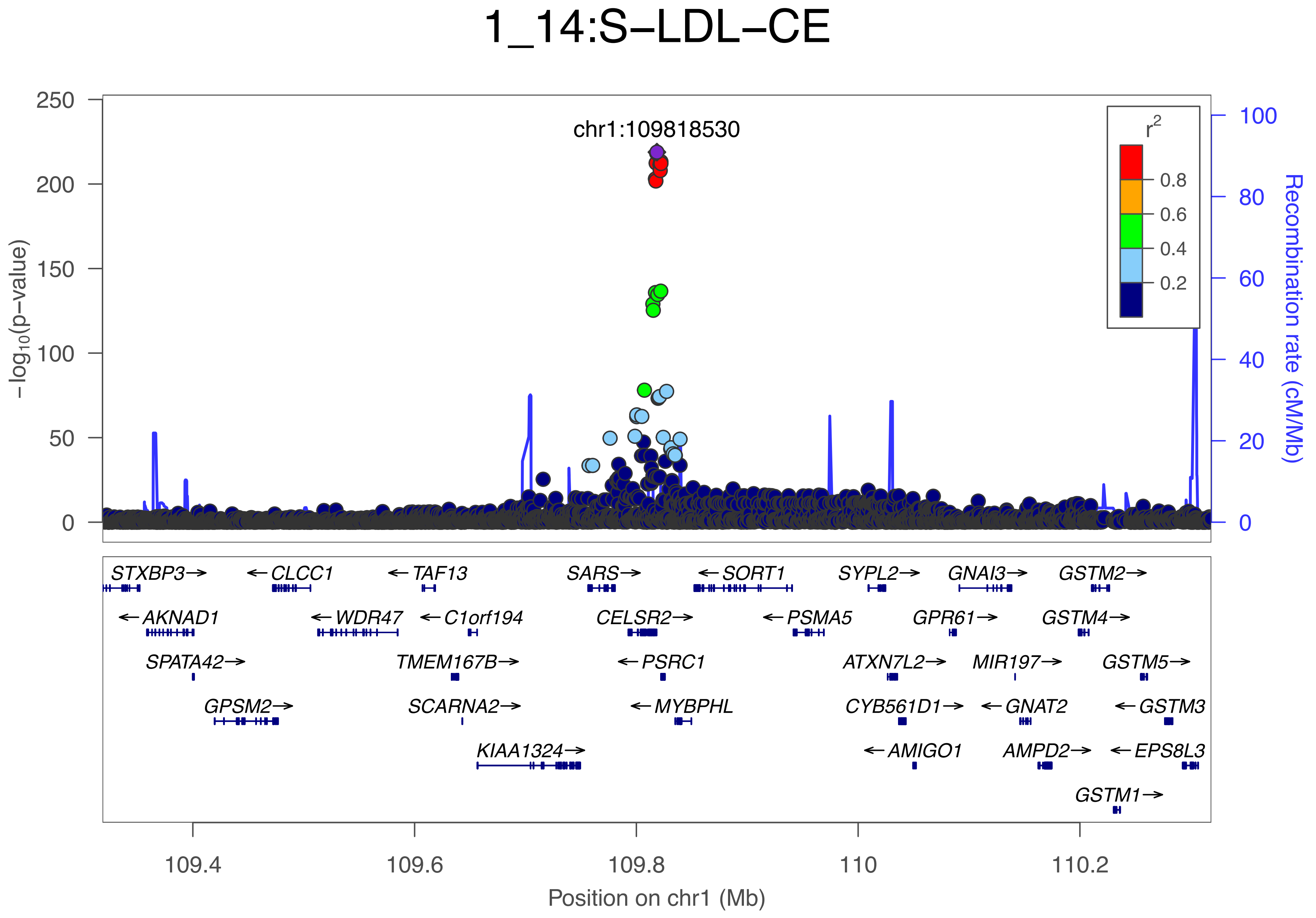

1_14:S−LDL−CE
chr1:109818530
−log10(p−value)
Recombination rate (cM/Mb)
r2
0.8
0.6
0.4
0.2
250
200
150
100
50
0
100
80
60
40
20
0
STXBP3→
←CLCC1
←TAF13
SARS→
←SORT1
SYPL2→
GNAI3→
GSTM2→
←AKNAD1
←WDR47
←C1orf194
CELSR2→
←PSMA5
GPR61→
GSTM4→
SPATA42→
TMEM167B→
←PSRC1
ATXN7L2→
MIR197→
GSTM5→
GPSM2→
SCARNA2→
←MYBPHL
CYB561D1→
←GNAT2
←GSTM3
KIAA1324→
←AMIGO1
AMPD2→
←EPS8L3
GSTM1→
109.4
109.6
109.8
110
110.2
Position on chr1 (Mb)

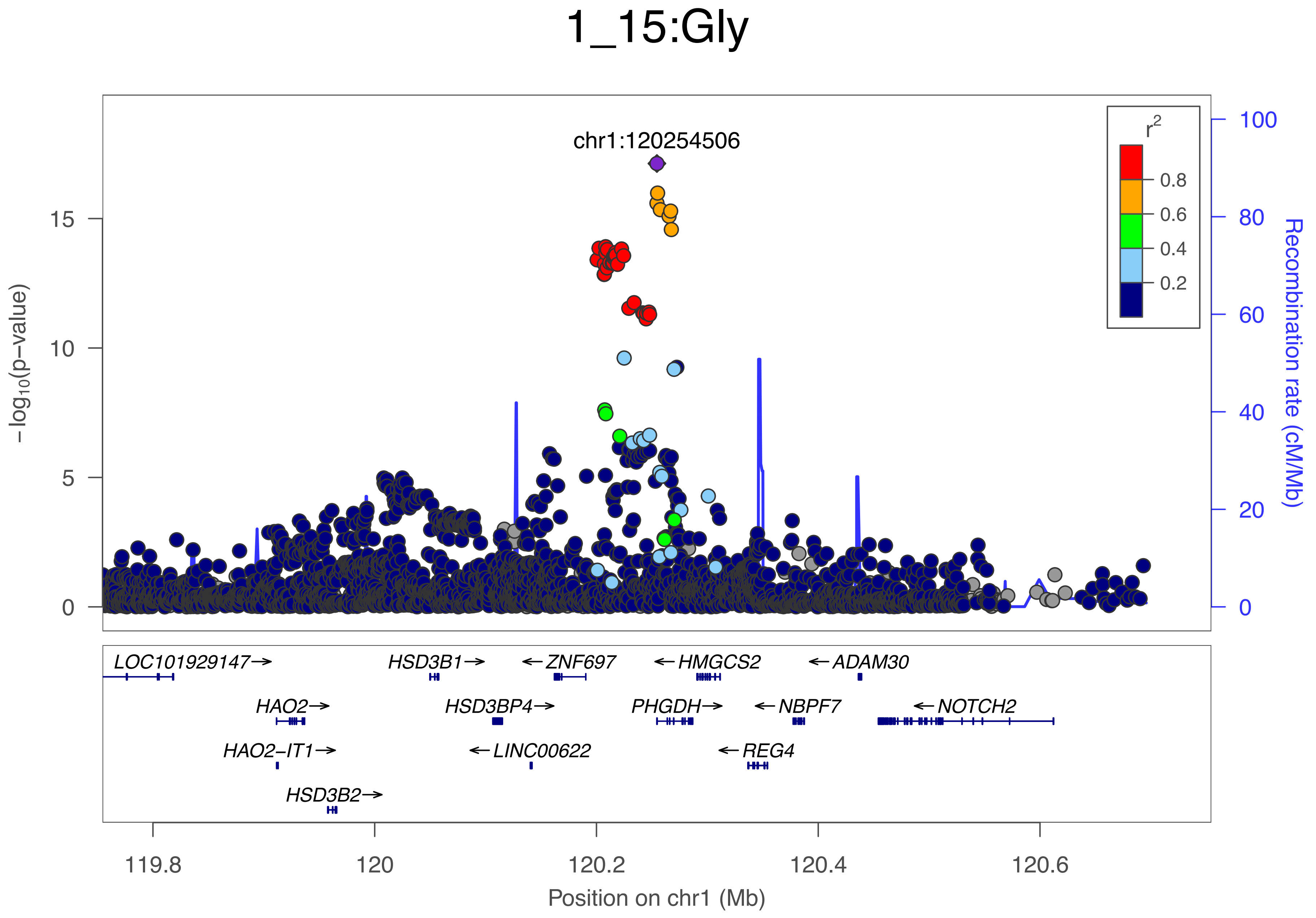
1_15:Gly
chr1:120254506
r2
0.8
0.6
0.4
0.2
−log10(p−value)
15
10
5
0
100
80
60
40
20
0
Recombination rate (cM/Mb)
LOC101929147→
HSD3B1→
←ZNF697
←HMGCS2
←ADAM30
HAO2→
HSD3BP4→
PHGDH→
←NBPF7
←NOTCH2
HAO2−IT1→
←LINC00622
←REG4
HSD3B2→
119.8
120
120.2
120.4
120.6
Position on chr1 (Mb)

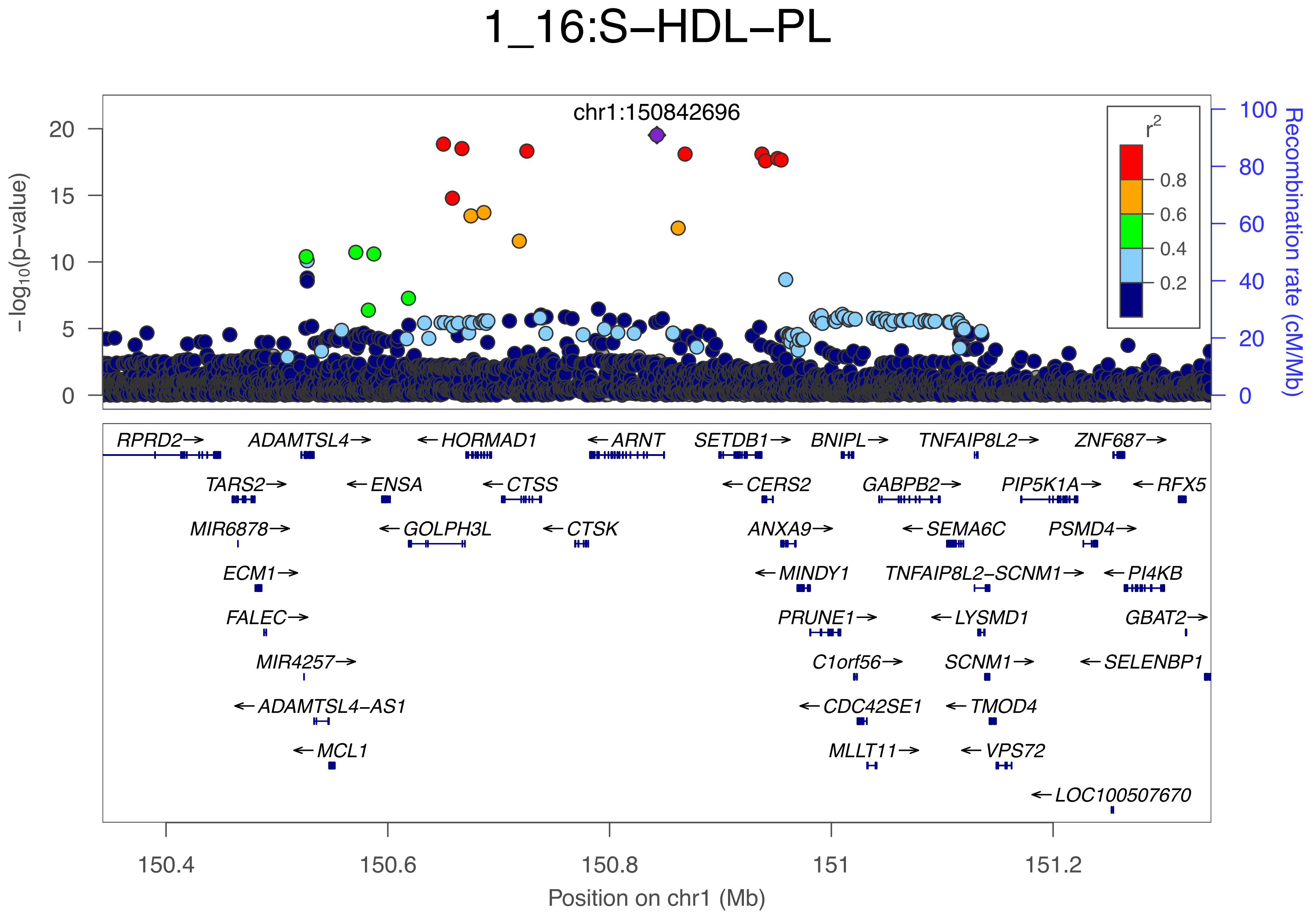

1_16:S−HDL−PL
chr1:150842696
−log10(p−value)
Recombination rate (cM/Mb)
r2
0.8
0.6
0.4
0.2
RPRD2→
ADAMTSL4→
←HORMAD1
←ARNT
SETDB1→
BNIPL→
TNFAIP8L2→
ZNF687→
TARS2→
←ENSA
←CTSS
←CERS2
GABPB2→
PIP5K1A→
←RFX5
MIR6878→
←GOLPH3L
←CTSK
ANXA9→
←SEMA6C
PSMD4→
ECM1→
←MINDY1
TNFAIP8L2−SCNM1→
←PI4KB
FALEC→
PRUNE1→
←LYSMD1
GBAT2→
MIR4257→
C1orf56→
SCNM1→
←SELENBP1
←ADAMTSL4−AS1
←CDC42SE1
←TMOD4
←MCL1
MLLT11→
←VPS72
←LOC100507670
150.4
150.6
150.8
151
151.2
Position on chr1 (Mb)

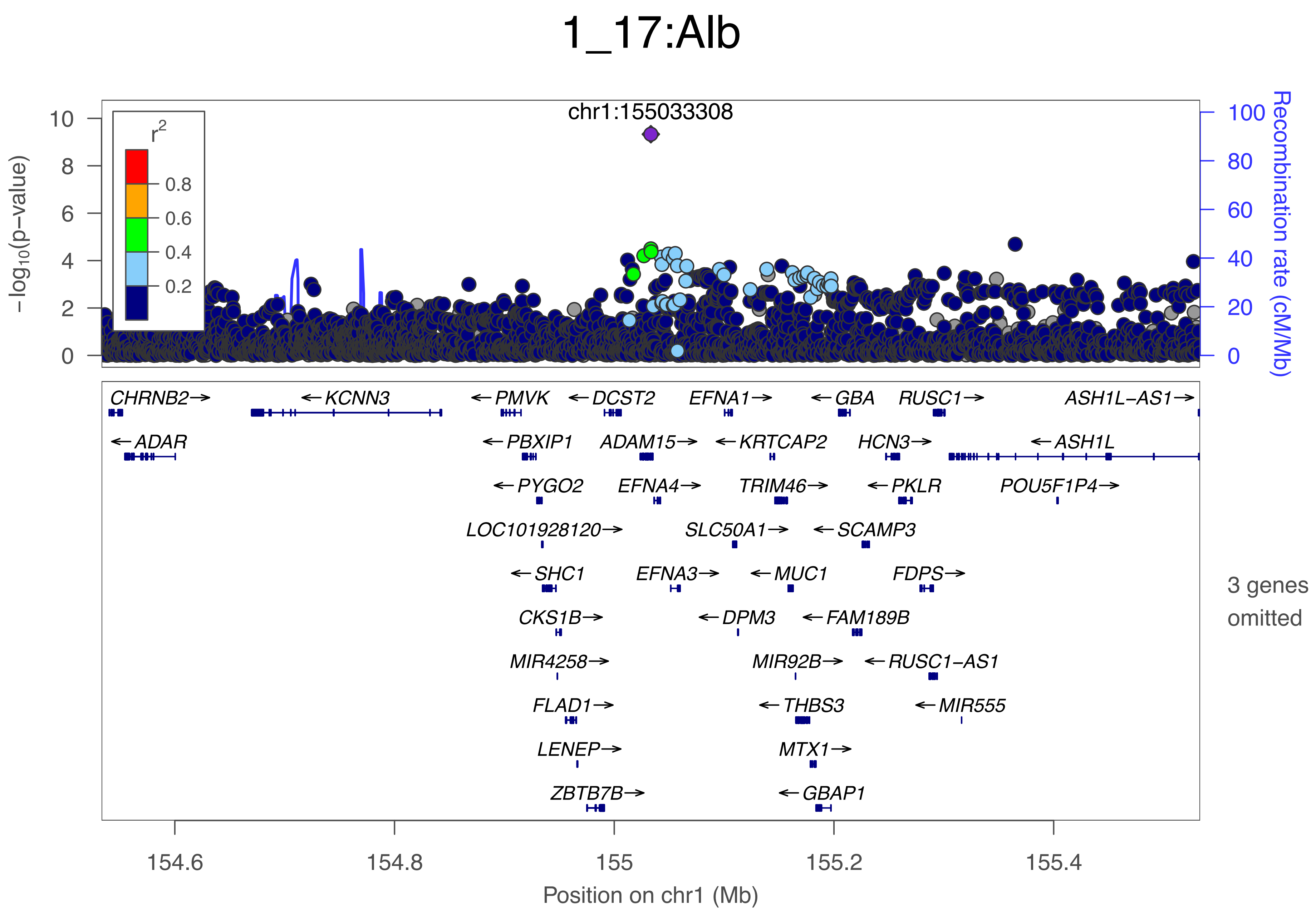
1_17:Alb
chr1:155033308
r2
0.8
0.6
0.4
0.2
−log10(p−value)
Recombination rate (cM/Mb)
0
2
4
6
8
10
0
20
40
60
80
100
CHRNB2→
←KCNN3
←PMVK
←DCST2
EFNA1→
←GBA
RUSC1→
ASH1L−AS1→
←ADAR
←PBXIP1
ADAM15→
←KRTCAP2
HCN3→
←ASH1L
←PYGO2
EFNA4→
TRIM46→
←PKLR
POU5F1P4→
LOC101928120→
SLC50A1→
←SCAMP3
←SHC1
EFNA3→
←MUC1
FDPS→
CKS1B→
←DPM3
←FAM189B
MIR4258→
MIR92B→
←RUSC1−AS1
FLAD1→
←THBS3
←MIR555
LENEP→
MTX1→
ZBTB7B→
←GBAP1
3 genes omitted
154.6
154.8
155
155.2
155.4
Position on chr1 (Mb)

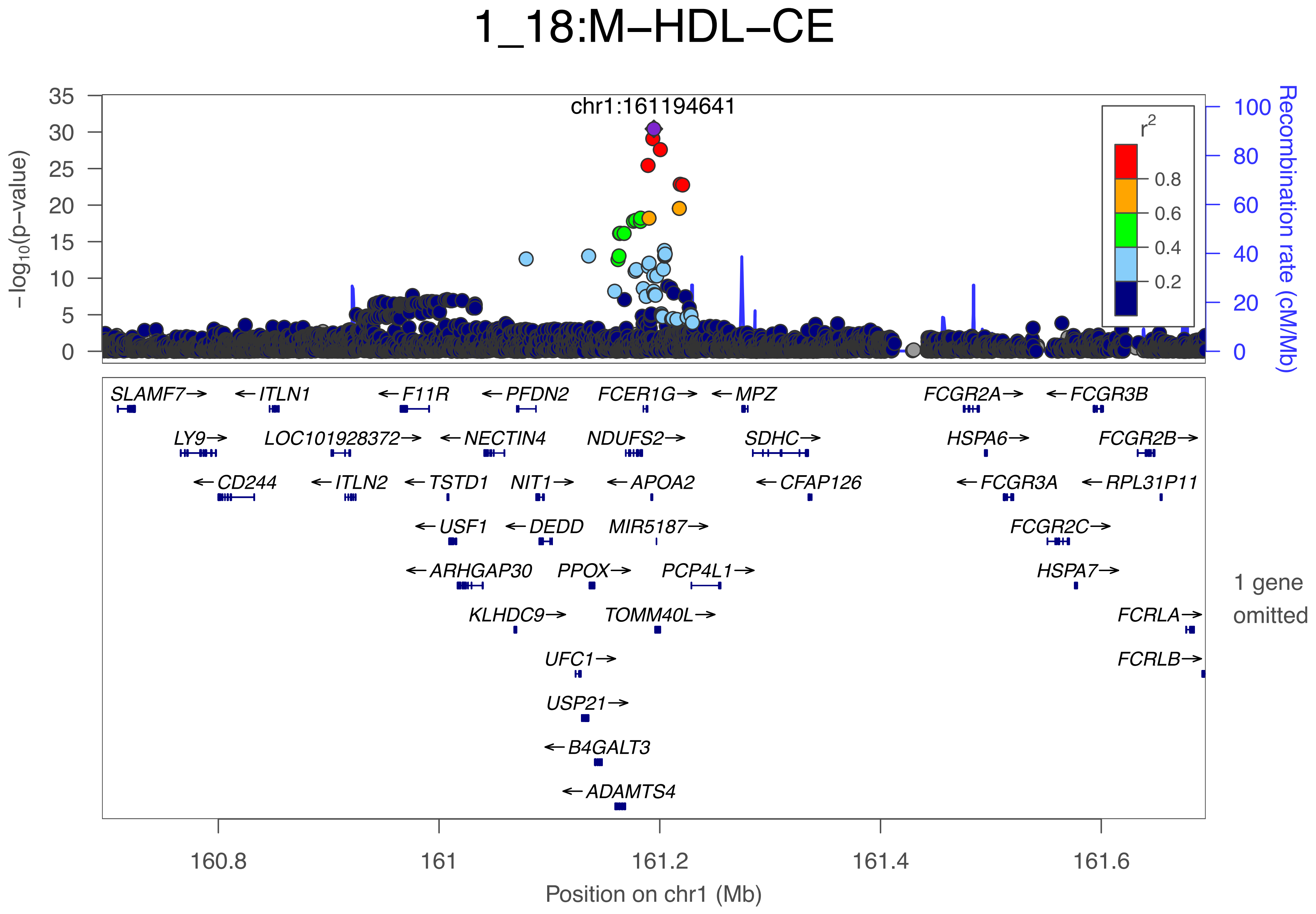

1_18:M−HDL−CE
chr1:161194641
−log10(p−value)
Recombination rate (cM/Mb)
r2
0.8
0.6
0.4
0.2
SLAMF7→
←ITLN1
←F11R
←PFDN2
FCER1G→
←MPZ
FCGR2A→
←FCGR3B
LY9→
LOC101928372→
←NECTIN4
NDUFS2→
SDHC→
HSPA6→
FCGR2B→
←CD244
←ITLN2
←TSTD1
NIT1→
←APOA2
←CFAP126
←FCGR3A
←RPL31P11
←USF1
←DEDD
MIR5187→
FCGR2C→
←ARHGAP30
PPOX→
PCP4L1→
HSPA7→
1 gene omitted
KLHDC9→
TOMM40L→
FCRLA→
UFC1→
FCRLB→
USP21→
←B4GALT3
←ADAMTS4
160.8
161
161.2
161.4
161.6
Position on chr1 (Mb)

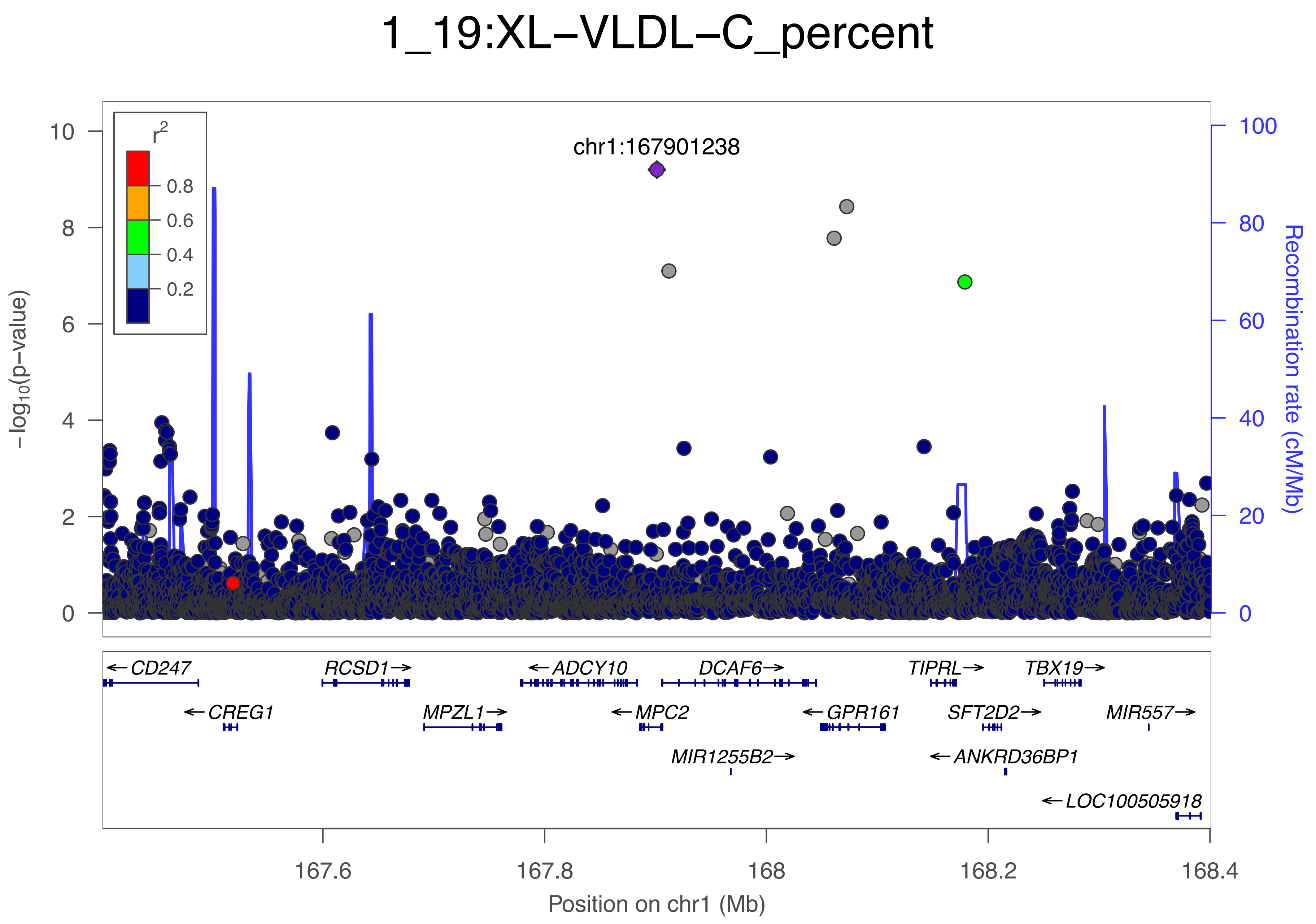

1_19:XL−VLDL−C_percent
r²
0.8
0.6
0.4
0.2
chr1:167901238
−log₁₀(p−value)
Recombination rate (cM/Mb)
0
2
4
6
8
10
0
20
40
60
80
100
CD247
CREG1
RCSD1
MPZL1
ADCY10
MPC2
DCAF6
MIR1255B2
GPR161
TIPRL
SFT2D2
ANKRD36BP1
TBX19
MIR557
LOC100505918
167.6
167.8
168
168.2
168.4
Position on chr1 (Mb)

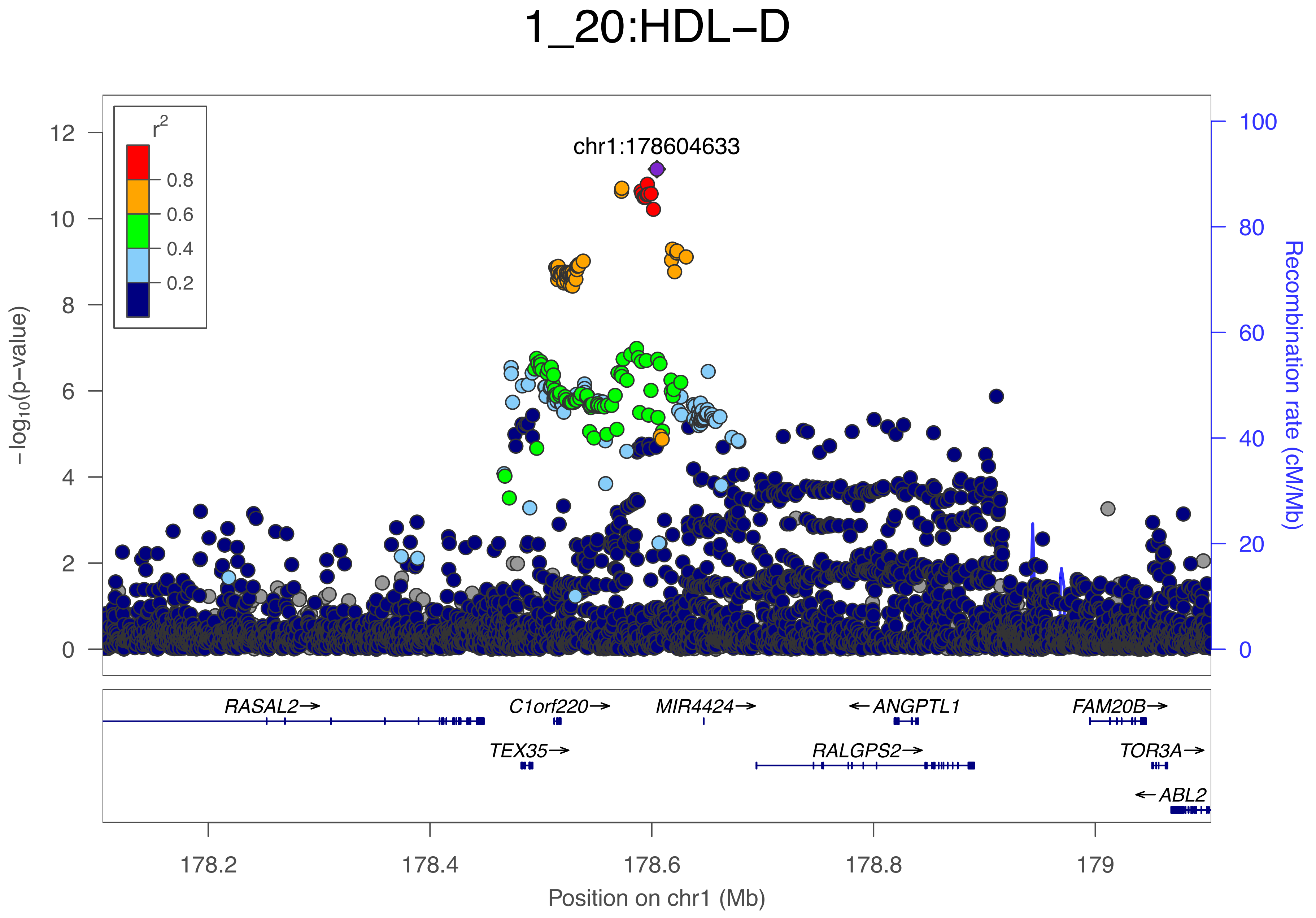
1_20:HDL−D
chr1:178604633
r²
0.8
0.6
0.4
0.2
−log10(p−value)
Recombination rate (cM/Mb)
0
2
4
6
8
10
12
20
40
60
80
100
RASAL2→
C1orf220→
MIR4424→
←ANGPTL1
FAM20B→
TEX35→
RALGPS2→
TOR3A→
←ABL2
178.2
178.4
178.6
178.8
179
Position on chr1 (Mb)

### 1_21:HDL–D

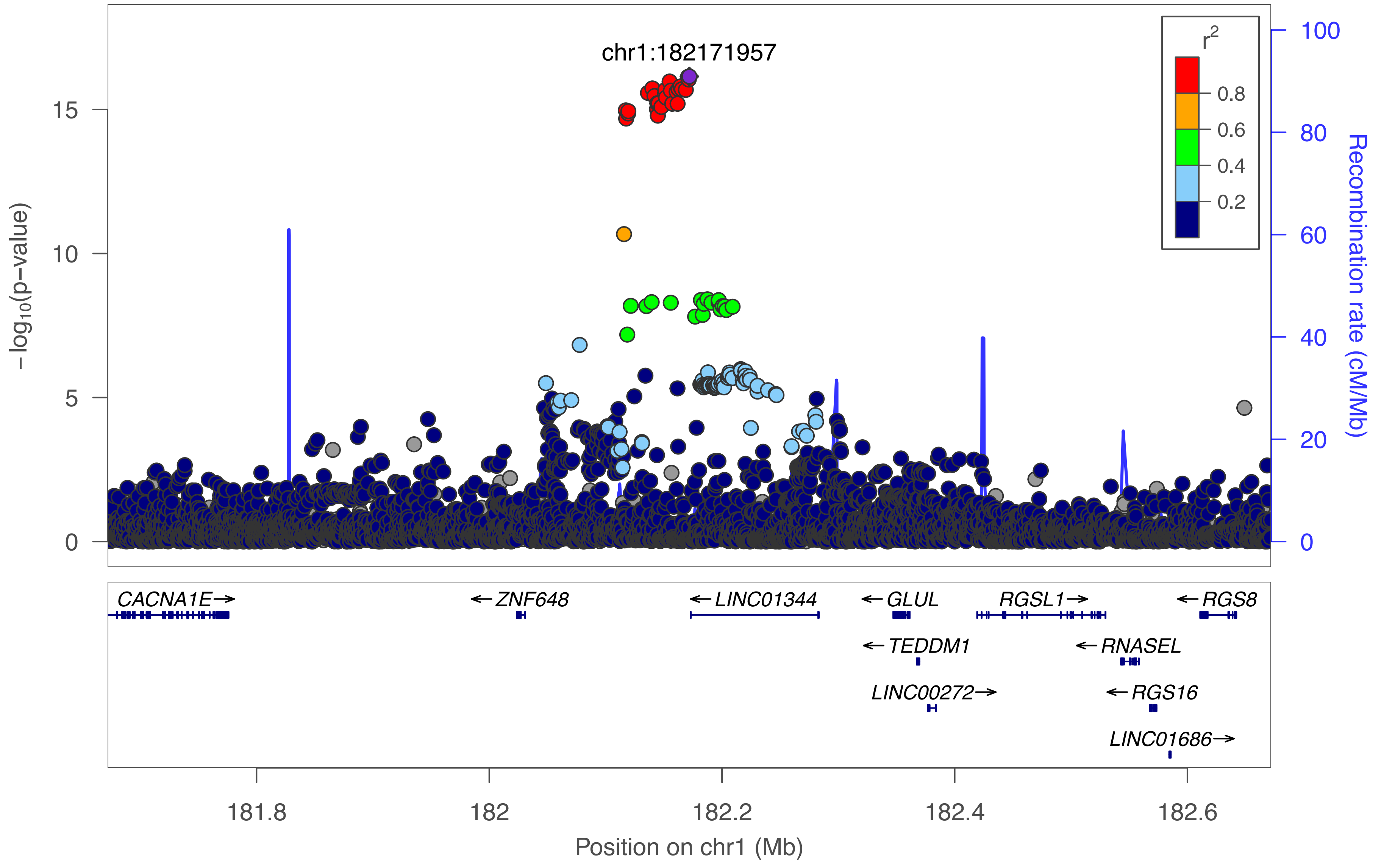

chr1:182171957
r²
0.8
0.6
0.4
0.2
−log10(p−value)
15
10
5
0
100
80
60
40
20
0
Recombination rate (cM/Mb)
CACNA1E→
←ZNF648
←LINC01344
←GLUL
RGSL1→
←RGS8
←TEDDM1
←RNASEL
LINC00272→
←RGS16
LINC01686→
181.8
182
182.2
182.4
182.6
Position on chr1 (Mb)

1_22:GlycA

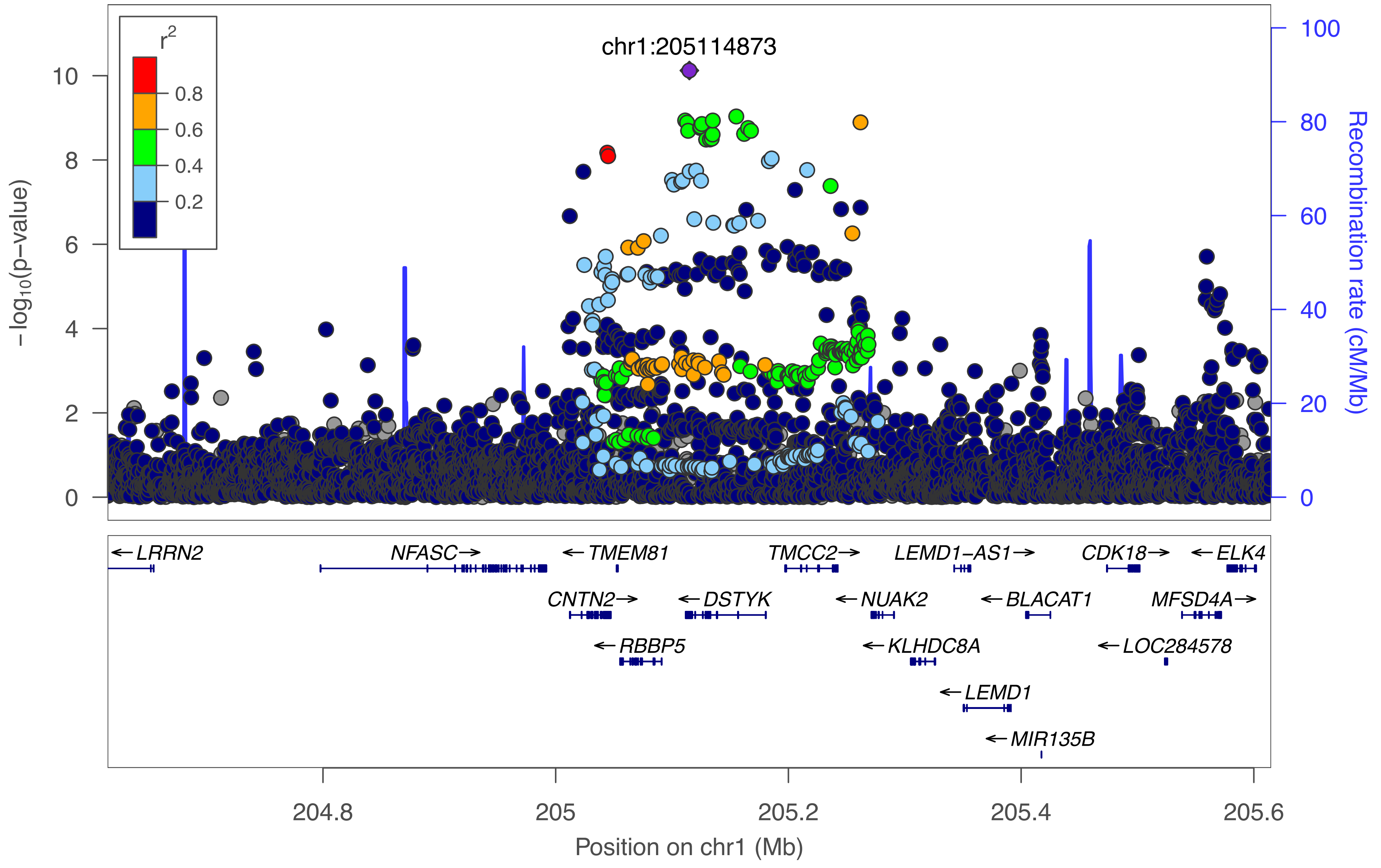

r2
0.8
0.6
0.4
0.2
chr1:205114873
−log10(p−value)
0
2
4
6
8
10
Recombination rate (cM/Mb)
0
20
40
60
80
100
LRRN2
NFASC
TMEM81
TMCC2
LEMD1−AS1
CDK18
ELK4
CNTN2
DSTYK
NUAK2
BLACAT1
MFSD4A
RBBP5
KLHDC8A
LOC284578
LEMD1
MIR135B
204.8
205
205.2
205.4
205.6
Position on chr1 (Mb)

1_23:Ala

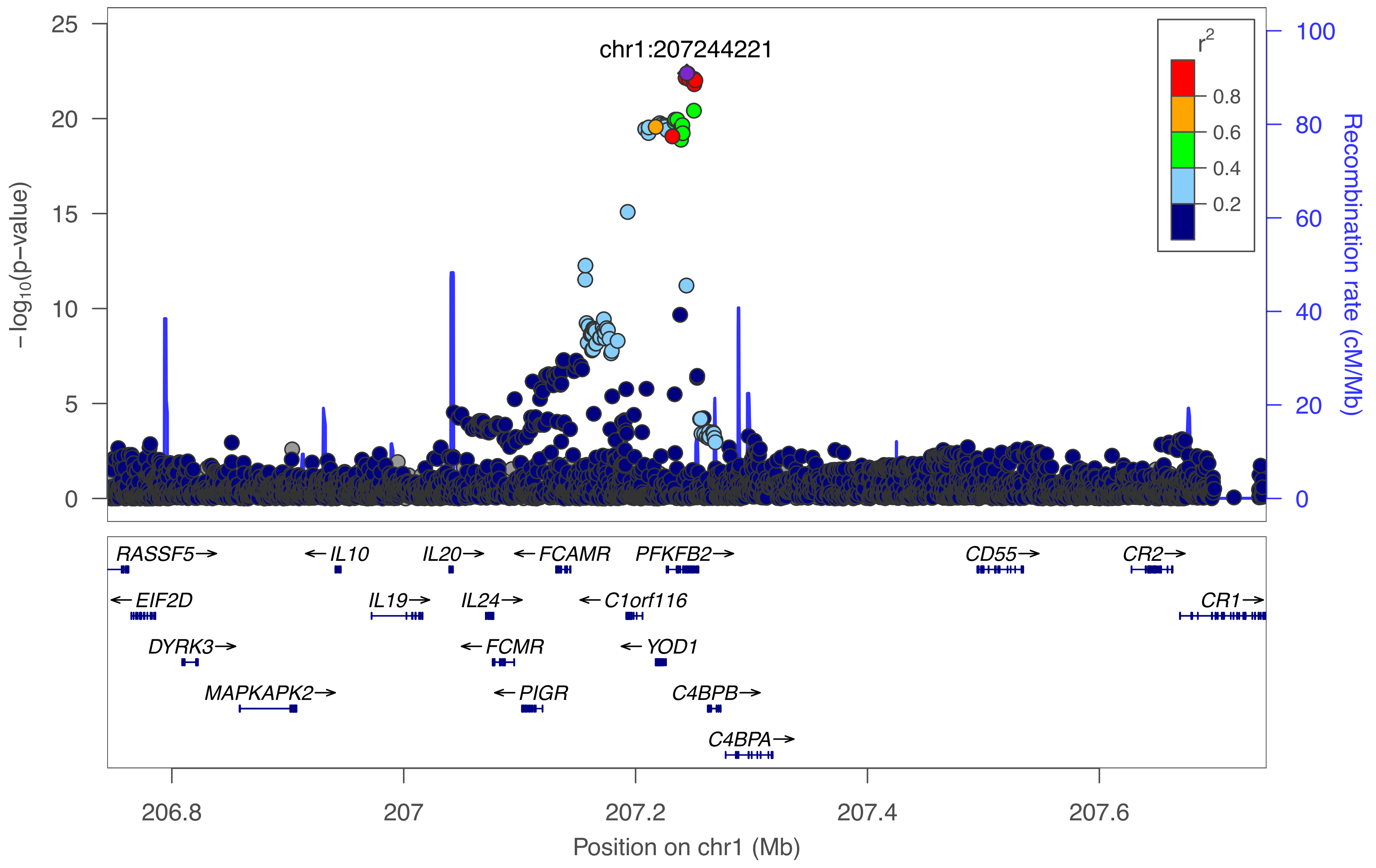

chr1:207244221
25
20
15
10
5
0
−log10(p−value)
100
80
60
40
20
0
Recombination rate (cM/Mb)
r2
0.8
0.6
0.4
0.2
RASSF5→
←IL10
IL20→
←FCAMR
PFKFB2→
CD55→
CR2→
←EIF2D
IL19→
IL24→
←C1orf116
CR1→
DYRK3→
←FCMR
←YOD1
MAPKAPK2→
←PIGR
C4BPB→
C4BPA→
206.8
207
207.2
207.4
207.6
Position on chr1 (Mb)

### 1_24:Gln

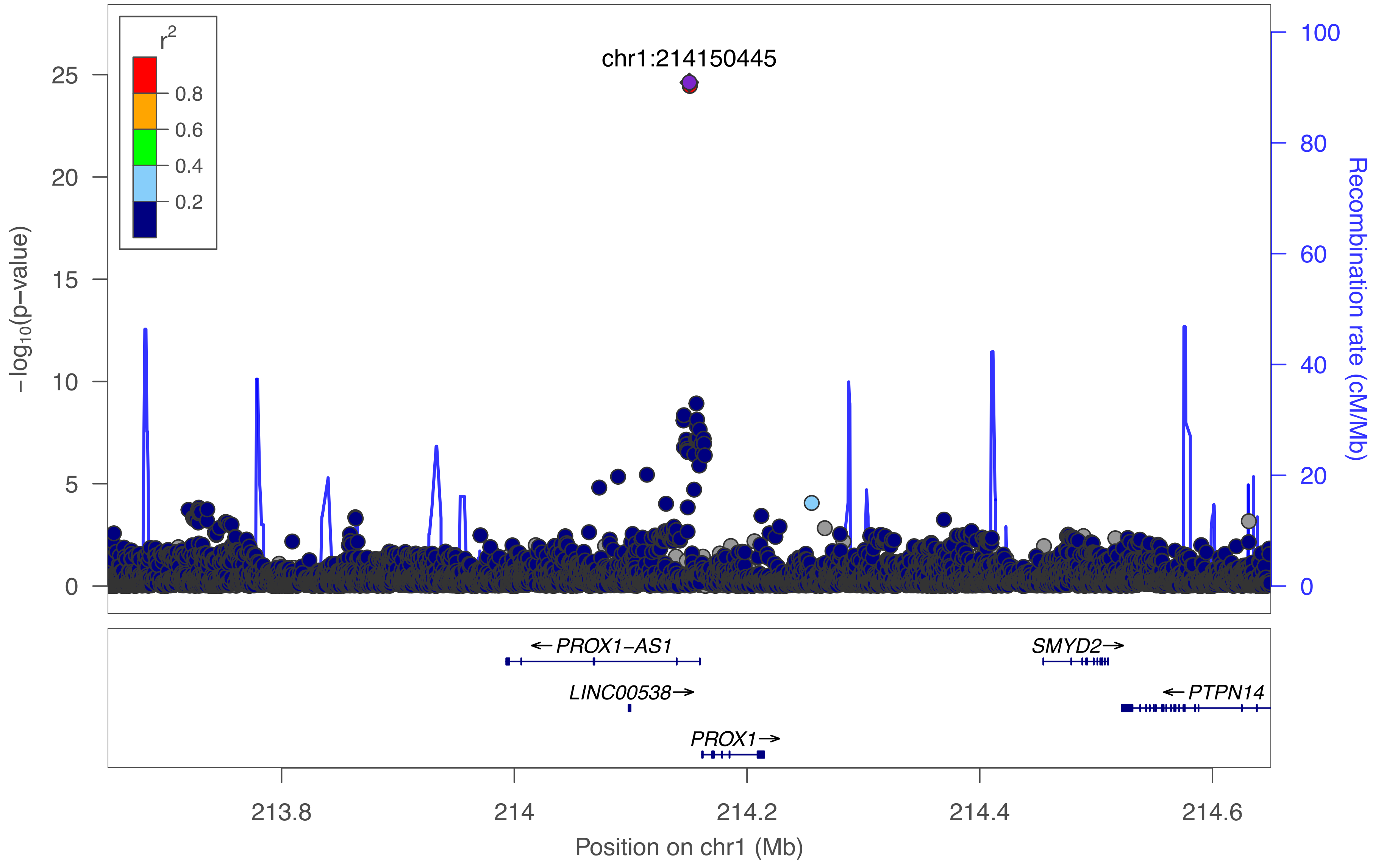

chr1:214150445
r2
0.8
0.6
0.4
0.2
−log10(p−value)
0
5
10
15
20
25
Recombination rate (cM/Mb)
0
20
40
60
80
100
← PROX1−AS1
LINC00538→
PROX1→
SMYD2→
← PTPN14
213.8
214
214.2
214.4
214.6
Position on chr1 (Mb)

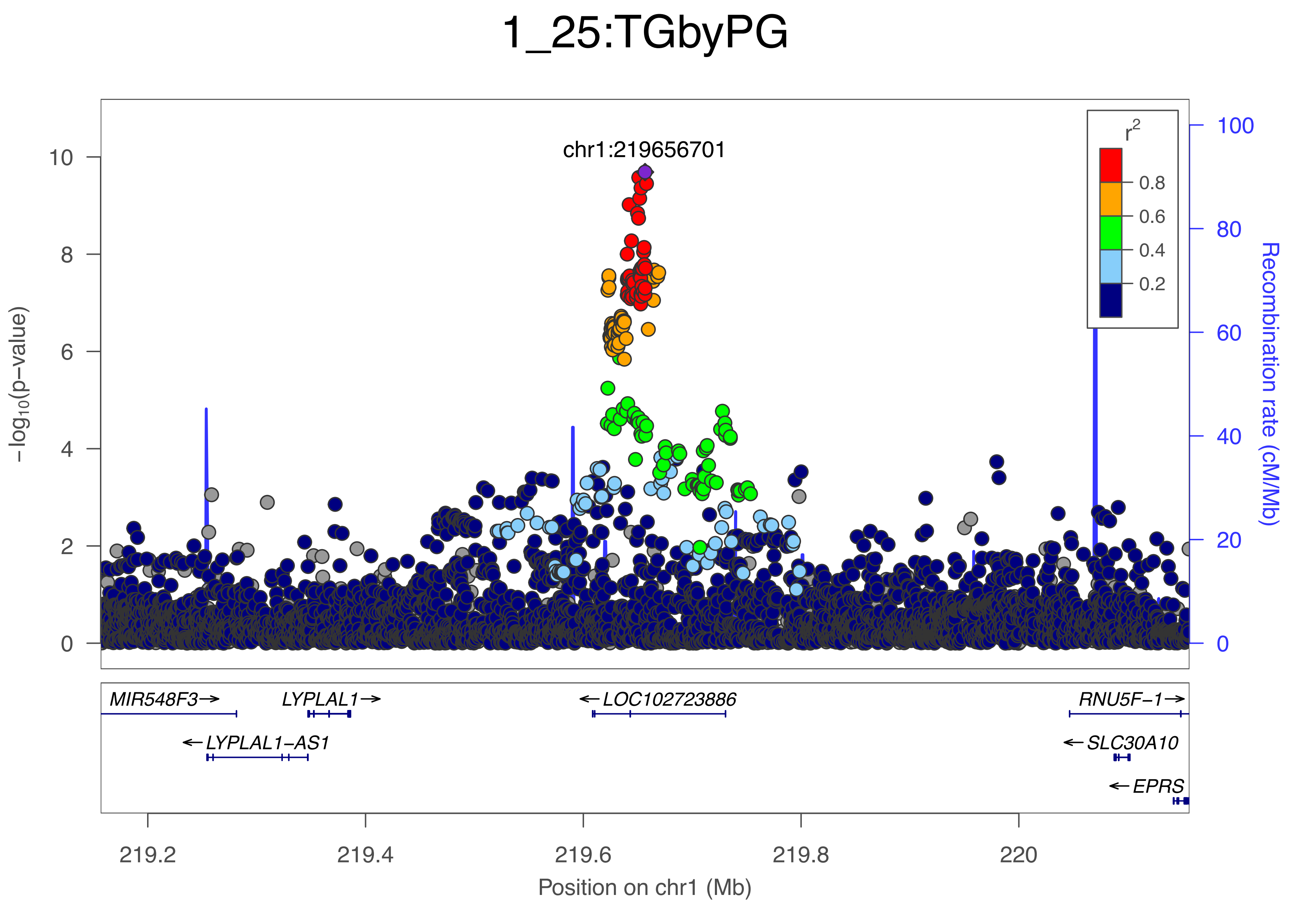

1_25:TGbyPG
chr1:219656701
r2
0.8
0.6
0.4
0.2
−log10(p−value)
Recombination rate (cM/Mb)
MIR548F3→
LYPLAL1→
←LOC102723886
RNU5F−1→
←LYPLAL1−AS1
←SLC30A10
←EPRS
219.2
219.4
219.6
219.8
220
Position on chr1 (Mb)

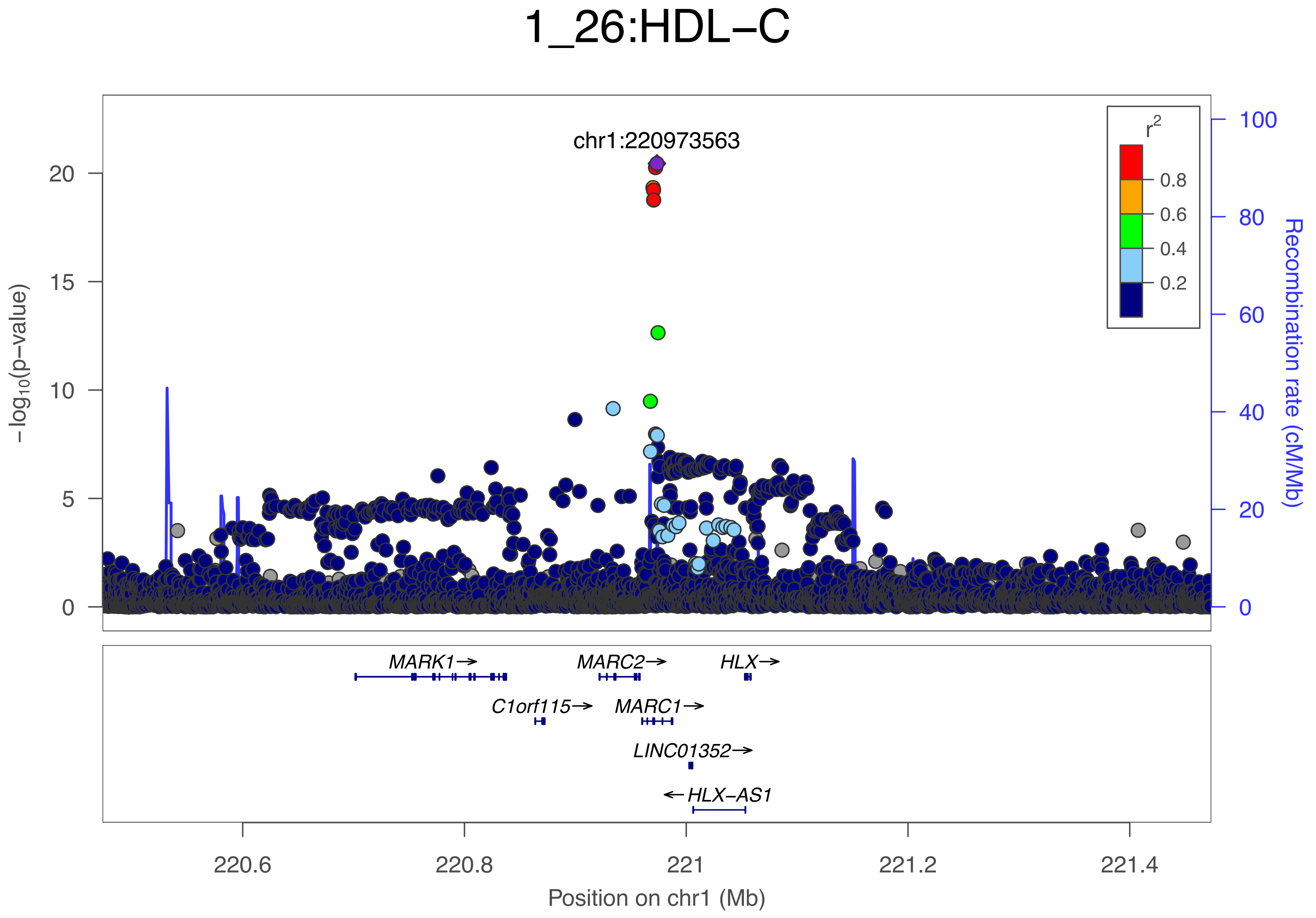
1_26:HDL−C
chr1:220973563
−log10(p−value)
Recombination rate (cM/Mb)
r2
0.8
0.6
0.4
0.2
0
5
10
15
20
0
20
40
60
80
100
MARK1→
MARC2→
HLX→
C1orf115→
MARC1→
LINC01352→
←HLX−AS1
220.6
220.8
221
221.2
221.4
Position on chr1 (Mb)

### 1_27:TGbyPG

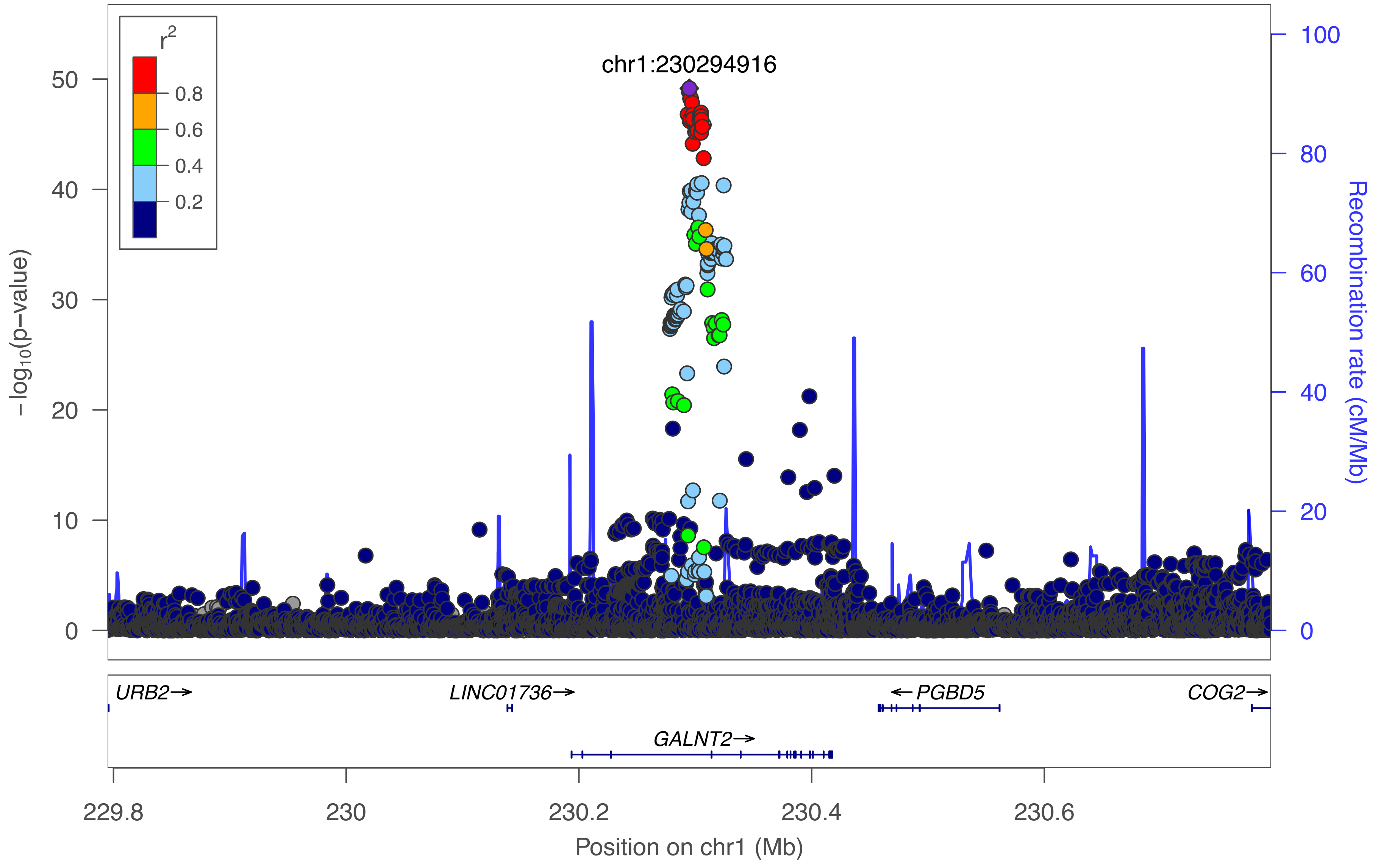

r²
0.8
0.6
0.4
0.2
chr1:230294916
−log10(p−value)
50
40
30
20
10
0
Recombination rate (cM/Mb)
100
80
60
40
20
0
URB2→
LINC01736→
←PGBD5
COG2→
GALNT2→
229.8
230
230.2
230.4
230.6
Position on chr1 (Mb)

### 1_28:M–LDL–FC

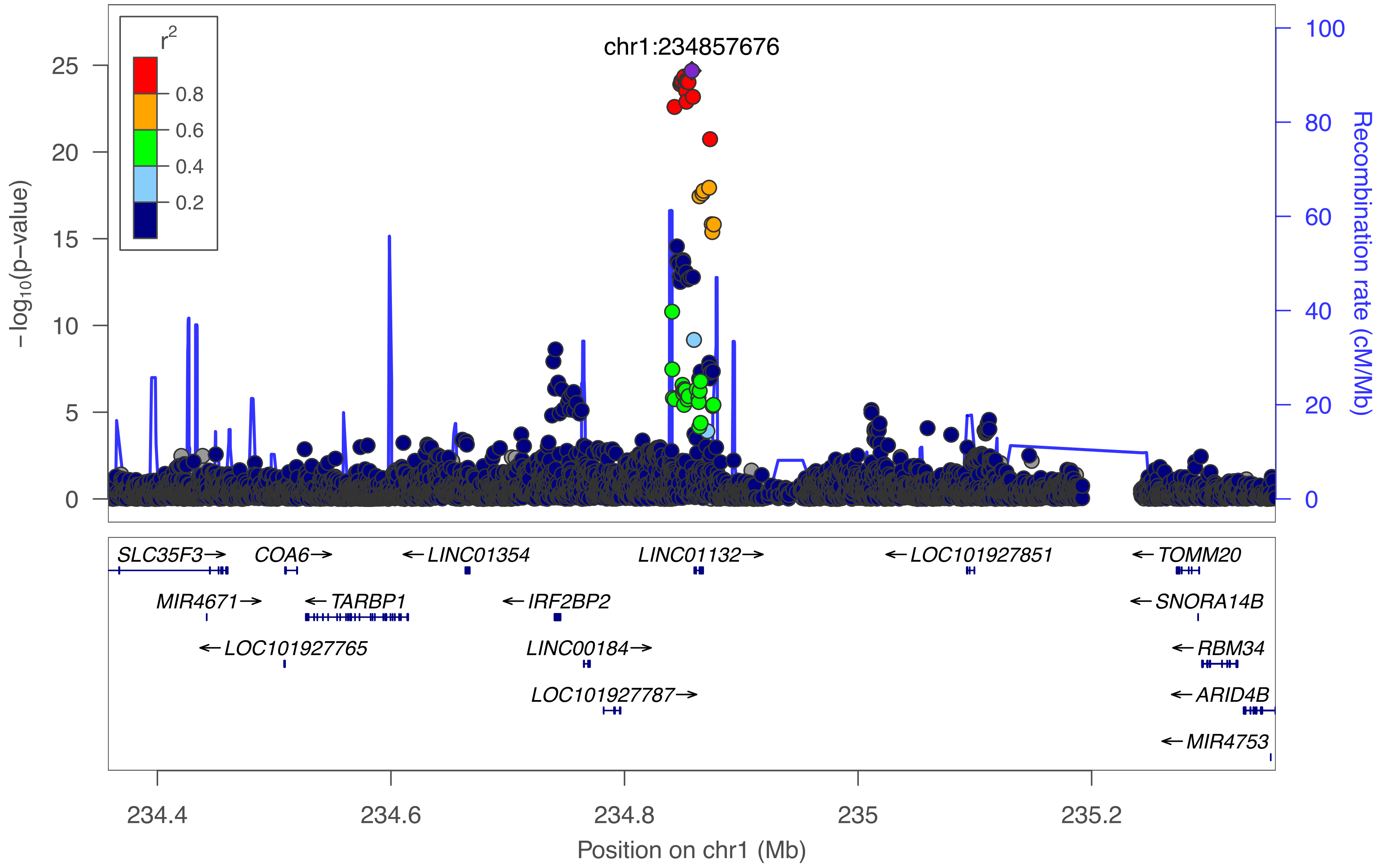

chr1:234857676
r²
0.8
0.6
0.4
0.2
−log10(p−value)
0
5
10
15
20
25
Recombination rate (cM/Mb)
0
20
40
60
80
100
SLC35F3→
COA6→
←LINC01354
LINC01132→
←LOC101927851
←TOMM20
MIR4671→
←TARBP1
←IRF2BP2
←SNORA14B
←LOC101927765
LINC00184→
←RBM34
LOC101927787→
←ARID4B
←MIR4753
234.4
234.6
234.8
235
235.2
Position on chr1 (Mb)

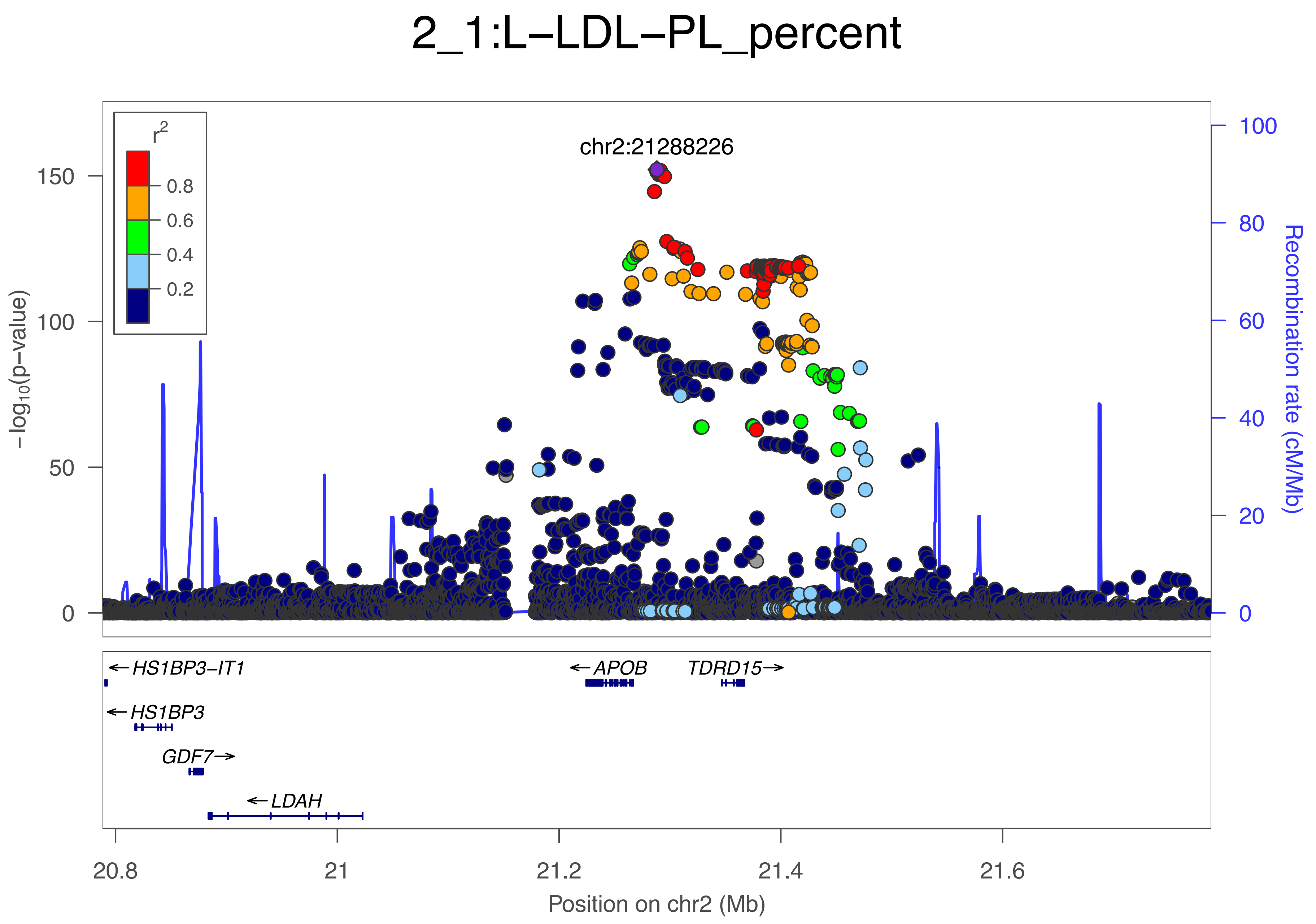
2_1:L−LDL−PL_percent
chr2:21288226
r2
0.8
0.6
0.4
0.2
150
100
50
0
−log10(p−value)
100
80
60
40
20
0
Recombination rate (cM/Mb)
←HS1BP3−IT1
←HS1BP3
GDF7→
←LDAH
←APOB
TDRD15→
20.8
21
21.2
21.4
21.6
Position on chr2 (Mb)

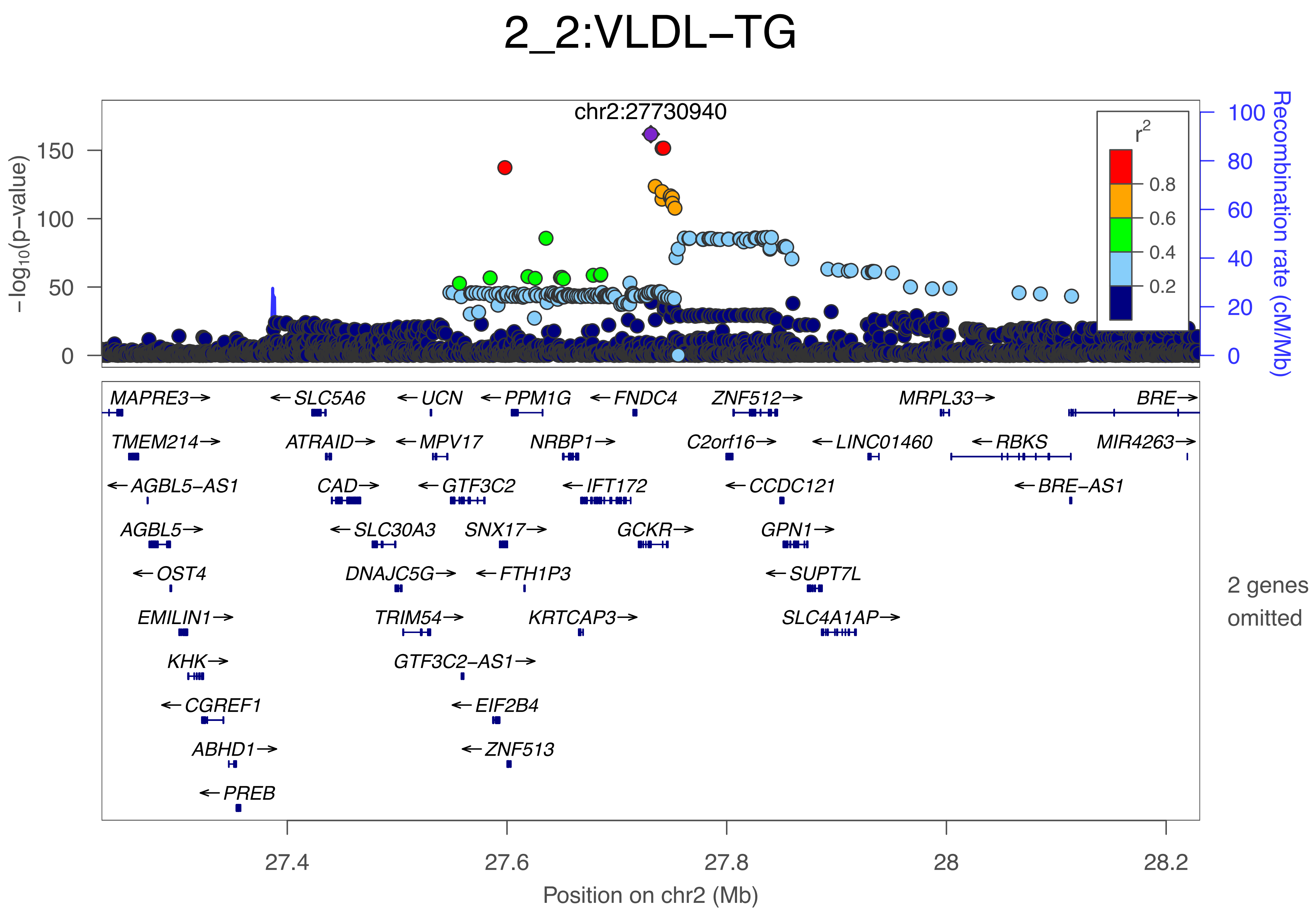
2_2:VLDL−TG
chr2:27730940
−log10(p−value)
150
100
50
0
Recombination rate (cM/Mb)
100
80
60
40
20
0
r2
0.8
0.6
0.4
0.2
MAPRE3→
←SLC5A6
←UCN
←PPM1G
←FNDC4
ZNF512→
MRPL33→
BRE→
TMEM214→
ATRAID→
←MPV17
NRBP1→
C2orf16→
←LINC01460
←RBKS
MIR4263→
←AGBL5−AS1
CAD→
←GTF3C2
←IFT172
←CCDC121
←BRE−AS1
AGBL5→
←SLC30A3
SNX17→
GCKR→
GPN1→
←OST4
DNAJC5G→
←FTH1P3
←SUPT7L
2 genes omitted
EMILIN1→
TRIM54→
KRTCAP3→
SLC4A1AP→
KHK→
GTF3C2−AS1→
←CGREF1
←EIF2B4
ABHD1→
←ZNF513
←PREB
27.4
27.6
27.8
28
28.2
Position on chr2 (Mb)

### 2_3:IDL−CE

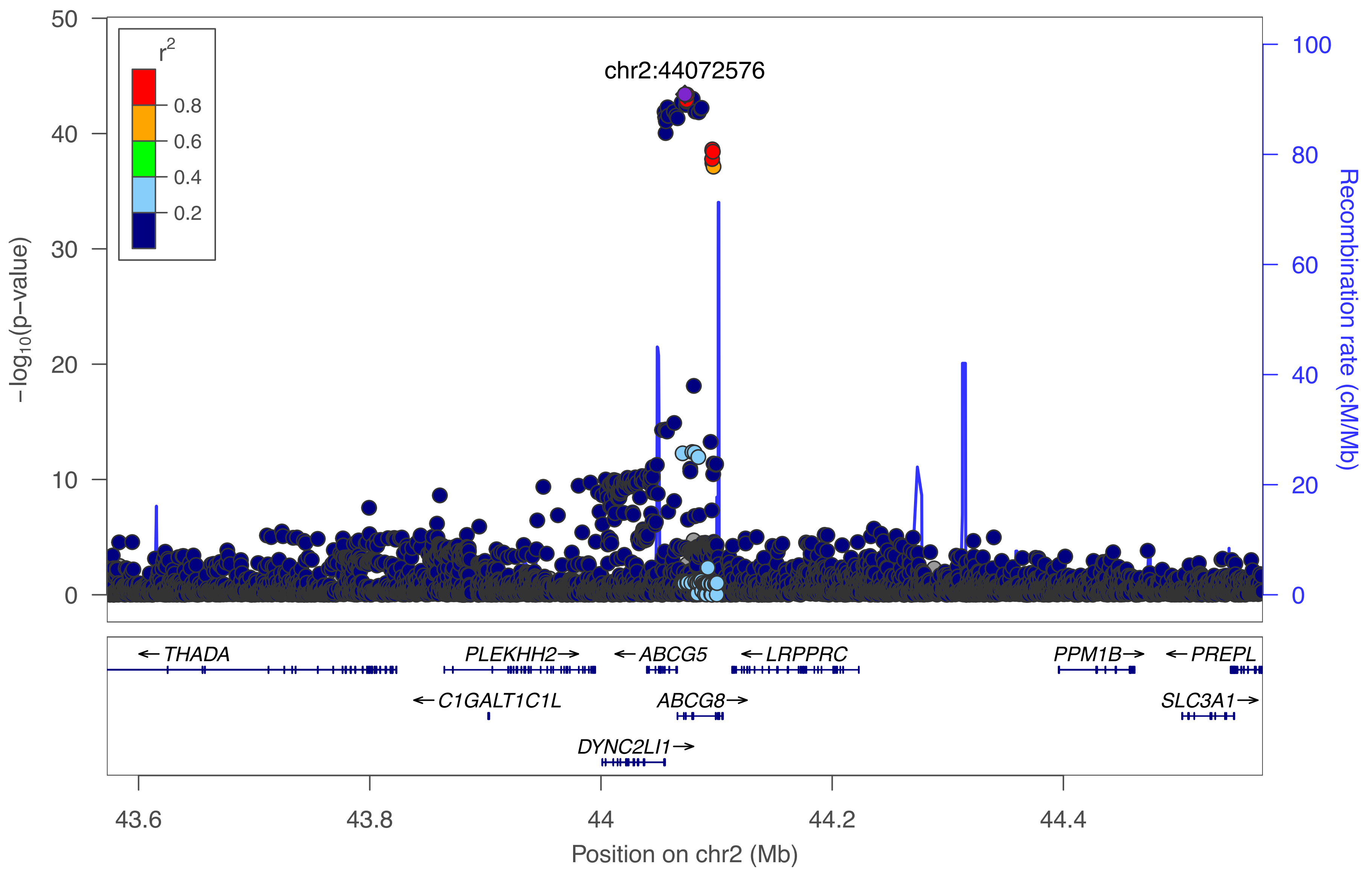

r²
0.8
0.6
0.4
0.2
chr2:44072576
−log10(p−value)
Recombination rate (cM/Mb)
50
40
30
20
10
0
100
80
60
40
20
0
← THADA
PLEKHH2→
← ABCG5
← LRPPRC
PPM1B→
← PREPL
← C1GALT1C1L
ABCG8→
SLC3A1→
DYNC2LI1→
43.6
43.8
44
44.2
44.4
Position on chr2 (Mb)

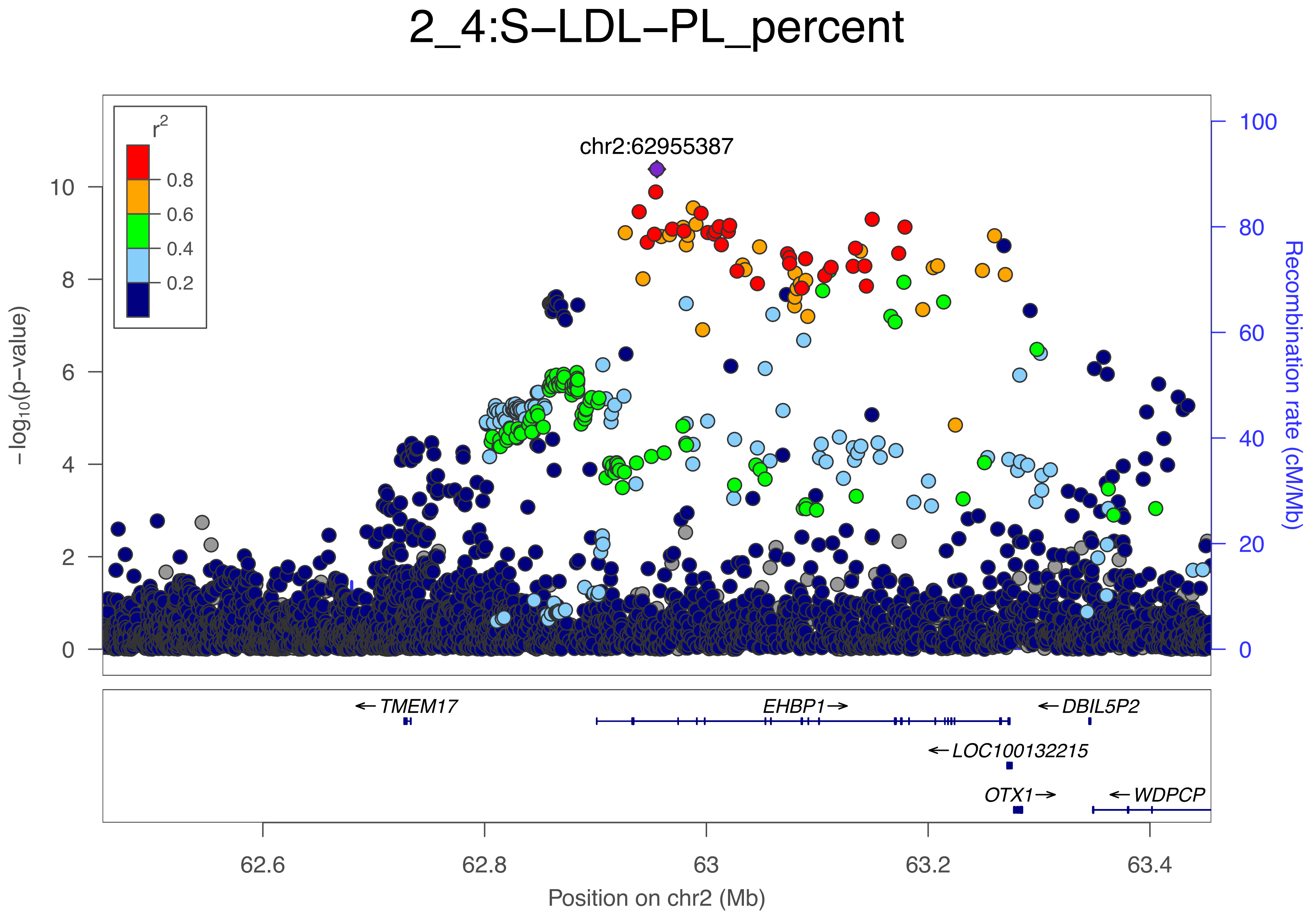

2_4:S−LDL−PL_percent
r2
0.8
0.6
0.4
0.2
chr2:62955387
−log10(p−value)
Recombination rate (cM/Mb)
0
2
4
6
8
10
0
20
40
60
80
100
←TMEM17
EHBP1→
←DBIL5P2
←LOC100132215
OTX1→
←WDPCP
62.6
62.8
63
63.2
63.4
Position on chr2 (Mb)

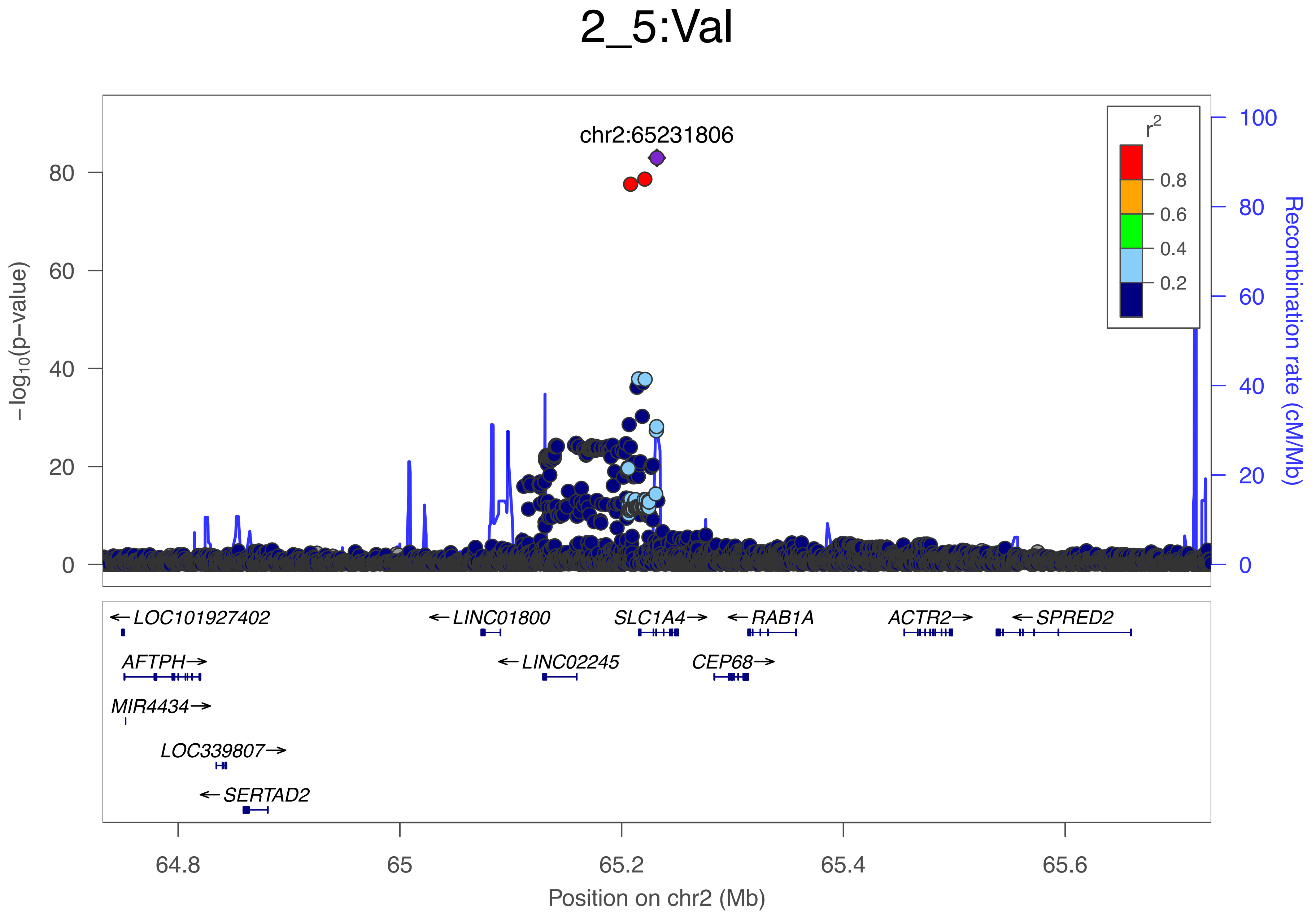

2_5:Val
chr2:65231806
r2
0.8
0.6
0.4
0.2
−log10(p−value)
0
20
40
60
80
Recombination rate (cM/Mb)
0
20
40
60
80
100
LOC101927402
LINC01800
SLC1A4
RAB1A
ACTR2
SPRED2
AFTPH
LINC02245
CEP68
MIR4434
LOC339807
SERTAD2
64.8
65
65.2
65.4
65.6
Position on chr2 (Mb)

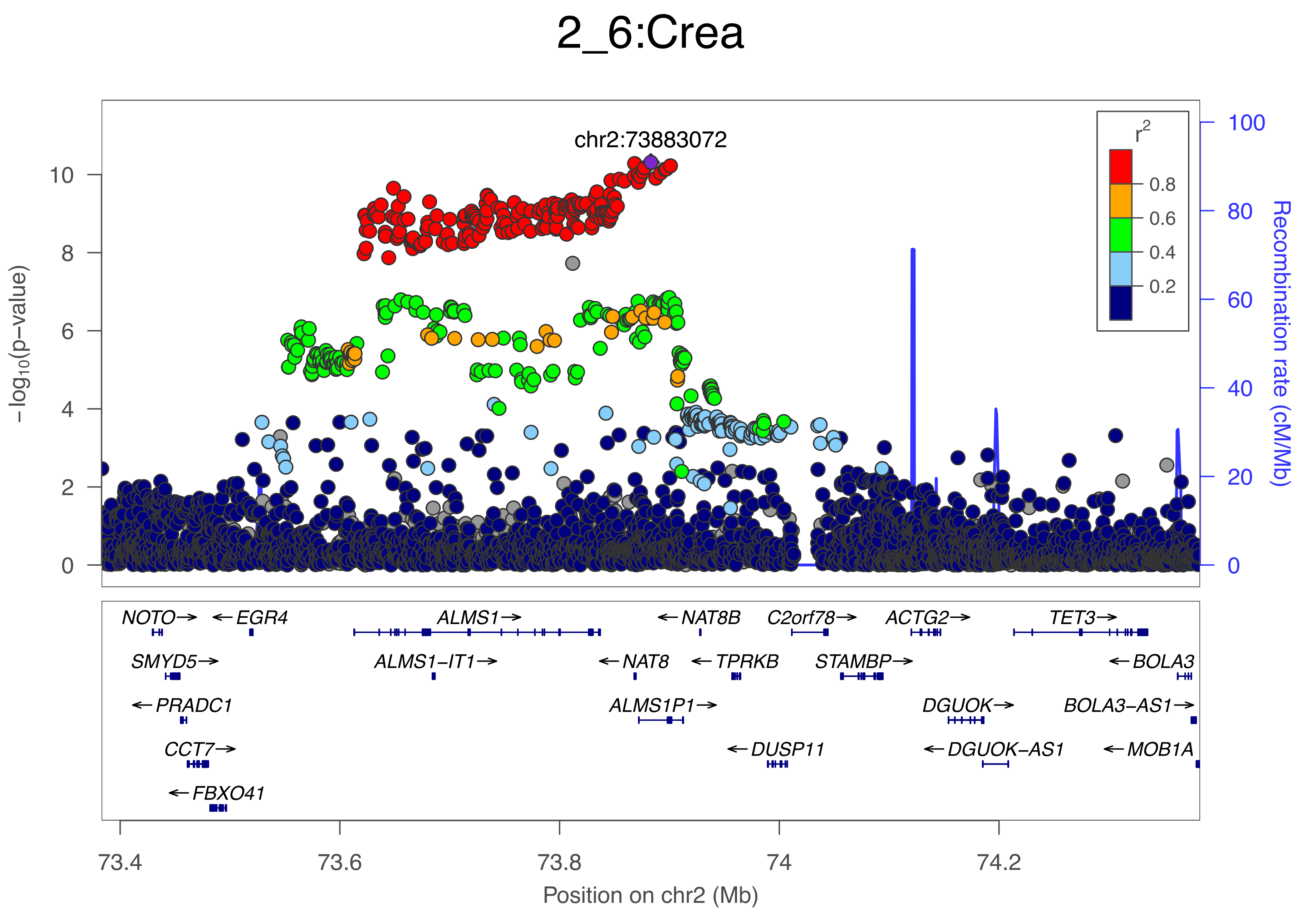

2_6:Crea
chr2:73883072
r2
0.8
0.6
0.4
0.2
−log10(p−value)
Recombination rate (cM/Mb)
NOTO→
←EGR4
ALMS1→
←NAT8B
C2orf78→
ACTG2→
TET3→
SMYD5→
ALMS1−IT1→
←NAT8
←TPRKB
STAMBP→
←BOLA3
←PRADC1
ALMS1P1→
DGUOK→
BOLA3−AS1→
CCT7→
←DUSP11
←DGUOK−AS1
←MOB1A
←FBXO41
73.4
73.6
73.8
74
74.2
Position on chr2 (Mb)

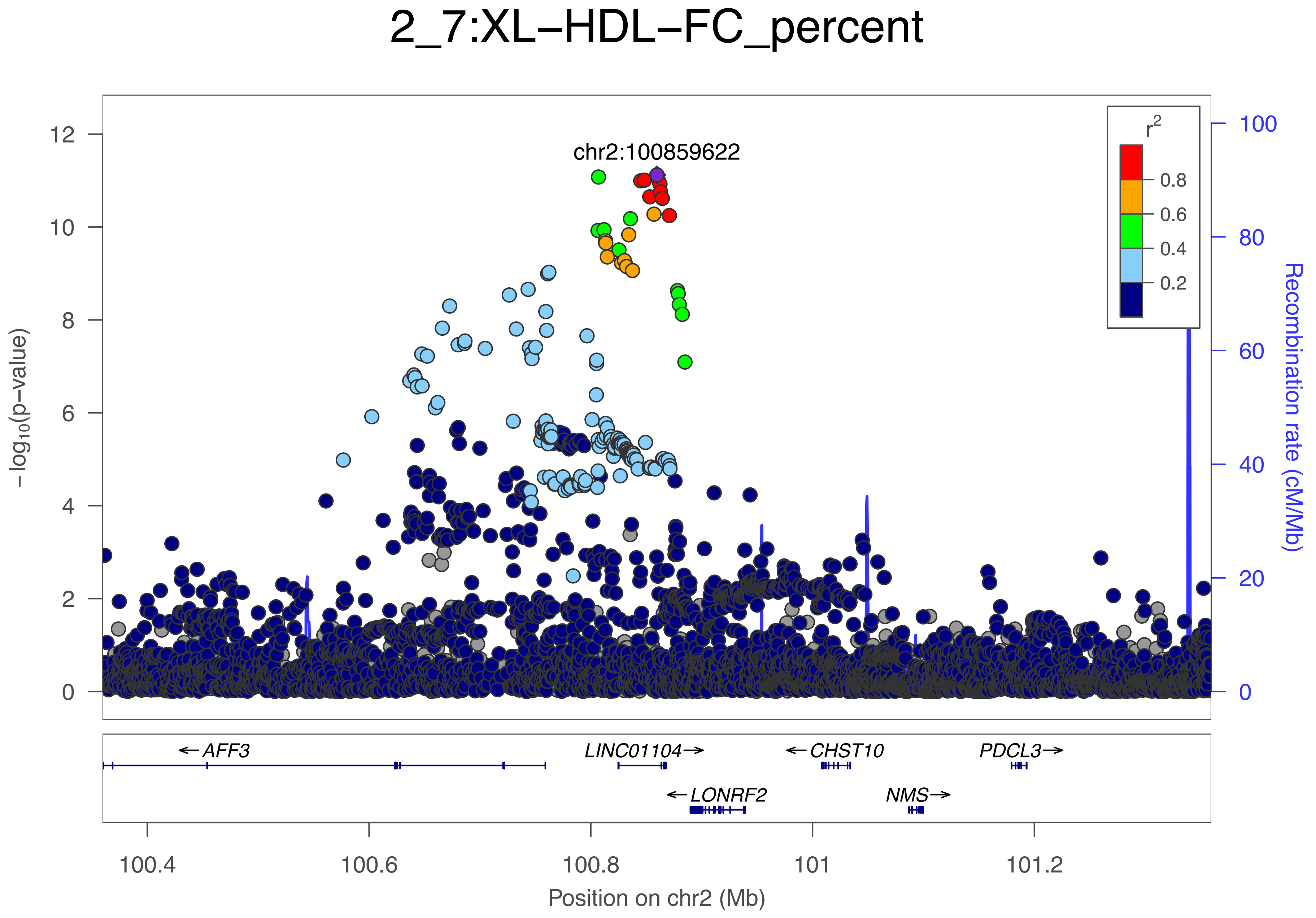

2_7:XL−HDL−FC_percent
chr2:100859622
r2
0.8
0.6
0.4
0.2
12
10
8
6
4
2
0
−log10(p−value)
100
80
60
40
20
0
Recombination rate (cM/Mb)
←AFF3
LINC01104→
←CHST10
PDCL3→
←LONRF2
NMS→
100.4
100.6
100.8
101
101.2
Position on chr2 (Mb)

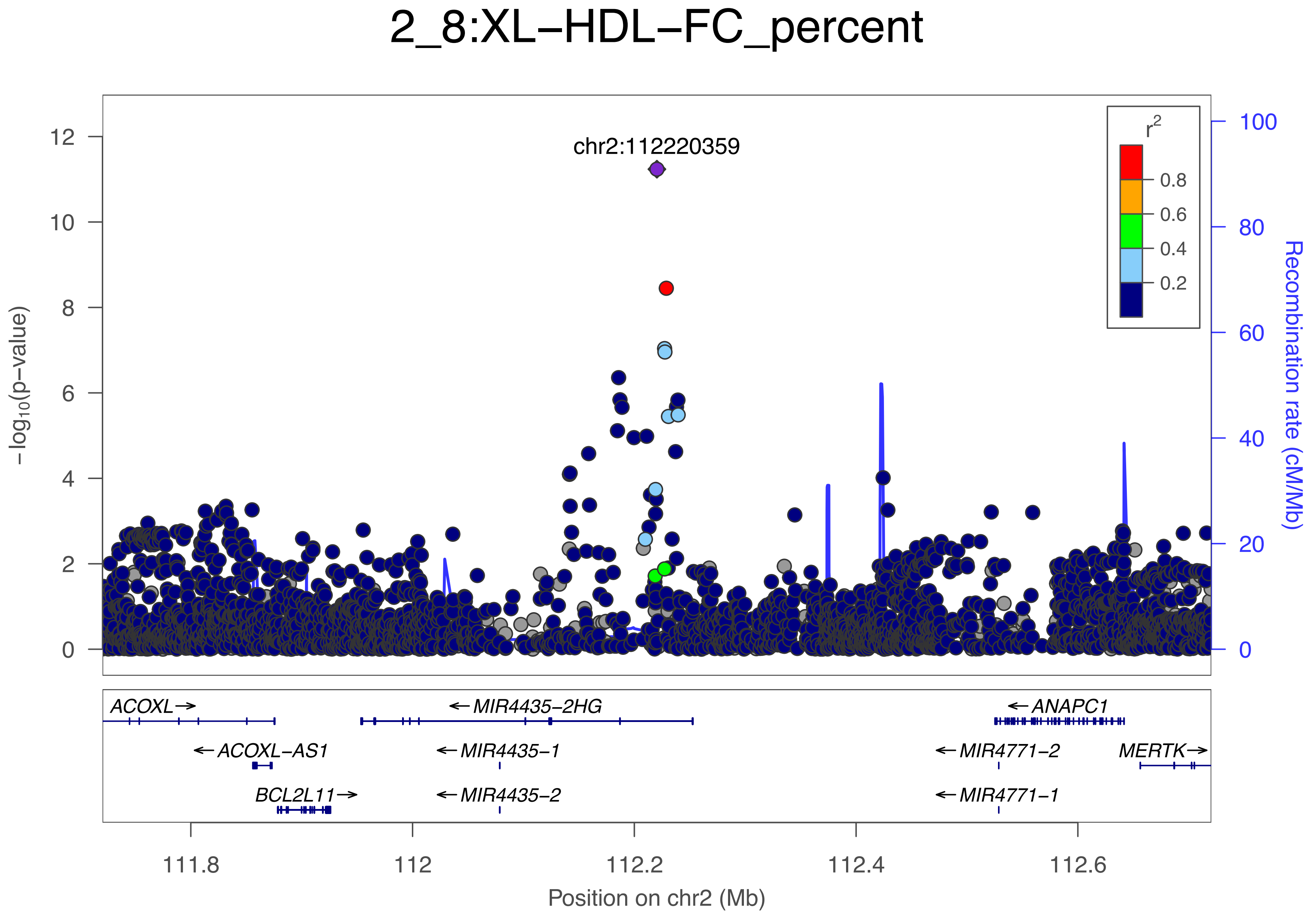

2_8:XL−HDL−FC_percent
chr2:112220359
r2
0.8
0.6
0.4
0.2
−log10(p−value)
Recombination rate (cM/Mb)
ACOXL→
←MIR4435−2HG
←ANAPC1
←ACOXL−AS1
←MIR4435−1
←MIR4771−2
MERTK→
BCL2L11→
←MIR4435−2
←MIR4771−1
111.8
112
112.2
112.4
112.6
Position on chr2 (Mb)

### 2_9:GlycA

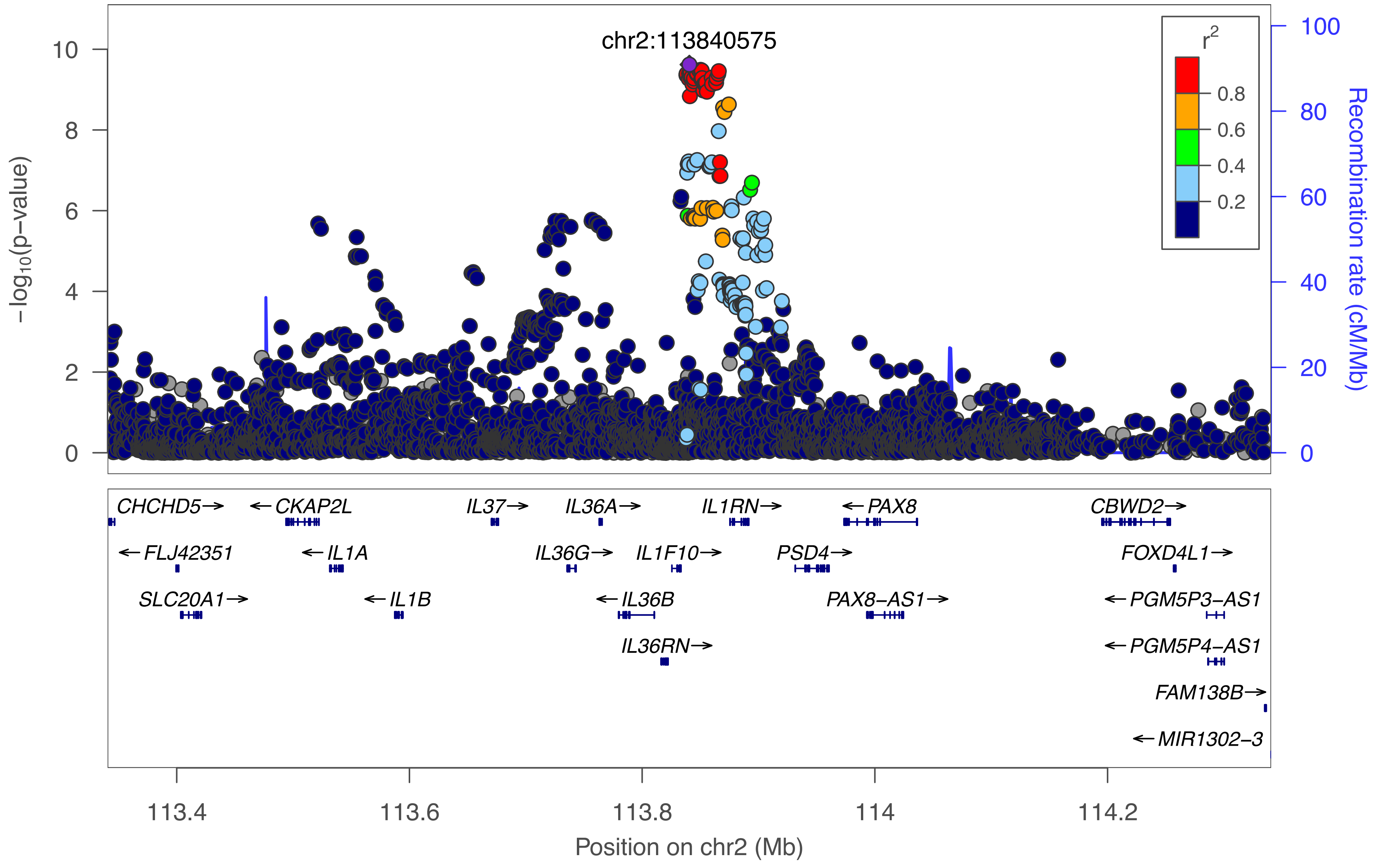

chr2:113840575
r²
0.8
0.6
0.4
0.2
−log10(p−value)
Recombination rate (cM/Mb)
0
2
4
6
8
10
0
20
40
60
80
100
CHCHD5→
←CKAP2L
IL37→
IL36A→
IL1RN→
←PAX8
CBWD2→
←FLJ42351
←IL1A
IL36G→
IL1F10→
PSD4→
FOXD4L1→
SLC20A1→
←IL1B
←IL36B
PAX8−AS1→
←PGM5P3−AS1
IL36RN→
←PGM5P4−AS1
FAM138B→
←MIR1302−3
113.4
113.6
113.8
114
114.2
Position on chr2 (Mb)

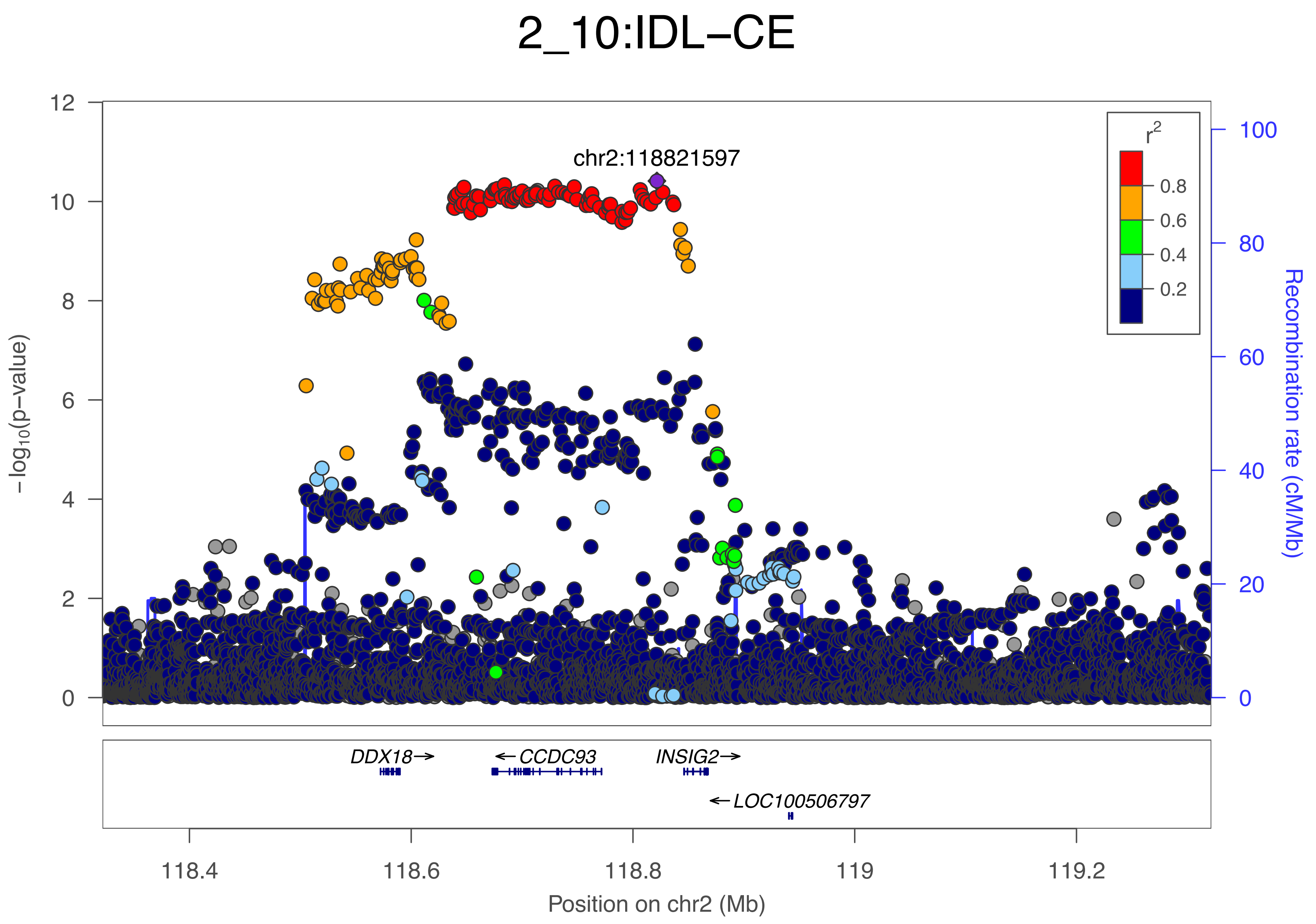

2_10:IDL−CE
chr2:118821597
−log10(p−value)
Recombination rate (cM/Mb)
r2
0.8
0.6
0.4
0.2
DDX18→
←CCDC93
INSIG2→
←LOC100506797
118.4
118.6
118.8
119
119.2
Position on chr2 (Mb)

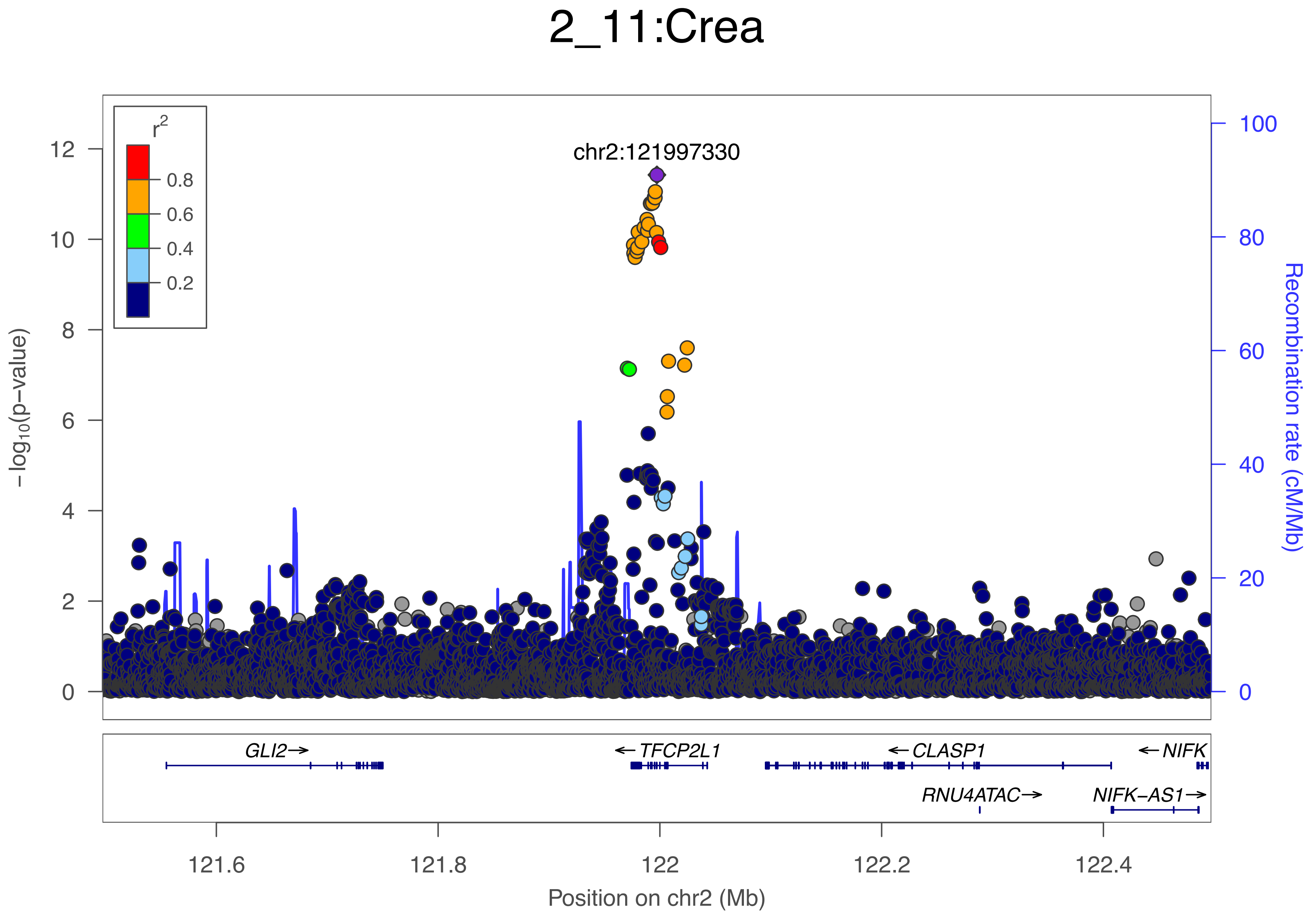
2_11:Crea
r2
0.8
0.6
0.4
0.2
chr2:121997330
−log10(p−value)
Recombination rate (cM/Mb)
GLI2→
←TFCP2L1
←CLASP1
←NIFK
RNU4ATAC→
NIFK−AS1→
121.6
121.8
122
122.2
122.4
Position on chr2 (Mb)

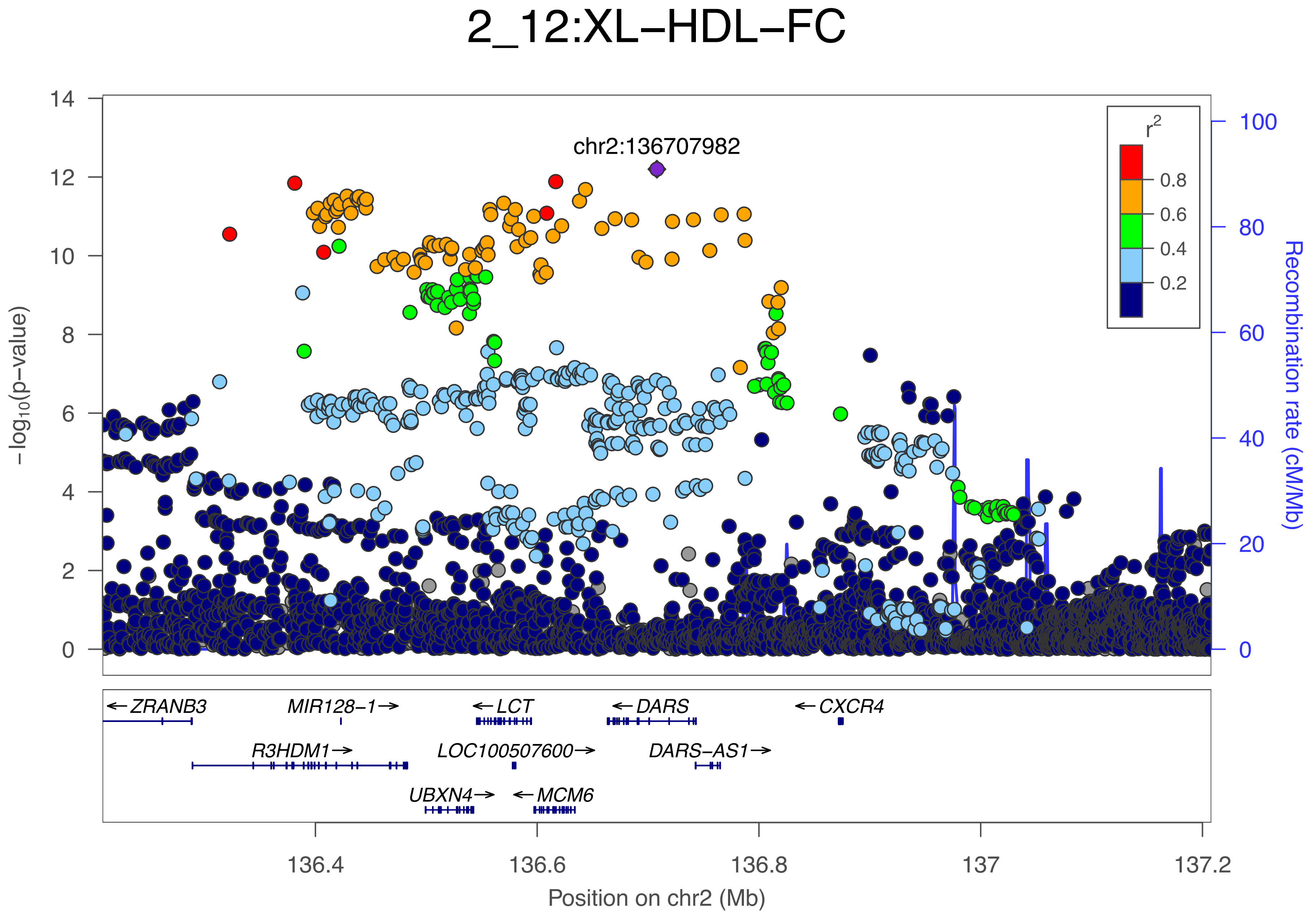

2_12:XL–HDL–FC
chr2:136707982
r2
0.8
0.6
0.4
0.2
–log10(p–value)
Recombination rate (cM/Mb)
ZRANB3
MIR128–1
LCT
DARS
CXCR4
R3HDM1
LOC100507600
DARS–AS1
UBXN4
MCM6
136.4
136.6
136.8
137
137.2
Position on chr2 (Mb)

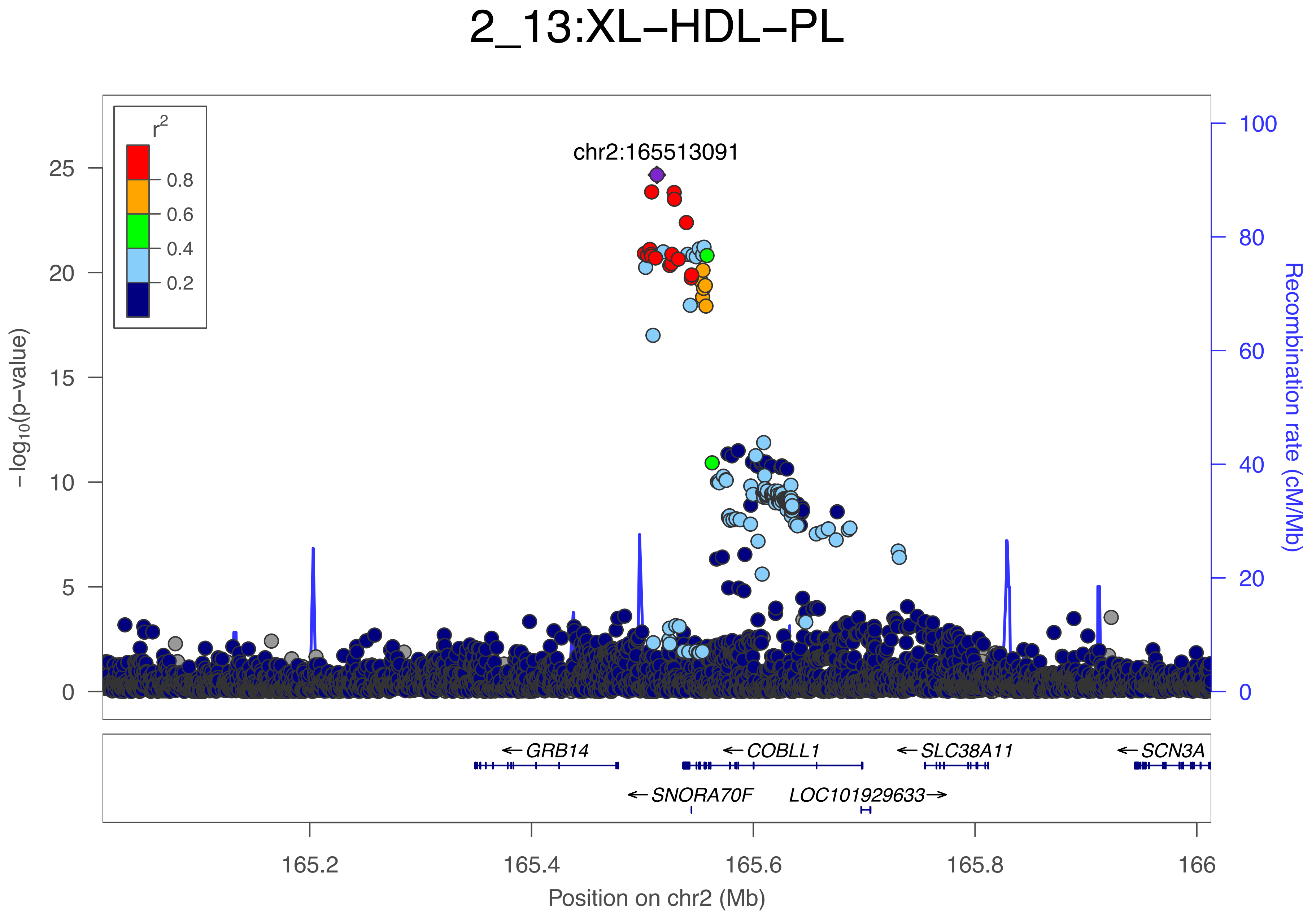
2_13:XL−HDL−PL
r2
0.8
0.6
0.4
0.2
chr2:165513091
−log10(p−value)
25
20
15
10
5
0
Recombination rate (cM/Mb)
100
80
60
40
20
0
← GRB14
← COBLL1
← SLC38A11
← SCN3A
← SNORA70F
LOC101929633 →
165.2
165.4
165.6
165.8
166
Position on chr2 (Mb)

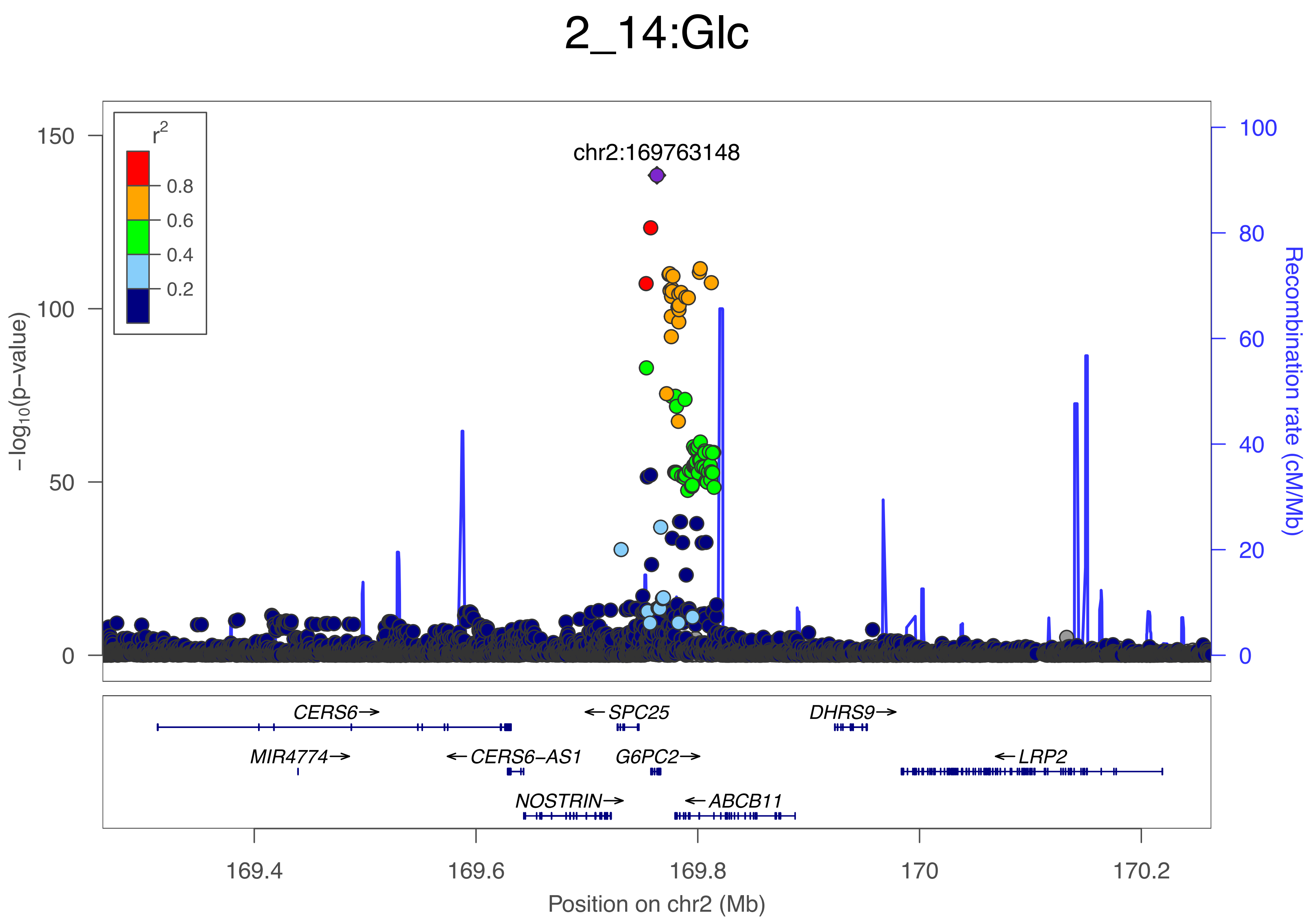

2_14:Glc
r2
0.8
0.6
0.4
0.2
chr2:169763148
150
100
50
0
−log10(p−value)
100
80
60
40
20
0
Recombination rate (cM/Mb)
CERS6→
←SPC25
DHRS9→
MIR4774→
←CERS6−AS1
G6PC2→
←LRP2
NOSTRIN→
←ABCB11
169.4
169.6
169.8
170
170.2
Position on chr2 (Mb)

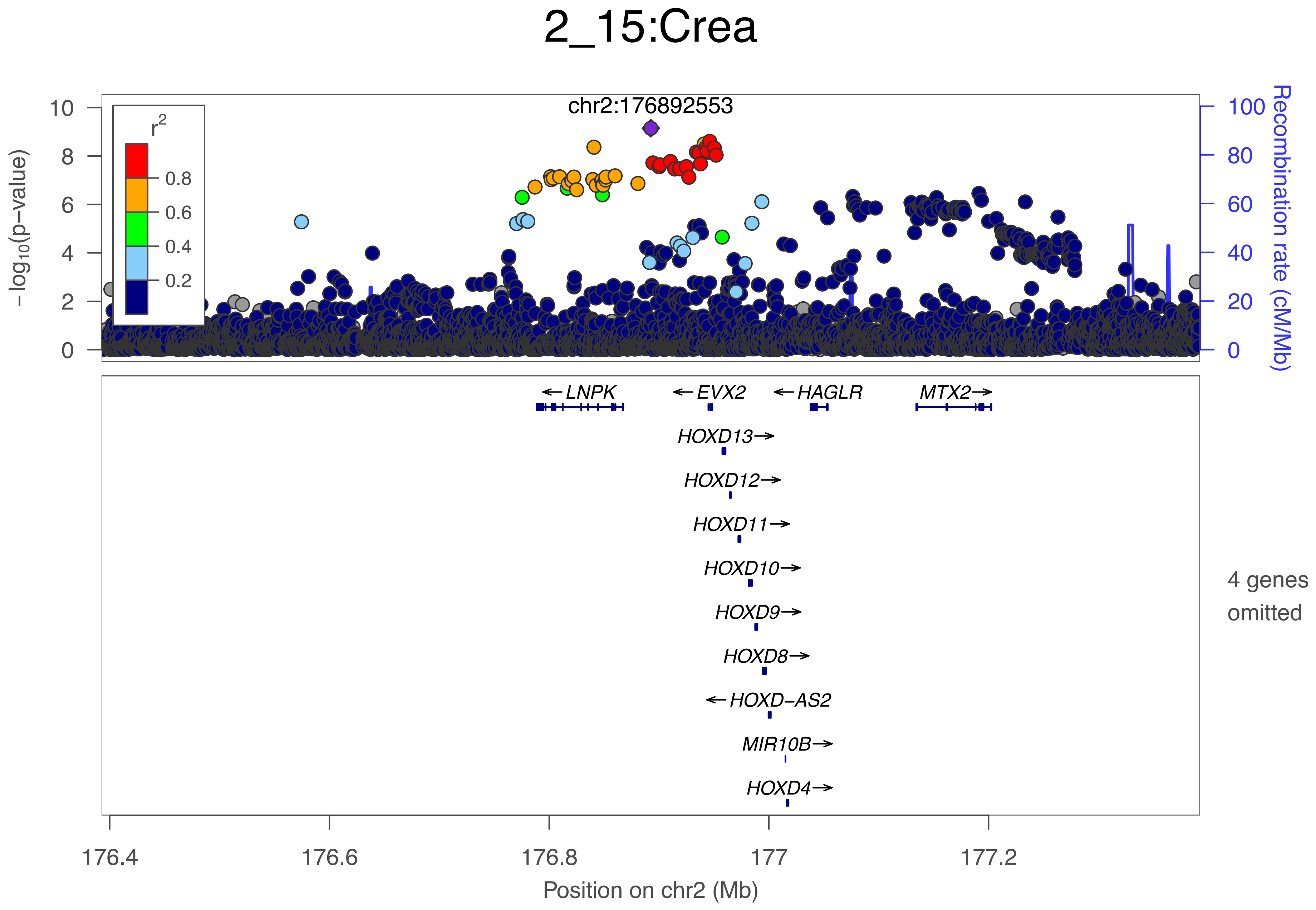
2_15:Crea
chr2:176892553
r2
0.8
0.6
0.4
0.2
−log10(p−value)
10
8
6
4
2
0
100
80
60
40
20
0
Recombination rate (cM/Mb)
← LNPK
← EVX2
← HAGLR
MTX2→
HOXD13→
HOXD12→
HOXD11→
HOXD10→
HOXD9→
HOXD8→
← HOXD−AS2
MIR10B→
HOXD4→
4 genes omitted
176.4
176.6
176.8
177
177.2
Position on chr2 (Mb)

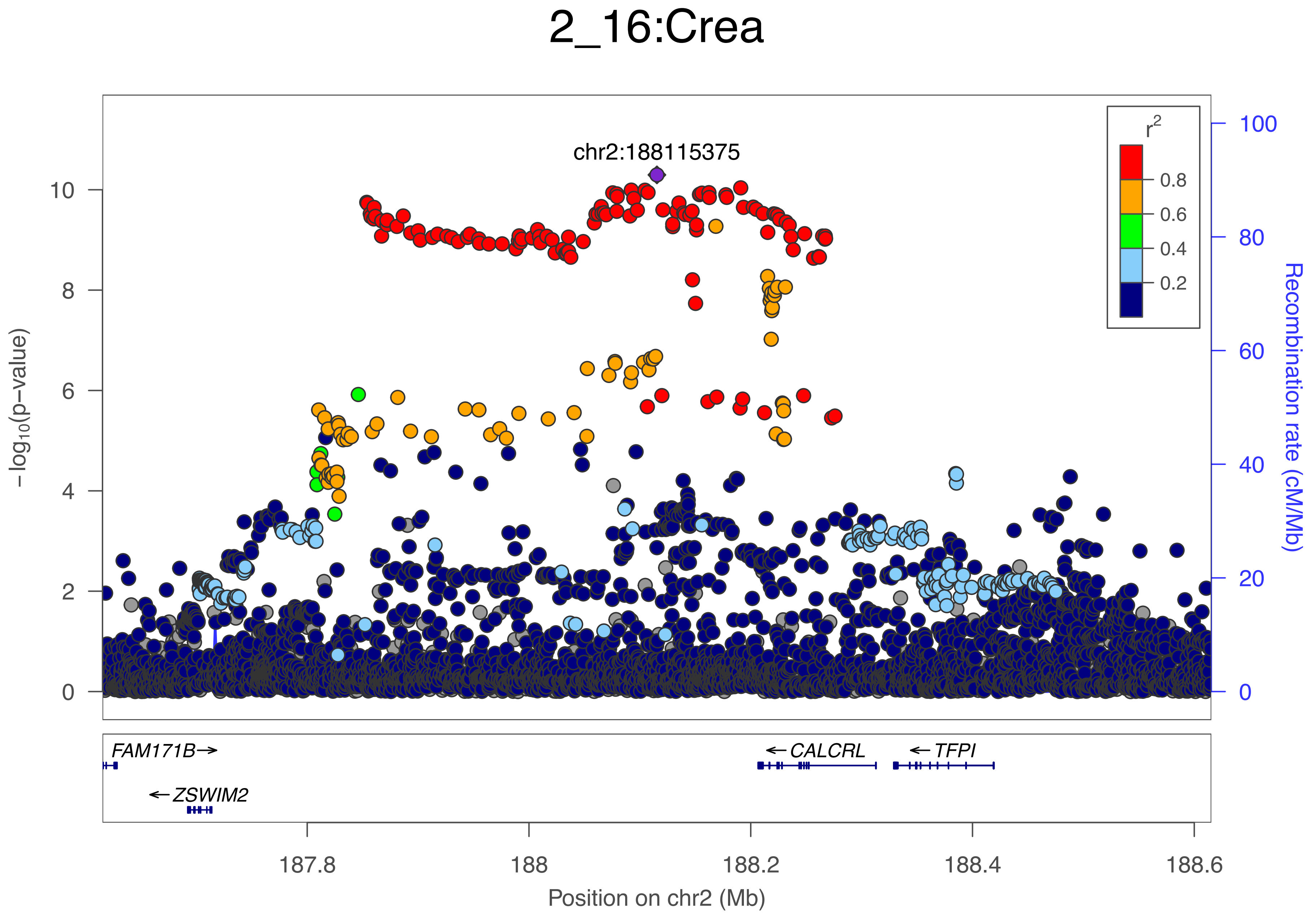

2_16:Crea
chr2:188115375
r2
0.8
0.6
0.4
0.2
−log10(p−value)
Recombination rate (cM/Mb)
FAM171B→
←ZSWIM2
←CALCRL
←TFPI
187.8
188
188.2
188.4
188.6
Position on chr2 (Mb)

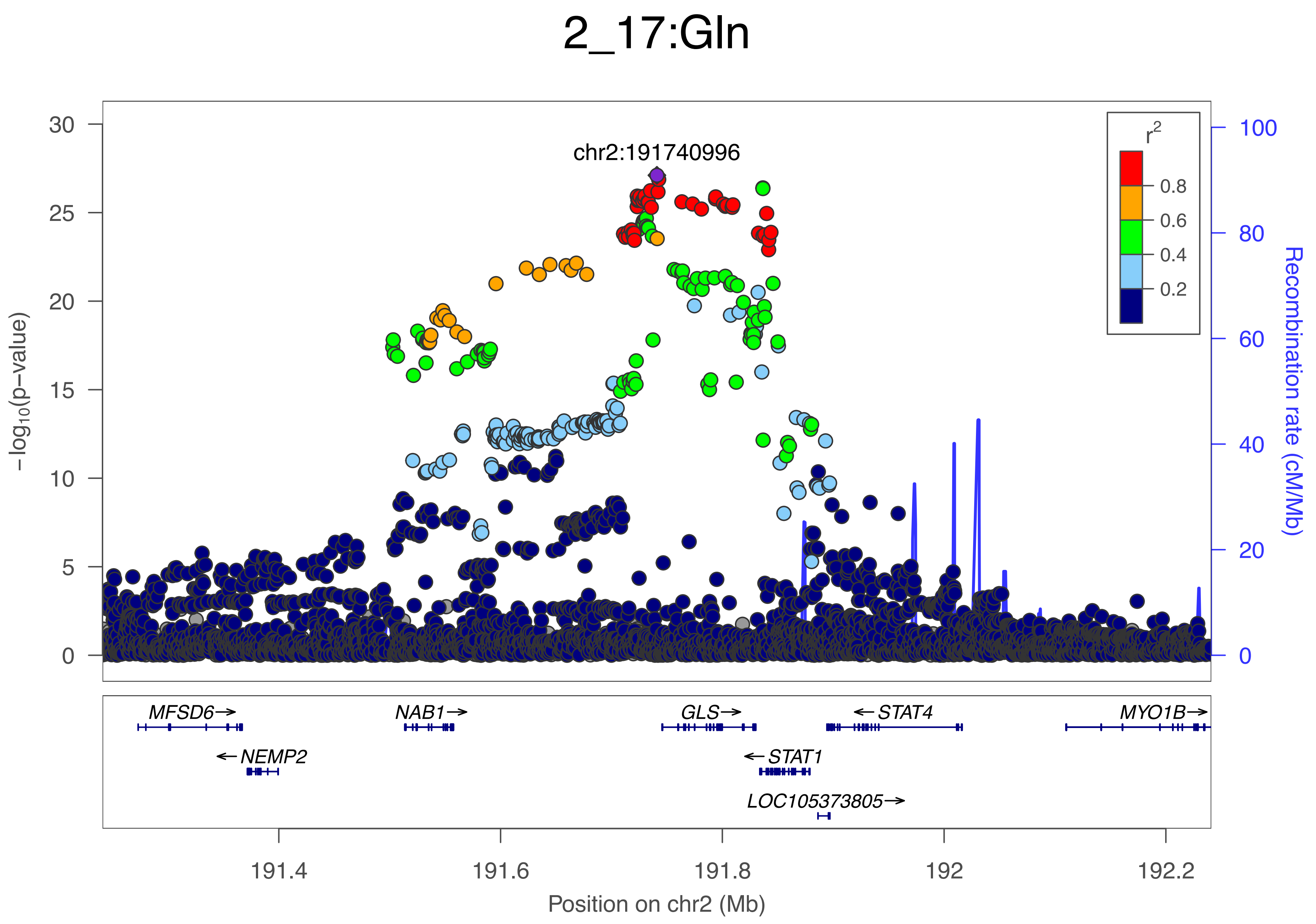

2_17:Gln
chr2:191740996
r2
0.8
0.6
0.4
0.2
30
25
20
15
10
5
0
−log10(p−value)
100
80
60
40
20
0
Recombination rate (cM/Mb)
MFSD6→
←NEMP2
NAB1→
GLS→
←STAT1
LOC105373805→
←STAT4
MYO1B→
191.4
191.6
191.8
192
192.2
Position on chr2 (Mb)

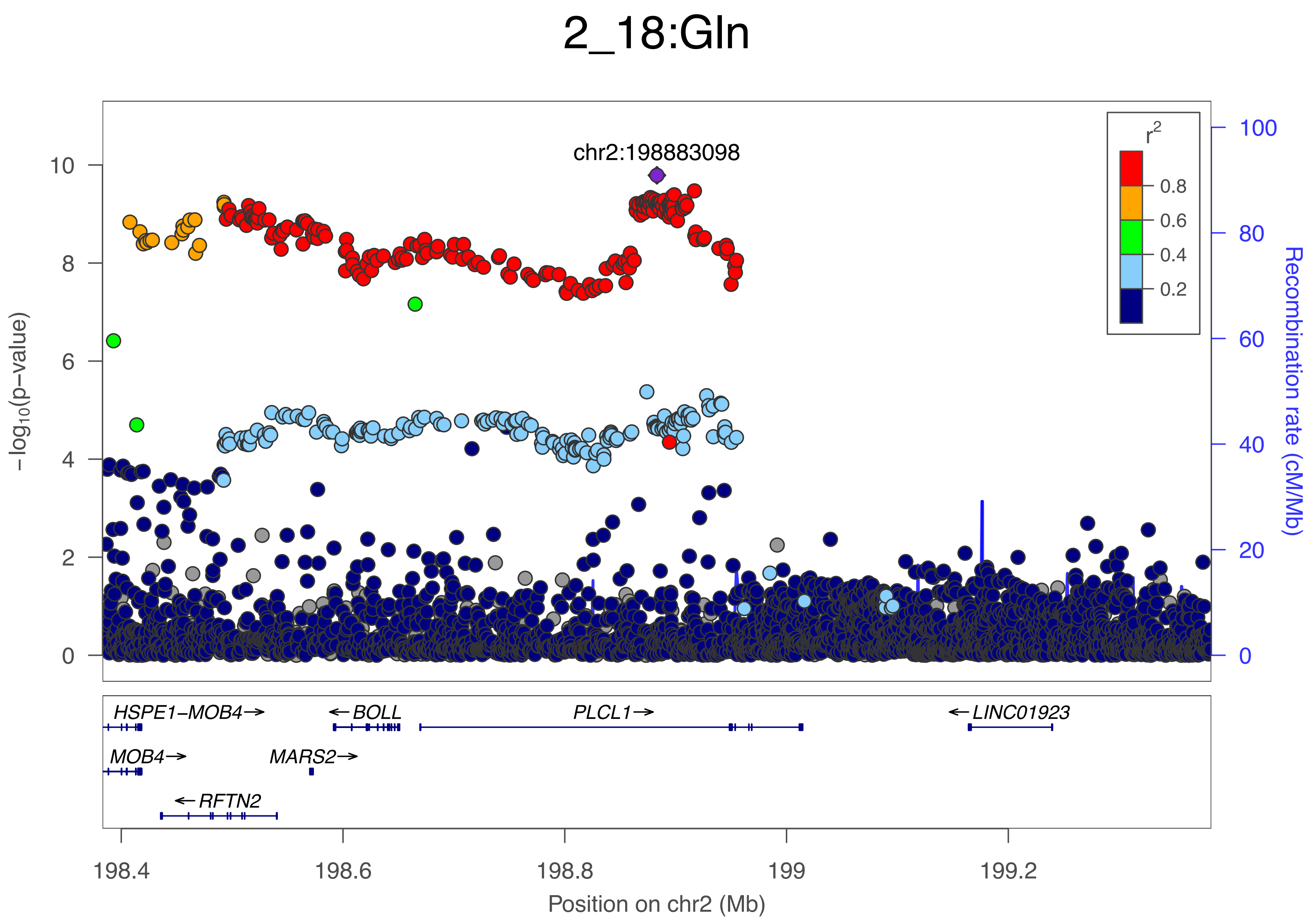

2_18:Gln
chr2:198883098
r²
0.8
0.6
0.4
0.2
−log10(p−value)
Recombination rate (cM/Mb)
HSPE1−MOB4→
←BOLL
PLCL1→
←LINC01923
MOB4→
MARS2→
←RFTN2
198.4
198.6
198.8
199
199.2
Position on chr2 (Mb)

### 2_19:PUFA

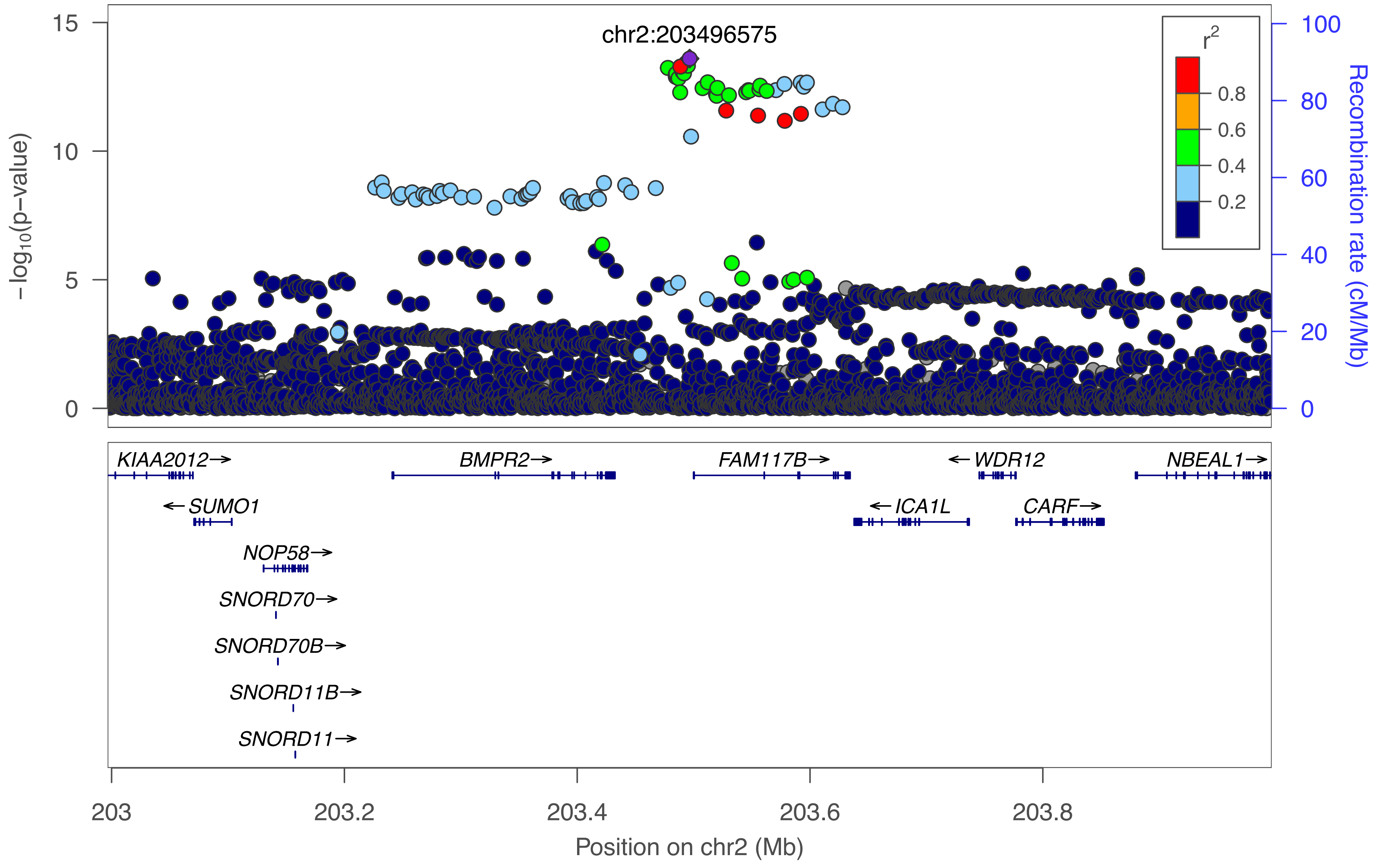

chr2:203496575
r²
0.8
0.6
0.4
0.2
−log10(p−value)
15
10
5
0
Recombination rate (cM/Mb)
100
80
60
40
20
0
KIAA2012→
BMPR2→
FAM117B→
←WDR12
NBEAL1→
←SUMO1
←ICA1L
CARF→
NOP58→
SNORD70→
SNORD70B→
SNORD11B→
SNORD11→
203
203.2
203.4
203.6
203.8
Position on chr2 (Mb)

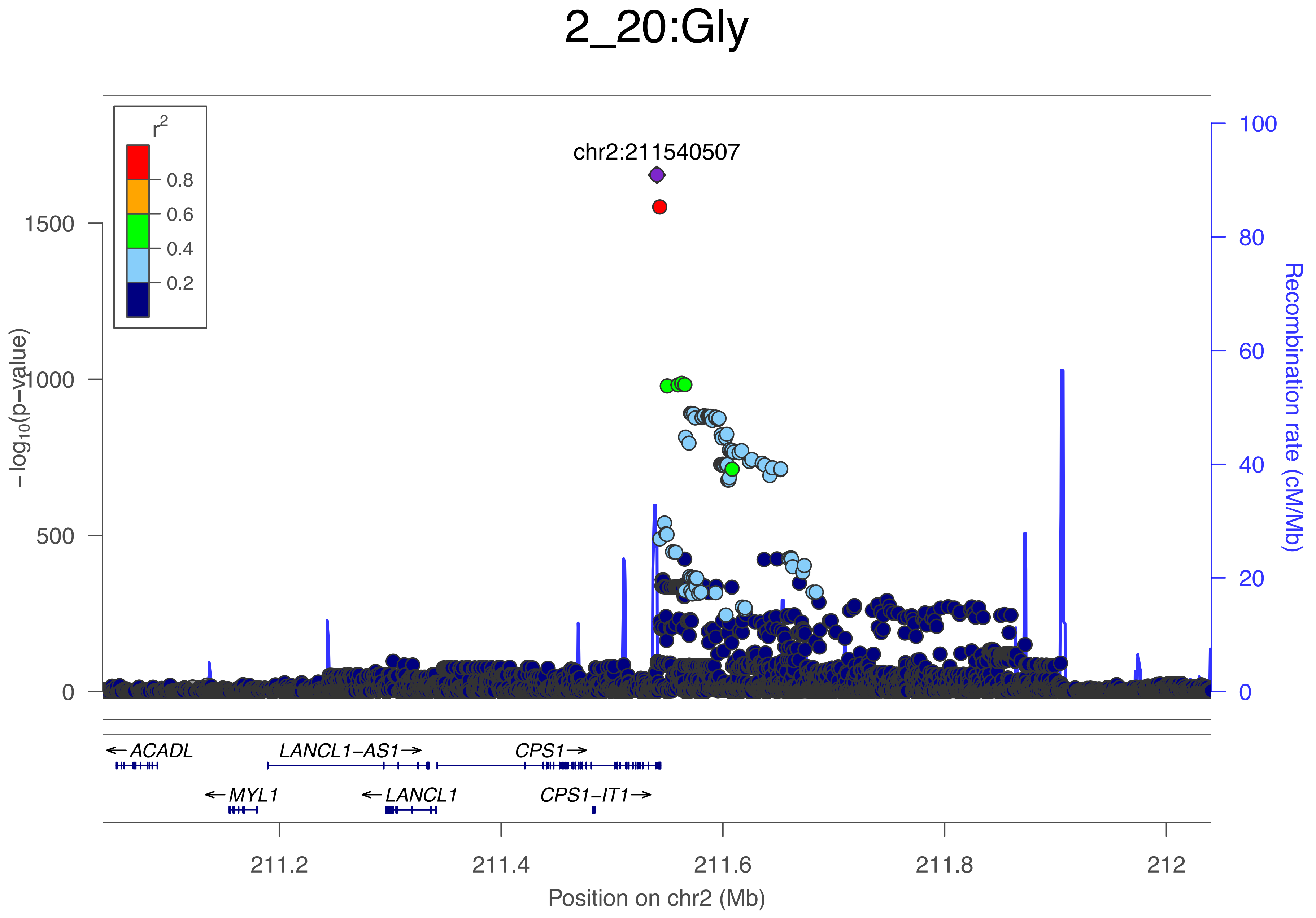

2_20:Gly
r2
0.8
0.6
0.4
0.2
chr2:211540507
1500
1000
500
0
−log10(p−value)
100
80
60
40
20
0
Recombination rate (cM/Mb)
←ACADL
LANCL1−AS1→
CPS1→
←MYL1
←LANCL1
CPS1−IT1→
211.2
211.4
211.6
211.8
212
Position on chr2 (Mb)

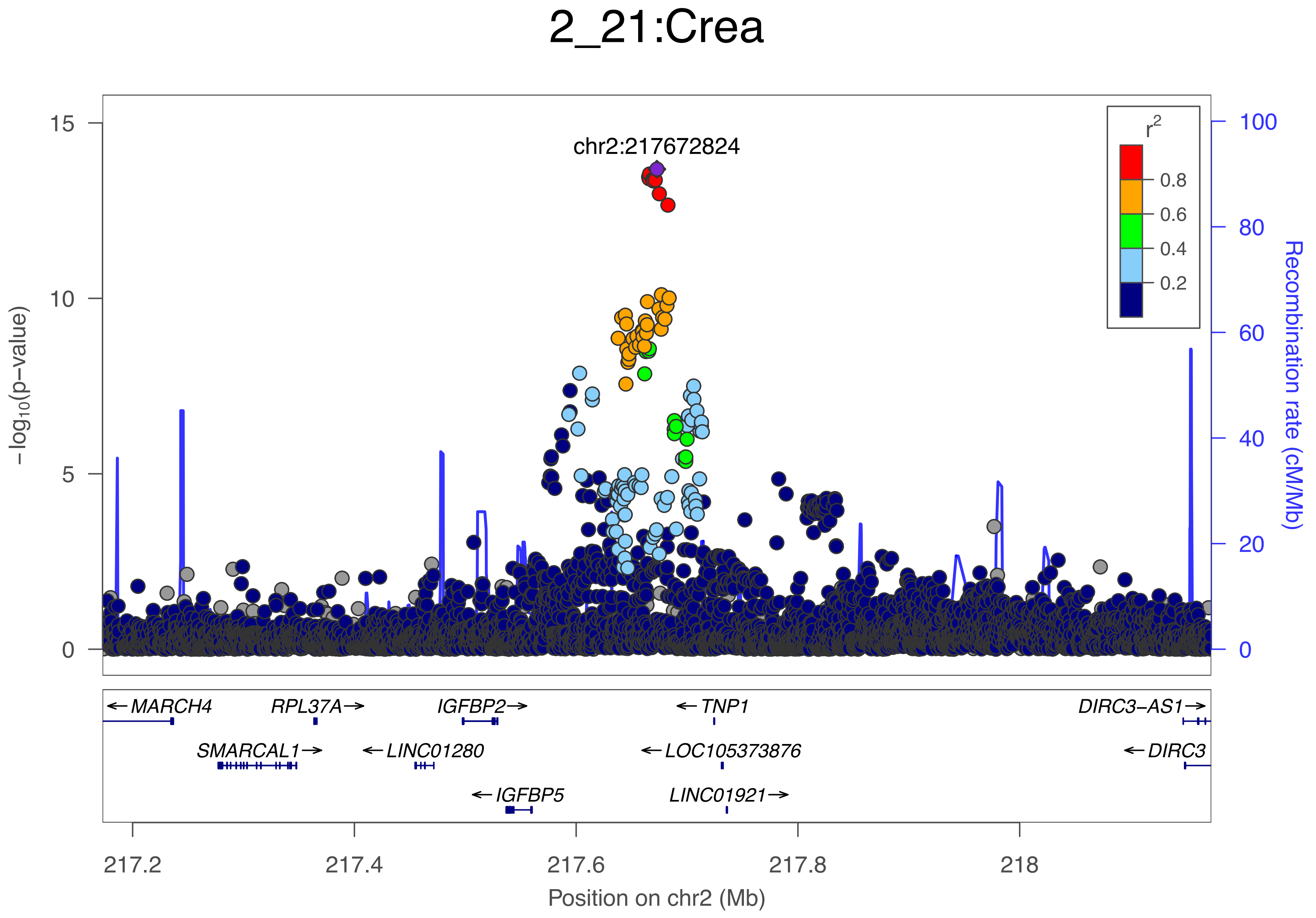

2_21:Crea
chr2:217672824
r2
0.8
0.6
0.4
0.2
15
10
5
0
100
80
60
40
20
0
−log10(p−value)
Recombination rate (cM/Mb)
←MARCH4
RPL37A→
IGFBP2→
←TNP1
DIRC3−AS1→
SMARCAL1→
←LINC01280
←LOC105373876
←DIRC3
←IGFBP5
LINC01921→
217.2
217.4
217.6
217.8
218
Position on chr2 (Mb)

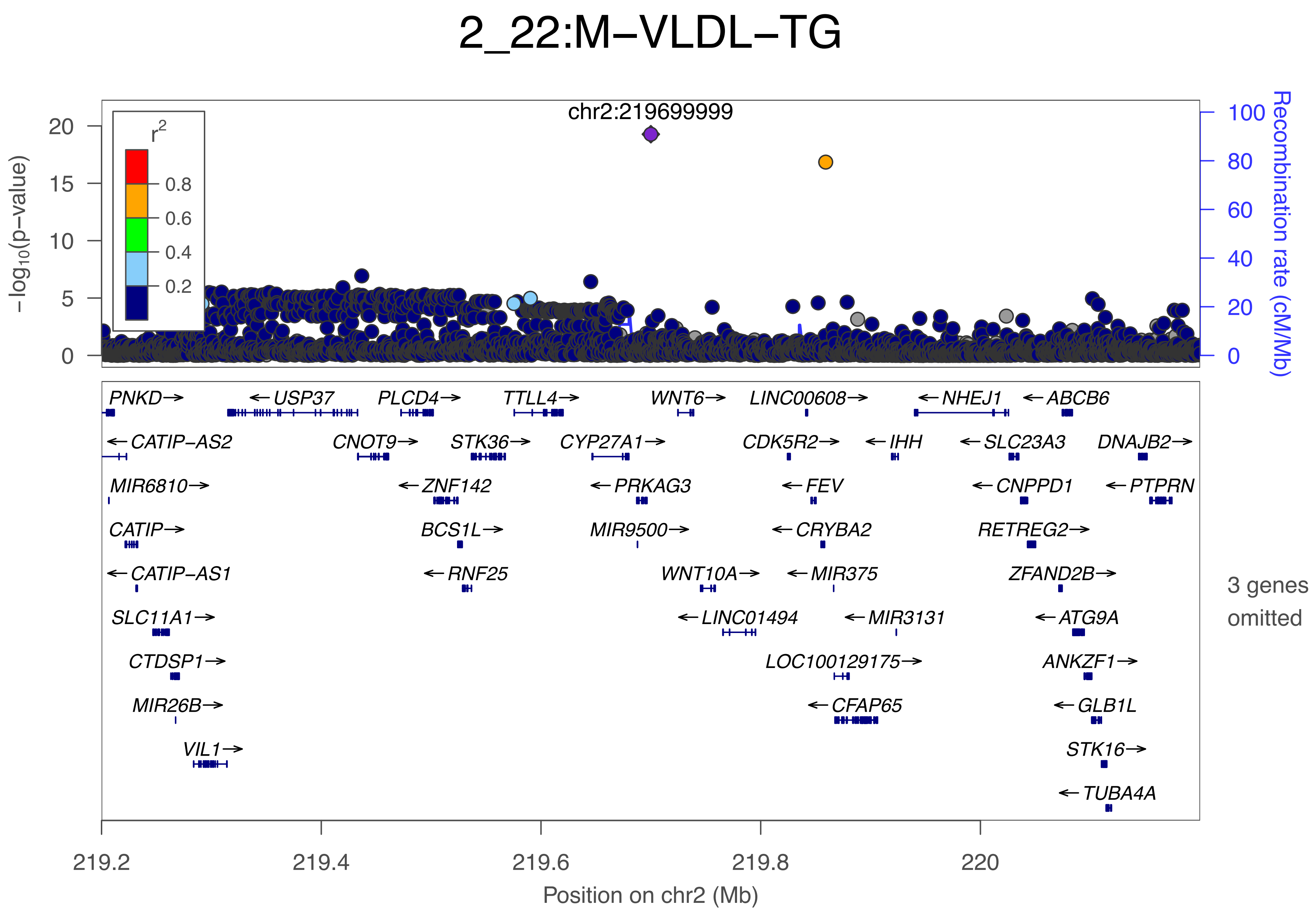
2_22:M−VLDL−TG
chr2:219699999
r2
0.8
0.6
0.4
0.2
−log10(p−value)
20
15
10
5
0
Recombination rate (cM/Mb)
100
80
60
40
20
0
PNKD→
←USP37
PLCD4→
TTLL4→
WNT6→
LINC00608→
←NHEJ1
←ABCB6
←CATIP−AS2
CNOT9→
STK36→
CYP27A1→
CDK5R2→
←IHH
←SLC23A3
DNAJB2→
MIR6810→
←ZNF142
←PRKAG3
←FEV
←CNPPD1
←PTPRN
CATIP→
BCS1L→
MIR9500→
←CRYBA2
RETREG2→
←CATIP−AS1
←RNF25
WNT10A→
←MIR375
ZFAND2B→
SLC11A1→
←LINC01494
←MIR3131
←ATG9A
CTDSP1→
LOC100129175→
ANKZF1→
MIR26B→
←CFAP65
←GLB1L
VIL1→
STK16→
←TUBA4A
3 genes omitted
219.2
219.4
219.6
219.8
220
Position on chr2 (Mb)

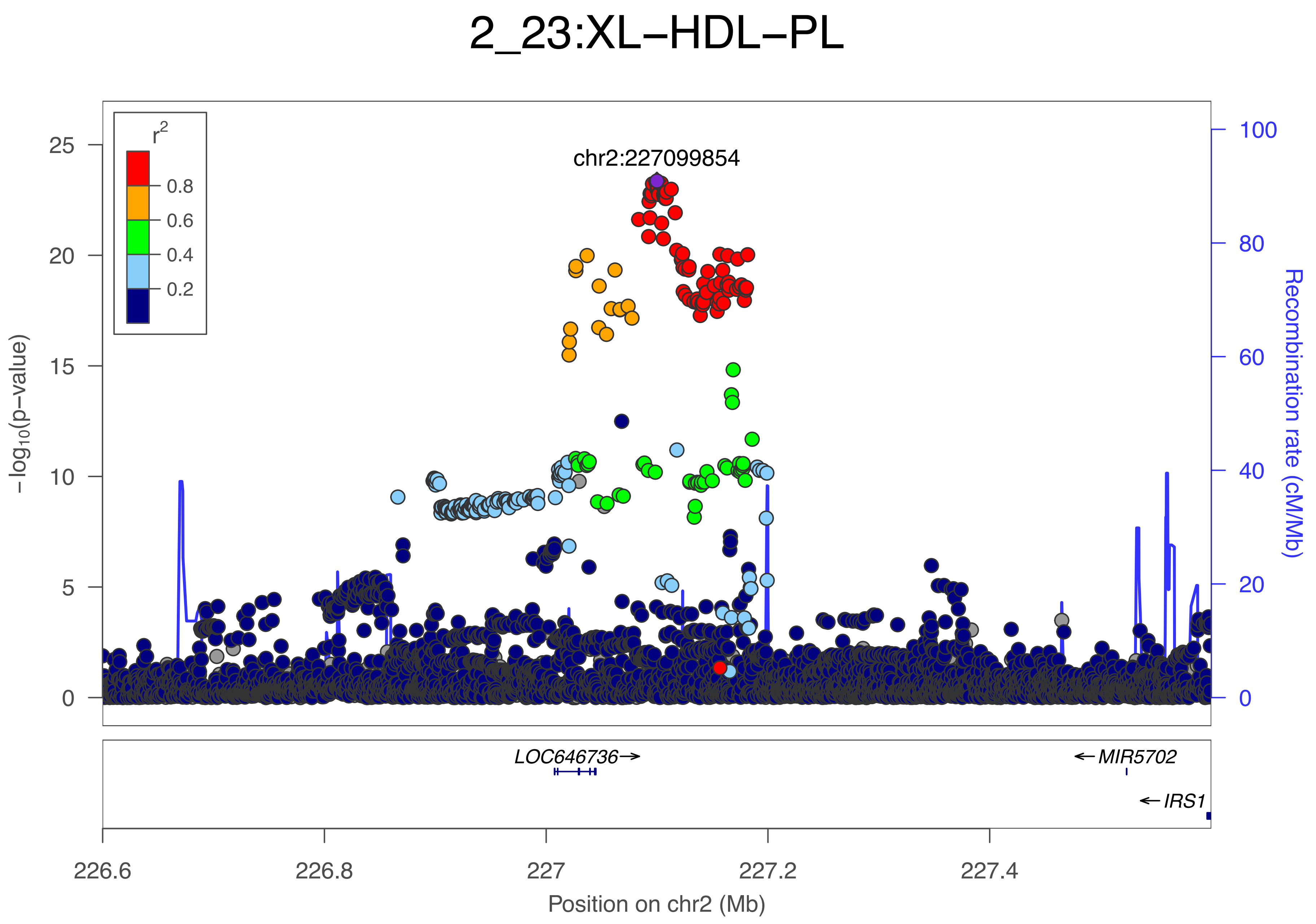

2_23:XL−HDL−PL
r2
0.8
0.6
0.4
0.2
chr2:227099854
−log10(p−value)
Recombination rate (cM/Mb)
25
20
15
10
5
0
100
80
60
40
20
0
LOC646736→
←MIR5702
←IRS1
226.6
226.8
227
227.2
227.4
Position on chr2 (Mb)

### 3_1:EstC

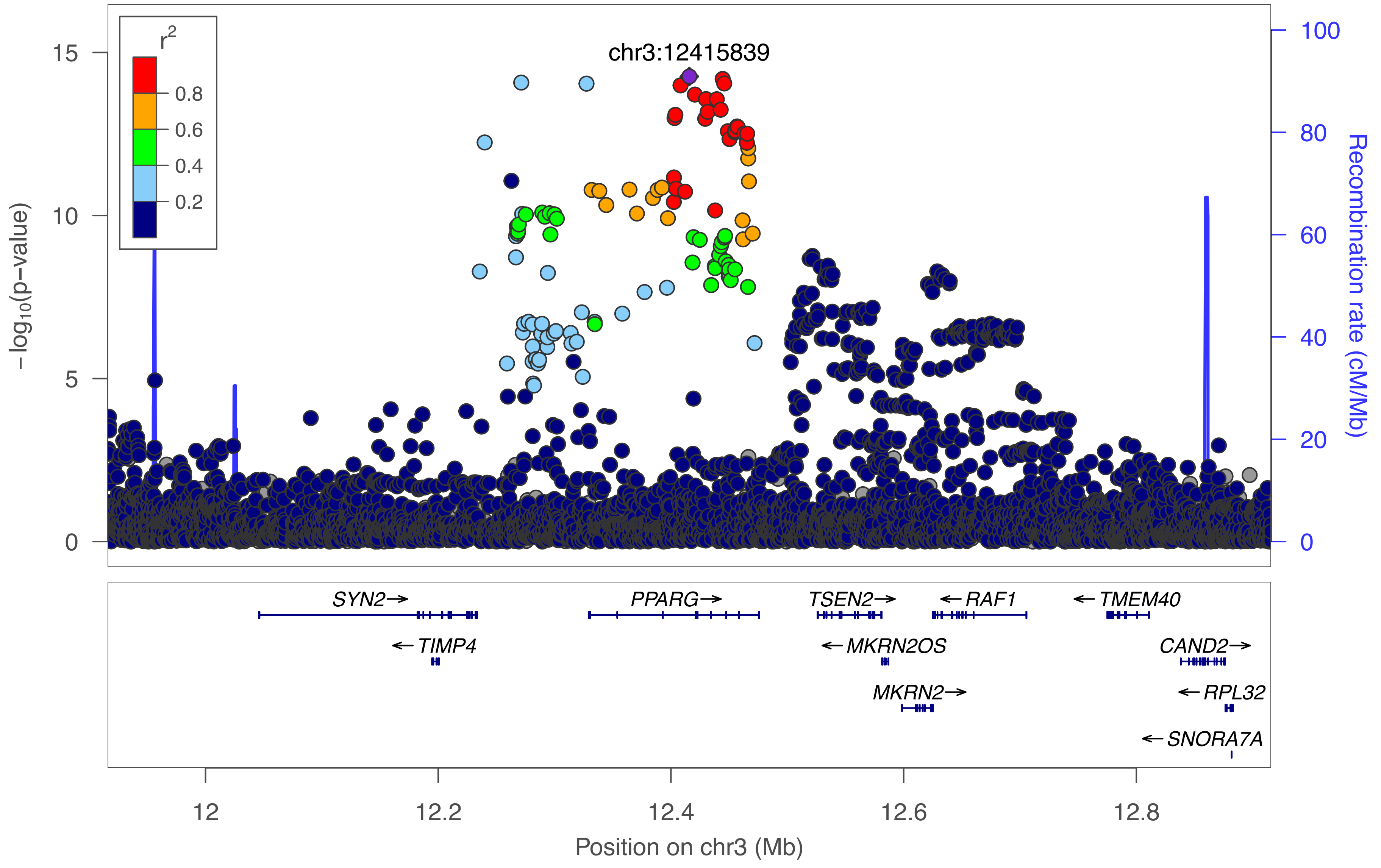

chr3:12415839
r2
0.8
0.6
0.4
0.2
15
10
5
0
−log10(p−value)
100
80
60
40
20
0
Recombination rate (cM/Mb)
SYN2→
TIMP4
PPARG→
TSEN2→
MKRN2OS
MKRN2→
RAF1
TMEM40
CAND2→
RPL32
SNORA7A
12
12.2
12.4
12.6
12.8
Position on chr3 (Mb)

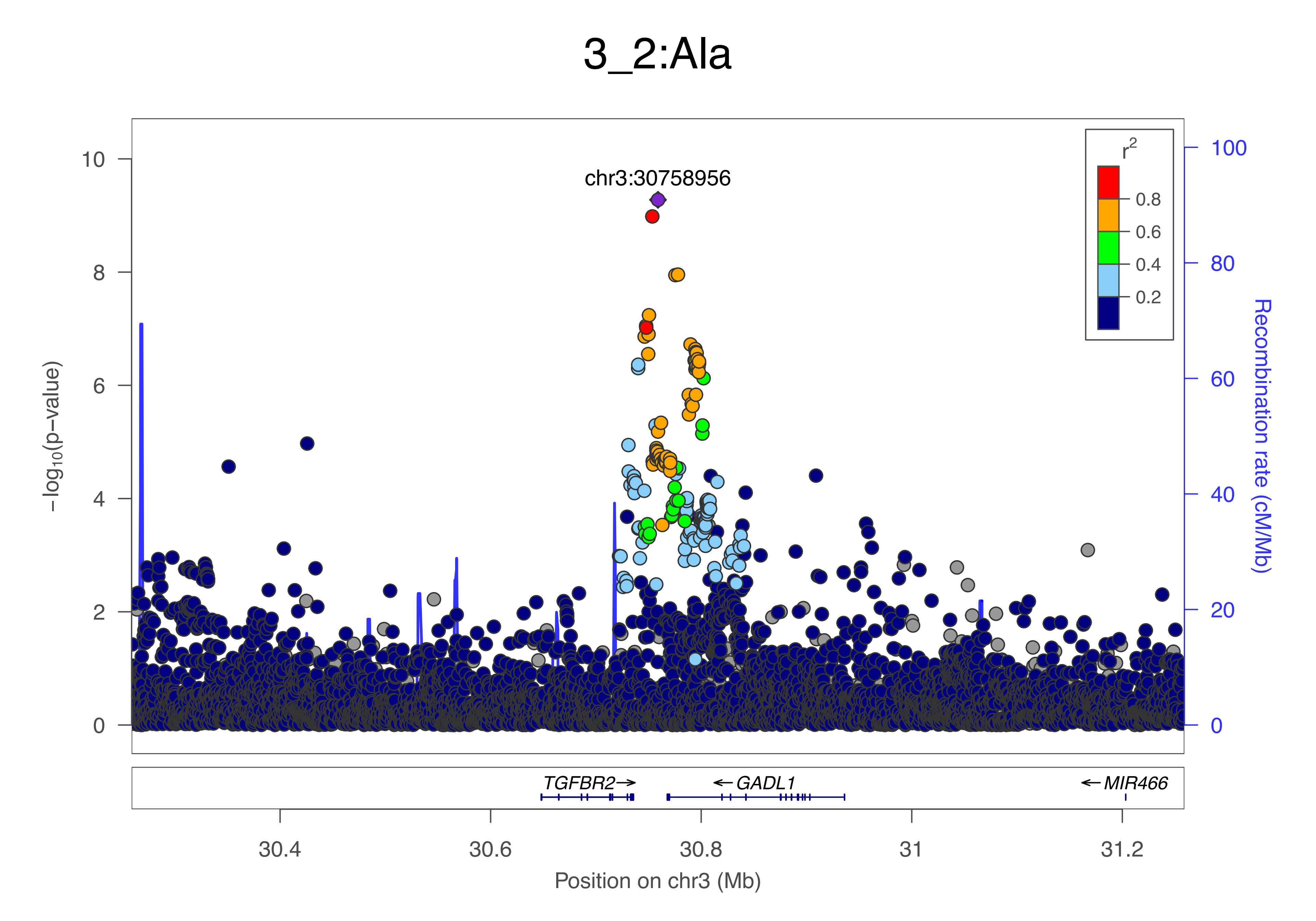

3_2:Ala
chr3:30758956
−log10(p−value)
Recombination rate (cM/Mb)
r2
0.8
0.6
0.4
0.2
10
8
6
4
2
0
100
80
60
40
20
0
TGFBR2→
←GADL1
←MIR466
30.4
30.6
30.8
31
31.2
Position on chr3 (Mb)

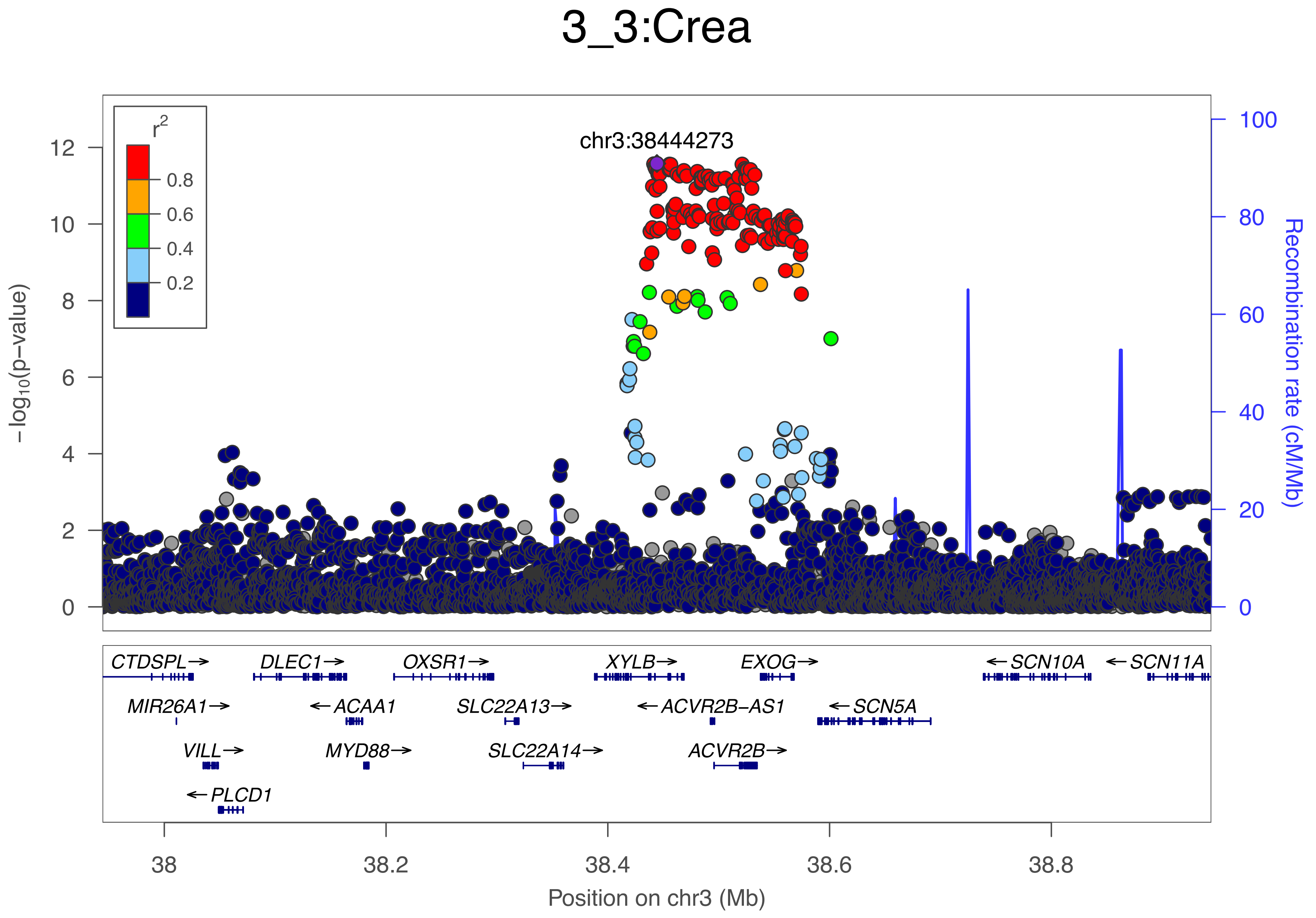
3_3:Crea
chr3:38444273
r2
0.8
0.6
0.4
0.2
−log10(p−value)
Recombination rate (cM/Mb)
0
2
4
6
8
10
12
0
20
40
60
80
100
CTDSPL→
DLEC1→
OXSR1→
XYLB→
EXOG→
←SCN10A
←SCN11A
MIR26A1→
←ACAA1
SLC22A13→
←ACVR2B−AS1
←SCN5A
VILL→
MYD88→
SLC22A14→
ACVR2B→
←PLCD1
38
38.2
38.4
38.6
38.8
Position on chr3 (Mb)

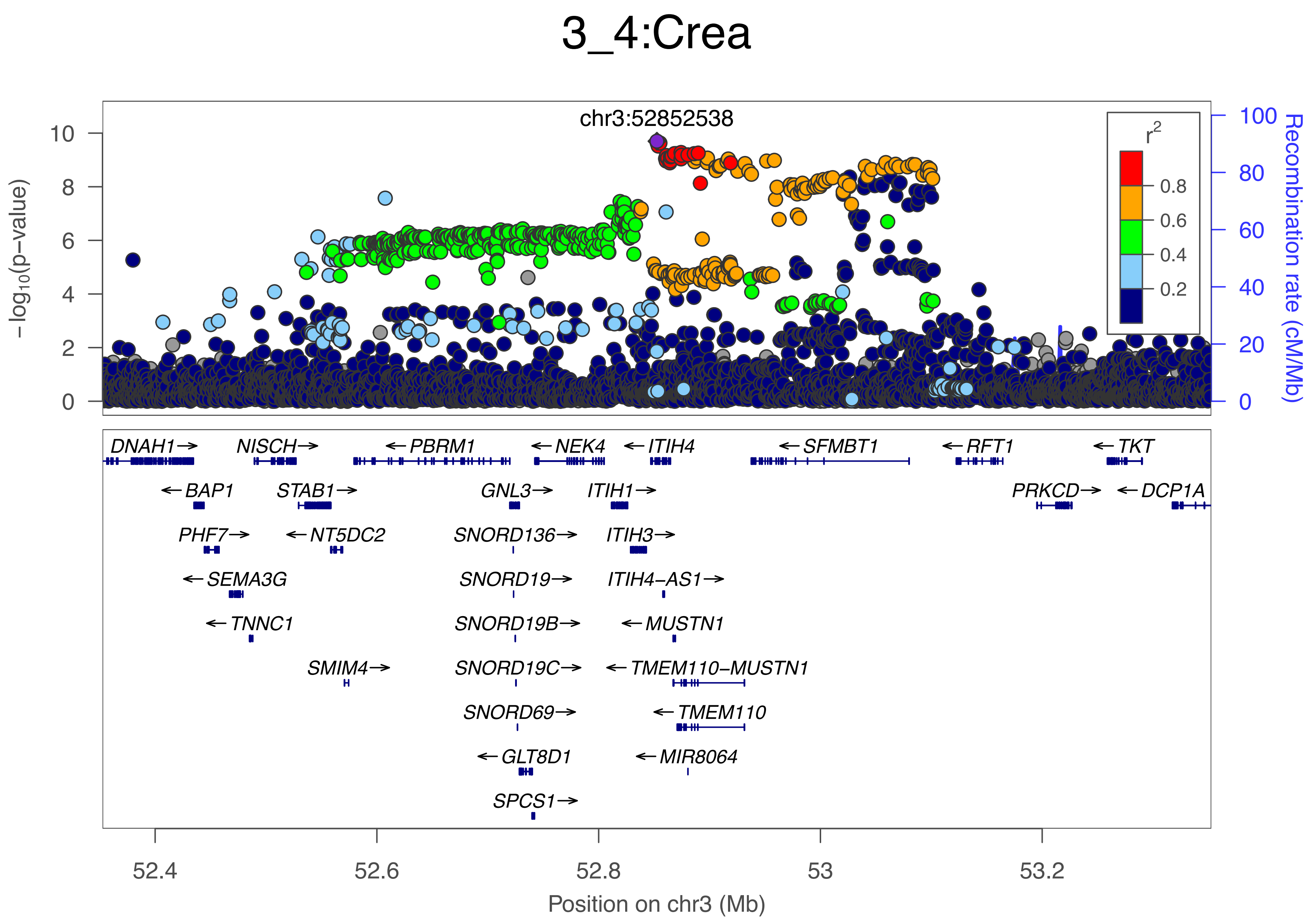

3_4:Crea
chr3:52852538
r²
0.8
0.6
0.4
0.2
−log10(p−value)
Recombination rate (cM/Mb)
DNAH1→
NISCH→
←PBRM1
←NEK4
←ITIH4
←SFMBT1
←RFT1
←TKT
←BAP1
STAB1→
GNL3→
ITIH1→
PRKCD→
←DCP1A
PHF7→
←NT5DC2
SNORD136→
ITIH3→
←SEMA3G
SNORD19→
ITIH4−AS1→
←TNNC1
SNORD19B→
←MUSTN1
SMIM4→
SNORD19C→
←TMEM110−MUSTN1
SNORD69→
←TMEM110
←GLT8D1
←MIR8064
SPCS1→
52.4
52.6
52.8
53
53.2
Position on chr3 (Mb)

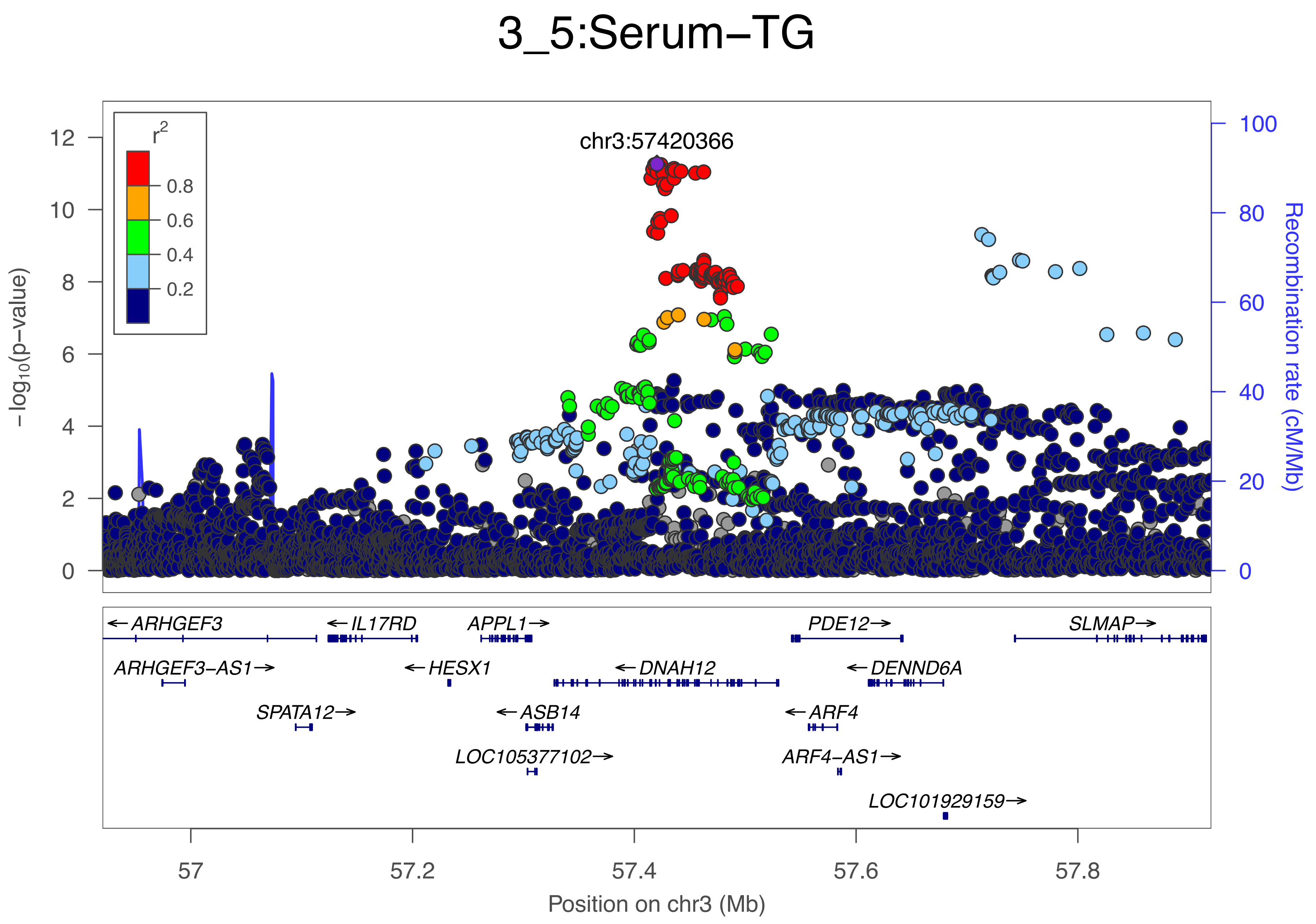

3_5:Serum−TG
chr3:57420366
r2
0.8
0.6
0.4
0.2
−log10(p−value)
Recombination rate (cM/Mb)
12
10
8
6
4
2
0
100
80
60
40
20
0
ARHGEF3
IL17RD
APPL1
PDE12
SLMAP
ARHGEF3−AS1
HESX1
DNAH12
DENND6A
SPATA12
ASB14
ARF4
LOC105377102
ARF4−AS1
LOC101929159
57
57.2
57.4
57.6
57.8
Position on chr3 (Mb)

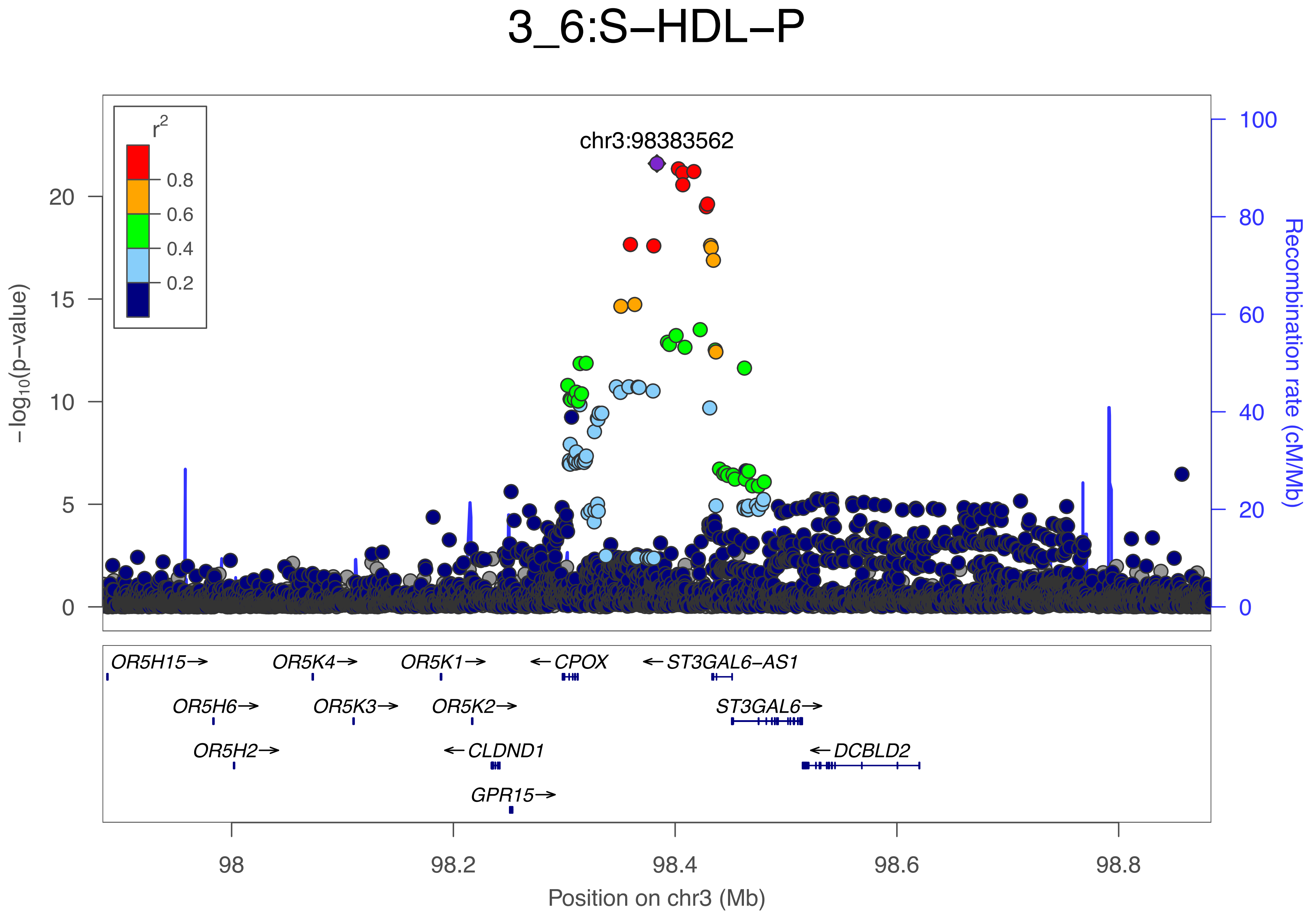

3_6:S−HDL−P
chr3:98383562
r2
0.8
0.6
0.4
0.2
−log10(p−value)
Recombination rate (cM/Mb)
0
5
10
15
20
0
20
40
60
80
100
OR5H15→
OR5K4→
OR5K1→
←CPOX
←ST3GAL6−AS1
OR5H6→
OR5K3→
OR5K2→
ST3GAL6→
OR5H2→
←CLDND1
←DCBLD2
GPR15→
98
98.2
98.4
98.6
98.8
Position on chr3 (Mb)

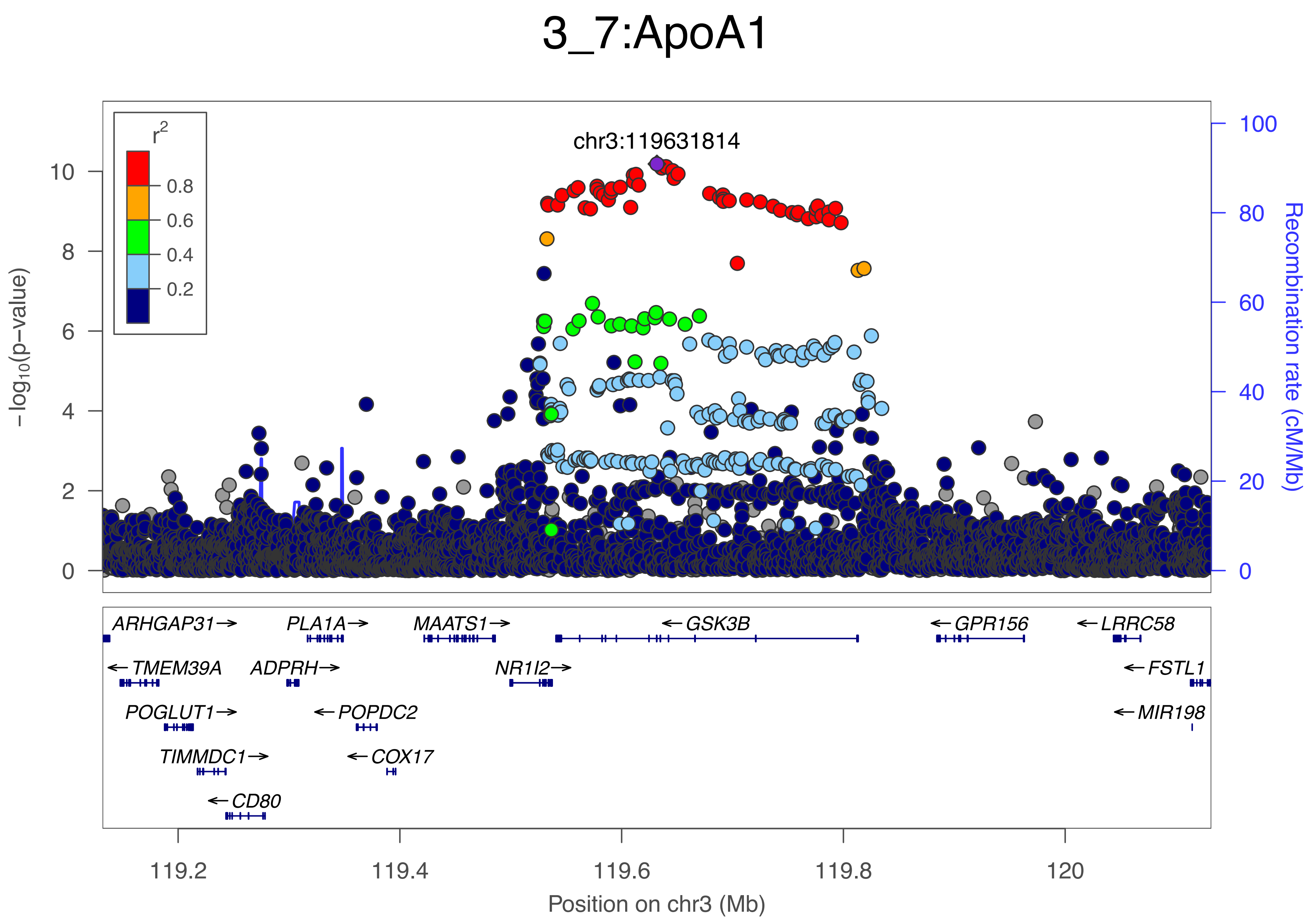
3_7:ApoA1
chr3:119631814
r2
0.8
0.6
0.4
0.2
−log10(p−value)
Recombination rate (cM/Mb)
ARHGAP31
PLA1A
MAATS1
GSK3B
GPR156
LRRC58
TMEM39A
ADPRH
NR1I2
FSTL1
POGLUT1
POPDC2
MIR198
TIMMDC1
COX17
CD80
119.2
119.4
119.6
119.8
120
Position on chr3 (Mb)

### 3_8:Ala

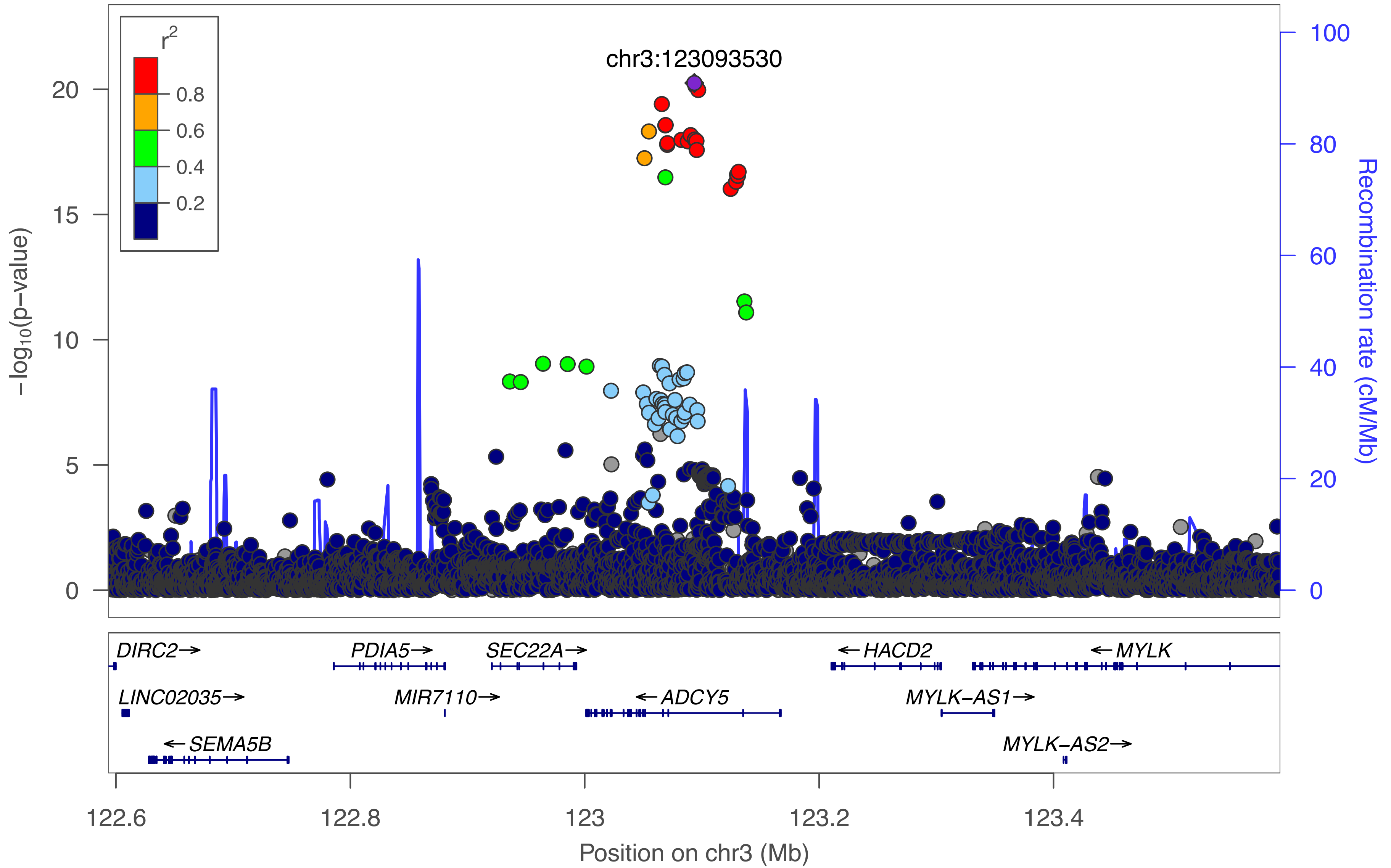

r2
0.8
0.6
0.4
0.2
chr3:123093530
−log10(p−value)
20
15
10
5
0
Recombination rate (cM/Mb)
100
80
60
40
20
0
DIRC2→
PDIA5→
SEC22A→
←HACD2
←MYLK
LINC02035→
MIR7110→
←ADCY5
MYLK−AS1→
←SEMA5B
MYLK−AS2→
122.6
122.8
123
123.2
123.4
Position on chr3 (Mb)

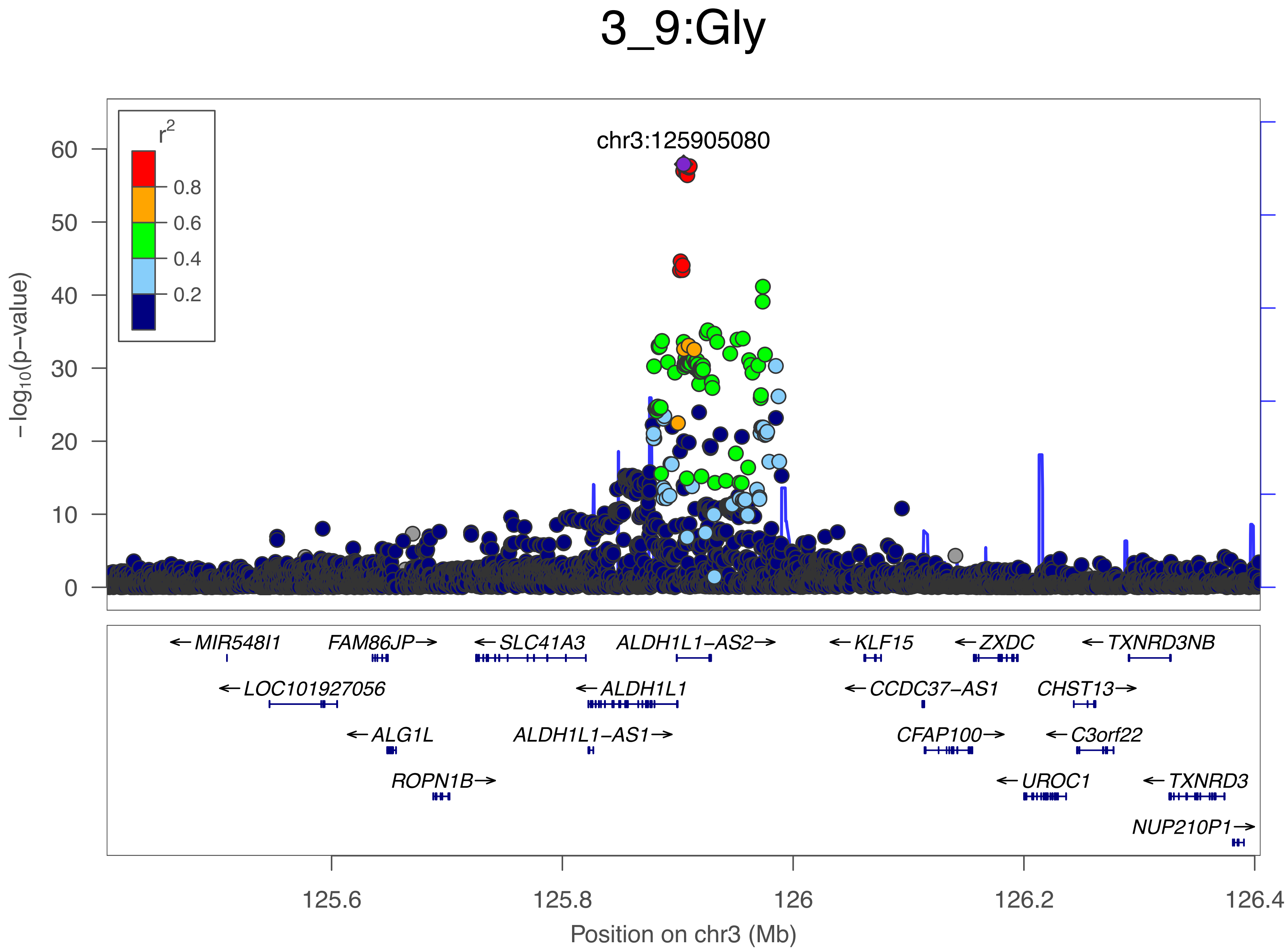

3_9:Gly
chr3:125905080
r2
0.8
0.6
0.4
0.2
60
50
40
30
20
10
0
-log10(p-value)
100
80
60
40
20
0
Recombination rate (cM/Mb)
← MIR548I1
FAM86JP →
← SLC41A3
ALDH1L1-AS2 →
← KLF15
← ZXDC
← TXNRD3NB
← LOC101927056
← ALDH1L1
← CCDC37-AS1
CHST13 →
← ALG1L
ALDH1L1-AS1 →
CFAP100 →
← C3orf22
ROPN1B →
← UROC1
← TXNRD3
NUP210P1 →
125.6
125.8
126
126.2
126.4
Position on chr3 (Mb)

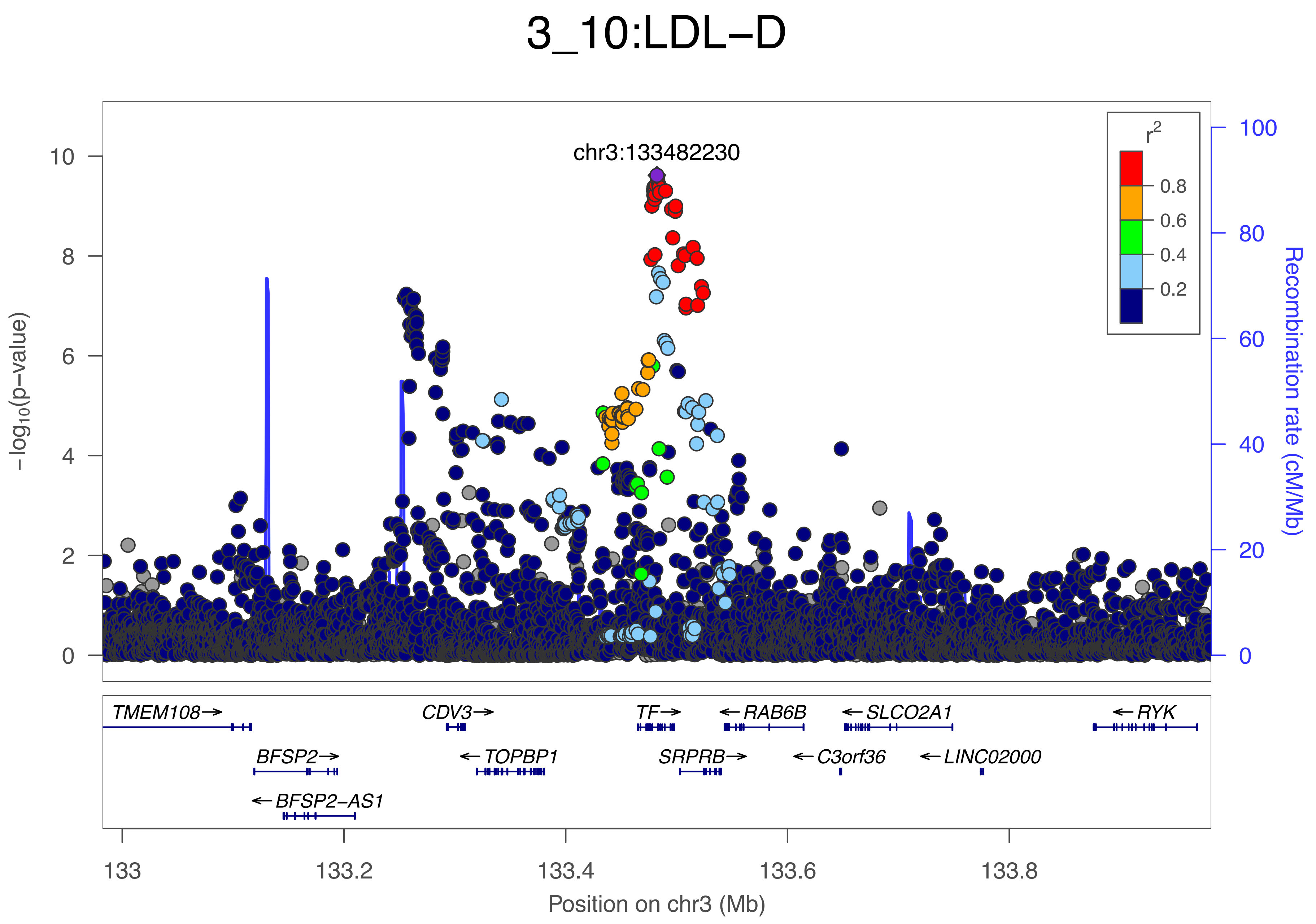
3_10:LDL−D
chr3:133482230
r²
0.8
0.6
0.4
0.2
−log₁₀(p−value)
Recombination rate (cM/Mb)
TMEM108→
CDV3→
TF→
←RAB6B
←SLCO2A1
←RYK
BFSP2→
←TOPBP1
SRPRB→
←C3orf36
←LINC02000
←BFSP2−AS1
133
133.2
133.4
133.6
133.8
Position on chr3 (Mb)

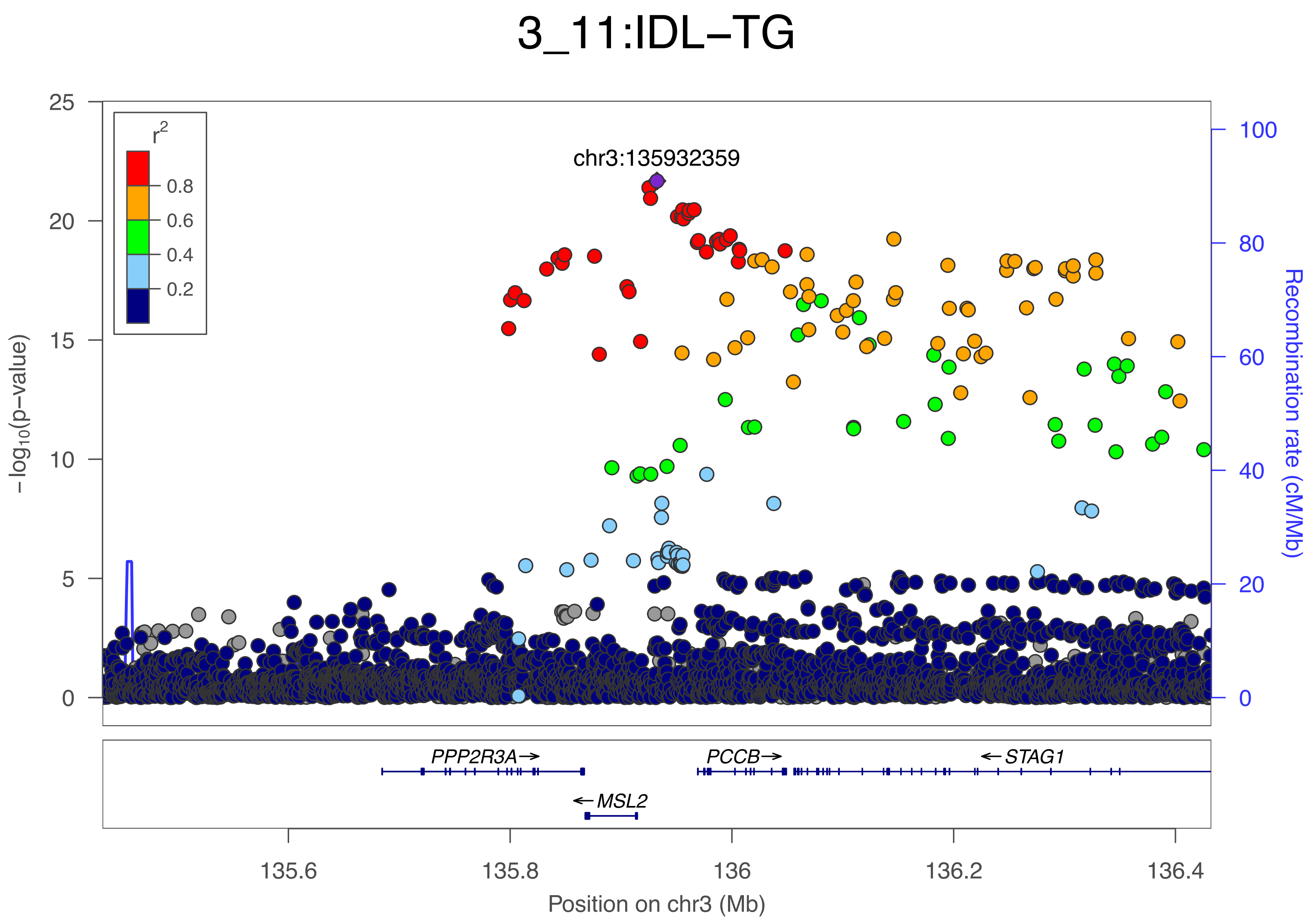
3_11:IDL−TG
r2
0.8
0.6
0.4
0.2
chr3:135932359
−log10(p−value)
Recombination rate (cM/Mb)
25
20
15
10
5
0
100
80
60
40
20
0
PPP2R3A→
PCCB→
←STAG1
←MSL2
135.6
135.8
136
136.2
136.4
Position on chr3 (Mb)

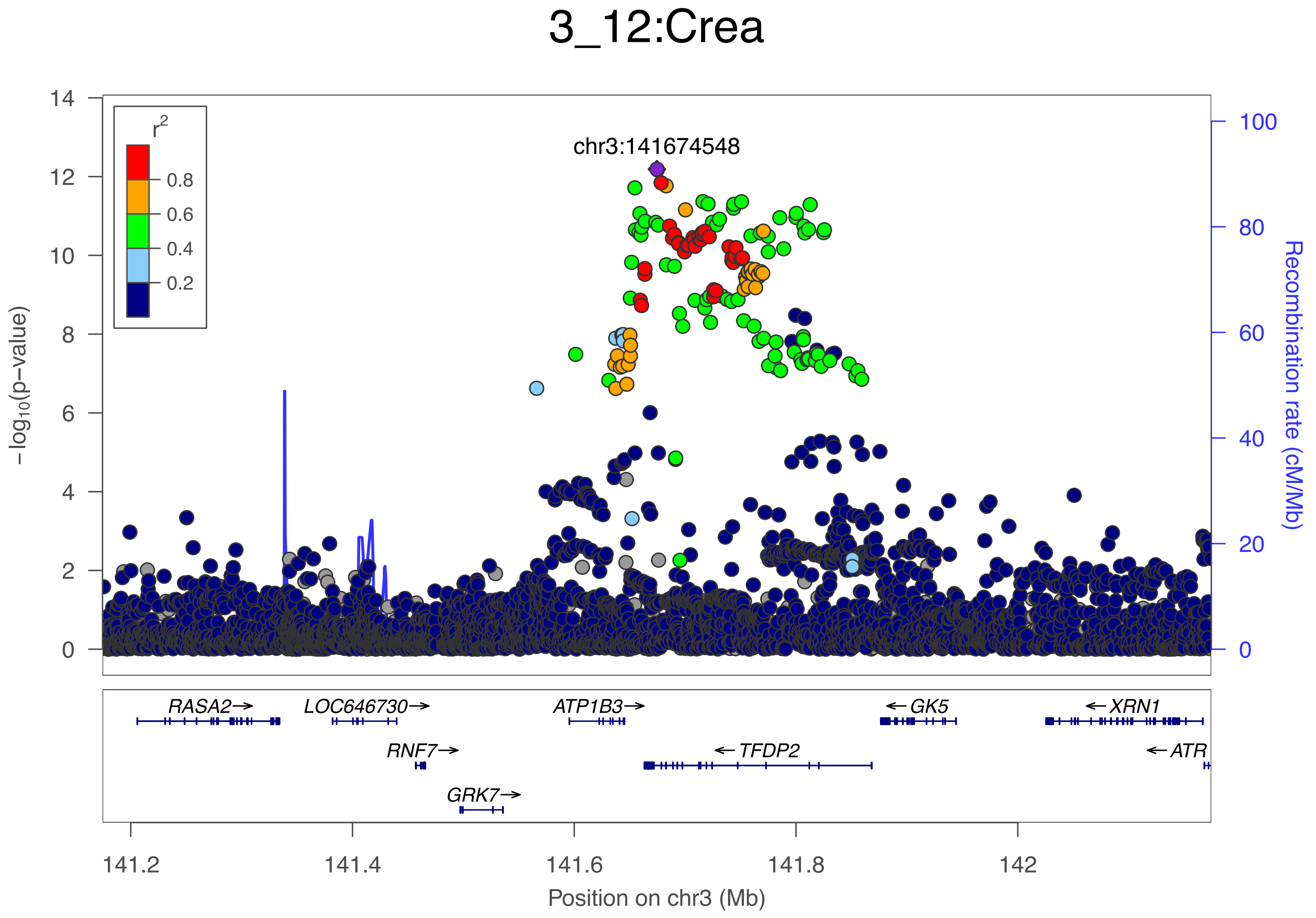
3_12:Crea
chr3:141674548
r2
0.8
0.6
0.4
0.2
14
12
10
8
6
4
2
0
−log10(p−value)
100
80
60
40
20
0
Recombination rate (cM/Mb)
RASA2→
LOC646730→
ATP1B3→
←GK5
←XRN1
RNF7→
←TFDP2
←ATR
GRK7→
141.2
141.4
141.6
141.8
142
Position on chr3 (Mb)

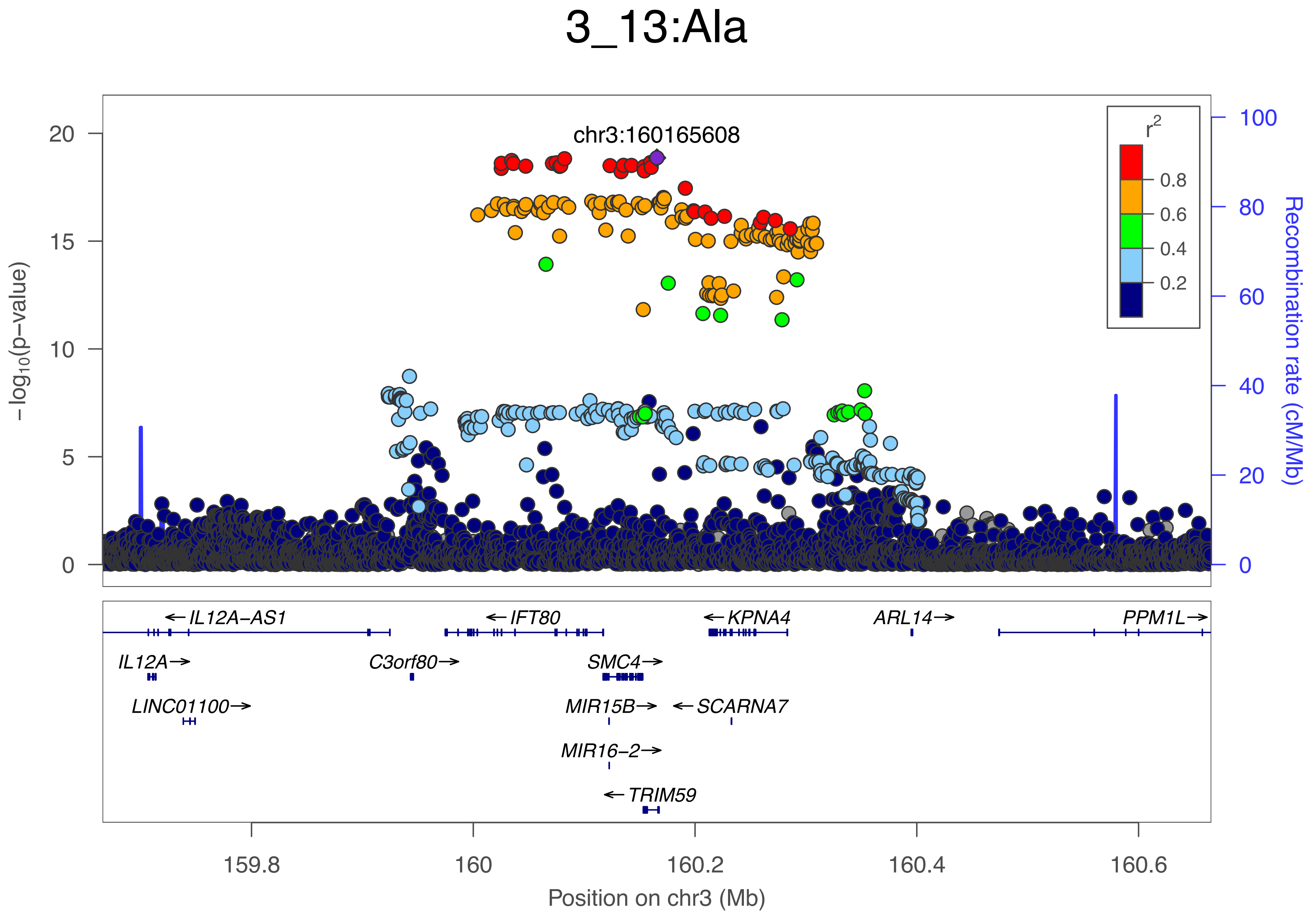

3_13:Ala
chr3:160165608
r2
0.8
0.6
0.4
0.2
20
15
10
5
0
100
80
60
40
20
0
−log10(p−value)
Recombination rate (cM/Mb)
←IL12A−AS1
←IFT80
←KPNA4
ARL14→
PPM1L→
IL12A→
C3orf80→
SMC4→
LINC01100→
MIR15B→
←SCARNA7
MIR16−2→
←TRIM59
159.8
160
160.2
160.4
160.6
Position on chr3 (Mb)

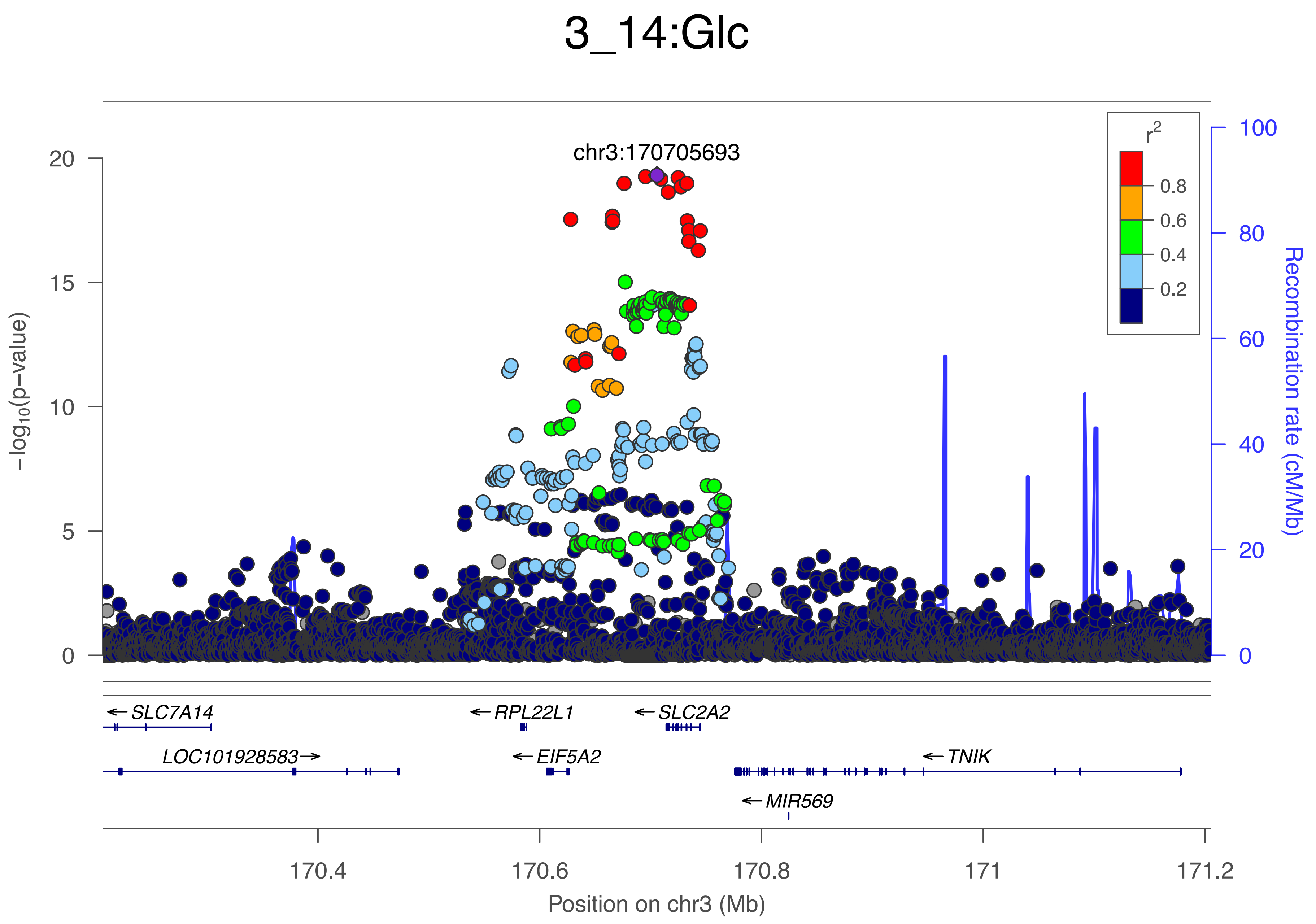

3_14:Glc
chr3:170705693
r2
0.8
0.6
0.4
0.2
−log10(p−value)
0
5
10
15
20
Recombination rate (cM/Mb)
0
20
40
60
80
100
SLC7A14
RPL22L1
SLC2A2
LOC101928583
EIF5A2
TNIK
MIR569
170.4
170.6
170.8
171
171.2
Position on chr3 (Mb)

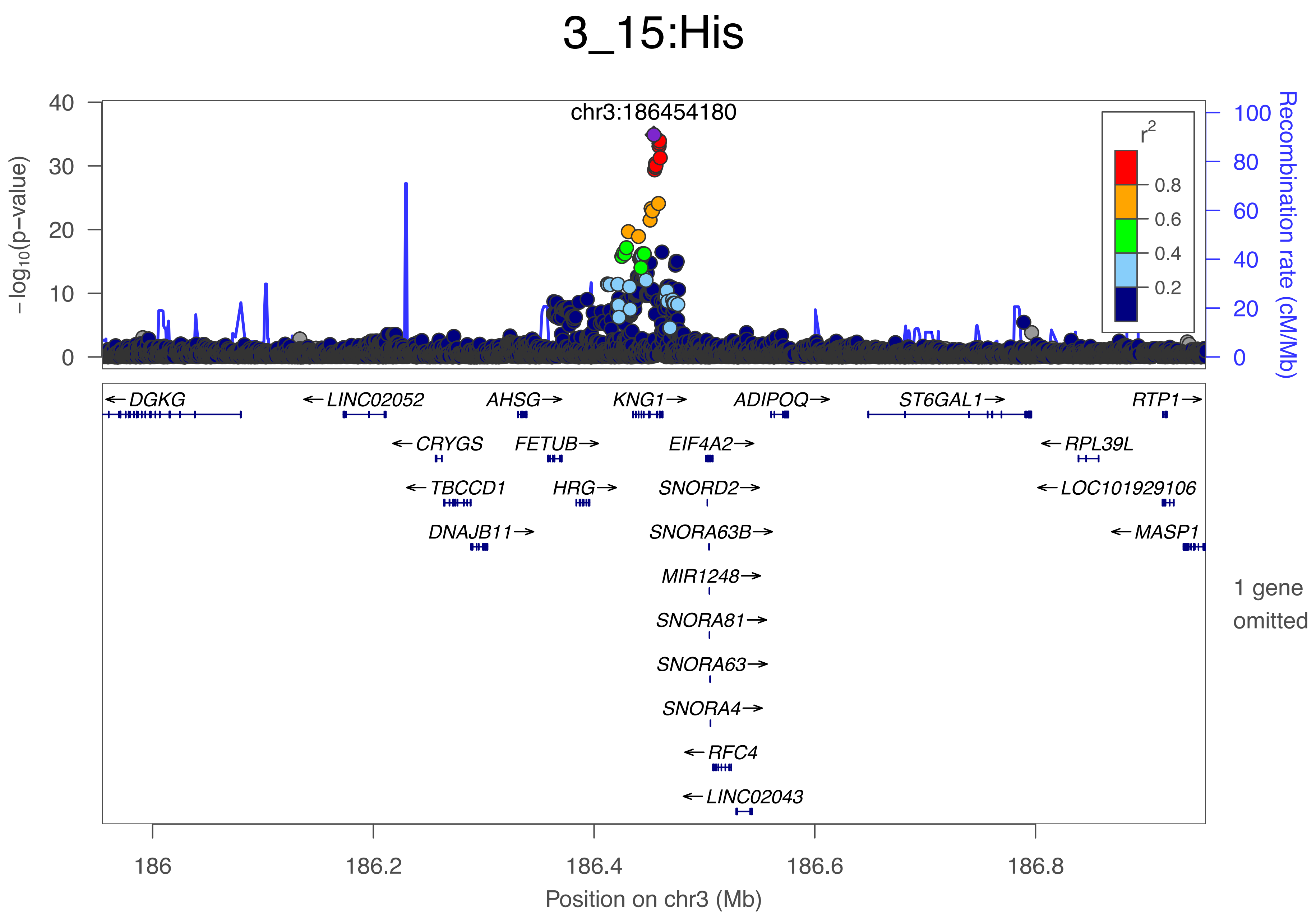

3_15:His
chr3:186454180
−log10(p−value)
Recombination rate (cM/Mb)
r2
0.8
0.6
0.4
0.2
DGKG
LINC02052
AHSG
KNG1
ADIPOQ
ST6GAL1
RTP1
CRYGS
FETUB
EIF4A2
RPL39L
TBCCD1
HRG
SNORD2
LOC101929106
DNAJB11
SNORA63B
MASP1
MIR1248
SNORA81
SNORA63
SNORA4
RFC4
LINC02043
1 gene omitted
Position on chr3 (Mb)

### 4_1:GlycA

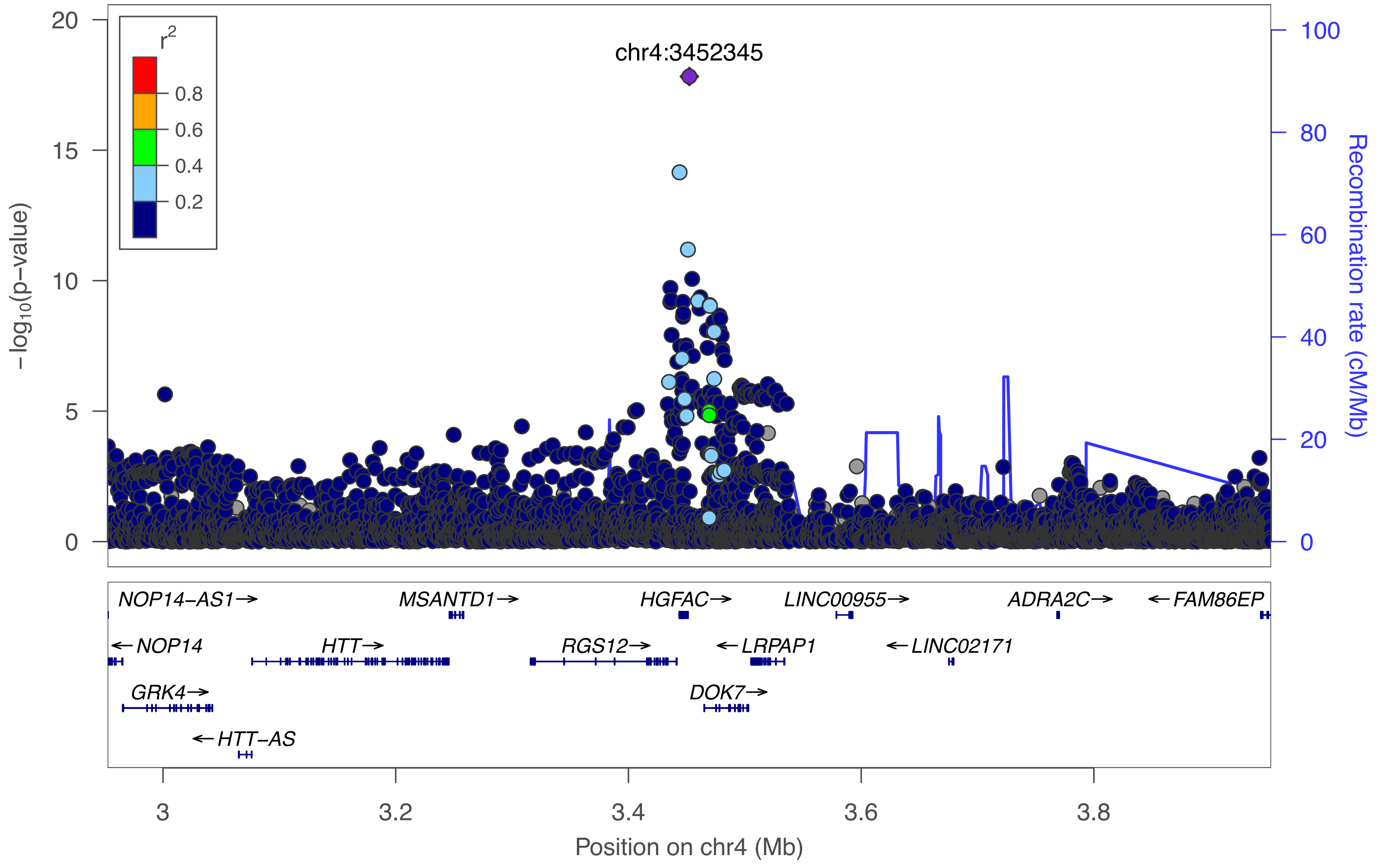

r2
0.8
0.6
0.4
0.2
chr4:3452345
20
15
10
5
0
−log10(p−value)
100
80
60
40
20
0
Recombination rate (cM/Mb)
NOP14−AS1→
MSANTD1→
HGFAC→
LINC00955→
ADRA2C→
←FAM86EP
←NOP14
HTT→
RGS12→
←LRPAP1
←LINC02171
GRK4→
DOK7→
←HTT−AS
3
3.2
3.4
3.6
3.8
Position on chr4 (Mb)

### 4_2:Alb

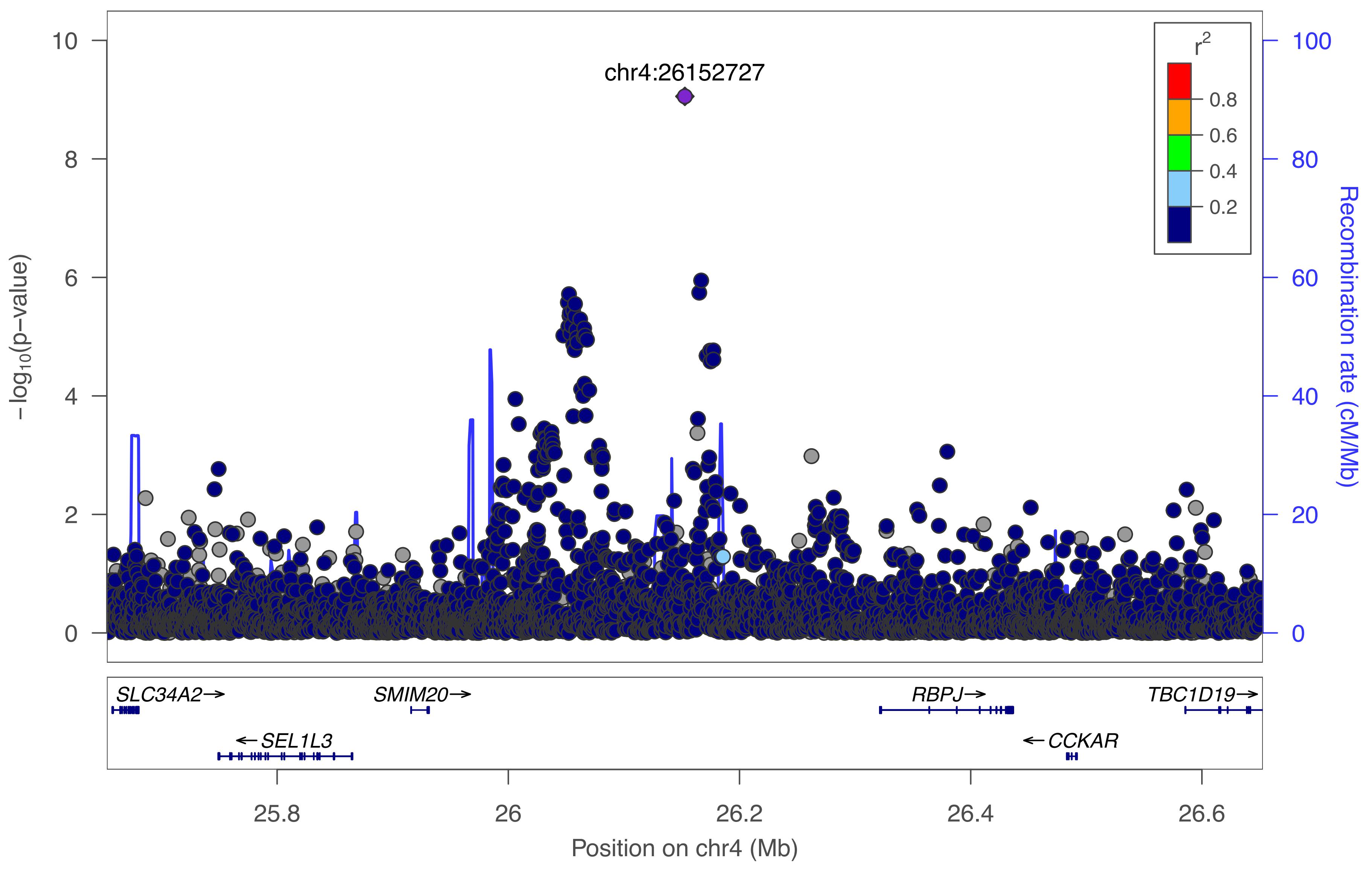

chr4:26152727
r²
0.8
0.6
0.4
0.2
10
8
6
4
2
0
−log10(p−value)
100
80
60
40
20
0
Recombination rate (cM/Mb)
SLC34A2→
SMIM20→
RBPJ→
TBC1D19→
←SEL1L3
←CCKAR
25.8
26
26.2
26.4
26.6
Position on chr4 (Mb)

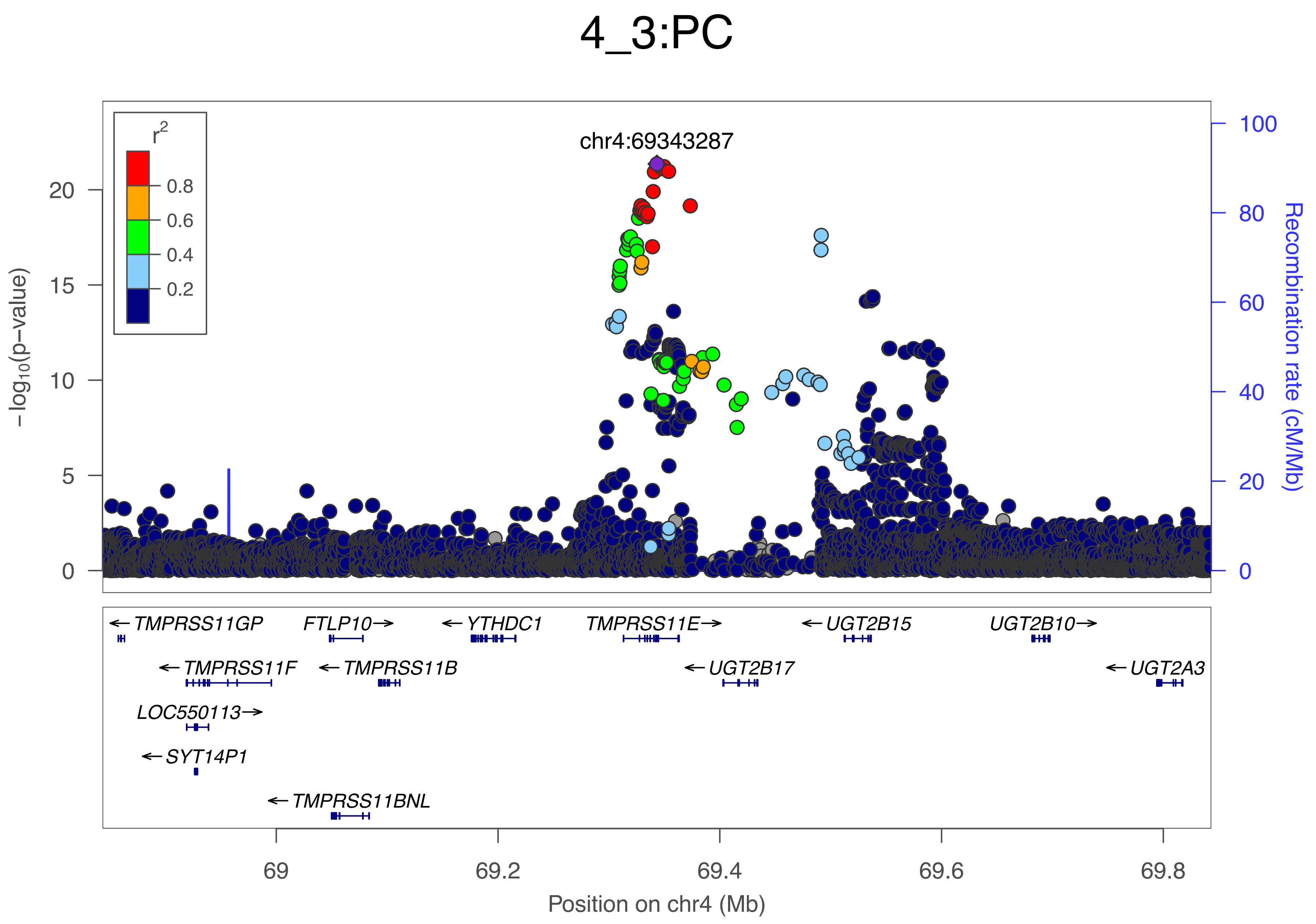

4_3:PC
chr4:69343287
r2
0.8
0.6
0.4
0.2
20
15
10
5
0
100
80
60
40
20
0
−log10(p−value)
Recombination rate (cM/Mb)
←TMPRSS11GP
FTLP10→
←YTHDC1
TMPRSS11E→
←UGT2B15
UGT2B10→
←TMPRSS11F
←TMPRSS11B
←UGT2B17
←UGT2A3
LOC550113→
←SYT14P1
←TMPRSS11BNL
69
69.2
69.4
69.6
69.8
Position on chr4 (Mb)

### 4_4:Alb

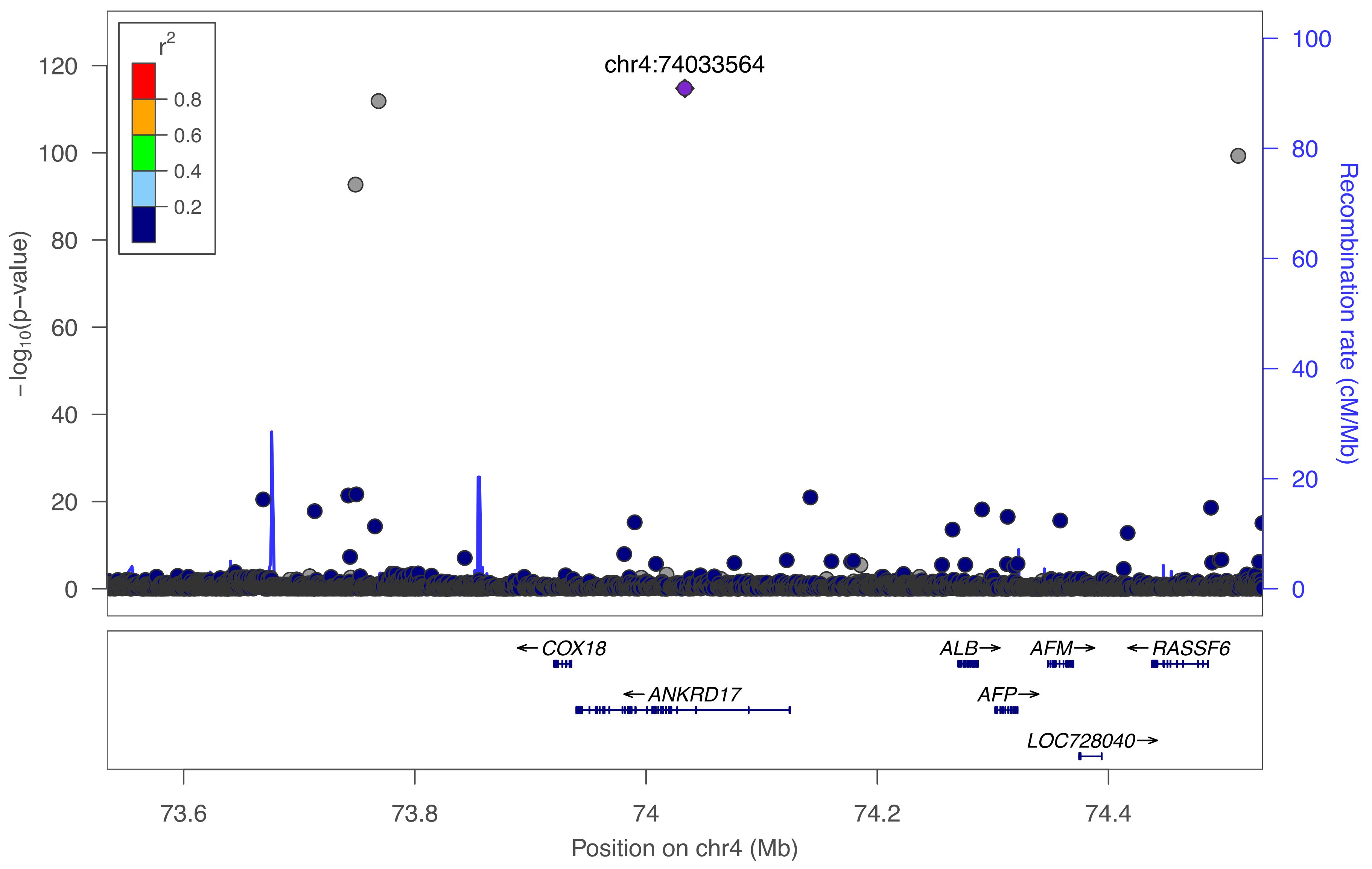
r2
0.8
0.6
0.4
0.2
chr4:74033564
120
100
80
60
40
20
0
−log10(p−value)
100
80
60
40
20
0
Recombination rate (cM/Mb)
← COX18
← ANKRD17
ALB →
AFM →
← RASSF6
AFP →
LOC728040 →
73.6
73.8
74
74.2
74.4
Position on chr4 (Mb)

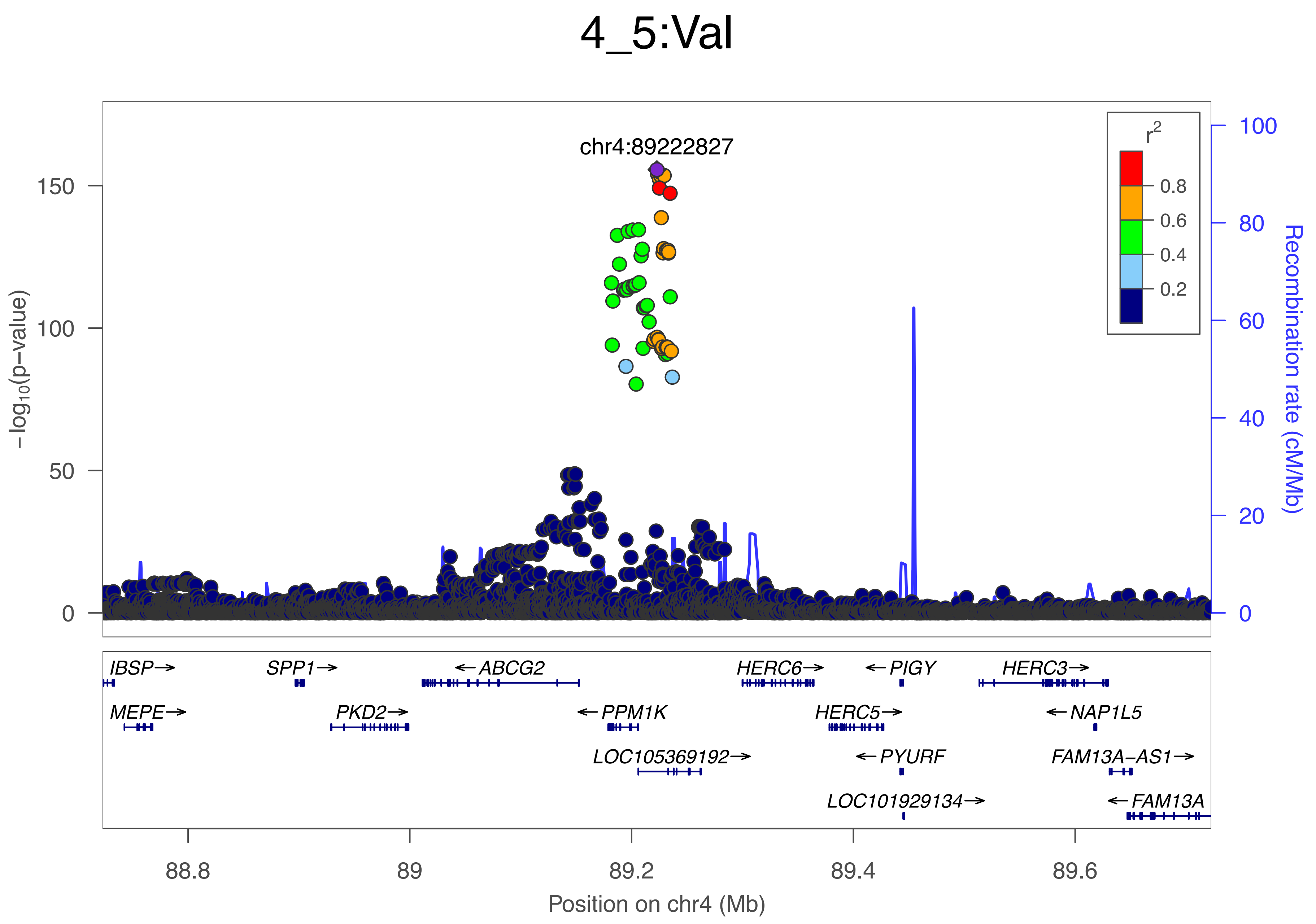

4_5:Val
chr4:89222827
−log10(p−value)
150
100
50
0
r2
0.8
0.6
0.4
0.2
100
80
60
40
20
0
Recombination rate (cM/Mb)
IBSP→
SPP1→
←ABCG2
HERC6→
←PIGY
HERC3→
MEPE→
PKD2→
←PPM1K
HERC5→
←NAP1L5
LOC105369192→
←PYURF
FAM13A−AS1→
LOC101929134→
←FAM13A
88.8
89
89.2
89.4
89.6
Position on chr4 (Mb)

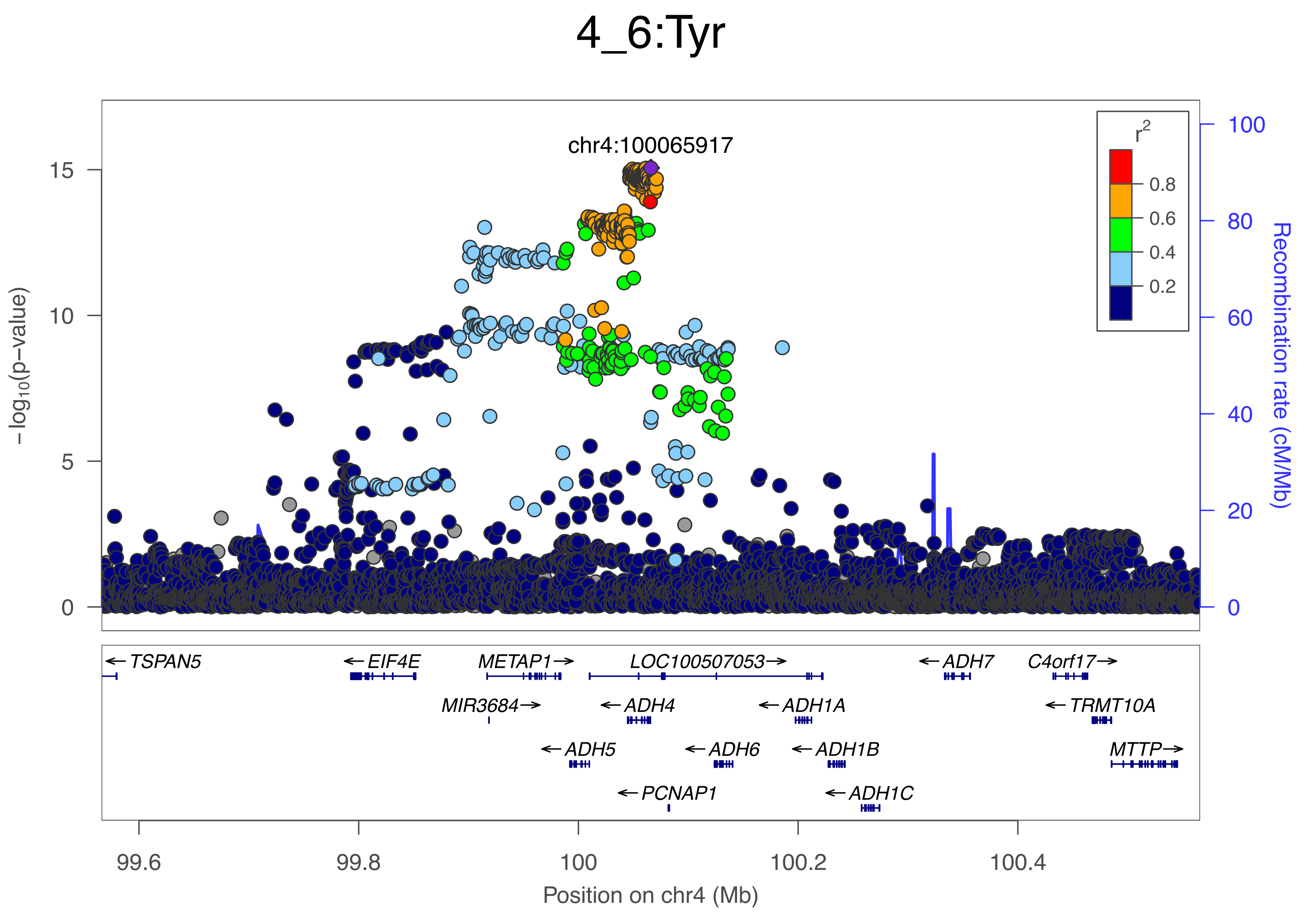
4_6:Tyr
chr4:100065917
−log10(p−value)
Recombination rate (cM/Mb)
r2
0.8
0.6
0.4
0.2
TSPAN5
EIF4E
METAP1
LOC100507053
ADH7
C4orf17
MIR3684
ADH4
ADH1A
TRMT10A
ADH5
ADH6
ADH1B
MTTP
PCNAP1
ADH1C
99.6
99.8
100
100.2
100.4
Position on chr4 (Mb)

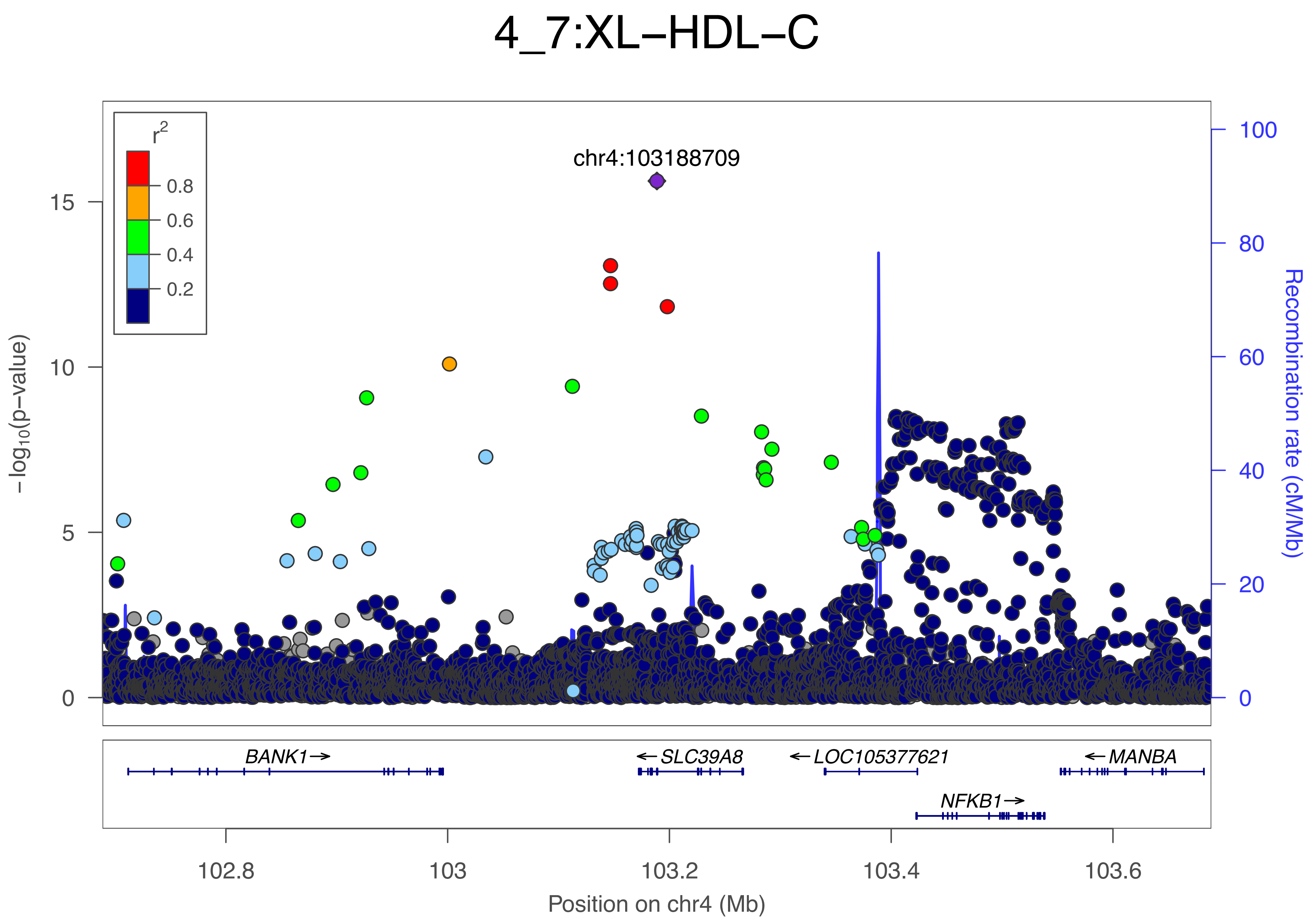

4_7:XL−HDL−C
r²
0.8
0.6
0.4
0.2
chr4:103188709
15
10
5
0
−log₁₀(p−value)
100
80
60
40
20
0
Recombination rate (cM/Mb)
BANK1→
←SLC39A8
←LOC105377621
←MANBA
NFKB1→
102.8
103
103.2
103.4
103.6
Position on chr4 (Mb)

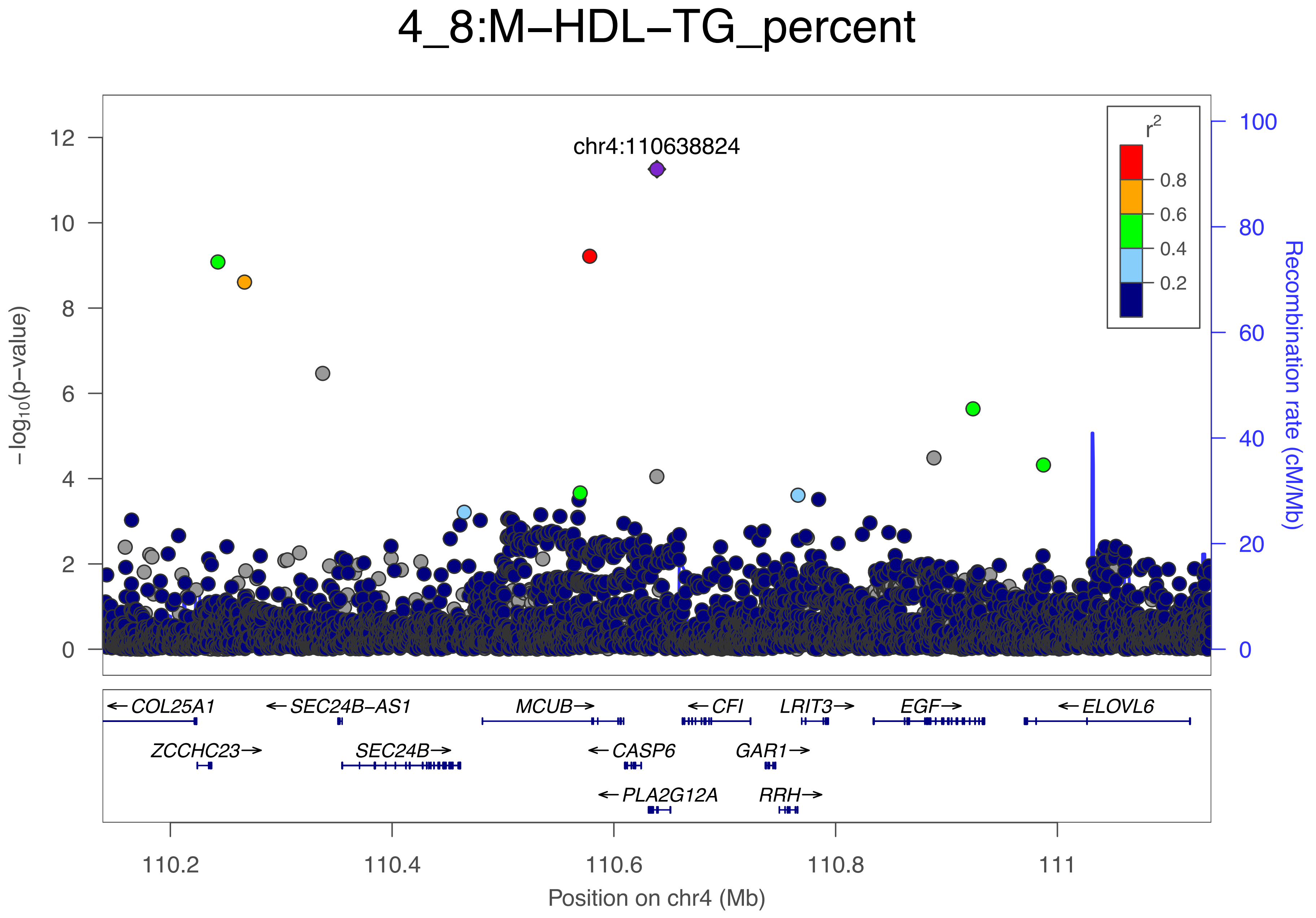

4_8:M−HDL−TG_percent
chr4:110638824
r2
0.8
0.6
0.4
0.2
−log10(p−value)
Recombination rate (cM/Mb)
12
10
8
6
4
2
0
100
80
60
40
20
0
← COL25A1
← SEC24B−AS1
MCUB →
← CFI
LRIT3 →
EGF →
← ELOVL6
ZCCHC23 →
SEC24B →
← CASP6
GAR1 →
← PLA2G12A
RRH →
110.2
110.4
110.6
110.8
111
Position on chr4 (Mb)

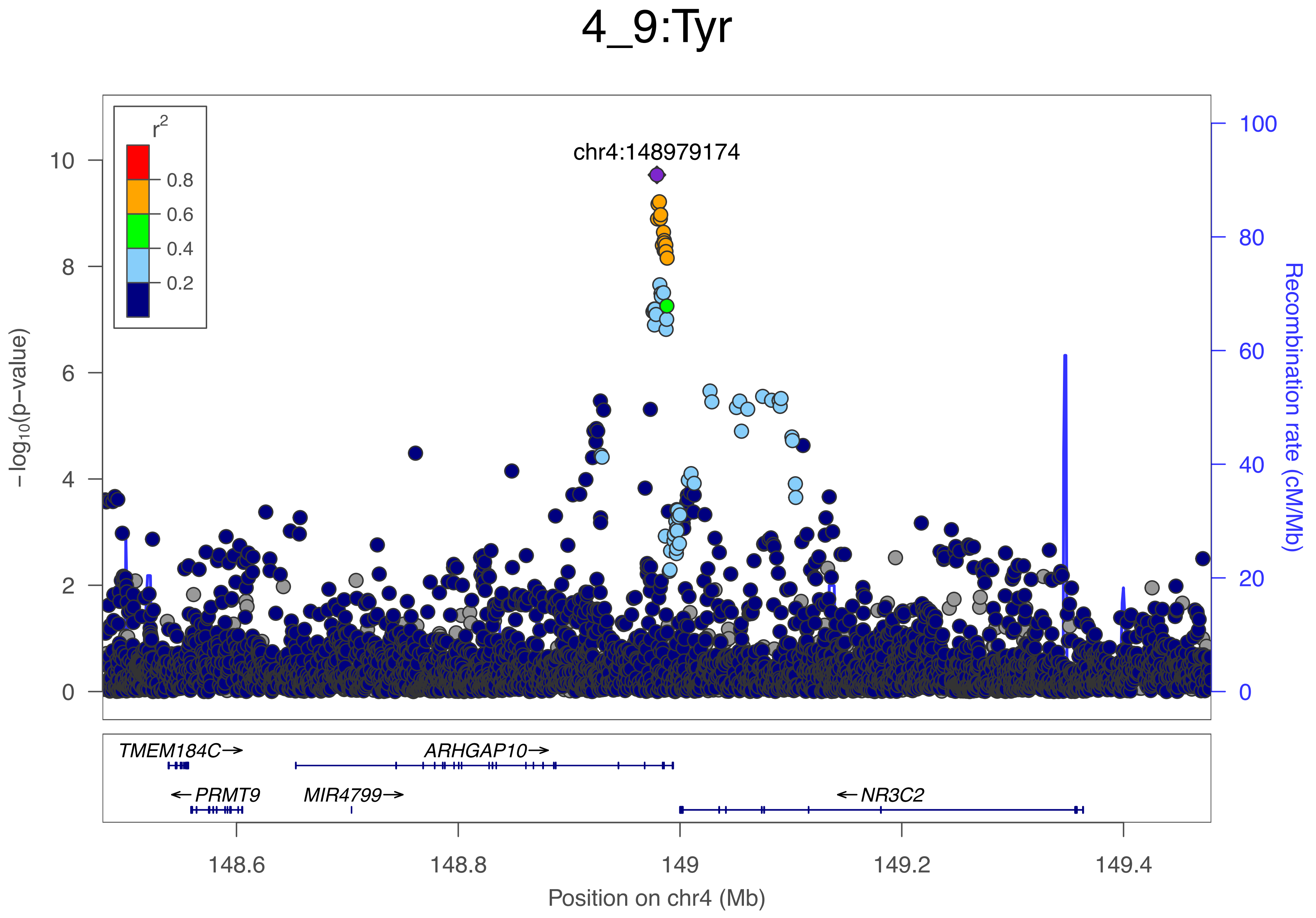

4_9:Tyr
chr4:148979174
r²
0.8
0.6
0.4
0.2
−log10(p−value)
Recombination rate (cM/Mb)
TMEM184C→
ARHGAP10→
←PRMT9
MIR4799→
←NR3C2
148.6
148.8
149
149.2
149.4
Position on chr4 (Mb)

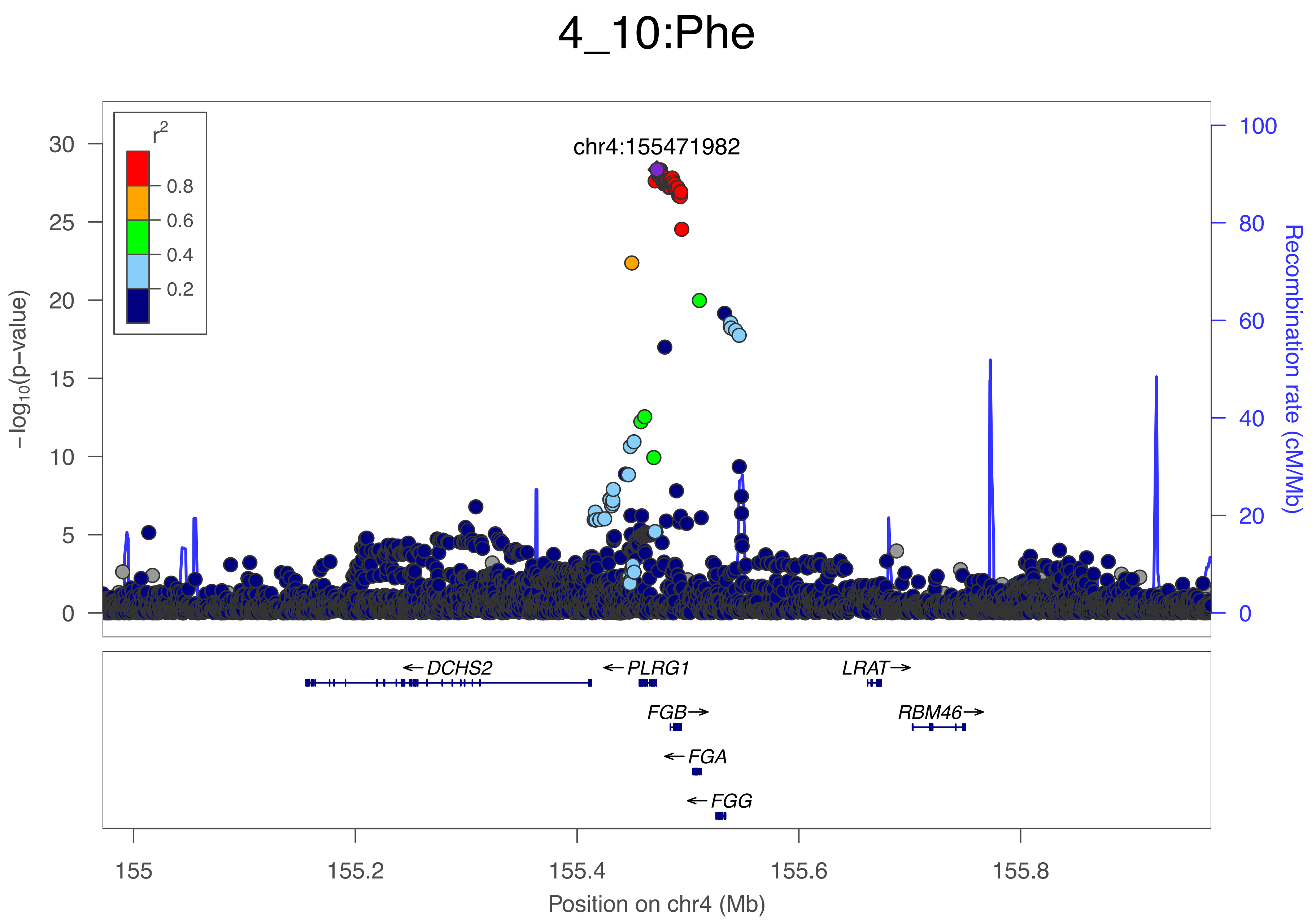

4_10:Phe
r2
0.8
0.6
0.4
0.2
chr4:155471982
−log10(p−value)
Recombination rate (cM/Mb)
DCHS2
PLRG1
LRAT
FGB
RBM46
FGA
FGG
155
155.2
155.4
155.6
155.8
Position on chr4 (Mb)

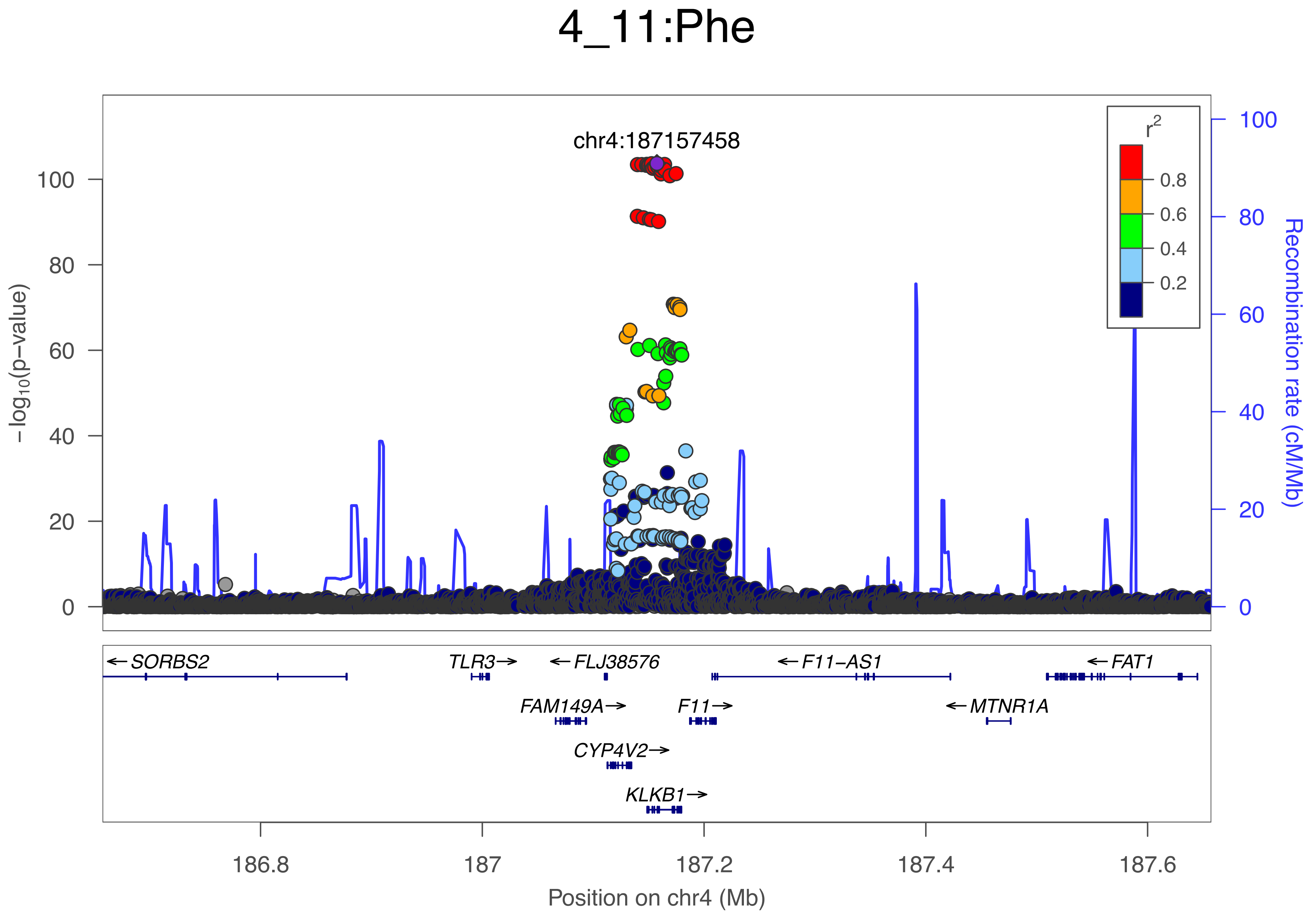

4_11:Phe
chr4:187157458
−log10(p−value)
100
80
60
40
20
0
Recombination rate (cM/Mb)
r2
0.8
0.6
0.4
0.2
← SORBS2
TLR3→
← FLJ38576
← F11−AS1
← FAT1
FAM149A→
F11→
← MTNR1A
CYP4V2→
KLKB1→
186.8
187
187.2
187.4
187.6
Position on chr4 (Mb)

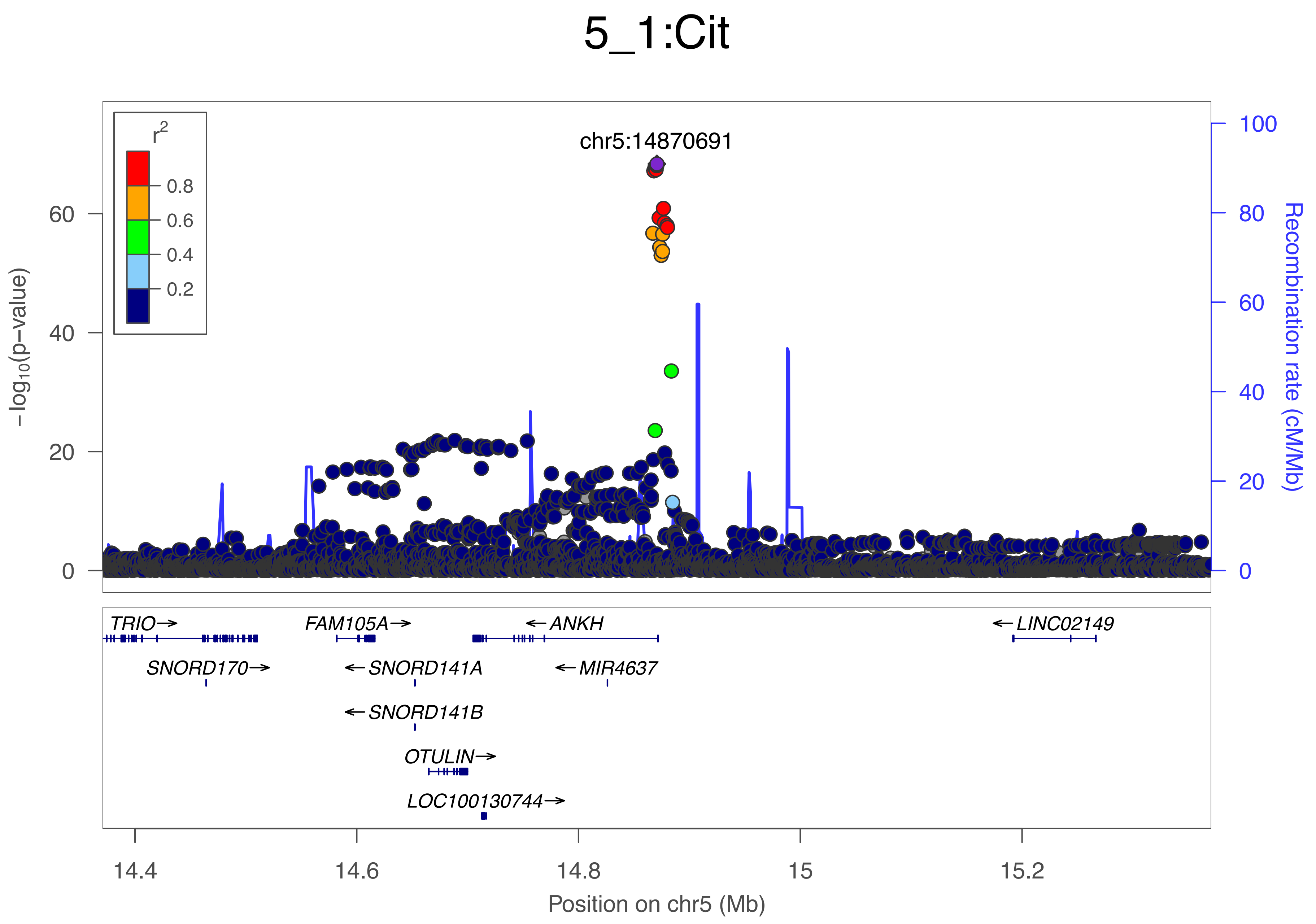

5_1:Cit
r2
0.8
0.6
0.4
0.2
chr5:14870691
−log10(p−value)
0
20
40
60
Recombination rate (cM/Mb)
0
20
40
60
80
100
TRIO→
FAM105A→
←ANKH
←LINC02149
SNORD170→
←SNORD141A
←MIR4637
←SNORD141B
OTULIN→
LOC100130744→
14.4
14.6
14.8
15
15.2
Position on chr5 (Mb)

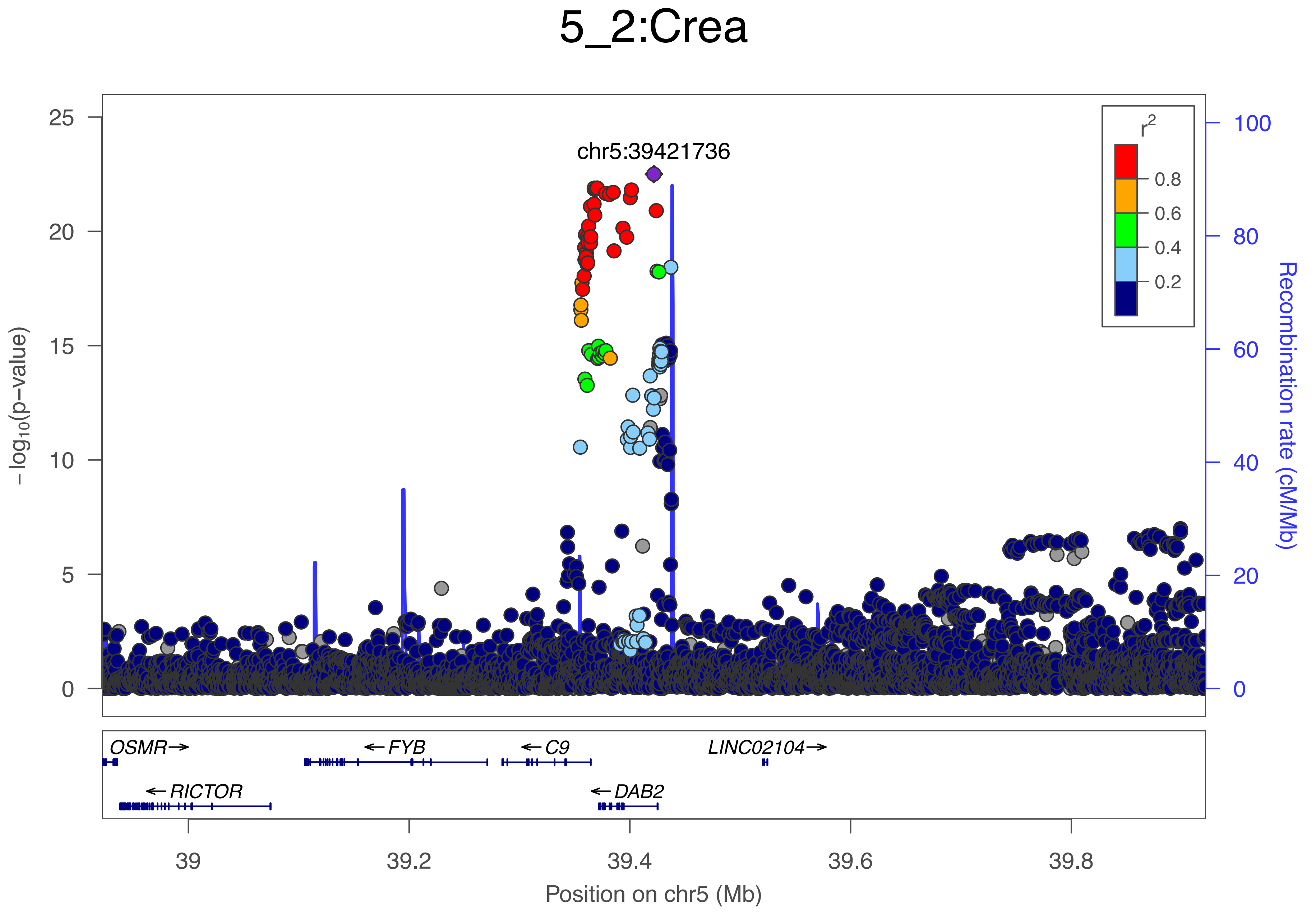
5_2:Crea
chr5:39421736
r2
0.8
0.6
0.4
0.2
−log10(p−value)
Recombination rate (cM/Mb)
25
20
15
10
5
0
100
80
60
40
20
0
OSMR→
←FYB
←C9
LINC02104→
←RICTOR
←DAB2
39
39.2
39.4
39.6
39.8
Position on chr5 (Mb)

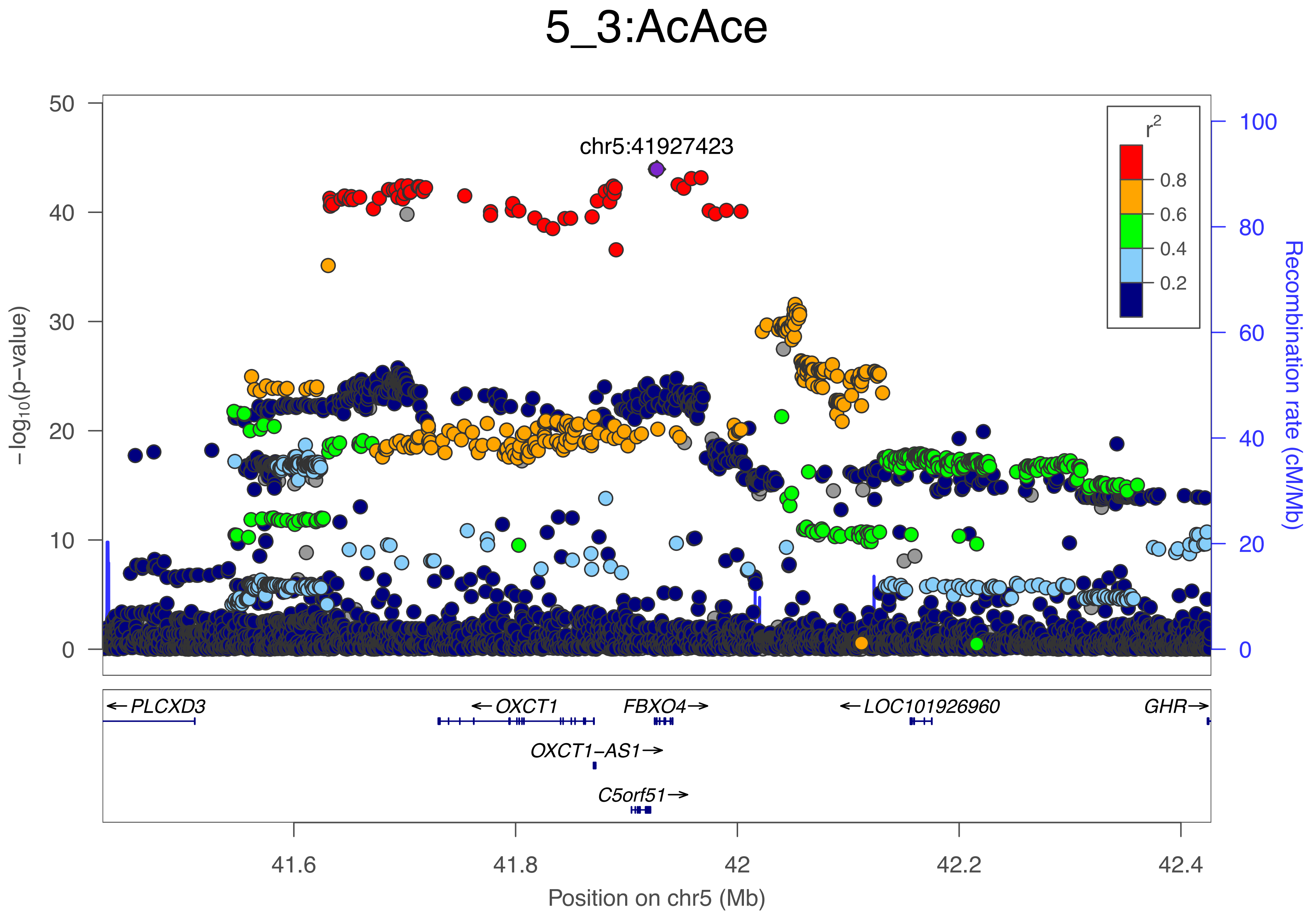
5_3:AcAce
chr5:41927423
r²
0.8
0.6
0.4
0.2
−log10(p−value)
Recombination rate (cM/Mb)
PLCXD3
OXCT1
FBXO4
LOC101926960
GHR
OXCT1−AS1
C5orf51
Position on chr5 (Mb)
41.6
41.8
42
42.2
42.4

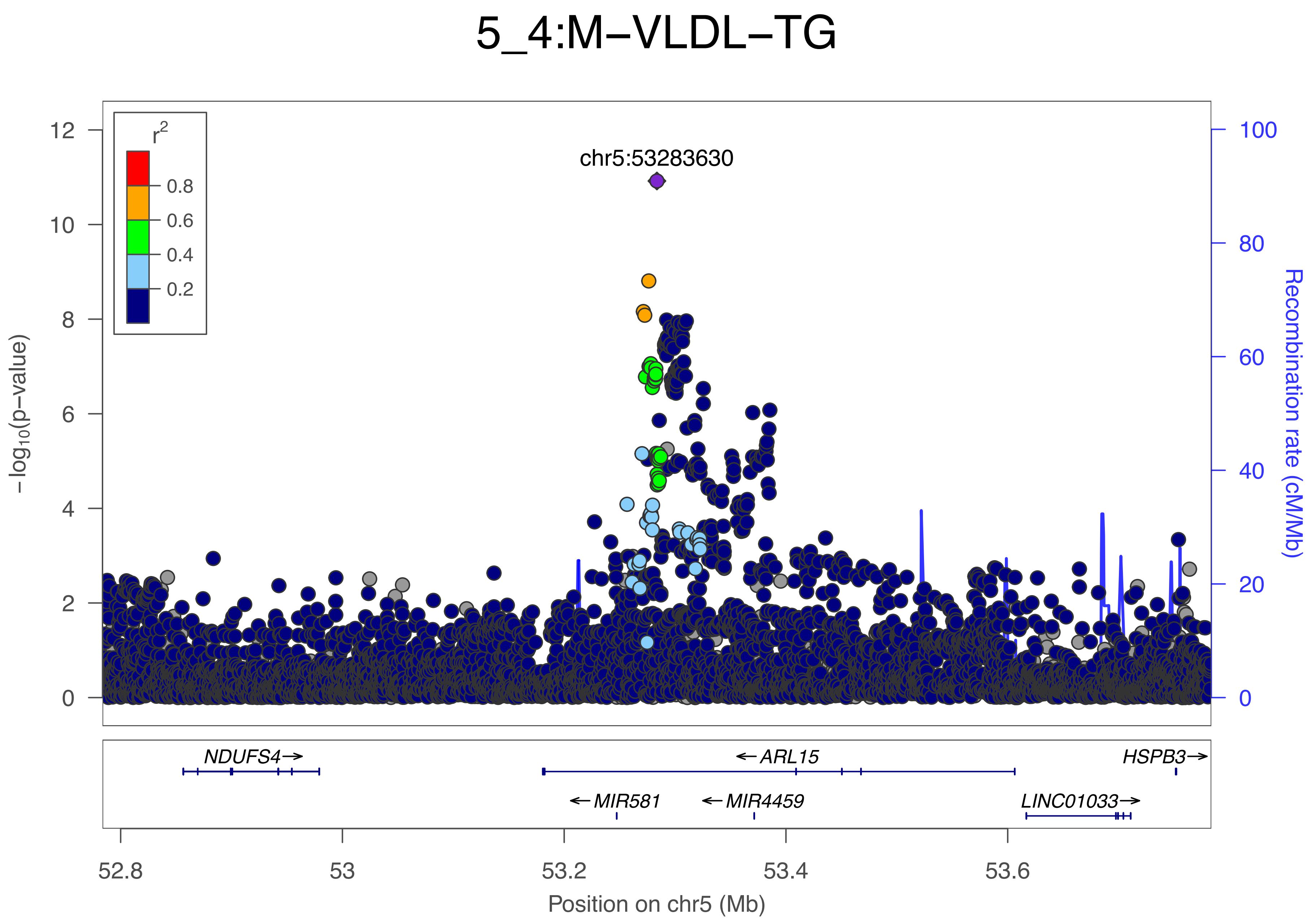
5_4:M−VLDL−TG
chr5:53283630
r2
0.8
0.6
0.4
0.2
−log10(p−value)
12
10
8
6
4
2
0
Recombination rate (cM/Mb)
100
80
60
40
20
0
NDUFS4→
←ARL15
HSPB3→
←MIR581
←MIR4459
LINC01033→
52.8
53
53.2
53.4
53.6
Position on chr5 (Mb)

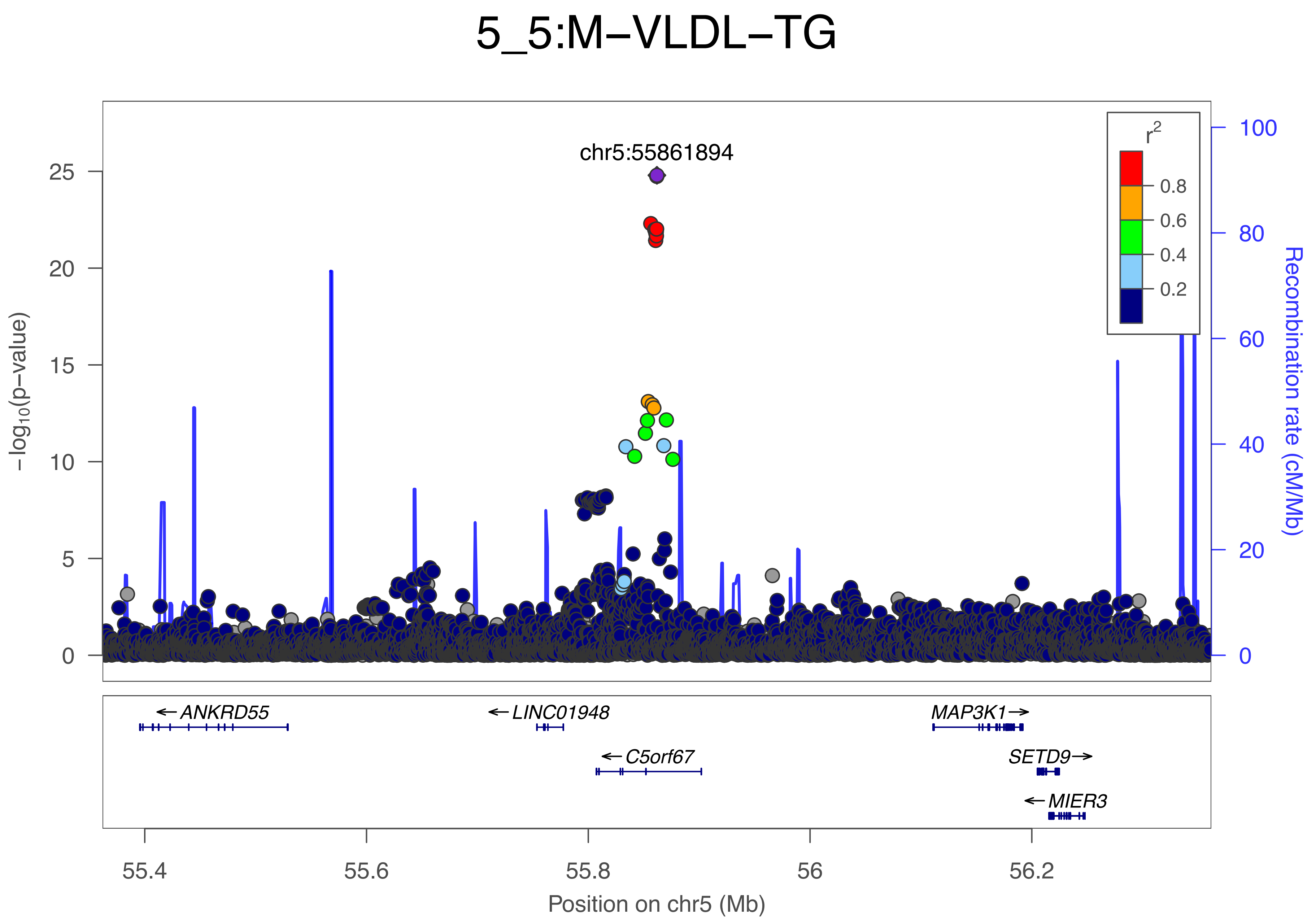
5_5:M−VLDL−TG
chr5:55861894
−log10(p−value)
Recombination rate (cM/Mb)
r2
0.8
0.6
0.4
0.2
ANKRD55
LINC01948
C5orf67
MAP3K1
SETD9
MIER3
55.4
55.6
55.8
56
56.2
Position on chr5 (Mb)

### 5_6:Crea

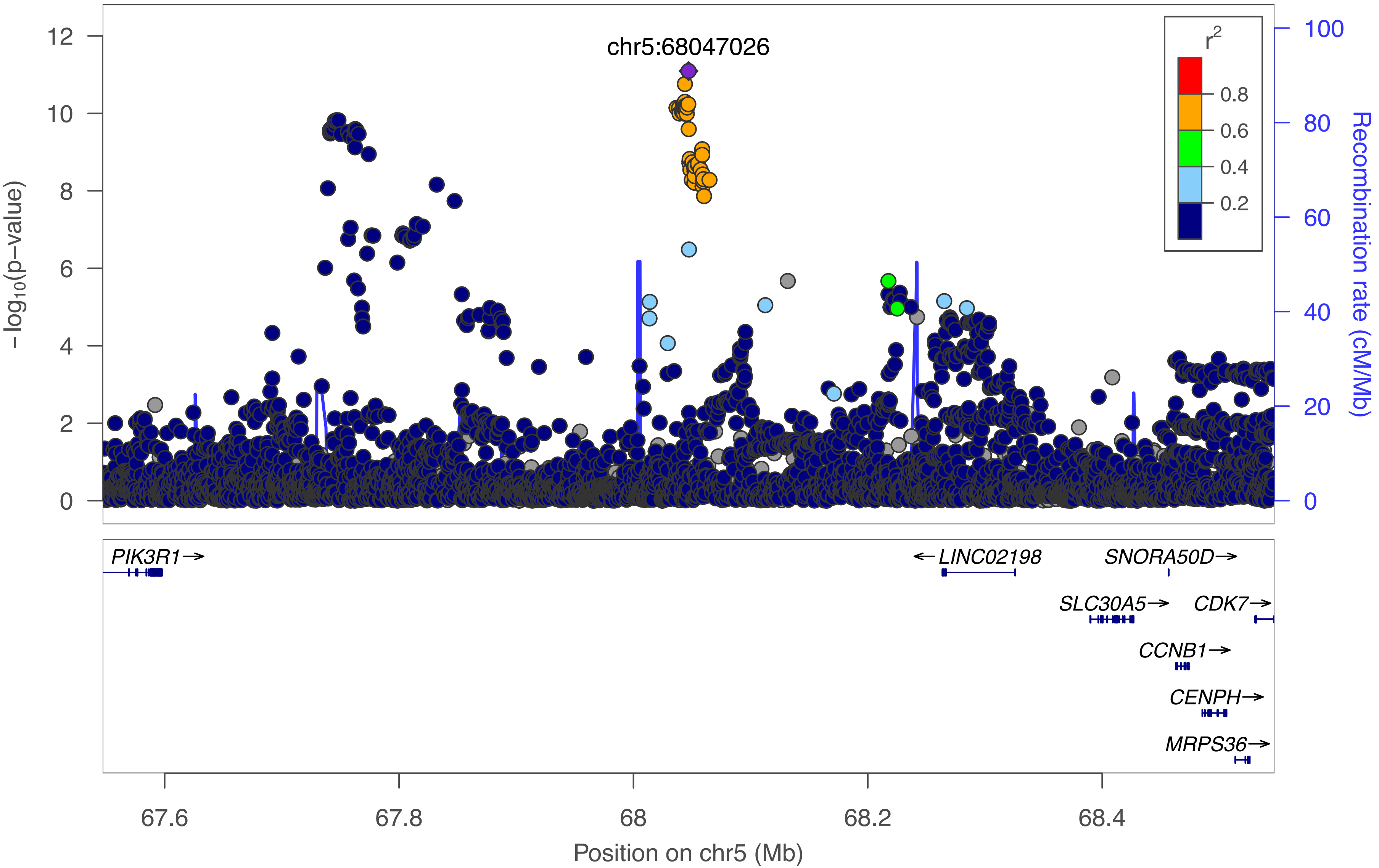

chr5:68047026
r2
0.8
0.6
0.4
0.2
−log10(p−value)
Recombination rate (cM/Mb)
PIK3R1→
←LINC02198
SNORA50D→
SLC30A5→
CDK7→
CCNB1→
CENPH→
MRPS36→
67.6
67.8
68
68.2
68.4
Position on chr5 (Mb)

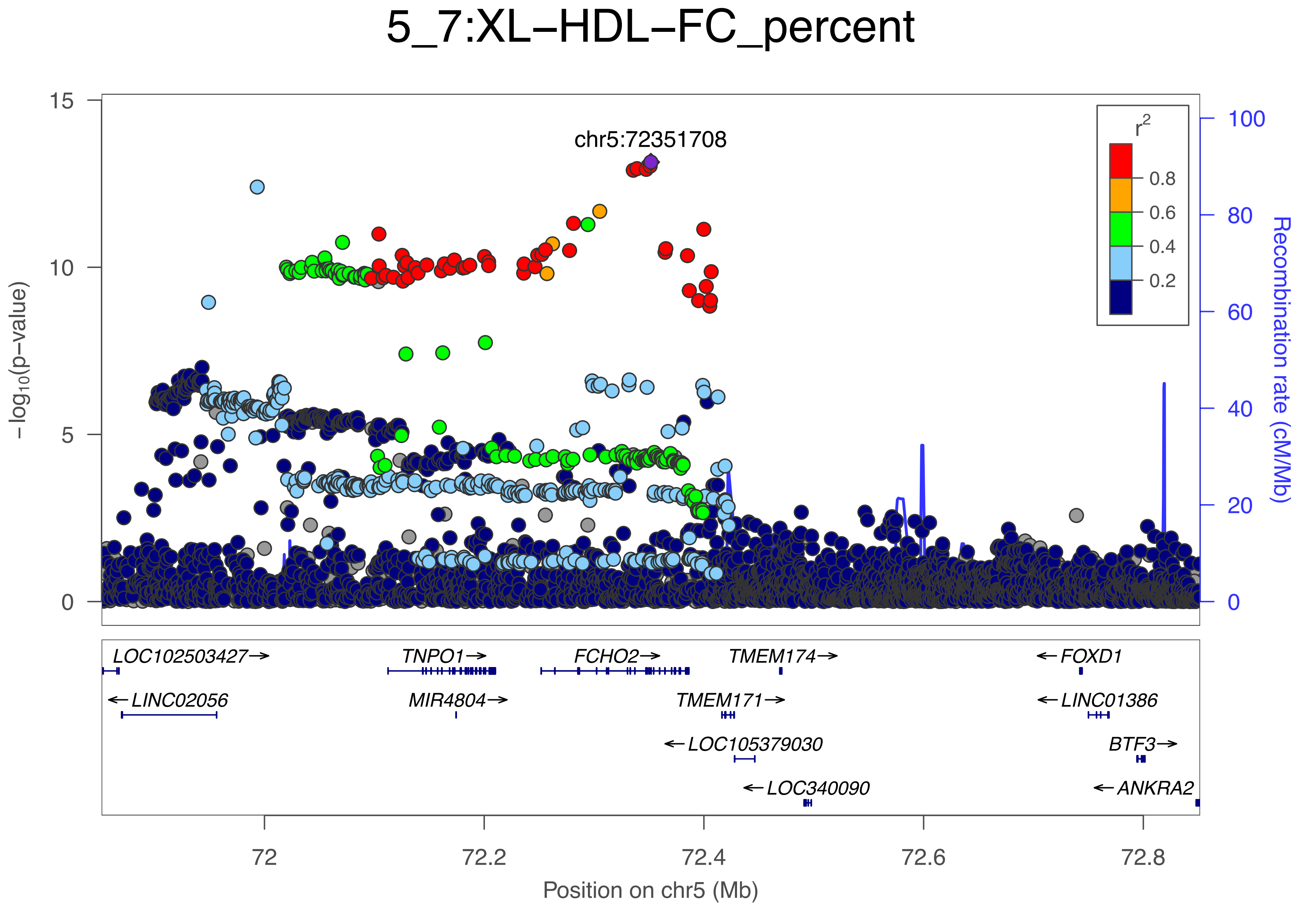

5_7:XL−HDL−FC_percent
chr5:72351708
$r^2$
0.8
0.6
0.4
0.2
−log10(p−value)
Recombination rate (cM/Mb)
0
5
10
15
0
20
40
60
80
100
LOC102503427→
TNPO1→
FCHO2→
TMEM174→
←FOXD1
←LINC02056
MIR4804→
TMEM171→
←LINC01386
←LOC105379030
BTF3→
←LOC340090
←ANKRA2
72
72.2
72.4
72.6
72.8
Position on chr5 (Mb)

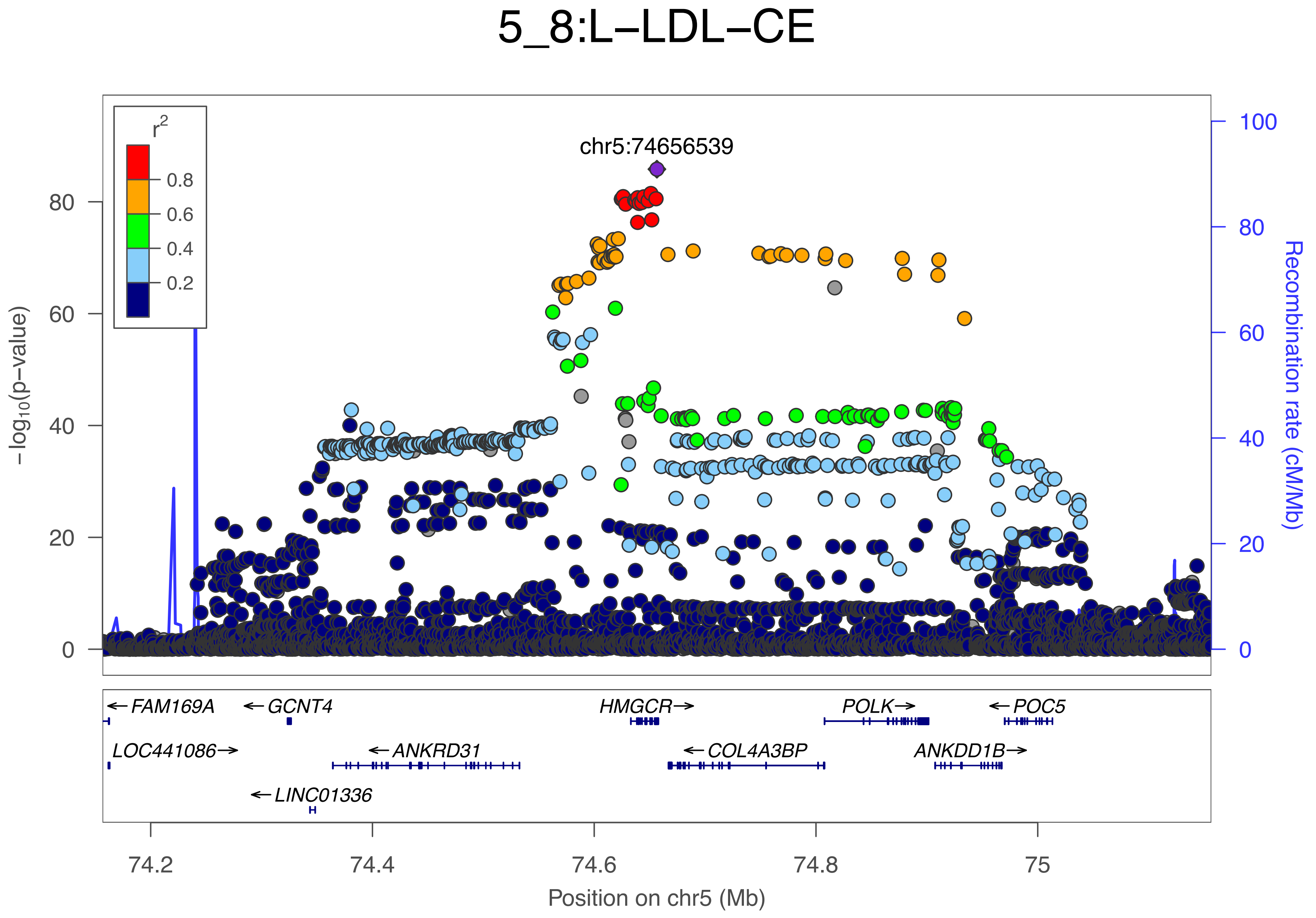

5_8:L−LDL−CE
r2
0.8
0.6
0.4
0.2
chr5:74656539
−log10(p−value)
Recombination rate (cM/Mb)
0
20
40
60
80
100
FAM169A
GCNT4
HMGCR
POLK
POC5
LOC441086
ANKRD31
COL4A3BP
ANKDD1B
LINC01336
74.2
74.4
74.6
74.8
75
Position on chr5 (Mb)

### 5_9:Glc

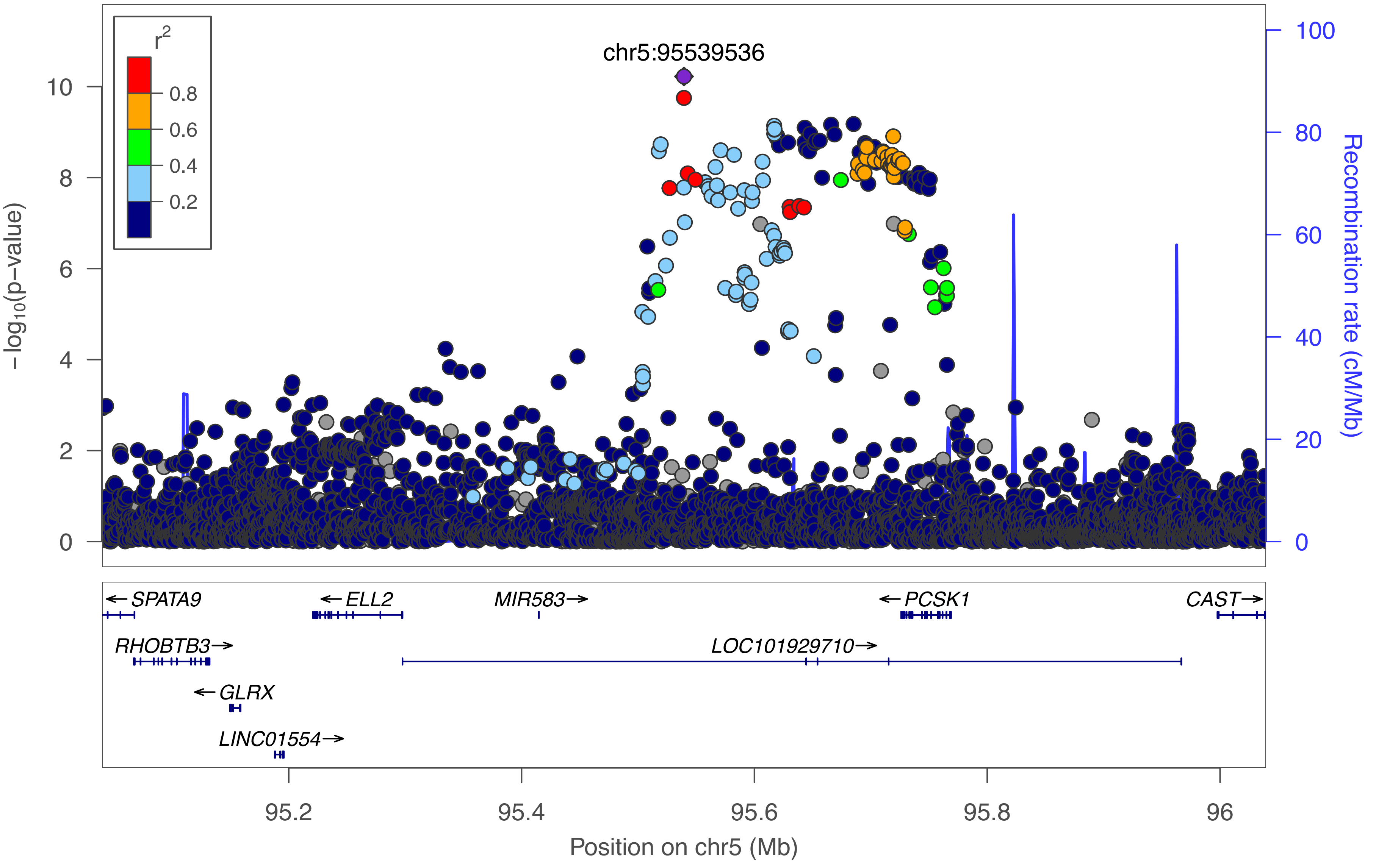

chr5:95539536
r2
0.8
0.6
0.4
0.2
10
8
6
4
2
0
-log10(p-value)
100
80
60
40
20
0
Recombination rate (cM/Mb)
← SPATA9
← ELL2
MIR583 →
← PCSK1
CAST →
RHOBTB3 →
LOC101929710 →
← GLRX
LINC01554 →
95.2
95.4
95.6
95.8
96
Position on chr5 (Mb)

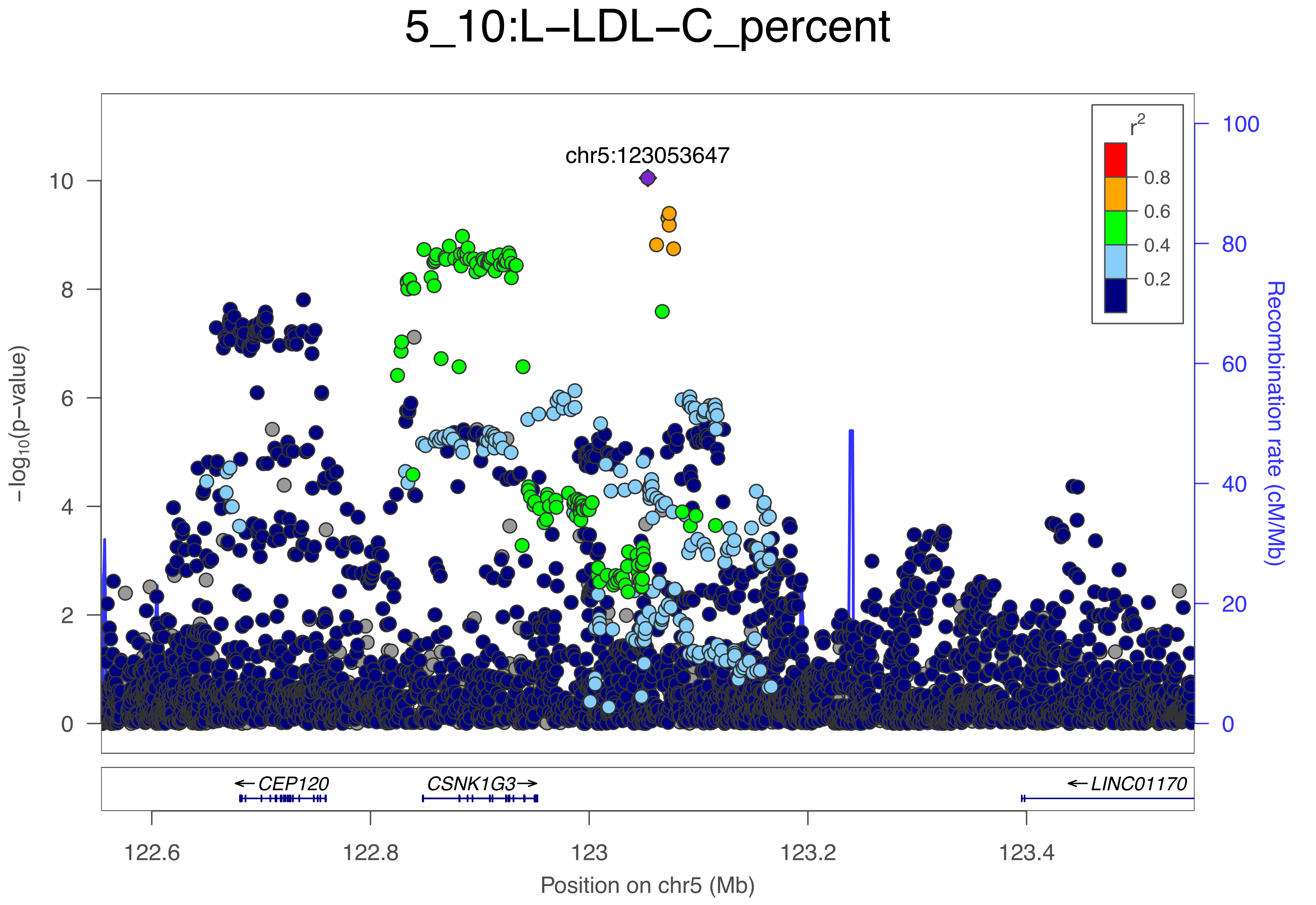

5_10:L−LDL−C_percent
chr5:123053647
−log10(p−value)
0
2
4
6
8
10
Recombination rate (cM/Mb)
0
20
40
60
80
100
r2
0.8
0.6
0.4
0.2
← CEP120
CSNK1G3 →
← LINC01170
122.6
122.8
123
123.2
123.4
Position on chr5 (Mb)

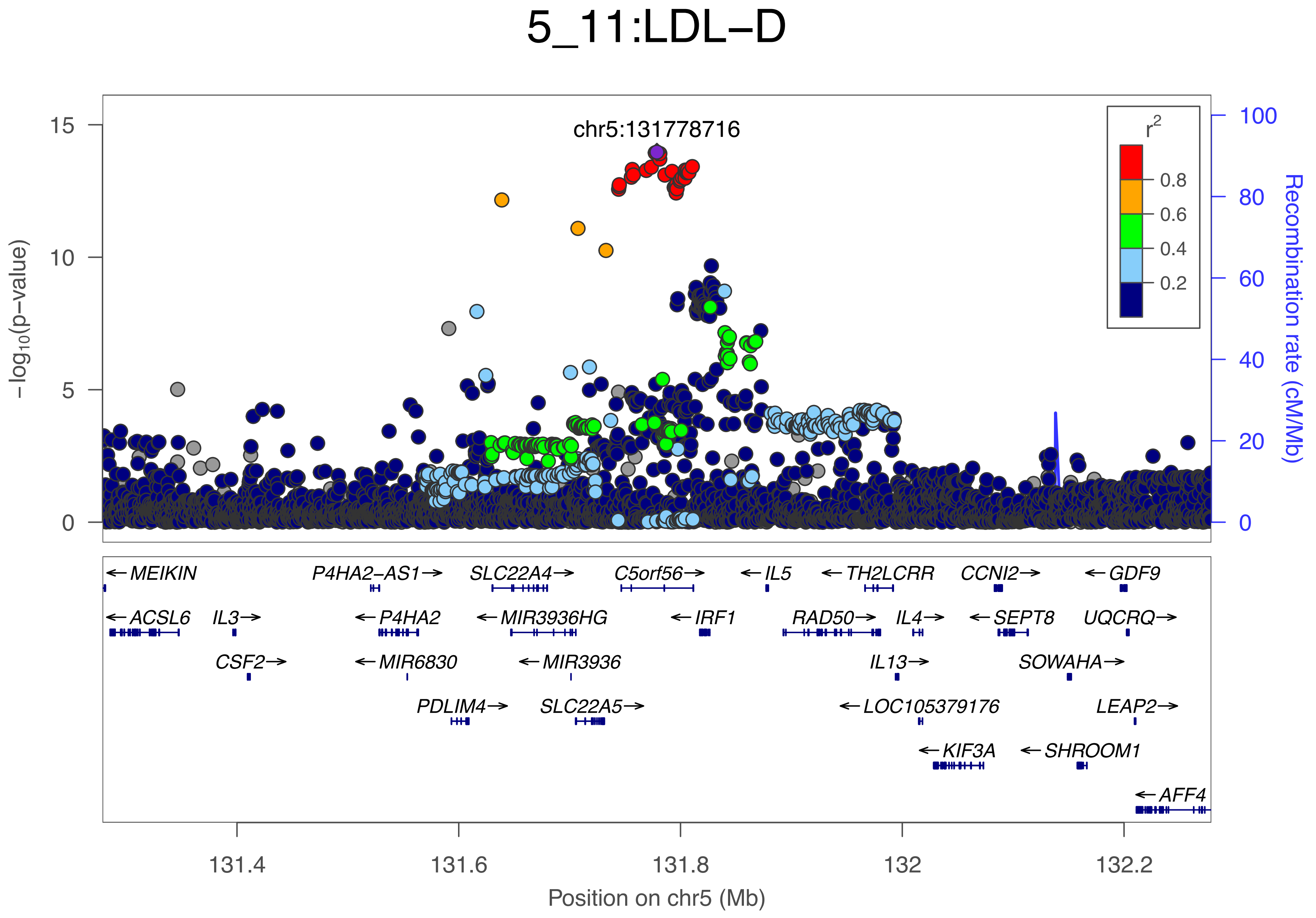

5_11:LDL−D
chr5:131778716
r2
0.8
0.6
0.4
0.2
−log10(p−value)
Recombination rate (cM/Mb)
15
10
5
0
100
80
60
40
20
0
←MEIKIN
P4HA2−AS1→
SLC22A4→
C5orf56→
←IL5
←TH2LCRR
CCNI2→
←GDF9
←ACSL6
IL3→
←P4HA2
←MIR3936HG
←IRF1
RAD50→
IL4→
←SEPT8
UQCRQ→
CSF2→
←MIR6830
←MIR3936
IL13→
SOWAHA→
PDLIM4→
SLC22A5→
←LOC105379176
LEAP2→
←KIF3A
←SHROOM1
←AFF4
131.4
131.6
131.8
132
132.2
Position on chr5 (Mb)

### 5_12:Ala

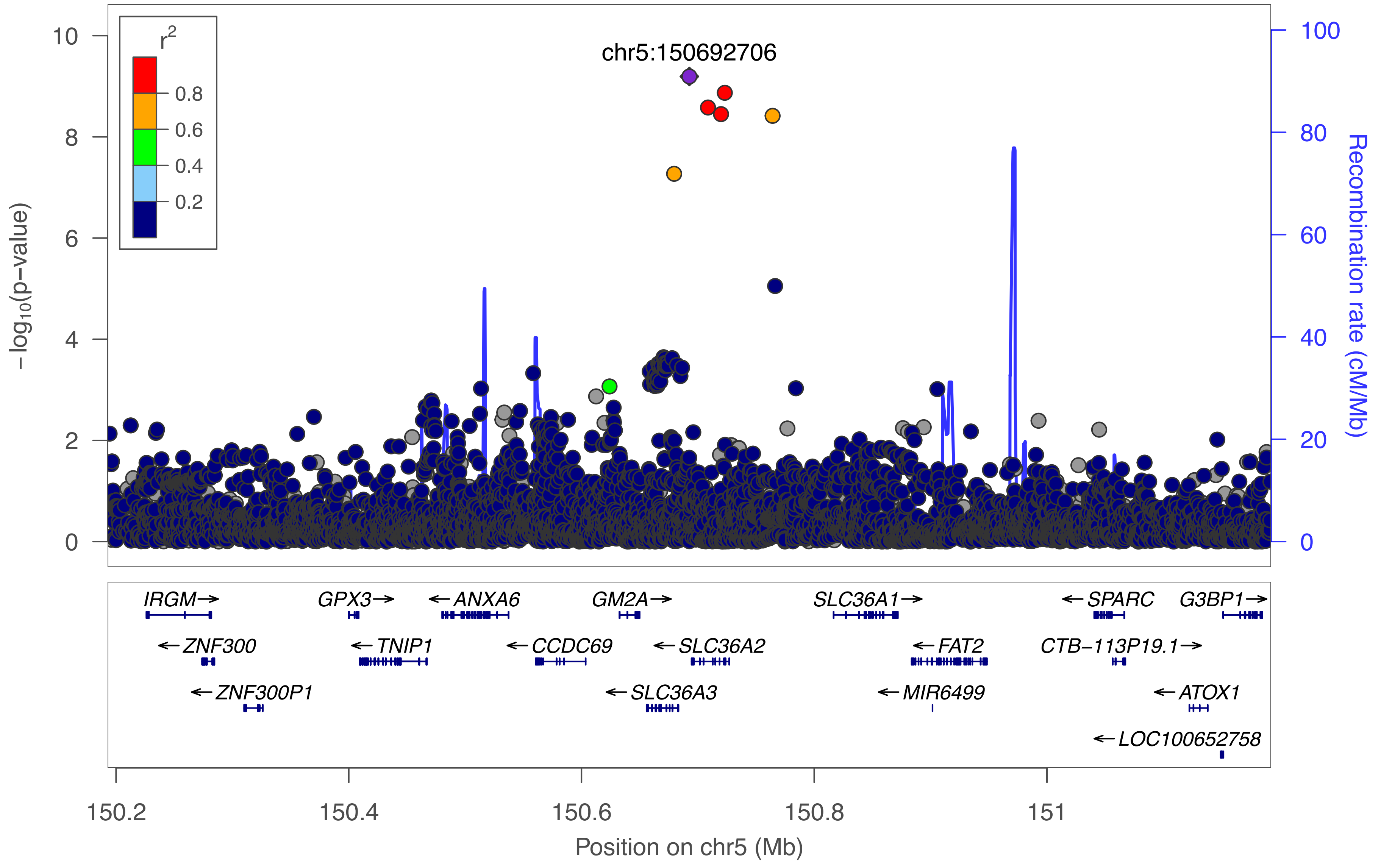

r²
0.8
0.6
0.4
0.2
chr5:150692706
−log10(p−value)
Recombination rate (cM/Mb)
0
2
4
6
8
10
0
20
40
60
80
100
IRGM→
GPX3→
←ANXA6
GM2A→
SLC36A1→
←SPARC
G3BP1→
←ZNF300
←TNIP1
←CCDC69
←SLC36A2
←FAT2
CTB−113P19.1→
←ZNF300P1
←SLC36A3
←MIR6499
←ATOX1
←LOC100652758
150.2
150.4
150.6
150.8
151
Position on chr5 (Mb)

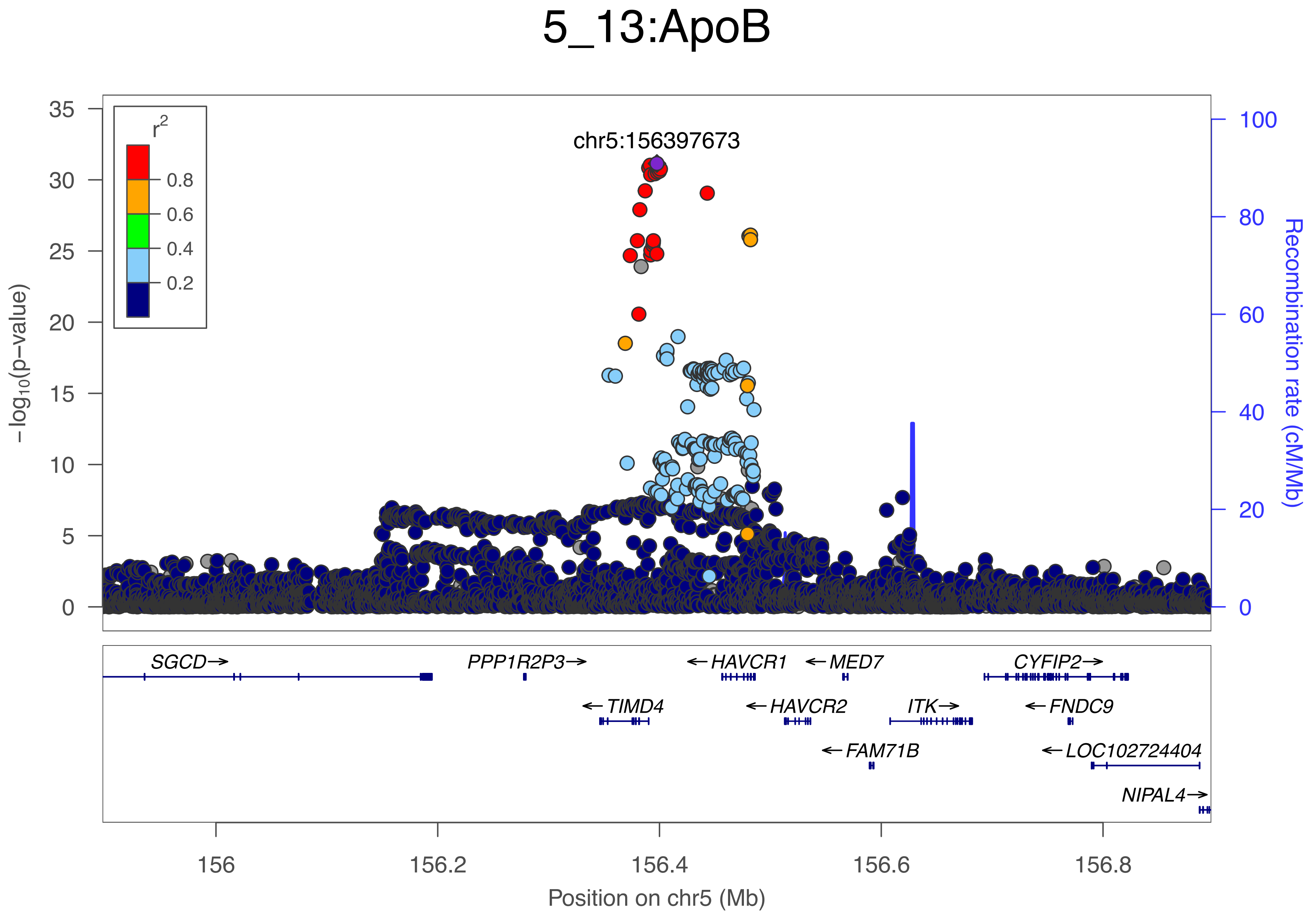

5_13:ApoB
chr5:156397673
r2
0.8
0.6
0.4
0.2
−log10(p−value)
Recombination rate (cM/Mb)
SGCD→
PPP1R2P3→
←HAVCR1
←MED7
CYFIP2→
←TIMD4
←HAVCR2
ITK→
←FNDC9
←FAM71B
←LOC102724404
NIPAL4→
156
156.2
156.4
156.6
156.8
Position on chr5 (Mb)

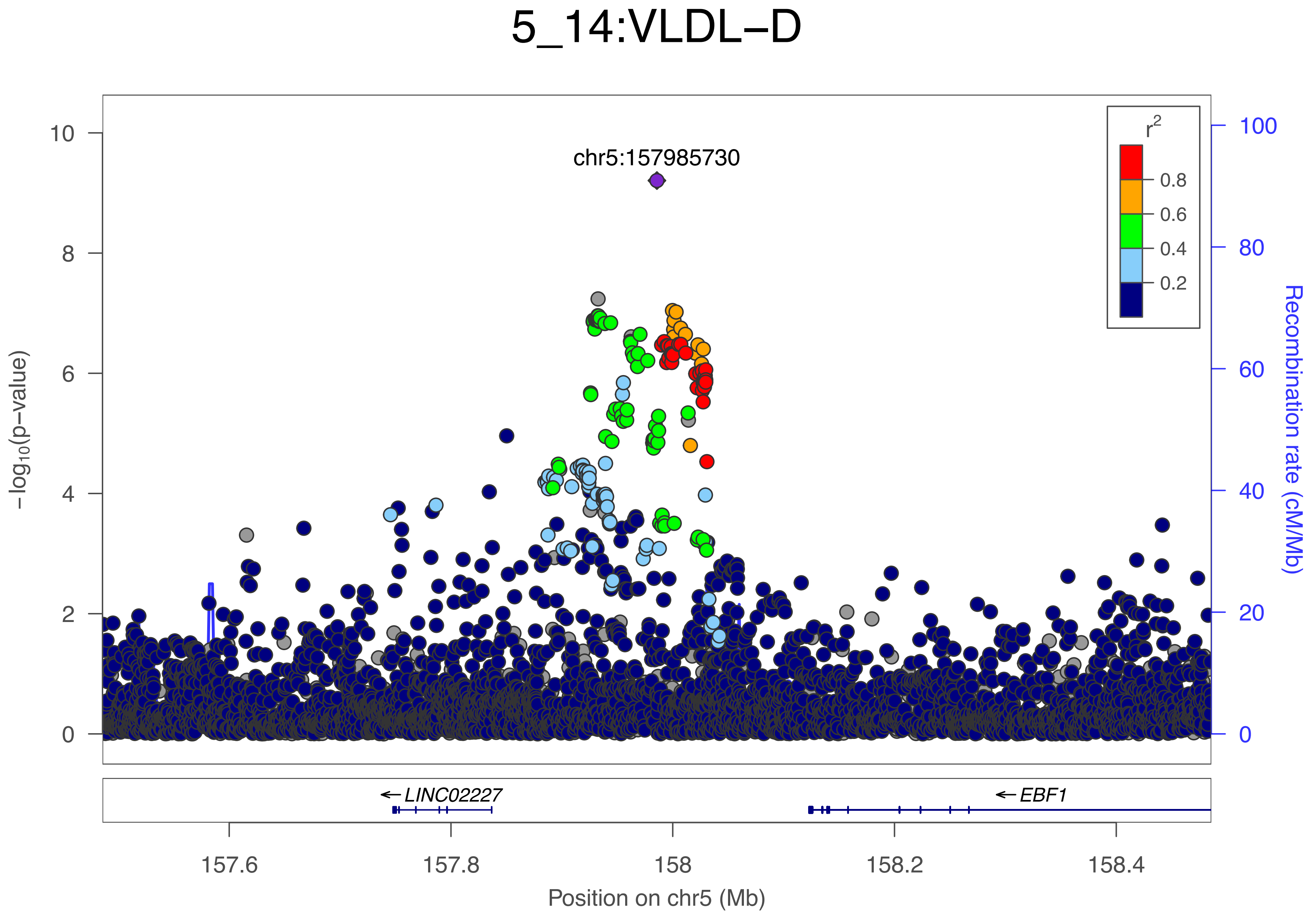

5_14:VLDL−D
chr5:157985730
r2
0.8
0.6
0.4
0.2
−log10(p−value)
10
8
6
4
2
0
Recombination rate (cM/Mb)
100
80
60
40
20
0
LINC02227
EBF1
157.6
157.8
158
158.2
158.4
Position on chr5 (Mb)

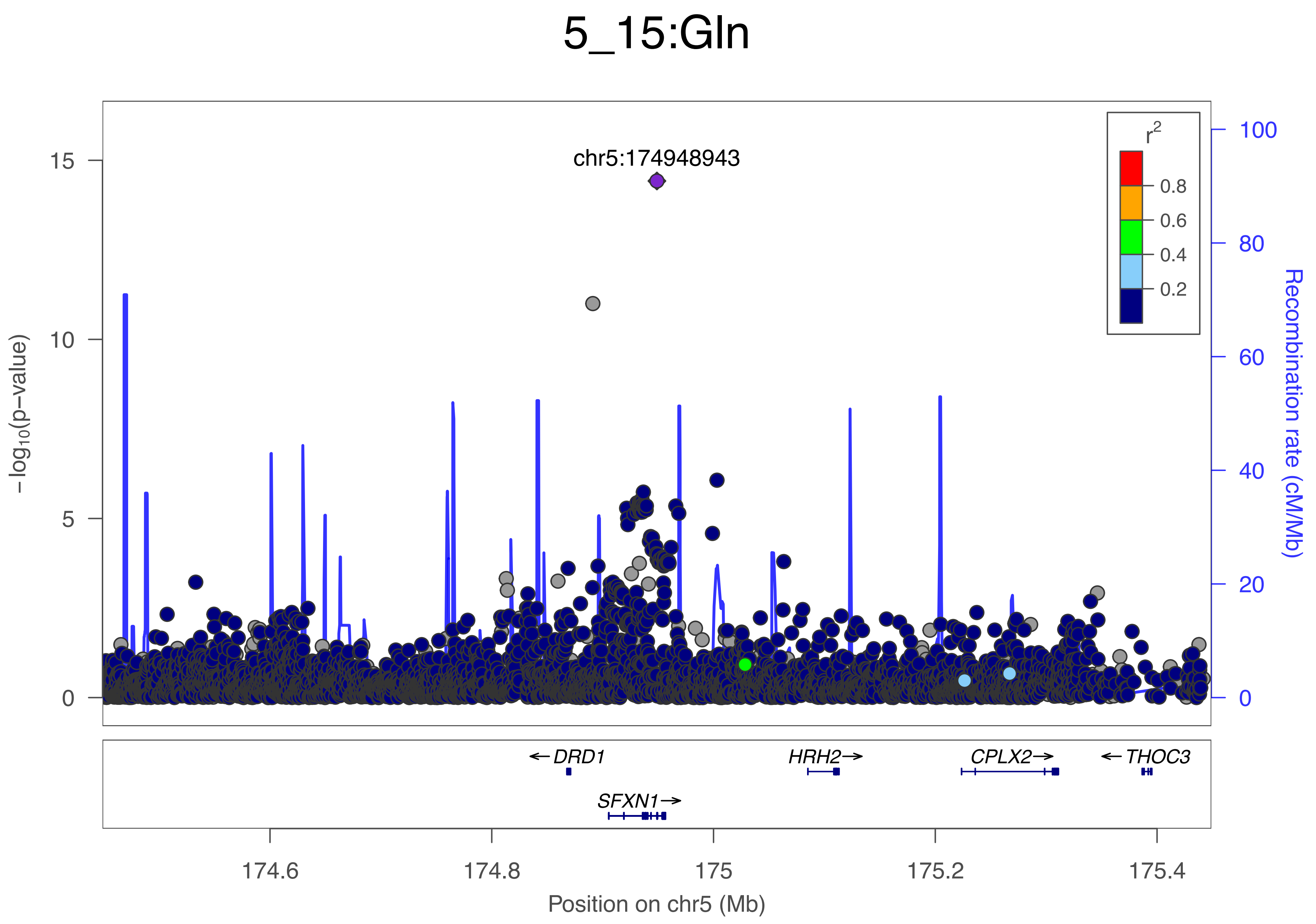

5_15:Gln
chr5:174948943
r2
0.8
0.6
0.4
0.2
15
10
5
0
−log10(p−value)
100
80
60
40
20
0
Recombination rate (cM/Mb)
← DRD1
HRH2→
CPLX2→
← THOC3
SFXN1→
174.6
174.8
175
175.2
175.4
Position on chr5 (Mb)

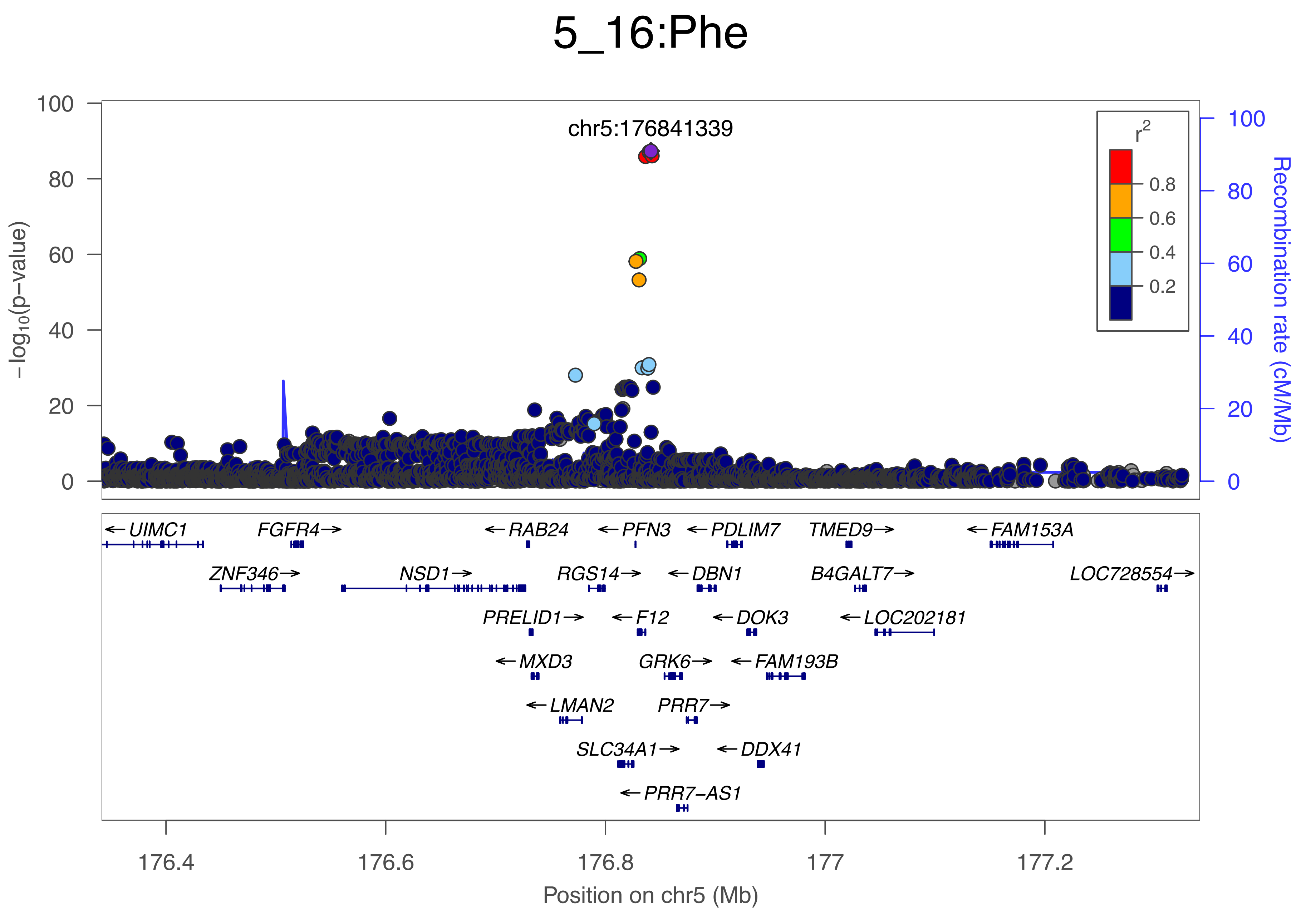

5_16:Phe
chr5:176841339
−log10(p−value)
Recombination rate (cM/Mb)
r2
0.8
0.6
0.4
0.2
0
20
40
60
80
100
UIMC1
FGFR4
RAB24
PFN3
PDLIM7
TMED9
FAM153A
ZNF346
NSD1
RGS14
DBN1
B4GALT7
LOC728554
PRELID1
F12
DOK3
LOC202181
MXD3
GRK6
FAM193B
LMAN2
PRR7
SLC34A1
DDX41
PRR7−AS1
176.4
176.6
176.8
177
177.2
Position on chr5 (Mb)

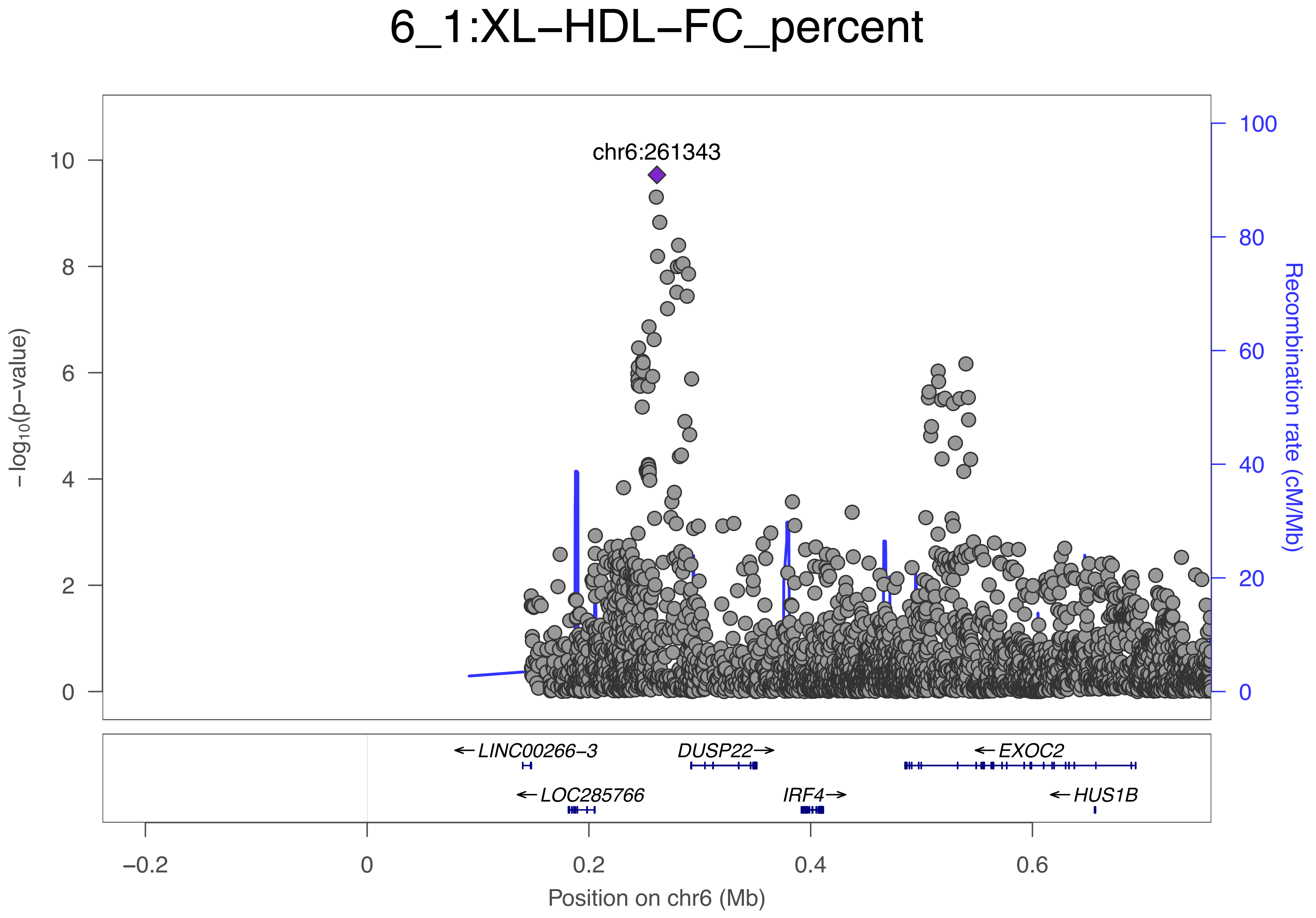

6_1:XL−HDL−FC_percent
chr6:261343
−log10(p−value)
Recombination rate (cM/Mb)
←LINC00266−3
DUSP22→
←EXOC2
←LOC285766
IRF4→
←HUS1B
Position on chr6 (Mb)

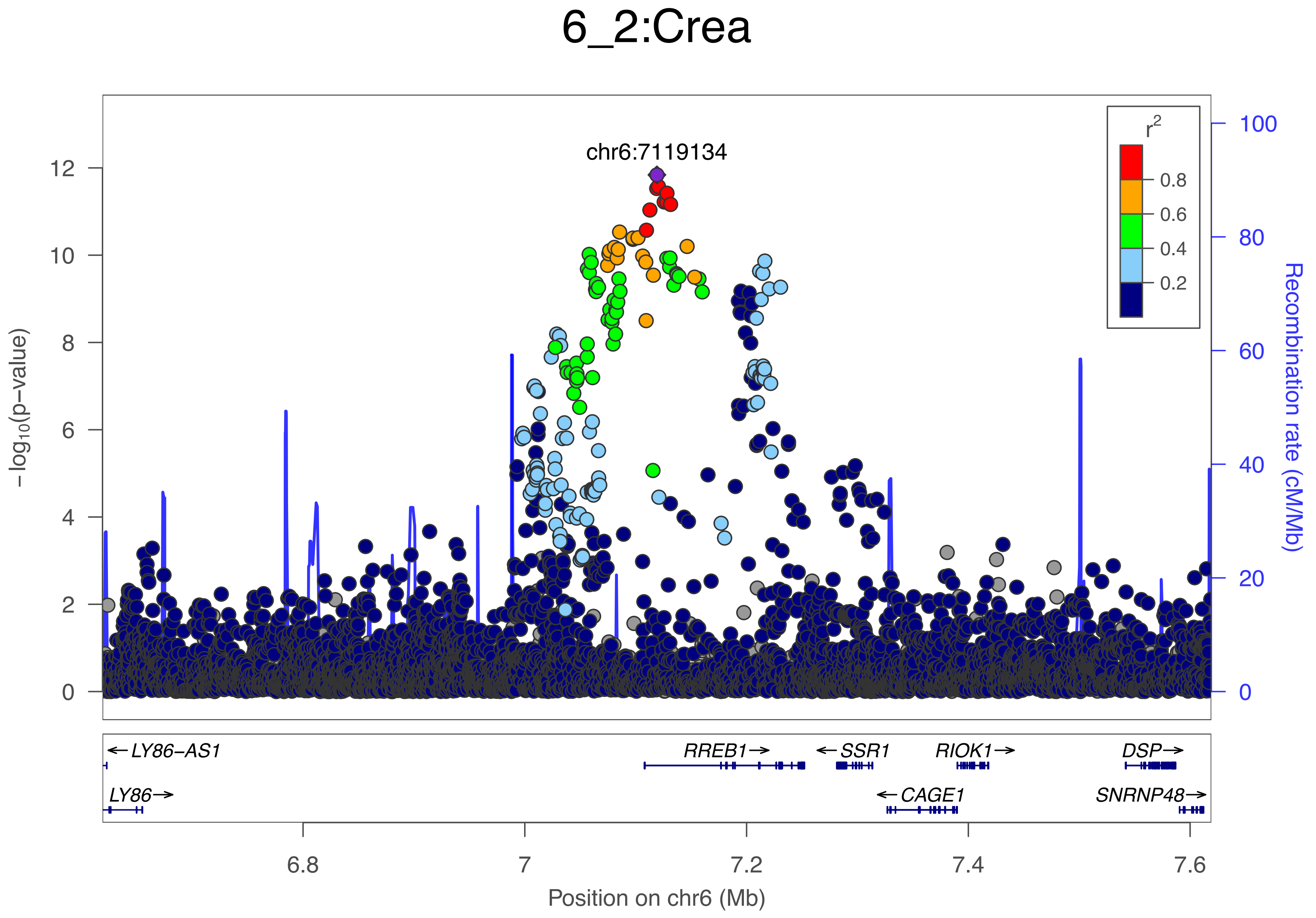

6_2:Crea
chr6:7119134
r²
0.8
0.6
0.4
0.2
−log10(p−value)
12
10
8
6
4
2
0
100
80
60
40
20
0
Recombination rate (cM/Mb)
← LY86−AS1
LY86 →
RREB1 →
← SSR1
RIOK1 →
DSP →
← CAGE1
SNRNP48 →
6.8
7
7.2
7.4
7.6
Position on chr6 (Mb)

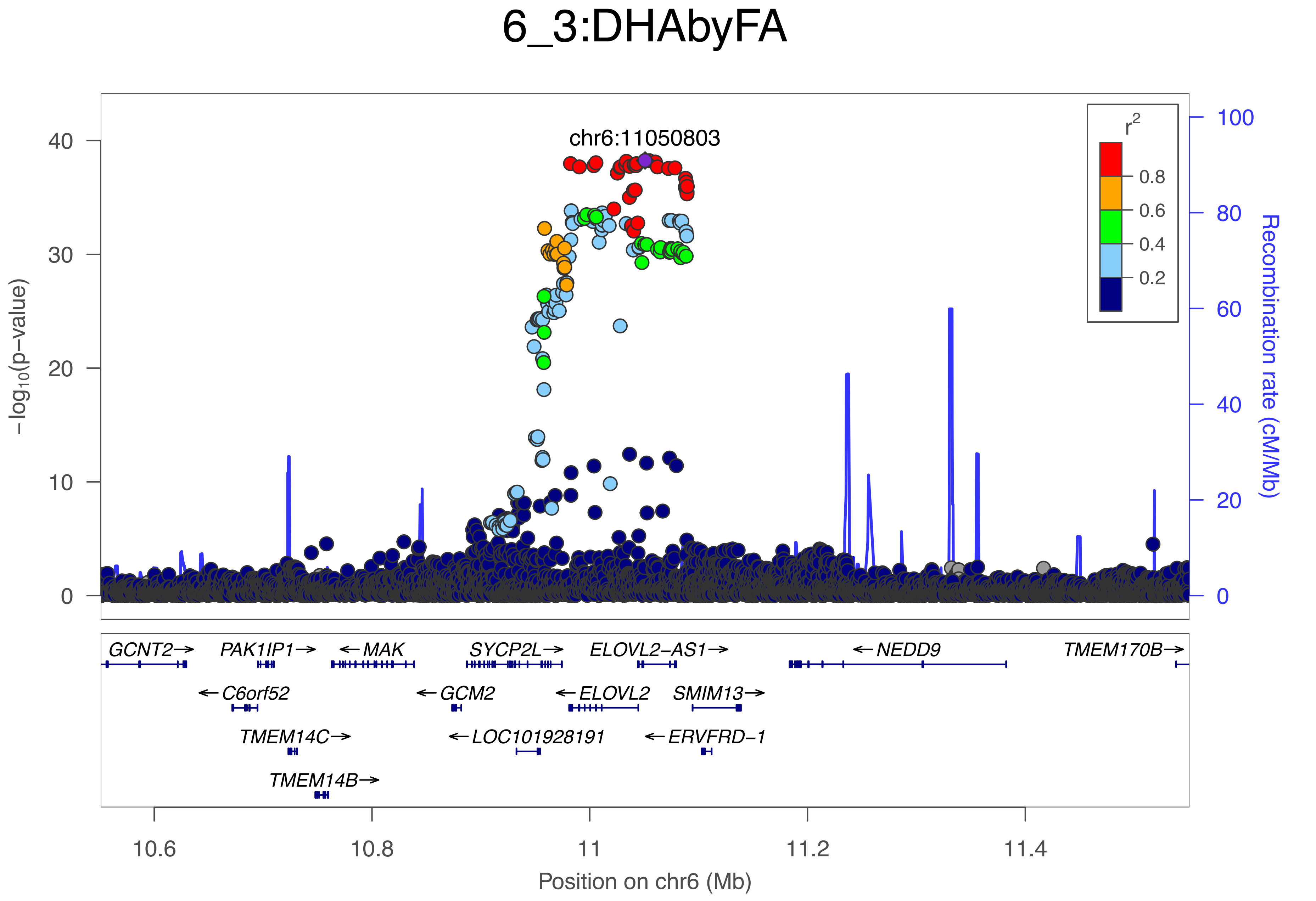
6_3:DHAbyFA
chr6:11050803
r2
0.8
0.6
0.4
0.2
-log10(p-value)
Recombination rate (cM/Mb)
0
10
20
30
40
60
80
100
GCNT2→
PAK1IP1→
← MAK
SYCP2L→
ELOVL2-AS1→
← NEDD9
TMEM170B→
← C6orf52
← GCM2
← ELOVL2
SMIM13→
TMEM14C→
← LOC101928191
← ERVFRD-1
TMEM14B→
10.6
10.8
11
11.2
11.4
Position on chr6 (Mb)

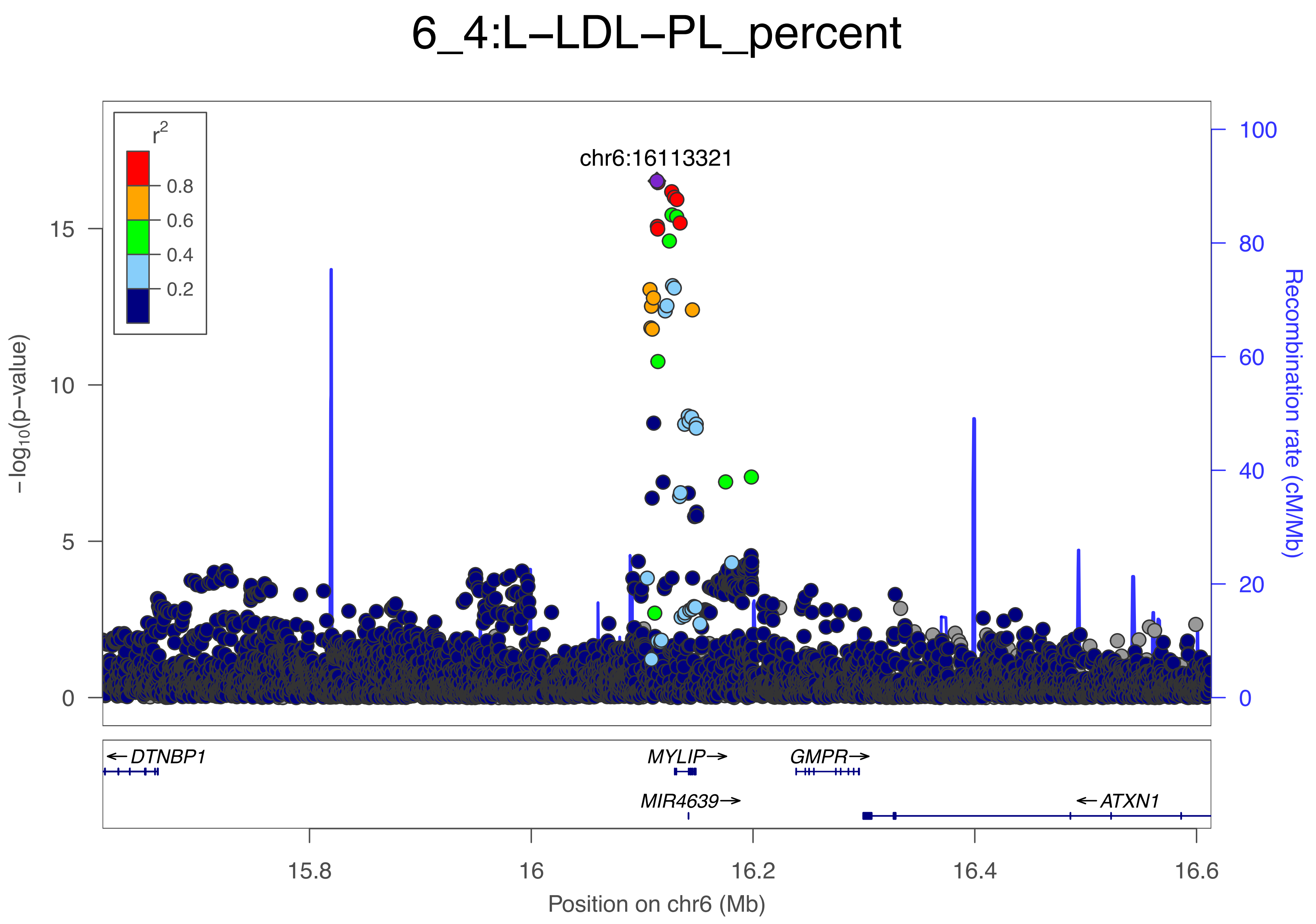
6_4:L−LDL−PL_percent
r2
0.8
0.6
0.4
0.2
chr6:16113321
15
10
5
0
−log10(p−value)
100
80
60
40
20
0
Recombination rate (cM/Mb)
← DTNBP1
MYLIP →
GMPR →
MIR4639 →
← ATXN1
15.8
16
16.2
16.4
16.6
Position on chr6 (Mb)

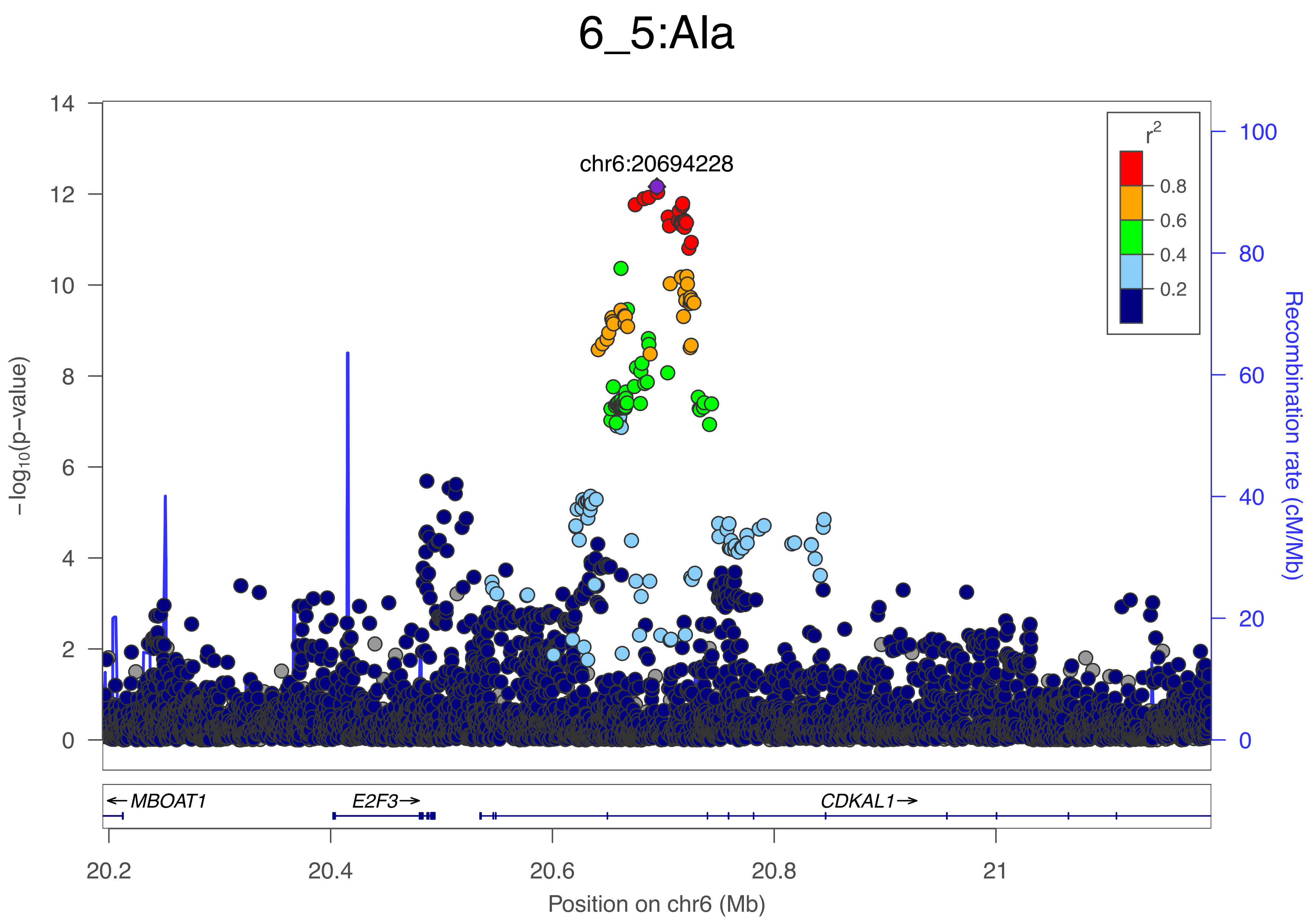

6_5:Ala
chr6:20694228
r2
0.8
0.6
0.4
0.2
−log10(p−value)
Recombination rate (cM/Mb)
14
12
10
8
6
4
2
0
100
80
60
40
20
0
← MBOAT1
E2F3→
CDKAL1→
20.2
20.4
20.6
20.8
21
Position on chr6 (Mb)

### 6_6:SFAbyFA

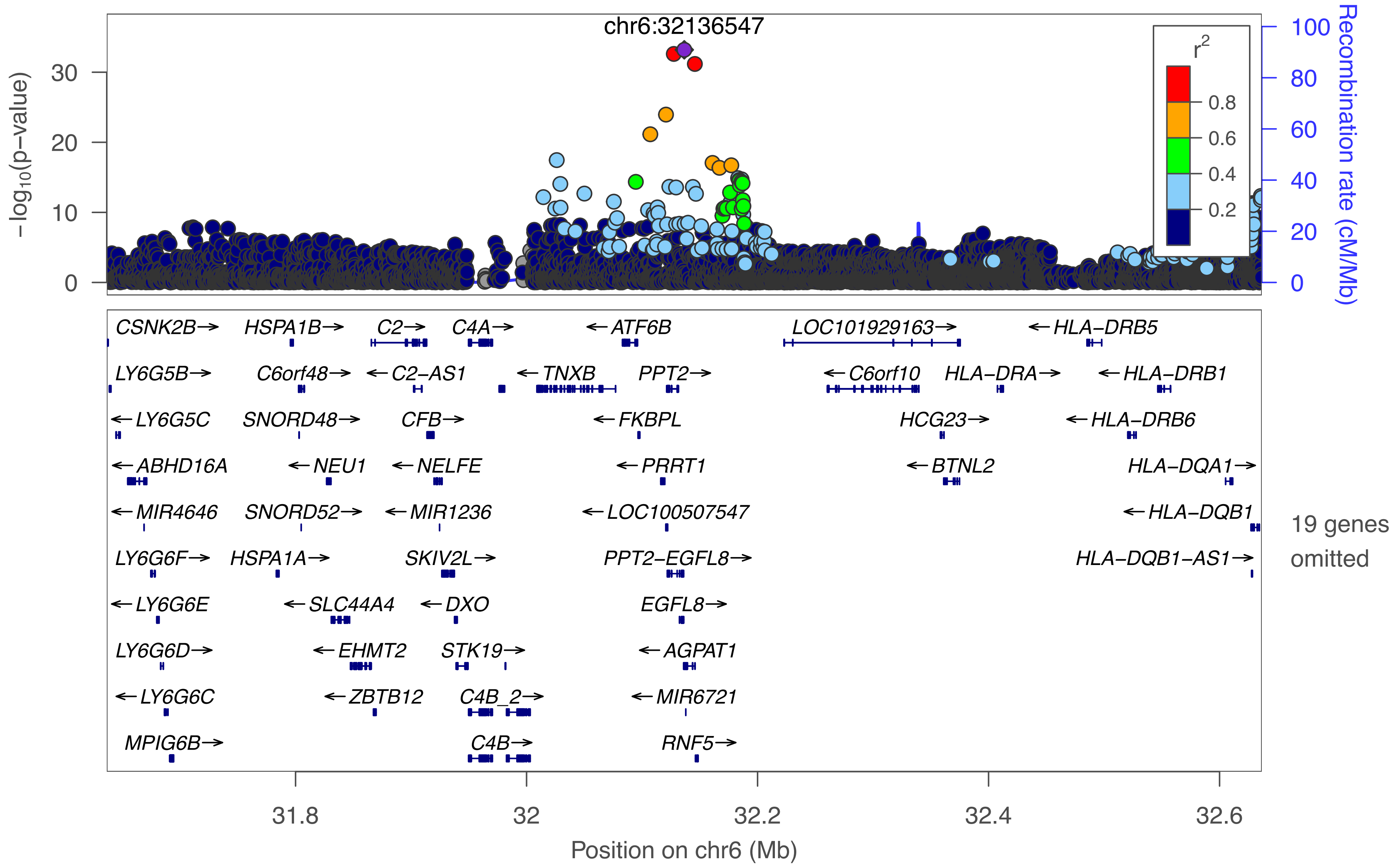

chr6:32136547
−log10(p−value)
Recombination rate (cM/Mb)
r2
0.8
0.6
0.4
0.2
0
10
20
30
0
20
40
60
80
100
CSNK2B→
HSPA1B→
C2→
C4A→
←ATF6B
LOC101929163→
←HLA−DRB5
LY6G5B→
C6orf48→
←C2−AS1
←TNXB
PPT2→
←C6orf10
HLA−DRA→
←HLA−DRB1
←LY6G5C
SNORD48→
CFB→
←FKBPL
HCG23→
←HLA−DRB6
←ABHD16A
←NEU1
←NELFE
←PRRT1
←BTNL2
HLA−DQA1→
←MIR4646
SNORD52→
←MIR1236
←LOC100507547
←HLA−DQB1
LY6G6F→
HSPA1A→
SKIV2L→
PPT2−EGFL8→
HLA−DQB1−AS1→
←LY6G6E
←SLC44A4
←DXO
EGFL8→
LY6G6D→
←EHMT2
STK19→
←AGPAT1
←LY6G6C
←ZBTB12
C4B_2→
←MIR6721
MPIG6B→
C4B→
RNF5→
19 genes omitted
31.8
32
32.2
32.4
32.6
Position on chr6 (Mb)

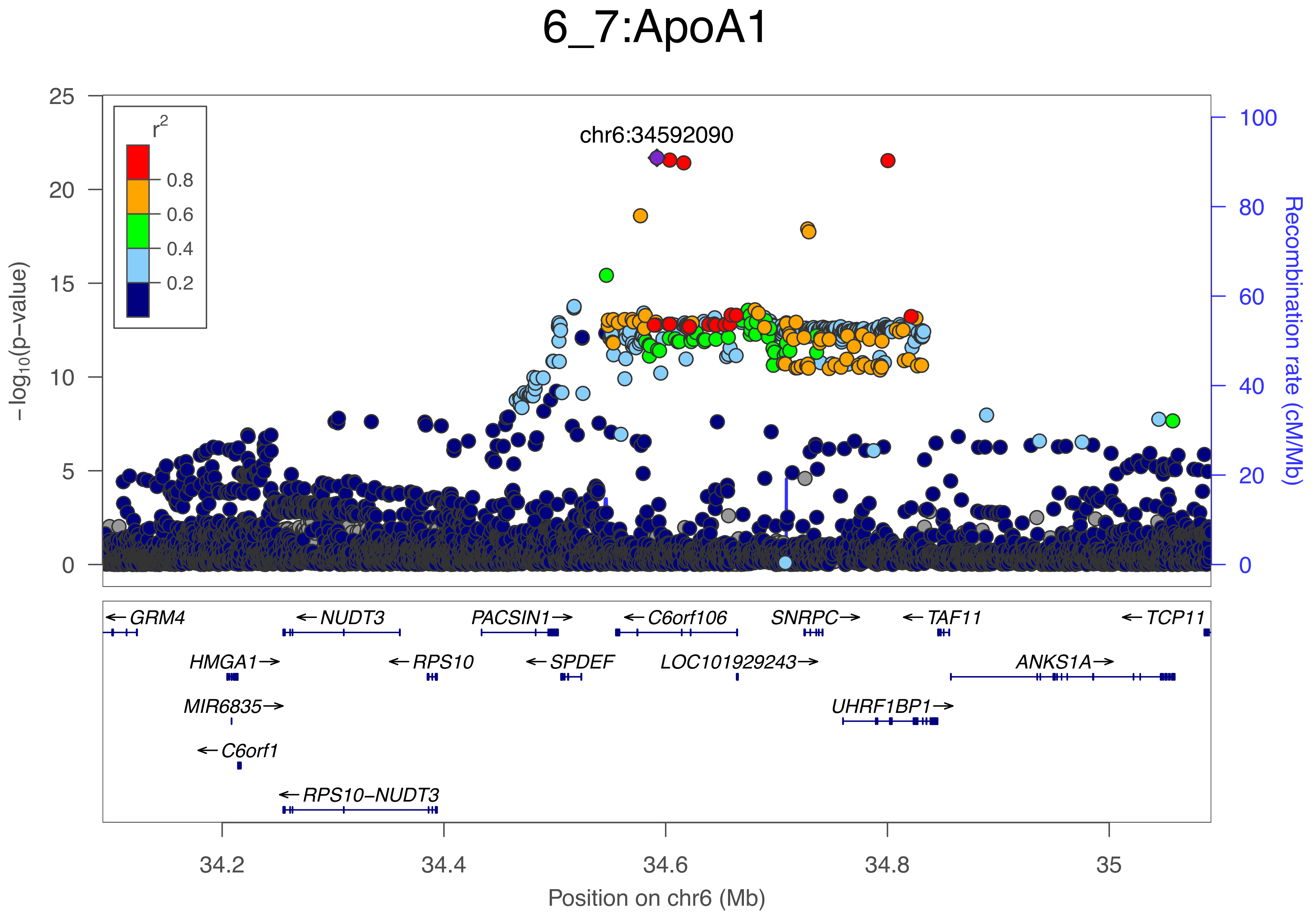

6_7:ApoA1
r²
0.8
0.6
0.4
0.2
chr6:34592090
−log10(p−value)
Recombination rate (cM/Mb)
GRM4
NUDT3
PACSIN1
C6orf106
SNRPC
TAF11
TCP11
HMGA1
RPS10
SPDEF
LOC101929243
ANKS1A
MIR6835
UHRF1BP1
C6orf1
RPS10−NUDT3
34.2
34.4
34.6
34.8
35
Position on chr6 (Mb)

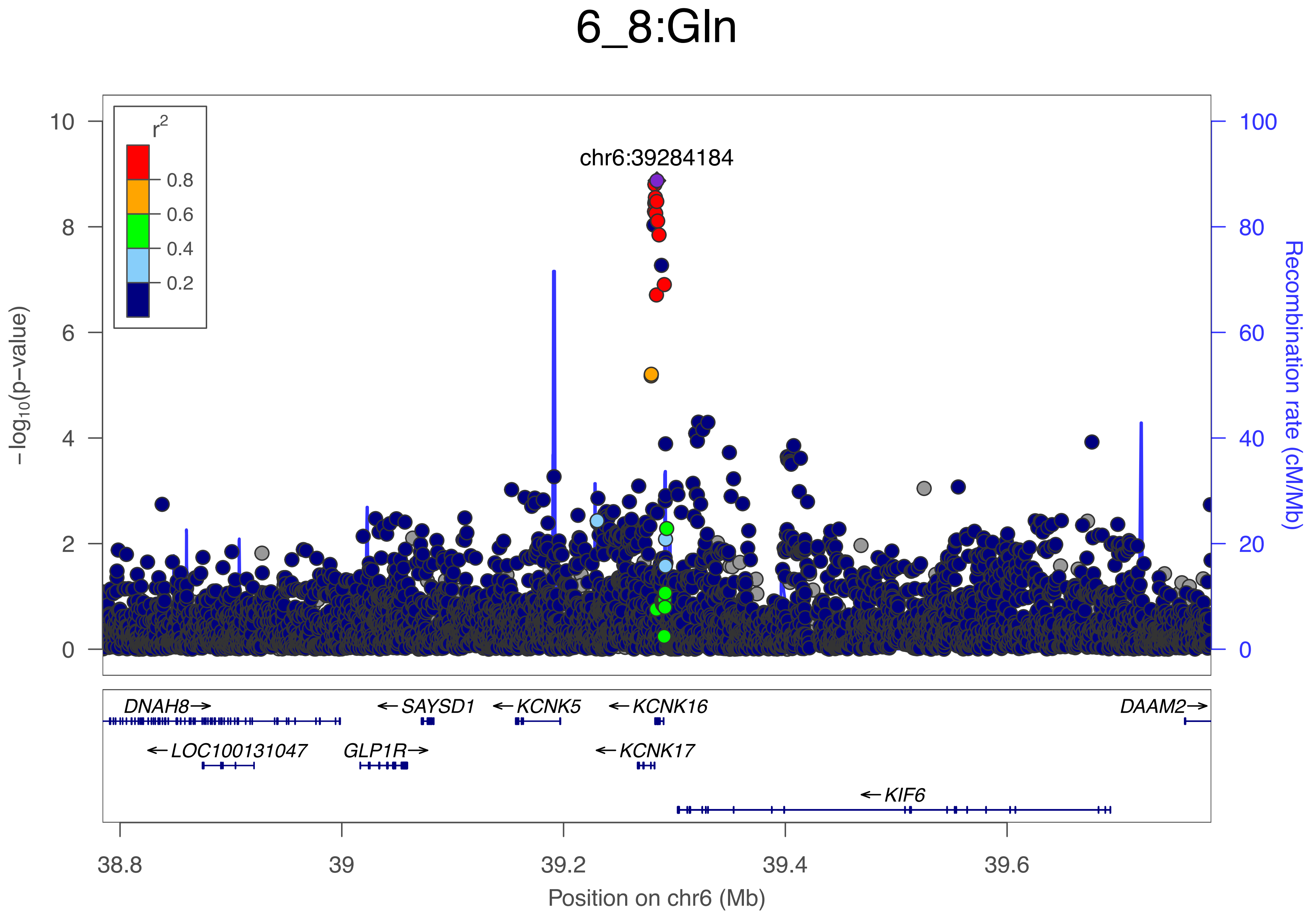
6_8:Gln
chr6:39284184
r2
0.8
0.6
0.4
0.2
10
8
6
4
2
0
−log10(p−value)
100
80
60
40
20
0
Recombination rate (cM/Mb)
DNAH8→
←SAYSD1
←KCNK5
←KCNK16
DAAM2→
←LOC100131047
GLP1R→
←KCNK17
←KIF6
38.8
39
39.2
39.4
39.6
Position on chr6 (Mb)

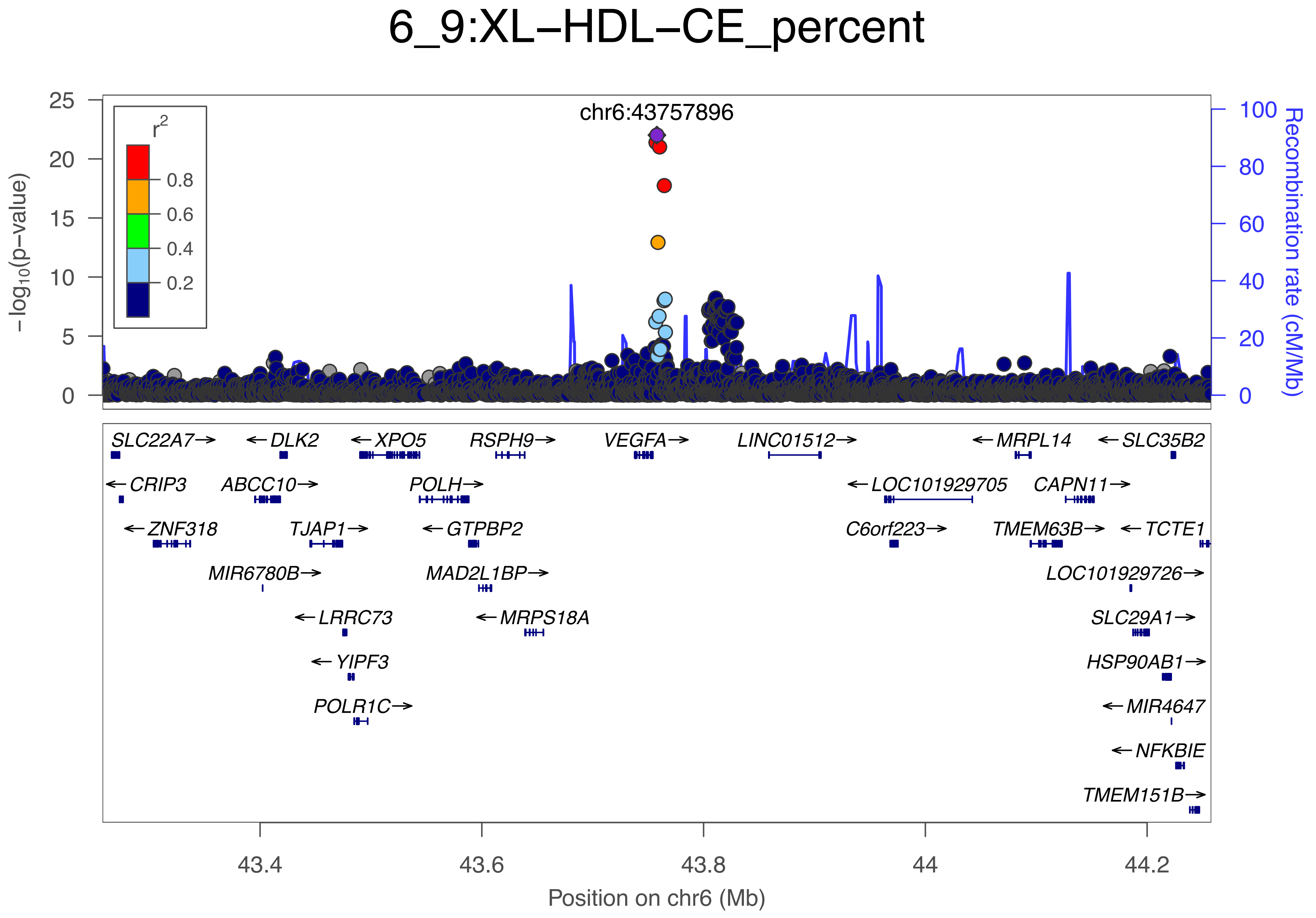
6_9:XL−HDL−CE_percent
chr6:43757896
r2
0.8
0.6
0.4
0.2
−log10(p−value)
Recombination rate (cM/Mb)
25
20
15
10
5
0
100
80
60
40
20
0
SLC22A7→
←DLK2
←XPO5
RSPH9→
VEGFA→
LINC01512→
←MRPL14
←SLC35B2
←CRIP3
ABCC10→
POLH→
←LOC101929705
CAPN11→
←ZNF318
TJAP1→
←GTPBP2
C6orf223→
TMEM63B→
←TCTE1
MIR6780B→
MAD2L1BP→
LOC101929726→
←LRRC73
←MRPS18A
SLC29A1→
←YIPF3
HSP90AB1→
POLR1C→
←MIR4647
←NFKBIE
TMEM151B→
43.4
43.6
43.8
44
44.2
Position on chr6 (Mb)

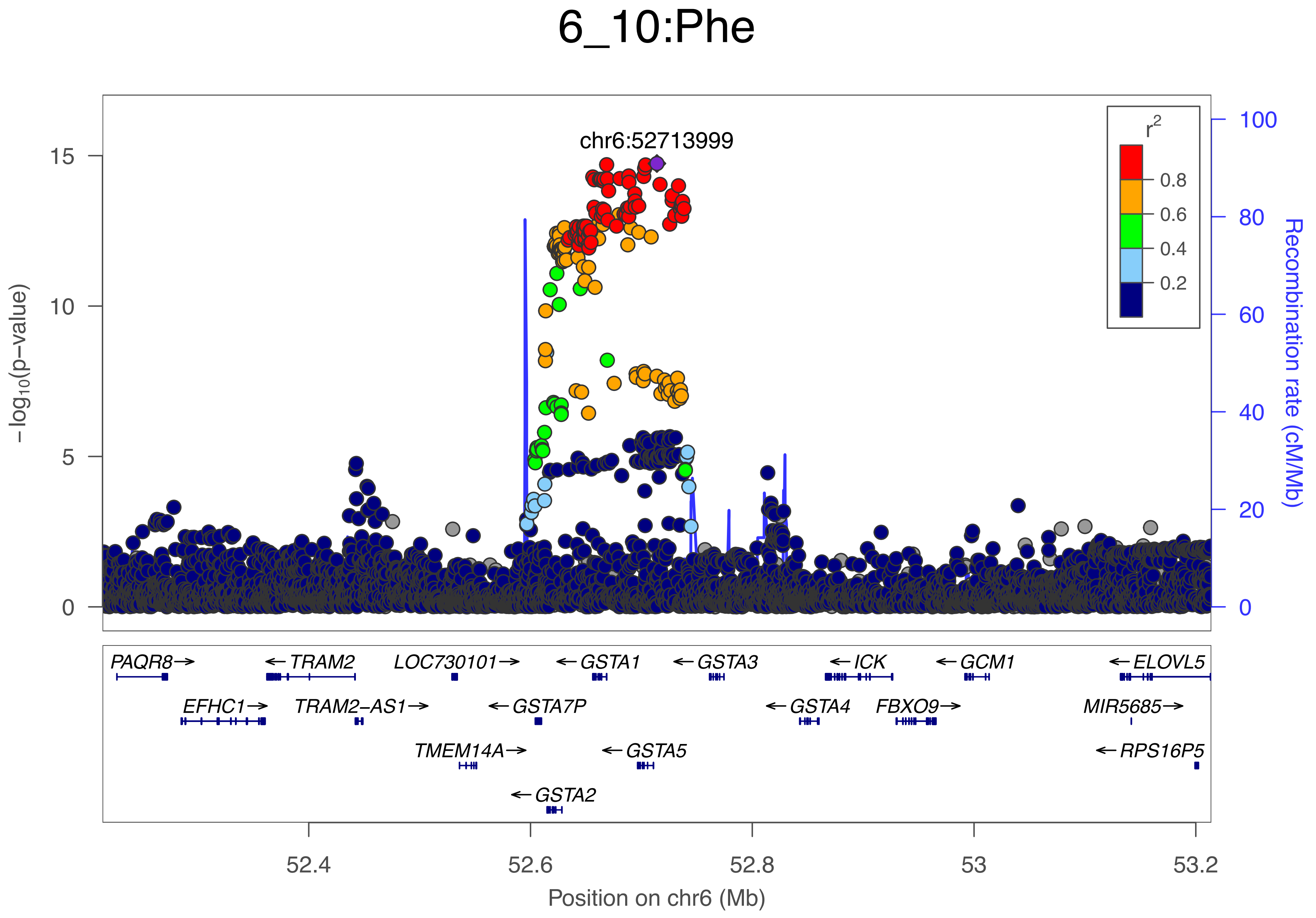

6_10:Phe
chr6:52713999
r2
0.8
0.6
0.4
0.2
−log10(p−value)
Recombination rate (cM/Mb)
15
10
5
0
100
80
60
40
20
0
PAQR8→
←TRAM2
LOC730101→
←GSTA1
←GSTA3
←ICK
←GCM1
←ELOVL5
EFHC1→
TRAM2−AS1→
←GSTA7P
←GSTA4
FBXO9→
MIR5685→
TMEM14A→
←GSTA5
←RPS16P5
←GSTA2
52.4
52.6
52.8
53
53.2
Position on chr6 (Mb)

### 6_11:TotFA

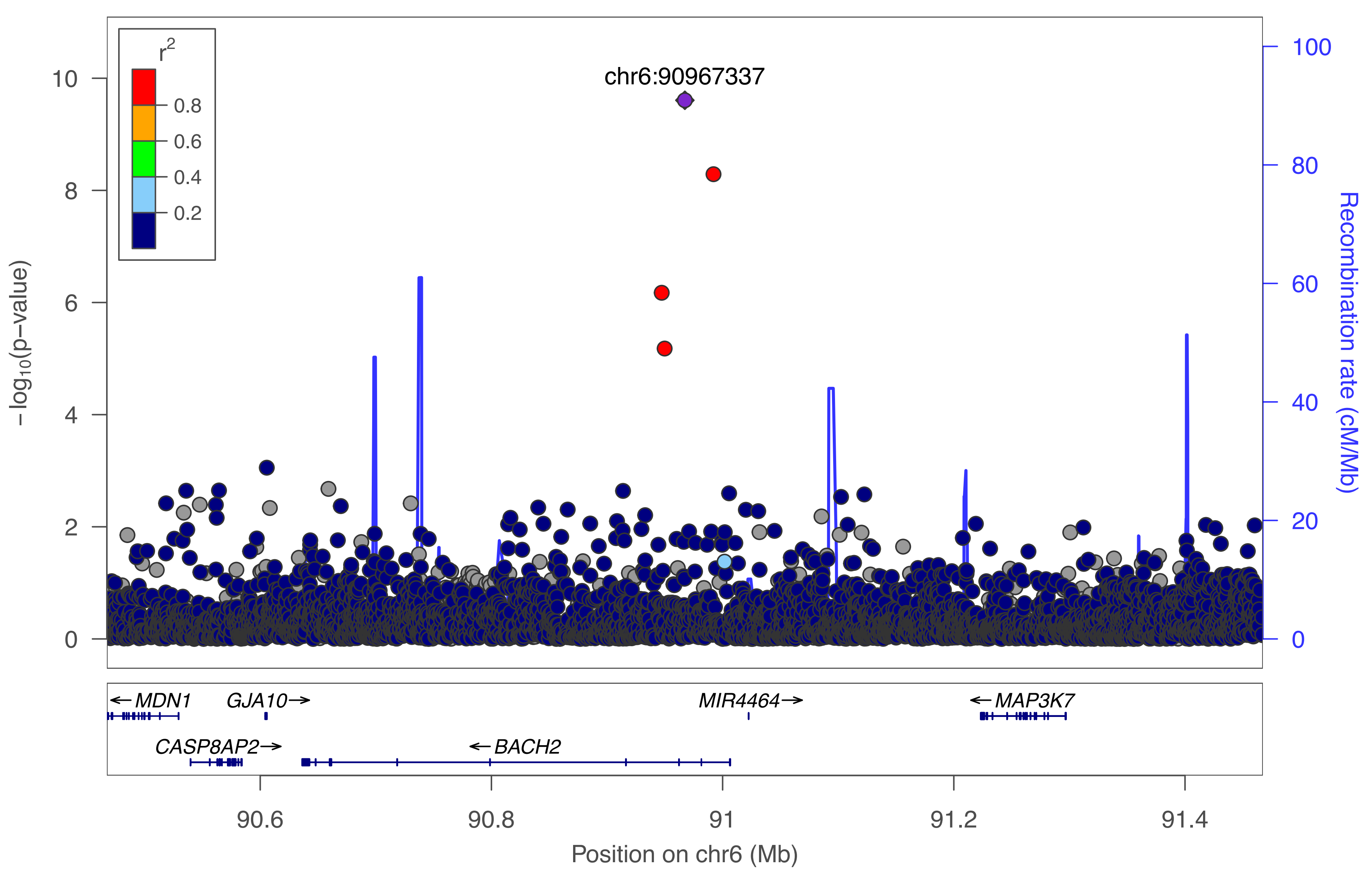

chr6:90967337
r2
0.8
0.6
0.4
0.2
−log10(p−value)
Recombination rate (cM/Mb)
MDN1
GJA10
MIR4464
MAP3K7
CASP8AP2
BACH2
Position on chr6 (Mb)

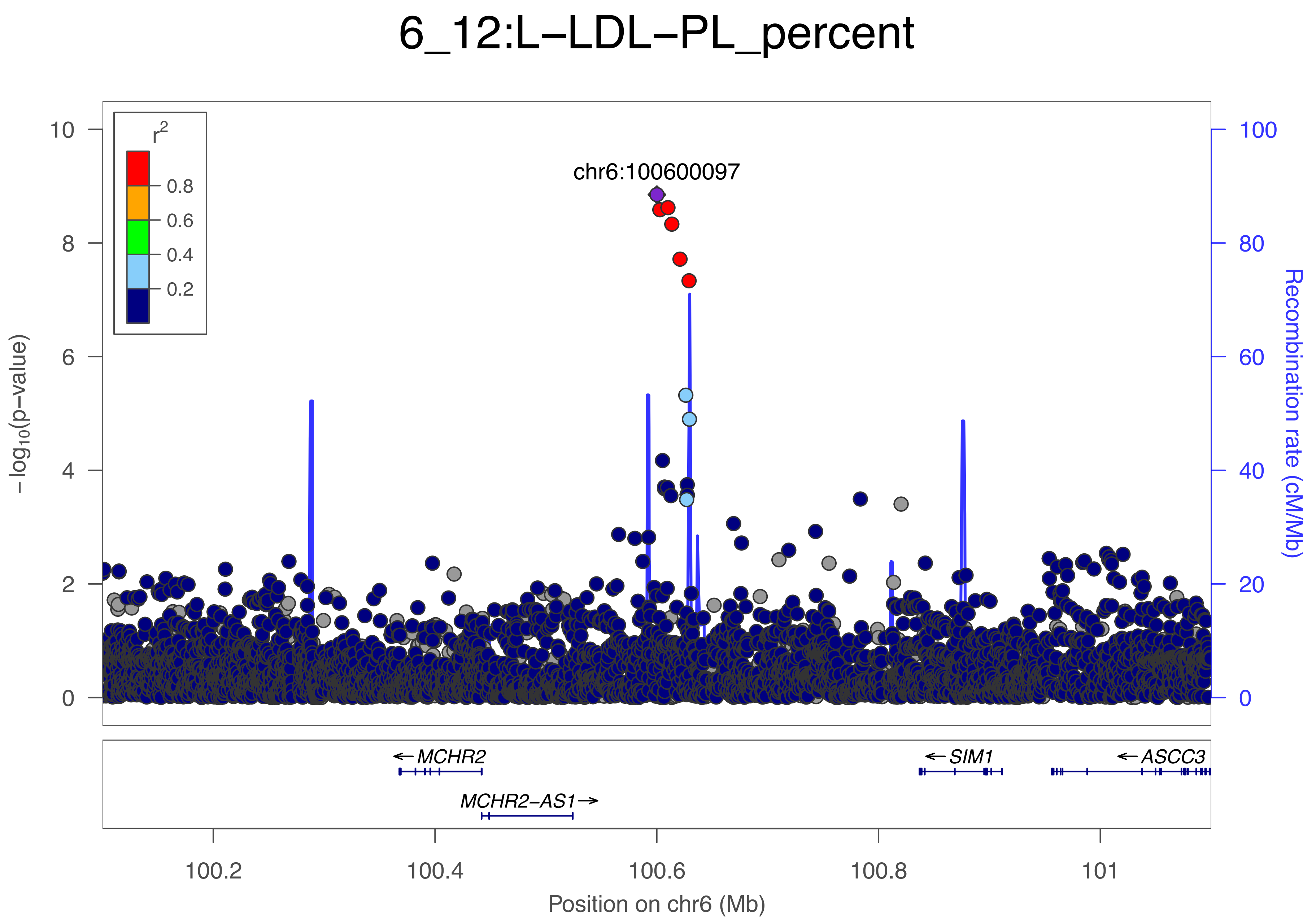

6_12:L−LDL−PL_percent
r2
0.8
0.6
0.4
0.2
chr6:100600097
−log10(p−value)
Recombination rate (cM/Mb)
10
8
6
4
2
0
100
80
60
40
20
0
MCHR2
MCHR2−AS1
SIM1
ASCC3
100.2
100.4
100.6
100.8
101
Position on chr6 (Mb)

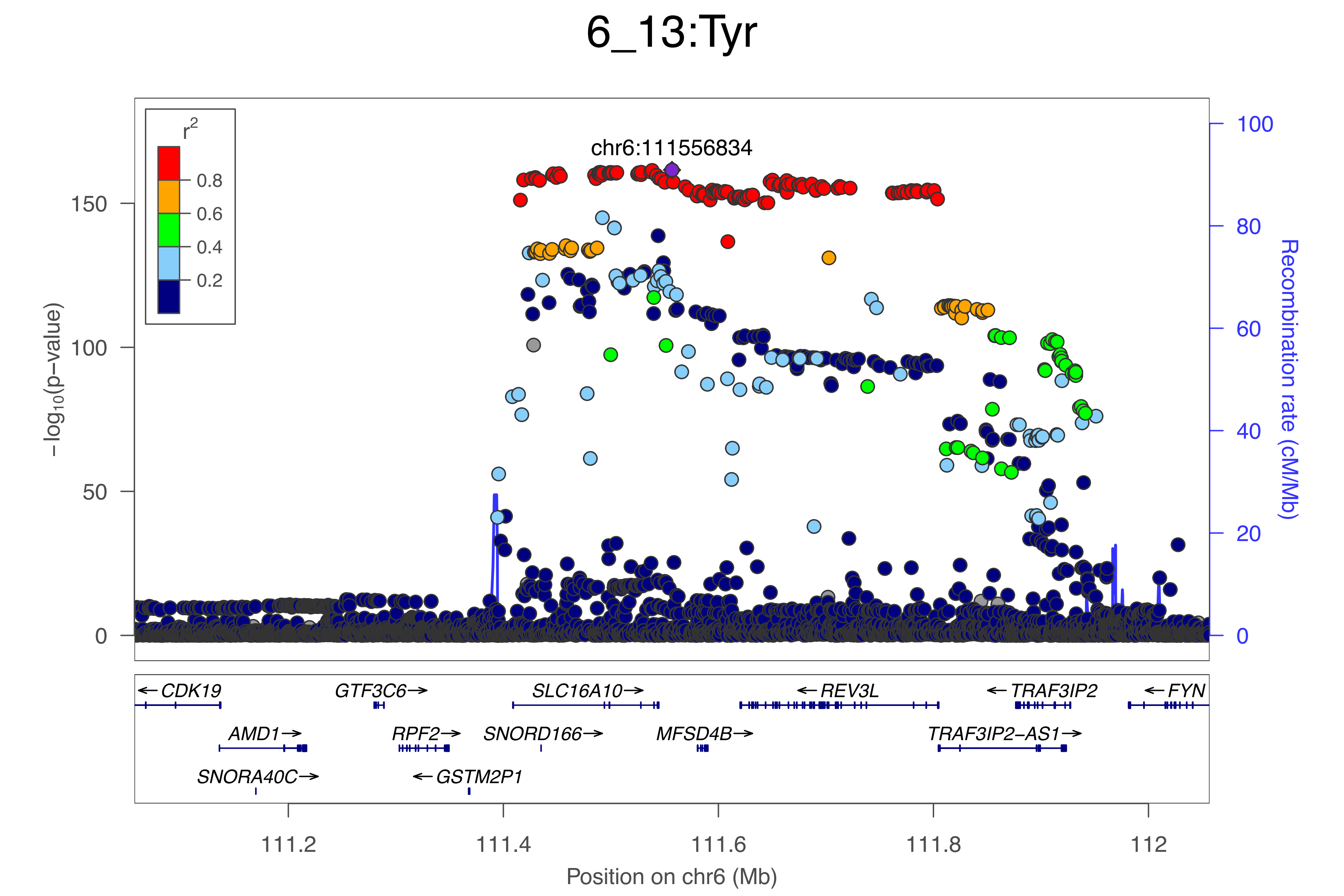

6_13:Tyr
chr6:111556834
r2
0.8
0.6
0.4
0.2
150
100
50
0
−log10(p−value)
100
80
60
40
20
0
Recombination rate (cM/Mb)
← CDK19
GTF3C6→
SLC16A10→
← REV3L
← TRAF3IP2
← FYN
AMD1→
RPF2→
SNORD166→
MFSD4B→
TRAF3IP2−AS1→
SNORA40C→
← GSTM2P1
111.2
111.4
111.6
111.8
112
Position on chr6 (Mb)

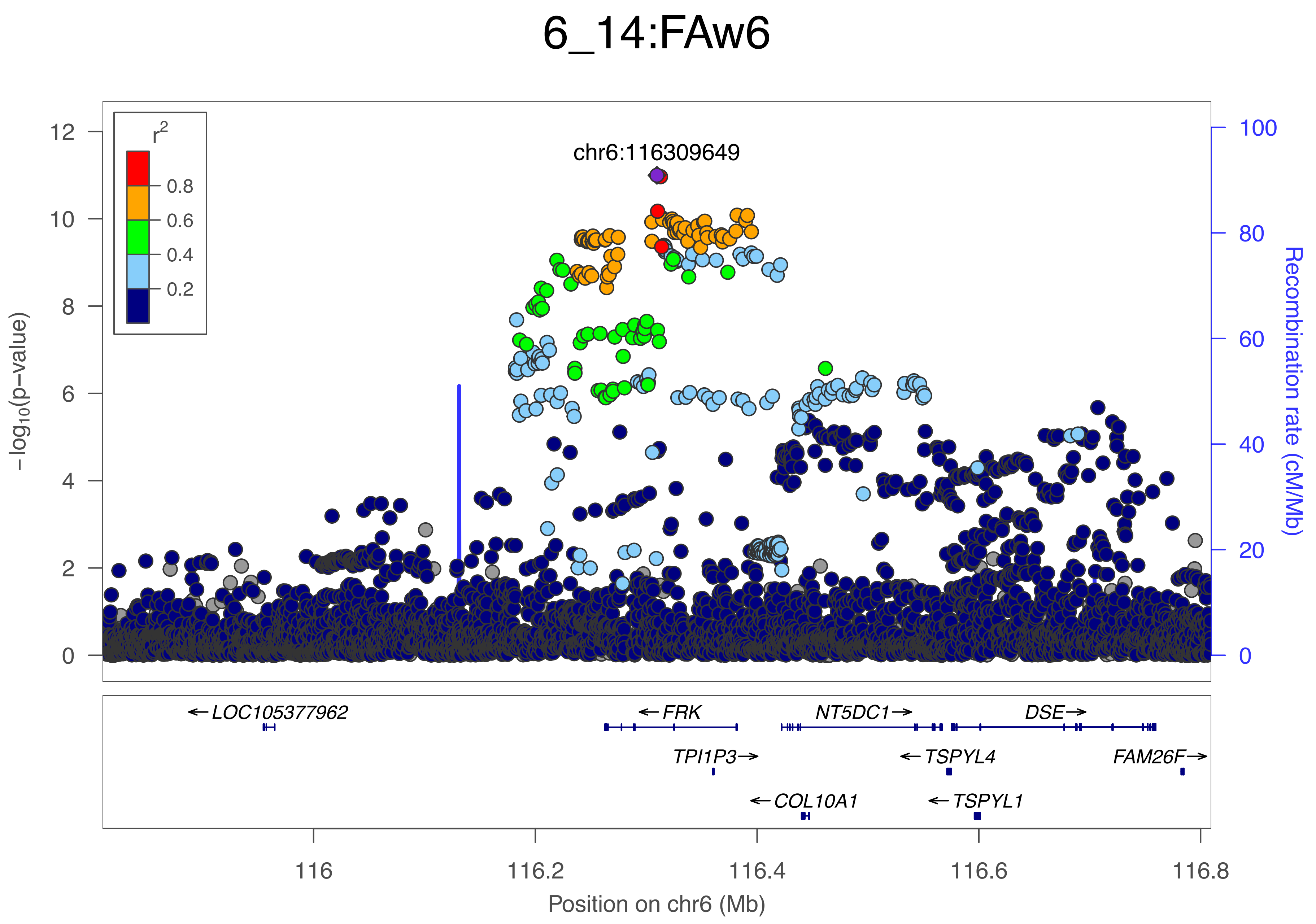
6_14:FAw6
chr6:116309649
r2
0.8
0.6
0.4
0.2
-log10(p-value)
Recombination rate (cM/Mb)
LOC105377962
FRK
NT5DC1
DSE
TPI1P3
TSPYL4
FAM26F
COL10A1
TSPYL1
116
116.2
116.4
116.6
116.8
Position on chr6 (Mb)

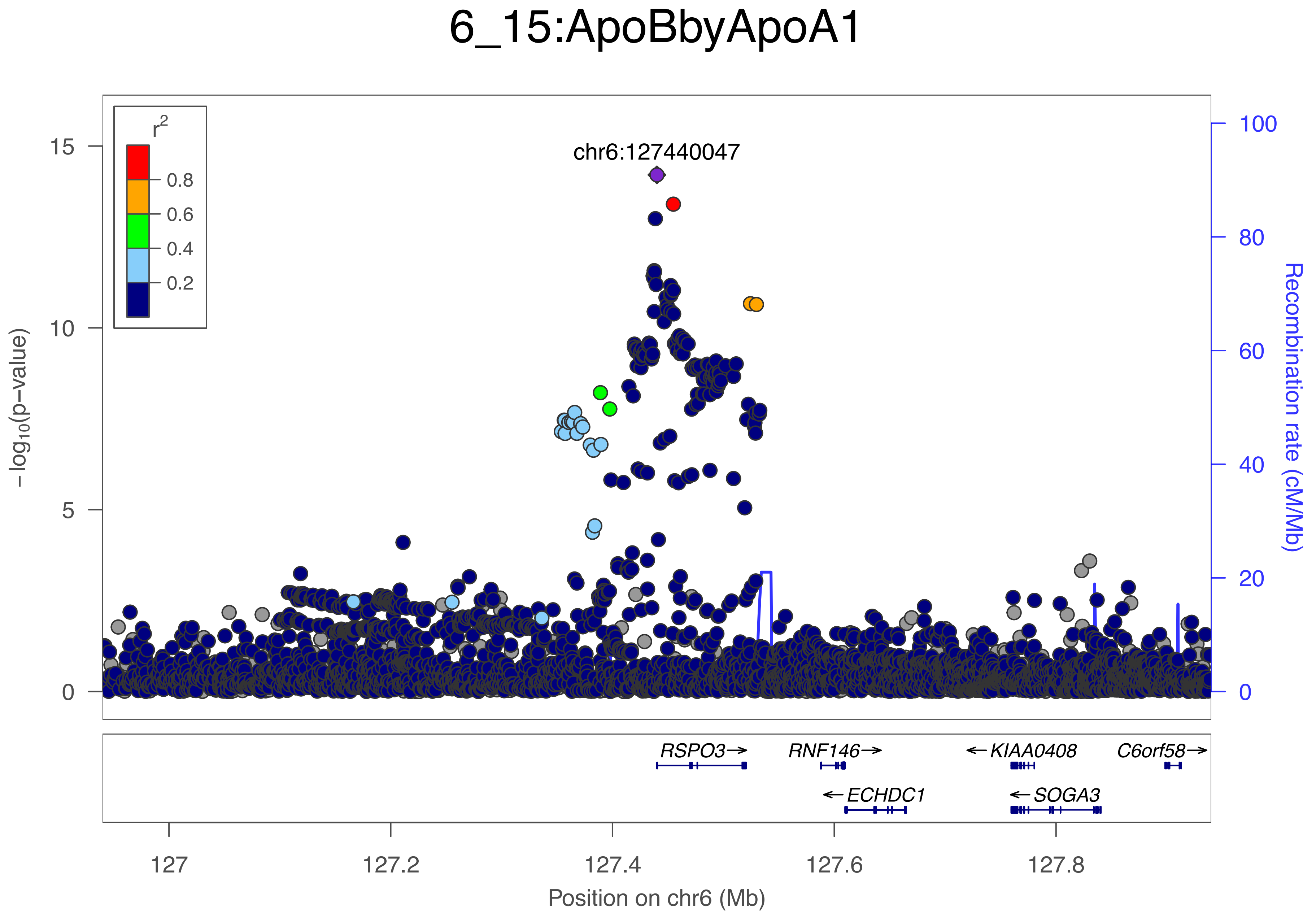

6_15:ApoBbyApoA1
chr6:127440047
r2
0.8
0.6
0.4
0.2
15
10
5
0
100
80
60
40
20
0
−log10(p−value)
Recombination rate (cM/Mb)
RSPO3→
RNF146→
←KIAA0408
C6orf58→
←ECHDC1
←SOGA3
127
127.2
127.4
127.6
127.8
Position on chr6 (Mb)

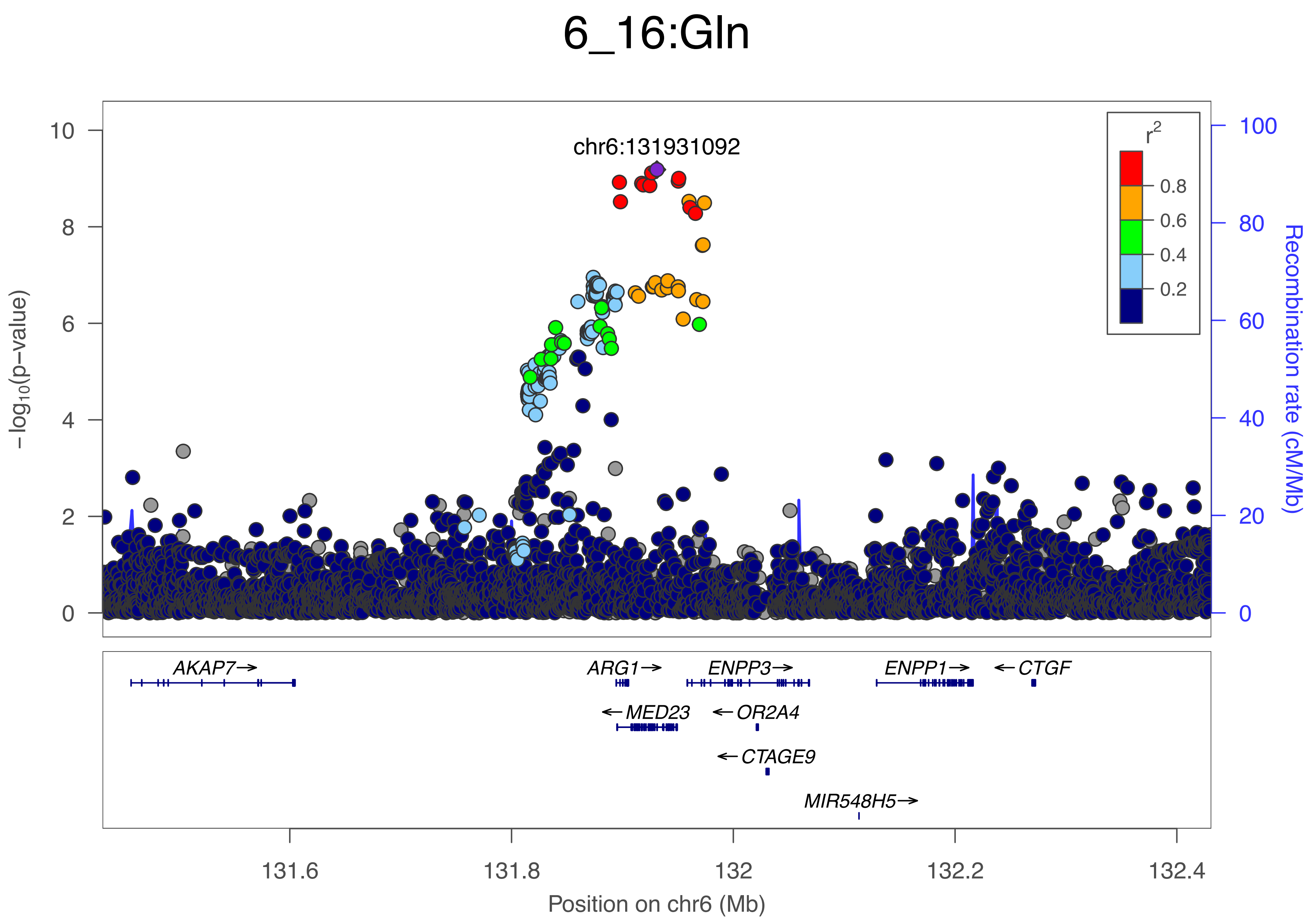

6_16:Gln
chr6:131931092
r2
0.8
0.6
0.4
0.2
−log10(p−value)
Recombination rate (cM/Mb)
10
8
6
4
2
0
100
80
60
40
20
0
AKAP7→
ARG1→
ENPP3→
ENPP1→
←CTGF
←MED23
←OR2A4
←CTAGE9
MIR548H5→
131.6
131.8
132
132.2
132.4
Position on chr6 (Mb)

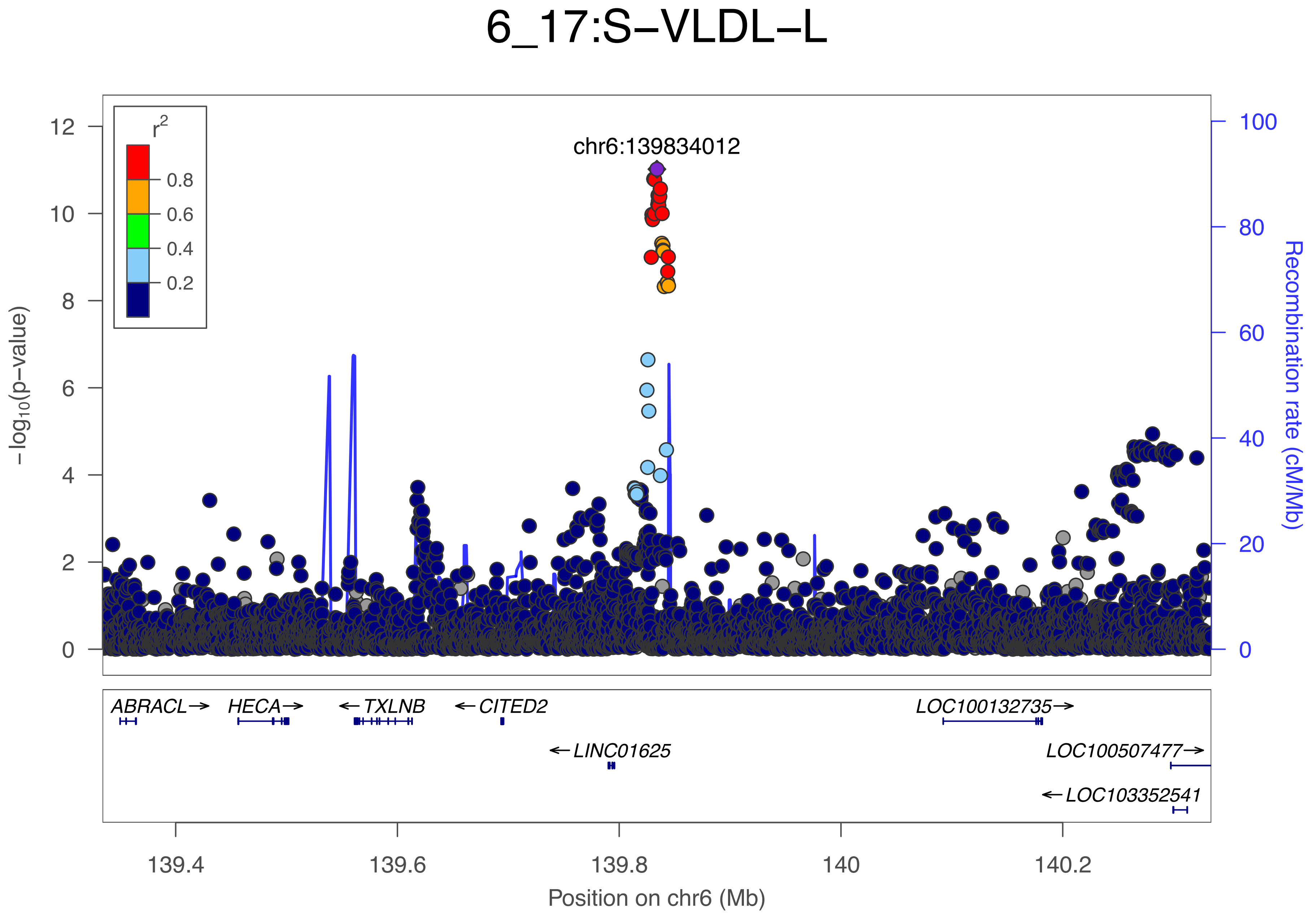
6_17:S−VLDL−L
chr6:139834012
r2
0.8
0.6
0.4
0.2
12
10
8
6
4
2
0
−log10(p−value)
100
80
60
40
20
0
Recombination rate (cM/Mb)
ABRACL→
HECA→
←TXLNB
←CITED2
←LINC01625
LOC100132735→
LOC100507477→
←LOC103352541
139.4
139.6
139.8
140
140.2
Position on chr6 (Mb)

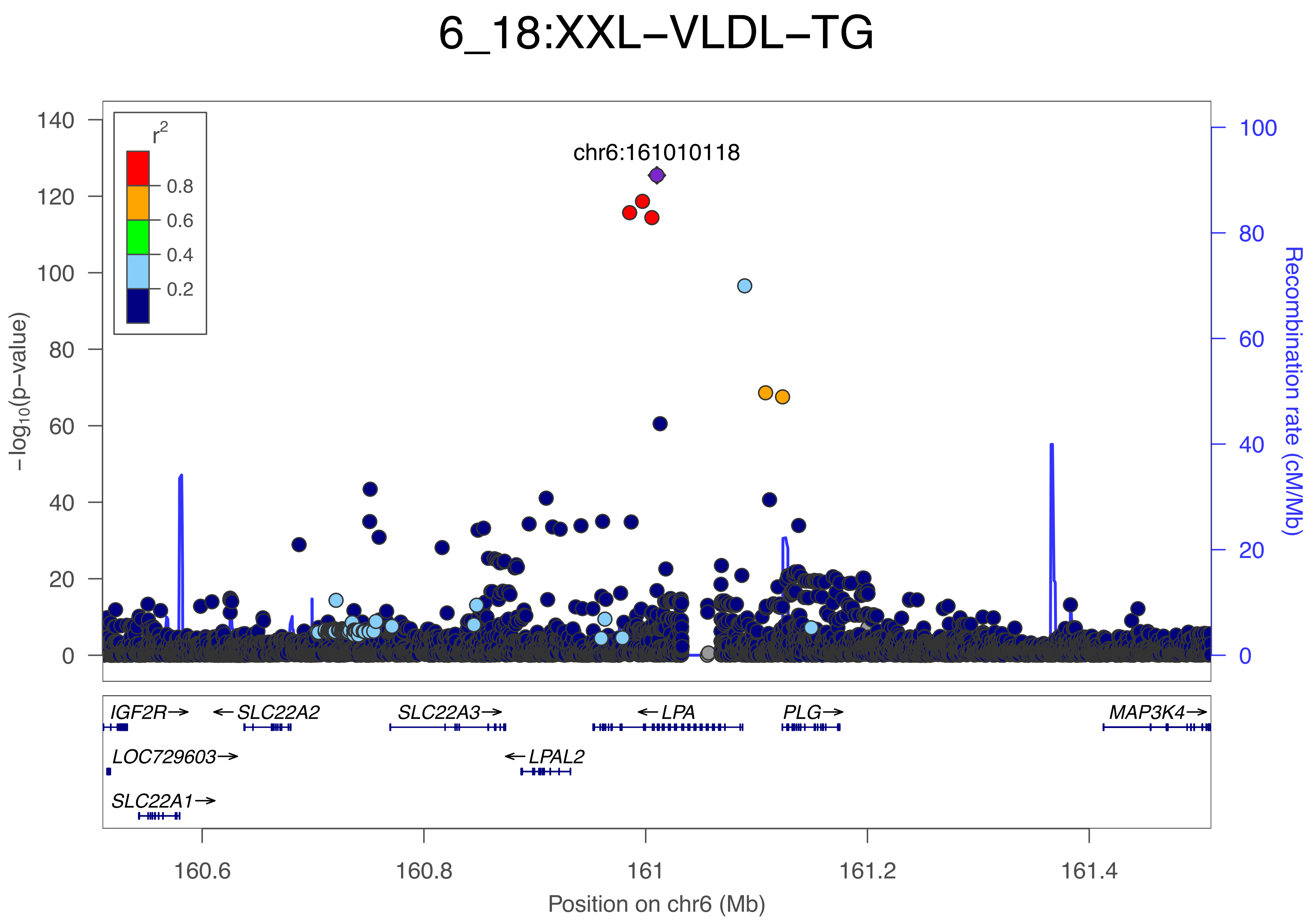
6_18:XXL−VLDL−TG
chr6:161010118
r2
0.8
0.6
0.4
0.2
−log10(p−value)
Recombination rate (cM/Mb)
140
120
100
80
60
40
20
0
100
80
60
40
20
0
IGF2R→
←SLC22A2
SLC22A3→
←LPA
PLG→
MAP3K4→
LOC729603→
←LPAL2
SLC22A1→
160.6
160.8
161
161.2
161.4
Position on chr6 (Mb)

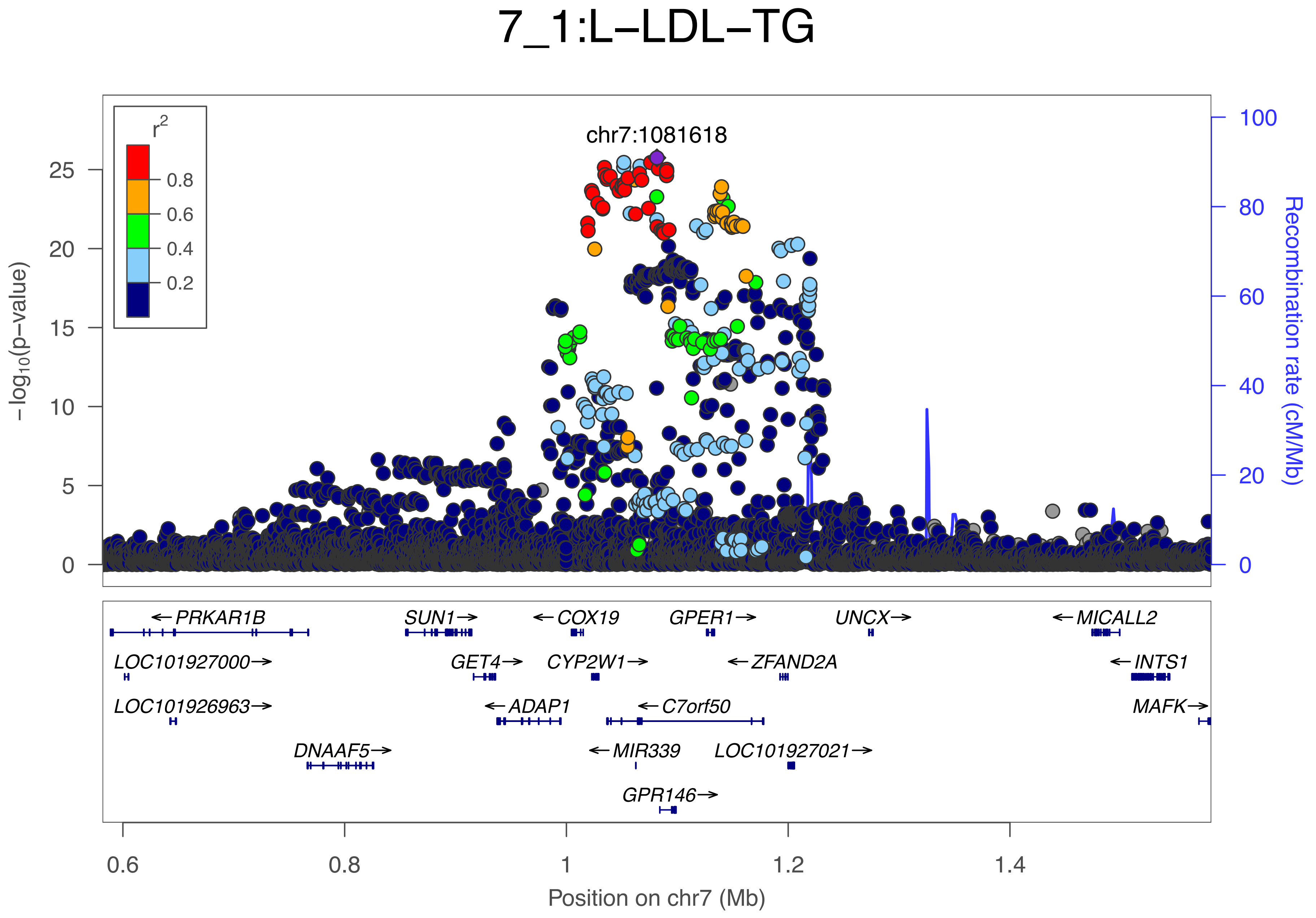

7_1:L−LDL−TG
chr7:1081618
r2
0.8
0.6
0.4
0.2
−log10(p−value)
Recombination rate (cM/Mb)
PRKAR1B
SUN1
COX19
GPER1
UNCX
MICALL2
LOC101927000
GET4
CYP2W1
ZFAND2A
INTS1
LOC101926963
ADAP1
C7orf50
MAFK
DNAAF5
MIR339
LOC101927021
GPR146
Position on chr7 (Mb)

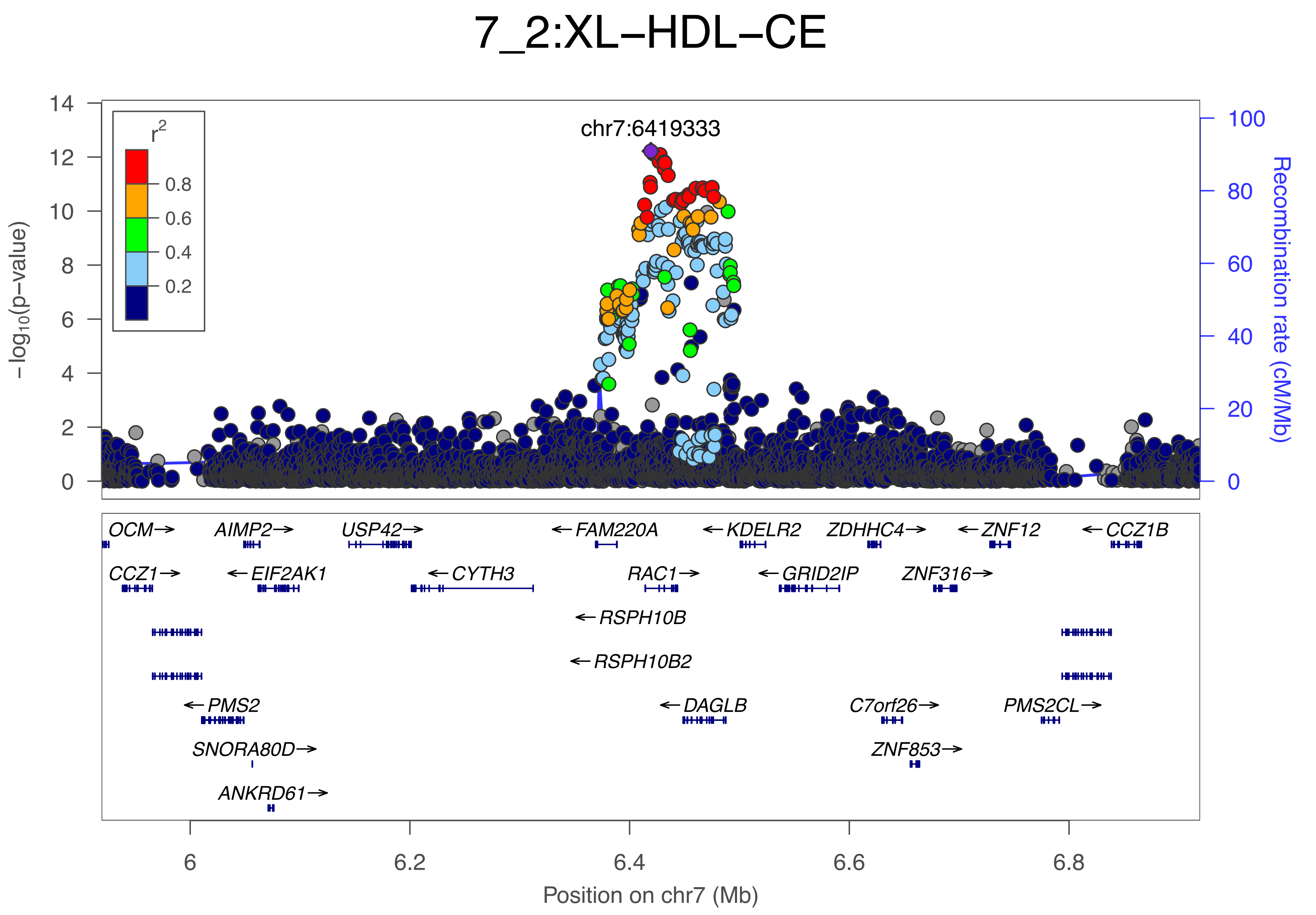
7_2:XL−HDL−CE
chr7:6419333
r2
0.8
0.6
0.4
0.2
−log10(p−value)
Recombination rate (cM/Mb)
OCM→
AIMP2→
USP42→
←FAM220A
←KDELR2
ZDHHC4→
←ZNF12
←CCZ1B
CCZ1→
←EIF2AK1
←CYTH3
RAC1→
←GRID2IP
ZNF316→
←RSPH10B
←RSPH10B2
←PMS2
←DAGLB
C7orf26→
PMS2CL→
SNORA80D→
ZNF853→
ANKRD61→
6
6.2
6.4
6.6
6.8
Position on chr7 (Mb)

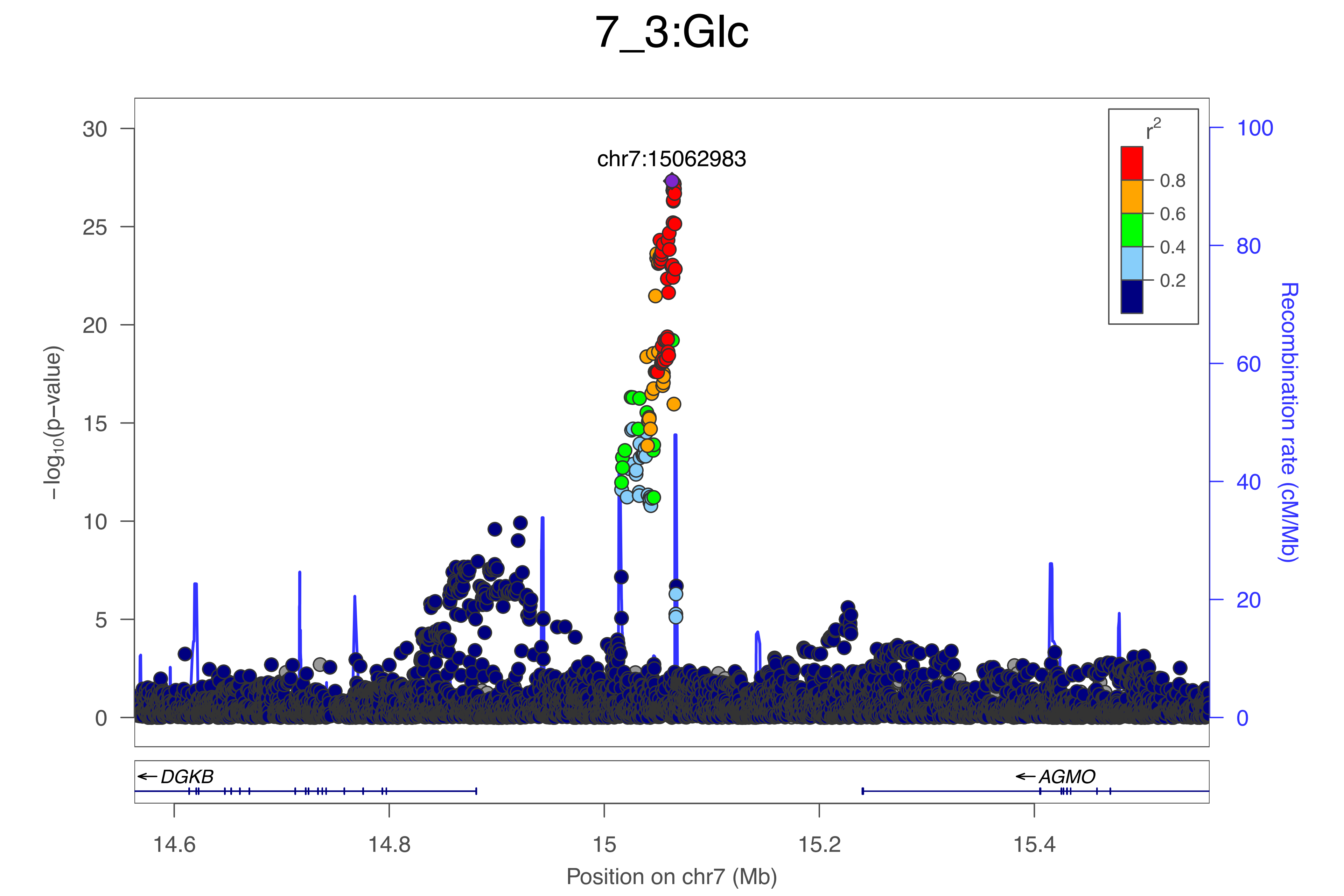

7_3:Glc
chr7:15062983
r2
0.8
0.6
0.4
0.2
−log10(p−value)
Recombination rate (cM/Mb)
30
25
20
15
10
5
0
100
80
60
40
20
0
DGKB
AGMO
14.6
14.8
15
15.2
15.4
Position on chr7 (Mb)

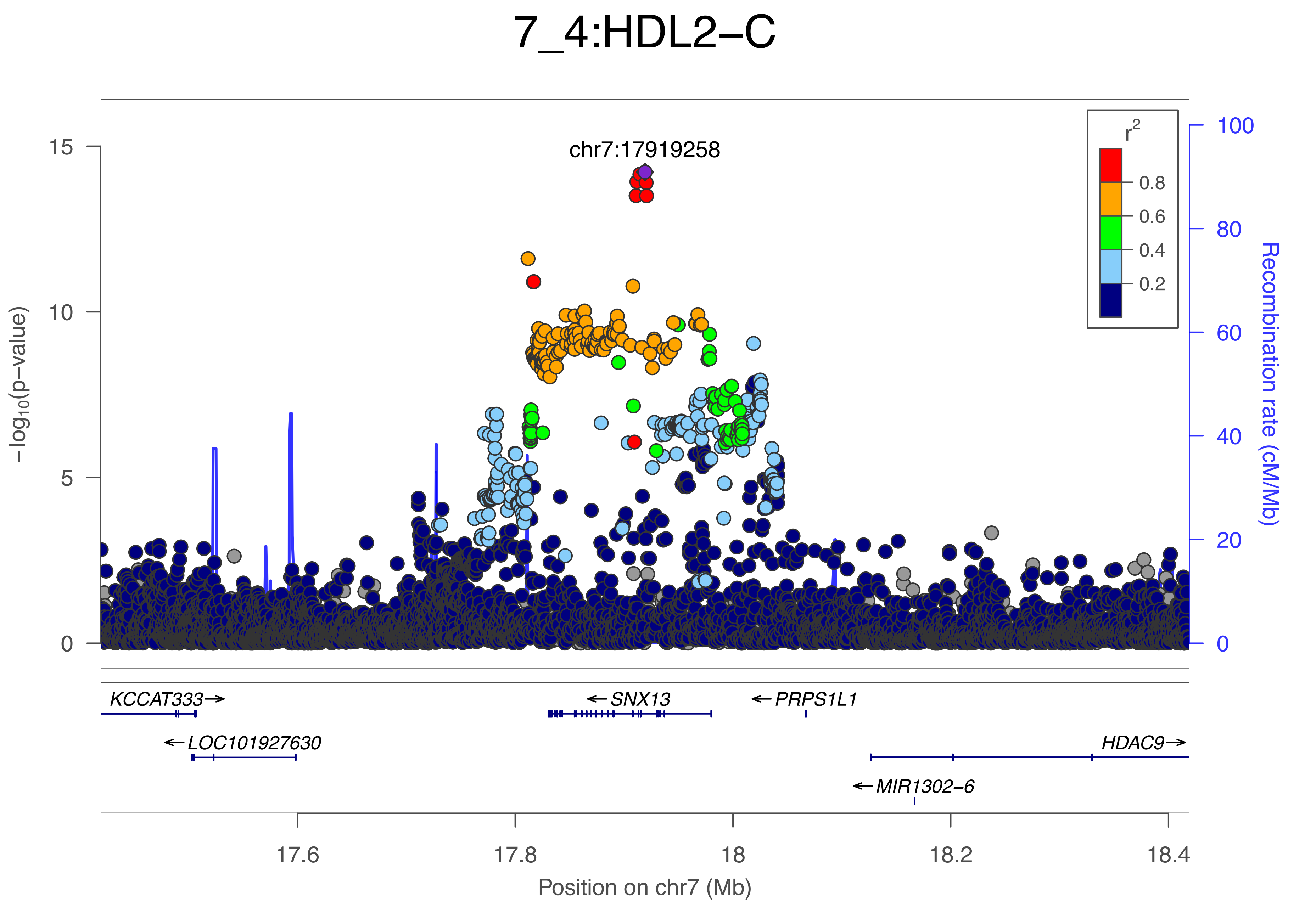

7_4:HDL2−C
chr7:17919258
r2
0.8
0.6
0.4
0.2
−log10(p−value)
Recombination rate (cM/Mb)
15
10
5
0
100
80
60
40
20
0
KCCAT333→
←SNX13
←PRPS1L1
←LOC101927630
HDAC9→
←MIR1302−6
17.6
17.8
18
18.2
18.4
Position on chr7 (Mb)

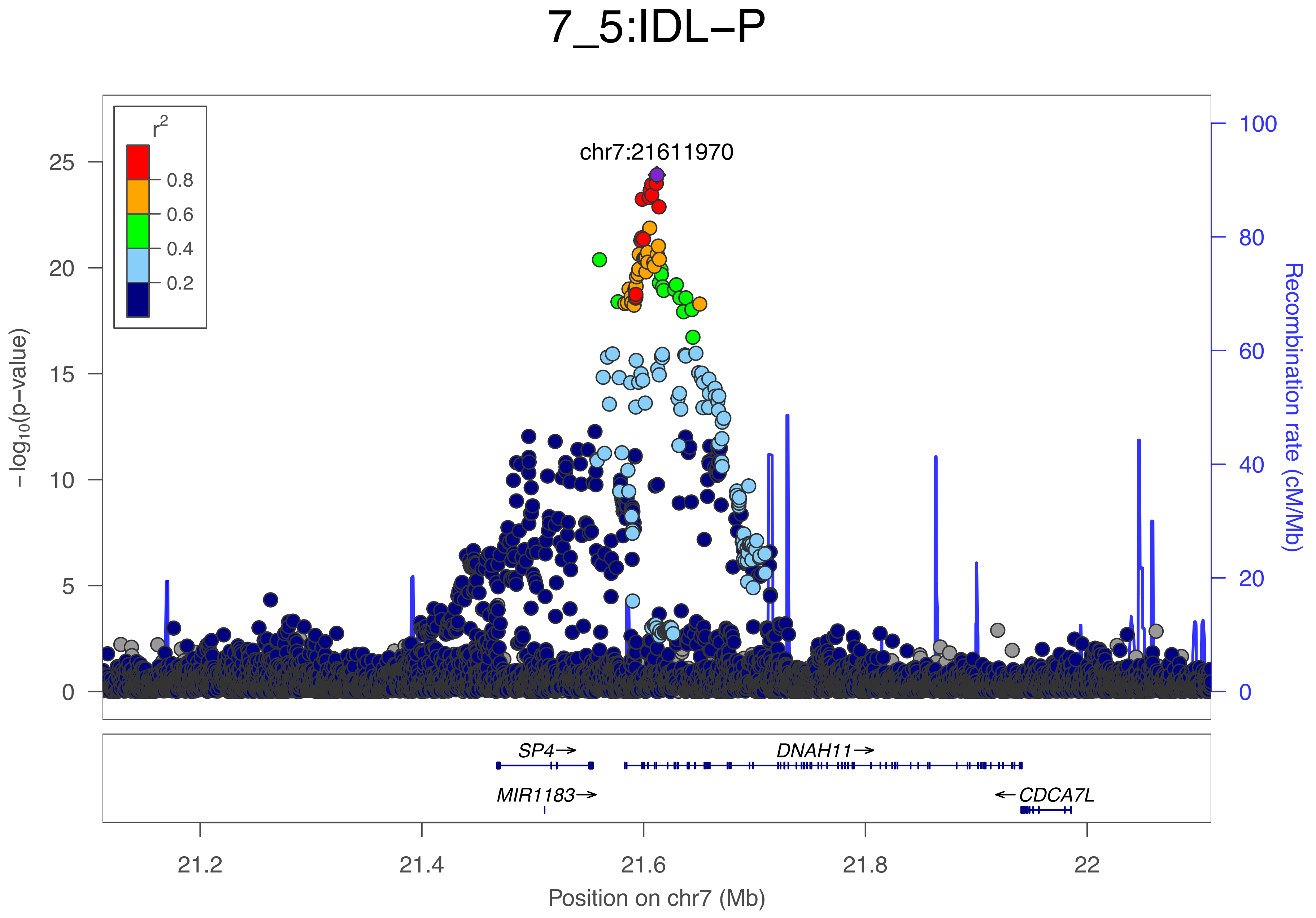

7_5:IDL−P
chr7:21611970
r2
0.8
0.6
0.4
0.2
25
20
15
10
5
0
−log10(p−value)
100
80
60
40
20
0
Recombination rate (cM/Mb)
SP4→
DNAH11→
MIR1183→
←CDCA7L
21.2
21.4
21.6
21.8
22
Position on chr7 (Mb)

### 7_6:S−LDL−C_percent

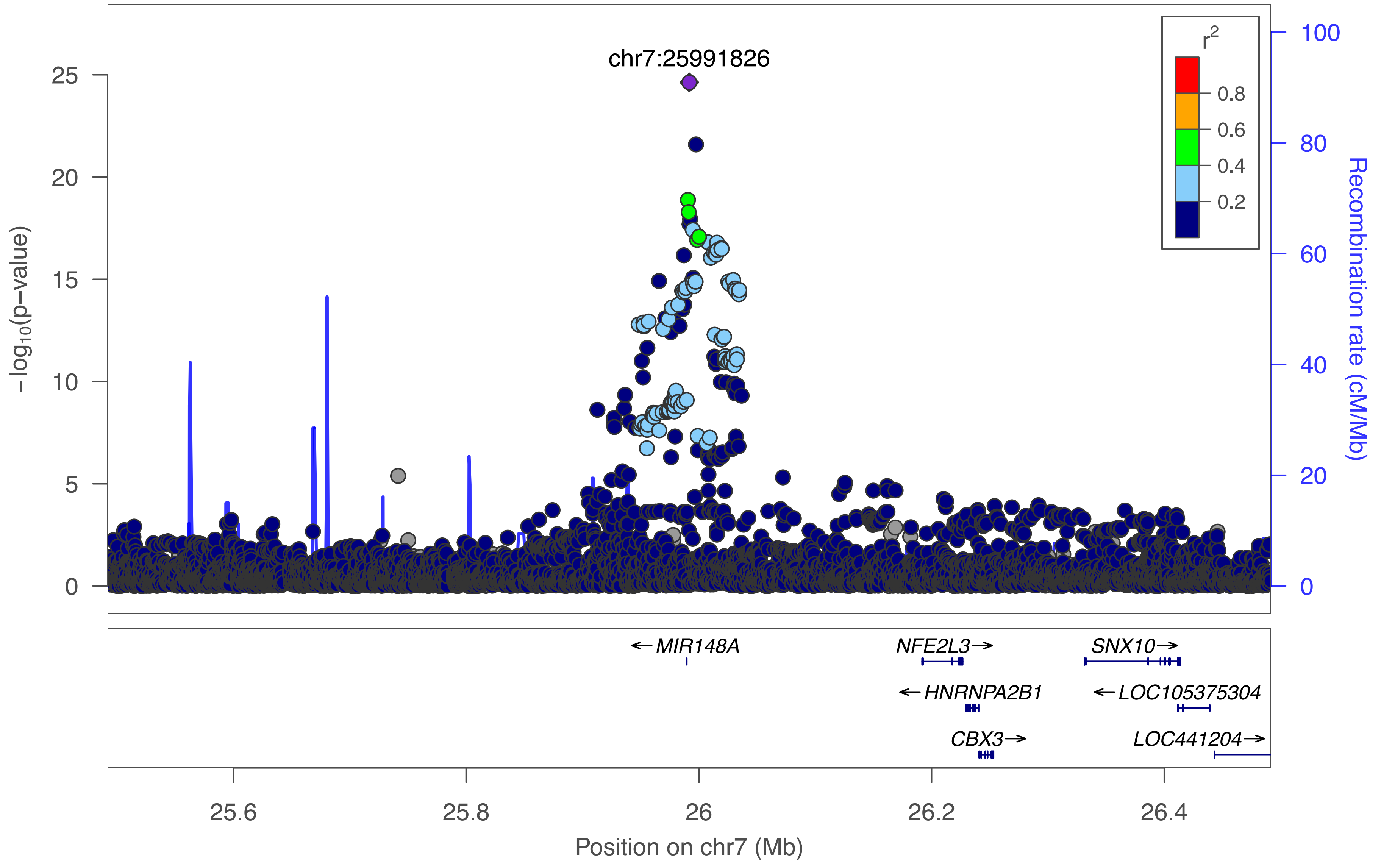

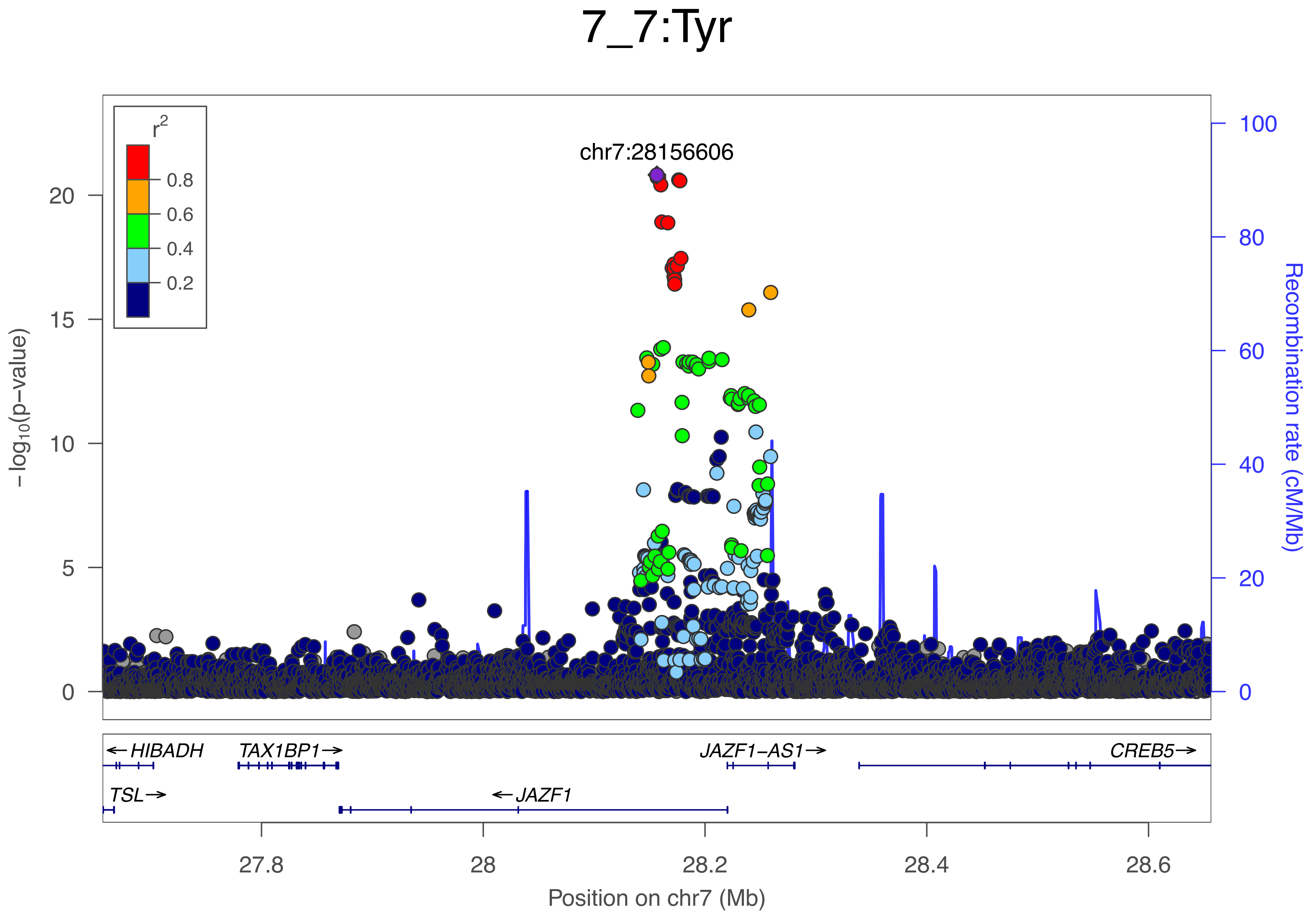

7_7:Tyr
r2
0.8
0.6
0.4
0.2
chr7:28156606
20
15
10
5
0
100
80
60
40
20
0
−log10(p−value)
Recombination rate (cM/Mb)
← HIBADH
TAX1BP1→
JAZF1−AS1→
CREB5→
TSL→
← JAZF1
27.8
28
28.2
28.4
28.6
Position on chr7 (Mb)

### 7_8:Glc

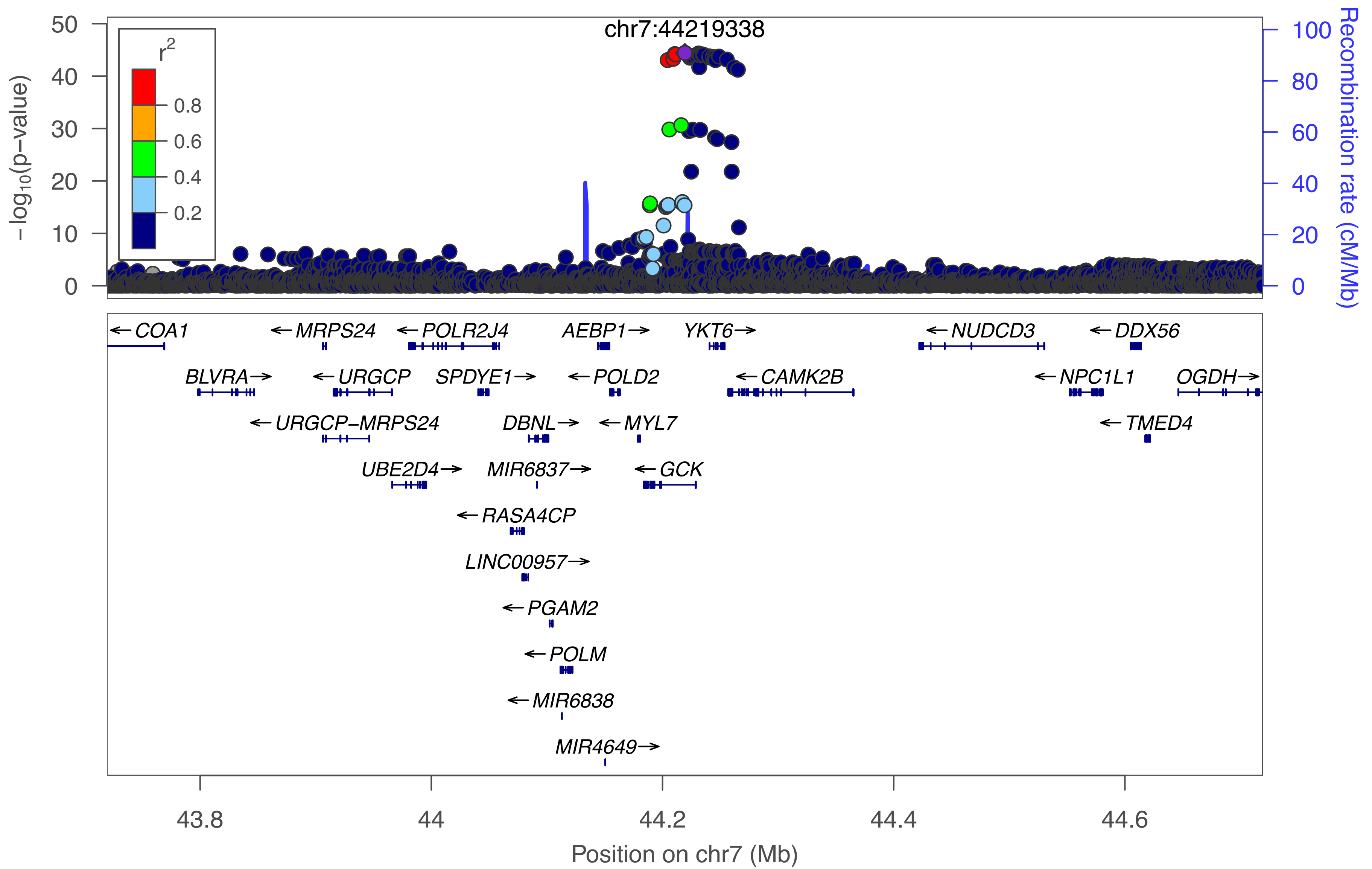

chr7:44219338
r2
0.8
0.6
0.4
0.2
−log10(p−value)
Recombination rate (cM/Mb)
COA1
MRPS24
POLR2J4
AEBP1
YKT6
NUDCD3
DDX56
BLVRA
URGCP
SPDYE1
POLD2
CAMK2B
NPC1L1
OGDH
URGCP−MRPS24
DBNL
MYL7
TMED4
UBE2D4
MIR6837
GCK
RASA4CP
LINC00957
PGAM2
POLM
MIR6838
MIR4649
Position on chr7 (Mb)

### 7_9:Crea

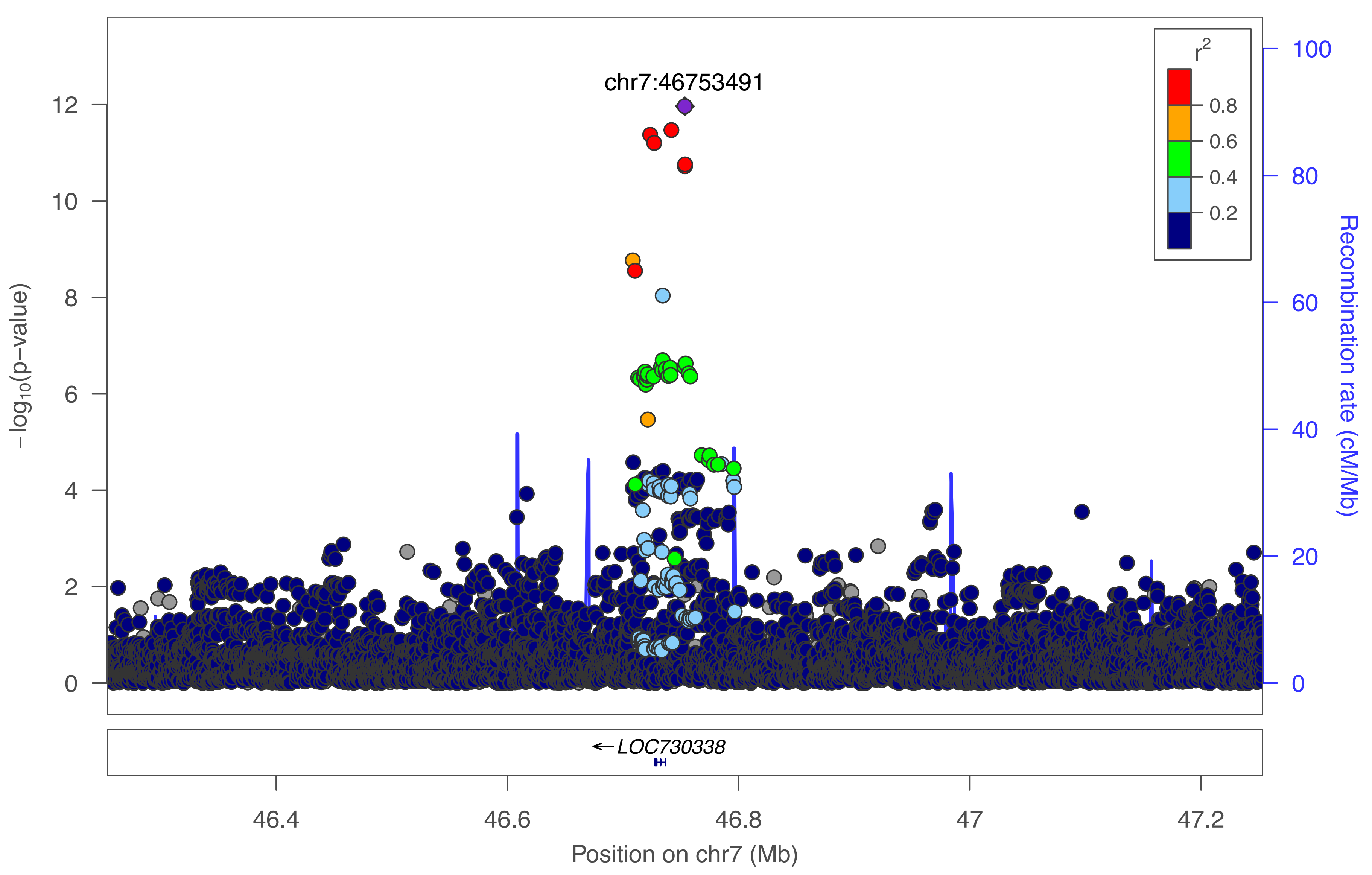

chr7:46753491
r²
0.8
0.6
0.4
0.2
−log10(p−value)
12
10
8
6
4
2
0
Recombination rate (cM/Mb)
100
80
60
40
20
0
← LOC730338
46.4
46.6
46.8
47
47.2
Position on chr7 (Mb)

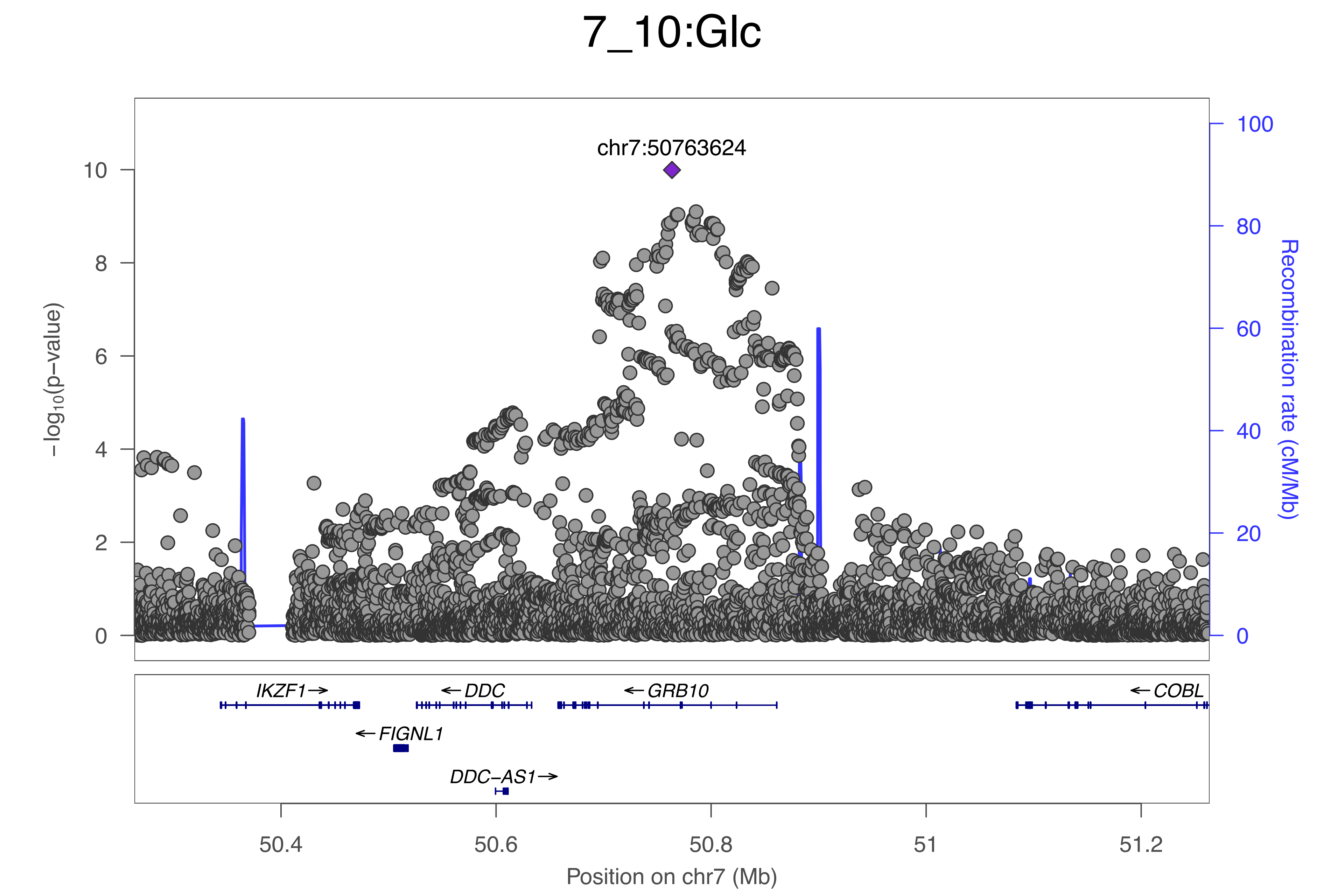
7_10:Glc
chr7:50763624
−log10(p−value)
Recombination rate (cM/Mb)
0
2
4
6
8
10
0
20
40
60
80
100
IKZF1→
←DDC
←GRB10
←COBL
←FIGNL1
DDC−AS1→
50.4
50.6
50.8
51
51.2
Position on chr7 (Mb)

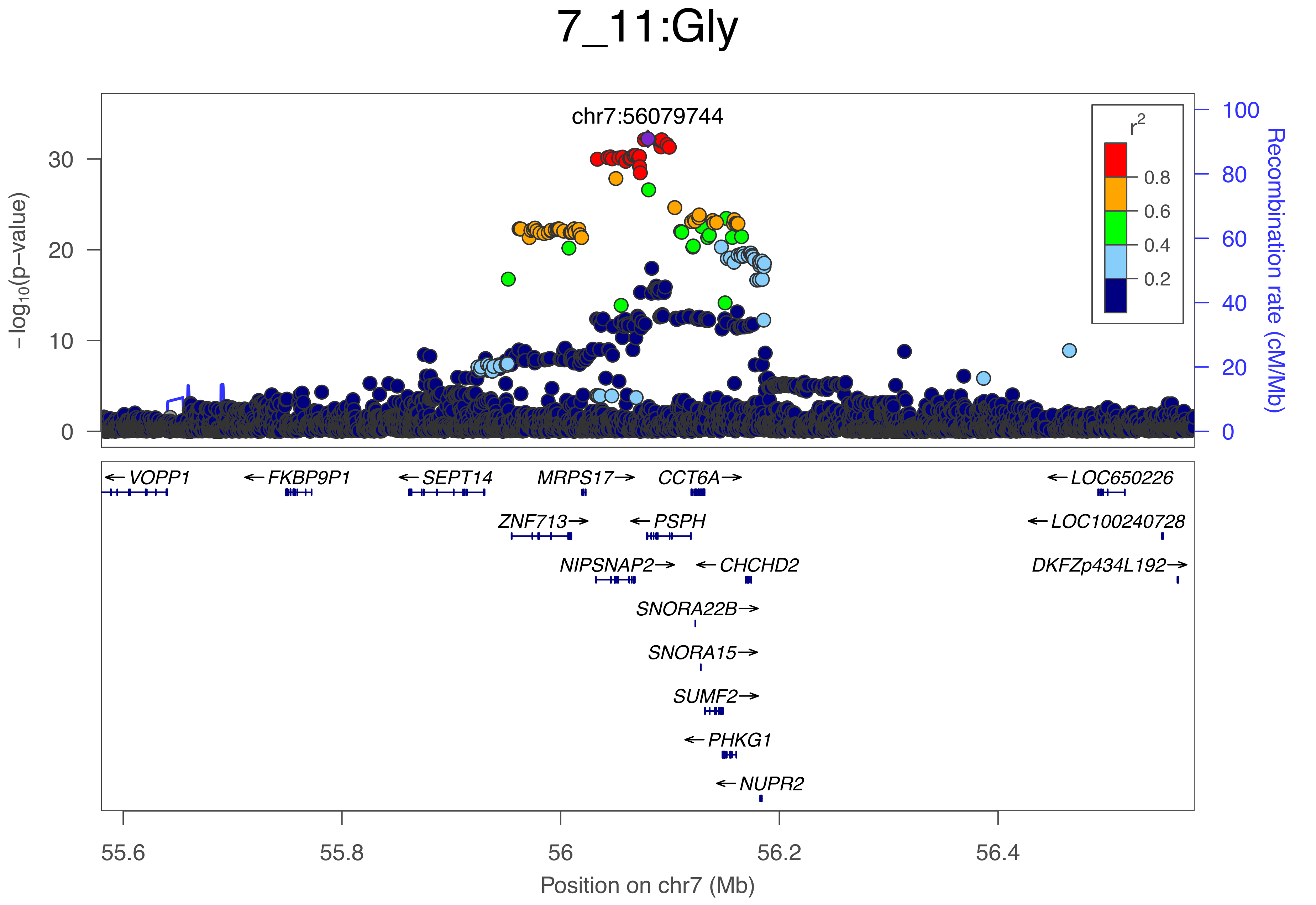

7_11:Gly
chr7:56079744
−log10(p−value)
30
20
10
0
r2
0.8
0.6
0.4
0.2
100
80
60
40
20
0
Recombination rate (cM/Mb)
VOPP1
FKBP9P1
SEPT14
MRPS17
CCT6A
LOC650226
ZNF713
PSPH
LOC100240728
NIPSNAP2
CHCHD2
DKFZp434L192
SNORA22B
SNORA15
SUMF2
PHKG1
NUPR2
55.6
55.8
56
56.2
56.4
Position on chr7 (Mb)

### 7_12:S−VLDL−TG

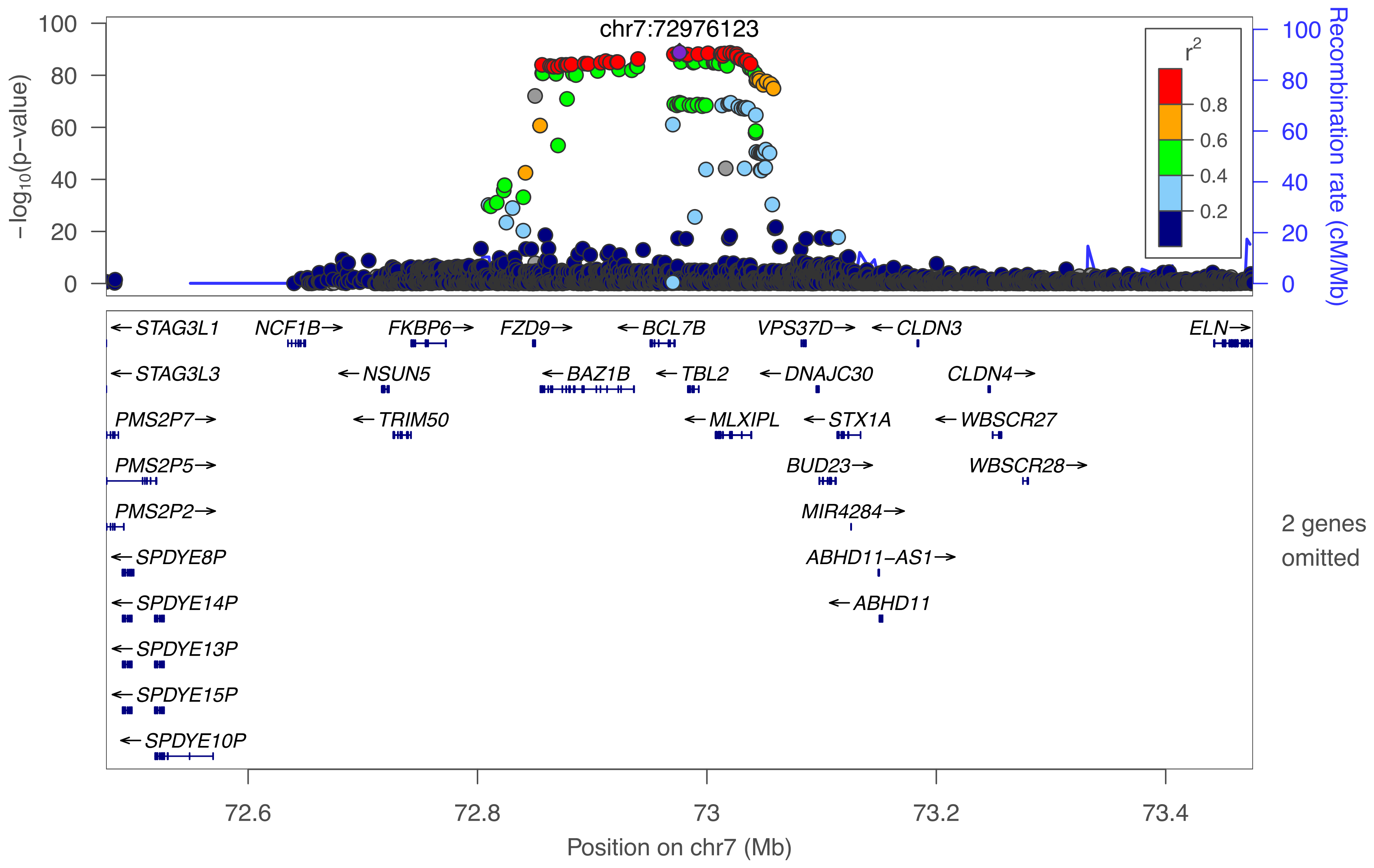
chr7:72976123
−log10(p−value)
Recombination rate (cM/Mb)
r2
0.8
0.6
0.4
0.2
STAG3L1
NCF1B
FKBP6
FZD9
BCL7B
VPS37D
CLDN3
ELN
STAG3L3
NSUN5
BAZ1B
TBL2
DNAJC30
CLDN4
PMS2P7
TRIM50
MLXIPL
STX1A
WBSCR27
PMS2P5
BUD23
WBSCR28
PMS2P2
MIR4284
SPDYE8P
ABHD11−AS1
SPDYE14P
ABHD11
SPDYE13P
SPDYE15P
SPDYE10P
2 genes omitted
72.6
72.8
73
73.2
73.4
Position on chr7 (Mb)

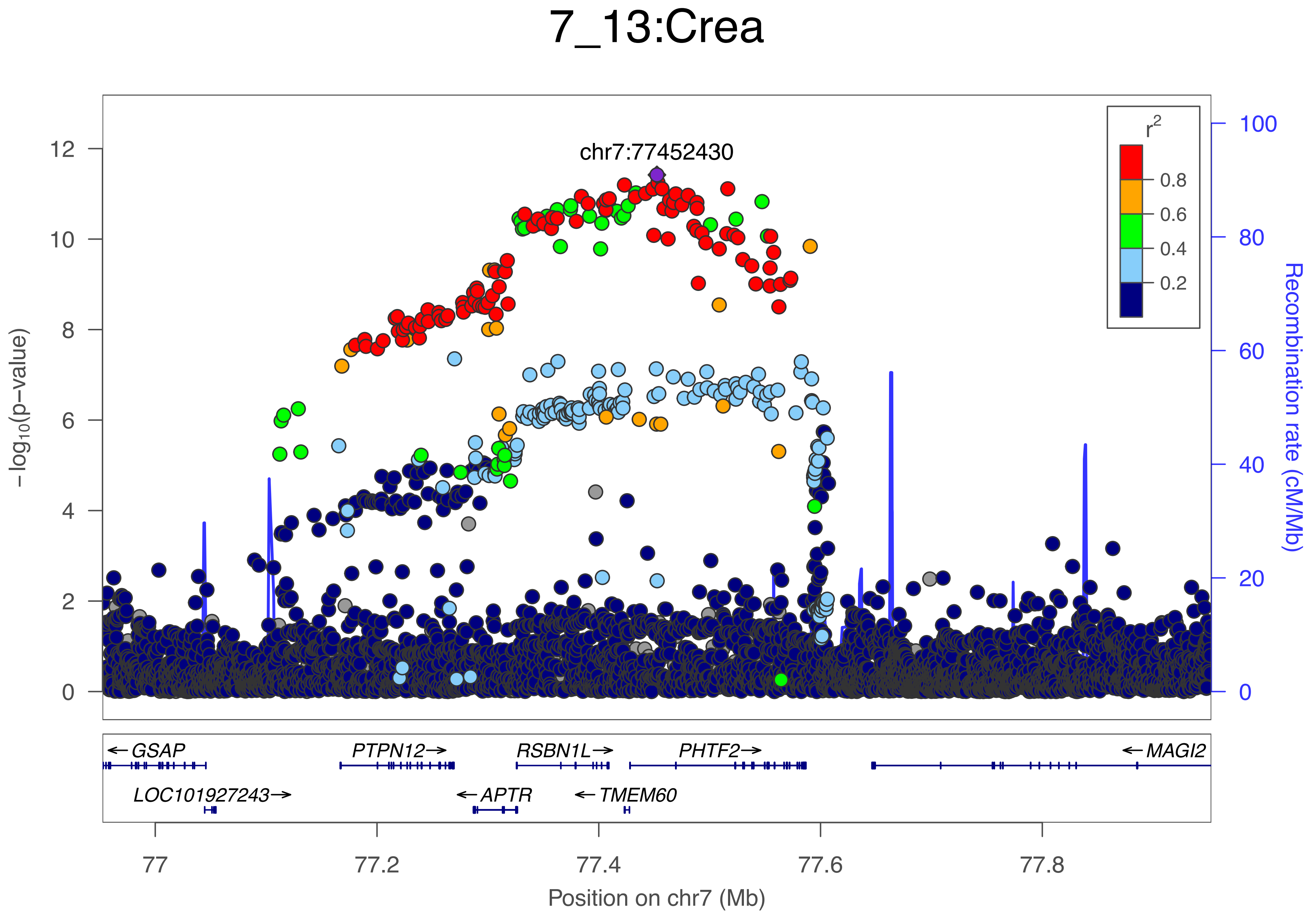
7_13:Crea
chr7:77452430
r2
0.8
0.6
0.4
0.2
-log10(p-value)
0
2
4
6
8
10
12
Recombination rate (cM/Mb)
0
20
40
60
80
100
← GSAP
PTPN12 →
RSBN1L →
PHTF2 →
← MAGI2
LOC101927243 →
← APTR
← TMEM60
77
77.2
77.4
77.6
77.8
Position on chr7 (Mb)

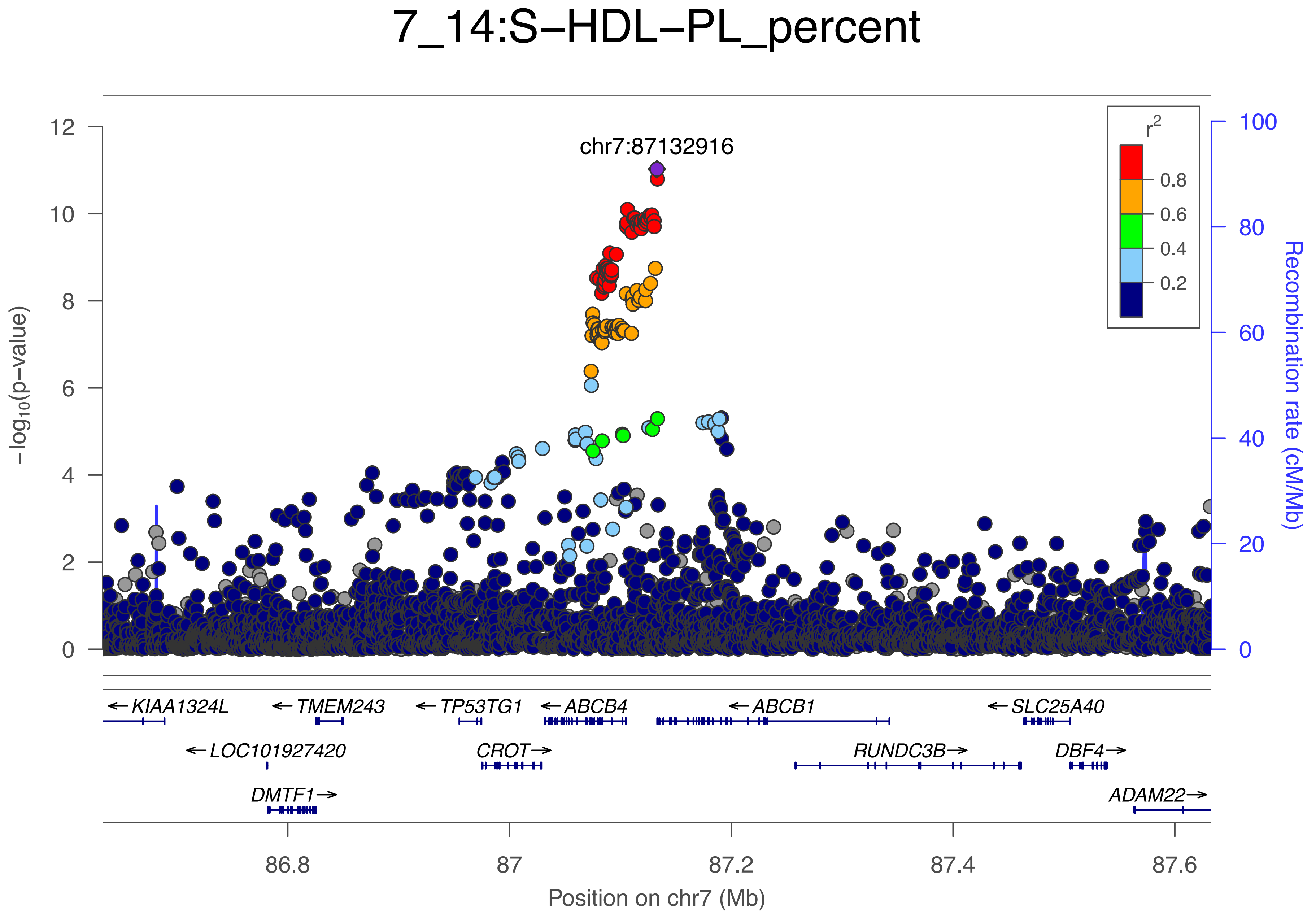

7_14:S−HDL−PL_percent
chr7:87132916
r²
0.8
0.6
0.4
0.2
−log10(p−value)
Recombination rate (cM/Mb)
12
10
8
6
4
2
0
100
80
60
40
20
0
KIAA1324L
TMEM243
TP53TG1
ABCB4
ABCB1
SLC25A40
LOC101927420
CROT
RUNDC3B
DBF4
DMTF1
ADAM22
86.8
87
87.2
87.4
87.6
Position on chr7 (Mb)

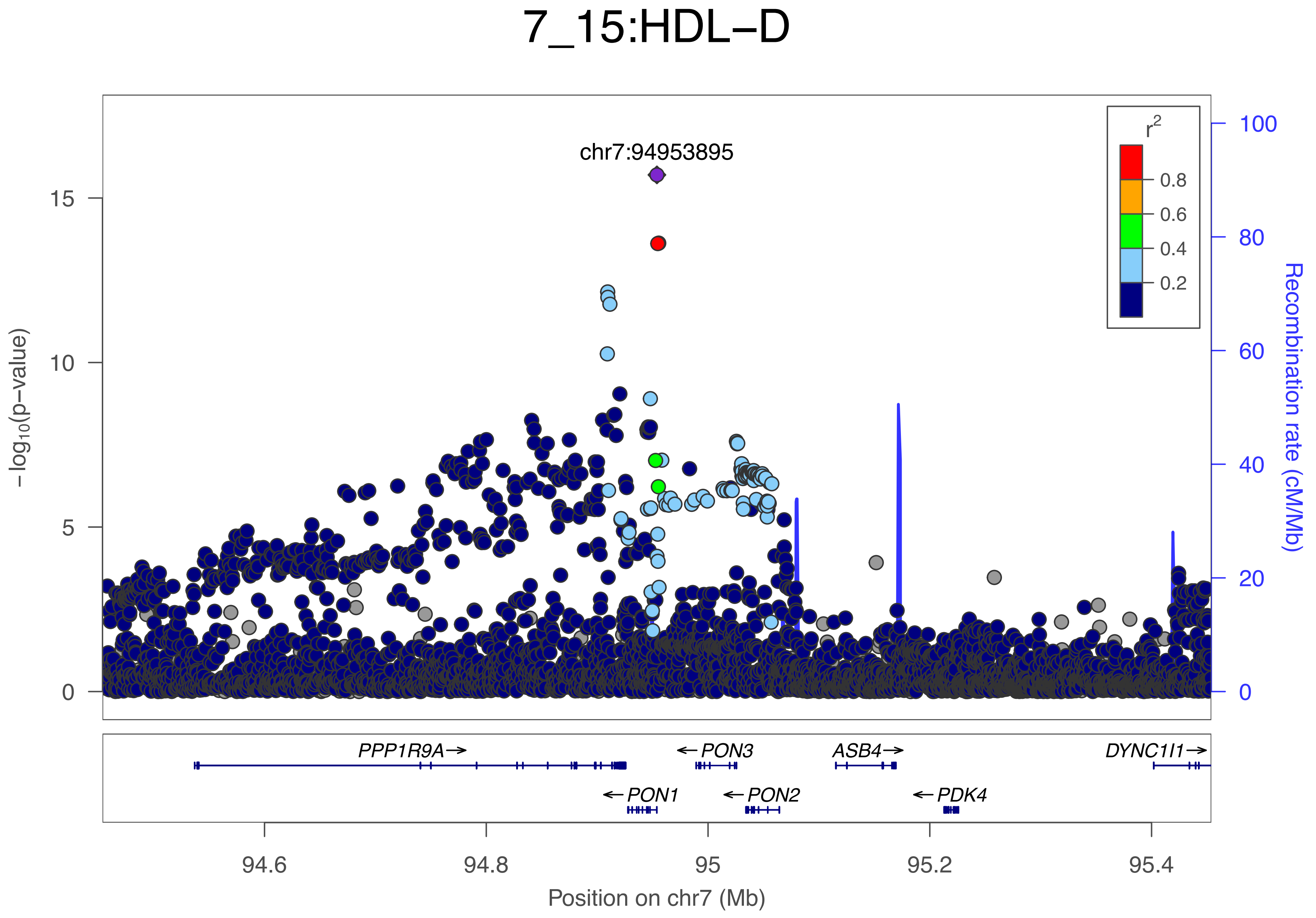
7_15:HDL−D
chr7:94953895
r2
0.8
0.6
0.4
0.2
15
10
5
0
−log10(p−value)
100
80
60
40
20
0
Recombination rate (cM/Mb)
PPP1R9A→
←PON3
ASB4→
DYNC1I1→
←PON1
←PON2
←PDK4
94.6
94.8
95
95.2
95.4
Position on chr7 (Mb)

### 7_16:ApoBbyApoA1

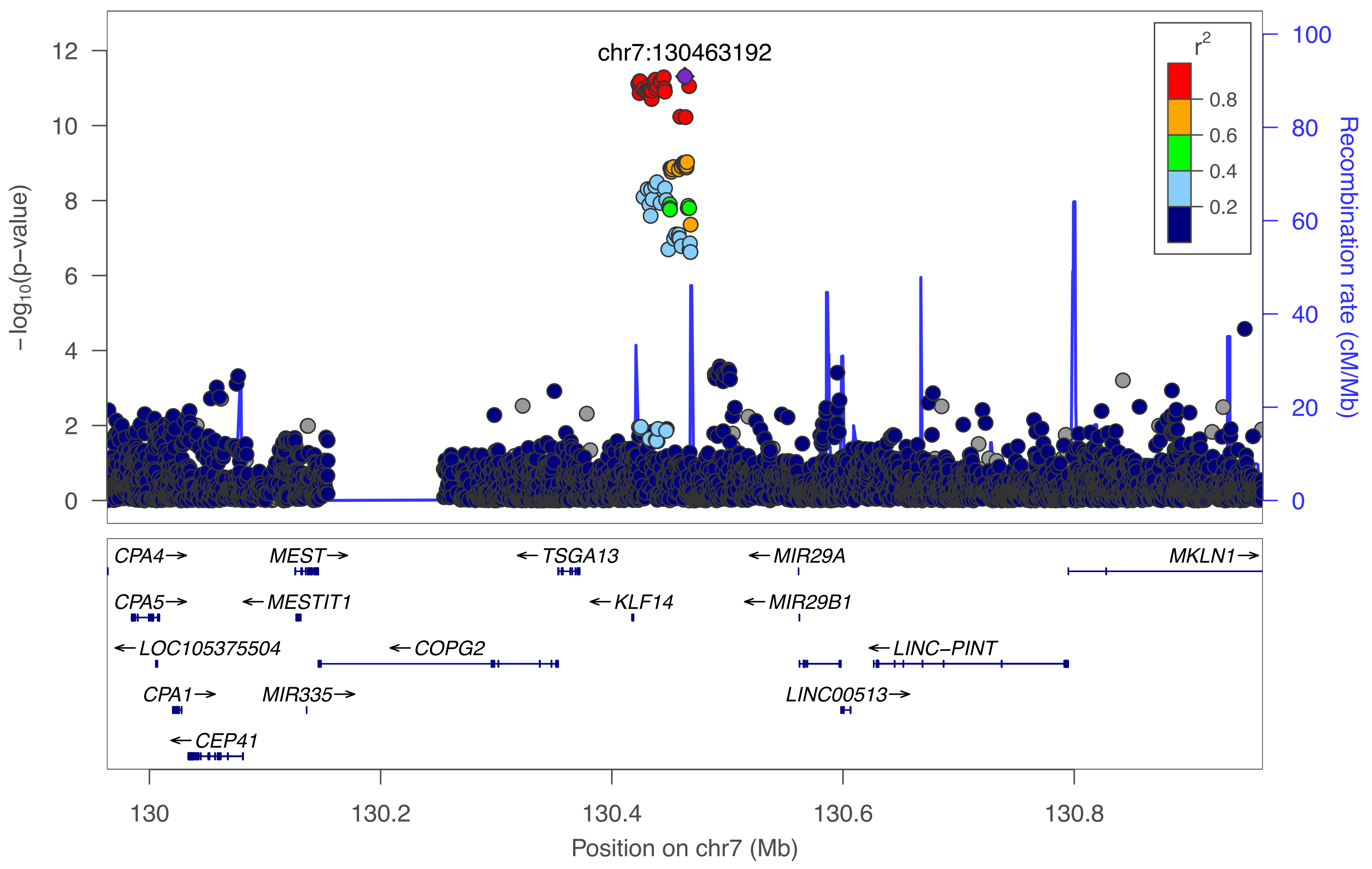

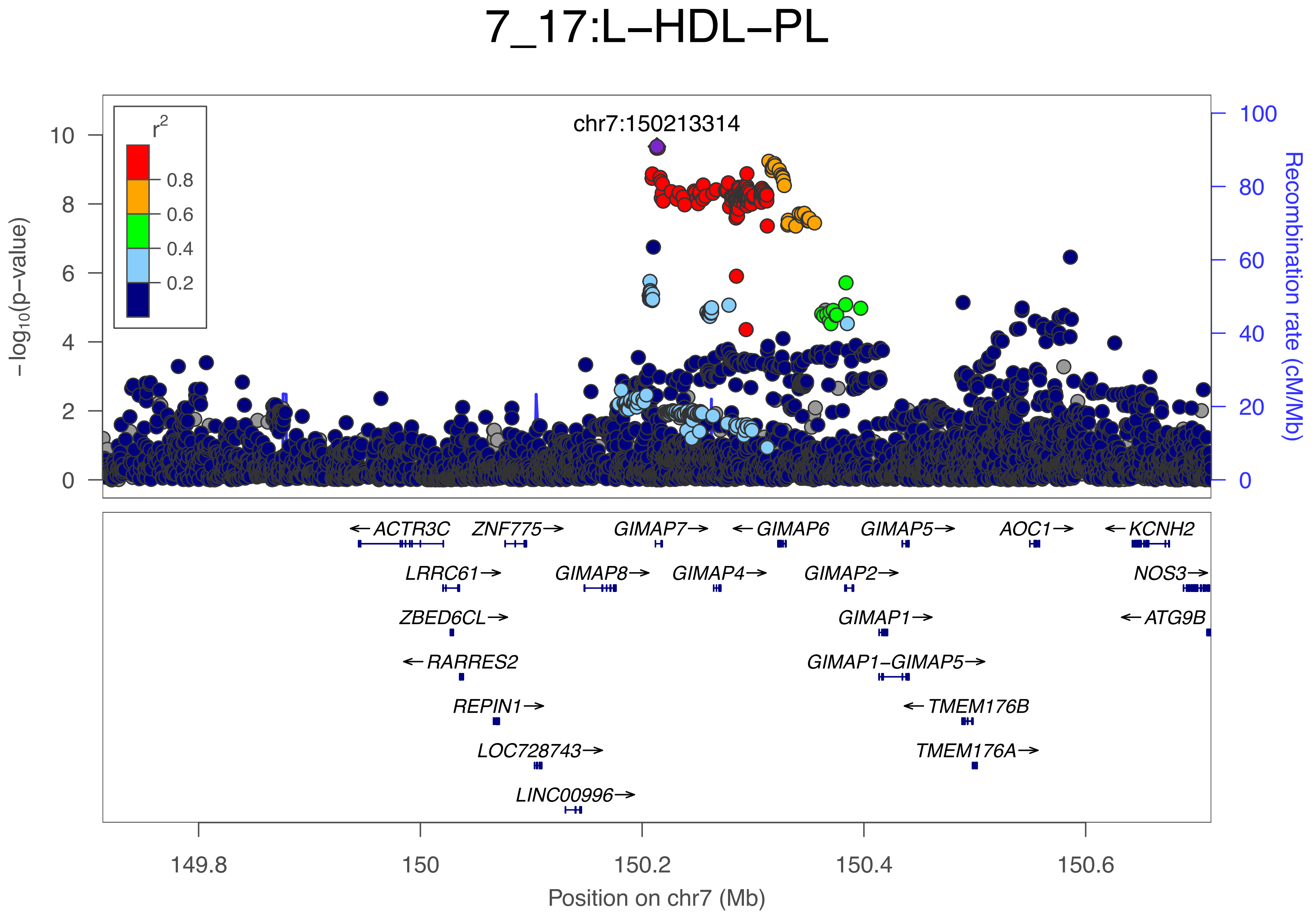
7_17:L–HDL–PL
chr7:150213314
r2
0.8
0.6
0.4
0.2
−log10(p−value)
Recombination rate (cM/Mb)
0
2
4
6
8
10
0
20
40
60
80
100
ACTR3C
ZNF775
GIMAP7
GIMAP6
GIMAP5
AOC1
KCNH2
LRRC61
GIMAP8
GIMAP4
GIMAP2
NOS3
ZBED6CL
GIMAP1
ATG9B
RARRES2
GIMAP1−GIMAP5
REPIN1
TMEM176B
LOC728743
TMEM176A
LINC00996
149.8
150
150.2
150.4
150.6
Position on chr7 (Mb)

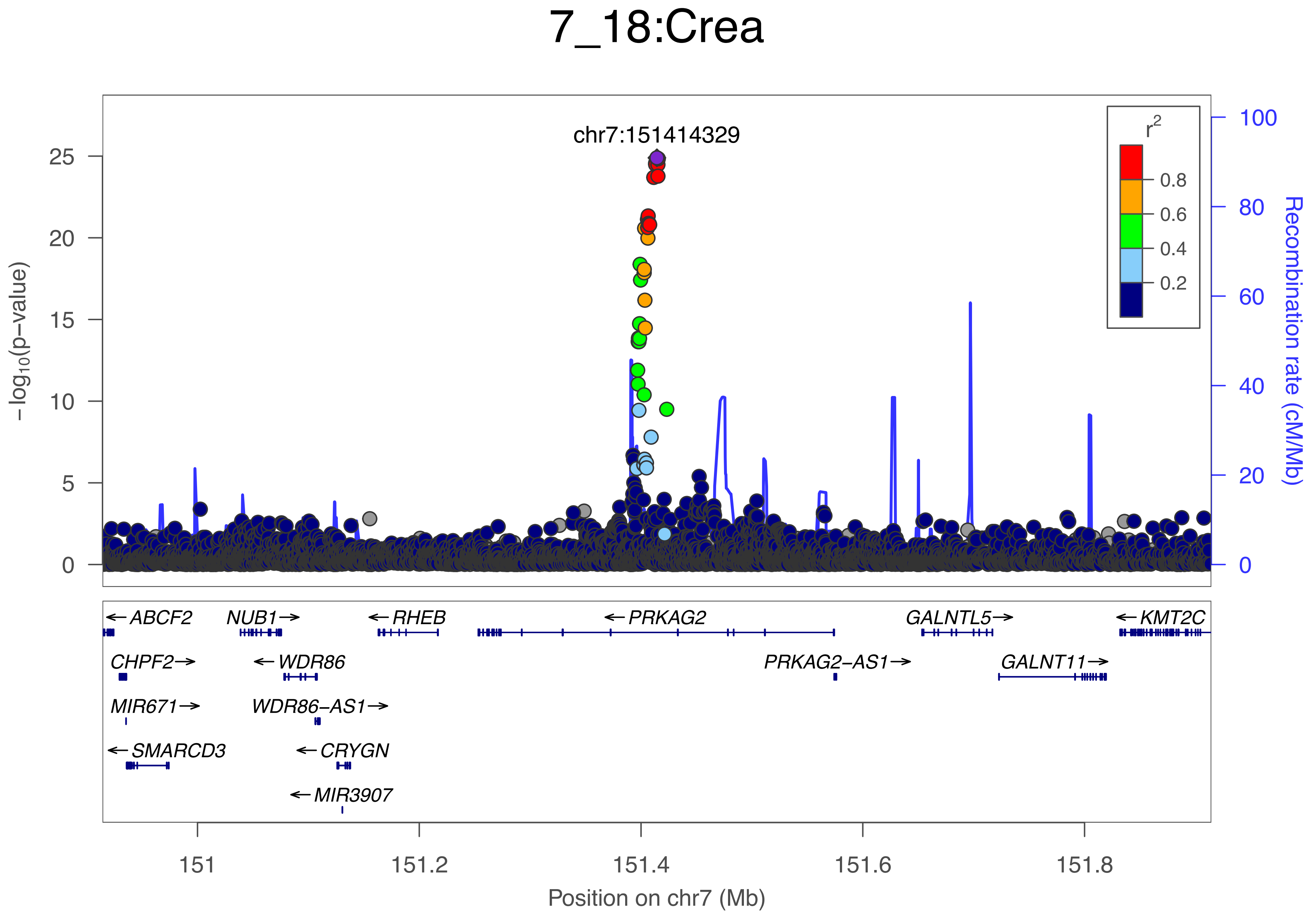
7_18:Crea
chr7:151414329
r2
0.8
0.6
0.4
0.2
−log10(p−value)
0
5
10
15
20
25
Recombination rate (cM/Mb)
0
20
40
60
80
100
←ABCF2
NUB1→
←RHEB
←PRKAG2
GALNTL5→
←KMT2C
CHPF2→
←WDR86
PRKAG2−AS1→
GALNT11→
MIR671→
WDR86−AS1→
←SMARCD3
←CRYGN
←MIR3907
151
151.2
151.4
151.6
151.8
Position on chr7 (Mb)

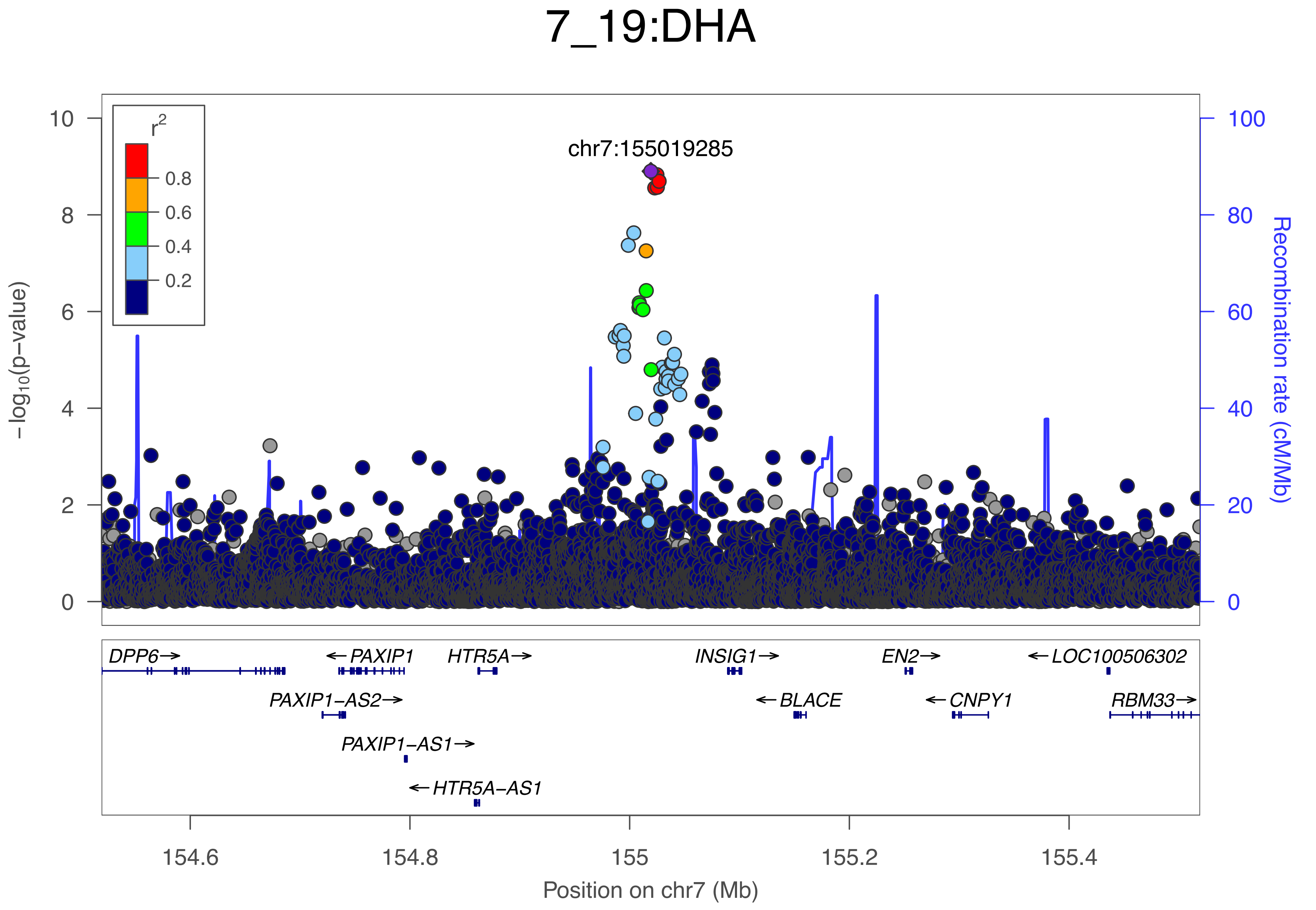

7_19:DHA
chr7:155019285
r2
0.8
0.6
0.4
0.2
−log10(p−value)
Recombination rate (cM/Mb)
0
2
4
6
8
10
0
20
40
60
80
100
DPP6→
←PAXIP1
HTR5A→
INSIG1→
EN2→
←LOC100506302
PAXIP1−AS2→
←BLACE
←CNPY1
RBM33→
PAXIP1−AS1→
←HTR5A−AS1
154.6
154.8
155
155.2
155.4
Position on chr7 (Mb)

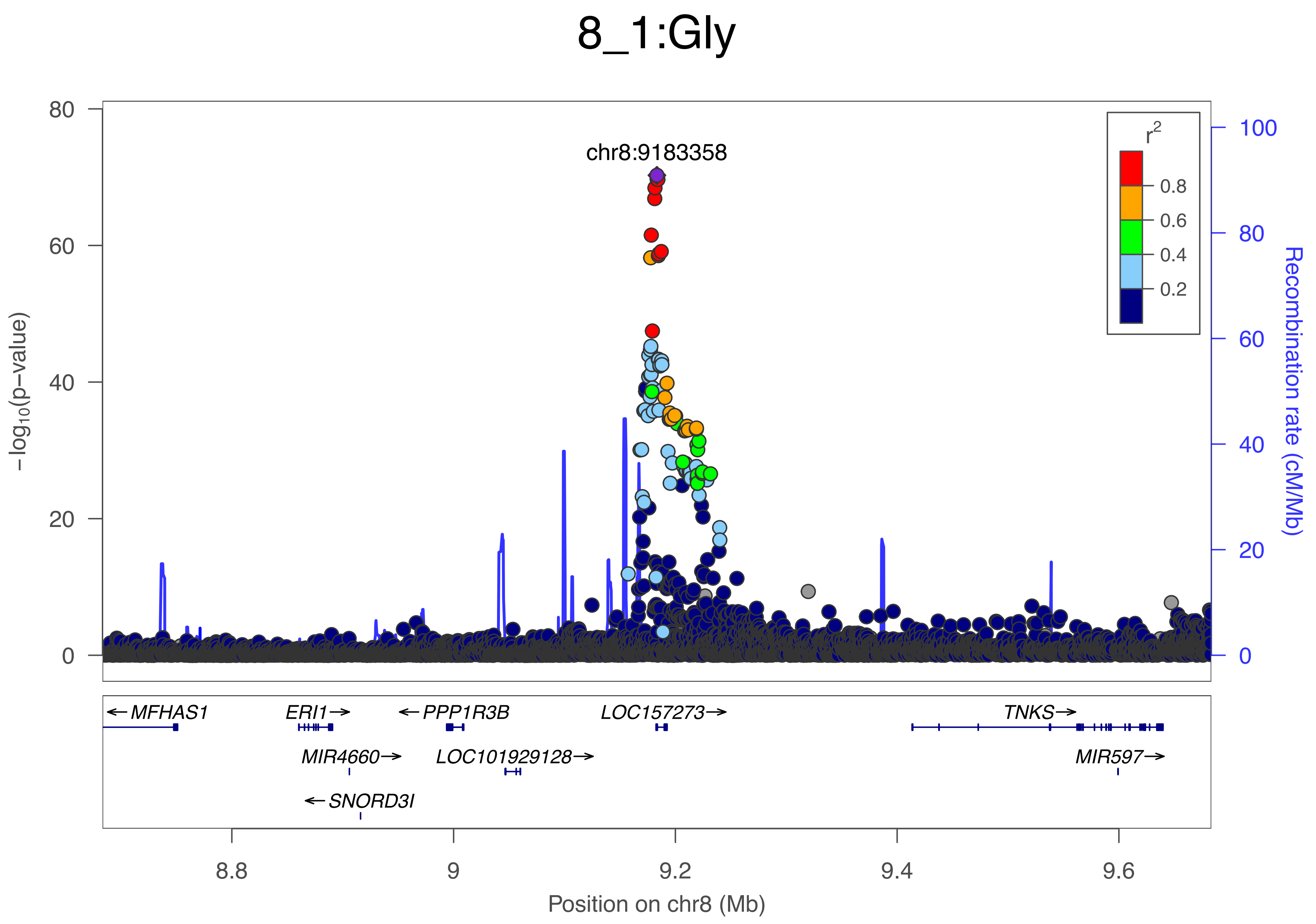

8_1:Gly
chr8:9183358
r²
0.8
0.6
0.4
0.2
−log10(p−value)
Recombination rate (cM/Mb)
80
60
40
20
0
100
80
60
40
20
0
← MFHAS1
ERI1→
← PPP1R3B
LOC157273→
TNKS→
MIR4660→
LOC101929128→
MIR597→
← SNORD3I
8.8
9
9.2
9.4
9.6
Position on chr8 (Mb)

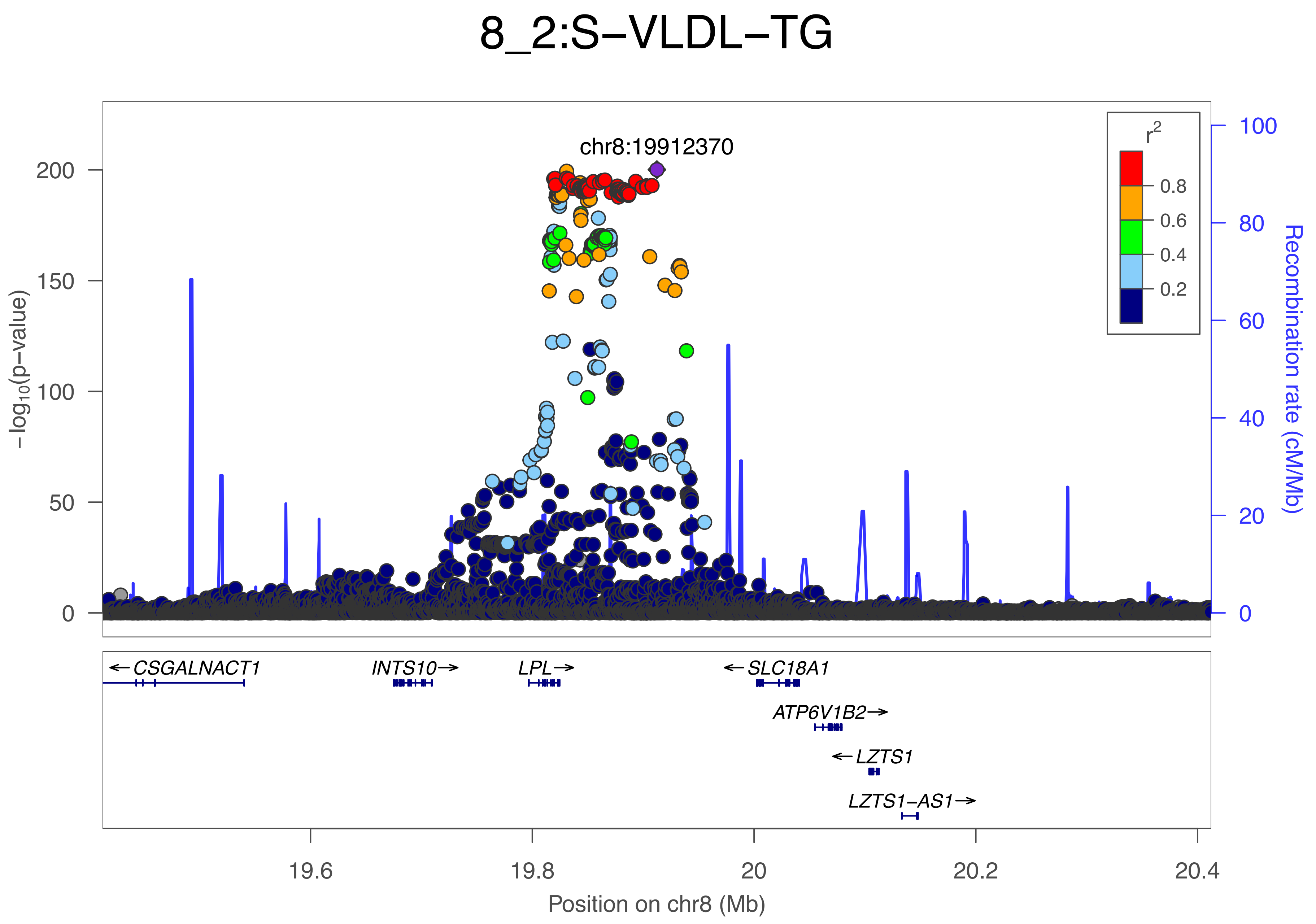

8_2:S−VLDL−TG
chr8:19912370
r²
0.8
0.6
0.4
0.2
−log10(p−value)
200
150
100
50
0
100
80
60
40
20
0
Recombination rate (cM/Mb)
← CSGALNACT1
INTS10 →
LPL →
← SLC18A1
ATP6V1B2 →
← LZTS1
LZTS1−AS1 →
19.6
19.8
20
20.2
20.4
Position on chr8 (Mb)

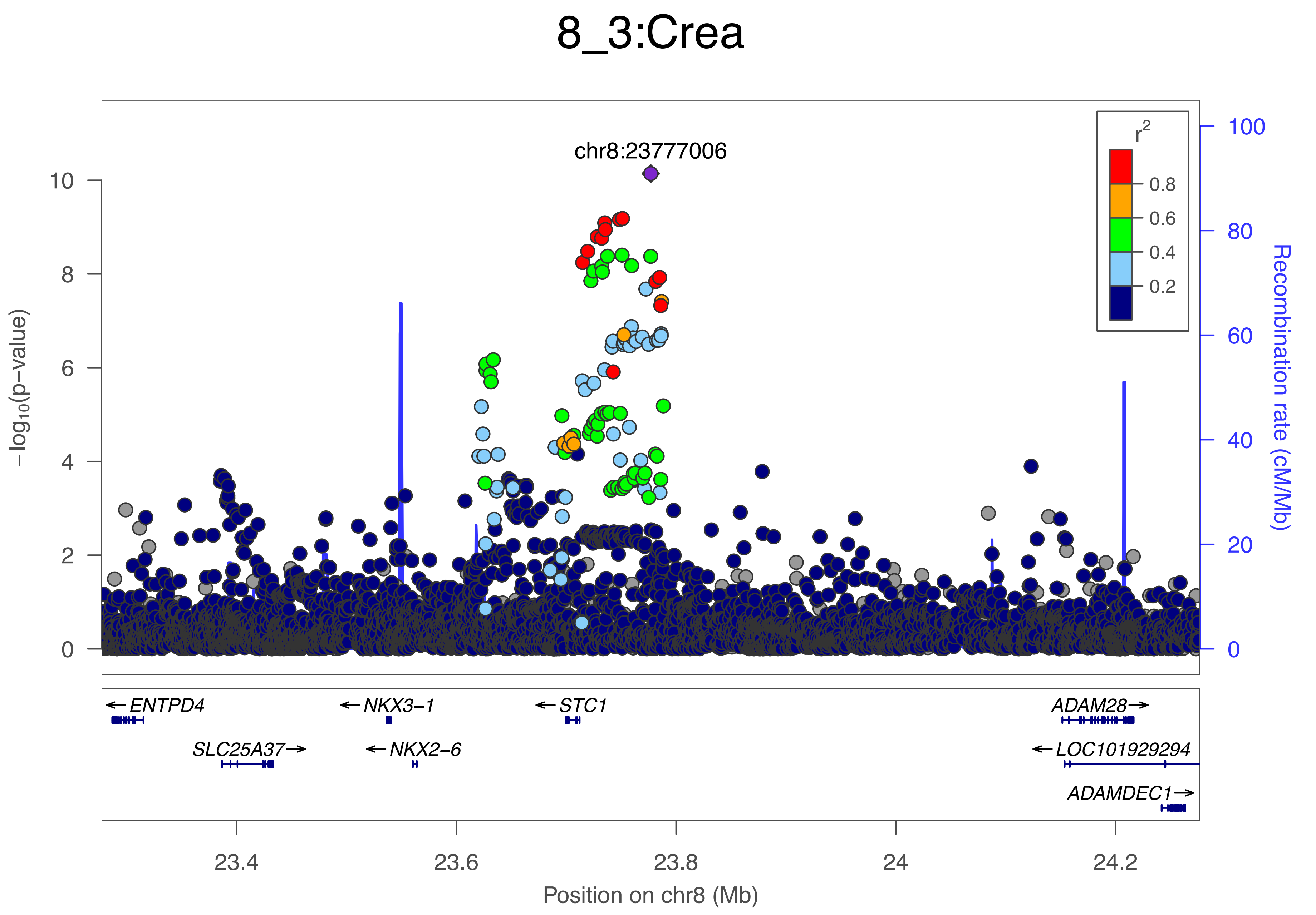

8_3:Crea
chr8:23777006
$r^2$
0.8
0.6
0.4
0.2
10
8
6
4
2
0
100
80
60
40
20
0
$-\log_{10}$(p-value)
Recombination rate (cM/Mb)
← ENTPD4
SLC25A37→
← NKX3-1
← NKX2-6
← STC1
ADAM28→
← LOC101929294
ADAMDEC1→
23.4
23.6
23.8
24
24.2
Position on chr8 (Mb)

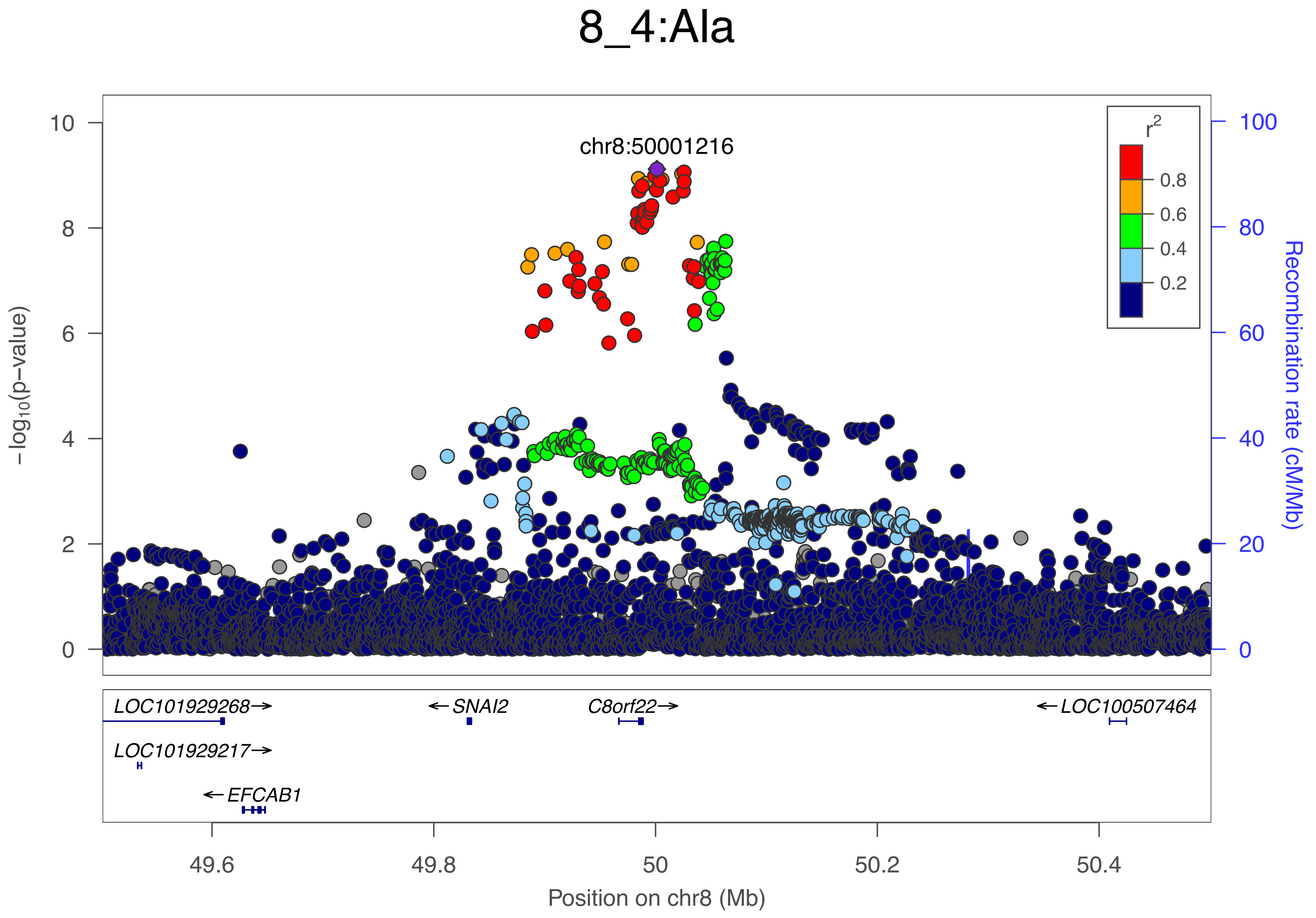

8_4:Ala
chr8:50001216
−log10(p−value)
Recombination rate (cM/Mb)
r2
0.8
0.6
0.4
0.2
0
2
4
6
8
10
20
40
60
80
100
LOC101929268→
←SNAI2
C8orf22→
←LOC100507464
LOC101929217→
←EFCAB1
49.6
49.8
50
50.2
50.4
Position on chr8 (Mb)

### 8_5:PUFA

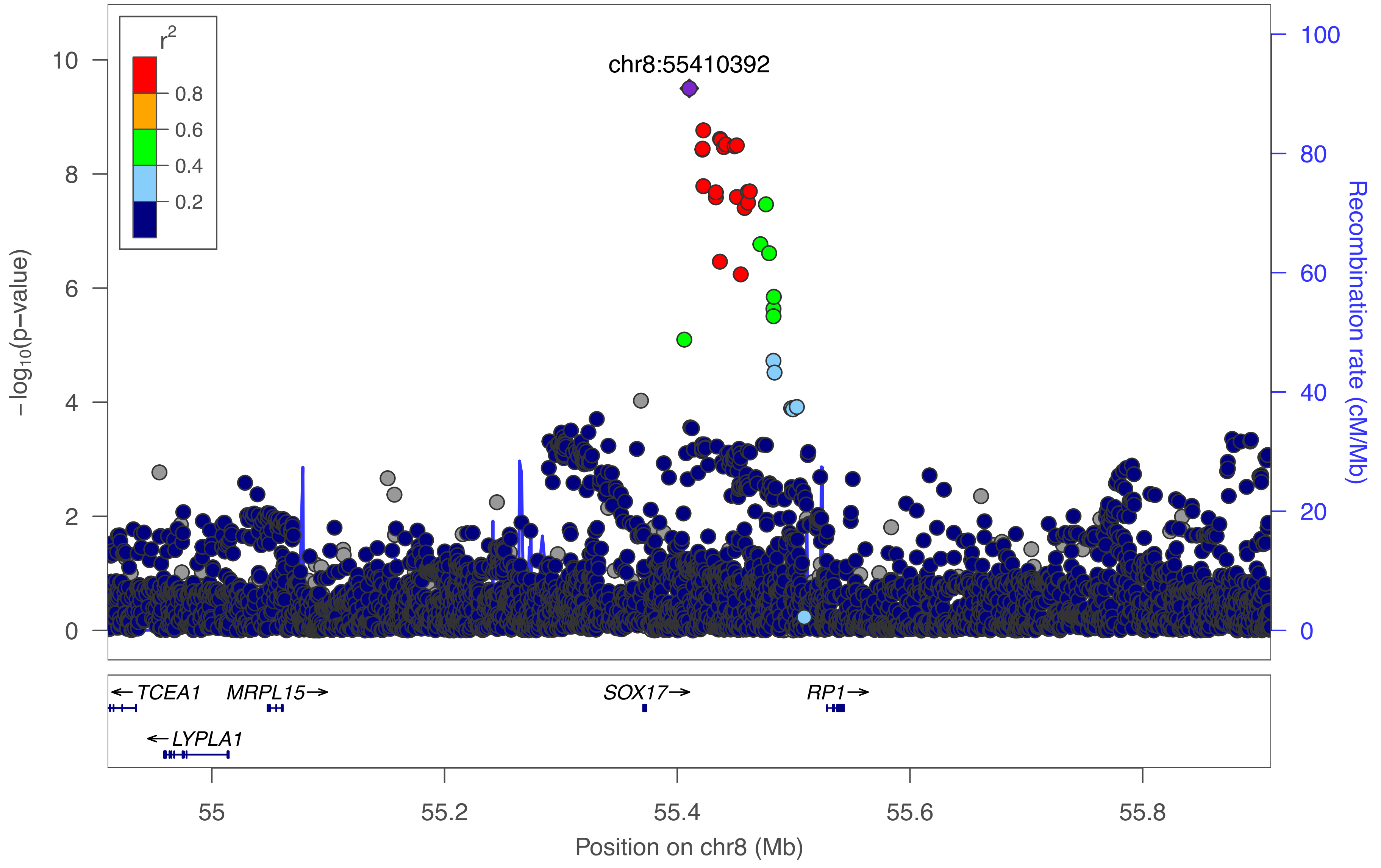

chr8:55410392
r2
0.8
0.6
0.4
0.2
10
8
6
4
2
0
−log10(p−value)
100
80
60
40
20
0
Recombination rate (cM/Mb)
← TCEA1
MRPL15→
SOX17→
RP1→
← LYPLA1
55
55.2
55.4
55.6
55.8
Position on chr8 (Mb)

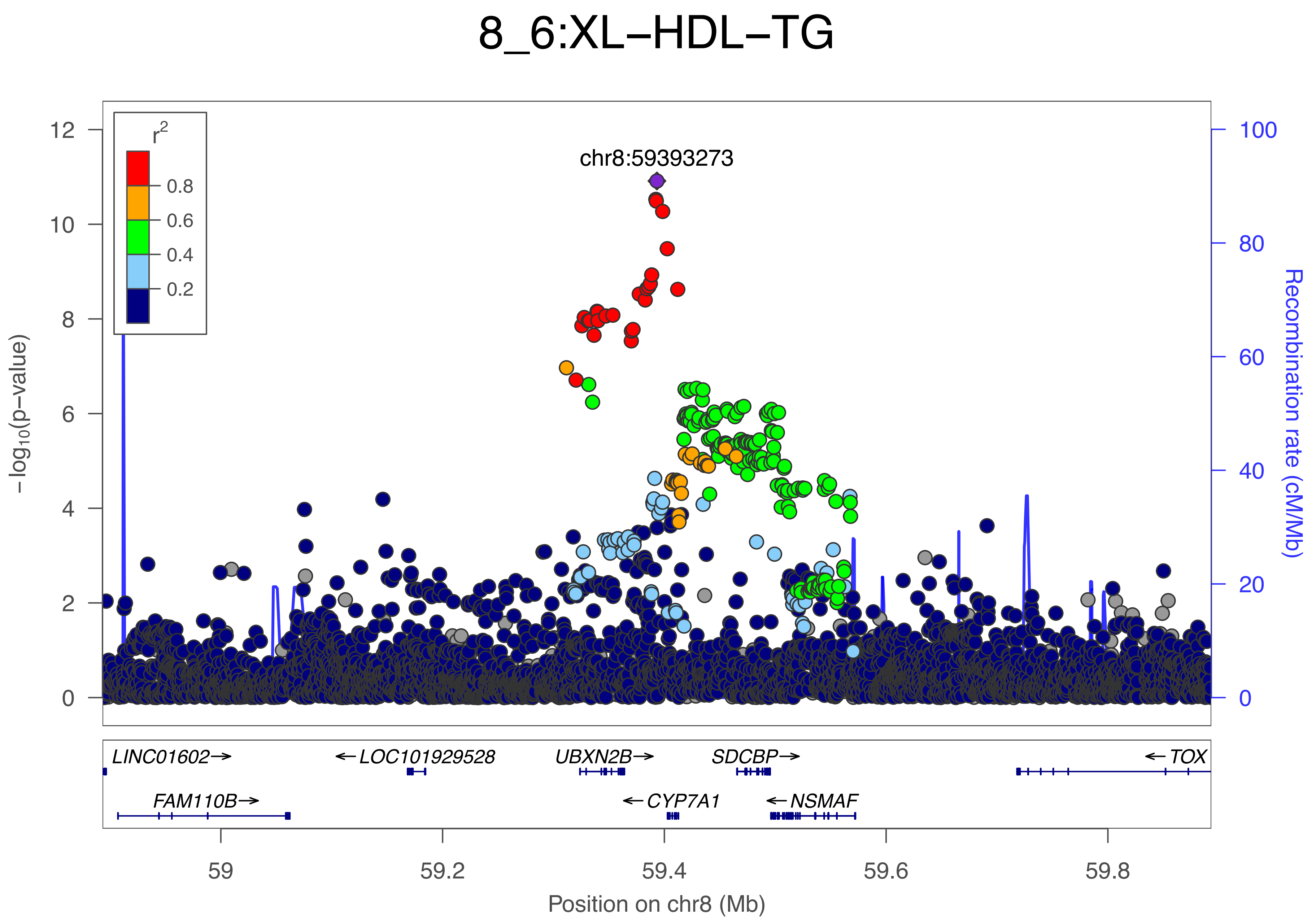

8_6:XL−HDL−TG
chr8:59393273
r2
0.8
0.6
0.4
0.2
−log10(p−value)
Recombination rate (cM/Mb)
12
10
8
6
4
2
0
100
80
60
40
20
0
LINC01602→
← LOC101929528
UBXN2B→
SDCBP→
← TOX
FAM110B→
← CYP7A1
← NSMAF
59
59.2
59.4
59.6
59.8
Position on chr8 (Mb)

### 8_7:XS−VLDL−PL

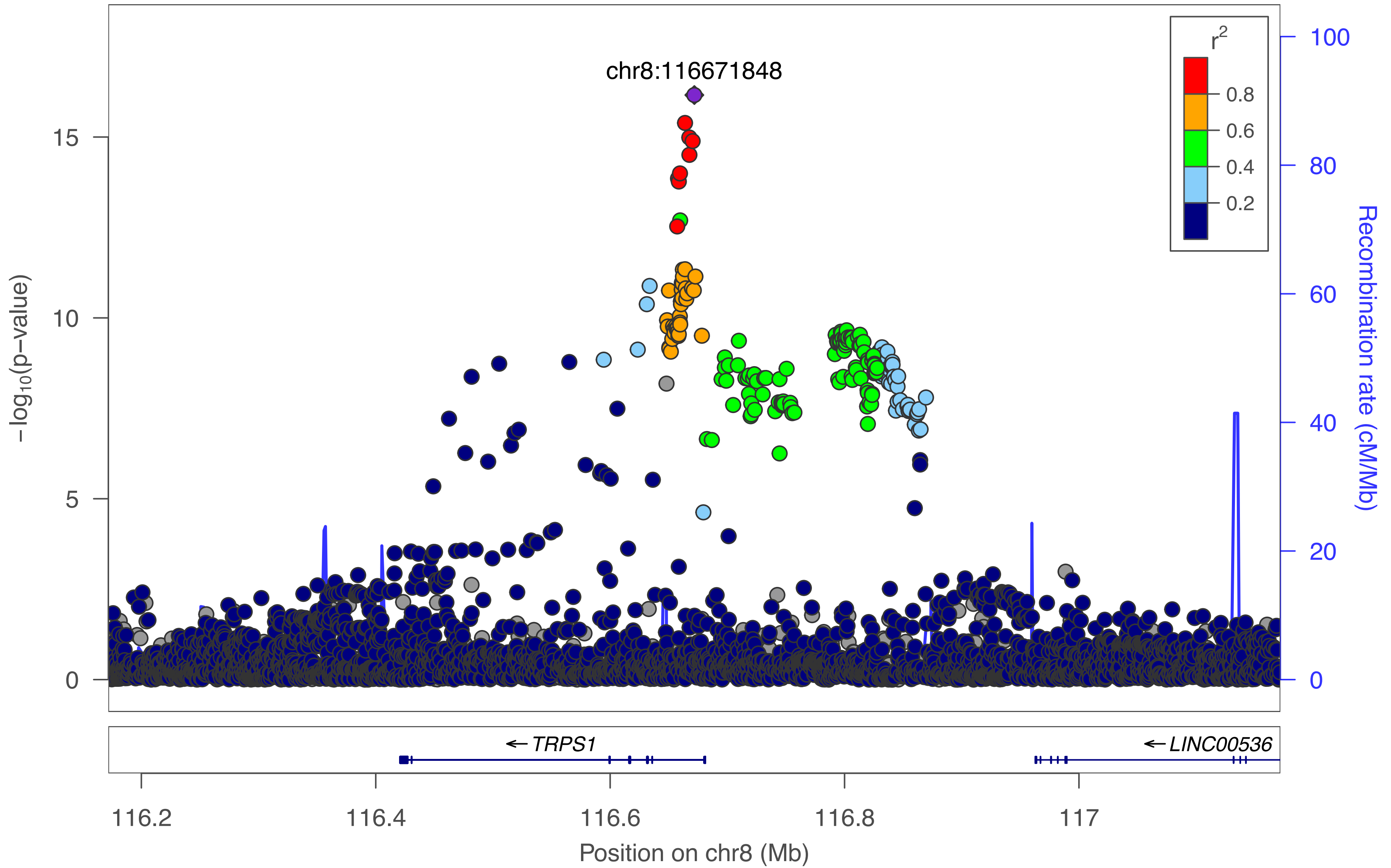

chr8:116671848
r²
0.8
0.6
0.4
0.2
15
10
5
0
−log₁₀(p−value)
100
80
60
40
20
0
Recombination rate (cM/Mb)
← TRPS1
← LINC00536
116.2
116.4
116.6
116.8
117
Position on chr8 (Mb)

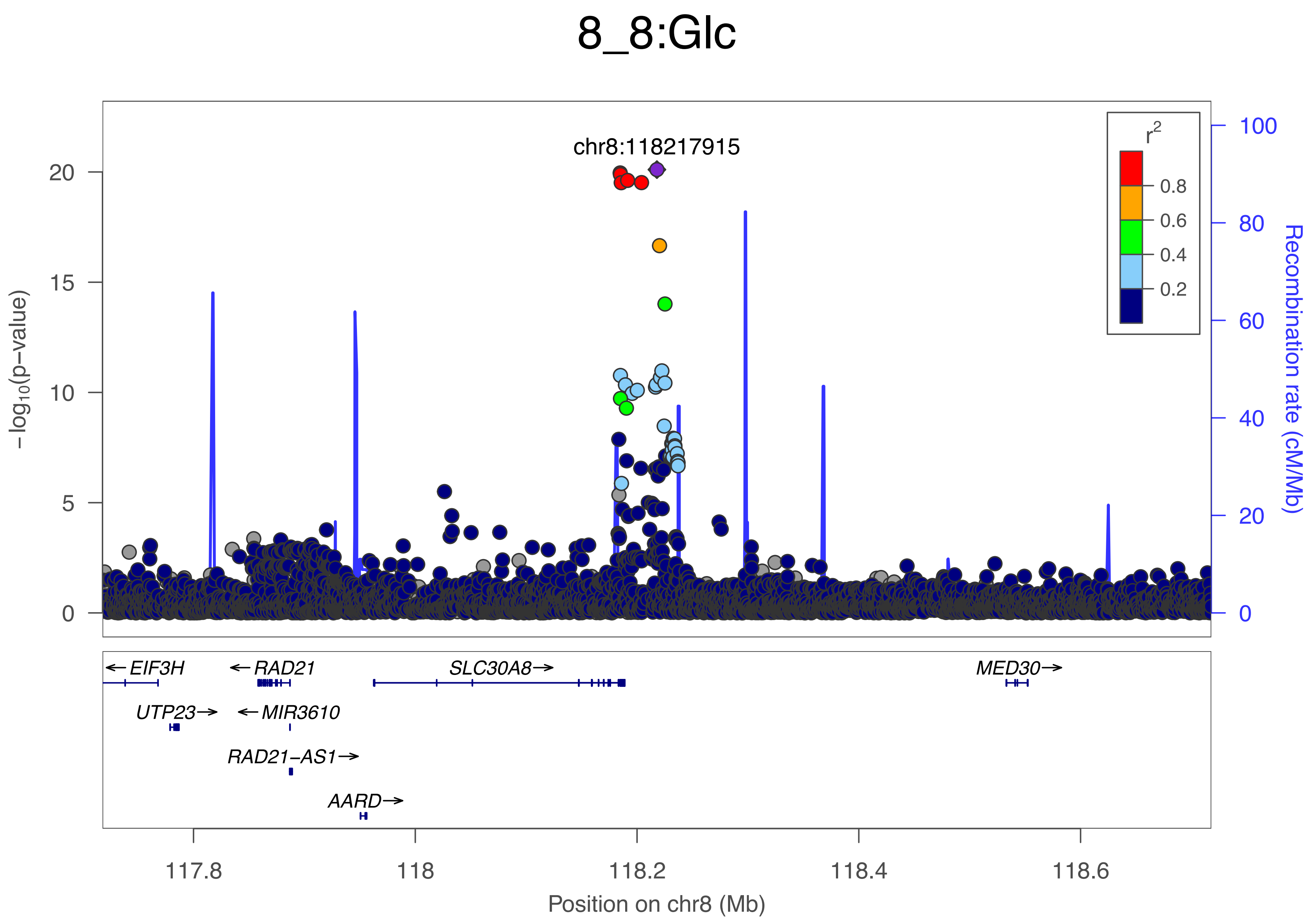
8_8:Glc
chr8:118217915
r2
0.8
0.6
0.4
0.2
−log10(p−value)
Recombination rate (cM/Mb)
← EIF3H
← RAD21
SLC30A8→
MED30→
UTP23→
← MIR3610
RAD21−AS1→
AARD→
117.8
118
118.2
118.4
118.6
Position on chr8 (Mb)

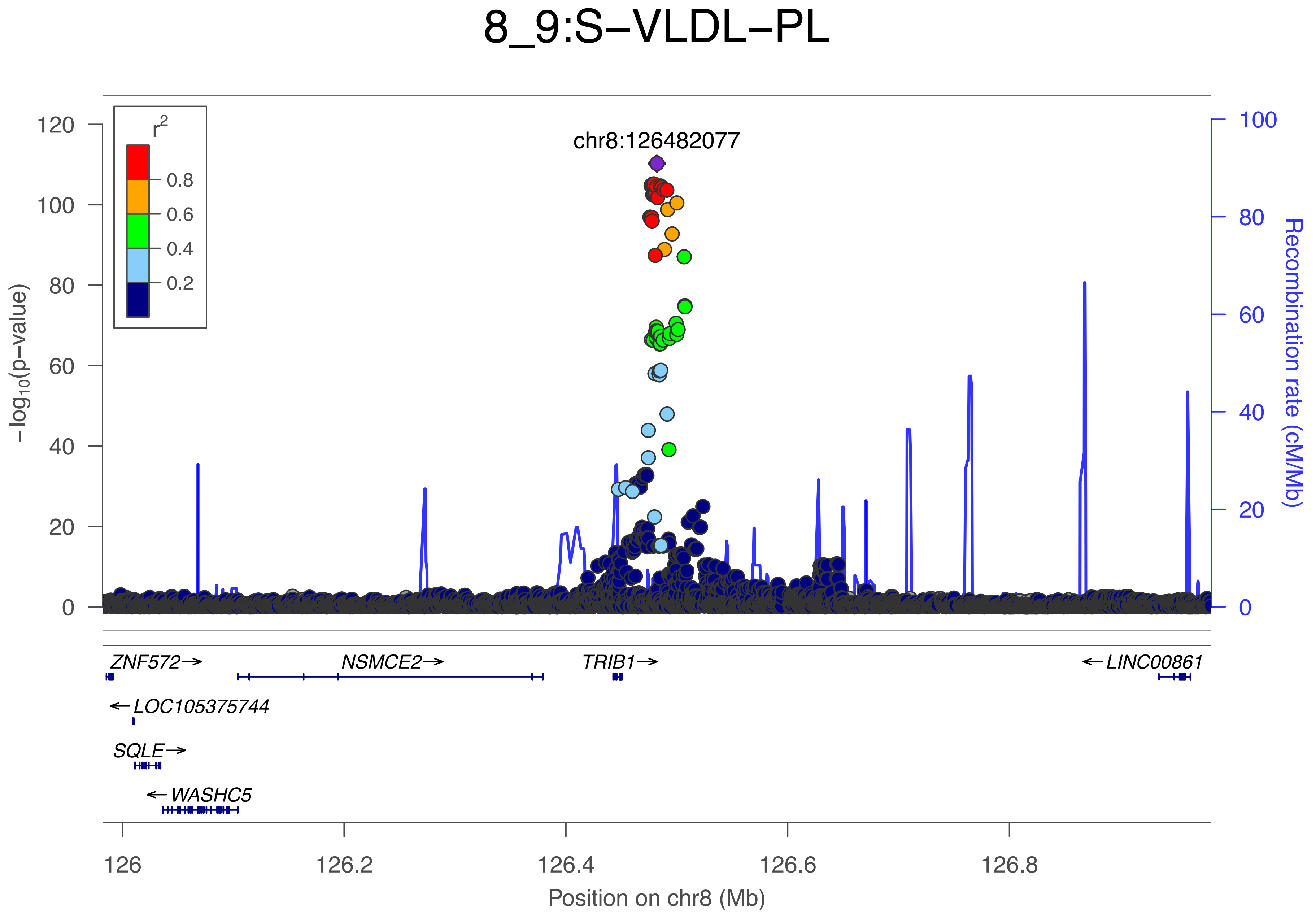

8_9:S−VLDL−PL
chr8:126482077
r2
0.8
0.6
0.4
0.2
−log10(p−value)
Recombination rate (cM/Mb)
ZNF572→
NSMCE2→
TRIB1→
←LINC00861
←LOC105375744
SQLE→
←WASHC5
126
126.2
126.4
126.6
126.8
Position on chr8 (Mb)

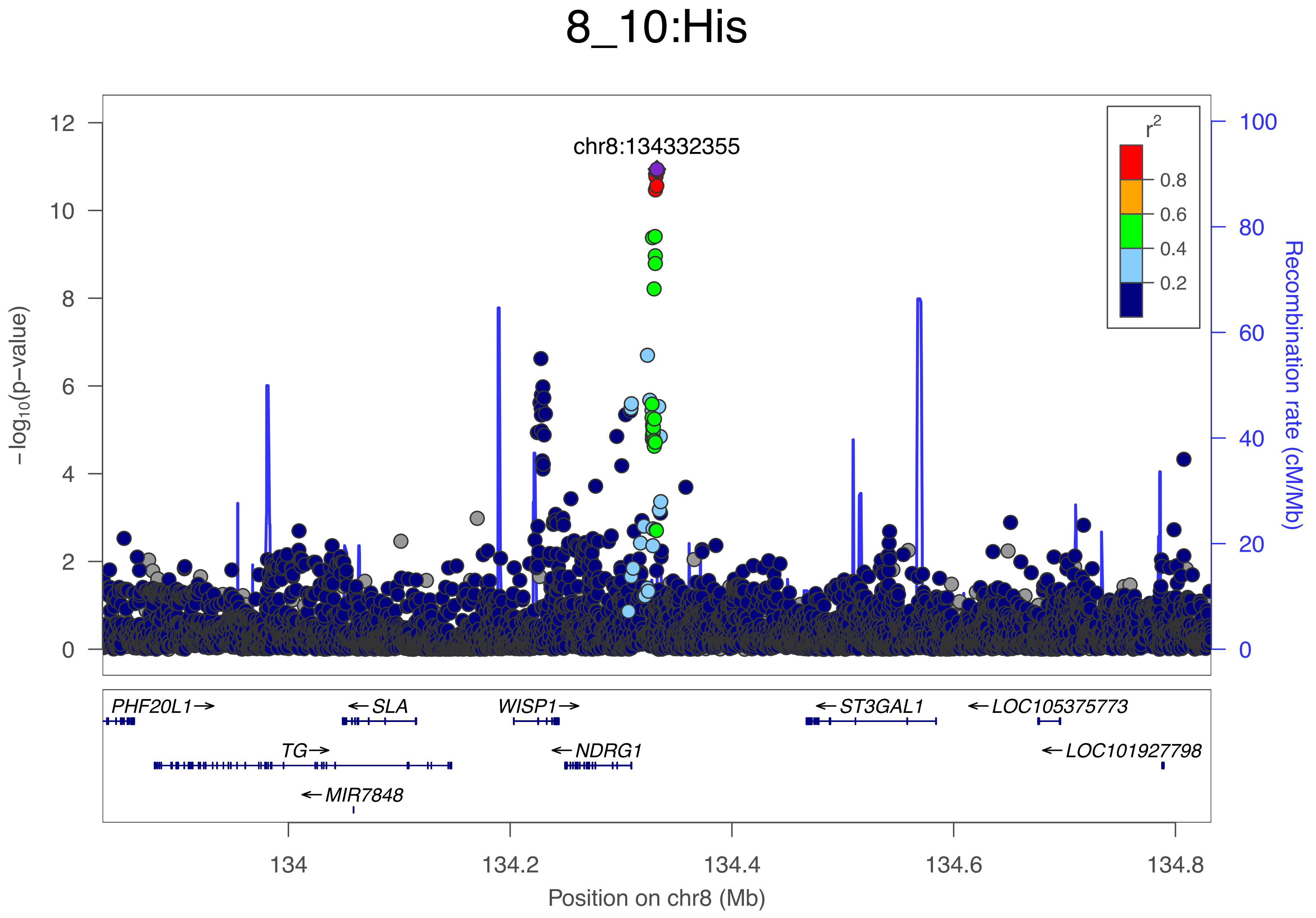
8_10:His
chr8:134332355
r2
0.8
0.6
0.4
0.2
−log10(p−value)
Recombination rate (cM/Mb)
PHF20L1→
←SLA
WISP1→
←ST3GAL1
←LOC105375773
TG→
←NDRG1
←LOC101927798
←MIR7848
134
134.2
134.4
134.6
134.8
Position on chr8 (Mb)

### 8_11:His

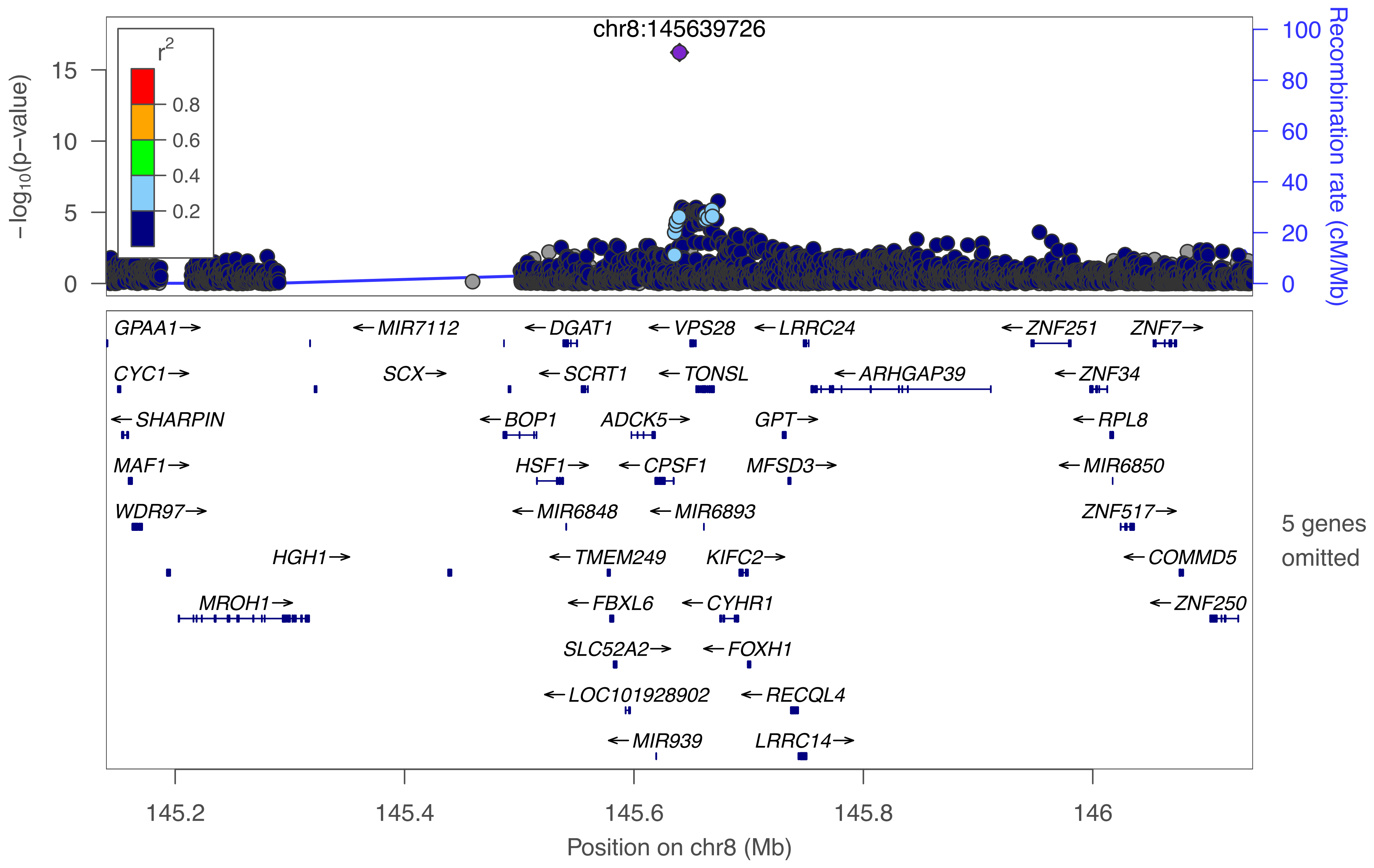

chr8:145639726
r2
0.8
0.6
0.4
0.2
−log10(p−value)
15
10
5
0
100
80
60
40
20
0
Recombination rate (cM/Mb)
GPAA1→
←MIR7112
←DGAT1
←VPS28
←LRRC24
←ZNF251
ZNF7→
CYC1→
SCX→
←SCRT1
←TONSL
←ARHGAP39
←ZNF34
←SHARPIN
←BOP1
ADCK5→
GPT→
←RPL8
MAF1→
HSF1→
←CPSF1
MFSD3→
←MIR6850
WDR97→
←MIR6848
←MIR6893
ZNF517→
HGH1→
←TMEM249
KIFC2→
←COMMD5
MROH1→
←FBXL6
←CYHR1
←ZNF250
SLC52A2→
←FOXH1
←LOC101928902
←RECQL4
←MIR939
LRRC14→
5 genes omitted
145.2
145.4
145.6
145.8
146
Position on chr8 (Mb)

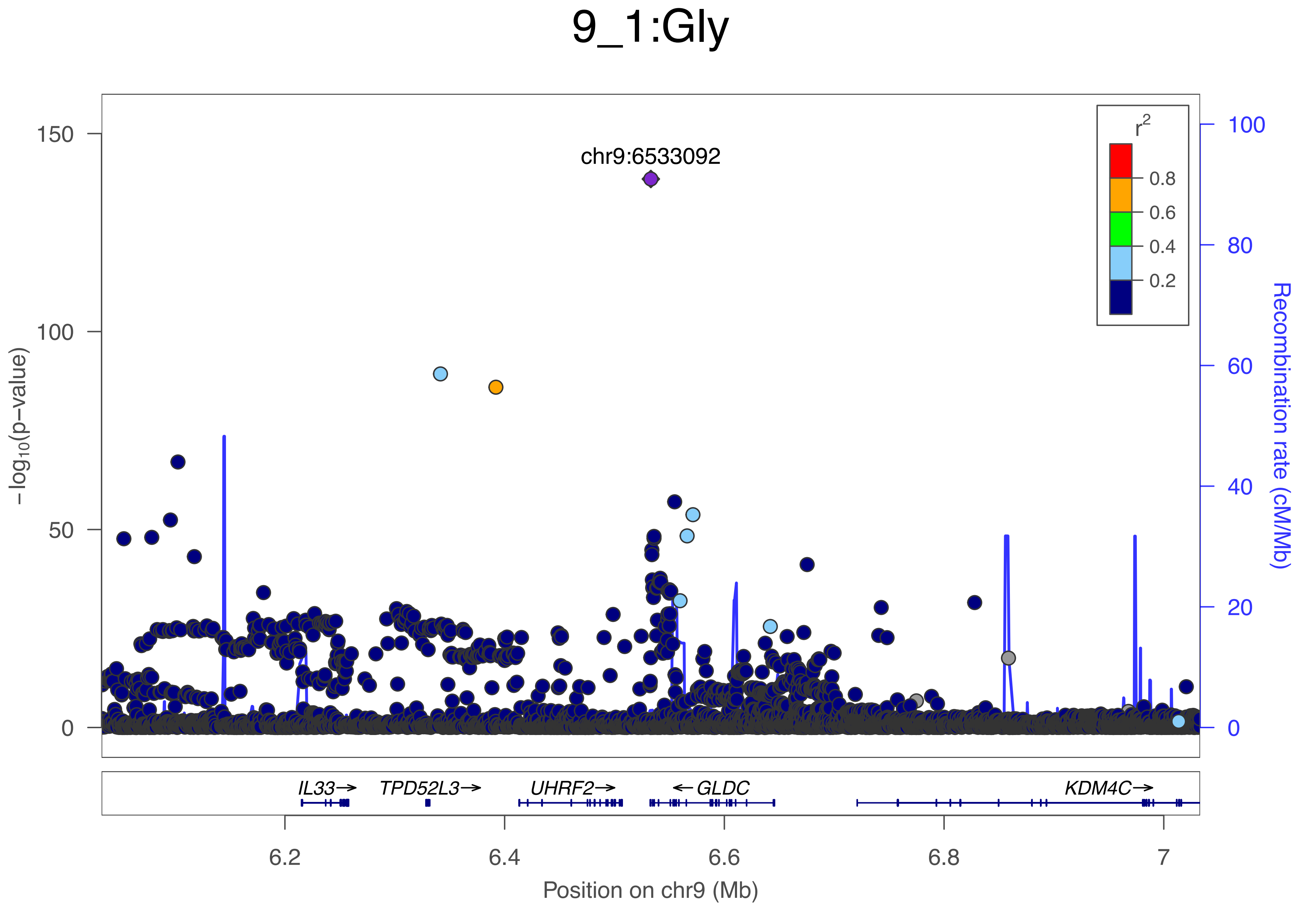

9_1:Gly
chr9:6533092
r2
0.8
0.6
0.4
0.2
150
100
50
0
100
80
60
40
20
0
−log10(p−value)
Recombination rate (cM/Mb)
IL33→
TPD52L3→
UHRF2→
←GLDC
KDM4C→
6.2
6.4
6.6
6.8
7
Position on chr9 (Mb)

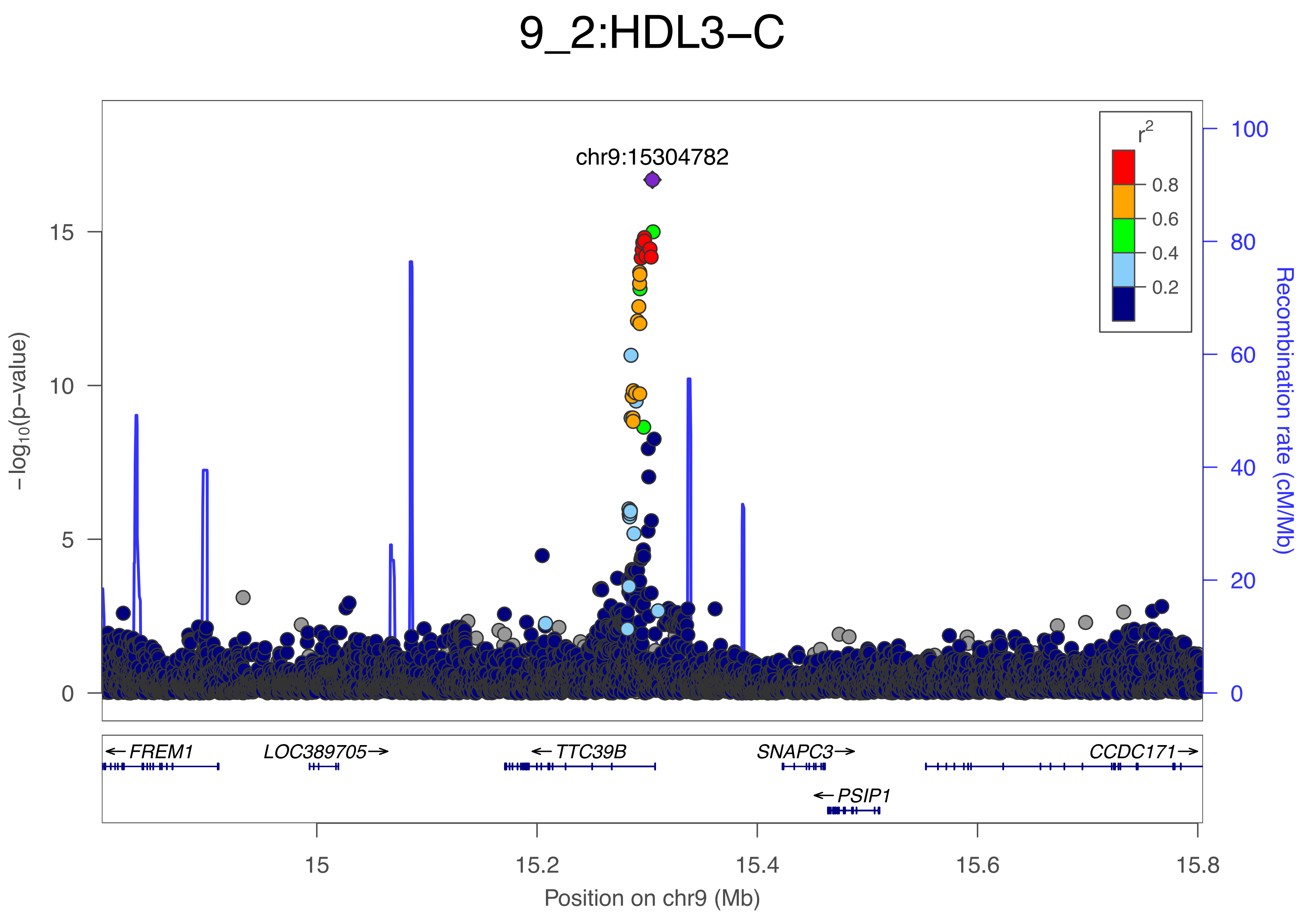

9_2:HDL3−C
chr9:15304782
r2
0.8
0.6
0.4
0.2
−log10(p−value)
0
5
10
15
Recombination rate (cM/Mb)
0
20
40
60
80
100
← FREM1
LOC389705→
← TTC39B
SNAPC3→
CCDC171→
← PSIP1
15
15.2
15.4
15.6
15.8
Position on chr9 (Mb)

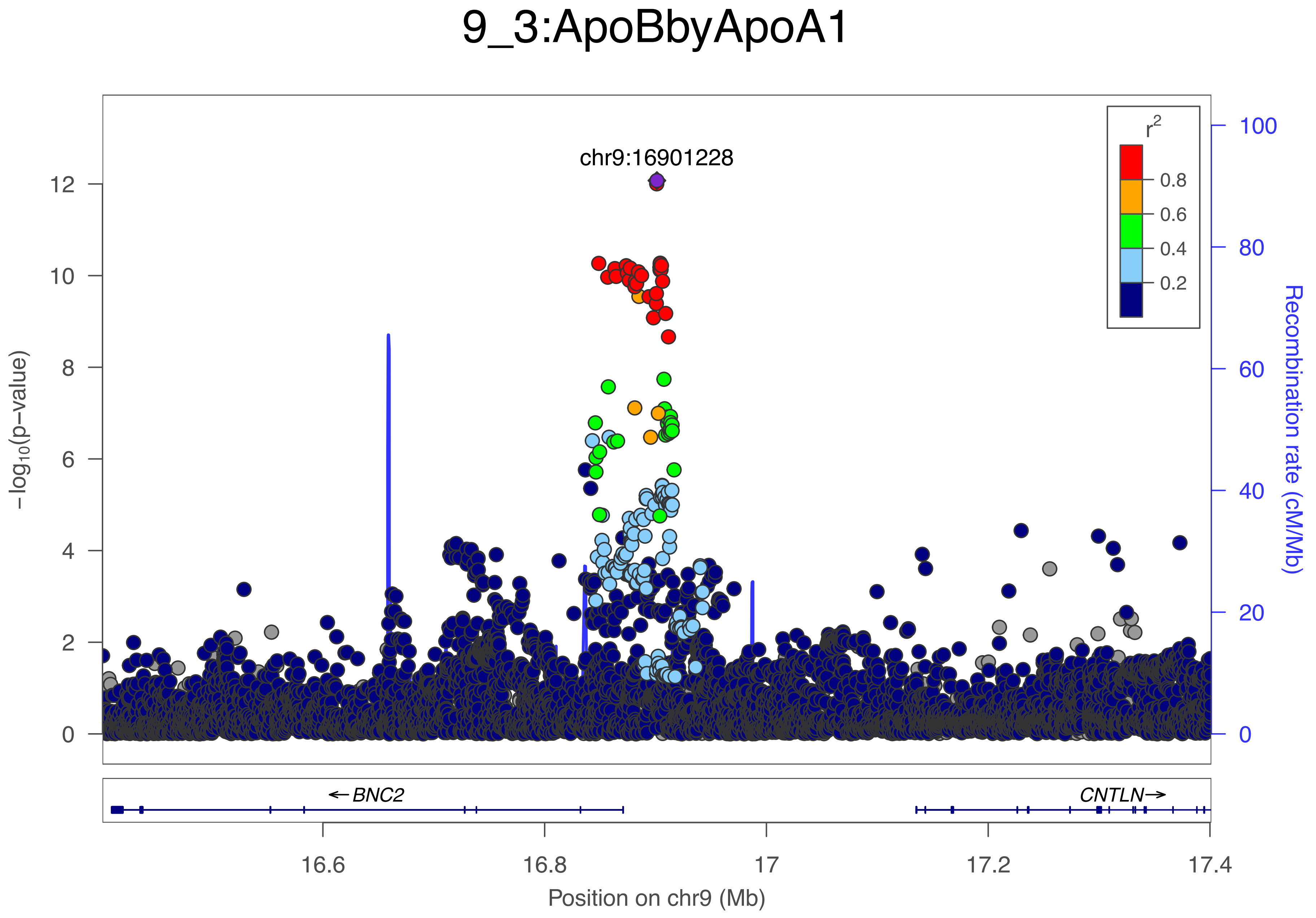
9_3:ApoBbyApoA1
chr9:16901228
r2
0.8
0.6
0.4
0.2
−log10(p−value)
Recombination rate (cM/Mb)
BNC2
CNTLN
Position on chr9 (Mb)
16.6
16.8
17
17.2
17.4
0
2
4
6
8
10
12
20
40
60
80
100

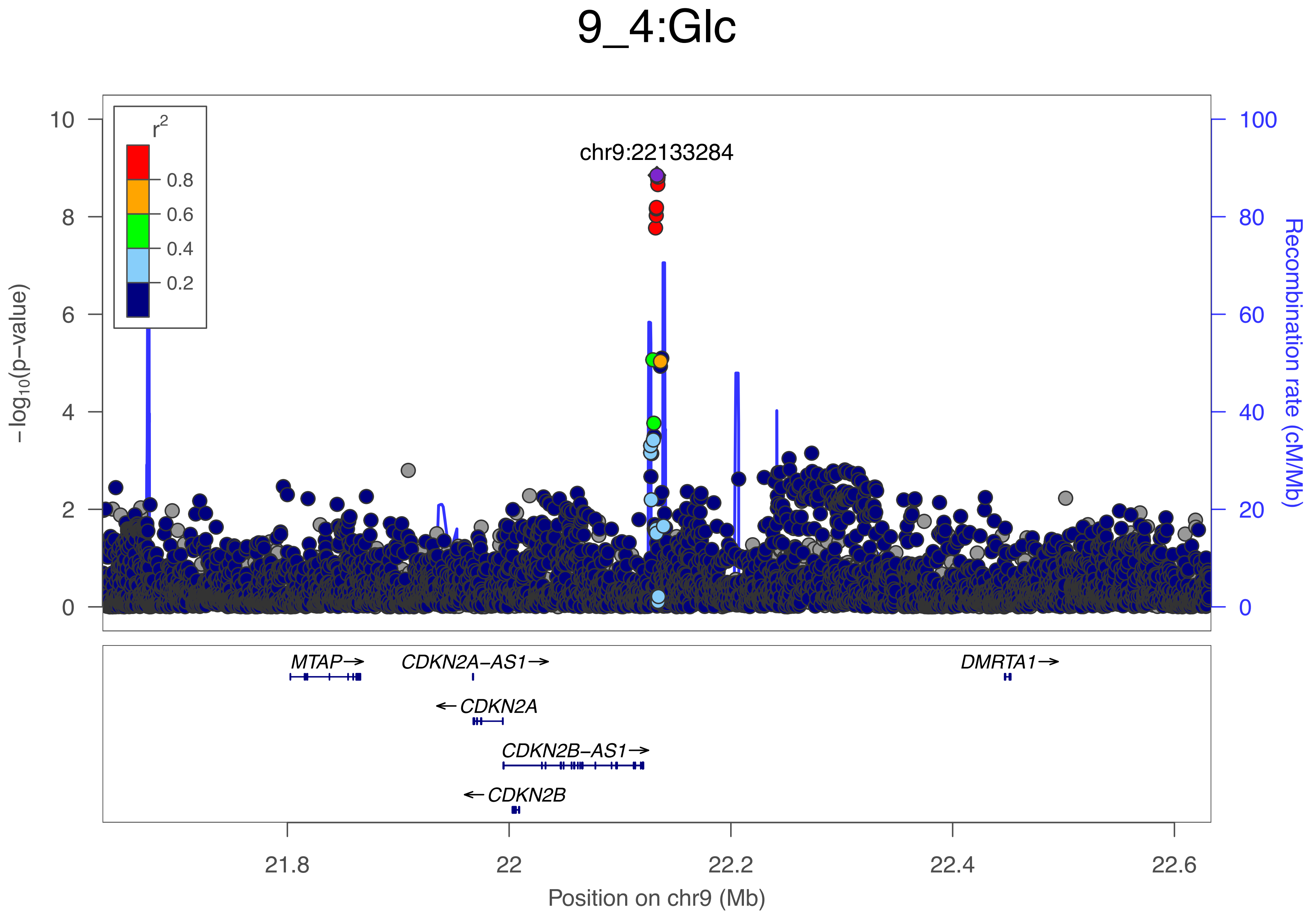

9_4:Glc
chr9:22133284
r2
0.8
0.6
0.4
0.2
−log10(p−value)
Recombination rate (cM/Mb)
0
2
4
6
8
10
0
20
40
60
80
100
MTAP→
CDKN2A−AS1→
←CDKN2A
CDKN2B−AS1→
←CDKN2B
DMRTA1→
21.8
22
22.2
22.4
22.6
Position on chr9 (Mb)

### 9_5:Crea

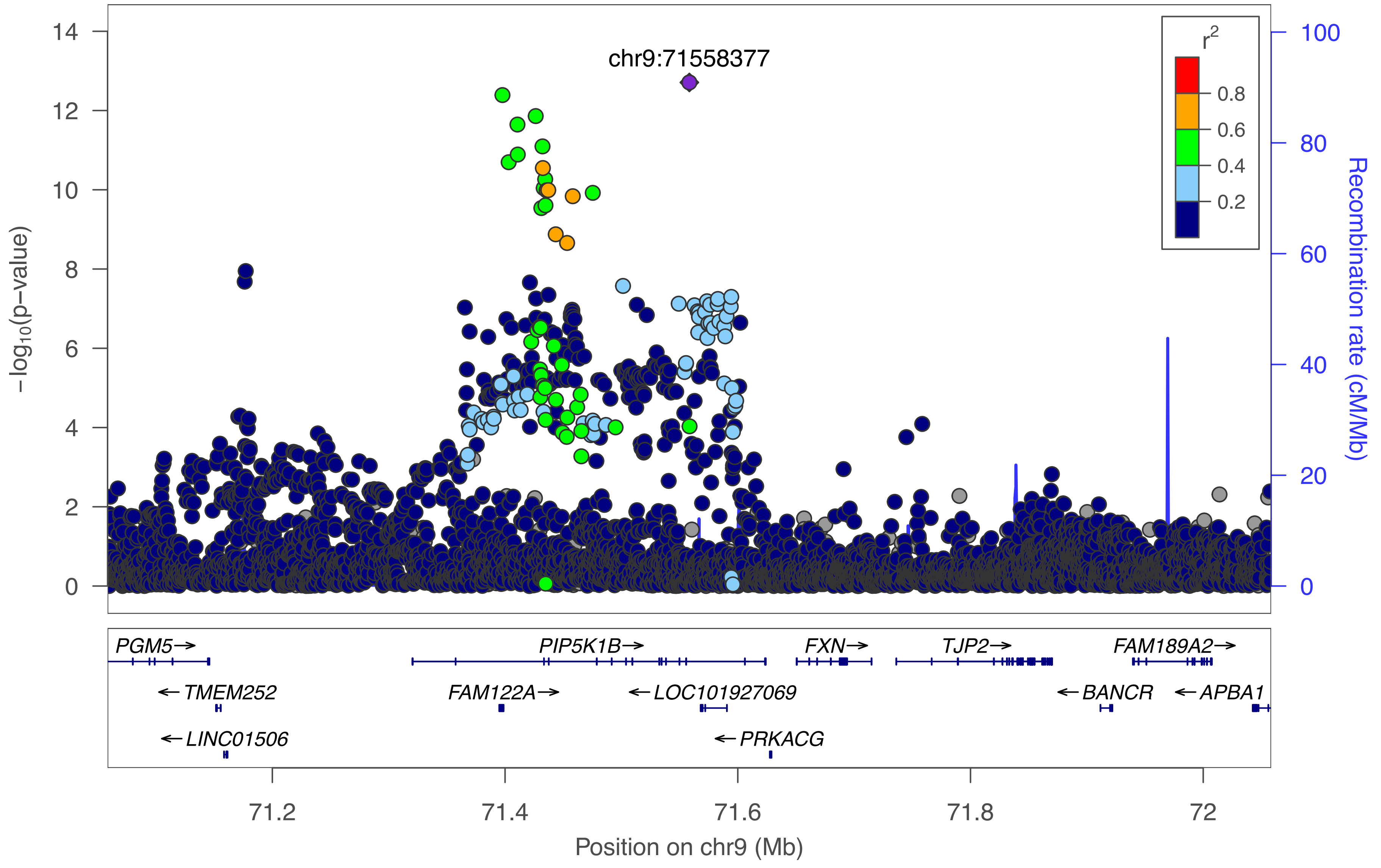

chr9:71558377
r2
0.8
0.6
0.4
0.2
−log10(p−value)
0
2
4
6
8
10
12
14
Recombination rate (cM/Mb)
0
20
40
60
80
100
PGM5→
PIP5K1B→
FXN→
TJP2→
FAM189A2→
←TMEM252
FAM122A→
←LOC101927069
←BANCR
←APBA1
←LINC01506
←PRKACG
71.2
71.4
71.6
71.8
72
Position on chr9 (Mb)

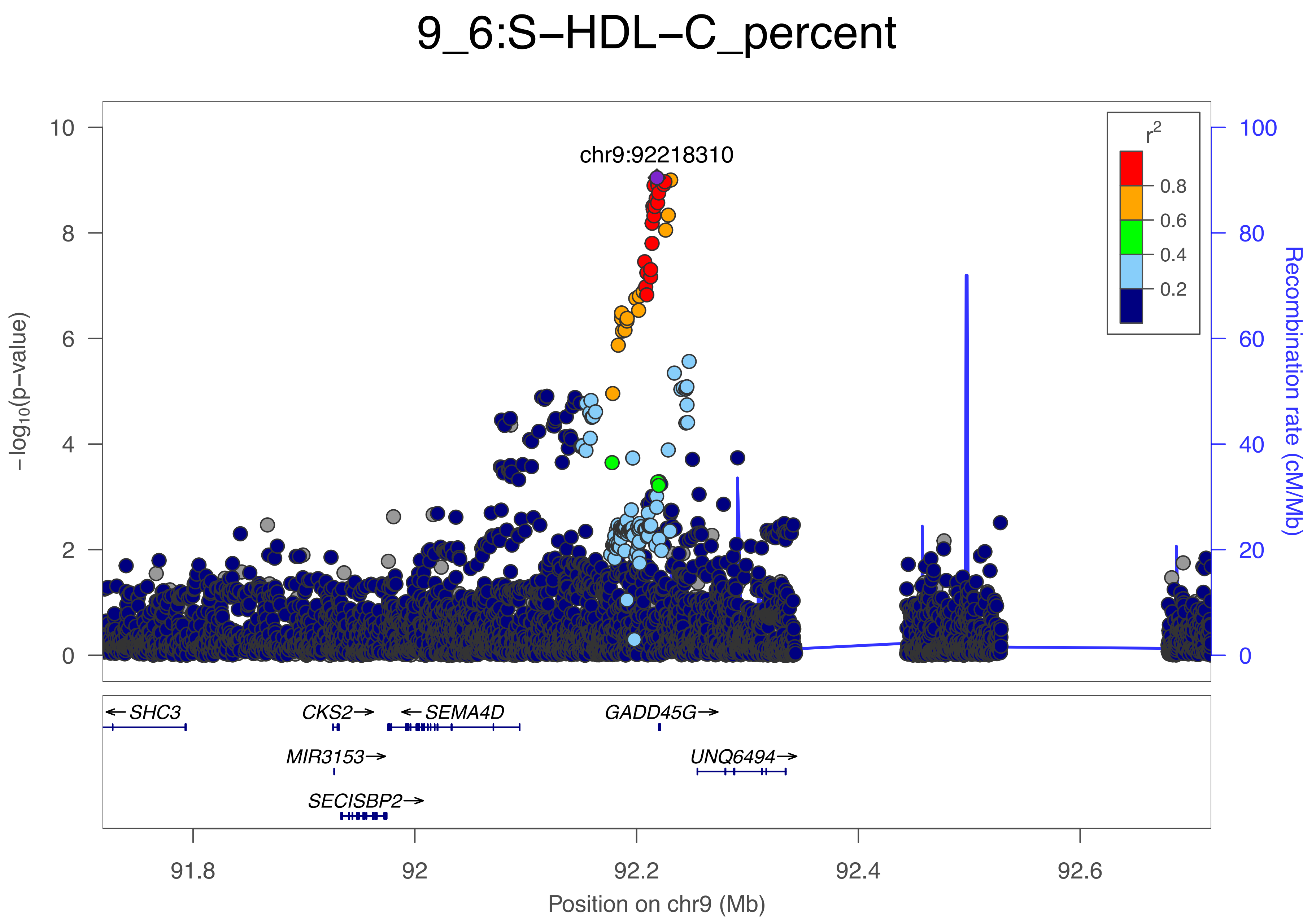
9_6:S−HDL−C_percent
chr9:92218310
r2
0.8
0.6
0.4
0.2
−log10(p−value)
Recombination rate (cM/Mb)
SHC3
CKS2
SEMA4D
GADD45G
MIR3153
UNQ6494
SECISBP2
91.8
92
92.2
92.4
92.6
Position on chr9 (Mb)

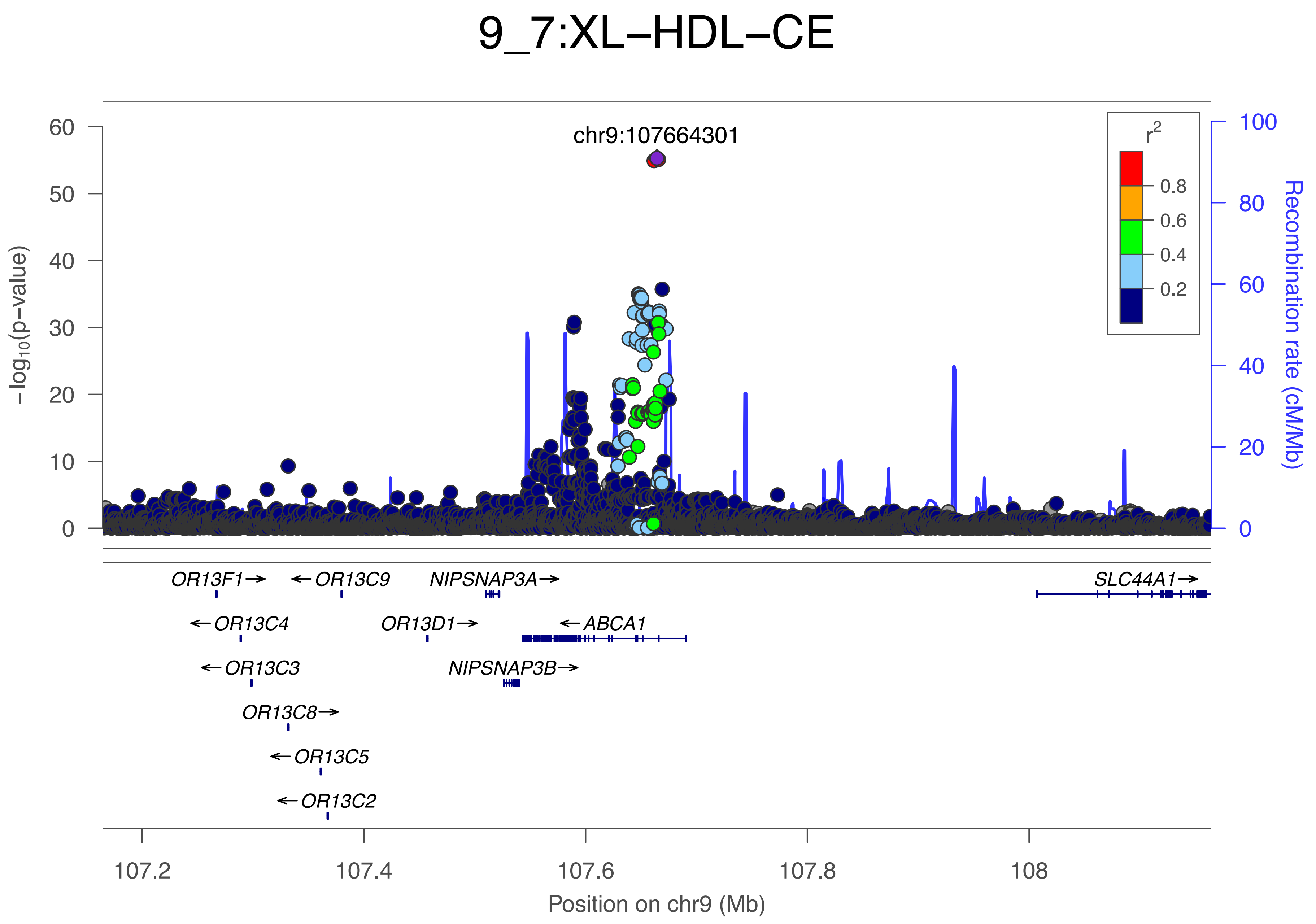

9_7:XL–HDL–CE
chr9:107664301
r2
0.8
0.6
0.4
0.2
60
50
40
30
20
10
0
−log10(p−value)
100
80
60
40
20
0
Recombination rate (cM/Mb)
OR13F1→
←OR13C9
NIPSNAP3A→
SLC44A1→
←OR13C4
OR13D1→
←ABCA1
←OR13C3
NIPSNAP3B→
OR13C8→
←OR13C5
←OR13C2
107.2
107.4
107.6
107.8
108
Position on chr9 (Mb)

### 9_8:GlycA

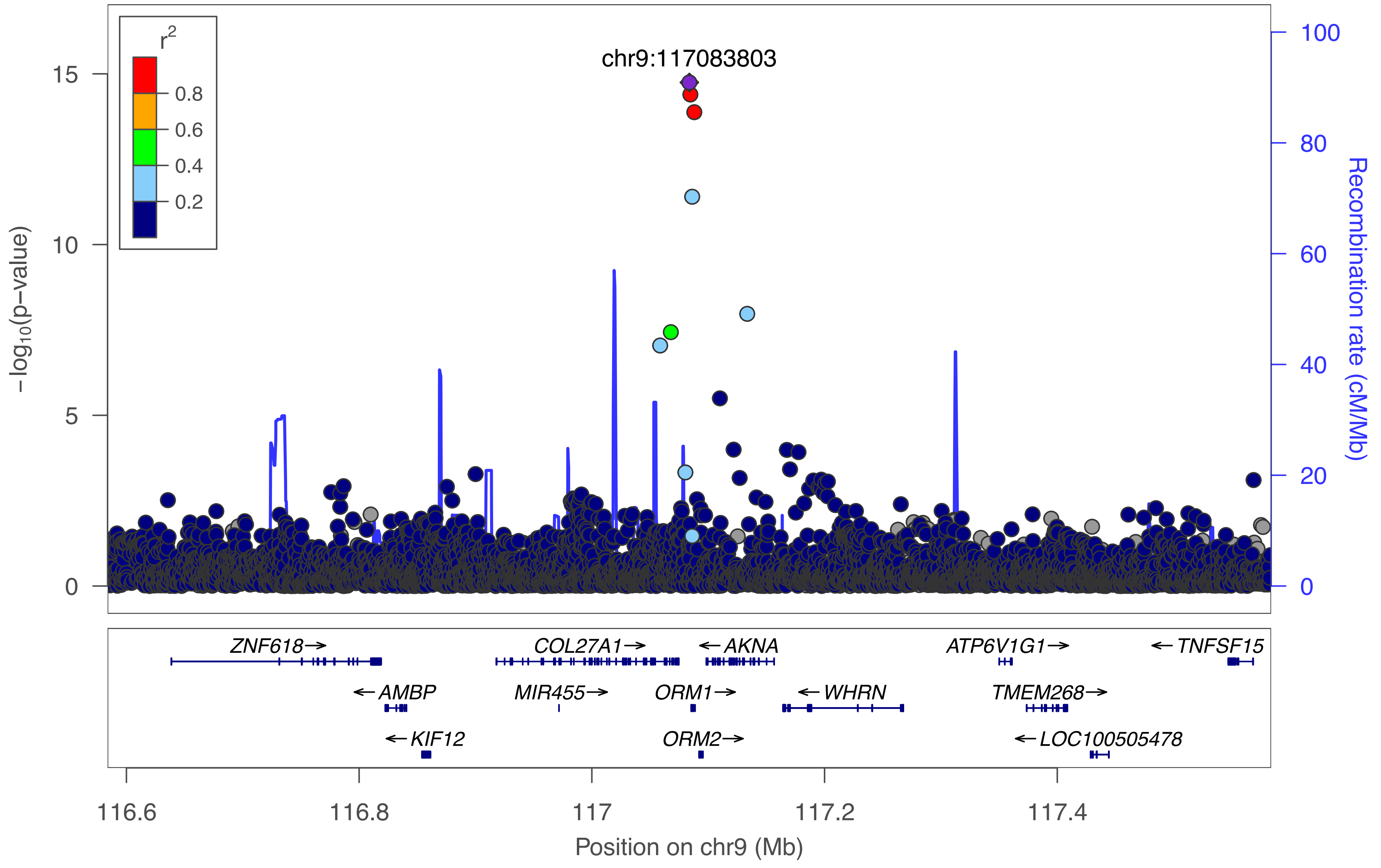

chr9:117083803
r2
0.8
0.6
0.4
0.2
15
10
5
0
−log10(p−value)
100
80
60
40
20
0
Recombination rate (cM/Mb)
ZNF618→
COL27A1→
←AKNA
ATP6V1G1→
←TNFSF15
←AMBP
MIR455→
ORM1→
←WHRN
TMEM268→
←KIF12
ORM2→
←LOC100505478
116.6
116.8
117
117.2
117.4
Position on chr9 (Mb)

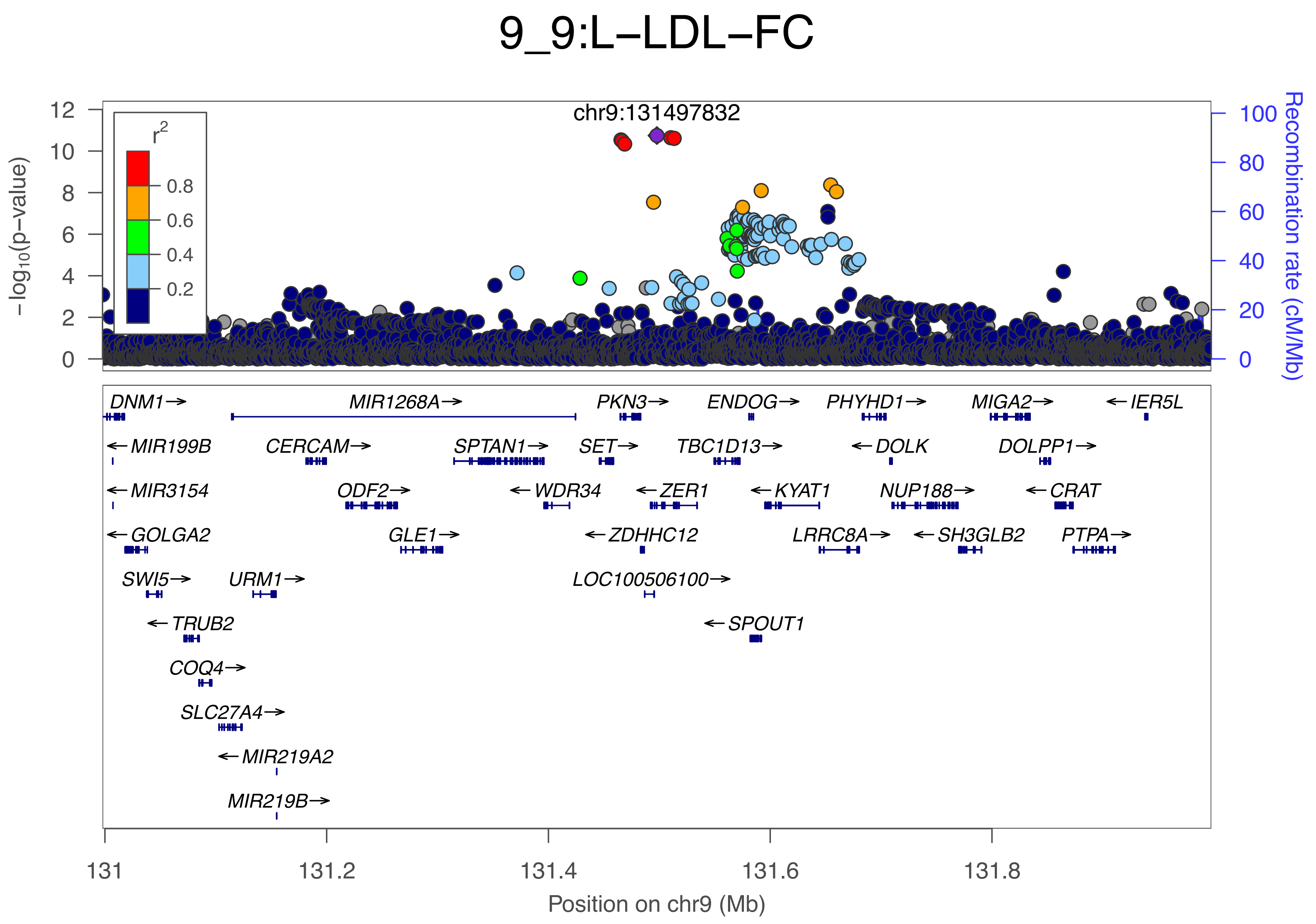

9_9:L−LDL−FC
chr9:131497832
r2
0.8
0.6
0.4
0.2
−log10(p−value)
Recombination rate (cM/Mb)
DNM1→
MIR1268A→
PKN3→
ENDOG→
PHYHD1→
MIGA2→
←IER5L
←MIR199B
CERCAM→
SPTAN1→
SET→
TBC1D13→
←DOLK
DOLPP1→
←MIR3154
ODF2→
←WDR34
←ZER1
←KYAT1
NUP188→
←CRAT
←GOLGA2
GLE1→
←ZDHHC12
LRRC8A→
←SH3GLB2
PTPA→
SWI5→
URM1→
LOC100506100→
←TRUB2
←SPOUT1
COQ4→
SLC27A4→
←MIR219A2
MIR219B→
131
131.2
131.4
131.6
131.8
Position on chr9 (Mb)

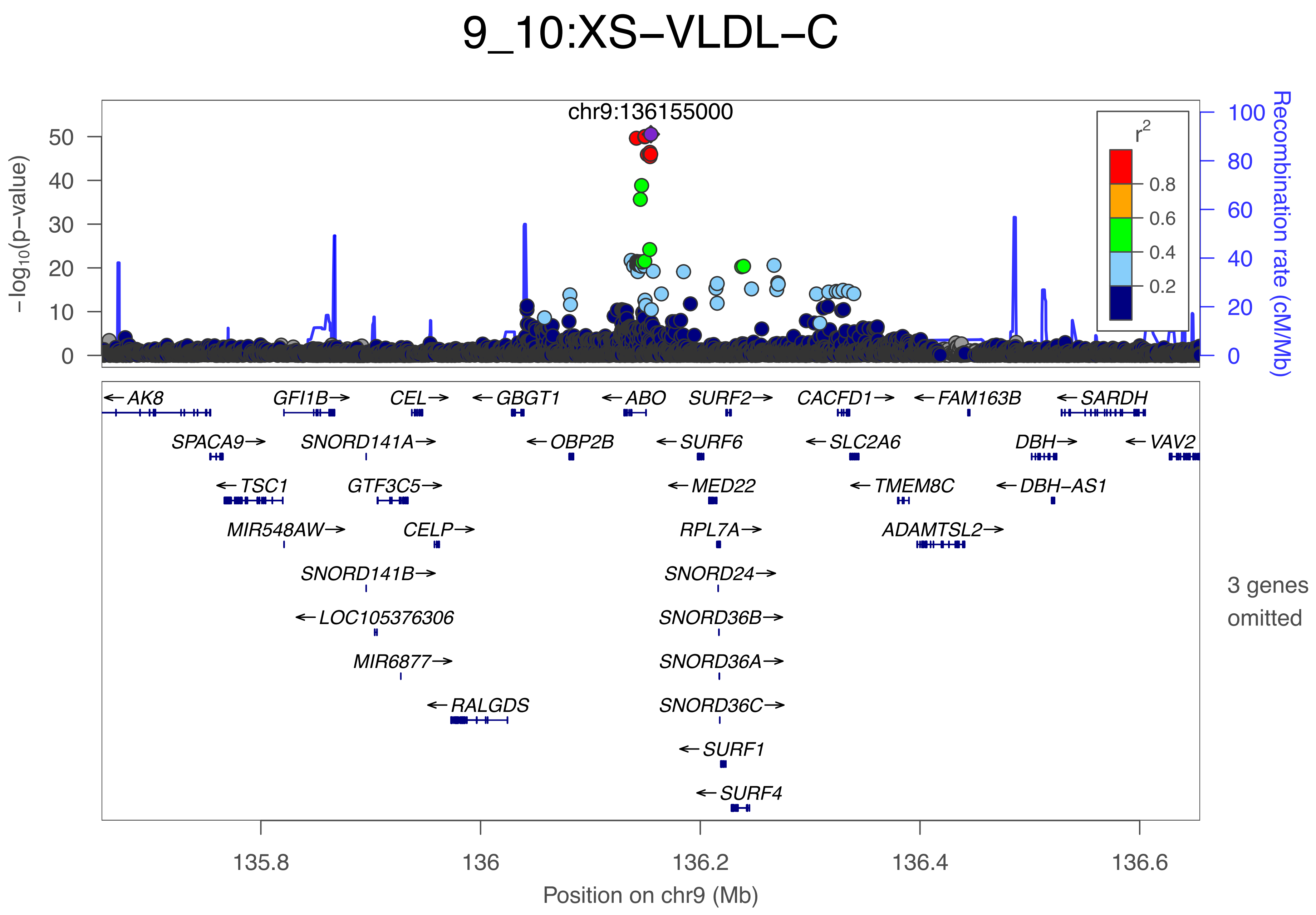
9_10:XS−VLDL−C
chr9:136155000
−log10(p−value)
Recombination rate (cM/Mb)
r2
0.8
0.6
0.4
0.2
AK8
GFI1B
CEL
GBGT1
ABO
SURF2
CACFD1
FAM163B
SARDH
SPACA9
SNORD141A
OBP2B
SURF6
SLC2A6
DBH
VAV2
TSC1
GTF3C5
MED22
TMEM8C
DBH−AS1
MIR548AW
CELP
RPL7A
ADAMTSL2
SNORD141B
SNORD24
LOC105376306
SNORD36B
MIR6877
SNORD36A
RALGDS
SNORD36C
SURF1
SURF4
3 genes omitted
135.8
136
136.2
136.4
136.6
Position on chr9 (Mb)

### 9_11:MUFAbyFA

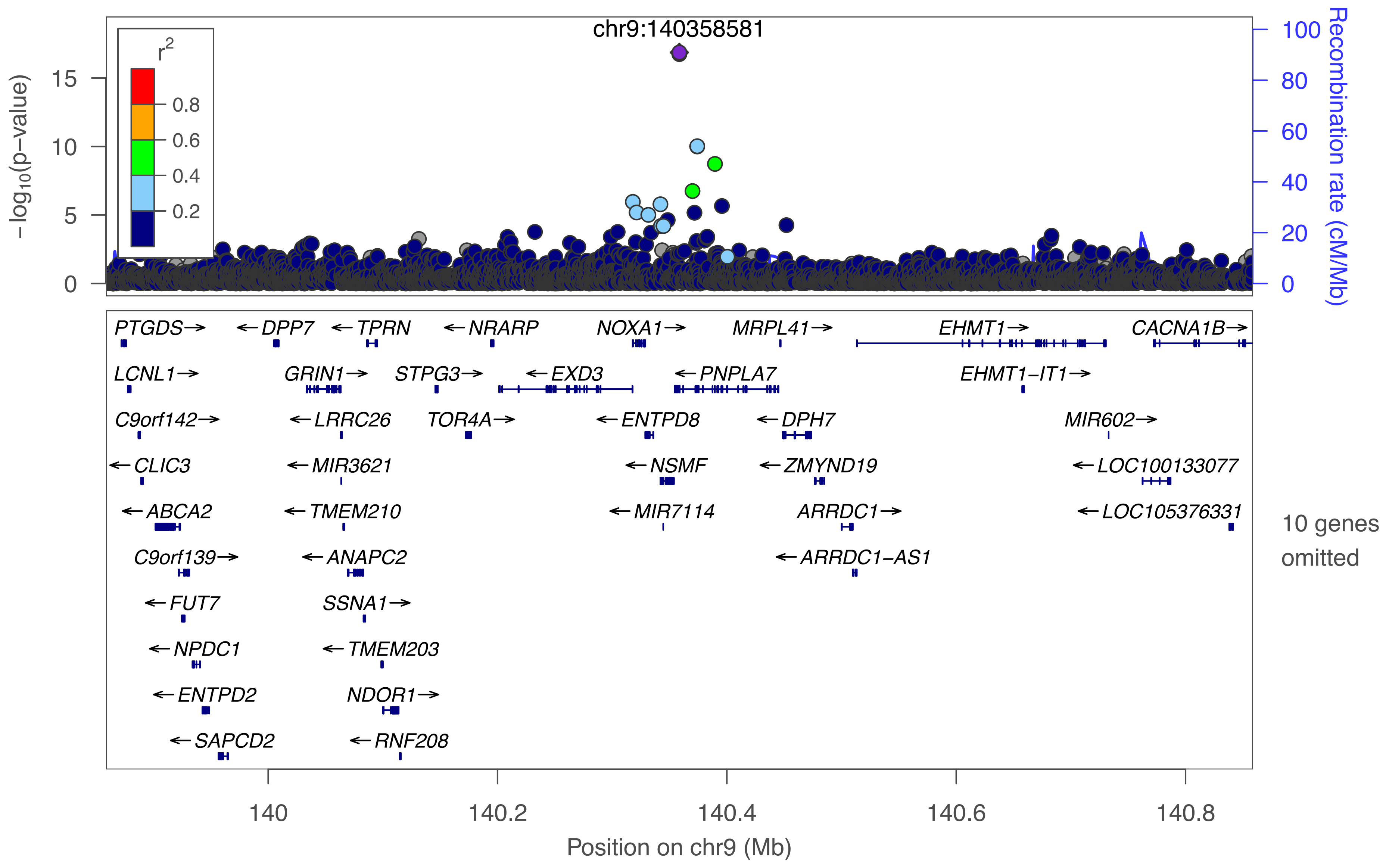

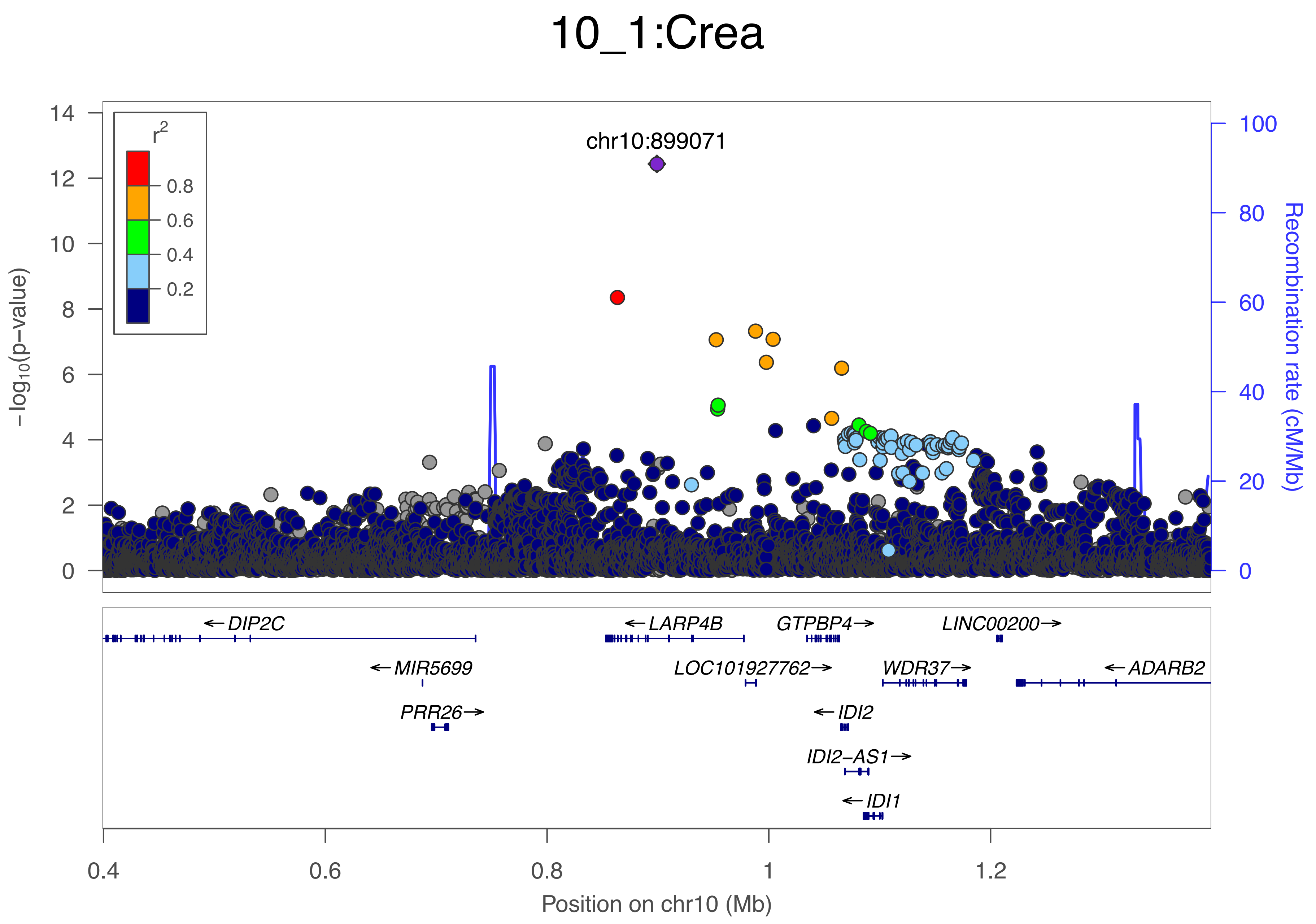

10_1:Crea
chr10:899071
r2
0.8
0.6
0.4
0.2
−log10(p−value)
Recombination rate (cM/Mb)
0
2
4
6
8
10
12
14
0
20
40
60
80
100
←DIP2C
←LARP4B
GTPBP4→
LINC00200→
←MIR5699
LOC101927762→
WDR37→
←ADARB2
PRR26→
←IDI2
IDI2−AS1→
←IDI1
0.4
0.6
0.8
1
1.2
Position on chr10 (Mb)

### 10_2:Lac

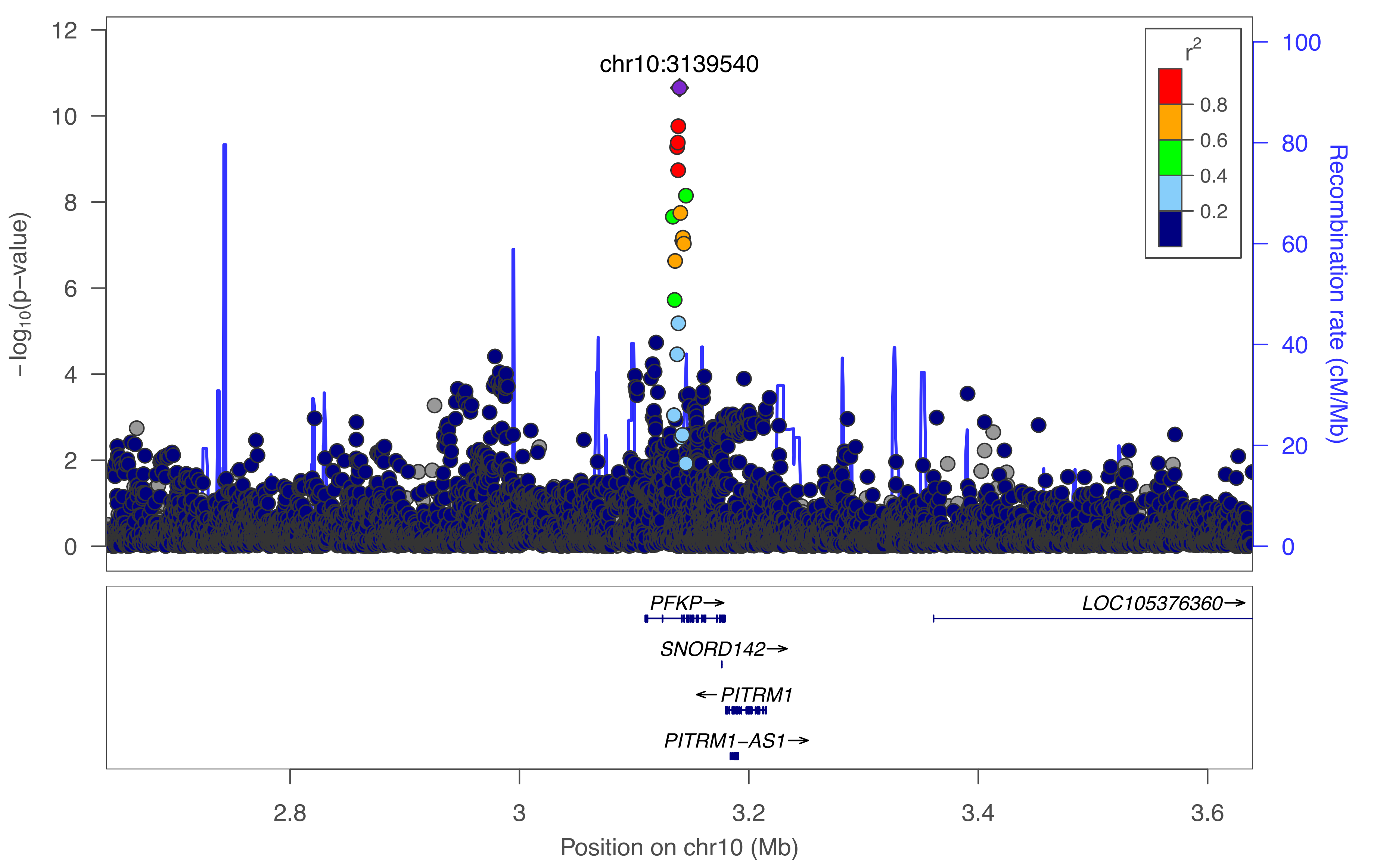
chr10:3139540
$-\log_{10}$(p−value)
Recombination rate (cM/Mb)
$r^2$
0.8
0.6
0.4
0.2
PFKP→
LOC105376360→
SNORD142→
← PITRM1
PITRM1−AS1→
2.8
3
3.2
3.4
3.6
Position on chr10 (Mb)

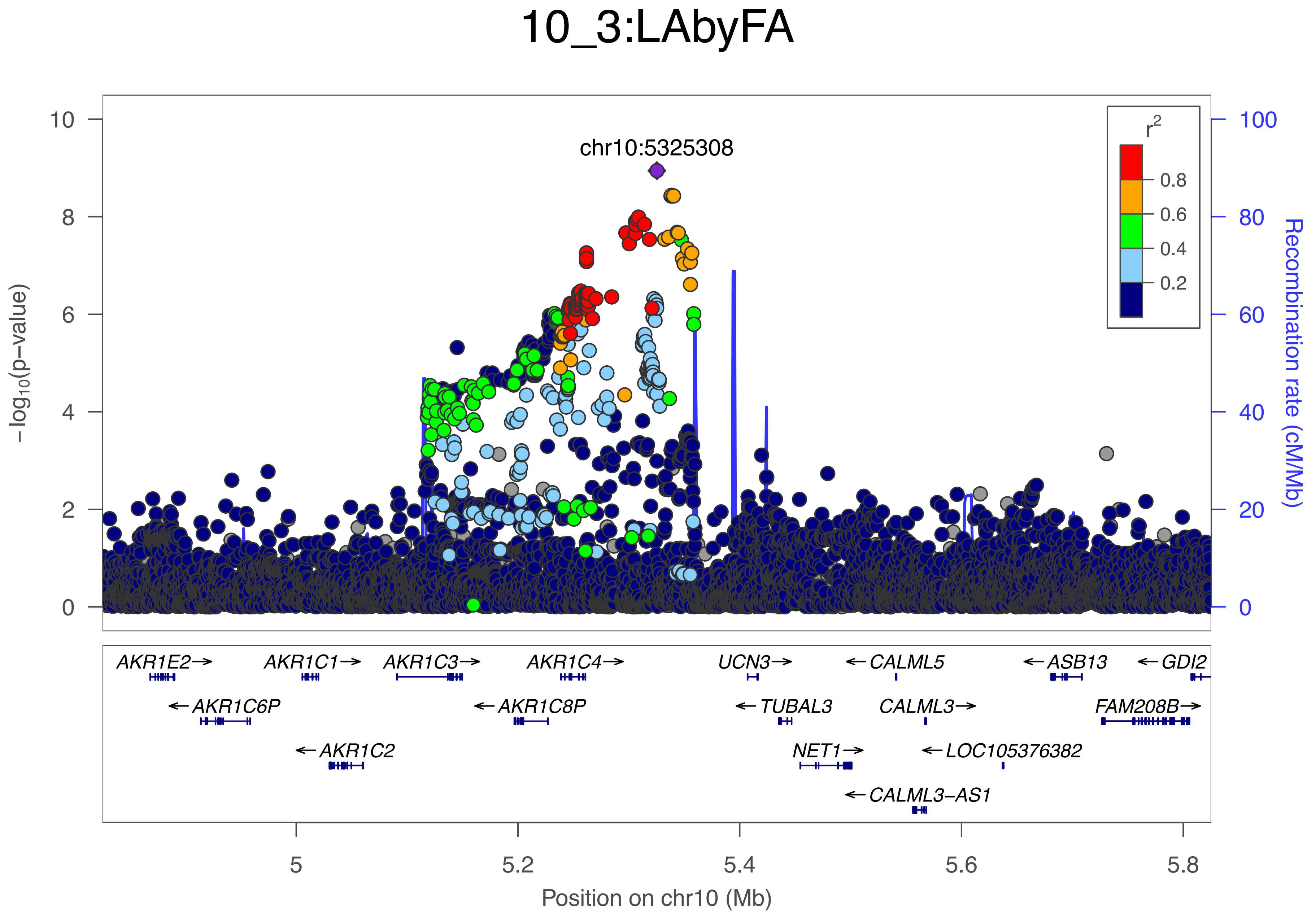

10_3:LAbyFA
chr10:5325308
r2
0.8
0.6
0.4
0.2
−log10(p−value)
Recombination rate (cM/Mb)
10
8
6
4
2
0
100
80
60
40
20
0
AKR1E2→
AKR1C1→
AKR1C3→
AKR1C4→
UCN3→
←CALML5
←ASB13
←GDI2
←AKR1C6P
←AKR1C8P
←TUBAL3
CALML3→
FAM208B→
←AKR1C2
NET1→
←LOC105376382
←CALML3−AS1
5
5.2
5.4
5.6
5.8
Position on chr10 (Mb)

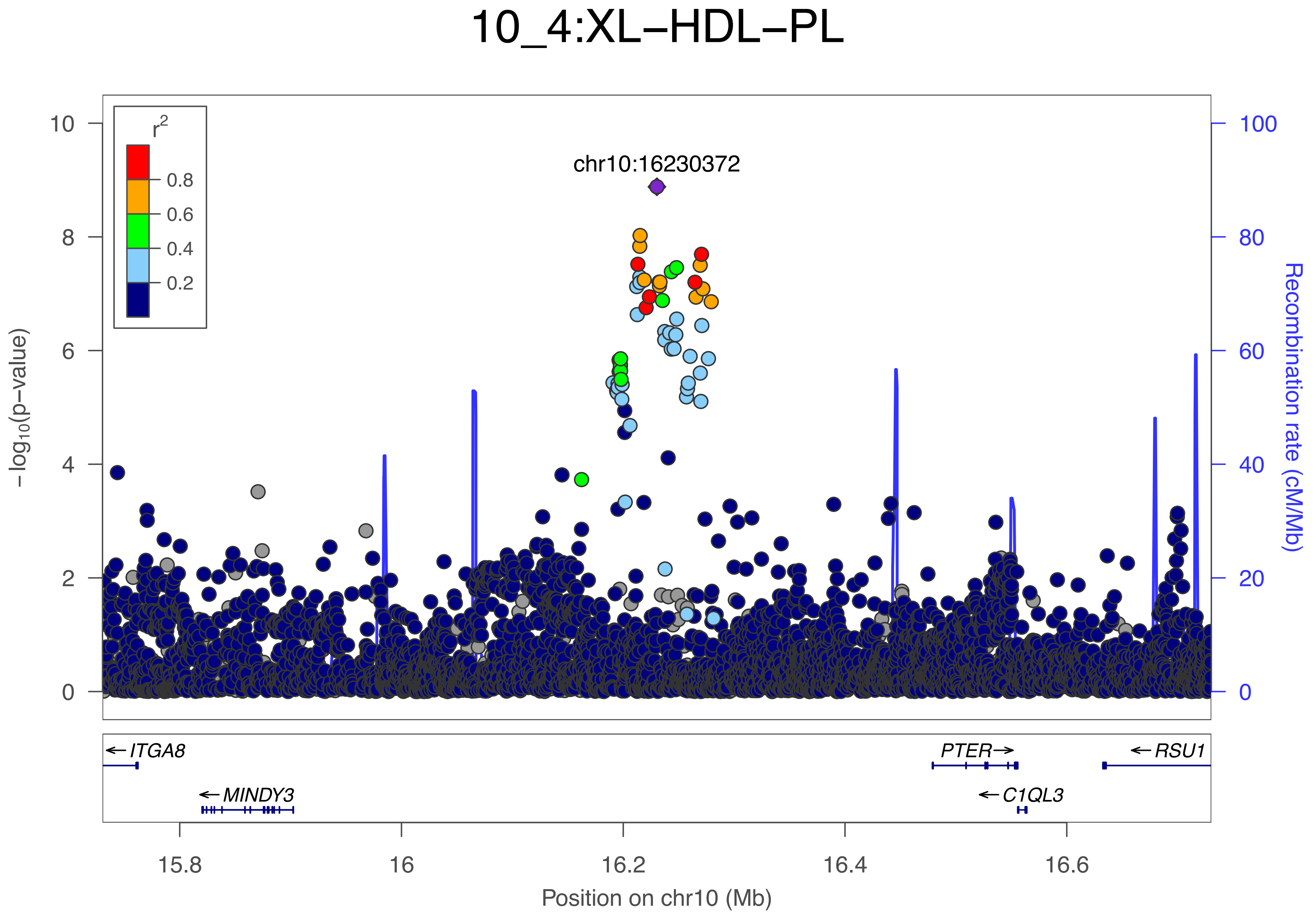

10_4:XL−HDL−PL
chr10:16230372
r2
0.8
0.6
0.4
0.2
−log10(p−value)
Recombination rate (cM/Mb)
ITGA8
MINDY3
PTER
C1QL3
RSU1
15.8
16
16.2
16.4
16.6
Position on chr10 (Mb)

### 10_5:ApoA1

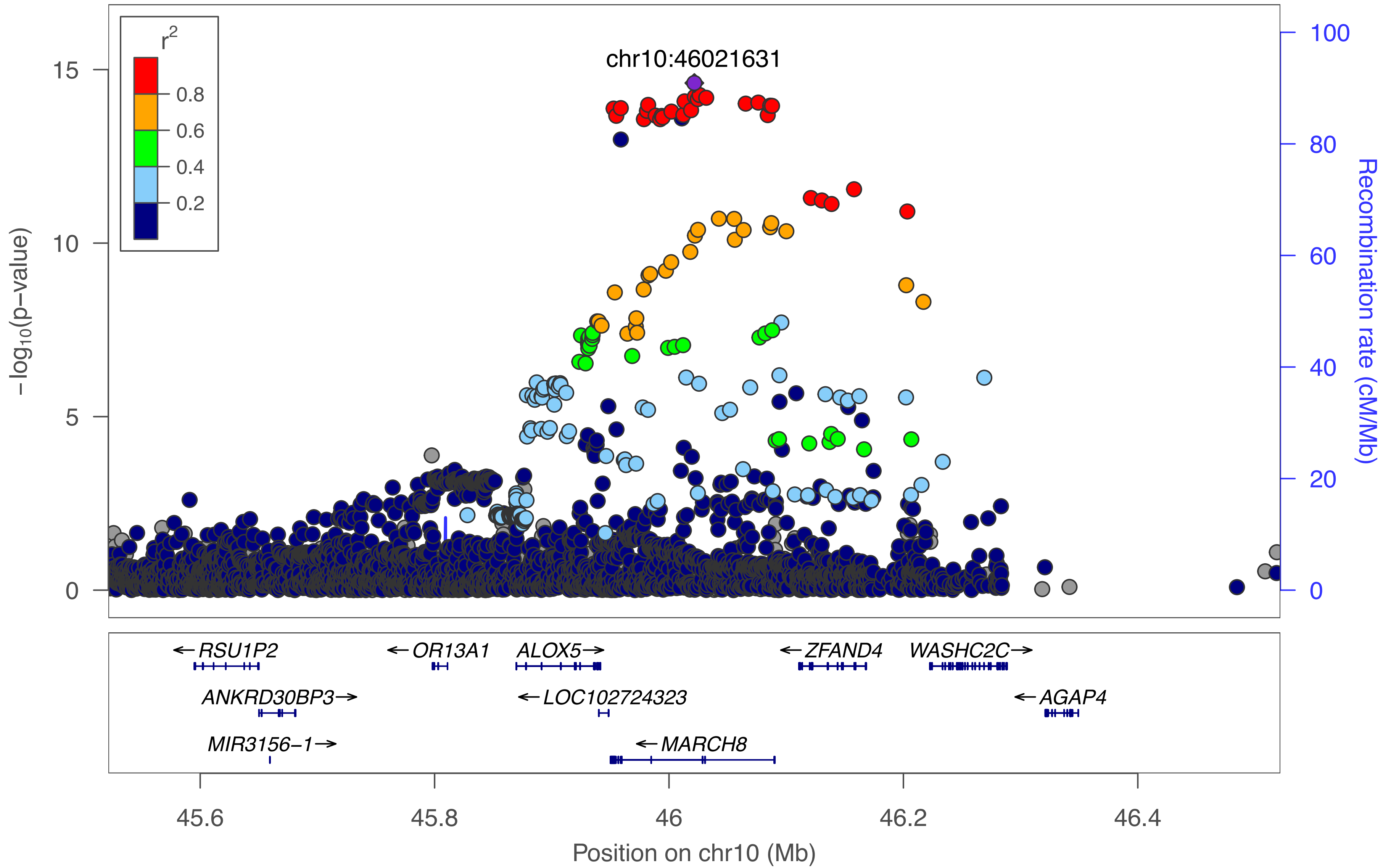

r2
0.8
0.6
0.4
0.2
chr10:46021631
-log10(p-value)
15
10
5
0
100
80
60
40
20
0
Recombination rate (cM/Mb)
RSU1P2
OR13A1
ALOX5
ZFAND4
WASHC2C
ANKRD30BP3
LOC102724323
AGAP4
MIR3156-1
MARCH8
45.6
45.8
46
46.2
46.4
Position on chr10 (Mb)

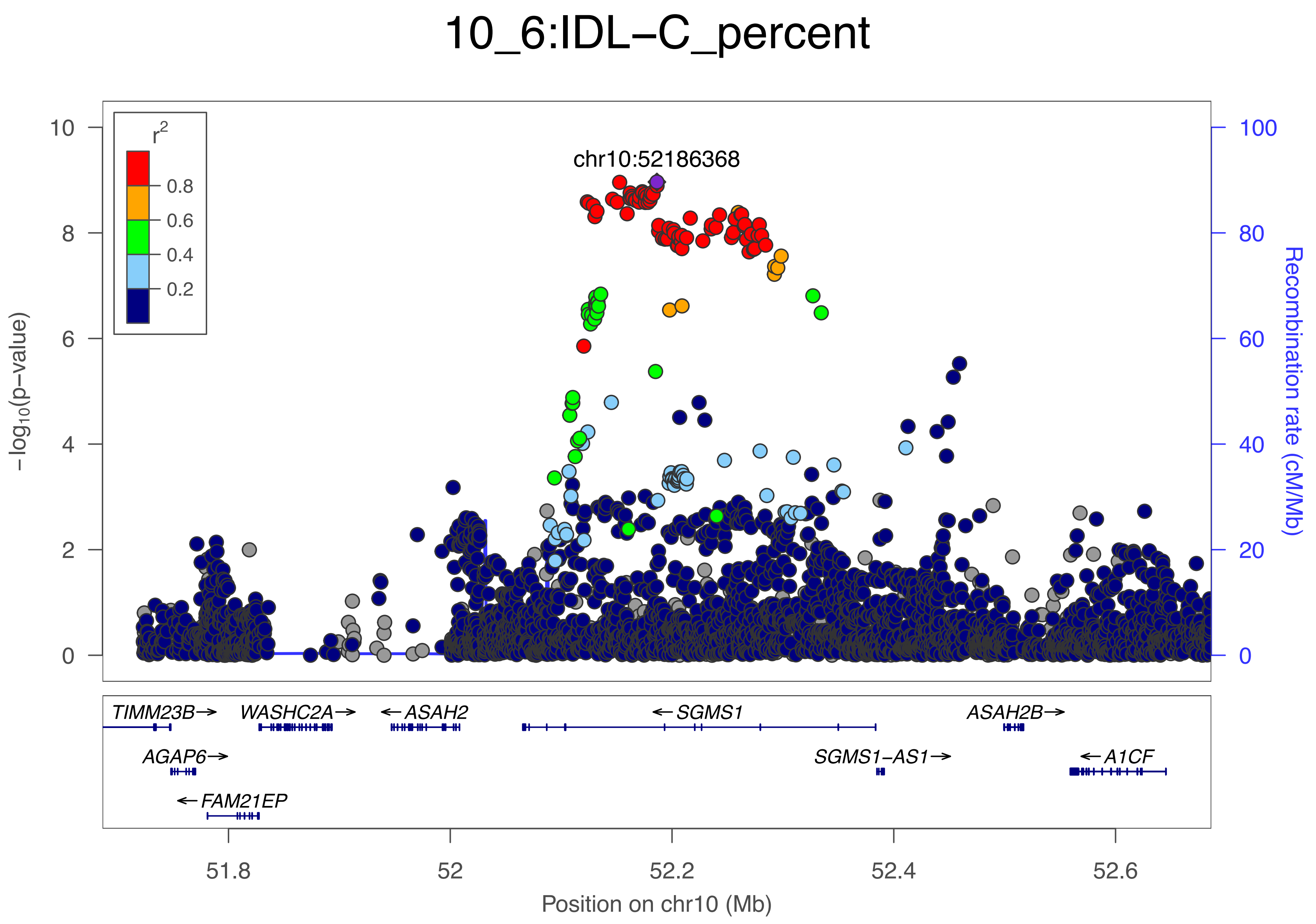

10_6:IDL−C_percent
chr10:52186368
r²
0.8
0.6
0.4
0.2
−log10(p−value)
Recombination rate (cM/Mb)
TIMM23B→
WASHC2A→
←ASAH2
←SGMS1
ASAH2B→
AGAP6→
SGMS1−AS1→
←A1CF
←FAM21EP
51.8
52
52.2
52.4
52.6
Position on chr10 (Mb)

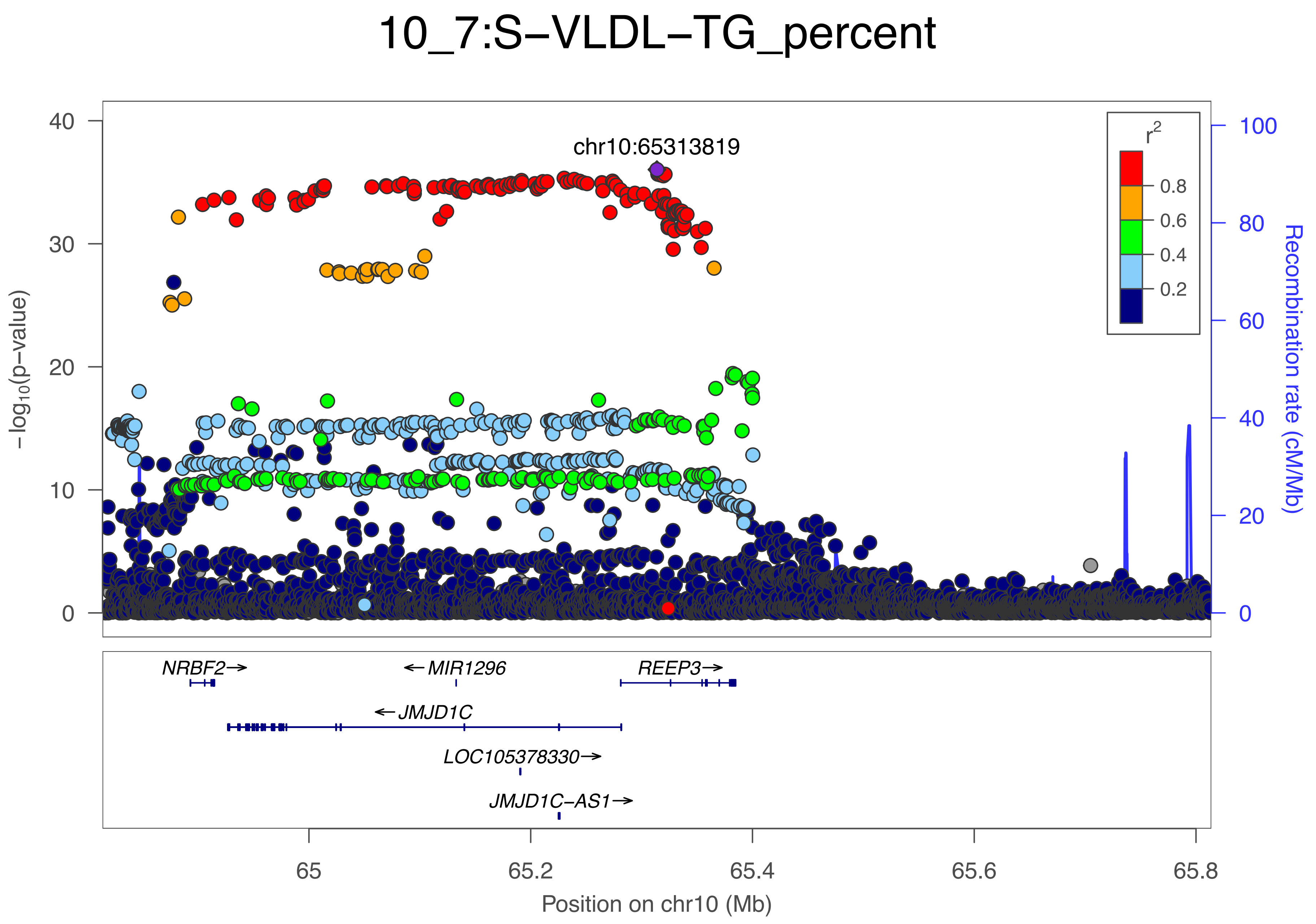

10_7:S−VLDL−TG_percent
chr10:65313819
r²
0.8
0.6
0.4
0.2
−log10(p−value)
Recombination rate (cM/Mb)
40
30
20
10
0
100
80
60
40
20
0
NRBF2→
←MIR1296
REEP3→
←JMJD1C
LOC105378330→
JMJD1C−AS1→
65
65.2
65.4
65.6
65.8
Position on chr10 (Mb)

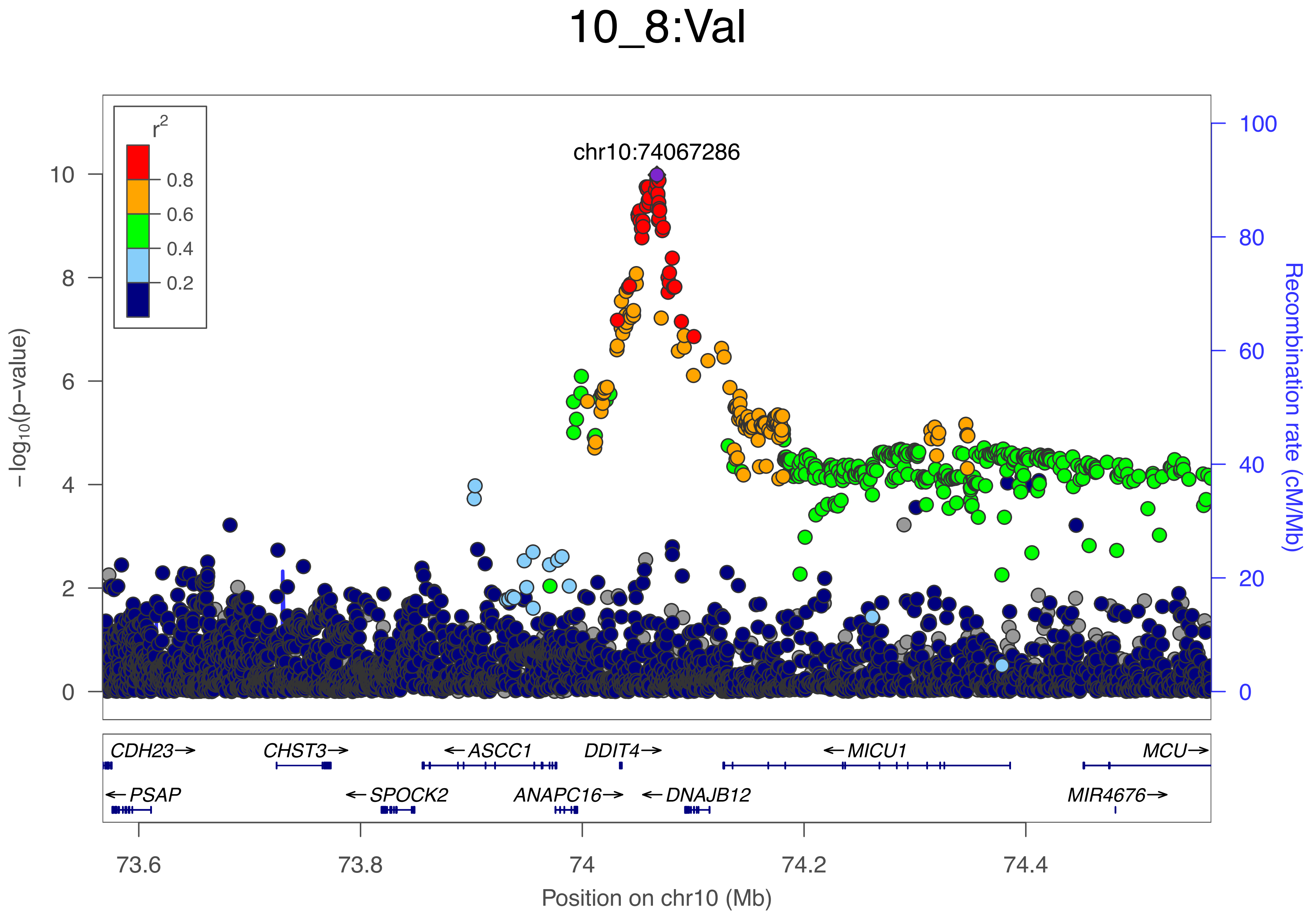

10_8:Val
r2
0.8
0.6
0.4
0.2
chr10:74067286
−log10(p−value)
Recombination rate (cM/Mb)
10
8
6
4
2
0
100
80
60
40
20
0
CDH23→
CHST3→
←ASCC1
DDIT4→
←MICU1
MCU→
←PSAP
←SPOCK2
ANAPC16→
←DNAJB12
MIR4676→
73.6
73.8
74
74.2
74.4
Position on chr10 (Mb)

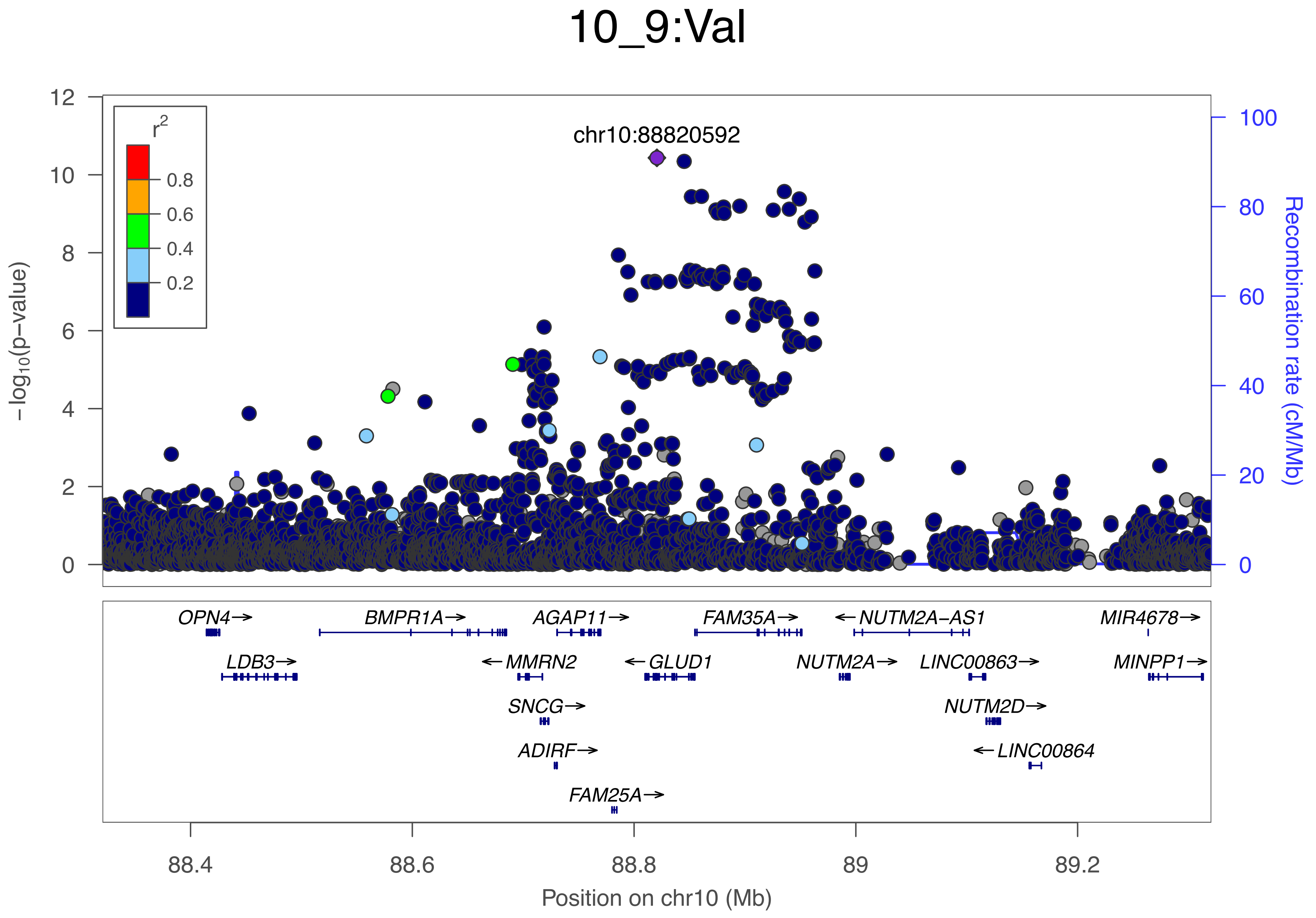

10_9:Val
chr10:88820592
r²
0.8
0.6
0.4
0.2
−log₁₀(p−value)
Recombination rate (cM/Mb)
OPN4→
BMPR1A→
AGAP11→
FAM35A→
←NUTM2A−AS1
MIR4678→
LDB3→
←MMRN2
←GLUD1
NUTM2A→
LINC00863→
MINPP1→
SNCG→
NUTM2D→
ADIRF→
←LINC00864
FAM25A→
88.4
88.6
88.8
89
89.2
Position on chr10 (Mb)

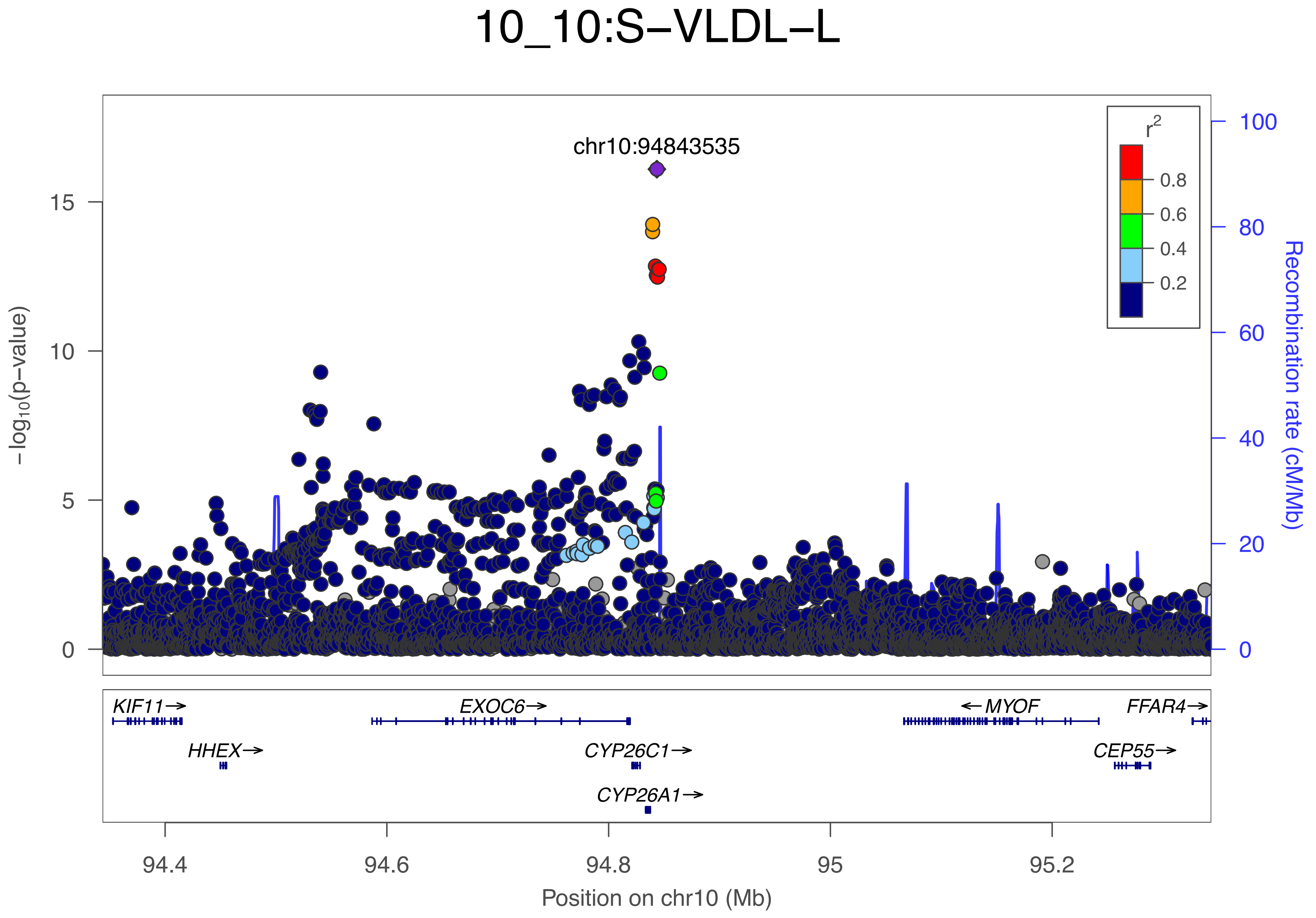
10_10:S−VLDL−L
chr10:94843535
−log10(p−value)
Recombination rate (cM/Mb)
r2
0.8
0.6
0.4
0.2
0
5
10
15
20
40
60
80
100
KIF11→
HHEX→
EXOC6→
CYP26C1→
CYP26A1→
←MYOF
FFAR4→
CEP55→
94.4
94.6
94.8
95
95.2
Position on chr10 (Mb)

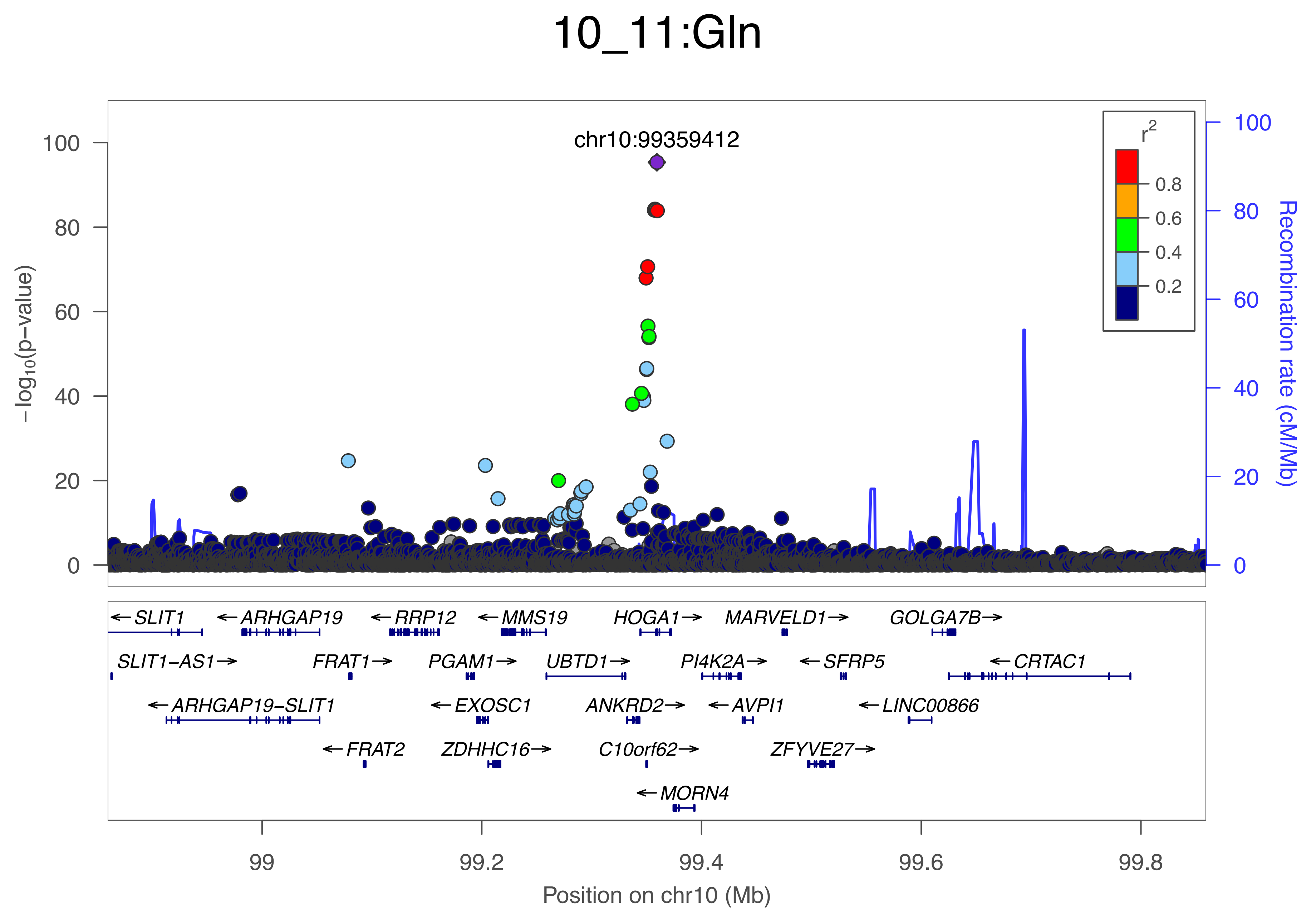

10_11:Gln
chr10:99359412
r2
0.8
0.6
0.4
0.2
−log10(p−value)
Recombination rate (cM/Mb)
100
80
60
40
20
0
SLIT1
ARHGAP19
RRP12
MMS19
HOGA1
MARVELD1
GOLGA7B
SLIT1−AS1
FRAT1
PGAM1
UBTD1
PI4K2A
SFRP5
CRTAC1
ARHGAP19−SLIT1
EXOSC1
ANKRD2
AVPI1
LINC00866
FRAT2
ZDHHC16
C10orf62
ZFYVE27
MORN4
99
99.2
99.4
99.6
99.8
Position on chr10 (Mb)

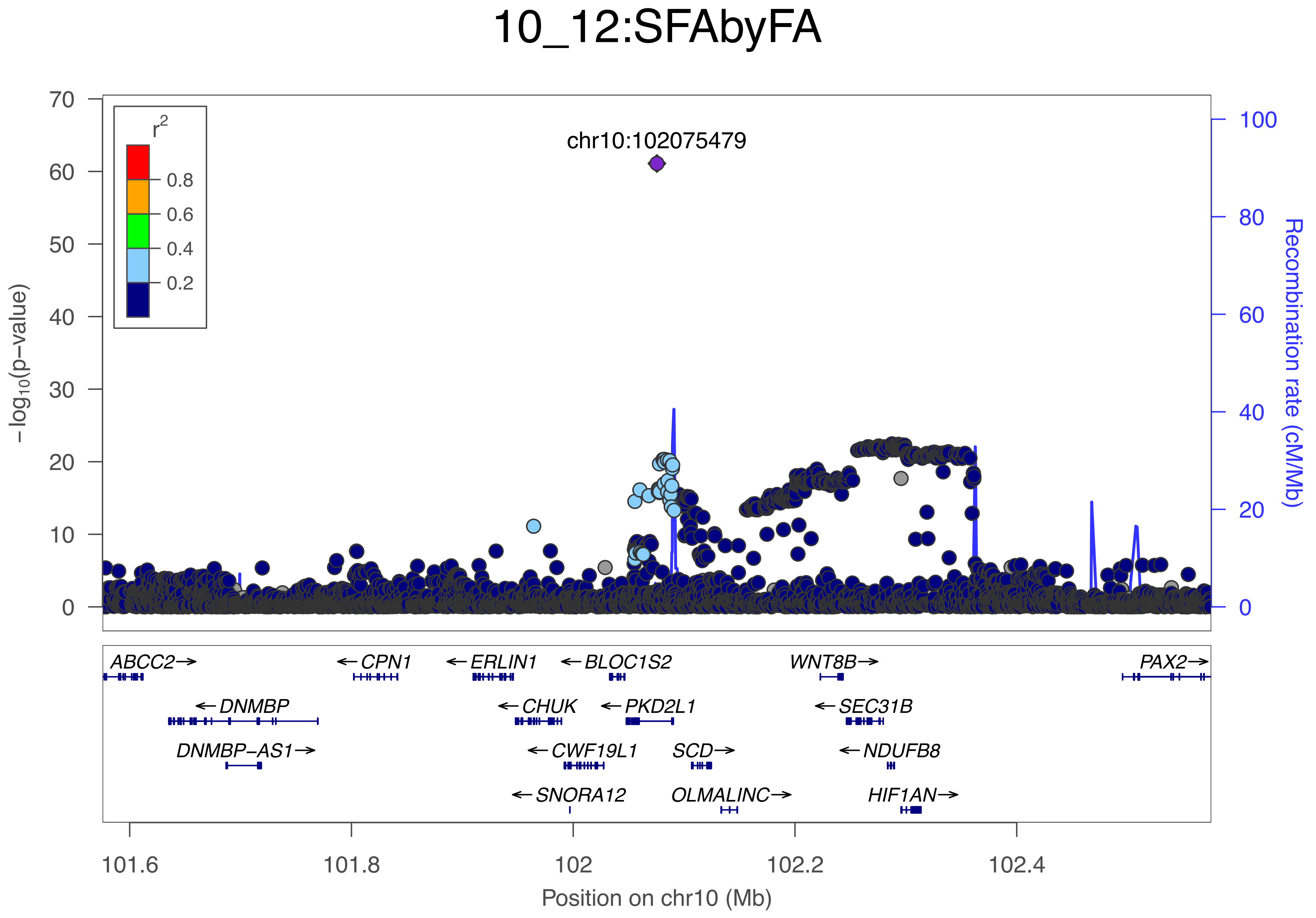
10_12:SFAbyFA
chr10:102075479
r2
0.8
0.6
0.4
0.2
−log10(p−value)
Recombination rate (cM/Mb)
0
10
20
30
40
50
60
70
0
20
40
60
80
100
ABCC2→
←CPN1
←ERLIN1
←BLOC1S2
WNT8B→
PAX2→
←DNMBP
←CHUK
←PKD2L1
←SEC31B
DNMBP−AS1→
←CWF19L1
SCD→
←NDUFB8
←SNORA12
OLMALINC→
HIF1AN→
101.6
101.8
102
102.2
102.4
Position on chr10 (Mb)

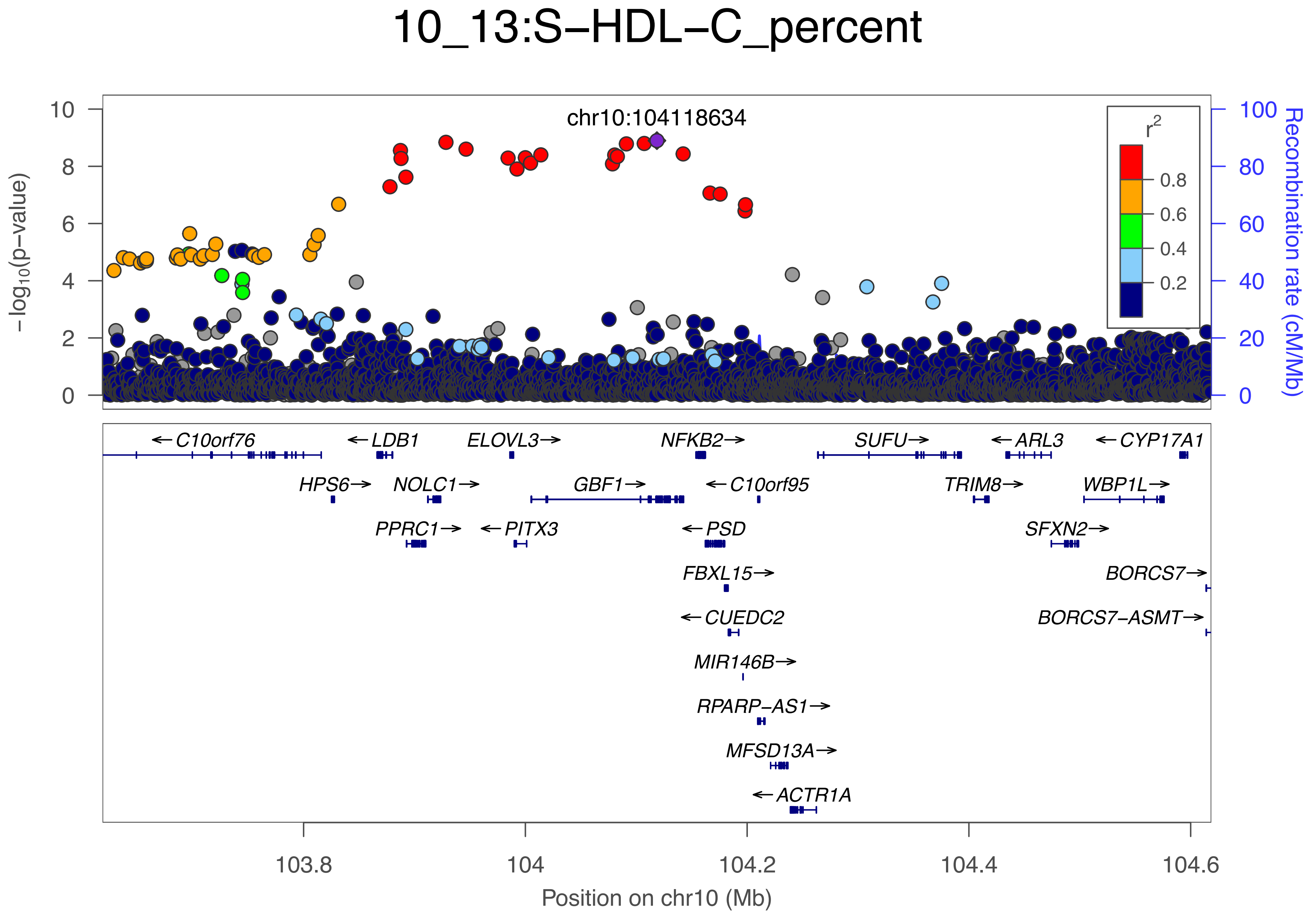
10_13:S−HDL−C_percent
chr10:104118634
−log10(p−value)
Recombination rate (cM/Mb)
r2
0.8
0.6
0.4
0.2
C10orf76
LDB1
ELOVL3
NFKB2
SUFU
ARL3
CYP17A1
HPS6
NOLC1
GBF1
C10orf95
TRIM8
WBP1L
PPRC1
PITX3
PSD
SFXN2
FBXL15
BORCS7
CUEDC2
BORCS7−ASMT
MIR146B
RPARP−AS1
MFSD13A
ACTR1A
103.8
104
104.2
104.4
104.6
Position on chr10 (Mb)

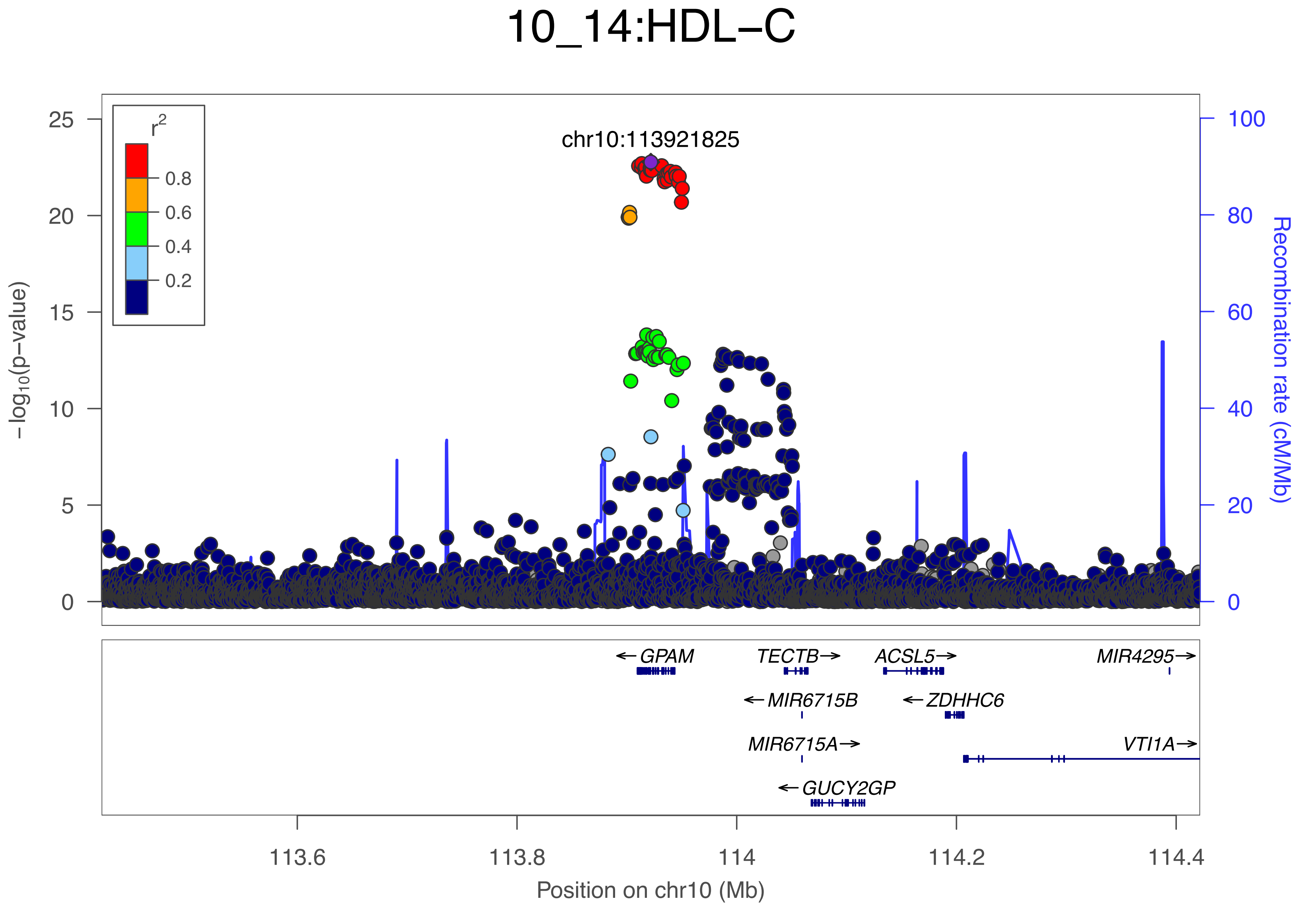
10_14:HDL−C
chr10:113921825
r2
0.8
0.6
0.4
0.2
−log10(p−value)
Recombination rate (cM/Mb)
GPAM
TECTB
ACSL5
MIR4295
MIR6715B
ZDHHC6
MIR6715A
VTI1A
GUCY2GP
113.6
113.8
114
114.2
114.4
Position on chr10 (Mb)

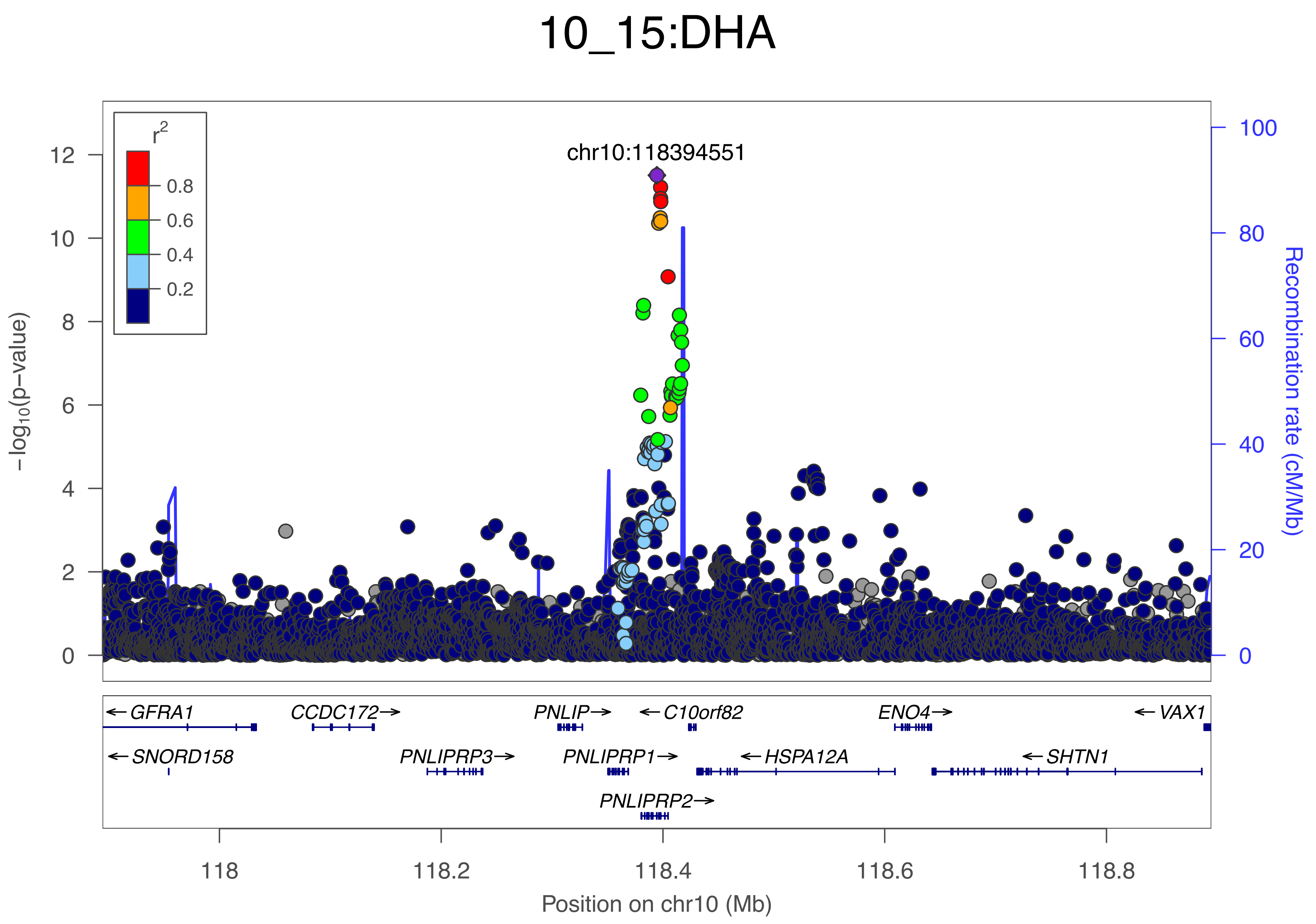

10_15:DHA
chr10:118394551
r2
0.8
0.6
0.4
0.2
-log10(p-value)
Recombination rate (cM/Mb)
GFRA1
CCDC172
PNLIP
C10orf82
ENO4
VAX1
SNORD158
PNLIPRP3
PNLIPRP1
HSPA12A
SHTN1
PNLIPRP2
Position on chr10 (Mb)

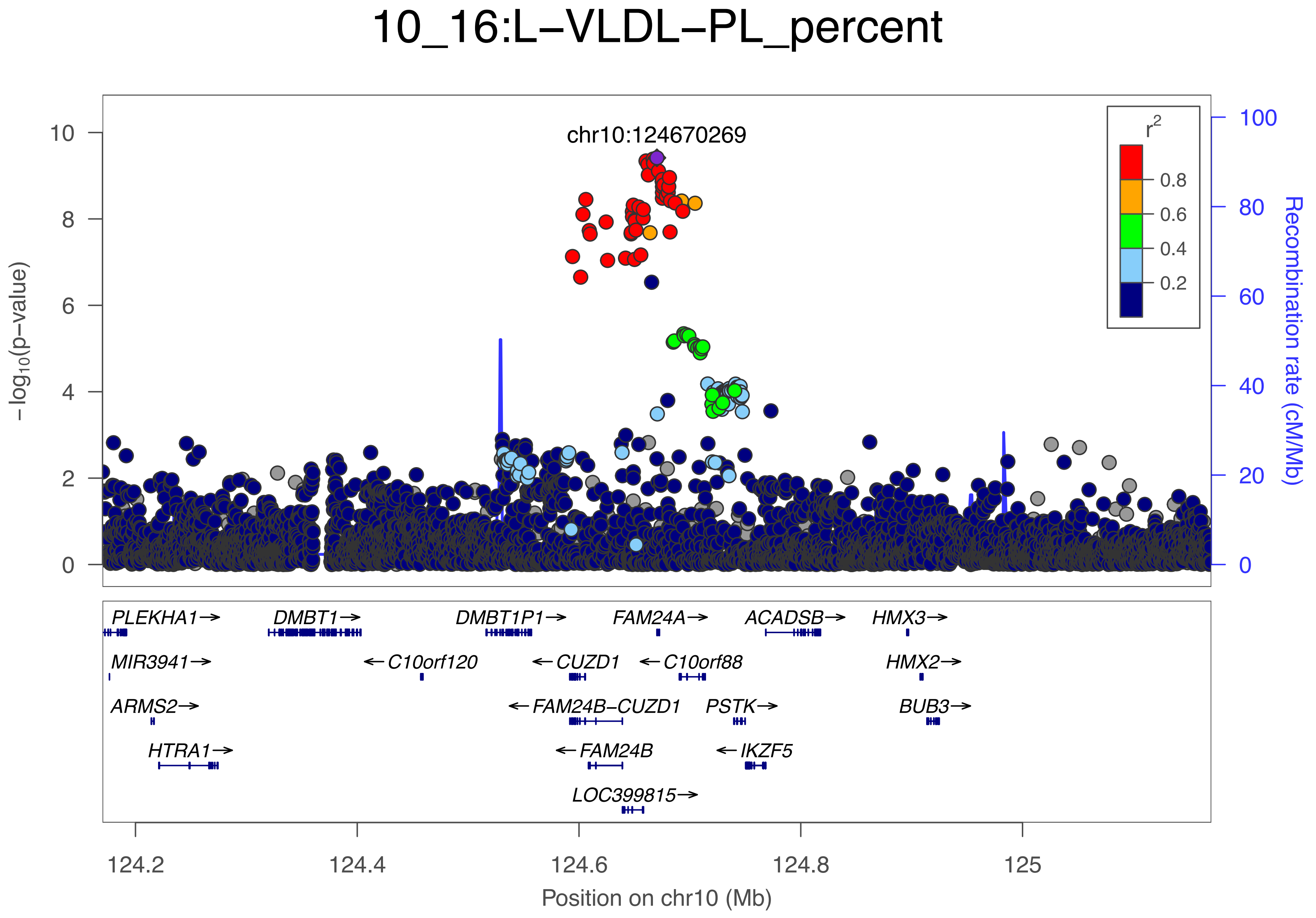
10_16:L−VLDL−PL_percent
chr10:124670269
r2
0.8
0.6
0.4
0.2
−log10(p−value)
Recombination rate (cM/Mb)
PLEKHA1→
DMBT1→
DMBT1P1→
FAM24A→
ACADSB→
HMX3→
MIR3941→
←C10orf120
←CUZD1
←C10orf88
HMX2→
ARMS2→
←FAM24B−CUZD1
PSTK→
BUB3→
HTRA1→
←FAM24B
←IKZF5
LOC399815→
124.2
124.4
124.6
124.8
125
Position on chr10 (Mb)

### 10_17:AcAce

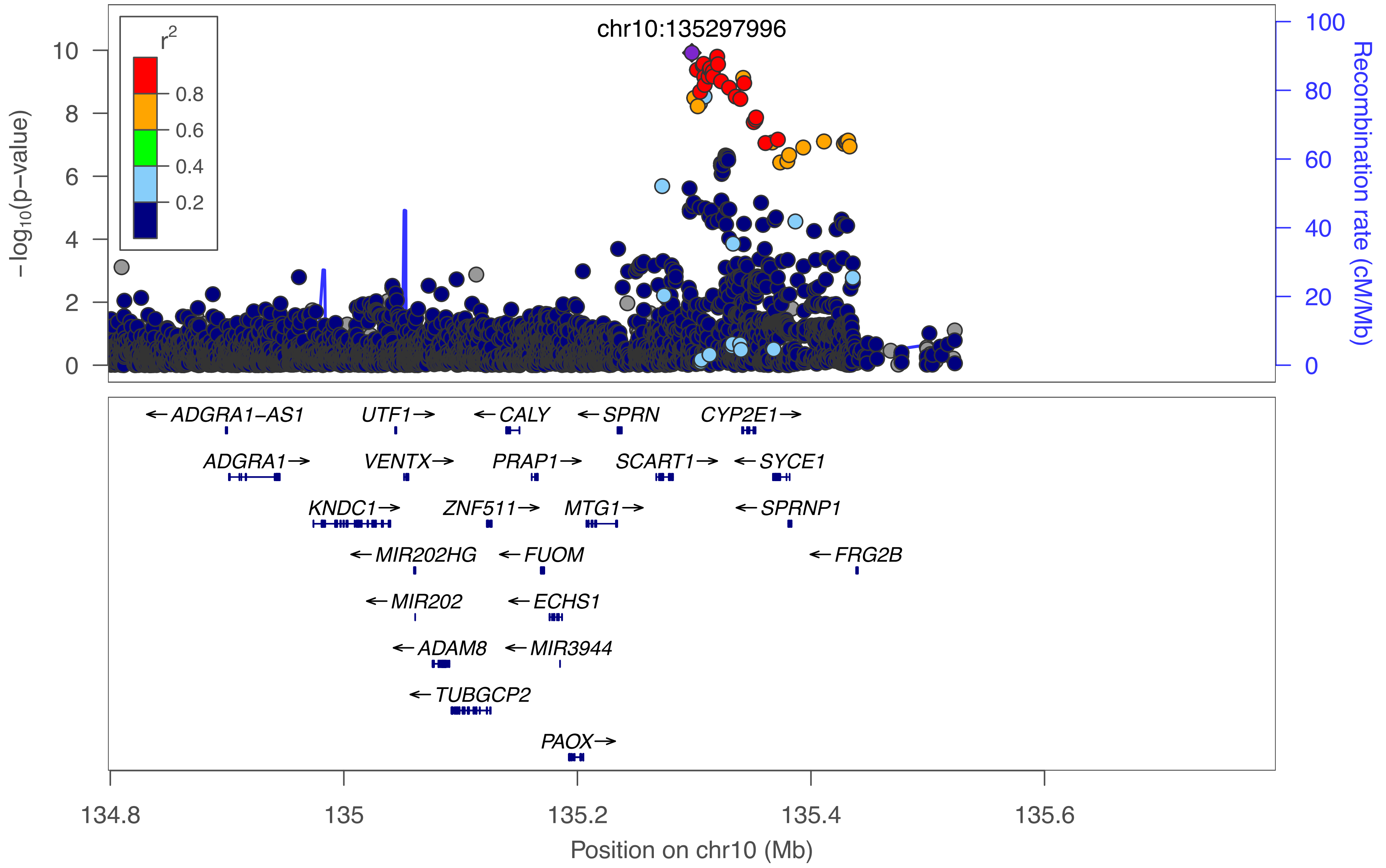

chr10:135297996
r²
0.8
0.6
0.4
0.2
−log₁₀(p−value)
Recombination rate (cM/Mb)
ADGRA1−AS1
UTF1
CALY
SPRN
CYP2E1
ADGRA1
VENTX
PRAP1
SCART1
SYCE1
KNDC1
ZNF511
MTG1
SPRNP1
MIR202HG
FUOM
FRG2B
MIR202
ECHS1
ADAM8
MIR3944
TUBGCP2
PAOX
134.8
135
135.2
135.4
135.6
Position on chr10 (Mb)

### 11_1:Glc

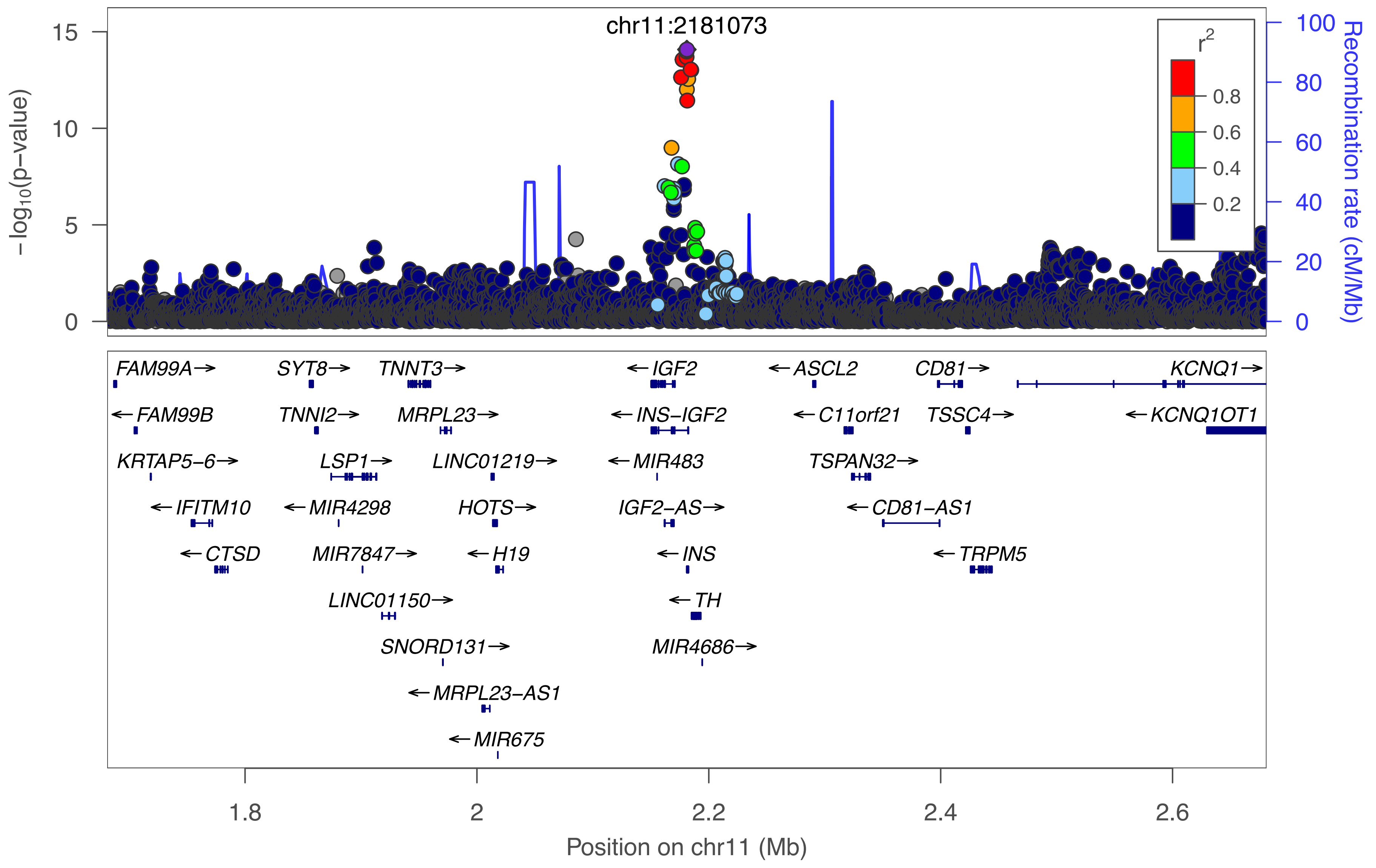

chr11:2181073
−log10(p−value)
Recombination rate (cM/Mb)
r2
0.8
0.6
0.4
0.2
FAM99A→
SYT8→
TNNT3→
←IGF2
←ASCL2
CD81→
KCNQ1→
←FAM99B
TNNI2→
MRPL23→
←INS−IGF2
←C11orf21
TSSC4→
←KCNQ1OT1
KRTAP5−6→
LSP1→
LINC01219→
←MIR483
TSPAN32→
←IFITM10
←MIR4298
HOTS→
IGF2−AS→
←CD81−AS1
←CTSD
MIR7847→
←H19
←INS
←TRPM5
LINC01150→
←TH
SNORD131→
MIR4686→
←MRPL23−AS1
←MIR675
1.8
2
2.2
2.4
2.6
Position on chr11 (Mb)

### 11_2:S−LDL−PL_percent

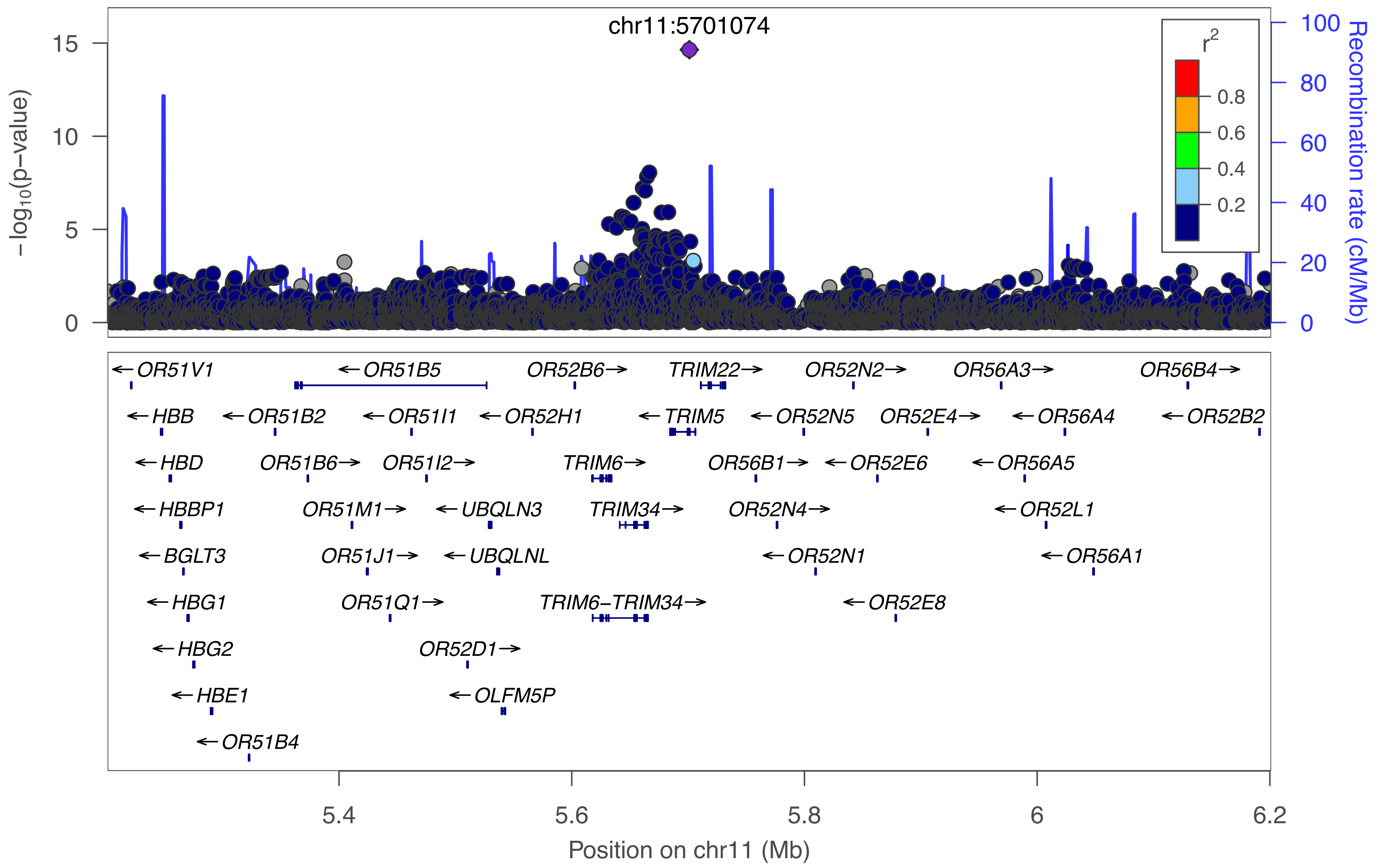

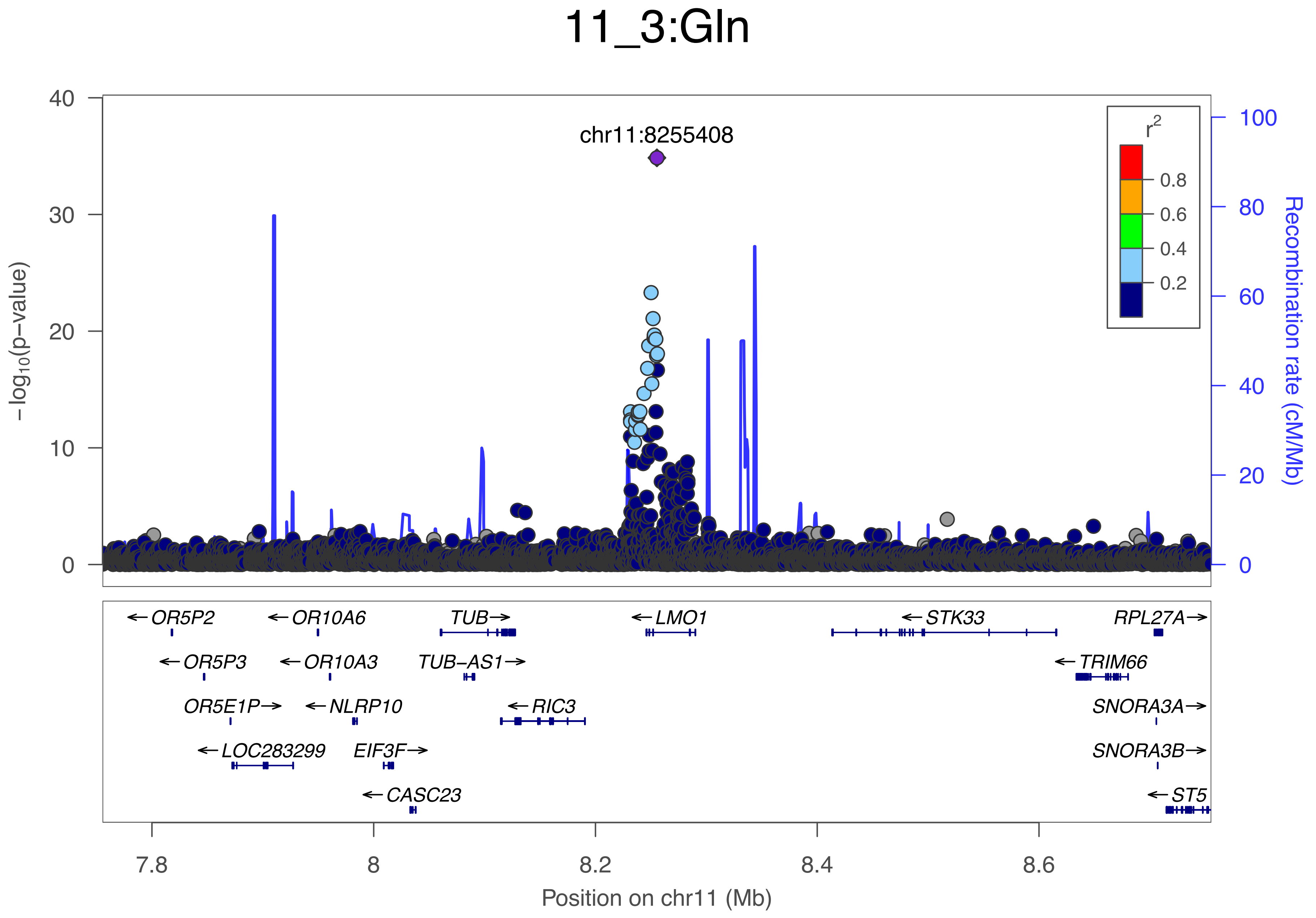
11_3:Gln
chr11:8255408
r2
0.8
0.6
0.4
0.2
−log10(p−value)
Recombination rate (cM/Mb)
0
10
20
30
40
0
20
40
60
80
100
OR5P2
OR10A6
TUB
LMO1
STK33
RPL27A
OR5P3
OR10A3
TUB−AS1
TRIM66
OR5E1P
NLRP10
RIC3
SNORA3A
LOC283299
EIF3F
SNORA3B
CASC23
ST5
7.8
8
8.2
8.4
8.6
Position on chr11 (Mb)

### 11_4:S−VLDL−TG

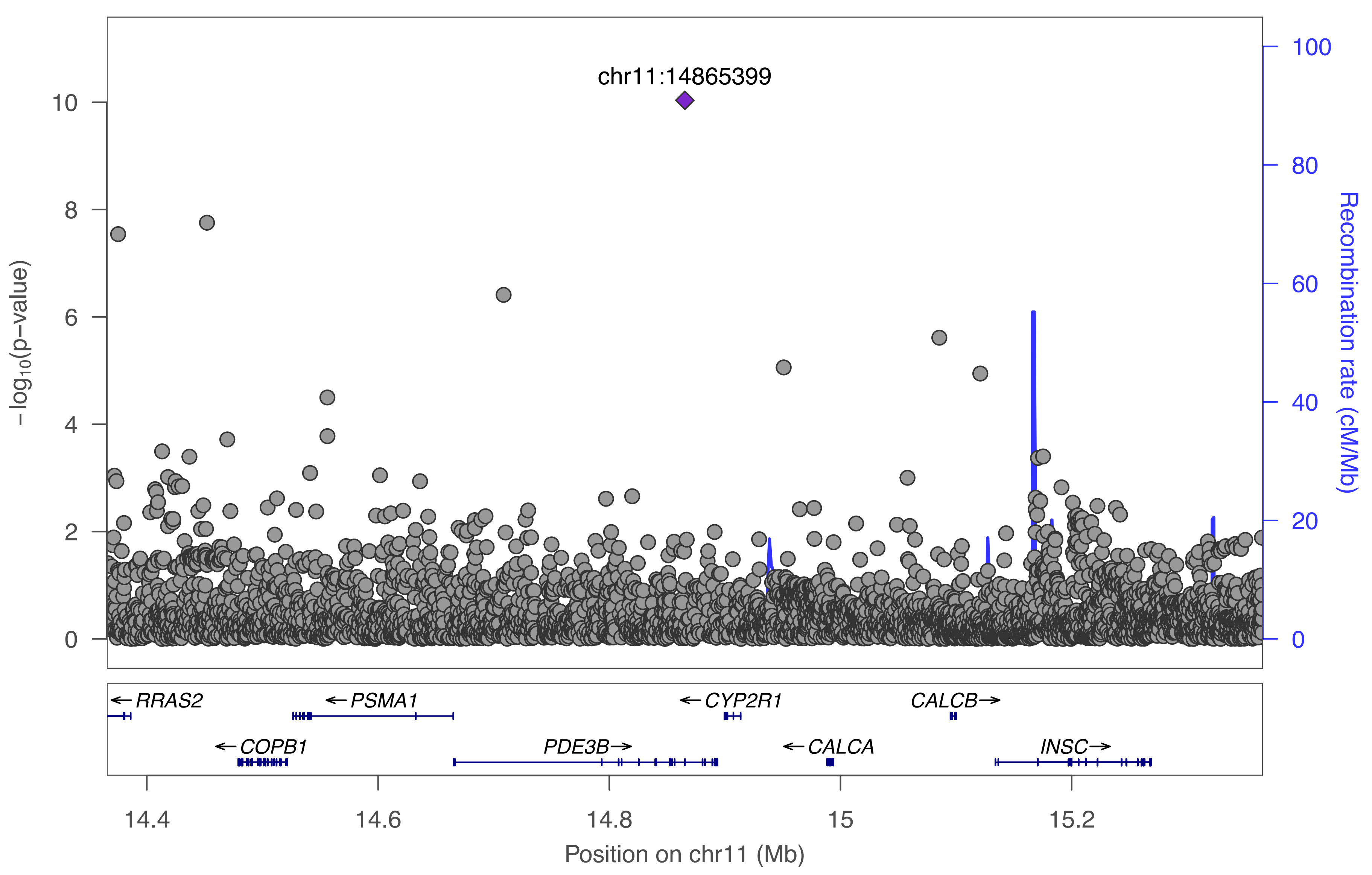

chr11:14865399
−log10(p−value)
Recombination rate (cM/Mb)
0
2
4
6
8
10
0
20
40
60
80
100
RRAS2
PSMA1
CYP2R1
CALCB
COPB1
PDE3B
CALCA
INSC
14.4
14.6
14.8
15
15.2
Position on chr11 (Mb)

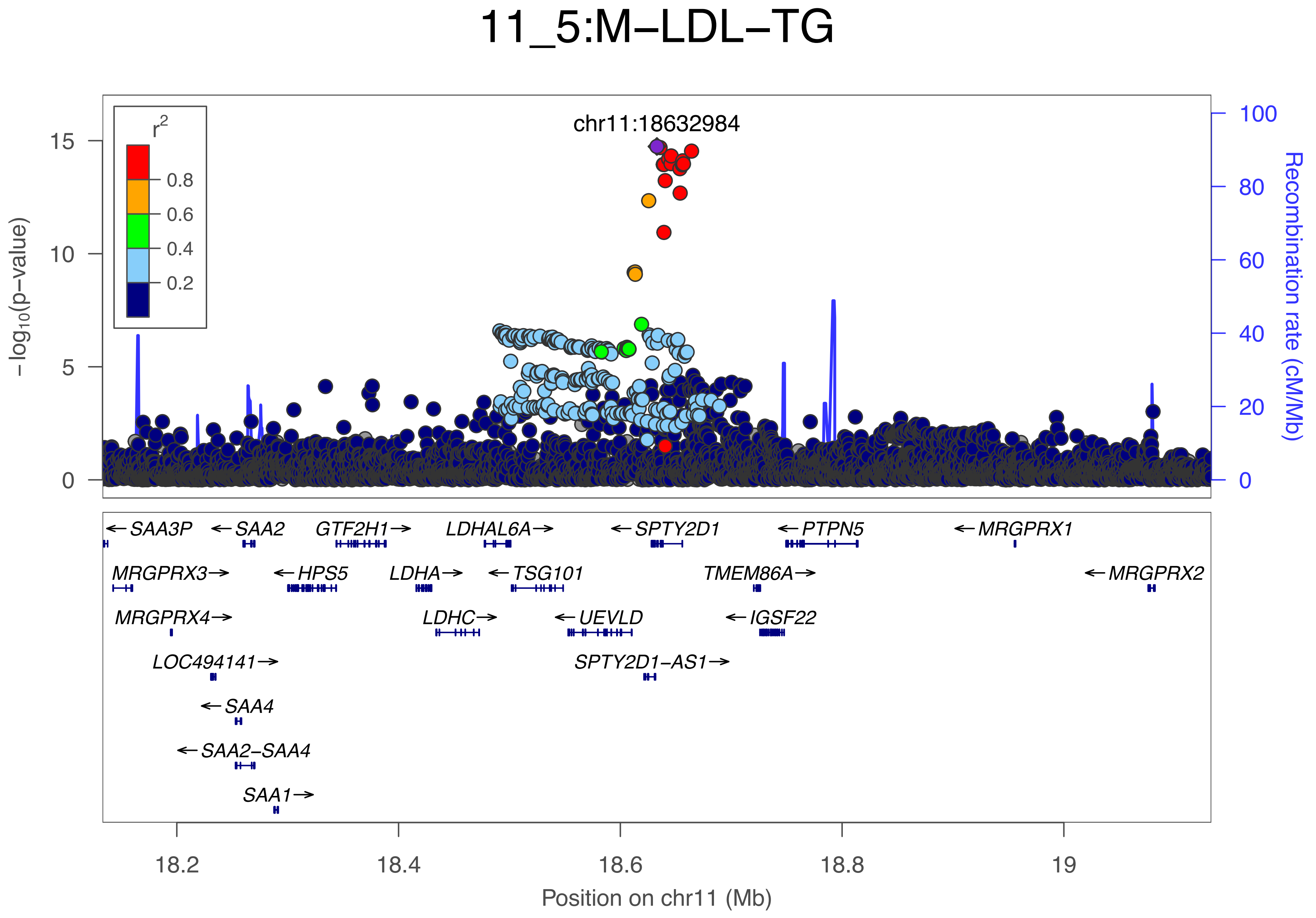
11_5:M–LDL–TG
chr11:18632984
r²
0.8
0.6
0.4
0.2
–log₁₀(p–value)
15
10
5
0
100
80
60
40
20
0
Recombination rate (cM/Mb)
←SAA3P
←SAA2
GTF2H1→
LDHAL6A→
←SPTY2D1
←PTPN5
←MRGPRX1
MRGPRX3→
←HPS5
LDHA→
←TSG101
TMEM86A→
←MRGPRX2
MRGPRX4→
LDHC→
←UEVLD
←IGSF22
LOC494141→
SPTY2D1–AS1→
←SAA4
←SAA2–SAA4
SAA1→
18.2
18.4
18.6
18.8
19
Position on chr11 (Mb)

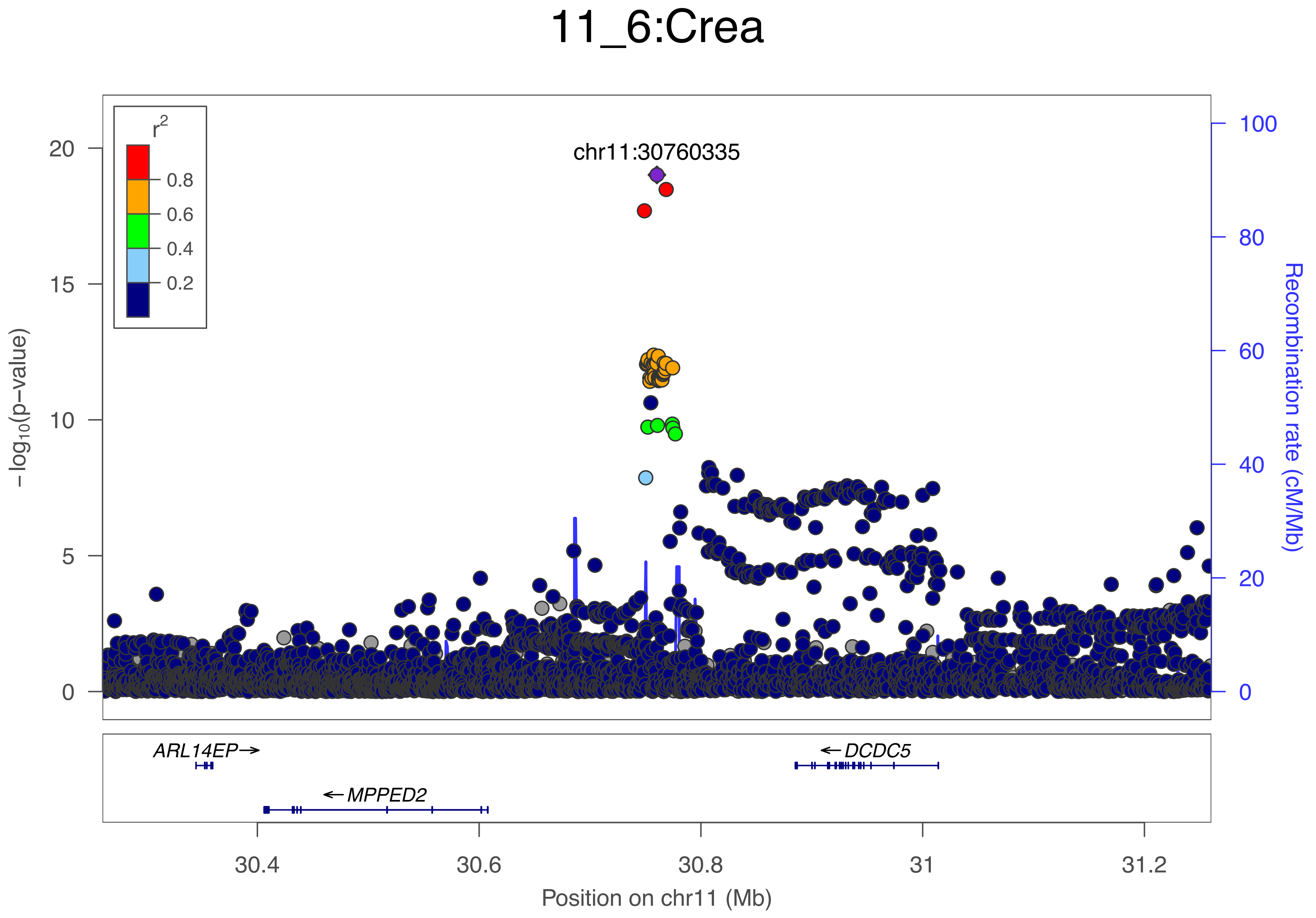

11_6:Crea
r2
0.8
0.6
0.4
0.2
chr11:30760335
20
15
10
5
0
−log10(p−value)
100
80
60
40
20
0
Recombination rate (cM/Mb)
ARL14EP→
←DCDC5
←MPPED2
30.4
30.6
30.8
31
31.2
Position on chr11 (Mb)

### 11_7:Ala

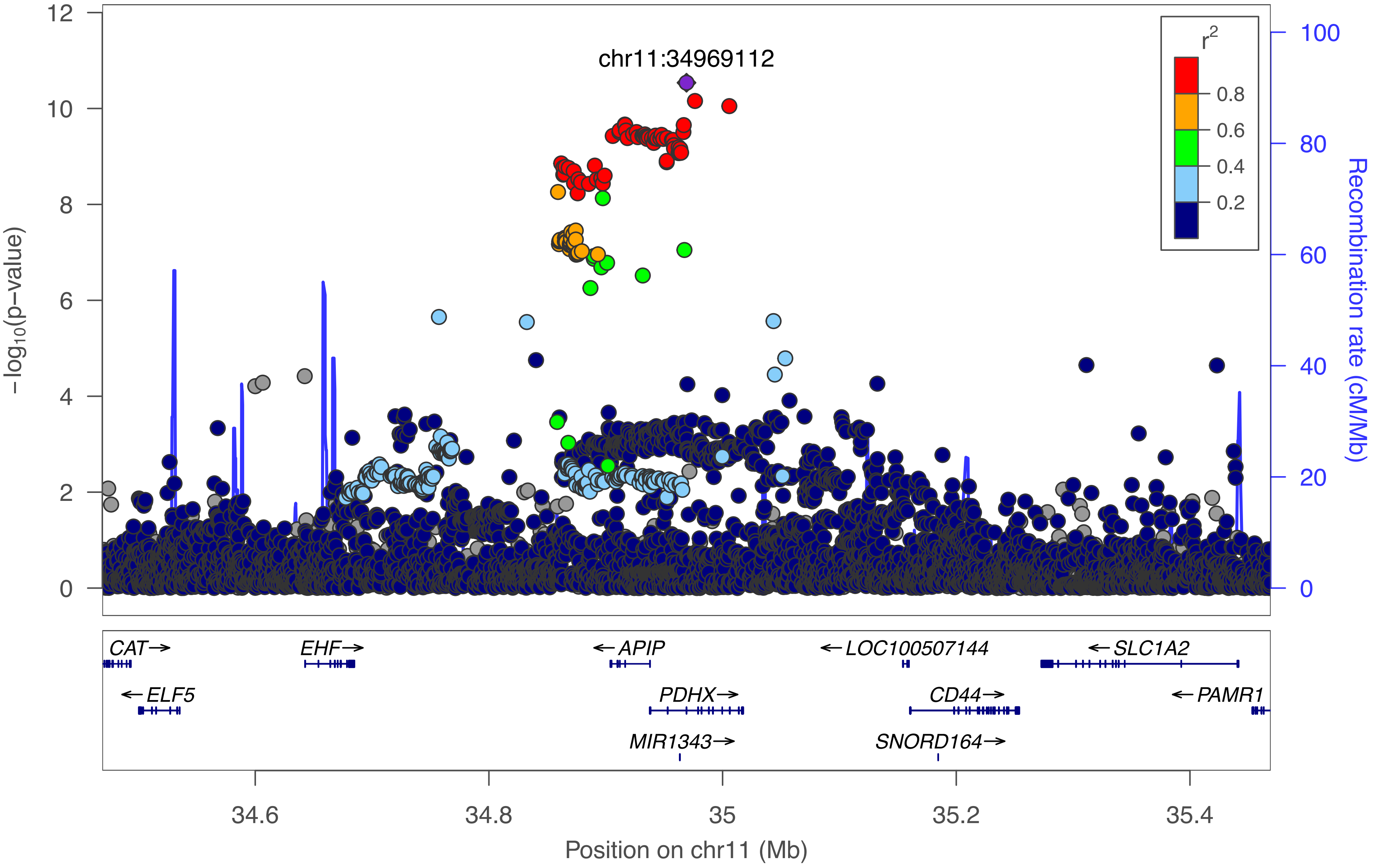
chr11:34969112
r²
0.8
0.6
0.4
0.2
−log10(p−value)
Recombination rate (cM/Mb)
CAT→
EHF→
←APIP
←LOC100507144
←SLC1A2
←ELF5
PDHX→
CD44→
←PAMR1
MIR1343→
SNORD164→
34.6
34.8
35
35.2
35.4
Position on chr11 (Mb)

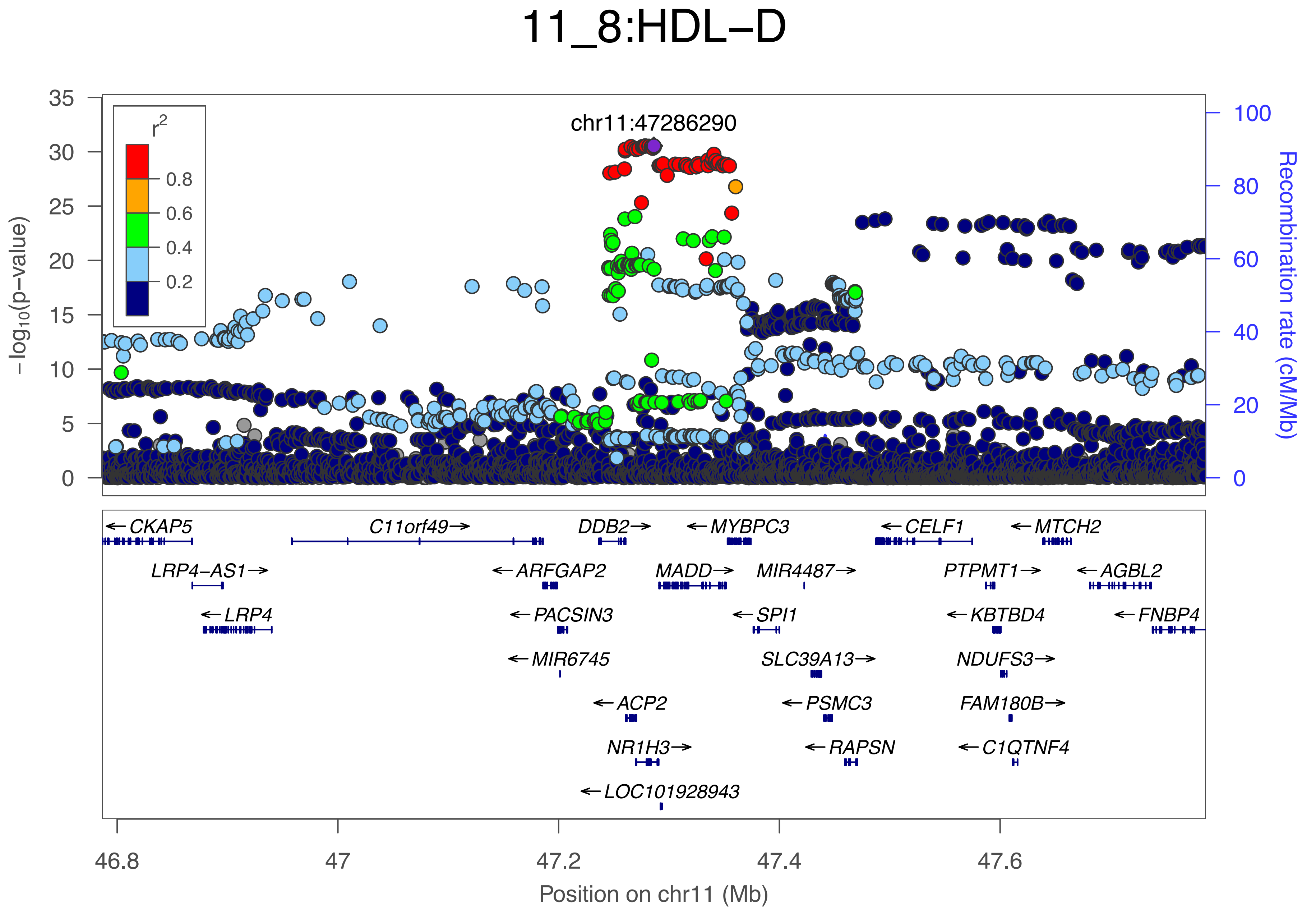

11_8:HDL−D
chr11:47286290
r2
0.8
0.6
0.4
0.2
35
30
25
20
15
10
5
0
−log10(p−value)
100
80
60
40
20
0
Recombination rate (cM/Mb)
←CKAP5
C11orf49→
DDB2→
←MYBPC3
←CELF1
←MTCH2
LRP4−AS1→
←ARFGAP2
MADD→
MIR4487→
PTPMT1→
←AGBL2
←LRP4
←PACSIN3
←SPI1
←KBTBD4
←FNBP4
←MIR6745
SLC39A13→
NDUFS3→
←ACP2
←PSMC3
FAM180B→
NR1H3→
←RAPSN
←C1QTNF4
←LOC101928943
46.8
47
47.2
47.4
47.6
Position on chr11 (Mb)

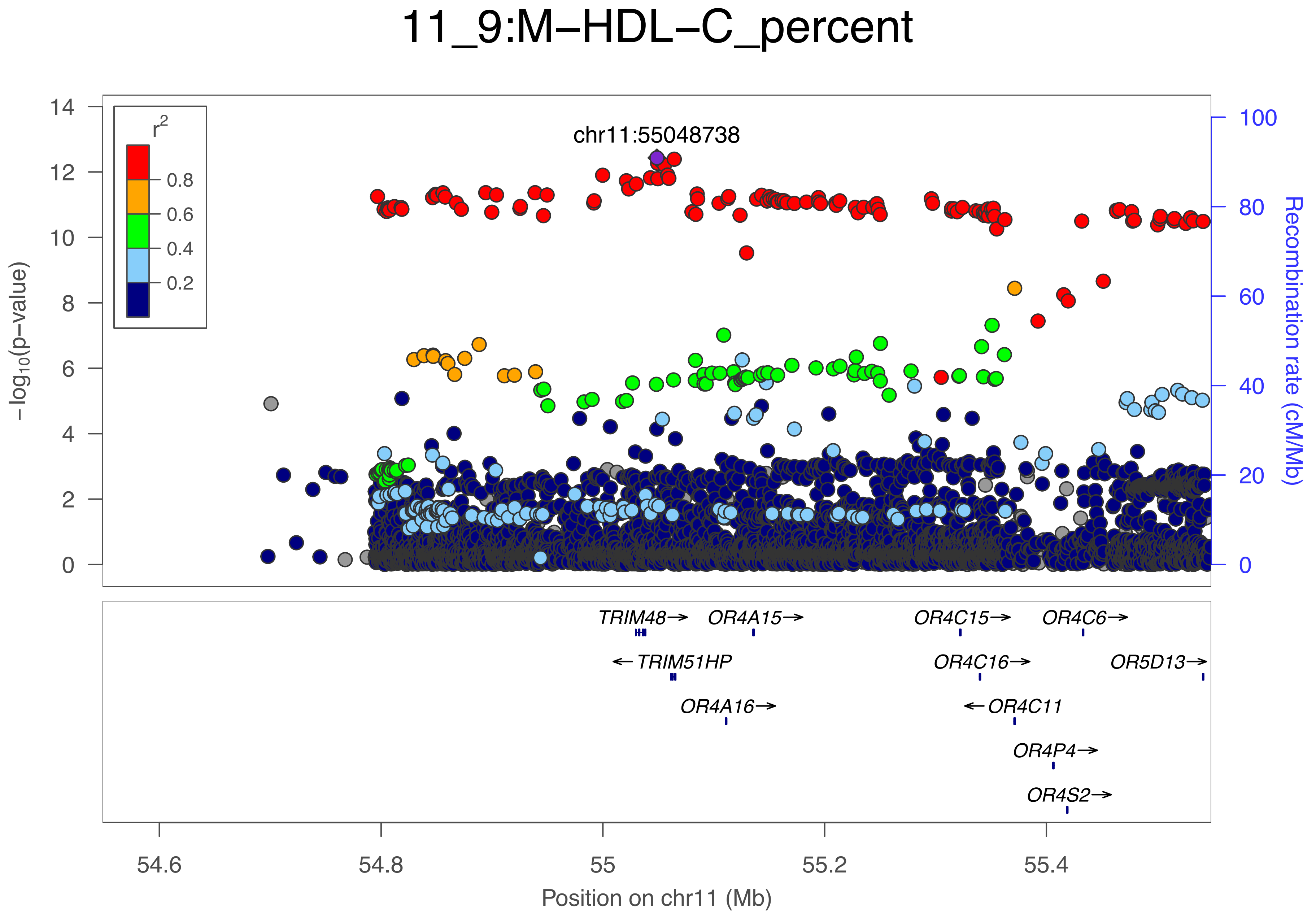

11_9:M−HDL−C_percent
chr11:55048738
r²
0.8
0.6
0.4
0.2
−log10(p−value)
Recombination rate (cM/Mb)
TRIM48→
OR4A15→
OR4C15→
OR4C6→
←TRIM51HP
OR4C16→
OR5D13→
OR4A16→
←OR4C11
OR4P4→
OR4S2→
54.6
54.8
55
55.2
55.4
Position on chr11 (Mb)

### 11_10:UnsatDeg

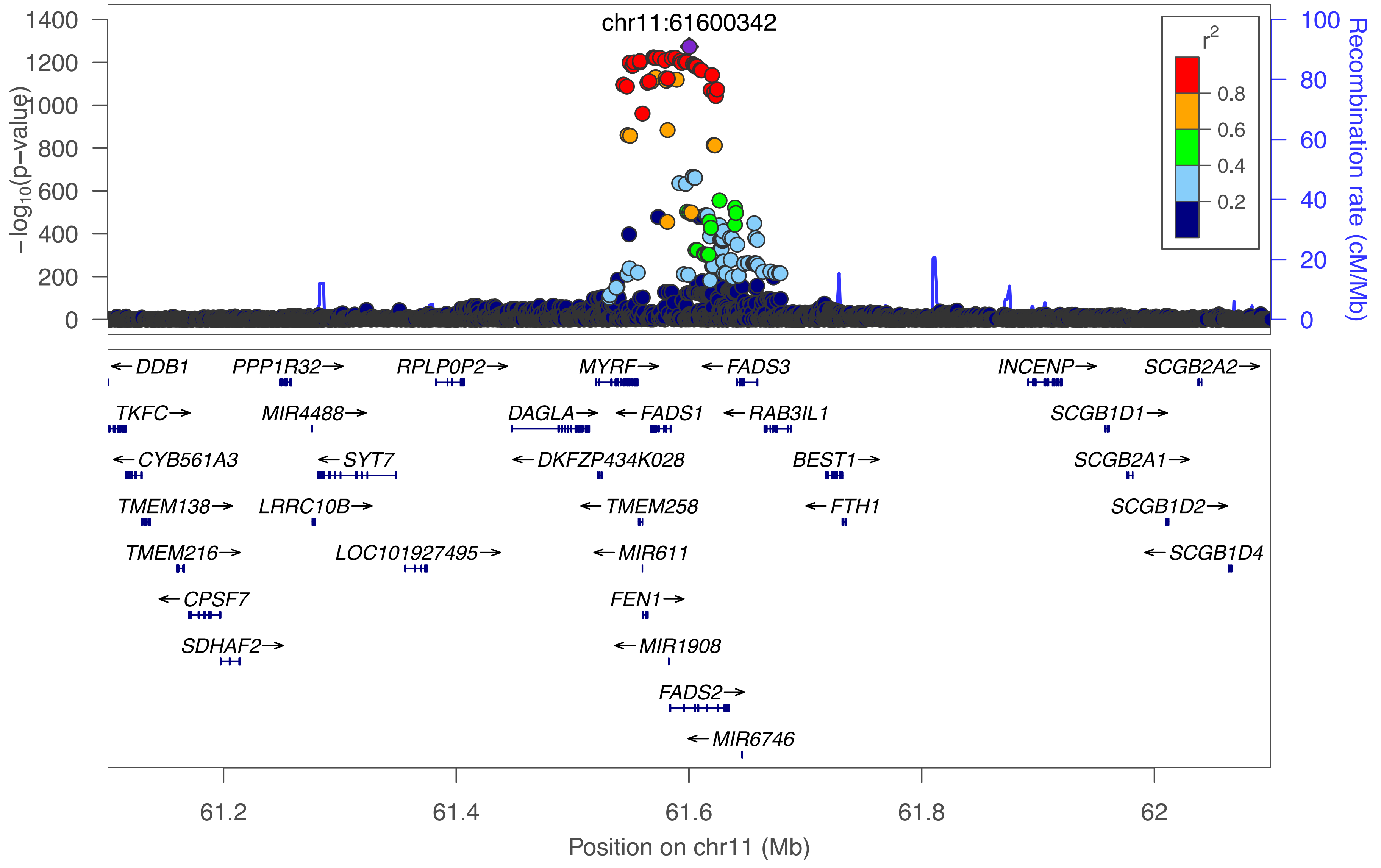

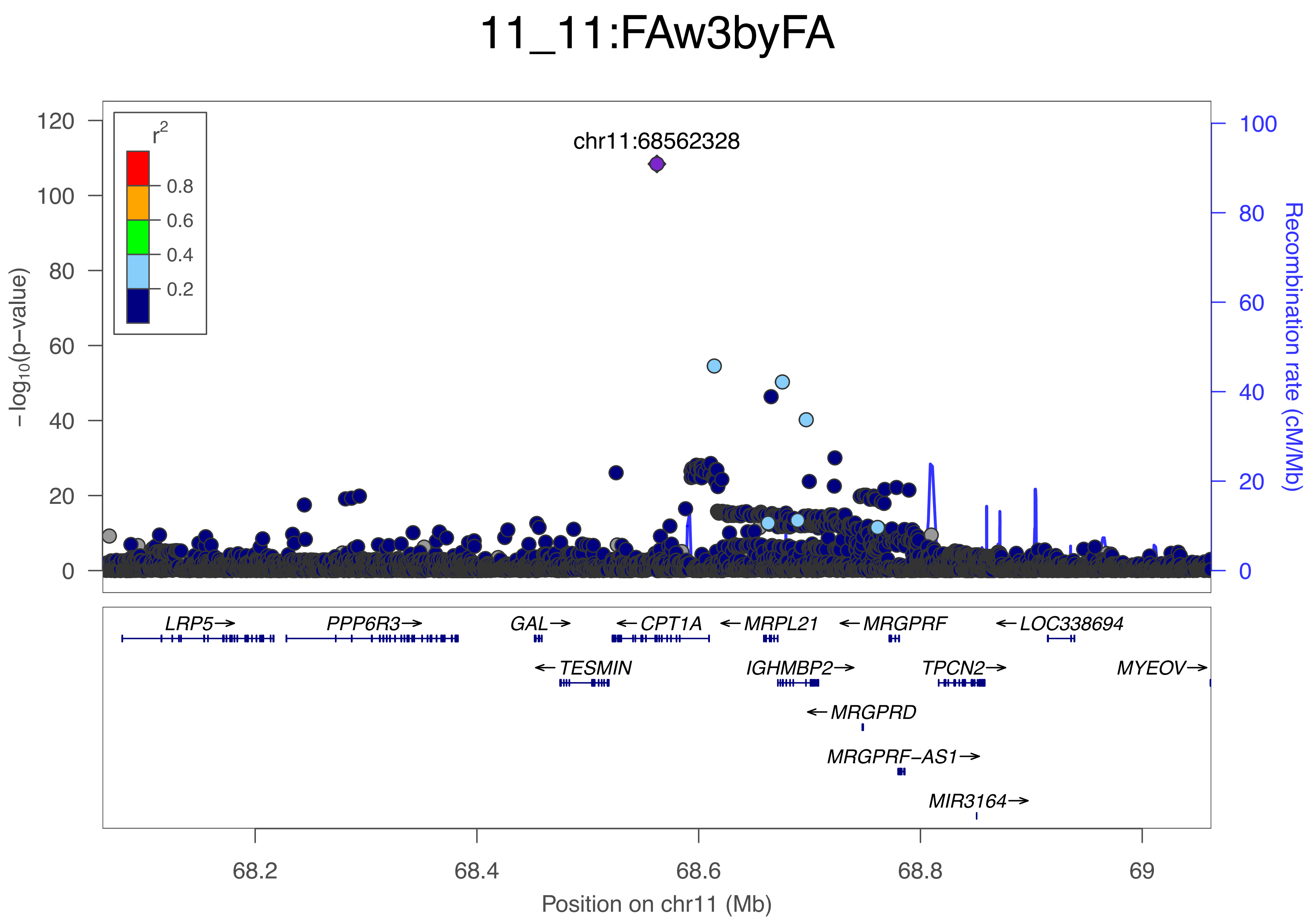
11_11:FAw3byFA
chr11:68562328
r2
0.8
0.6
0.4
0.2
−log10(p−value)
120
100
80
60
40
20
0
Recombination rate (cM/Mb)
100
80
60
40
20
0
LRP5→
PPP6R3→
GAL→
←CPT1A
←MRPL21
←MRGPRF
←LOC338694
←TESMIN
IGHMBP2→
TPCN2→
MYEOV→
←MRGPRD
MRGPRF−AS1→
MIR3164→
68.2
68.4
68.6
68.8
69
Position on chr11 (Mb)

### 11_12:Ala

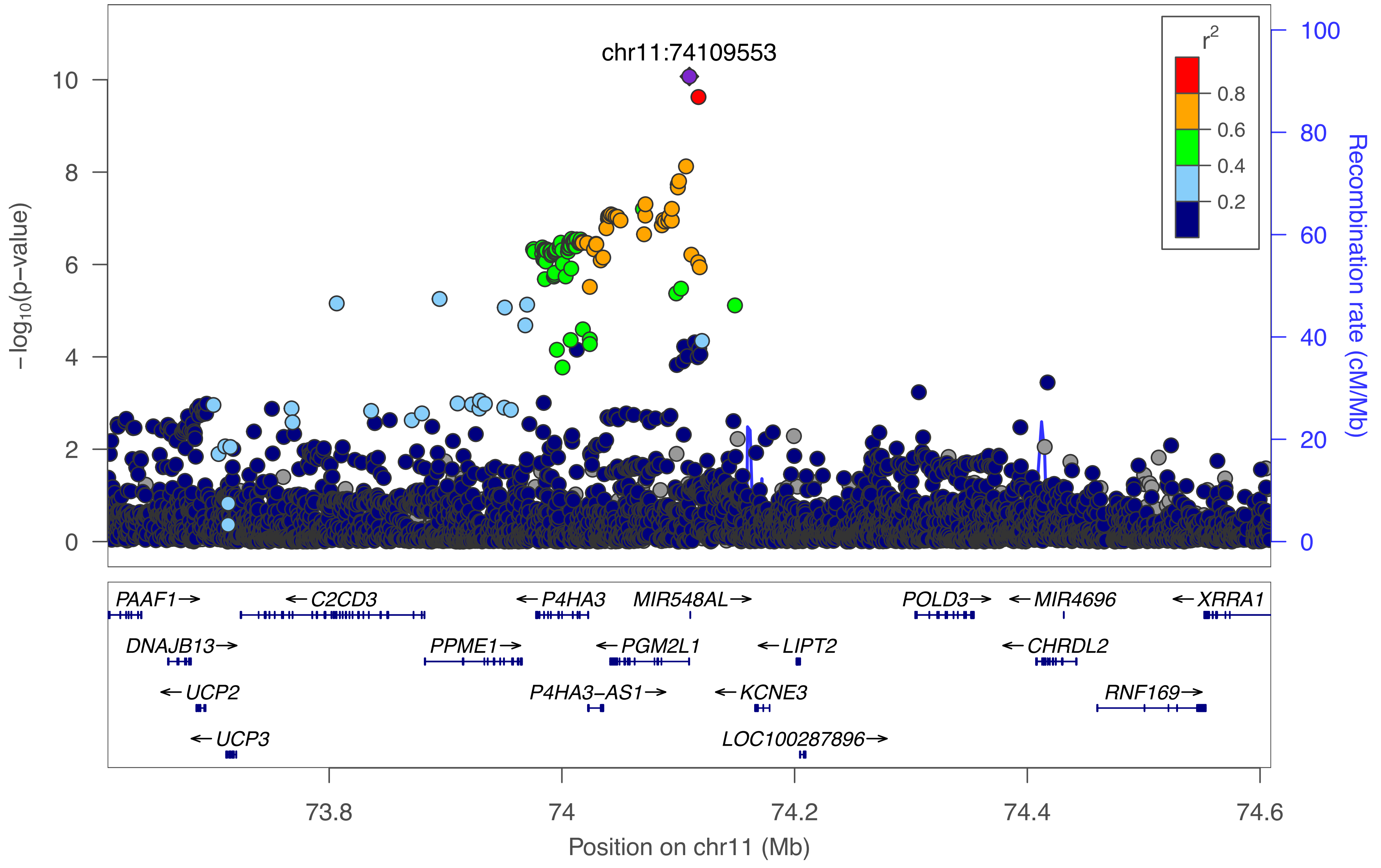

chr11:74109553
r2
0.8
0.6
0.4
0.2
−log10(p−value)
0
2
4
6
8
10
Recombination rate (cM/Mb)
0
20
40
60
80
100
PAAF1→
←C2CD3
←P4HA3
MIR548AL→
POLD3→
←MIR4696
←XRRA1
DNAJB13→
PPME1→
←PGM2L1
←LIPT2
←CHRDL2
←UCP2
P4HA3−AS1→
←KCNE3
RNF169→
←UCP3
LOC100287896→
73.8
74
74.2
74.4
74.6
Position on chr11 (Mb)

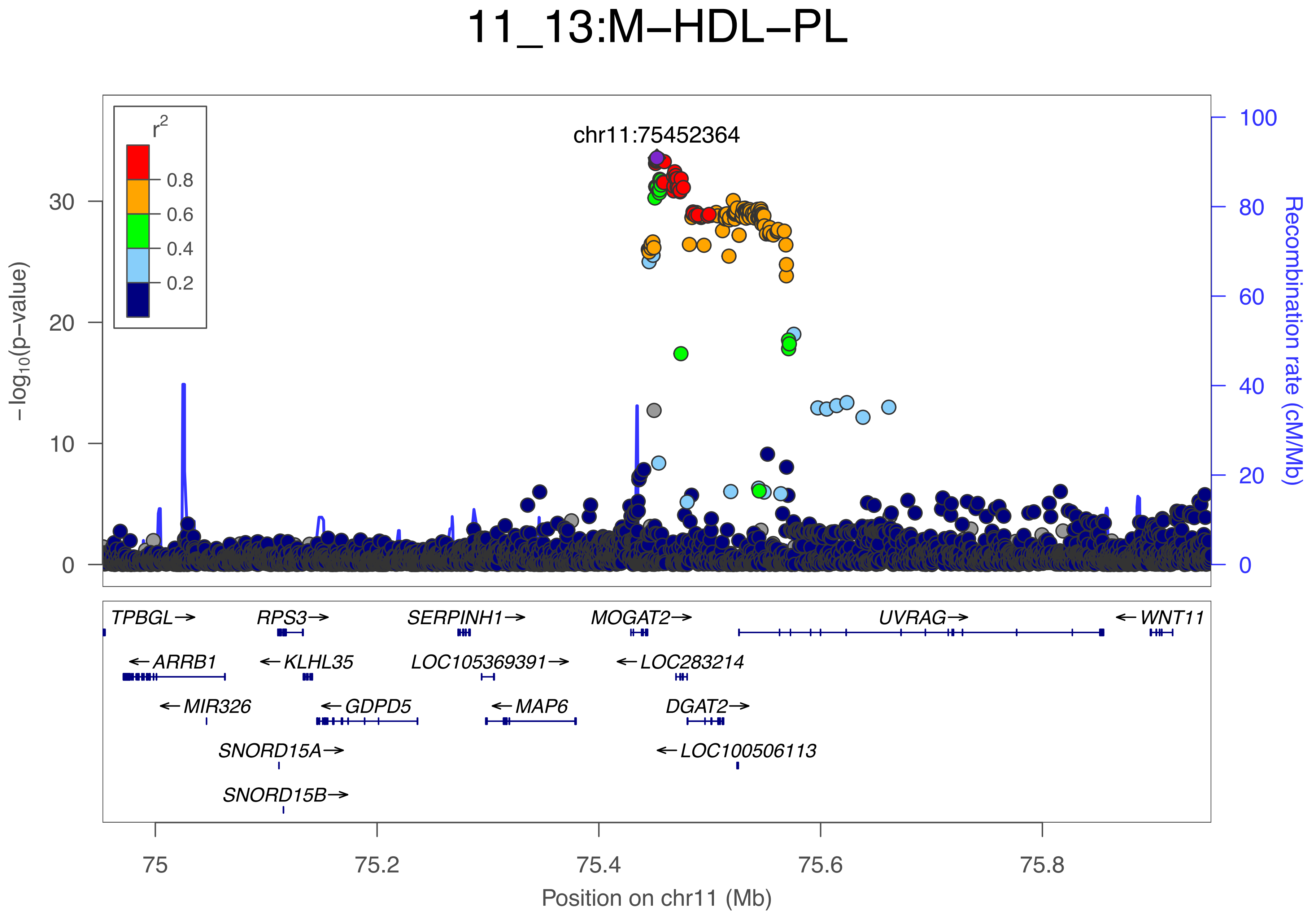
11_13:M−HDL−PL
chr11:75452364
r2
0.8
0.6
0.4
0.2
−log10(p−value)
Recombination rate (cM/Mb)
TPBGL→
RPS3→
SERPINH1→
MOGAT2→
UVRAG→
←WNT11
←ARRB1
←KLHL35
LOC105369391→
←LOC283214
←MIR326
←GDPD5
←MAP6
DGAT2→
SNORD15A→
←LOC100506113
SNORD15B→
Position on chr11 (Mb)

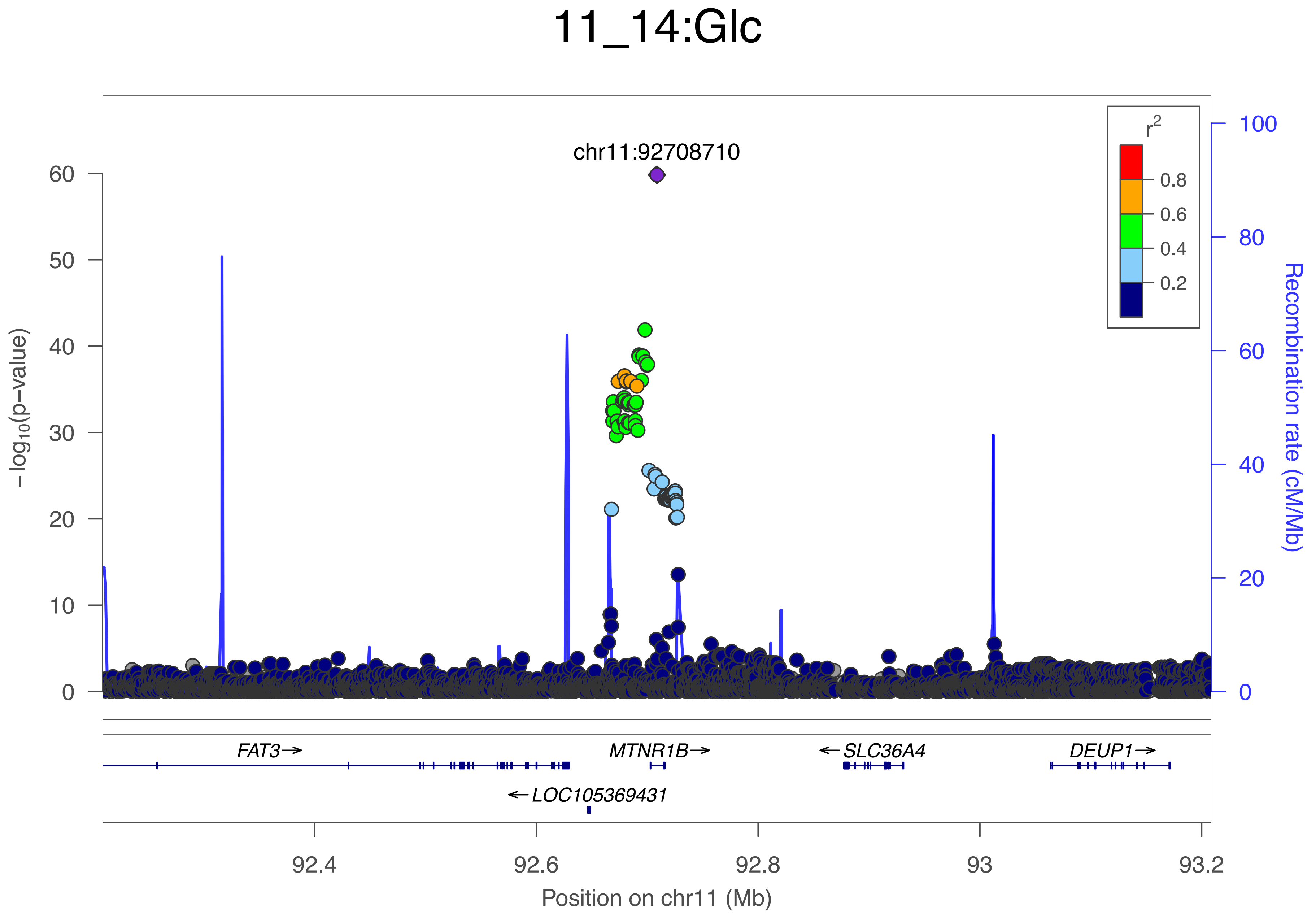

11_14:Glc
chr11:92708710
−log10(p−value)
0
10
20
30
40
50
60
r2
0.8
0.6
0.4
0.2
100
80
60
40
20
0
Recombination rate (cM/Mb)
FAT3→
MTNR1B→
←SLC36A4
DEUP1→
←LOC105369431
92.4
92.6
92.8
93
93.2
Position on chr11 (Mb)

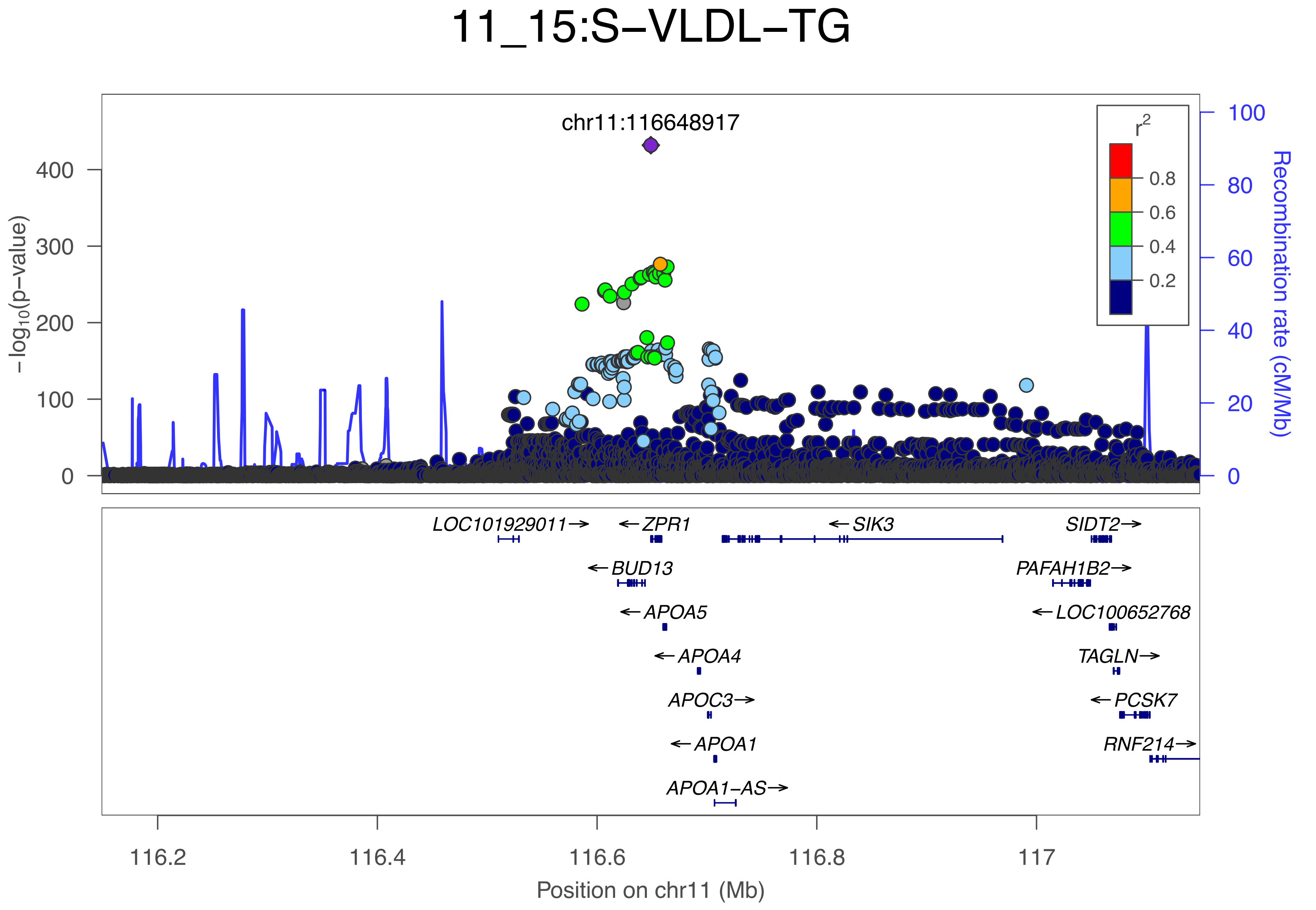

11_15:S−VLDL−TG
chr11:116648917
$-\log_{10}$(p−value)
0
100
200
300
400
Recombination rate (cM/Mb)
0
20
40
60
80
100
$r^2$
0.8
0.6
0.4
0.2
LOC101929011→
←ZPR1
←SIK3
SIDT2→
←BUD13
PAFAH1B2→
←APOA5
←LOC100652768
←APOA4
TAGLN→
APOC3→
←PCSK7
←APOA1
RNF214→
APOA1−AS→
116.2
116.4
116.6
116.8
117
Position on chr11 (Mb)

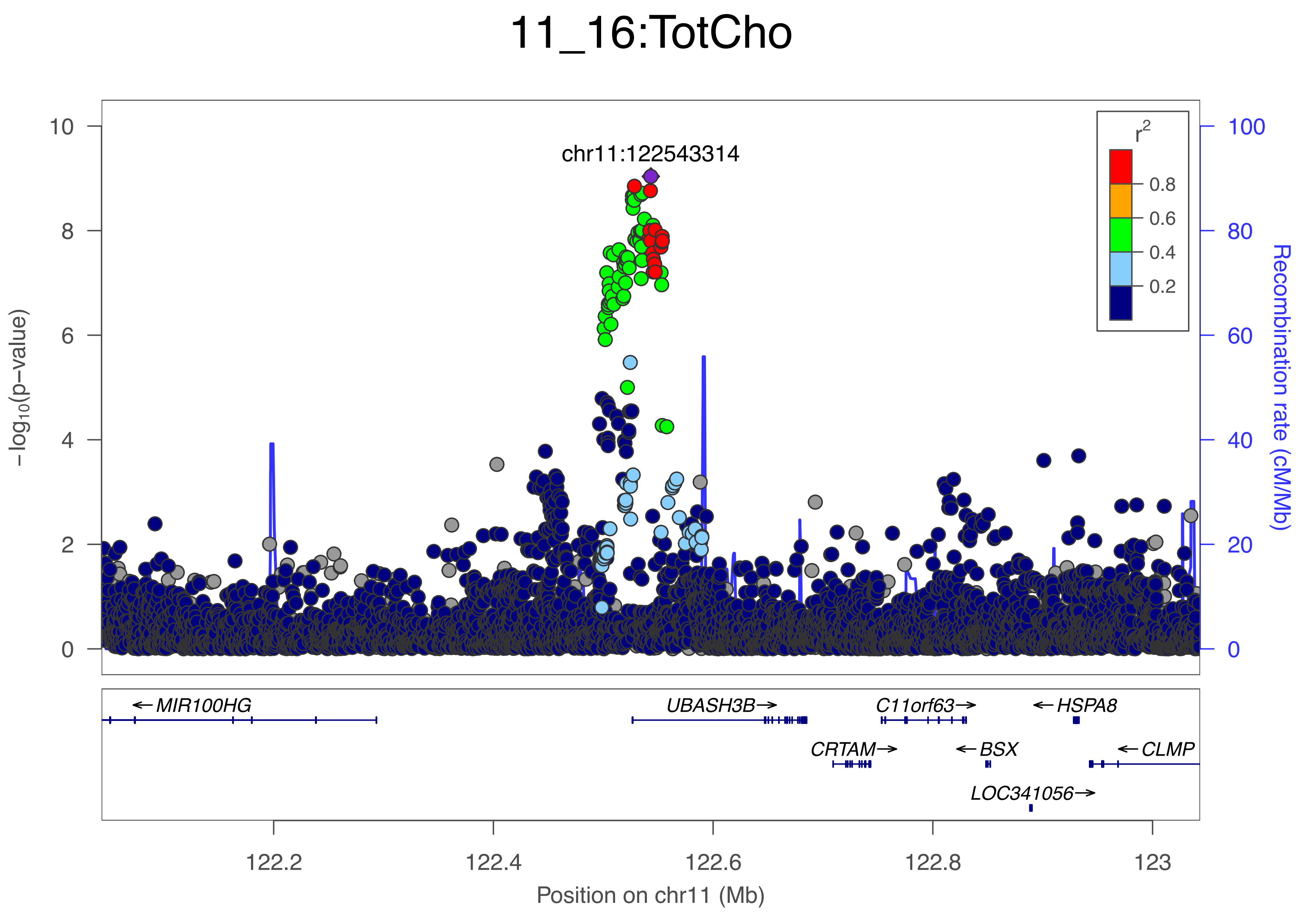
11_16:TotCho
chr11:122543314
r2
0.8
0.6
0.4
0.2
-log10(p-value)
Recombination rate (cM/Mb)
10
8
6
4
2
0
100
80
60
40
20
0
MIR100HG
UBASH3B
C11orf63
HSPA8
CRTAM
BSX
CLMP
LOC341056
122.2
122.4
122.6
122.8
123
Position on chr11 (Mb)

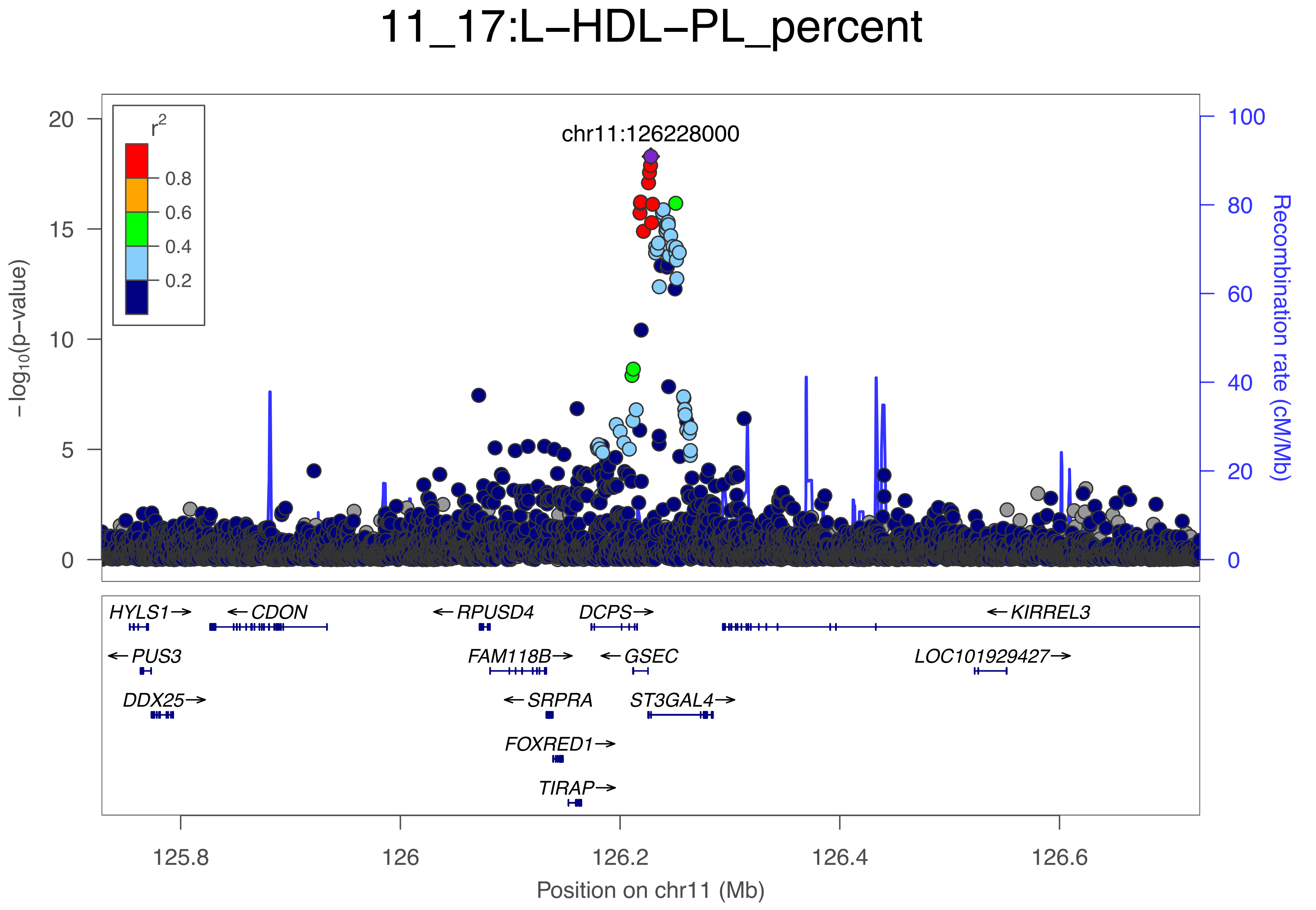

11_17:L−HDL−PL_percent
chr11:126228000
r2
0.8
0.6
0.4
0.2
−log10(p−value)
Recombination rate (cM/Mb)
20
15
10
5
0
100
80
60
40
20
0
HYLS1→
←CDON
←RPUSD4
DCPS→
←KIRREL3
←PUS3
FAM118B→
←GSEC
LOC101929427→
DDX25→
←SRPRA
ST3GAL4→
FOXRED1→
TIRAP→
125.8
126
126.2
126.4
126.6
Position on chr11 (Mb)

### 11_18:XS−VLDL−C

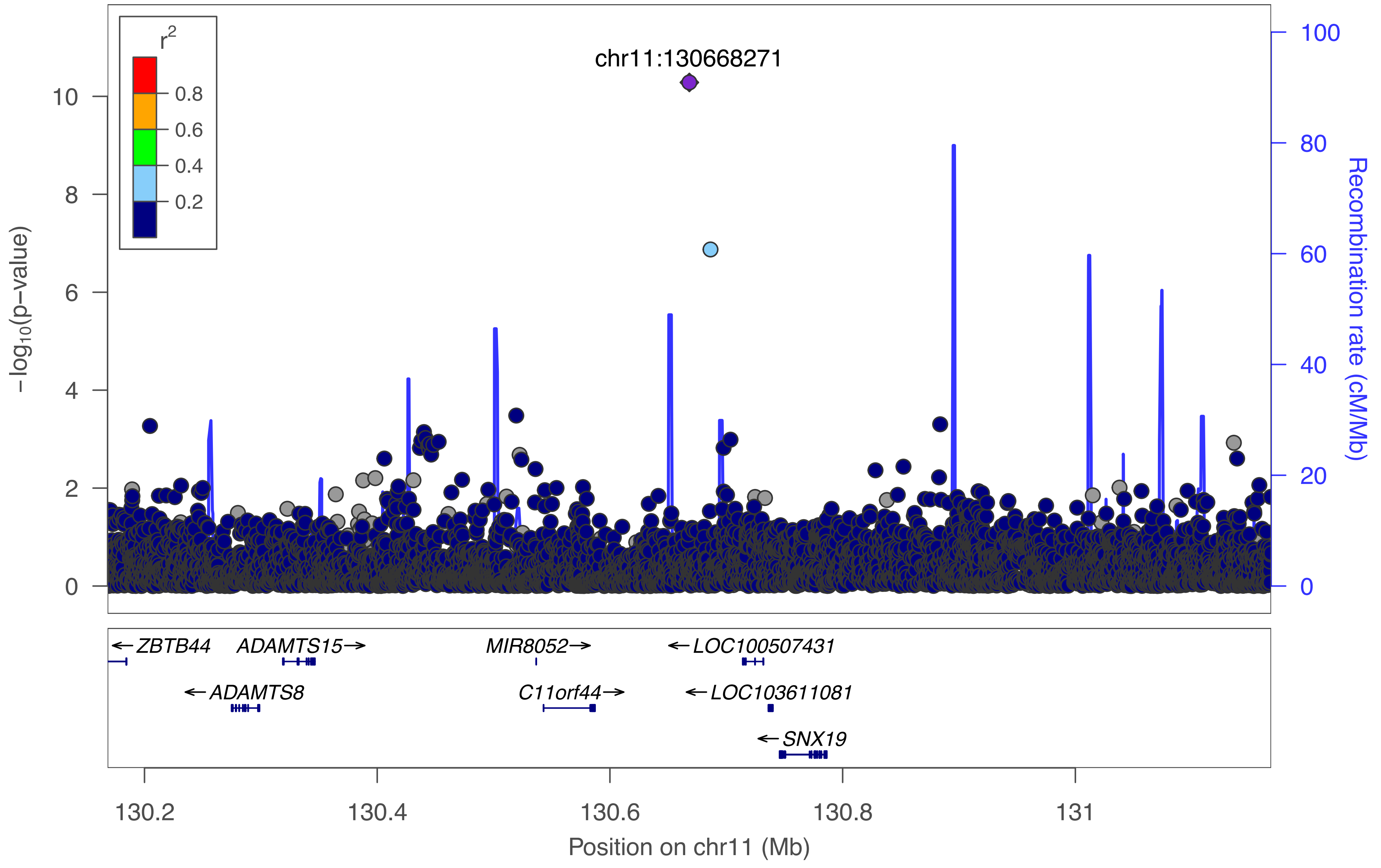

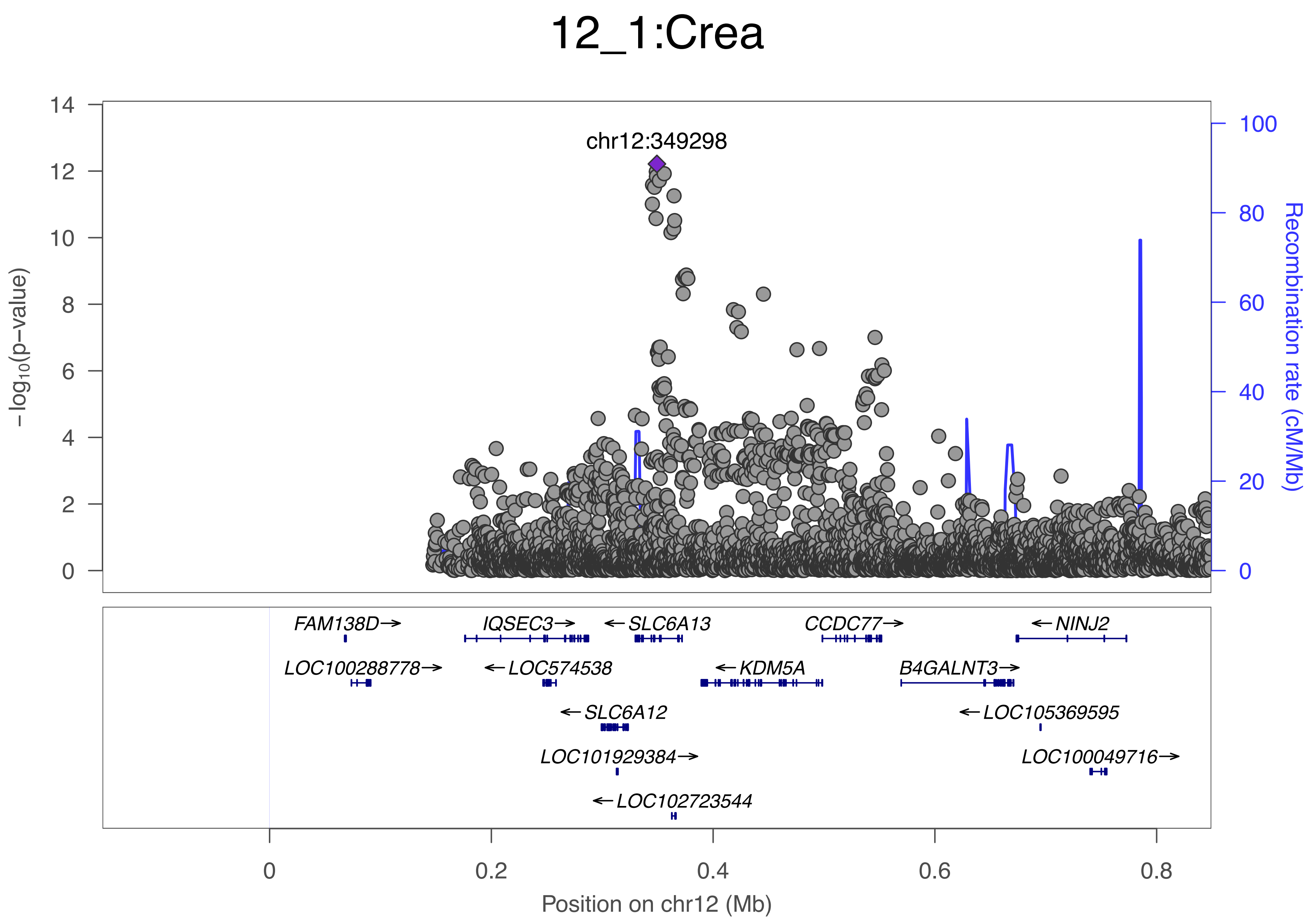

12_1:Crea
chr12:349298
−log10(p−value)
Recombination rate (cM/Mb)
0
2
4
6
8
10
12
14
0
20
40
60
80
100
FAM138D→
IQSEC3→
←SLC6A13
CCDC77→
←NINJ2
LOC100288778→
←LOC574538
←KDM5A
B4GALNT3→
←SLC6A12
←LOC105369595
LOC101929384→
LOC100049716→
←LOC102723544
0
0.2
0.4
0.6
0.8
Position on chr12 (Mb)

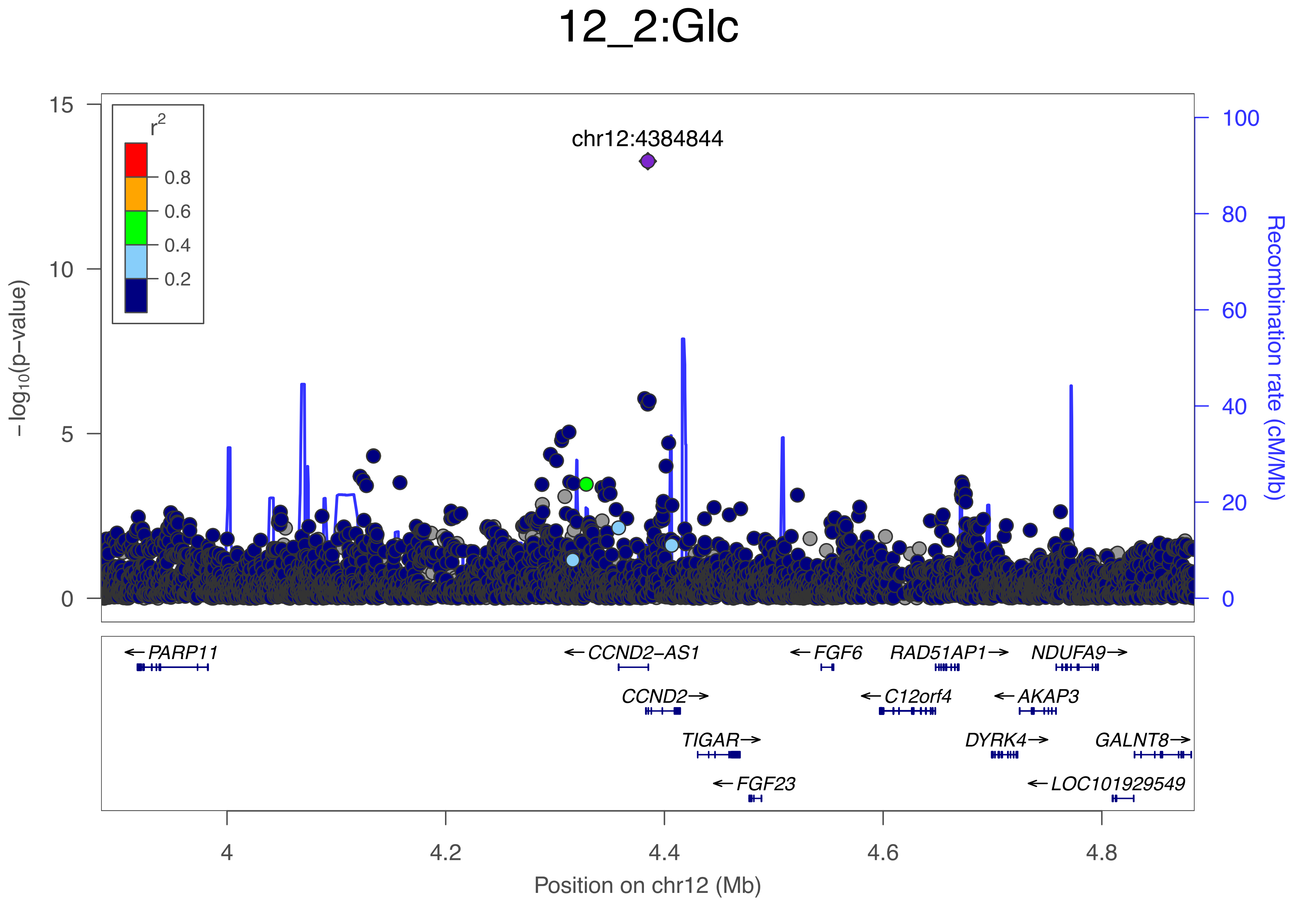
12_2:Glc
chr12:4384844
r2
0.8
0.6
0.4
0.2
15
10
5
0
−log10(p−value)
100
80
60
40
20
0
Recombination rate (cM/Mb)
←PARP11
←CCND2−AS1
←FGF6
RAD51AP1→
NDUFA9→
CCND2→
←C12orf4
←AKAP3
TIGAR→
DYRK4→
GALNT8→
←FGF23
←LOC101929549
4
4.2
4.4
4.6
4.8
Position on chr12 (Mb)

### 12_3:PUFA

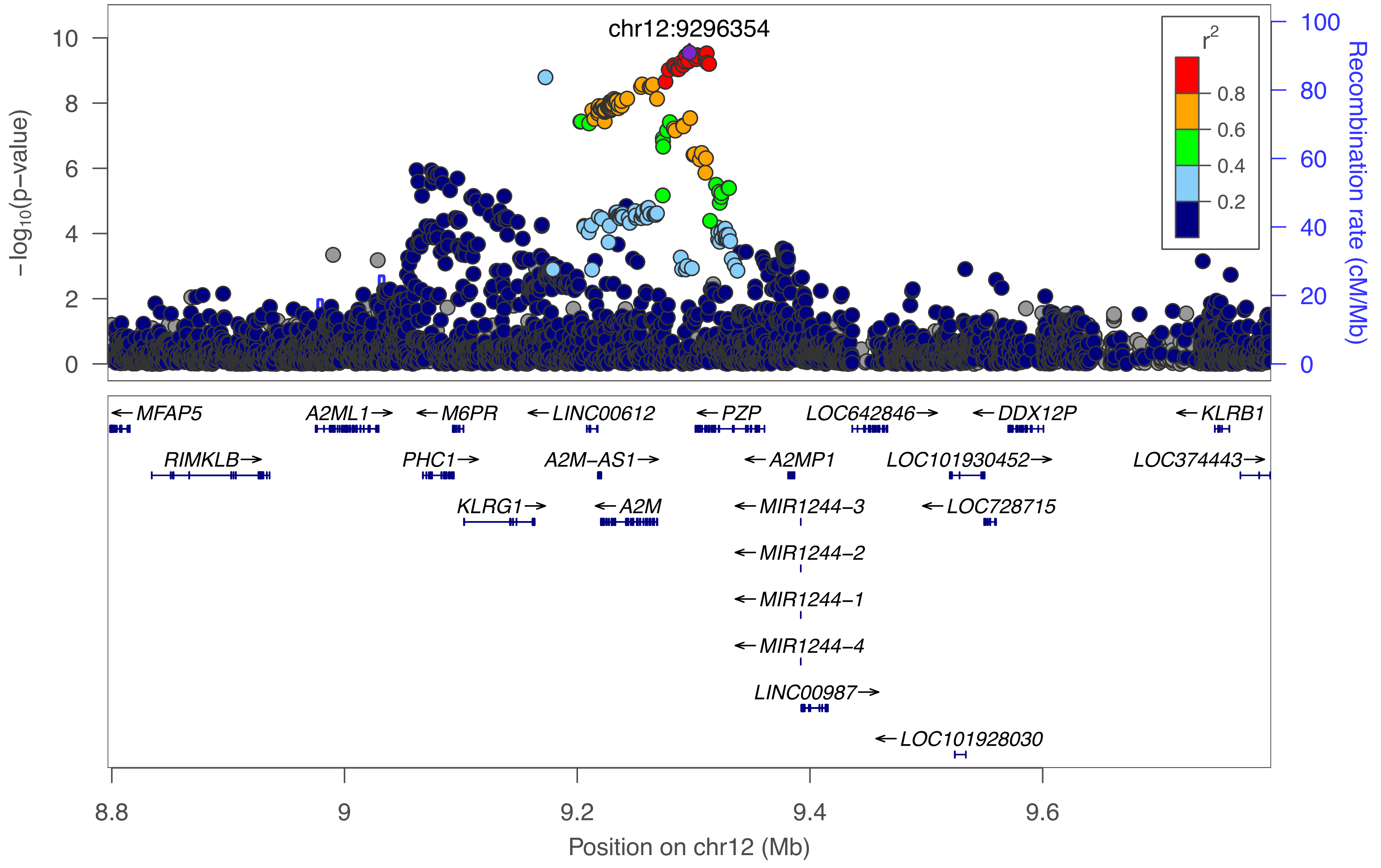

chr12:9296354
r^2
0.8
0.6
0.4
0.2
10
8
6
4
2
0
-log10(p-value)
100
80
60
40
20
0
Recombination rate (cM/Mb)
← MFAP5
A2ML1 →
← M6PR
← LINC00612
← PZP
LOC642846 →
← DDX12P
← KLRB1
RIMKLB →
PHC1 →
A2M-AS1 →
← A2MP1
LOC101930452 →
LOC374443 →
KLRG1 →
← A2M
← MIR1244-3
← LOC728715
← MIR1244-2
← MIR1244-1
← MIR1244-4
LINC00987 →
← LOC101928030
8.8
9
9.2
9.4
9.6
Position on chr12 (Mb)

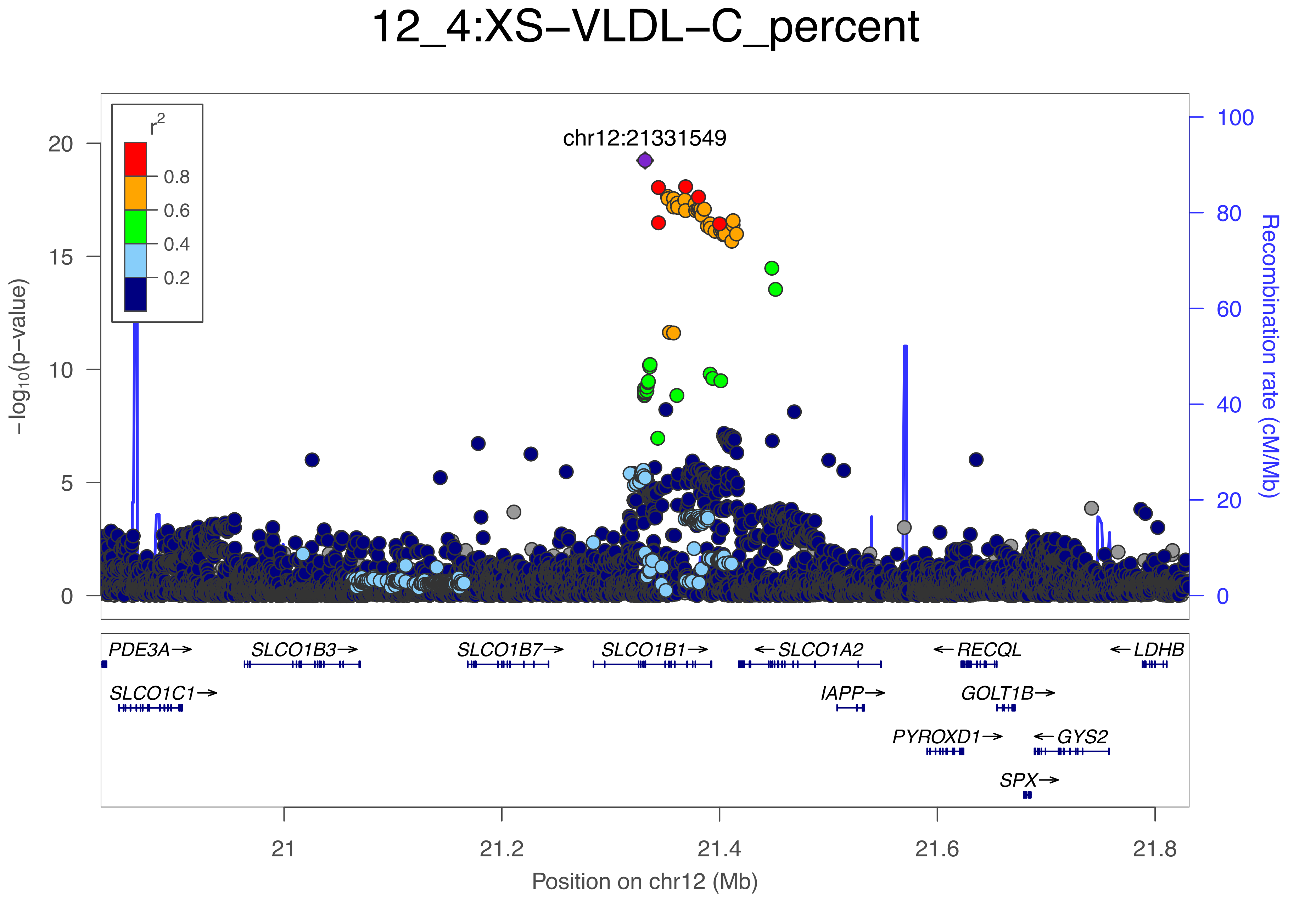

12_4:XS−VLDL−C_percent
chr12:21331549
r2
0.8
0.6
0.4
0.2
−log10(p−value)
Recombination rate (cM/Mb)
0
5
10
15
20
0
20
40
60
80
100
PDE3A→
SLCO1C1→
SLCO1B3→
SLCO1B7→
SLCO1B1→
←SLCO1A2
IAPP→
PYROXD1→
←RECQL
GOLT1B→
SPX→
←GYS2
←LDHB
21
21.2
21.4
21.6
21.8
Position on chr12 (Mb)

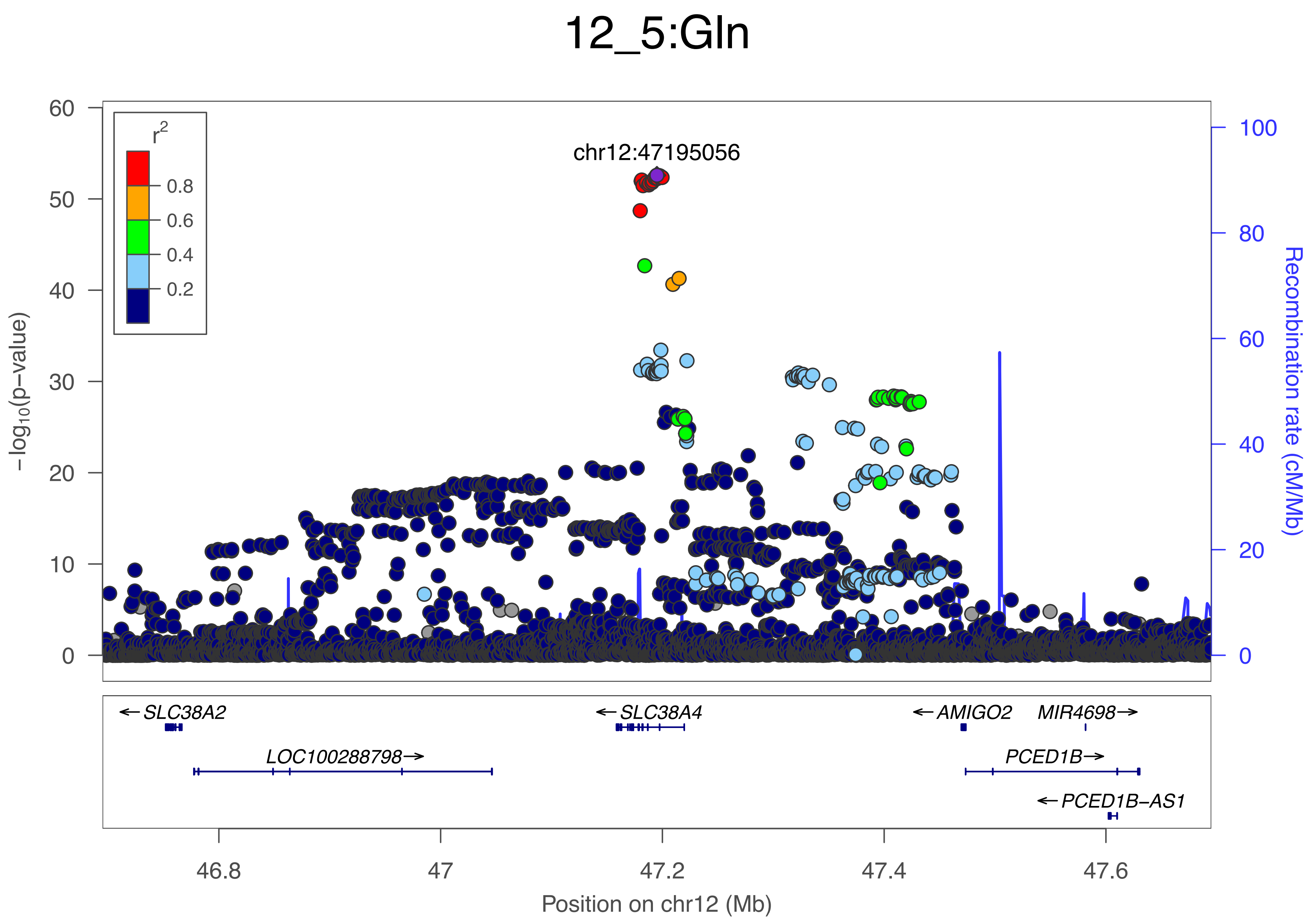

12_5:Gln
chr12:47195056
r²
0.8
0.6
0.4
0.2
−log₁₀(p−value)
Recombination rate (cM/Mb)
0
10
20
30
40
50
60
0
20
40
60
80
100
←SLC38A2
←SLC38A4
←AMIGO2
MIR4698→
LOC100288798→
PCED1B→
←PCED1B−AS1
46.8
47
47.2
47.4
47.6
Position on chr12 (Mb)

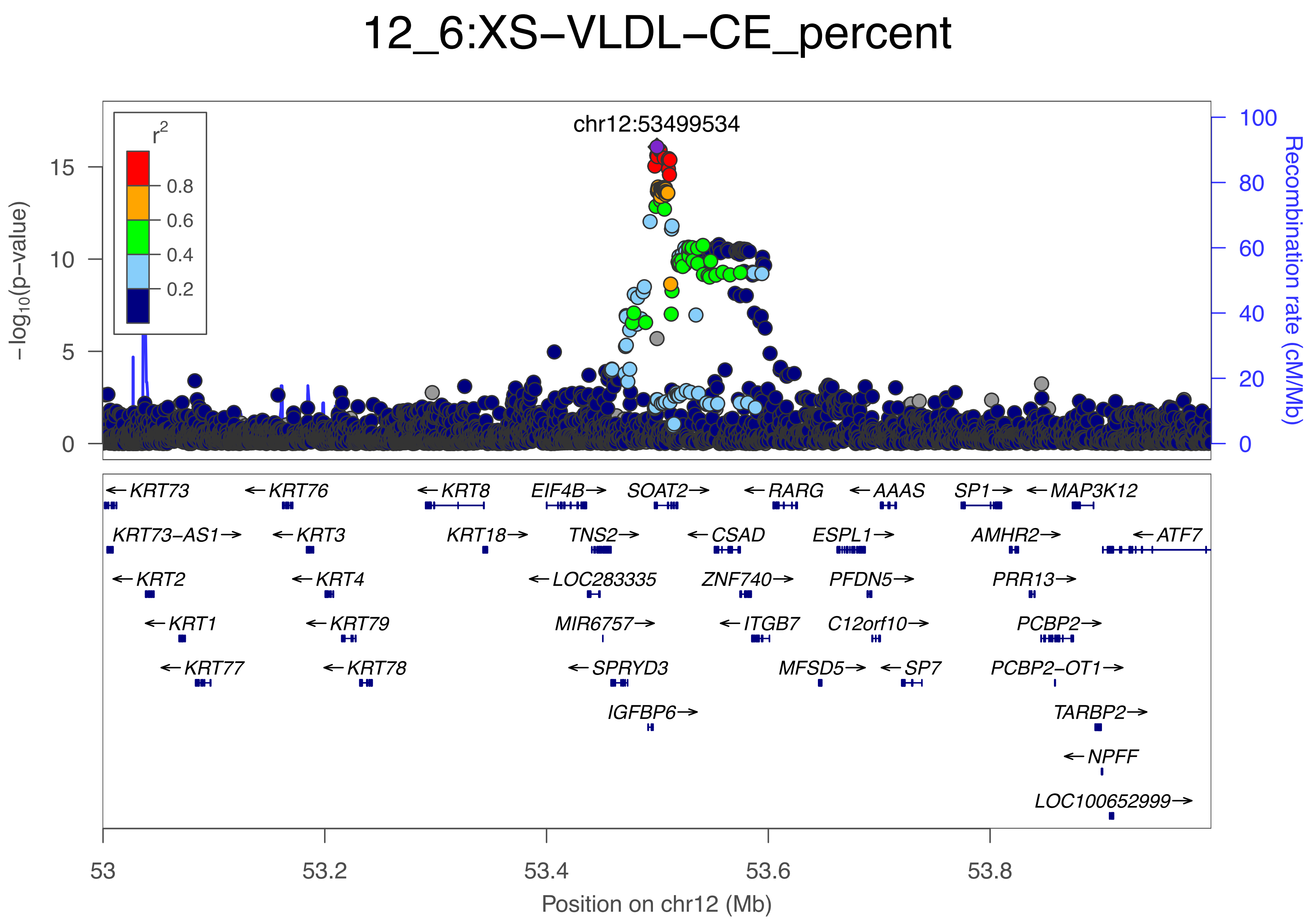
12_6:XS−VLDL−CE_percent
chr12:53499534
r2
0.8
0.6
0.4
0.2
−log10(p−value)
Recombination rate (cM/Mb)
Position on chr12 (Mb)
53
53.2
53.4
53.6
53.8
KRT73
KRT73−AS1
KRT2
KRT1
KRT77
KRT76
KRT3
KRT4
KRT79
KRT78
KRT8
KRT18
EIF4B
TNS2
LOC283335
MIR6757
SPRYD3
IGFBP6
SOAT2
CSAD
ZNF740
ITGB7
RARG
MFSD5
ESPL1
PFDN5
C12orf10
AAAS
SP7
SP1
AMHR2
PRR13
PCBP2
PCBP2−OT1
MAP3K12
TARBP2
NPFF
LOC100652999
ATF7

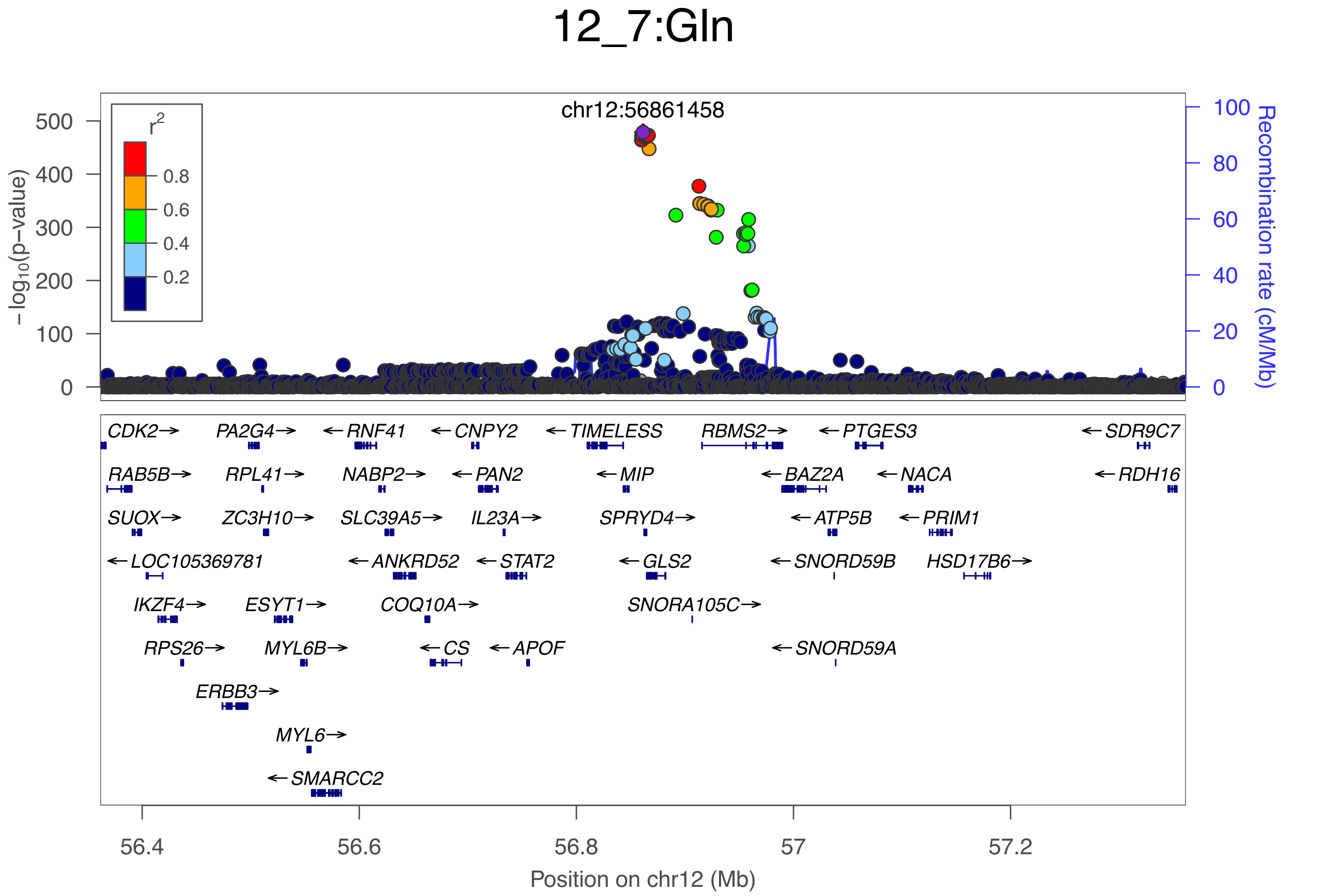

12_7:Gln
chr12:56861458
r2
0.8
0.6
0.4
0.2
−log10(p−value)
500
400
300
200
100
0
Recombination rate (cM/Mb)
100
80
60
40
20
0
CDK2→
PA2G4→
←RNF41
←CNPY2
←TIMELESS
RBMS2→
←PTGES3
←SDR9C7
RAB5B→
RPL41→
NABP2→
←PAN2
←MIP
←BAZ2A
←NACA
←RDH16
SUOX→
ZC3H10→
SLC39A5→
IL23A→
SPRYD4→
←ATP5B
←PRIM1
←LOC105369781
←ANKRD52
←STAT2
←GLS2
←SNORD59B
HSD17B6→
IKZF4→
ESYT1→
COQ10A→
SNORA105C→
RPS26→
MYL6B→
←CS
←APOF
←SNORD59A
ERBB3→
MYL6→
←SMARCC2
56.4
56.6
56.8
57
57.2
Position on chr12 (Mb)

### 12_8:Gln

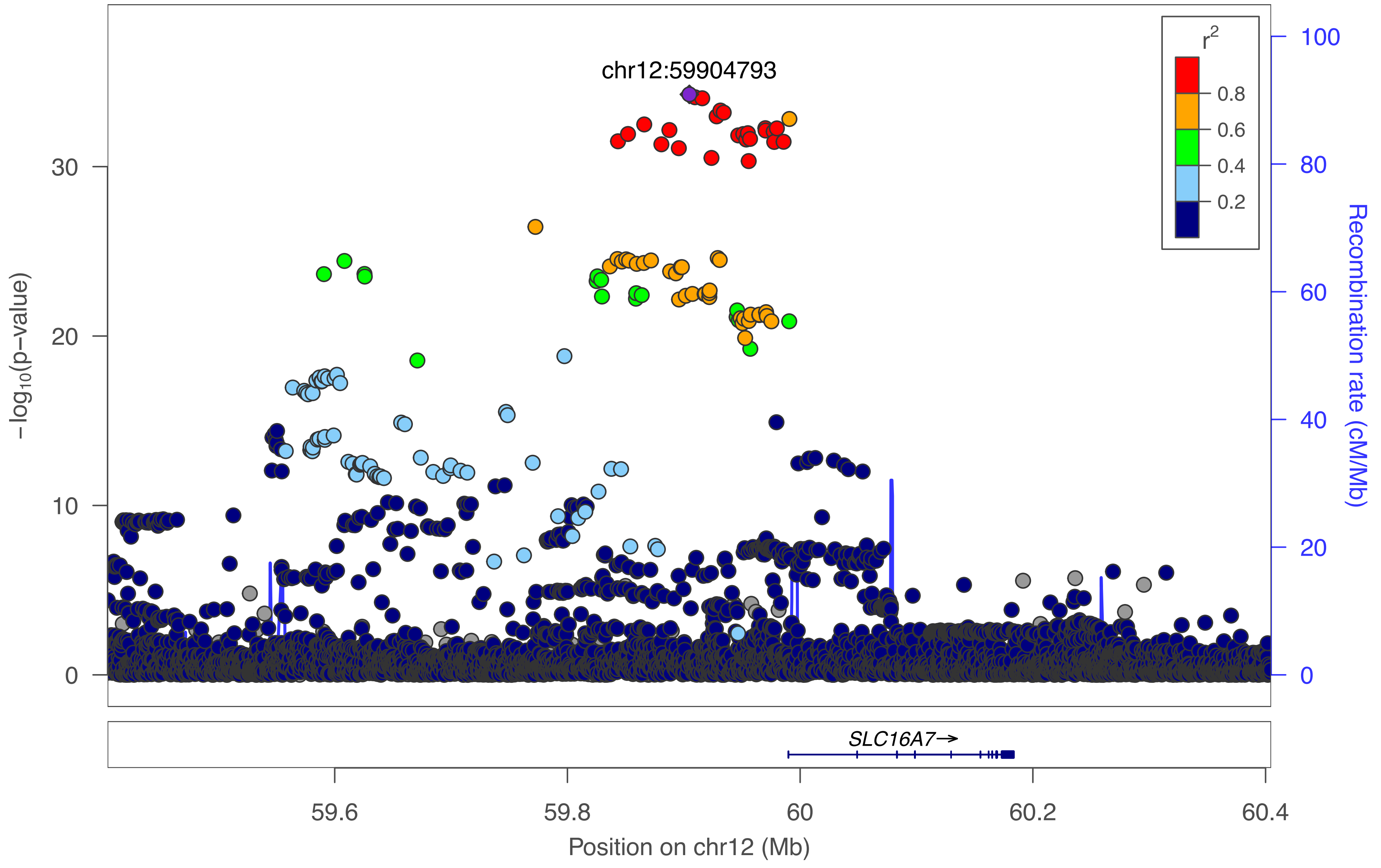

chr12:59904793
r²
0.8
0.6
0.4
0.2
30
20
10
0
−log₁₀(p−value)
100
80
60
40
20
0
Recombination rate (cM/Mb)
SLC16A7→
59.6
59.8
60
60.2
60.4
Position on chr12 (Mb)

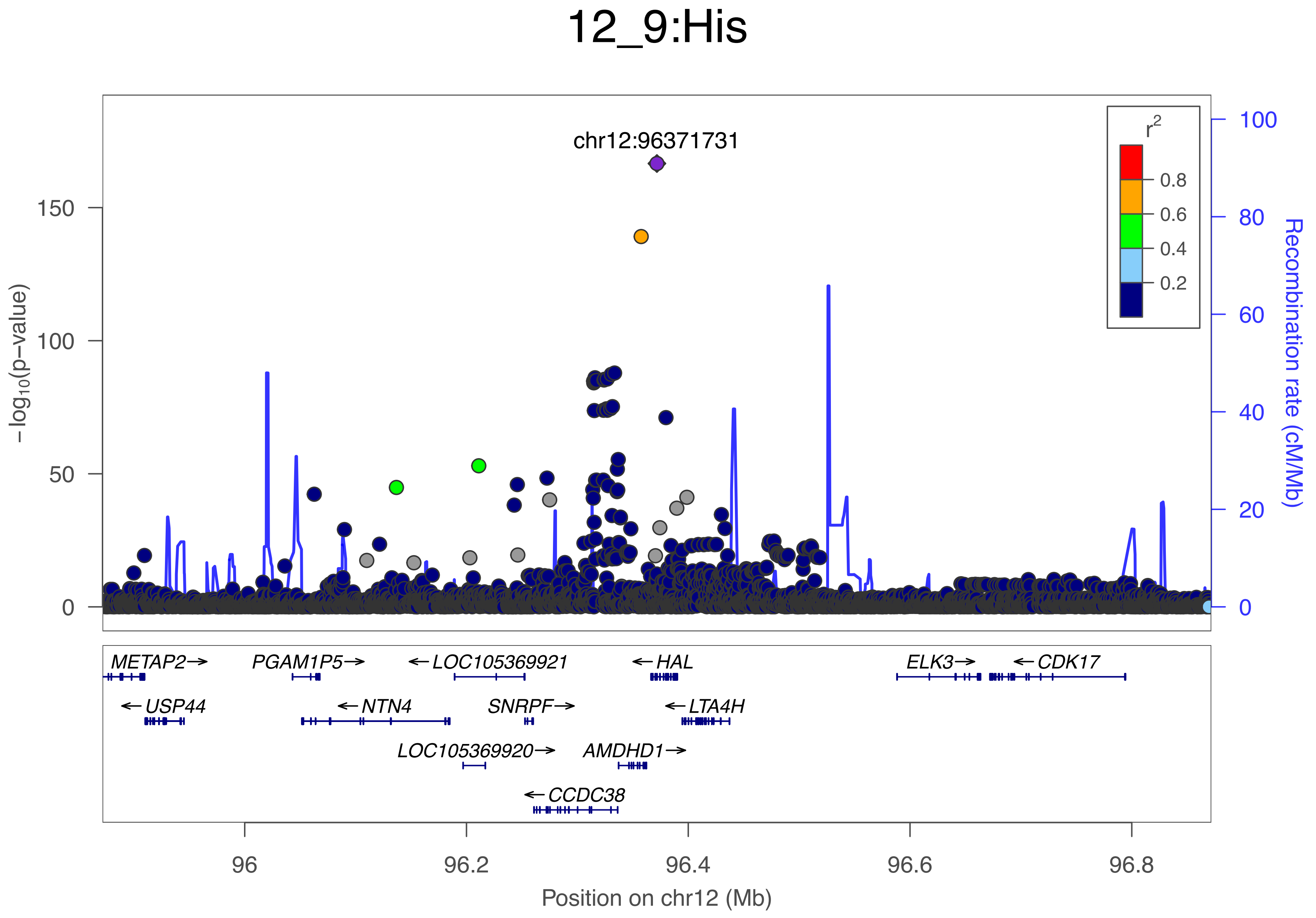

12_9:His
chr12:96371731
r²
0.8
0.6
0.4
0.2
150
100
50
0
100
80
60
40
20
0
−log₁₀(p−value)
Recombination rate (cM/Mb)
METAP2→
PGAM1P5→
←LOC105369921
←HAL
ELK3→
←CDK17
←USP44
←NTN4
SNRPF→
←LTA4H
LOC105369920→
AMDHD1→
←CCDC38
96
96.2
96.4
96.6
96.8
Position on chr12 (Mb)

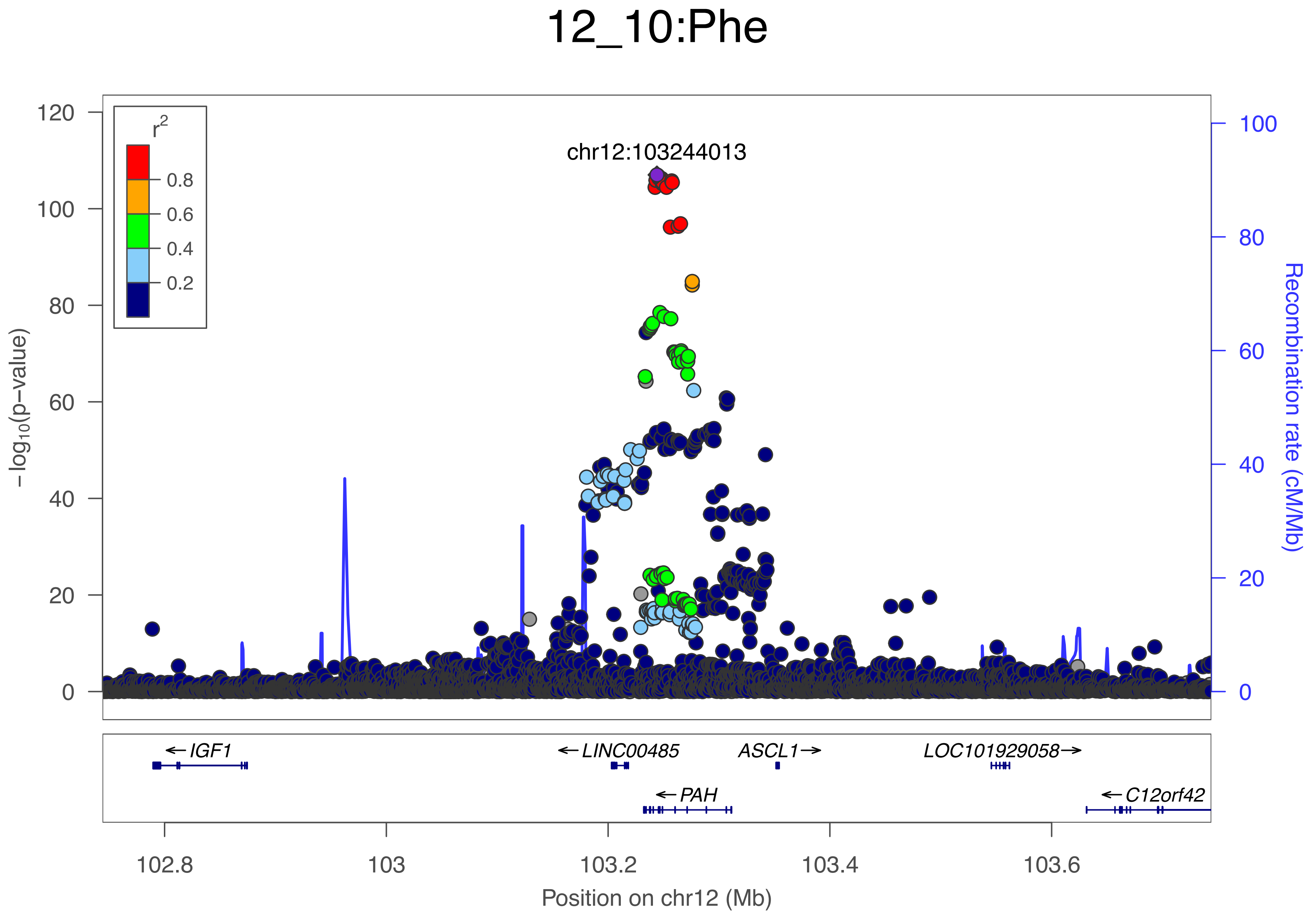
12_10:Phe
chr12:103244013
r2
0.8
0.6
0.4
0.2
120
100
80
60
40
20
0
−log10(p−value)
100
80
60
40
20
0
Recombination rate (cM/Mb)
← IGF1
← LINC00485
ASCL1 →
LOC101929058 →
← PAH
← C12orf42
102.8
103
103.2
103.4
103.6
Position on chr12 (Mb)

### 12_11:HDL−D

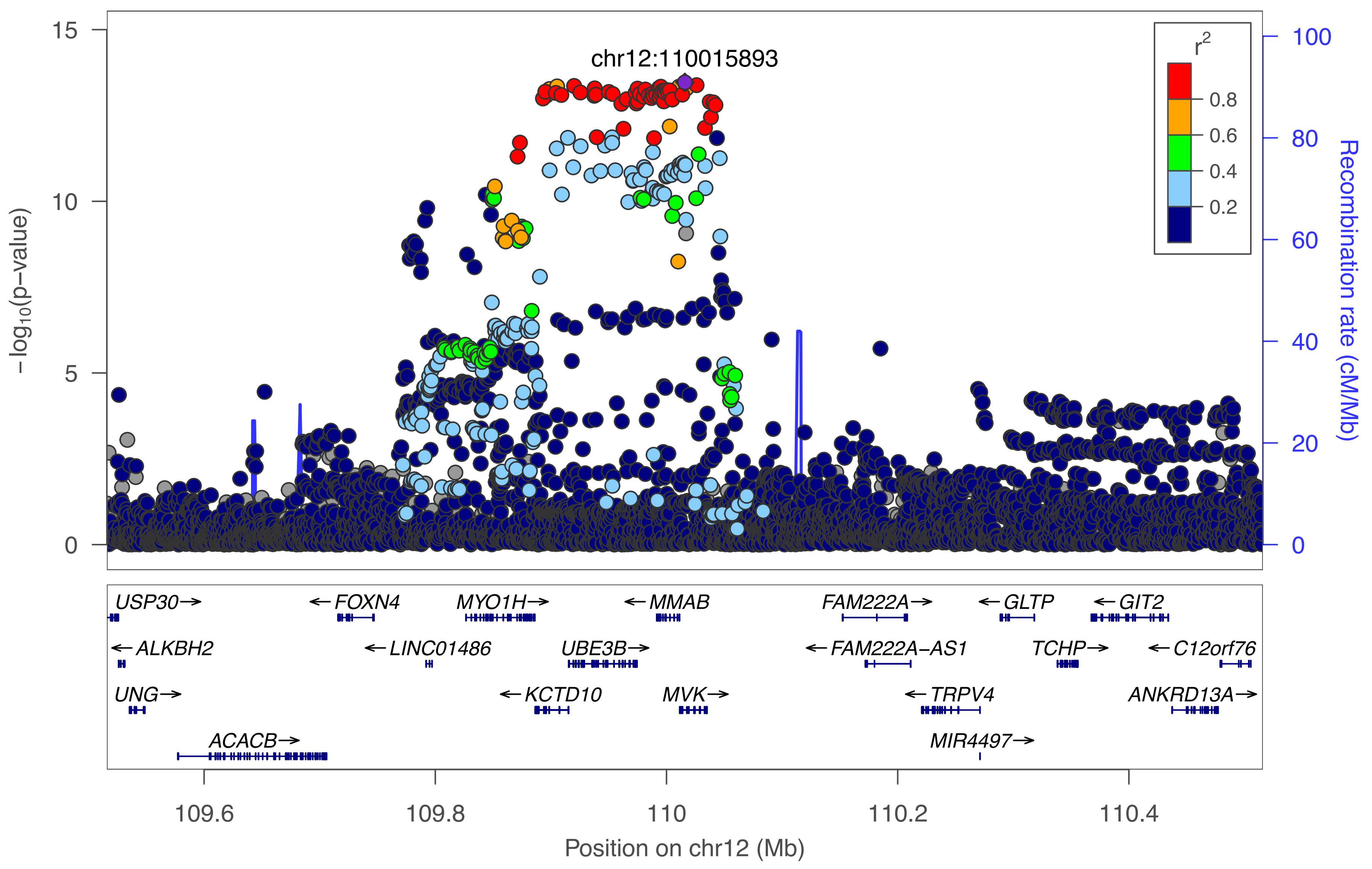

chr12:110015893
r2
0.8
0.6
0.4
0.2
−log10(p−value)
15
10
5
0
Recombination rate (cM/Mb)
100
80
60
40
20
0
USP30→
←FOXN4
MYO1H→
←MMAB
FAM222A→
←GLTP
←GIT2
←ALKBH2
←LINC01486
UBE3B→
←FAM222A-AS1
TCHP→
←C12orf76
UNG→
←KCTD10
MVK→
←TRPV4
ANKRD13A→
ACACB→
MIR4497→
109.6
109.8
110
110.2
110.4
Position on chr12 (Mb)

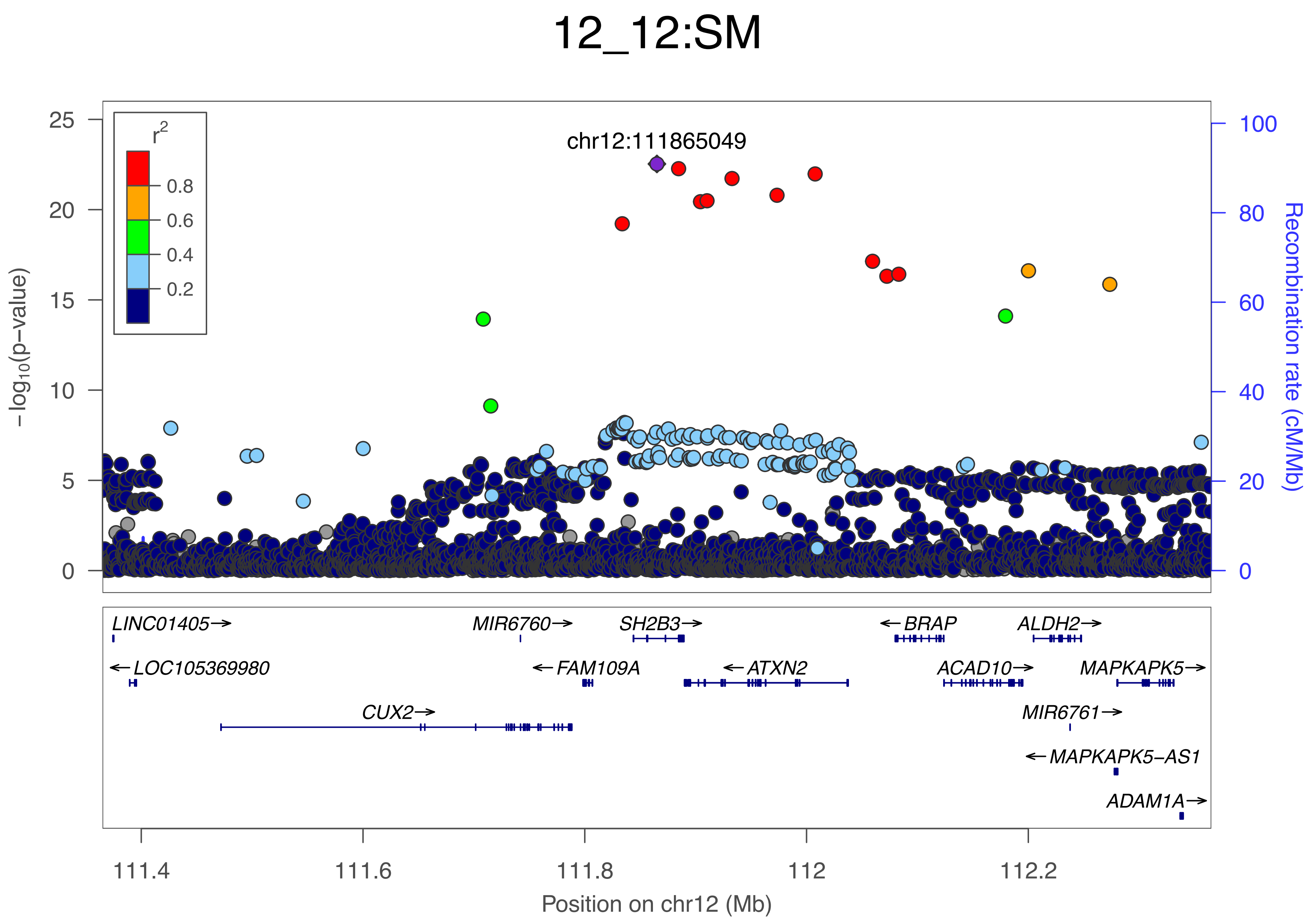

12_12:SM
chr12:111865049
r2
0.8
0.6
0.4
0.2
25
20
15
10
5
0
−log10(p−value)
100
80
60
40
20
0
Recombination rate (cM/Mb)
LINC01405→
MIR6760→
SH2B3→
←BRAP
ALDH2→
←LOC105369980
←FAM109A
←ATXN2
ACAD10→
MAPKAPK5→
CUX2→
MIR6761→
←MAPKAPK5−AS1
ADAM1A→
111.4
111.6
111.8
112
112.2
Position on chr12 (Mb)

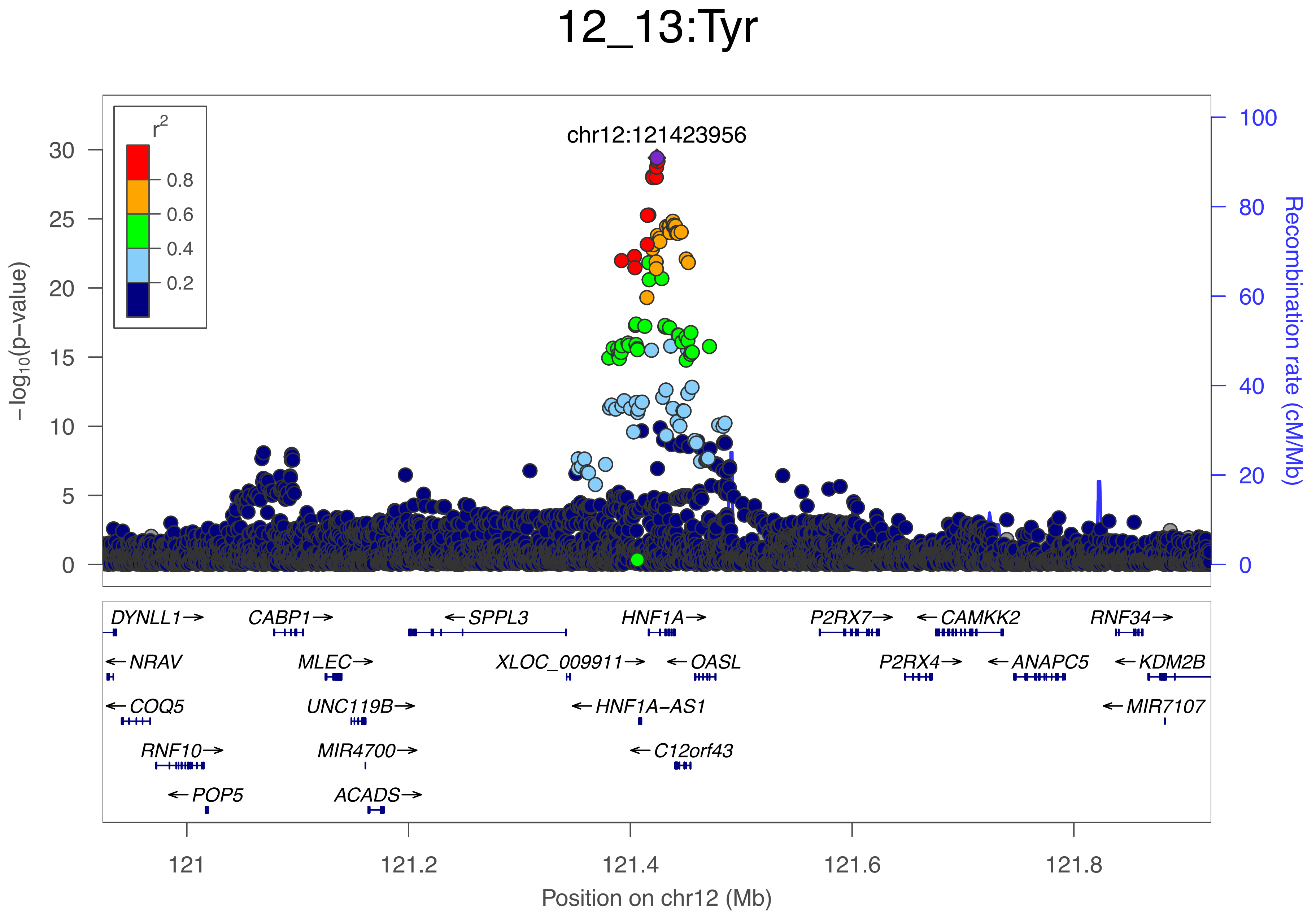
12_13:Tyr
chr12:121423956
r2
0.8
0.6
0.4
0.2
−log10(p−value)
Recombination rate (cM/Mb)
DYNLL1→
CABP1→
←SPPL3
HNF1A→
P2RX7→
←CAMKK2
RNF34→
←NRAV
MLEC→
XLOC_009911→
←OASL
P2RX4→
←ANAPC5
←KDM2B
←COQ5
UNC119B→
←HNF1A−AS1
←MIR7107
RNF10→
MIR4700→
←C12orf43
←POP5
ACADS→
121
121.2
121.4
121.6
121.8
Position on chr12 (Mb)

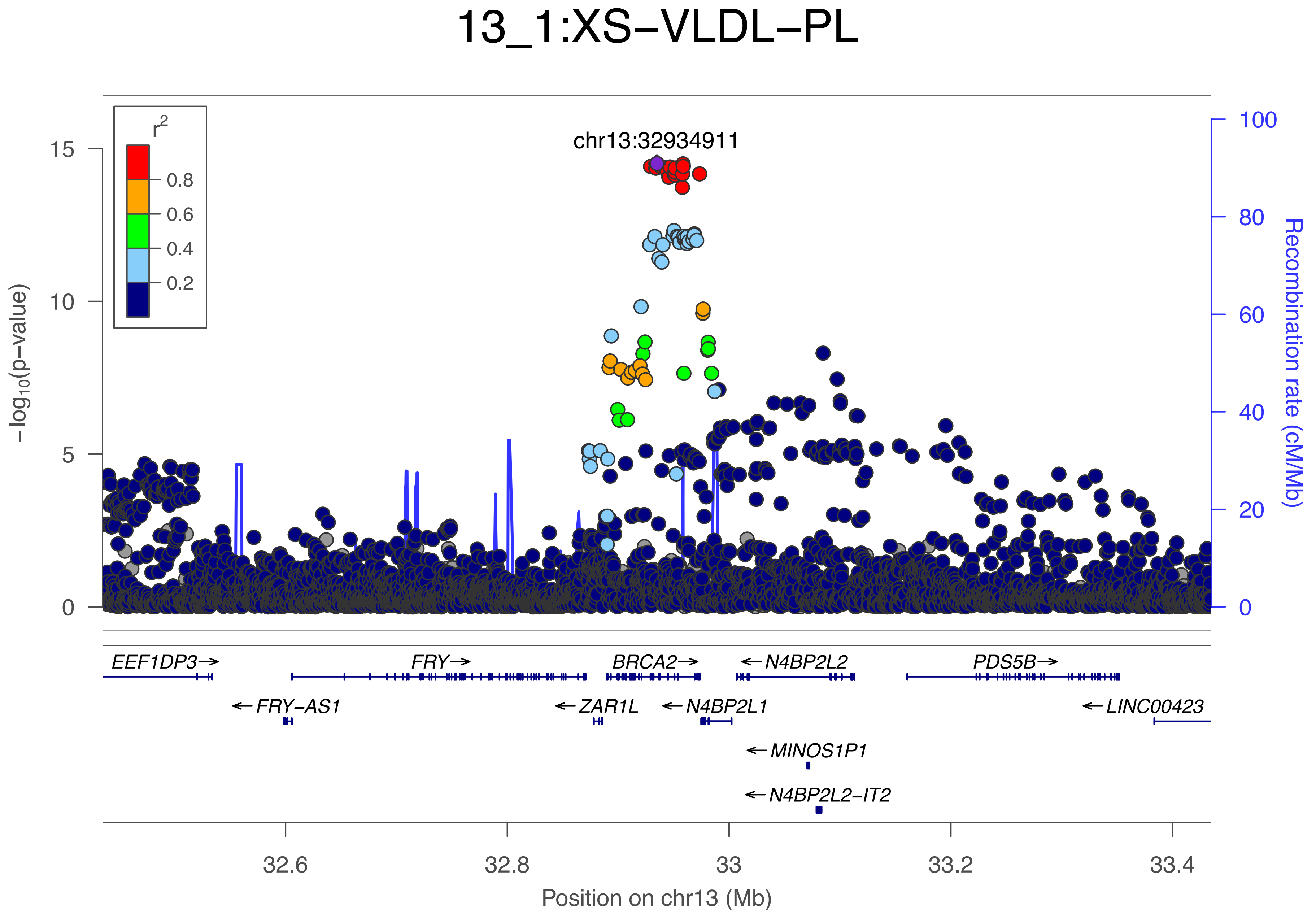

13_1:XS−VLDL−PL
r²
0.8
0.6
0.4
0.2
chr13:32934911
15
10
5
0
−log₁₀(p−value)
100
80
60
40
20
0
Recombination rate (cM/Mb)
EEF1DP3→
FRY→
BRCA2→
←N4BP2L2
PDS5B→
←FRY−AS1
←ZAR1L
←N4BP2L1
←LINC00423
←MINOS1P1
←N4BP2L2−IT2
32.6
32.8
33
33.2
33.4
Position on chr13 (Mb)

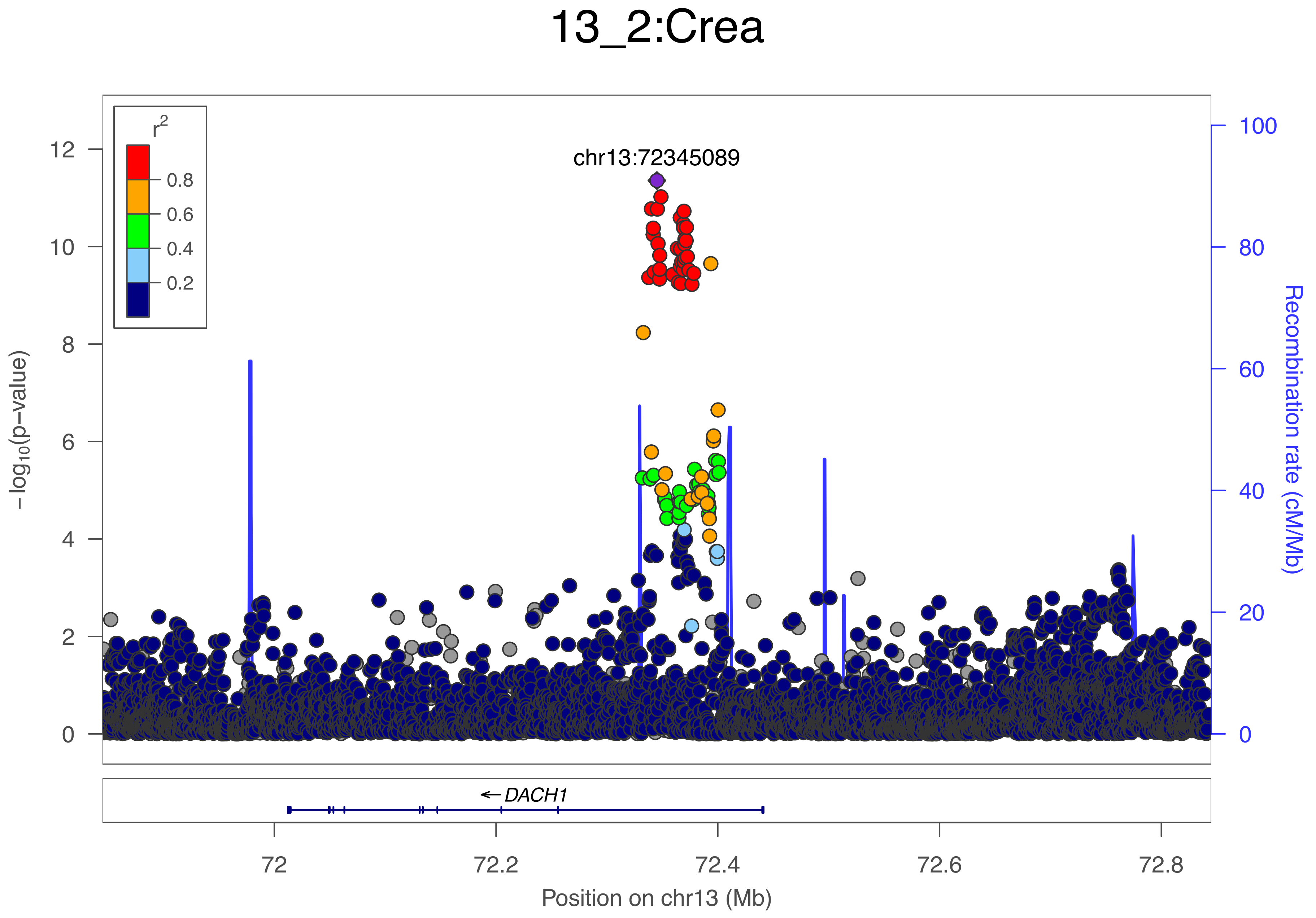

13_2:Crea
chr13:72345089
r²
0.8
0.6
0.4
0.2
−log10(p−value)
0
2
4
6
8
10
12
Recombination rate (cM/Mb)
0
20
40
60
80
100
← DACH1
72
72.2
72.4
72.6
72.8
Position on chr13 (Mb)

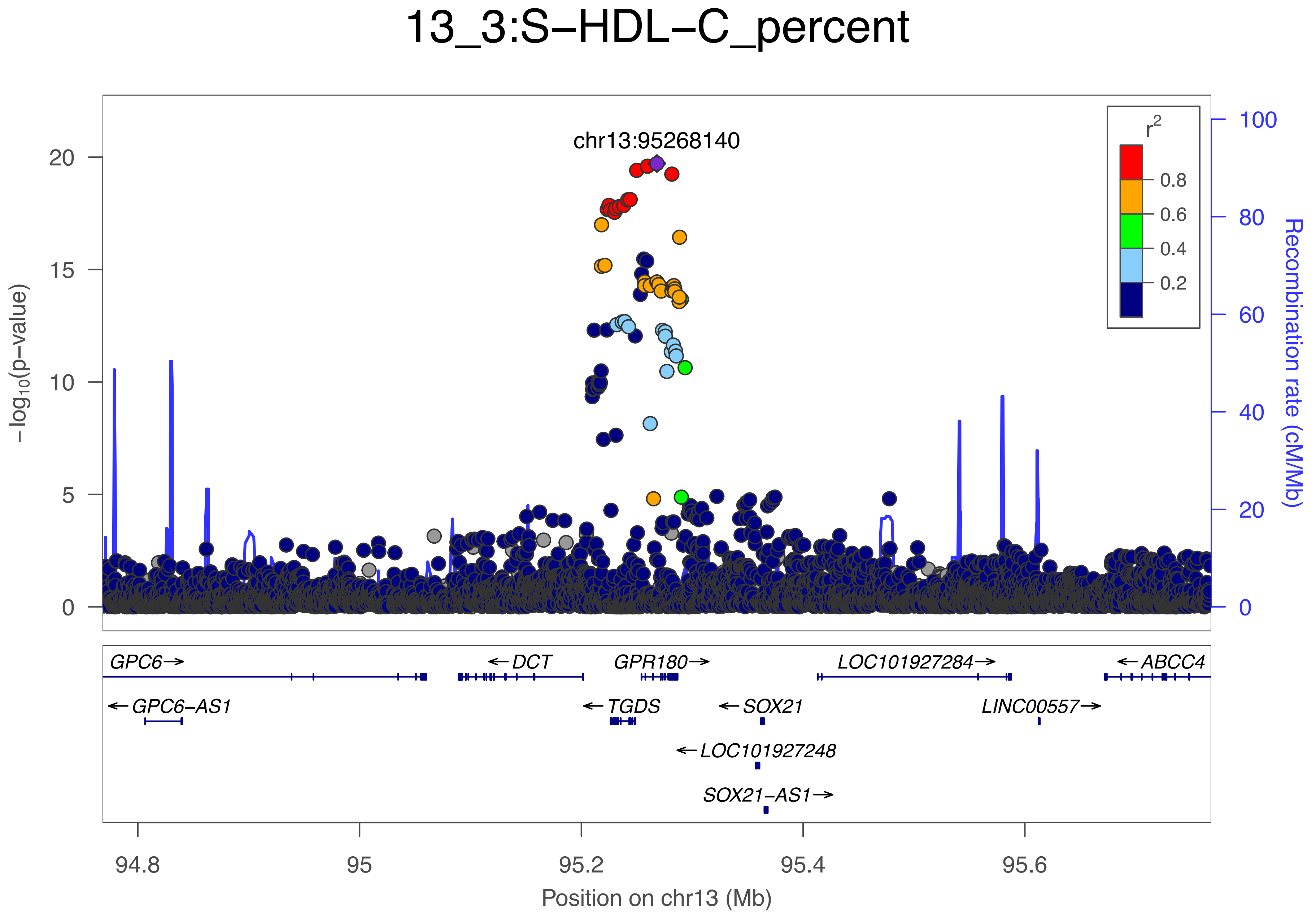

13_3:S−HDL−C_percent
chr13:95268140
r2
0.8
0.6
0.4
0.2
−log10(p−value)
Recombination rate (cM/Mb)
GPC6→
←DCT
GPR180→
LOC101927284→
←ABCC4
←GPC6−AS1
←TGDS
←SOX21
LINC00557→
←LOC101927248
SOX21−AS1→
Position on chr13 (Mb)

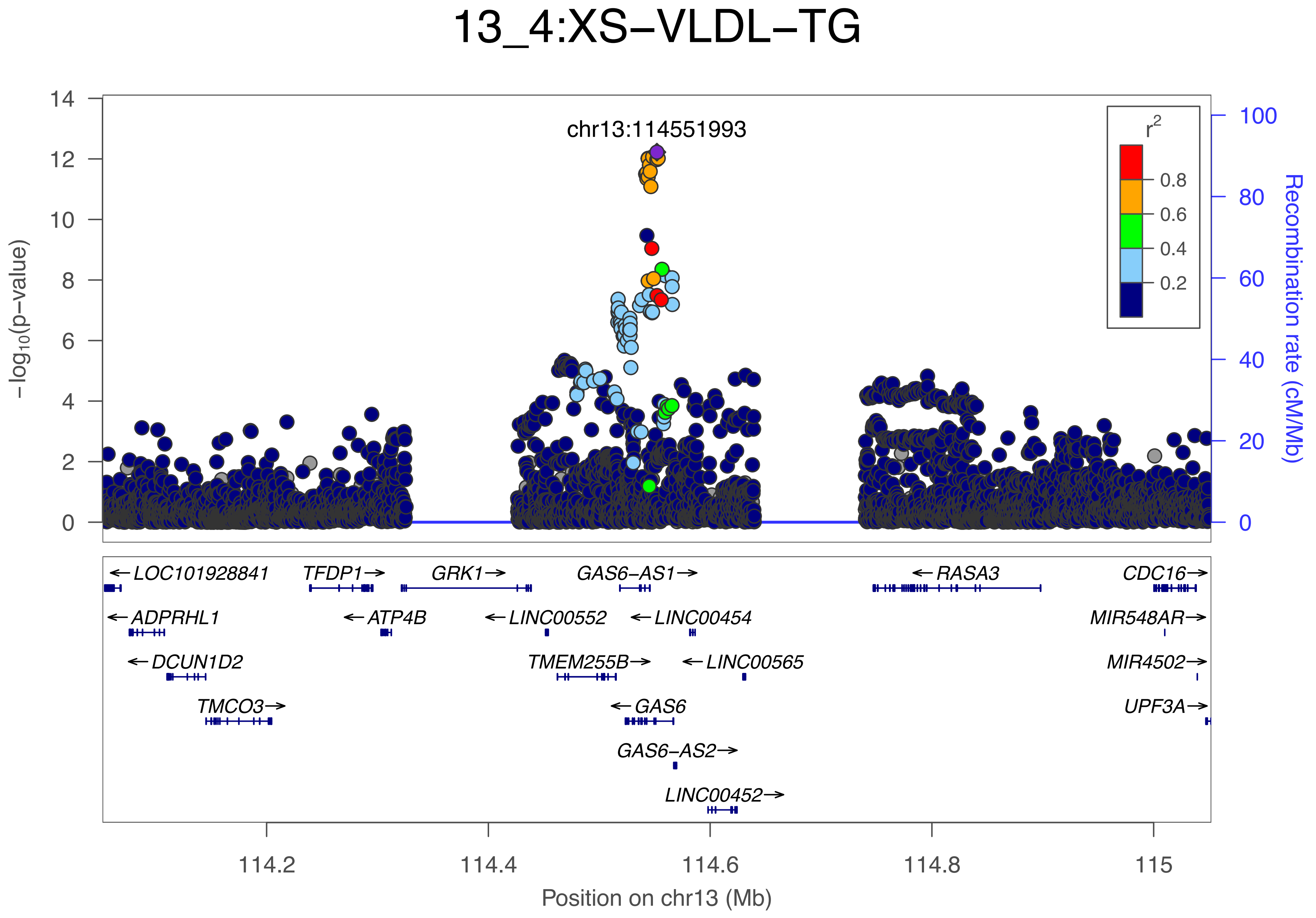
13_4:XS−VLDL−TG
chr13:114551993
−log10(p−value)
Recombination rate (cM/Mb)
r2
0.8
0.6
0.4
0.2
0
2
4
6
8
10
12
14
0
20
40
60
80
100
←LOC101928841
TFDP1→
GRK1→
GAS6−AS1→
←RASA3
CDC16→
←ADPRHL1
←ATP4B
←LINC00552
←LINC00454
MIR548AR→
←DCUN1D2
TMEM255B→
←LINC00565
MIR4502→
TMCO3→
←GAS6
UPF3A→
GAS6−AS2→
LINC00452→
114.2
114.4
114.6
114.8
115
Position on chr13 (Mb)

### 14_1:His

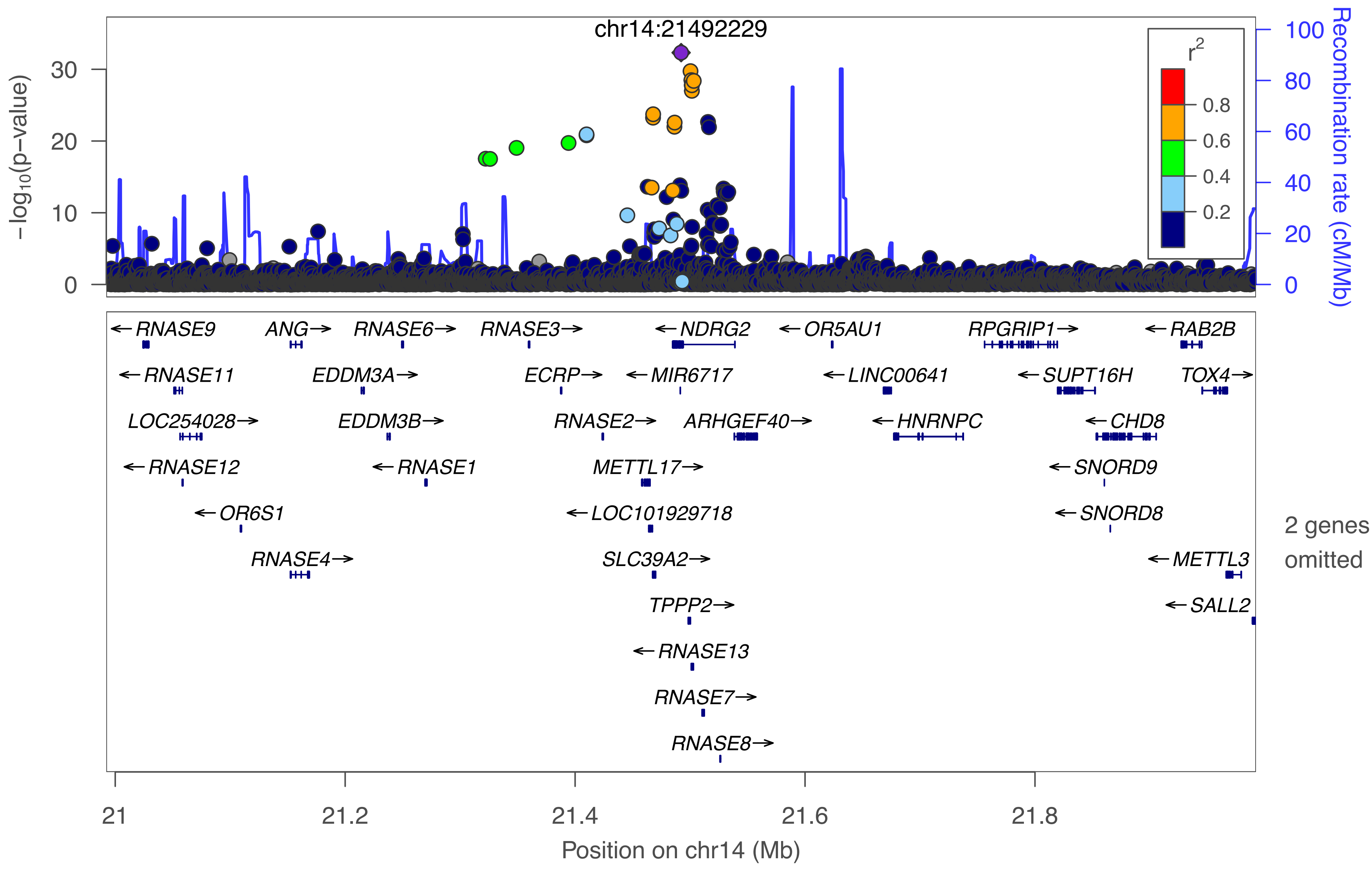

chr14:21492229
−log10(p−value)
0
10
20
30
Recombination rate (cM/Mb)
0
20
40
60
80
100
r2
0.8
0.6
0.4
0.2
RNASE9
ANG
RNASE6
RNASE3
NDRG2
OR5AU1
RPGRIP1
RAB2B
RNASE11
EDDM3A
ECRP
MIR6717
LINC00641
SUPT16H
TOX4
LOC254028
EDDM3B
RNASE2
ARHGEF40
HNRNPC
CHD8
RNASE12
RNASE1
METTL17
SNORD9
OR6S1
LOC101929718
SNORD8
RNASE4
SLC39A2
METTL3
TPPP2
SALL2
RNASE13
RNASE7
RNASE8
2 genes omitted
21
21.2
21.4
21.6
21.8
Position on chr14 (Mb)

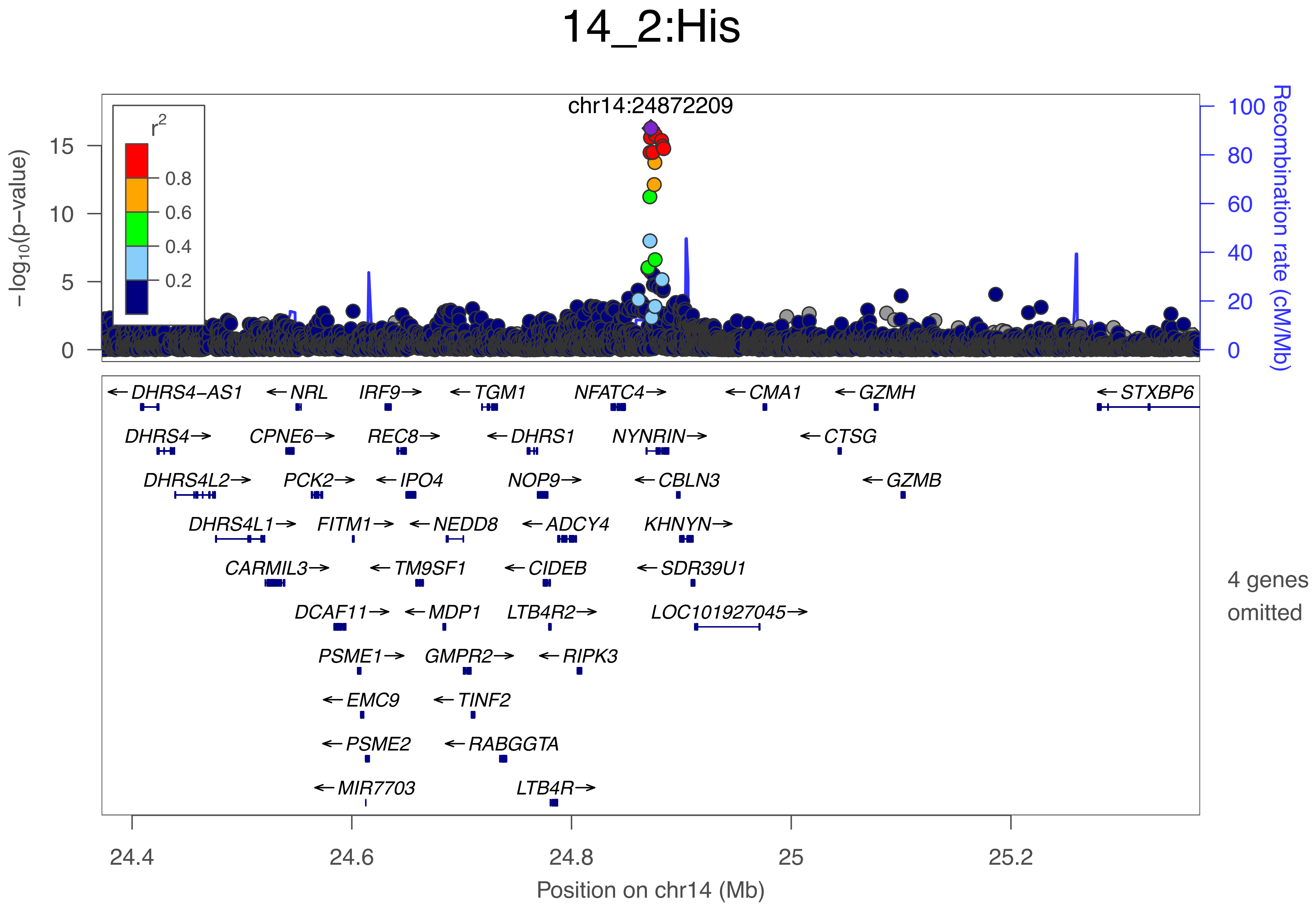

14_2:His
chr14:24872209
r2
0.8
0.6
0.4
0.2
−log10(p−value)
Recombination rate (cM/Mb)
DHRS4−AS1
NRL
IRF9
TGM1
NFATC4
CMA1
GZMH
STXBP6
DHRS4
CPNE6
REC8
DHRS1
NYNRIN
CTSG
DHRS4L2
PCK2
IPO4
NOP9
CBLN3
GZMB
DHRS4L1
FITM1
NEDD8
ADCY4
KHNYN
CARMIL3
TM9SF1
CIDEB
SDR39U1
DCAF11
MDP1
LTB4R2
LOC101927045
PSME1
GMPR2
RIPK3
EMC9
TINF2
PSME2
RABGGTA
MIR7703
LTB4R
4 genes omitted
Position on chr14 (Mb)

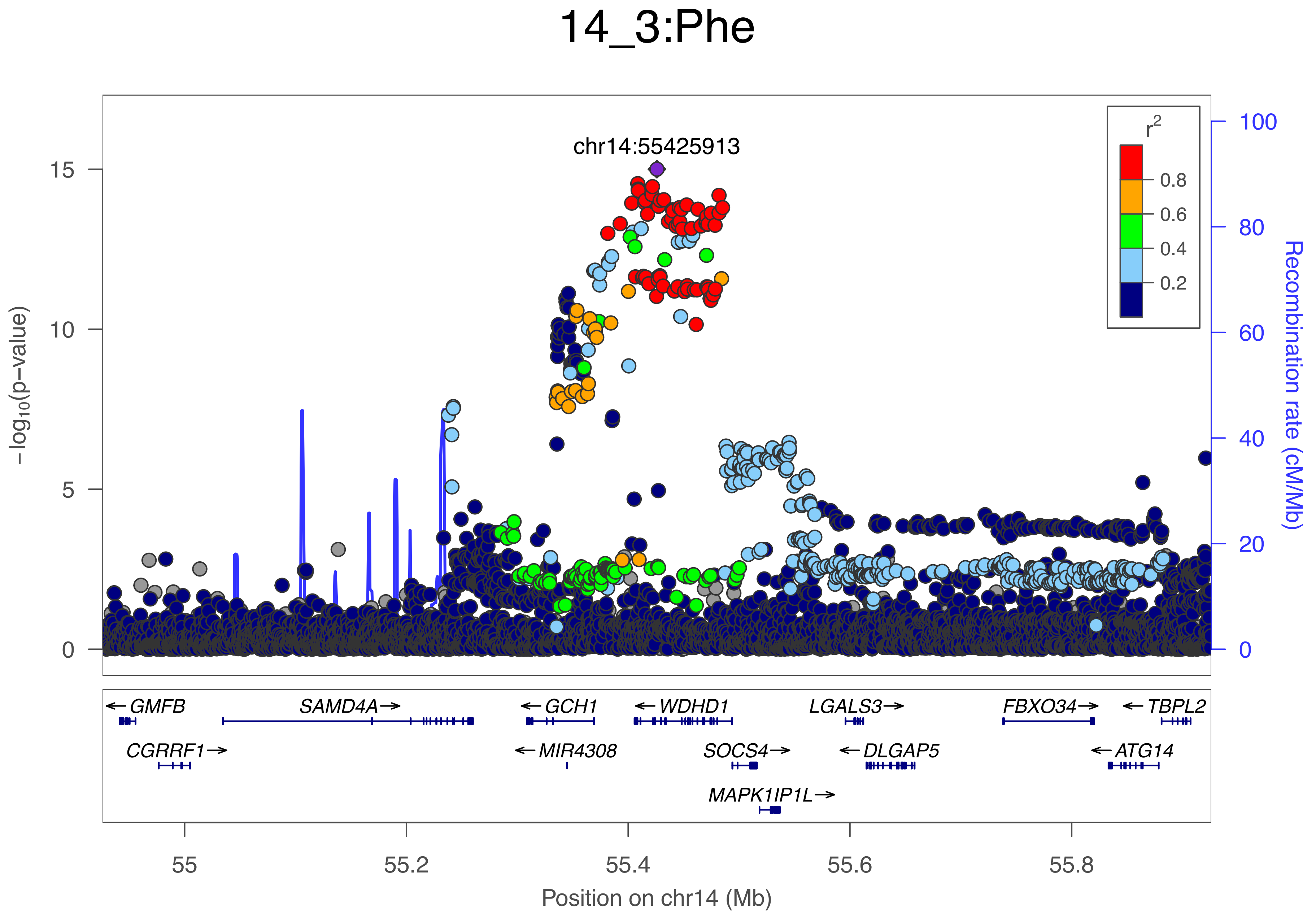
14_3:Phe
chr14:55425913
r²
0.8
0.6
0.4
0.2
15
10
5
0
−log10(p−value)
100
80
60
40
20
0
Recombination rate (cM/Mb)
← GMFB
SAMD4A →
← GCH1
← WDHD1
LGALS3 →
FBXO34 →
← TBPL2
CGRRF1 →
← MIR4308
SOCS4 →
← DLGAP5
← ATG14
MAPK1IP1L →
55
55.2
55.4
55.6
55.8
Position on chr14 (Mb)

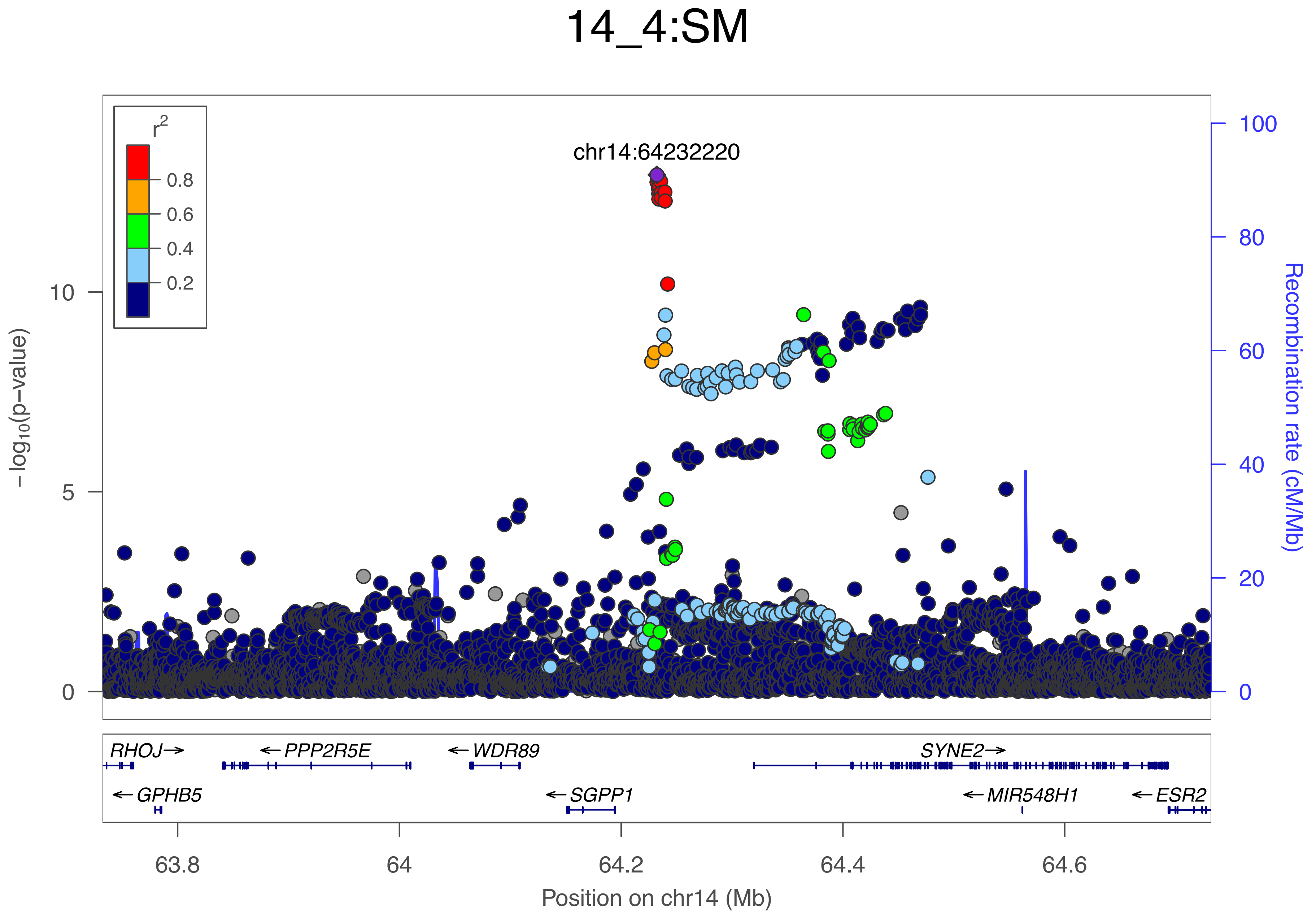

14_4:SM
chr14:64232220
r2
0.8
0.6
0.4
0.2
−log10(p−value)
10
5
0
Recombination rate (cM/Mb)
100
80
60
40
20
0
RHOJ→
←PPP2R5E
←WDR89
SYNE2→
←GPHB5
←SGPP1
←MIR548H1
←ESR2
63.8
64
64.2
64.4
64.6
Position on chr14 (Mb)

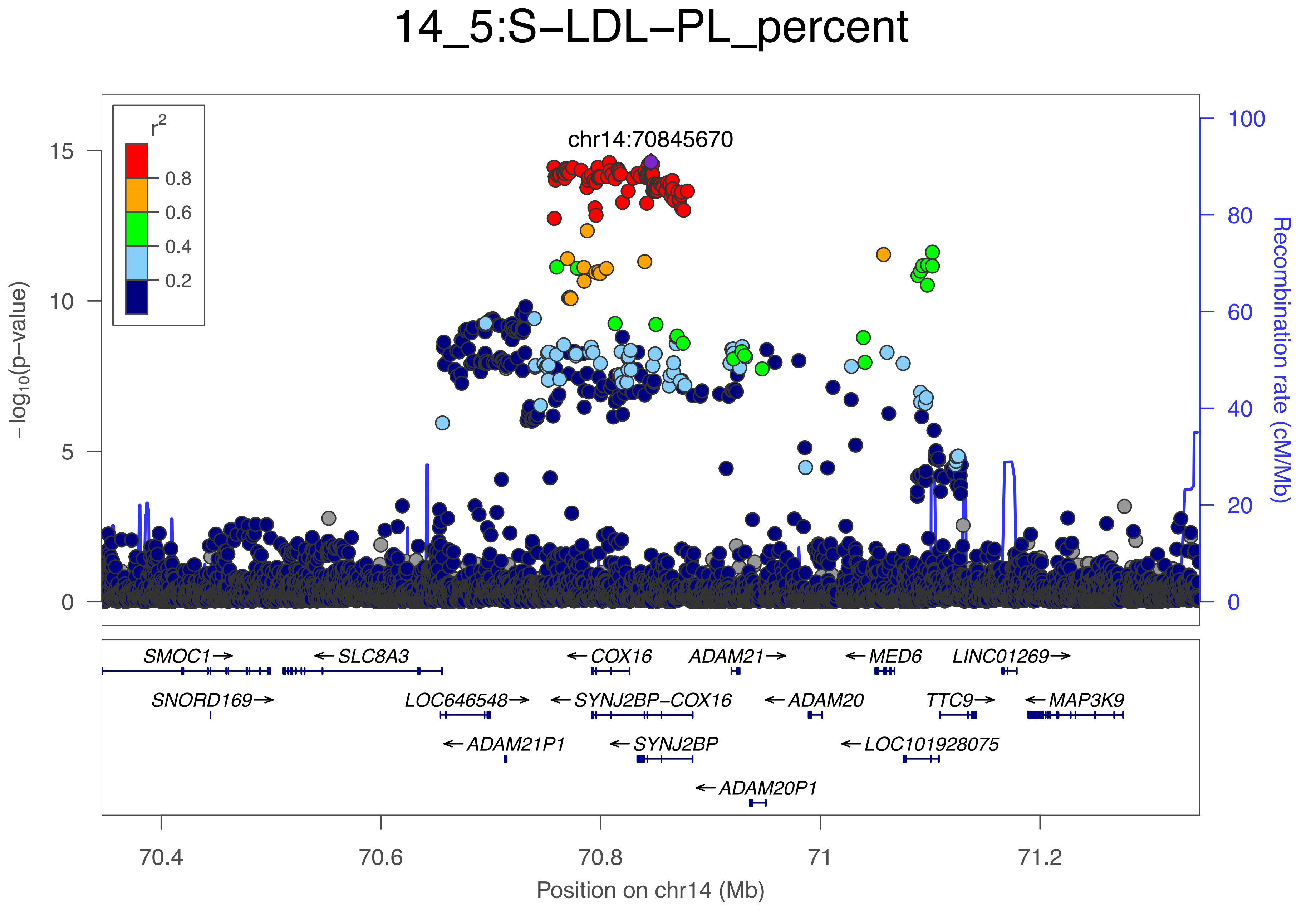

14_5:S−LDL−PL_percent
r2
0.8
0.6
0.4
0.2
chr14:70845670
−log10(p−value)
Recombination rate (cM/Mb)
0
5
10
15
0
20
40
60
80
100
SMOC1→
←SLC8A3
←COX16
ADAM21→
←MED6
LINC01269→
SNORD169→
LOC646548→
←SYNJ2BP−COX16
←ADAM20
TTC9→
←MAP3K9
←ADAM21P1
←SYNJ2BP
←LOC101928075
←ADAM20P1
70.4
70.6
70.8
71
71.2
Position on chr14 (Mb)

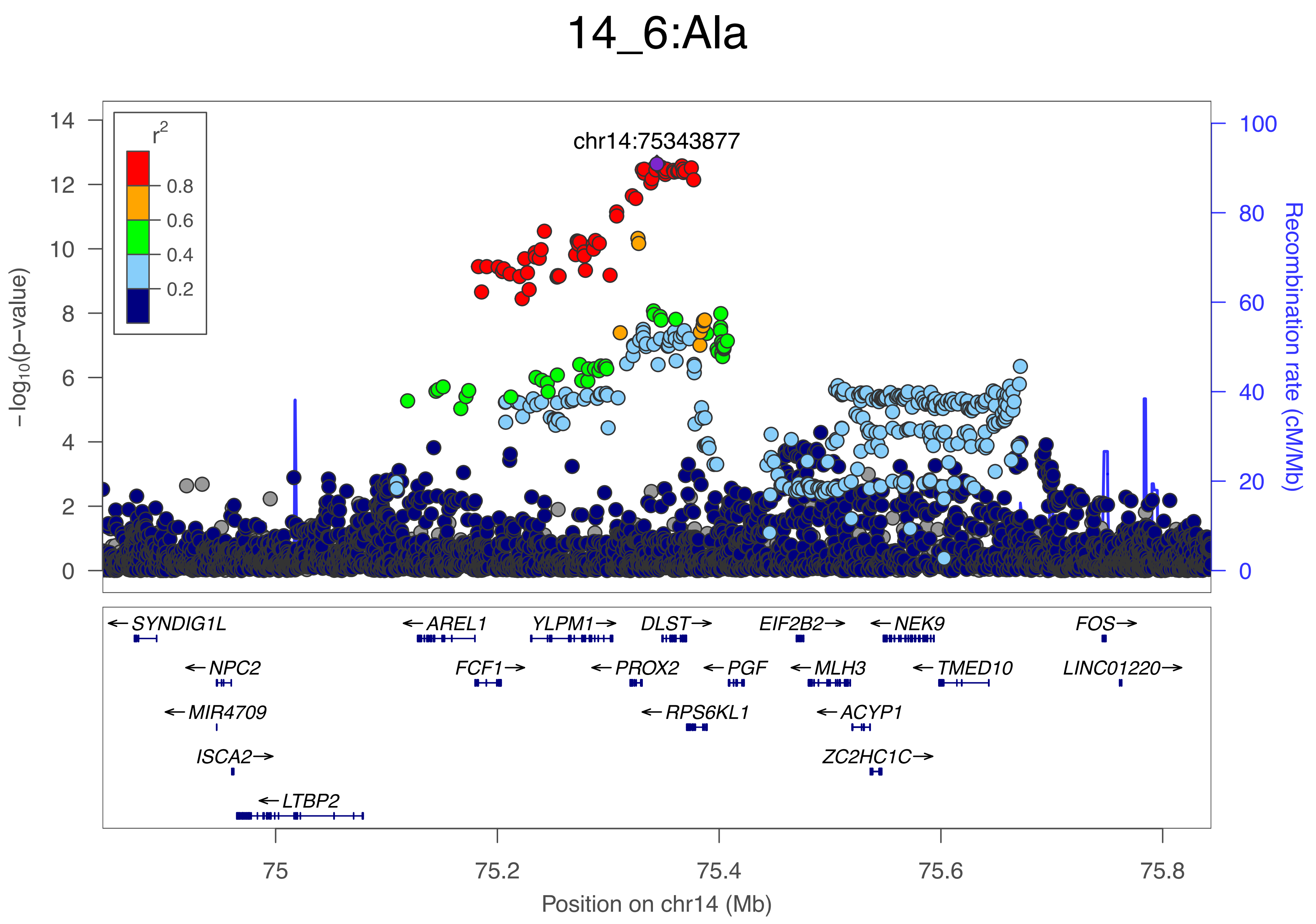
14_6:Ala
chr14:75343877
r2
0.8
0.6
0.4
0.2
−log10(p−value)
Recombination rate (cM/Mb)
SYNDIG1L
NPC2
MIR4709
ISCA2
LTBP2
AREL1
FCF1
YLPM1
PROX2
DLST
RPS6KL1
PGF
EIF2B2
MLH3
ACYP1
ZC2HC1C
NEK9
TMED10
FOS
LINC01220
Position on chr14 (Mb)

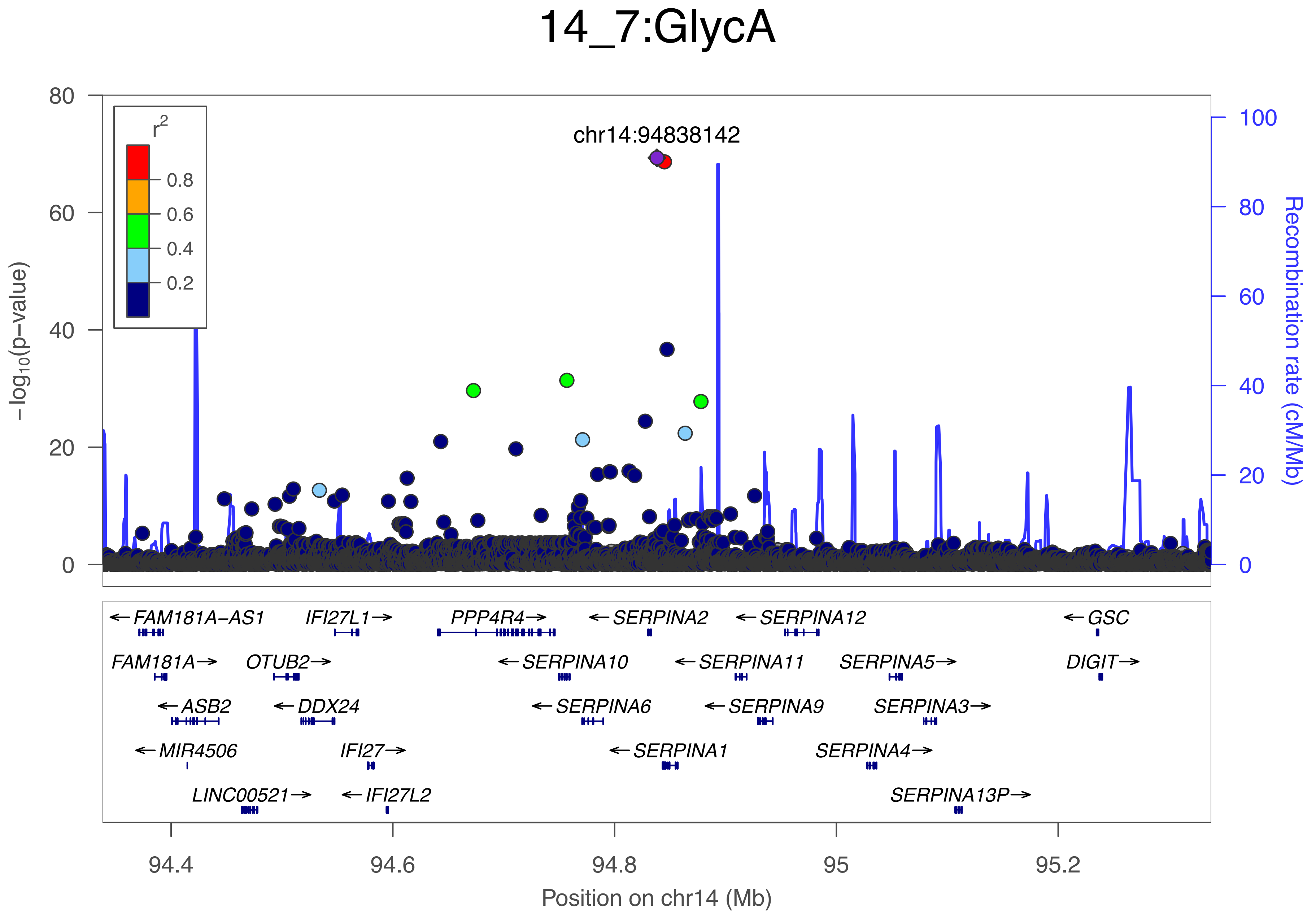

14_7:GlycA
chr14:94838142
r2
0.8
0.6
0.4
0.2
0
20
40
60
80
−log10(p−value)
100
Recombination rate (cM/Mb)
←FAM181A−AS1
IFI27L1→
PPP4R4→
←SERPINA2
←SERPINA12
←GSC
FAM181A→
OTUB2→
←SERPINA10
←SERPINA11
SERPINA5→
DIGIT→
←ASB2
←DDX24
←SERPINA6
←SERPINA9
SERPINA3→
←MIR4506
IFI27→
←SERPINA1
SERPINA4→
LINC00521→
←IFI27L2
SERPINA13P→
94.4
94.6
94.8
95
95.2
Position on chr14 (Mb)

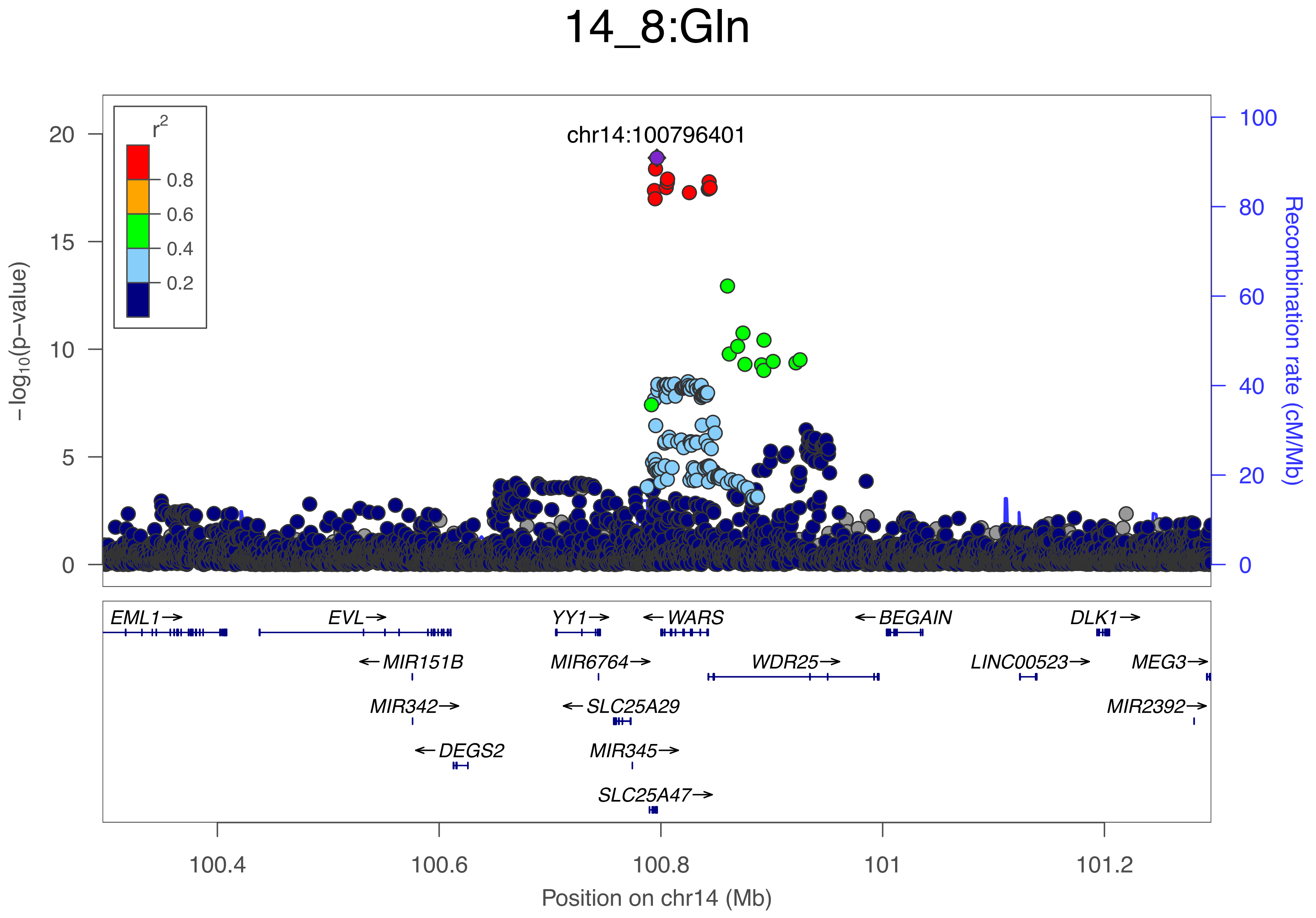

14_8:Gln
chr14:100796401
r2
0.8
0.6
0.4
0.2
−log10(p−value)
Recombination rate (cM/Mb)
EML1→
EVL→
YY1→
←WARS
←BEGAIN
DLK1→
←MIR151B
MIR6764→
WDR25→
LINC00523→
MEG3→
MIR342→
←SLC25A29
MIR2392→
←DEGS2
MIR345→
SLC25A47→
100.4
100.6
100.8
101
101.2
Position on chr14 (Mb)

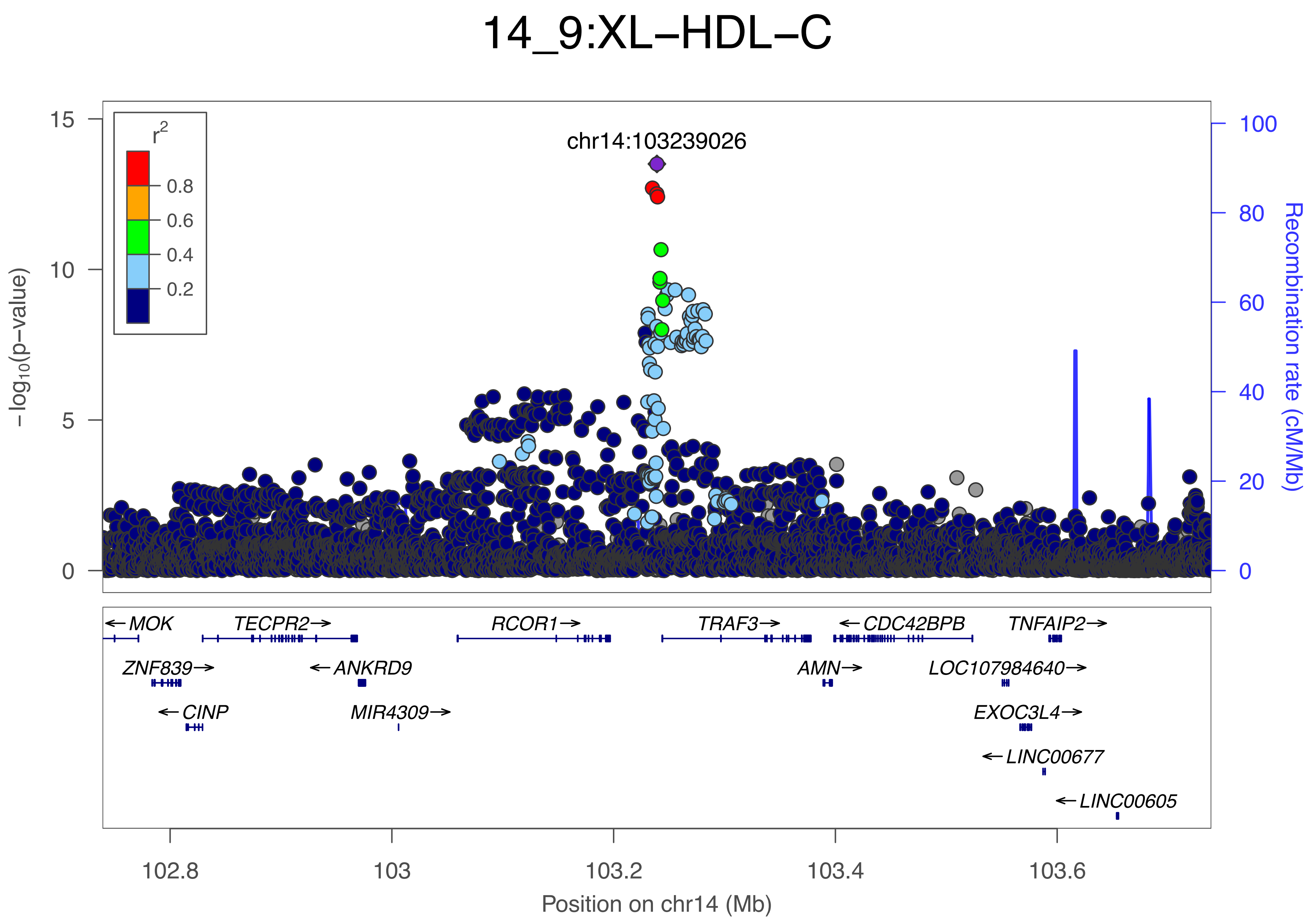

14_9:XL−HDL−C
chr14:103239026
r2
0.8
0.6
0.4
0.2
−log10(p−value)
Recombination rate (cM/Mb)
15
10
5
0
100
80
60
40
20
0
MOK
TECPR2
RCOR1
TRAF3
CDC42BPB
TNFAIP2
ZNF839
ANKRD9
AMN
LOC107984640
CINP
MIR4309
EXOC3L4
LINC00677
LINC00605
102.8
103
103.2
103.4
103.6
Position on chr14 (Mb)

### 14_10:LDL−D

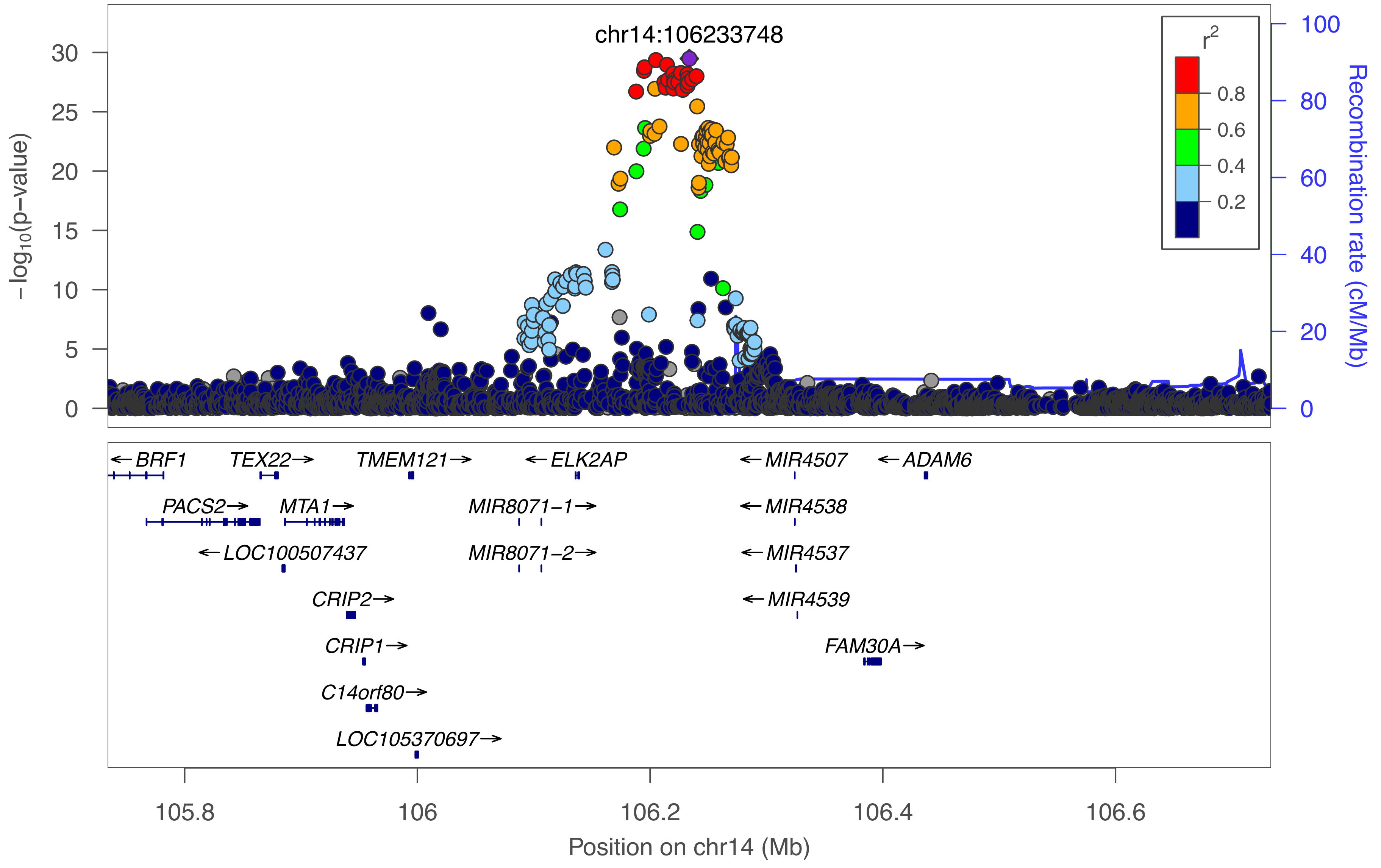

chr14:106233748
−log10(p−value)
30
25
20
15
10
5
0
r2
0.8
0.6
0.4
0.2
100
80
60
40
20
0
Recombination rate (cM/Mb)
← BRF1
TEX22 →
TMEM121 →
← ELK2AP
← MIR4507
← ADAM6
PACS2 →
MTA1 →
MIR8071−1 →
← MIR4538
← LOC100507437
MIR8071−2 →
← MIR4537
CRIP2 →
← MIR4539
CRIP1 →
FAM30A →
C14orf80 →
LOC105370697 →
105.8
106
106.2
106.4
106.6
Position on chr14 (Mb)

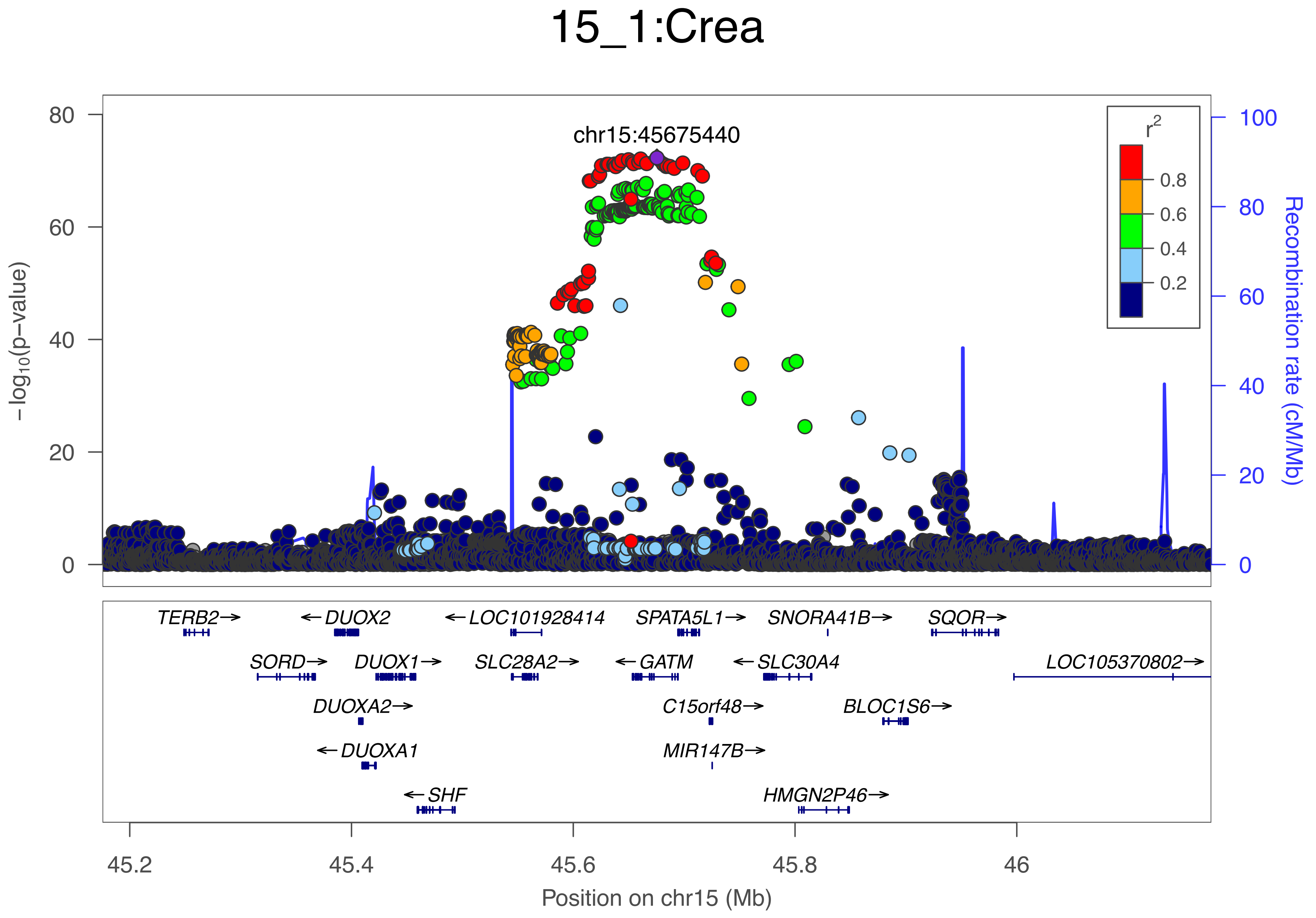

15_1:Crea
chr15:45675440
−log10(p−value)
Recombination rate (cM/Mb)
r2
0.8
0.6
0.4
0.2
0
20
40
60
80
100
TERB2→
←DUOX2
←LOC101928414
SPATA5L1→
SNORA41B→
SQOR→
SORD→
DUOX1→
SLC28A2→
←GATM
←SLC30A4
LOC105370802→
DUOXA2→
C15orf48→
BLOC1S6→
←DUOXA1
MIR147B→
←SHF
HMGN2P46→
45.2
45.4
45.6
45.8
46
Position on chr15 (Mb)

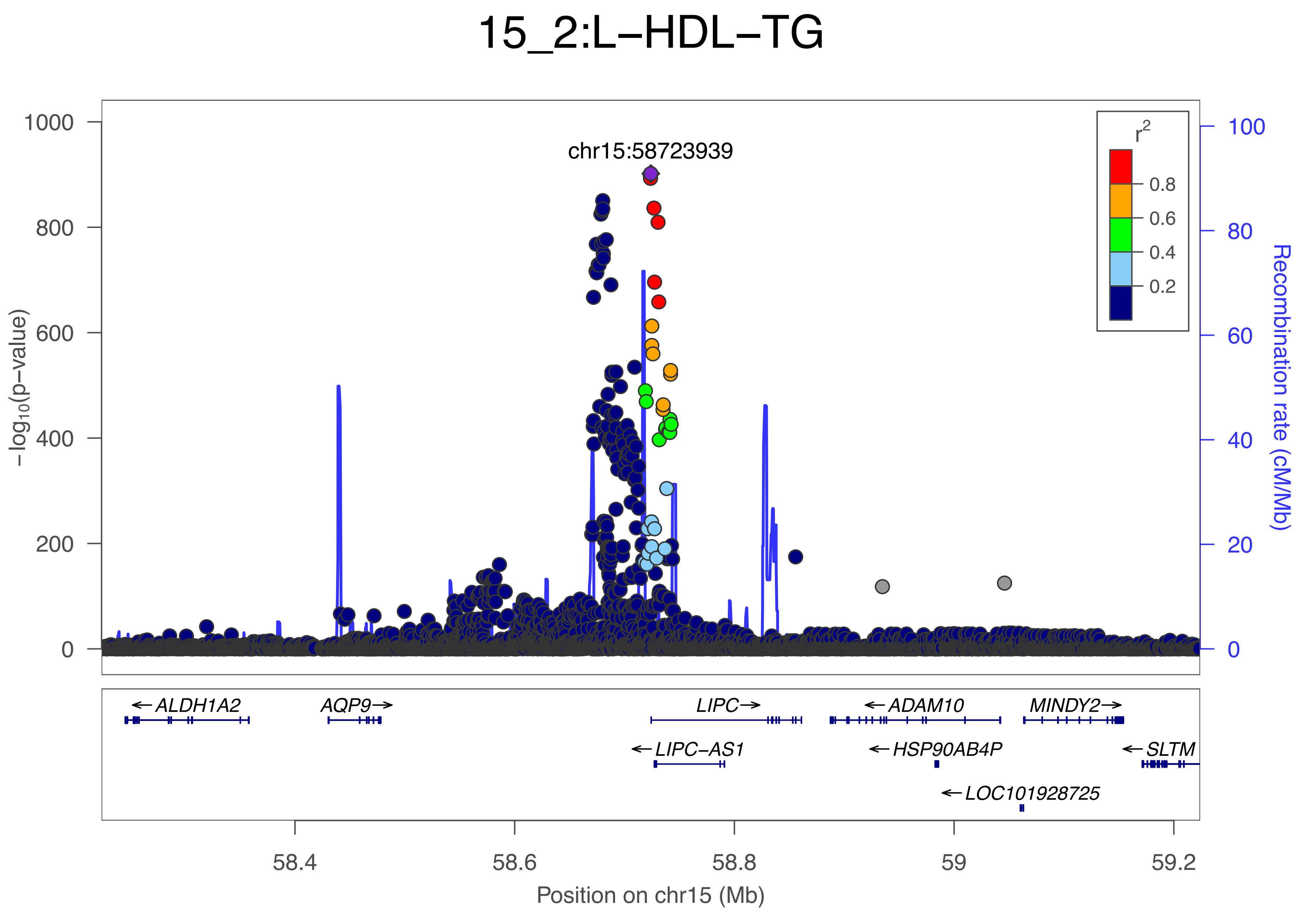

15_2:L−HDL−TG
chr15:58723939
−log10(p−value)
1000
800
600
400
200
0
Recombination rate (cM/Mb)
100
80
60
40
20
0
r2
0.8
0.6
0.4
0.2
ALDH1A2
AQP9
LIPC
ADAM10
MINDY2
LIPC−AS1
HSP90AB4P
SLTM
LOC101928725
58.4
58.6
58.8
59
59.2
Position on chr15 (Mb)

### 15_3:Crea

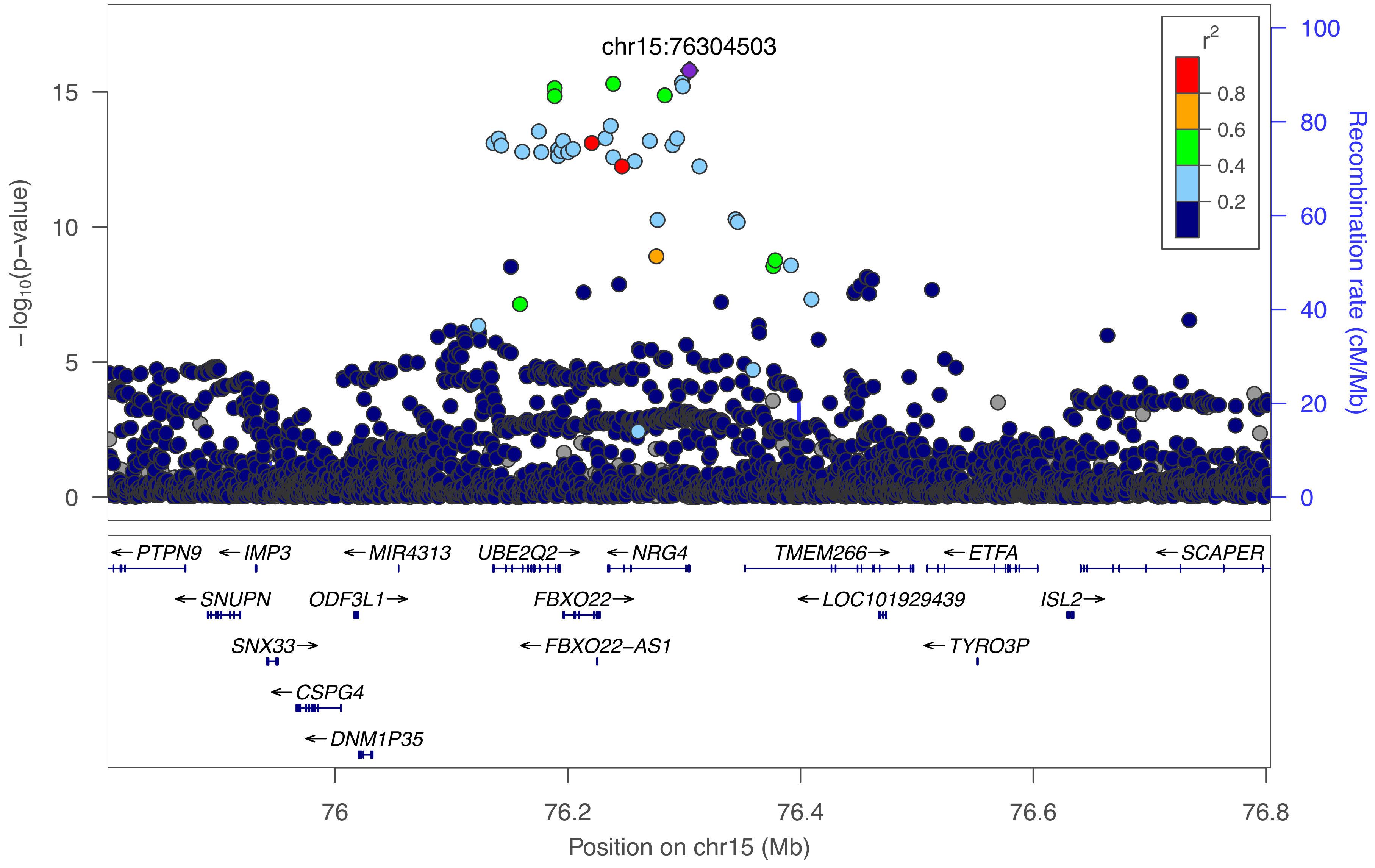
chr15:76304503
r²
0.8
0.6
0.4
0.2
−log10(p−value)
Recombination rate (cM/Mb)
PTPN9
IMP3
MIR4313
UBE2Q2
NRG4
TMEM266
ETFA
SCAPER
SNUPN
ODF3L1
FBXO22
LOC101929439
ISL2
SNX33
FBXO22−AS1
TYRO3P
CSPG4
DNM1P35
Position on chr15 (Mb)

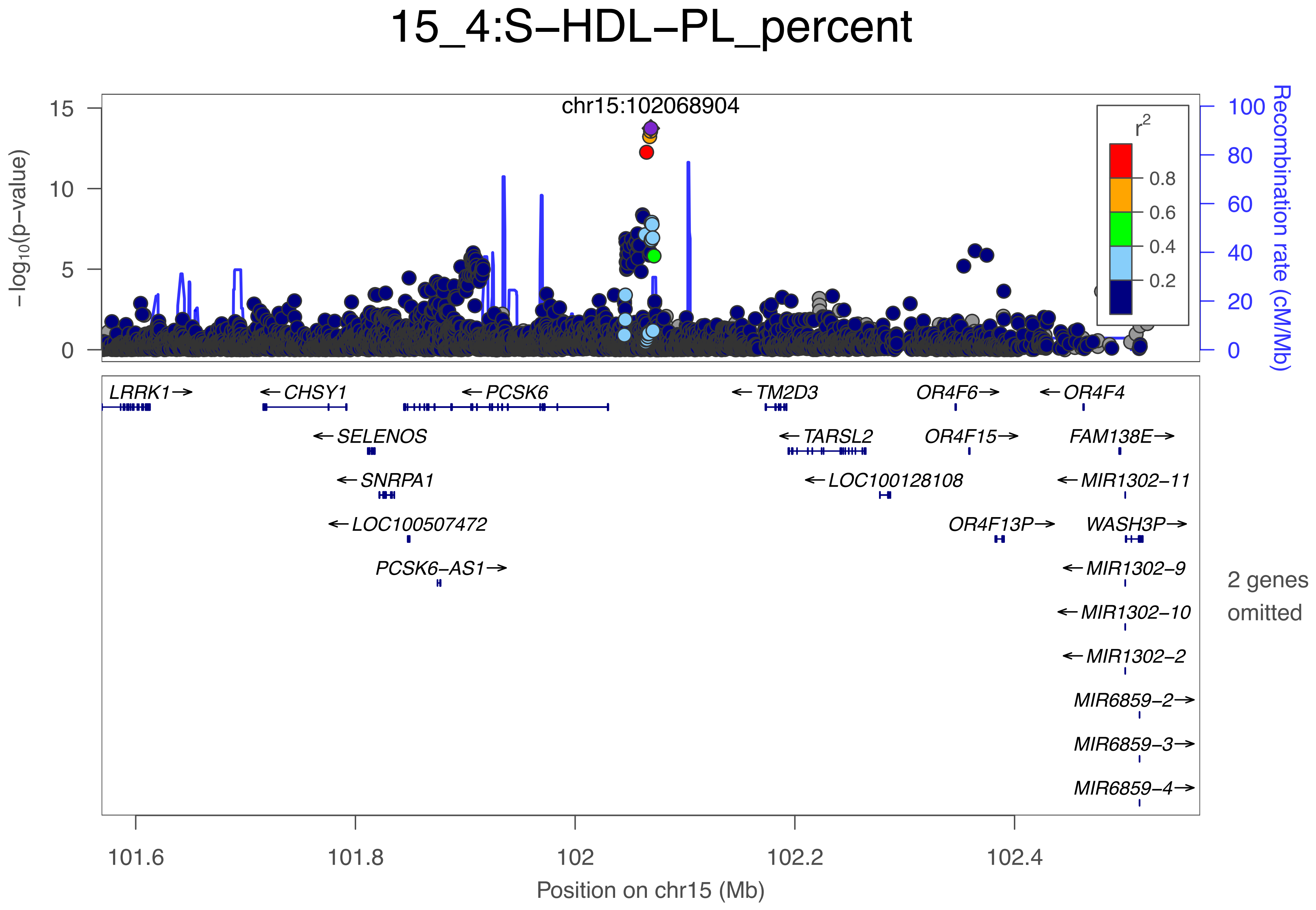

15_4:S−HDL−PL_percent
chr15:102068904
−log10(p−value)
Recombination rate (cM/Mb)
r2
0.8
0.6
0.4
0.2
LRRK1→
←CHSY1
←PCSK6
←TM2D3
OR4F6→
←OR4F4
←SELENOS
←TARSL2
OR4F15→
FAM138E→
←SNRPA1
←LOC100128108
←MIR1302−11
←LOC100507472
OR4F13P→
WASH3P→
PCSK6−AS1→
←MIR1302−9
←MIR1302−10
←MIR1302−2
MIR6859−2→
MIR6859−3→
MIR6859−4→
2 genes omitted
101.6
101.8
102
102.2
102.4
Position on chr15 (Mb)

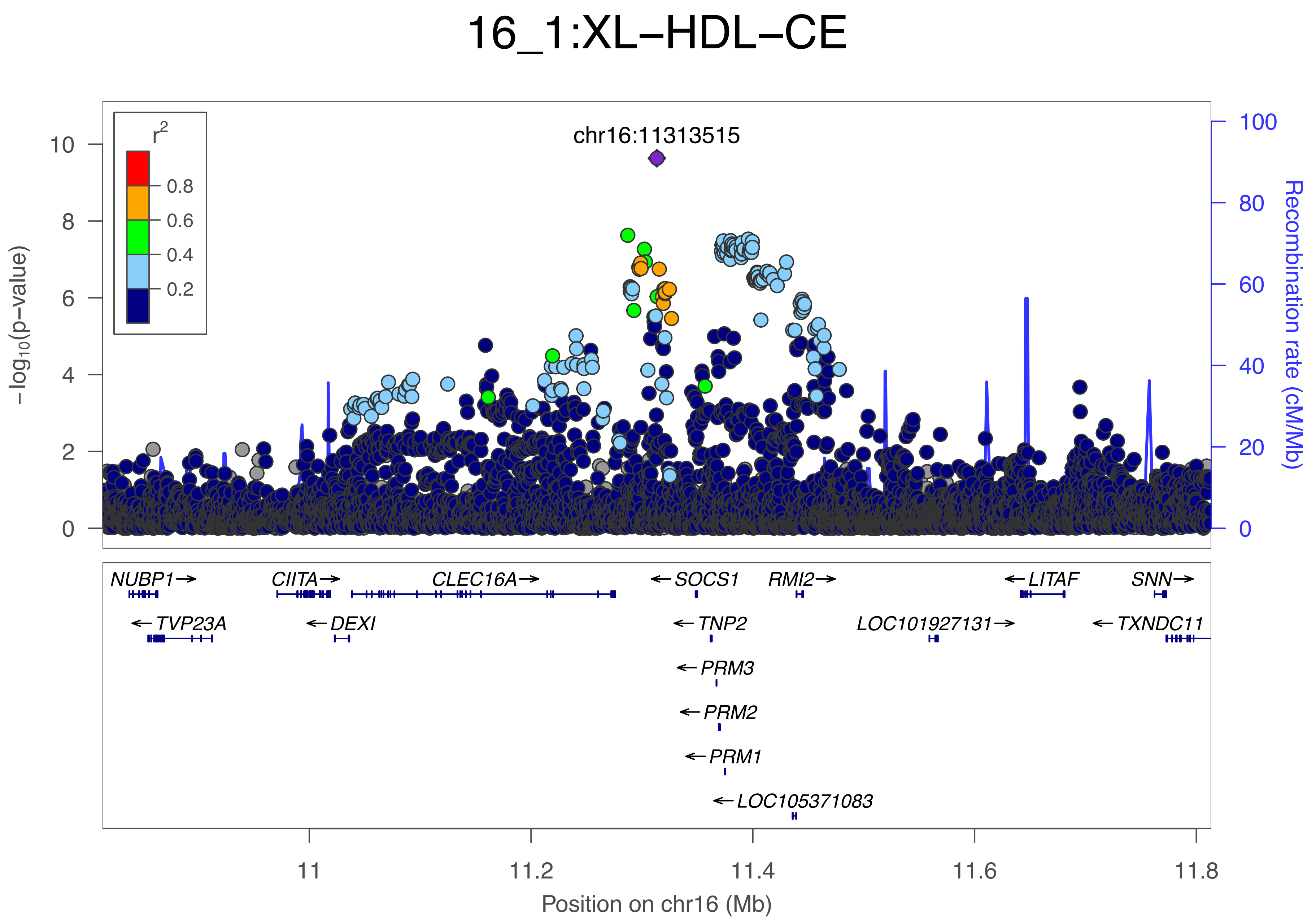

16_1:XL−HDL−CE
chr16:11313515
r2
0.8
0.6
0.4
0.2
−log10(p−value)
Recombination rate (cM/Mb)
0
2
4
6
8
10
0
20
40
60
80
100
NUBP1→
CIITA→
CLEC16A→
←SOCS1
RMI2→
←LITAF
SNN→
←TVP23A
←DEXI
←TNP2
LOC101927131→
←TXNDC11
←PRM3
←PRM2
←PRM1
←LOC105371083
11
11.2
11.4
11.6
11.8
Position on chr16 (Mb)

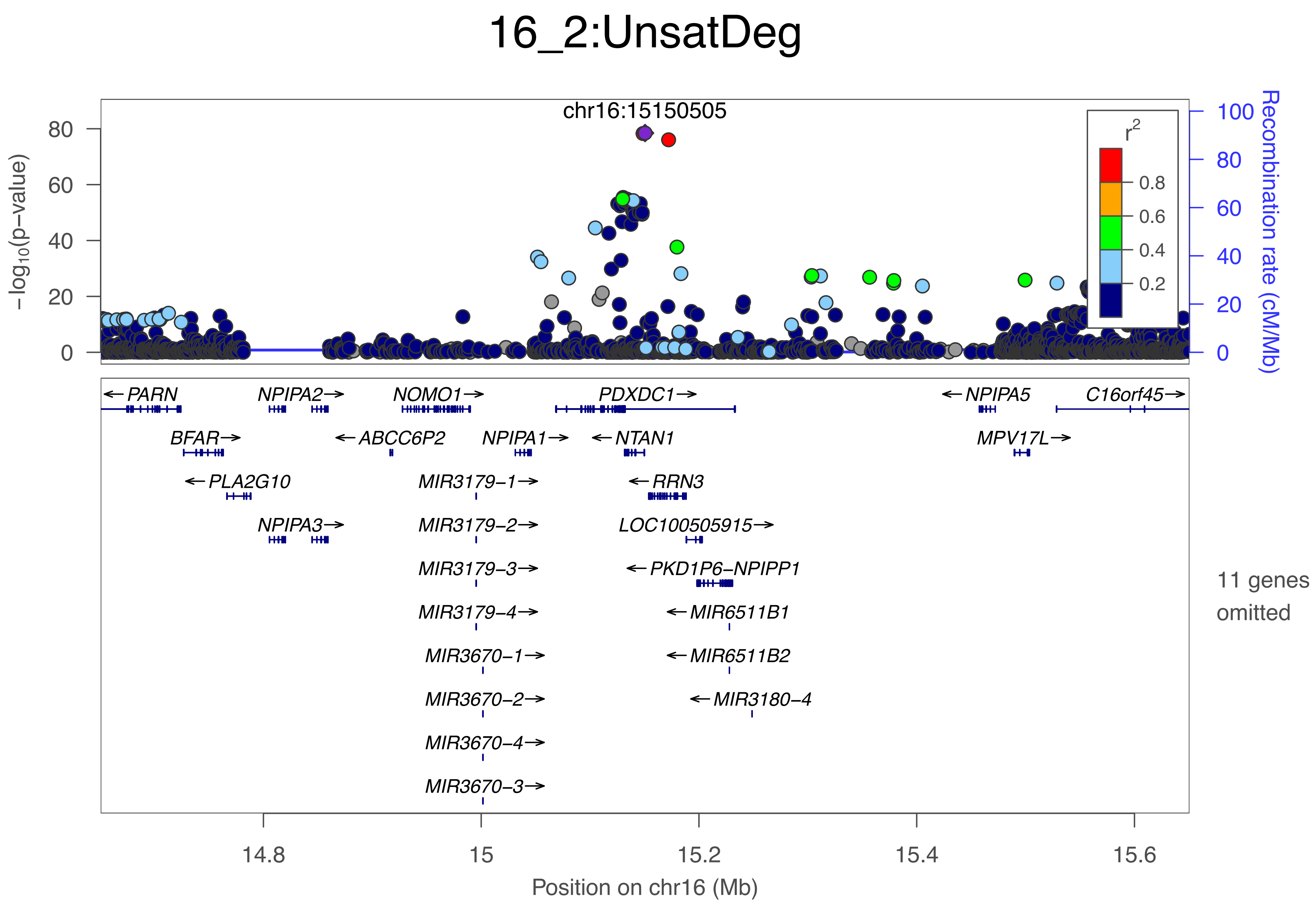

16_2:UnsatDeg
chr16:15150505
−log10(p−value)
Recombination rate (cM/Mb)
r2
0.8
0.6
0.4
0.2
80
60
40
20
0
100
80
60
40
20
0
PARN
NPIPA2
NOMO1
PDXDC1
NPIPA5
C16orf45
BFAR
ABCC6P2
NPIPA1
NTAN1
MPV17L
PLA2G10
MIR3179−1
RRN3
NPIPA3
MIR3179−2
LOC100505915
MIR3179−3
PKD1P6−NPIPP1
MIR3179−4
MIR6511B1
MIR3670−1
MIR6511B2
MIR3670−2
MIR3180−4
MIR3670−4
MIR3670−3
11 genes omitted
14.8
15
15.2
15.4
15.6
Position on chr16 (Mb)

### 16_3:Crea

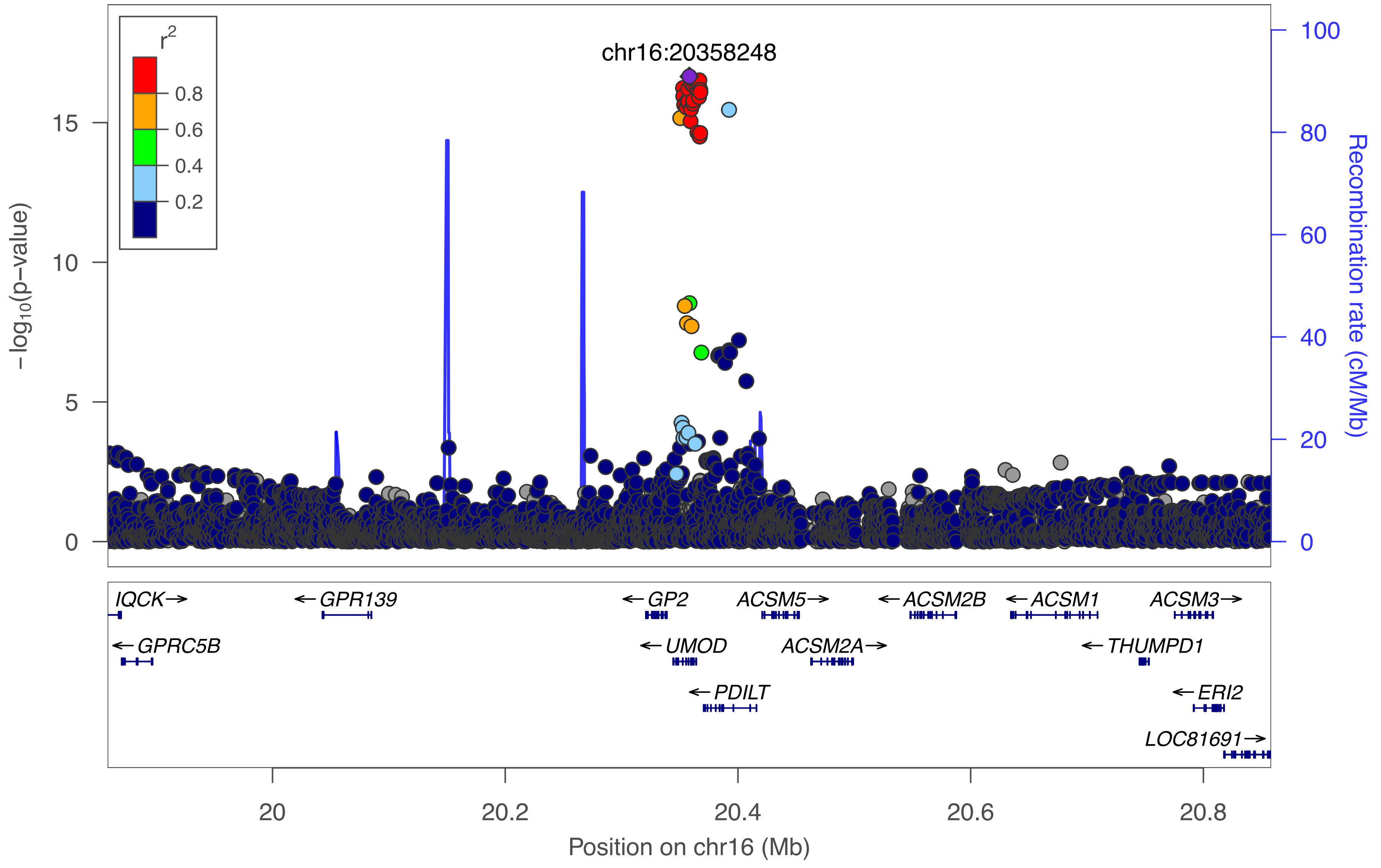

chr16:20358248
r2
0.8
0.6
0.4
0.2
15
10
5
0
−log10(p−value)
100
80
60
40
20
0
Recombination rate (cM/Mb)
IQCK→
←GPRC5B
←GPR139
←GP2
←UMOD
←PDILT
ACSM5→
ACSM2A→
←ACSM2B
←ACSM1
←THUMPD1
ACSM3→
←ERI2
LOC81691→
20
20.2
20.4
20.6
20.8
Position on chr16 (Mb)

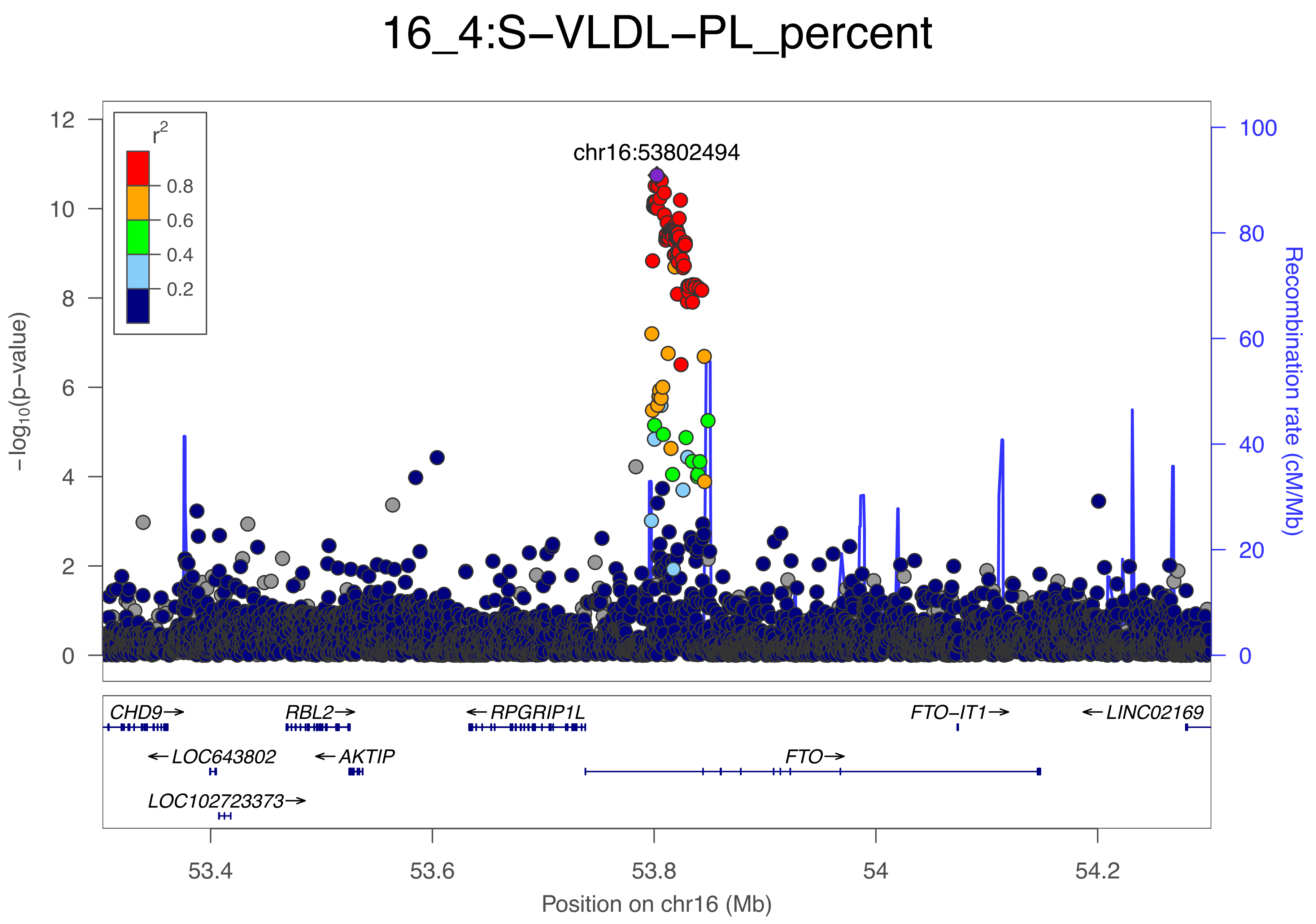

16_4:S−VLDL−PL_percent
chr16:53802494
r2
0.8
0.6
0.4
0.2
−log10(p−value)
Recombination rate (cM/Mb)
CHD9→
RBL2→
←RPGRIP1L
FTO−IT1→
←LINC02169
←LOC643802
←AKTIP
FTO→
LOC102723373→
53.4
53.6
53.8
54
54.2
Position on chr16 (Mb)

### 16_5:HDL−C

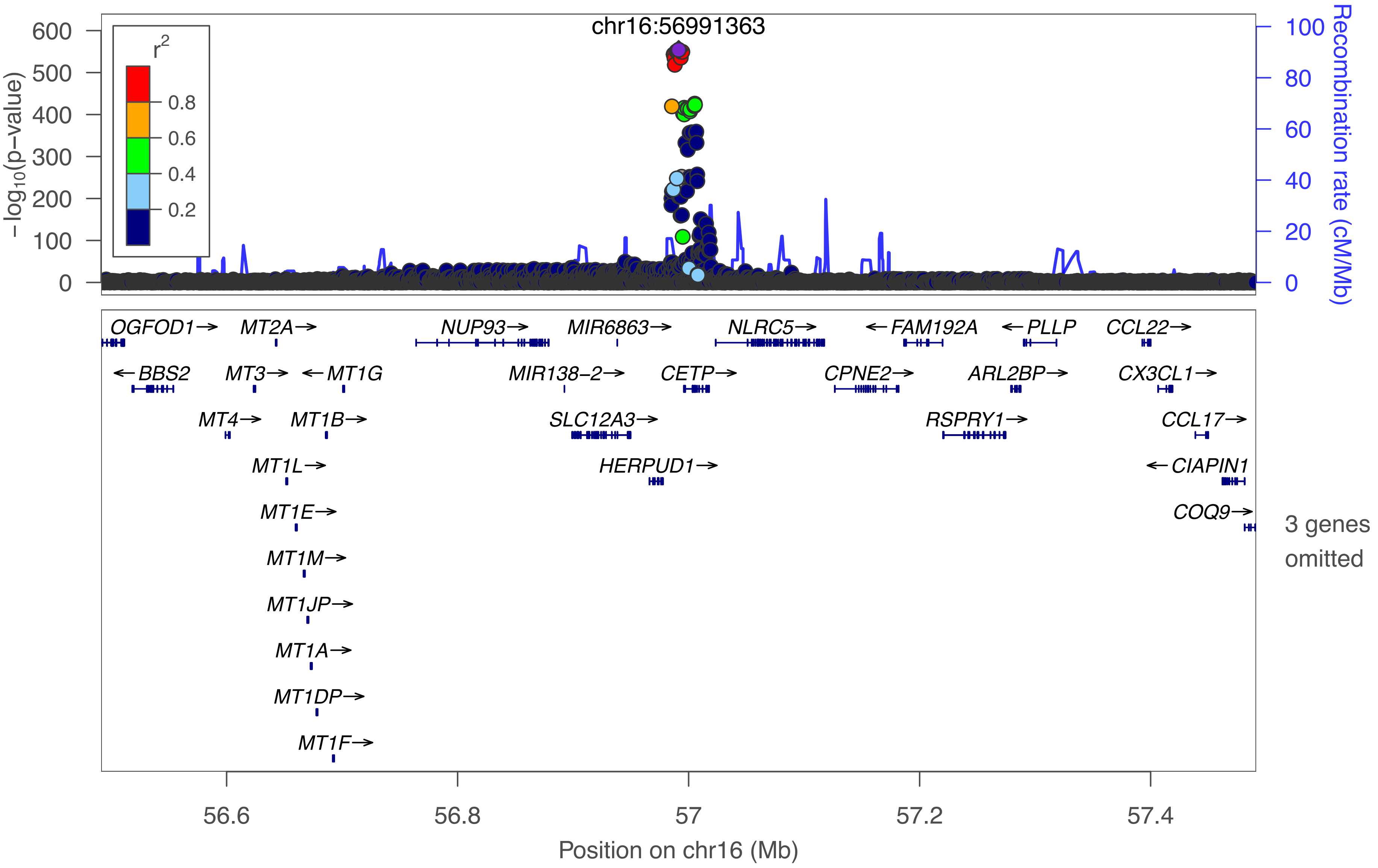

chr16:56991363
r²
0.8
0.6
0.4
0.2
−log10(p−value)
600
500
400
300
200
100
0
Recombination rate (cM/Mb)
100
80
60
40
20
0
OGFOD1→
MT2A→
NUP93→
MIR6863→
NLRC5→
←FAM192A
←PLLP
CCL22→
←BBS2
MT3→
←MT1G
MIR138−2→
CETP→
CPNE2→
ARL2BP→
CX3CL1→
MT4→
MT1B→
SLC12A3→
RSPRY1→
CCL17→
MT1L→
HERPUD1→
←CIAPIN1
MT1E→
COQ9→
MT1M→
MT1JP→
MT1A→
MT1DP→
MT1F→
3 genes omitted
56.6
56.8
57
57.2
57.4
Position on chr16 (Mb)

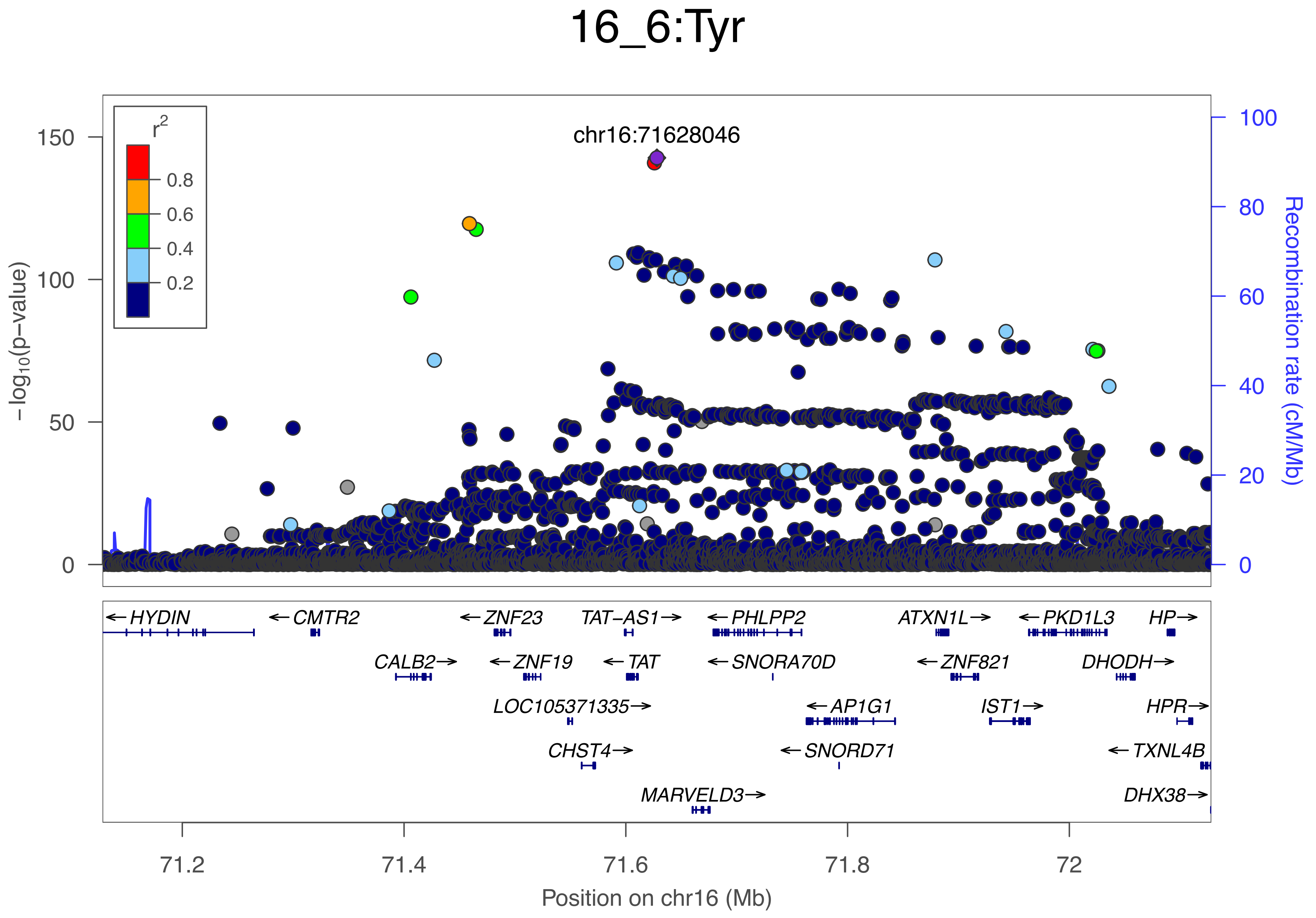

16_6:Tyr
chr16:71628046
r2
0.8
0.6
0.4
0.2
150
100
50
0
−log10(p−value)
100
80
60
40
20
0
Recombination rate (cM/Mb)
←HYDIN
←CMTR2
←ZNF23
TAT−AS1→
←PHLPP2
ATXN1L→
←PKD1L3
HP→
CALB2→
←ZNF19
←TAT
←SNORA70D
←ZNF821
DHODH→
LOC105371335→
←AP1G1
IST1→
HPR→
CHST4→
←SNORD71
←TXNL4B
MARVELD3→
DHX38→
71.2
71.4
71.6
71.8
72
Position on chr16 (Mb)

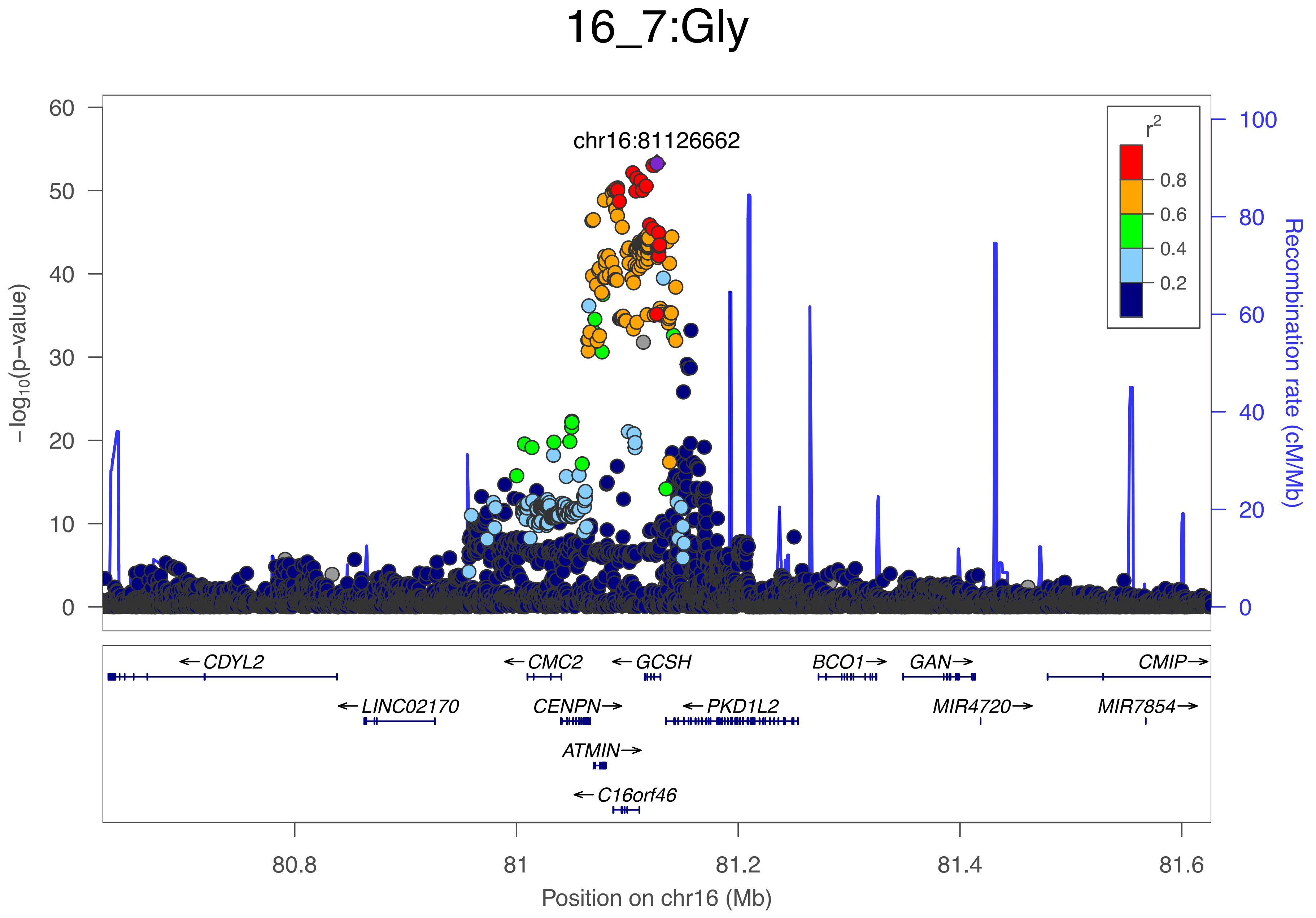

16_7:Gly
chr16:81126662
r2
0.8
0.6
0.4
0.2
60
50
40
30
20
10
0
−log10(p−value)
100
80
60
40
20
0
Recombination rate (cM/Mb)
←CDYL2
←LINC02170
←CMC2
CENPN→
ATMIN→
←C16orf46
←GCSH
←PKD1L2
BCO1→
GAN→
MIR4720→
CMIP→
MIR7854→
80.8
81
81.2
81.4
81.6
Position on chr16 (Mb)

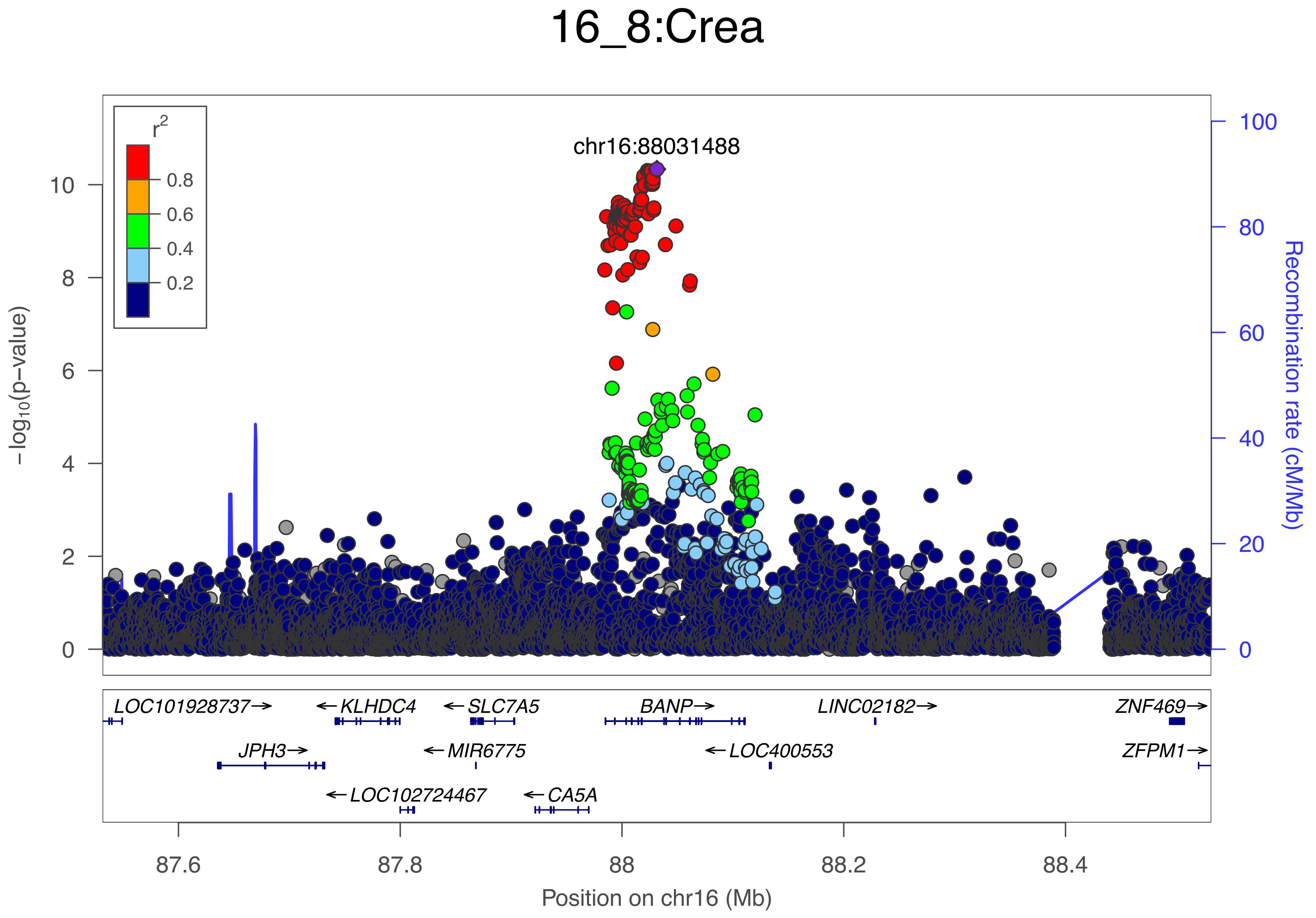
16_8:Crea
chr16:88031488
r2
0.8
0.6
0.4
0.2
−log10(p−value)
Recombination rate (cM/Mb)
0
2
4
6
8
10
0
20
40
60
80
100
LOC101928737→
←KLHDC4
←SLC7A5
BANP→
LINC02182→
ZNF469→
JPH3→
←MIR6775
←LOC400553
ZFPM1→
←LOC102724467
←CA5A
87.6
87.8
88
88.2
88.4
Position on chr16 (Mb)

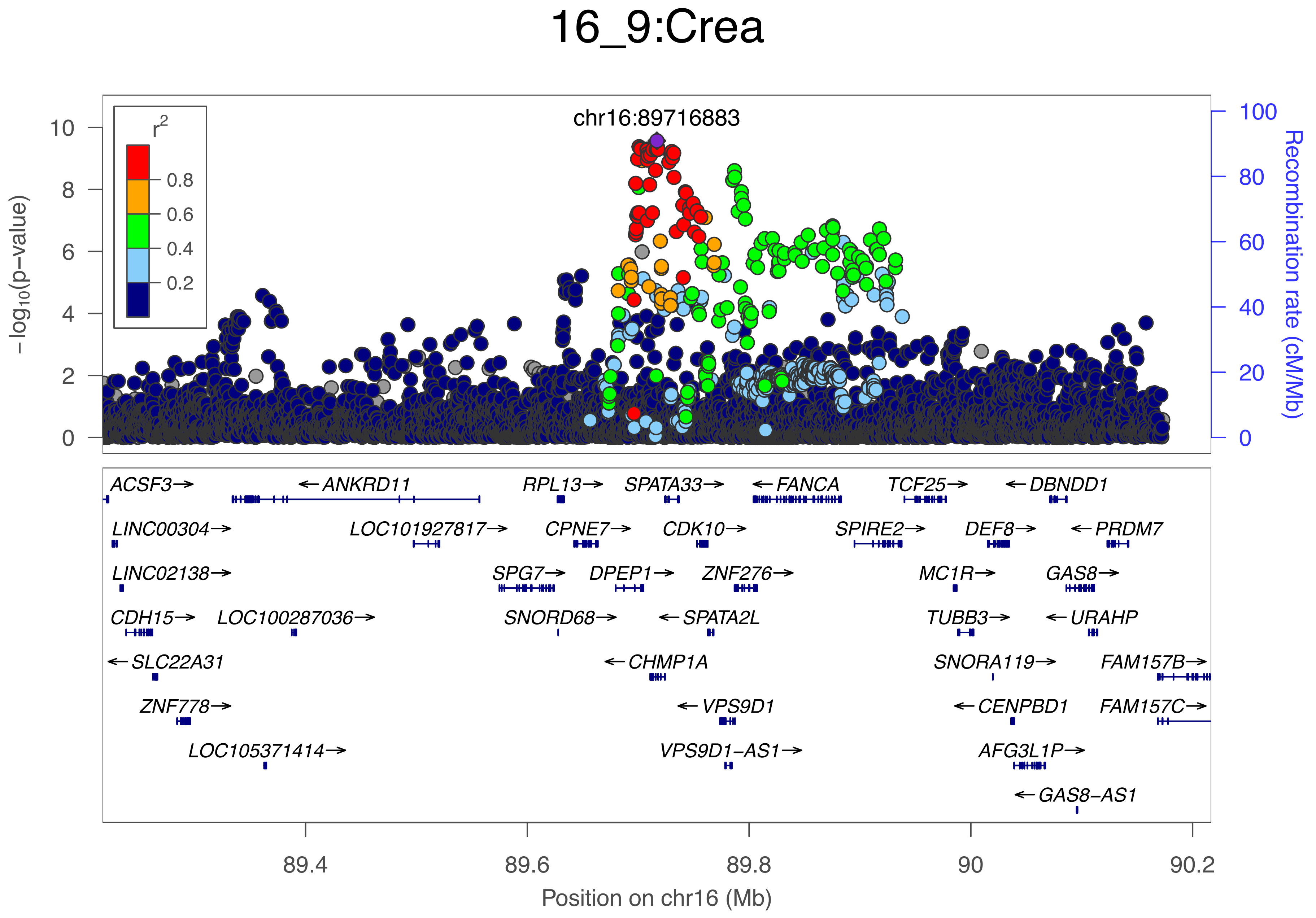
16_9:Crea
chr16:89716883
r2
0.8
0.6
0.4
0.2
−log10(p−value)
Recombination rate (cM/Mb)
0
2
4
6
8
10
0
20
40
60
80
100
ACSF3→
←ANKRD11
RPL13→
SPATA33→
←FANCA
TCF25→
←DBNDD1
LINC00304→
LOC101927817→
CPNE7→
CDK10→
SPIRE2→
DEF8→
←PRDM7
LINC02138→
SPG7→
DPEP1→
ZNF276→
MC1R→
GAS8→
CDH15→
LOC100287036→
SNORD68→
←SPATA2L
TUBB3→
←URAHP
←SLC22A31
←CHMP1A
SNORA119→
FAM157B→
ZNF778→
←VPS9D1
←CENPBD1
FAM157C→
LOC105371414→
VPS9D1−AS1→
AFG3L1P→
←GAS8−AS1
89.4
89.6
89.8
90
90.2
Position on chr16 (Mb)

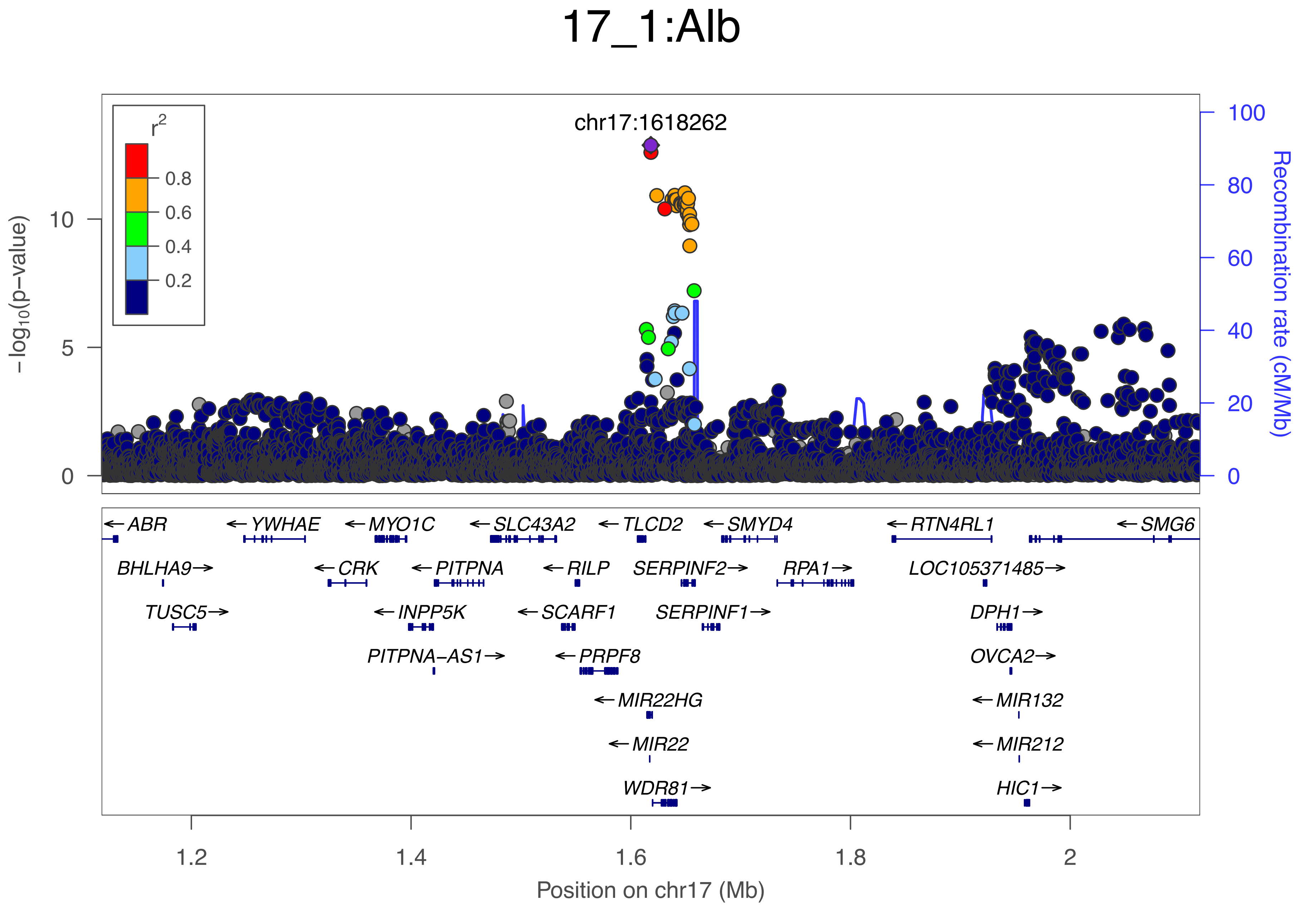
17_1:Alb
chr17:1618262
r2
0.8
0.6
0.4
0.2
10
5
0
−log10(p−value)
100
80
60
40
20
0
Recombination rate (cM/Mb)
←ABR
←YWHAE
←MYO1C
←SLC43A2
←TLCD2
←SMYD4
←RTN4RL1
←SMG6
BHLHA9→
←CRK
←PITPNA
←RILP
SERPINF2→
RPA1→
LOC105371485→
TUSC5→
←INPP5K
←SCARF1
SERPINF1→
DPH1→
PITPNA−AS1→
←PRPF8
OVCA2→
←MIR22HG
←MIR132
←MIR22
←MIR212
WDR81→
HIC1→
1.2
1.4
1.6
1.8
2
Position on chr17 (Mb)

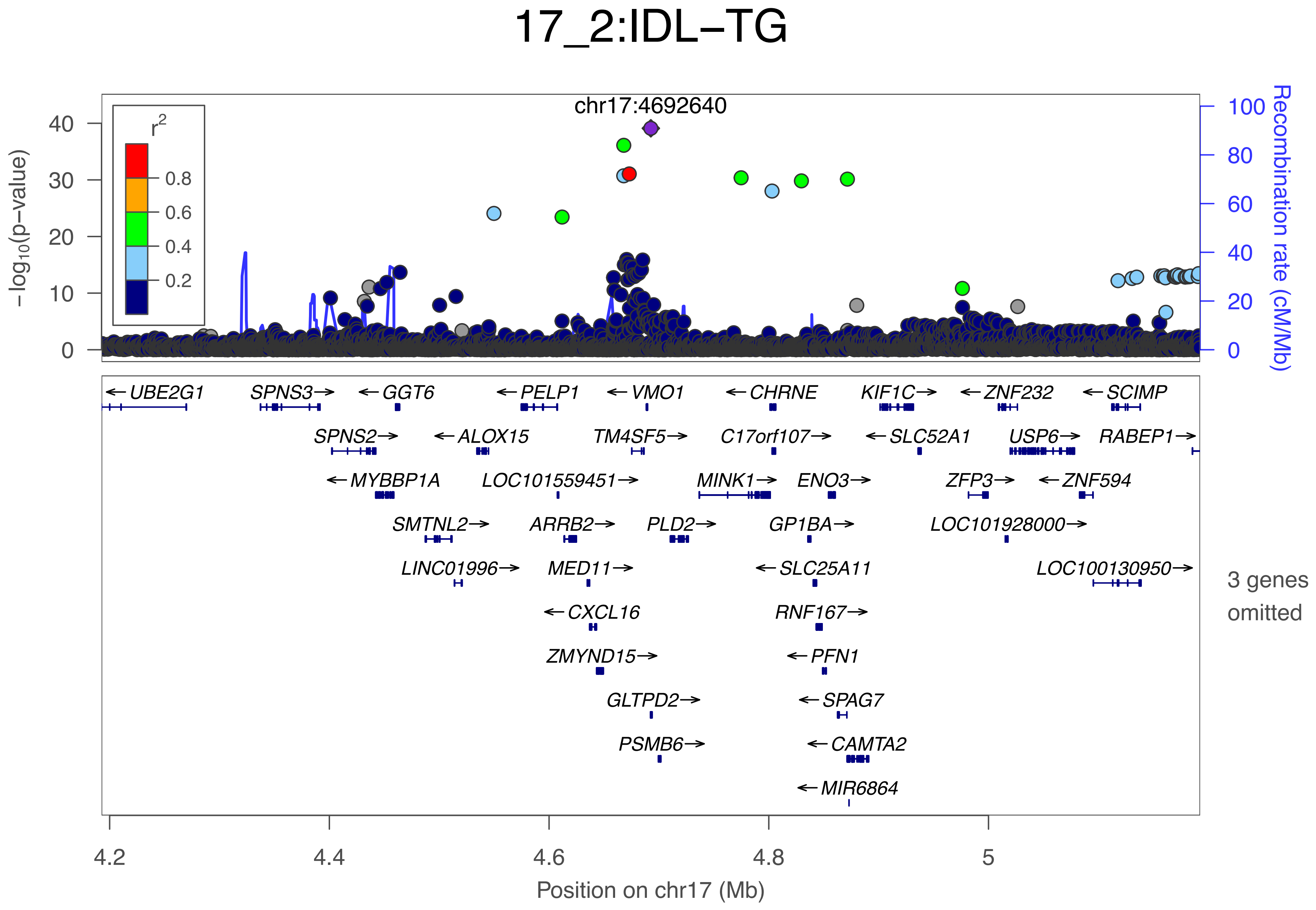

17_2:IDL−TG
chr17:4692640
r2
0.8
0.6
0.4
0.2
−log10(p−value)
0
10
20
30
40
Recombination rate (cM/Mb)
0
20
40
60
80
100
← UBE2G1
SPNS3 →
← GGT6
← PELP1
← VMO1
← CHRNE
KIF1C →
← ZNF232
← SCIMP
SPNS2 →
← ALOX15
TM4SF5 →
C17orf107 →
← SLC52A1
USP6 →
RABEP1 →
← MYBBP1A
LOC101559451 →
MINK1 →
ENO3 →
ZFP3 →
← ZNF594
SMTNL2 →
ARRB2 →
PLD2 →
GP1BA →
LOC101928000 →
LINC01996 →
MED11 →
← SLC25A11
LOC100130950 →
← CXCL16
RNF167 →
ZMYND15 →
← PFN1
GLTPD2 →
← SPAG7
PSMB6 →
← CAMTA2
← MIR6864
3 genes omitted
4.2
4.4
4.6
4.8
5
Position on chr17 (Mb)

### 17_3:Cit

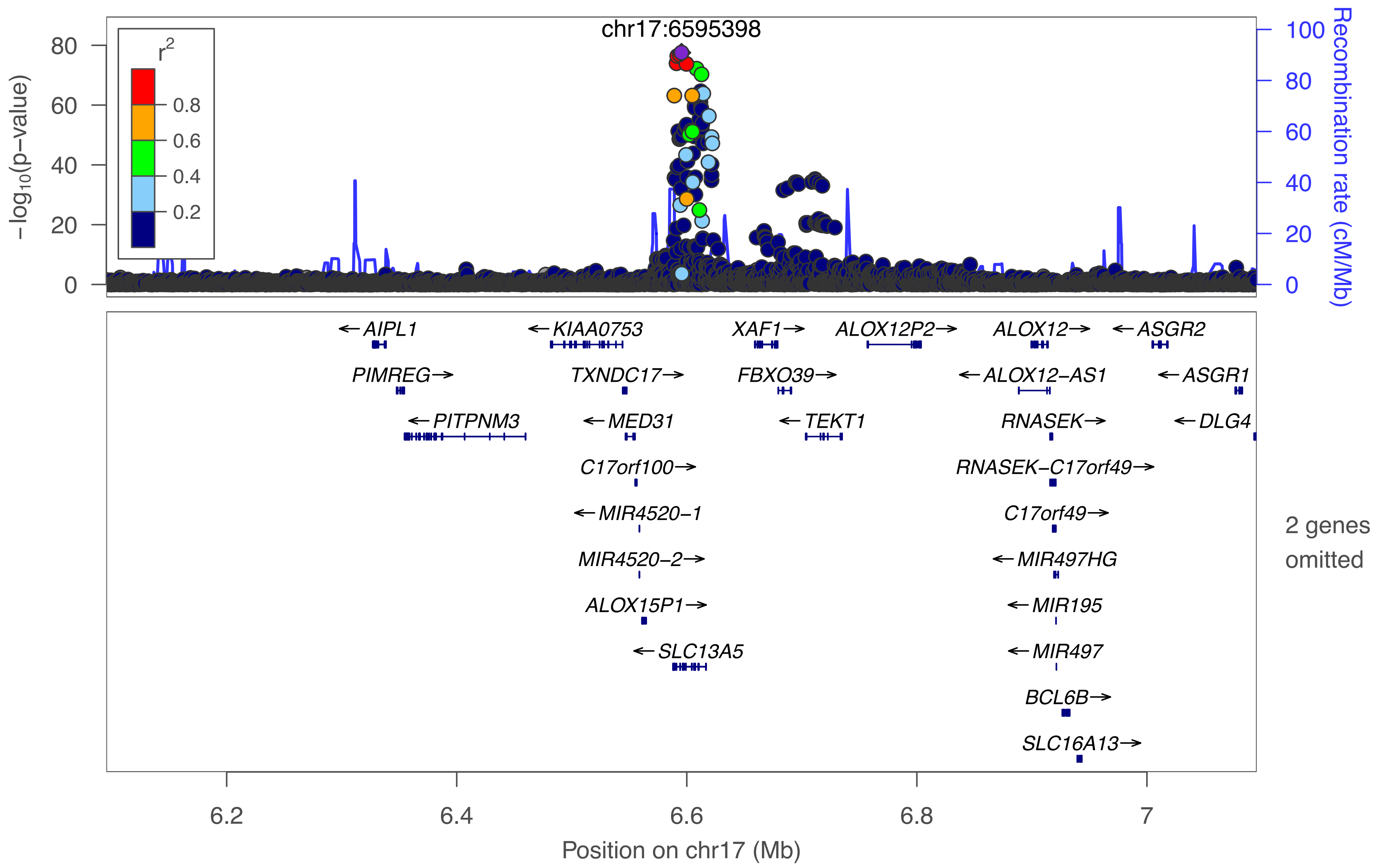

chr17:6595398
r2
0.8
0.6
0.4
0.2
80
60
40
20
0
−log10(p−value)
100
80
60
40
20
0
Recombination rate (cM/Mb)
←AIPL1
←KIAA0753
XAF1→
ALOX12P2→
ALOX12→
←ASGR2
PIMREG→
TXNDC17→
FBXO39→
←ALOX12−AS1
←ASGR1
←PITPNM3
←MED31
←TEKT1
RNASEK→
←DLG4
C17orf100→
RNASEK−C17orf49→
←MIR4520−1
C17orf49→
MIR4520−2→
←MIR497HG
ALOX15P1→
←MIR195
←SLC13A5
←MIR497
BCL6B→
SLC16A13→
2 genes omitted
6.2
6.4
6.6
6.8
7
Position on chr17 (Mb)

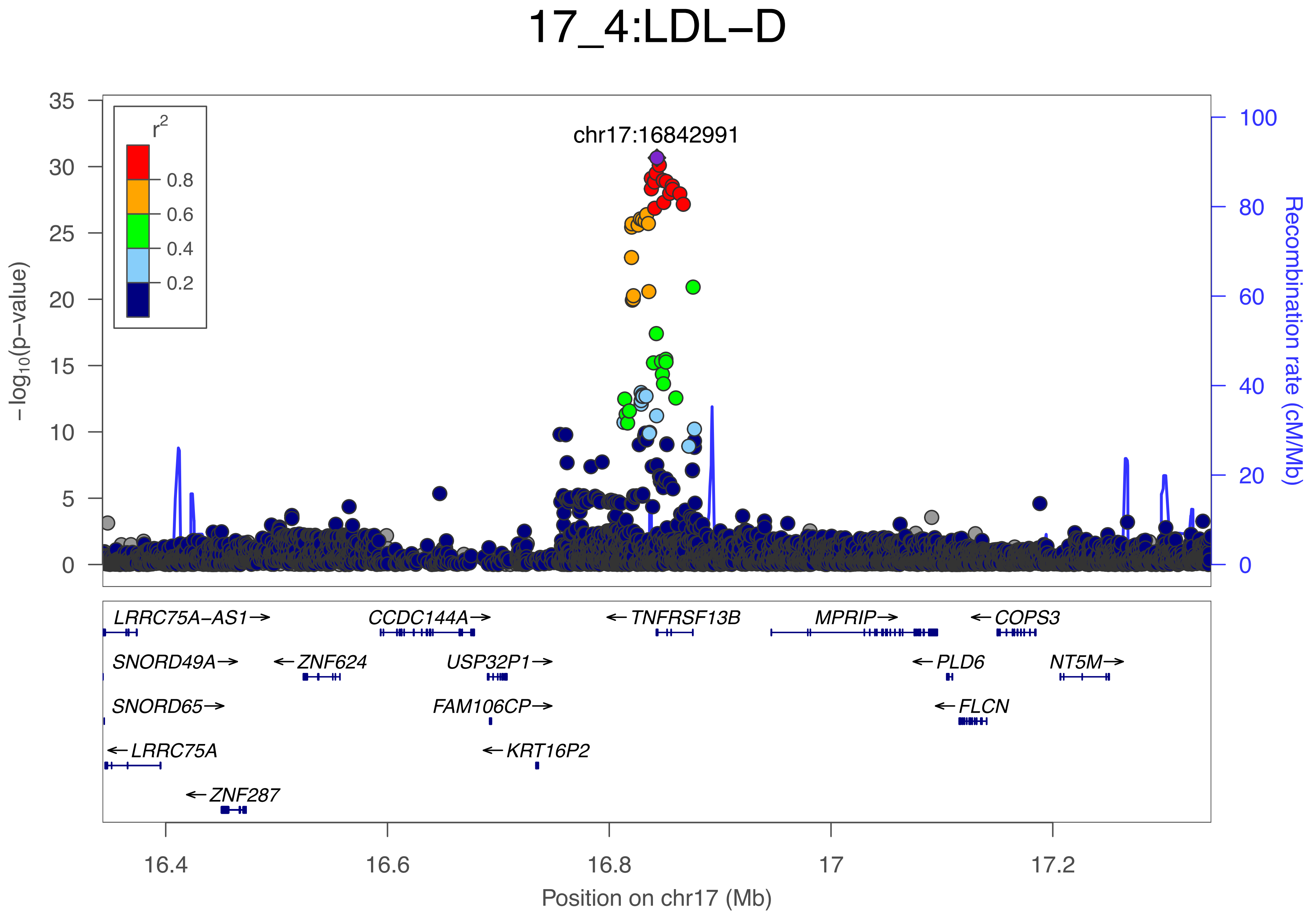
17_4:LDL−D
chr17:16842991
r2
0.8
0.6
0.4
0.2
−log10(p−value)
Recombination rate (cM/Mb)
0
5
10
15
20
25
30
35
0
20
40
60
80
100
LRRC75A−AS1→
CCDC144A→
←TNFRSF13B
MPRIP→
←COPS3
SNORD49A→
←ZNF624
USP32P1→
←PLD6
NT5M→
SNORD65→
FAM106CP→
←FLCN
←LRRC75A
←KRT16P2
←ZNF287
16.4
16.6
16.8
17
17.2
Position on chr17 (Mb)

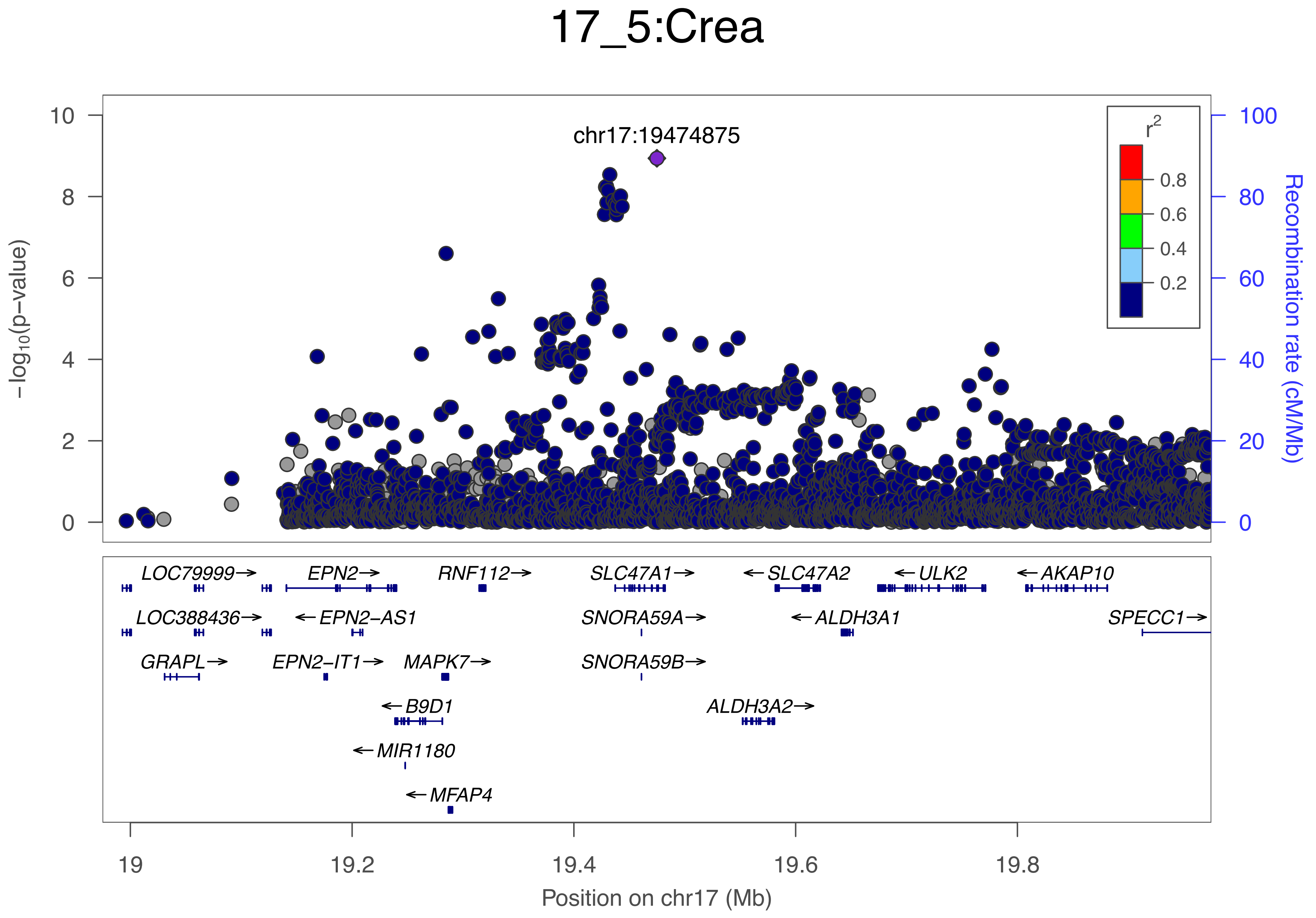
17_5:Crea
chr17:19474875
−log10(p−value)
Recombination rate (cM/Mb)
r2
0.8
0.6
0.4
0.2
LOC79999→
EPN2→
RNF112→
SLC47A1→
←SLC47A2
←ULK2
←AKAP10
LOC388436→
←EPN2−AS1
SNORA59A→
←ALDH3A1
SPECC1→
GRAPL→
EPN2−IT1→
MAPK7→
SNORA59B→
←B9D1
ALDH3A2→
←MIR1180
←MFAP4
Position on chr17 (Mb)

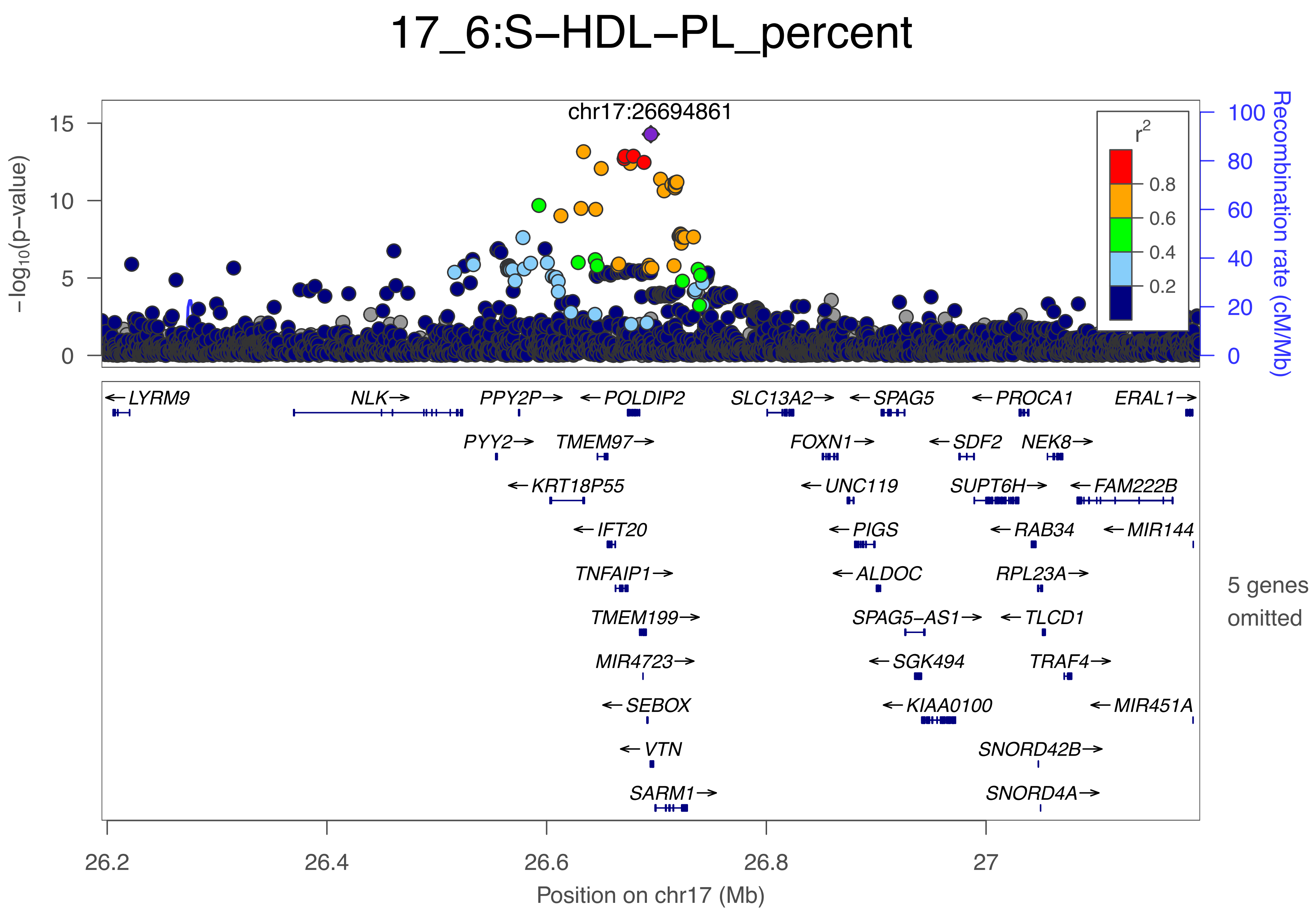
17_6:S–HDL–PL_percent
chr17:26694861
−log10(p−value)
Recombination rate (cM/Mb)
r2
0.8
0.6
0.4
0.2
0
5
10
15
0
20
40
60
80
100
LYRM9
NLK
PPY2P
POLDIP2
SLC13A2
SPAG5
PROCA1
ERAL1
PYY2
TMEM97
FOXN1
SDF2
NEK8
KRT18P55
UNC119
SUPT6H
FAM222B
IFT20
PIGS
RAB34
MIR144
TNFAIP1
ALDOC
RPL23A
TMEM199
SPAG5−AS1
TLCD1
MIR4723
SGK494
TRAF4
SEBOX
KIAA0100
MIR451A
VTN
SNORD42B
SARM1
SNORD4A
5 genes omitted
26.2
26.4
26.6
26.8
27
Position on chr17 (Mb)

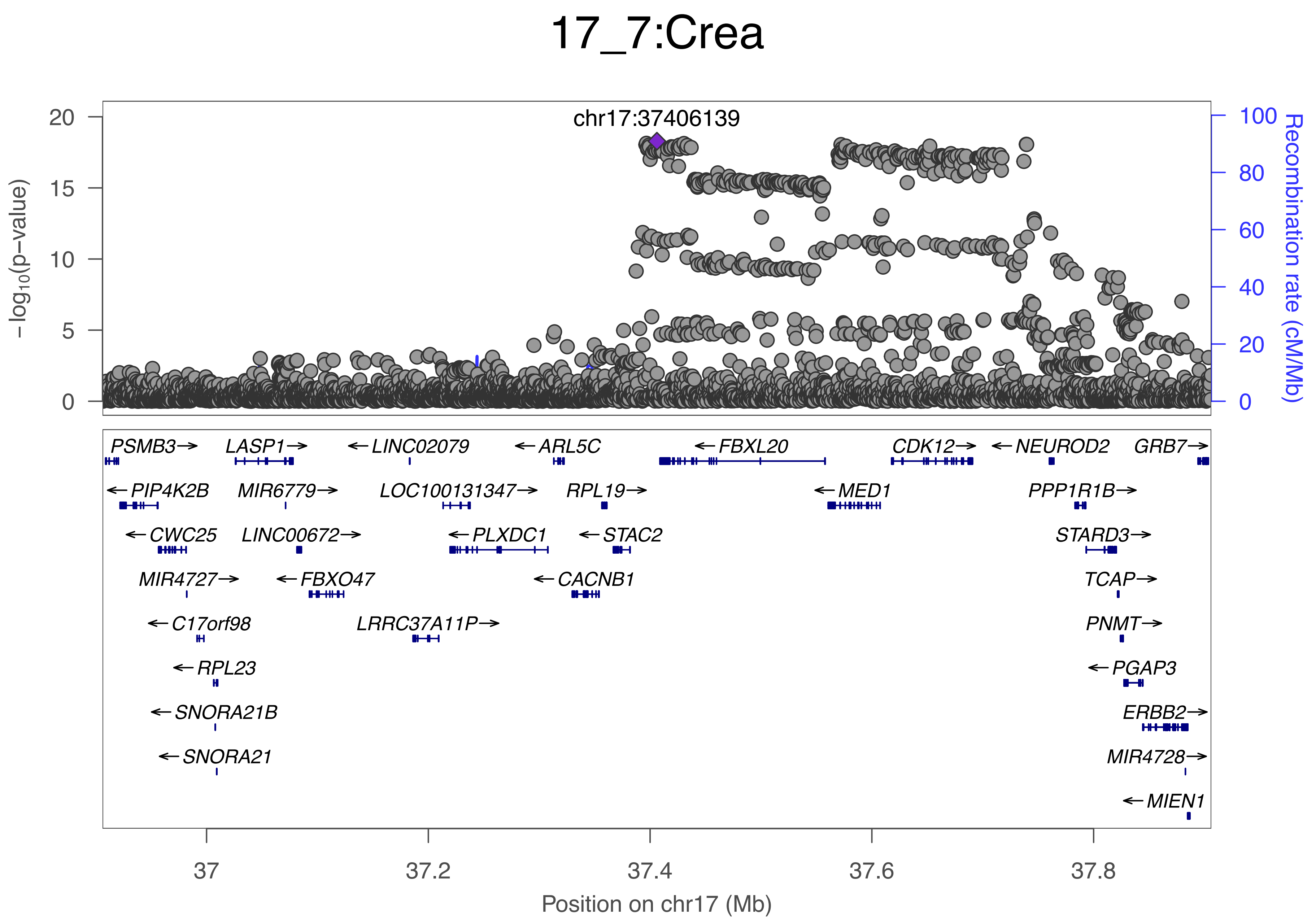
17_7:Crea
chr17:37406139
−log10(p−value)
Recombination rate (cM/Mb)
20
15
10
5
0
100
80
60
40
20
0
PSMB3→
LASP1→
←LINC02079
←ARL5C
←FBXL20
CDK12→
←NEUROD2
GRB7→
←PIP4K2B
MIR6779→
LOC100131347→
RPL19→
←MED1
PPP1R1B→
←CWC25
LINC00672→
←PLXDC1
←STAC2
STARD3→
MIR4727→
←FBXO47
←CACNB1
TCAP→
←C17orf98
LRRC37A11P→
PNMT→
←RPL23
←PGAP3
←SNORA21B
ERBB2→
←SNORA21
MIR4728→
←MIEN1
37
37.2
37.4
37.6
37.8
Position on chr17 (Mb)

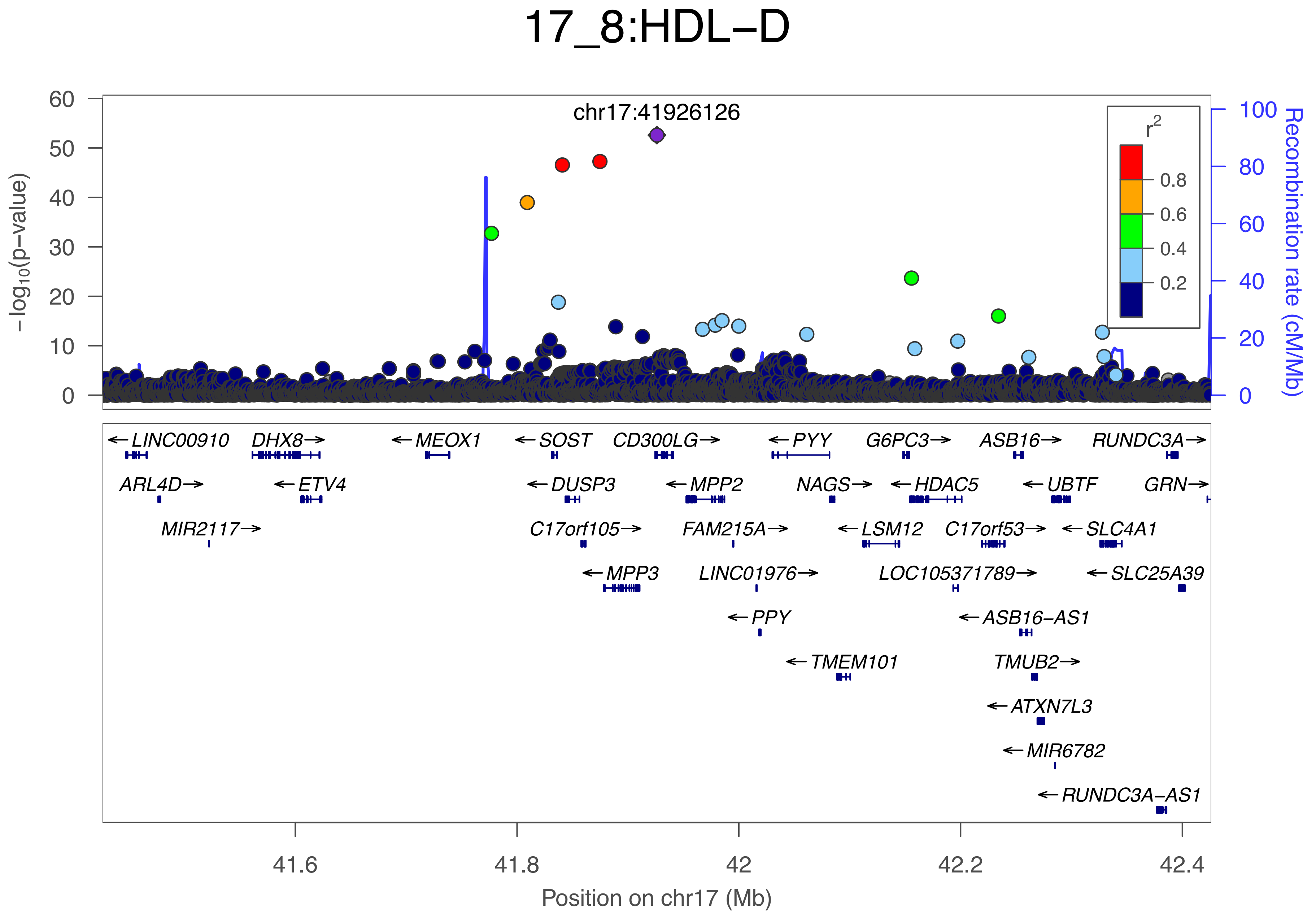
17_8:HDL−D
chr17:41926126
−log10(p−value)
Recombination rate (cM/Mb)
r2
0.8
0.6
0.4
0.2
LINC00910
DHX8
MEOX1
SOST
CD300LG
PYY
G6PC3
ASB16
RUNDC3A
ARL4D
ETV4
DUSP3
MPP2
NAGS
HDAC5
UBTF
GRN
MIR2117
C17orf105
FAM215A
LSM12
C17orf53
SLC4A1
MPP3
LINC01976
LOC105371789
SLC25A39
PPY
ASB16−AS1
TMEM101
TMUB2
ATXN7L3
MIR6782
RUNDC3A−AS1
41.6
41.8
42
42.2
42.4
Position on chr17 (Mb)

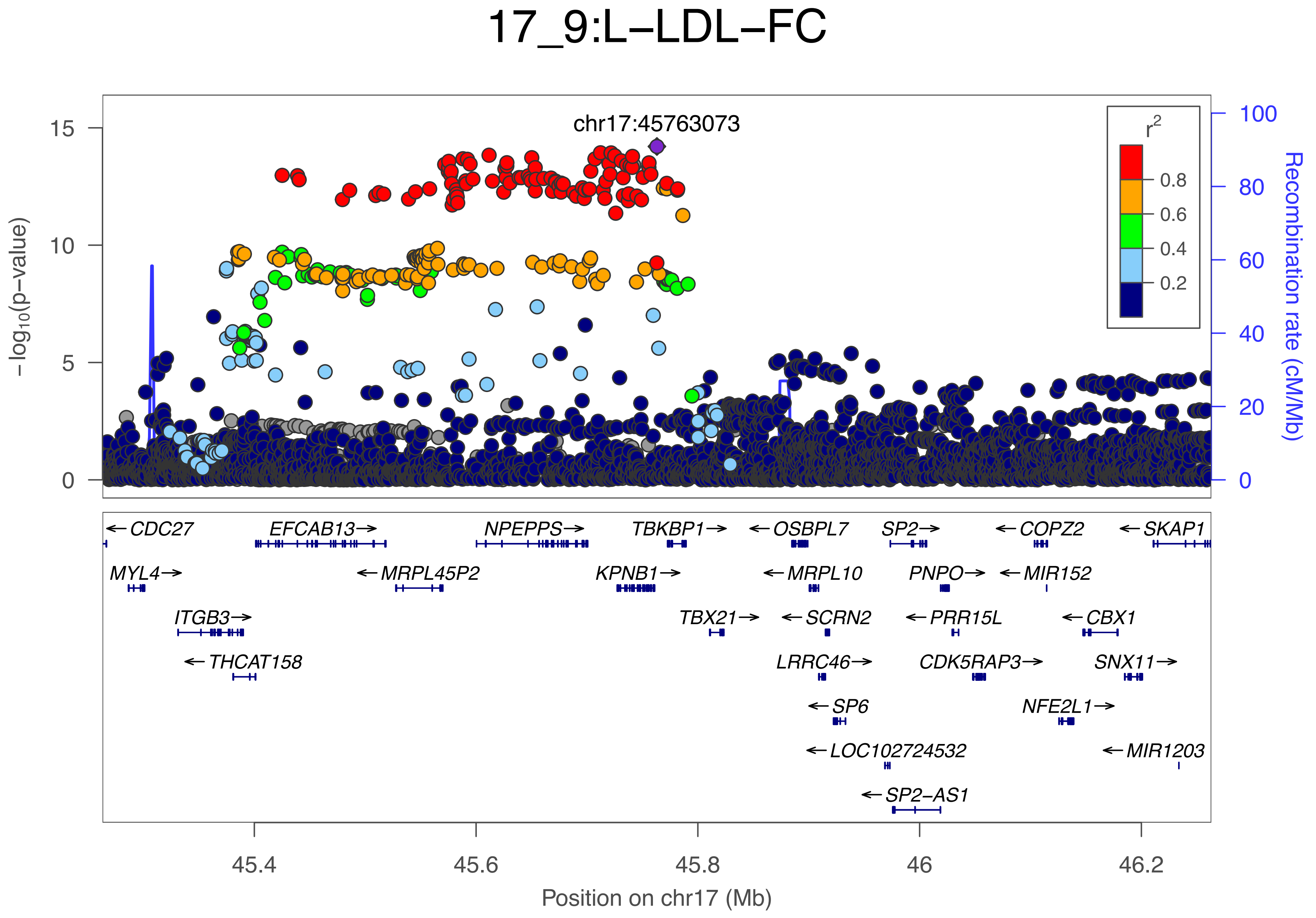

17_9:L−LDL−FC
chr17:45763073
r2
0.8
0.6
0.4
0.2
−log10(p−value)
Recombination rate (cM/Mb)
15
10
5
0
100
80
60
40
20
0
CDC27
EFCAB13
NPEPPS
TBKBP1
OSBPL7
SP2
COPZ2
SKAP1
MYL4
MRPL45P2
KPNB1
MRPL10
PNPO
MIR152
ITGB3
TBX21
SCRN2
PRR15L
CBX1
THCAT158
LRRC46
CDK5RAP3
SNX11
SP6
NFE2L1
LOC102724532
MIR1203
SP2−AS1
45.4
45.6
45.8
46
46.2
Position on chr17 (Mb)

### 17_10:S−LDL−FC_percent

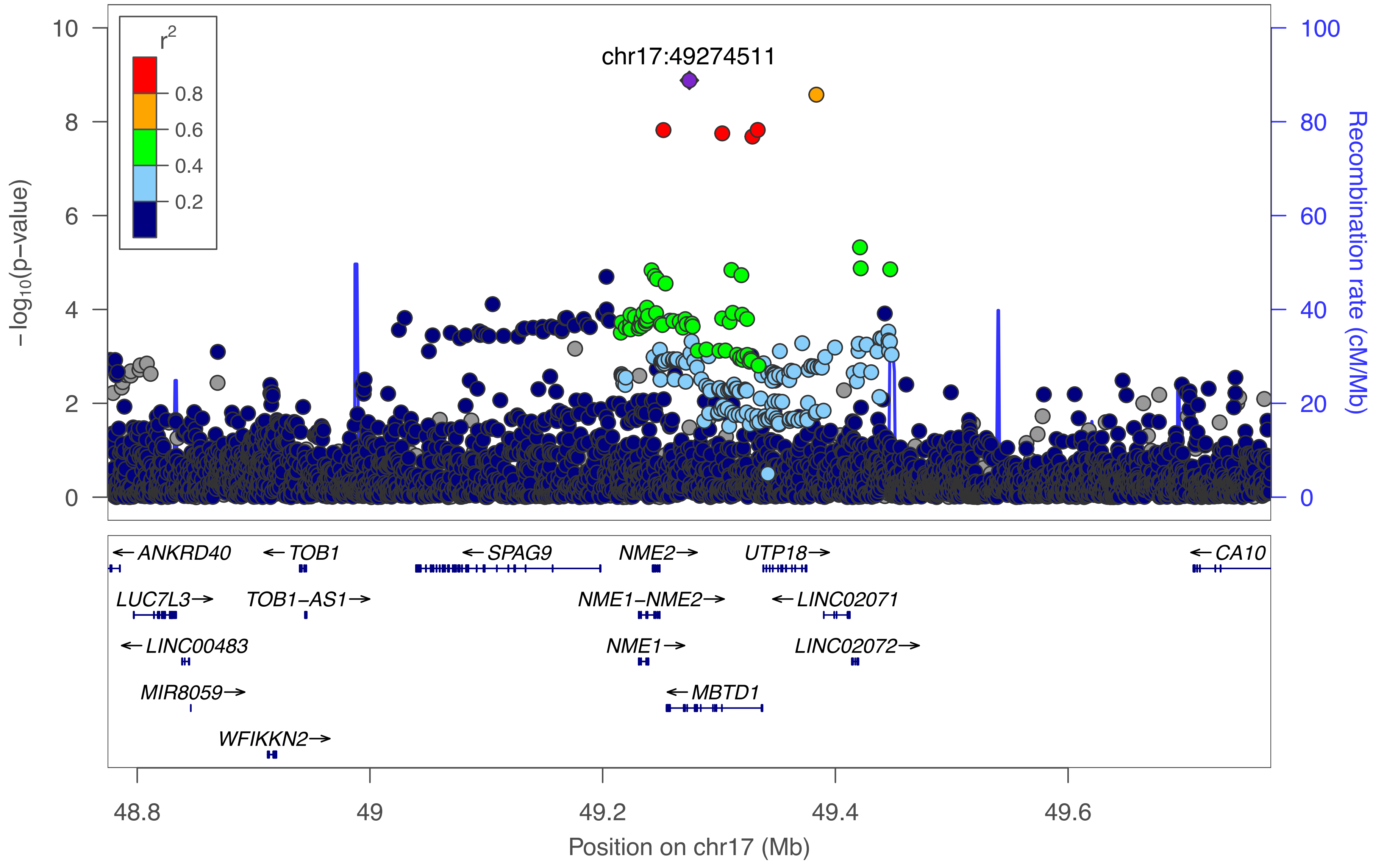

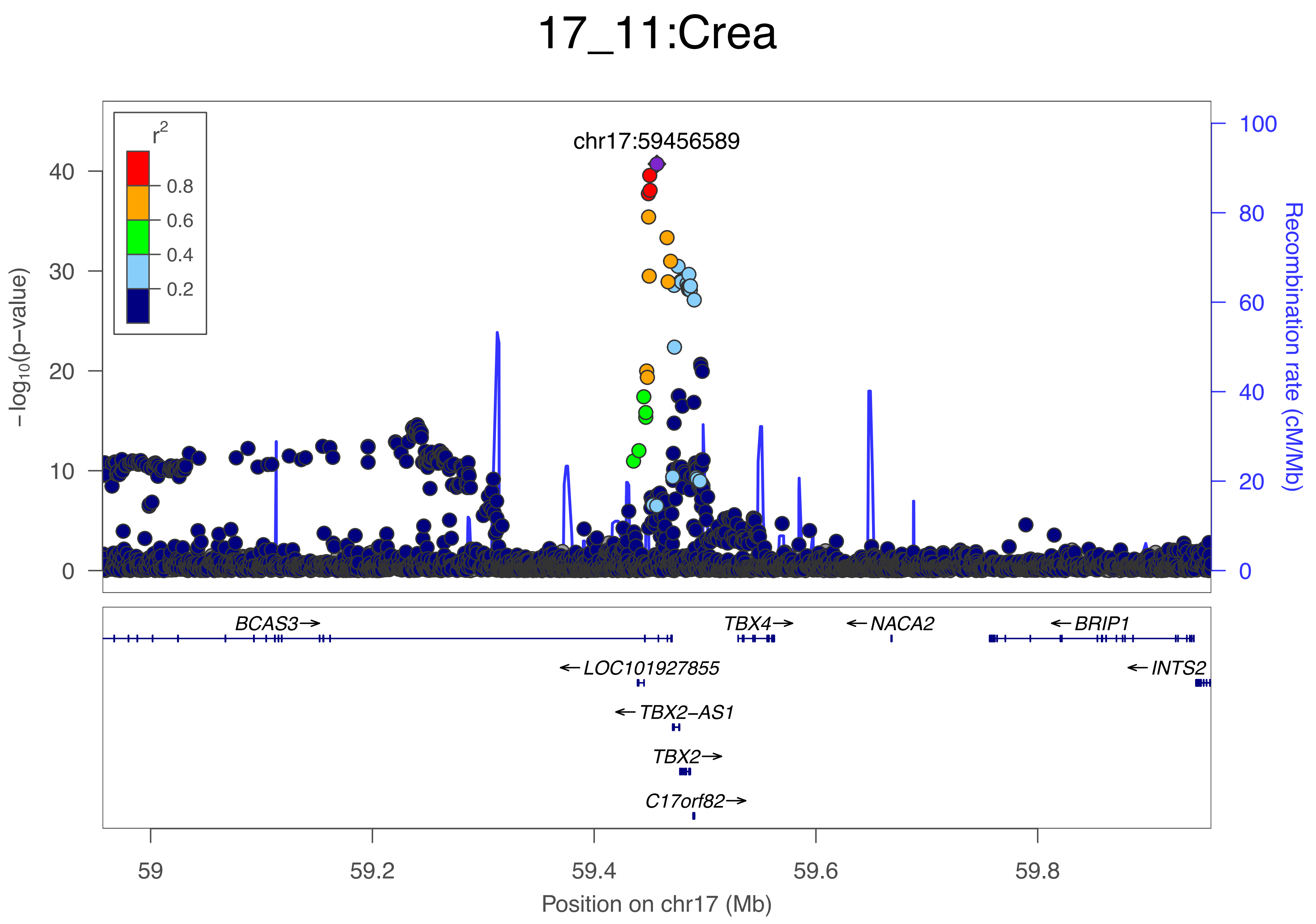
17_11:Crea
chr17:59456589
r2
0.8
0.6
0.4
0.2
−log10(p−value)
0
10
20
30
40
Recombination rate (cM/Mb)
0
20
40
60
80
100
BCAS3→
TBX4→
←NACA2
←BRIP1
←LOC101927855
←INTS2
←TBX2−AS1
TBX2→
C17orf82→
59
59.2
59.4
59.6
59.8
Position on chr17 (Mb)

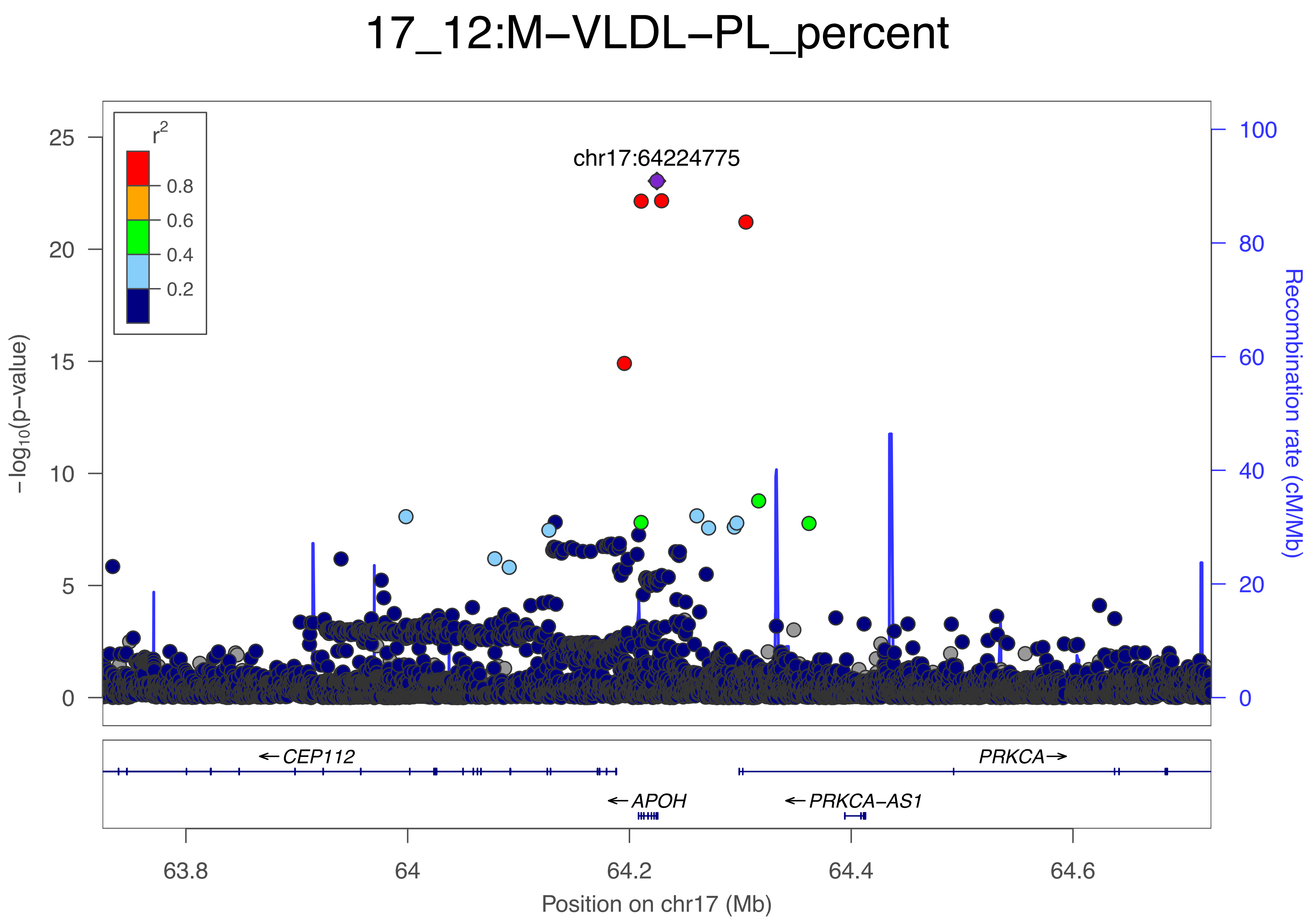

17_12:M−VLDL−PL_percent
r2
0.8
0.6
0.4
0.2
chr17:64224775
−log10(p−value)
0
5
10
15
20
25
Recombination rate (cM/Mb)
0
20
40
60
80
100
← CEP112
PRKCA→
← APOH
← PRKCA−AS1
63.8
64
64.2
64.4
64.6
Position on chr17 (Mb)

### 17_13:L−LDL−FC

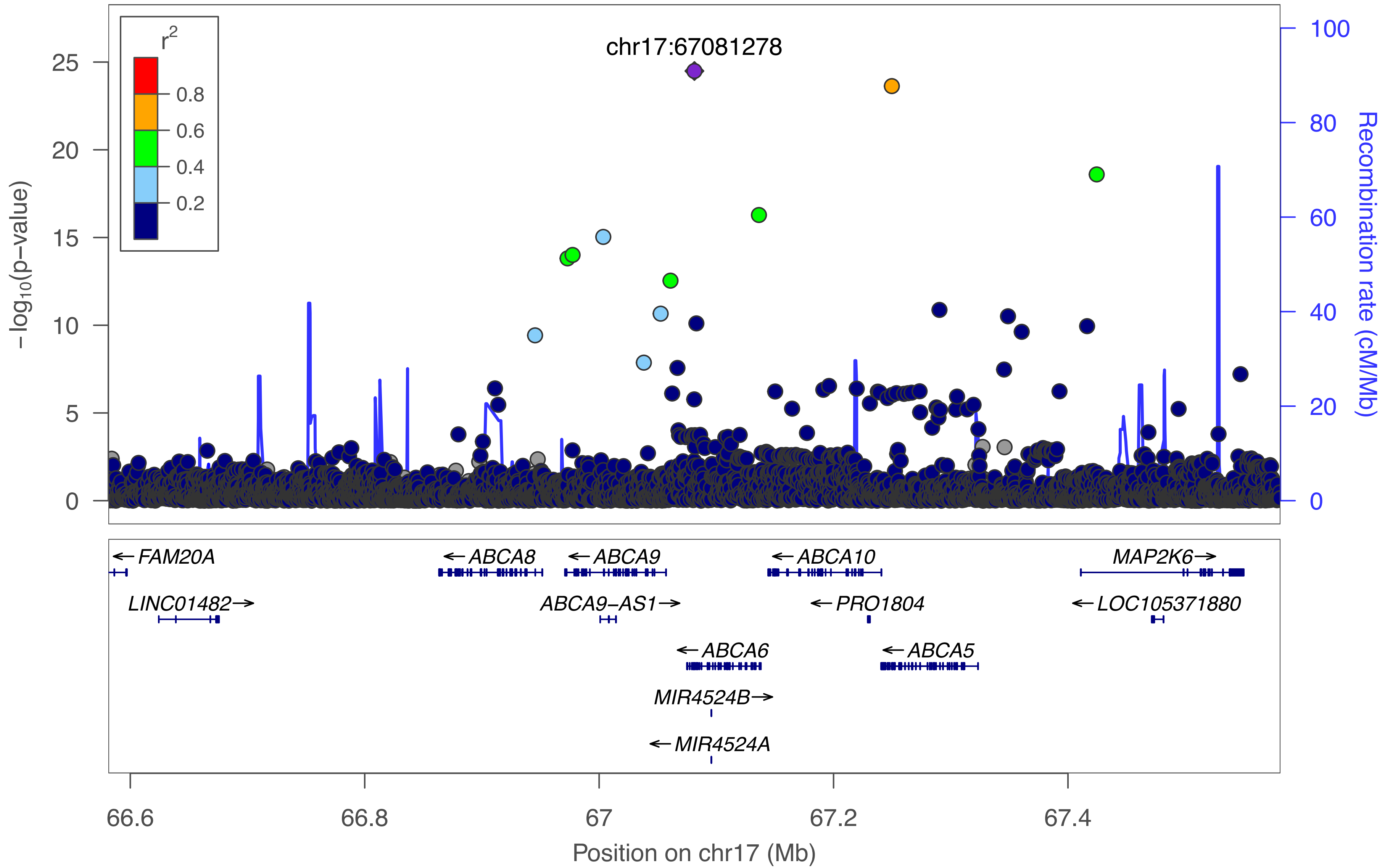

r2
0.8
0.6
0.4
0.2
chr17:67081278
−log10(p−value)
25
20
15
10
5
0
100
80
60
40
20
0
Recombination rate (cM/Mb)
← FAM20A
← ABCA8
← ABCA9
← ABCA10
MAP2K6 →
LINC01482 →
ABCA9−AS1 →
← PRO1804
← LOC105371880
← ABCA6
← ABCA5
MIR4524B →
← MIR4524A
66.6
66.8
67
67.2
67.4
Position on chr17 (Mb)

### 17_14:ApoBbyApoA1

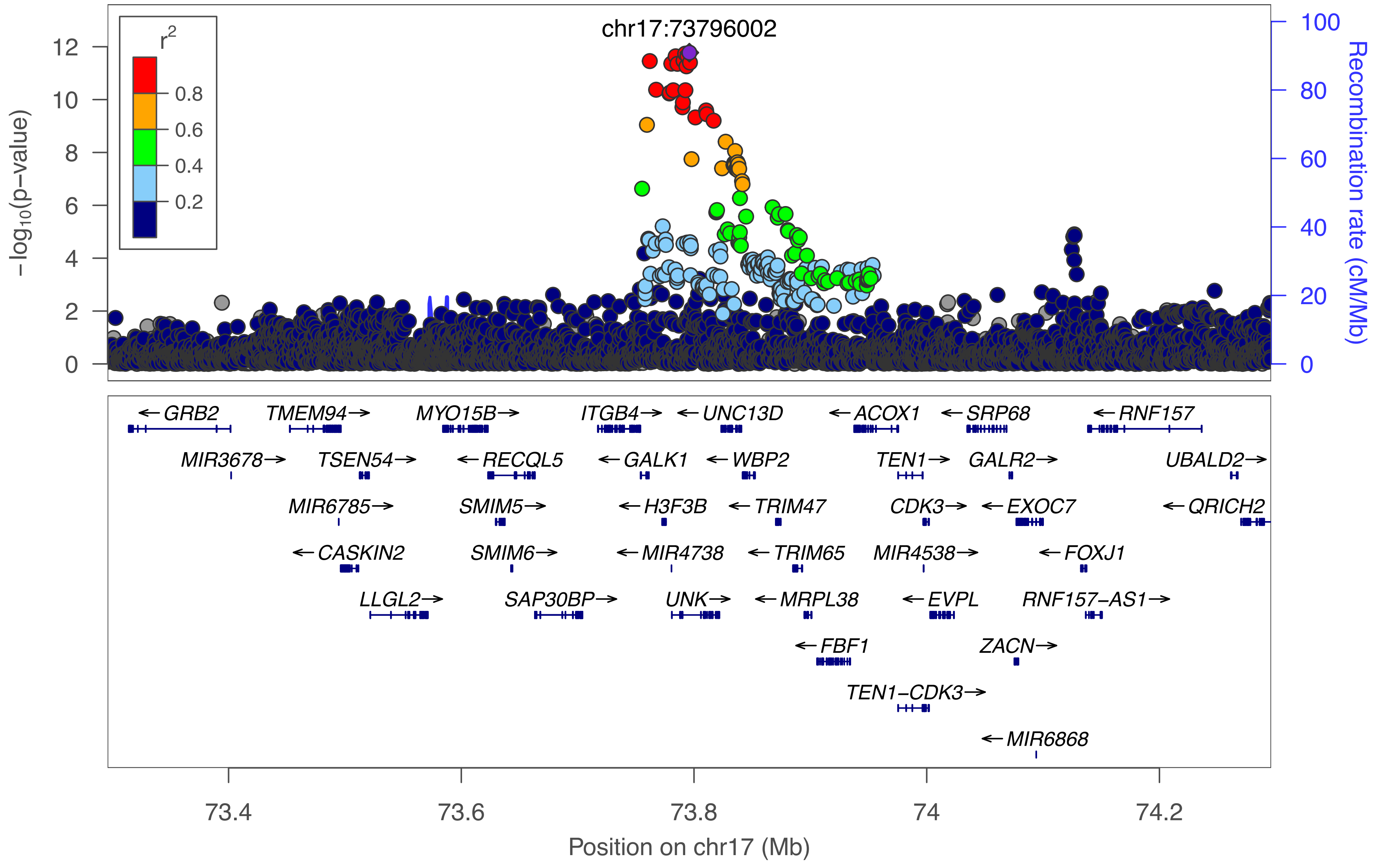

### 17_15:S−VLDL−CE_percent

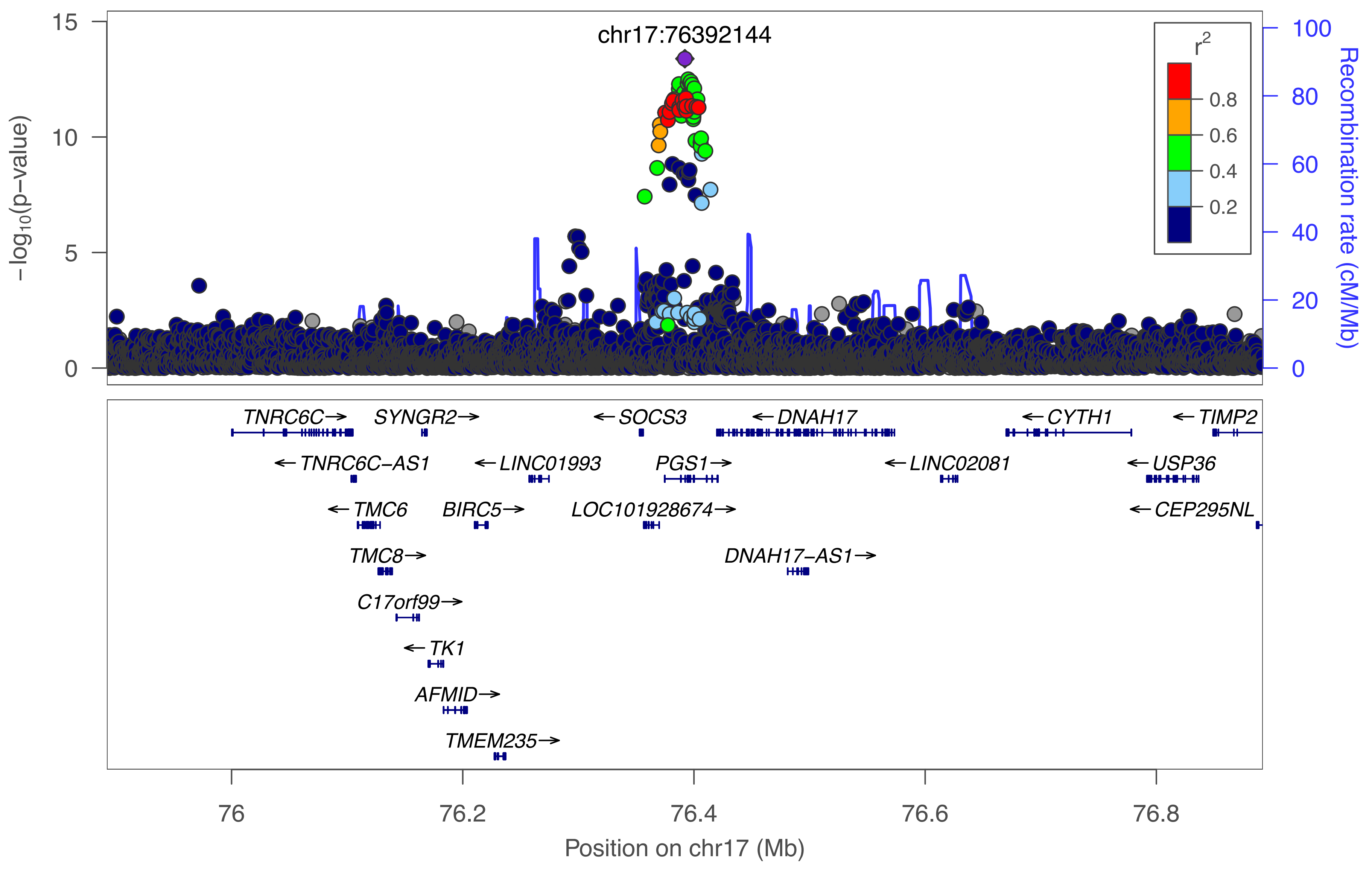

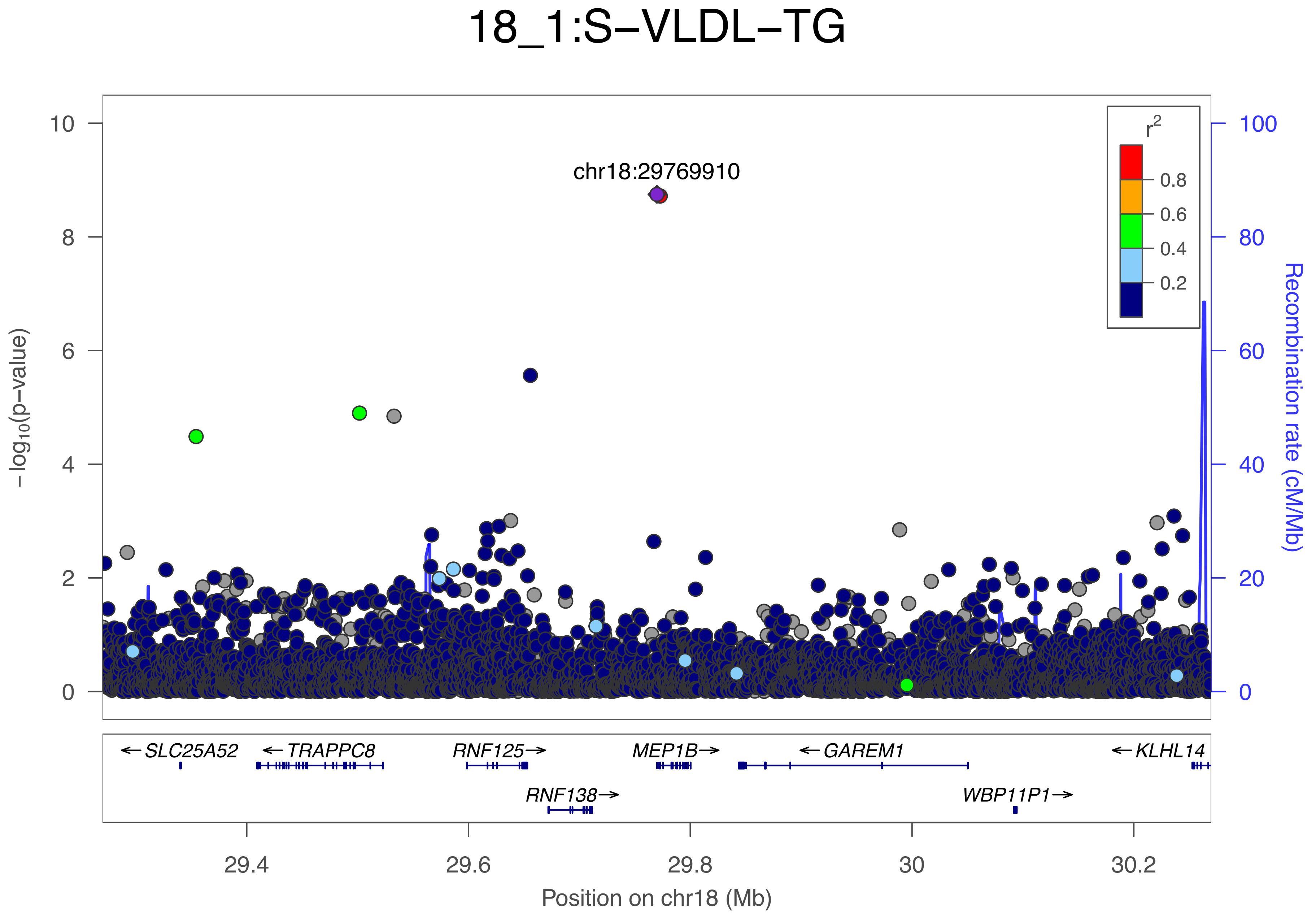
18_1:S−VLDL−TG
chr18:29769910
r²
0.8
0.6
0.4
0.2
−log₁₀(p−value)
Recombination rate (cM/Mb)
10
8
6
4
2
0
100
80
60
40
20
0
←SLC25A52
←TRAPPC8
RNF125→
MEP1B→
←GAREM1
←KLHL14
RNF138→
WBP11P1→
29.4
29.6
29.8
30
30.2
Position on chr18 (Mb)

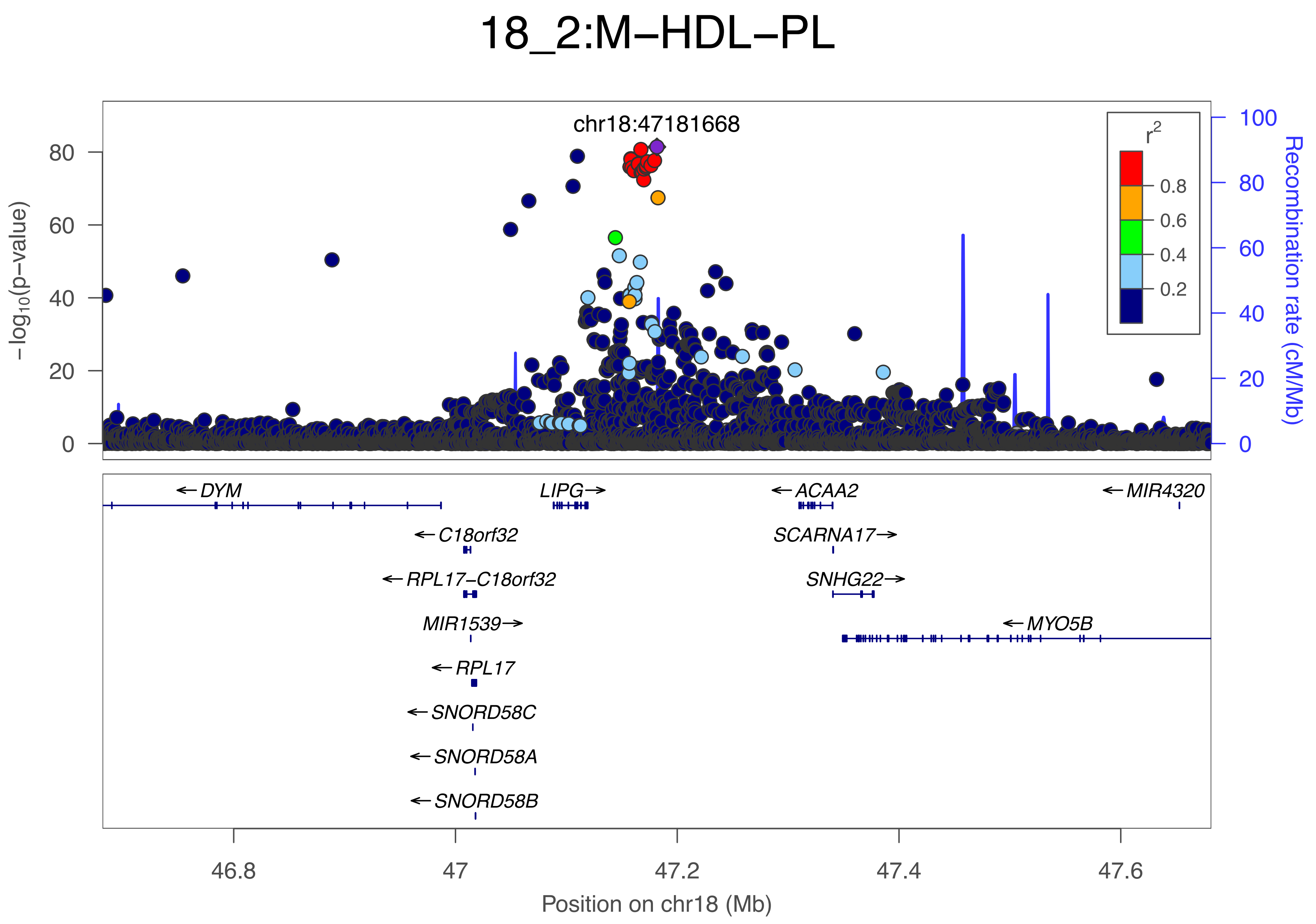

18_2:M−HDL−PL
chr18:47181668
−log10(p−value)
Recombination rate (cM/Mb)
r2
0.8
0.6
0.4
0.2
0
20
40
60
80
100
DYM
LIPG
ACAA2
MIR4320
C18orf32
SCARNA17
RPL17−C18orf32
SNHG22
MIR1539
MYO5B
RPL17
SNORD58C
SNORD58A
SNORD58B
46.8
47
47.2
47.4
47.6
Position on chr18 (Mb)

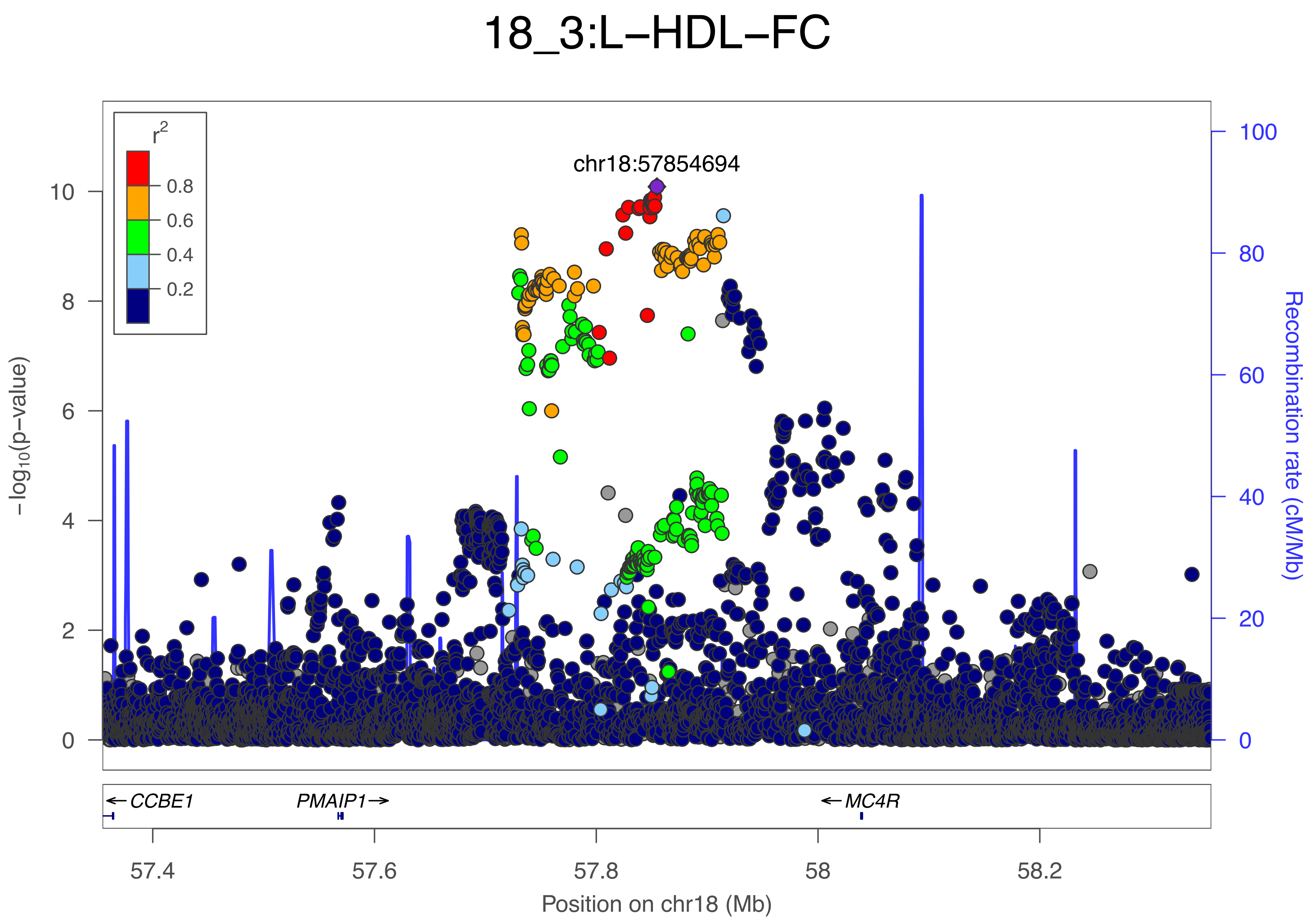

18_3:L−HDL−FC
r2
0.8
0.6
0.4
0.2
chr18:57854694
−log10(p−value)
Recombination rate (cM/Mb)
0
2
4
6
8
10
0
20
40
60
80
100
CCBE1
PMAIP1
MC4R
57.4
57.6
57.8
58
58.2
Position on chr18 (Mb)

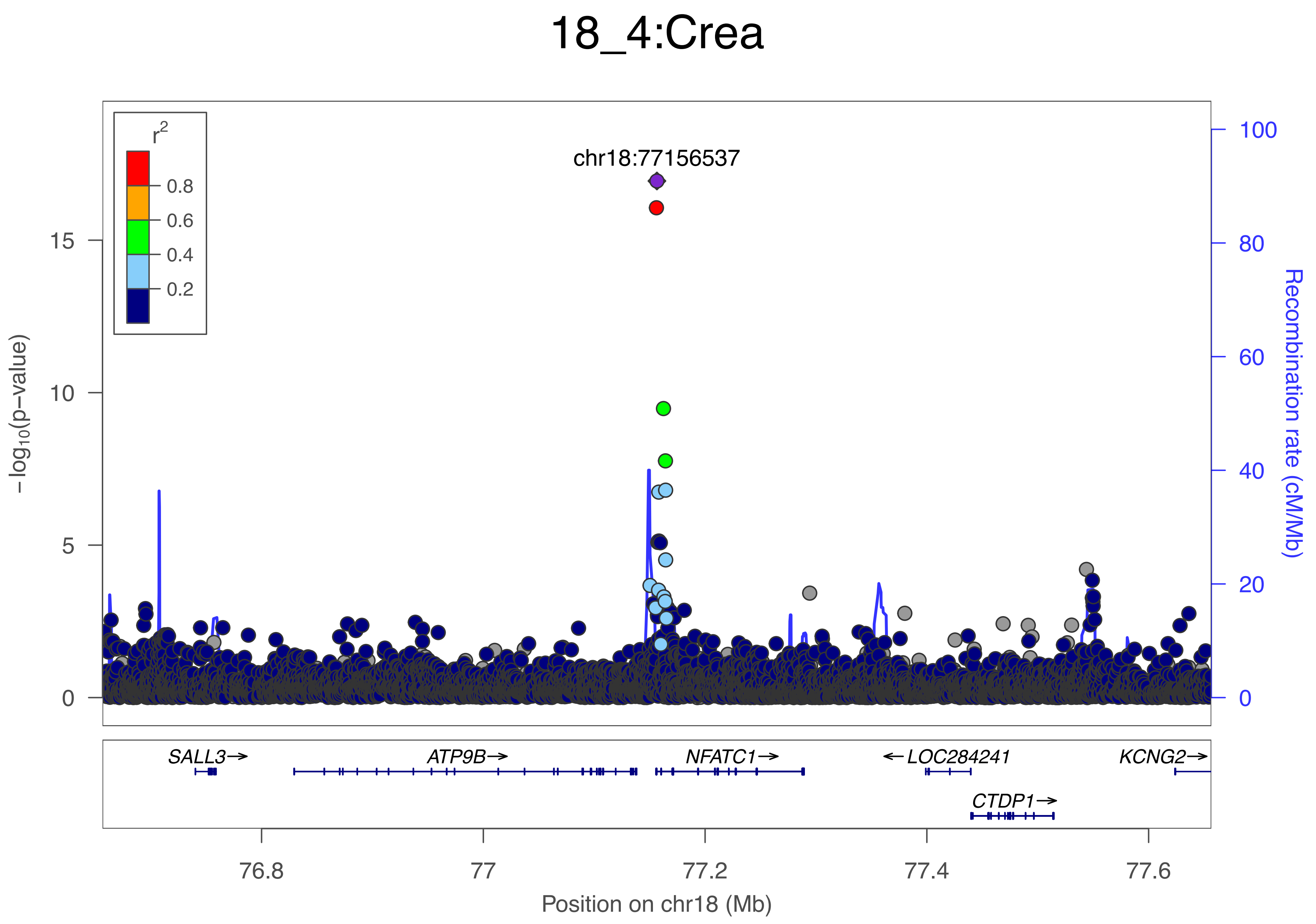
18_4:Crea
chr18:77156537
r2
0.8
0.6
0.4
0.2
15
10
5
0
−log10(p−value)
100
80
60
40
20
0
Recombination rate (cM/Mb)
SALL3→
ATP9B→
NFATC1→
←LOC284241
KCNG2→
CTDP1→
76.8
77
77.2
77.4
77.6
Position on chr18 (Mb)

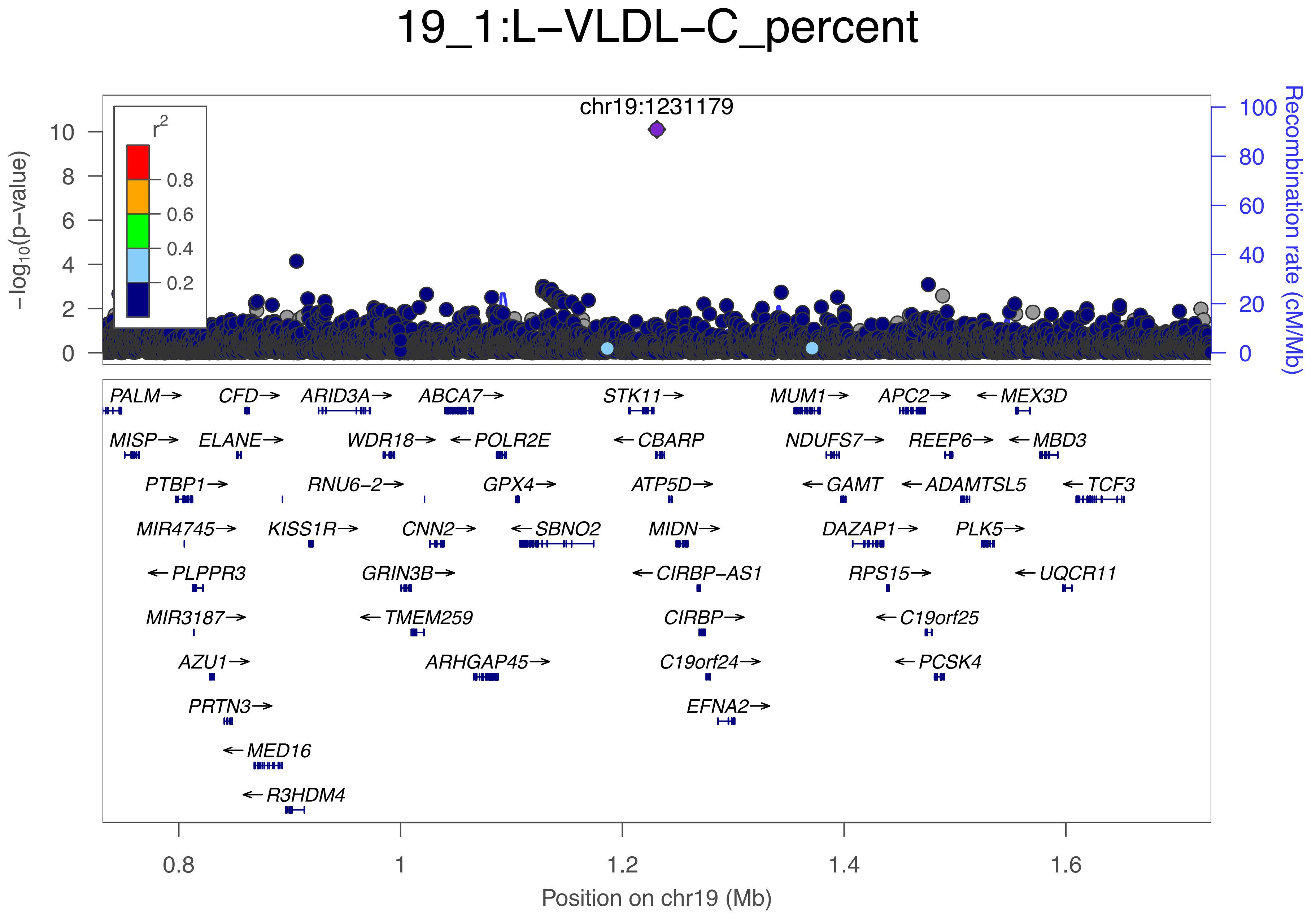

19_1:L−VLDL−C_percent
chr19:1231179
r2
0.8
0.6
0.4
0.2
−log10(p−value)
Recombination rate (cM/Mb)
PALM→
CFD→
ARID3A→
ABCA7→
STK11→
MUM1→
APC2→
←MEX3D
MISP→
ELANE→
WDR18→
←POLR2E
←CBARP
NDUFS7→
REEP6→
←MBD3
PTBP1→
RNU6−2→
GPX4→
ATP5D→
←GAMT
←ADAMTSL5
←TCF3
MIR4745→
KISS1R→
CNN2→
←SBNO2
MIDN→
DAZAP1→
PLK5→
←PLPPR3
GRIN3B→
←CIRBP−AS1
RPS15→
←UQCR11
MIR3187→
←TMEM259
CIRBP→
←C19orf25
AZU1→
ARHGAP45→
C19orf24→
←PCSK4
PRTN3→
EFNA2→
←MED16
←R3HDM4
Position on chr19 (Mb)

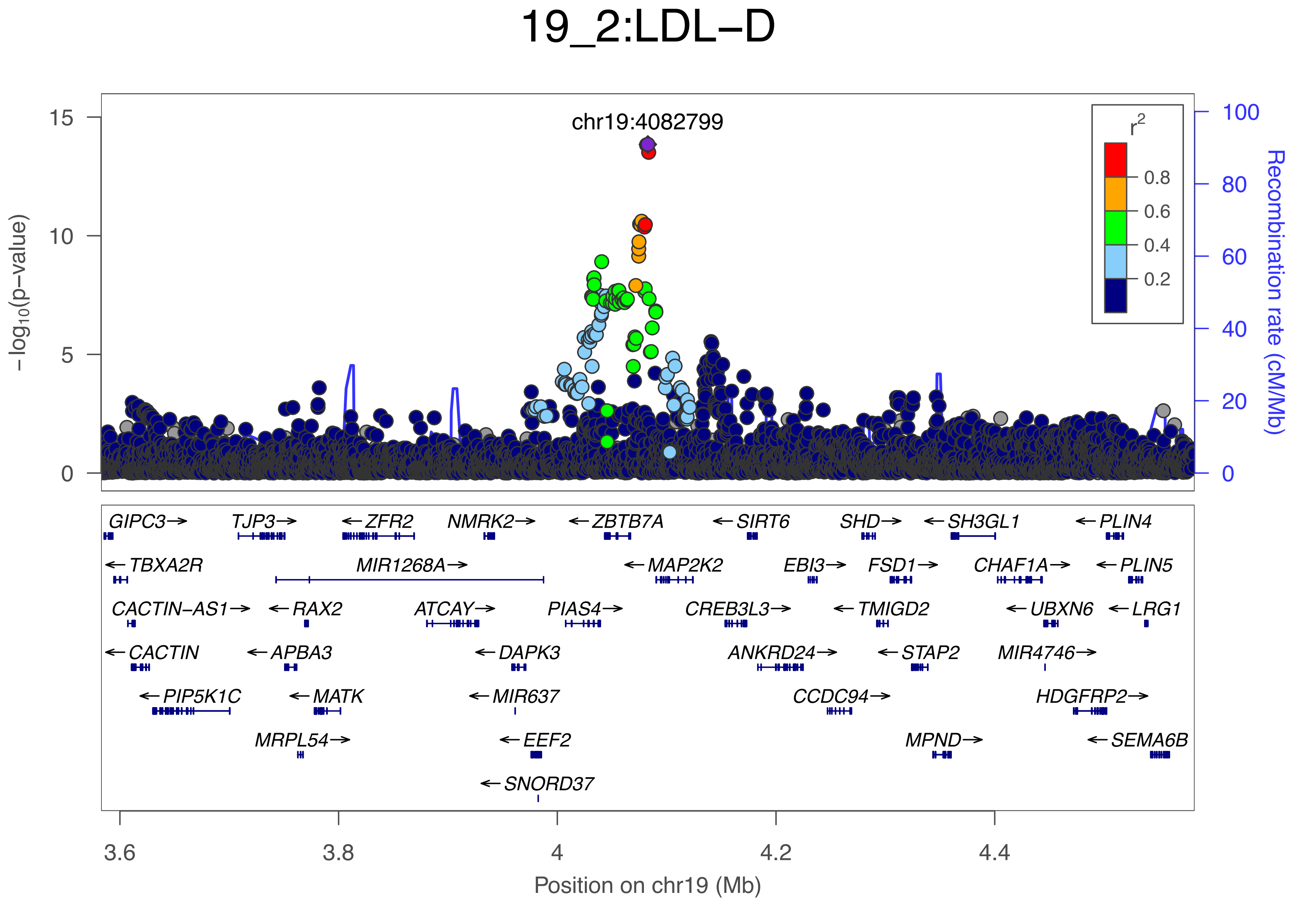
19_2:LDL−D
chr19:4082799
r2
0.8
0.6
0.4
0.2
−log10(p−value)
Recombination rate (cM/Mb)
GIPC3→
TJP3→
←ZFR2
NMRK2→
←ZBTB7A
←SIRT6
SHD→
←SH3GL1
←PLIN4
←TBXA2R
MIR1268A→
←MAP2K2
EBI3→
FSD1→
CHAF1A→
←PLIN5
CACTIN−AS1→
←RAX2
ATCAY→
PIAS4→
CREB3L3→
←TMIGD2
←UBXN6
←LRG1
←CACTIN
←APBA3
←DAPK3
ANKRD24→
←STAP2
MIR4746→
←PIP5K1C
←MATK
←MIR637
CCDC94→
HDGFRP2→
MRPL54→
←EEF2
MPND→
←SEMA6B
←SNORD37
Position on chr19 (Mb)

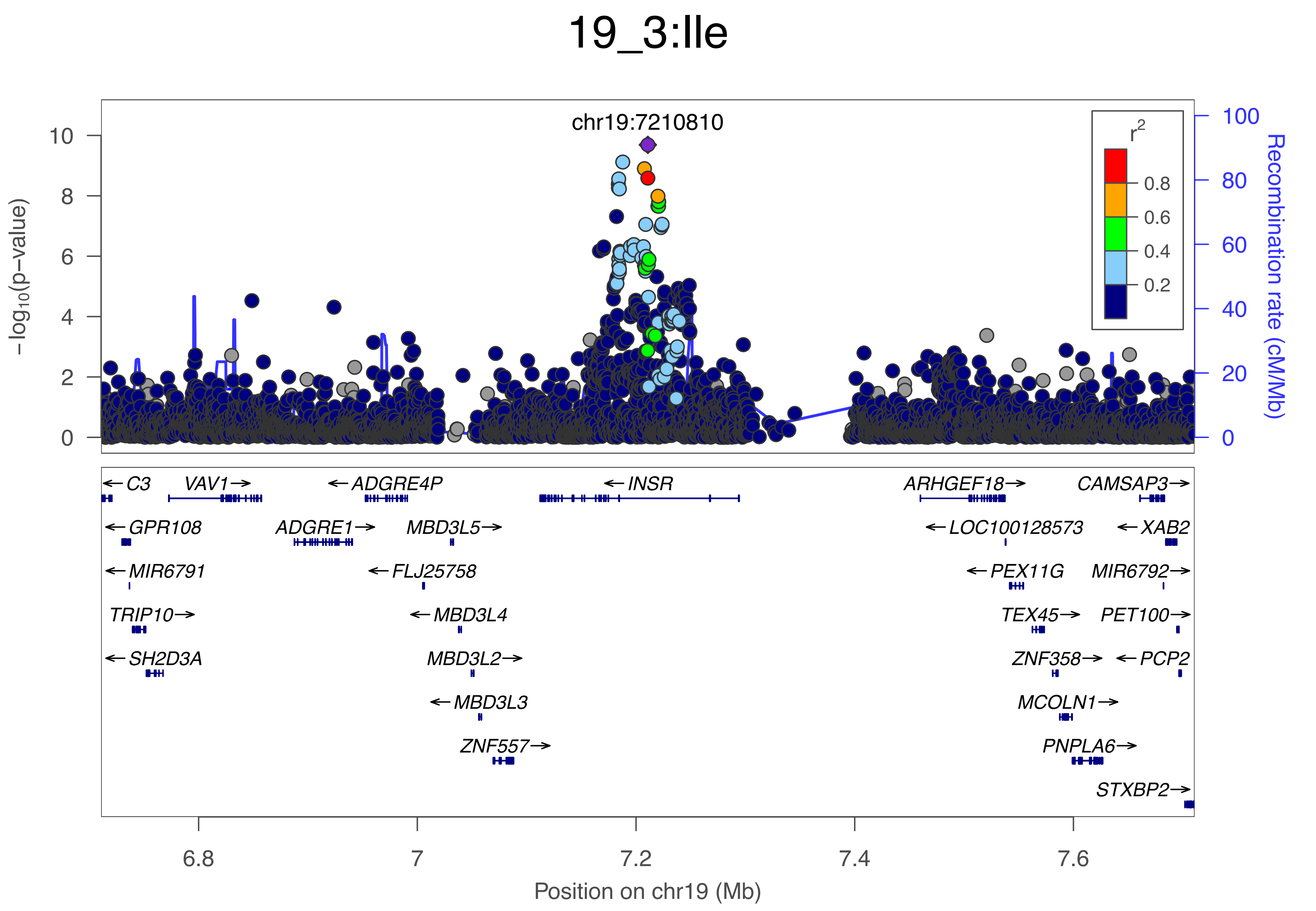

19_3:Ile
chr19:7210810
−log10(p−value)
Recombination rate (cM/Mb)
r2
0.8
0.6
0.4
0.2
C3
VAV1
ADGRE4P
INSR
ARHGEF18
CAMSAP3
GPR108
ADGRE1
MBD3L5
LOC100128573
XAB2
MIR6791
FLJ25758
PEX11G
MIR6792
TRIP10
MBD3L4
TEX45
PET100
SH2D3A
MBD3L2
ZNF358
PCP2
MBD3L3
MCOLN1
ZNF557
PNPLA6
STXBP2
Position on chr19 (Mb)

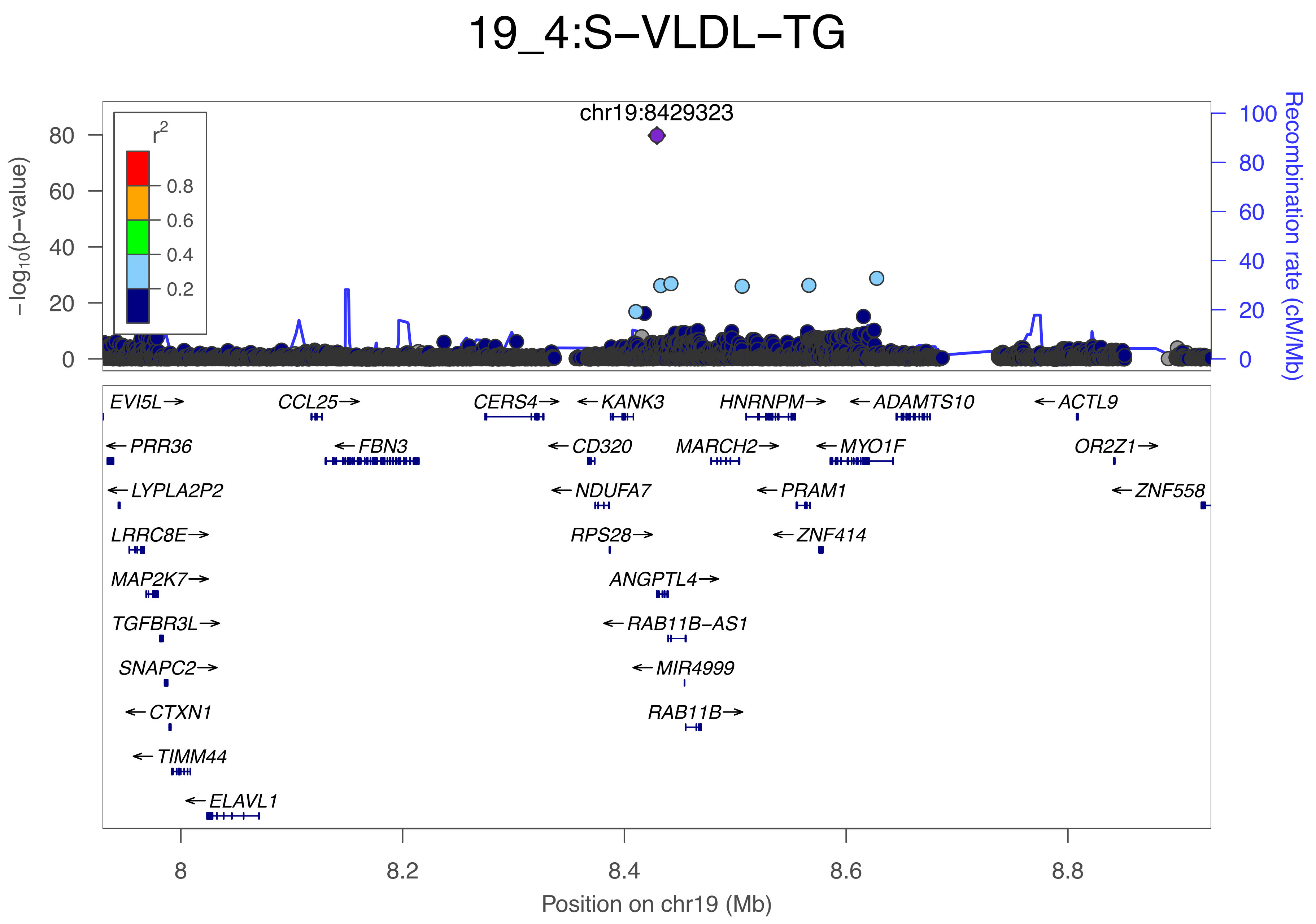

19_4:S−VLDL−TG
chr19:8429323
r2
0.8
0.6
0.4
0.2
−log10(p−value)
80
60
40
20
0
100
80
60
40
20
0
Recombination rate (cM/Mb)
EVI5L→
←PRR36
←LYPLA2P2
LRRC8E→
MAP2K7→
TGFBR3L→
SNAPC2→
←CTXN1
←TIMM44
←ELAVL1
CCL25→
←FBN3
CERS4→
←CD320
←NDUFA7
RPS28→
←KANK3
ANGPTL4→
←RAB11B−AS1
←MIR4999
RAB11B→
HNRNPM→
MARCH2→
←PRAM1
←ZNF414
←ADAMTS10
←MYO1F
←ACTL9
OR2Z1→
←ZNF558
8
8.2
8.4
8.6
8.8
Position on chr19 (Mb)

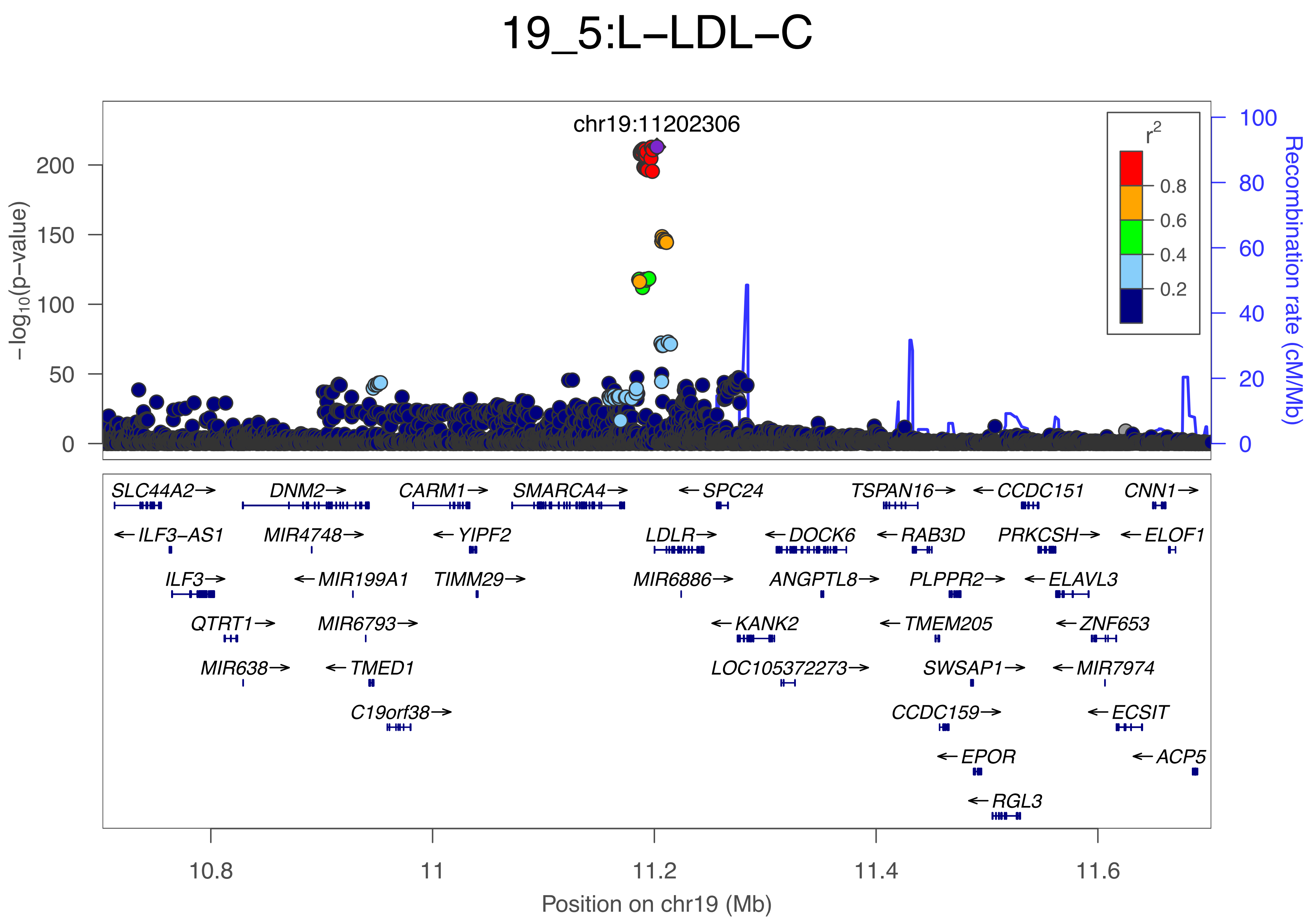
19_5:L−LDL−C
chr19:11202306
r2
0.8
0.6
0.4
0.2
−log10(p−value)
200
150
100
50
0
Recombination rate (cM/Mb)
100
80
60
40
20
0
SLC44A2→
DNM2→
CARM1→
SMARCA4→
←SPC24
TSPAN16→
←CCDC151
CNN1→
←ILF3−AS1
MIR4748→
←YIPF2
LDLR→
←DOCK6
←RAB3D
PRKCSH→
←ELOF1
ILF3→
←MIR199A1
TIMM29→
MIR6886→
ANGPTL8→
PLPPR2→
←ELAVL3
QTRT1→
MIR6793→
←KANK2
←TMEM205
←ZNF653
MIR638→
←TMED1
LOC105372273→
SWSAP1→
←MIR7974
C19orf38→
CCDC159→
←ECSIT
←EPOR
←ACP5
←RGL3
10.8
11
11.2
11.4
11.6
Position on chr19 (Mb)

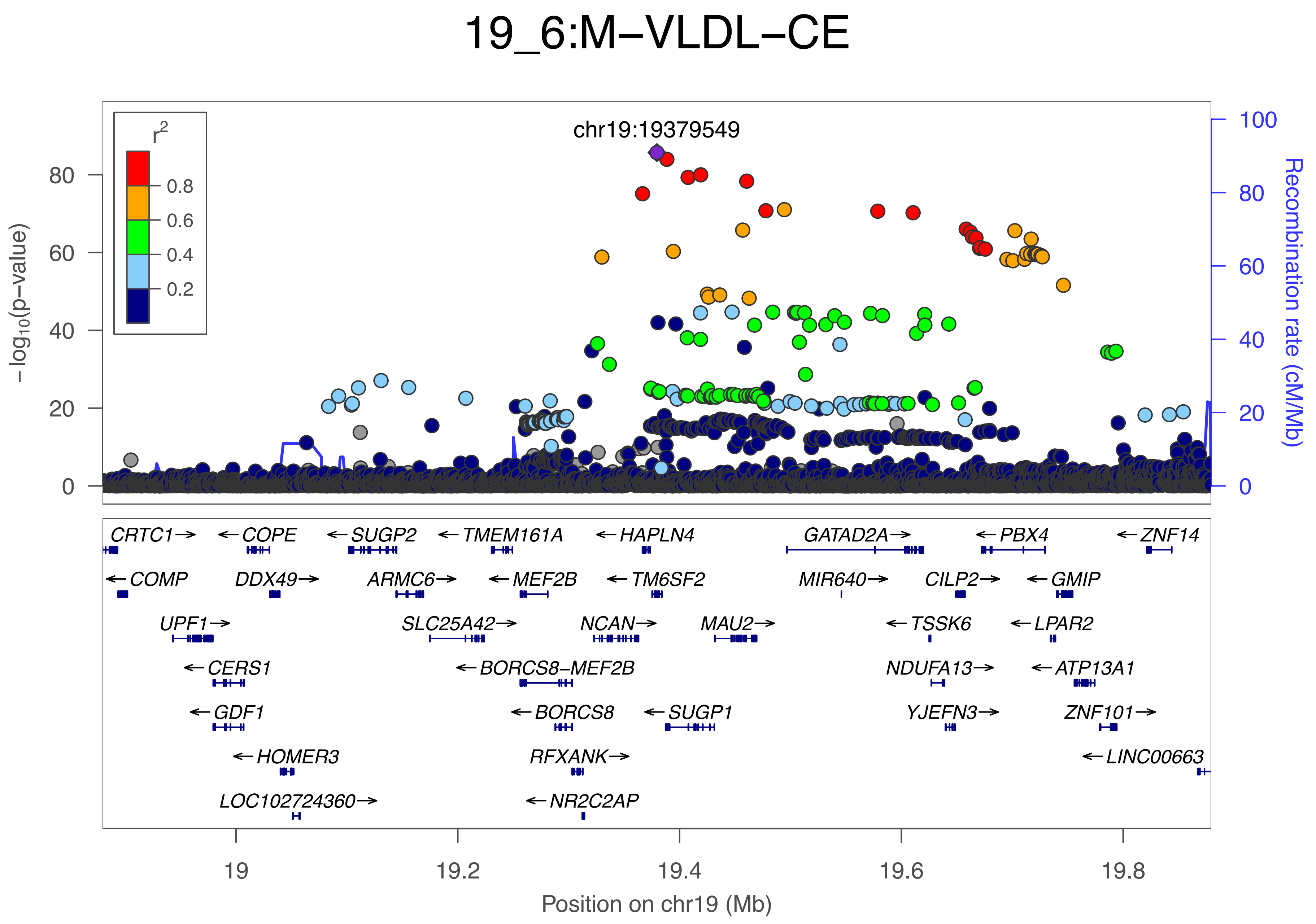

19_6:M−VLDL−CE
chr19:19379549
r2
0.8
0.6
0.4
0.2
−log10(p−value)
Recombination rate (cM/Mb)
0
20
40
60
80
100
CRTC1→
←COPE
←SUGP2
←TMEM161A
←HAPLN4
GATAD2A→
←PBX4
←ZNF14
←COMP
DDX49→
ARMC6→
←MEF2B
←TM6SF2
MIR640→
CILP2→
←GMIP
UPF1→
SLC25A42→
NCAN→
MAU2→
←TSSK6
←LPAR2
←CERS1
←BORCS8−MEF2B
NDUFA13→
←ATP13A1
←GDF1
←BORCS8
←SUGP1
YJEFN3→
ZNF101→
←HOMER3
RFXANK→
←LINC00663
LOC102724360→
←NR2C2AP
19
19.2
19.4
19.6
19.8
Position on chr19 (Mb)

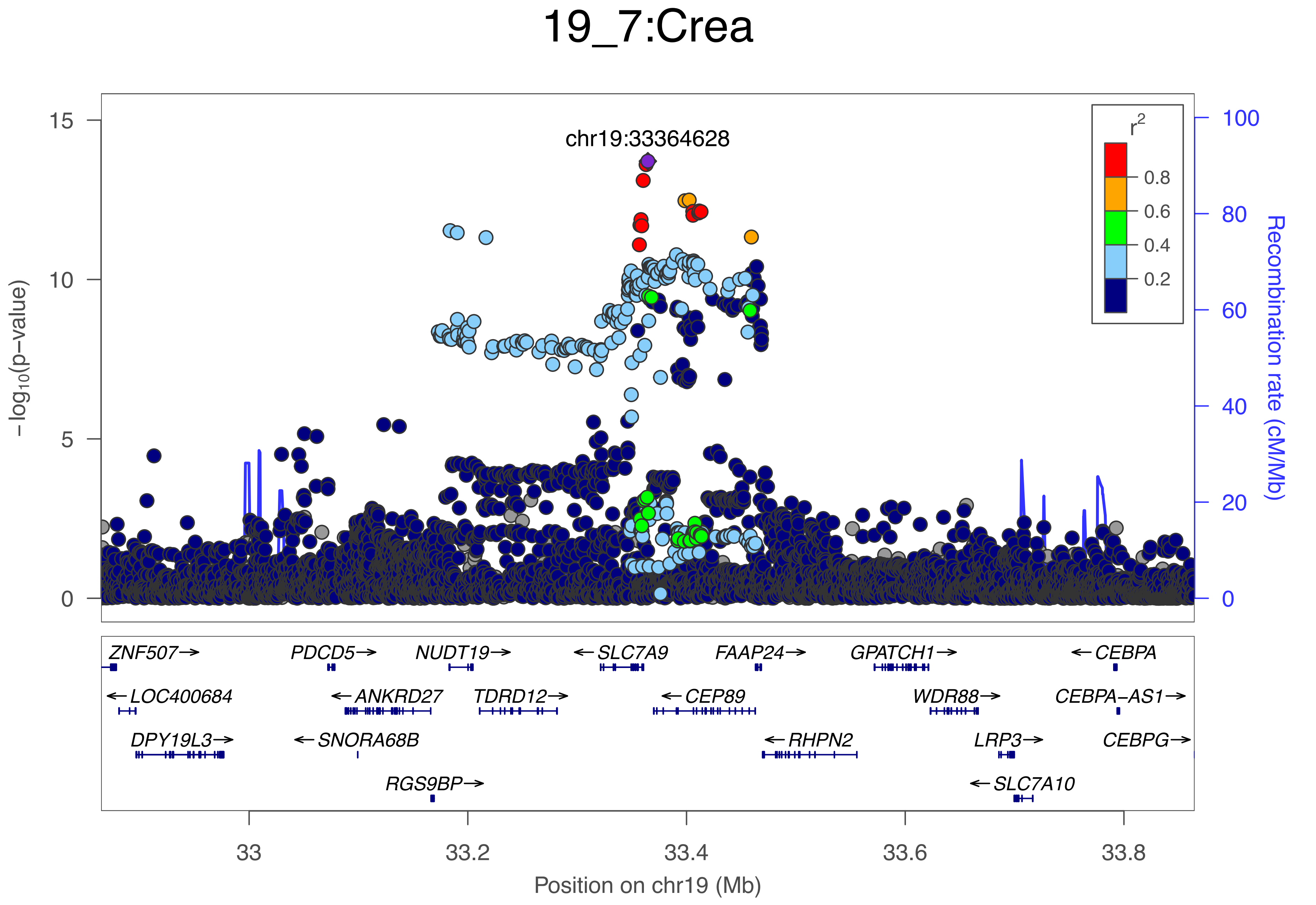

19_7:Crea
chr19:33364628
r2
0.8
0.6
0.4
0.2
−log10(p−value)
Recombination rate (cM/Mb)
15
10
5
0
100
80
60
40
20
0
ZNF507→
PDCD5→
NUDT19→
←SLC7A9
FAAP24→
GPATCH1→
←CEBPA
←LOC400684
←ANKRD27
TDRD12→
←CEP89
WDR88→
CEBPA−AS1→
DPY19L3→
←SNORA68B
←RHPN2
LRP3→
CEBPG→
RGS9BP→
←SLC7A10
33
33.2
33.4
33.6
33.8
Position on chr19 (Mb)

### 19_8:Alb

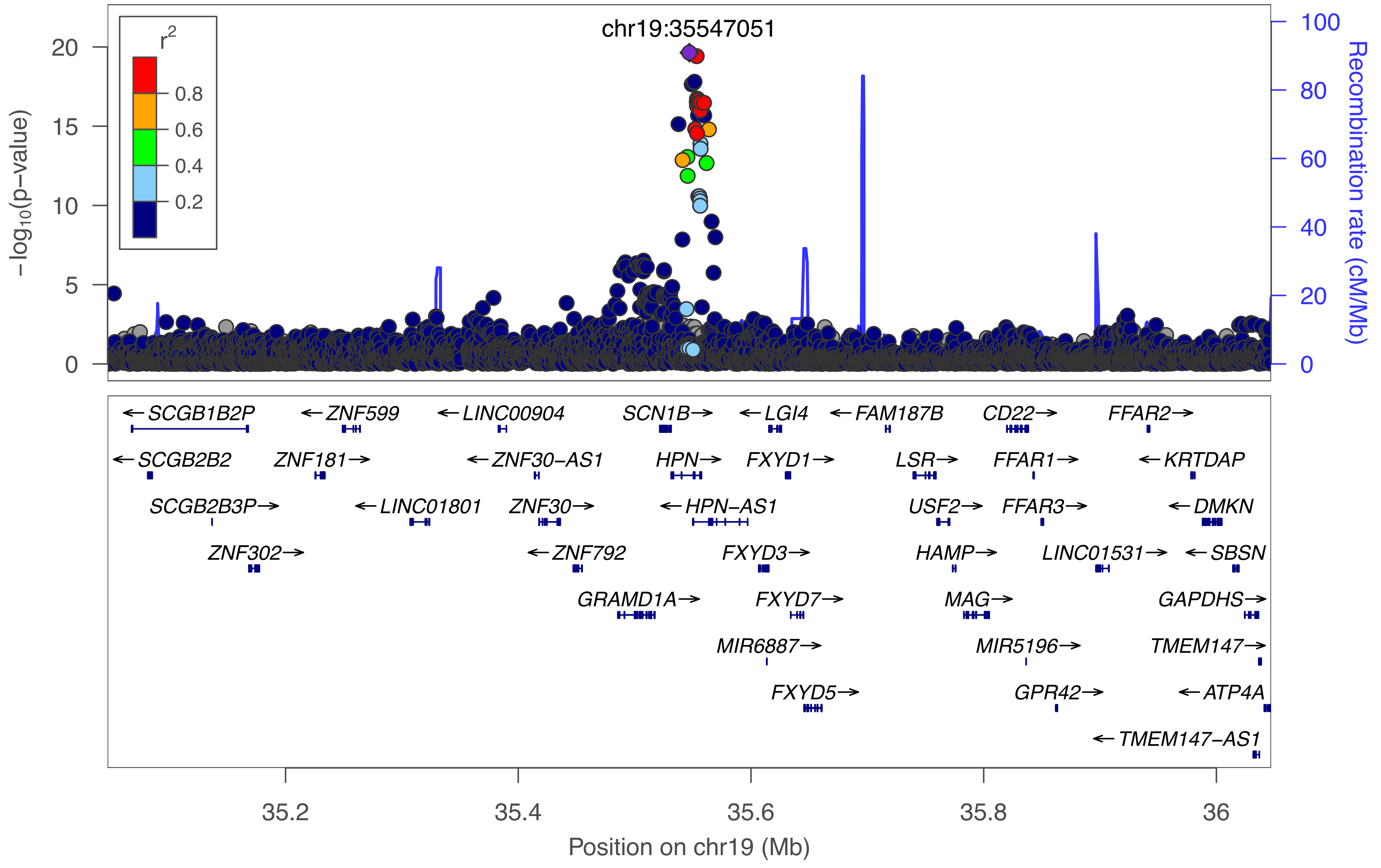

chr19:35547051
r²
0.8
0.6
0.4
0.2
−log10(p−value)
Recombination rate (cM/Mb)
0
5
10
15
20
0
20
40
60
80
100
←SCGB1B2P
←ZNF599
←LINC00904
SCN1B→
←LGI4
←FAM187B
CD22→
FFAR2→
←SCGB2B2
ZNF181→
←ZNF30−AS1
HPN→
FXYD1→
LSR→
FFAR1→
←KRTDAP
SCGB2B3P→
←LINC01801
ZNF30→
←HPN−AS1
USF2→
FFAR3→
←DMKN
ZNF302→
←ZNF792
FXYD3→
HAMP→
LINC01531→
←SBSN
GRAMD1A→
FXYD7→
MAG→
GAPDHS→
MIR6887→
MIR5196→
TMEM147→
FXYD5→
GPR42→
←ATP4A
←TMEM147−AS1
35.2
35.4
35.6
35.8
36
Position on chr19 (Mb)

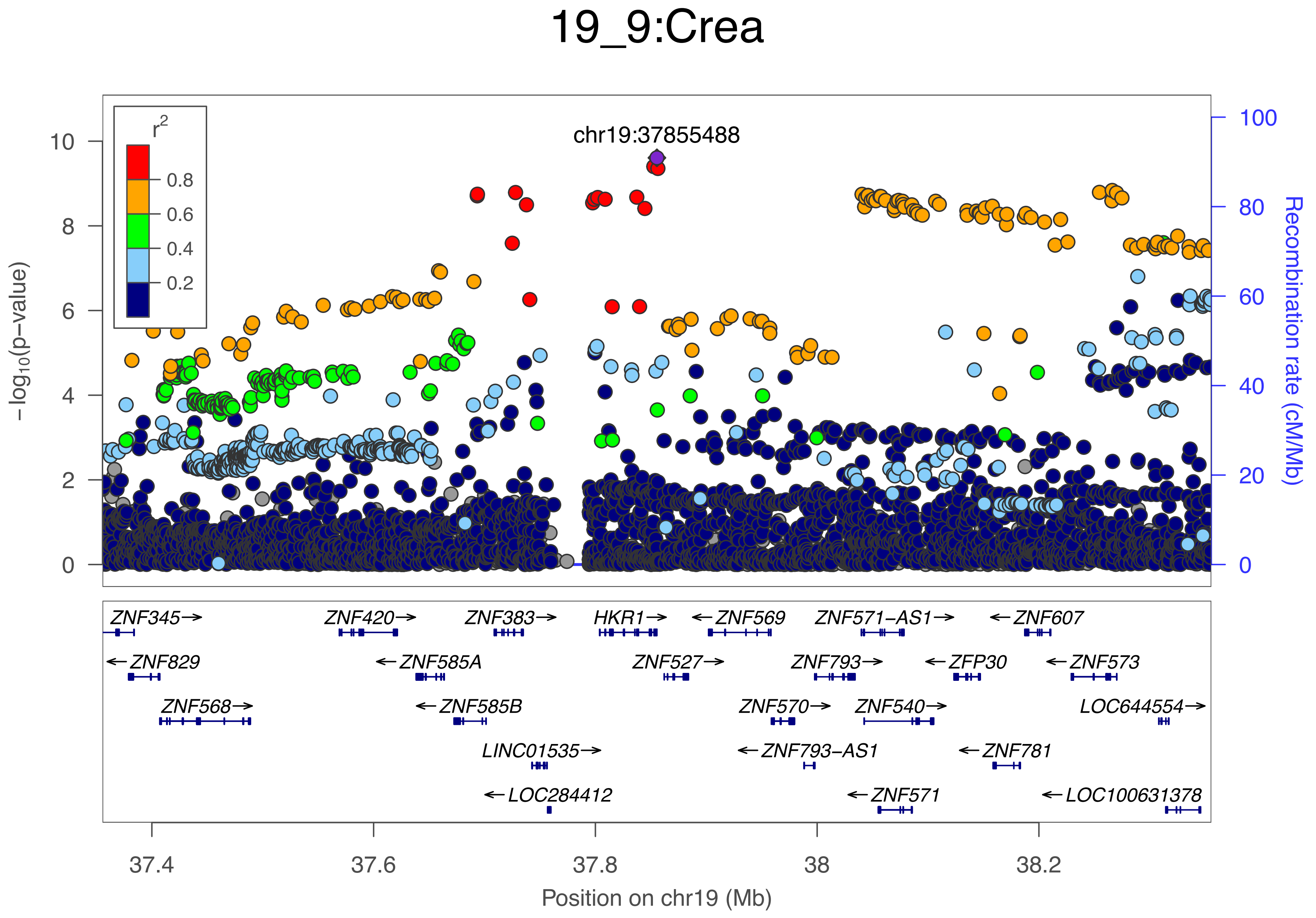
19_9:Crea
chr19:37855488
r2
0.8
0.6
0.4
0.2
-log10(p-value)
Recombination rate (cM/Mb)
10
8
6
4
2
0
100
80
60
40
20
0
ZNF345→
ZNF420→
ZNF383→
HKR1→
←ZNF569
ZNF571-AS1→
←ZNF607
←ZNF829
←ZNF585A
ZNF527→
ZNF793→
←ZFP30
←ZNF573
ZNF568→
←ZNF585B
ZNF570→
ZNF540→
LOC644554→
LINC01535→
←ZNF793-AS1
←ZNF781
←LOC284412
←ZNF571
←LOC100631378
37.4
37.6
37.8
38
38.2
Position on chr19 (Mb)

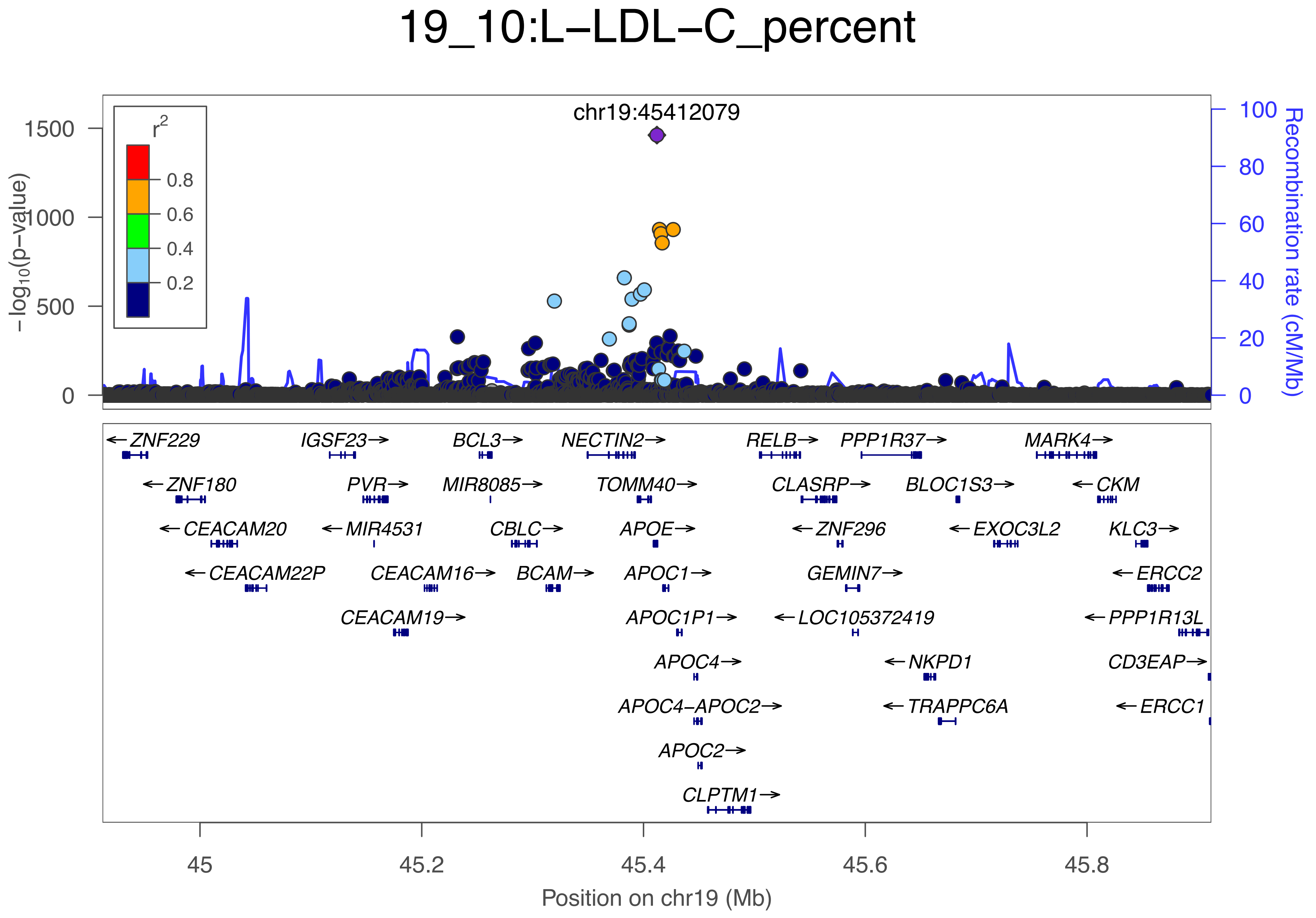

19_10:L−LDL−C_percent
chr19:45412079
r2
0.8
0.6
0.4
0.2
−log10(p−value)
1500
1000
500
0
Recombination rate (cM/Mb)
100
80
60
40
20
0
ZNF229
IGSF23
BCL3
NECTIN2
RELB
PPP1R37
MARK4
ZNF180
PVR
MIR8085
TOMM40
CLASRP
BLOC1S3
CKM
CEACAM20
MIR4531
CBLC
APOE
ZNF296
EXOC3L2
KLC3
CEACAM22P
CEACAM16
BCAM
APOC1
GEMIN7
ERCC2
CEACAM19
APOC1P1
LOC105372419
PPP1R13L
APOC4
NKPD1
CD3EAP
APOC4−APOC2
TRAPPC6A
ERCC1
APOC2
CLPTM1
45
45.2
45.4
45.6
45.8
Position on chr19 (Mb)

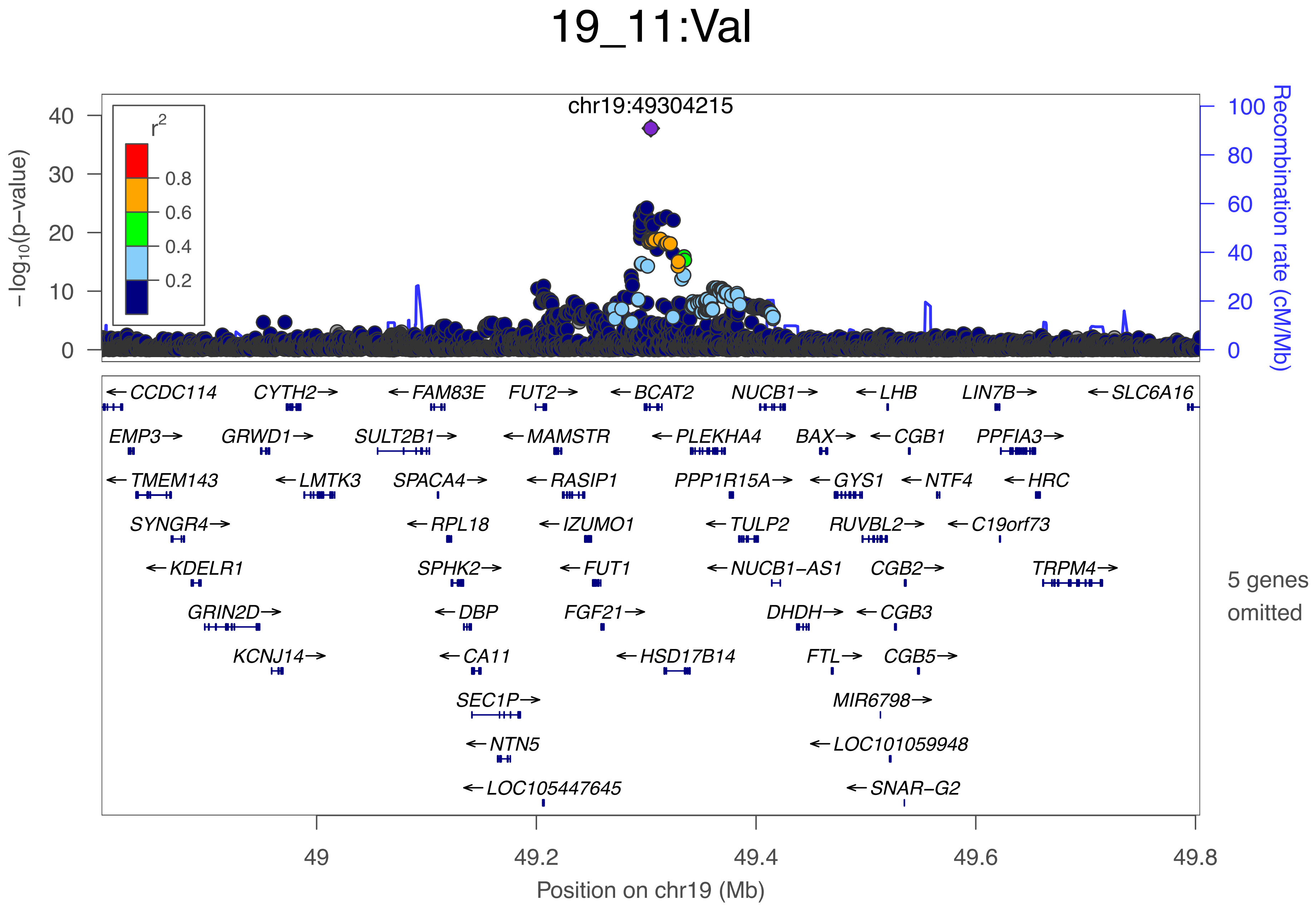
19_11:Val
chr19:49304215
r2
0.8
0.6
0.4
0.2
−log10(p−value)
0
10
20
30
40
Recombination rate (cM/Mb)
0
20
40
60
80
100
←CCDC114
CYTH2→
←FAM83E
FUT2→
←BCAT2
NUCB1→
←LHB
LIN7B→
←SLC6A16
EMP3→
GRWD1→
SULT2B1→
←MAMSTR
←PLEKHA4
BAX→
←CGB1
PPFIA3→
←TMEM143
←LMTK3
SPACA4→
←RASIP1
PPP1R15A→
←GYS1
←NTF4
←HRC
SYNGR4→
←RPL18
←IZUMO1
←TULP2
RUVBL2→
←C19orf73
←KDELR1
SPHK2→
←FUT1
←NUCB1−AS1
CGB2→
TRPM4→
GRIN2D→
←DBP
FGF21→
DHDH→
←CGB3
KCNJ14→
←CA11
←HSD17B14
FTL→
CGB5→
SEC1P→
MIR6798→
←NTN5
←LOC101059948
←LOC105447645
←SNAR−G2
5 genes omitted
49
49.2
49.4
49.6
49.8
Position on chr19 (Mb)

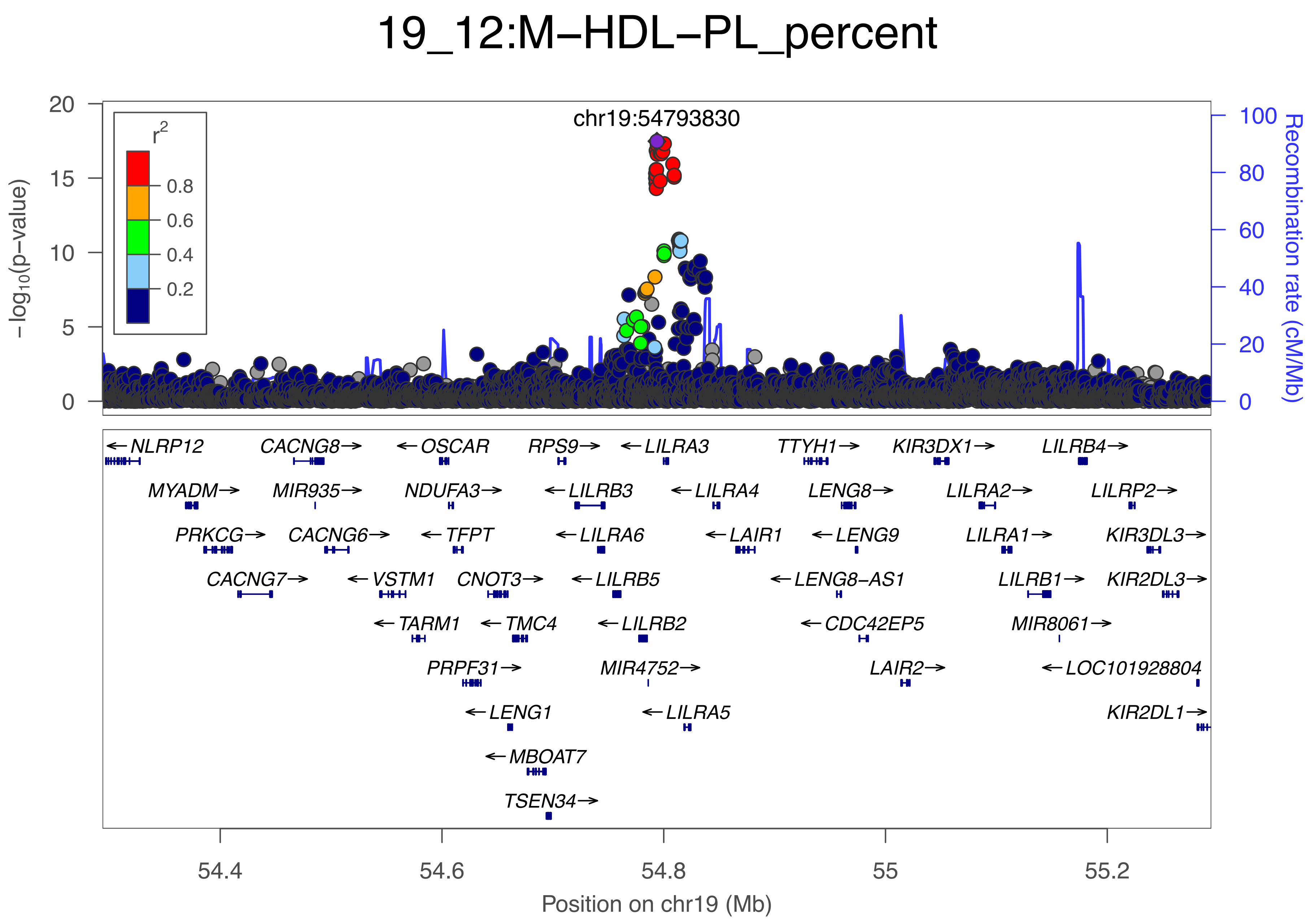

19_12:M−HDL−PL_percent
chr19:54793830
r²
0.8
0.6
0.4
0.2
−log10(p−value)
Recombination rate (cM/Mb)
20
15
10
5
0
100
80
60
40
20
0
←NLRP12
CACNG8→
←OSCAR
RPS9→
←LILRA3
TTYH1→
KIR3DX1→
LILRB4→
MYADM→
MIR935→
NDUFA3→
←LILRB3
←LILRA4
LENG8→
LILRA2→
LILRP2→
PRKCG→
CACNG6→
←TFPT
←LILRA6
←LAIR1
←LENG9
LILRA1→
KIR3DL3→
CACNG7→
←VSTM1
CNOT3→
←LILRB5
←LENG8−AS1
LILRB1→
KIR2DL3→
←TARM1
←TMC4
←LILRB2
←CDC42EP5
MIR8061→
PRPF31→
MIR4752→
LAIR2→
←LOC101928804
←LENG1
←LILRA5
KIR2DL1→
←MBOAT7
TSEN34→
54.4
54.6
54.8
55
55.2
Position on chr19 (Mb)

### 19_13:Serum−TG

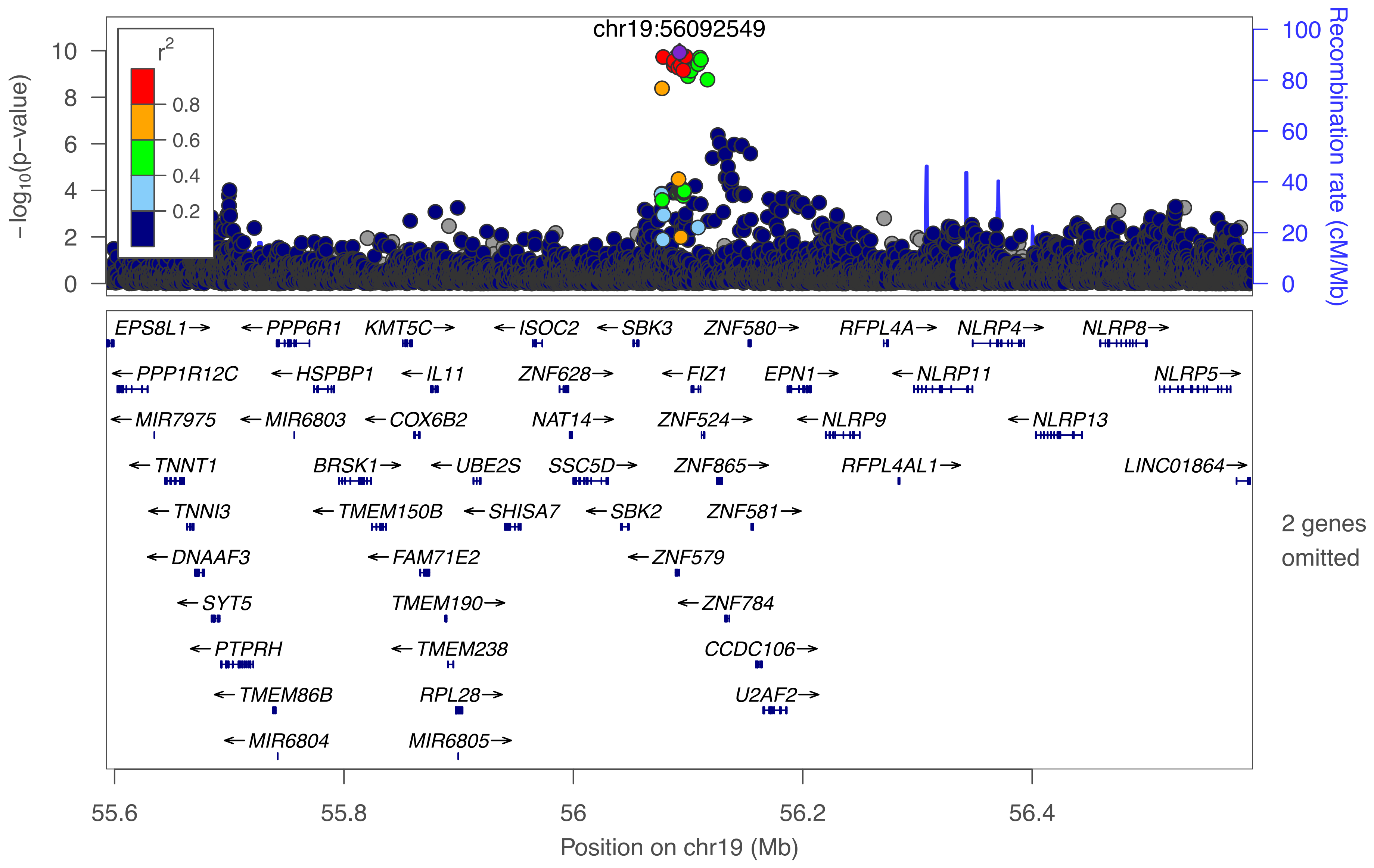

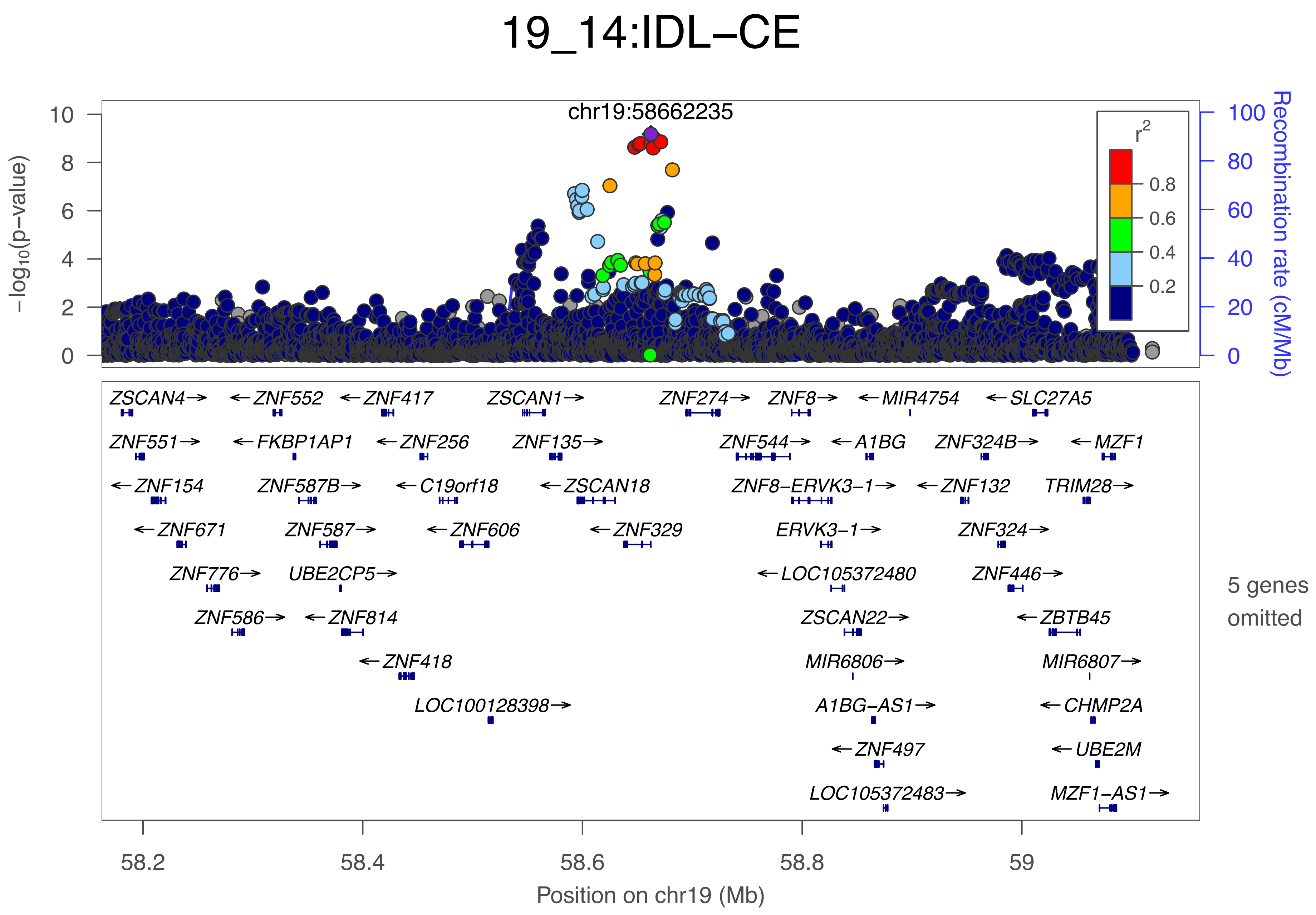
19_14:IDL−CE
chr19:58662235
−log10(p−value)
Recombination rate (cM/Mb)
r2
0.8
0.6
0.4
0.2
0
2
4
6
8
10
0
20
40
60
80
100
ZSCAN4→
←ZNF552
←ZNF417
ZSCAN1→
ZNF274→
ZNF8→
←MIR4754
←SLC27A5
ZNF551→
←FKBP1AP1
←ZNF256
ZNF135→
ZNF544→
←A1BG
ZNF324B→
←MZF1
←ZNF154
ZNF587B→
←C19orf18
←ZSCAN18
ZNF8−ERVK3−1→
←ZNF132
TRIM28→
←ZNF671
ZNF587→
←ZNF606
←ZNF329
ERVK3−1→
ZNF324→
ZNF776→
UBE2CP5→
←LOC105372480
ZNF446→
ZNF586→
←ZNF814
ZSCAN22→
←ZBTB45
←ZNF418
MIR6806→
MIR6807→
LOC100128398→
A1BG−AS1→
←CHMP2A
←ZNF497
←UBE2M
LOC105372483→
MZF1−AS1→
5 genes omitted
58.2
58.4
58.6
58.8
59
Position on chr19 (Mb)

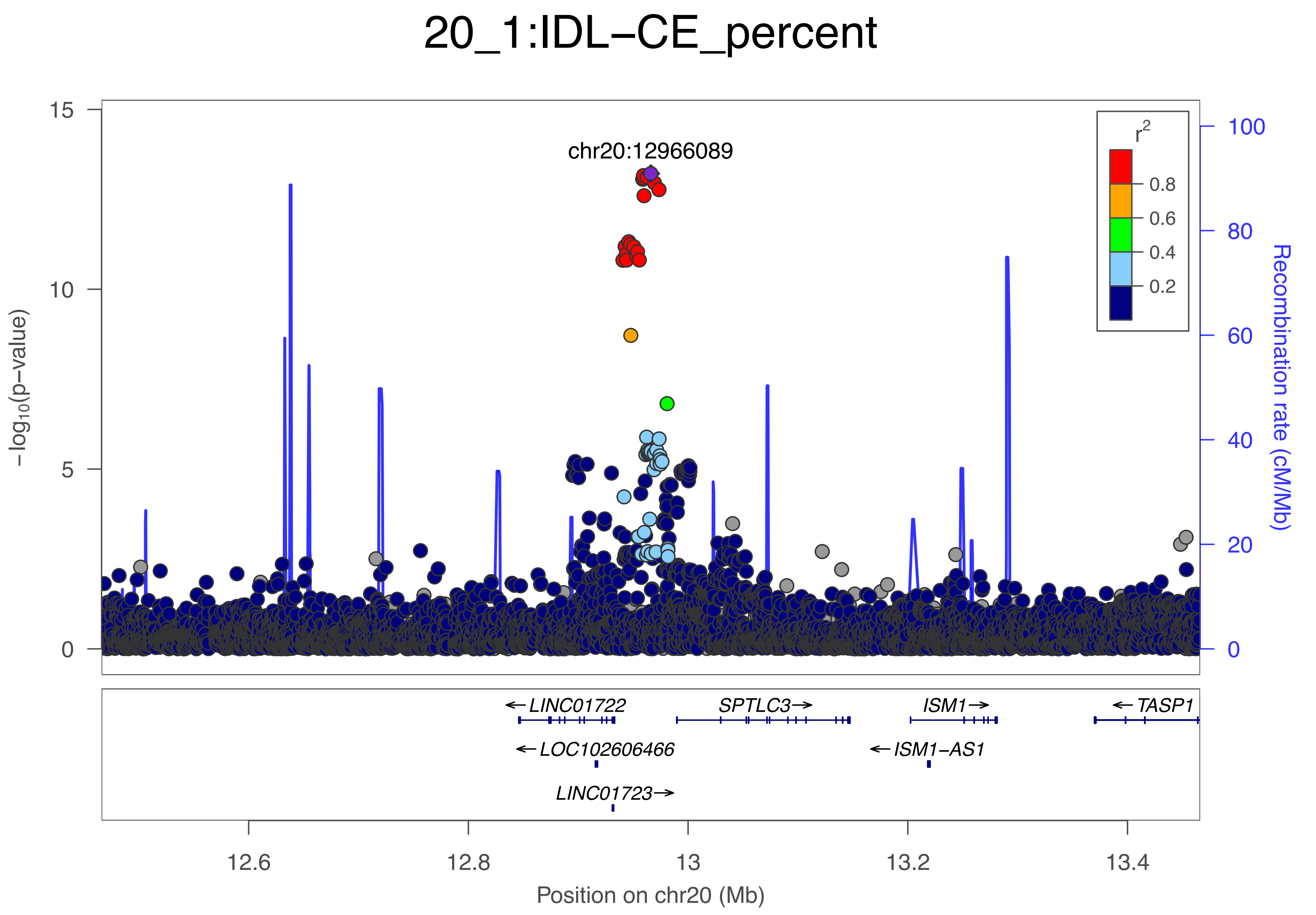

20_1:IDL−CE_percent
chr20:12966089
−log10(p−value)
15
10
5
0
r2
0.8
0.6
0.4
0.2
100
80
60
40
20
0
Recombination rate (cM/Mb)
←LINC01722
SPTLC3→
ISM1→
←TASP1
←LOC102606466
←ISM1−AS1
LINC01723→
12.6
12.8
13
13.2
13.4
Position on chr20 (Mb)

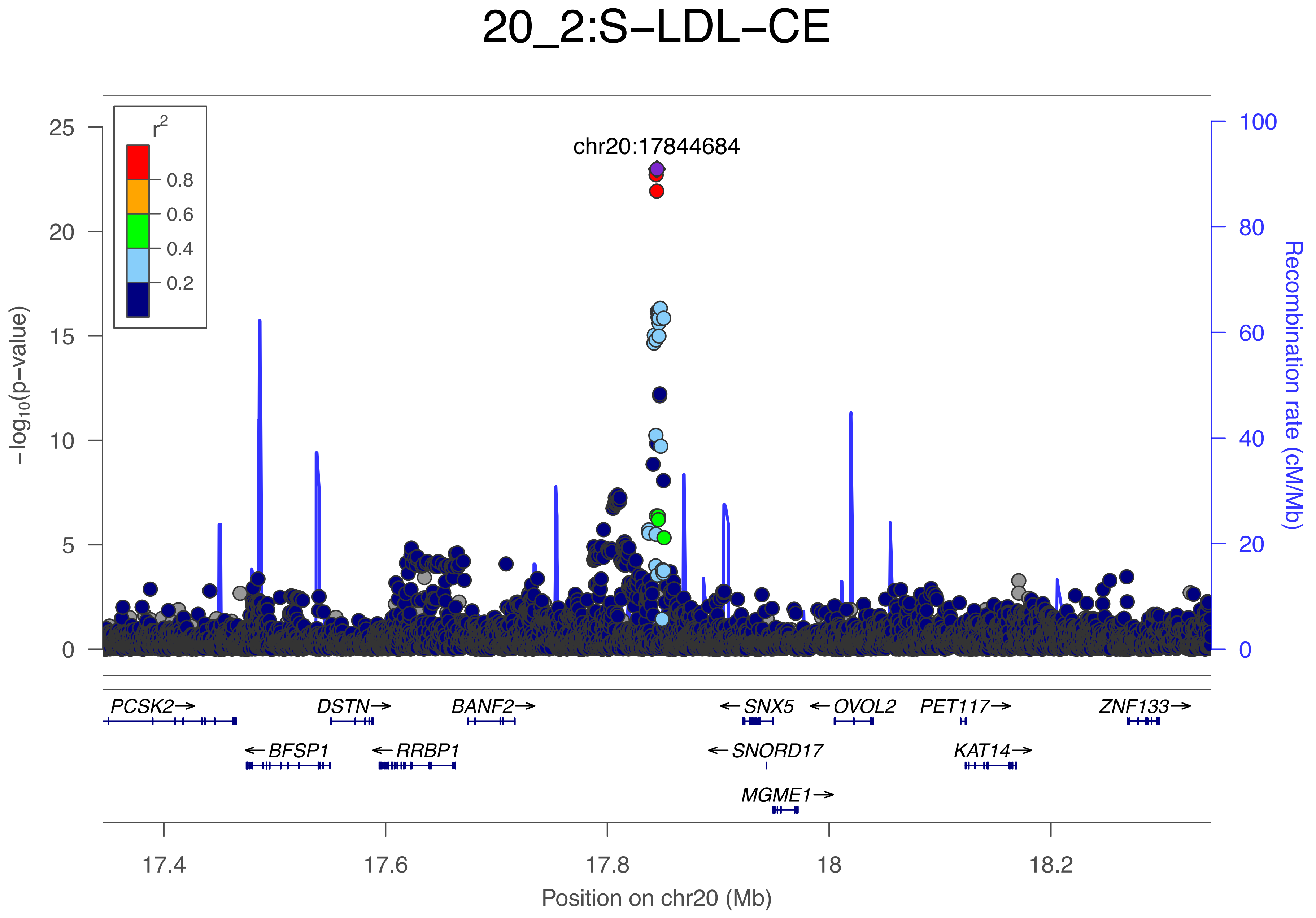

20_2:S−LDL−CE
r2
0.8
0.6
0.4
0.2
chr20:17844684
−log10(p−value)
Recombination rate (cM/Mb)
0
5
10
15
20
25
0
20
40
60
80
100
PCSK2→
DSTN→
BANF2→
←SNX5
←OVOL2
PET117→
ZNF133→
←BFSP1
←RRBP1
←SNORD17
KAT14→
MGME1→
17.4
17.6
17.8
18
18.2
Position on chr20 (Mb)

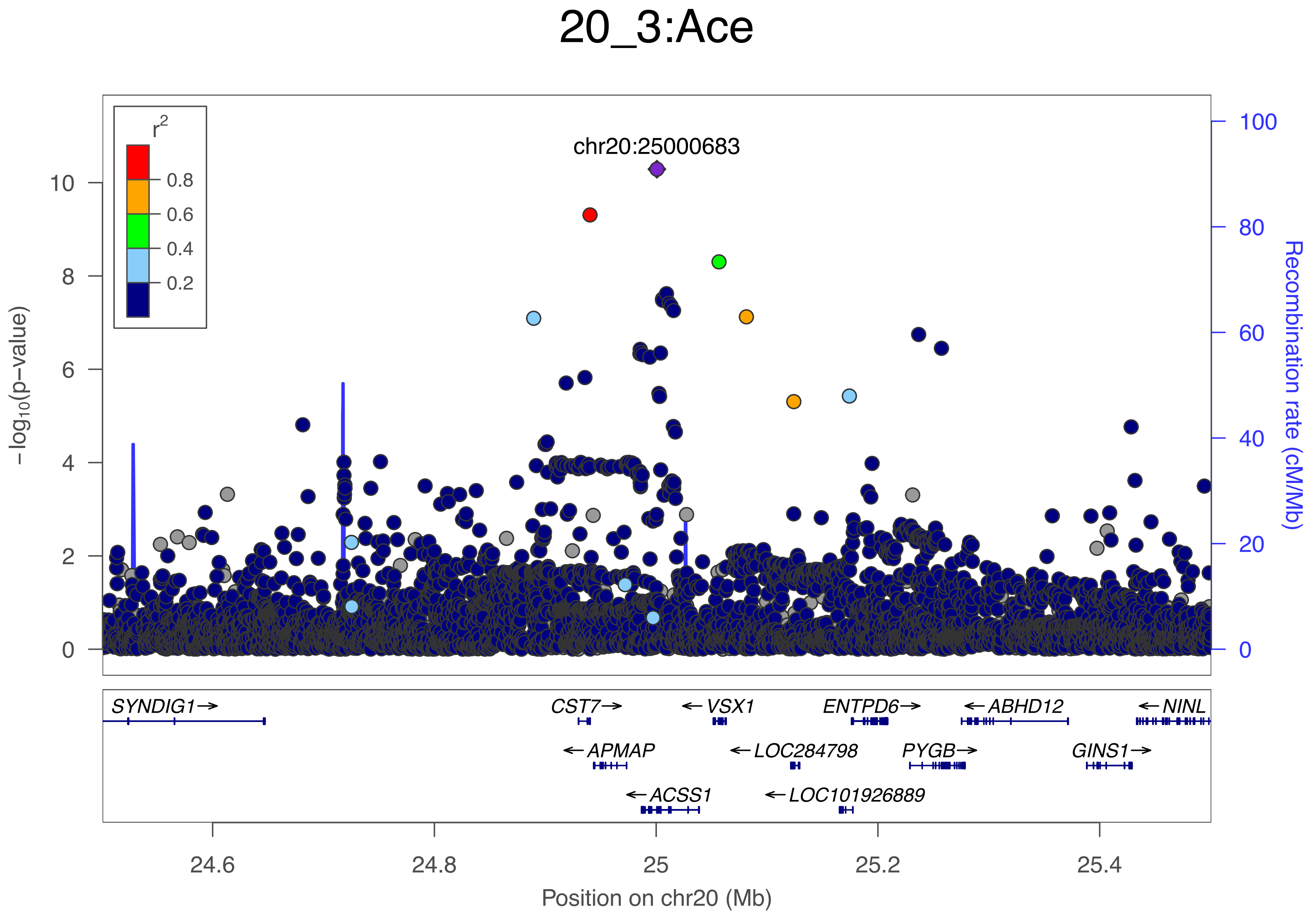

20_3:Ace
chr20:25000683
r2
0.8
0.6
0.4
0.2
−log10(p−value)
Recombination rate (cM/Mb)
0
2
4
6
8
10
0
20
40
60
80
100
SYNDIG1→
CST7→
←VSX1
ENTPD6→
←ABHD12
←NINL
←APMAP
←LOC284798
PYGB→
GINS1→
←ACSS1
←LOC101926889
24.6
24.8
25
25.2
25.4
Position on chr20 (Mb)

### 20_4:UnsatDeg

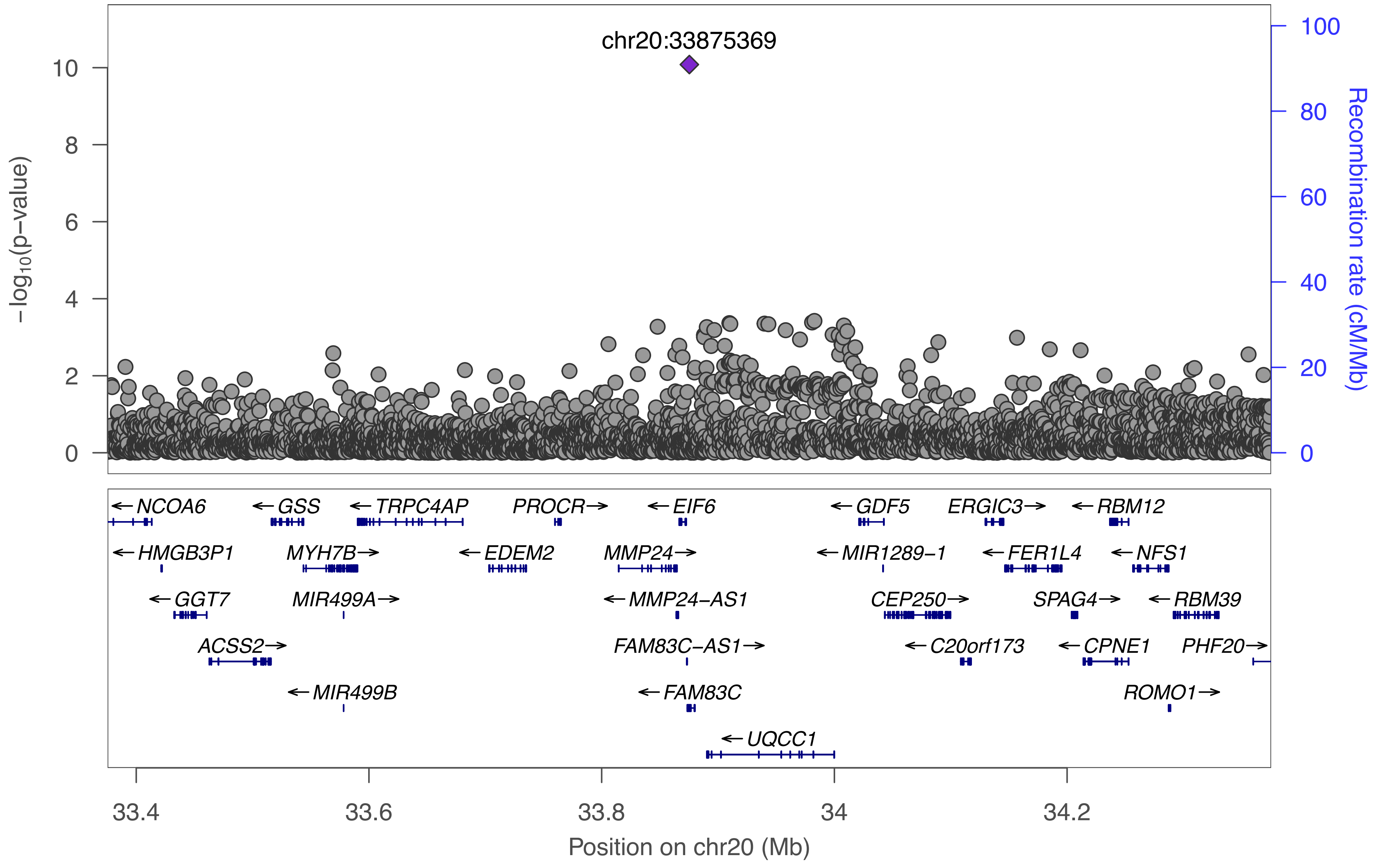

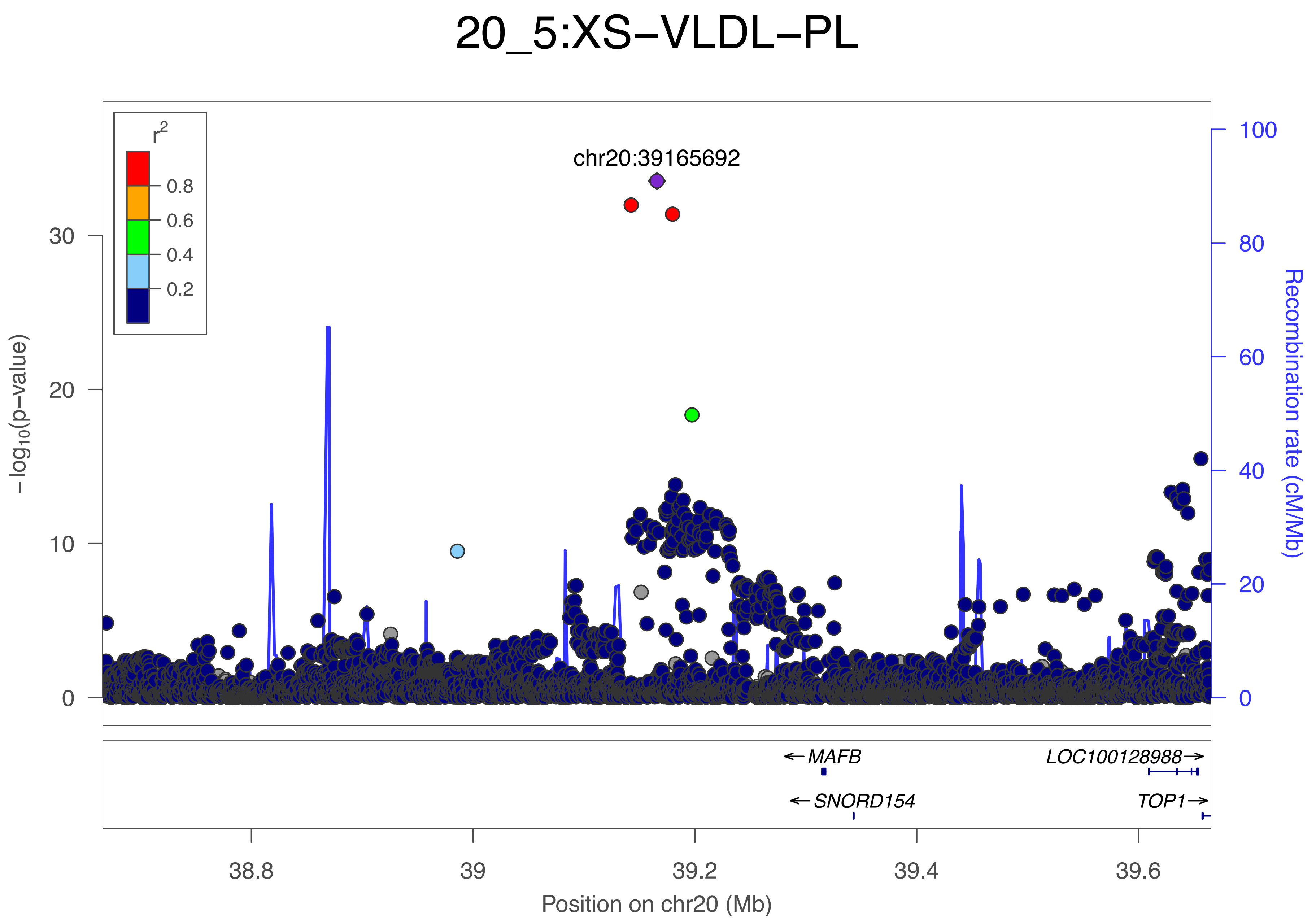

20_5:XS−VLDL−PL
chr20:39165692
r2
0.8
0.6
0.4
0.2
−log10(p−value)
0
10
20
30
Recombination rate (cM/Mb)
0
20
40
60
80
100
MAFB
LOC100128988
SNORD154
TOP1
38.8
39
39.2
39.4
39.6
Position on chr20 (Mb)

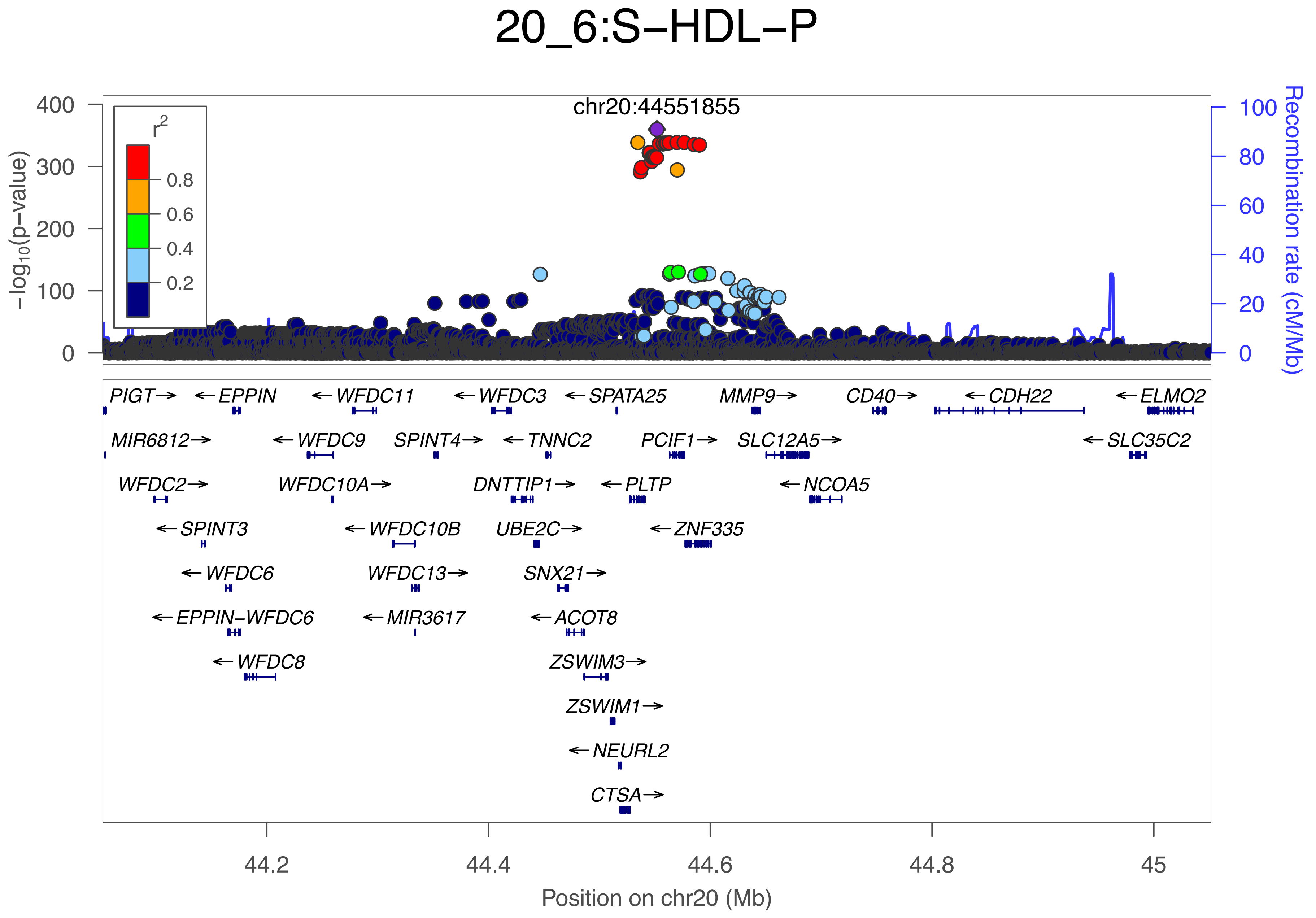

20_6:S−HDL−P
chr20:44551855
r2
0.8
0.6
0.4
0.2
−log10(p−value)
Recombination rate (cM/Mb)
400
300
200
100
0
100
80
60
40
20
0
PIGT→
←EPPIN
←WFDC11
←WFDC3
←SPATA25
MMP9→
CD40→
←CDH22
←ELMO2
MIR6812→
←WFDC9
SPINT4→
←TNNC2
PCIF1→
SLC12A5→
←SLC35C2
WFDC2→
WFDC10A→
DNTTIP1→
←PLTP
←NCOA5
←SPINT3
←WFDC10B
UBE2C→
←ZNF335
←WFDC6
WFDC13→
SNX21→
←EPPIN−WFDC6
←MIR3617
←ACOT8
←WFDC8
ZSWIM3→
ZSWIM1→
←NEURL2
CTSA→
44.2
44.4
44.6
44.8
45
Position on chr20 (Mb)

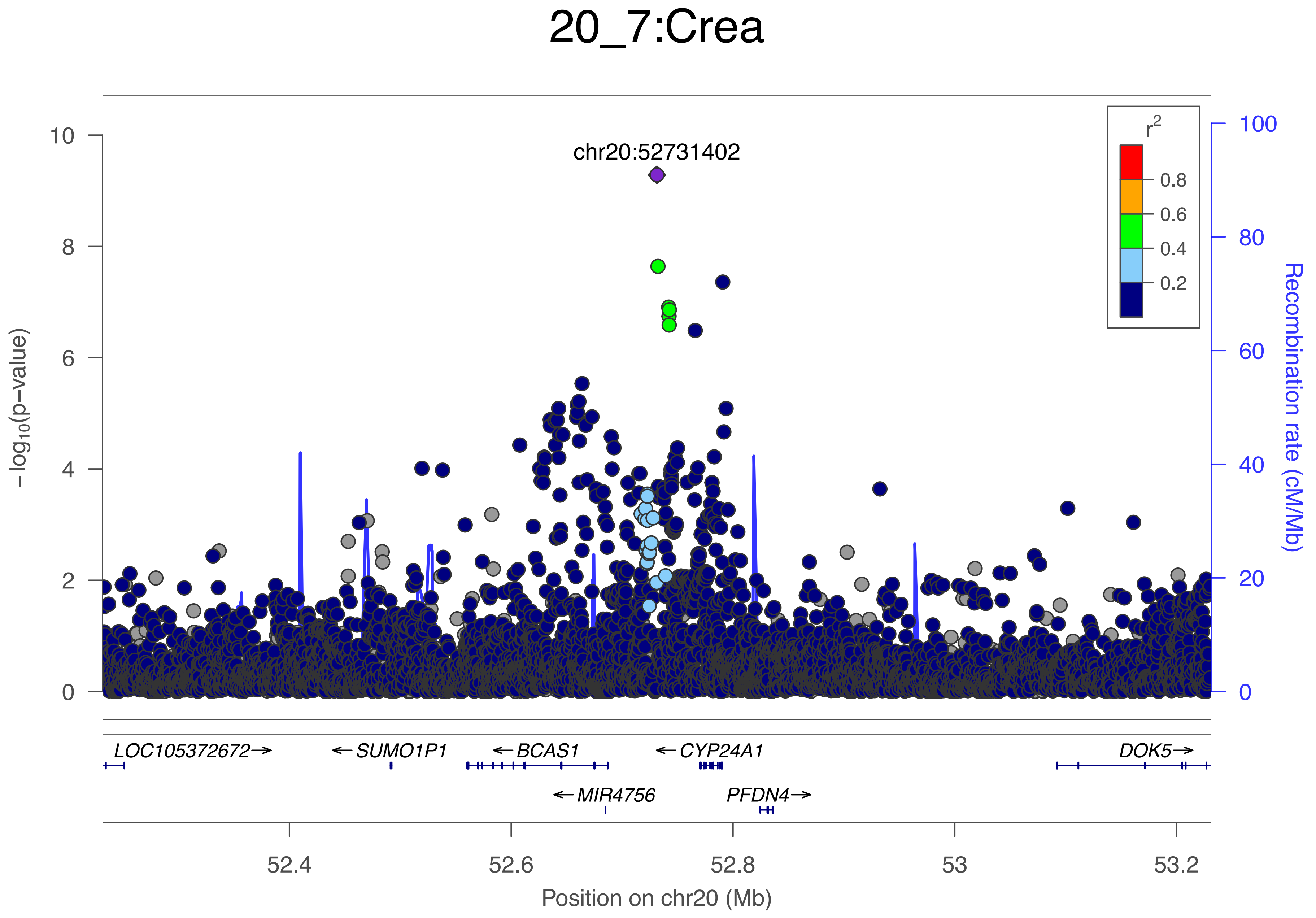
20_7:Crea
chr20:52731402
r2
0.8
0.6
0.4
0.2
-log10(p-value)
Recombination rate (cM/Mb)
10
8
6
4
2
0
100
80
60
40
20
0
LOC105372672→
←SUMO1P1
←BCAS1
←CYP24A1
DOK5→
←MIR4756
PFDN4→
52.4
52.6
52.8
53
53.2
Position on chr20 (Mb)

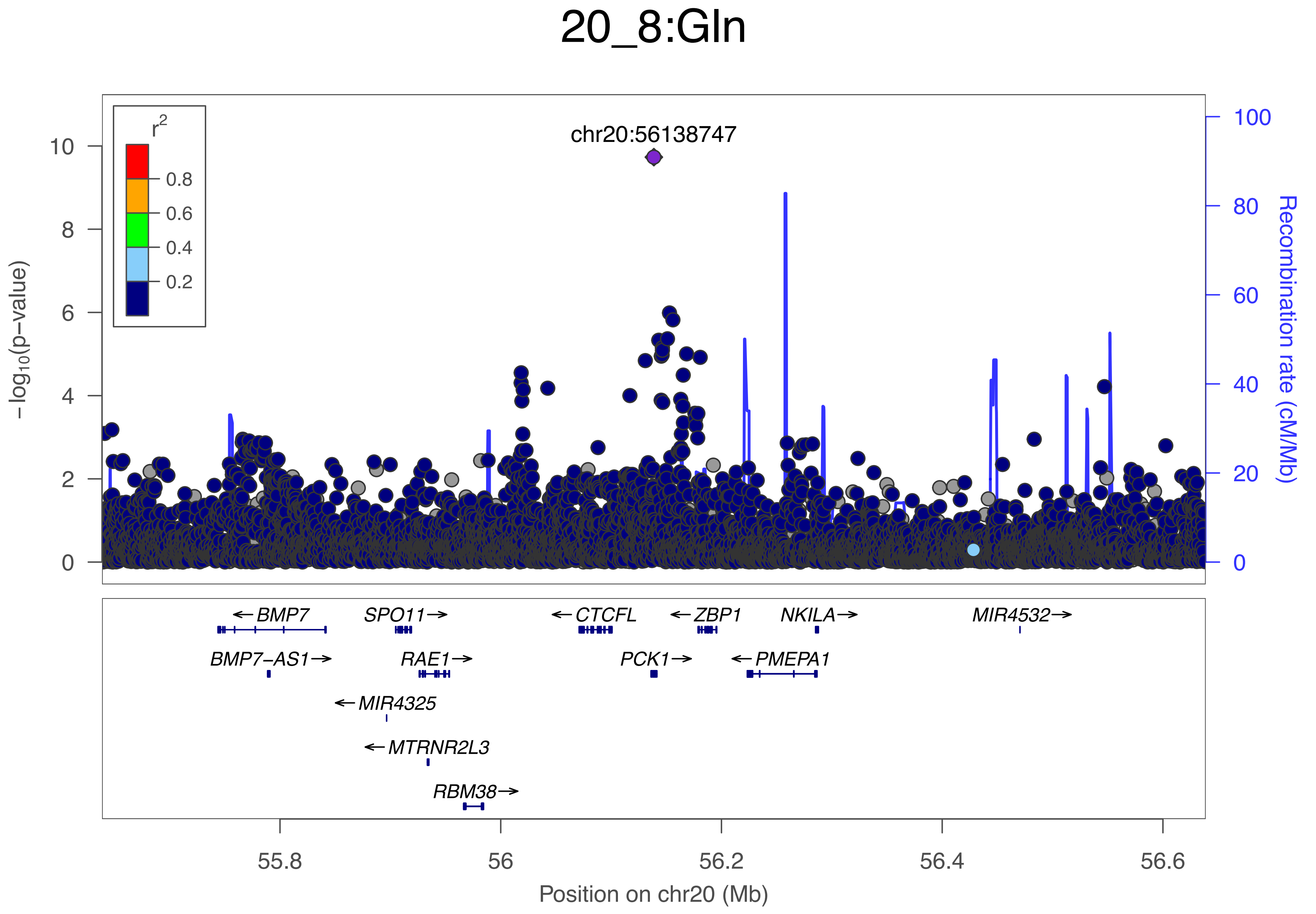

20_8:Gln
chr20:56138747
r2
0.8
0.6
0.4
0.2
−log10(p−value)
Recombination rate (cM/Mb)
0
2
4
6
8
10
0
20
40
60
80
100
BMP7
SPO11
CTCFL
ZBP1
NKILA
MIR4532
BMP7−AS1
RAE1
PCK1
PMEPA1
MIR4325
MTRNR2L3
RBM38
55.8
56
56.2
56.4
56.6
Position on chr20 (Mb)

### 20_9:Phe

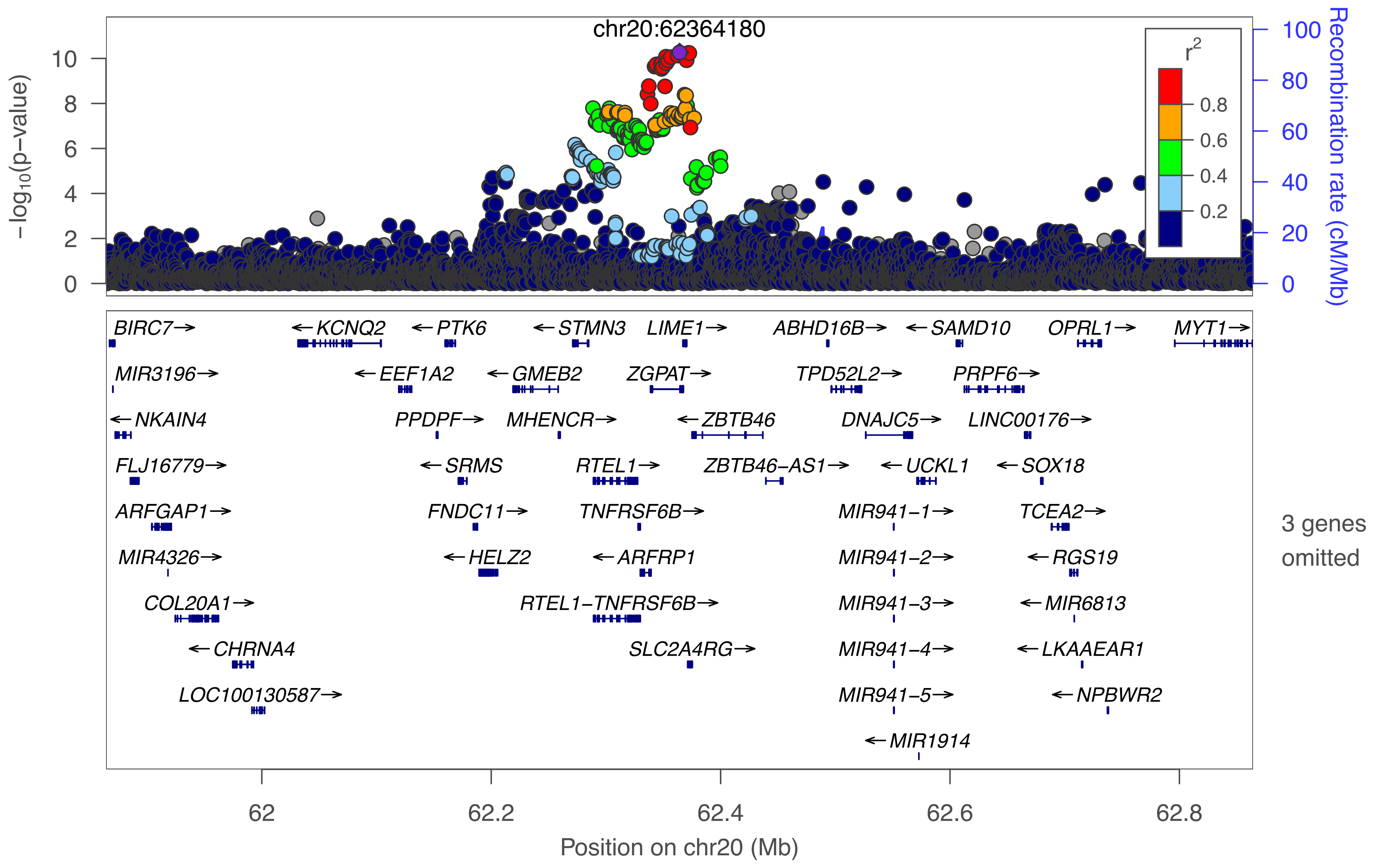

chr20:62364180
−log10(p−value)
0
2
4
6
8
10
Recombination rate (cM/Mb)
0
20
40
60
80
100
r2
0.2
0.4
0.6
0.8
BIRC7→
←KCNQ2
←PTK6
←STMN3
LIME1→
ABHD16B→
←SAMD10
OPRL1→
MYT1→
MIR3196→
←EEF1A2
←GMEB2
ZGPAT→
TPD52L2→
PRPF6→
←NKAIN4
PPDPF→
MHENCR→
←ZBTB46
DNAJC5→
LINC00176→
FLJ16779→
←SRMS
RTEL1→
ZBTB46−AS1→
←UCKL1
←SOX18
ARFGAP1→
FNDC11→
TNFRSF6B→
MIR941−1→
TCEA2→
MIR4326→
←HELZ2
←ARFRP1
MIR941−2→
←RGS19
COL20A1→
RTEL1−TNFRSF6B→
MIR941−3→
←MIR6813
←CHRNA4
SLC2A4RG→
MIR941−4→
←LKAAEAR1
LOC100130587→
MIR941−5→
←NPBWR2
←MIR1914
3 genes omitted
62
62.2
62.4
62.6
62.8
Position on chr20 (Mb)

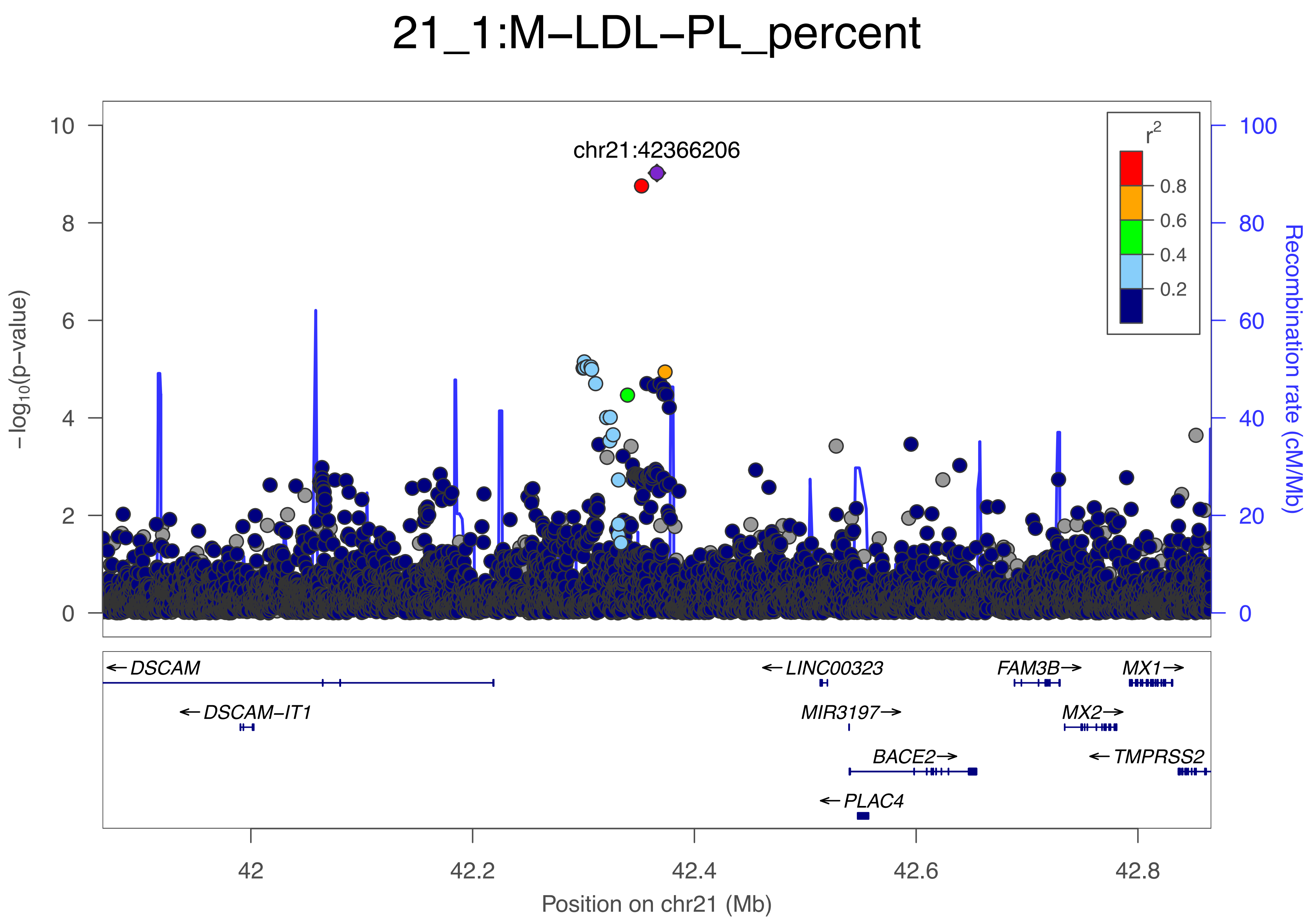
21_1:M−LDL−PL_percent
chr21:42366206
r2
0.8
0.6
0.4
0.2
−log10(p−value)
Recombination rate (cM/Mb)
0
2
4
6
8
10
20
40
60
80
100
DSCAM
DSCAM−IT1
LINC00323
MIR3197
BACE2
PLAC4
FAM3B
MX2
MX1
TMPRSS2
42
42.2
42.4
42.6
42.8
Position on chr21 (Mb)

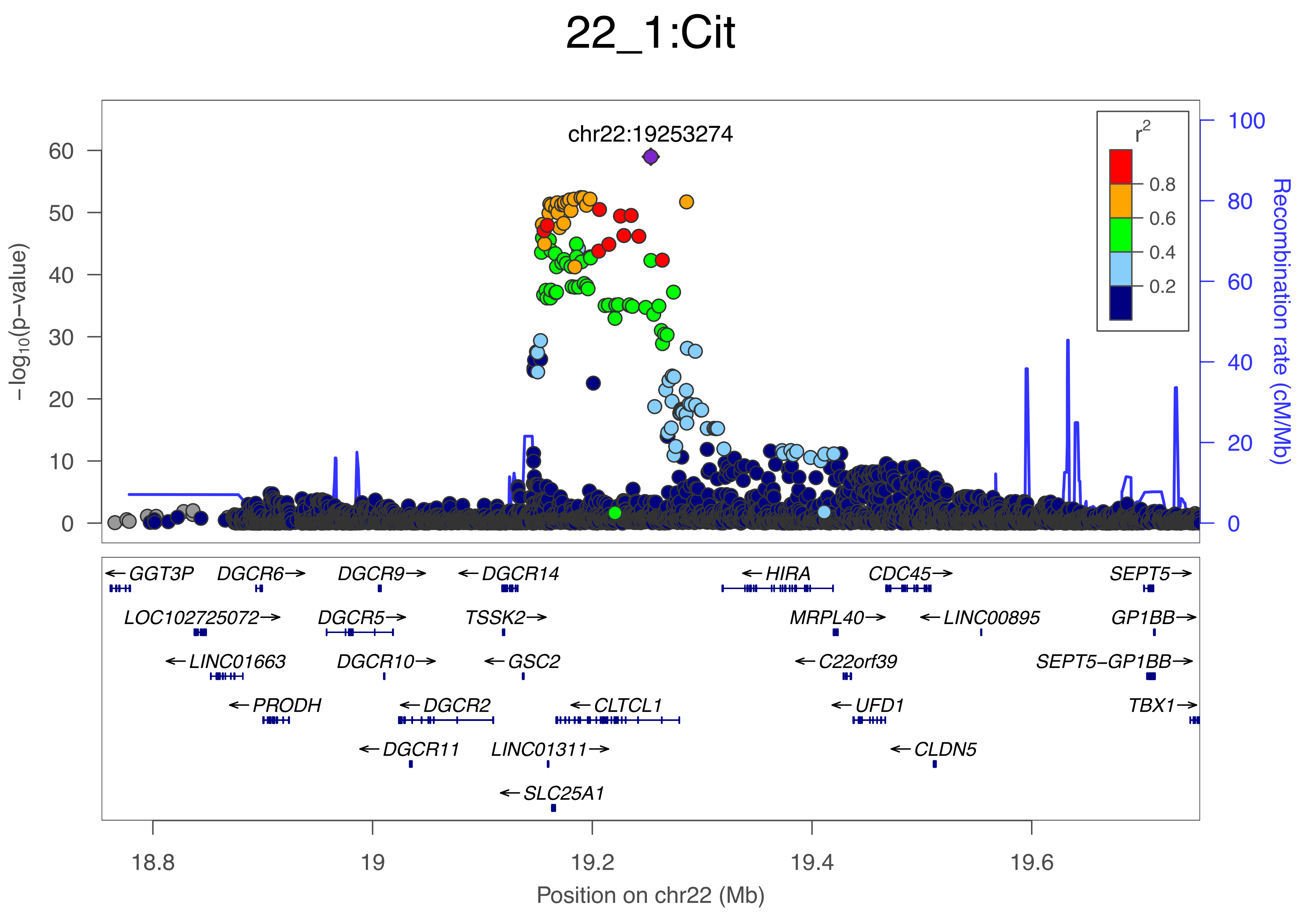
22_1:Cit
chr22:19253274
−log10(p−value)
Recombination rate (cM/Mb)
r2
0.8
0.6
0.4
0.2
GGT3P
DGCR6
DGCR9
DGCR14
HIRA
CDC45
SEPT5
LOC102725072
DGCR5
TSSK2
MRPL40
LINC00895
GP1BB
LINC01663
DGCR10
GSC2
C22orf39
SEPT5−GP1BB
PRODH
DGCR2
CLTCL1
UFD1
TBX1
DGCR11
LINC01311
CLDN5
SLC25A1
18.8
19
19.2
19.4
19.6
Position on chr22 (Mb)

### 22_2:ApoA1

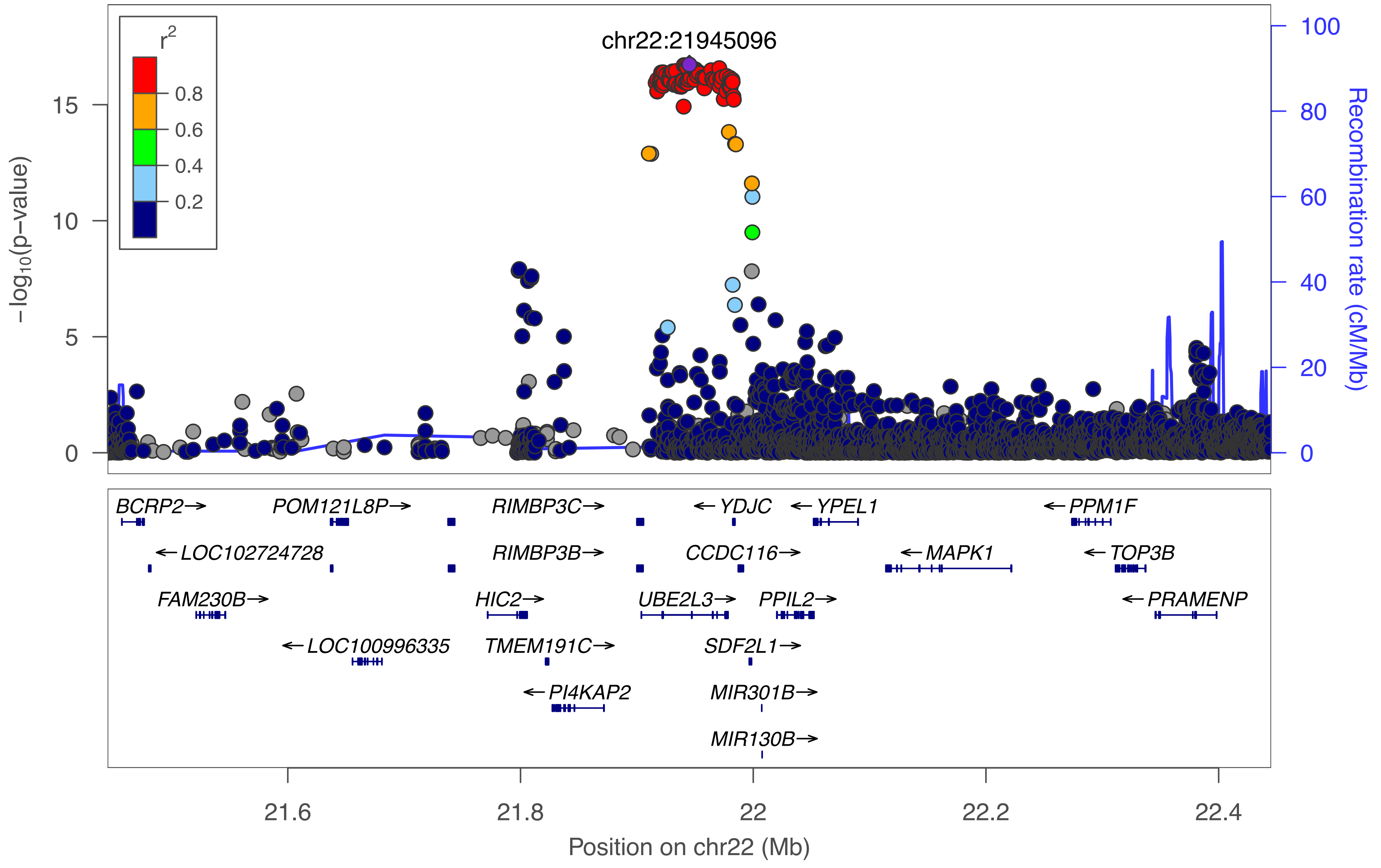

r2
0.8
0.6
0.4
0.2
chr22:21945096
−log10(p−value)
Recombination rate (cM/Mb)
BCRP2→
POM121L8P→
RIMBP3C→
←YDJC
←YPEL1
←PPM1F
←LOC102724728
RIMBP3B→
CCDC116→
←MAPK1
←TOP3B
FAM230B→
HIC2→
UBE2L3→
PPIL2→
←PRAMENP
←LOC100996335
TMEM191C→
SDF2L1→
←PI4KAP2
MIR301B→
MIR130B→
21.6
21.8
22
22.2
22.4
Position on chr22 (Mb)

### 22_3:XL−HDL−FC_percent

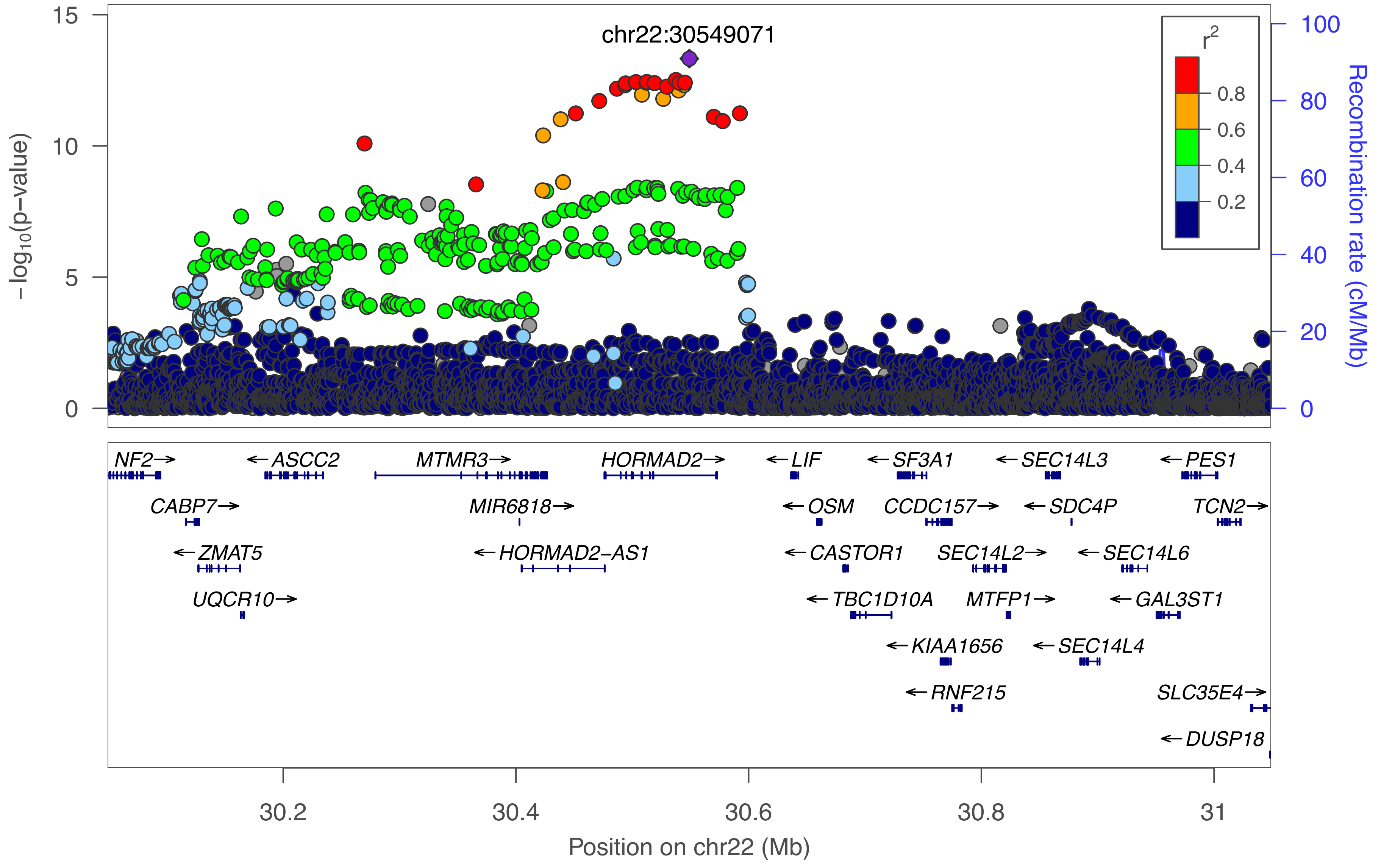

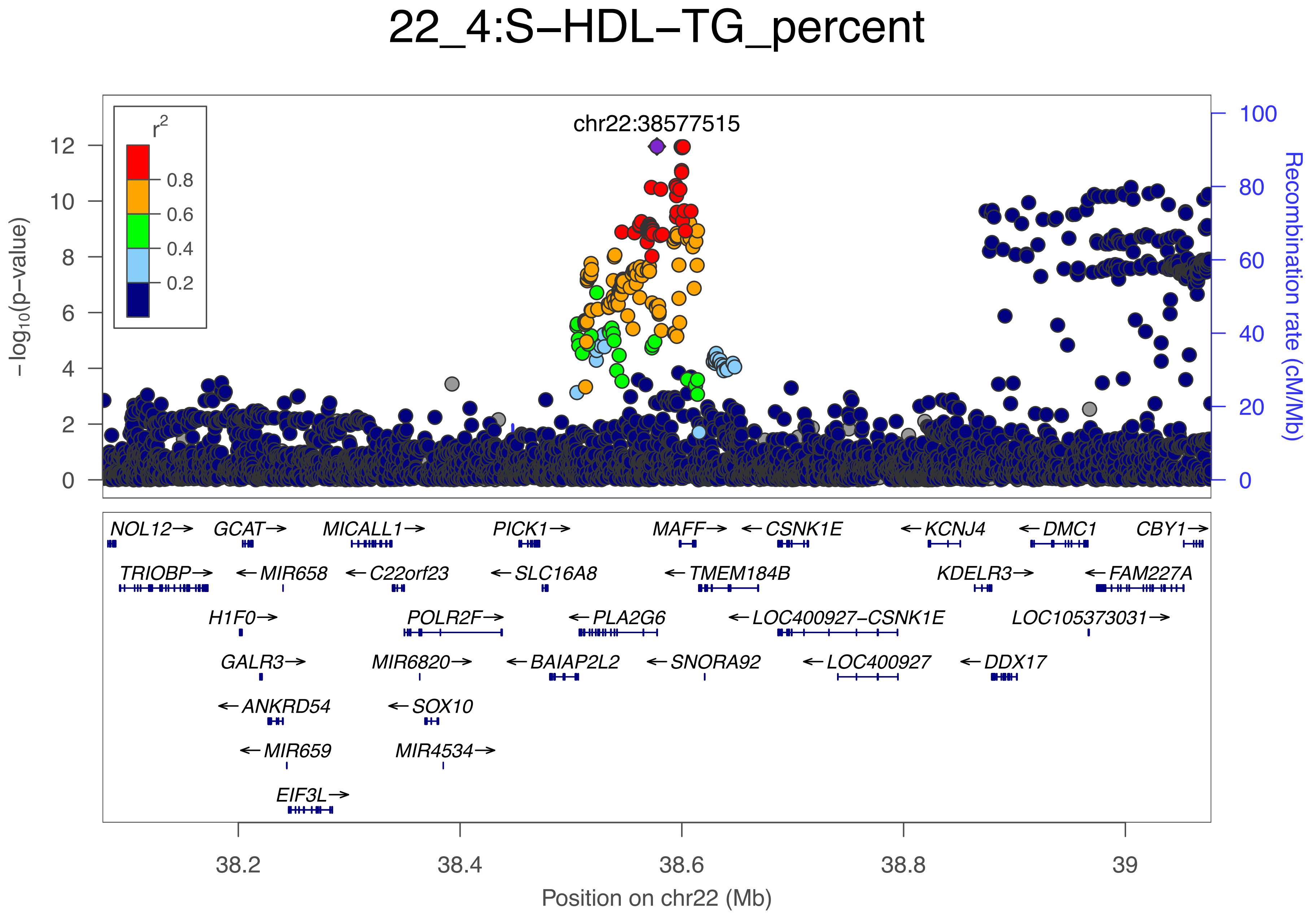

22_4:S−HDL−TG_percent
chr22:38577515
r²
0.8
0.6
0.4
0.2
−log10(p−value)
Recombination rate (cM/Mb)
NOL12→
GCAT→
MICALL1→
PICK1→
MAFF→
←CSNK1E
←KCNJ4
←DMC1
CBY1→
TRIOBP→
←MIR658
←C22orf23
←SLC16A8
←TMEM184B
KDELR3→
←FAM227A
H1F0→
POLR2F→
←PLA2G6
←LOC400927−CSNK1E
LOC105373031→
GALR3→
MIR6820→
←BAIAP2L2
←SNORA92
←LOC400927
←DDX17
←ANKRD54
←SOX10
←MIR659
MIR4534→
EIF3L→
38.2
38.4
38.6
38.8
39
Position on chr22 (Mb)

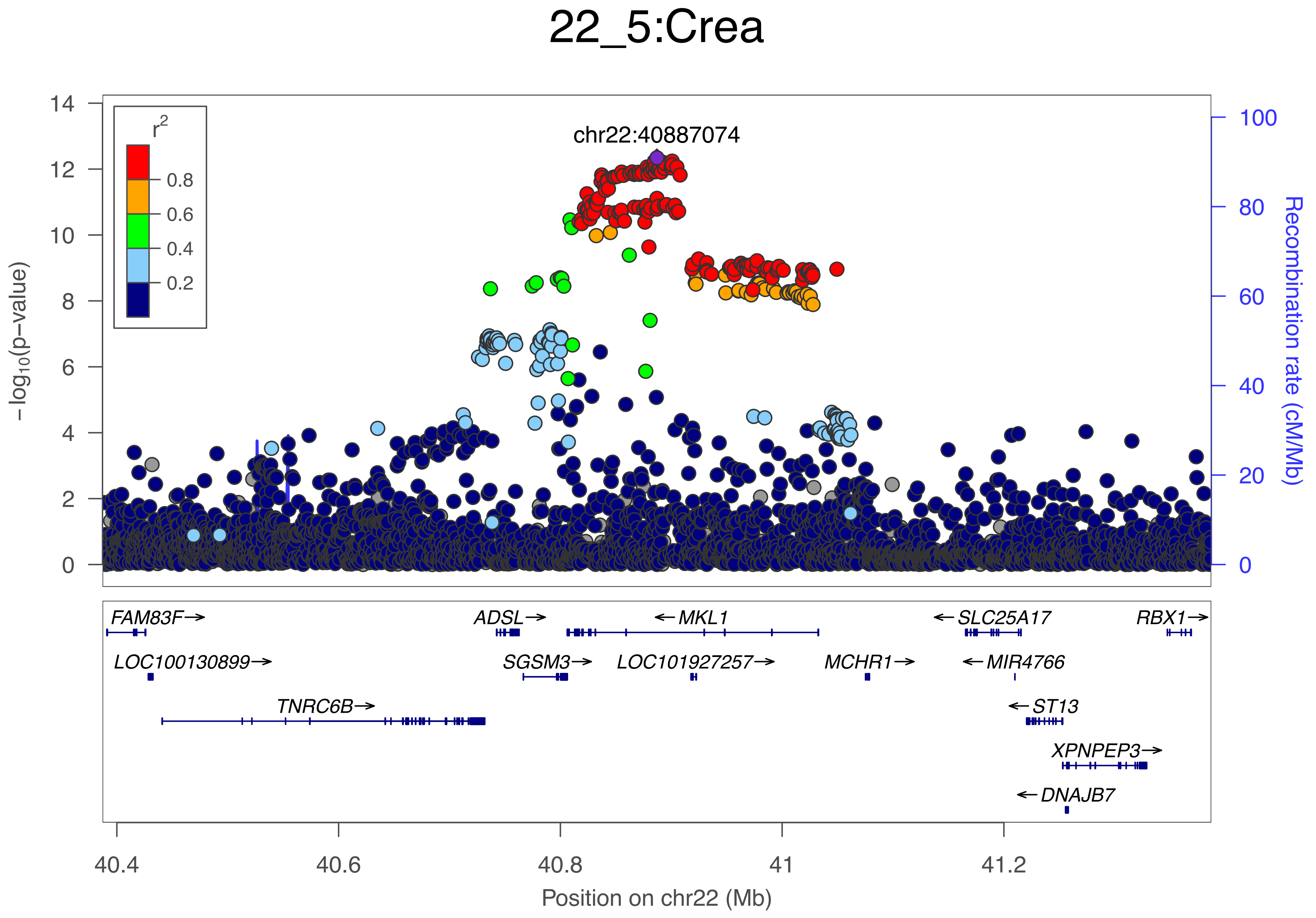

22_5:Crea
chr22:40887074
r2
0.8
0.6
0.4
0.2
−log10(p−value)
Recombination rate (cM/Mb)
FAM83F→
ADSL→
←MKL1
←SLC25A17
RBX1→
LOC100130899→
SGSM3→
LOC101927257→
MCHR1→
←MIR4766
TNRC6B→
←ST13
XPNPEP3→
←DNAJB7
Position on chr22 (Mb)

### 22_6:IDL−CE_percent

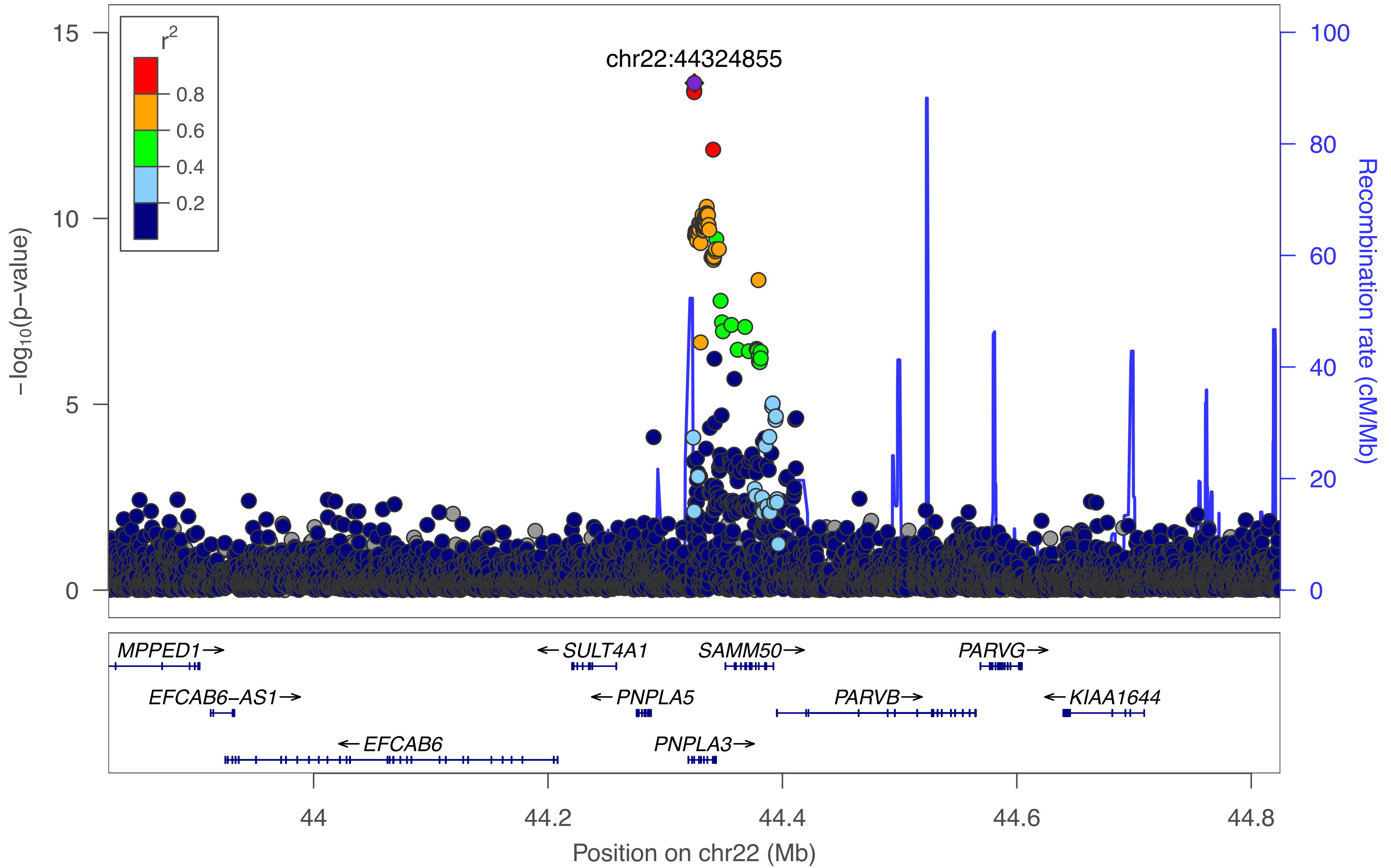
