## Supplementary Figure S3 for "Genome-wide characterization of circulating metabolic biomarkers reveals substantial pleiotropy and novel disease pathways"

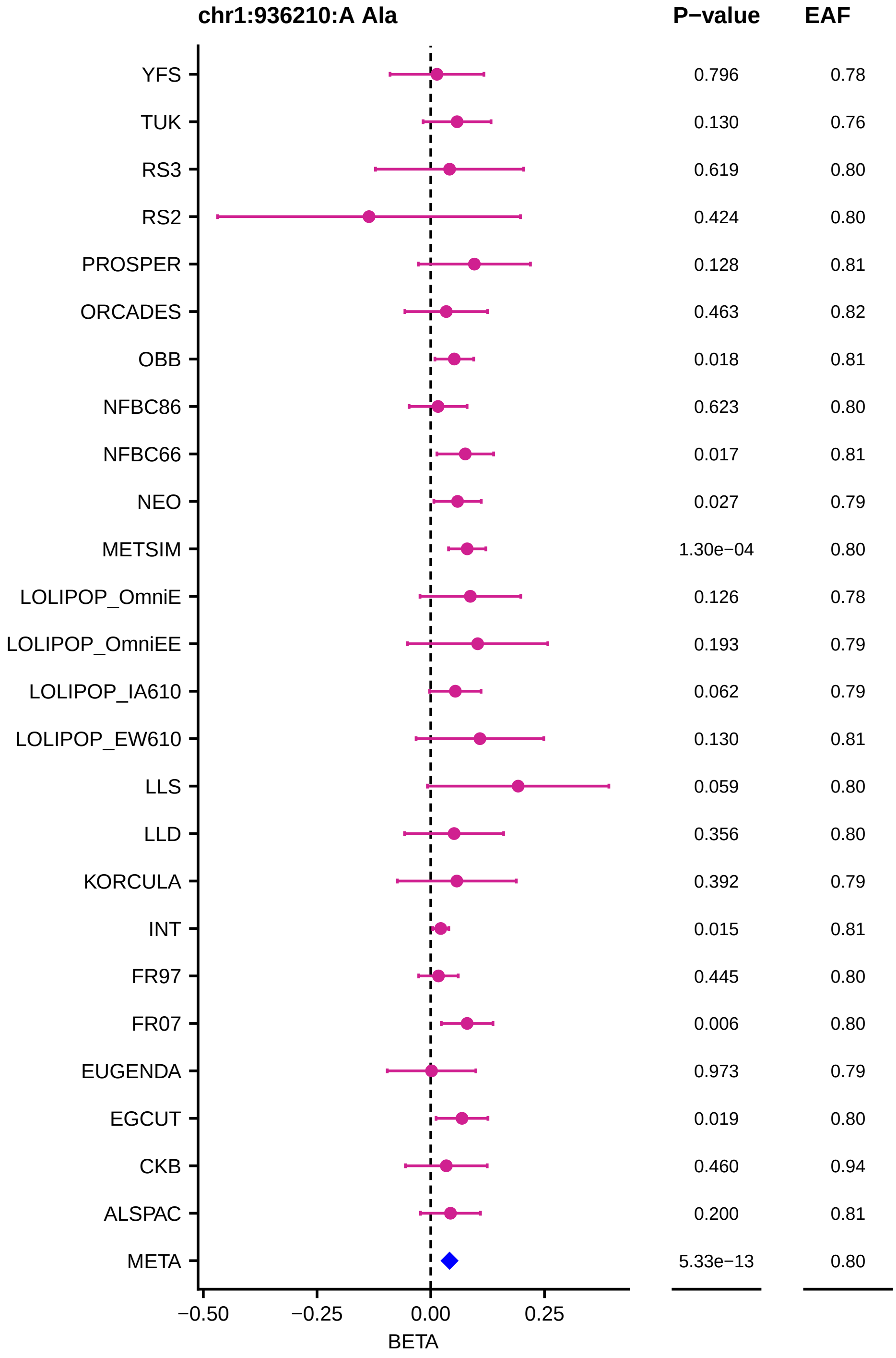
chr1:936210:A Ala
P−value
EAF
YFS 0.796 0.78
TUK 0.130 0.76
RS3 0.619 0.80
RS2 0.424 0.80
PROSPER 0.128 0.81
ORCADES 0.463 0.82
OBB 0.018 0.81
NFBC86 0.623 0.80
NFBC66 0.017 0.81
NEO 0.027 0.79
METSIM 1.30e−04 0.80
LOLIPOP_OmniE 0.126 0.78
LOLIPOP_OmniEE 0.193 0.79
LOLIPOP_IA610 0.062 0.79
LOLIPOP_EW610 0.130 0.81
LLS 0.059 0.80
LLD 0.356 0.80
KORCULA 0.392 0.79
INT 0.015 0.81
FR97 0.445 0.80
FR07 0.006 0.80
EUGENDA 0.973 0.79
EGCUT 0.019 0.80
CKB 0.460 0.94
ALSPAC 0.200 0.81
META 5.33e−13 0.80
−0.50
−0.25
0.00
0.25
BETA

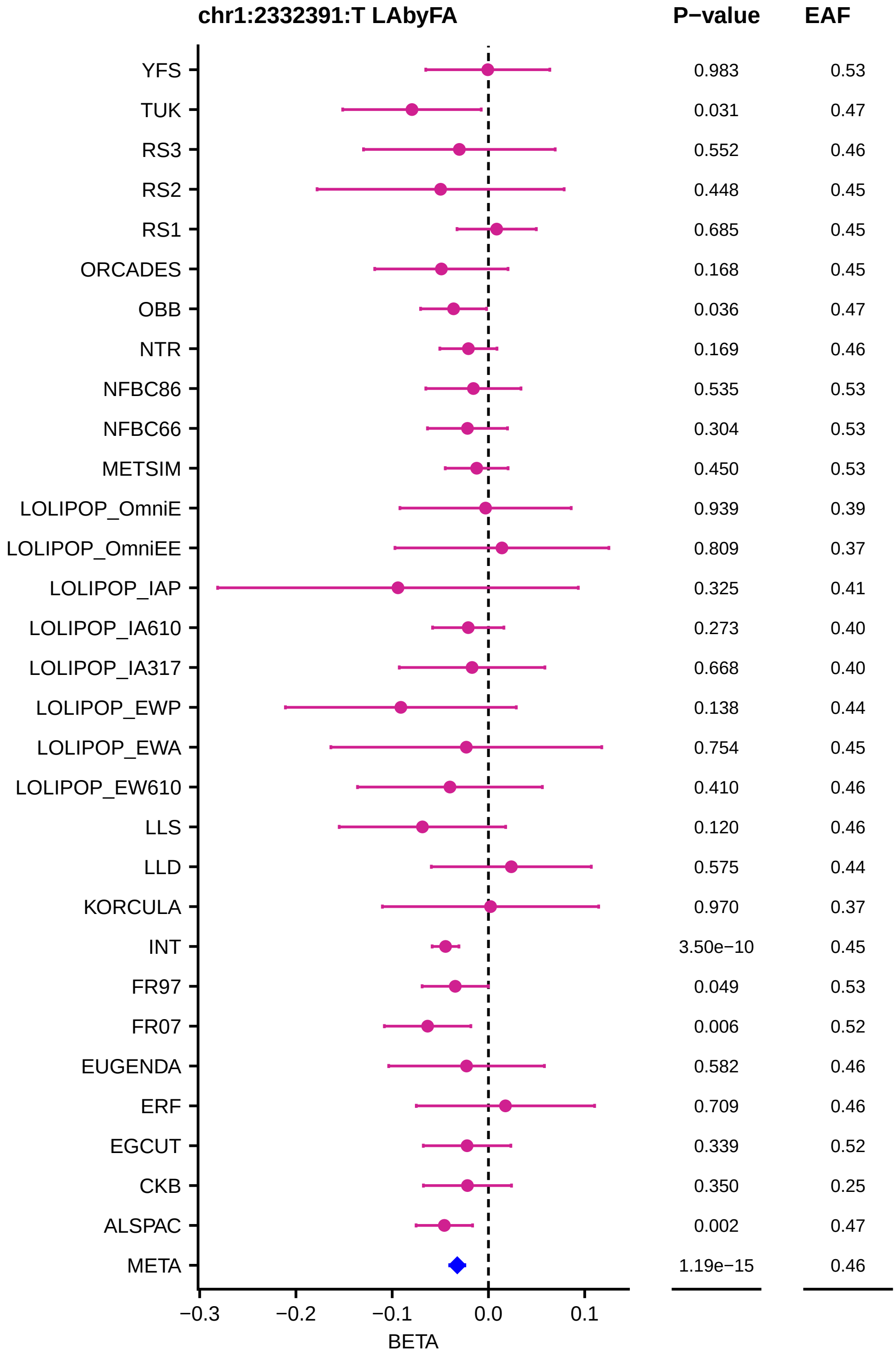
chr1:2332391:T LAbyFA
P−value
EAF
YFS 0.983 0.53
TUK 0.031 0.47
RS3 0.552 0.46
RS2 0.448 0.45
RS1 0.685 0.45
ORCADES 0.168 0.45
OBB 0.036 0.47
NTR 0.169 0.46
NFBC86 0.535 0.53
NFBC66 0.304 0.53
METSIM 0.450 0.53
LOLIPOP_OmniE 0.939 0.39
LOLIPOP_OmniEE 0.809 0.37
LOLIPOP_IAP 0.325 0.41
LOLIPOP_IA610 0.273 0.40
LOLIPOP_IA317 0.668 0.40
LOLIPOP_EWP 0.138 0.44
LOLIPOP_EWA 0.754 0.45
LOLIPOP_EW610 0.410 0.46
LLS 0.120 0.46
LLD 0.575 0.44
KORCULA 0.970 0.37
INT 3.50e−10 0.45
FR97 0.049 0.53
FR07 0.006 0.52
EUGENDA 0.582 0.46
ERF 0.709 0.46
EGCUT 0.339 0.52
CKB 0.350 0.25
ALSPAC 0.002 0.47
META 1.19e−15 0.46
−0.3
−0.2
−0.1
0.0
0.1
BETA

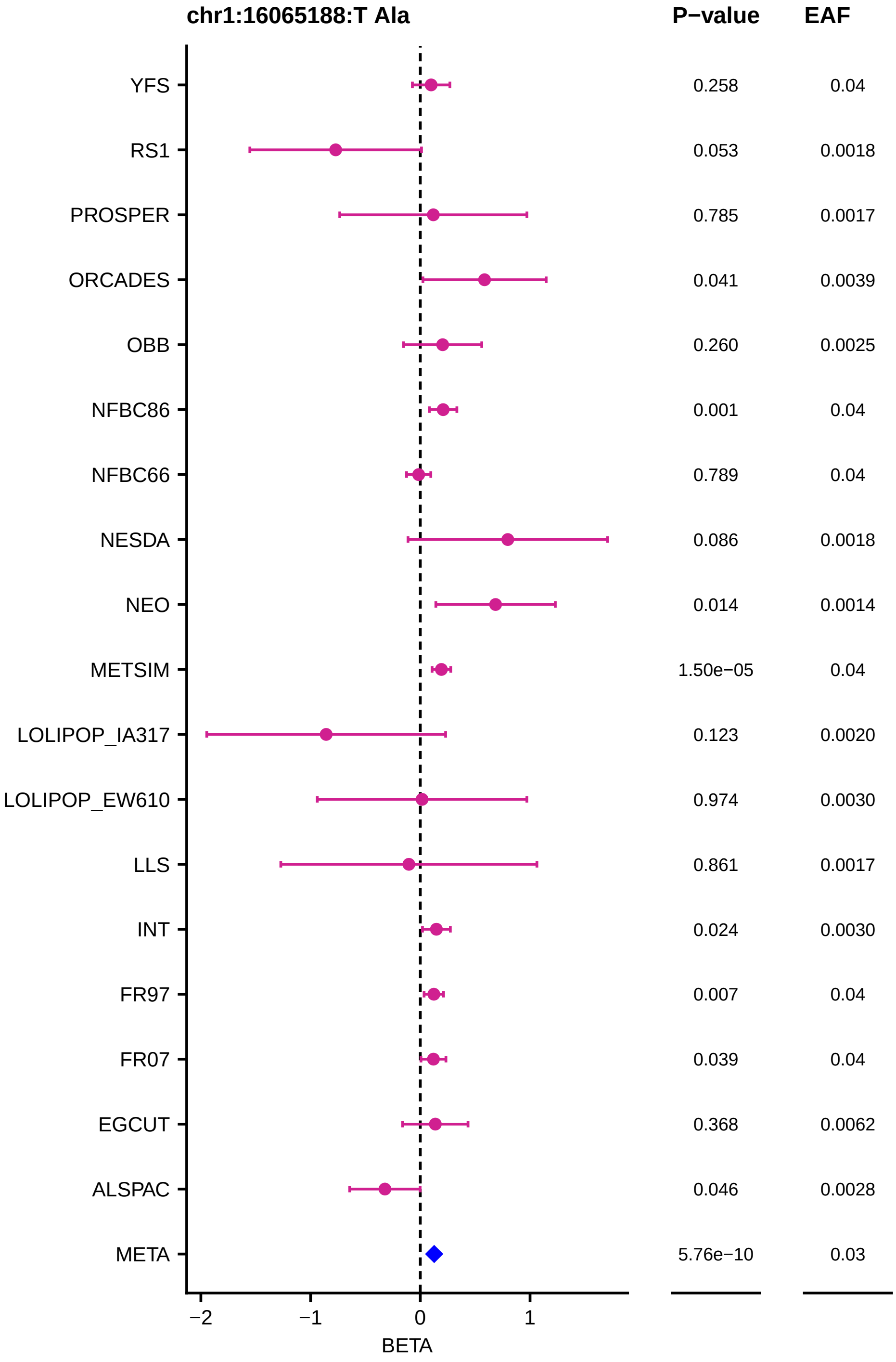

chr1:16065188:T Ala
P−value
EAF
YFS
RS1
PROSPER
ORCADES
OBB
NFBC86
NFBC66
NESDA
NEO
METSIM
LOLIPOP_IA317
LOLIPOP_EW610
LLS
INT
FR97
FR07
EGCUT
ALSPAC
META
0.258
0.053
0.785
0.041
0.260
0.001
0.789
0.086
0.014
1.50e−05
0.123
0.974
0.861
0.024
0.007
0.039
0.368
0.046
5.76e−10
0.04
0.0018
0.0017
0.0039
0.0025
0.04
0.04
0.0018
0.0014
0.04
0.0020
0.0030
0.0017
0.0030
0.04
0.04
0.0062
0.0028
0.03
−2
−1
0
1
BETA

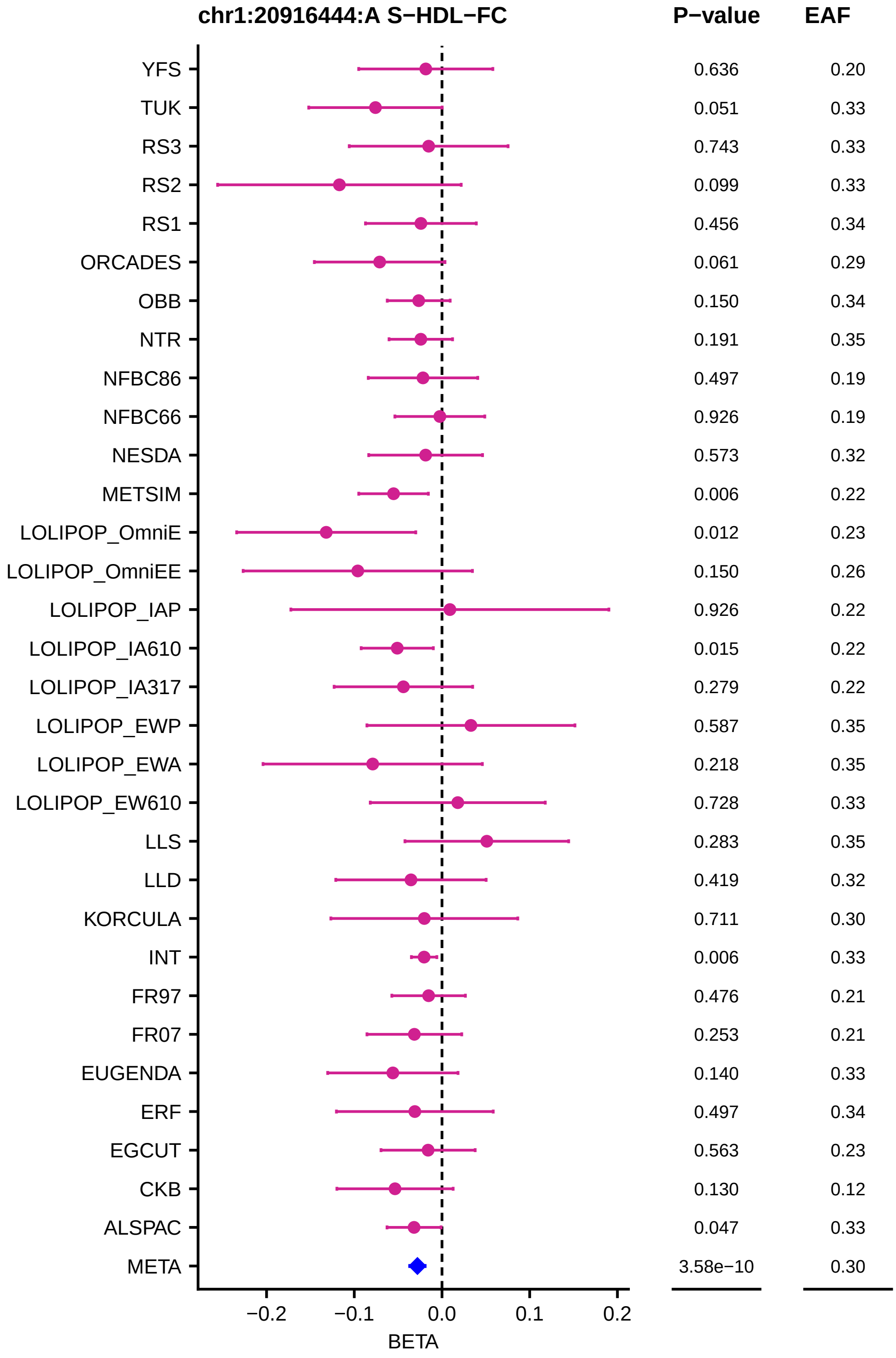
chr1:20916444:A S−HDL−FC
P−value
EAF
YFS 0.636 0.20
TUK 0.051 0.33
RS3 0.743 0.33
RS2 0.099 0.33
RS1 0.456 0.34
ORCADES 0.061 0.29
OBB 0.150 0.34
NTR 0.191 0.35
NFBC86 0.497 0.19
NFBC66 0.926 0.19
NESDA 0.573 0.32
METSIM 0.006 0.22
LOLIPOP_OmniE 0.012 0.23
LOLIPOP_OmniEE 0.150 0.26
LOLIPOP_IAP 0.926 0.22
LOLIPOP_IA610 0.015 0.22
LOLIPOP_IA317 0.279 0.22
LOLIPOP_EWP 0.587 0.35
LOLIPOP_EWA 0.218 0.35
LOLIPOP_EW610 0.728 0.33
LLS 0.283 0.35
LLD 0.419 0.32
KORCULA 0.711 0.30
INT 0.006 0.33
FR97 0.476 0.21
FR07 0.253 0.21
EUGENDA 0.140 0.33
ERF 0.497 0.34
EGCUT 0.563 0.23
CKB 0.130 0.12
ALSPAC 0.047 0.33
META 3.58e−10 0.30
−0.2
−0.1
0.0
0.1
0.2
BETA

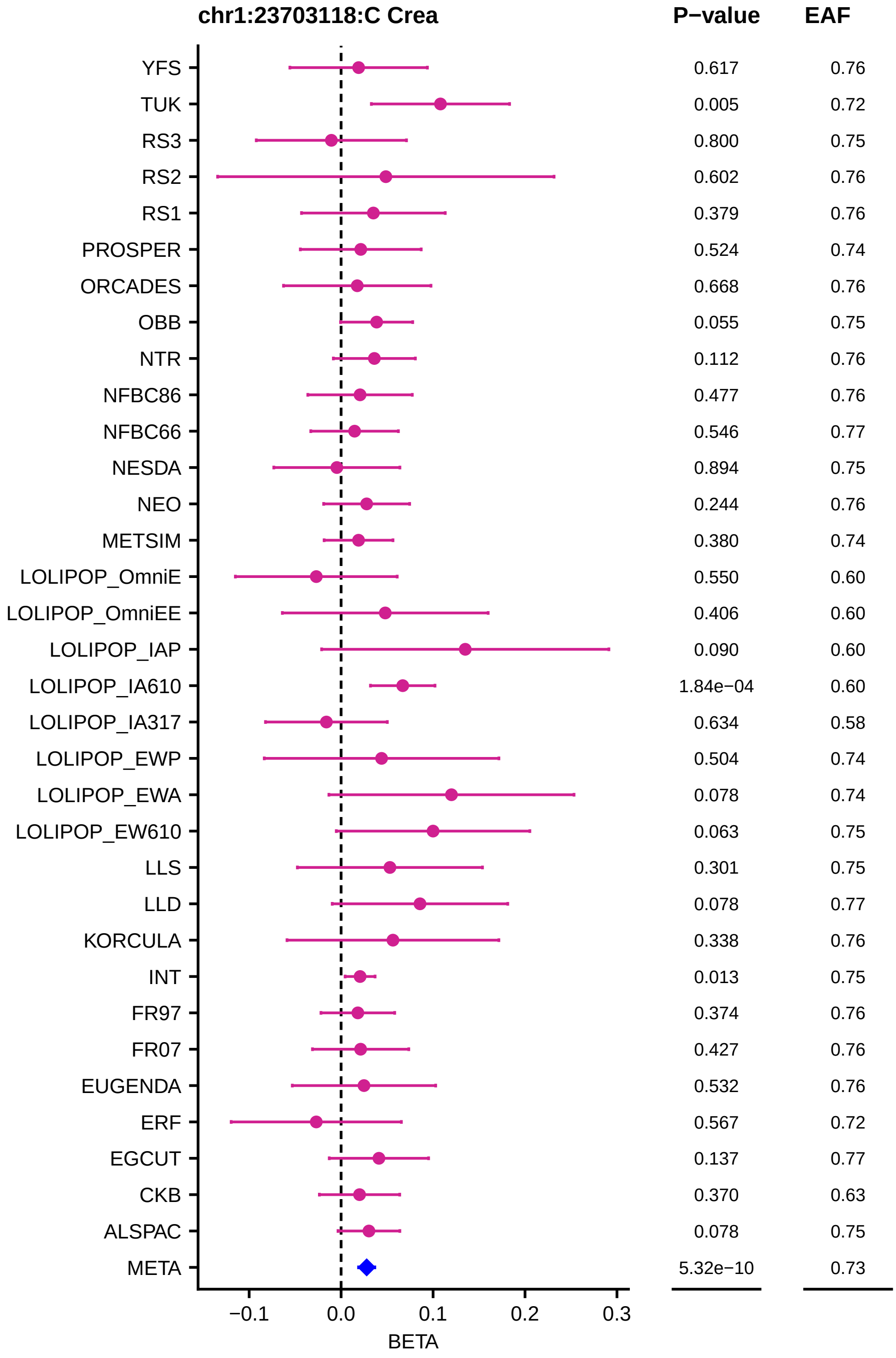
chr1:23703118:C Crea
P−value
EAF
YFS
TUK
RS3
RS2
RS1
PROSPER
ORCADES
OBB
NTR
NFBC86
NFBC66
NESDA
NEO
METSIM
LOLIPOP_OmniE
LOLIPOP_OmniEE
LOLIPOP_IAP
LOLIPOP_IA610
LOLIPOP_IA317
LOLIPOP_EWP
LOLIPOP_EWA
LOLIPOP_EW610
LLS
LLD
KORCULA
INT
FR97
FR07
EUGENDA
ERF
EGCUT
CKB
ALSPAC
META
0.617
0.005
0.800
0.602
0.379
0.524
0.668
0.055
0.112
0.477
0.546
0.894
0.244
0.380
0.550
0.406
0.090
1.84e−04
0.634
0.504
0.078
0.063
0.301
0.078
0.338
0.013
0.374
0.427
0.532
0.567
0.137
0.370
0.078
5.32e−10
0.76
0.72
0.75
0.76
0.76
0.74
0.76
0.75
0.76
0.76
0.77
0.75
0.76
0.74
0.60
0.60
0.60
0.60
0.58
0.74
0.74
0.75
0.75
0.77
0.76
0.75
0.76
0.76
0.76
0.72
0.77
0.63
0.75
0.73
−0.1
0.0
0.1
0.2
0.3
BETA

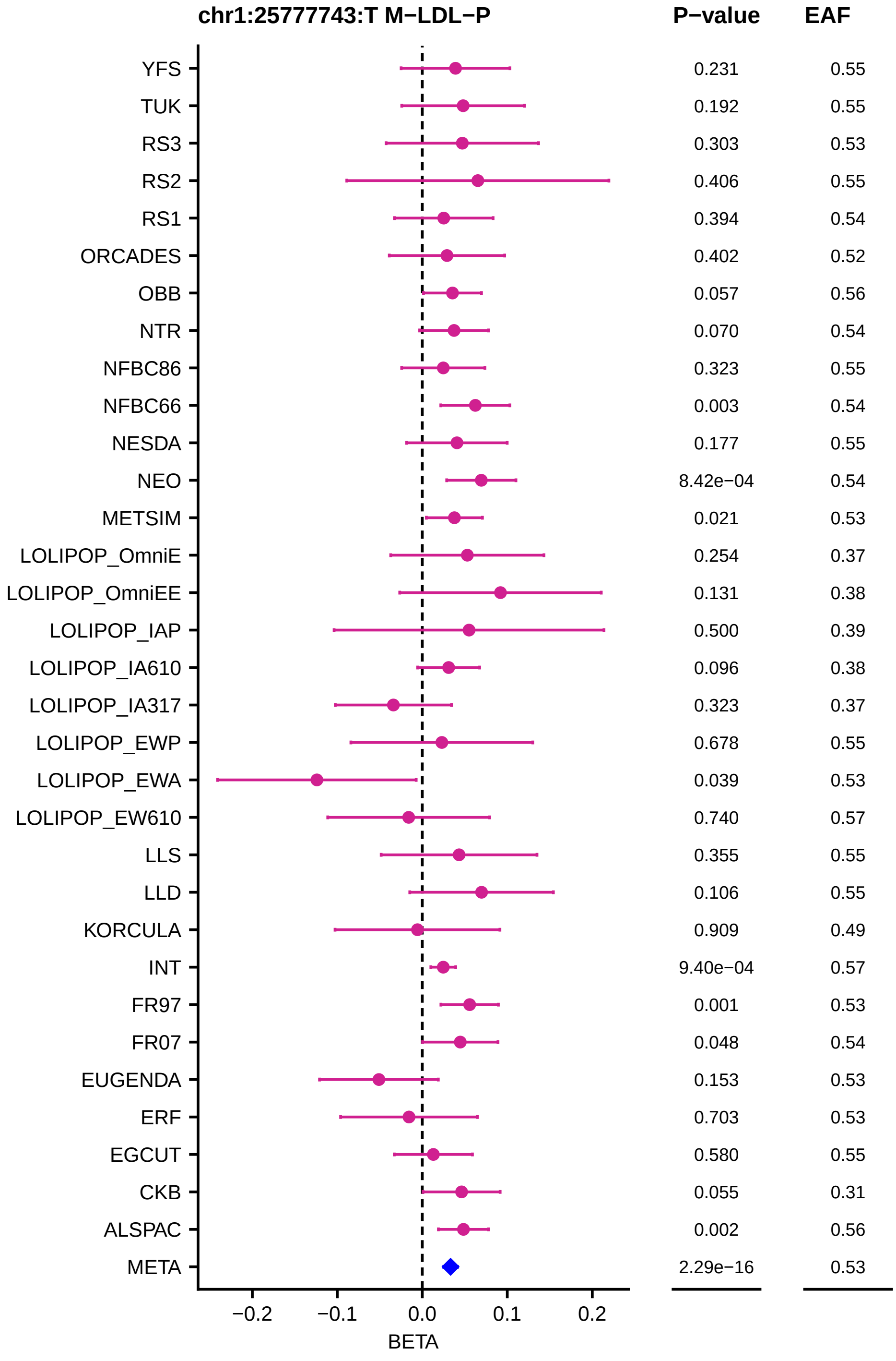
chr1:25777743:T M−LDL−P
P−value
EAF
YFS 0.231 0.55
TUK 0.192 0.55
RS3 0.303 0.53
RS2 0.406 0.55
RS1 0.394 0.54
ORCADES 0.402 0.52
OBB 0.057 0.56
NTR 0.070 0.54
NFBC86 0.323 0.55
NFBC66 0.003 0.54
NESDA 0.177 0.55
NEO 8.42e−04 0.54
METSIM 0.021 0.53
LOLIPOP_OmniE 0.254 0.37
LOLIPOP_OmniEE 0.131 0.38
LOLIPOP_IAP 0.500 0.39
LOLIPOP_IA610 0.096 0.38
LOLIPOP_IA317 0.323 0.37
LOLIPOP_EWP 0.678 0.55
LOLIPOP_EWA 0.039 0.53
LOLIPOP_EW610 0.740 0.57
LLS 0.355 0.55
LLD 0.106 0.55
KORCULA 0.909 0.49
INT 9.40e−04 0.57
FR97 0.001 0.53
FR07 0.048 0.54
EUGENDA 0.153 0.53
ERF 0.703 0.53
EGCUT 0.580 0.55
CKB 0.055 0.31
ALSPAC 0.002 0.56
META 2.29e−16 0.53
−0.2
−0.1
0.0
0.1
0.2
BETA

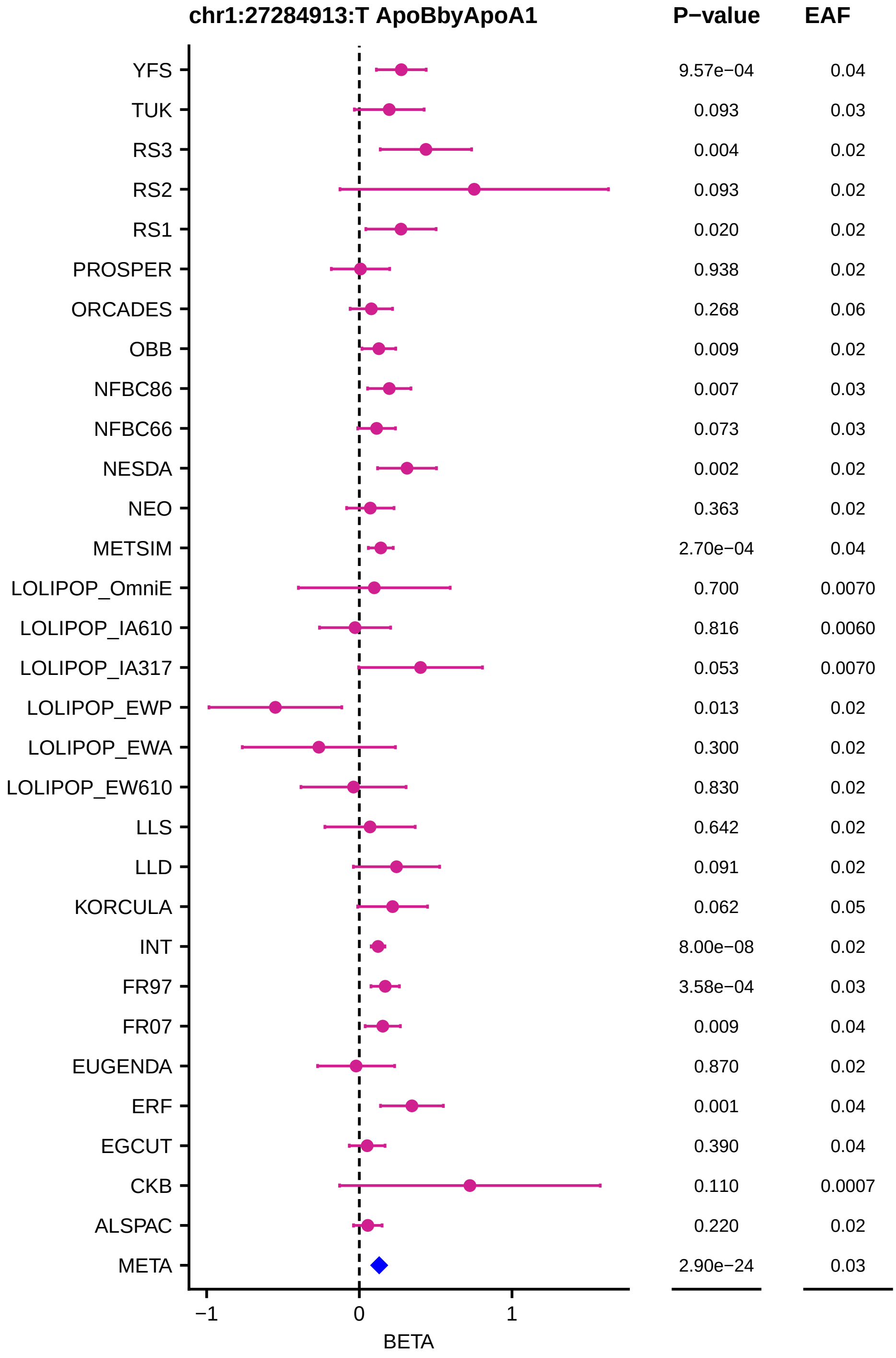
chr1:27284913:T ApoBbyApoA1
P−value
EAF
YFS
TUK
RS3
RS2
RS1
PROSPER
ORCADES
OBB
NFBC86
NFBC66
NESDA
NEO
METSIM
LOLIPOP_OmniE
LOLIPOP_IA610
LOLIPOP_IA317
LOLIPOP_EWP
LOLIPOP_EWA
LOLIPOP_EW610
LLS
LLD
KORCULA
INT
FR97
FR07
EUGENDA
ERF
EGCUT
CKB
ALSPAC
META
9.57e−04
0.093
0.004
0.093
0.020
0.938
0.268
0.009
0.007
0.073
0.002
0.363
2.70e−04
0.700
0.816
0.053
0.013
0.300
0.830
0.642
0.091
0.062
8.00e−08
3.58e−04
0.009
0.870
0.001
0.390
0.110
0.220
2.90e−24
0.04
0.03
0.02
0.02
0.02
0.02
0.06
0.02
0.03
0.03
0.02
0.02
0.04
0.0070
0.0060
0.0070
0.02
0.02
0.02
0.02
0.02
0.05
0.02
0.03
0.04
0.02
0.04
0.04
0.0007
0.02
0.03
−1
0
1
BETA

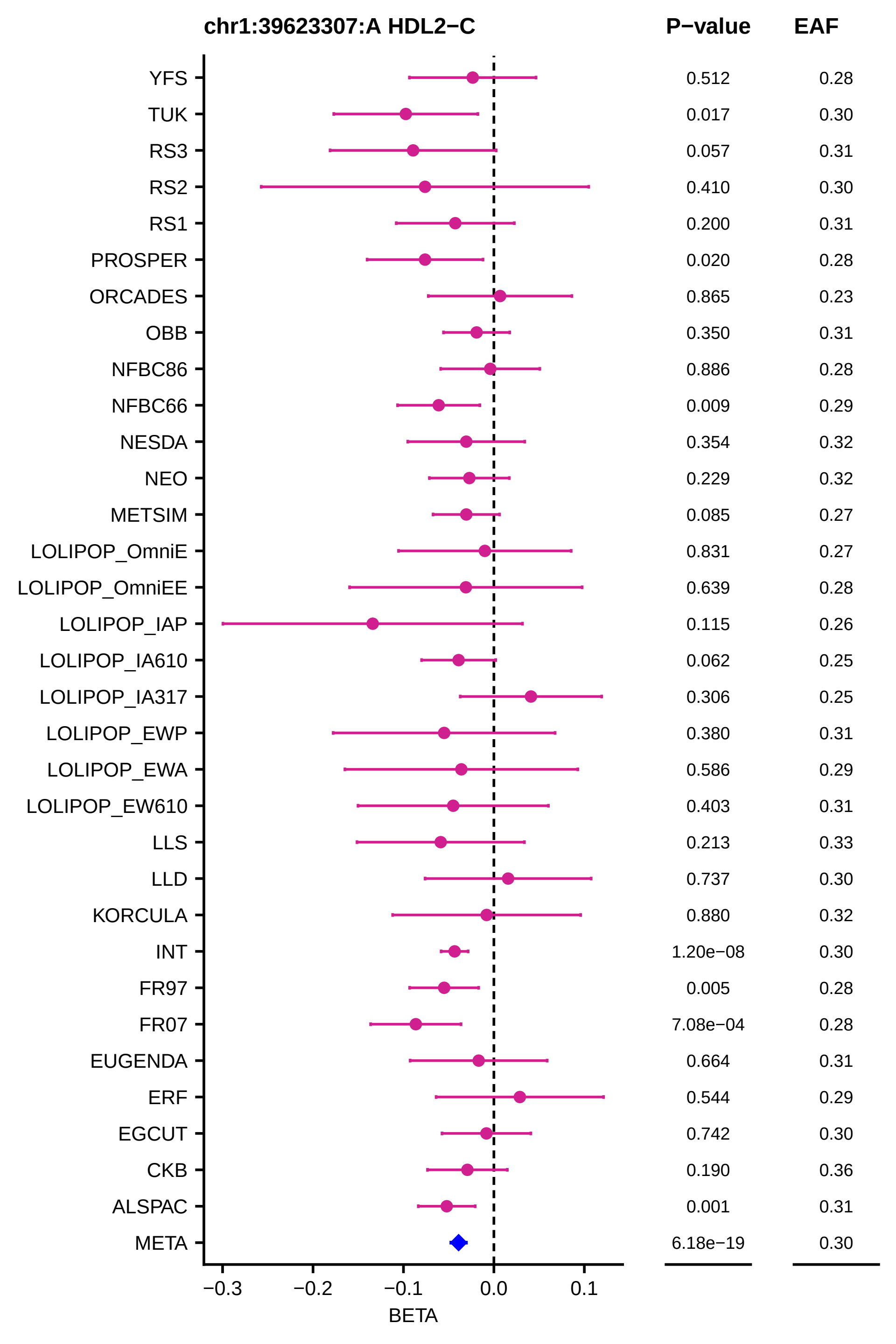
chr1:39623307:A HDL2−C
P−value
EAF
YFS 0.512 0.28
TUK 0.017 0.30
RS3 0.057 0.31
RS2 0.410 0.30
RS1 0.200 0.31
PROSPER 0.020 0.28
ORCADES 0.865 0.23
OBB 0.350 0.31
NFBC86 0.886 0.28
NFBC66 0.009 0.29
NESDA 0.354 0.32
NEO 0.229 0.32
METSIM 0.085 0.27
LOLIPOP_OmniE 0.831 0.27
LOLIPOP_OmniEE 0.639 0.28
LOLIPOP_IAP 0.115 0.26
LOLIPOP_IA610 0.062 0.25
LOLIPOP_IA317 0.306 0.25
LOLIPOP_EWP 0.380 0.31
LOLIPOP_EWA 0.586 0.29
LOLIPOP_EW610 0.403 0.31
LLS 0.213 0.33
LLD 0.737 0.30
KORCULA 0.880 0.32
INT 1.20e−08 0.30
FR97 0.005 0.28
FR07 7.08e−04 0.28
EUGENDA 0.664 0.31
ERF 0.544 0.29
EGCUT 0.742 0.30
CKB 0.190 0.36
ALSPAC 0.001 0.31
META 6.18e−19 0.30
−0.3
−0.2
−0.1
0.0
0.1
BETA

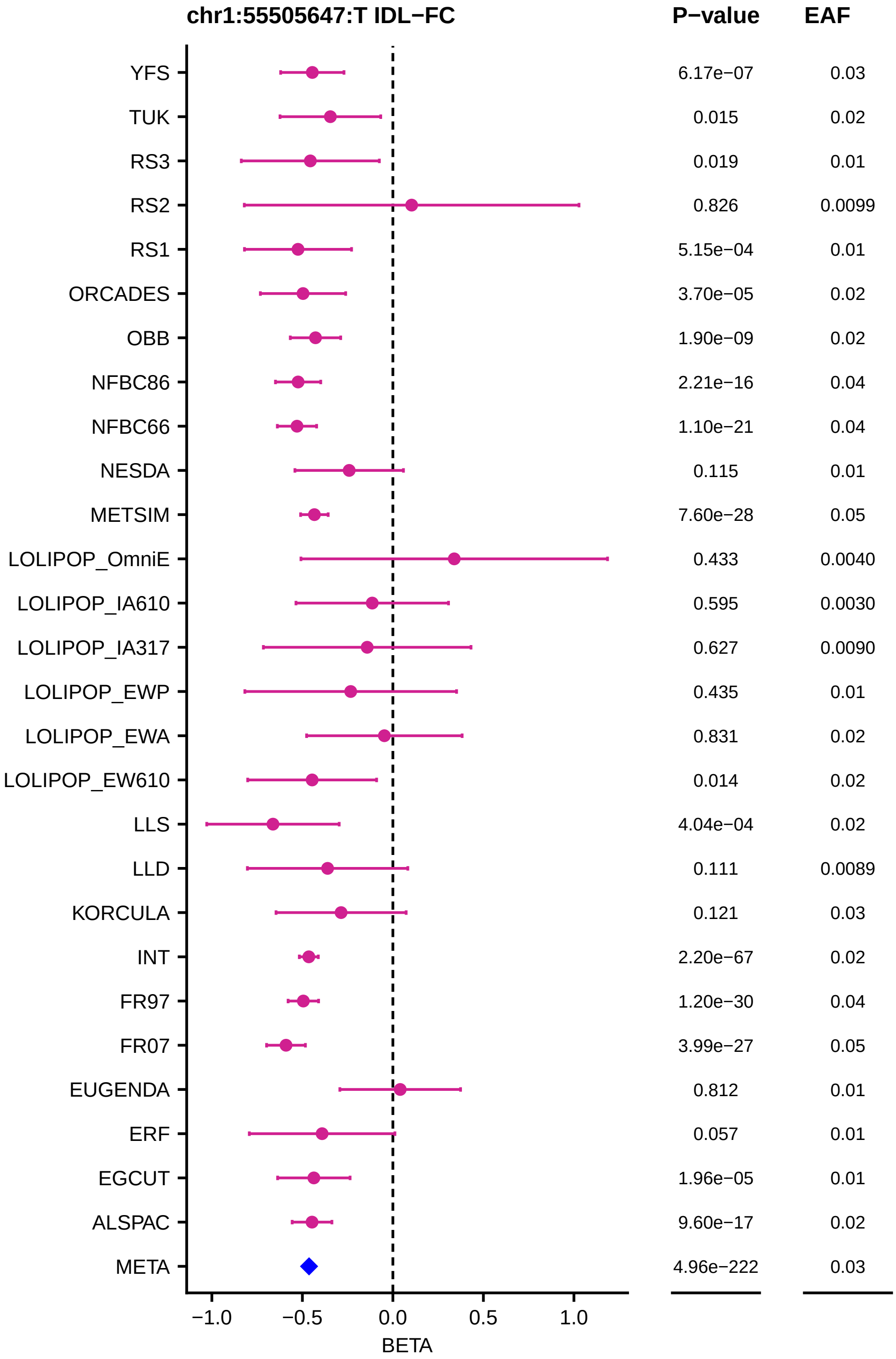

chr1:55505647:T IDL−FC
P−value
EAF
YFS 6.17e−07 0.03
TUK 0.015 0.02
RS3 0.019 0.01
RS2 0.826 0.0099
RS1 5.15e−04 0.01
ORCADES 3.70e−05 0.02
OBB 1.90e−09 0.02
NFBC86 2.21e−16 0.04
NFBC66 1.10e−21 0.04
NESDA 0.115 0.01
METSIM 7.60e−28 0.05
LOLIPOP_OmniE 0.433 0.0040
LOLIPOP_IA610 0.595 0.0030
LOLIPOP_IA317 0.627 0.0090
LOLIPOP_EWP 0.435 0.01
LOLIPOP_EWA 0.831 0.02
LOLIPOP_EW610 0.014 0.02
LLS 4.04e−04 0.02
LLD 0.111 0.0089
KORCULA 0.121 0.03
INT 2.20e−67 0.02
FR97 1.20e−30 0.04
FR07 3.99e−27 0.05
EUGENDA 0.812 0.01
ERF 0.057 0.01
EGCUT 1.96e−05 0.01
ALSPAC 9.60e−17 0.02
META 4.96e−222 0.03
−1.0
−0.5
0.0
0.5
1.0
BETA

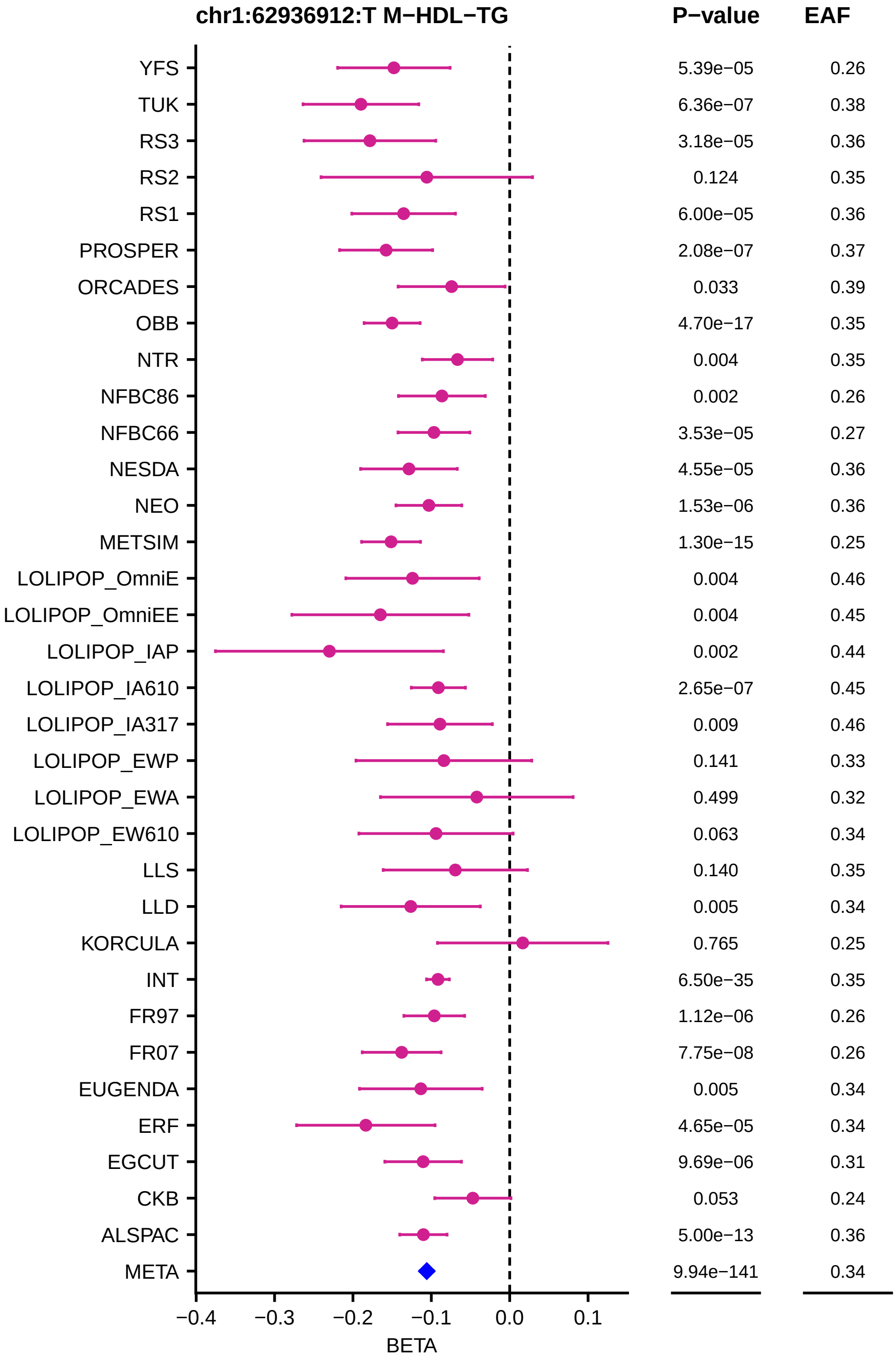
chr1:62936912:T M−HDL−TG
P−value
EAF
YFS 5.39e−05 0.26
TUK 6.36e−07 0.38
RS3 3.18e−05 0.36
RS2 0.124 0.35
RS1 6.00e−05 0.36
PROSPER 2.08e−07 0.37
ORCADES 0.033 0.39
OBB 4.70e−17 0.35
NTR 0.004 0.35
NFBC86 0.002 0.26
NFBC66 3.53e−05 0.27
NESDA 4.55e−05 0.36
NEO 1.53e−06 0.36
METSIM 1.30e−15 0.25
LOLIPOP_OmniE 0.004 0.46
LOLIPOP_OmniEE 0.004 0.45
LOLIPOP_IAP 0.002 0.44
LOLIPOP_IA610 2.65e−07 0.45
LOLIPOP_IA317 0.009 0.46
LOLIPOP_EWP 0.141 0.33
LOLIPOP_EWA 0.499 0.32
LOLIPOP_EW610 0.063 0.34
LLS 0.140 0.35
LLD 0.005 0.34
KORCULA 0.765 0.25
INT 6.50e−35 0.35
FR97 1.12e−06 0.26
FR07 7.75e−08 0.26
EUGENDA 0.005 0.34
ERF 4.65e−05 0.34
EGCUT 9.69e−06 0.31
CKB 0.053 0.24
ALSPAC 5.00e−13 0.36
META 9.94e−141 0.34
−0.4 −0.3 −0.2 −0.1 0.0 0.1
BETA

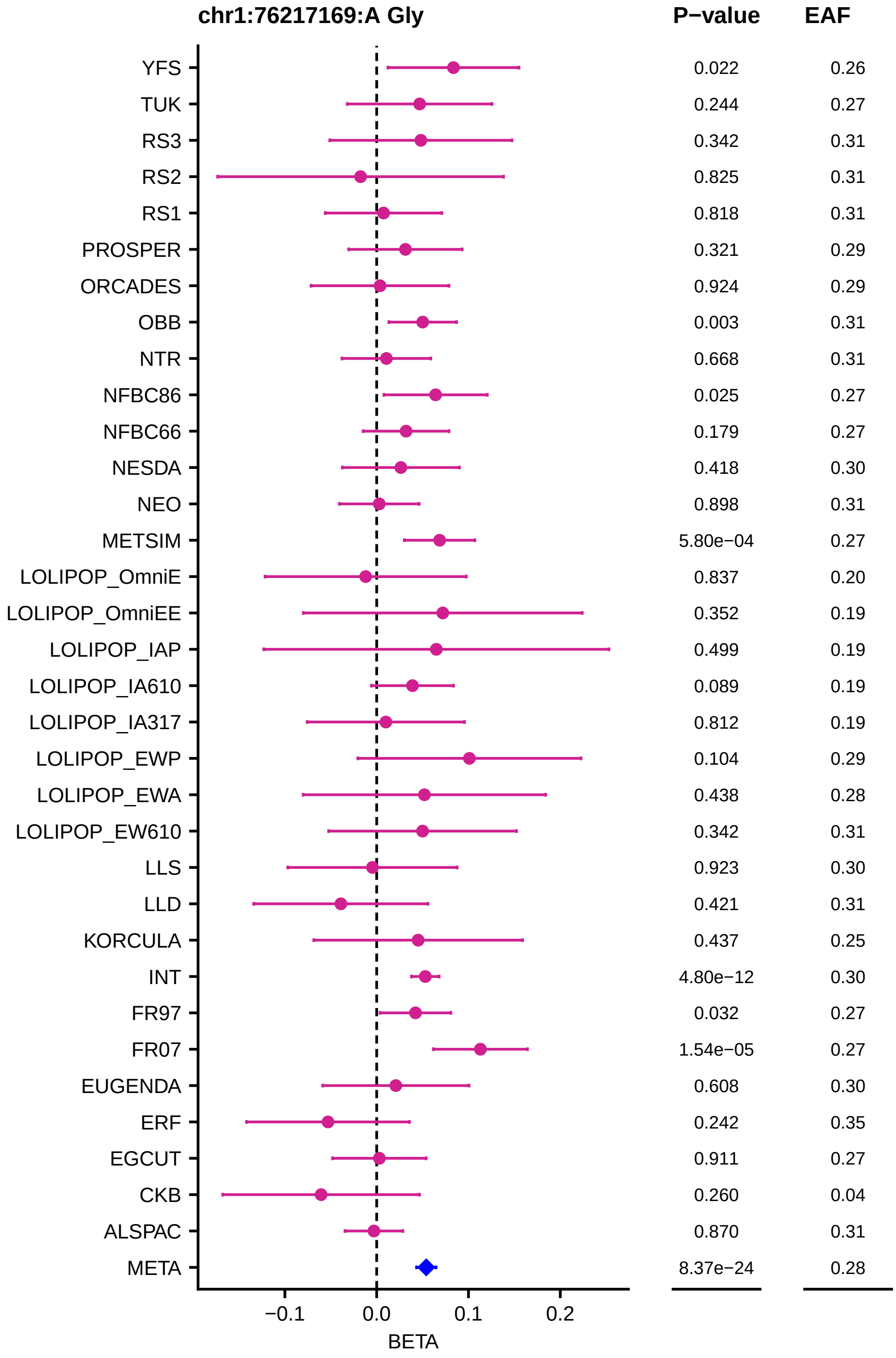

chr1:76217169:A Gly
P−value
EAF
YFS 0.022 0.26
TUK 0.244 0.27
RS3 0.342 0.31
RS2 0.825 0.31
RS1 0.818 0.31
PROSPER 0.321 0.29
ORCADES 0.924 0.29
OBB 0.003 0.31
NTR 0.668 0.31
NFBC86 0.025 0.27
NFBC66 0.179 0.27
NESDA 0.418 0.30
NEO 0.898 0.31
METSIM 5.80e−04 0.27
LOLIPOP_OmniE 0.837 0.20
LOLIPOP_OmniEE 0.352 0.19
LOLIPOP_IAP 0.499 0.19
LOLIPOP_IA610 0.089 0.19
LOLIPOP_IA317 0.812 0.19
LOLIPOP_EWP 0.104 0.29
LOLIPOP_EWA 0.438 0.28
LOLIPOP_EW610 0.342 0.31
LLS 0.923 0.30
LLD 0.421 0.31
KORCULA 0.437 0.25
INT 4.80e−12 0.30
FR97 0.032 0.27
FR07 1.54e−05 0.27
EUGENDA 0.608 0.30
ERF 0.242 0.35
EGCUT 0.911 0.27
CKB 0.260 0.04
ALSPAC 0.870 0.31
META 8.37e−24 0.28
−0.1
0.0
0.1
0.2
BETA

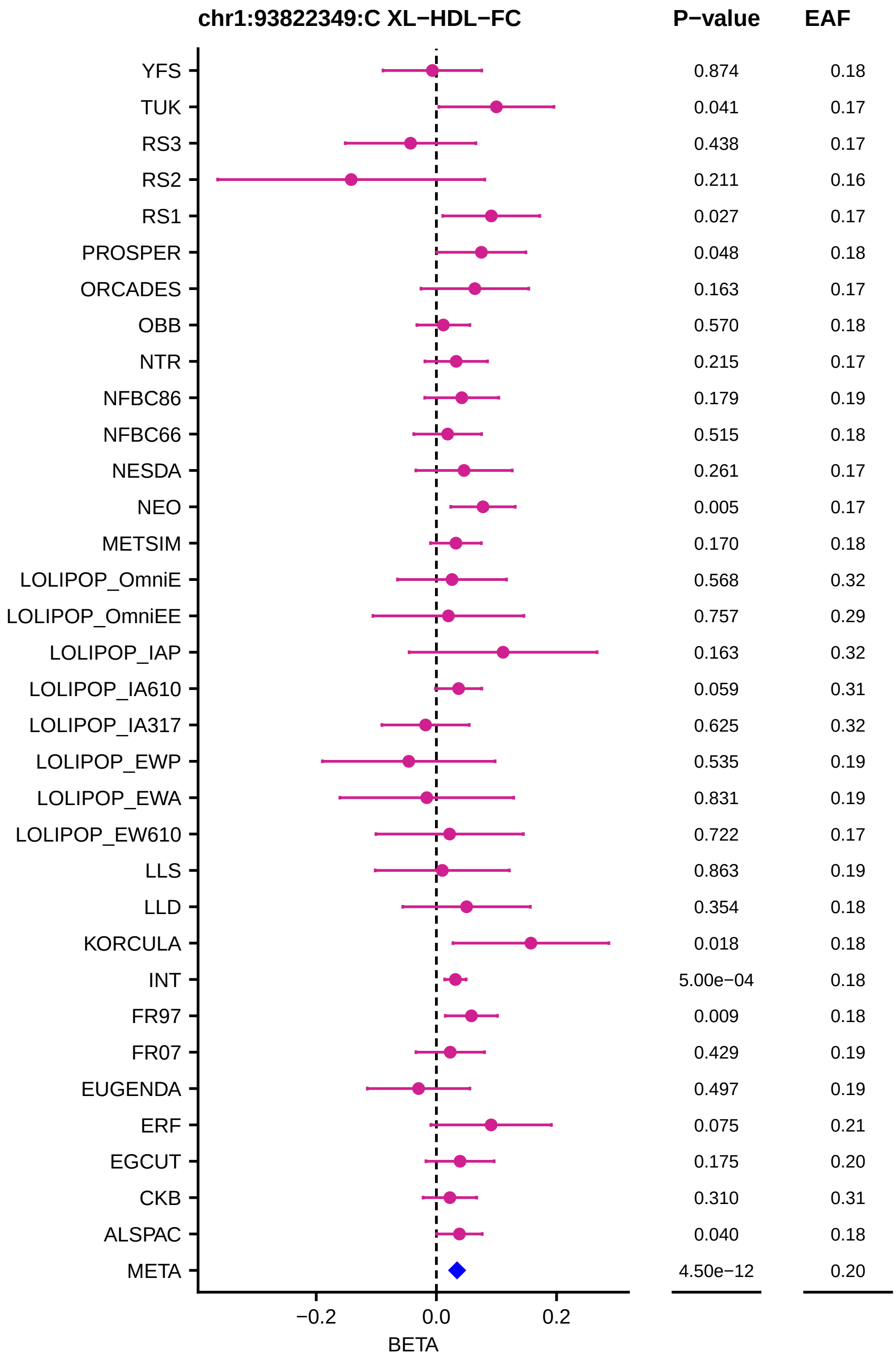
chr1:93822349:C XL−HDL−FC
P−value
EAF
YFS 0.874 0.18
TUK 0.041 0.17
RS3 0.438 0.17
RS2 0.211 0.16
RS1 0.027 0.17
PROSPER 0.048 0.18
ORCADES 0.163 0.17
OBB 0.570 0.18
NTR 0.215 0.17
NFBC86 0.179 0.19
NFBC66 0.515 0.18
NESDA 0.261 0.17
NEO 0.005 0.17
METSIM 0.170 0.18
LOLIPOP_OmniE 0.568 0.32
LOLIPOP_OmniEE 0.757 0.29
LOLIPOP_IAP 0.163 0.32
LOLIPOP_IA610 0.059 0.31
LOLIPOP_IA317 0.625 0.32
LOLIPOP_EWP 0.535 0.19
LOLIPOP_EWA 0.831 0.19
LOLIPOP_EW610 0.722 0.17
LLS 0.863 0.19
LLD 0.354 0.18
KORCULA 0.018 0.18
INT 5.00e−04 0.18
FR97 0.009 0.18
FR07 0.429 0.19
EUGENDA 0.497 0.19
ERF 0.075 0.21
EGCUT 0.175 0.20
CKB 0.310 0.31
ALSPAC 0.040 0.18
META 4.50e−12 0.20
−0.2
0.0
0.2
BETA

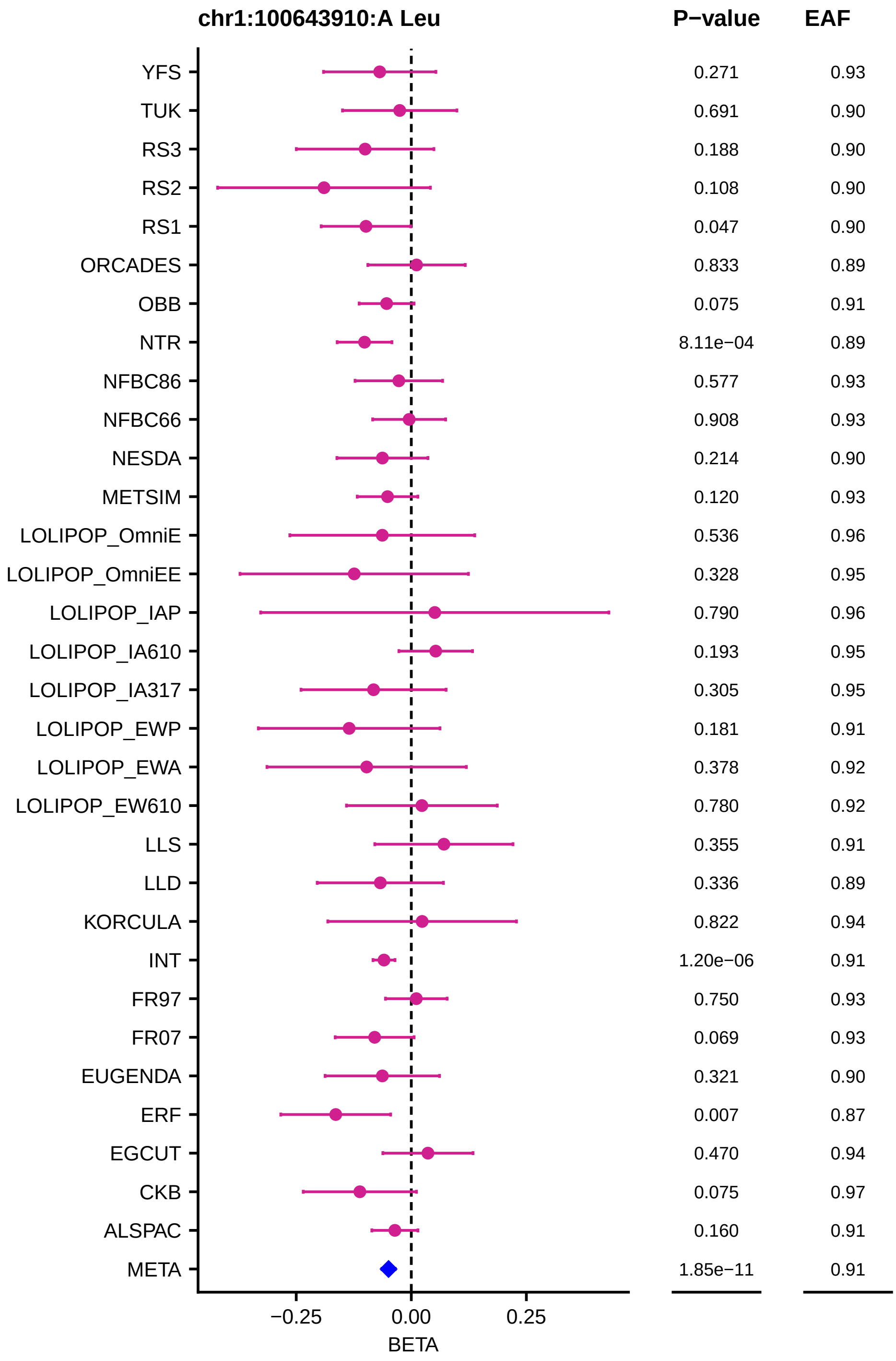
chr1:100643910:A Leu
P−value
EAF
YFS 0.271 0.93
TUK 0.691 0.90
RS3 0.188 0.90
RS2 0.108 0.90
RS1 0.047 0.90
ORCADES 0.833 0.89
OBB 0.075 0.91
NTR 8.11e−04 0.89
NFBC86 0.577 0.93
NFBC66 0.908 0.93
NESDA 0.214 0.90
METSIM 0.120 0.93
LOLIPOP_OmniE 0.536 0.96
LOLIPOP_OmniEE 0.328 0.95
LOLIPOP_IAP 0.790 0.96
LOLIPOP_IA610 0.193 0.95
LOLIPOP_IA317 0.305 0.95
LOLIPOP_EWP 0.181 0.91
LOLIPOP_EWA 0.378 0.92
LOLIPOP_EW610 0.780 0.92
LLS 0.355 0.91
LLD 0.336 0.89
KORCULA 0.822 0.94
INT 1.20e−06 0.91
FR97 0.750 0.93
FR07 0.069 0.93
EUGENDA 0.321 0.90
ERF 0.007 0.87
EGCUT 0.470 0.94
CKB 0.075 0.97
ALSPAC 0.160 0.91
META 1.85e−11 0.91
−0.25
0.00
0.25
BETA

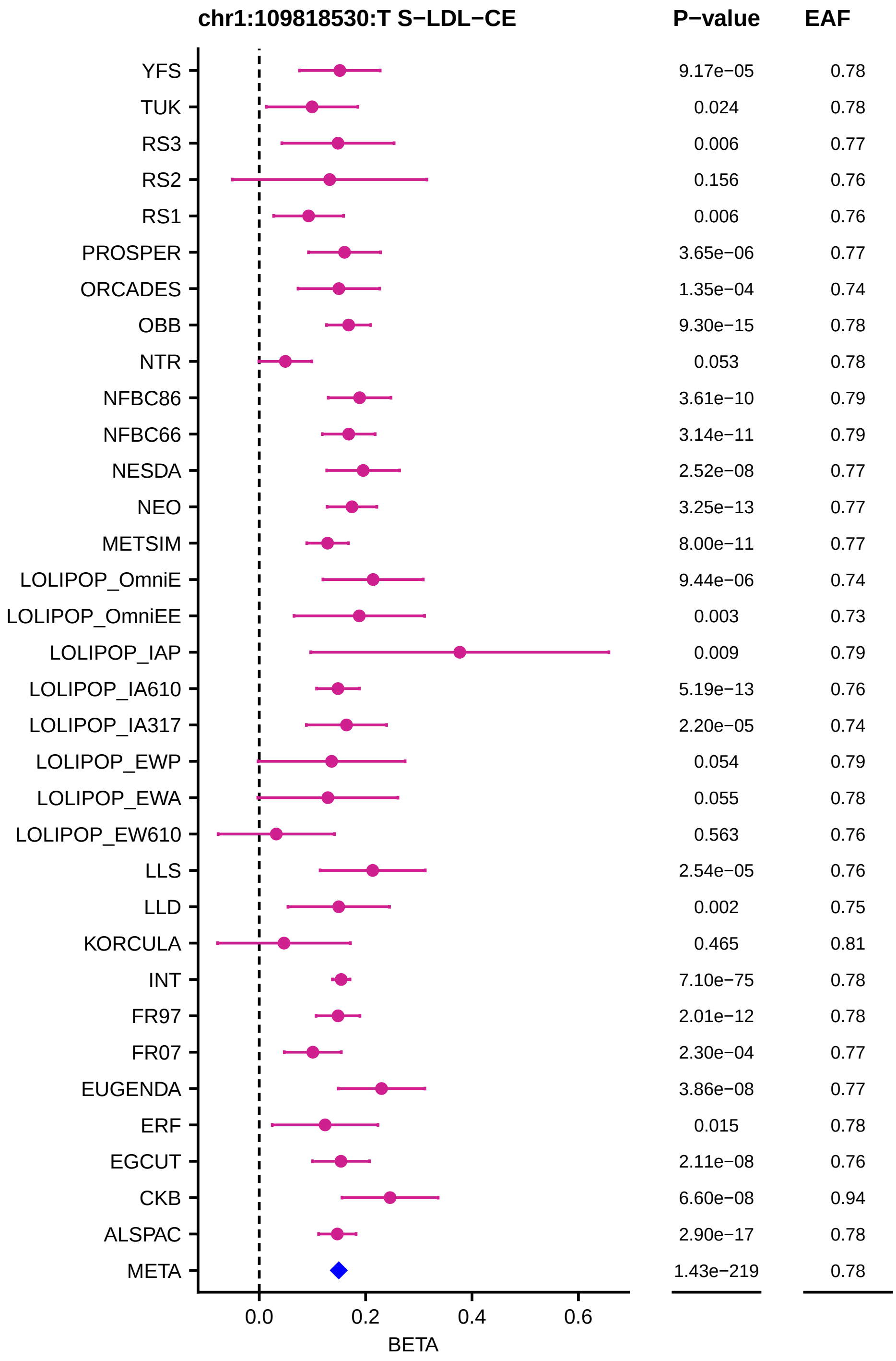

chr1:109818530:T S−LDL−CE
P−value
EAF
YFS 9.17e−05 0.78
TUK 0.024 0.78
RS3 0.006 0.77
RS2 0.156 0.76
RS1 0.006 0.76
PROSPER 3.65e−06 0.77
ORCADES 1.35e−04 0.74
OBB 9.30e−15 0.78
NTR 0.053 0.78
NFBC86 3.61e−10 0.79
NFBC66 3.14e−11 0.79
NESDA 2.52e−08 0.77
NEO 3.25e−13 0.77
METSIM 8.00e−11 0.77
LOLIPOP_OmniE 9.44e−06 0.74
LOLIPOP_OmniEE 0.003 0.73
LOLIPOP_IAP 0.009 0.79
LOLIPOP_IA610 5.19e−13 0.76
LOLIPOP_IA317 2.20e−05 0.74
LOLIPOP_EWP 0.054 0.79
LOLIPOP_EWA 0.055 0.78
LOLIPOP_EW610 0.563 0.76
LLS 2.54e−05 0.76
LLD 0.002 0.75
KORCULA 0.465 0.81
INT 7.10e−75 0.78
FR97 2.01e−12 0.78
FR07 2.30e−04 0.77
EUGENDA 3.86e−08 0.77
ERF 0.015 0.78
EGCUT 2.11e−08 0.76
CKB 6.60e−08 0.94
ALSPAC 2.90e−17 0.78
META 1.43e−219 0.78
0.0
0.2
0.4
0.6
BETA

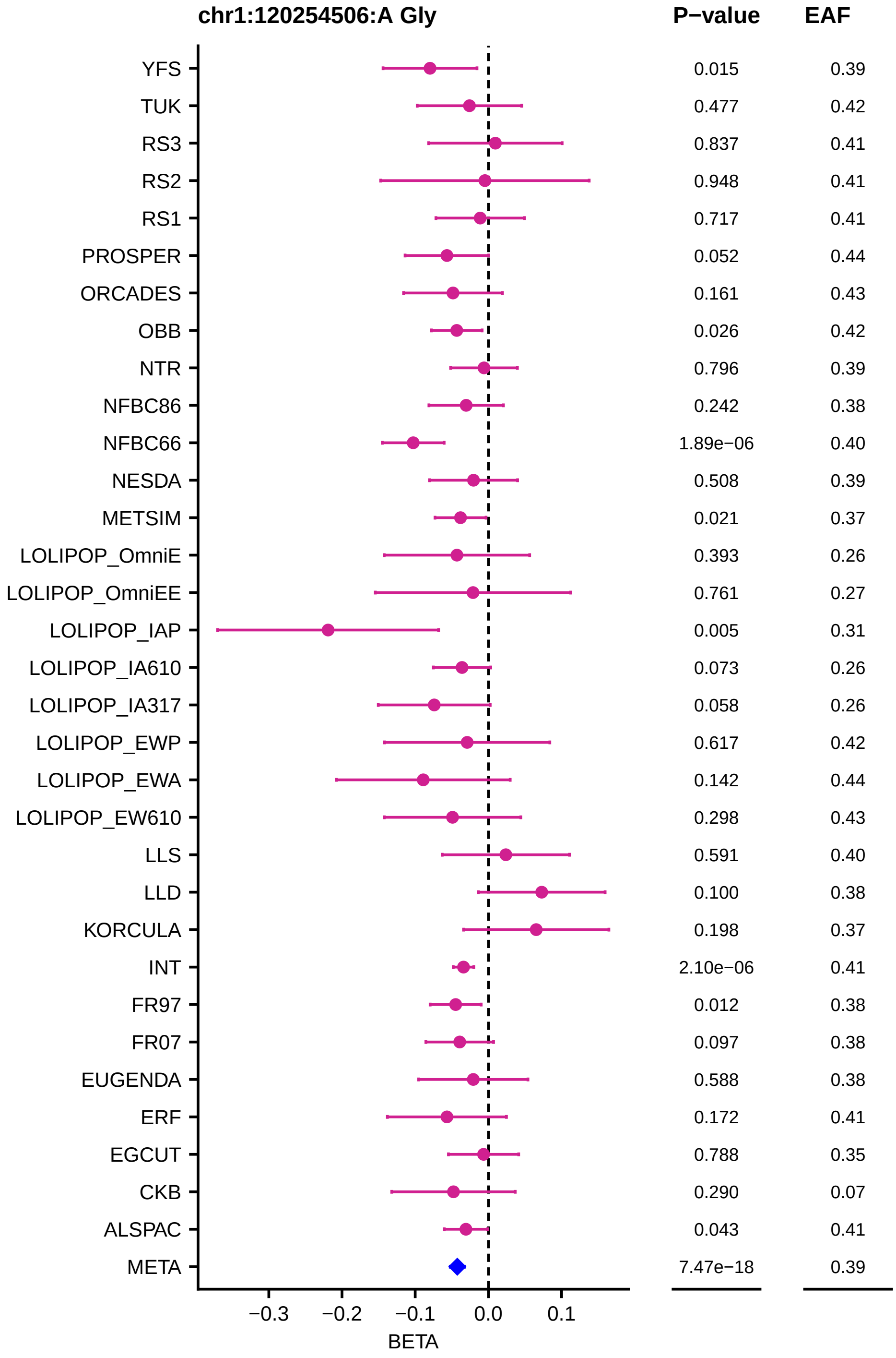
chr1:120254506:A Gly
P−value
EAF
YFS 0.015 0.39
TUK 0.477 0.42
RS3 0.837 0.41
RS2 0.948 0.41
RS1 0.717 0.41
PROSPER 0.052 0.44
ORCADES 0.161 0.43
OBB 0.026 0.42
NTR 0.796 0.39
NFBC86 0.242 0.38
NFBC66 1.89e−06 0.40
NESDA 0.508 0.39
METSIM 0.021 0.37
LOLIPOP_OmniE 0.393 0.26
LOLIPOP_OmniEE 0.761 0.27
LOLIPOP_IAP 0.005 0.31
LOLIPOP_IA610 0.073 0.26
LOLIPOP_IA317 0.058 0.26
LOLIPOP_EWP 0.617 0.42
LOLIPOP_EWA 0.142 0.44
LOLIPOP_EW610 0.298 0.43
LLS 0.591 0.40
LLD 0.100 0.38
KORCULA 0.198 0.37
INT 2.10e−06 0.41
FR97 0.012 0.38
FR07 0.097 0.38
EUGENDA 0.588 0.38
ERF 0.172 0.41
EGCUT 0.788 0.35
CKB 0.290 0.07
ALSPAC 0.043 0.41
META 7.47e−18 0.39
−0.3 −0.2 −0.1 0.0 0.1
BETA

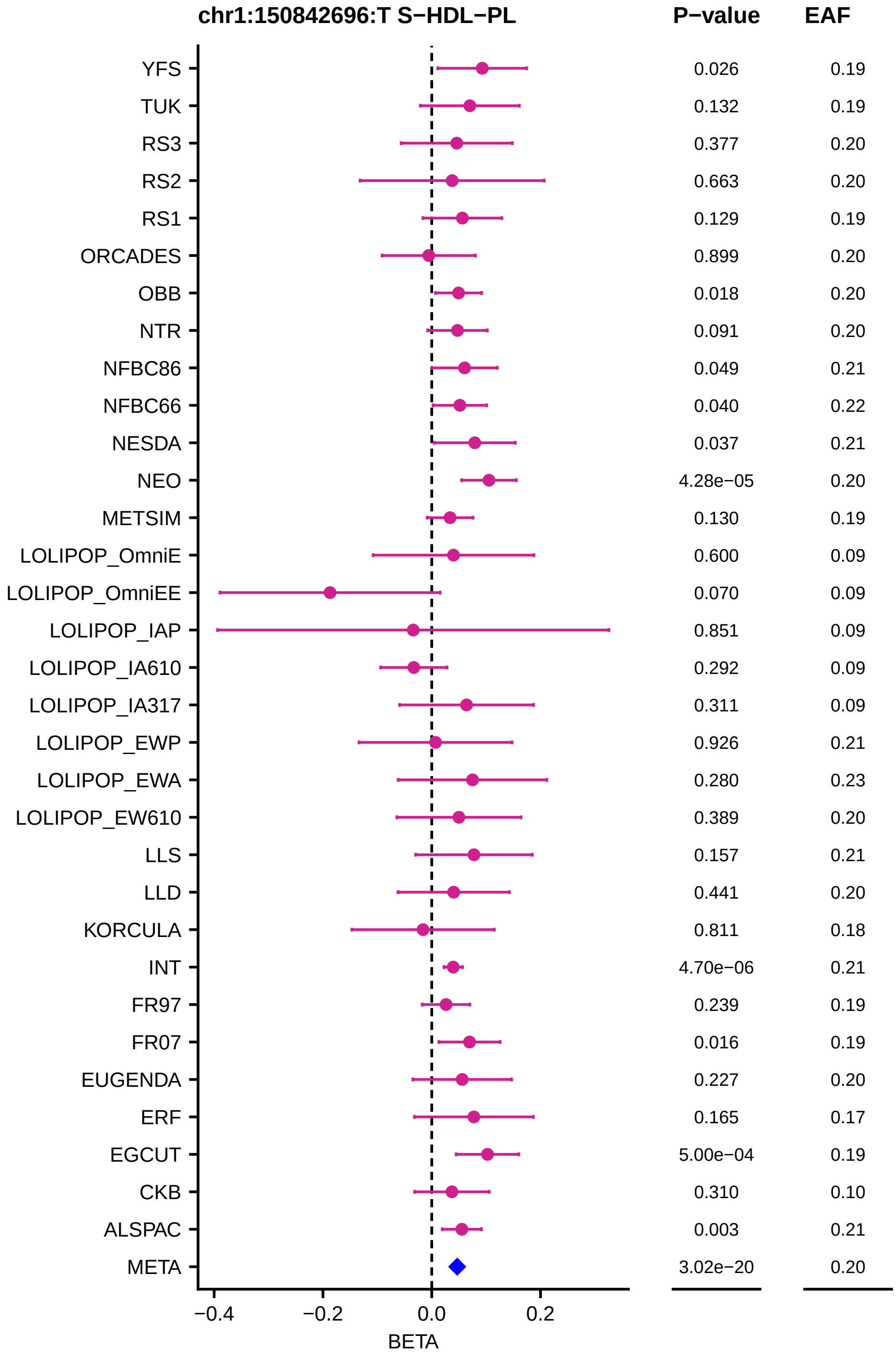
chr1:150842696:T S−HDL−PL
P−value
EAF
YFS 0.026 0.19
TUK 0.132 0.19
RS3 0.377 0.20
RS2 0.663 0.20
RS1 0.129 0.19
ORCADES 0.899 0.20
OBB 0.018 0.20
NTR 0.091 0.20
NFBC86 0.049 0.21
NFBC66 0.040 0.22
NESDA 0.037 0.21
NEO 4.28e−05 0.20
METSIM 0.130 0.19
LOLIPOP_OmniE 0.600 0.09
LOLIPOP_OmniEE 0.070 0.09
LOLIPOP_IAP 0.851 0.09
LOLIPOP_IA610 0.292 0.09
LOLIPOP_IA317 0.311 0.09
LOLIPOP_EWP 0.926 0.21
LOLIPOP_EWA 0.280 0.23
LOLIPOP_EW610 0.389 0.20
LLS 0.157 0.21
LLD 0.441 0.20
KORCULA 0.811 0.18
INT 4.70e−06 0.21
FR97 0.239 0.19
FR07 0.016 0.19
EUGENDA 0.227 0.20
ERF 0.165 0.17
EGCUT 5.00e−04 0.19
CKB 0.310 0.10
ALSPAC 0.003 0.21
META 3.02e−20 0.20
−0.4
−0.2
0.0
0.2
BETA

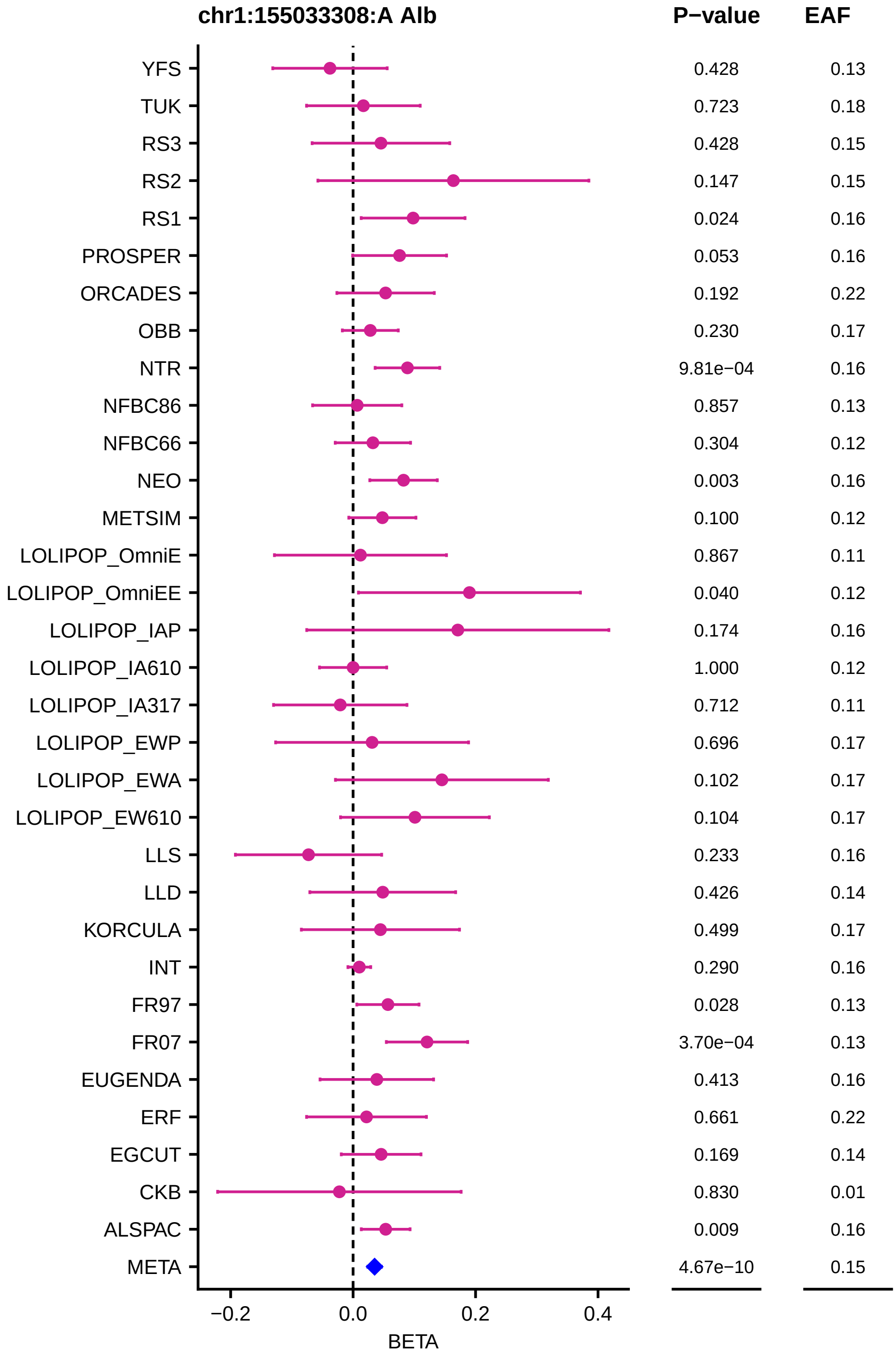
chr1:155033308:A Alb
P−value
EAF
YFS 0.428 0.13
TUK 0.723 0.18
RS3 0.428 0.15
RS2 0.147 0.15
RS1 0.024 0.16
PROSPER 0.053 0.16
ORCADES 0.192 0.22
OBB 0.230 0.17
NTR 9.81e−04 0.16
NFBC86 0.857 0.13
NFBC66 0.304 0.12
NEO 0.003 0.16
METSIM 0.100 0.12
LOLIPOP_OmniE 0.867 0.11
LOLIPOP_OmniEE 0.040 0.12
LOLIPOP_IAP 0.174 0.16
LOLIPOP_IA610 1.000 0.12
LOLIPOP_IA317 0.712 0.11
LOLIPOP_EWP 0.696 0.17
LOLIPOP_EWA 0.102 0.17
LOLIPOP_EW610 0.104 0.17
LLS 0.233 0.16
LLD 0.426 0.14
KORCULA 0.499 0.17
INT 0.290 0.16
FR97 0.028 0.13
FR07 3.70e−04 0.13
EUGENDA 0.413 0.16
ERF 0.661 0.22
EGCUT 0.169 0.14
CKB 0.830 0.01
ALSPAC 0.009 0.16
META 4.67e−10 0.15
−0.2
0.0
0.2
0.4
BETA

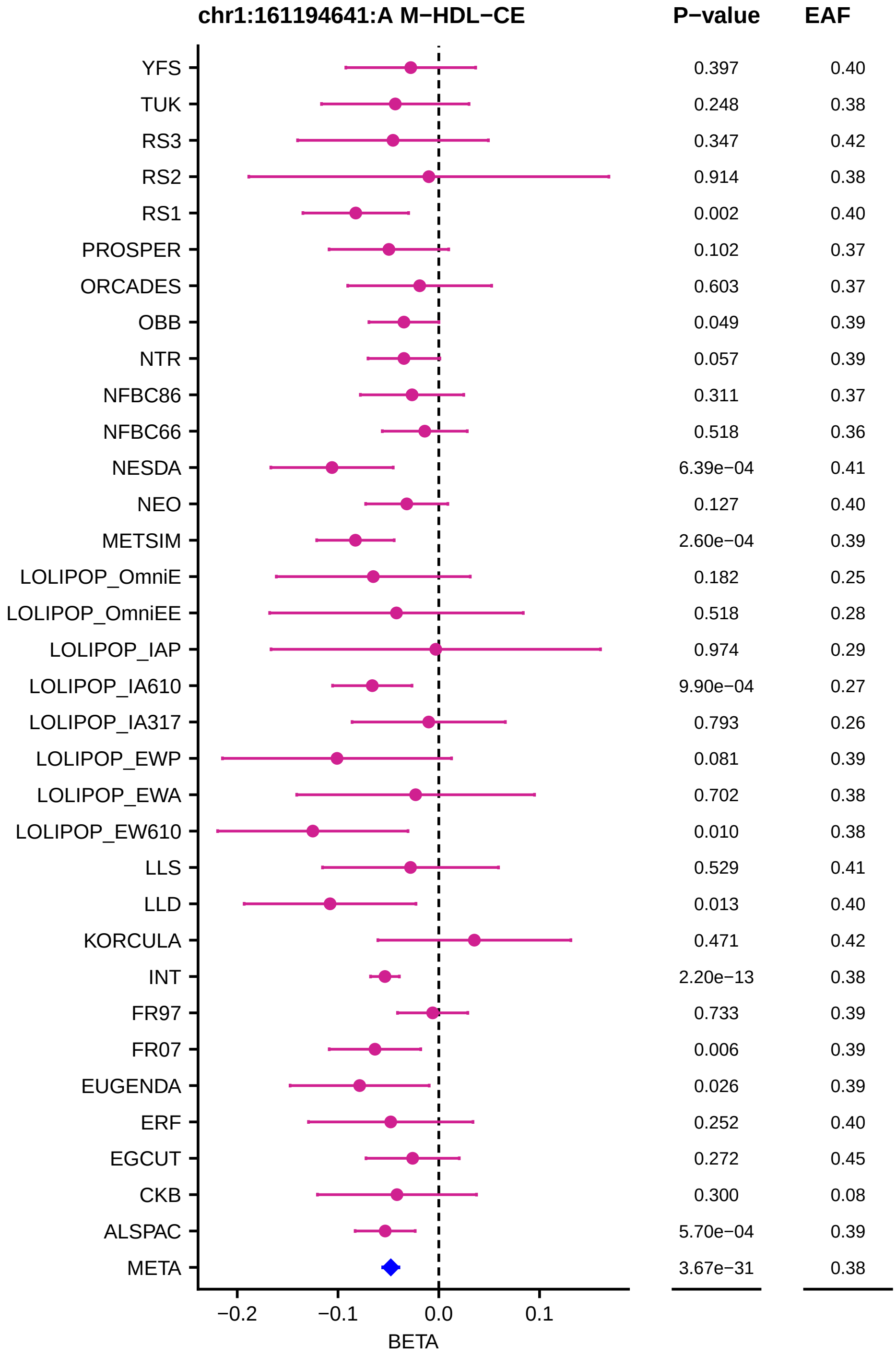
chr1:161194641:A M−HDL−CE
P−value
EAF
YFS 0.397 0.40
TUK 0.248 0.38
RS3 0.347 0.42
RS2 0.914 0.38
RS1 0.002 0.40
PROSPER 0.102 0.37
ORCADES 0.603 0.37
OBB 0.049 0.39
NTR 0.057 0.39
NFBC86 0.311 0.37
NFBC66 0.518 0.36
NESDA 6.39e−04 0.41
NEO 0.127 0.40
METSIM 2.60e−04 0.39
LOLIPOP_OmniE 0.182 0.25
LOLIPOP_OmniEE 0.518 0.28
LOLIPOP_IAP 0.974 0.29
LOLIPOP_IA610 9.90e−04 0.27
LOLIPOP_IA317 0.793 0.26
LOLIPOP_EWP 0.081 0.39
LOLIPOP_EWA 0.702 0.38
LOLIPOP_EW610 0.010 0.38
LLS 0.529 0.41
LLD 0.013 0.40
KORCULA 0.471 0.42
INT 2.20e−13 0.38
FR97 0.733 0.39
FR07 0.006 0.39
EUGENDA 0.026 0.39
ERF 0.252 0.40
EGCUT 0.272 0.45
CKB 0.300 0.08
ALSPAC 5.70e−04 0.39
META 3.67e−31 0.38
−0.2
−0.1
0.0
0.1
BETA

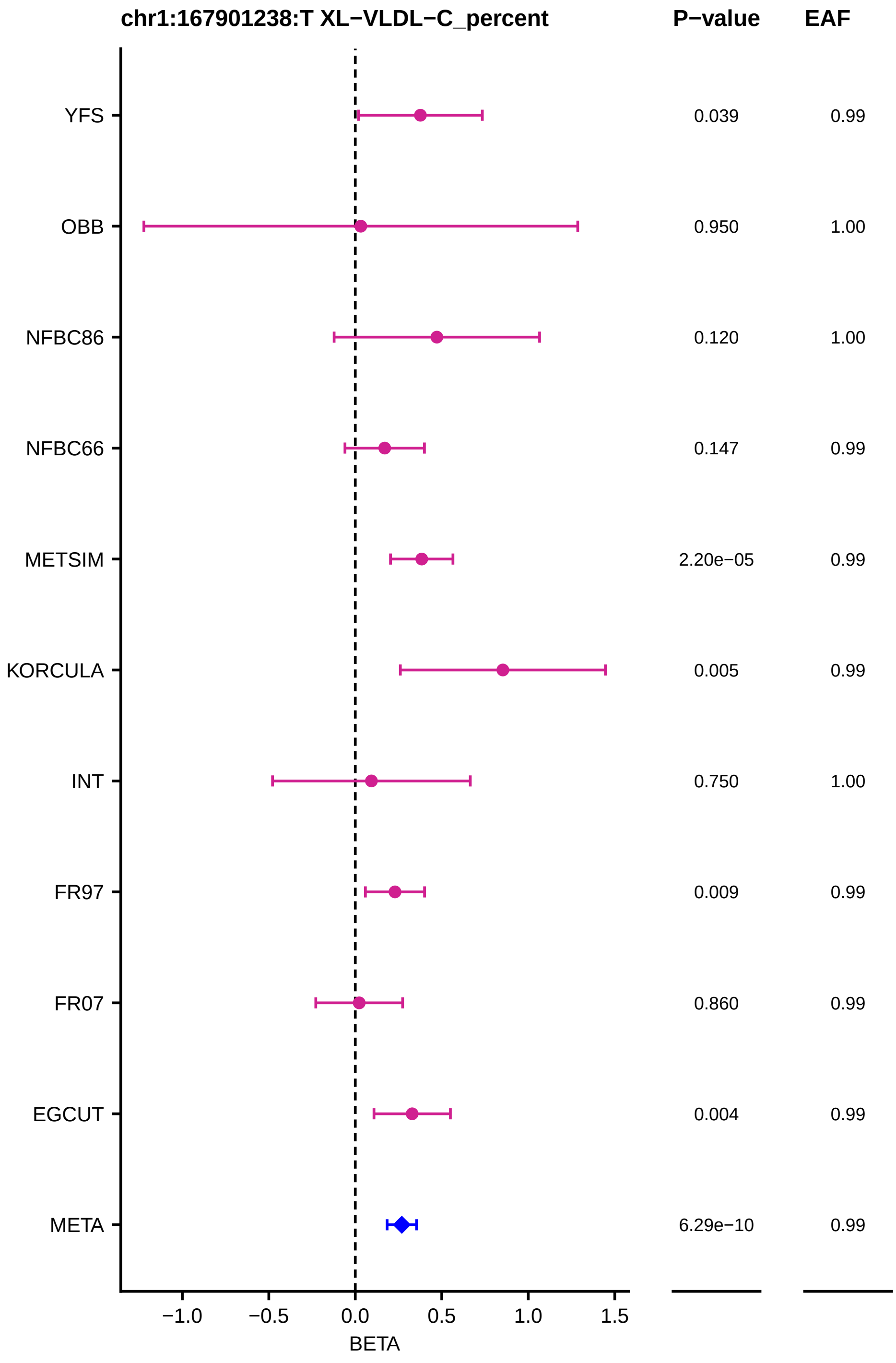
chr1:167901238:T XL−VLDL−C_percent
P−value
EAF
YFS
OBB
NFBC86
NFBC66
METSIM
KORCULA
INT
FR97
FR07
EGCUT
META
0.039
0.950
0.120
0.147
2.20e−05
0.005
0.750
0.009
0.860
0.004
6.29e−10
0.99
1.00
1.00
0.99
0.99
0.99
1.00
0.99
0.99
0.99
0.99
−1.0
−0.5
0.0
0.5
1.0
1.5
BETA

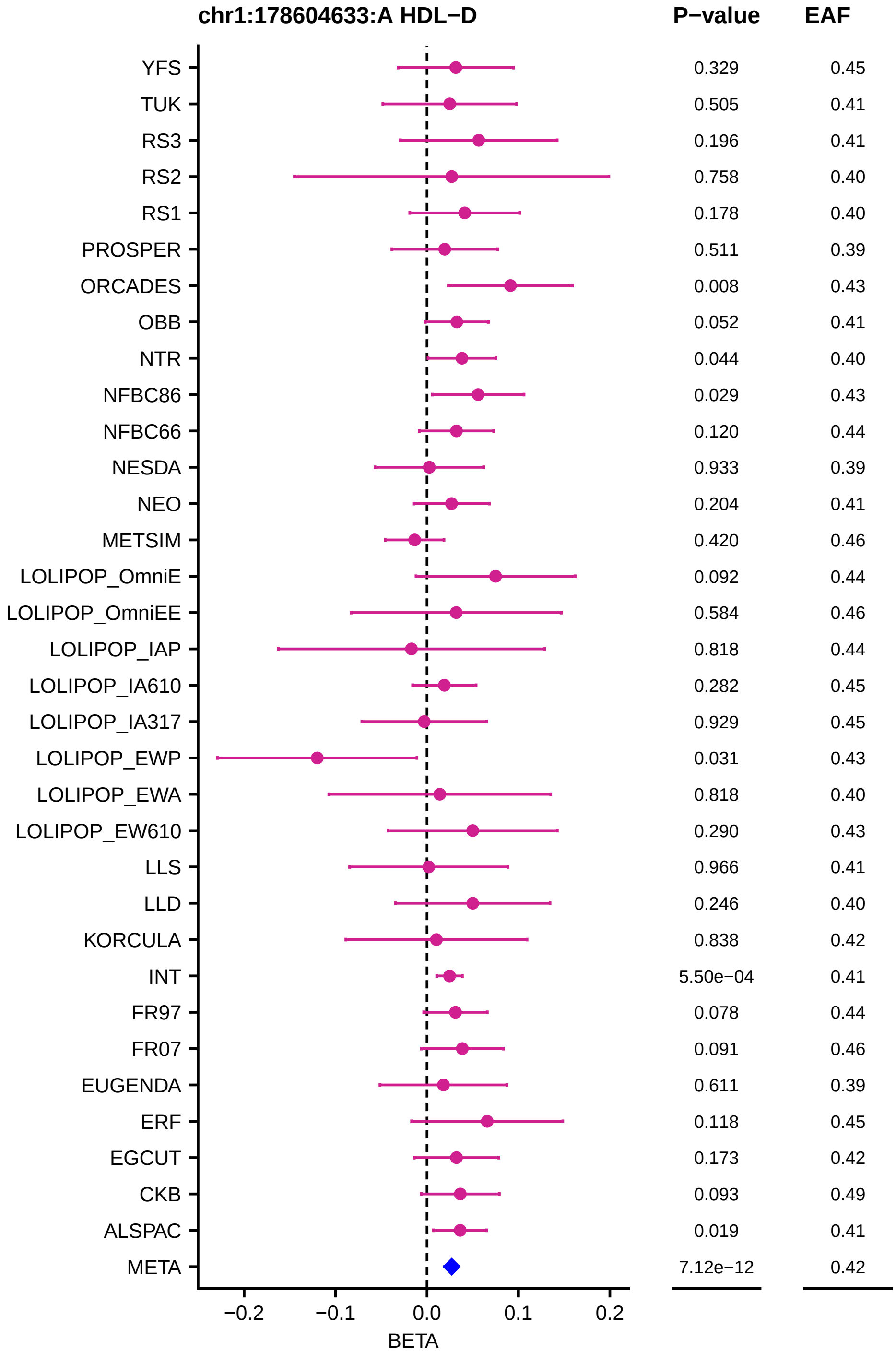
chr1:178604633:A HDL−D
P−value
EAF
YFS 0.329 0.45
TUK 0.505 0.41
RS3 0.196 0.41
RS2 0.758 0.40
RS1 0.178 0.40
PROSPER 0.511 0.39
ORCADES 0.008 0.43
OBB 0.052 0.41
NTR 0.044 0.40
NFBC86 0.029 0.43
NFBC66 0.120 0.44
NESDA 0.933 0.39
NEO 0.204 0.41
METSIM 0.420 0.46
LOLIPOP_OmniE 0.092 0.44
LOLIPOP_OmniEE 0.584 0.46
LOLIPOP_IAP 0.818 0.44
LOLIPOP_IA610 0.282 0.45
LOLIPOP_IA317 0.929 0.45
LOLIPOP_EWP 0.031 0.43
LOLIPOP_EWA 0.818 0.40
LOLIPOP_EW610 0.290 0.43
LLS 0.966 0.41
LLD 0.246 0.40
KORCULA 0.838 0.42
INT 5.50e−04 0.41
FR97 0.078 0.44
FR07 0.091 0.46
EUGENDA 0.611 0.39
ERF 0.118 0.45
EGCUT 0.173 0.42
CKB 0.093 0.49
ALSPAC 0.019 0.41
META 7.12e−12 0.42
−0.2 −0.1 0.0 0.1 0.2
BETA

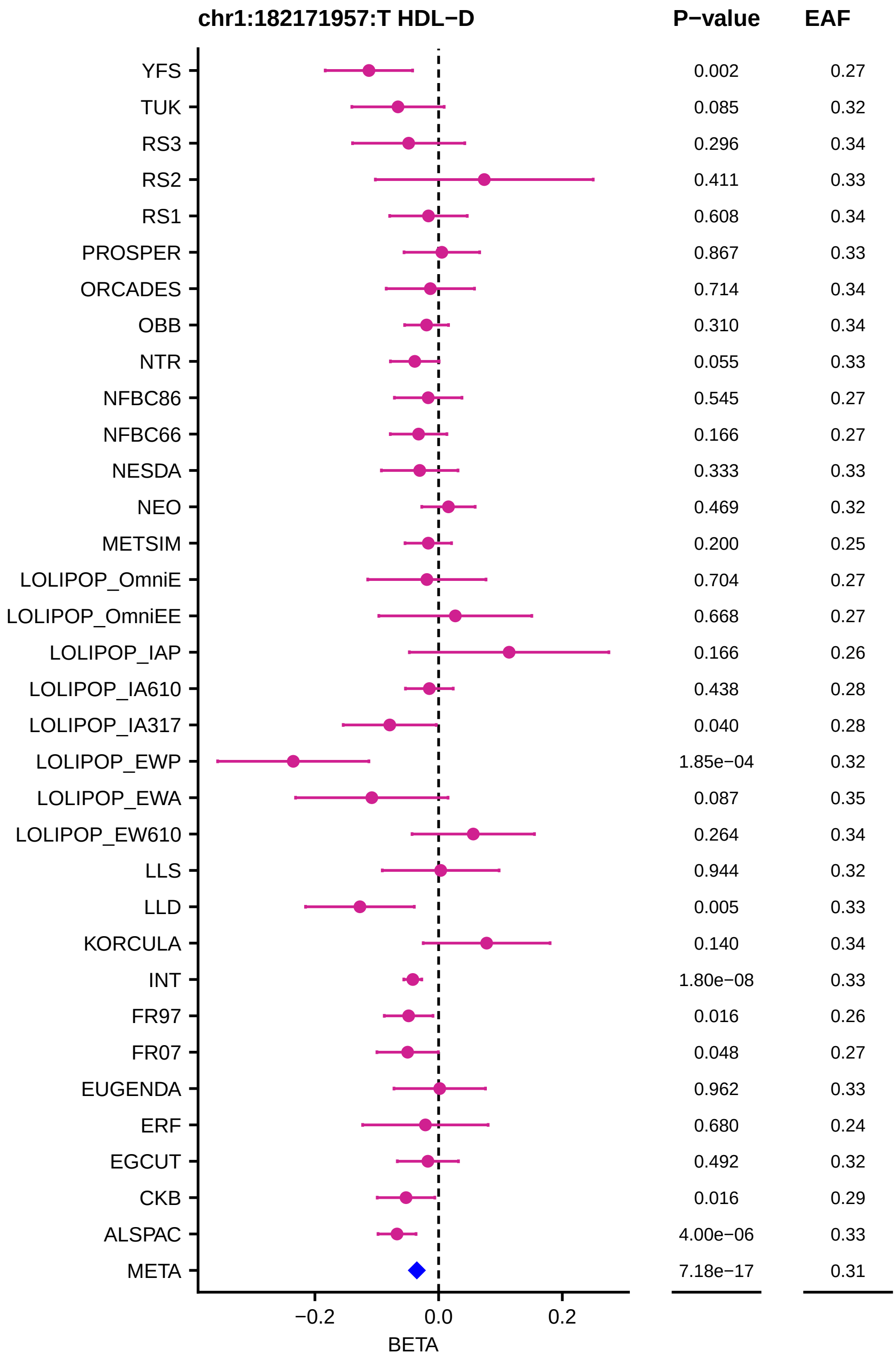
chr1:182171957:T HDL−D
P−value
EAF
YFS 0.002 0.27
TUK 0.085 0.32
RS3 0.296 0.34
RS2 0.411 0.33
RS1 0.608 0.34
PROSPER 0.867 0.33
ORCADES 0.714 0.34
OBB 0.310 0.34
NTR 0.055 0.33
NFBC86 0.545 0.27
NFBC66 0.166 0.27
NESDA 0.333 0.33
NEO 0.469 0.32
METSIM 0.200 0.25
LOLIPOP_OmniE 0.704 0.27
LOLIPOP_OmniEE 0.668 0.27
LOLIPOP_IAP 0.166 0.26
LOLIPOP_IA610 0.438 0.28
LOLIPOP_IA317 0.040 0.28
LOLIPOP_EWP 1.85e−04 0.32
LOLIPOP_EWA 0.087 0.35
LOLIPOP_EW610 0.264 0.34
LLS 0.944 0.32
LLD 0.005 0.33
KORCULA 0.140 0.34
INT 1.80e−08 0.33
FR97 0.016 0.26
FR07 0.048 0.27
EUGENDA 0.962 0.33
ERF 0.680 0.24
EGCUT 0.492 0.32
CKB 0.016 0.29
ALSPAC 4.00e−06 0.33
META 7.18e−17 0.31
−0.2
0.0
0.2
BETA

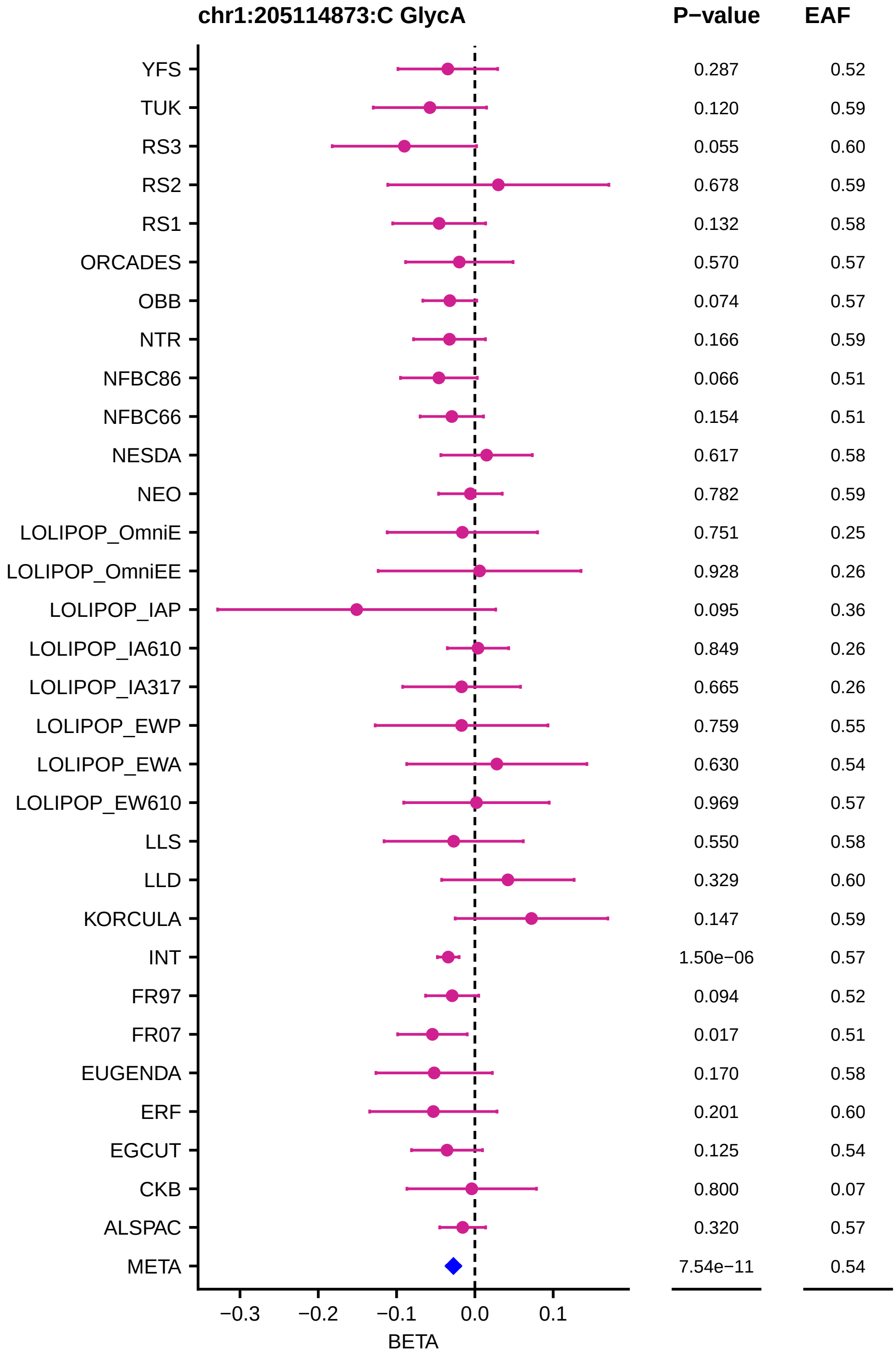
chr1:205114873:C GlycA
P−value
EAF
YFS
TUK
RS3
RS2
RS1
ORCADES
OBB
NTR
NFBC86
NFBC66
NESDA
NEO
LOLIPOP_OmniE
LOLIPOP_OmniEE
LOLIPOP_IAP
LOLIPOP_IA610
LOLIPOP_IA317
LOLIPOP_EWP
LOLIPOP_EWA
LOLIPOP_EW610
LLS
LLD
KORCULA
INT
FR97
FR07
EUGENDA
ERF
EGCUT
CKB
ALSPAC
META
0.287 0.52
0.120 0.59
0.055 0.60
0.678 0.59
0.132 0.58
0.570 0.57
0.074 0.57
0.166 0.59
0.066 0.51
0.154 0.51
0.617 0.58
0.782 0.59
0.751 0.25
0.928 0.26
0.095 0.36
0.849 0.26
0.665 0.26
0.759 0.55
0.630 0.54
0.969 0.57
0.550 0.58
0.329 0.60
0.147 0.59
1.50e−06 0.57
0.094 0.52
0.017 0.51
0.170 0.58
0.201 0.60
0.125 0.54
0.800 0.07
0.320 0.57
7.54e−11 0.54
−0.3
−0.2
−0.1
0.0
0.1
BETA

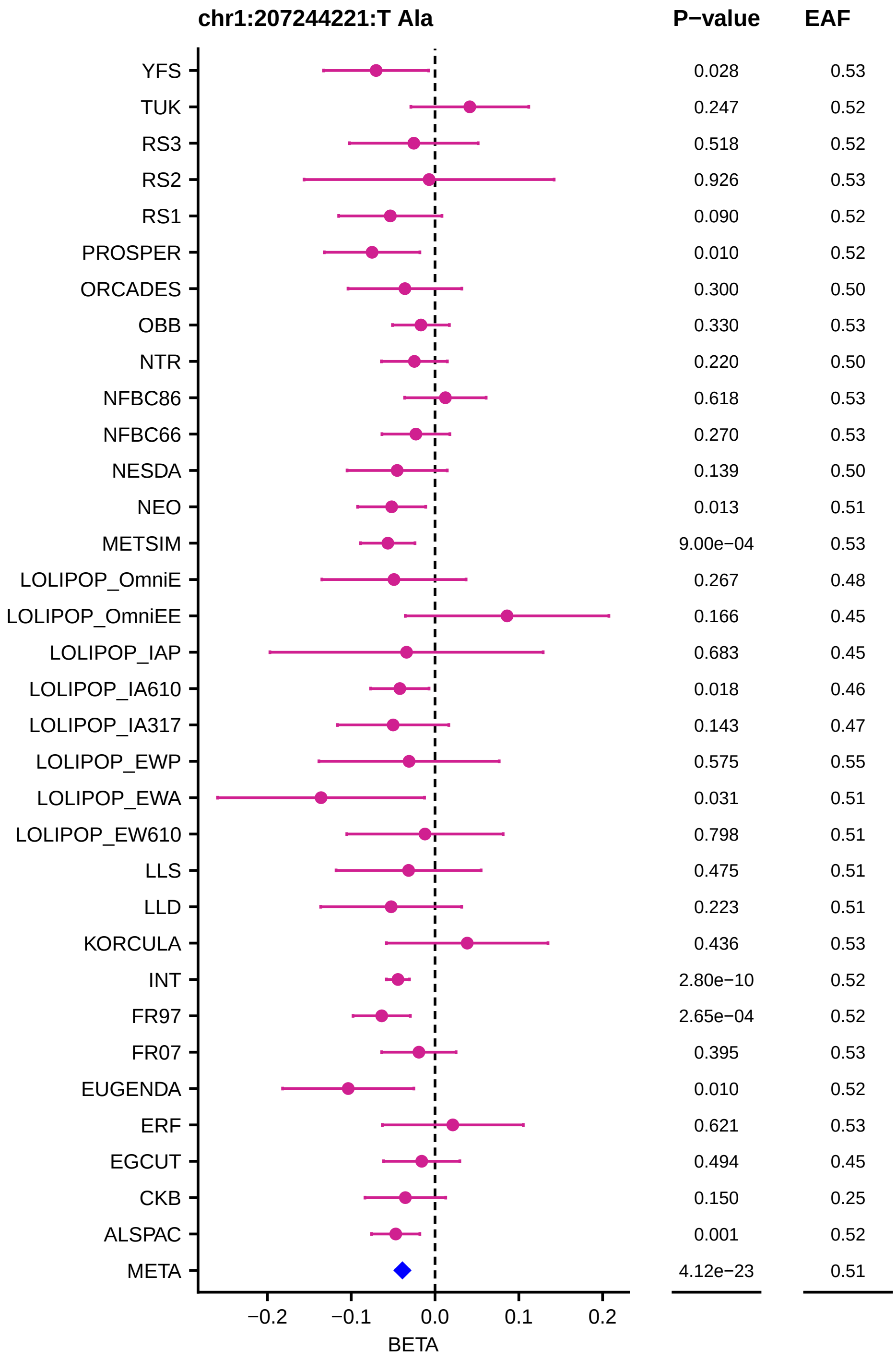
chr1:207244221:T Ala
P−value
EAF
YFS 0.028 0.53
TUK 0.247 0.52
RS3 0.518 0.52
RS2 0.926 0.53
RS1 0.090 0.52
PROSPER 0.010 0.52
ORCADES 0.300 0.50
OBB 0.330 0.53
NTR 0.220 0.50
NFBC86 0.618 0.53
NFBC66 0.270 0.53
NESDA 0.139 0.50
NEO 0.013 0.51
METSIM 9.00e−04 0.53
LOLIPOP_OmniE 0.267 0.48
LOLIPOP_OmniEE 0.166 0.45
LOLIPOP_IAP 0.683 0.45
LOLIPOP_IA610 0.018 0.46
LOLIPOP_IA317 0.143 0.47
LOLIPOP_EWP 0.575 0.55
LOLIPOP_EWA 0.031 0.51
LOLIPOP_EW610 0.798 0.51
LLS 0.475 0.51
LLD 0.223 0.51
KORCULA 0.436 0.53
INT 2.80e−10 0.52
FR97 2.65e−04 0.52
FR07 0.395 0.53
EUGENDA 0.010 0.52
ERF 0.621 0.53
EGCUT 0.494 0.45
CKB 0.150 0.25
ALSPAC 0.001 0.52
META 4.12e−23 0.51
−0.2
−0.1
0.0
0.1
0.2
BETA

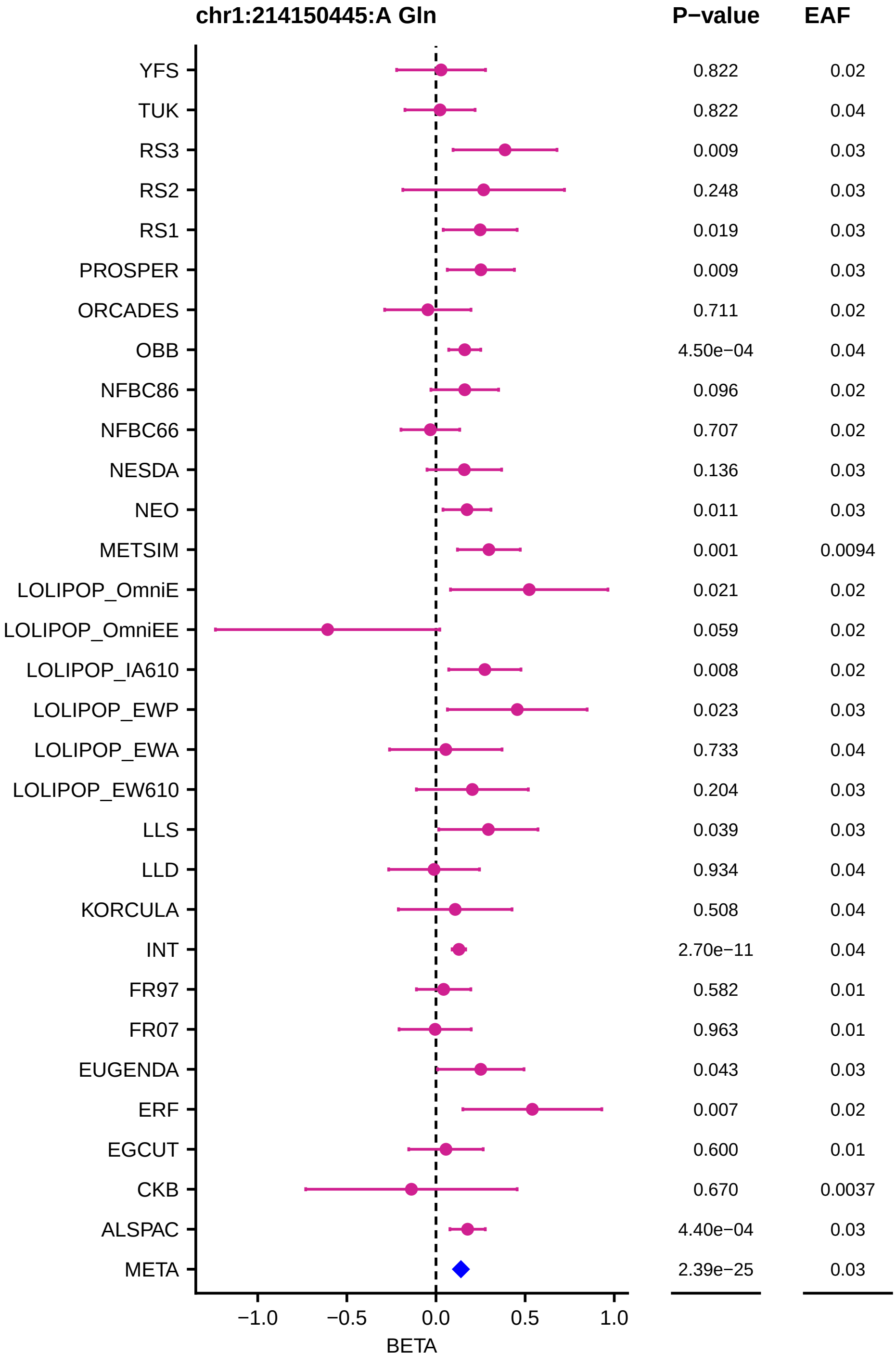
chr1:214150445:A Gln
P−value
EAF
YFS 0.822 0.02
TUK 0.822 0.04
RS3 0.009 0.03
RS2 0.248 0.03
RS1 0.019 0.03
PROSPER 0.009 0.03
ORCADES 0.711 0.02
OBB 4.50e−04 0.04
NFBC86 0.096 0.02
NFBC66 0.707 0.02
NESDA 0.136 0.03
NEO 0.011 0.03
METSIM 0.001 0.0094
LOLIPOP_OmniE 0.021 0.02
LOLIPOP_OmniEE 0.059 0.02
LOLIPOP_IA610 0.008 0.02
LOLIPOP_EWP 0.023 0.03
LOLIPOP_EWA 0.733 0.04
LOLIPOP_EW610 0.204 0.03
LLS 0.039 0.03
LLD 0.934 0.04
KORCULA 0.508 0.04
INT 2.70e−11 0.04
FR97 0.582 0.01
FR07 0.963 0.01
EUGENDA 0.043 0.03
ERF 0.007 0.02
EGCUT 0.600 0.01
CKB 0.670 0.0037
ALSPAC 4.40e−04 0.03
META 2.39e−25 0.03
−1.0
−0.5
0.0
0.5
1.0
BETA

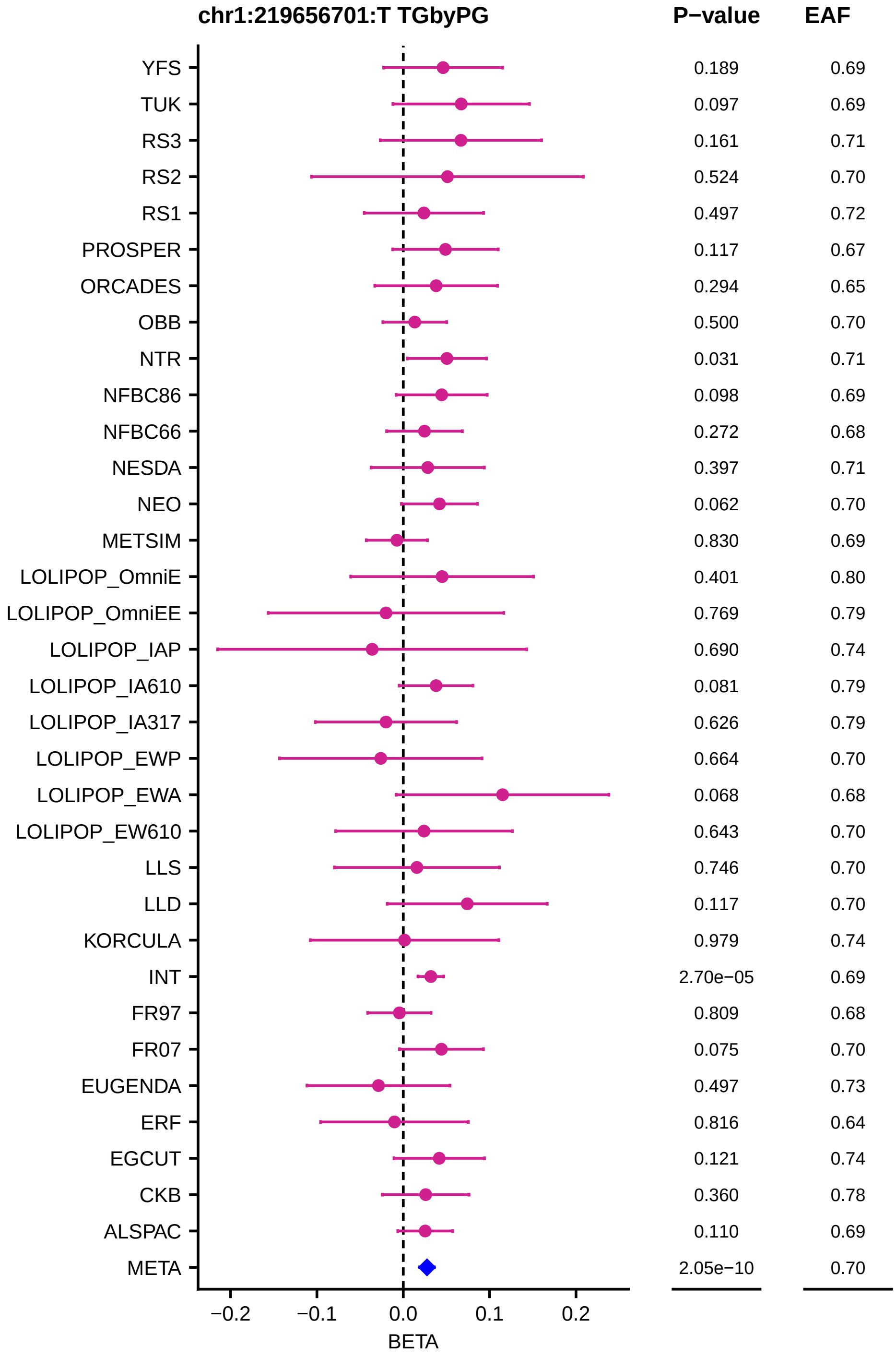

chr1:219656701:T TGbyPG
P−value
EAF
YFS 0.189 0.69
TUK 0.097 0.69
RS3 0.161 0.71
RS2 0.524 0.70
RS1 0.497 0.72
PROSPER 0.117 0.67
ORCADES 0.294 0.65
OBB 0.500 0.70
NTR 0.031 0.71
NFBC86 0.098 0.69
NFBC66 0.272 0.68
NESDA 0.397 0.71
NEO 0.062 0.70
METSIM 0.830 0.69
LOLIPOP_OmniE 0.401 0.80
LOLIPOP_OmniEE 0.769 0.79
LOLIPOP_IAP 0.690 0.74
LOLIPOP_IA610 0.081 0.79
LOLIPOP_IA317 0.626 0.79
LOLIPOP_EWP 0.664 0.70
LOLIPOP_EWA 0.068 0.68
LOLIPOP_EW610 0.643 0.70
LLS 0.746 0.70
LLD 0.117 0.70
KORCULA 0.979 0.74
INT 2.70e−05 0.69
FR97 0.809 0.68
FR07 0.075 0.70
EUGENDA 0.497 0.73
ERF 0.816 0.64
EGCUT 0.121 0.74
CKB 0.360 0.78
ALSPAC 0.110 0.69
META 2.05e−10 0.70
−0.2
−0.1
0.0
0.1
0.2
BETA

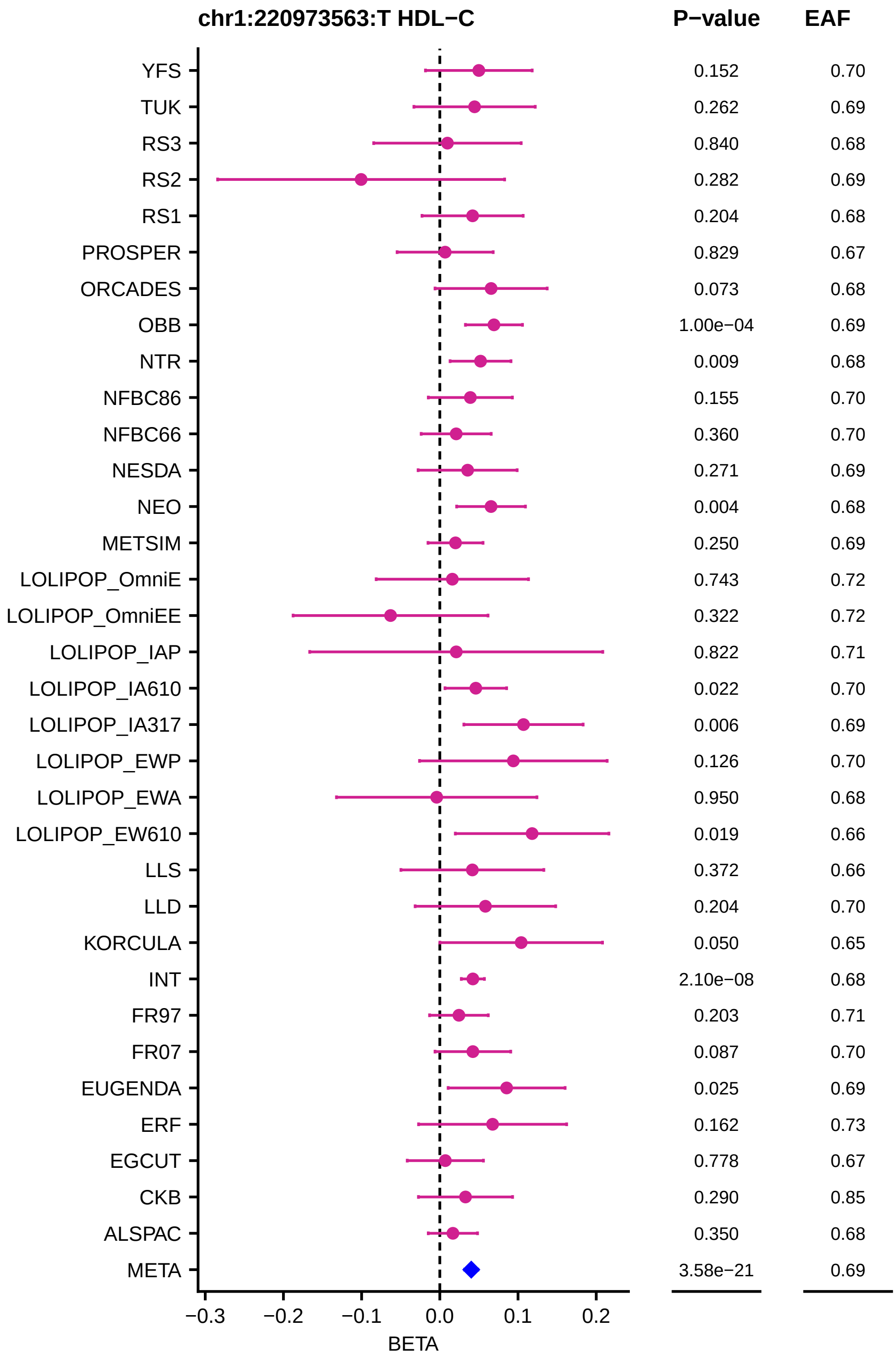
chr1:220973563:T HDL−C
P−value
EAF
YFS 0.152 0.70
TUK 0.262 0.69
RS3 0.840 0.68
RS2 0.282 0.69
RS1 0.204 0.68
PROSPER 0.829 0.67
ORCADES 0.073 0.68
OBB 1.00e−04 0.69
NTR 0.009 0.68
NFBC86 0.155 0.70
NFBC66 0.360 0.70
NESDA 0.271 0.69
NEO 0.004 0.68
METSIM 0.250 0.69
LOLIPOP_OmniE 0.743 0.72
LOLIPOP_OmniEE 0.322 0.72
LOLIPOP_IAP 0.822 0.71
LOLIPOP_IA610 0.022 0.70
LOLIPOP_IA317 0.006 0.69
LOLIPOP_EWP 0.126 0.70
LOLIPOP_EWA 0.950 0.68
LOLIPOP_EW610 0.019 0.66
LLS 0.372 0.66
LLD 0.204 0.70
KORCULA 0.050 0.65
INT 2.10e−08 0.68
FR97 0.203 0.71
FR07 0.087 0.70
EUGENDA 0.025 0.69
ERF 0.162 0.73
EGCUT 0.778 0.67
CKB 0.290 0.85
ALSPAC 0.350 0.68
META 3.58e−21 0.69
−0.3 −0.2 −0.1 0.0 0.1 0.2
BETA

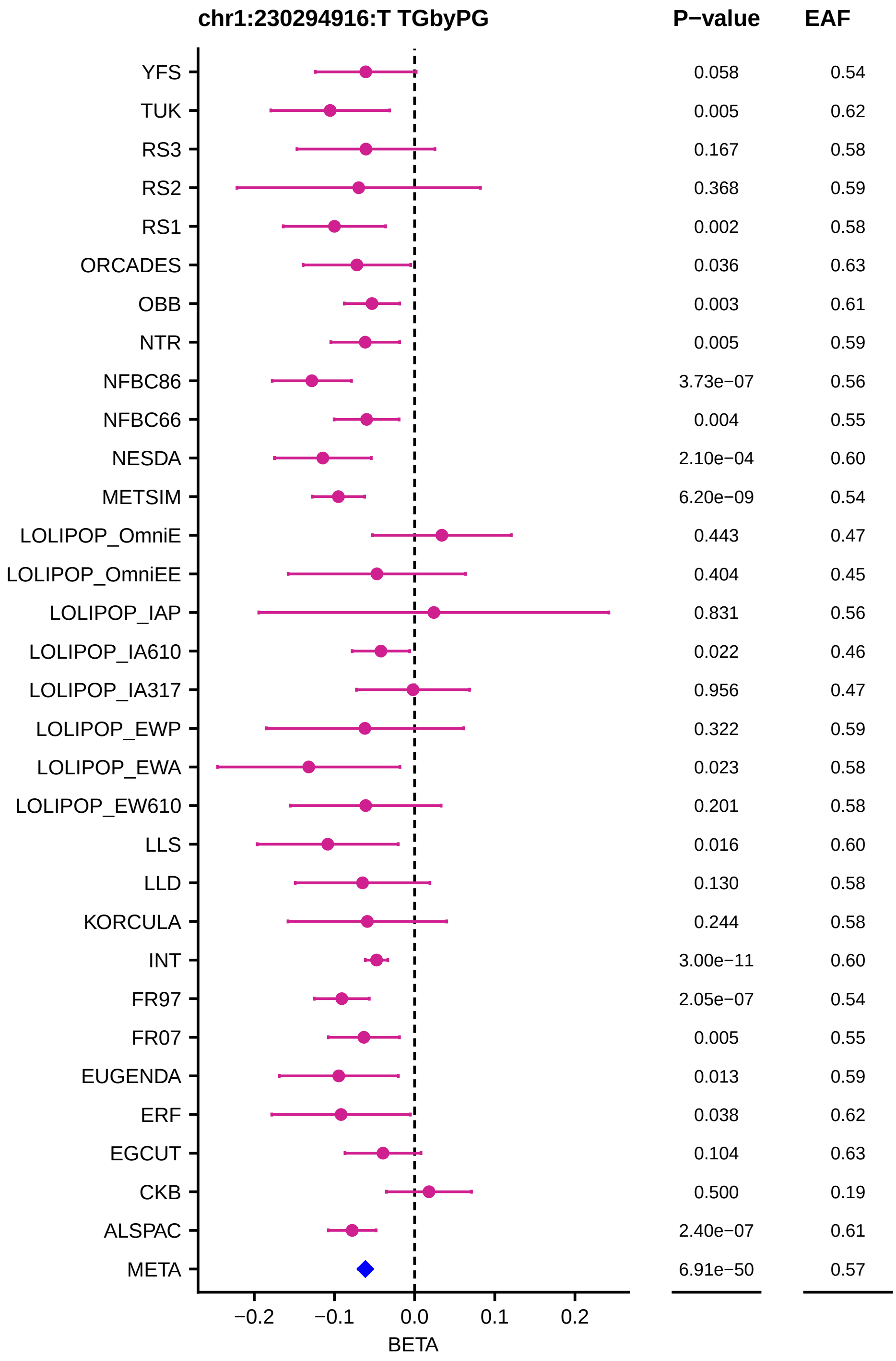
chr1:230294916:T TGbyPG
P−value
EAF
YFS 0.058 0.54
TUK 0.005 0.62
RS3 0.167 0.58
RS2 0.368 0.59
RS1 0.002 0.58
ORCADES 0.036 0.63
OBB 0.003 0.61
NTR 0.005 0.59
NFBC86 3.73e−07 0.56
NFBC66 0.004 0.55
NESDA 2.10e−04 0.60
METSIM 6.20e−09 0.54
LOLIPOP_OmniE 0.443 0.47
LOLIPOP_OmniEE 0.404 0.45
LOLIPOP_IAP 0.831 0.56
LOLIPOP_IA610 0.022 0.46
LOLIPOP_IA317 0.956 0.47
LOLIPOP_EWP 0.322 0.59
LOLIPOP_EWA 0.023 0.58
LOLIPOP_EW610 0.201 0.58
LLS 0.016 0.60
LLD 0.130 0.58
KORCULA 0.244 0.58
INT 3.00e−11 0.60
FR97 2.05e−07 0.54
FR07 0.005 0.55
EUGENDA 0.013 0.59
ERF 0.038 0.62
EGCUT 0.104 0.63
CKB 0.500 0.19
ALSPAC 2.40e−07 0.61
META 6.91e−50 0.57
−0.2
−0.1
0.0
0.1
0.2
BETA

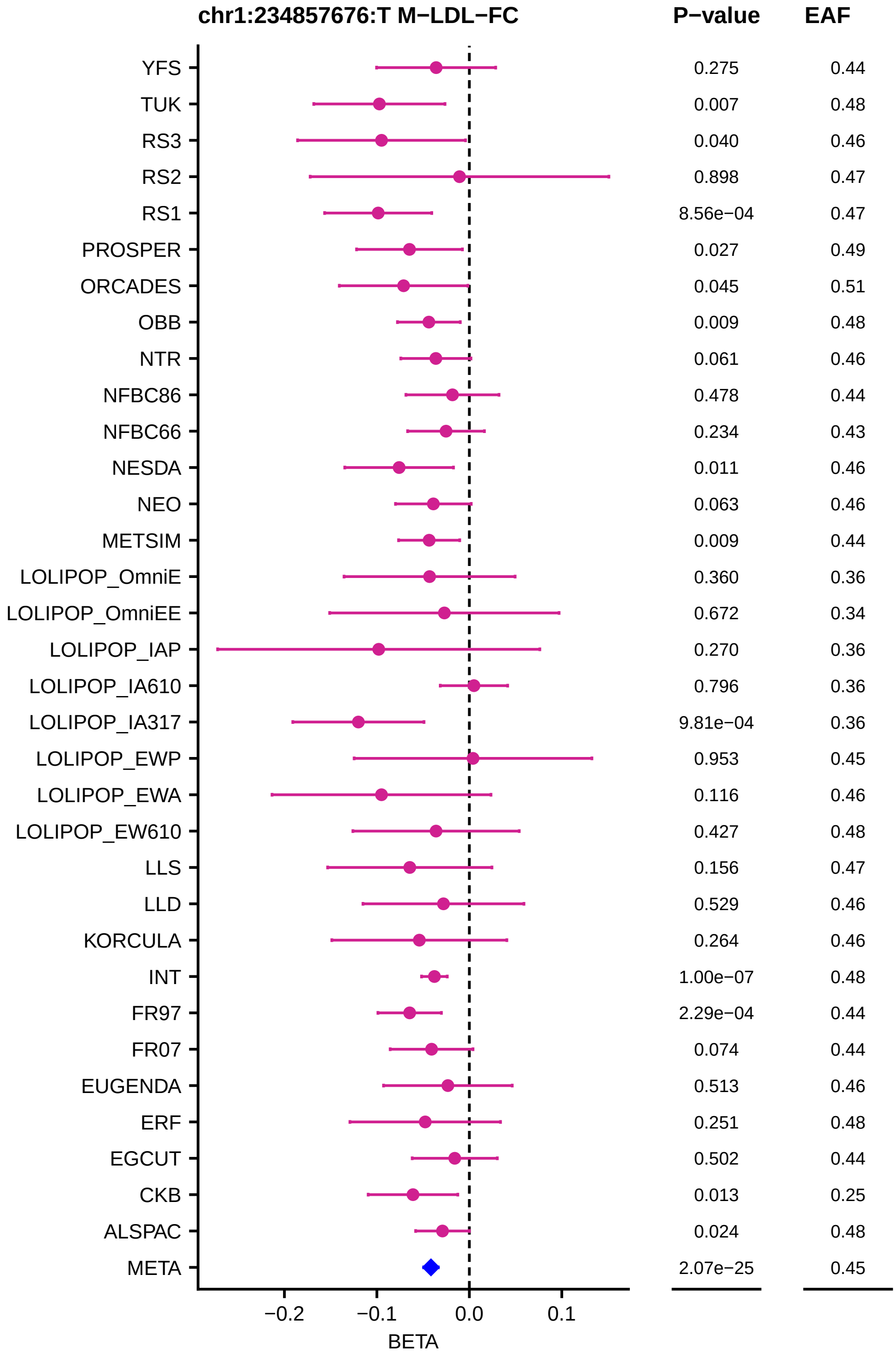
chr1:234857676:T M−LDL−FC
P−value
EAF
YFS 0.275 0.44
TUK 0.007 0.48
RS3 0.040 0.46
RS2 0.898 0.47
RS1 8.56e−04 0.47
PROSPER 0.027 0.49
ORCADES 0.045 0.51
OBB 0.009 0.48
NTR 0.061 0.46
NFBC86 0.478 0.44
NFBC66 0.234 0.43
NESDA 0.011 0.46
NEO 0.063 0.46
METSIM 0.009 0.44
LOLIPOP_OmniE 0.360 0.36
LOLIPOP_OmniEE 0.672 0.34
LOLIPOP_IAP 0.270 0.36
LOLIPOP_IA610 0.796 0.36
LOLIPOP_IA317 9.81e−04 0.36
LOLIPOP_EWP 0.953 0.45
LOLIPOP_EWA 0.116 0.46
LOLIPOP_EW610 0.427 0.48
LLS 0.156 0.47
LLD 0.529 0.46
KORCULA 0.264 0.46
INT 1.00e−07 0.48
FR97 2.29e−04 0.44
FR07 0.074 0.44
EUGENDA 0.513 0.46
ERF 0.251 0.48
EGCUT 0.502 0.44
CKB 0.013 0.25
ALSPAC 0.024 0.48
META 2.07e−25 0.45
−0.2
−0.1
0.0
0.1
BETA

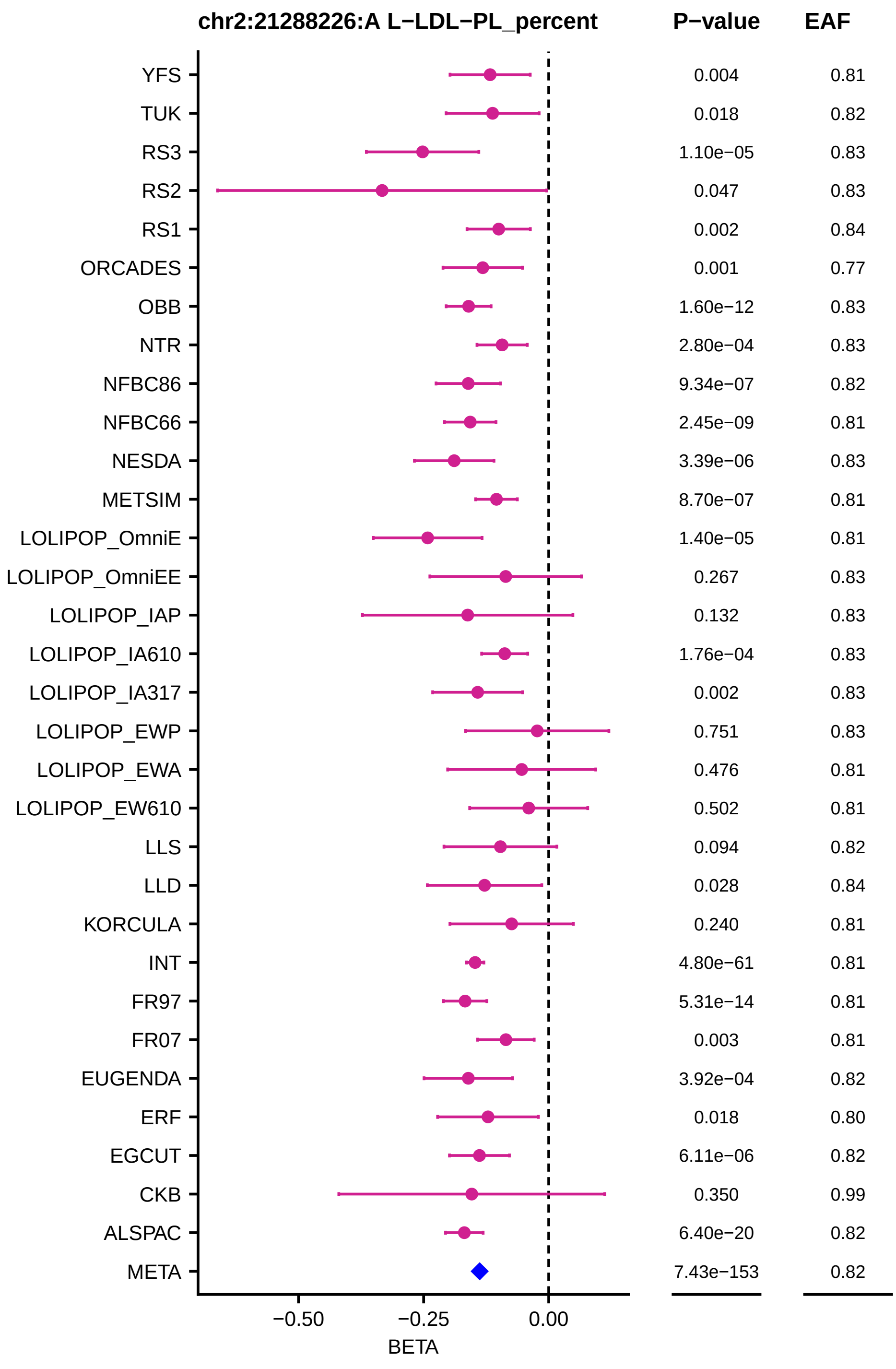
chr2:21288226:A L−LDL−PL_percent
P−value
EAF
YFS 0.004 0.81
TUK 0.018 0.82
RS3 1.10e−05 0.83
RS2 0.047 0.83
RS1 0.002 0.84
ORCADES 0.001 0.77
OBB 1.60e−12 0.83
NTR 2.80e−04 0.83
NFBC86 9.34e−07 0.82
NFBC66 2.45e−09 0.81
NESDA 3.39e−06 0.83
METSIM 8.70e−07 0.81
LOLIPOP_OmniE 1.40e−05 0.81
LOLIPOP_OmniEE 0.267 0.83
LOLIPOP_IAP 0.132 0.83
LOLIPOP_IA610 1.76e−04 0.83
LOLIPOP_IA317 0.002 0.83
LOLIPOP_EWP 0.751 0.83
LOLIPOP_EWA 0.476 0.81
LOLIPOP_EW610 0.502 0.81
LLS 0.094 0.82
LLD 0.028 0.84
KORCULA 0.240 0.81
INT 4.80e−61 0.81
FR97 5.31e−14 0.81
FR07 0.003 0.81
EUGENDA 3.92e−04 0.82
ERF 0.018 0.80
EGCUT 6.11e−06 0.82
CKB 0.350 0.99
ALSPAC 6.40e−20 0.82
META 7.43e−153 0.82
−0.50
−0.25
0.00
BETA

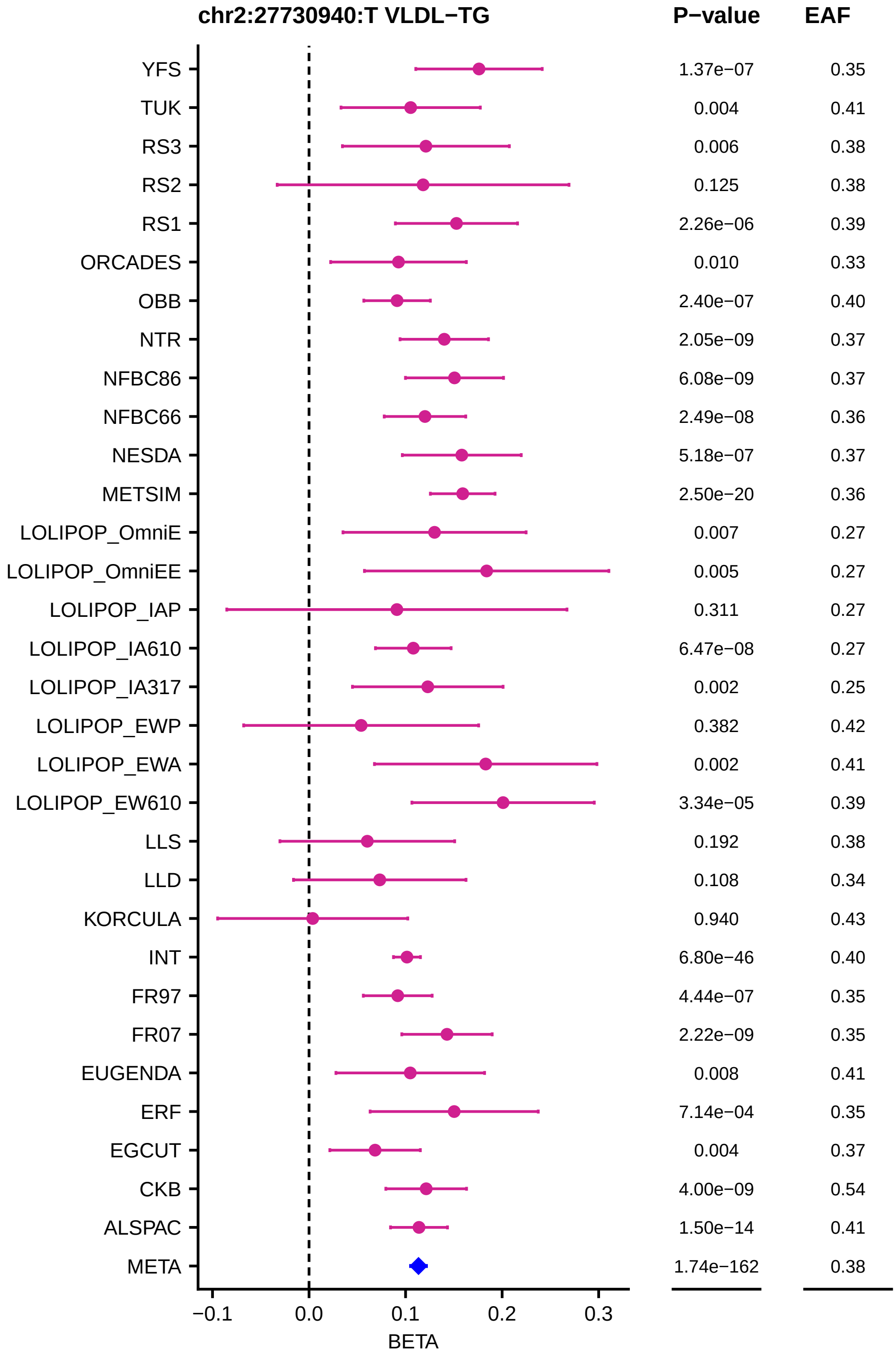
chr2:27730940:T VLDL−TG
P−value
EAF
YFS
TUK
RS3
RS2
RS1
ORCADES
OBB
NTR
NFBC86
NFBC66
NESDA
METSIM
LOLIPOP_OmniE
LOLIPOP_OmniEE
LOLIPOP_IAP
LOLIPOP_IA610
LOLIPOP_IA317
LOLIPOP_EWP
LOLIPOP_EWA
LOLIPOP_EW610
LLS
LLD
KORCULA
INT
FR97
FR07
EUGENDA
ERF
EGCUT
CKB
ALSPAC
META
1.37e−07
0.004
0.006
0.125
2.26e−06
0.010
2.40e−07
2.05e−09
6.08e−09
2.49e−08
5.18e−07
2.50e−20
0.007
0.005
0.311
6.47e−08
0.002
0.382
0.002
3.34e−05
0.192
0.108
0.940
6.80e−46
4.44e−07
2.22e−09
0.008
7.14e−04
0.004
4.00e−09
1.50e−14
1.74e−162
0.35
0.41
0.38
0.38
0.39
0.33
0.40
0.37
0.37
0.36
0.37
0.36
0.27
0.27
0.27
0.27
0.25
0.42
0.41
0.39
0.38
0.34
0.43
0.40
0.35
0.35
0.41
0.35
0.37
0.54
0.41
0.38
−0.1
0.0
0.1
0.2
0.3
BETA

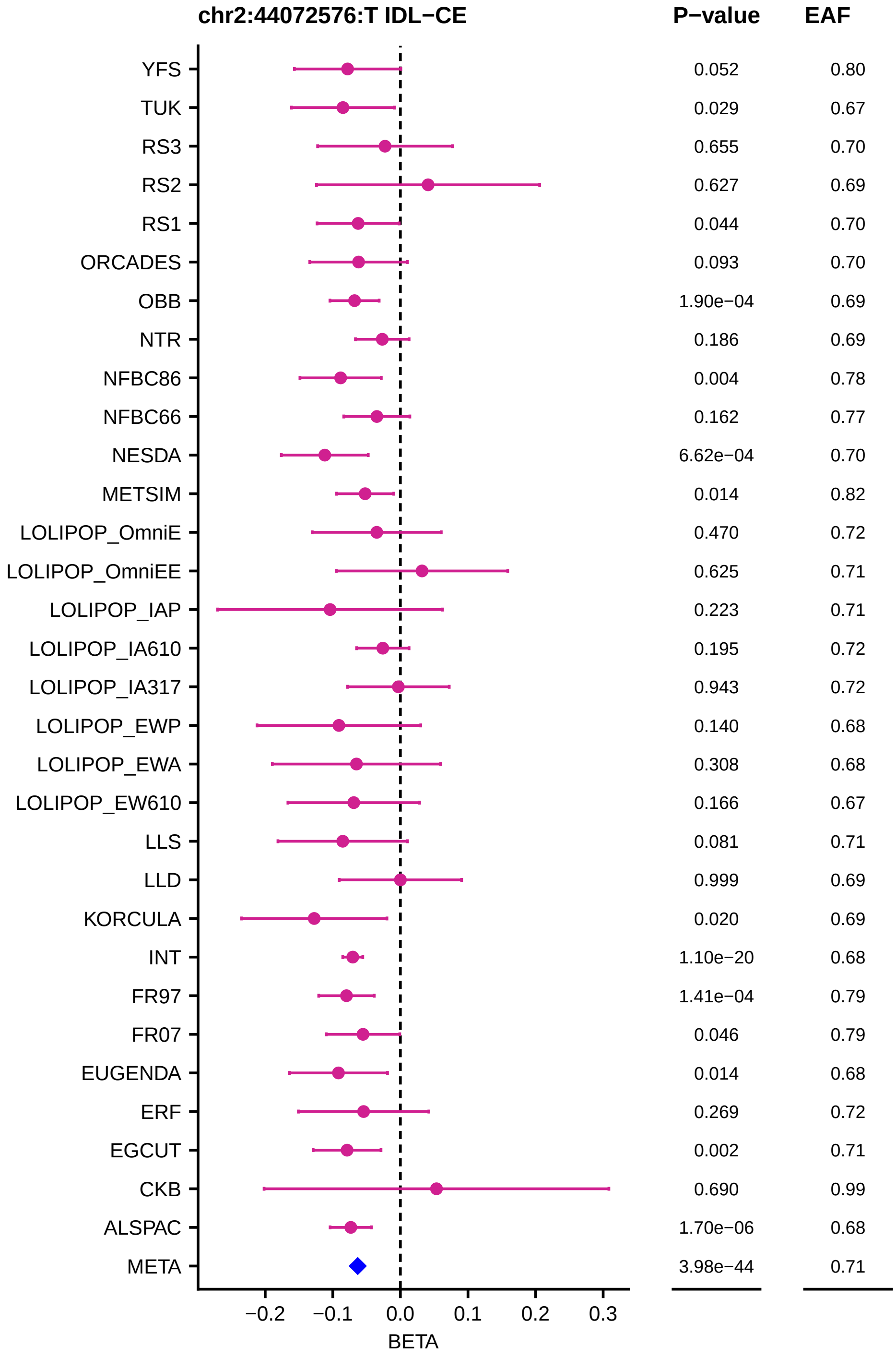
chr2:44072576:T IDL−CE
P−value
EAF
YFS 0.052 0.80
TUK 0.029 0.67
RS3 0.655 0.70
RS2 0.627 0.69
RS1 0.044 0.70
ORCADES 0.093 0.70
OBB 1.90e−04 0.69
NTR 0.186 0.69
NFBC86 0.004 0.78
NFBC66 0.162 0.77
NESDA 6.62e−04 0.70
METSIM 0.014 0.82
LOLIPOP_OmniE 0.470 0.72
LOLIPOP_OmniEE 0.625 0.71
LOLIPOP_IAP 0.223 0.71
LOLIPOP_IA610 0.195 0.72
LOLIPOP_IA317 0.943 0.72
LOLIPOP_EWP 0.140 0.68
LOLIPOP_EWA 0.308 0.68
LOLIPOP_EW610 0.166 0.67
LLS 0.081 0.71
LLD 0.999 0.69
KORCULA 0.020 0.69
INT 1.10e−20 0.68
FR97 1.41e−04 0.79
FR07 0.046 0.79
EUGENDA 0.014 0.68
ERF 0.269 0.72
EGCUT 0.002 0.71
CKB 0.690 0.99
ALSPAC 1.70e−06 0.68
META 3.98e−44 0.71
−0.2 −0.1 0.0 0.1 0.2 0.3
BETA

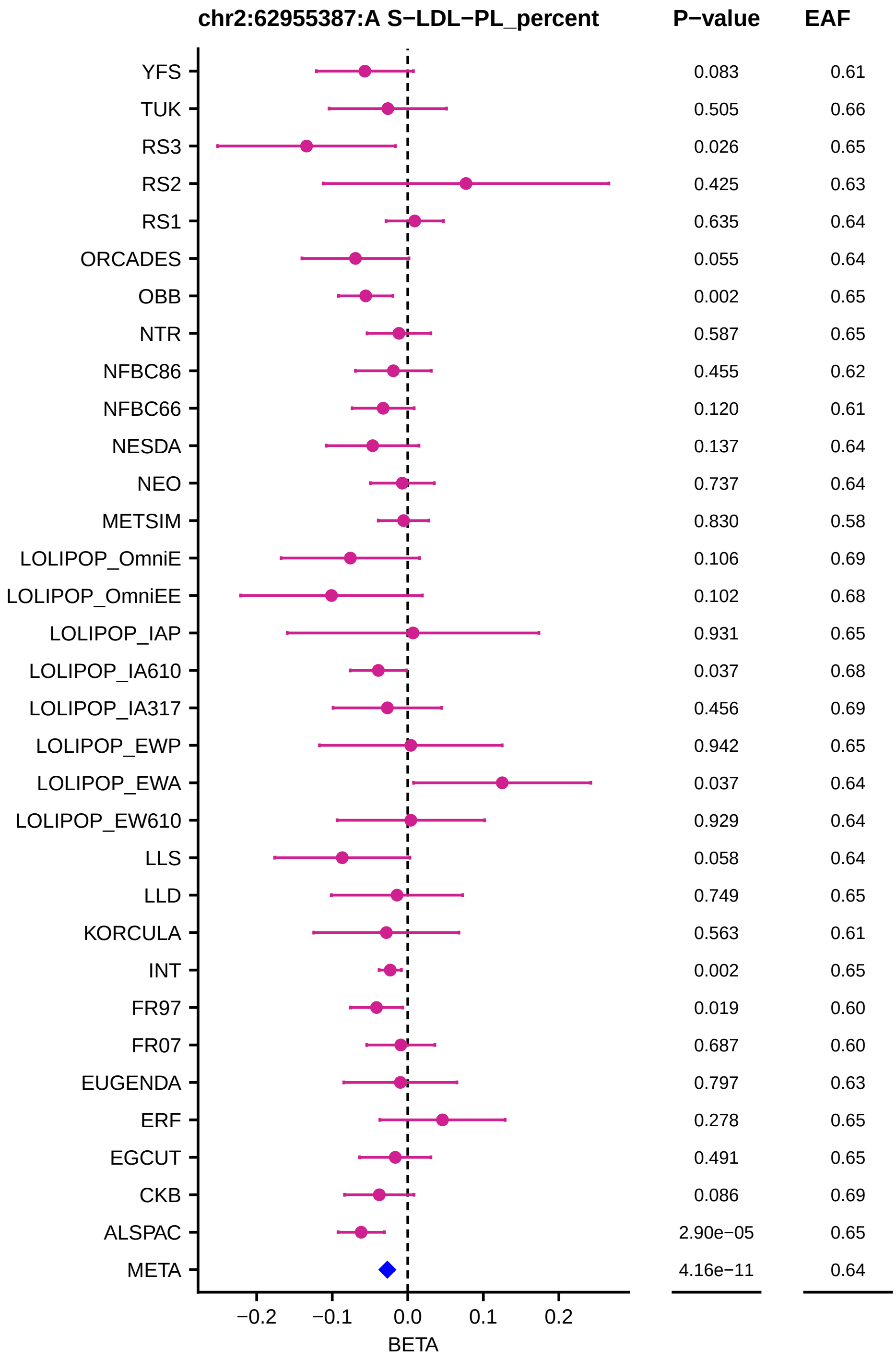
chr2:62955387:A S−LDL−PL_percent
P−value
EAF
YFS
TUK
RS3
RS2
RS1
ORCADES
OBB
NTR
NFBC86
NFBC66
NESDA
NEO
METSIM
LOLIPOP_OmniE
LOLIPOP_OmniEE
LOLIPOP_IAP
LOLIPOP_IA610
LOLIPOP_IA317
LOLIPOP_EWP
LOLIPOP_EWA
LOLIPOP_EW610
LLS
LLD
KORCULA
INT
FR97
FR07
EUGENDA
ERF
EGCUT
CKB
ALSPAC
META
0.083 0.61
0.505 0.66
0.026 0.65
0.425 0.63
0.635 0.64
0.055 0.64
0.002 0.65
0.587 0.65
0.455 0.62
0.120 0.61
0.137 0.64
0.737 0.64
0.830 0.58
0.106 0.69
0.102 0.68
0.931 0.65
0.037 0.68
0.456 0.69
0.942 0.65
0.037 0.64
0.929 0.64
0.058 0.64
0.749 0.65
0.563 0.61
0.002 0.65
0.019 0.60
0.687 0.60
0.797 0.63
0.278 0.65
0.491 0.65
0.086 0.69
2.90e−05 0.65
4.16e−11 0.64
−0.2
−0.1
0.0
0.1
0.2
BETA

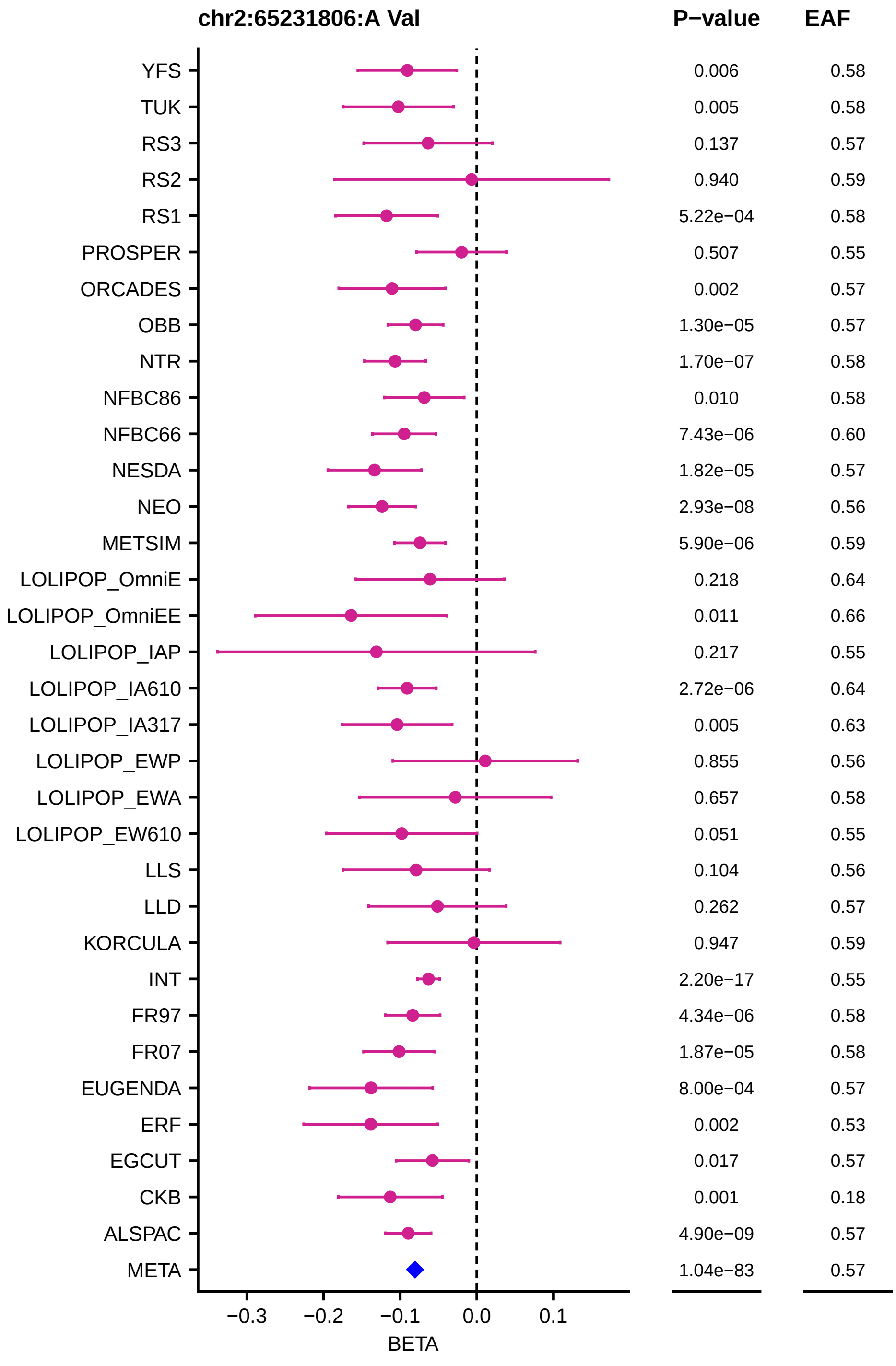
chr2:65231806:A Val
P−value
EAF
YFS 0.006 0.58
TUK 0.005 0.58
RS3 0.137 0.57
RS2 0.940 0.59
RS1 5.22e−04 0.58
PROSPER 0.507 0.55
ORCADES 0.002 0.57
OBB 1.30e−05 0.57
NTR 1.70e−07 0.58
NFBC86 0.010 0.58
NFBC66 7.43e−06 0.60
NESDA 1.82e−05 0.57
NEO 2.93e−08 0.56
METSIM 5.90e−06 0.59
LOLIPOP_OmniE 0.218 0.64
LOLIPOP_OmniEE 0.011 0.66
LOLIPOP_IAP 0.217 0.55
LOLIPOP_IA610 2.72e−06 0.64
LOLIPOP_IA317 0.005 0.63
LOLIPOP_EWP 0.855 0.56
LOLIPOP_EWA 0.657 0.58
LOLIPOP_EW610 0.051 0.55
LLS 0.104 0.56
LLD 0.262 0.57
KORCULA 0.947 0.59
INT 2.20e−17 0.55
FR97 4.34e−06 0.58
FR07 1.87e−05 0.58
EUGENDA 8.00e−04 0.57
ERF 0.002 0.53
EGCUT 0.017 0.57
CKB 0.001 0.18
ALSPAC 4.90e−09 0.57
META 1.04e−83 0.57
−0.3
−0.2
−0.1
0.0
0.1
BETA

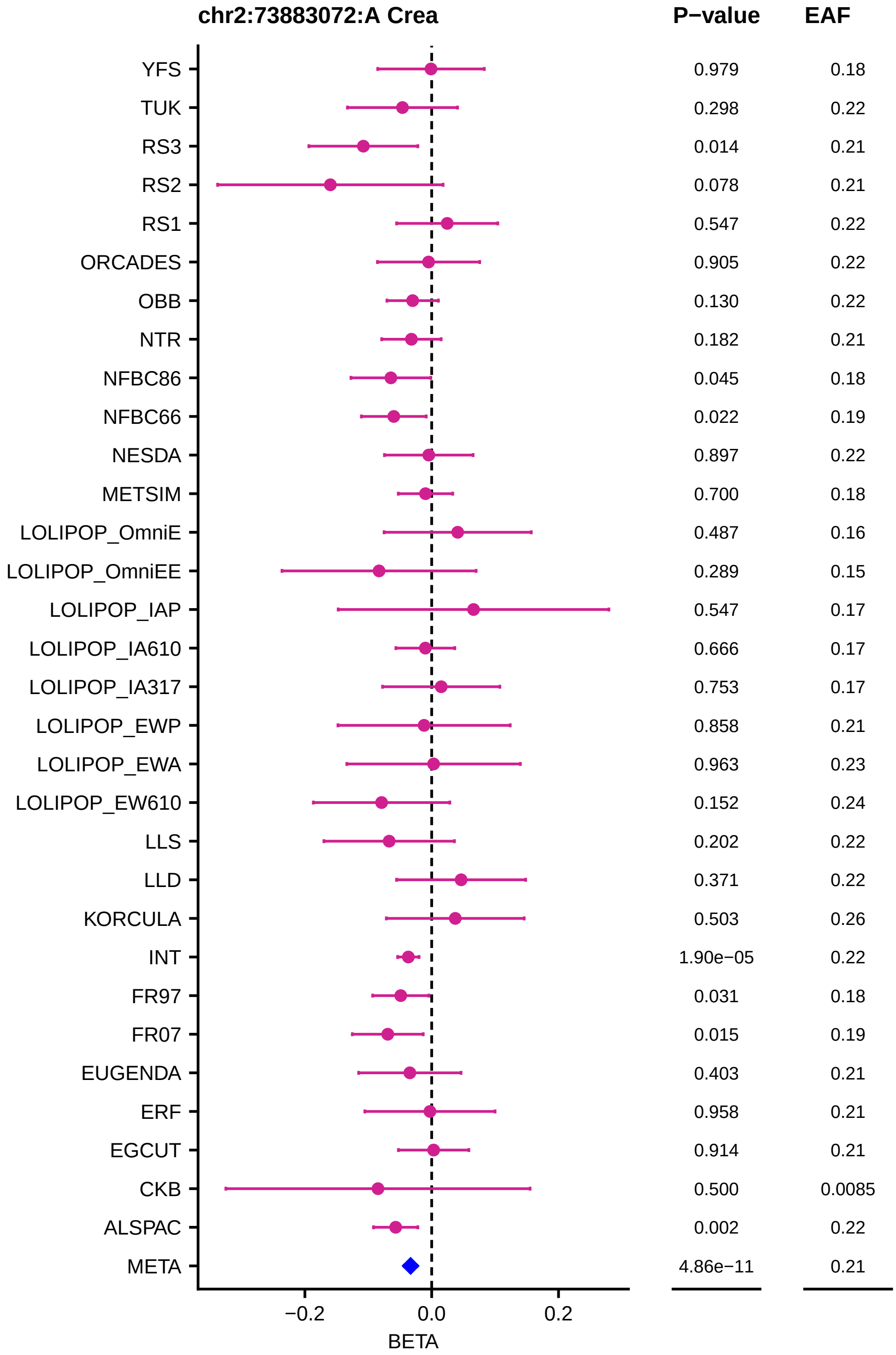
chr2:73883072:A Crea
P−value
EAF
YFS 0.979 0.18
TUK 0.298 0.22
RS3 0.014 0.21
RS2 0.078 0.21
RS1 0.547 0.22
ORCADES 0.905 0.22
OBB 0.130 0.22
NTR 0.182 0.21
NFBC86 0.045 0.18
NFBC66 0.022 0.19
NESDA 0.897 0.22
METSIM 0.700 0.18
LOLIPOP_OmniE 0.487 0.16
LOLIPOP_OmniEE 0.289 0.15
LOLIPOP_IAP 0.547 0.17
LOLIPOP_IA610 0.666 0.17
LOLIPOP_IA317 0.753 0.17
LOLIPOP_EWP 0.858 0.21
LOLIPOP_EWA 0.963 0.23
LOLIPOP_EW610 0.152 0.24
LLS 0.202 0.22
LLD 0.371 0.22
KORCULA 0.503 0.26
INT 1.90e−05 0.22
FR97 0.031 0.18
FR07 0.015 0.19
EUGENDA 0.403 0.21
ERF 0.958 0.21
EGCUT 0.914 0.21
CKB 0.500 0.0085
ALSPAC 0.002 0.22
META 4.86e−11 0.21
−0.2
0.0
0.2
BETA

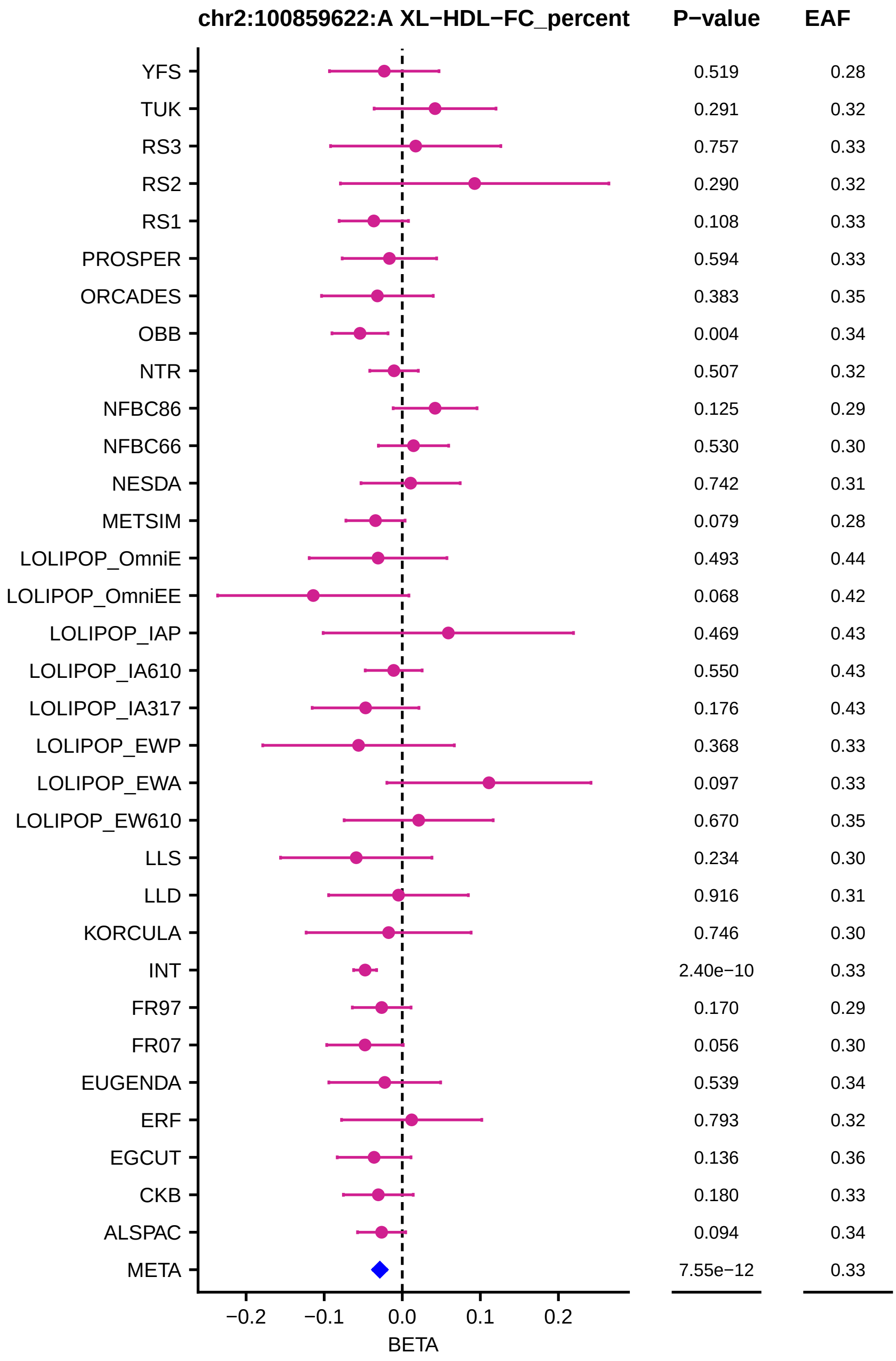
chr2:100859622:A XL−HDL−FC_percent
P−value
EAF
YFS 0.519 0.28
TUK 0.291 0.32
RS3 0.757 0.33
RS2 0.290 0.32
RS1 0.108 0.33
PROSPER 0.594 0.33
ORCADES 0.383 0.35
OBB 0.004 0.34
NTR 0.507 0.32
NFBC86 0.125 0.29
NFBC66 0.530 0.30
NESDA 0.742 0.31
METSIM 0.079 0.28
LOLIPOP_OmniE 0.493 0.44
LOLIPOP_OmniEE 0.068 0.42
LOLIPOP_IAP 0.469 0.43
LOLIPOP_IA610 0.550 0.43
LOLIPOP_IA317 0.176 0.43
LOLIPOP_EWP 0.368 0.33
LOLIPOP_EWA 0.097 0.33
LOLIPOP_EW610 0.670 0.35
LLS 0.234 0.30
LLD 0.916 0.31
KORCULA 0.746 0.30
INT 2.40e−10 0.33
FR97 0.170 0.29
FR07 0.056 0.30
EUGENDA 0.539 0.34
ERF 0.793 0.32
EGCUT 0.136 0.36
CKB 0.180 0.33
ALSPAC 0.094 0.34
META 7.55e−12 0.33
−0.2
−0.1
0.0
0.1
0.2
BETA

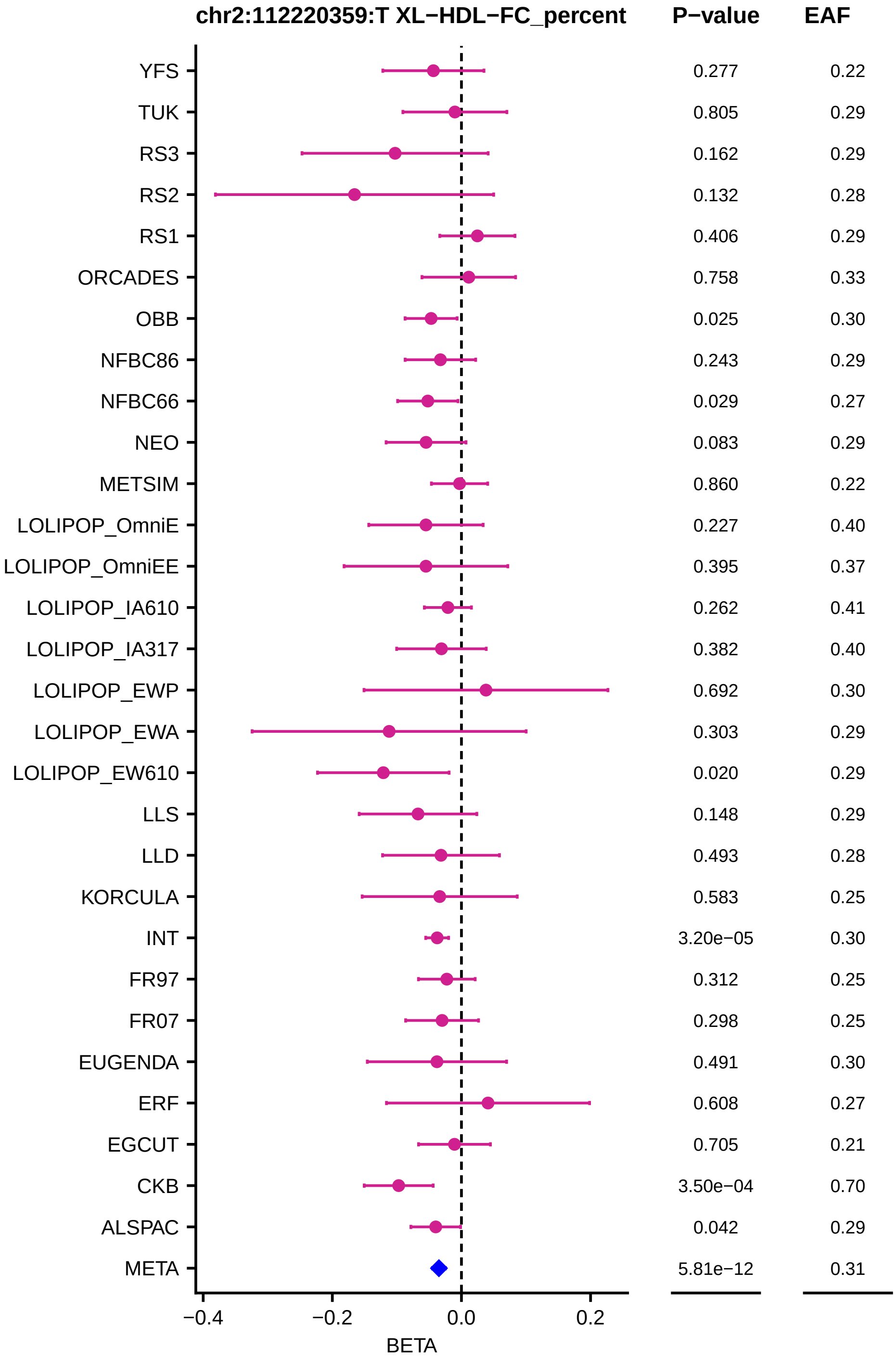

chr2:112220359:T XL−HDL−FC_percent
P−value
EAF
YFS 0.277 0.22
TUK 0.805 0.29
RS3 0.162 0.29
RS2 0.132 0.28
RS1 0.406 0.29
ORCADES 0.758 0.33
OBB 0.025 0.30
NFBC86 0.243 0.29
NFBC66 0.029 0.27
NEO 0.083 0.29
METSIM 0.860 0.22
LOLIPOP_OmniE 0.227 0.40
LOLIPOP_OmniEE 0.395 0.37
LOLIPOP_IA610 0.262 0.41
LOLIPOP_IA317 0.382 0.40
LOLIPOP_EWP 0.692 0.30
LOLIPOP_EWA 0.303 0.29
LOLIPOP_EW610 0.020 0.29
LLS 0.148 0.29
LLD 0.493 0.28
KORCULA 0.583 0.25
INT 3.20e−05 0.30
FR97 0.312 0.25
FR07 0.298 0.25
EUGENDA 0.491 0.30
ERF 0.608 0.27
EGCUT 0.705 0.21
CKB 3.50e−04 0.70
ALSPAC 0.042 0.29
META 5.81e−12 0.31
−0.4
−0.2
0.0
0.2
BETA

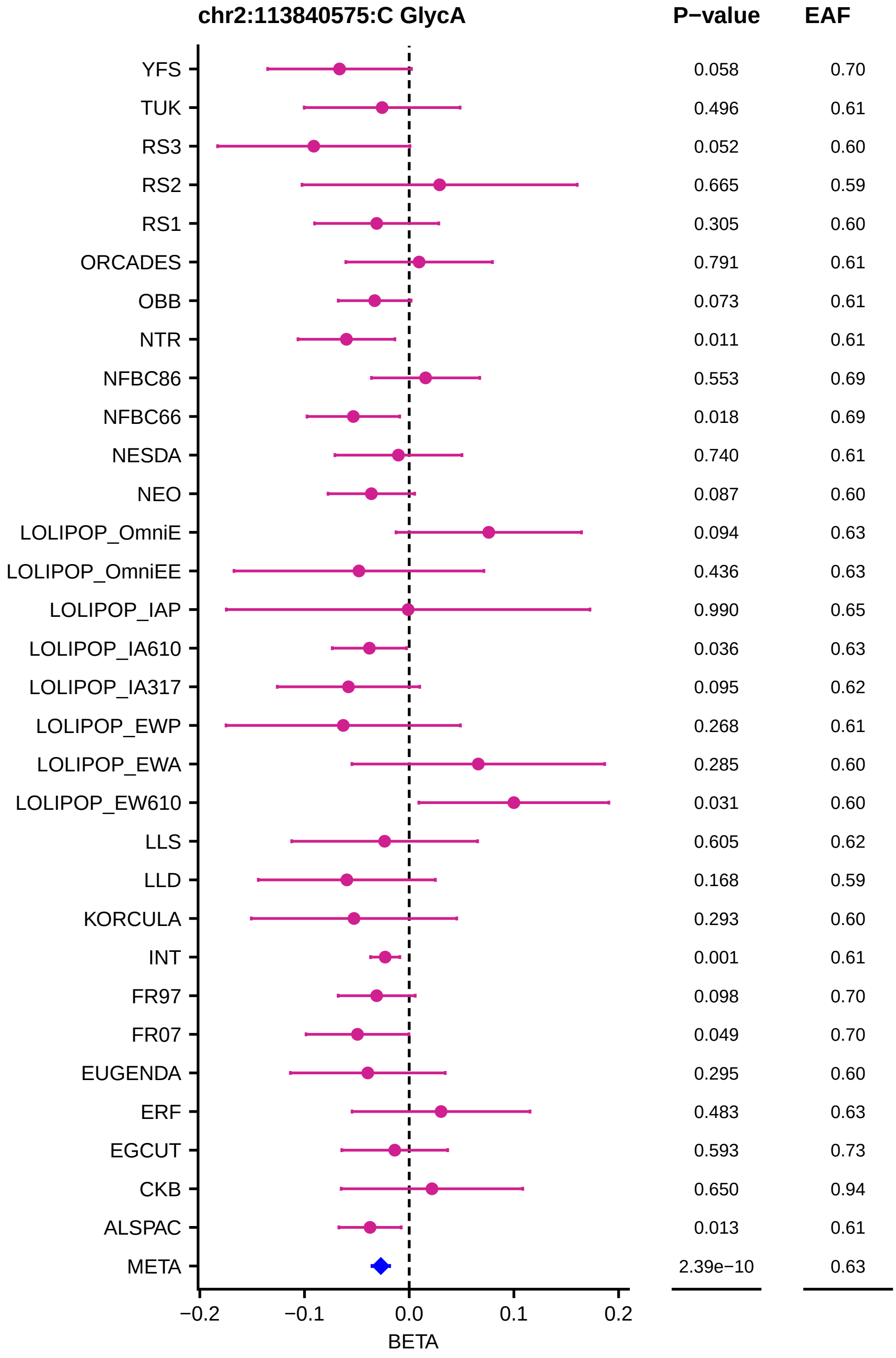
chr2:113840575:C GlycA
P−value
EAF
YFS 0.058 0.70
TUK 0.496 0.61
RS3 0.052 0.60
RS2 0.665 0.59
RS1 0.305 0.60
ORCADES 0.791 0.61
OBB 0.073 0.61
NTR 0.011 0.61
NFBC86 0.553 0.69
NFBC66 0.018 0.69
NESDA 0.740 0.61
NEO 0.087 0.60
LOLIPOP_OmniE 0.094 0.63
LOLIPOP_OmniEE 0.436 0.63
LOLIPOP_IAP 0.990 0.65
LOLIPOP_IA610 0.036 0.63
LOLIPOP_IA317 0.095 0.62
LOLIPOP_EWP 0.268 0.61
LOLIPOP_EWA 0.285 0.60
LOLIPOP_EW610 0.031 0.60
LLS 0.605 0.62
LLD 0.168 0.59
KORCULA 0.293 0.60
INT 0.001 0.61
FR97 0.098 0.70
FR07 0.049 0.70
EUGENDA 0.295 0.60
ERF 0.483 0.63
EGCUT 0.593 0.73
CKB 0.650 0.94
ALSPAC 0.013 0.61
META 2.39e−10 0.63
−0.2
−0.1
0.0
0.1
0.2
BETA

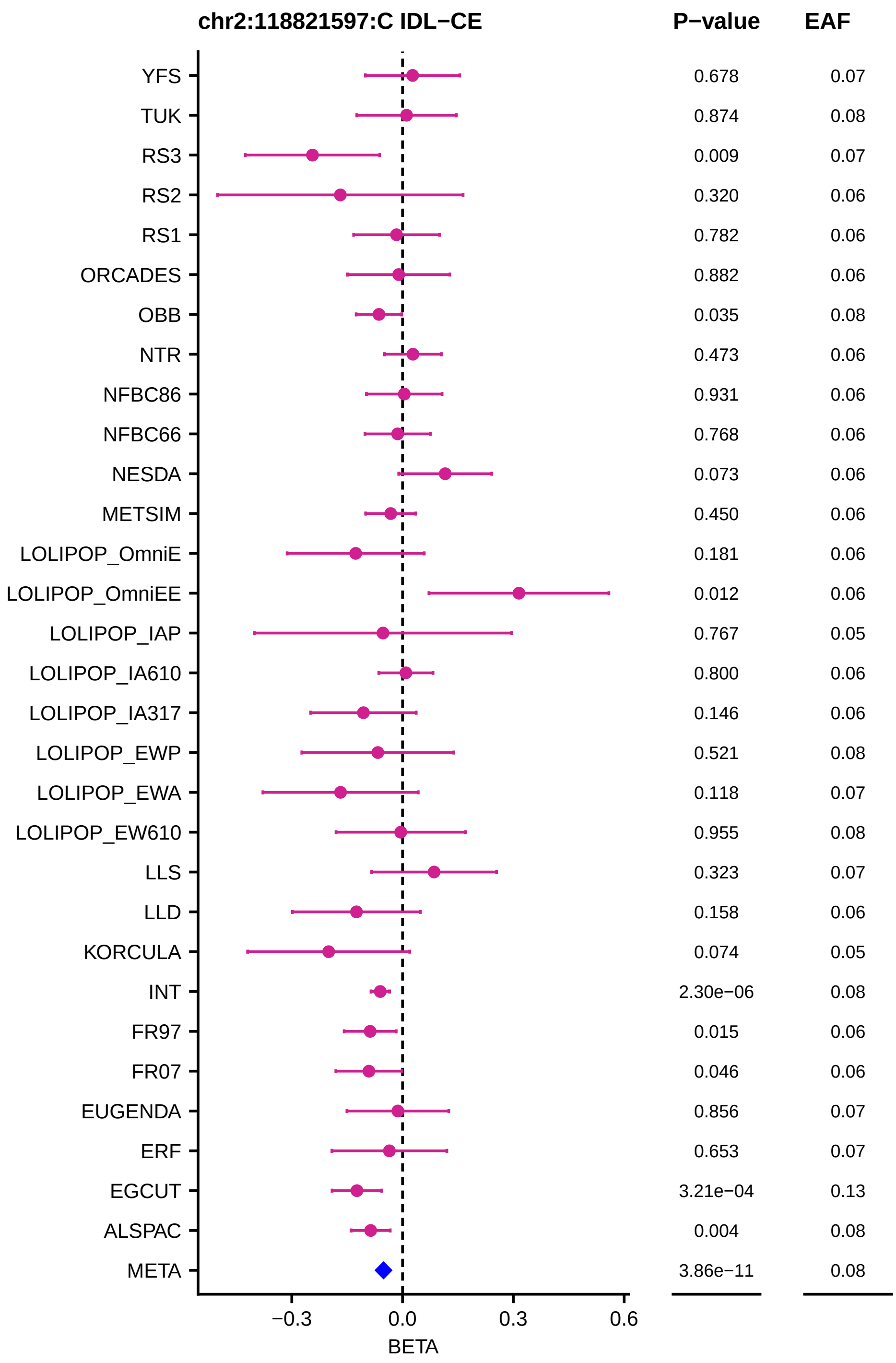
chr2:118821597:C IDL−CE
P−value
EAF
YFS 0.678 0.07
TUK 0.874 0.08
RS3 0.009 0.07
RS2 0.320 0.06
RS1 0.782 0.06
ORCADES 0.882 0.06
OBB 0.035 0.08
NTR 0.473 0.06
NFBC86 0.931 0.06
NFBC66 0.768 0.06
NESDA 0.073 0.06
METSIM 0.450 0.06
LOLIPOP_OmniE 0.181 0.06
LOLIPOP_OmniEE 0.012 0.06
LOLIPOP_IAP 0.767 0.05
LOLIPOP_IA610 0.800 0.06
LOLIPOP_IA317 0.146 0.06
LOLIPOP_EWP 0.521 0.08
LOLIPOP_EWA 0.118 0.07
LOLIPOP_EW610 0.955 0.08
LLS 0.323 0.07
LLD 0.158 0.06
KORCULA 0.074 0.05
INT 2.30e−06 0.08
FR97 0.015 0.06
FR07 0.046 0.06
EUGENDA 0.856 0.07
ERF 0.653 0.07
EGCUT 3.21e−04 0.13
ALSPAC 0.004 0.08
META 3.86e−11 0.08
−0.3
0.0
0.3
0.6
BETA

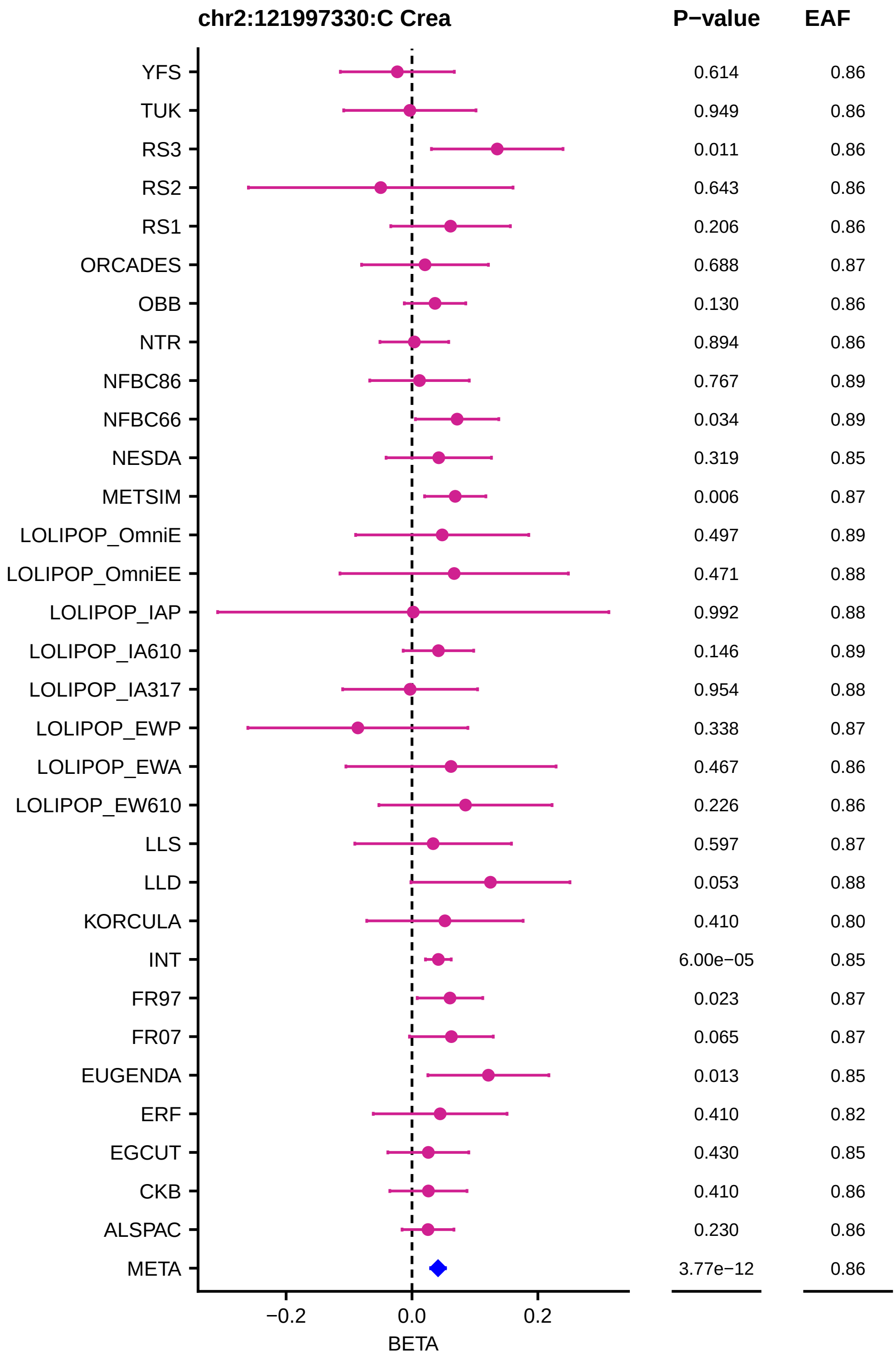

chr2:121997330:C Crea
P−value
EAF
YFS
TUK
RS3
RS2
RS1
ORCADES
OBB
NTR
NFBC86
NFBC66
NESDA
METSIM
LOLIPOP_OmniE
LOLIPOP_OmniEE
LOLIPOP_IAP
LOLIPOP_IA610
LOLIPOP_IA317
LOLIPOP_EWP
LOLIPOP_EWA
LOLIPOP_EW610
LLS
LLD
KORCULA
INT
FR97
FR07
EUGENDA
ERF
EGCUT
CKB
ALSPAC
META
0.614 0.86
0.949 0.86
0.011 0.86
0.643 0.86
0.206 0.86
0.688 0.87
0.130 0.86
0.894 0.86
0.767 0.89
0.034 0.89
0.319 0.85
0.006 0.87
0.497 0.89
0.471 0.88
0.992 0.88
0.146 0.89
0.954 0.88
0.338 0.87
0.467 0.86
0.226 0.86
0.597 0.87
0.053 0.88
0.410 0.80
6.00e−05 0.85
0.023 0.87
0.065 0.87
0.013 0.85
0.410 0.82
0.430 0.85
0.410 0.86
0.230 0.86
3.77e−12 0.86
−0.2
0.0
0.2
BETA

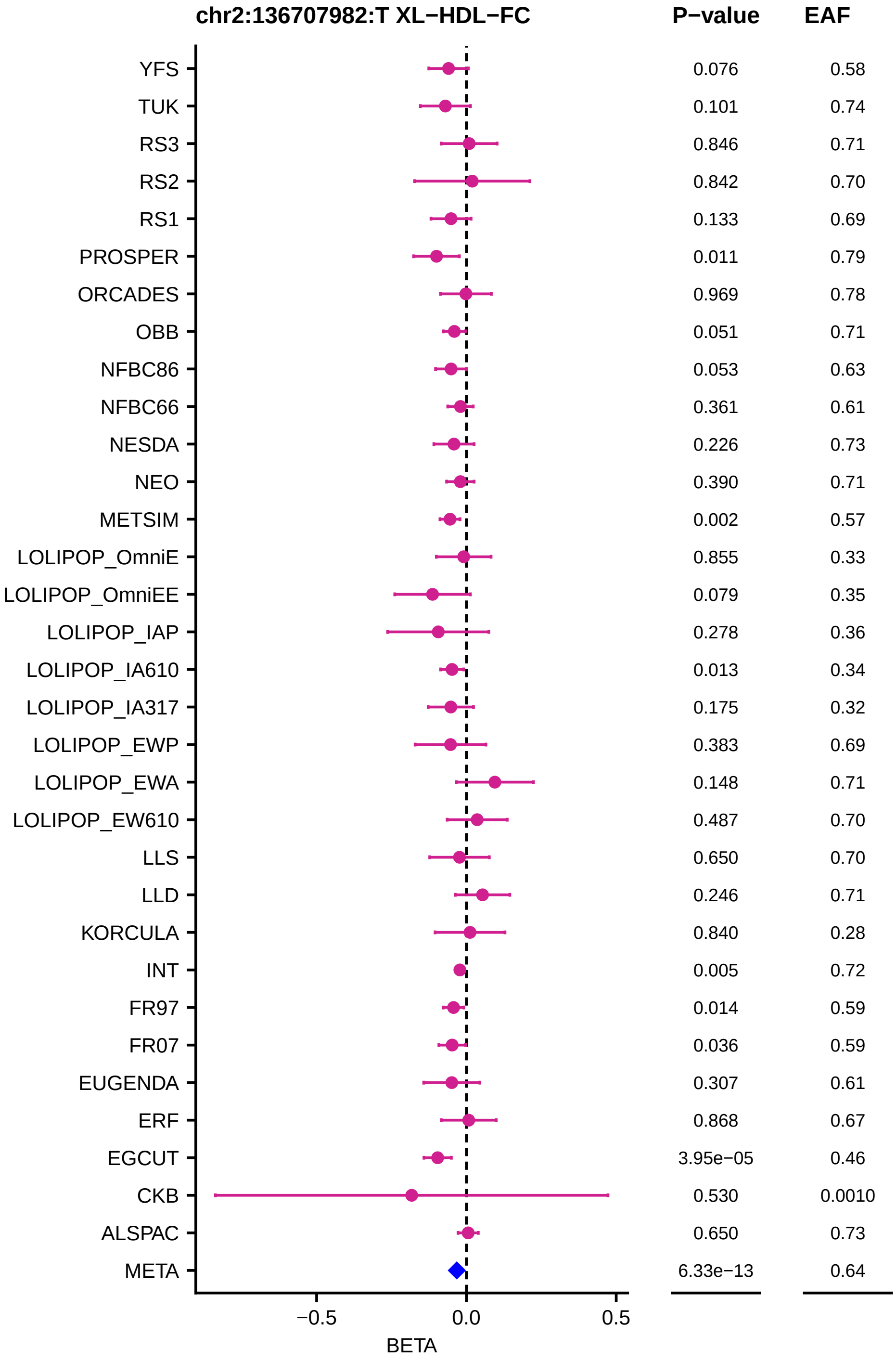
chr2:136707982:T XL−HDL−FC
P−value
EAF
YFS 0.076 0.58
TUK 0.101 0.74
RS3 0.846 0.71
RS2 0.842 0.70
RS1 0.133 0.69
PROSPER 0.011 0.79
ORCADES 0.969 0.78
OBB 0.051 0.71
NFBC86 0.053 0.63
NFBC66 0.361 0.61
NESDA 0.226 0.73
NEO 0.390 0.71
METSIM 0.002 0.57
LOLIPOP_OmniE 0.855 0.33
LOLIPOP_OmniEE 0.079 0.35
LOLIPOP_IAP 0.278 0.36
LOLIPOP_IA610 0.013 0.34
LOLIPOP_IA317 0.175 0.32
LOLIPOP_EWP 0.383 0.69
LOLIPOP_EWA 0.148 0.71
LOLIPOP_EW610 0.487 0.70
LLS 0.650 0.70
LLD 0.246 0.71
KORCULA 0.840 0.28
INT 0.005 0.72
FR97 0.014 0.59
FR07 0.036 0.59
EUGENDA 0.307 0.61
ERF 0.868 0.67
EGCUT 3.95e−05 0.46
CKB 0.530 0.0010
ALSPAC 0.650 0.73
META 6.33e−13 0.64
−0.5
0.0
0.5
BETA

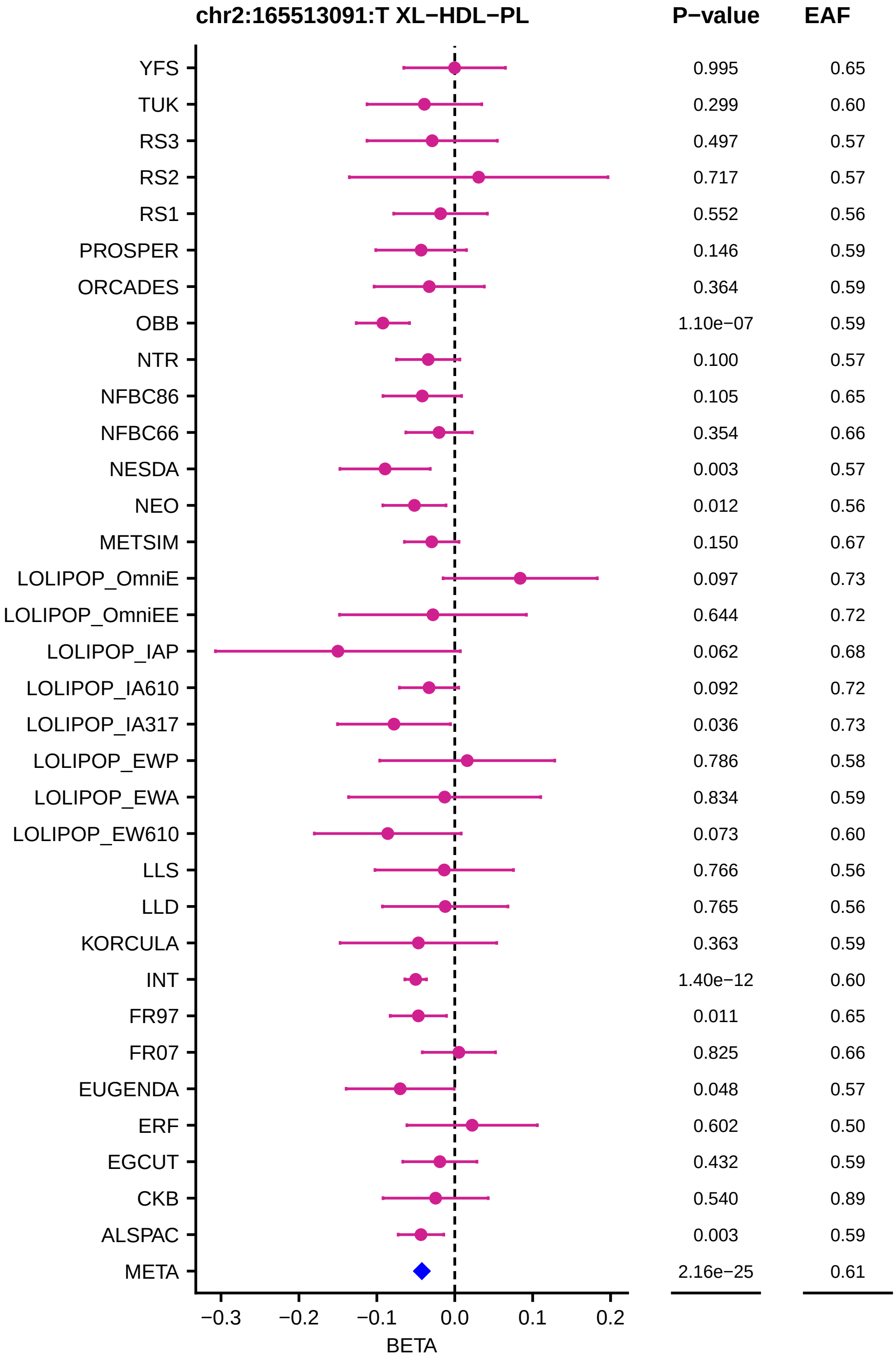
chr2:165513091:T XL−HDL−PL
P−value
EAF
YFS 0.995 0.65
TUK 0.299 0.60
RS3 0.497 0.57
RS2 0.717 0.57
RS1 0.552 0.56
PROSPER 0.146 0.59
ORCADES 0.364 0.59
OBB 1.10e−07 0.59
NTR 0.100 0.57
NFBC86 0.105 0.65
NFBC66 0.354 0.66
NESDA 0.003 0.57
NEO 0.012 0.56
METSIM 0.150 0.67
LOLIPOP_OmniE 0.097 0.73
LOLIPOP_OmniEE 0.644 0.72
LOLIPOP_IAP 0.062 0.68
LOLIPOP_IA610 0.092 0.72
LOLIPOP_IA317 0.036 0.73
LOLIPOP_EWP 0.786 0.58
LOLIPOP_EWA 0.834 0.59
LOLIPOP_EW610 0.073 0.60
LLS 0.766 0.56
LLD 0.765 0.56
KORCULA 0.363 0.59
INT 1.40e−12 0.60
FR97 0.011 0.65
FR07 0.825 0.66
EUGENDA 0.048 0.57
ERF 0.602 0.50
EGCUT 0.432 0.59
CKB 0.540 0.89
ALSPAC 0.003 0.59
META 2.16e−25 0.61
−0.3
−0.2
−0.1
0.0
0.1
0.2
BETA

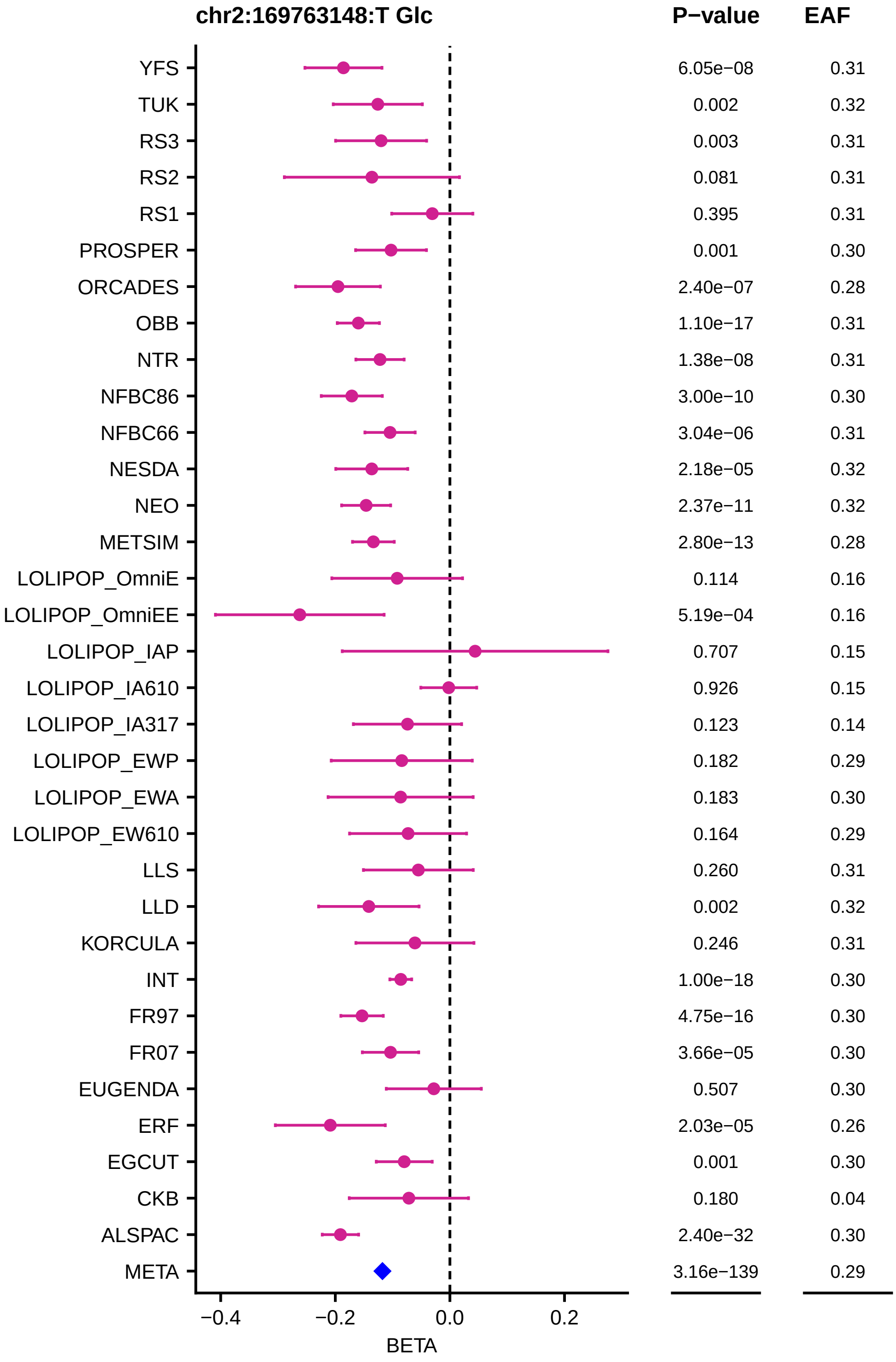
chr2:169763148:T Glc
P−value
EAF
YFS
TUK
RS3
RS2
RS1
PROSPER
ORCADES
OBB
NTR
NFBC86
NFBC66
NESDA
NEO
METSIM
LOLIPOP_OmniE
LOLIPOP_OmniEE
LOLIPOP_IAP
LOLIPOP_IA610
LOLIPOP_IA317
LOLIPOP_EWP
LOLIPOP_EWA
LOLIPOP_EW610
LLS
LLD
KORCULA
INT
FR97
FR07
EUGENDA
ERF
EGCUT
CKB
ALSPAC
META
6.05e−08
0.002
0.003
0.081
0.395
0.001
2.40e−07
1.10e−17
1.38e−08
3.00e−10
3.04e−06
2.18e−05
2.37e−11
2.80e−13
0.114
5.19e−04
0.707
0.926
0.123
0.182
0.183
0.164
0.260
0.002
0.246
1.00e−18
4.75e−16
3.66e−05
0.507
2.03e−05
0.001
0.180
2.40e−32
3.16e−139
0.31
0.32
0.31
0.31
0.31
0.30
0.28
0.31
0.31
0.30
0.31
0.32
0.32
0.28
0.16
0.16
0.15
0.15
0.14
0.29
0.30
0.29
0.31
0.32
0.31
0.30
0.30
0.30
0.30
0.26
0.30
0.04
0.30
0.29
−0.4
−0.2
0.0
0.2
BETA

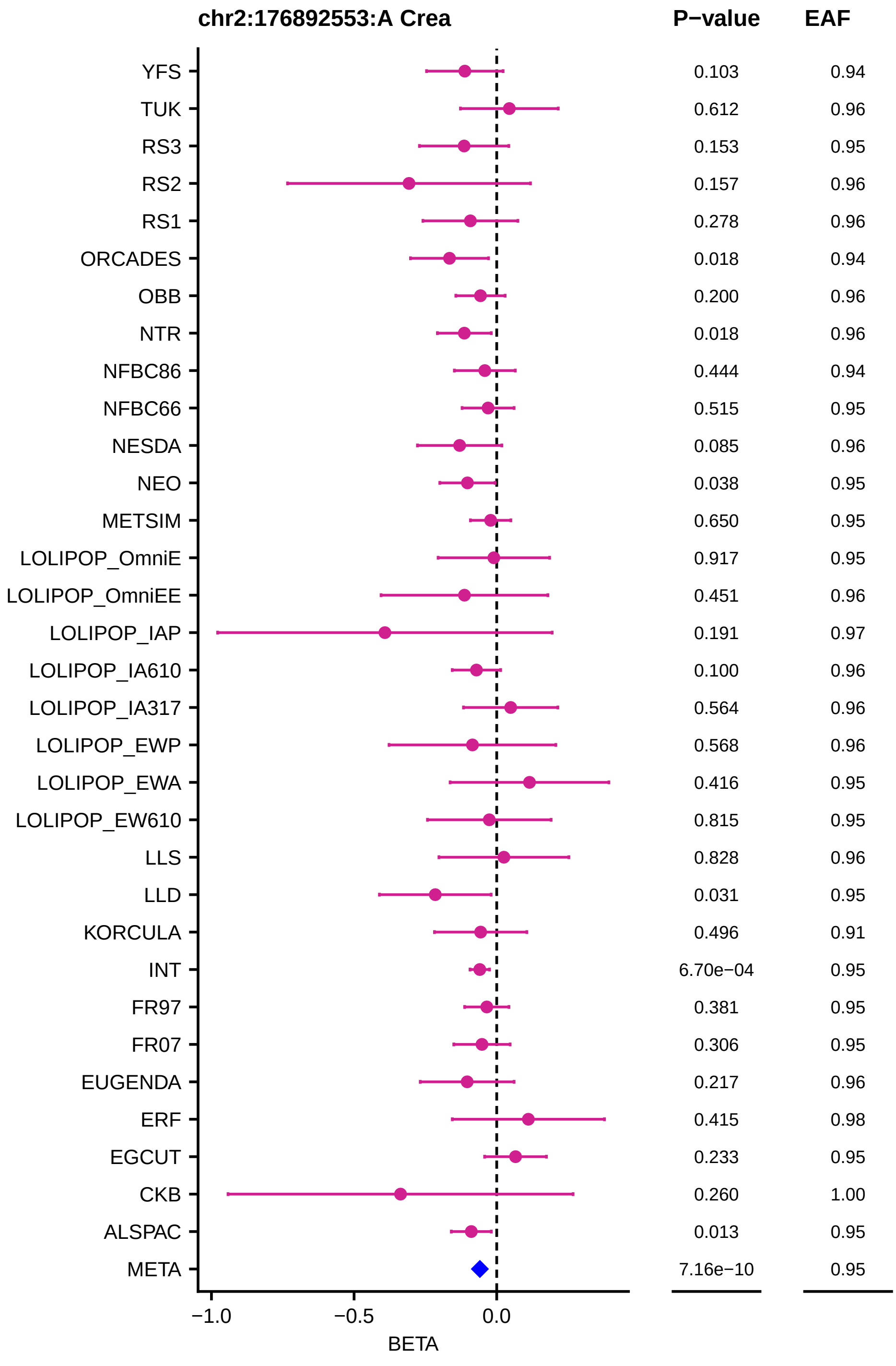
chr2:176892553:A Crea
P−value
EAF
YFS 0.103 0.94
TUK 0.612 0.96
RS3 0.153 0.95
RS2 0.157 0.96
RS1 0.278 0.96
ORCADES 0.018 0.94
OBB 0.200 0.96
NTR 0.018 0.96
NFBC86 0.444 0.94
NFBC66 0.515 0.95
NESDA 0.085 0.96
NEO 0.038 0.95
METSIM 0.650 0.95
LOLIPOP_OmniE 0.917 0.95
LOLIPOP_OmniEE 0.451 0.96
LOLIPOP_IAP 0.191 0.97
LOLIPOP_IA610 0.100 0.96
LOLIPOP_IA317 0.564 0.96
LOLIPOP_EWP 0.568 0.96
LOLIPOP_EWA 0.416 0.95
LOLIPOP_EW610 0.815 0.95
LLS 0.828 0.96
LLD 0.031 0.95
KORCULA 0.496 0.91
INT 6.70e−04 0.95
FR97 0.381 0.95
FR07 0.306 0.95
EUGENDA 0.217 0.96
ERF 0.415 0.98
EGCUT 0.233 0.95
CKB 0.260 1.00
ALSPAC 0.013 0.95
META 7.16e−10 0.95
−1.0
−0.5
0.0
BETA

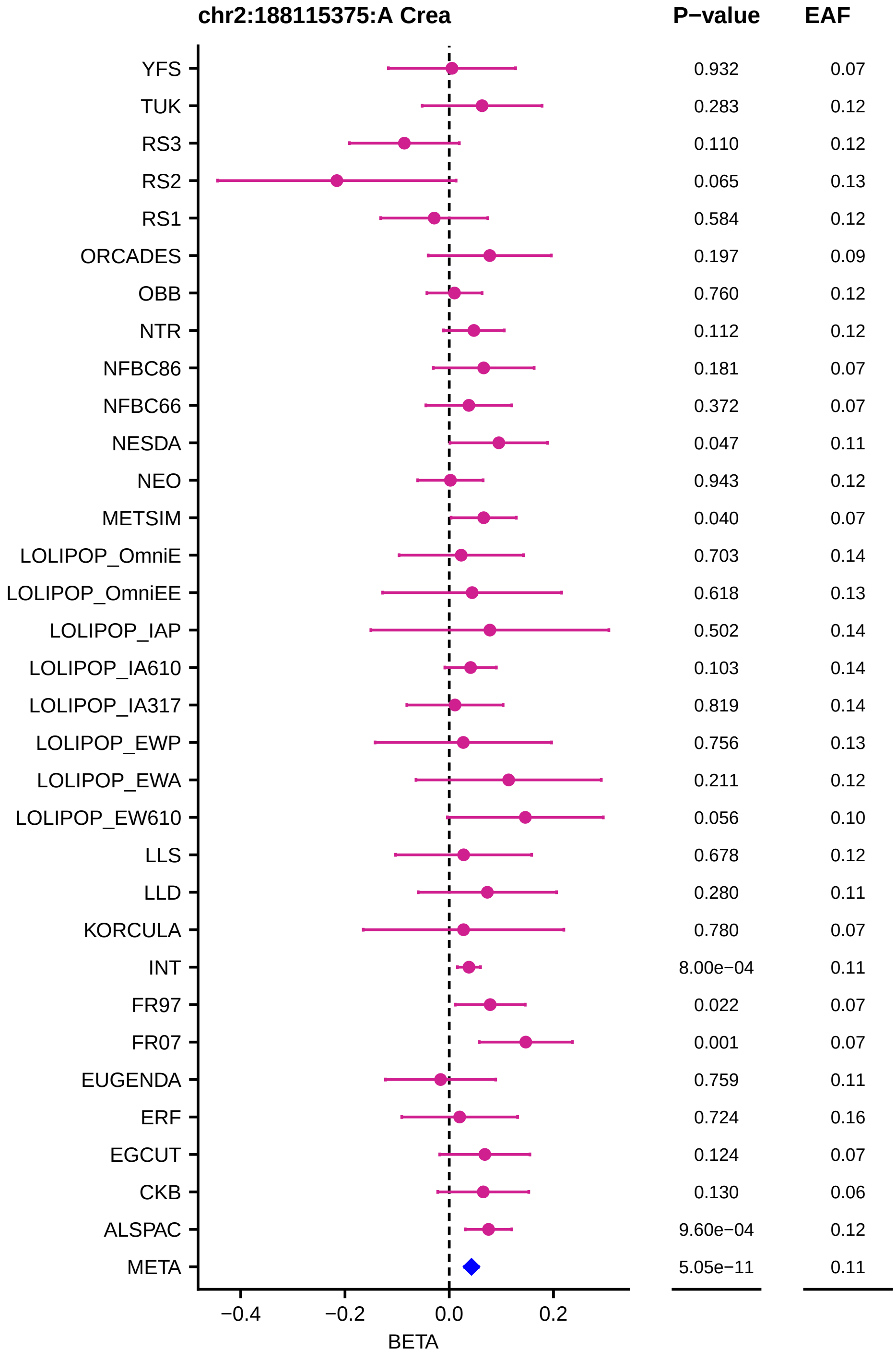
chr2:188115375:A Crea
P−value
EAF
YFS 0.932 0.07
TUK 0.283 0.12
RS3 0.110 0.12
RS2 0.065 0.13
RS1 0.584 0.12
ORCADES 0.197 0.09
OBB 0.760 0.12
NTR 0.112 0.12
NFBC86 0.181 0.07
NFBC66 0.372 0.07
NESDA 0.047 0.11
NEO 0.943 0.12
METSIM 0.040 0.07
LOLIPOP_OmniE 0.703 0.14
LOLIPOP_OmniEE 0.618 0.13
LOLIPOP_IAP 0.502 0.14
LOLIPOP_IA610 0.103 0.14
LOLIPOP_IA317 0.819 0.14
LOLIPOP_EWP 0.756 0.13
LOLIPOP_EWA 0.211 0.12
LOLIPOP_EW610 0.056 0.10
LLS 0.678 0.12
LLD 0.280 0.11
KORCULA 0.780 0.07
INT 8.00e−04 0.11
FR97 0.022 0.07
FR07 0.001 0.07
EUGENDA 0.759 0.11
ERF 0.724 0.16
EGCUT 0.124 0.07
CKB 0.130 0.06
ALSPAC 9.60e−04 0.12
META 5.05e−11 0.11
−0.4
−0.2
0.0
0.2
BETA

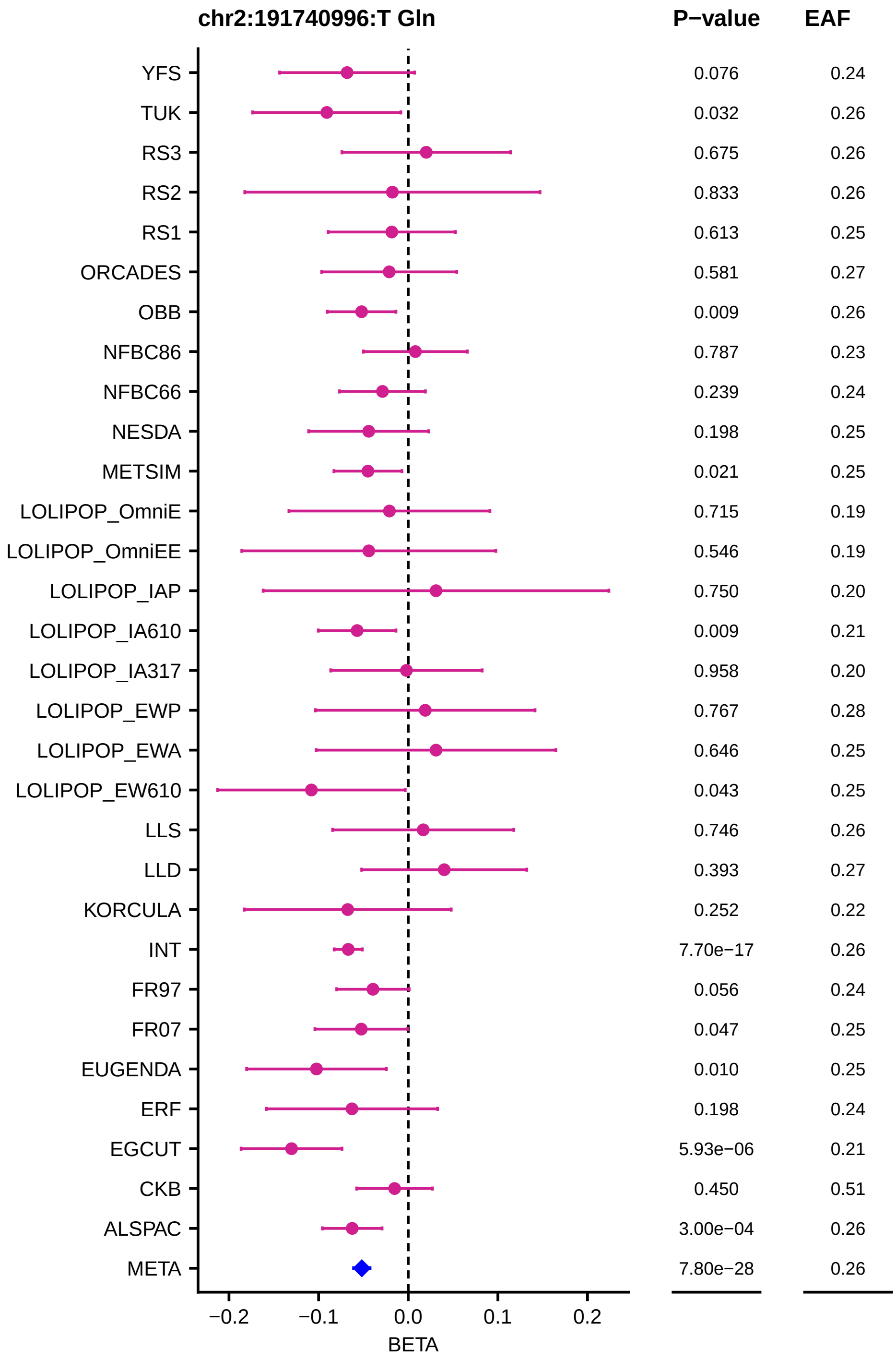

chr2:191740996:T Gln
P−value
EAF
YFS 0.076 0.24
TUK 0.032 0.26
RS3 0.675 0.26
RS2 0.833 0.26
RS1 0.613 0.25
ORCADES 0.581 0.27
OBB 0.009 0.26
NFBC86 0.787 0.23
NFBC66 0.239 0.24
NESDA 0.198 0.25
METSIM 0.021 0.25
LOLIPOP_OmniE 0.715 0.19
LOLIPOP_OmniEE 0.546 0.19
LOLIPOP_IAP 0.750 0.20
LOLIPOP_IA610 0.009 0.21
LOLIPOP_IA317 0.958 0.20
LOLIPOP_EWP 0.767 0.28
LOLIPOP_EWA 0.646 0.25
LOLIPOP_EW610 0.043 0.25
LLS 0.746 0.26
LLD 0.393 0.27
KORCULA 0.252 0.22
INT 7.70e−17 0.26
FR97 0.056 0.24
FR07 0.047 0.25
EUGENDA 0.010 0.25
ERF 0.198 0.24
EGCUT 5.93e−06 0.21
CKB 0.450 0.51
ALSPAC 3.00e−04 0.26
META 7.80e−28 0.26
−0.2
−0.1
0.0
0.1
0.2
BETA

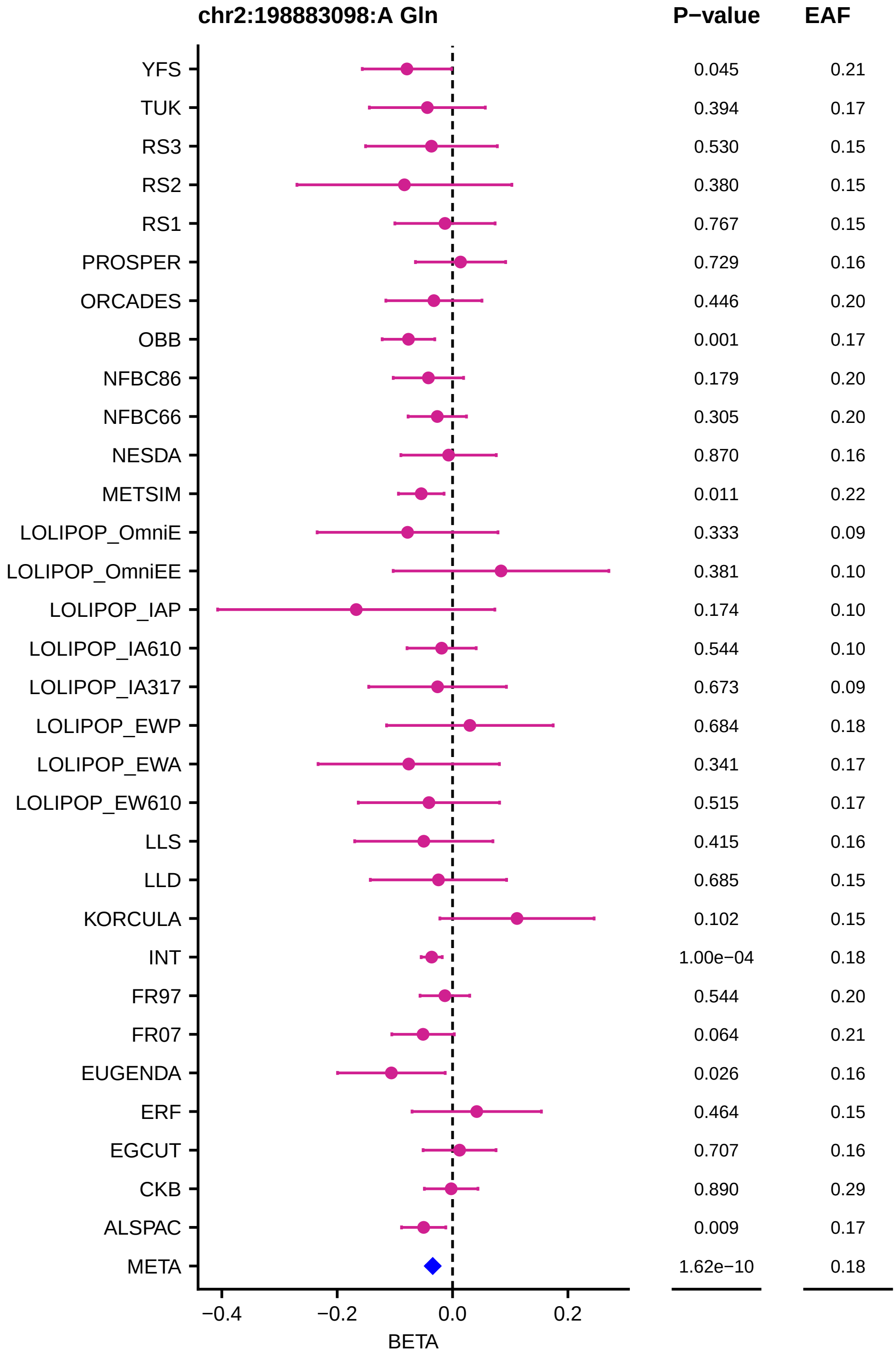

chr2:198883098:A Gln
P−value
EAF
YFS 0.045 0.21
TUK 0.394 0.17
RS3 0.530 0.15
RS2 0.380 0.15
RS1 0.767 0.15
PROSPER 0.729 0.16
ORCADES 0.446 0.20
OBB 0.001 0.17
NFBC86 0.179 0.20
NFBC66 0.305 0.20
NESDA 0.870 0.16
METSIM 0.011 0.22
LOLIPOP_OmniE 0.333 0.09
LOLIPOP_OmniEE 0.381 0.10
LOLIPOP_IAP 0.174 0.10
LOLIPOP_IA610 0.544 0.10
LOLIPOP_IA317 0.673 0.09
LOLIPOP_EWP 0.684 0.18
LOLIPOP_EWA 0.341 0.17
LOLIPOP_EW610 0.515 0.17
LLS 0.415 0.16
LLD 0.685 0.15
KORCULA 0.102 0.15
INT 1.00e−04 0.18
FR97 0.544 0.20
FR07 0.064 0.21
EUGENDA 0.026 0.16
ERF 0.464 0.15
EGCUT 0.707 0.16
CKB 0.890 0.29
ALSPAC 0.009 0.17
META 1.62e−10 0.18
−0.4
−0.2
0.0
0.2
BETA

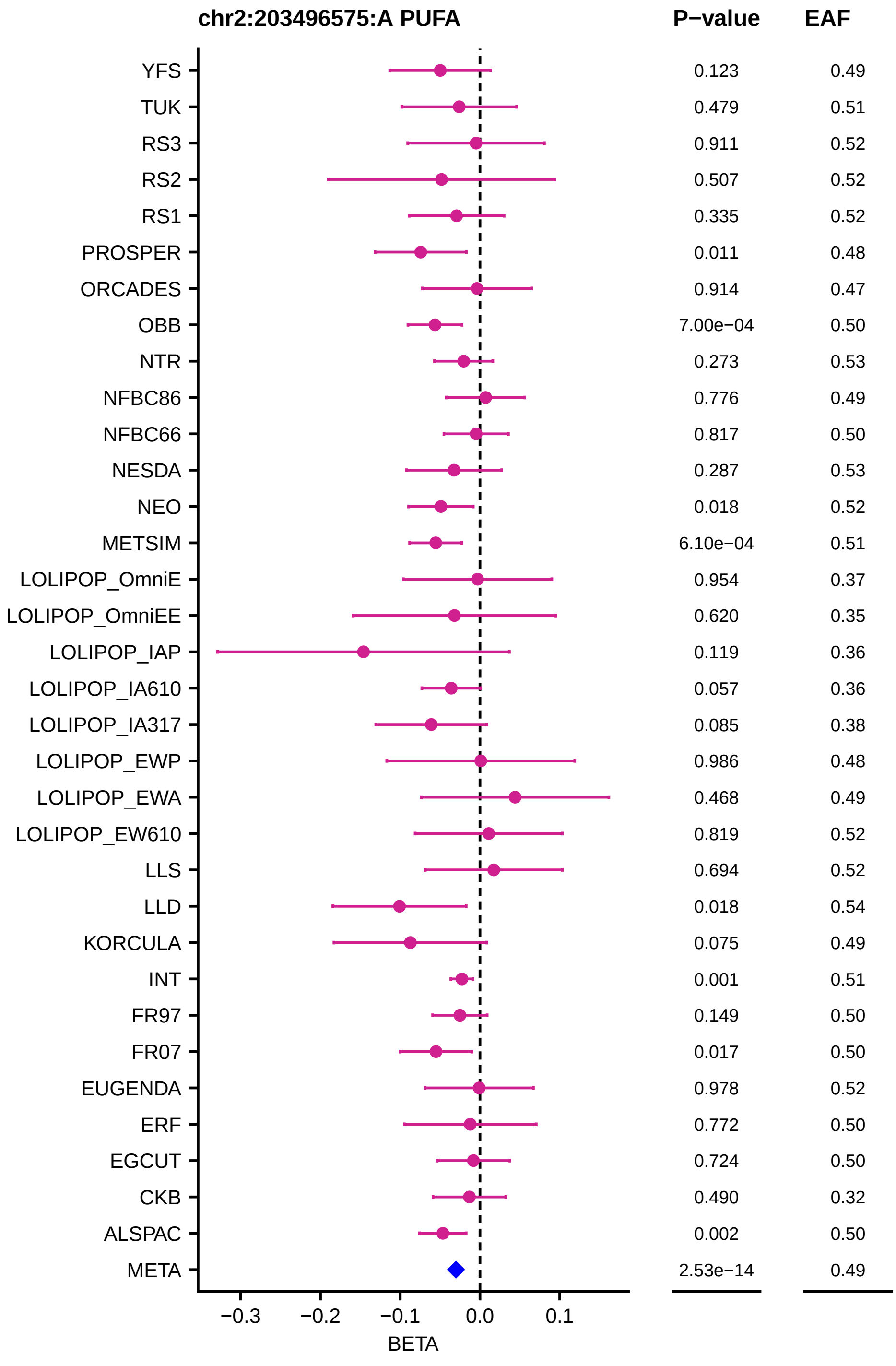
chr2:203496575:A PUFA
P−value
EAF
YFS 0.123 0.49
TUK 0.479 0.51
RS3 0.911 0.52
RS2 0.507 0.52
RS1 0.335 0.52
PROSPER 0.011 0.48
ORCADES 0.914 0.47
OBB 7.00e−04 0.50
NTR 0.273 0.53
NFBC86 0.776 0.49
NFBC66 0.817 0.50
NESDA 0.287 0.53
NEO 0.018 0.52
METSIM 6.10e−04 0.51
LOLIPOP_OmniE 0.954 0.37
LOLIPOP_OmniEE 0.620 0.35
LOLIPOP_IAP 0.119 0.36
LOLIPOP_IA610 0.057 0.36
LOLIPOP_IA317 0.085 0.38
LOLIPOP_EWP 0.986 0.48
LOLIPOP_EWA 0.468 0.49
LOLIPOP_EW610 0.819 0.52
LLS 0.694 0.52
LLD 0.018 0.54
KORCULA 0.075 0.49
INT 0.001 0.51
FR97 0.149 0.50
FR07 0.017 0.50
EUGENDA 0.978 0.52
ERF 0.772 0.50
EGCUT 0.724 0.50
CKB 0.490 0.32
ALSPAC 0.002 0.50
META 2.53e−14 0.49
−0.3
−0.2
−0.1
0.0
0.1
BETA

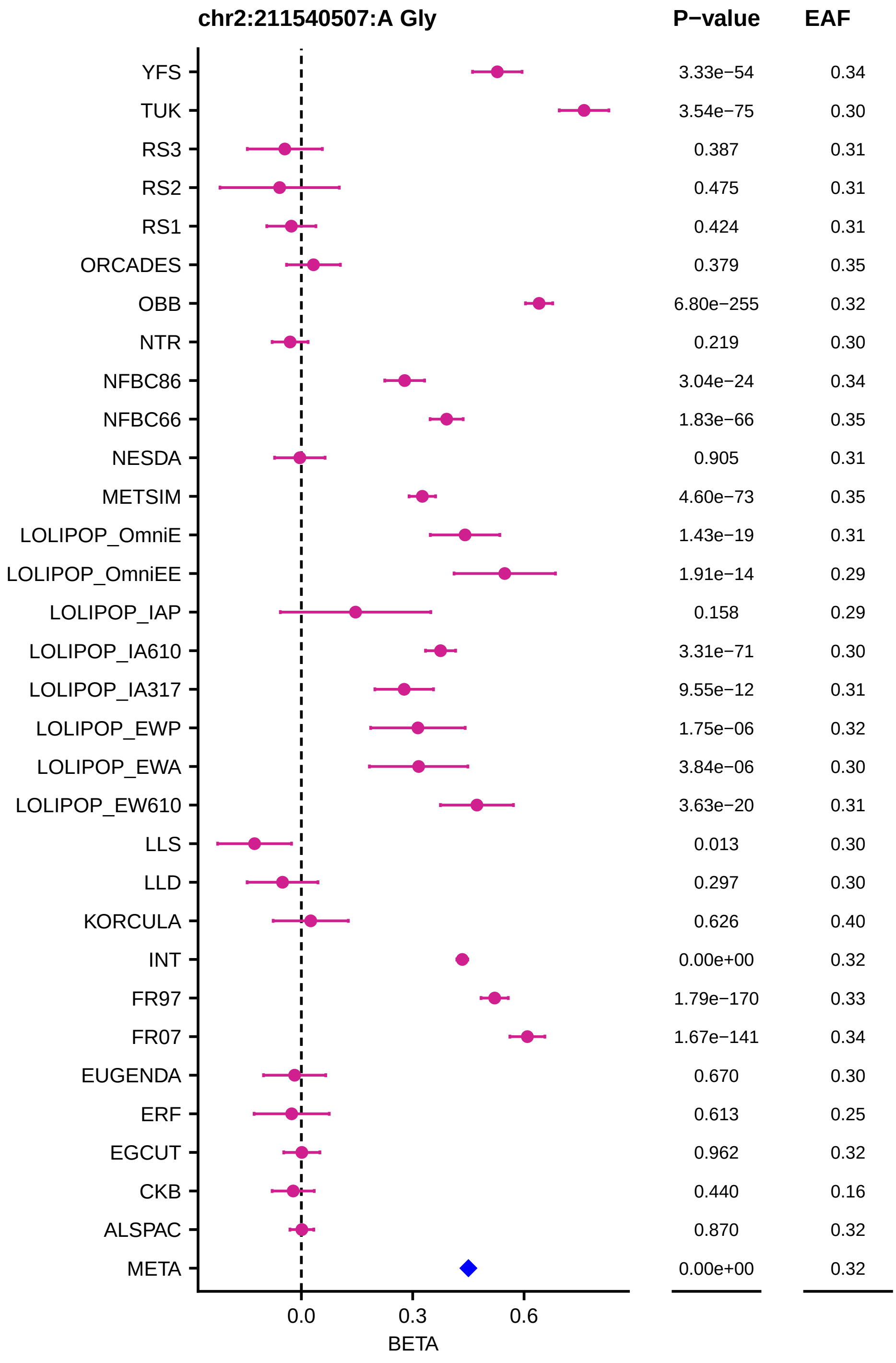
chr2:211540507:A Gly
P−value
EAF
YFS
TUK
RS3
RS2
RS1
ORCADES
OBB
NTR
NFBC86
NFBC66
NESDA
METSIM
LOLIPOP_OmniE
LOLIPOP_OmniEE
LOLIPOP_IAP
LOLIPOP_IA610
LOLIPOP_IA317
LOLIPOP_EWP
LOLIPOP_EWA
LOLIPOP_EW610
LLS
LLD
KORCULA
INT
FR97
FR07
EUGENDA
ERF
EGCUT
CKB
ALSPAC
META
3.33e−54 0.34
3.54e−75 0.30
0.387 0.31
0.475 0.31
0.424 0.31
0.379 0.35
6.80e−255 0.32
0.219 0.30
3.04e−24 0.34
1.83e−66 0.35
0.905 0.31
4.60e−73 0.35
1.43e−19 0.31
1.91e−14 0.29
0.158 0.29
3.31e−71 0.30
9.55e−12 0.31
1.75e−06 0.32
3.84e−06 0.30
3.63e−20 0.31
0.013 0.30
0.297 0.30
0.626 0.40
0.00e+00 0.32
1.79e−170 0.33
1.67e−141 0.34
0.670 0.30
0.613 0.25
0.962 0.32
0.440 0.16
0.870 0.32
0.00e+00 0.32
0.0
0.3
0.6
BETA

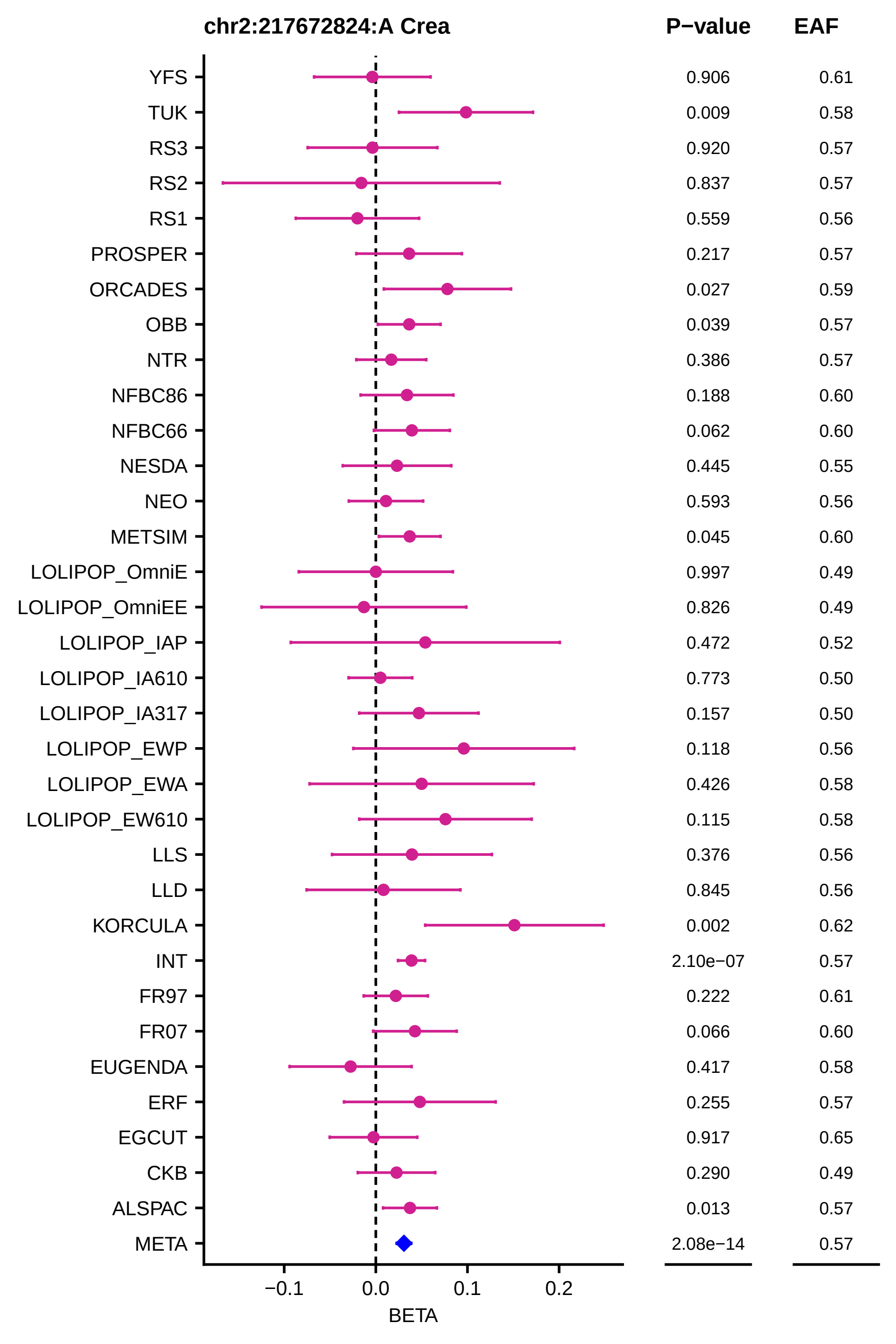
chr2:217672824:A Crea
P−value
EAF
YFS 0.906 0.61
TUK 0.009 0.58
RS3 0.920 0.57
RS2 0.837 0.57
RS1 0.559 0.56
PROSPER 0.217 0.57
ORCADES 0.027 0.59
OBB 0.039 0.57
NTR 0.386 0.57
NFBC86 0.188 0.60
NFBC66 0.062 0.60
NESDA 0.445 0.55
NEO 0.593 0.56
METSIM 0.045 0.60
LOLIPOP_OmniE 0.997 0.49
LOLIPOP_OmniEE 0.826 0.49
LOLIPOP_IAP 0.472 0.52
LOLIPOP_IA610 0.773 0.50
LOLIPOP_IA317 0.157 0.50
LOLIPOP_EWP 0.118 0.56
LOLIPOP_EWA 0.426 0.58
LOLIPOP_EW610 0.115 0.58
LLS 0.376 0.56
LLD 0.845 0.56
KORCULA 0.002 0.62
INT 2.10e−07 0.57
FR97 0.222 0.61
FR07 0.066 0.60
EUGENDA 0.417 0.58
ERF 0.255 0.57
EGCUT 0.917 0.65
CKB 0.290 0.49
ALSPAC 0.013 0.57
META 2.08e−14 0.57
−0.1
0.0
0.1
0.2
BETA

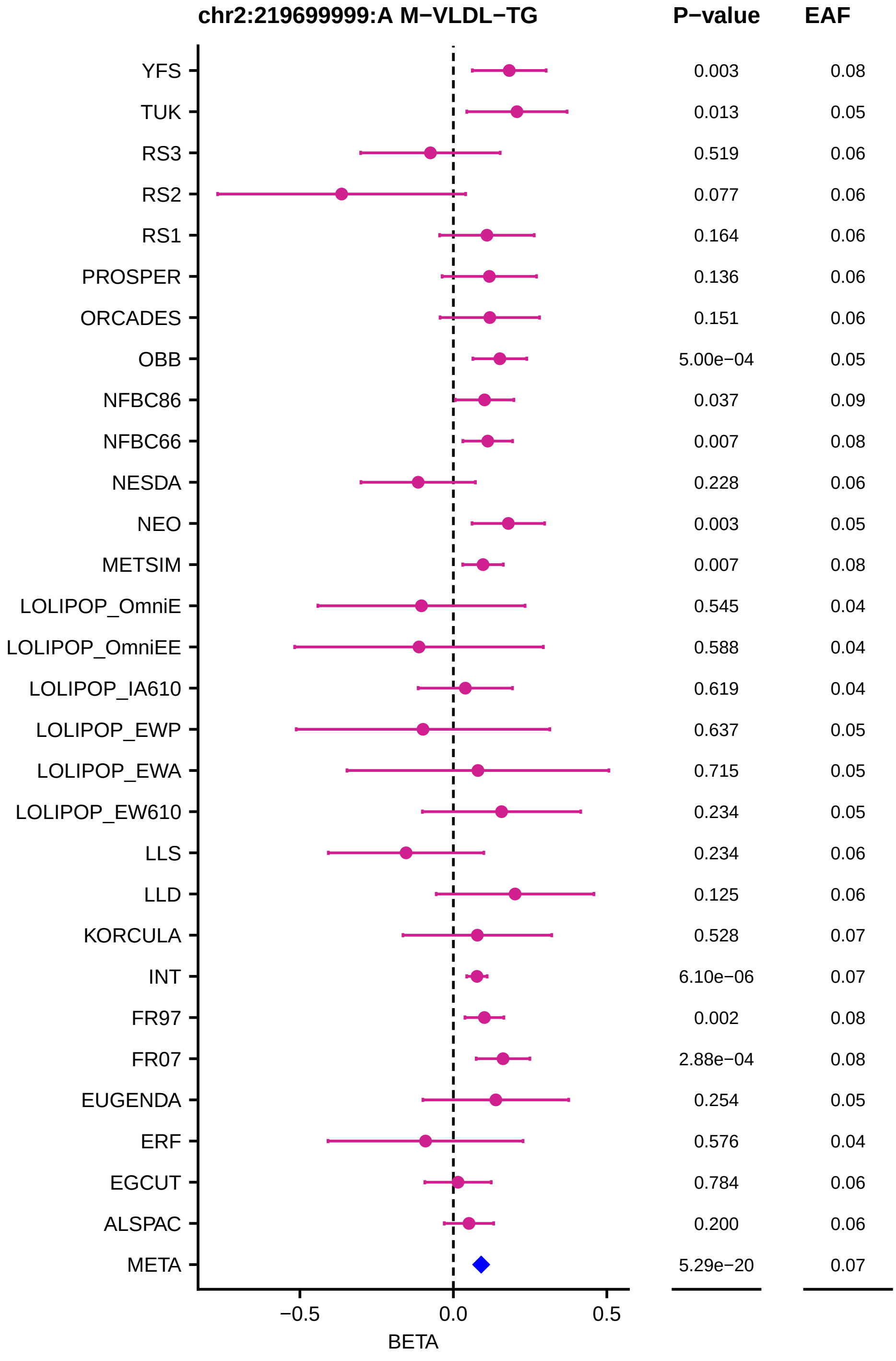
chr2:219699999:A M−VLDL−TG
P−value
EAF
YFS 0.003 0.08
TUK 0.013 0.05
RS3 0.519 0.06
RS2 0.077 0.06
RS1 0.164 0.06
PROSPER 0.136 0.06
ORCADES 0.151 0.06
OBB 5.00e−04 0.05
NFBC86 0.037 0.09
NFBC66 0.007 0.08
NESDA 0.228 0.06
NEO 0.003 0.05
METSIM 0.007 0.08
LOLIPOP_OmniE 0.545 0.04
LOLIPOP_OmniEE 0.588 0.04
LOLIPOP_IA610 0.619 0.04
LOLIPOP_EWP 0.637 0.05
LOLIPOP_EWA 0.715 0.05
LOLIPOP_EW610 0.234 0.05
LLS 0.234 0.06
LLD 0.125 0.06
KORCULA 0.528 0.07
INT 6.10e−06 0.07
FR97 0.002 0.08
FR07 2.88e−04 0.08
EUGENDA 0.254 0.05
ERF 0.576 0.04
EGCUT 0.784 0.06
ALSPAC 0.200 0.06
META 5.29e−20 0.07
−0.5
0.0
0.5
BETA

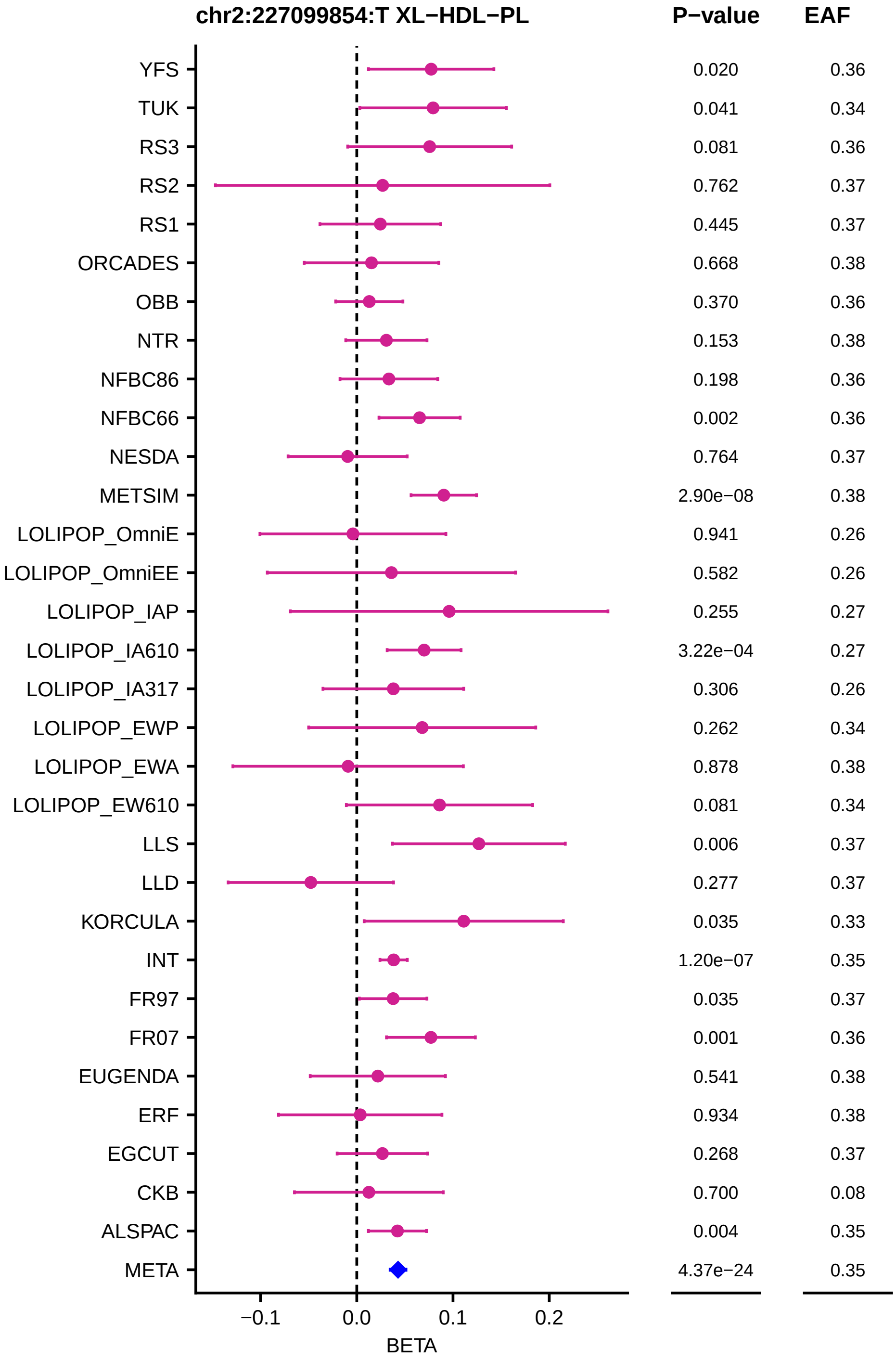
chr2:227099854:T XL−HDL−PL
P−value
EAF
YFS 0.020 0.36
TUK 0.041 0.34
RS3 0.081 0.36
RS2 0.762 0.37
RS1 0.445 0.37
ORCADES 0.668 0.38
OBB 0.370 0.36
NTR 0.153 0.38
NFBC86 0.198 0.36
NFBC66 0.002 0.36
NESDA 0.764 0.37
METSIM 2.90e−08 0.38
LOLIPOP_OmniE 0.941 0.26
LOLIPOP_OmniEE 0.582 0.26
LOLIPOP_IAP 0.255 0.27
LOLIPOP_IA610 3.22e−04 0.27
LOLIPOP_IA317 0.306 0.26
LOLIPOP_EWP 0.262 0.34
LOLIPOP_EWA 0.878 0.38
LOLIPOP_EW610 0.081 0.34
LLS 0.006 0.37
LLD 0.277 0.37
KORCULA 0.035 0.33
INT 1.20e−07 0.35
FR97 0.035 0.37
FR07 0.001 0.36
EUGENDA 0.541 0.38
ERF 0.934 0.38
EGCUT 0.268 0.37
CKB 0.700 0.08
ALSPAC 0.004 0.35
META 4.37e−24 0.35
−0.1
0.0
0.1
0.2
BETA

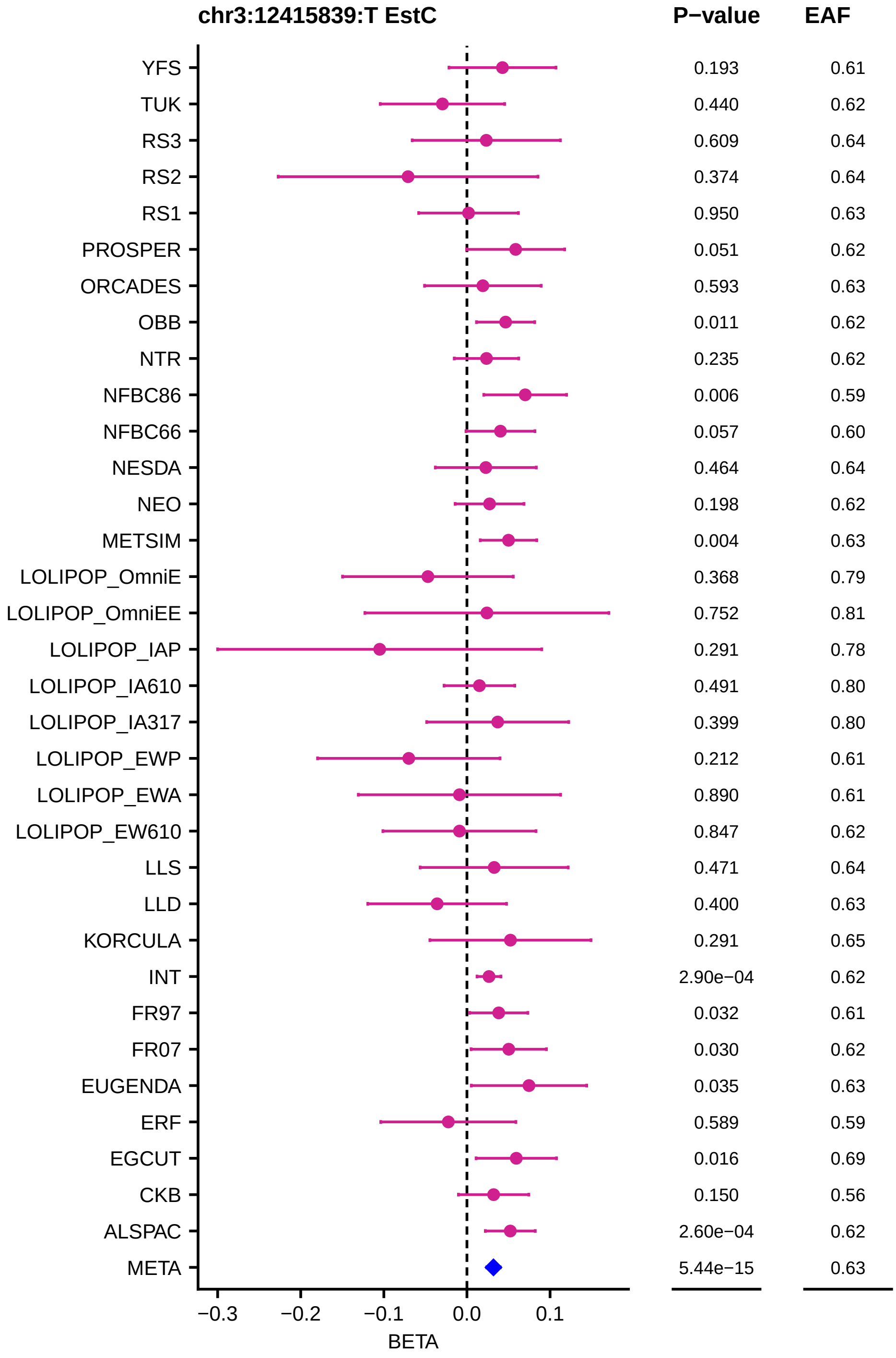
chr3:12415839:T EstC
P−value
EAF
YFS 0.193 0.61
TUK 0.440 0.62
RS3 0.609 0.64
RS2 0.374 0.64
RS1 0.950 0.63
PROSPER 0.051 0.62
ORCADES 0.593 0.63
OBB 0.011 0.62
NTR 0.235 0.62
NFBC86 0.006 0.59
NFBC66 0.057 0.60
NESDA 0.464 0.64
NEO 0.198 0.62
METSIM 0.004 0.63
LOLIPOP_OmniE 0.368 0.79
LOLIPOP_OmniEE 0.752 0.81
LOLIPOP_IAP 0.291 0.78
LOLIPOP_IA610 0.491 0.80
LOLIPOP_IA317 0.399 0.80
LOLIPOP_EWP 0.212 0.61
LOLIPOP_EWA 0.890 0.61
LOLIPOP_EW610 0.847 0.62
LLS 0.471 0.64
LLD 0.400 0.63
KORCULA 0.291 0.65
INT 2.90e−04 0.62
FR97 0.032 0.61
FR07 0.030 0.62
EUGENDA 0.035 0.63
ERF 0.589 0.59
EGCUT 0.016 0.69
CKB 0.150 0.56
ALSPAC 2.60e−04 0.62
META 5.44e−15 0.63
−0.3 −0.2 −0.1 0.0 0.1
BETA

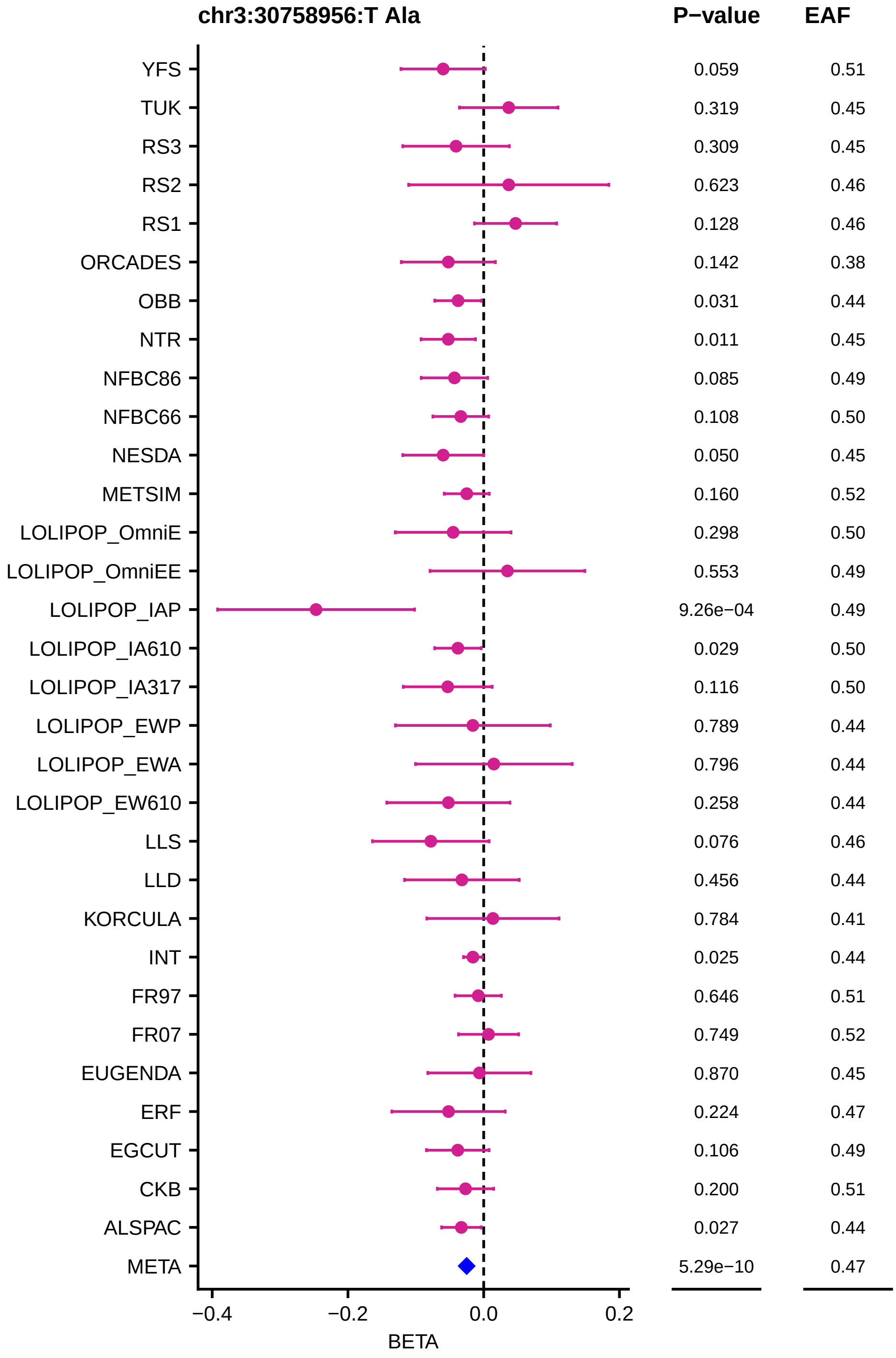
chr3:30758956:T Ala
P−value
EAF
YFS 0.059 0.51
TUK 0.319 0.45
RS3 0.309 0.45
RS2 0.623 0.46
RS1 0.128 0.46
ORCADES 0.142 0.38
OBB 0.031 0.44
NTR 0.011 0.45
NFBC86 0.085 0.49
NFBC66 0.108 0.50
NESDA 0.050 0.45
METSIM 0.160 0.52
LOLIPOP_OmniE 0.298 0.50
LOLIPOP_OmniEE 0.553 0.49
LOLIPOP_IAP 9.26e−04 0.49
LOLIPOP_IA610 0.029 0.50
LOLIPOP_IA317 0.116 0.50
LOLIPOP_EWP 0.789 0.44
LOLIPOP_EWA 0.796 0.44
LOLIPOP_EW610 0.258 0.44
LLS 0.076 0.46
LLD 0.456 0.44
KORCULA 0.784 0.41
INT 0.025 0.44
FR97 0.646 0.51
FR07 0.749 0.52
EUGENDA 0.870 0.45
ERF 0.224 0.47
EGCUT 0.106 0.49
CKB 0.200 0.51
ALSPAC 0.027 0.44
META 5.29e−10 0.47
−0.4
−0.2
0.0
0.2
BETA

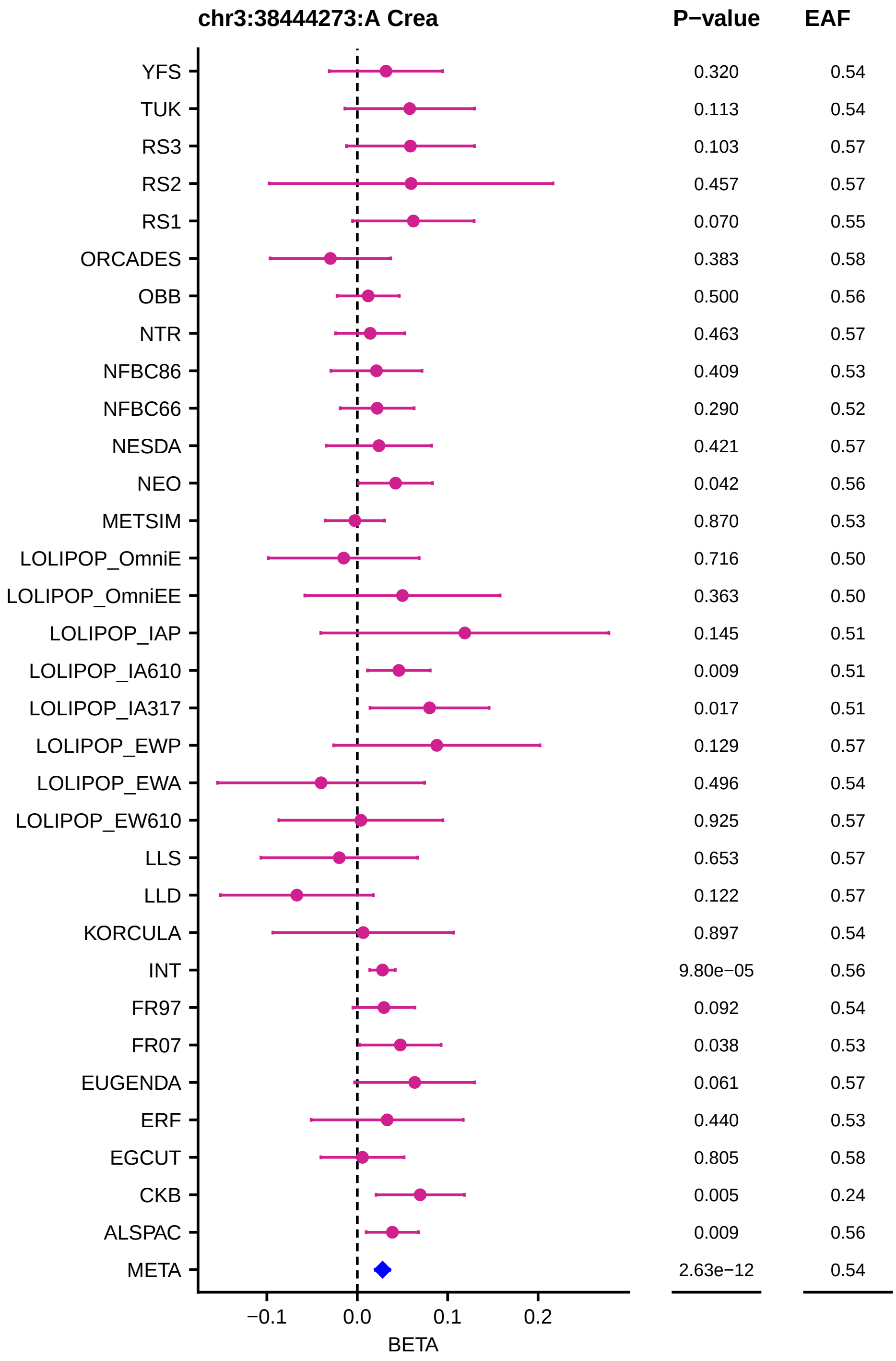

chr3:38444273:A Crea
P−value
EAF
YFS 0.320 0.54
TUK 0.113 0.54
RS3 0.103 0.57
RS2 0.457 0.57
RS1 0.070 0.55
ORCADES 0.383 0.58
OBB 0.500 0.56
NTR 0.463 0.57
NFBC86 0.409 0.53
NFBC66 0.290 0.52
NESDA 0.421 0.57
NEO 0.042 0.56
METSIM 0.870 0.53
LOLIPOP_OmniE 0.716 0.50
LOLIPOP_OmniEE 0.363 0.50
LOLIPOP_IAP 0.145 0.51
LOLIPOP_IA610 0.009 0.51
LOLIPOP_IA317 0.017 0.51
LOLIPOP_EWP 0.129 0.57
LOLIPOP_EWA 0.496 0.54
LOLIPOP_EW610 0.925 0.57
LLS 0.653 0.57
LLD 0.122 0.57
KORCULA 0.897 0.54
INT 9.80e−05 0.56
FR97 0.092 0.54
FR07 0.038 0.53
EUGENDA 0.061 0.57
ERF 0.440 0.53
EGCUT 0.805 0.58
CKB 0.005 0.24
ALSPAC 0.009 0.56
META 2.63e−12 0.54
−0.1
0.0
0.1
0.2
BETA

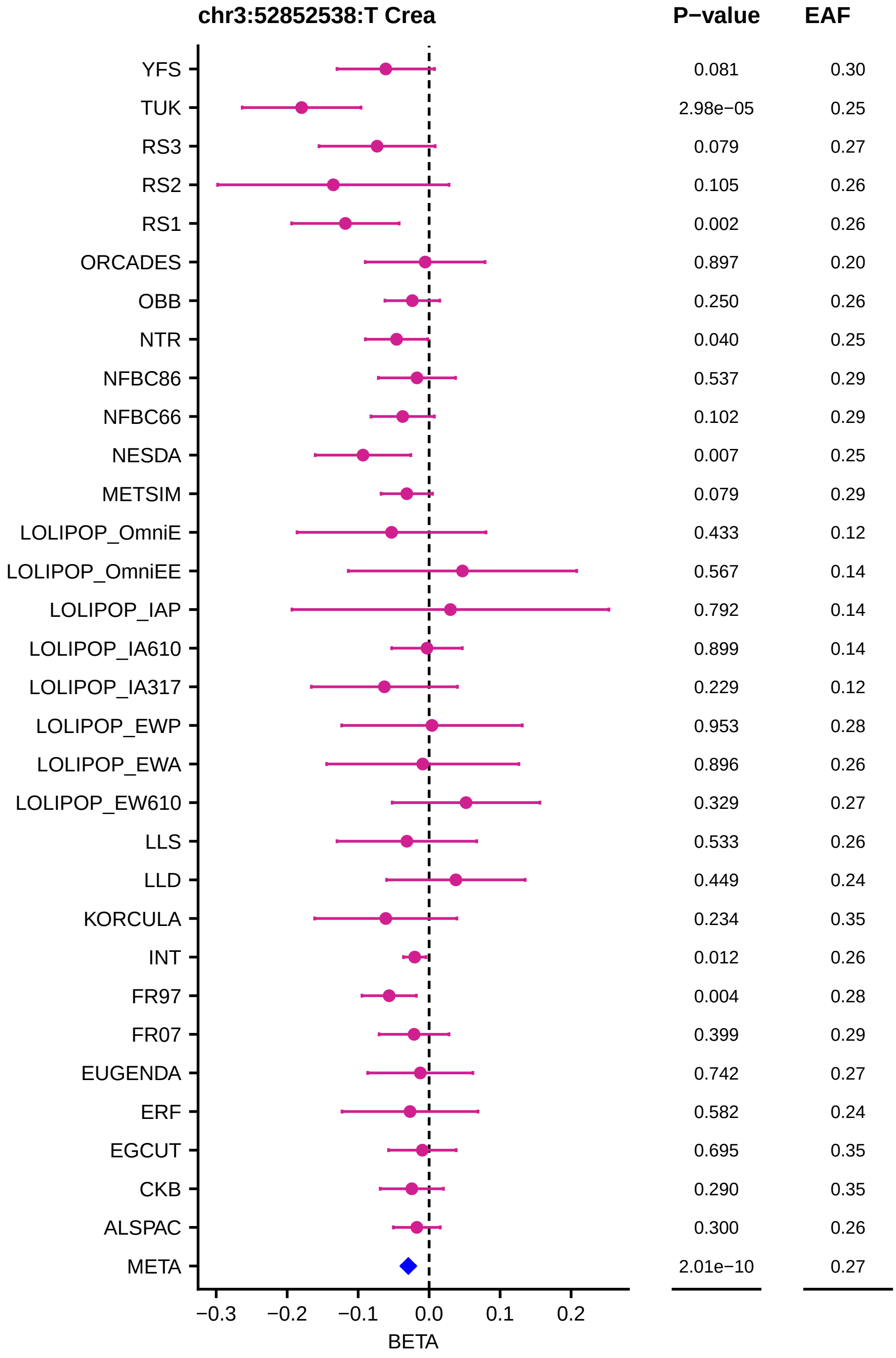

chr3:52852538:T Crea
P−value
EAF
YFS
TUK
RS3
RS2
RS1
ORCADES
OBB
NTR
NFBC86
NFBC66
NESDA
METSIM
LOLIPOP_OmniE
LOLIPOP_OmniEE
LOLIPOP_IAP
LOLIPOP_IA610
LOLIPOP_IA317
LOLIPOP_EWP
LOLIPOP_EWA
LOLIPOP_EW610
LLS
LLD
KORCULA
INT
FR97
FR07
EUGENDA
ERF
EGCUT
CKB
ALSPAC
META
0.081 0.30
2.98e−05 0.25
0.079 0.27
0.105 0.26
0.002 0.26
0.897 0.20
0.250 0.26
0.040 0.25
0.537 0.29
0.102 0.29
0.007 0.25
0.079 0.29
0.433 0.12
0.567 0.14
0.792 0.14
0.899 0.14
0.229 0.12
0.953 0.28
0.896 0.26
0.329 0.27
0.533 0.26
0.449 0.24
0.234 0.35
0.012 0.26
0.004 0.28
0.399 0.29
0.742 0.27
0.582 0.24
0.695 0.35
0.290 0.35
0.300 0.26
2.01e−10 0.27
−0.3
−0.2
−0.1
0.0
0.1
0.2
BETA

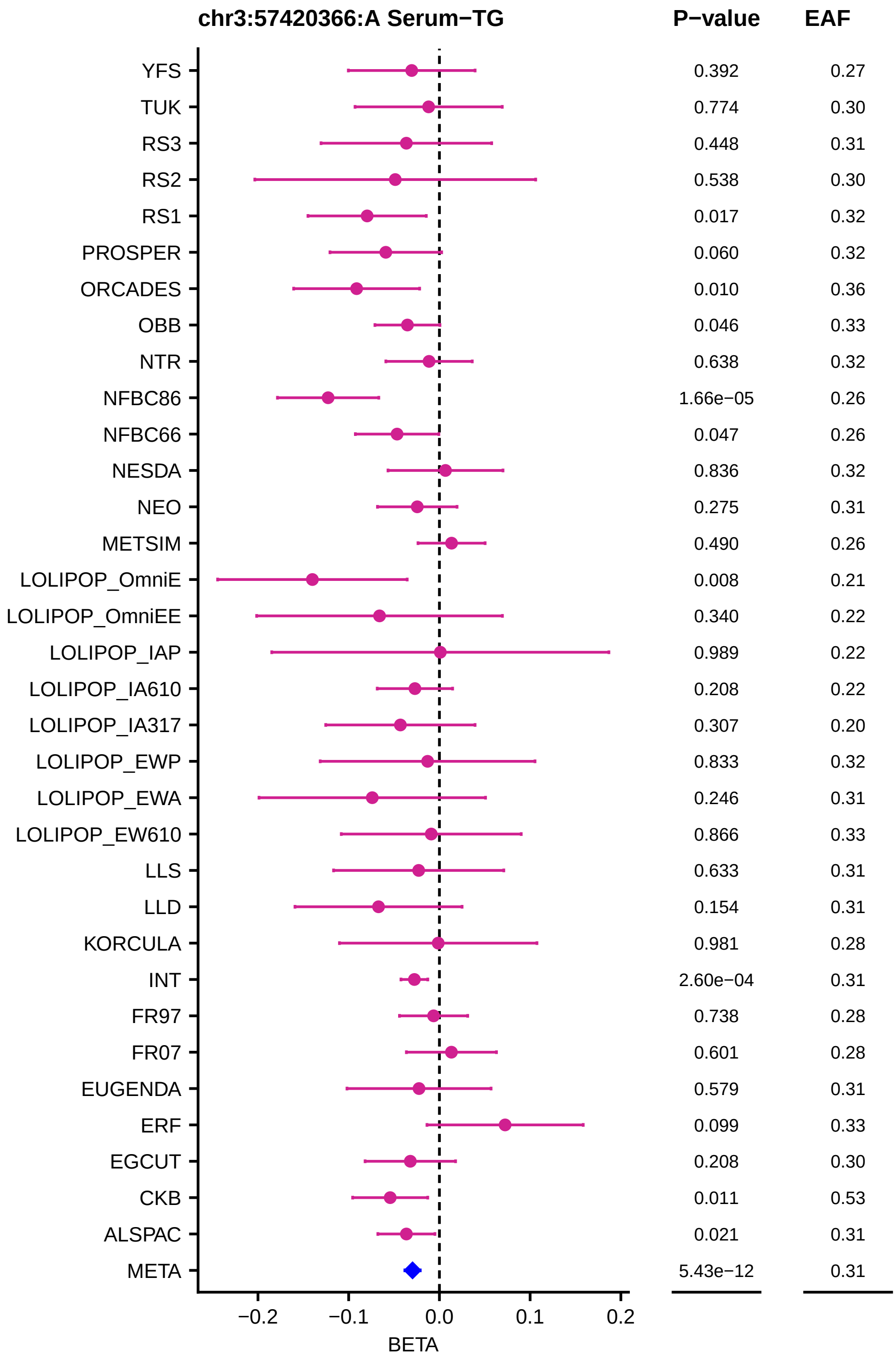

chr3:57420366:A Serum−TG
P−value
EAF
YFS 0.392 0.27
TUK 0.774 0.30
RS3 0.448 0.31
RS2 0.538 0.30
RS1 0.017 0.32
PROSPER 0.060 0.32
ORCADES 0.010 0.36
OBB 0.046 0.33
NTR 0.638 0.32
NFBC86 1.66e−05 0.26
NFBC66 0.047 0.26
NESDA 0.836 0.32
NEO 0.275 0.31
METSIM 0.490 0.26
LOLIPOP_OmniE 0.008 0.21
LOLIPOP_OmniEE 0.340 0.22
LOLIPOP_IAP 0.989 0.22
LOLIPOP_IA610 0.208 0.22
LOLIPOP_IA317 0.307 0.20
LOLIPOP_EWP 0.833 0.32
LOLIPOP_EWA 0.246 0.31
LOLIPOP_EW610 0.866 0.33
LLS 0.633 0.31
LLD 0.154 0.31
KORCULA 0.981 0.28
INT 2.60e−04 0.31
FR97 0.738 0.28
FR07 0.601 0.28
EUGENDA 0.579 0.31
ERF 0.099 0.33
EGCUT 0.208 0.30
CKB 0.011 0.53
ALSPAC 0.021 0.31
META 5.43e−12 0.31
−0.2
−0.1
0.0
0.1
0.2
BETA

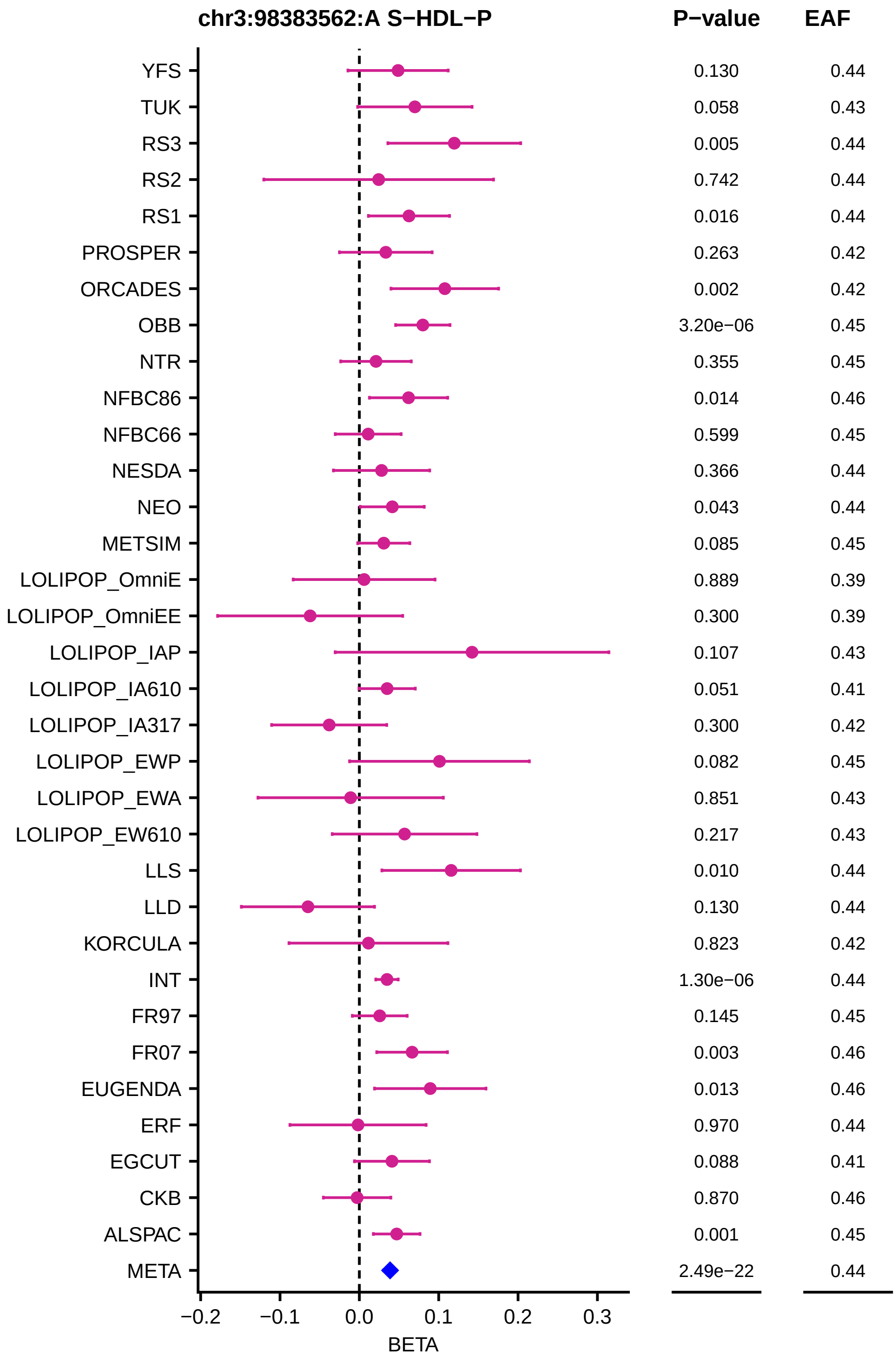

chr3:98383562:A S−HDL−P
P−value
EAF
YFS
TUK
RS3
RS2
RS1
PROSPER
ORCADES
OBB
NTR
NFBC86
NFBC66
NESDA
NEO
METSIM
LOLIPOP_OmniE
LOLIPOP_OmniEE
LOLIPOP_IAP
LOLIPOP_IA610
LOLIPOP_IA317
LOLIPOP_EWP
LOLIPOP_EWA
LOLIPOP_EW610
LLS
LLD
KORCULA
INT
FR97
FR07
EUGENDA
ERF
EGCUT
CKB
ALSPAC
META
0.130 0.44
0.058 0.43
0.005 0.44
0.742 0.44
0.016 0.44
0.263 0.42
0.002 0.42
3.20e−06 0.45
0.355 0.45
0.014 0.46
0.599 0.45
0.366 0.44
0.043 0.44
0.085 0.45
0.889 0.39
0.300 0.39
0.107 0.43
0.051 0.41
0.300 0.42
0.082 0.45
0.851 0.43
0.217 0.43
0.010 0.44
0.130 0.44
0.823 0.42
1.30e−06 0.44
0.145 0.45
0.003 0.46
0.013 0.46
0.970 0.44
0.088 0.41
0.870 0.46
0.001 0.45
2.49e−22 0.44
−0.2
−0.1
0.0
0.1
0.2
0.3
BETA

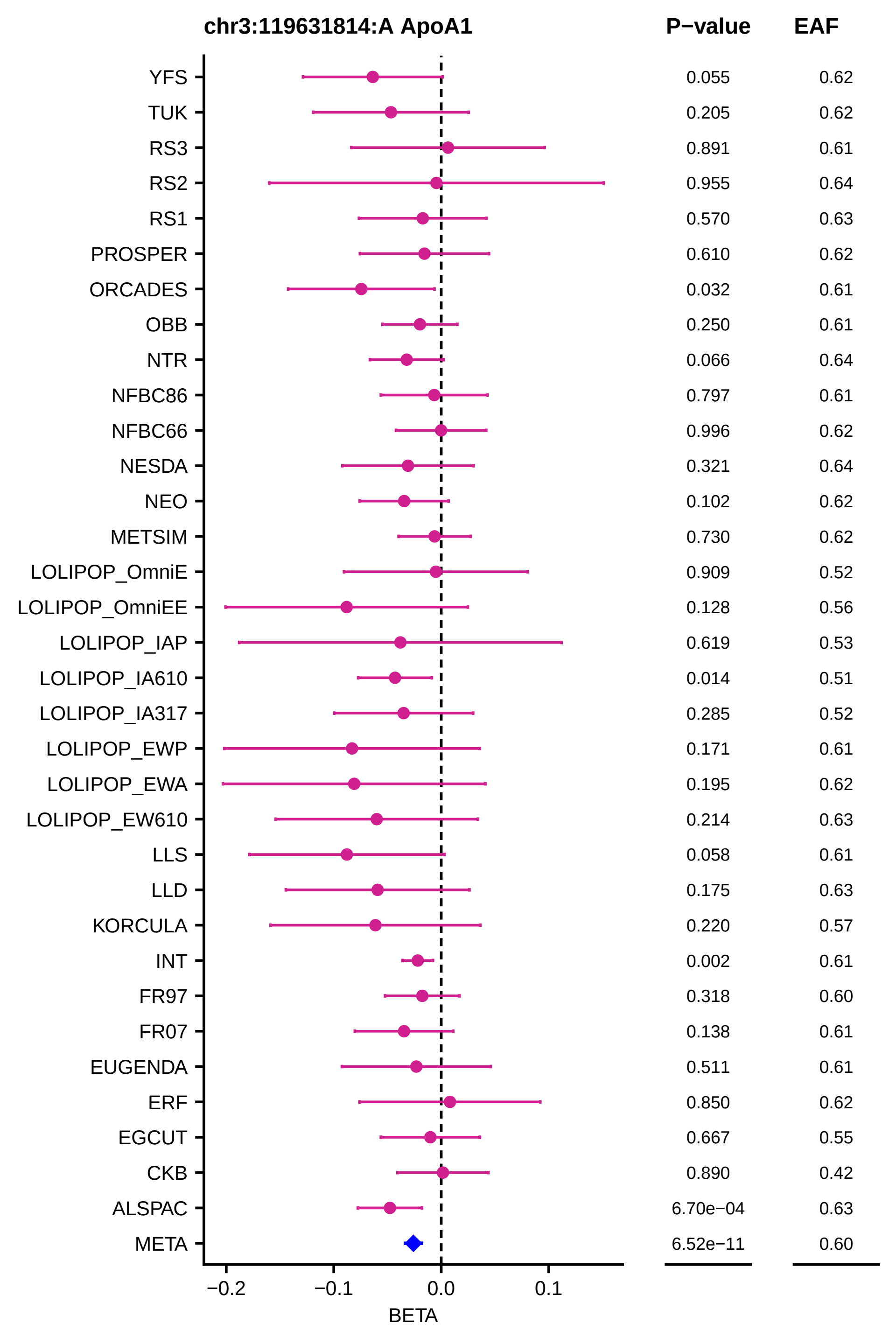
chr3:119631814:A ApoA1
P−value
EAF
YFS 0.055 0.62
TUK 0.205 0.62
RS3 0.891 0.61
RS2 0.955 0.64
RS1 0.570 0.63
PROSPER 0.610 0.62
ORCADES 0.032 0.61
OBB 0.250 0.61
NTR 0.066 0.64
NFBC86 0.797 0.61
NFBC66 0.996 0.62
NESDA 0.321 0.64
NEO 0.102 0.62
METSIM 0.730 0.62
LOLIPOP_OmniE 0.909 0.52
LOLIPOP_OmniEE 0.128 0.56
LOLIPOP_IAP 0.619 0.53
LOLIPOP_IA610 0.014 0.51
LOLIPOP_IA317 0.285 0.52
LOLIPOP_EWP 0.171 0.61
LOLIPOP_EWA 0.195 0.62
LOLIPOP_EW610 0.214 0.63
LLS 0.058 0.61
LLD 0.175 0.63
KORCULA 0.220 0.57
INT 0.002 0.61
FR97 0.318 0.60
FR07 0.138 0.61
EUGENDA 0.511 0.61
ERF 0.850 0.62
EGCUT 0.667 0.55
CKB 0.890 0.42
ALSPAC 6.70e−04 0.63
META 6.52e−11 0.60
−0.2
−0.1
0.0
0.1
BETA

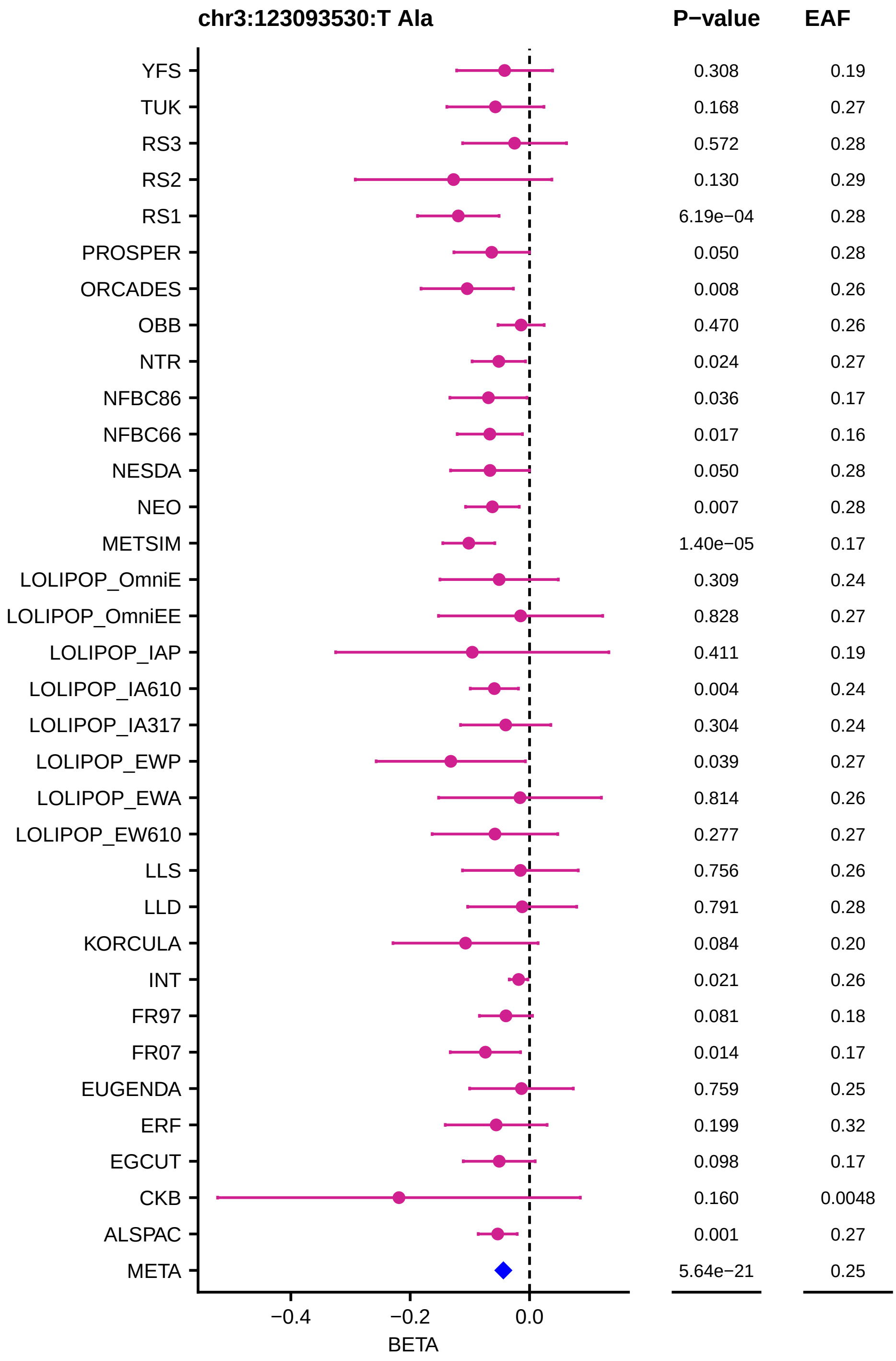
chr3:123093530:T Ala
P−value
EAF
YFS 0.308 0.19
TUK 0.168 0.27
RS3 0.572 0.28
RS2 0.130 0.29
RS1 6.19e−04 0.28
PROSPER 0.050 0.28
ORCADES 0.008 0.26
OBB 0.470 0.26
NTR 0.024 0.27
NFBC86 0.036 0.17
NFBC66 0.017 0.16
NESDA 0.050 0.28
NEO 0.007 0.28
METSIM 1.40e−05 0.17
LOLIPOP_OmniE 0.309 0.24
LOLIPOP_OmniEE 0.828 0.27
LOLIPOP_IAP 0.411 0.19
LOLIPOP_IA610 0.004 0.24
LOLIPOP_IA317 0.304 0.24
LOLIPOP_EWP 0.039 0.27
LOLIPOP_EWA 0.814 0.26
LOLIPOP_EW610 0.277 0.27
LLS 0.756 0.26
LLD 0.791 0.28
KORCULA 0.084 0.20
INT 0.021 0.26
FR97 0.081 0.18
FR07 0.014 0.17
EUGENDA 0.759 0.25
ERF 0.199 0.32
EGCUT 0.098 0.17
CKB 0.160 0.0048
ALSPAC 0.001 0.27
META 5.64e−21 0.25
−0.4
−0.2
0.0
BETA

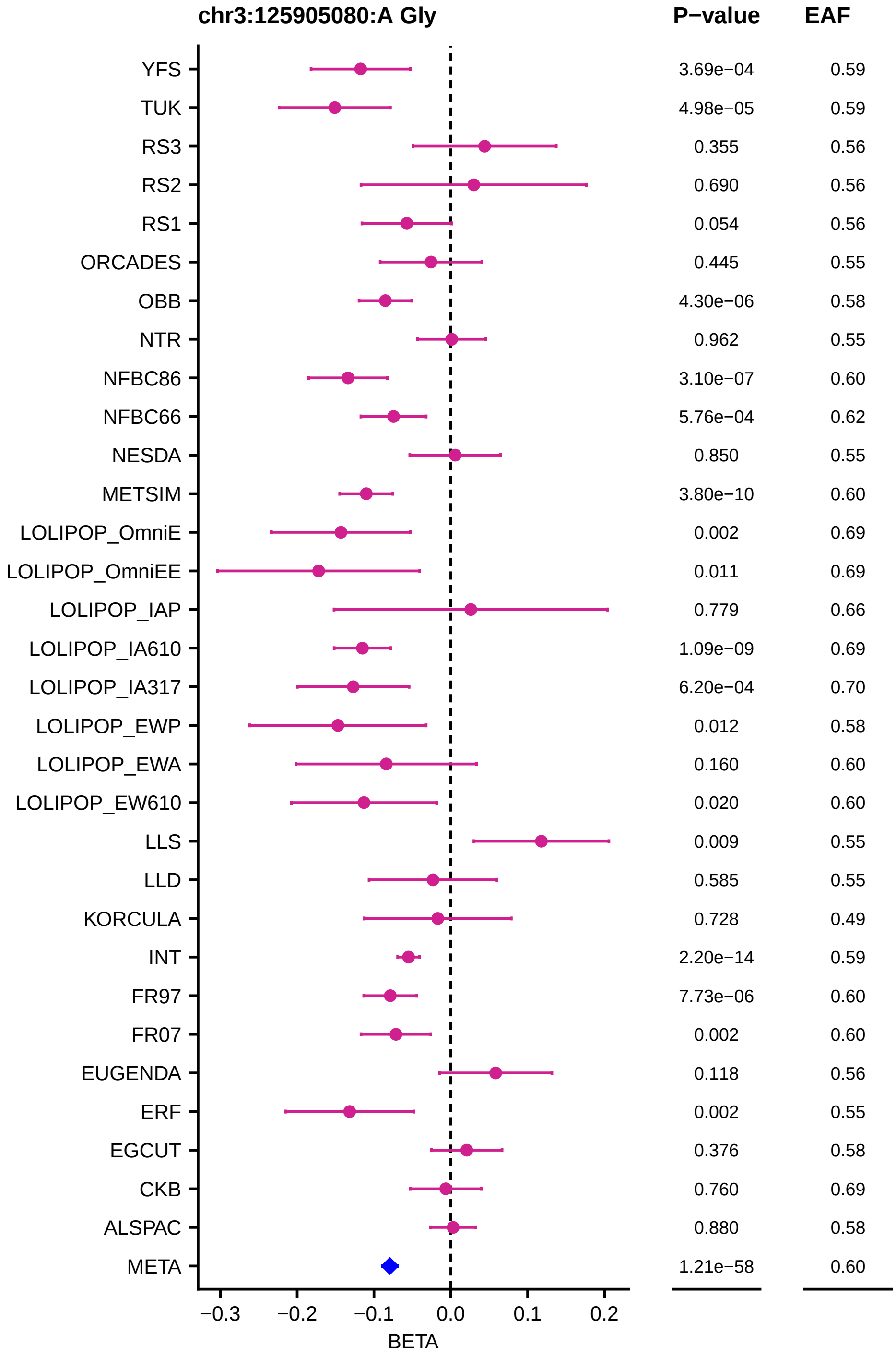
chr3:125905080:A Gly
P−value
EAF
YFS 3.69e−04 0.59
TUK 4.98e−05 0.59
RS3 0.355 0.56
RS2 0.690 0.56
RS1 0.054 0.56
ORCADES 0.445 0.55
OBB 4.30e−06 0.58
NTR 0.962 0.55
NFBC86 3.10e−07 0.60
NFBC66 5.76e−04 0.62
NESDA 0.850 0.55
METSIM 3.80e−10 0.60
LOLIPOP_OmniE 0.002 0.69
LOLIPOP_OmniEE 0.011 0.69
LOLIPOP_IAP 0.779 0.66
LOLIPOP_IA610 1.09e−09 0.69
LOLIPOP_IA317 6.20e−04 0.70
LOLIPOP_EWP 0.012 0.58
LOLIPOP_EWA 0.160 0.60
LOLIPOP_EW610 0.020 0.60
LLS 0.009 0.55
LLD 0.585 0.55
KORCULA 0.728 0.49
INT 2.20e−14 0.59
FR97 7.73e−06 0.60
FR07 0.002 0.60
EUGENDA 0.118 0.56
ERF 0.002 0.55
EGCUT 0.376 0.58
CKB 0.760 0.69
ALSPAC 0.880 0.58
META 1.21e−58 0.60
−0.3 −0.2 −0.1 0.0 0.1 0.2
BETA

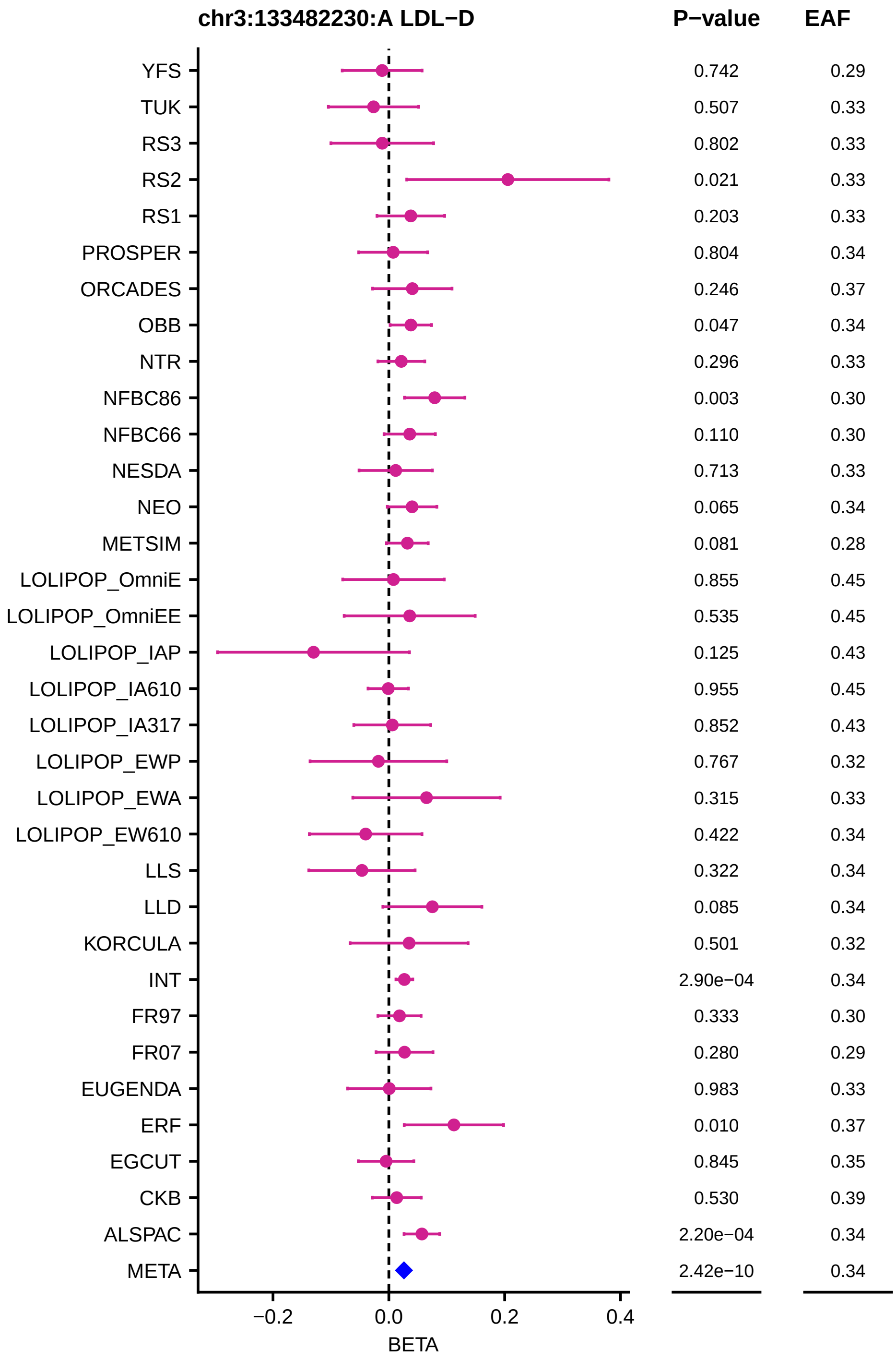
chr3:133482230:A LDL−D
P−value
EAF
YFS 0.742 0.29
TUK 0.507 0.33
RS3 0.802 0.33
RS2 0.021 0.33
RS1 0.203 0.33
PROSPER 0.804 0.34
ORCADES 0.246 0.37
OBB 0.047 0.34
NTR 0.296 0.33
NFBC86 0.003 0.30
NFBC66 0.110 0.30
NESDA 0.713 0.33
NEO 0.065 0.34
METSIM 0.081 0.28
LOLIPOP_OmniE 0.855 0.45
LOLIPOP_OmniEE 0.535 0.45
LOLIPOP_IAP 0.125 0.43
LOLIPOP_IA610 0.955 0.45
LOLIPOP_IA317 0.852 0.43
LOLIPOP_EWP 0.767 0.32
LOLIPOP_EWA 0.315 0.33
LOLIPOP_EW610 0.422 0.34
LLS 0.322 0.34
LLD 0.085 0.34
KORCULA 0.501 0.32
INT 2.90e−04 0.34
FR97 0.333 0.30
FR07 0.280 0.29
EUGENDA 0.983 0.33
ERF 0.010 0.37
EGCUT 0.845 0.35
CKB 0.530 0.39
ALSPAC 2.20e−04 0.34
META 2.42e−10 0.34
−0.2
0.0
0.2
0.4
BETA

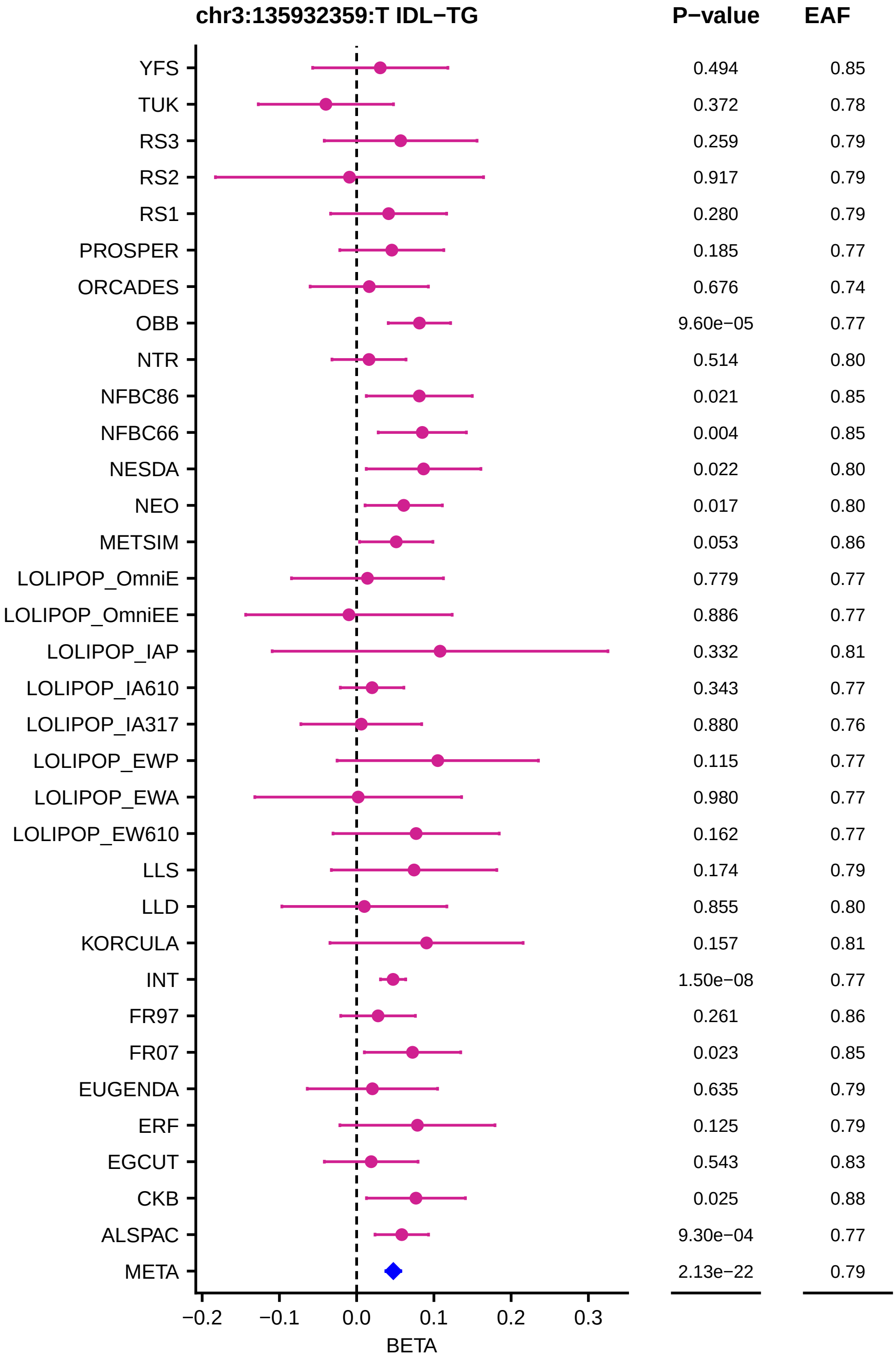
chr3:135932359:T IDL−TG
P−value
EAF
YFS 0.494 0.85
TUK 0.372 0.78
RS3 0.259 0.79
RS2 0.917 0.79
RS1 0.280 0.79
PROSPER 0.185 0.77
ORCADES 0.676 0.74
OBB 9.60e−05 0.77
NTR 0.514 0.80
NFBC86 0.021 0.85
NFBC66 0.004 0.85
NESDA 0.022 0.80
NEO 0.017 0.80
METSIM 0.053 0.86
LOLIPOP_OmniE 0.779 0.77
LOLIPOP_OmniEE 0.886 0.77
LOLIPOP_IAP 0.332 0.81
LOLIPOP_IA610 0.343 0.77
LOLIPOP_IA317 0.880 0.76
LOLIPOP_EWP 0.115 0.77
LOLIPOP_EWA 0.980 0.77
LOLIPOP_EW610 0.162 0.77
LLS 0.174 0.79
LLD 0.855 0.80
KORCULA 0.157 0.81
INT 1.50e−08 0.77
FR97 0.261 0.86
FR07 0.023 0.85
EUGENDA 0.635 0.79
ERF 0.125 0.79
EGCUT 0.543 0.83
CKB 0.025 0.88
ALSPAC 9.30e−04 0.77
META 2.13e−22 0.79
−0.2
−0.1
0.0
0.1
0.2
0.3
BETA

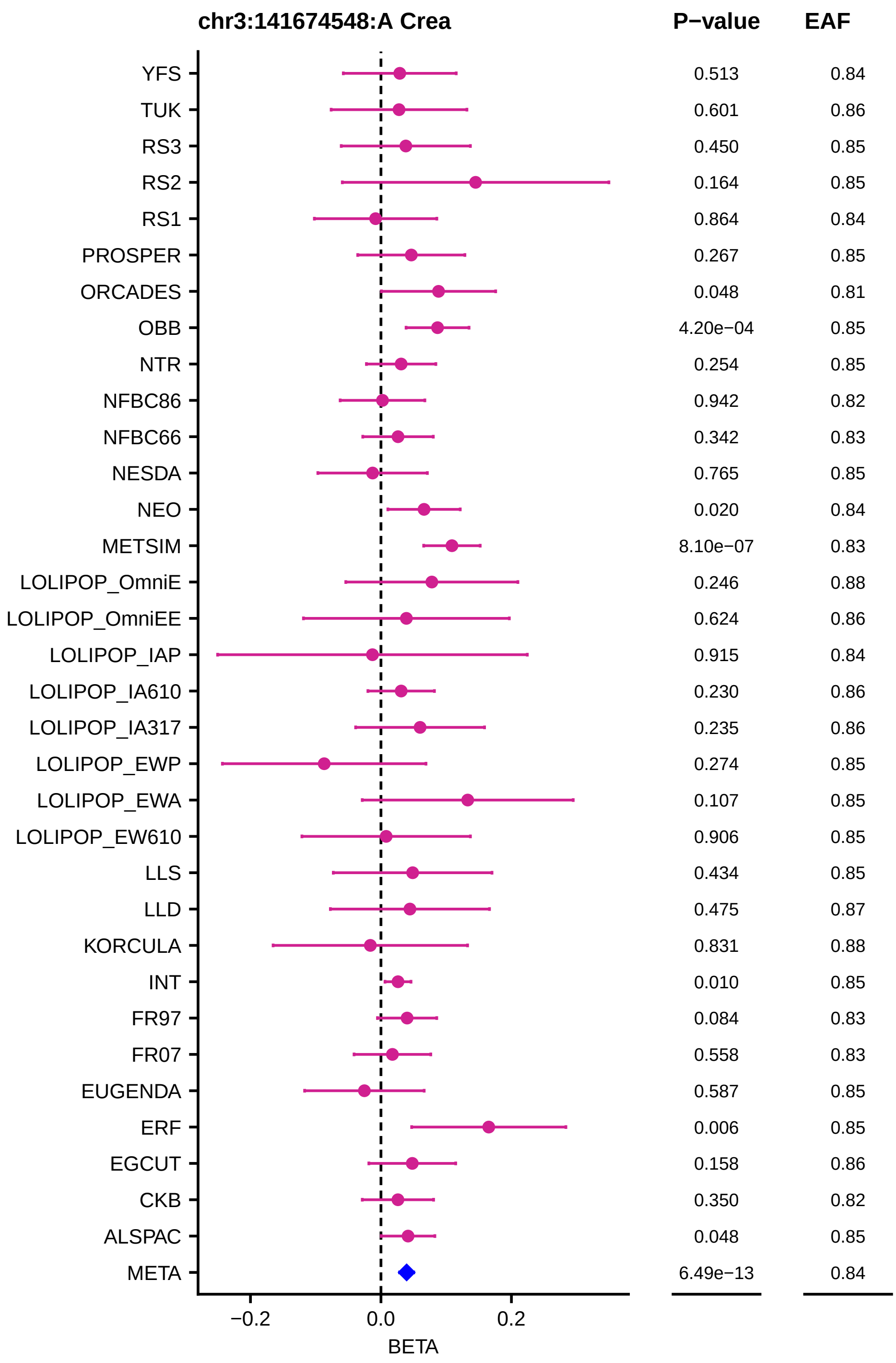
chr3:141674548:A Crea
P−value
EAF
YFS 0.513 0.84
TUK 0.601 0.86
RS3 0.450 0.85
RS2 0.164 0.85
RS1 0.864 0.84
PROSPER 0.267 0.85
ORCADES 0.048 0.81
OBB 4.20e−04 0.85
NTR 0.254 0.85
NFBC86 0.942 0.82
NFBC66 0.342 0.83
NESDA 0.765 0.85
NEO 0.020 0.84
METSIM 8.10e−07 0.83
LOLIPOP_OmniE 0.246 0.88
LOLIPOP_OmniEE 0.624 0.86
LOLIPOP_IAP 0.915 0.84
LOLIPOP_IA610 0.230 0.86
LOLIPOP_IA317 0.235 0.86
LOLIPOP_EWP 0.274 0.85
LOLIPOP_EWA 0.107 0.85
LOLIPOP_EW610 0.906 0.85
LLS 0.434 0.85
LLD 0.475 0.87
KORCULA 0.831 0.88
INT 0.010 0.85
FR97 0.084 0.83
FR07 0.558 0.83
EUGENDA 0.587 0.85
ERF 0.006 0.85
EGCUT 0.158 0.86
CKB 0.350 0.82
ALSPAC 0.048 0.85
META 6.49e−13 0.84
−0.2
0.0
0.2
BETA

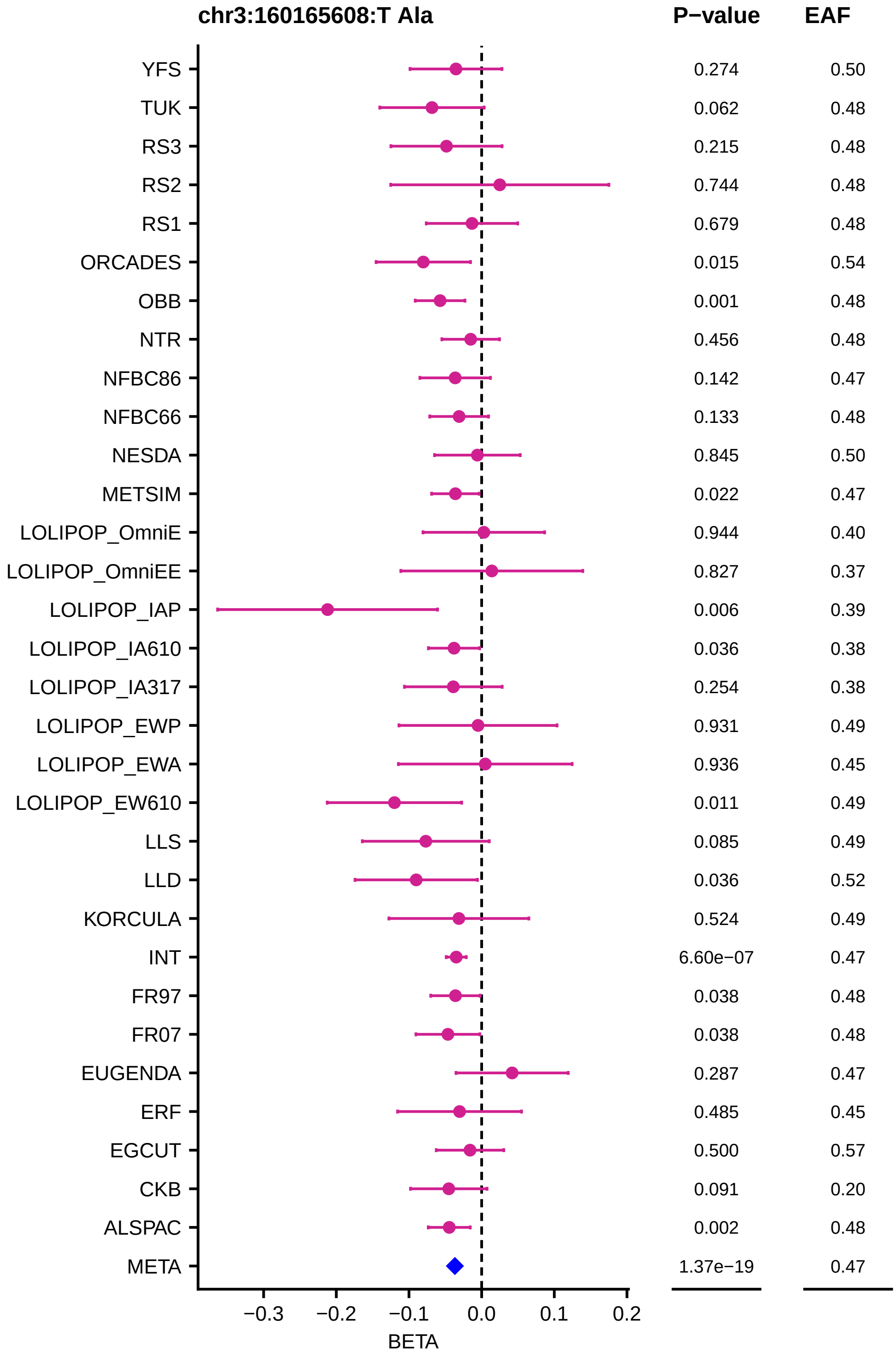

chr3:160165608:T Ala
P−value
EAF
YFS 0.274 0.50
TUK 0.062 0.48
RS3 0.215 0.48
RS2 0.744 0.48
RS1 0.679 0.48
ORCADES 0.015 0.54
OBB 0.001 0.48
NTR 0.456 0.48
NFBC86 0.142 0.47
NFBC66 0.133 0.48
NESDA 0.845 0.50
METSIM 0.022 0.47
LOLIPOP_OmniE 0.944 0.40
LOLIPOP_OmniEE 0.827 0.37
LOLIPOP_IAP 0.006 0.39
LOLIPOP_IA610 0.036 0.38
LOLIPOP_IA317 0.254 0.38
LOLIPOP_EWP 0.931 0.49
LOLIPOP_EWA 0.936 0.45
LOLIPOP_EW610 0.011 0.49
LLS 0.085 0.49
LLD 0.036 0.52
KORCULA 0.524 0.49
INT 6.60e−07 0.47
FR97 0.038 0.48
FR07 0.038 0.48
EUGENDA 0.287 0.47
ERF 0.485 0.45
EGCUT 0.500 0.57
CKB 0.091 0.20
ALSPAC 0.002 0.48
META 1.37e−19 0.47
−0.3 −0.2 −0.1 0.0 0.1 0.2
BETA

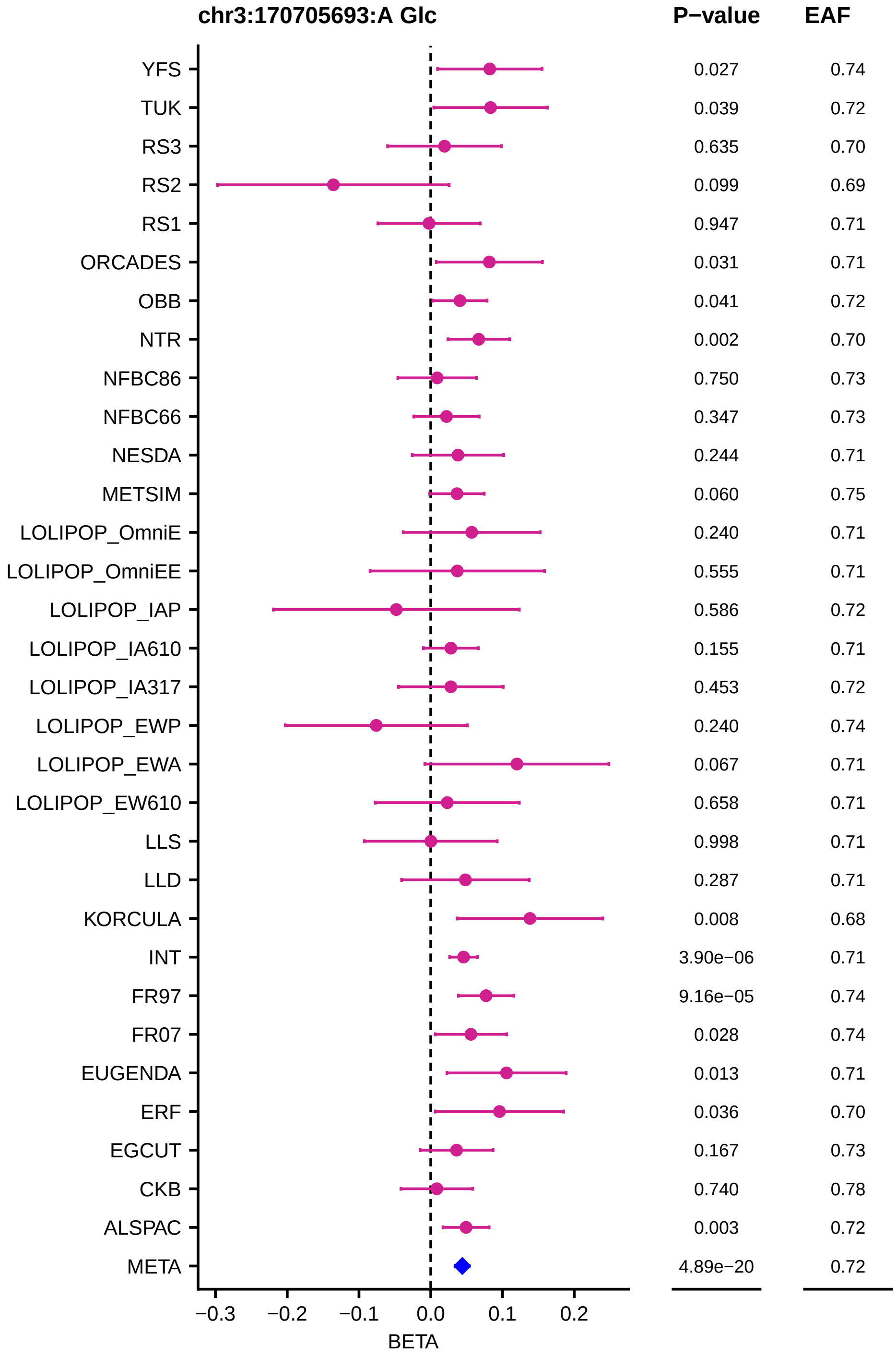
chr3:170705693:A Glc
P−value
EAF
YFS 0.027 0.74
TUK 0.039 0.72
RS3 0.635 0.70
RS2 0.099 0.69
RS1 0.947 0.71
ORCADES 0.031 0.71
OBB 0.041 0.72
NTR 0.002 0.70
NFBC86 0.750 0.73
NFBC66 0.347 0.73
NESDA 0.244 0.71
METSIM 0.060 0.75
LOLIPOP_OmniE 0.240 0.71
LOLIPOP_OmniEE 0.555 0.71
LOLIPOP_IAP 0.586 0.72
LOLIPOP_IA610 0.155 0.71
LOLIPOP_IA317 0.453 0.72
LOLIPOP_EWP 0.240 0.74
LOLIPOP_EWA 0.067 0.71
LOLIPOP_EW610 0.658 0.71
LLS 0.998 0.71
LLD 0.287 0.71
KORCULA 0.008 0.68
INT 3.90e−06 0.71
FR97 9.16e−05 0.74
FR07 0.028 0.74
EUGENDA 0.013 0.71
ERF 0.036 0.70
EGCUT 0.167 0.73
CKB 0.740 0.78
ALSPAC 0.003 0.72
META 4.89e−20 0.72
−0.3 −0.2 −0.1 0.0 0.1 0.2
BETA

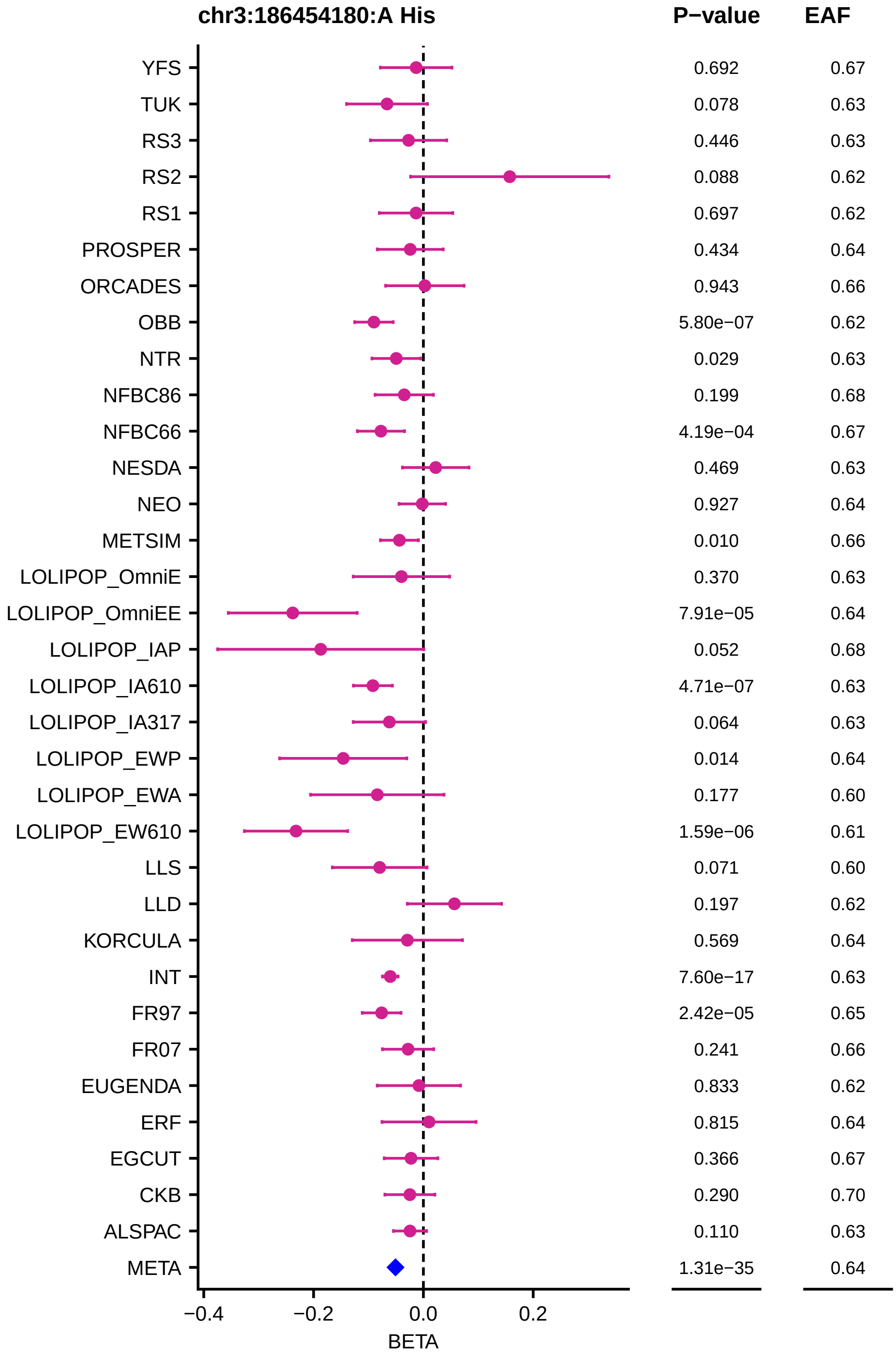
chr3:186454180:A His
P−value
EAF
YFS 0.692 0.67
TUK 0.078 0.63
RS3 0.446 0.63
RS2 0.088 0.62
RS1 0.697 0.62
PROSPER 0.434 0.64
ORCADES 0.943 0.66
OBB 5.80e−07 0.62
NTR 0.029 0.63
NFBC86 0.199 0.68
NFBC66 4.19e−04 0.67
NESDA 0.469 0.63
NEO 0.927 0.64
METSIM 0.010 0.66
LOLIPOP_OmniE 0.370 0.63
LOLIPOP_OmniEE 7.91e−05 0.64
LOLIPOP_IAP 0.052 0.68
LOLIPOP_IA610 4.71e−07 0.63
LOLIPOP_IA317 0.064 0.63
LOLIPOP_EWP 0.014 0.64
LOLIPOP_EWA 0.177 0.60
LOLIPOP_EW610 1.59e−06 0.61
LLS 0.071 0.60
LLD 0.197 0.62
KORCULA 0.569 0.64
INT 7.60e−17 0.63
FR97 2.42e−05 0.65
FR07 0.241 0.66
EUGENDA 0.833 0.62
ERF 0.815 0.64
EGCUT 0.366 0.67
CKB 0.290 0.70
ALSPAC 0.110 0.63
META 1.31e−35 0.64
−0.4
−0.2
0.0
0.2
BETA

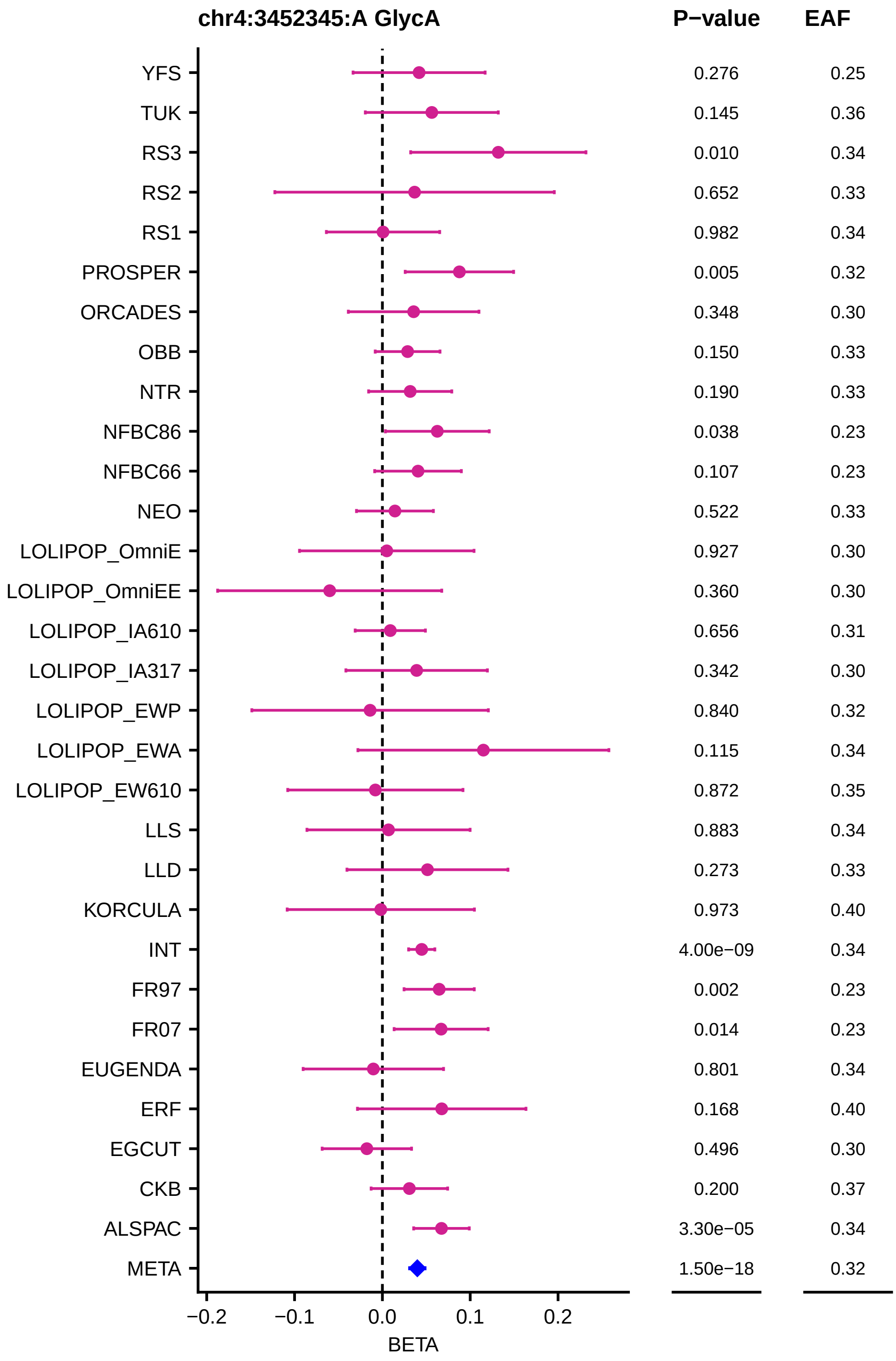
chr4:3452345:A GlycA
P−value
EAF
YFS 0.276 0.25
TUK 0.145 0.36
RS3 0.010 0.34
RS2 0.652 0.33
RS1 0.982 0.34
PROSPER 0.005 0.32
ORCADES 0.348 0.30
OBB 0.150 0.33
NTR 0.190 0.33
NFBC86 0.038 0.23
NFBC66 0.107 0.23
NEO 0.522 0.33
LOLIPOP_OmniE 0.927 0.30
LOLIPOP_OmniEE 0.360 0.30
LOLIPOP_IA610 0.656 0.31
LOLIPOP_IA317 0.342 0.30
LOLIPOP_EWP 0.840 0.32
LOLIPOP_EWA 0.115 0.34
LOLIPOP_EW610 0.872 0.35
LLS 0.883 0.34
LLD 0.273 0.33
KORCULA 0.973 0.40
INT 4.00e−09 0.34
FR97 0.002 0.23
FR07 0.014 0.23
EUGENDA 0.801 0.34
ERF 0.168 0.40
EGCUT 0.496 0.30
CKB 0.200 0.37
ALSPAC 3.30e−05 0.34
META 1.50e−18 0.32
−0.2 −0.1 0.0 0.1 0.2
BETA

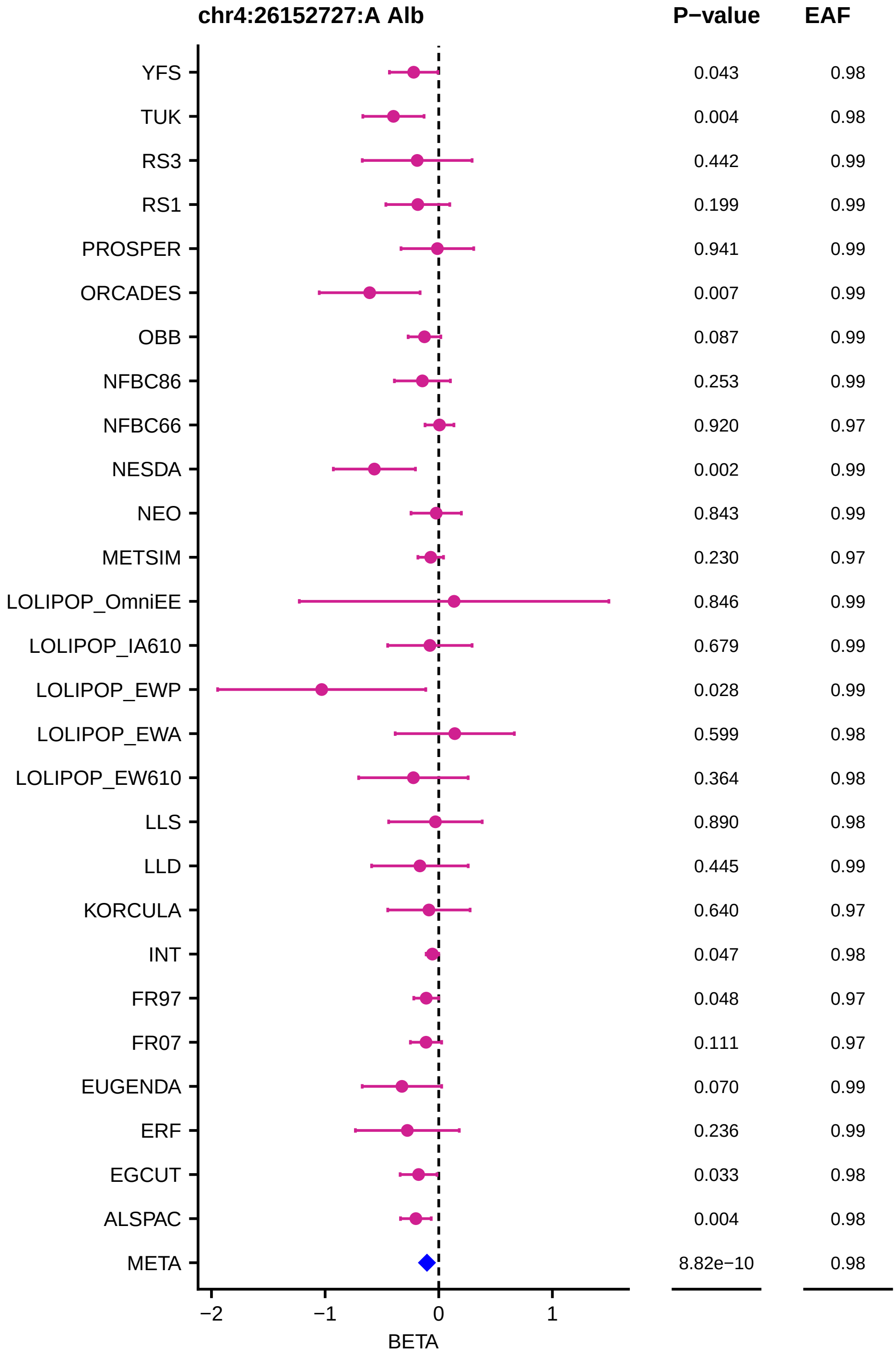

chr4:26152727:A Alb
P−value
EAF
YFS
TUK
RS3
RS1
PROSPER
ORCADES
OBB
NFBC86
NFBC66
NESDA
NEO
METSIM
LOLIPOP_OmniEE
LOLIPOP_IA610
LOLIPOP_EWP
LOLIPOP_EWA
LOLIPOP_EW610
LLS
LLD
KORCULA
INT
FR97
FR07
EUGENDA
ERF
EGCUT
ALSPAC
META
0.043 0.98
0.004 0.98
0.442 0.99
0.199 0.99
0.941 0.99
0.007 0.99
0.087 0.99
0.253 0.99
0.920 0.97
0.002 0.99
0.843 0.99
0.230 0.97
0.846 0.99
0.679 0.99
0.028 0.99
0.599 0.98
0.364 0.98
0.890 0.98
0.445 0.99
0.640 0.97
0.047 0.98
0.048 0.97
0.111 0.97
0.070 0.99
0.236 0.99
0.033 0.98
0.004 0.98
8.82e−10 0.98
−2
−1
0
1
BETA

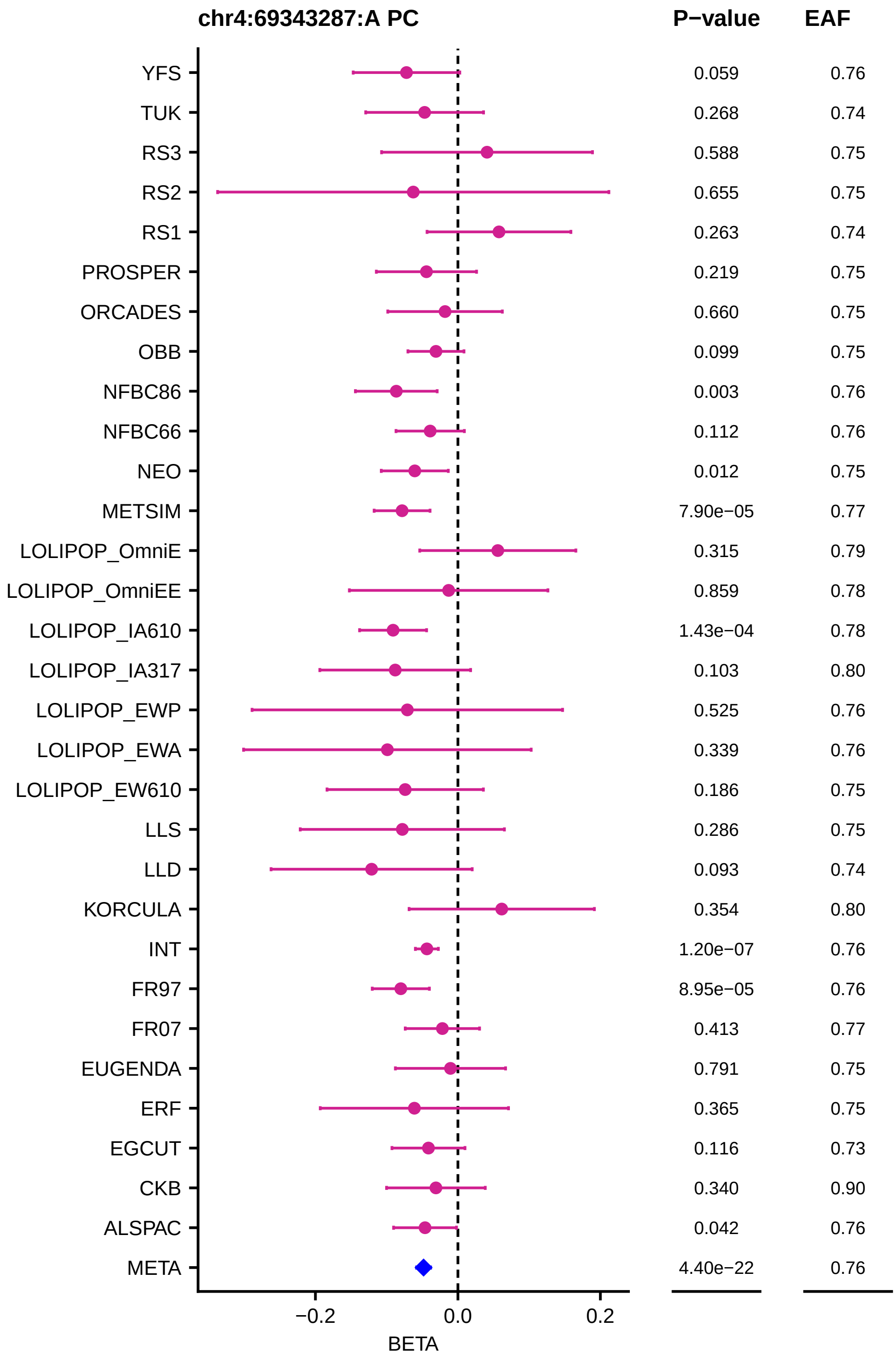
chr4:69343287:A PC
P−value
EAF
YFS 0.059 0.76
TUK 0.268 0.74
RS3 0.588 0.75
RS2 0.655 0.75
RS1 0.263 0.74
PROSPER 0.219 0.75
ORCADES 0.660 0.75
OBB 0.099 0.75
NFBC86 0.003 0.76
NFBC66 0.112 0.76
NEO 0.012 0.75
METSIM 7.90e−05 0.77
LOLIPOP_OmniE 0.315 0.79
LOLIPOP_OmniEE 0.859 0.78
LOLIPOP_IA610 1.43e−04 0.78
LOLIPOP_IA317 0.103 0.80
LOLIPOP_EWP 0.525 0.76
LOLIPOP_EWA 0.339 0.76
LOLIPOP_EW610 0.186 0.75
LLS 0.286 0.75
LLD 0.093 0.74
KORCULA 0.354 0.80
INT 1.20e−07 0.76
FR97 8.95e−05 0.76
FR07 0.413 0.77
EUGENDA 0.791 0.75
ERF 0.365 0.75
EGCUT 0.116 0.73
CKB 0.340 0.90
ALSPAC 0.042 0.76
META 4.40e−22 0.76
−0.2
0.0
0.2
BETA

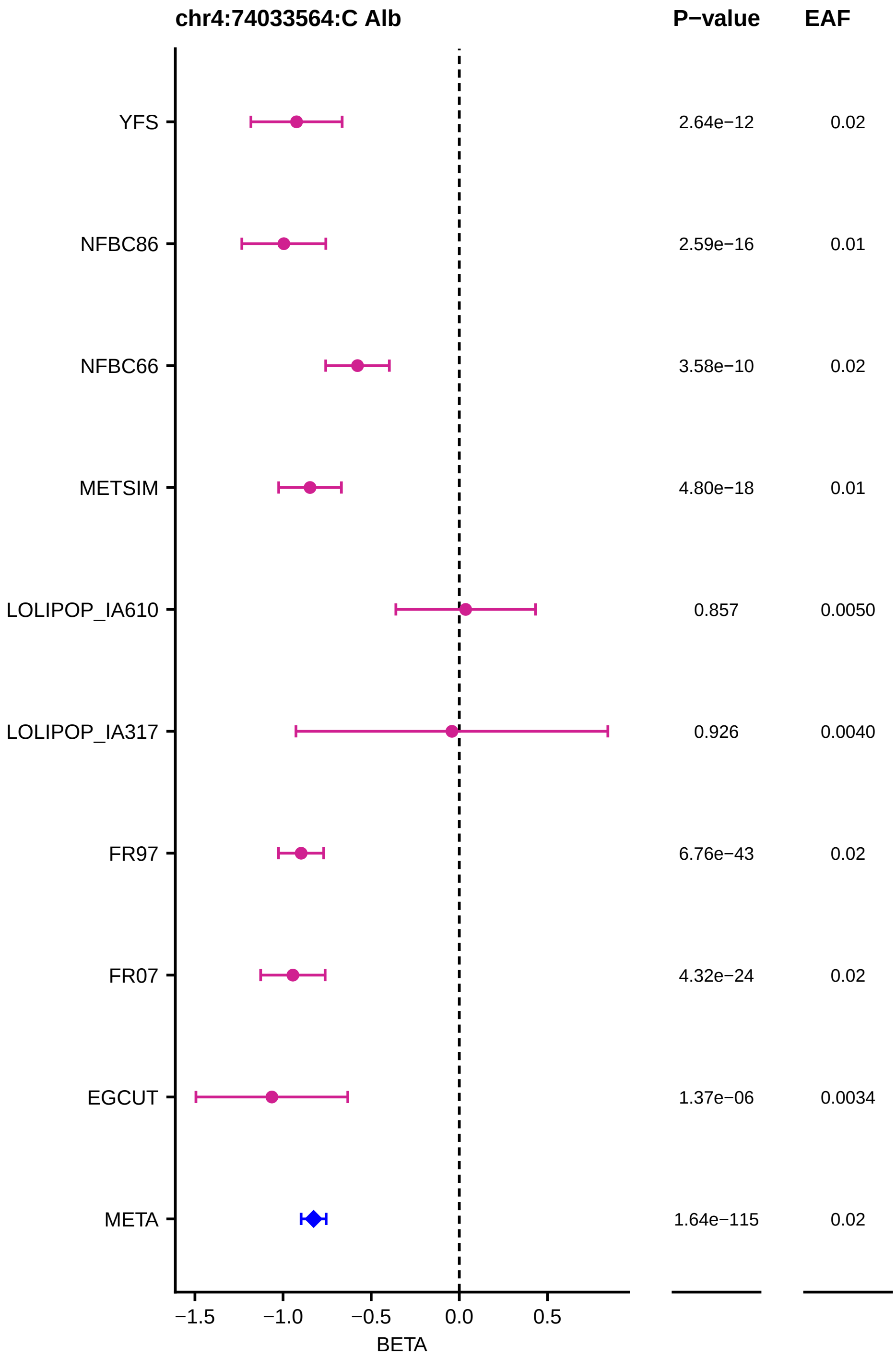

chr4:74033564:C Alb
P−value
EAF
YFS
2.64e−12
0.02
NFBC86
2.59e−16
0.01
NFBC66
3.58e−10
0.02
METSIM
4.80e−18
0.01
LOLIPOP_IA610
0.857
0.0050
LOLIPOP_IA317
0.926
0.0040
FR97
6.76e−43
0.02
FR07
4.32e−24
0.02
EGCUT
1.37e−06
0.0034
META
1.64e−115
0.02
−1.5
−1.0
−0.5
0.0
0.5
BETA

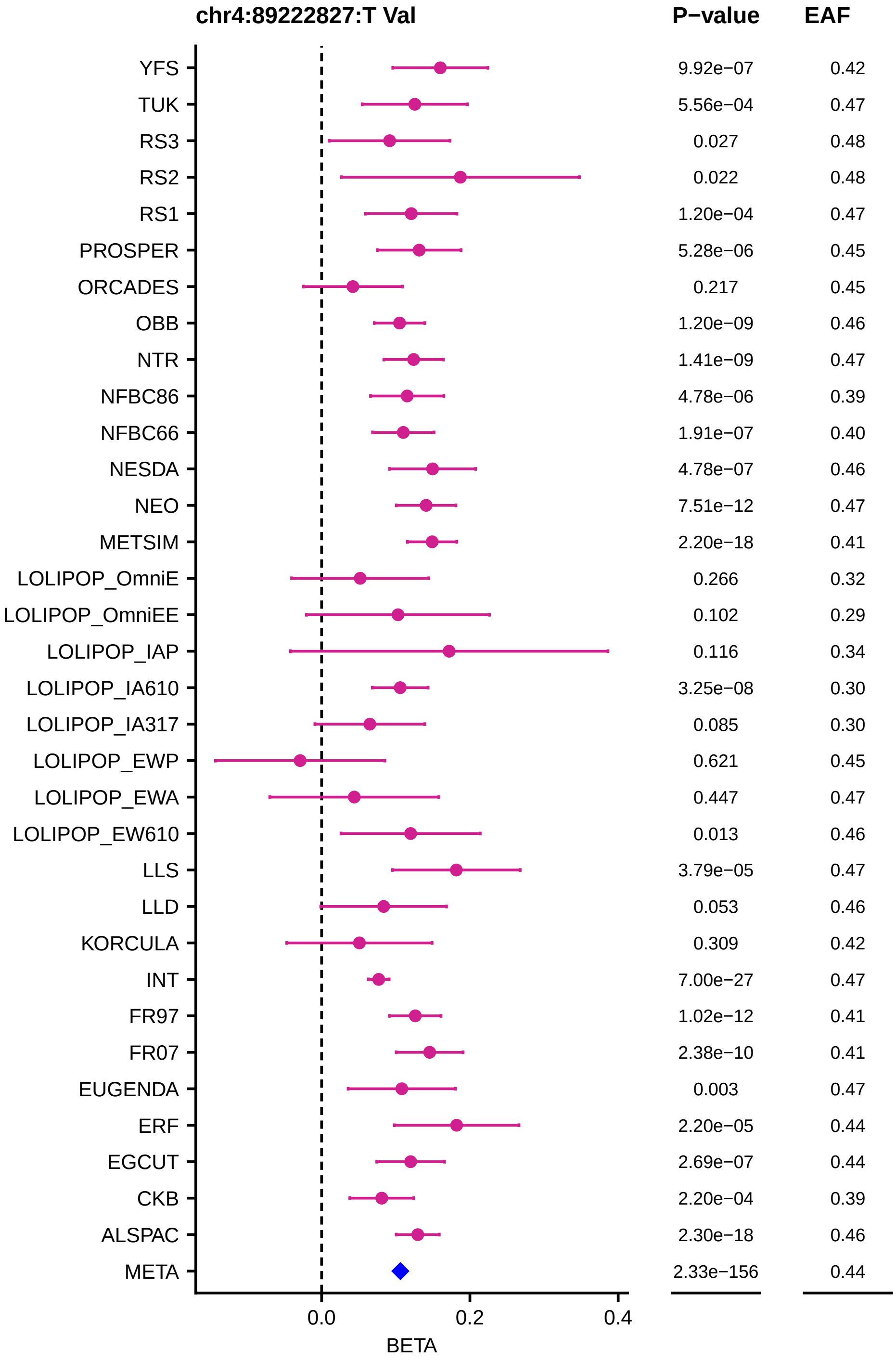
chr4:89222827:T Val
P−value
EAF
YFS 9.92e−07 0.42
TUK 5.56e−04 0.47
RS3 0.027 0.48
RS2 0.022 0.48
RS1 1.20e−04 0.47
PROSPER 5.28e−06 0.45
ORCADES 0.217 0.45
OBB 1.20e−09 0.46
NTR 1.41e−09 0.47
NFBC86 4.78e−06 0.39
NFBC66 1.91e−07 0.40
NESDA 4.78e−07 0.46
NEO 7.51e−12 0.47
METSIM 2.20e−18 0.41
LOLIPOP_OmniE 0.266 0.32
LOLIPOP_OmniEE 0.102 0.29
LOLIPOP_IAP 0.116 0.34
LOLIPOP_IA610 3.25e−08 0.30
LOLIPOP_IA317 0.085 0.30
LOLIPOP_EWP 0.621 0.45
LOLIPOP_EWA 0.447 0.47
LOLIPOP_EW610 0.013 0.46
LLS 3.79e−05 0.47
LLD 0.053 0.46
KORCULA 0.309 0.42
INT 7.00e−27 0.47
FR97 1.02e−12 0.41
FR07 2.38e−10 0.41
EUGENDA 0.003 0.47
ERF 2.20e−05 0.44
EGCUT 2.69e−07 0.44
CKB 2.20e−04 0.39
ALSPAC 2.30e−18 0.46
META 2.33e−156 0.44
0.0
0.2
0.4
BETA

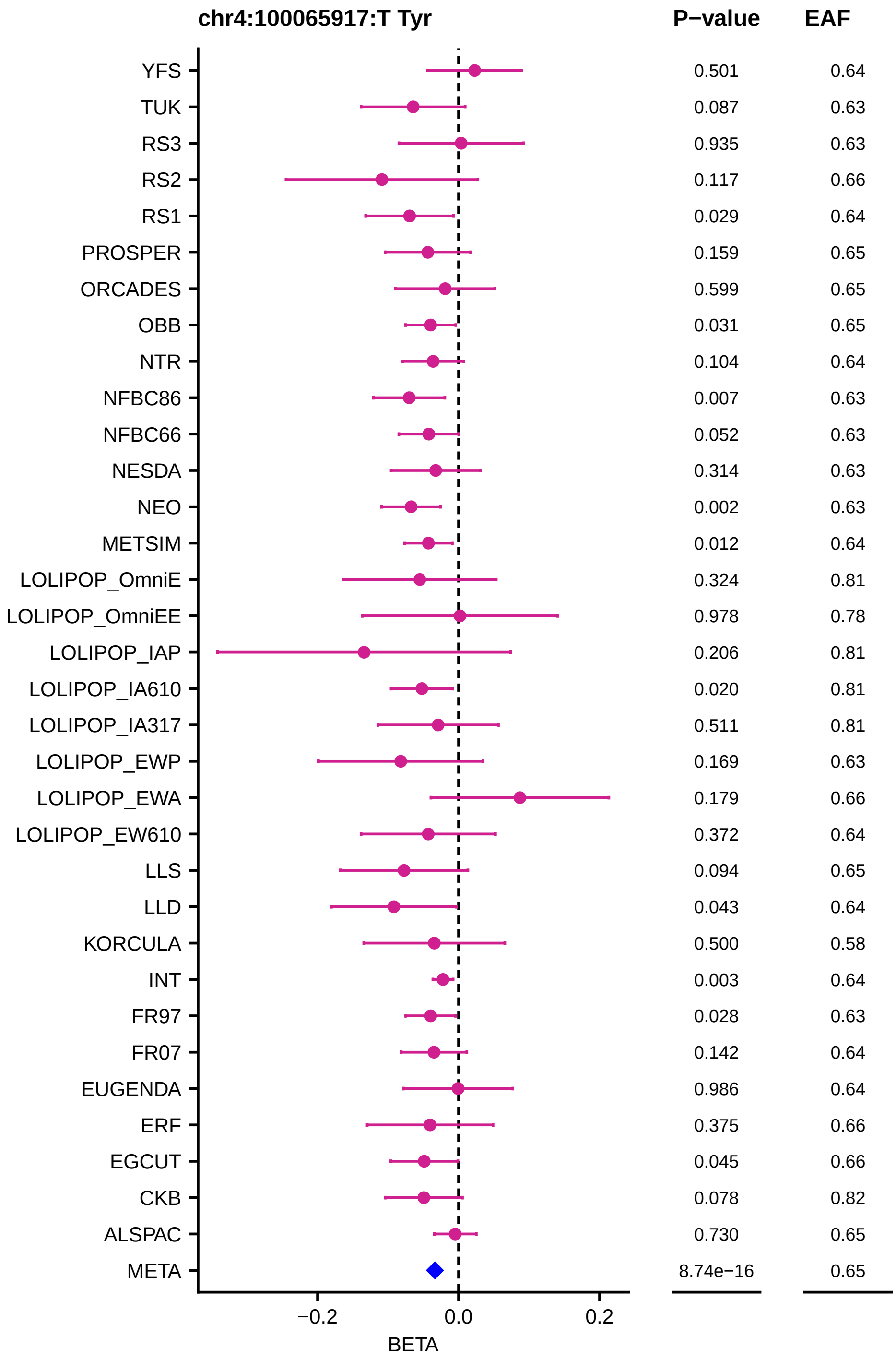
chr4:100065917:T Tyr
P−value
EAF
YFS 0.501 0.64
TUK 0.087 0.63
RS3 0.935 0.63
RS2 0.117 0.66
RS1 0.029 0.64
PROSPER 0.159 0.65
ORCADES 0.599 0.65
OBB 0.031 0.65
NTR 0.104 0.64
NFBC86 0.007 0.63
NFBC66 0.052 0.63
NESDA 0.314 0.63
NEO 0.002 0.63
METSIM 0.012 0.64
LOLIPOP_OmniE 0.324 0.81
LOLIPOP_OmniEE 0.978 0.78
LOLIPOP_IAP 0.206 0.81
LOLIPOP_IA610 0.020 0.81
LOLIPOP_IA317 0.511 0.81
LOLIPOP_EWP 0.169 0.63
LOLIPOP_EWA 0.179 0.66
LOLIPOP_EW610 0.372 0.64
LLS 0.094 0.65
LLD 0.043 0.64
KORCULA 0.500 0.58
INT 0.003 0.64
FR97 0.028 0.63
FR07 0.142 0.64
EUGENDA 0.986 0.64
ERF 0.375 0.66
EGCUT 0.045 0.66
CKB 0.078 0.82
ALSPAC 0.730 0.65
META 8.74e−16 0.65
−0.2
0.0
0.2
BETA

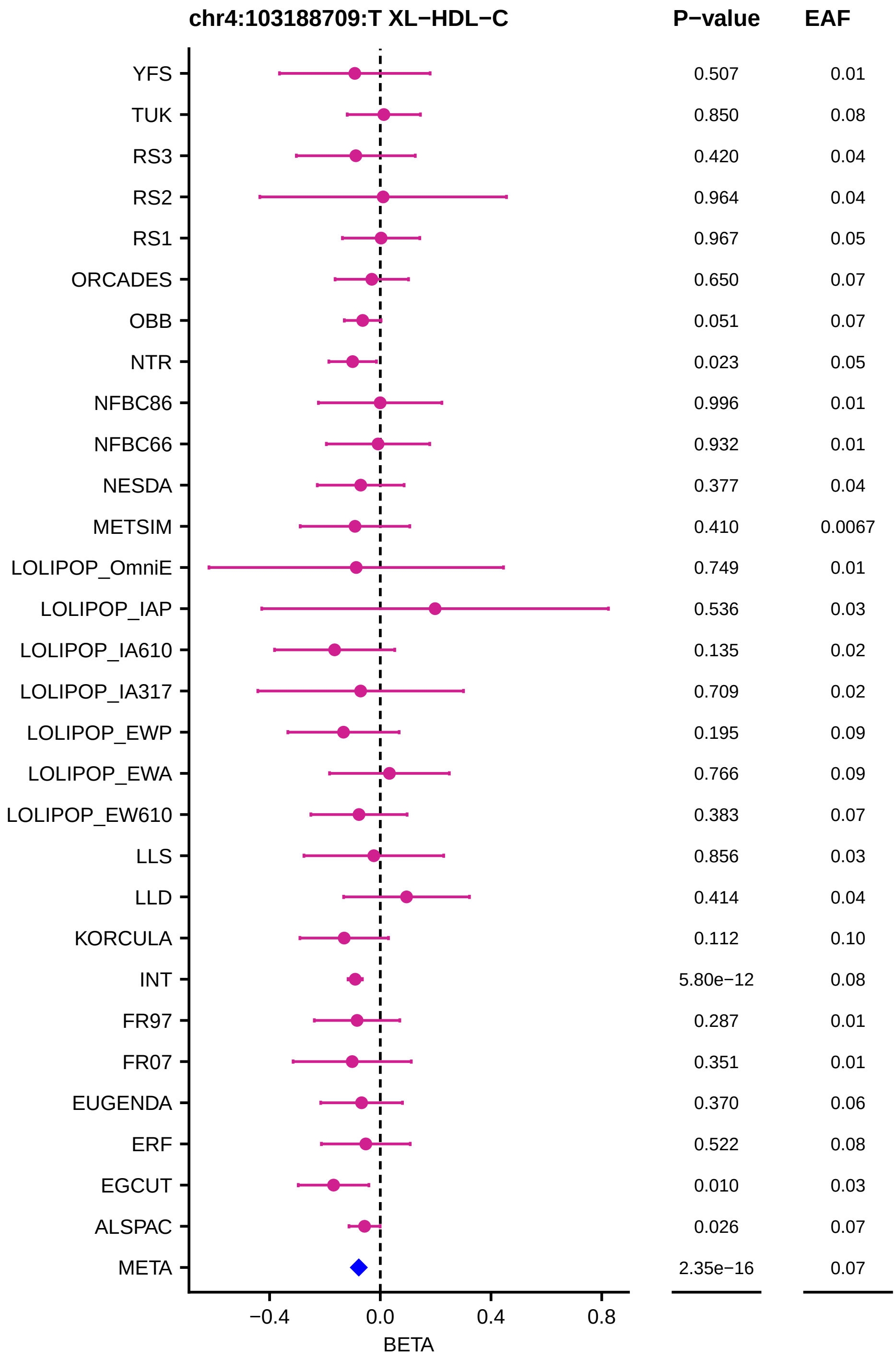
chr4:103188709:T XL−HDL−C
P−value
EAF
YFS 0.507 0.01
TUK 0.850 0.08
RS3 0.420 0.04
RS2 0.964 0.04
RS1 0.967 0.05
ORCADES 0.650 0.07
OBB 0.051 0.07
NTR 0.023 0.05
NFBC86 0.996 0.01
NFBC66 0.932 0.01
NESDA 0.377 0.04
METSIM 0.410 0.0067
LOLIPOP_OmniE 0.749 0.01
LOLIPOP_IAP 0.536 0.03
LOLIPOP_IA610 0.135 0.02
LOLIPOP_IA317 0.709 0.02
LOLIPOP_EWP 0.195 0.09
LOLIPOP_EWA 0.766 0.09
LOLIPOP_EW610 0.383 0.07
LLS 0.856 0.03
LLD 0.414 0.04
KORCULA 0.112 0.10
INT 5.80e−12 0.08
FR97 0.287 0.01
FR07 0.351 0.01
EUGENDA 0.370 0.06
ERF 0.522 0.08
EGCUT 0.010 0.03
ALSPAC 0.026 0.07
META 2.35e−16 0.07
−0.4
0.0
0.4
0.8
BETA

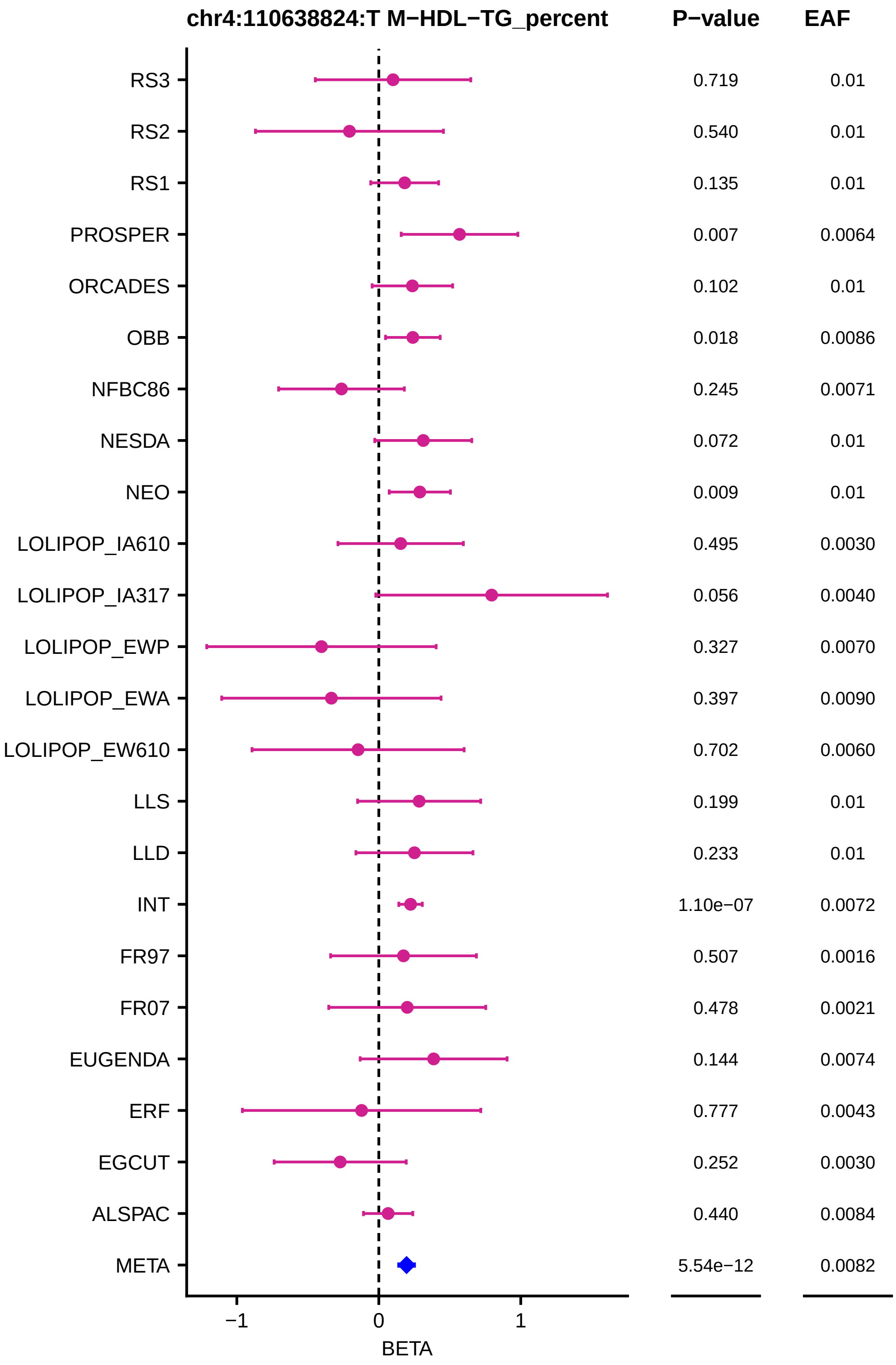
chr4:110638824:T M−HDL−TG_percent
P−value
EAF
RS3
0.719
0.01
RS2
0.540
0.01
RS1
0.135
0.01
PROSPER
0.007
0.0064
ORCADES
0.102
0.01
OBB
0.018
0.0086
NFBC86
0.245
0.0071
NESDA
0.072
0.01
NEO
0.009
0.01
LOLIPOP_IA610
0.495
0.0030
LOLIPOP_IA317
0.056
0.0040
LOLIPOP_EWP
0.327
0.0070
LOLIPOP_EWA
0.397
0.0090
LOLIPOP_EW610
0.702
0.0060
LLS
0.199
0.01
LLD
0.233
0.01
INT
1.10e−07
0.0072
FR97
0.507
0.0016
FR07
0.478
0.0021
EUGENDA
0.144
0.0074
ERF
0.777
0.0043
EGCUT
0.252
0.0030
ALSPAC
0.440
0.0084
META
5.54e−12
0.0082
−1
0
1
BETA

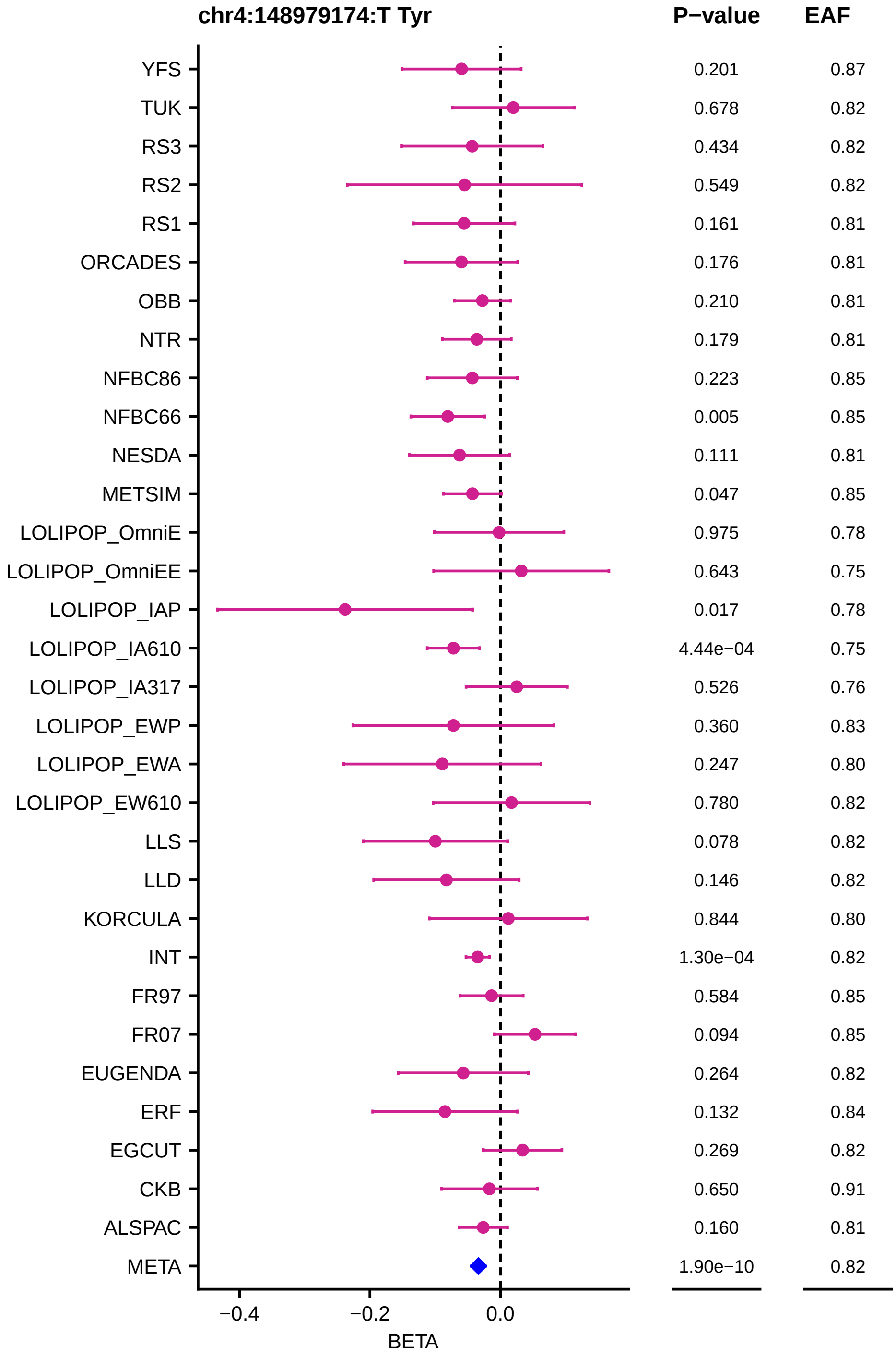
chr4:148979174:T Tyr
P−value
EAF
YFS
TUK
RS3
RS2
RS1
ORCADES
OBB
NTR
NFBC86
NFBC66
NESDA
METSIM
LOLIPOP_OmniE
LOLIPOP_OmniEE
LOLIPOP_IAP
LOLIPOP_IA610
LOLIPOP_IA317
LOLIPOP_EWP
LOLIPOP_EWA
LOLIPOP_EW610
LLS
LLD
KORCULA
INT
FR97
FR07
EUGENDA
ERF
EGCUT
CKB
ALSPAC
META
0.201 0.87
0.678 0.82
0.434 0.82
0.549 0.82
0.161 0.81
0.176 0.81
0.210 0.81
0.179 0.81
0.223 0.85
0.005 0.85
0.111 0.81
0.047 0.85
0.975 0.78
0.643 0.75
0.017 0.78
4.44e−04 0.75
0.526 0.76
0.360 0.83
0.247 0.80
0.780 0.82
0.078 0.82
0.146 0.82
0.844 0.80
1.30e−04 0.82
0.584 0.85
0.094 0.85
0.264 0.82
0.132 0.84
0.269 0.82
0.650 0.91
0.160 0.81
1.90e−10 0.82
−0.4
−0.2
0.0
BETA

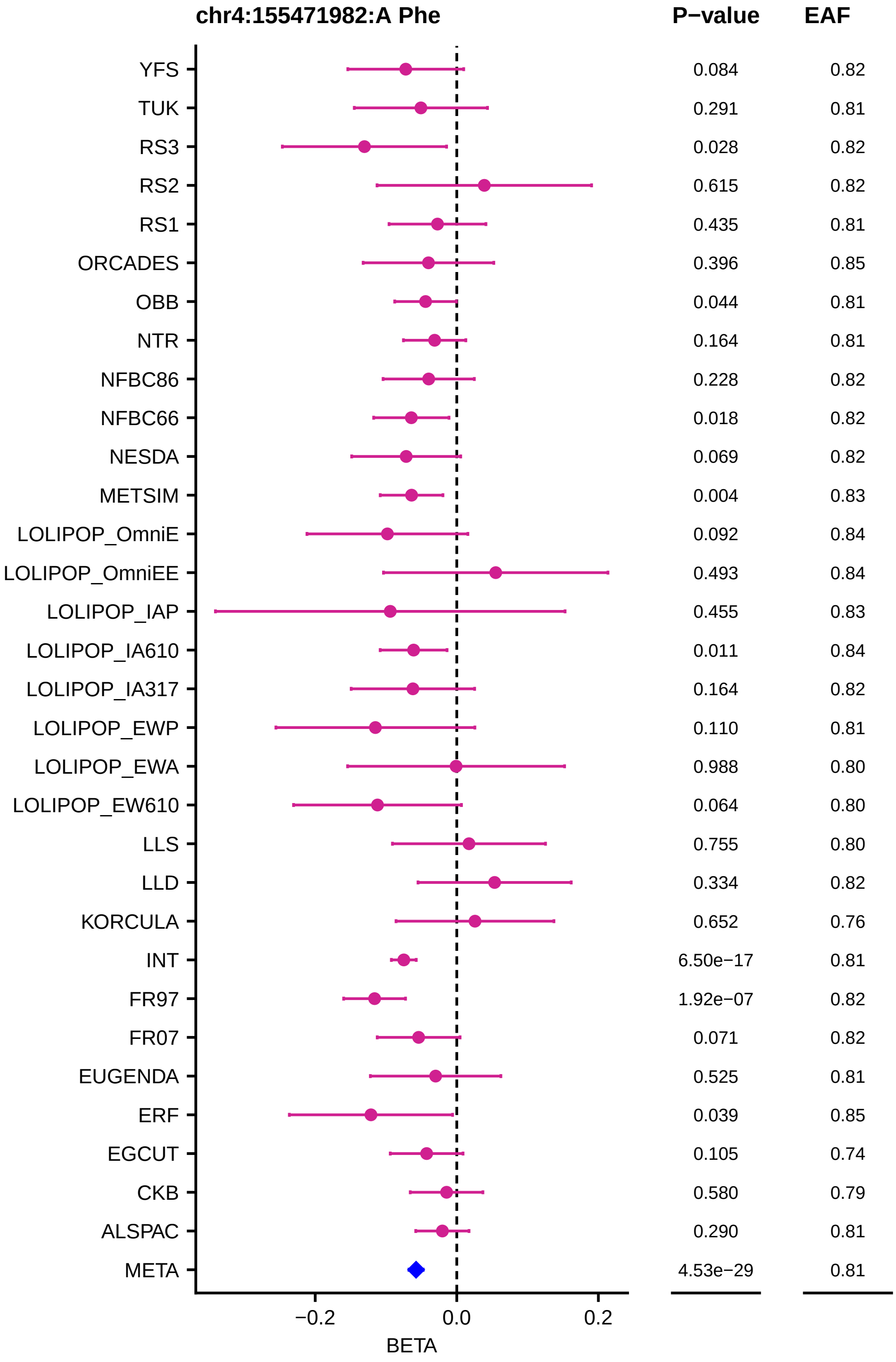
chr4:155471982:A Phe
P−value
EAF
YFS 0.084 0.82
TUK 0.291 0.81
RS3 0.028 0.82
RS2 0.615 0.82
RS1 0.435 0.81
ORCADES 0.396 0.85
OBB 0.044 0.81
NTR 0.164 0.81
NFBC86 0.228 0.82
NFBC66 0.018 0.82
NESDA 0.069 0.82
METSIM 0.004 0.83
LOLIPOP_OmniE 0.092 0.84
LOLIPOP_OmniEE 0.493 0.84
LOLIPOP_IAP 0.455 0.83
LOLIPOP_IA610 0.011 0.84
LOLIPOP_IA317 0.164 0.82
LOLIPOP_EWP 0.110 0.81
LOLIPOP_EWA 0.988 0.80
LOLIPOP_EW610 0.064 0.80
LLS 0.755 0.80
LLD 0.334 0.82
KORCULA 0.652 0.76
INT 6.50e−17 0.81
FR97 1.92e−07 0.82
FR07 0.071 0.82
EUGENDA 0.525 0.81
ERF 0.039 0.85
EGCUT 0.105 0.74
CKB 0.580 0.79
ALSPAC 0.290 0.81
META 4.53e−29 0.81
−0.2
0.0
0.2
BETA

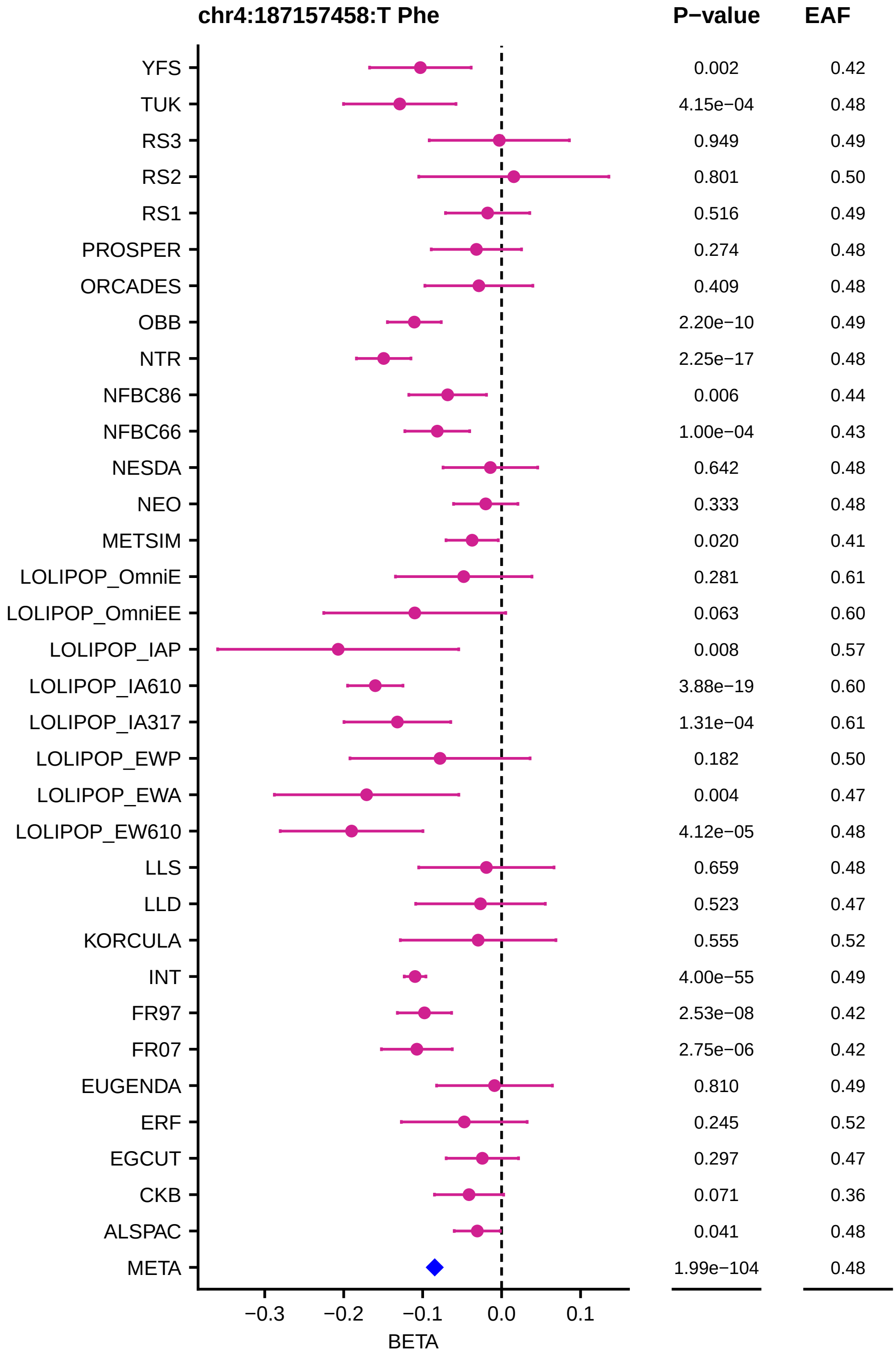
chr4:187157458:T Phe
P−value
EAF
YFS 0.002 0.42
TUK 4.15e−04 0.48
RS3 0.949 0.49
RS2 0.801 0.50
RS1 0.516 0.49
PROSPER 0.274 0.48
ORCADES 0.409 0.48
OBB 2.20e−10 0.49
NTR 2.25e−17 0.48
NFBC86 0.006 0.44
NFBC66 1.00e−04 0.43
NESDA 0.642 0.48
NEO 0.333 0.48
METSIM 0.020 0.41
LOLIPOP_OmniE 0.281 0.61
LOLIPOP_OmniEE 0.063 0.60
LOLIPOP_IAP 0.008 0.57
LOLIPOP_IA610 3.88e−19 0.60
LOLIPOP_IA317 1.31e−04 0.61
LOLIPOP_EWP 0.182 0.50
LOLIPOP_EWA 0.004 0.47
LOLIPOP_EW610 4.12e−05 0.48
LLS 0.659 0.48
LLD 0.523 0.47
KORCULA 0.555 0.52
INT 4.00e−55 0.49
FR97 2.53e−08 0.42
FR07 2.75e−06 0.42
EUGENDA 0.810 0.49
ERF 0.245 0.52
EGCUT 0.297 0.47
CKB 0.071 0.36
ALSPAC 0.041 0.48
META 1.99e−104 0.48
−0.3
−0.2
−0.1
0.0
0.1
BETA

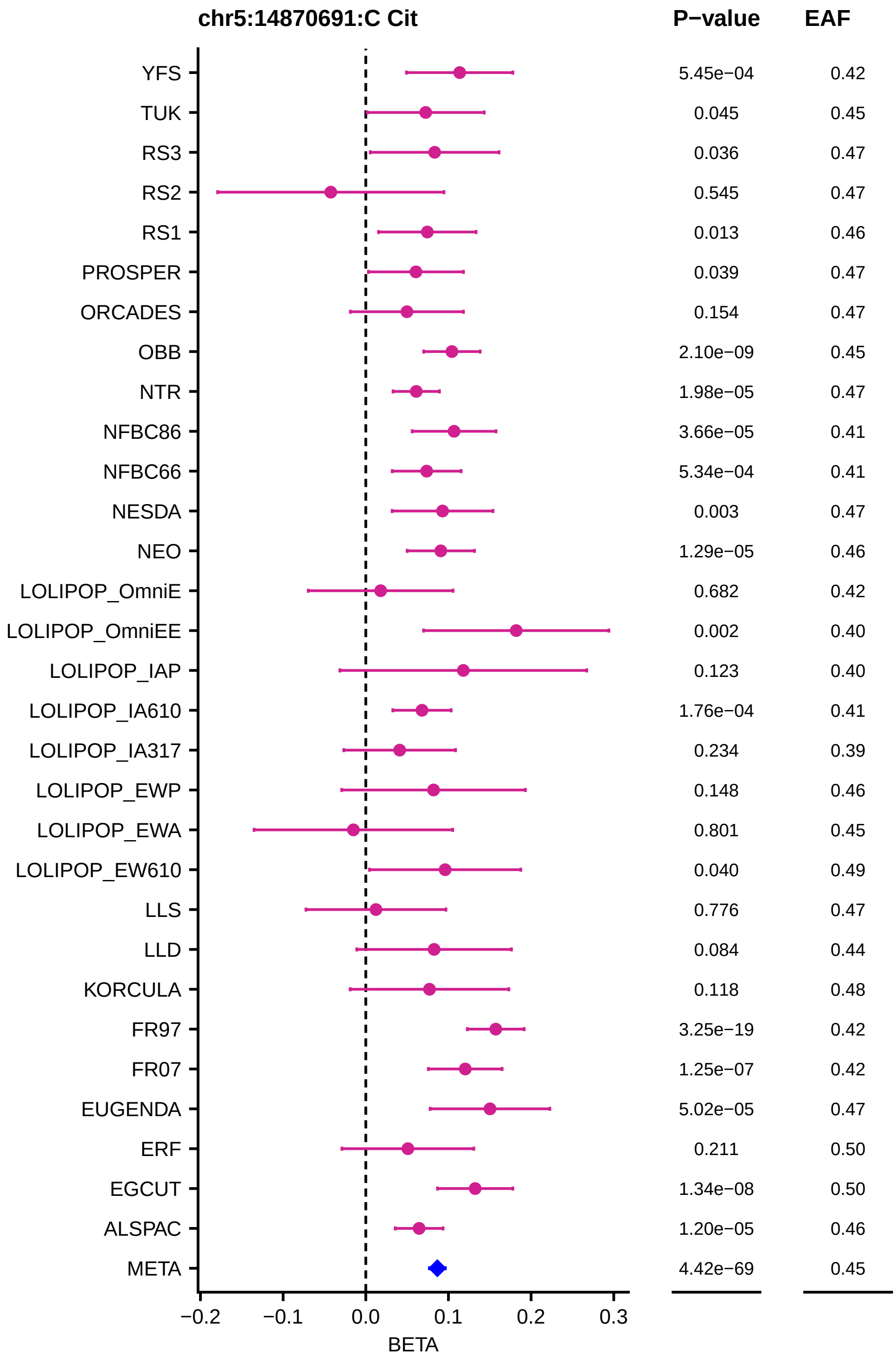
chr5:14870691:C Cit
P−value
EAF
YFS 5.45e−04 0.42
TUK 0.045 0.45
RS3 0.036 0.47
RS2 0.545 0.47
RS1 0.013 0.46
PROSPER 0.039 0.47
ORCADES 0.154 0.47
OBB 2.10e−09 0.45
NTR 1.98e−05 0.47
NFBC86 3.66e−05 0.41
NFBC66 5.34e−04 0.41
NESDA 0.003 0.47
NEO 1.29e−05 0.46
LOLIPOP_OmniE 0.682 0.42
LOLIPOP_OmniEE 0.002 0.40
LOLIPOP_IAP 0.123 0.40
LOLIPOP_IA610 1.76e−04 0.41
LOLIPOP_IA317 0.234 0.39
LOLIPOP_EWP 0.148 0.46
LOLIPOP_EWA 0.801 0.45
LOLIPOP_EW610 0.040 0.49
LLS 0.776 0.47
LLD 0.084 0.44
KORCULA 0.118 0.48
FR97 3.25e−19 0.42
FR07 1.25e−07 0.42
EUGENDA 5.02e−05 0.47
ERF 0.211 0.50
EGCUT 1.34e−08 0.50
ALSPAC 1.20e−05 0.46
META 4.42e−69 0.45
−0.2 −0.1 0.0 0.1 0.2 0.3
BETA

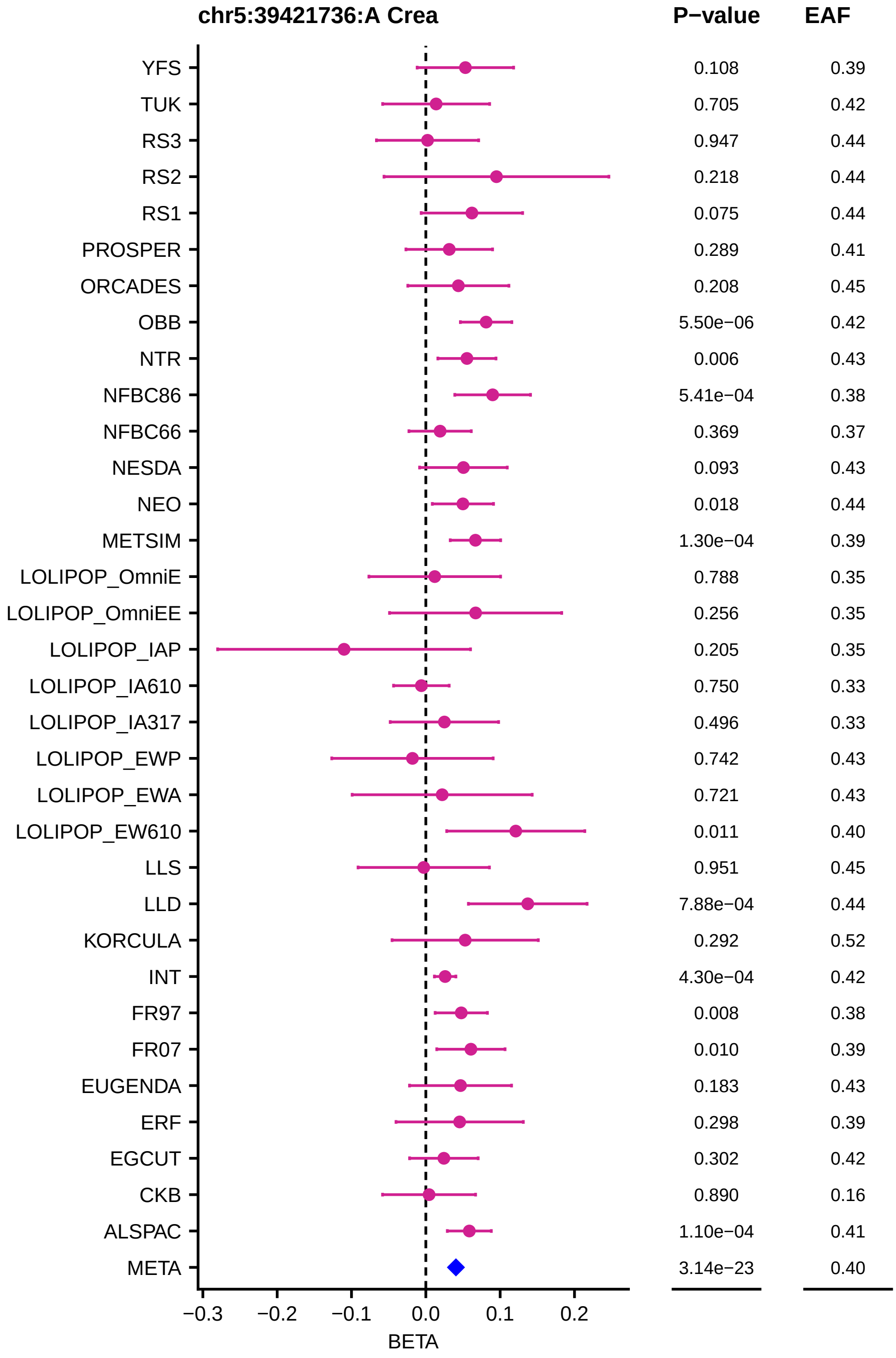
chr5:39421736:A Crea
P−value
EAF
YFS 0.108 0.39
TUK 0.705 0.42
RS3 0.947 0.44
RS2 0.218 0.44
RS1 0.075 0.44
PROSPER 0.289 0.41
ORCADES 0.208 0.45
OBB 5.50e−06 0.42
NTR 0.006 0.43
NFBC86 5.41e−04 0.38
NFBC66 0.369 0.37
NESDA 0.093 0.43
NEO 0.018 0.44
METSIM 1.30e−04 0.39
LOLIPOP_OmniE 0.788 0.35
LOLIPOP_OmniEE 0.256 0.35
LOLIPOP_IAP 0.205 0.35
LOLIPOP_IA610 0.750 0.33
LOLIPOP_IA317 0.496 0.33
LOLIPOP_EWP 0.742 0.43
LOLIPOP_EWA 0.721 0.43
LOLIPOP_EW610 0.011 0.40
LLS 0.951 0.45
LLD 7.88e−04 0.44
KORCULA 0.292 0.52
INT 4.30e−04 0.42
FR97 0.008 0.38
FR07 0.010 0.39
EUGENDA 0.183 0.43
ERF 0.298 0.39
EGCUT 0.302 0.42
CKB 0.890 0.16
ALSPAC 1.10e−04 0.41
META 3.14e−23 0.40
−0.3 −0.2 −0.1 0.0 0.1 0.2
BETA

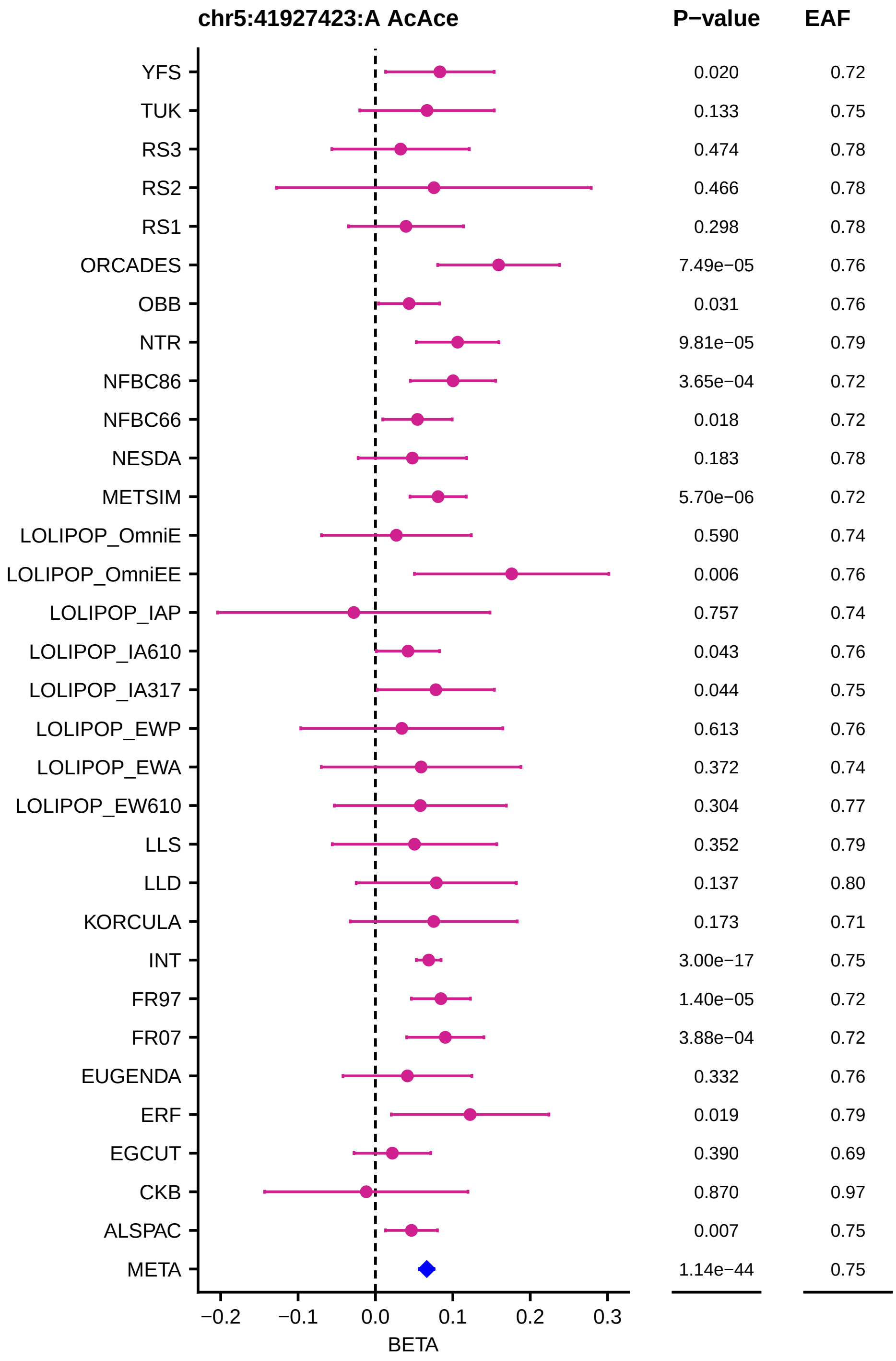
chr5:41927423:A AcAce
P−value
EAF
YFS 0.020 0.72
TUK 0.133 0.75
RS3 0.474 0.78
RS2 0.466 0.78
RS1 0.298 0.78
ORCADES 7.49e−05 0.76
OBB 0.031 0.76
NTR 9.81e−05 0.79
NFBC86 3.65e−04 0.72
NFBC66 0.018 0.72
NESDA 0.183 0.78
METSIM 5.70e−06 0.72
LOLIPOP_OmniE 0.590 0.74
LOLIPOP_OmniEE 0.006 0.76
LOLIPOP_IAP 0.757 0.74
LOLIPOP_IA610 0.043 0.76
LOLIPOP_IA317 0.044 0.75
LOLIPOP_EWP 0.613 0.76
LOLIPOP_EWA 0.372 0.74
LOLIPOP_EW610 0.304 0.77
LLS 0.352 0.79
LLD 0.137 0.80
KORCULA 0.173 0.71
INT 3.00e−17 0.75
FR97 1.40e−05 0.72
FR07 3.88e−04 0.72
EUGENDA 0.332 0.76
ERF 0.019 0.79
EGCUT 0.390 0.69
CKB 0.870 0.97
ALSPAC 0.007 0.75
META 1.14e−44 0.75
−0.2
−0.1
0.0
0.1
0.2
0.3
BETA

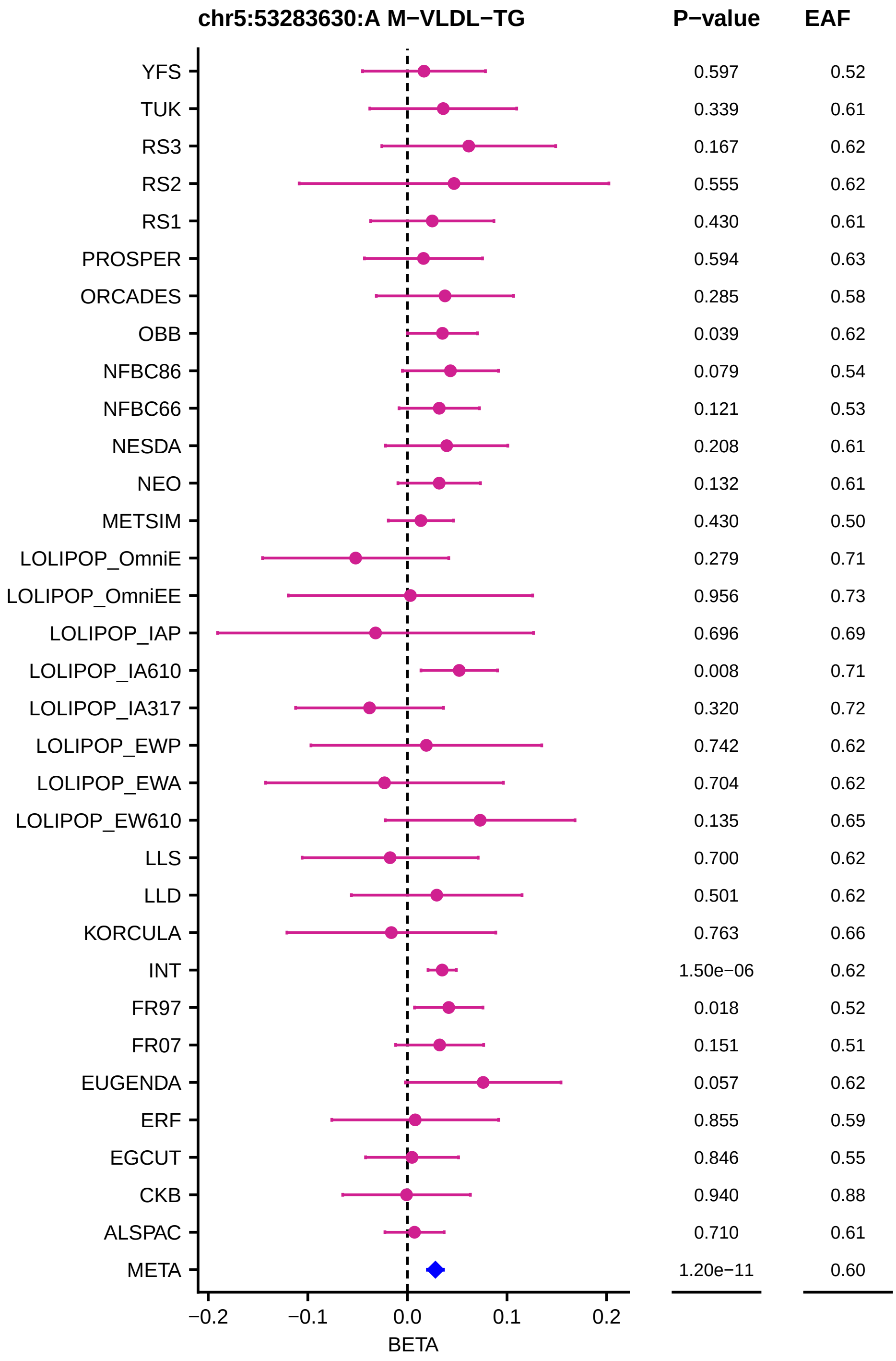
chr5:53283630:A M−VLDL−TG
P−value
EAF
YFS
TUK
RS3
RS2
RS1
PROSPER
ORCADES
OBB
NFBC86
NFBC66
NESDA
NEO
METSIM
LOLIPOP_OmniE
LOLIPOP_OmniEE
LOLIPOP_IAP
LOLIPOP_IA610
LOLIPOP_IA317
LOLIPOP_EWP
LOLIPOP_EWA
LOLIPOP_EW610
LLS
LLD
KORCULA
INT
FR97
FR07
EUGENDA
ERF
EGCUT
CKB
ALSPAC
META
0.597 0.52
0.339 0.61
0.167 0.62
0.555 0.62
0.430 0.61
0.594 0.63
0.285 0.58
0.039 0.62
0.079 0.54
0.121 0.53
0.208 0.61
0.132 0.61
0.430 0.50
0.279 0.71
0.956 0.73
0.696 0.69
0.008 0.71
0.320 0.72
0.742 0.62
0.704 0.62
0.135 0.65
0.700 0.62
0.501 0.62
0.763 0.66
1.50e−06 0.62
0.018 0.52
0.151 0.51
0.057 0.62
0.855 0.59
0.846 0.55
0.940 0.88
0.710 0.61
1.20e−11 0.60
−0.2
−0.1
0.0
0.1
0.2
BETA

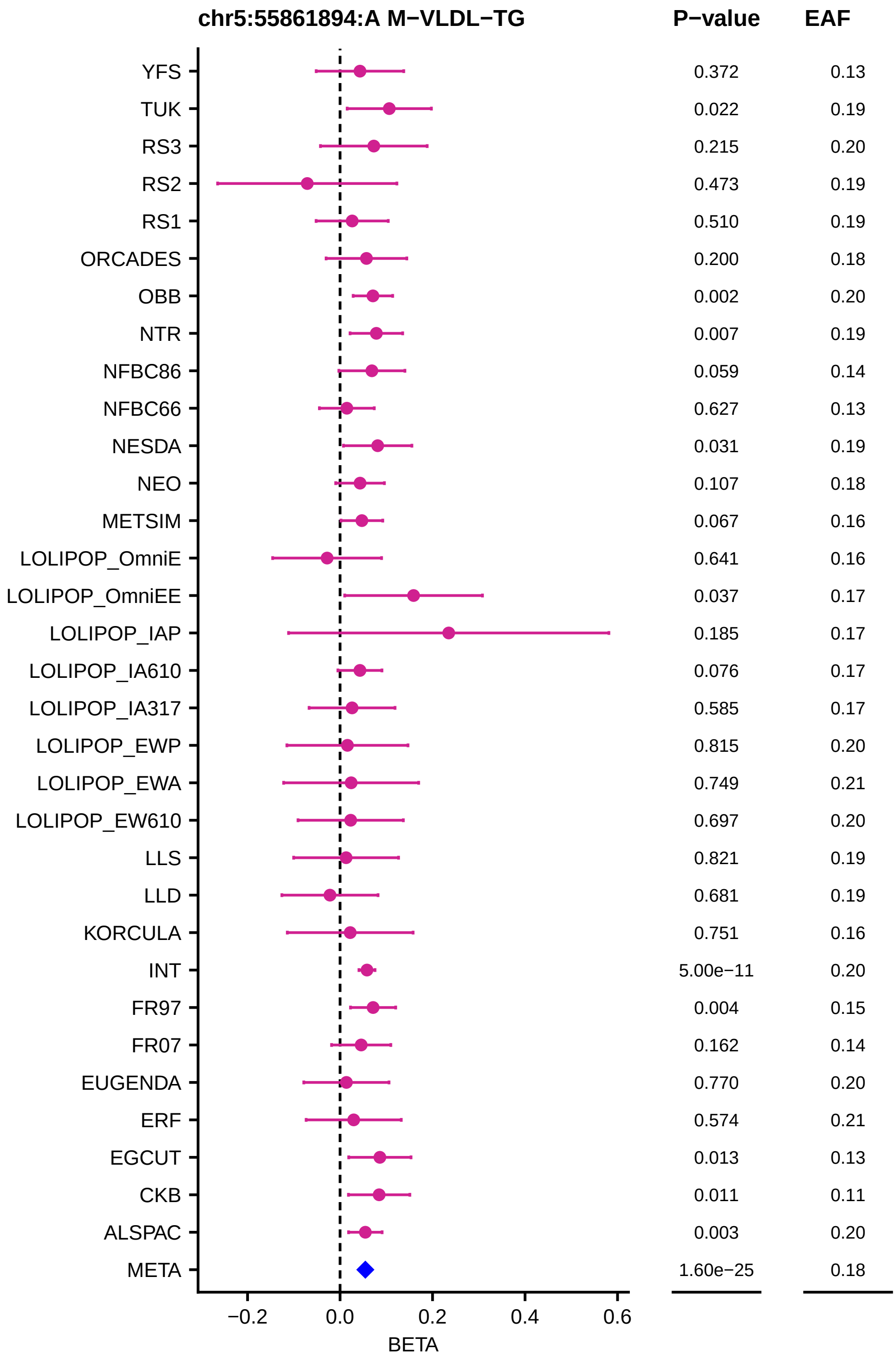
chr5:55861894:A M−VLDL−TG
P−value
EAF
YFS
TUK
RS3
RS2
RS1
ORCADES
OBB
NTR
NFBC86
NFBC66
NESDA
NEO
METSIM
LOLIPOP_OmniE
LOLIPOP_OmniEE
LOLIPOP_IAP
LOLIPOP_IA610
LOLIPOP_IA317
LOLIPOP_EWP
LOLIPOP_EWA
LOLIPOP_EW610
LLS
LLD
KORCULA
INT
FR97
FR07
EUGENDA
ERF
EGCUT
CKB
ALSPAC
META
0.372 0.13
0.022 0.19
0.215 0.20
0.473 0.19
0.510 0.19
0.200 0.18
0.002 0.20
0.007 0.19
0.059 0.14
0.627 0.13
0.031 0.19
0.107 0.18
0.067 0.16
0.641 0.16
0.037 0.17
0.185 0.17
0.076 0.17
0.585 0.17
0.815 0.20
0.749 0.21
0.697 0.20
0.821 0.19
0.681 0.19
0.751 0.16
5.00e−11 0.20
0.004 0.15
0.162 0.14
0.770 0.20
0.574 0.21
0.013 0.13
0.011 0.11
0.003 0.20
1.60e−25 0.18
−0.2
0.0
0.2
0.4
0.6
BETA

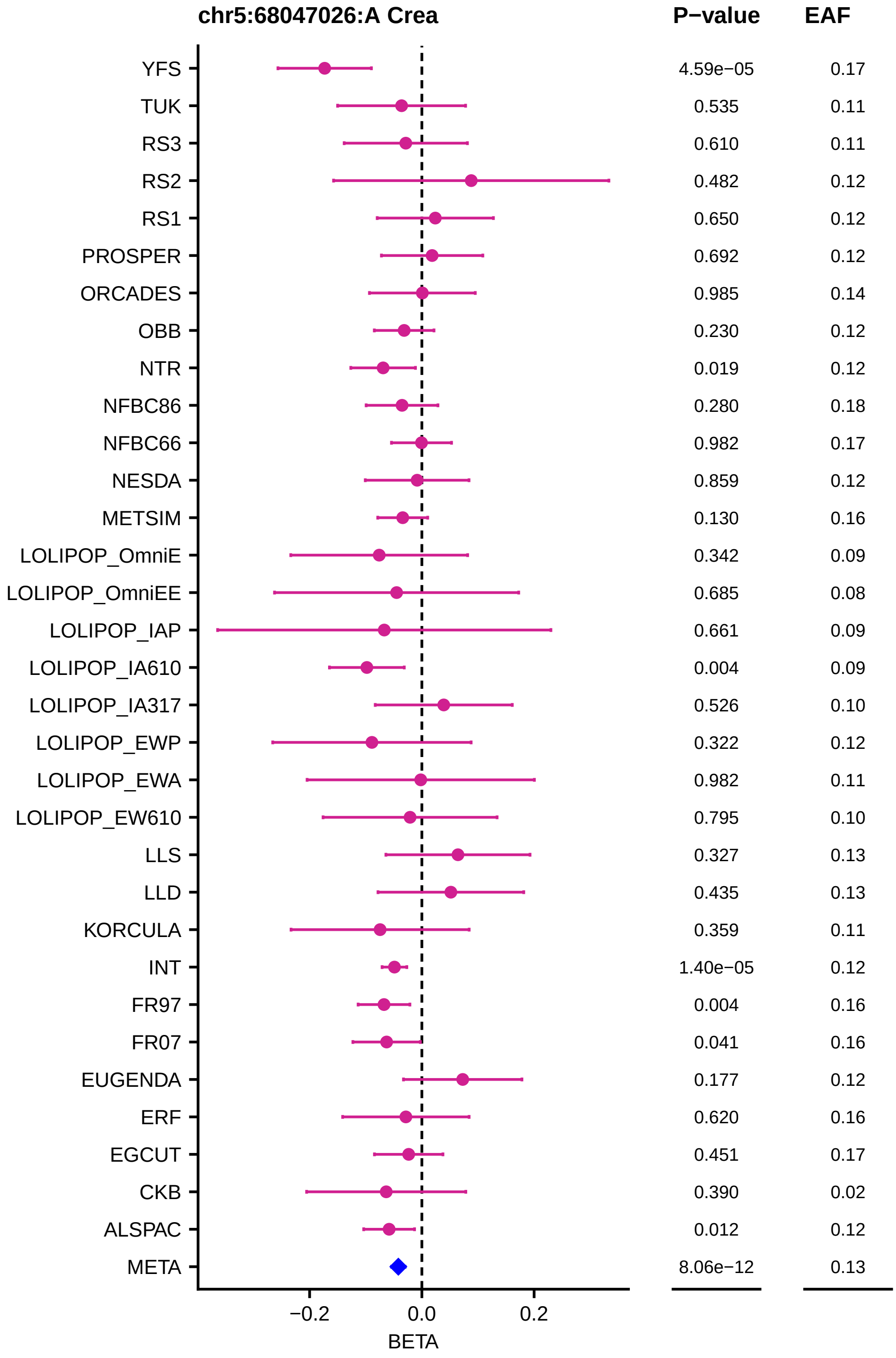

chr5:68047026:A Crea
P−value
EAF
YFS 4.59e−05 0.17
TUK 0.535 0.11
RS3 0.610 0.11
RS2 0.482 0.12
RS1 0.650 0.12
PROSPER 0.692 0.12
ORCADES 0.985 0.14
OBB 0.230 0.12
NTR 0.019 0.12
NFBC86 0.280 0.18
NFBC66 0.982 0.17
NESDA 0.859 0.12
METSIM 0.130 0.16
LOLIPOP_OmniE 0.342 0.09
LOLIPOP_OmniEE 0.685 0.08
LOLIPOP_IAP 0.661 0.09
LOLIPOP_IA610 0.004 0.09
LOLIPOP_IA317 0.526 0.10
LOLIPOP_EWP 0.322 0.12
LOLIPOP_EWA 0.982 0.11
LOLIPOP_EW610 0.795 0.10
LLS 0.327 0.13
LLD 0.435 0.13
KORCULA 0.359 0.11
INT 1.40e−05 0.12
FR97 0.004 0.16
FR07 0.041 0.16
EUGENDA 0.177 0.12
ERF 0.620 0.16
EGCUT 0.451 0.17
CKB 0.390 0.02
ALSPAC 0.012 0.12
META 8.06e−12 0.13
−0.2
0.0
0.2
BETA

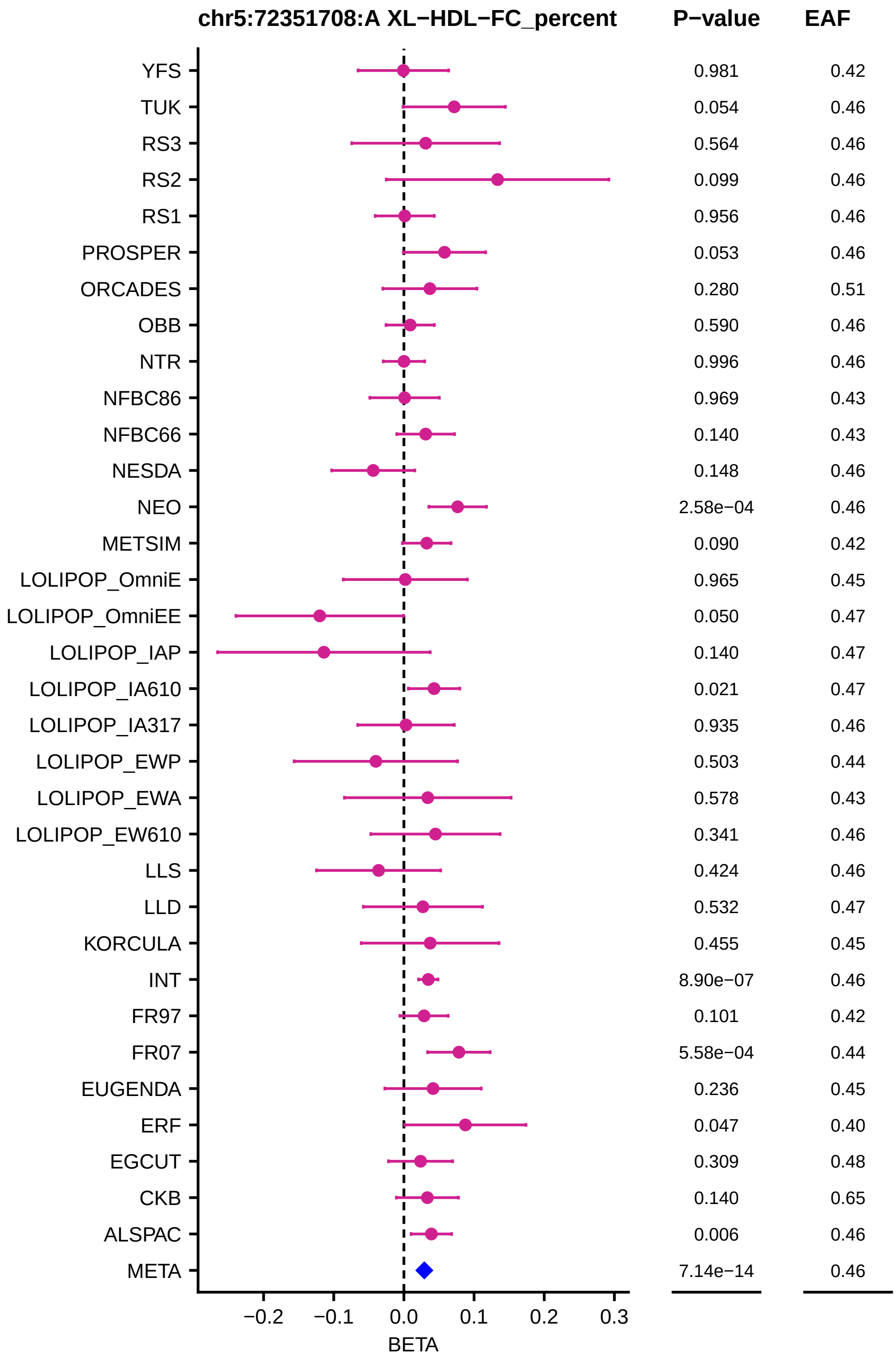
chr5:72351708:A XL−HDL−FC_percent
P−value
EAF
YFS 0.981 0.42
TUK 0.054 0.46
RS3 0.564 0.46
RS2 0.099 0.46
RS1 0.956 0.46
PROSPER 0.053 0.46
ORCADES 0.280 0.51
OBB 0.590 0.46
NTR 0.996 0.46
NFBC86 0.969 0.43
NFBC66 0.140 0.43
NESDA 0.148 0.46
NEO 2.58e−04 0.46
METSIM 0.090 0.42
LOLIPOP_OmniE 0.965 0.45
LOLIPOP_OmniEE 0.050 0.47
LOLIPOP_IAP 0.140 0.47
LOLIPOP_IA610 0.021 0.47
LOLIPOP_IA317 0.935 0.46
LOLIPOP_EWP 0.503 0.44
LOLIPOP_EWA 0.578 0.43
LOLIPOP_EW610 0.341 0.46
LLS 0.424 0.46
LLD 0.532 0.47
KORCULA 0.455 0.45
INT 8.90e−07 0.46
FR97 0.101 0.42
FR07 5.58e−04 0.44
EUGENDA 0.236 0.45
ERF 0.047 0.40
EGCUT 0.309 0.48
CKB 0.140 0.65
ALSPAC 0.006 0.46
META 7.14e−14 0.46
−0.2 −0.1 0.0 0.1 0.2 0.3
BETA

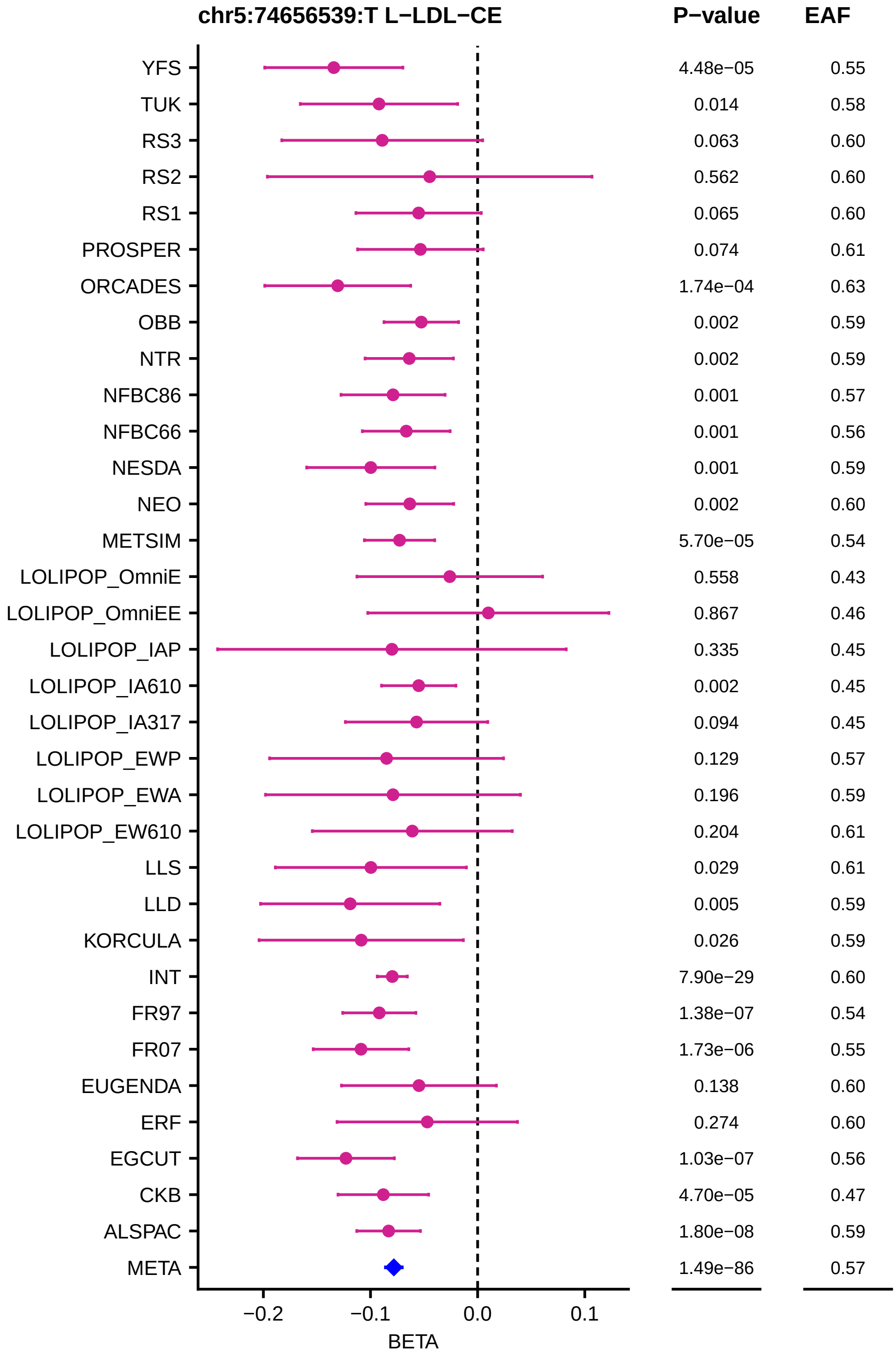
chr5:74656539:T L−LDL−CE
P−value
EAF
YFS
TUK
RS3
RS2
RS1
PROSPER
ORCADES
OBB
NTR
NFBC86
NFBC66
NESDA
NEO
METSIM
LOLIPOP_OmniE
LOLIPOP_OmniEE
LOLIPOP_IAP
LOLIPOP_IA610
LOLIPOP_IA317
LOLIPOP_EWP
LOLIPOP_EWA
LOLIPOP_EW610
LLS
LLD
KORCULA
INT
FR97
FR07
EUGENDA
ERF
EGCUT
CKB
ALSPAC
META
4.48e−05
0.014
0.063
0.562
0.065
0.074
1.74e−04
0.002
0.002
0.001
0.001
0.001
0.002
5.70e−05
0.558
0.867
0.335
0.002
0.094
0.129
0.196
0.204
0.029
0.005
0.026
7.90e−29
1.38e−07
1.73e−06
0.138
0.274
1.03e−07
4.70e−05
1.80e−08
1.49e−86
0.55
0.58
0.60
0.60
0.60
0.61
0.63
0.59
0.59
0.57
0.56
0.59
0.60
0.54
0.43
0.46
0.45
0.45
0.45
0.57
0.59
0.61
0.61
0.59
0.59
0.60
0.54
0.55
0.60
0.60
0.56
0.47
0.59
0.57
−0.2
−0.1
0.0
0.1
BETA

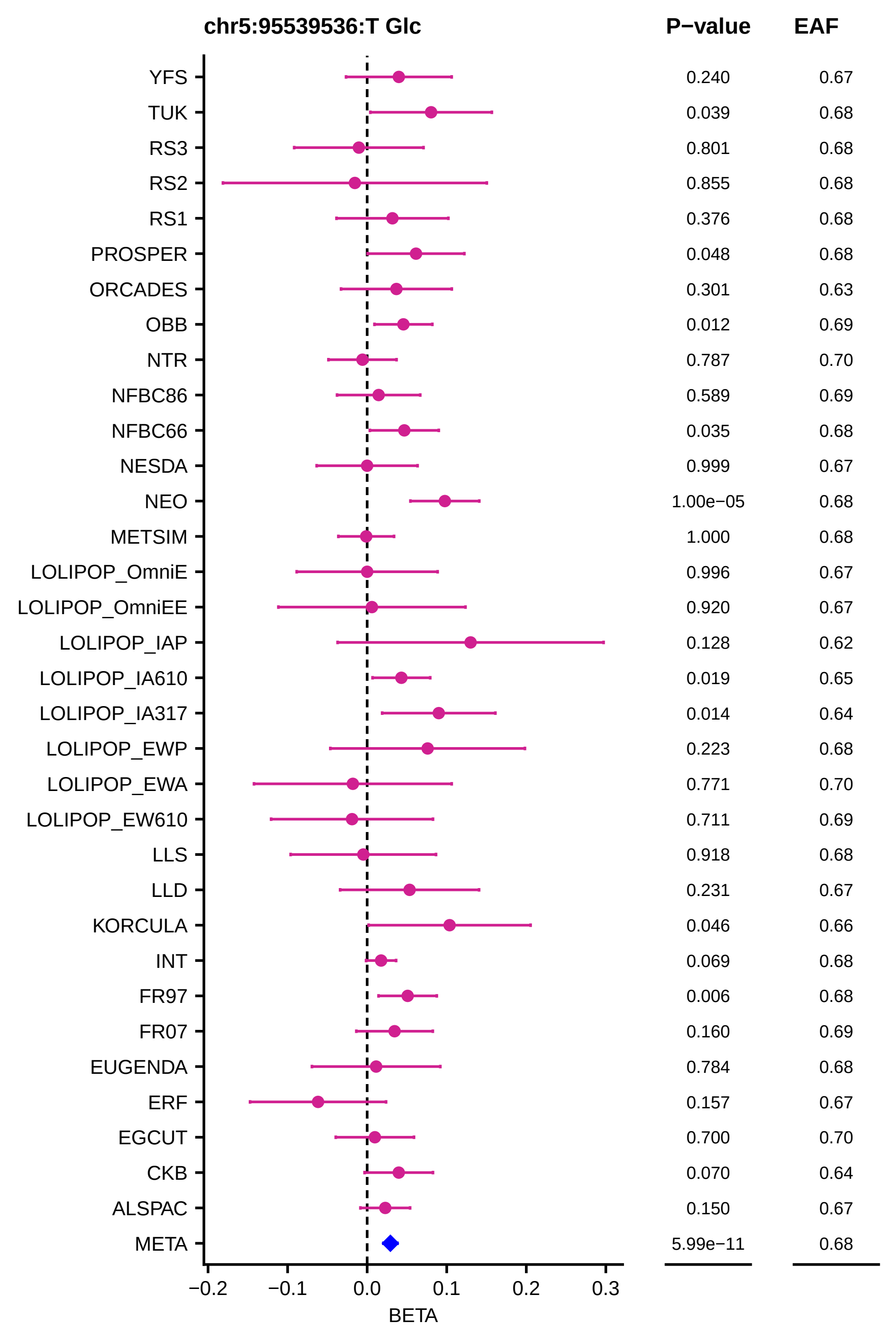

chr5:95539536:T Glc
P−value
EAF
YFS 0.240 0.67
TUK 0.039 0.68
RS3 0.801 0.68
RS2 0.855 0.68
RS1 0.376 0.68
PROSPER 0.048 0.68
ORCADES 0.301 0.63
OBB 0.012 0.69
NTR 0.787 0.70
NFBC86 0.589 0.69
NFBC66 0.035 0.68
NESDA 0.999 0.67
NEO 1.00e−05 0.68
METSIM 1.000 0.68
LOLIPOP_OmniE 0.996 0.67
LOLIPOP_OmniEE 0.920 0.67
LOLIPOP_IAP 0.128 0.62
LOLIPOP_IA610 0.019 0.65
LOLIPOP_IA317 0.014 0.64
LOLIPOP_EWP 0.223 0.68
LOLIPOP_EWA 0.771 0.70
LOLIPOP_EW610 0.711 0.69
LLS 0.918 0.68
LLD 0.231 0.67
KORCULA 0.046 0.66
INT 0.069 0.68
FR97 0.006 0.68
FR07 0.160 0.69
EUGENDA 0.784 0.68
ERF 0.157 0.67
EGCUT 0.700 0.70
CKB 0.070 0.64
ALSPAC 0.150 0.67
META 5.99e−11 0.68
−0.2 −0.1 0.0 0.1 0.2 0.3
BETA

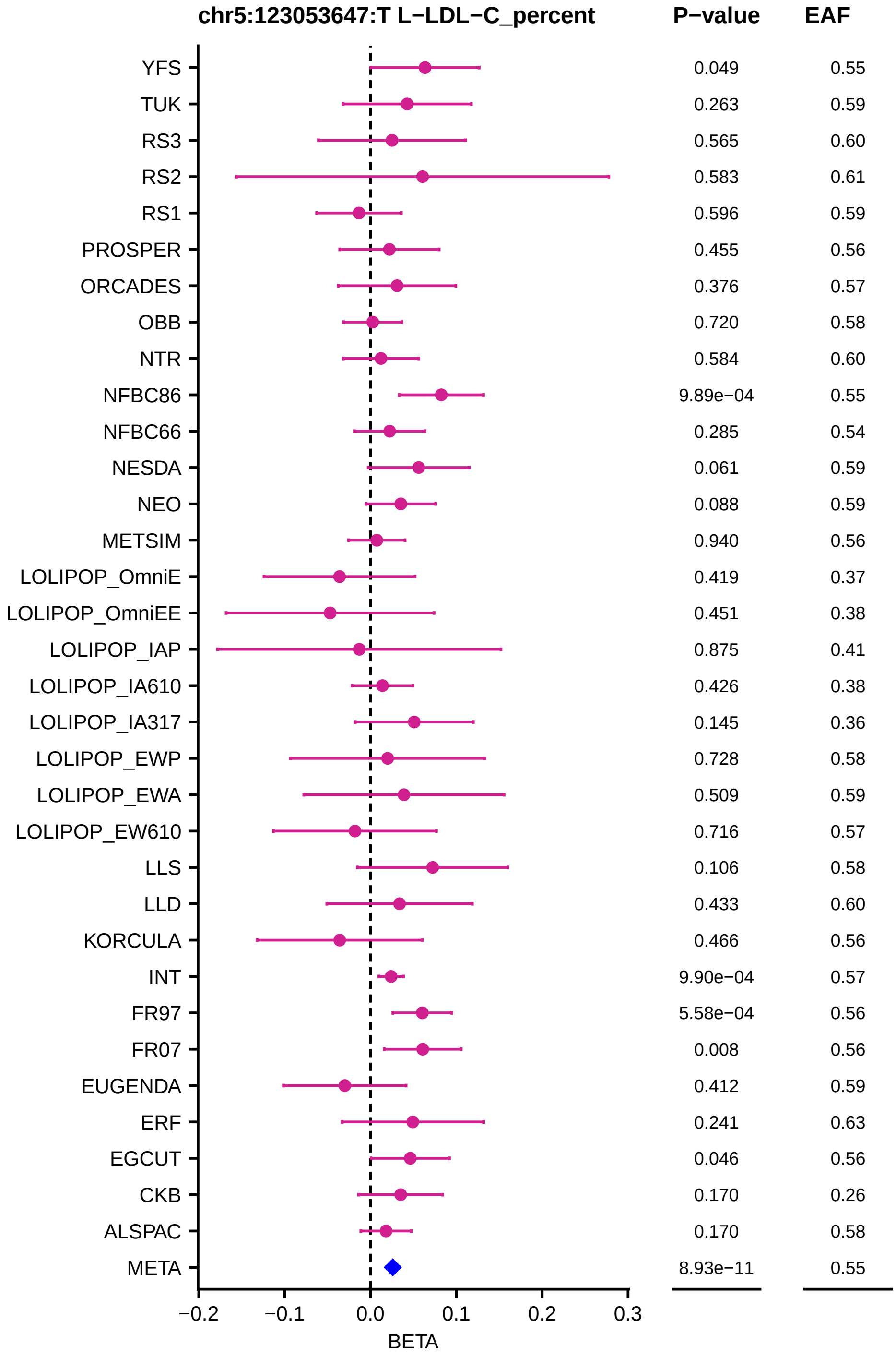
chr5:123053647:T L−LDL−C_percent
P−value
EAF
YFS 0.049 0.55
TUK 0.263 0.59
RS3 0.565 0.60
RS2 0.583 0.61
RS1 0.596 0.59
PROSPER 0.455 0.56
ORCADES 0.376 0.57
OBB 0.720 0.58
NTR 0.584 0.60
NFBC86 9.89e−04 0.55
NFBC66 0.285 0.54
NESDA 0.061 0.59
NEO 0.088 0.59
METSIM 0.940 0.56
LOLIPOP_OmniE 0.419 0.37
LOLIPOP_OmniEE 0.451 0.38
LOLIPOP_IAP 0.875 0.41
LOLIPOP_IA610 0.426 0.38
LOLIPOP_IA317 0.145 0.36
LOLIPOP_EWP 0.728 0.58
LOLIPOP_EWA 0.509 0.59
LOLIPOP_EW610 0.716 0.57
LLS 0.106 0.58
LLD 0.433 0.60
KORCULA 0.466 0.56
INT 9.90e−04 0.57
FR97 5.58e−04 0.56
FR07 0.008 0.56
EUGENDA 0.412 0.59
ERF 0.241 0.63
EGCUT 0.046 0.56
CKB 0.170 0.26
ALSPAC 0.170 0.58
META 8.93e−11 0.55
−0.2 −0.1 0.0 0.1 0.2 0.3
BETA

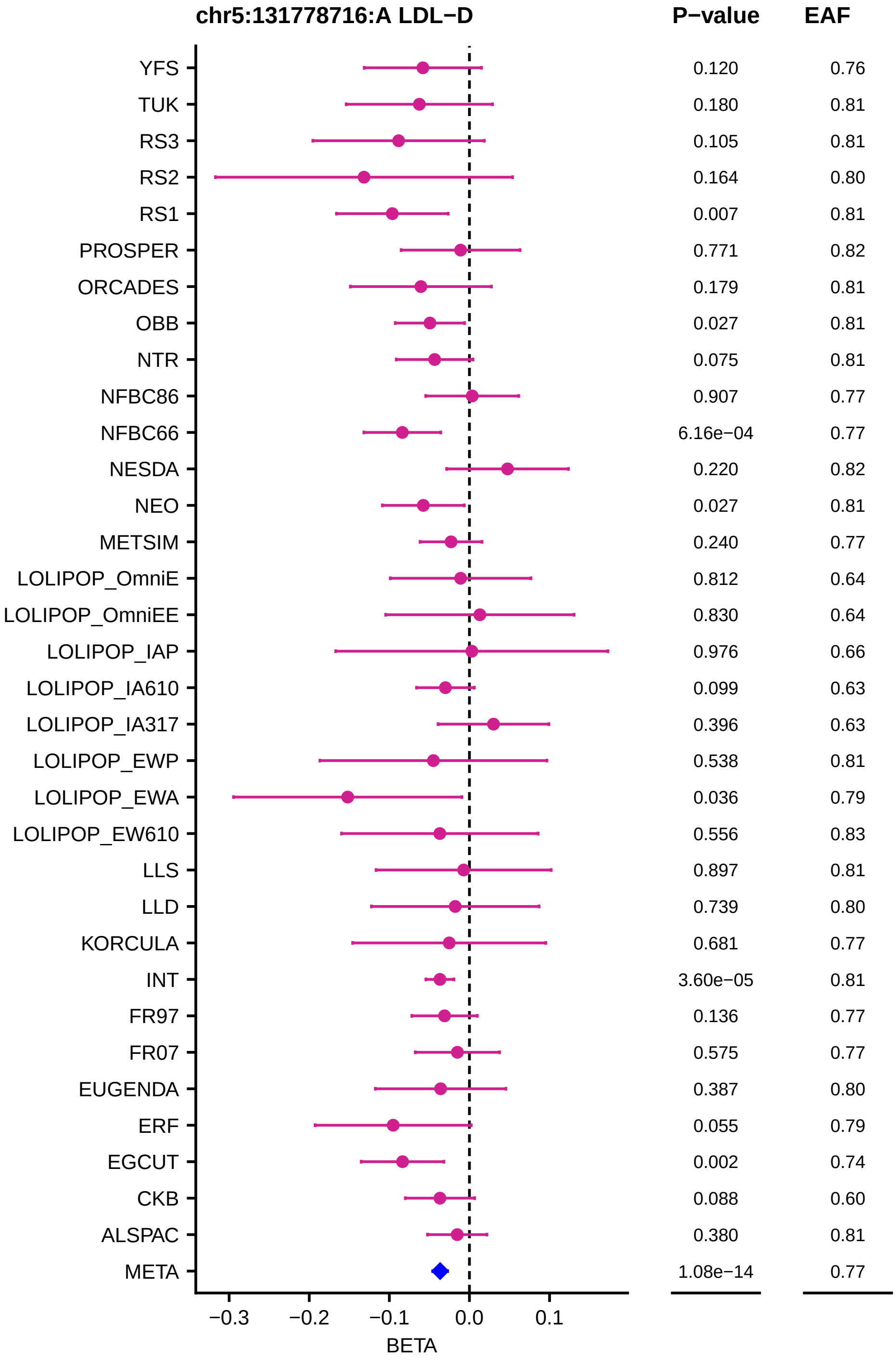
chr5:131778716:A LDL−D
P−value
EAF
YFS 0.120 0.76
TUK 0.180 0.81
RS3 0.105 0.81
RS2 0.164 0.80
RS1 0.007 0.81
PROSPER 0.771 0.82
ORCADES 0.179 0.81
OBB 0.027 0.81
NTR 0.075 0.81
NFBC86 0.907 0.77
NFBC66 6.16e−04 0.77
NESDA 0.220 0.82
NEO 0.027 0.81
METSIM 0.240 0.77
LOLIPOP_OmniE 0.812 0.64
LOLIPOP_OmniEE 0.830 0.64
LOLIPOP_IAP 0.976 0.66
LOLIPOP_IA610 0.099 0.63
LOLIPOP_IA317 0.396 0.63
LOLIPOP_EWP 0.538 0.81
LOLIPOP_EWA 0.036 0.79
LOLIPOP_EW610 0.556 0.83
LLS 0.897 0.81
LLD 0.739 0.80
KORCULA 0.681 0.77
INT 3.60e−05 0.81
FR97 0.136 0.77
FR07 0.575 0.77
EUGENDA 0.387 0.80
ERF 0.055 0.79
EGCUT 0.002 0.74
CKB 0.088 0.60
ALSPAC 0.380 0.81
META 1.08e−14 0.77
−0.3
−0.2
−0.1
0.0
0.1
BETA

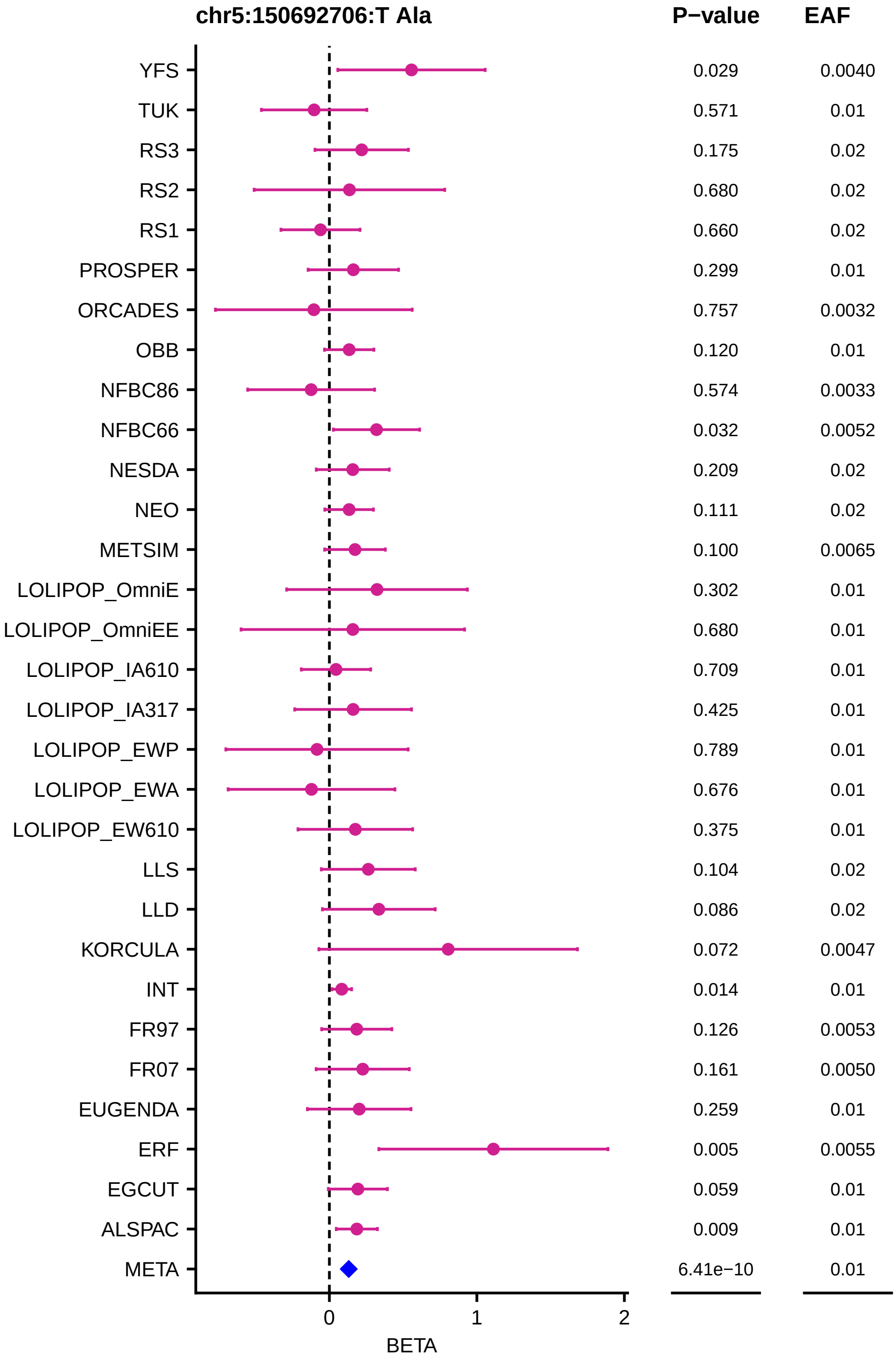
chr5:150692706:T Ala
P−value
EAF
YFS 0.029 0.0040
TUK 0.571 0.01
RS3 0.175 0.02
RS2 0.680 0.02
RS1 0.660 0.02
PROSPER 0.299 0.01
ORCADES 0.757 0.0032
OBB 0.120 0.01
NFBC86 0.574 0.0033
NFBC66 0.032 0.0052
NESDA 0.209 0.02
NEO 0.111 0.02
METSIM 0.100 0.0065
LOLIPOP_OmniE 0.302 0.01
LOLIPOP_OmniEE 0.680 0.01
LOLIPOP_IA610 0.709 0.01
LOLIPOP_IA317 0.425 0.01
LOLIPOP_EWP 0.789 0.01
LOLIPOP_EWA 0.676 0.01
LOLIPOP_EW610 0.375 0.01
LLS 0.104 0.02
LLD 0.086 0.02
KORCULA 0.072 0.0047
INT 0.014 0.01
FR97 0.126 0.0053
FR07 0.161 0.0050
EUGENDA 0.259 0.01
ERF 0.005 0.0055
EGCUT 0.059 0.01
ALSPAC 0.009 0.01
META 6.41e−10 0.01
0
1
2
BETA

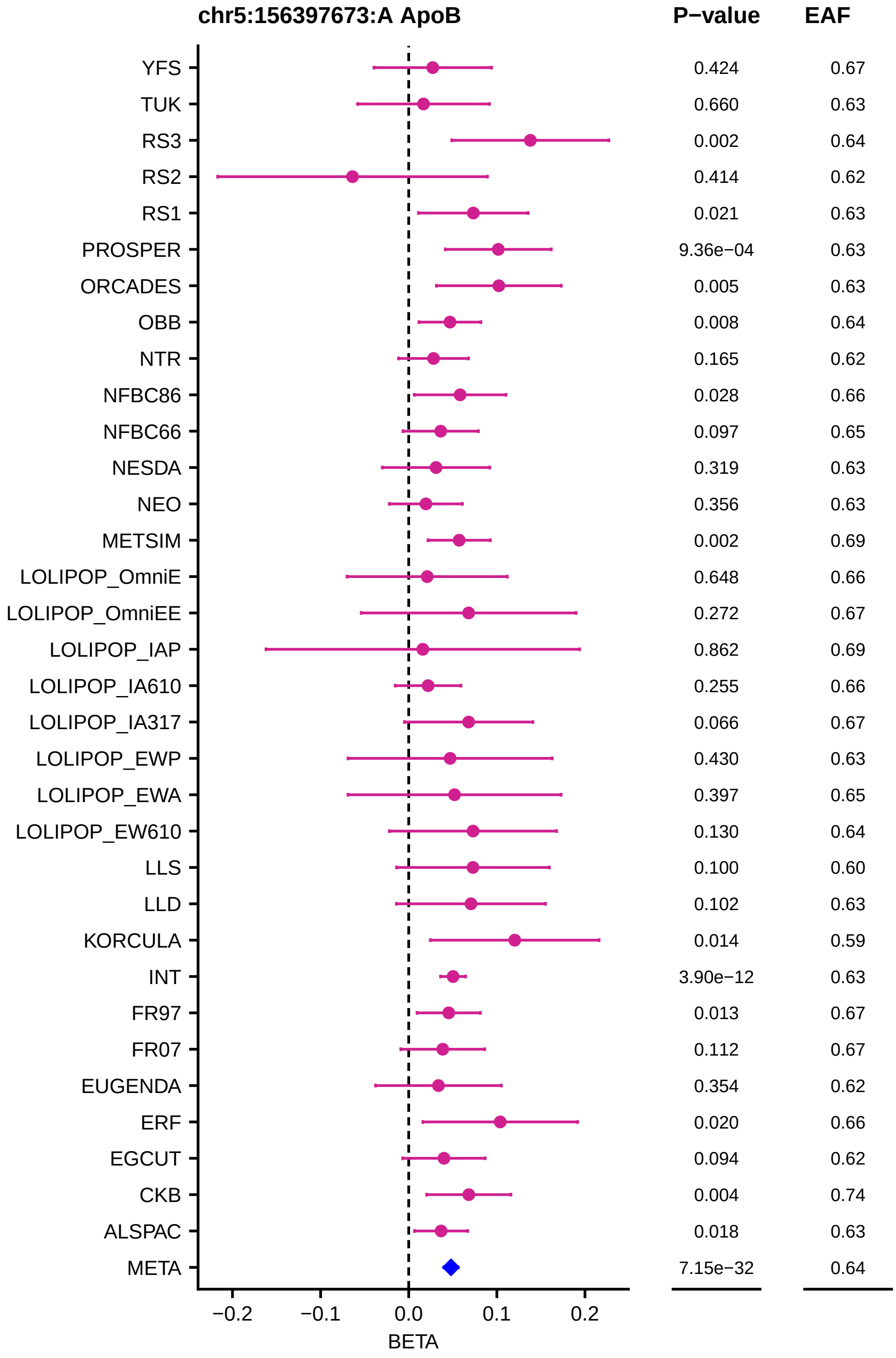

chr5:156397673:A ApoB
P−value
EAF
YFS 0.424 0.67
TUK 0.660 0.63
RS3 0.002 0.64
RS2 0.414 0.62
RS1 0.021 0.63
PROSPER 9.36e−04 0.63
ORCADES 0.005 0.63
OBB 0.008 0.64
NTR 0.165 0.62
NFBC86 0.028 0.66
NFBC66 0.097 0.65
NESDA 0.319 0.63
NEO 0.356 0.63
METSIM 0.002 0.69
LOLIPOP_OmniE 0.648 0.66
LOLIPOP_OmniEE 0.272 0.67
LOLIPOP_IAP 0.862 0.69
LOLIPOP_IA610 0.255 0.66
LOLIPOP_IA317 0.066 0.67
LOLIPOP_EWP 0.430 0.63
LOLIPOP_EWA 0.397 0.65
LOLIPOP_EW610 0.130 0.64
LLS 0.100 0.60
LLD 0.102 0.63
KORCULA 0.014 0.59
INT 3.90e−12 0.63
FR97 0.013 0.67
FR07 0.112 0.67
EUGENDA 0.354 0.62
ERF 0.020 0.66
EGCUT 0.094 0.62
CKB 0.004 0.74
ALSPAC 0.018 0.63
META 7.15e−32 0.64
−0.2
−0.1
0.0
0.1
0.2
BETA

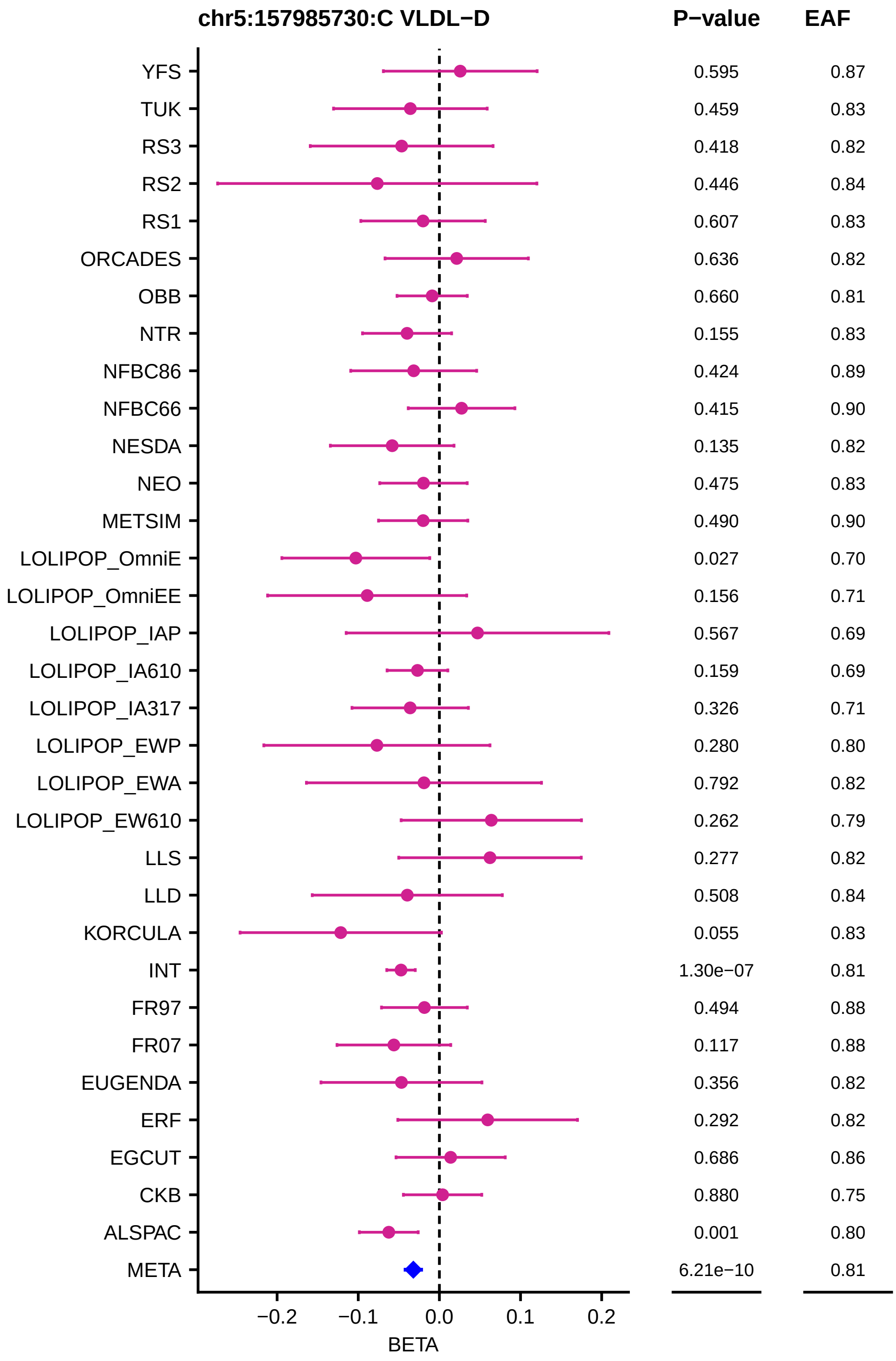
chr5:157985730:C VLDL−D
P−value
EAF
YFS 0.595 0.87
TUK 0.459 0.83
RS3 0.418 0.82
RS2 0.446 0.84
RS1 0.607 0.83
ORCADES 0.636 0.82
OBB 0.660 0.81
NTR 0.155 0.83
NFBC86 0.424 0.89
NFBC66 0.415 0.90
NESDA 0.135 0.82
NEO 0.475 0.83
METSIM 0.490 0.90
LOLIPOP_OmniE 0.027 0.70
LOLIPOP_OmniEE 0.156 0.71
LOLIPOP_IAP 0.567 0.69
LOLIPOP_IA610 0.159 0.69
LOLIPOP_IA317 0.326 0.71
LOLIPOP_EWP 0.280 0.80
LOLIPOP_EWA 0.792 0.82
LOLIPOP_EW610 0.262 0.79
LLS 0.277 0.82
LLD 0.508 0.84
KORCULA 0.055 0.83
INT 1.30e−07 0.81
FR97 0.494 0.88
FR07 0.117 0.88
EUGENDA 0.356 0.82
ERF 0.292 0.82
EGCUT 0.686 0.86
CKB 0.880 0.75
ALSPAC 0.001 0.80
META 6.21e−10 0.81
−0.2
−0.1
0.0
0.1
0.2
BETA

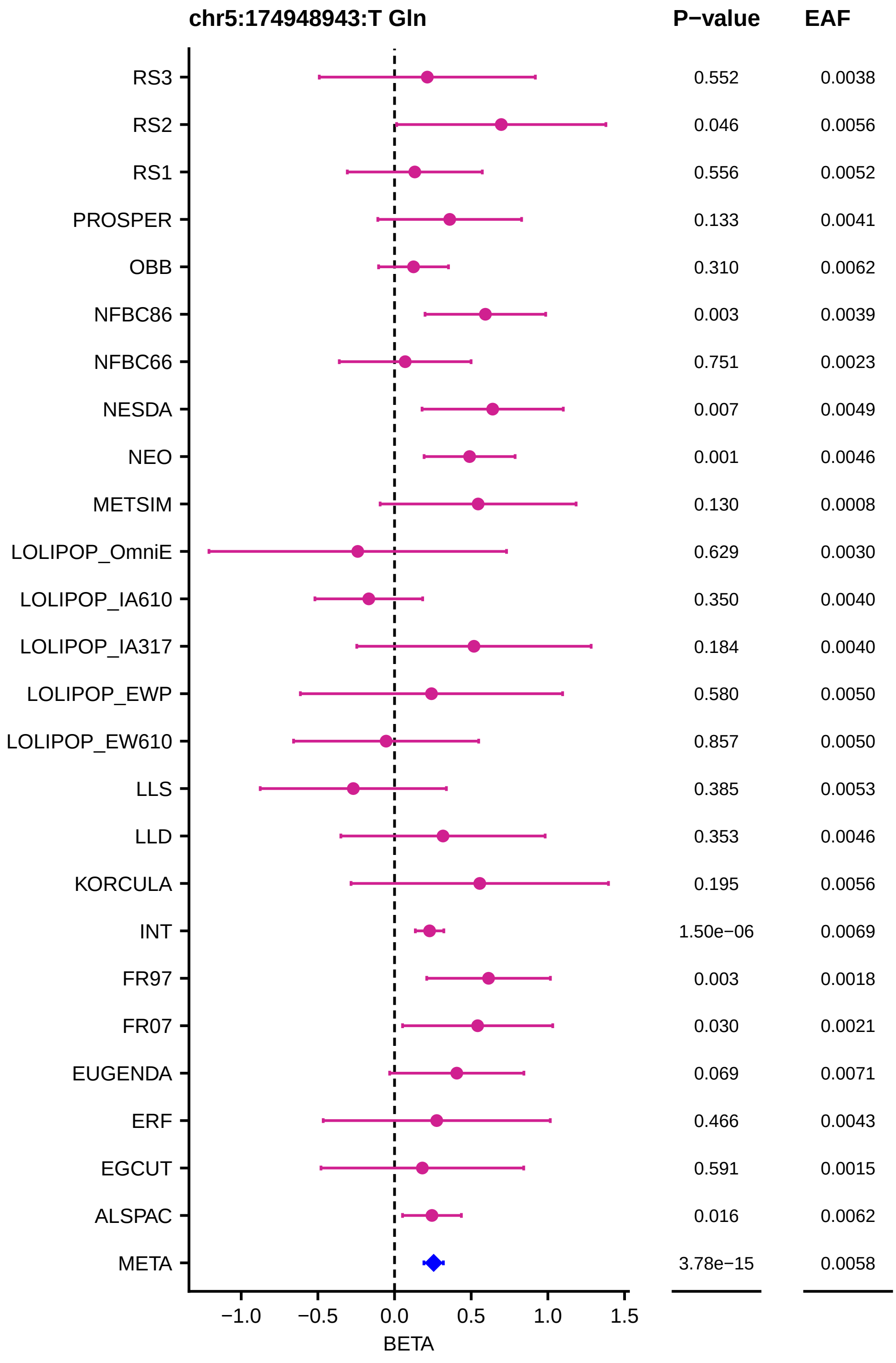
chr5:174948943:T Gln
P−value
EAF
RS3 0.552 0.0038
RS2 0.046 0.0056
RS1 0.556 0.0052
PROSPER 0.133 0.0041
OBB 0.310 0.0062
NFBC86 0.003 0.0039
NFBC66 0.751 0.0023
NESDA 0.007 0.0049
NEO 0.001 0.0046
METSIM 0.130 0.0008
LOLIPOP_OmniE 0.629 0.0030
LOLIPOP_IA610 0.350 0.0040
LOLIPOP_IA317 0.184 0.0040
LOLIPOP_EWP 0.580 0.0050
LOLIPOP_EW610 0.857 0.0050
LLS 0.385 0.0053
LLD 0.353 0.0046
KORCULA 0.195 0.0056
INT 1.50e−06 0.0069
FR97 0.003 0.0018
FR07 0.030 0.0021
EUGENDA 0.069 0.0071
ERF 0.466 0.0043
EGCUT 0.591 0.0015
ALSPAC 0.016 0.0062
META 3.78e−15 0.0058
−1.0
−0.5
0.0
0.5
1.0
1.5
BETA

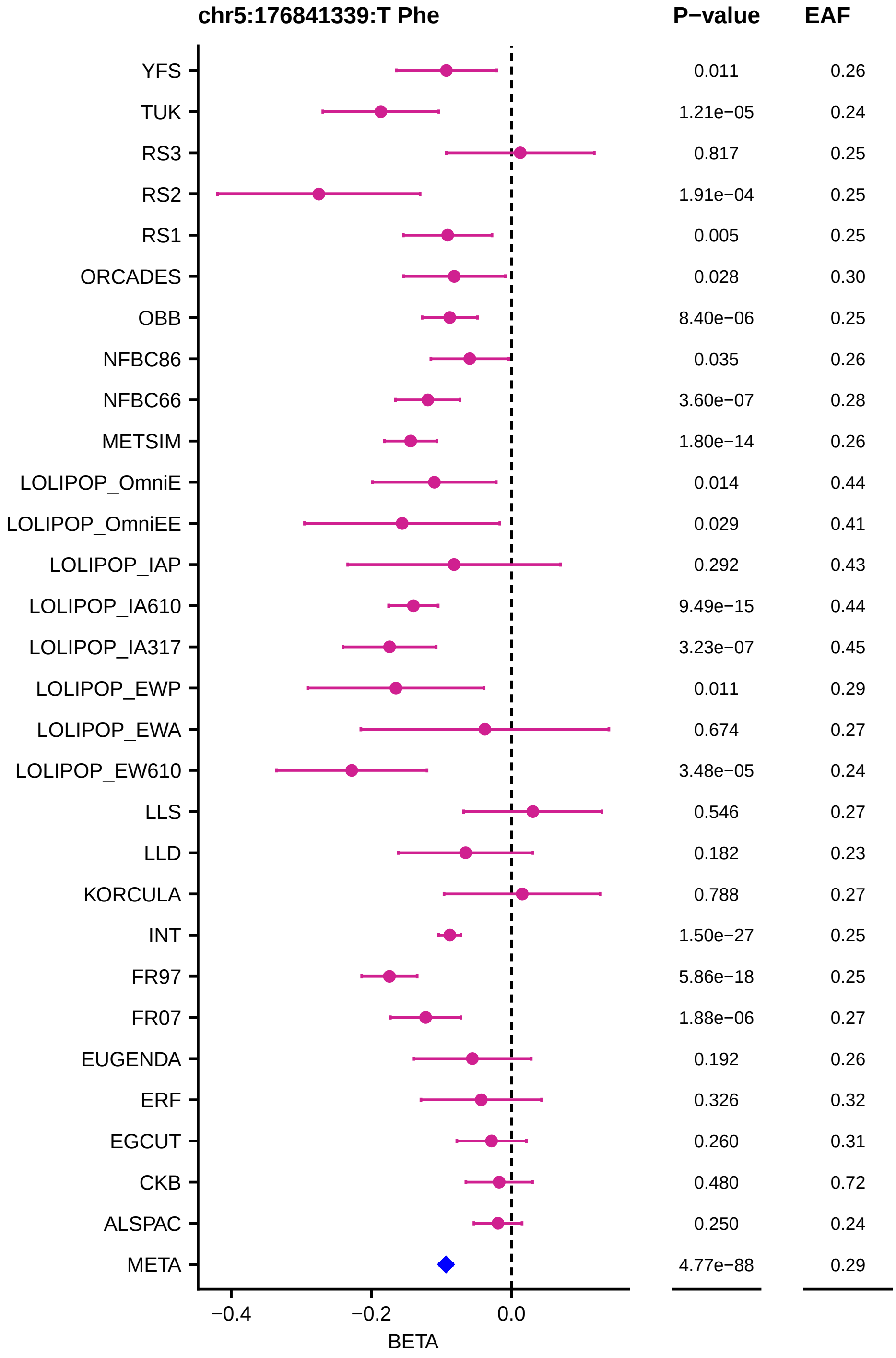
chr5:176841339:T Phe
P−value
EAF
YFS
TUK
RS3
RS2
RS1
ORCADES
OBB
NFBC86
NFBC66
METSIM
LOLIPOP_OmniE
LOLIPOP_OmniEE
LOLIPOP_IAP
LOLIPOP_IA610
LOLIPOP_IA317
LOLIPOP_EWP
LOLIPOP_EWA
LOLIPOP_EW610
LLS
LLD
KORCULA
INT
FR97
FR07
EUGENDA
ERF
EGCUT
CKB
ALSPAC
META
0.011 0.26
1.21e−05 0.24
0.817 0.25
1.91e−04 0.25
0.005 0.25
0.028 0.30
8.40e−06 0.25
0.035 0.26
3.60e−07 0.28
1.80e−14 0.26
0.014 0.44
0.029 0.41
0.292 0.43
9.49e−15 0.44
3.23e−07 0.45
0.011 0.29
0.674 0.27
3.48e−05 0.24
0.546 0.27
0.182 0.23
0.788 0.27
1.50e−27 0.25
5.86e−18 0.25
1.88e−06 0.27
0.192 0.26
0.326 0.32
0.260 0.31
0.480 0.72
0.250 0.24
4.77e−88 0.29
−0.4
−0.2
0.0
BETA

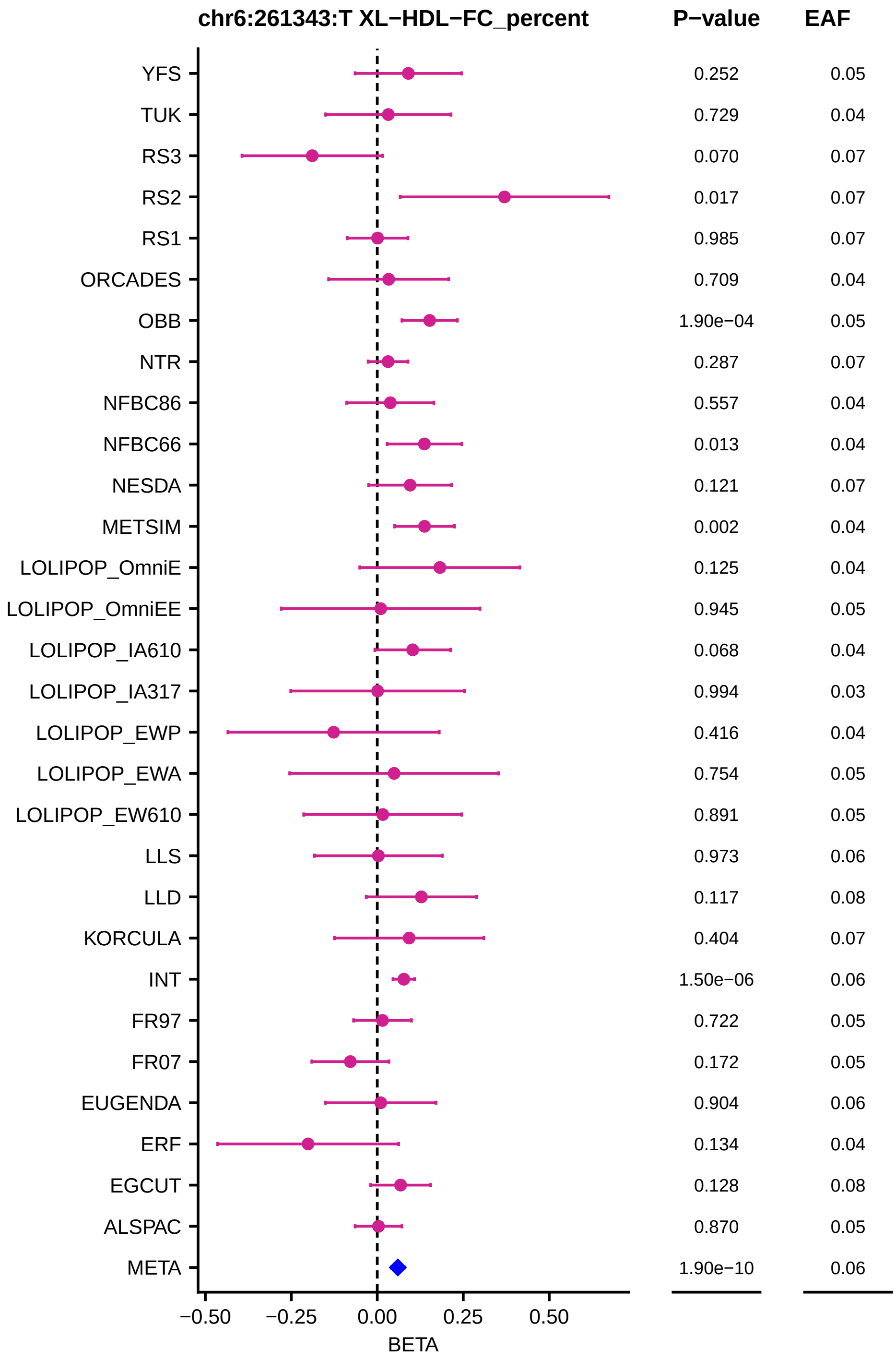
chr6:261343:T XL−HDL−FC_percent
P−value
EAF
YFS
0.252
0.05
TUK
0.729
0.04
RS3
0.070
0.07
RS2
0.017
0.07
RS1
0.985
0.07
ORCADES
0.709
0.04
OBB
1.90e−04
0.05
NTR
0.287
0.07
NFBC86
0.557
0.04
NFBC66
0.013
0.04
NESDA
0.121
0.07
METSIM
0.002
0.04
LOLIPOP_OmniE
0.125
0.04
LOLIPOP_OmniEE
0.945
0.05
LOLIPOP_IA610
0.068
0.04
LOLIPOP_IA317
0.994
0.03
LOLIPOP_EWP
0.416
0.04
LOLIPOP_EWA
0.754
0.05
LOLIPOP_EW610
0.891
0.05
LLS
0.973
0.06
LLD
0.117
0.08
KORCULA
0.404
0.07
INT
1.50e−06
0.06
FR97
0.722
0.05
FR07
0.172
0.05
EUGENDA
0.904
0.06
ERF
0.134
0.04
EGCUT
0.128
0.08
ALSPAC
0.870
0.05
META
1.90e−10
0.06
−0.50
−0.25
0.00
0.25
0.50
BETA

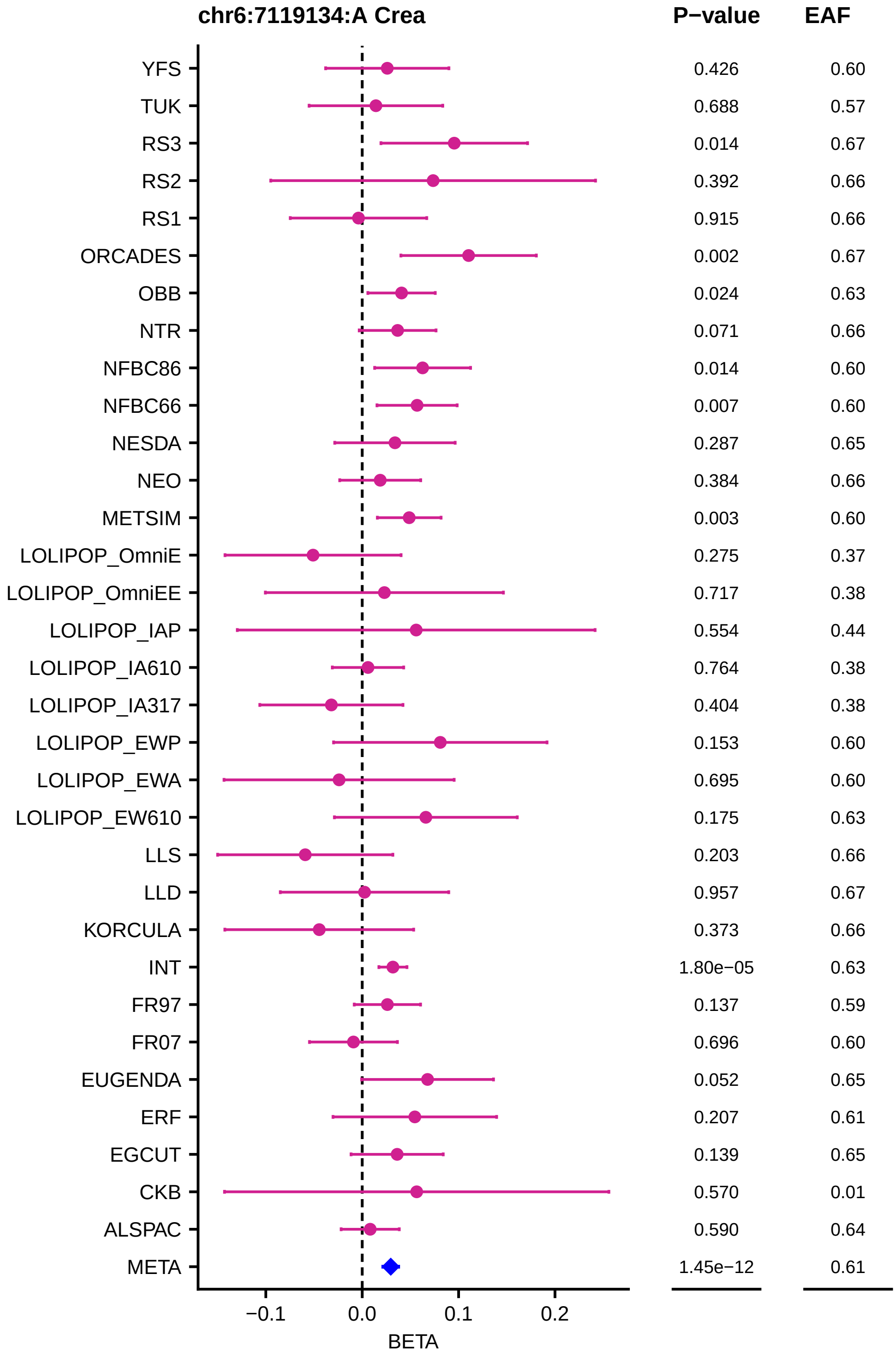

chr6:7119134:A Crea
P−value
EAF
YFS
TUK
RS3
RS2
RS1
ORCADES
OBB
NTR
NFBC86
NFBC66
NESDA
NEO
METSIM
LOLIPOP_OmniE
LOLIPOP_OmniEE
LOLIPOP_IAP
LOLIPOP_IA610
LOLIPOP_IA317
LOLIPOP_EWP
LOLIPOP_EWA
LOLIPOP_EW610
LLS
LLD
KORCULA
INT
FR97
FR07
EUGENDA
ERF
EGCUT
CKB
ALSPAC
META
0.426 0.60
0.688 0.57
0.014 0.67
0.392 0.66
0.915 0.66
0.002 0.67
0.024 0.63
0.071 0.66
0.014 0.60
0.007 0.60
0.287 0.65
0.384 0.66
0.003 0.60
0.275 0.37
0.717 0.38
0.554 0.44
0.764 0.38
0.404 0.38
0.153 0.60
0.695 0.60
0.175 0.63
0.203 0.66
0.957 0.67
0.373 0.66
1.80e−05 0.63
0.137 0.59
0.696 0.60
0.052 0.65
0.207 0.61
0.139 0.65
0.570 0.01
0.590 0.64
1.45e−12 0.61
−0.1
0.0
0.1
0.2
BETA

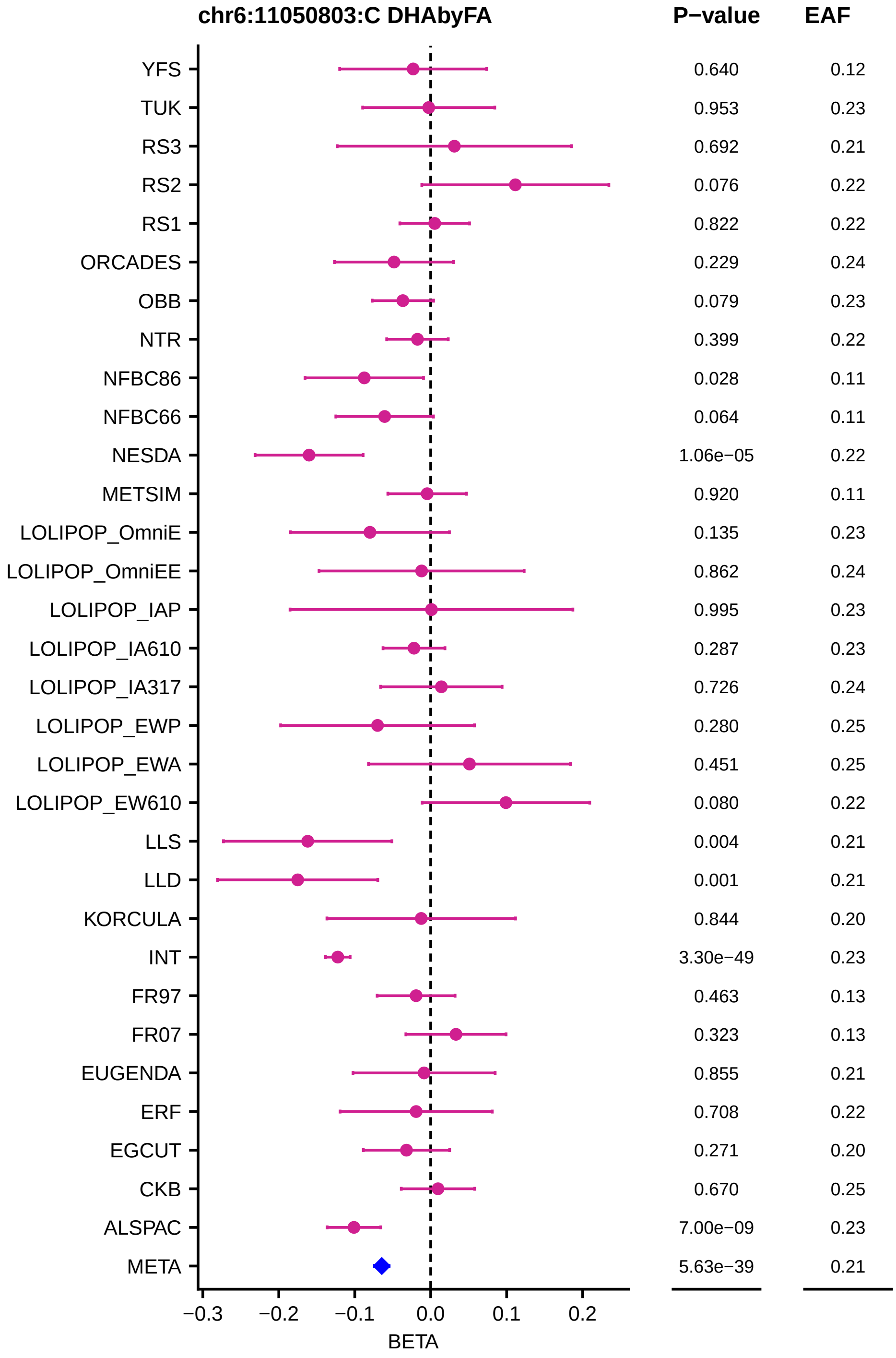

chr6:11050803:C DHAbyFA
P−value
EAF
YFS 0.640 0.12
TUK 0.953 0.23
RS3 0.692 0.21
RS2 0.076 0.22
RS1 0.822 0.22
ORCADES 0.229 0.24
OBB 0.079 0.23
NTR 0.399 0.22
NFBC86 0.028 0.11
NFBC66 0.064 0.11
NESDA 1.06e−05 0.22
METSIM 0.920 0.11
LOLIPOP_OmniE 0.135 0.23
LOLIPOP_OmniEE 0.862 0.24
LOLIPOP_IAP 0.995 0.23
LOLIPOP_IA610 0.287 0.23
LOLIPOP_IA317 0.726 0.24
LOLIPOP_EWP 0.280 0.25
LOLIPOP_EWA 0.451 0.25
LOLIPOP_EW610 0.080 0.22
LLS 0.004 0.21
LLD 0.001 0.21
KORCULA 0.844 0.20
INT 3.30e−49 0.23
FR97 0.463 0.13
FR07 0.323 0.13
EUGENDA 0.855 0.21
ERF 0.708 0.22
EGCUT 0.271 0.20
CKB 0.670 0.25
ALSPAC 7.00e−09 0.23
META 5.63e−39 0.21
−0.3 −0.2 −0.1 0.0 0.1 0.2
BETA

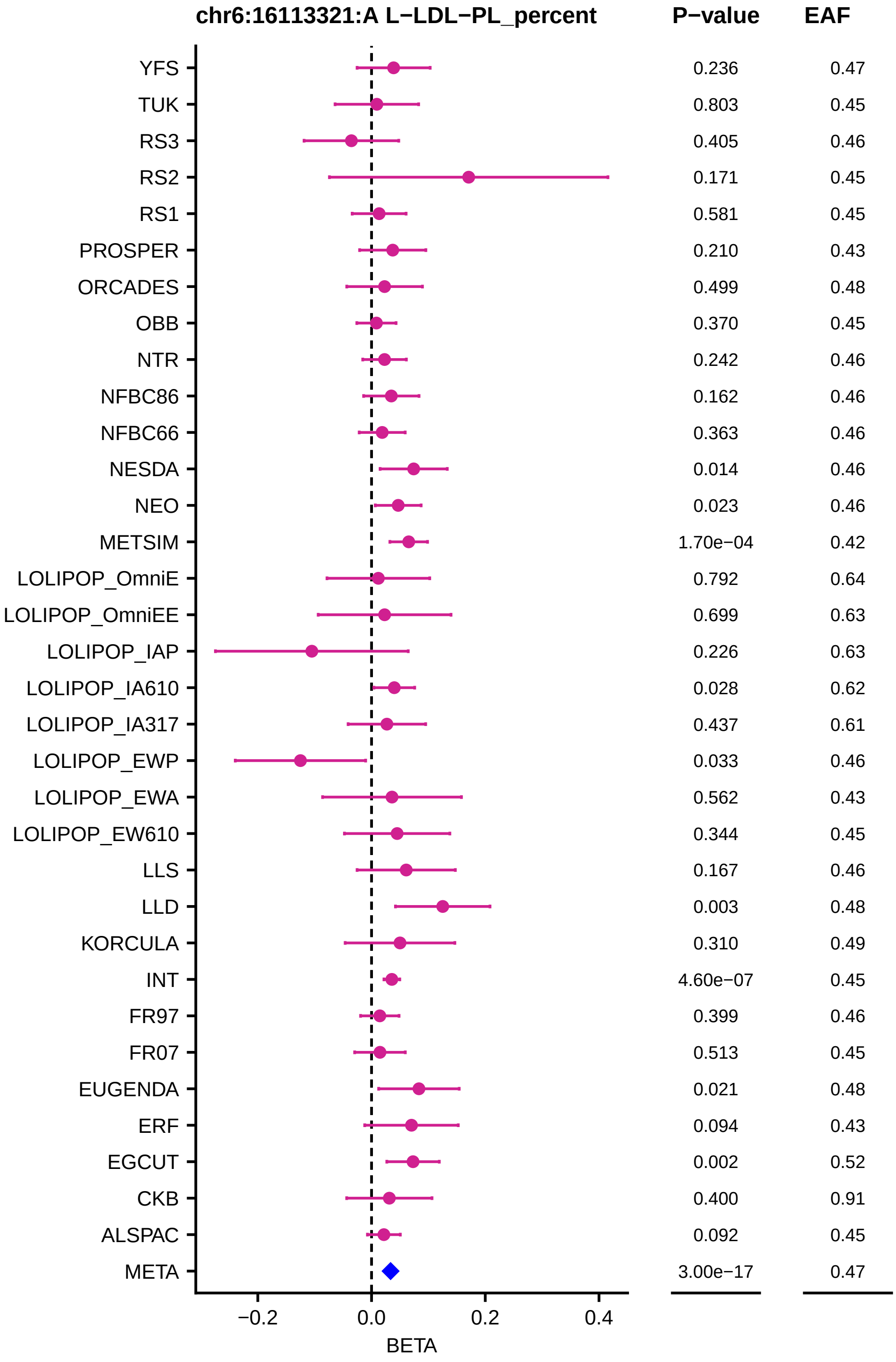
chr6:16113321:A L−LDL−PL_percent
P−value
EAF
YFS 0.236 0.47
TUK 0.803 0.45
RS3 0.405 0.46
RS2 0.171 0.45
RS1 0.581 0.45
PROSPER 0.210 0.43
ORCADES 0.499 0.48
OBB 0.370 0.45
NTR 0.242 0.46
NFBC86 0.162 0.46
NFBC66 0.363 0.46
NESDA 0.014 0.46
NEO 0.023 0.46
METSIM 1.70e−04 0.42
LOLIPOP_OmniE 0.792 0.64
LOLIPOP_OmniEE 0.699 0.63
LOLIPOP_IAP 0.226 0.63
LOLIPOP_IA610 0.028 0.62
LOLIPOP_IA317 0.437 0.61
LOLIPOP_EWP 0.033 0.46
LOLIPOP_EWA 0.562 0.43
LOLIPOP_EW610 0.344 0.45
LLS 0.167 0.46
LLD 0.003 0.48
KORCULA 0.310 0.49
INT 4.60e−07 0.45
FR97 0.399 0.46
FR07 0.513 0.45
EUGENDA 0.021 0.48
ERF 0.094 0.43
EGCUT 0.002 0.52
CKB 0.400 0.91
ALSPAC 0.092 0.45
META 3.00e−17 0.47
−0.2
0.0
0.2
0.4
BETA

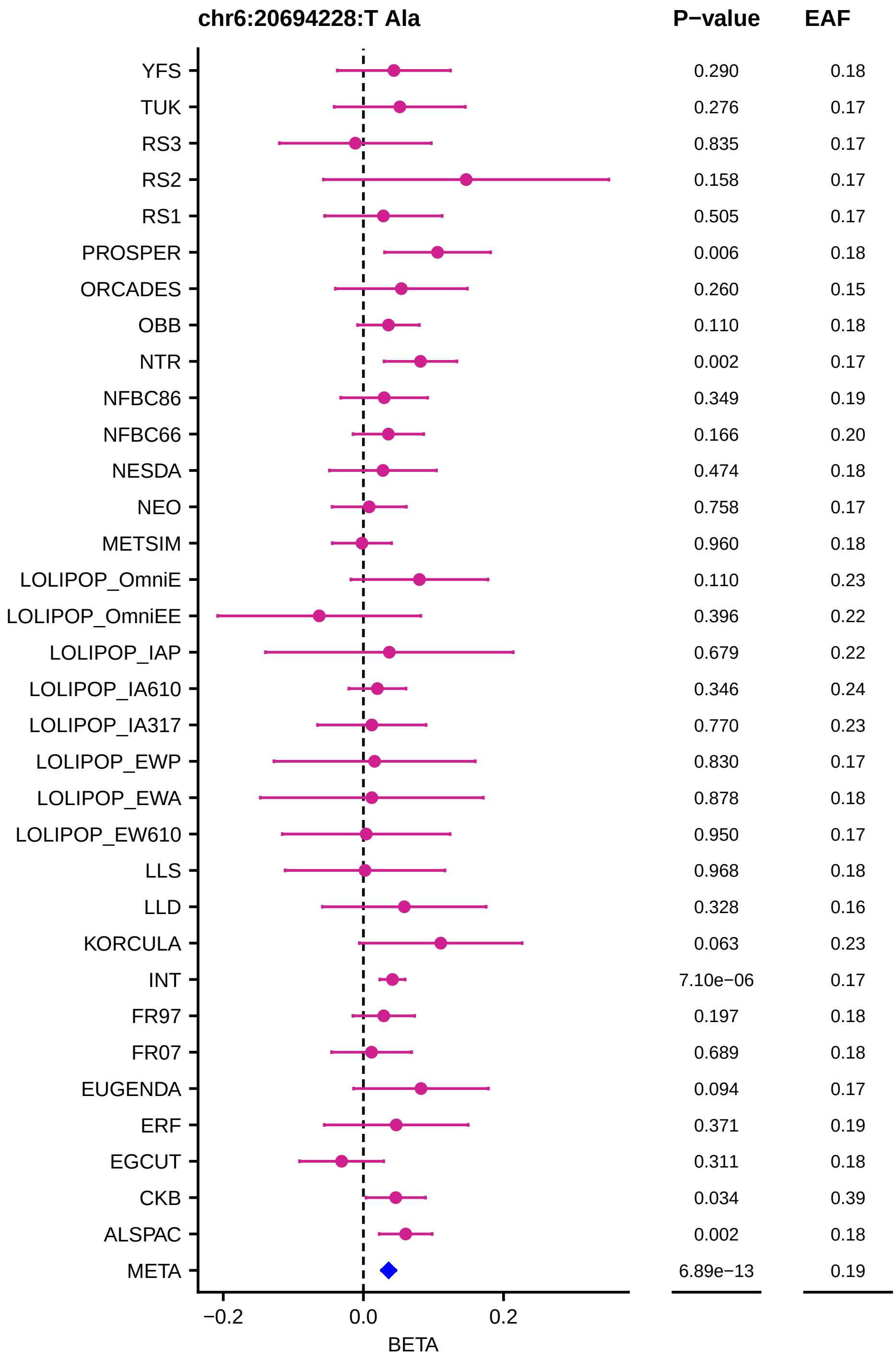
chr6:20694228:T Ala
P−value
EAF
YFS 0.290 0.18
TUK 0.276 0.17
RS3 0.835 0.17
RS2 0.158 0.17
RS1 0.505 0.17
PROSPER 0.006 0.18
ORCADES 0.260 0.15
OBB 0.110 0.18
NTR 0.002 0.17
NFBC86 0.349 0.19
NFBC66 0.166 0.20
NESDA 0.474 0.18
NEO 0.758 0.17
METSIM 0.960 0.18
LOLIPOP_OmniE 0.110 0.23
LOLIPOP_OmniEE 0.396 0.22
LOLIPOP_IAP 0.679 0.22
LOLIPOP_IA610 0.346 0.24
LOLIPOP_IA317 0.770 0.23
LOLIPOP_EWP 0.830 0.17
LOLIPOP_EWA 0.878 0.18
LOLIPOP_EW610 0.950 0.17
LLS 0.968 0.18
LLD 0.328 0.16
KORCULA 0.063 0.23
INT 7.10e−06 0.17
FR97 0.197 0.18
FR07 0.689 0.18
EUGENDA 0.094 0.17
ERF 0.371 0.19
EGCUT 0.311 0.18
CKB 0.034 0.39
ALSPAC 0.002 0.18
META 6.89e−13 0.19
−0.2
0.0
0.2
BETA

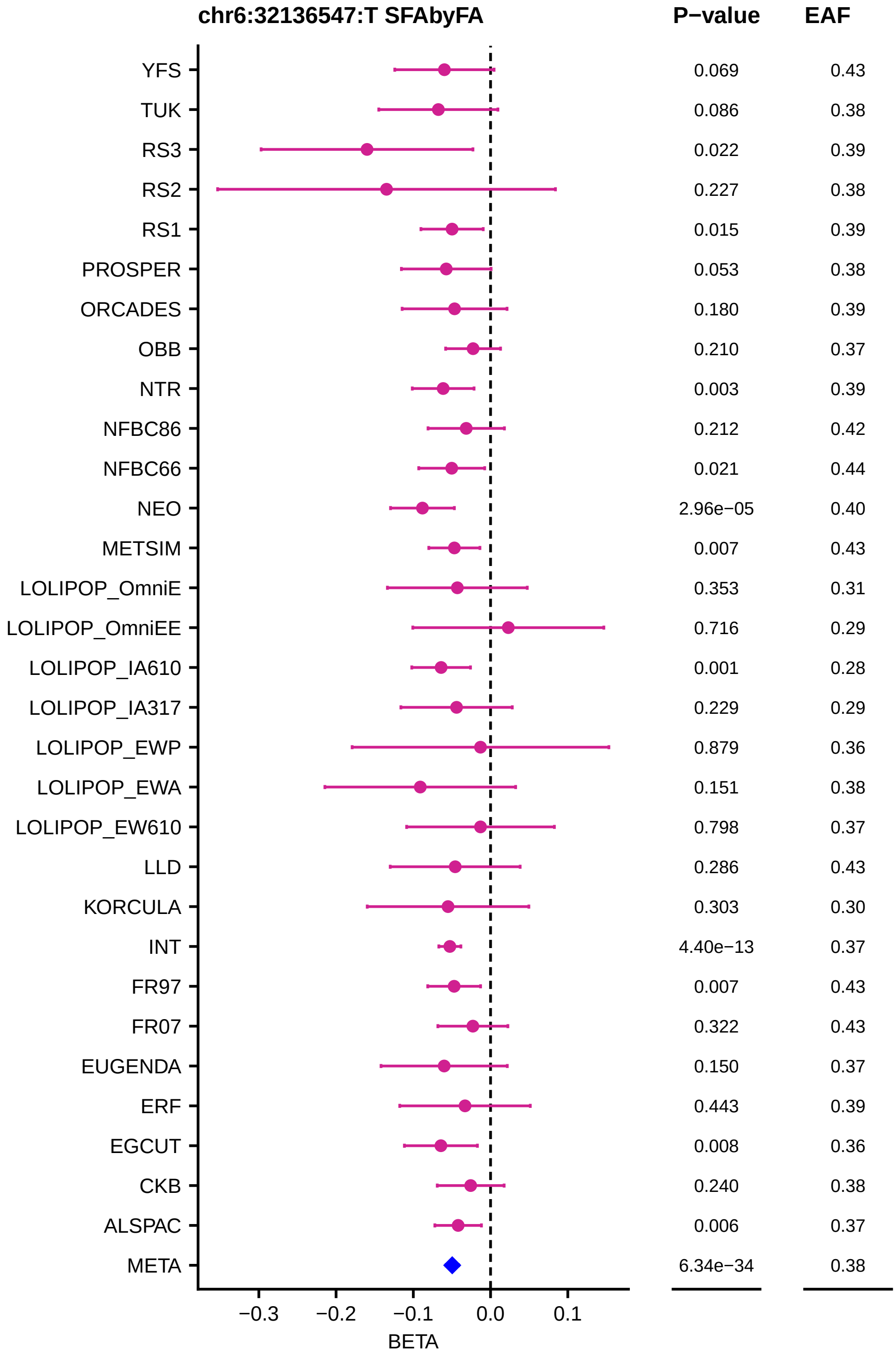
chr6:32136547:T SFAbyFA
P−value
EAF
YFS 0.069 0.43
TUK 0.086 0.38
RS3 0.022 0.39
RS2 0.227 0.38
RS1 0.015 0.39
PROSPER 0.053 0.38
ORCADES 0.180 0.39
OBB 0.210 0.37
NTR 0.003 0.39
NFBC86 0.212 0.42
NFBC66 0.021 0.44
NEO 2.96e−05 0.40
METSIM 0.007 0.43
LOLIPOP_OmniE 0.353 0.31
LOLIPOP_OmniEE 0.716 0.29
LOLIPOP_IA610 0.001 0.28
LOLIPOP_IA317 0.229 0.29
LOLIPOP_EWP 0.879 0.36
LOLIPOP_EWA 0.151 0.38
LOLIPOP_EW610 0.798 0.37
LLD 0.286 0.43
KORCULA 0.303 0.30
INT 4.40e−13 0.37
FR97 0.007 0.43
FR07 0.322 0.43
EUGENDA 0.150 0.37
ERF 0.443 0.39
EGCUT 0.008 0.36
CKB 0.240 0.38
ALSPAC 0.006 0.37
META 6.34e−34 0.38
−0.3
−0.2
−0.1
0.0
0.1
BETA

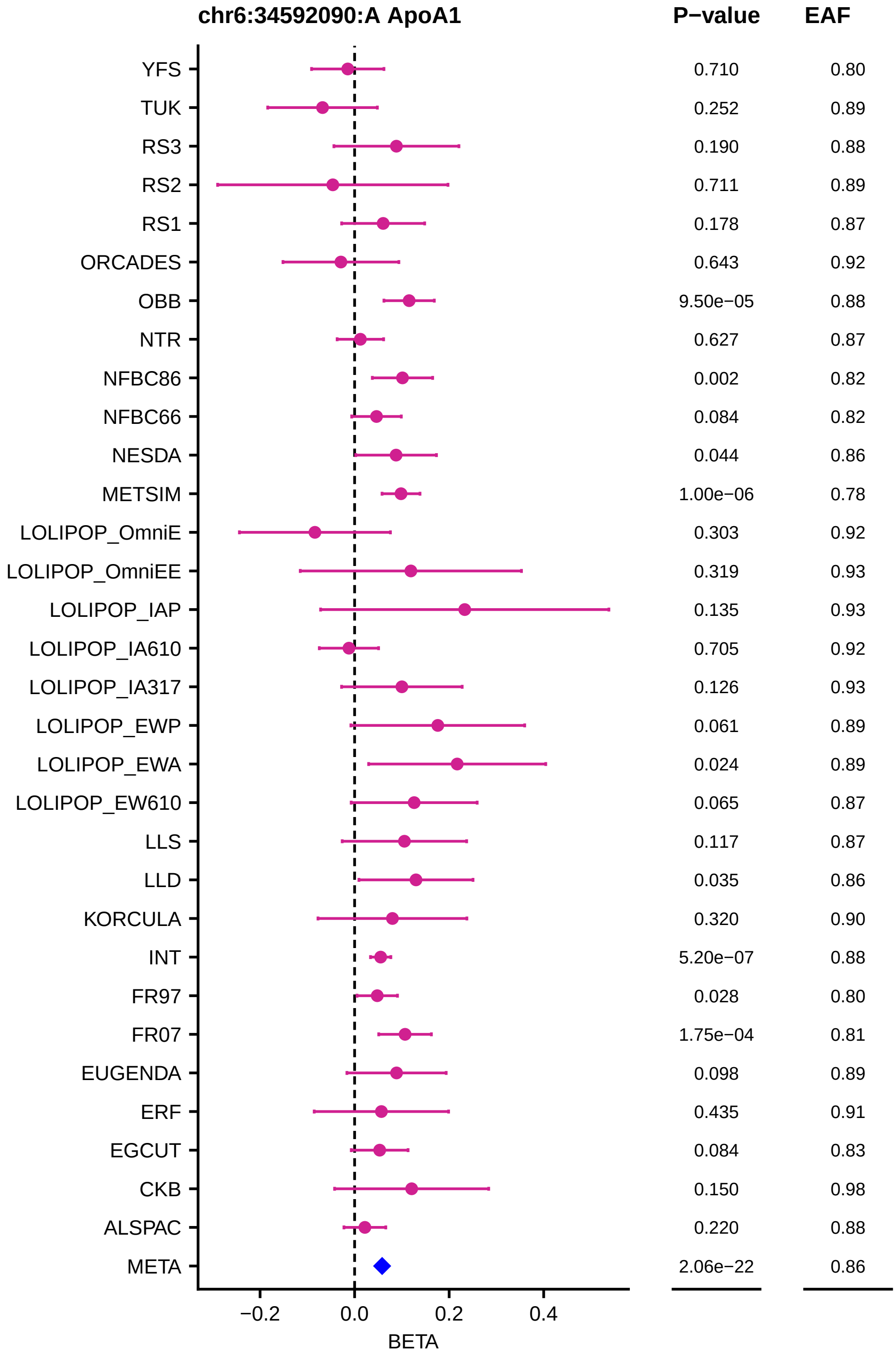
chr6:34592090:A ApoA1
P−value
EAF
YFS 0.710 0.80
TUK 0.252 0.89
RS3 0.190 0.88
RS2 0.711 0.89
RS1 0.178 0.87
ORCADES 0.643 0.92
OBB 9.50e−05 0.88
NTR 0.627 0.87
NFBC86 0.002 0.82
NFBC66 0.084 0.82
NESDA 0.044 0.86
METSIM 1.00e−06 0.78
LOLIPOP_OmniE 0.303 0.92
LOLIPOP_OmniEE 0.319 0.93
LOLIPOP_IAP 0.135 0.93
LOLIPOP_IA610 0.705 0.92
LOLIPOP_IA317 0.126 0.93
LOLIPOP_EWP 0.061 0.89
LOLIPOP_EWA 0.024 0.89
LOLIPOP_EW610 0.065 0.87
LLS 0.117 0.87
LLD 0.035 0.86
KORCULA 0.320 0.90
INT 5.20e−07 0.88
FR97 0.028 0.80
FR07 1.75e−04 0.81
EUGENDA 0.098 0.89
ERF 0.435 0.91
EGCUT 0.084 0.83
CKB 0.150 0.98
ALSPAC 0.220 0.88
META 2.06e−22 0.86
−0.2
0.0
0.2
0.4
BETA

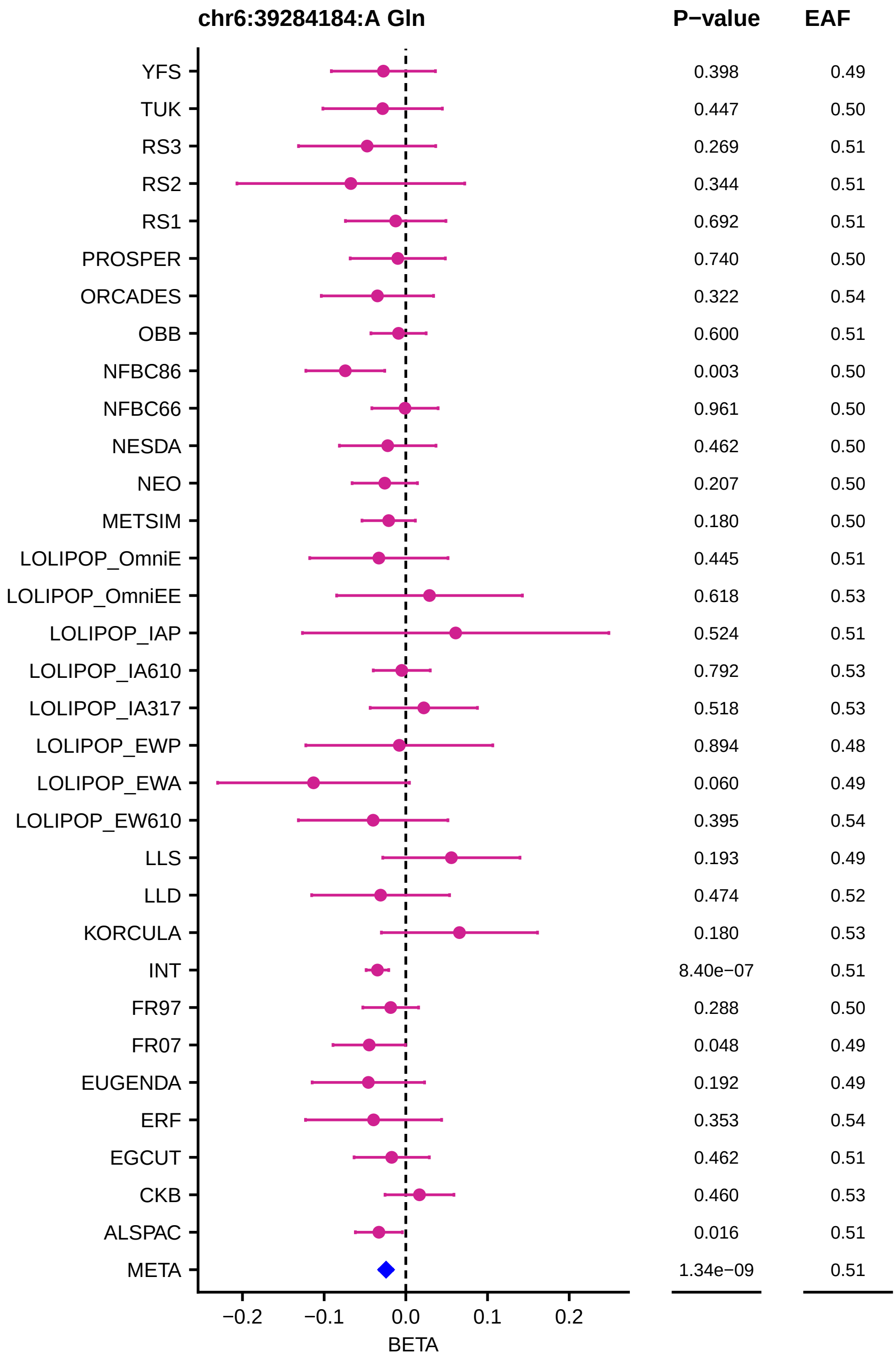
chr6:39284184:A Gln
P−value
EAF
YFS 0.398 0.49
TUK 0.447 0.50
RS3 0.269 0.51
RS2 0.344 0.51
RS1 0.692 0.51
PROSPER 0.740 0.50
ORCADES 0.322 0.54
OBB 0.600 0.51
NFBC86 0.003 0.50
NFBC66 0.961 0.50
NESDA 0.462 0.50
NEO 0.207 0.50
METSIM 0.180 0.50
LOLIPOP_OmniE 0.445 0.51
LOLIPOP_OmniEE 0.618 0.53
LOLIPOP_IAP 0.524 0.51
LOLIPOP_IA610 0.792 0.53
LOLIPOP_IA317 0.518 0.53
LOLIPOP_EWP 0.894 0.48
LOLIPOP_EWA 0.060 0.49
LOLIPOP_EW610 0.395 0.54
LLS 0.193 0.49
LLD 0.474 0.52
KORCULA 0.180 0.53
INT 8.40e−07 0.51
FR97 0.288 0.50
FR07 0.048 0.49
EUGENDA 0.192 0.49
ERF 0.353 0.54
EGCUT 0.462 0.51
CKB 0.460 0.53
ALSPAC 0.016 0.51
META 1.34e−09 0.51
−0.2
−0.1
0.0
0.1
0.2
BETA

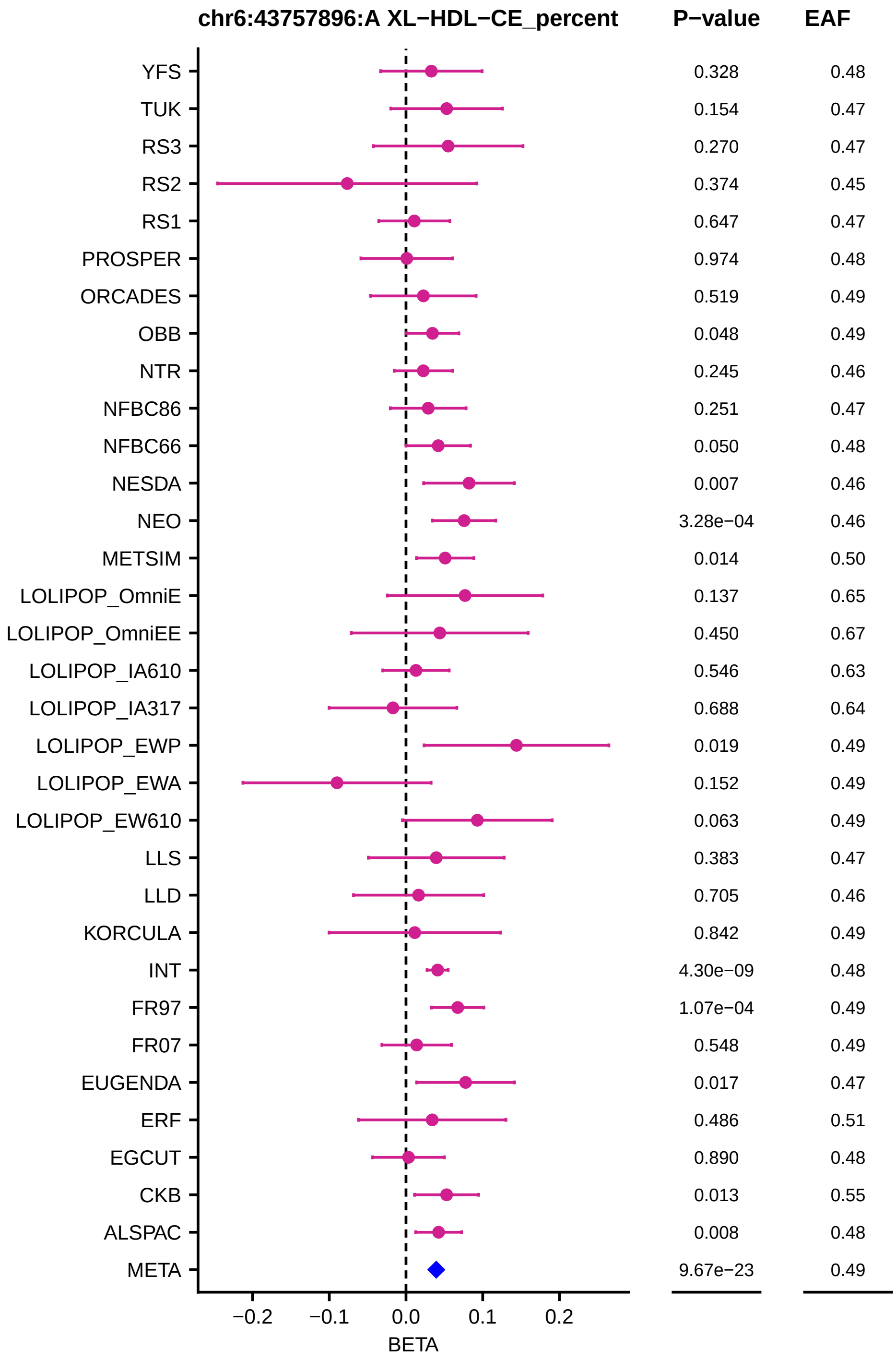
chr6:43757896:A XL−HDL−CE_percent
P−value
EAF
YFS 0.328 0.48
TUK 0.154 0.47
RS3 0.270 0.47
RS2 0.374 0.45
RS1 0.647 0.47
PROSPER 0.974 0.48
ORCADES 0.519 0.49
OBB 0.048 0.49
NTR 0.245 0.46
NFBC86 0.251 0.47
NFBC66 0.050 0.48
NESDA 0.007 0.46
NEO 3.28e−04 0.46
METSIM 0.014 0.50
LOLIPOP_OmniE 0.137 0.65
LOLIPOP_OmniEE 0.450 0.67
LOLIPOP_IA610 0.546 0.63
LOLIPOP_IA317 0.688 0.64
LOLIPOP_EWP 0.019 0.49
LOLIPOP_EWA 0.152 0.49
LOLIPOP_EW610 0.063 0.49
LLS 0.383 0.47
LLD 0.705 0.46
KORCULA 0.842 0.49
INT 4.30e−09 0.48
FR97 1.07e−04 0.49
FR07 0.548 0.49
EUGENDA 0.017 0.47
ERF 0.486 0.51
EGCUT 0.890 0.48
CKB 0.013 0.55
ALSPAC 0.008 0.48
META 9.67e−23 0.49
−0.2
−0.1
0.0
0.1
0.2
BETA

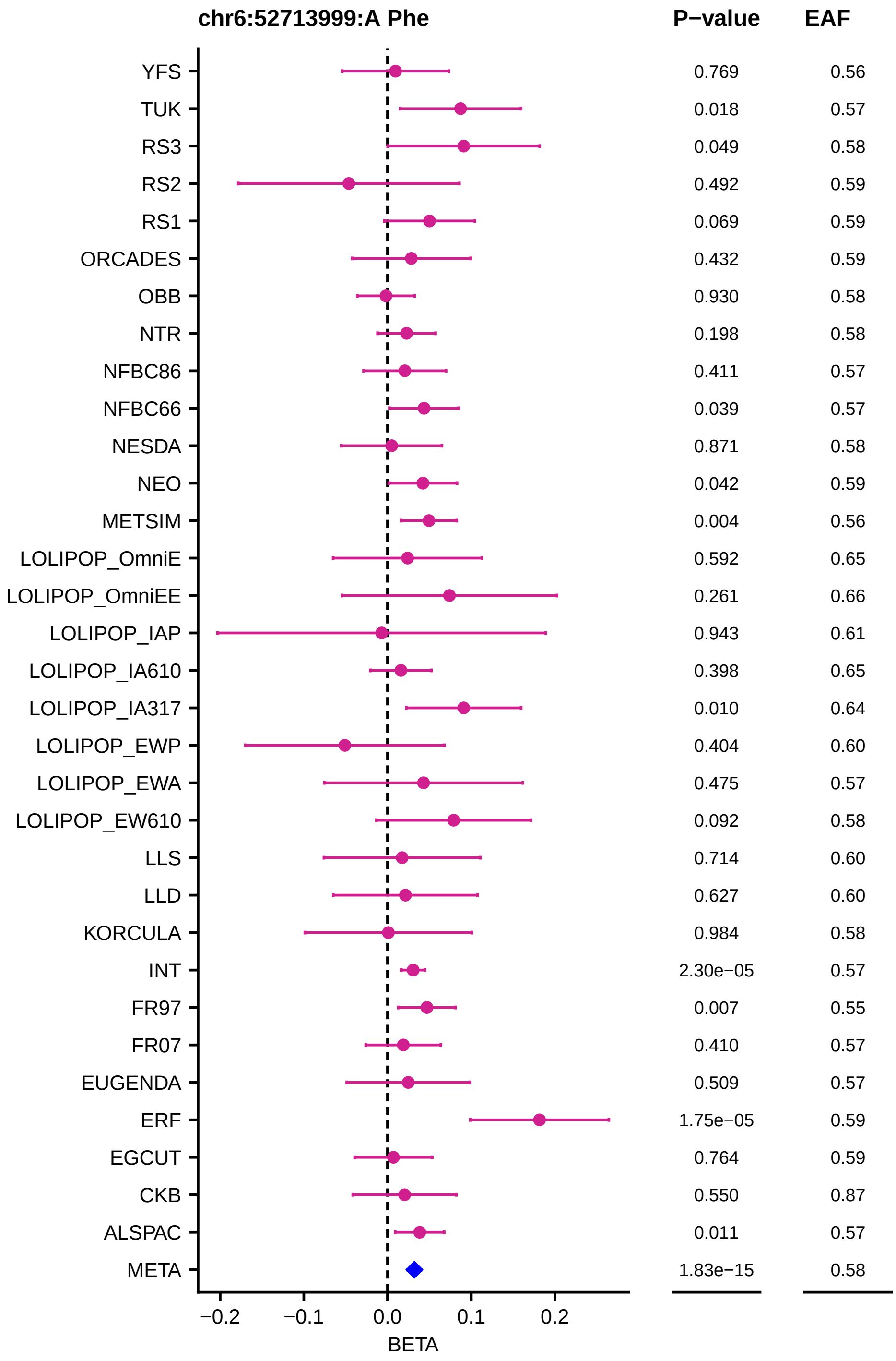

chr6:52713999:A Phe
P−value
EAF
YFS 0.769 0.56
TUK 0.018 0.57
RS3 0.049 0.58
RS2 0.492 0.59
RS1 0.069 0.59
ORCADES 0.432 0.59
OBB 0.930 0.58
NTR 0.198 0.58
NFBC86 0.411 0.57
NFBC66 0.039 0.57
NESDA 0.871 0.58
NEO 0.042 0.59
METSIM 0.004 0.56
LOLIPOP_OmniE 0.592 0.65
LOLIPOP_OmniEE 0.261 0.66
LOLIPOP_IAP 0.943 0.61
LOLIPOP_IA610 0.398 0.65
LOLIPOP_IA317 0.010 0.64
LOLIPOP_EWP 0.404 0.60
LOLIPOP_EWA 0.475 0.57
LOLIPOP_EW610 0.092 0.58
LLS 0.714 0.60
LLD 0.627 0.60
KORCULA 0.984 0.58
INT 2.30e−05 0.57
FR97 0.007 0.55
FR07 0.410 0.57
EUGENDA 0.509 0.57
ERF 1.75e−05 0.59
EGCUT 0.764 0.59
CKB 0.550 0.87
ALSPAC 0.011 0.57
META 1.83e−15 0.58
−0.2
−0.1
0.0
0.1
0.2
BETA

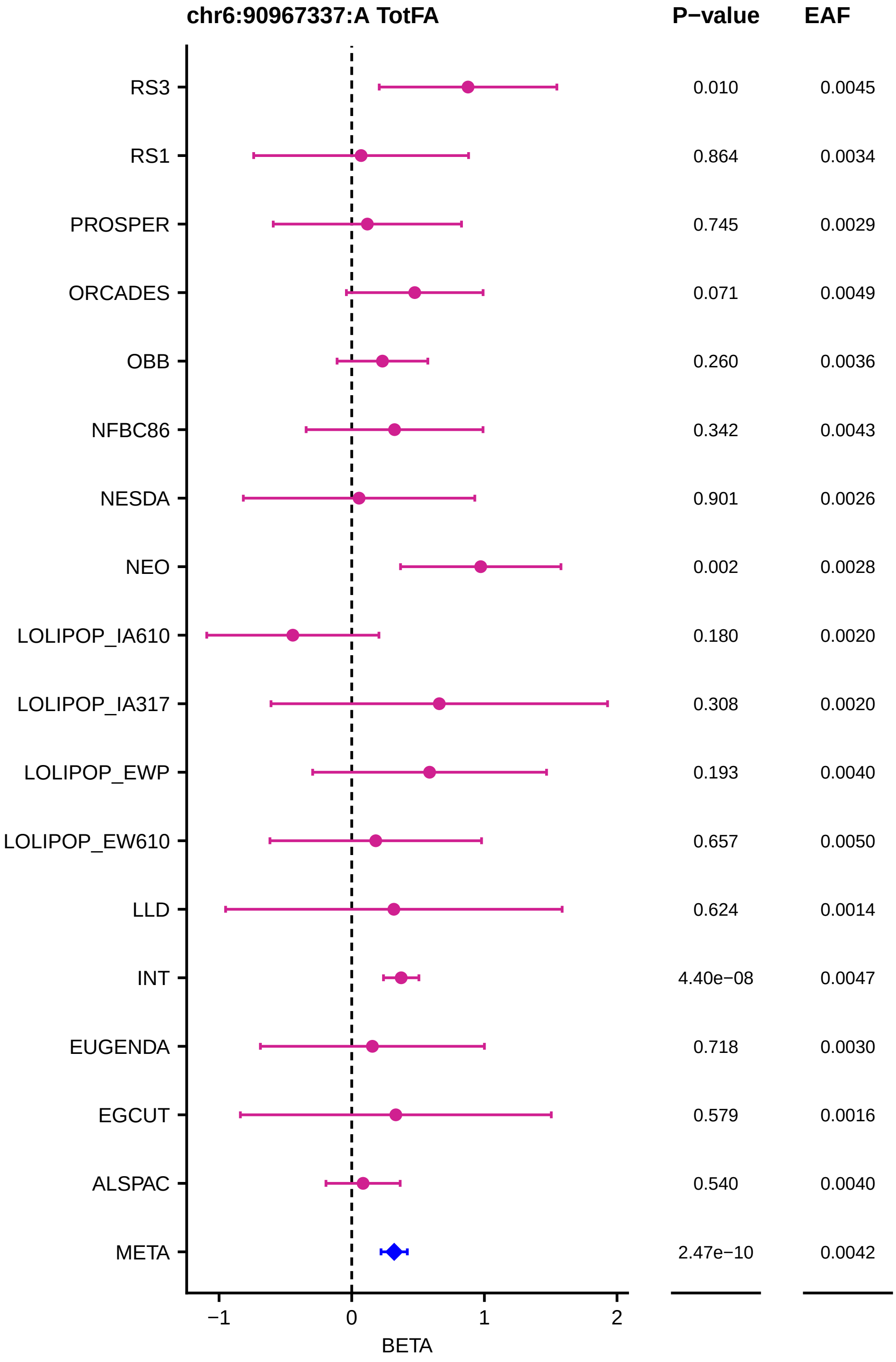

chr6:90967337:A TotFA
P−value
EAF
RS3 0.010 0.0045
RS1 0.864 0.0034
PROSPER 0.745 0.0029
ORCADES 0.071 0.0049
OBB 0.260 0.0036
NFBC86 0.342 0.0043
NESDA 0.901 0.0026
NEO 0.002 0.0028
LOLIPOP_IA610 0.180 0.0020
LOLIPOP_IA317 0.308 0.0020
LOLIPOP_EWP 0.193 0.0040
LOLIPOP_EW610 0.657 0.0050
LLD 0.624 0.0014
INT 4.40e−08 0.0047
EUGENDA 0.718 0.0030
EGCUT 0.579 0.0016
ALSPAC 0.540 0.0040
META 2.47e−10 0.0042
−1
0
1
2
BETA

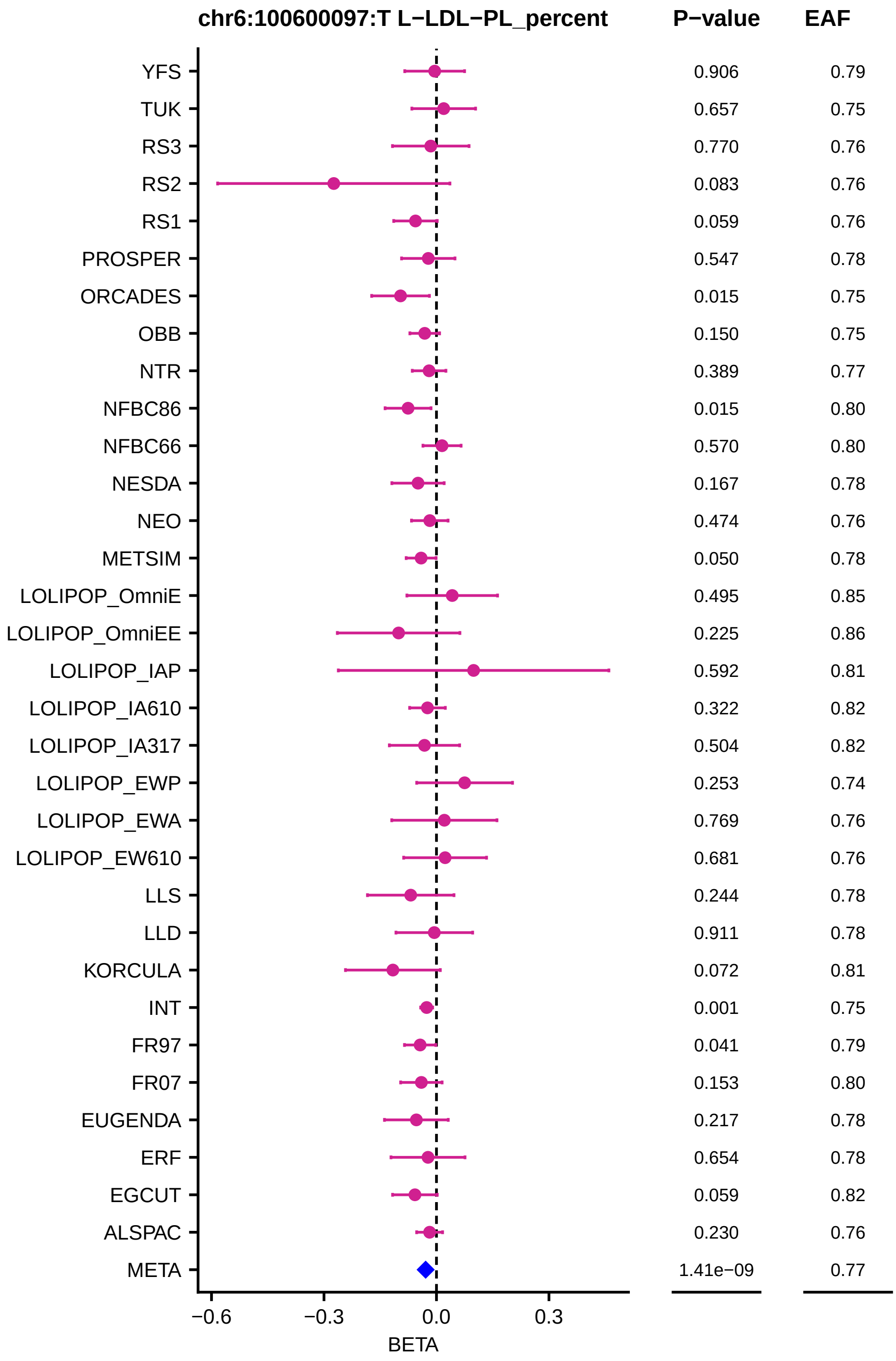

chr6:100600097:T L−LDL−PL_percent
P−value
EAF
YFS
TUK
RS3
RS2
RS1
PROSPER
ORCADES
OBB
NTR
NFBC86
NFBC66
NESDA
NEO
METSIM
LOLIPOP_OmniE
LOLIPOP_OmniEE
LOLIPOP_IAP
LOLIPOP_IA610
LOLIPOP_IA317
LOLIPOP_EWP
LOLIPOP_EWA
LOLIPOP_EW610
LLS
LLD
KORCULA
INT
FR97
FR07
EUGENDA
ERF
EGCUT
ALSPAC
META
0.906 0.79
0.657 0.75
0.770 0.76
0.083 0.76
0.059 0.76
0.547 0.78
0.015 0.75
0.150 0.75
0.389 0.77
0.015 0.80
0.570 0.80
0.167 0.78
0.474 0.76
0.050 0.78
0.495 0.85
0.225 0.86
0.592 0.81
0.322 0.82
0.504 0.82
0.253 0.74
0.769 0.76
0.681 0.76
0.244 0.78
0.911 0.78
0.072 0.81
0.001 0.75
0.041 0.79
0.153 0.80
0.217 0.78
0.654 0.78
0.059 0.82
0.230 0.76
1.41e−09 0.77
−0.6
−0.3
0.0
0.3
BETA

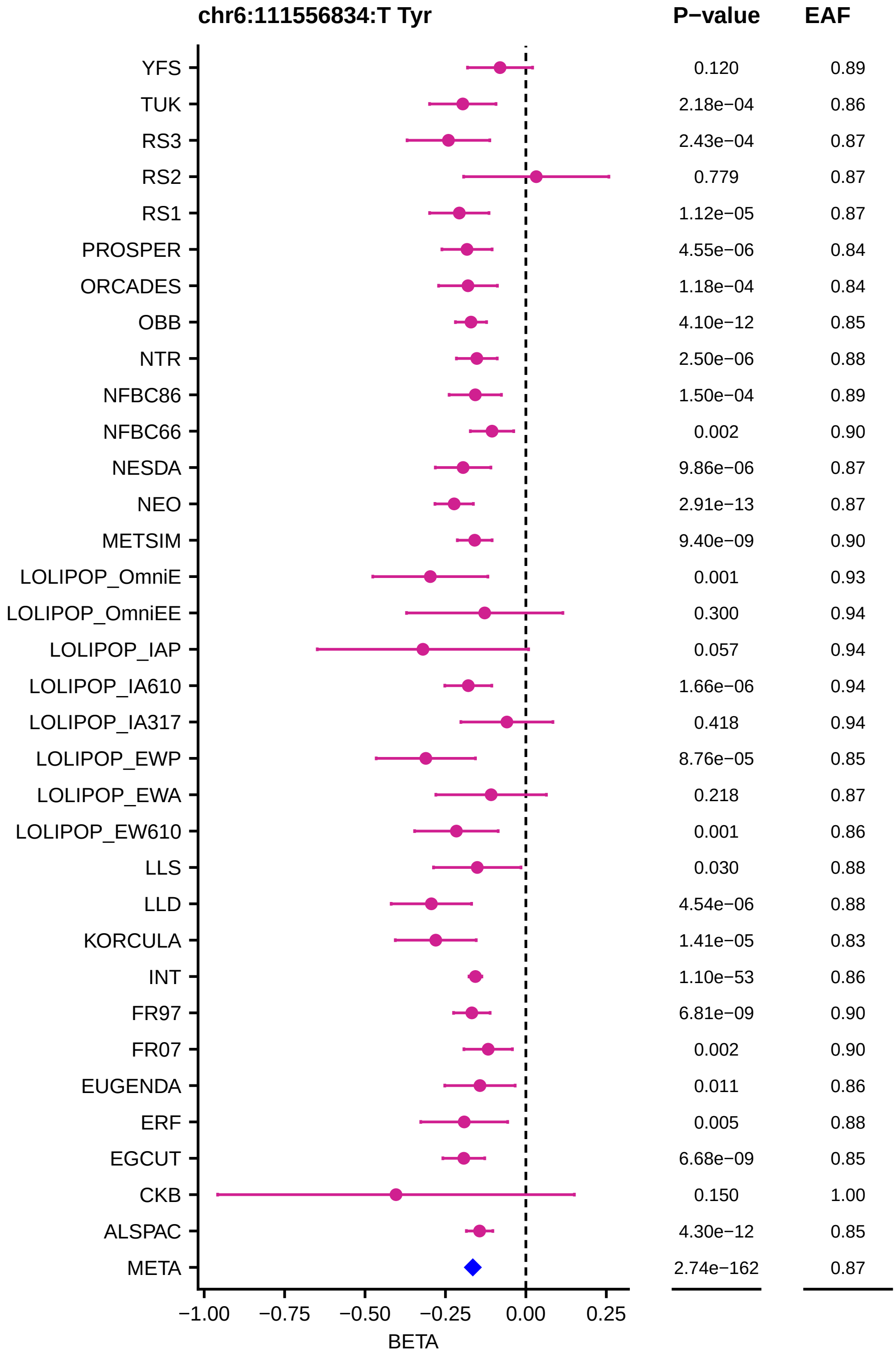
chr6:111556834:T Tyr
P−value
EAF
YFS 0.120 0.89
TUK 2.18e−04 0.86
RS3 2.43e−04 0.87
RS2 0.779 0.87
RS1 1.12e−05 0.87
PROSPER 4.55e−06 0.84
ORCADES 1.18e−04 0.84
OBB 4.10e−12 0.85
NTR 2.50e−06 0.88
NFBC86 1.50e−04 0.89
NFBC66 0.002 0.90
NESDA 9.86e−06 0.87
NEO 2.91e−13 0.87
METSIM 9.40e−09 0.90
LOLIPOP_OmniE 0.001 0.93
LOLIPOP_OmniEE 0.300 0.94
LOLIPOP_IAP 0.057 0.94
LOLIPOP_IA610 1.66e−06 0.94
LOLIPOP_IA317 0.418 0.94
LOLIPOP_EWP 8.76e−05 0.85
LOLIPOP_EWA 0.218 0.87
LOLIPOP_EW610 0.001 0.86
LLS 0.030 0.88
LLD 4.54e−06 0.88
KORCULA 1.41e−05 0.83
INT 1.10e−53 0.86
FR97 6.81e−09 0.90
FR07 0.002 0.90
EUGENDA 0.011 0.86
ERF 0.005 0.88
EGCUT 6.68e−09 0.85
CKB 0.150 1.00
ALSPAC 4.30e−12 0.85
META 2.74e−162 0.87
−1.00 −0.75 −0.50 −0.25 0.00 0.25
BETA

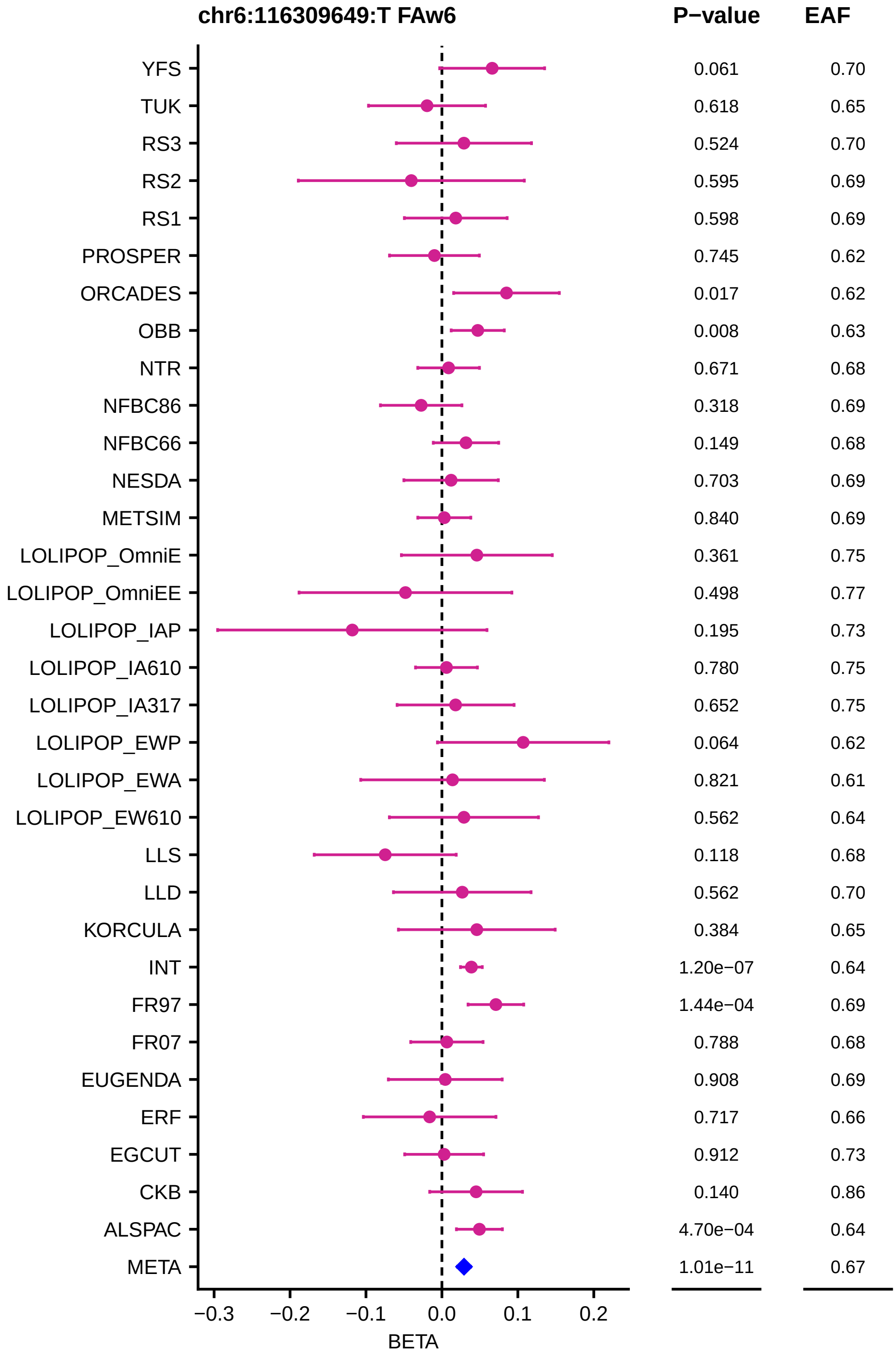
chr6:116309649:T FAw6
P−value
EAF
YFS 0.061 0.70
TUK 0.618 0.65
RS3 0.524 0.70
RS2 0.595 0.69
RS1 0.598 0.69
PROSPER 0.745 0.62
ORCADES 0.017 0.62
OBB 0.008 0.63
NTR 0.671 0.68
NFBC86 0.318 0.69
NFBC66 0.149 0.68
NESDA 0.703 0.69
METSIM 0.840 0.69
LOLIPOP_OmniE 0.361 0.75
LOLIPOP_OmniEE 0.498 0.77
LOLIPOP_IAP 0.195 0.73
LOLIPOP_IA610 0.780 0.75
LOLIPOP_IA317 0.652 0.75
LOLIPOP_EWP 0.064 0.62
LOLIPOP_EWA 0.821 0.61
LOLIPOP_EW610 0.562 0.64
LLS 0.118 0.68
LLD 0.562 0.70
KORCULA 0.384 0.65
INT 1.20e−07 0.64
FR97 1.44e−04 0.69
FR07 0.788 0.68
EUGENDA 0.908 0.69
ERF 0.717 0.66
EGCUT 0.912 0.73
CKB 0.140 0.86
ALSPAC 4.70e−04 0.64
META 1.01e−11 0.67
−0.3 −0.2 −0.1 0.0 0.1 0.2
BETA

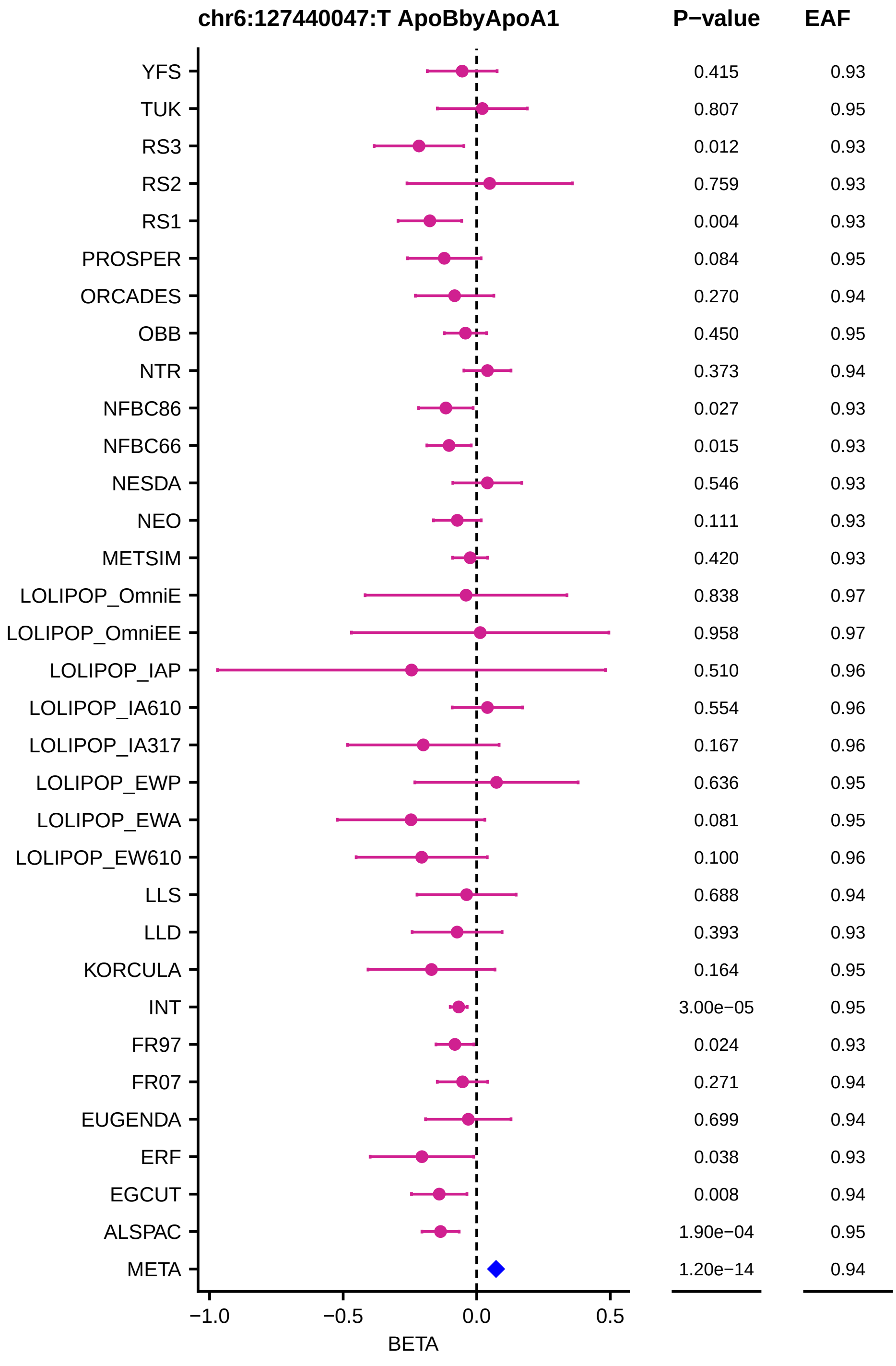
chr6:127440047:T ApoBbyApoA1
P−value
EAF
YFS
TUK
RS3
RS2
RS1
PROSPER
ORCADES
OBB
NTR
NFBC86
NFBC66
NESDA
NEO
METSIM
LOLIPOP_OmniE
LOLIPOP_OmniEE
LOLIPOP_IAP
LOLIPOP_IA610
LOLIPOP_IA317
LOLIPOP_EWP
LOLIPOP_EWA
LOLIPOP_EW610
LLS
LLD
KORCULA
INT
FR97
FR07
EUGENDA
ERF
EGCUT
ALSPAC
META
0.415 0.93
0.807 0.95
0.012 0.93
0.759 0.93
0.004 0.93
0.084 0.95
0.270 0.94
0.450 0.95
0.373 0.94
0.027 0.93
0.015 0.93
0.546 0.93
0.111 0.93
0.420 0.93
0.838 0.97
0.958 0.97
0.510 0.96
0.554 0.96
0.167 0.96
0.636 0.95
0.081 0.95
0.100 0.96
0.688 0.94
0.393 0.93
0.164 0.95
3.00e−05 0.95
0.024 0.93
0.271 0.94
0.699 0.94
0.038 0.93
0.008 0.94
1.90e−04 0.95
1.20e−14 0.94
−1.0
−0.5
0.0
0.5
BETA

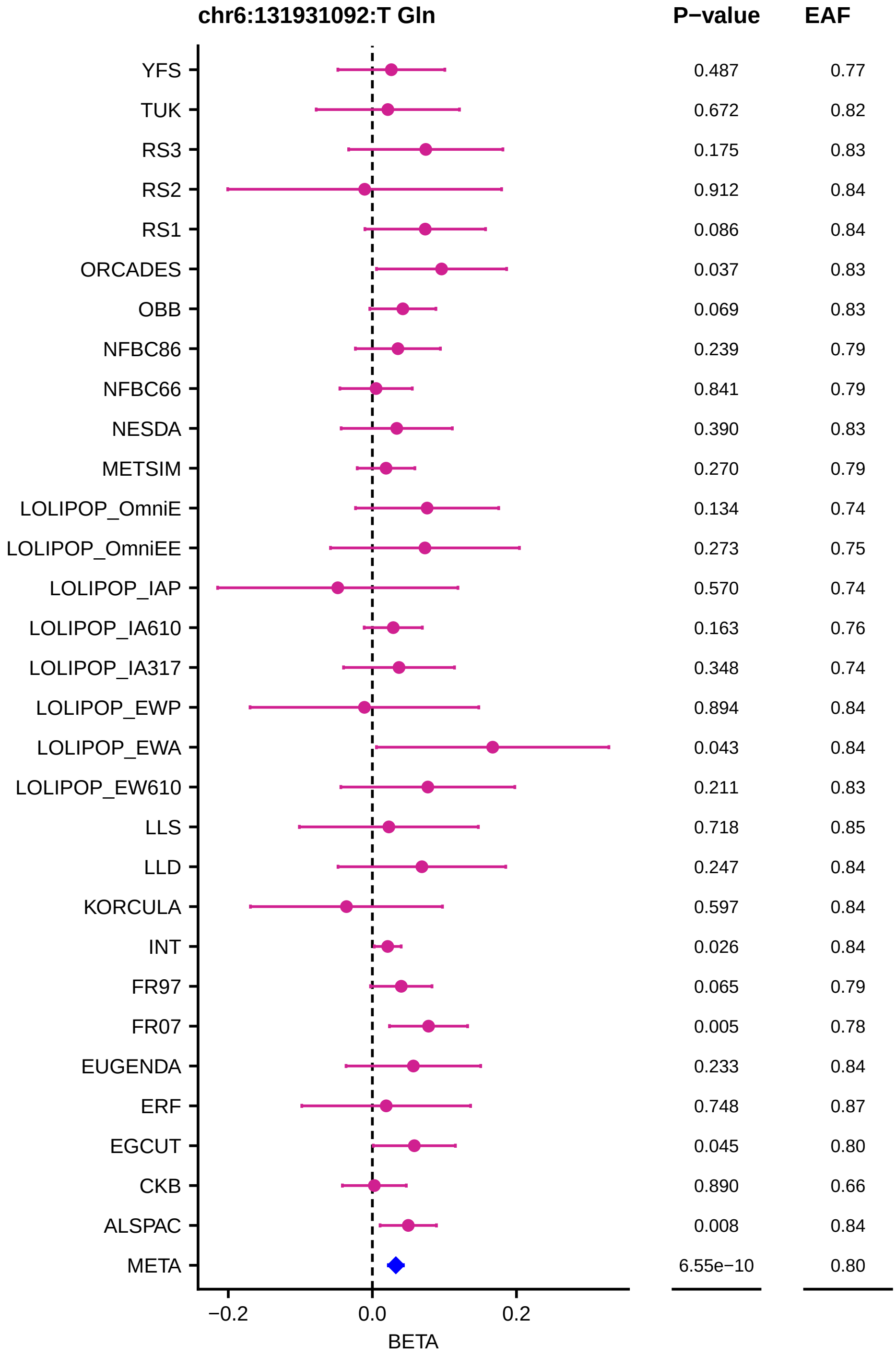
chr6:131931092:T Gln
P−value
EAF
YFS
TUK
RS3
RS2
RS1
ORCADES
OBB
NFBC86
NFBC66
NESDA
METSIM
LOLIPOP_OmniE
LOLIPOP_OmniEE
LOLIPOP_IAP
LOLIPOP_IA610
LOLIPOP_IA317
LOLIPOP_EWP
LOLIPOP_EWA
LOLIPOP_EW610
LLS
LLD
KORCULA
INT
FR97
FR07
EUGENDA
ERF
EGCUT
CKB
ALSPAC
META
0.487 0.77
0.672 0.82
0.175 0.83
0.912 0.84
0.086 0.84
0.037 0.83
0.069 0.83
0.239 0.79
0.841 0.79
0.390 0.83
0.270 0.79
0.134 0.74
0.273 0.75
0.570 0.74
0.163 0.76
0.348 0.74
0.894 0.84
0.043 0.84
0.211 0.83
0.718 0.85
0.247 0.84
0.597 0.84
0.026 0.84
0.065 0.79
0.005 0.78
0.233 0.84
0.748 0.87
0.045 0.80
0.890 0.66
0.008 0.84
6.55e−10 0.80
−0.2
0.0
0.2
BETA

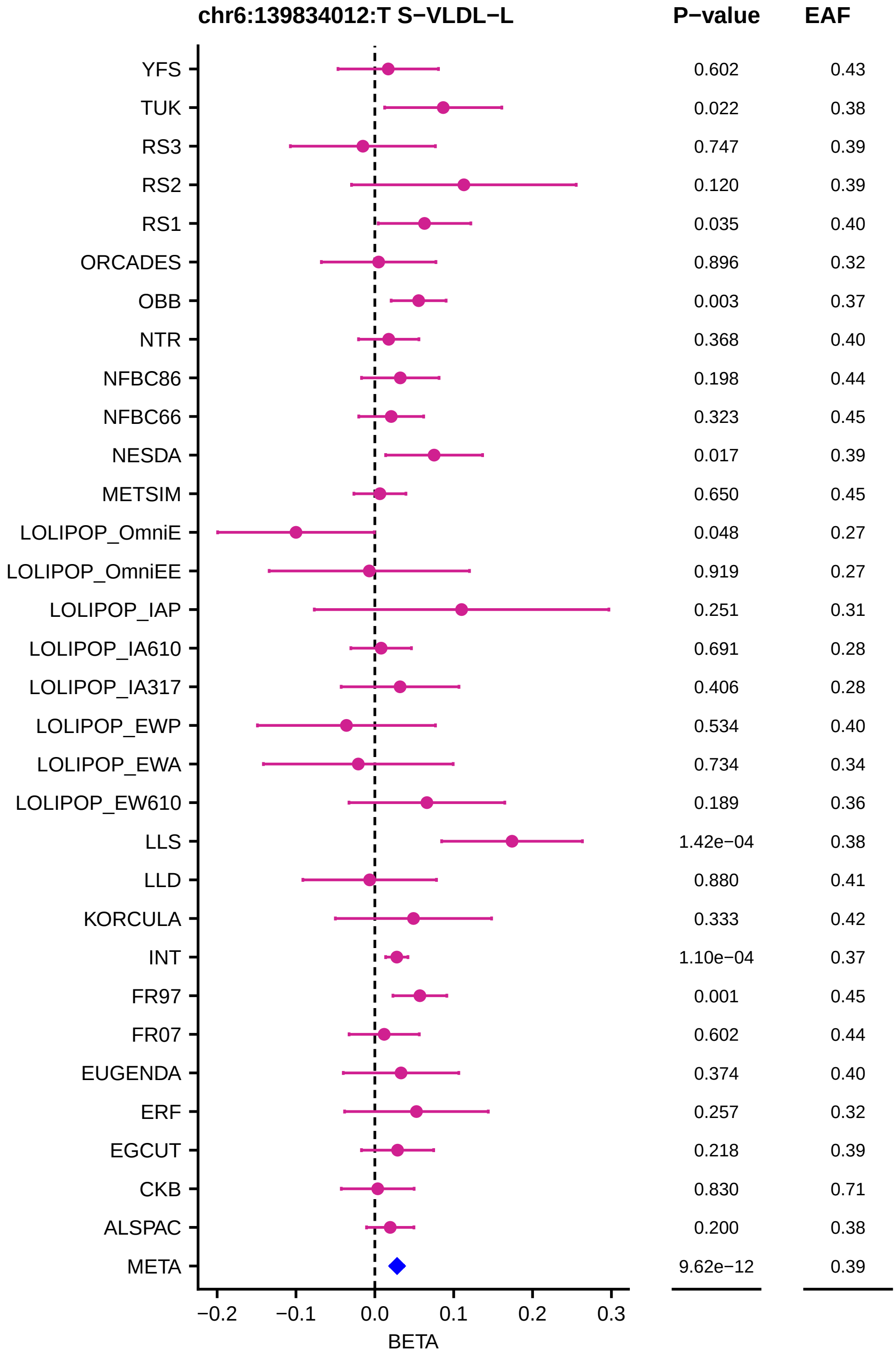
chr6:139834012:T S−VLDL−L
P−value
EAF
YFS 0.602 0.43
TUK 0.022 0.38
RS3 0.747 0.39
RS2 0.120 0.39
RS1 0.035 0.40
ORCADES 0.896 0.32
OBB 0.003 0.37
NTR 0.368 0.40
NFBC86 0.198 0.44
NFBC66 0.323 0.45
NESDA 0.017 0.39
METSIM 0.650 0.45
LOLIPOP_OmniE 0.048 0.27
LOLIPOP_OmniEE 0.919 0.27
LOLIPOP_IAP 0.251 0.31
LOLIPOP_IA610 0.691 0.28
LOLIPOP_IA317 0.406 0.28
LOLIPOP_EWP 0.534 0.40
LOLIPOP_EWA 0.734 0.34
LOLIPOP_EW610 0.189 0.36
LLS 1.42e−04 0.38
LLD 0.880 0.41
KORCULA 0.333 0.42
INT 1.10e−04 0.37
FR97 0.001 0.45
FR07 0.602 0.44
EUGENDA 0.374 0.40
ERF 0.257 0.32
EGCUT 0.218 0.39
CKB 0.830 0.71
ALSPAC 0.200 0.38
META 9.62e−12 0.39
−0.2 −0.1 0.0 0.1 0.2 0.3
BETA

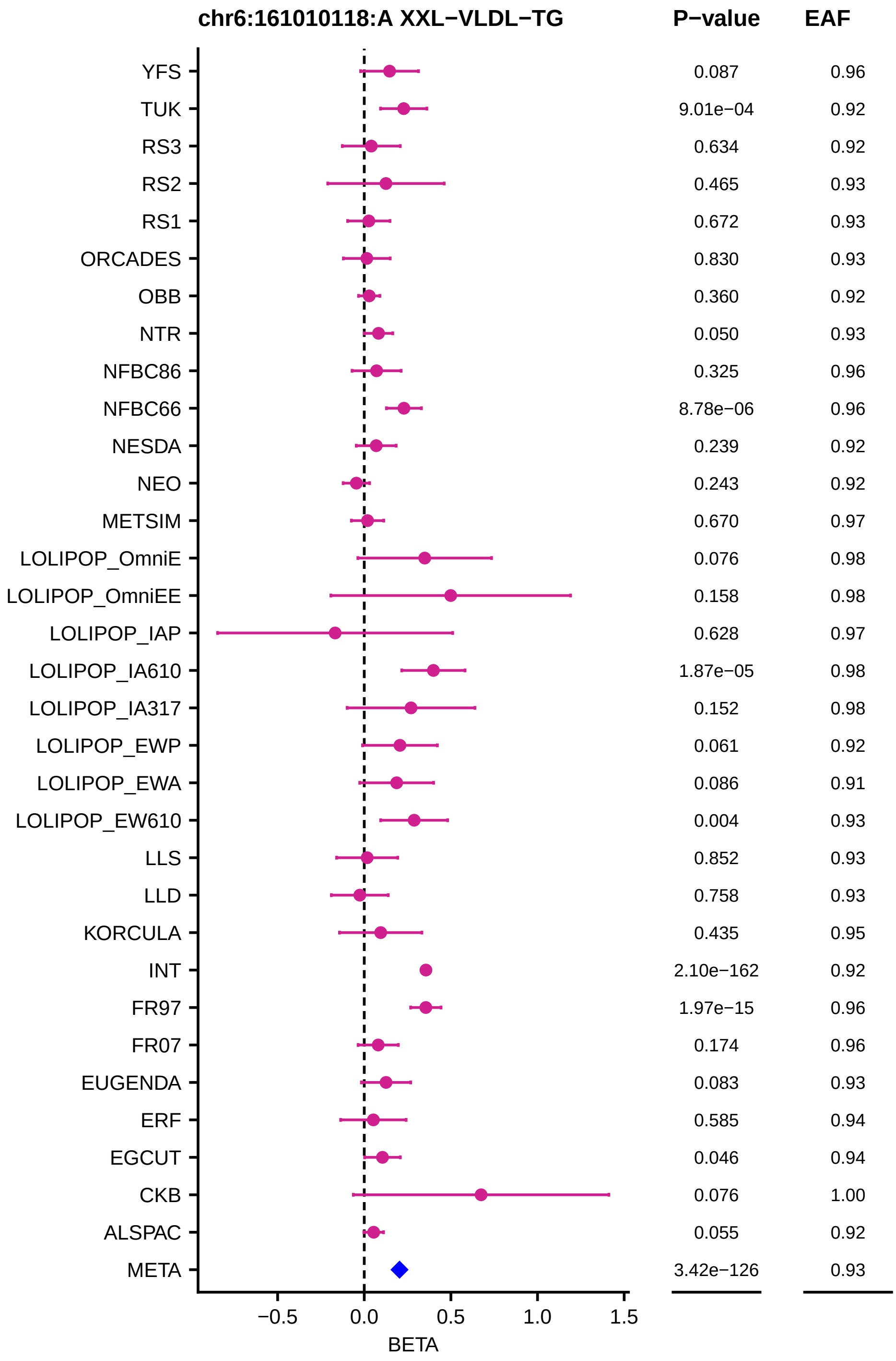
chr6:161010118:A XXL−VLDL−TG
P−value
EAF
YFS 0.087 0.96
TUK 9.01e−04 0.92
RS3 0.634 0.92
RS2 0.465 0.93
RS1 0.672 0.93
ORCADES 0.830 0.93
OBB 0.360 0.92
NTR 0.050 0.93
NFBC86 0.325 0.96
NFBC66 8.78e−06 0.96
NESDA 0.239 0.92
NEO 0.243 0.92
METSIM 0.670 0.97
LOLIPOP_OmniE 0.076 0.98
LOLIPOP_OmniEE 0.158 0.98
LOLIPOP_IAP 0.628 0.97
LOLIPOP_IA610 1.87e−05 0.98
LOLIPOP_IA317 0.152 0.98
LOLIPOP_EWP 0.061 0.92
LOLIPOP_EWA 0.086 0.91
LOLIPOP_EW610 0.004 0.93
LLS 0.852 0.93
LLD 0.758 0.93
KORCULA 0.435 0.95
INT 2.10e−162 0.92
FR97 1.97e−15 0.96
FR07 0.174 0.96
EUGENDA 0.083 0.93
ERF 0.585 0.94
EGCUT 0.046 0.94
CKB 0.076 1.00
ALSPAC 0.055 0.92
META 3.42e−126 0.93
−0.5
0.0
0.5
1.0
1.5
BETA

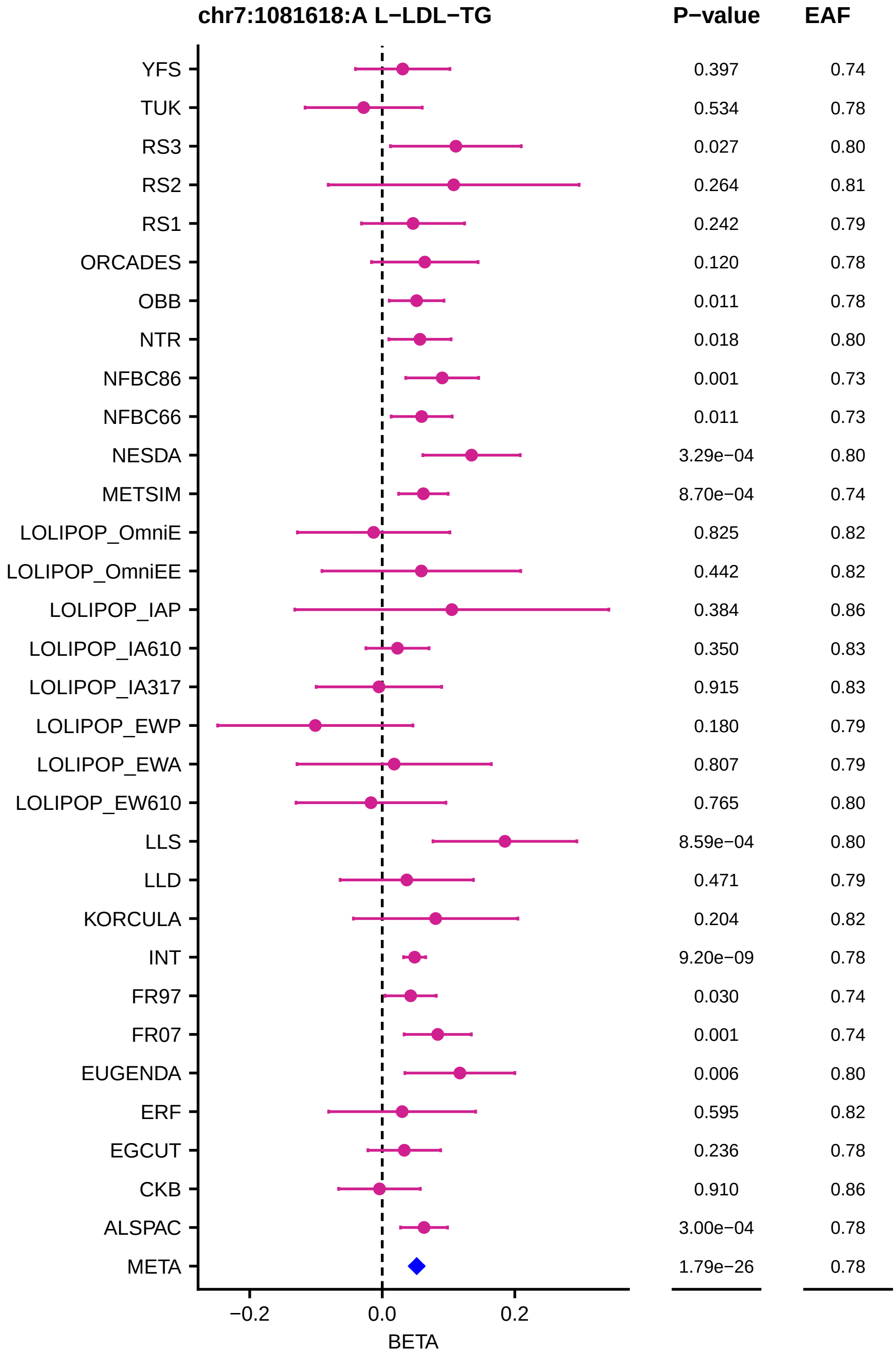
chr7:1081618:A L−LDL−TG
P−value
EAF
YFS 0.397 0.74
TUK 0.534 0.78
RS3 0.027 0.80
RS2 0.264 0.81
RS1 0.242 0.79
ORCADES 0.120 0.78
OBB 0.011 0.78
NTR 0.018 0.80
NFBC86 0.001 0.73
NFBC66 0.011 0.73
NESDA 3.29e−04 0.80
METSIM 8.70e−04 0.74
LOLIPOP_OmniE 0.825 0.82
LOLIPOP_OmniEE 0.442 0.82
LOLIPOP_IAP 0.384 0.86
LOLIPOP_IA610 0.350 0.83
LOLIPOP_IA317 0.915 0.83
LOLIPOP_EWP 0.180 0.79
LOLIPOP_EWA 0.807 0.79
LOLIPOP_EW610 0.765 0.80
LLS 8.59e−04 0.80
LLD 0.471 0.79
KORCULA 0.204 0.82
INT 9.20e−09 0.78
FR97 0.030 0.74
FR07 0.001 0.74
EUGENDA 0.006 0.80
ERF 0.595 0.82
EGCUT 0.236 0.78
CKB 0.910 0.86
ALSPAC 3.00e−04 0.78
META 1.79e−26 0.78
−0.2
0.0
0.2
BETA

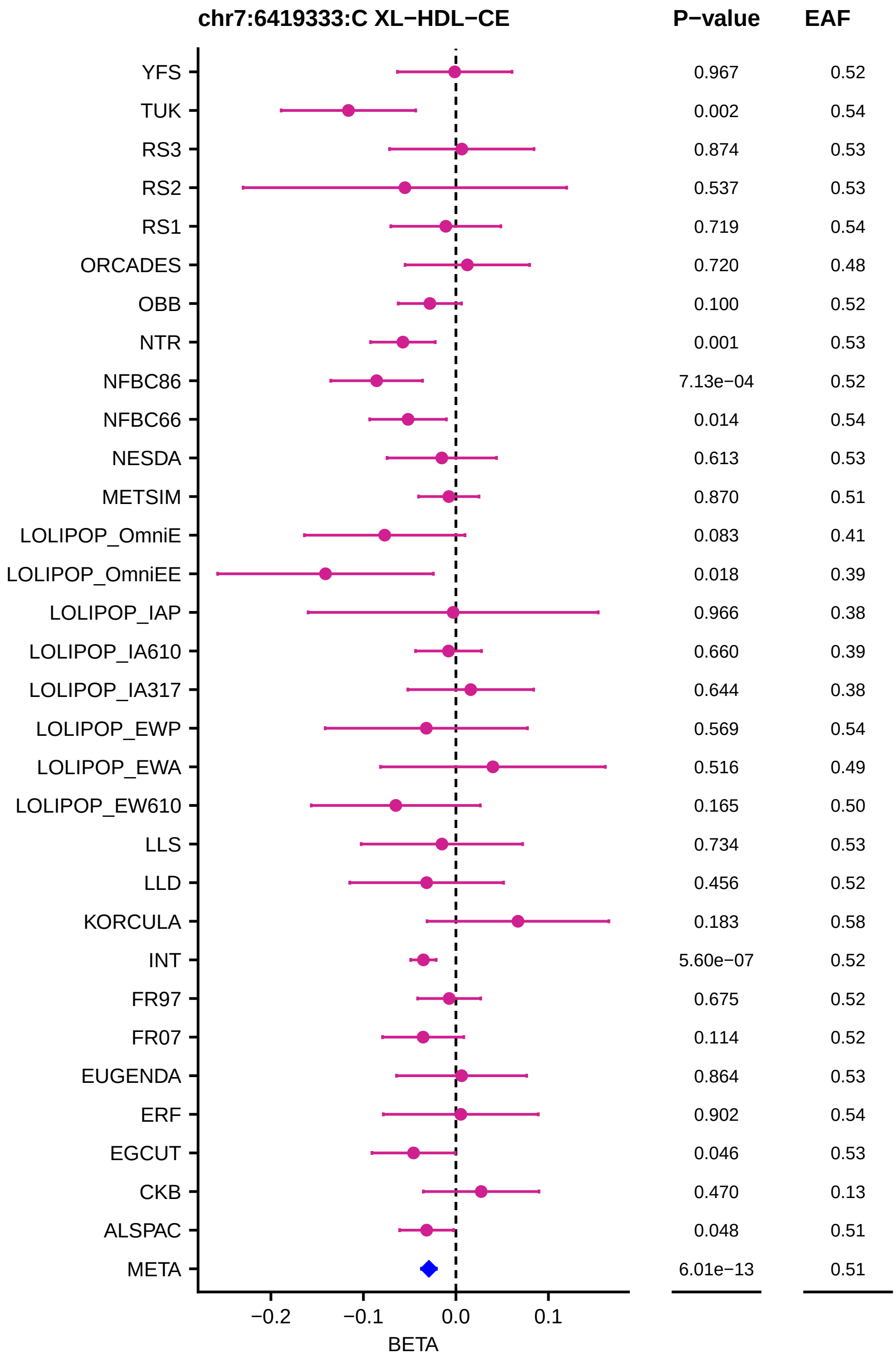
chr7:6419333:C XL−HDL−CE
P−value
EAF
YFS 0.967 0.52
TUK 0.002 0.54
RS3 0.874 0.53
RS2 0.537 0.53
RS1 0.719 0.54
ORCADES 0.720 0.48
OBB 0.100 0.52
NTR 0.001 0.53
NFBC86 7.13e−04 0.52
NFBC66 0.014 0.54
NESDA 0.613 0.53
METSIM 0.870 0.51
LOLIPOP_OmniE 0.083 0.41
LOLIPOP_OmniEE 0.018 0.39
LOLIPOP_IAP 0.966 0.38
LOLIPOP_IA610 0.660 0.39
LOLIPOP_IA317 0.644 0.38
LOLIPOP_EWP 0.569 0.54
LOLIPOP_EWA 0.516 0.49
LOLIPOP_EW610 0.165 0.50
LLS 0.734 0.53
LLD 0.456 0.52
KORCULA 0.183 0.58
INT 5.60e−07 0.52
FR97 0.675 0.52
FR07 0.114 0.52
EUGENDA 0.864 0.53
ERF 0.902 0.54
EGCUT 0.046 0.53
CKB 0.470 0.13
ALSPAC 0.048 0.51
META 6.01e−13 0.51
−0.2
−0.1
0.0
0.1
BETA

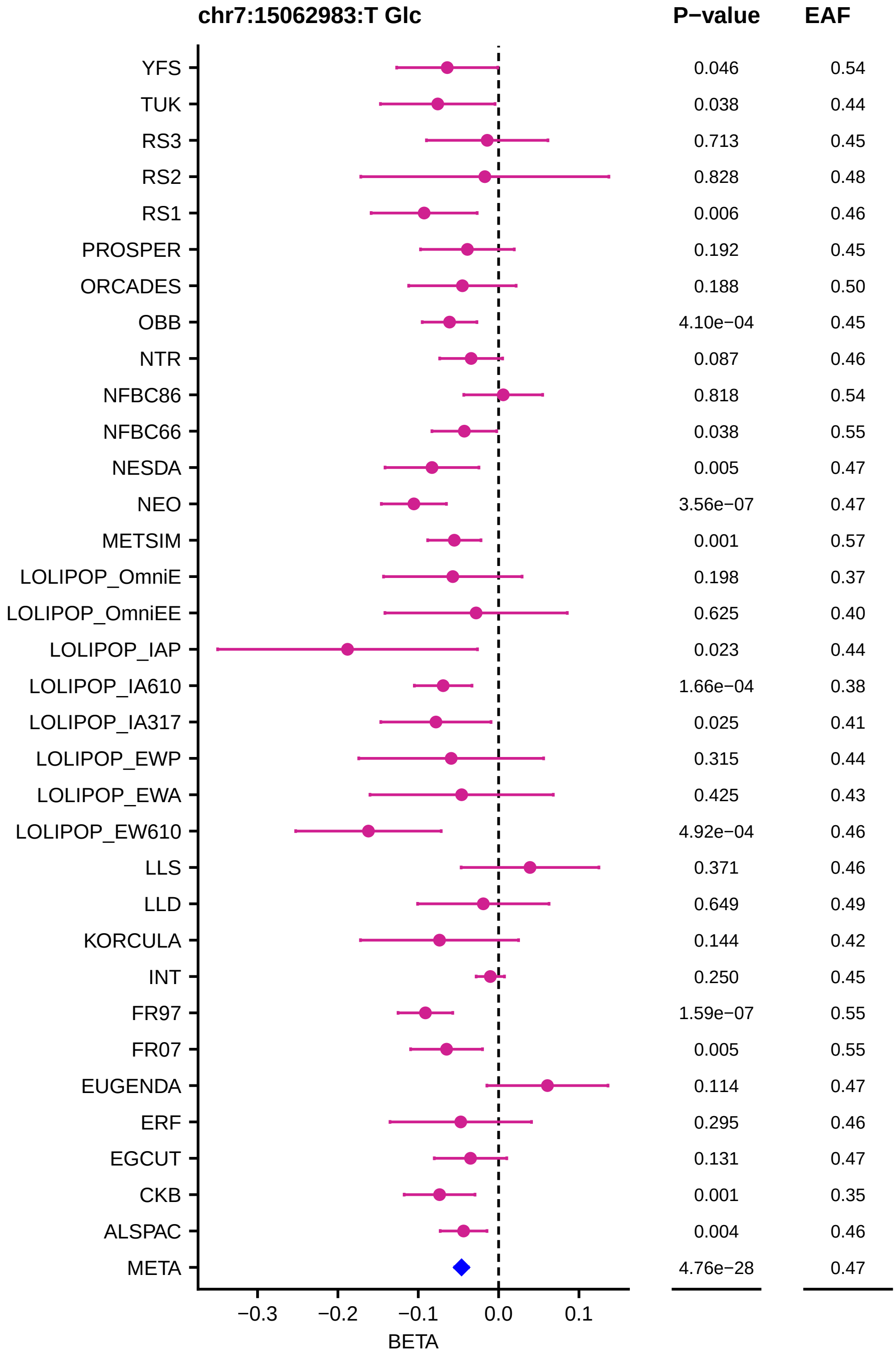
chr7:15062983:T Glc
P−value
EAF
YFS 0.046 0.54
TUK 0.038 0.44
RS3 0.713 0.45
RS2 0.828 0.48
RS1 0.006 0.46
PROSPER 0.192 0.45
ORCADES 0.188 0.50
OBB 4.10e−04 0.45
NTR 0.087 0.46
NFBC86 0.818 0.54
NFBC66 0.038 0.55
NESDA 0.005 0.47
NEO 3.56e−07 0.47
METSIM 0.001 0.57
LOLIPOP_OmniE 0.198 0.37
LOLIPOP_OmniEE 0.625 0.40
LOLIPOP_IAP 0.023 0.44
LOLIPOP_IA610 1.66e−04 0.38
LOLIPOP_IA317 0.025 0.41
LOLIPOP_EWP 0.315 0.44
LOLIPOP_EWA 0.425 0.43
LOLIPOP_EW610 4.92e−04 0.46
LLS 0.371 0.46
LLD 0.649 0.49
KORCULA 0.144 0.42
INT 0.250 0.45
FR97 1.59e−07 0.55
FR07 0.005 0.55
EUGENDA 0.114 0.47
ERF 0.295 0.46
EGCUT 0.131 0.47
CKB 0.001 0.35
ALSPAC 0.004 0.46
META 4.76e−28 0.47
−0.3
−0.2
−0.1
0.0
0.1
BETA

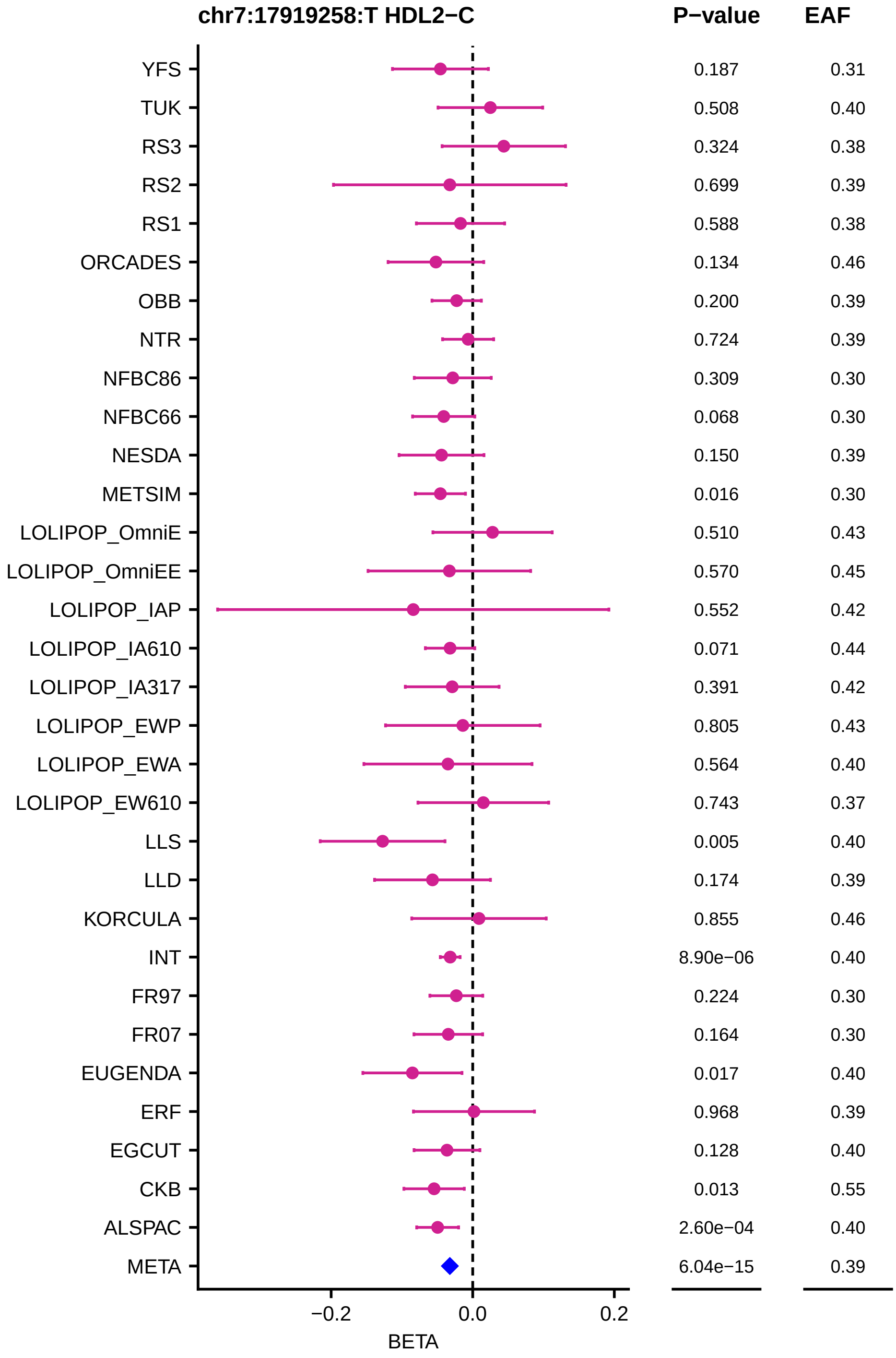

chr7:17919258:T HDL2−C
P−value
EAF
YFS 0.187 0.31
TUK 0.508 0.40
RS3 0.324 0.38
RS2 0.699 0.39
RS1 0.588 0.38
ORCADES 0.134 0.46
OBB 0.200 0.39
NTR 0.724 0.39
NFBC86 0.309 0.30
NFBC66 0.068 0.30
NESDA 0.150 0.39
METSIM 0.016 0.30
LOLIPOP_OmniE 0.510 0.43
LOLIPOP_OmniEE 0.570 0.45
LOLIPOP_IAP 0.552 0.42
LOLIPOP_IA610 0.071 0.44
LOLIPOP_IA317 0.391 0.42
LOLIPOP_EWP 0.805 0.43
LOLIPOP_EWA 0.564 0.40
LOLIPOP_EW610 0.743 0.37
LLS 0.005 0.40
LLD 0.174 0.39
KORCULA 0.855 0.46
INT 8.90e−06 0.40
FR97 0.224 0.30
FR07 0.164 0.30
EUGENDA 0.017 0.40
ERF 0.968 0.39
EGCUT 0.128 0.40
CKB 0.013 0.55
ALSPAC 2.60e−04 0.40
META 6.04e−15 0.39
−0.2
0.0
0.2
BETA

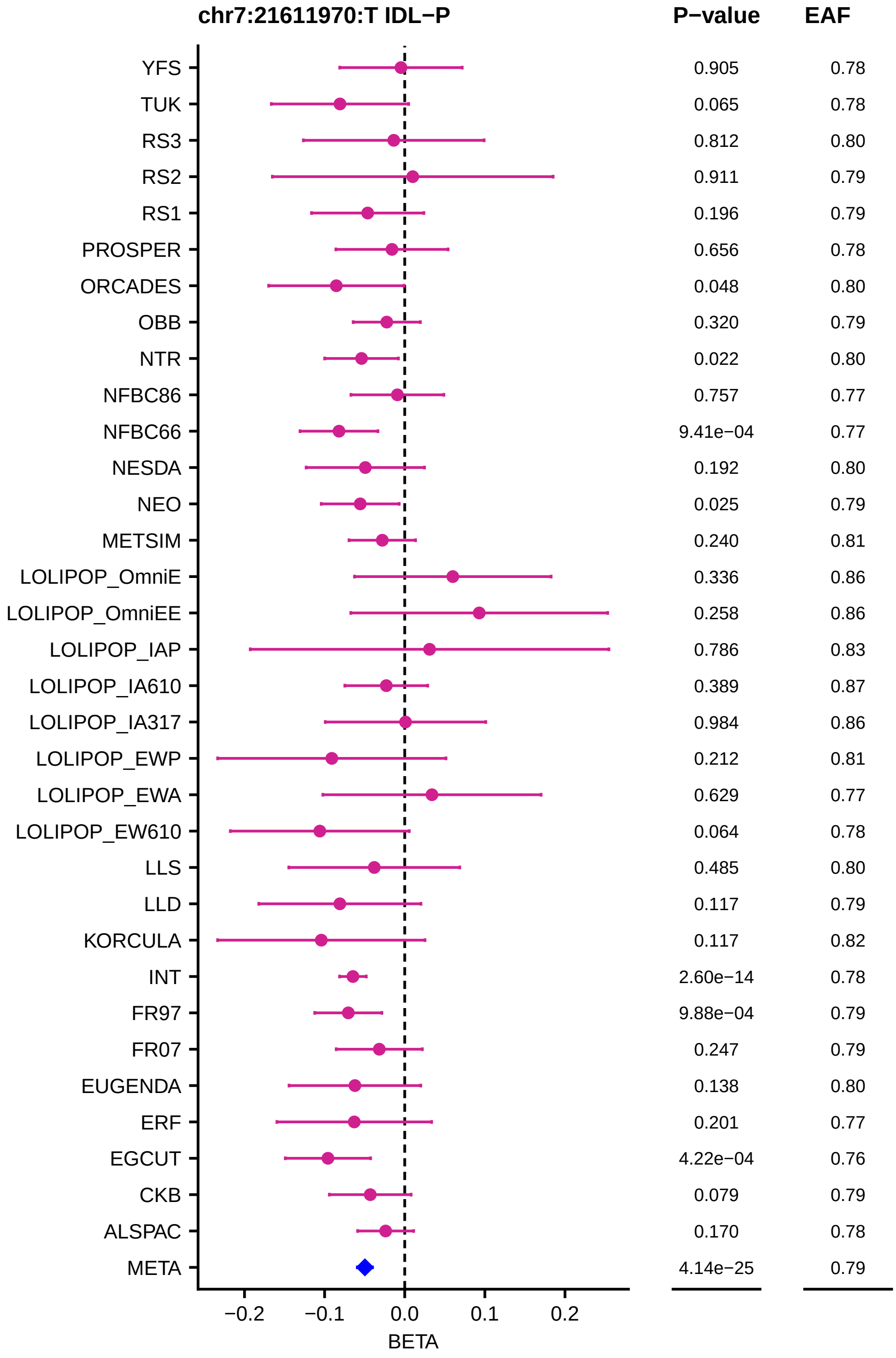

chr7:21611970:T IDL−P
P−value
EAF
YFS 0.905 0.78
TUK 0.065 0.78
RS3 0.812 0.80
RS2 0.911 0.79
RS1 0.196 0.79
PROSPER 0.656 0.78
ORCADES 0.048 0.80
OBB 0.320 0.79
NTR 0.022 0.80
NFBC86 0.757 0.77
NFBC66 9.41e−04 0.77
NESDA 0.192 0.80
NEO 0.025 0.79
METSIM 0.240 0.81
LOLIPOP_OmniE 0.336 0.86
LOLIPOP_OmniEE 0.258 0.86
LOLIPOP_IAP 0.786 0.83
LOLIPOP_IA610 0.389 0.87
LOLIPOP_IA317 0.984 0.86
LOLIPOP_EWP 0.212 0.81
LOLIPOP_EWA 0.629 0.77
LOLIPOP_EW610 0.064 0.78
LLS 0.485 0.80
LLD 0.117 0.79
KORCULA 0.117 0.82
INT 2.60e−14 0.78
FR97 9.88e−04 0.79
FR07 0.247 0.79
EUGENDA 0.138 0.80
ERF 0.201 0.77
EGCUT 4.22e−04 0.76
CKB 0.079 0.79
ALSPAC 0.170 0.78
META 4.14e−25 0.79
−0.2
−0.1
0.0
0.1
0.2
BETA

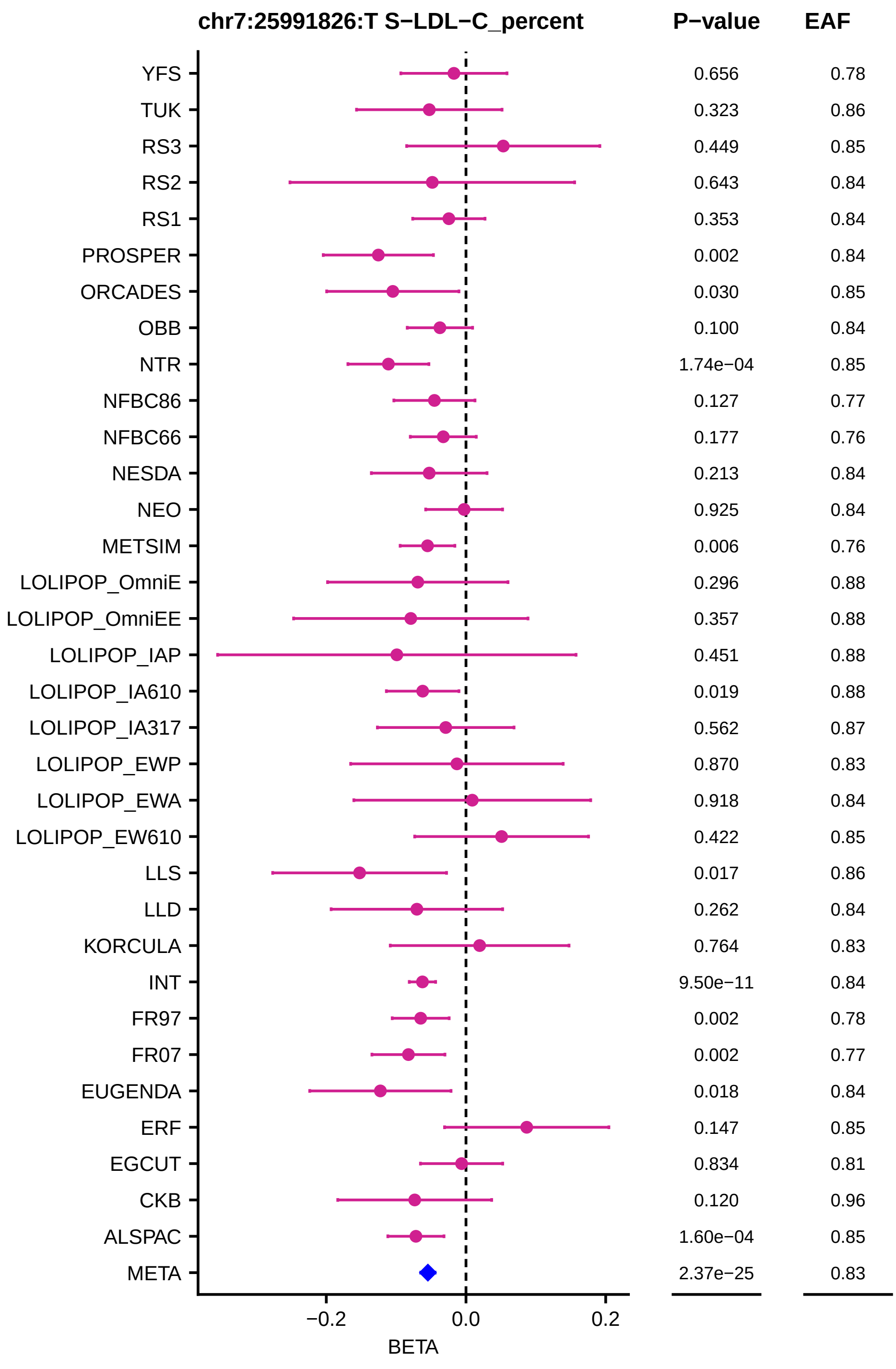
chr7:25991826:T S−LDL−C_percent
P−value
EAF
YFS 0.656 0.78
TUK 0.323 0.86
RS3 0.449 0.85
RS2 0.643 0.84
RS1 0.353 0.84
PROSPER 0.002 0.84
ORCADES 0.030 0.85
OBB 0.100 0.84
NTR 1.74e−04 0.85
NFBC86 0.127 0.77
NFBC66 0.177 0.76
NESDA 0.213 0.84
NEO 0.925 0.84
METSIM 0.006 0.76
LOLIPOP_OmniE 0.296 0.88
LOLIPOP_OmniEE 0.357 0.88
LOLIPOP_IAP 0.451 0.88
LOLIPOP_IA610 0.019 0.88
LOLIPOP_IA317 0.562 0.87
LOLIPOP_EWP 0.870 0.83
LOLIPOP_EWA 0.918 0.84
LOLIPOP_EW610 0.422 0.85
LLS 0.017 0.86
LLD 0.262 0.84
KORCULA 0.764 0.83
INT 9.50e−11 0.84
FR97 0.002 0.78
FR07 0.002 0.77
EUGENDA 0.018 0.84
ERF 0.147 0.85
EGCUT 0.834 0.81
CKB 0.120 0.96
ALSPAC 1.60e−04 0.85
META 2.37e−25 0.83
−0.2
0.0
0.2
BETA

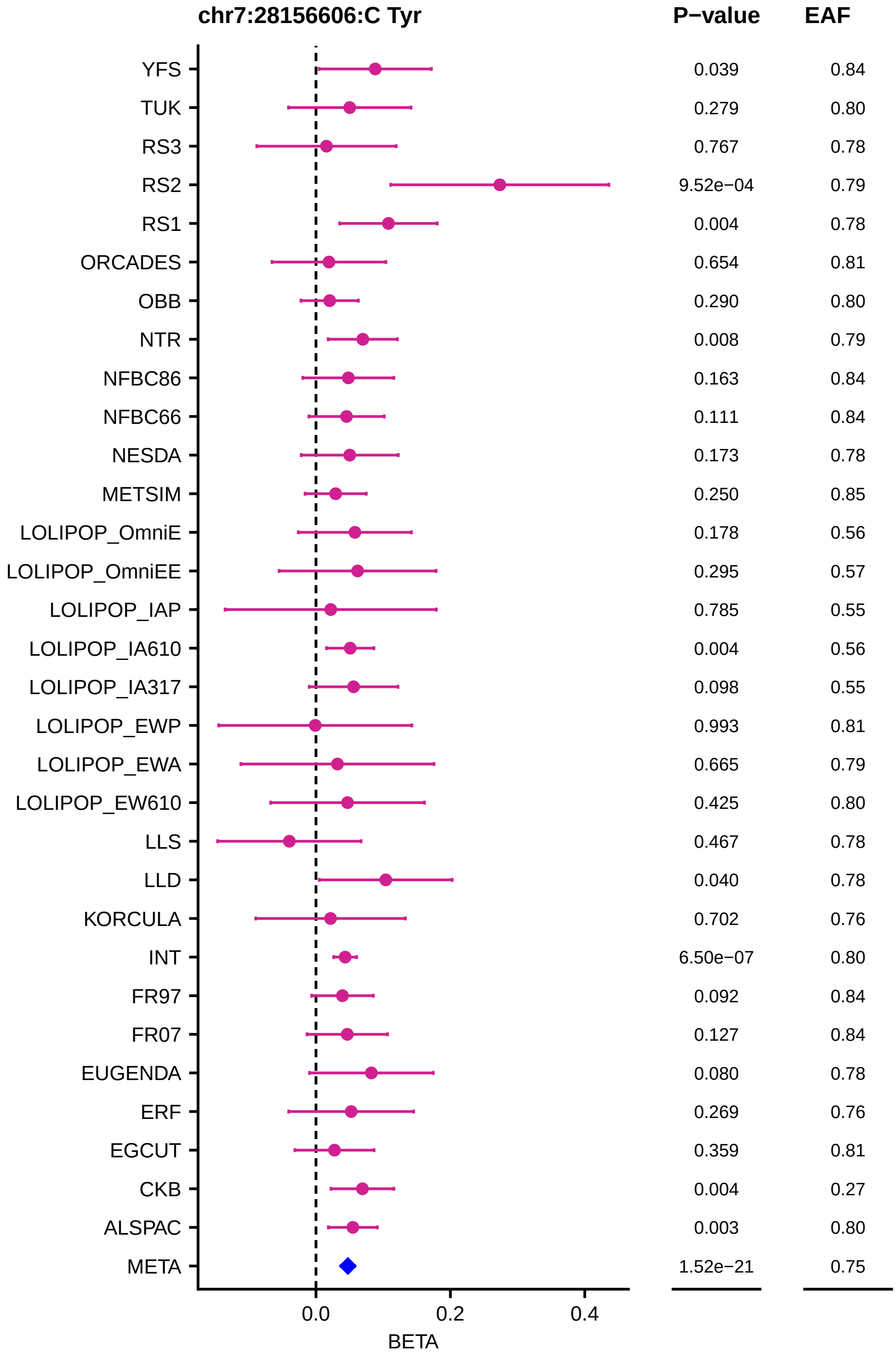
chr7:28156606:C Tyr
P−value
EAF
YFS 0.039 0.84
TUK 0.279 0.80
RS3 0.767 0.78
RS2 9.52e−04 0.79
RS1 0.004 0.78
ORCADES 0.654 0.81
OBB 0.290 0.80
NTR 0.008 0.79
NFBC86 0.163 0.84
NFBC66 0.111 0.84
NESDA 0.173 0.78
METSIM 0.250 0.85
LOLIPOP_OmniE 0.178 0.56
LOLIPOP_OmniEE 0.295 0.57
LOLIPOP_IAP 0.785 0.55
LOLIPOP_IA610 0.004 0.56
LOLIPOP_IA317 0.098 0.55
LOLIPOP_EWP 0.993 0.81
LOLIPOP_EWA 0.665 0.79
LOLIPOP_EW610 0.425 0.80
LLS 0.467 0.78
LLD 0.040 0.78
KORCULA 0.702 0.76
INT 6.50e−07 0.80
FR97 0.092 0.84
FR07 0.127 0.84
EUGENDA 0.080 0.78
ERF 0.269 0.76
EGCUT 0.359 0.81
CKB 0.004 0.27
ALSPAC 0.003 0.80
META 1.52e−21 0.75
0.0
0.2
0.4
BETA

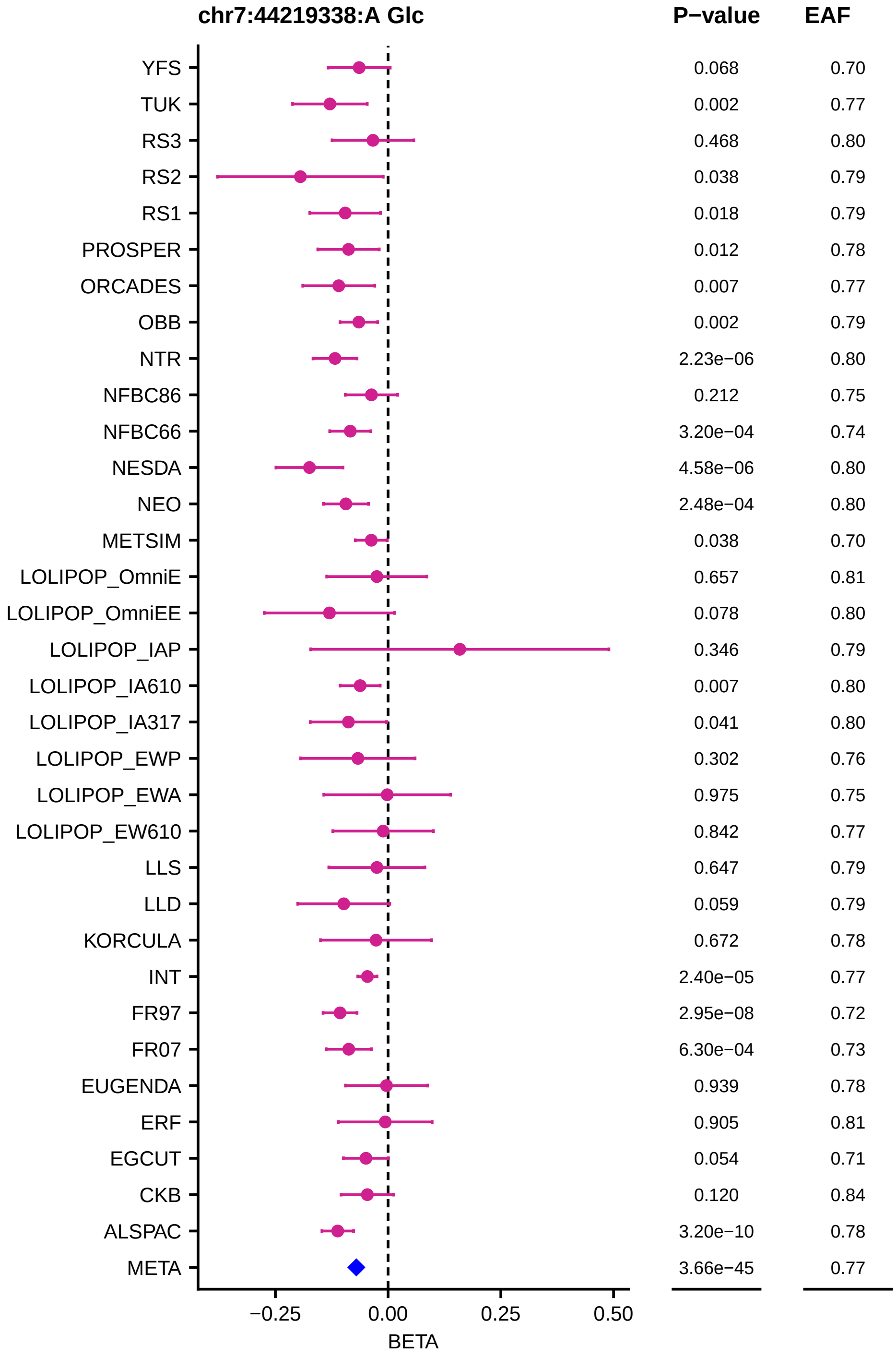

chr7:44219338:A Glc
P−value
EAF
YFS 0.068 0.70
TUK 0.002 0.77
RS3 0.468 0.80
RS2 0.038 0.79
RS1 0.018 0.79
PROSPER 0.012 0.78
ORCADES 0.007 0.77
OBB 0.002 0.79
NTR 2.23e−06 0.80
NFBC86 0.212 0.75
NFBC66 3.20e−04 0.74
NESDA 4.58e−06 0.80
NEO 2.48e−04 0.80
METSIM 0.038 0.70
LOLIPOP_OmniE 0.657 0.81
LOLIPOP_OmniEE 0.078 0.80
LOLIPOP_IAP 0.346 0.79
LOLIPOP_IA610 0.007 0.80
LOLIPOP_IA317 0.041 0.80
LOLIPOP_EWP 0.302 0.76
LOLIPOP_EWA 0.975 0.75
LOLIPOP_EW610 0.842 0.77
LLS 0.647 0.79
LLD 0.059 0.79
KORCULA 0.672 0.78
INT 2.40e−05 0.77
FR97 2.95e−08 0.72
FR07 6.30e−04 0.73
EUGENDA 0.939 0.78
ERF 0.905 0.81
EGCUT 0.054 0.71
CKB 0.120 0.84
ALSPAC 3.20e−10 0.78
META 3.66e−45 0.77
−0.25
0.00
0.25
0.50
BETA

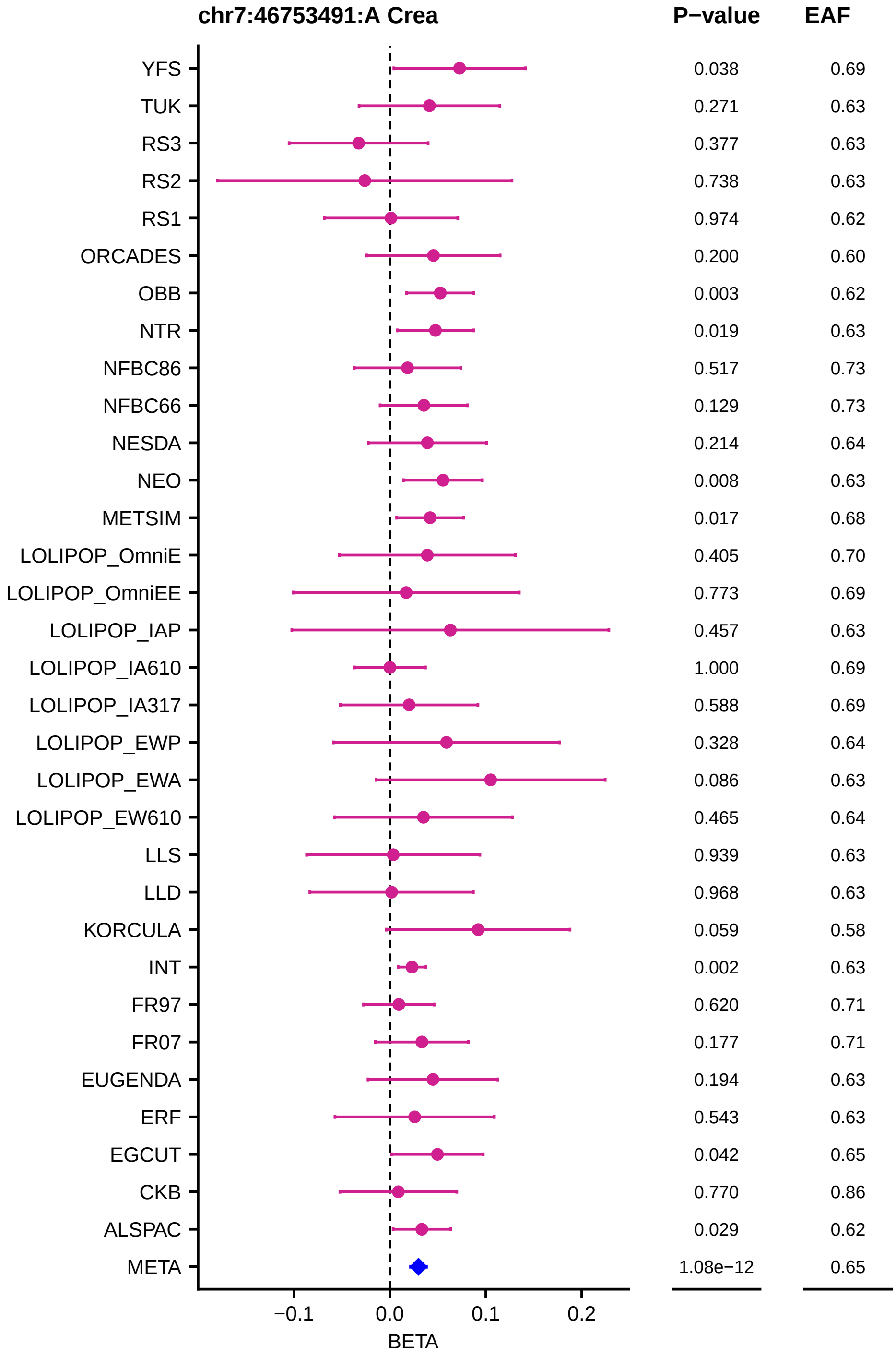
chr7:46753491:A Crea
P−value
EAF
YFS 0.038 0.69
TUK 0.271 0.63
RS3 0.377 0.63
RS2 0.738 0.63
RS1 0.974 0.62
ORCADES 0.200 0.60
OBB 0.003 0.62
NTR 0.019 0.63
NFBC86 0.517 0.73
NFBC66 0.129 0.73
NESDA 0.214 0.64
NEO 0.008 0.63
METSIM 0.017 0.68
LOLIPOP_OmniE 0.405 0.70
LOLIPOP_OmniEE 0.773 0.69
LOLIPOP_IAP 0.457 0.63
LOLIPOP_IA610 1.000 0.69
LOLIPOP_IA317 0.588 0.69
LOLIPOP_EWP 0.328 0.64
LOLIPOP_EWA 0.086 0.63
LOLIPOP_EW610 0.465 0.64
LLS 0.939 0.63
LLD 0.968 0.63
KORCULA 0.059 0.58
INT 0.002 0.63
FR97 0.620 0.71
FR07 0.177 0.71
EUGENDA 0.194 0.63
ERF 0.543 0.63
EGCUT 0.042 0.65
CKB 0.770 0.86
ALSPAC 0.029 0.62
META 1.08e−12 0.65
−0.1
0.0
0.1
0.2
BETA

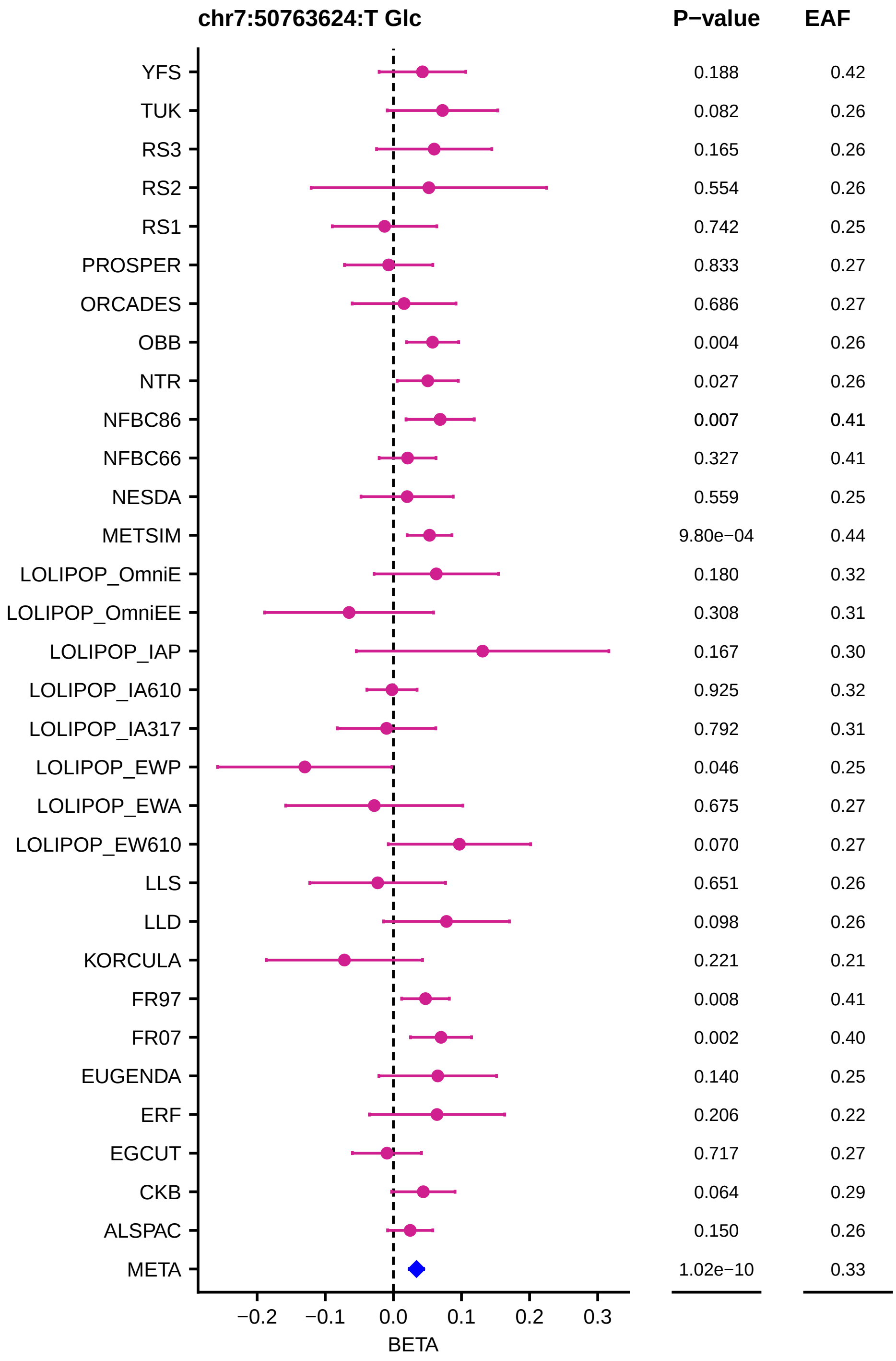

chr7:50763624:T Glc
P−value
EAF
YFS
TUK
RS3
RS2
RS1
PROSPER
ORCADES
OBB
NTR
NFBC86
NFBC66
NESDA
METSIM
LOLIPOP_OmniE
LOLIPOP_OmniEE
LOLIPOP_IAP
LOLIPOP_IA610
LOLIPOP_IA317
LOLIPOP_EWP
LOLIPOP_EWA
LOLIPOP_EW610
LLS
LLD
KORCULA
FR97
FR07
EUGENDA
ERF
EGCUT
CKB
ALSPAC
META
0.188 0.42
0.082 0.26
0.165 0.26
0.554 0.26
0.742 0.25
0.833 0.27
0.686 0.27
0.004 0.26
0.027 0.26
0.007 0.41
0.327 0.41
0.559 0.25
9.80e−04 0.44
0.180 0.32
0.308 0.31
0.167 0.30
0.925 0.32
0.792 0.31
0.046 0.25
0.675 0.27
0.070 0.27
0.651 0.26
0.098 0.26
0.221 0.21
0.008 0.41
0.002 0.40
0.140 0.25
0.206 0.22
0.717 0.27
0.064 0.29
0.150 0.26
1.02e−10 0.33
−0.2
−0.1
0.0
0.1
0.2
0.3
BETA

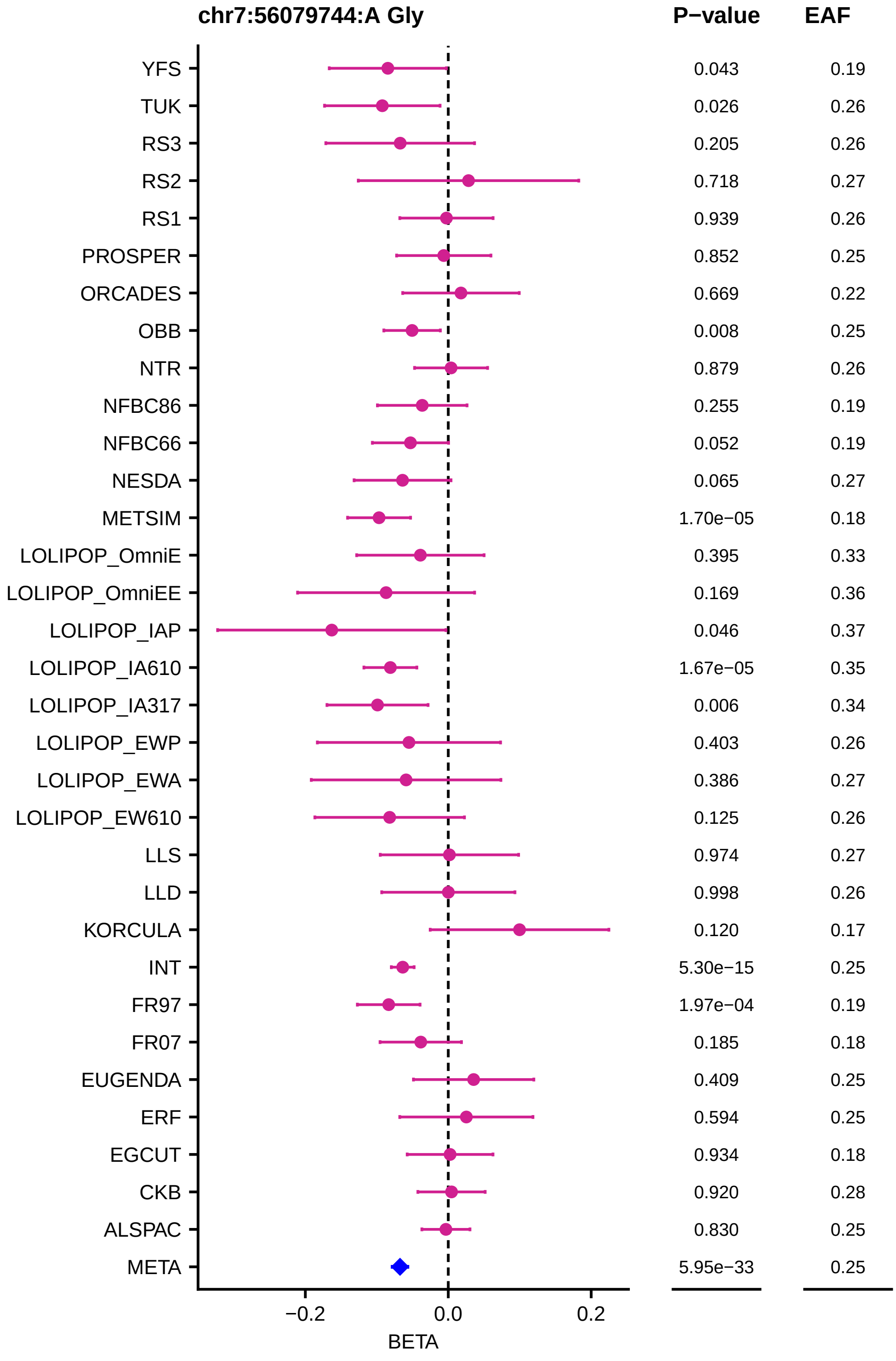
chr7:56079744:A Gly
P−value
EAF
YFS 0.043 0.19
TUK 0.026 0.26
RS3 0.205 0.26
RS2 0.718 0.27
RS1 0.939 0.26
PROSPER 0.852 0.25
ORCADES 0.669 0.22
OBB 0.008 0.25
NTR 0.879 0.26
NFBC86 0.255 0.19
NFBC66 0.052 0.19
NESDA 0.065 0.27
METSIM 1.70e−05 0.18
LOLIPOP_OmniE 0.395 0.33
LOLIPOP_OmniEE 0.169 0.36
LOLIPOP_IAP 0.046 0.37
LOLIPOP_IA610 1.67e−05 0.35
LOLIPOP_IA317 0.006 0.34
LOLIPOP_EWP 0.403 0.26
LOLIPOP_EWA 0.386 0.27
LOLIPOP_EW610 0.125 0.26
LLS 0.974 0.27
LLD 0.998 0.26
KORCULA 0.120 0.17
INT 5.30e−15 0.25
FR97 1.97e−04 0.19
FR07 0.185 0.18
EUGENDA 0.409 0.25
ERF 0.594 0.25
EGCUT 0.934 0.18
CKB 0.920 0.28
ALSPAC 0.830 0.25
META 5.95e−33 0.25
−0.2
0.0
0.2
BETA

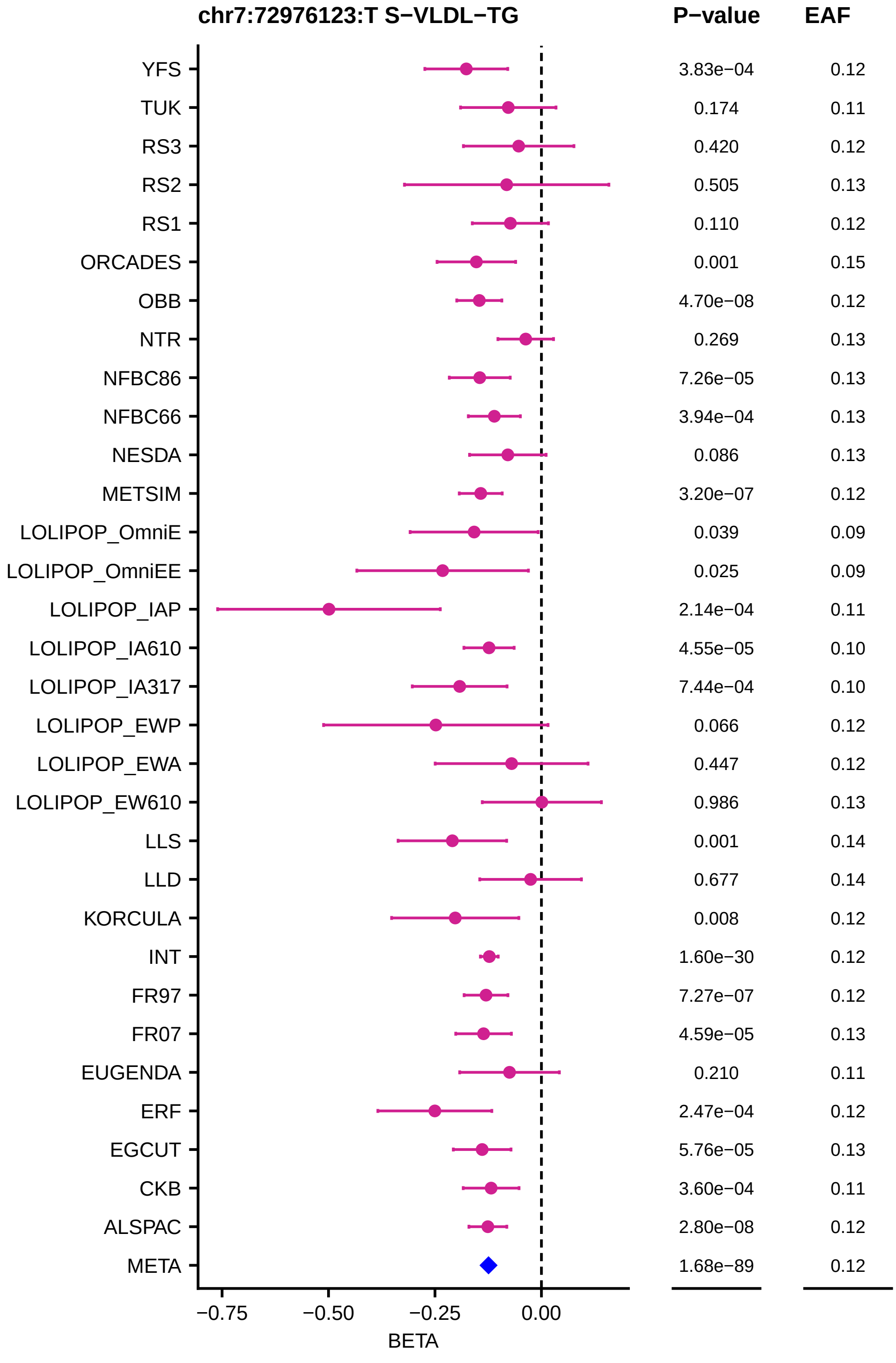

chr7:72976123:T S−VLDL−TG
P−value
EAF
YFS 3.83e−04 0.12
TUK 0.174 0.11
RS3 0.420 0.12
RS2 0.505 0.13
RS1 0.110 0.12
ORCADES 0.001 0.15
OBB 4.70e−08 0.12
NTR 0.269 0.13
NFBC86 7.26e−05 0.13
NFBC66 3.94e−04 0.13
NESDA 0.086 0.13
METSIM 3.20e−07 0.12
LOLIPOP_OmniE 0.039 0.09
LOLIPOP_OmniEE 0.025 0.09
LOLIPOP_IAP 2.14e−04 0.11
LOLIPOP_IA610 4.55e−05 0.10
LOLIPOP_IA317 7.44e−04 0.10
LOLIPOP_EWP 0.066 0.12
LOLIPOP_EWA 0.447 0.12
LOLIPOP_EW610 0.986 0.13
LLS 0.001 0.14
LLD 0.677 0.14
KORCULA 0.008 0.12
INT 1.60e−30 0.12
FR97 7.27e−07 0.12
FR07 4.59e−05 0.13
EUGENDA 0.210 0.11
ERF 2.47e−04 0.12
EGCUT 5.76e−05 0.13
CKB 3.60e−04 0.11
ALSPAC 2.80e−08 0.12
META 1.68e−89 0.12
−0.75
−0.50
−0.25
0.00
BETA

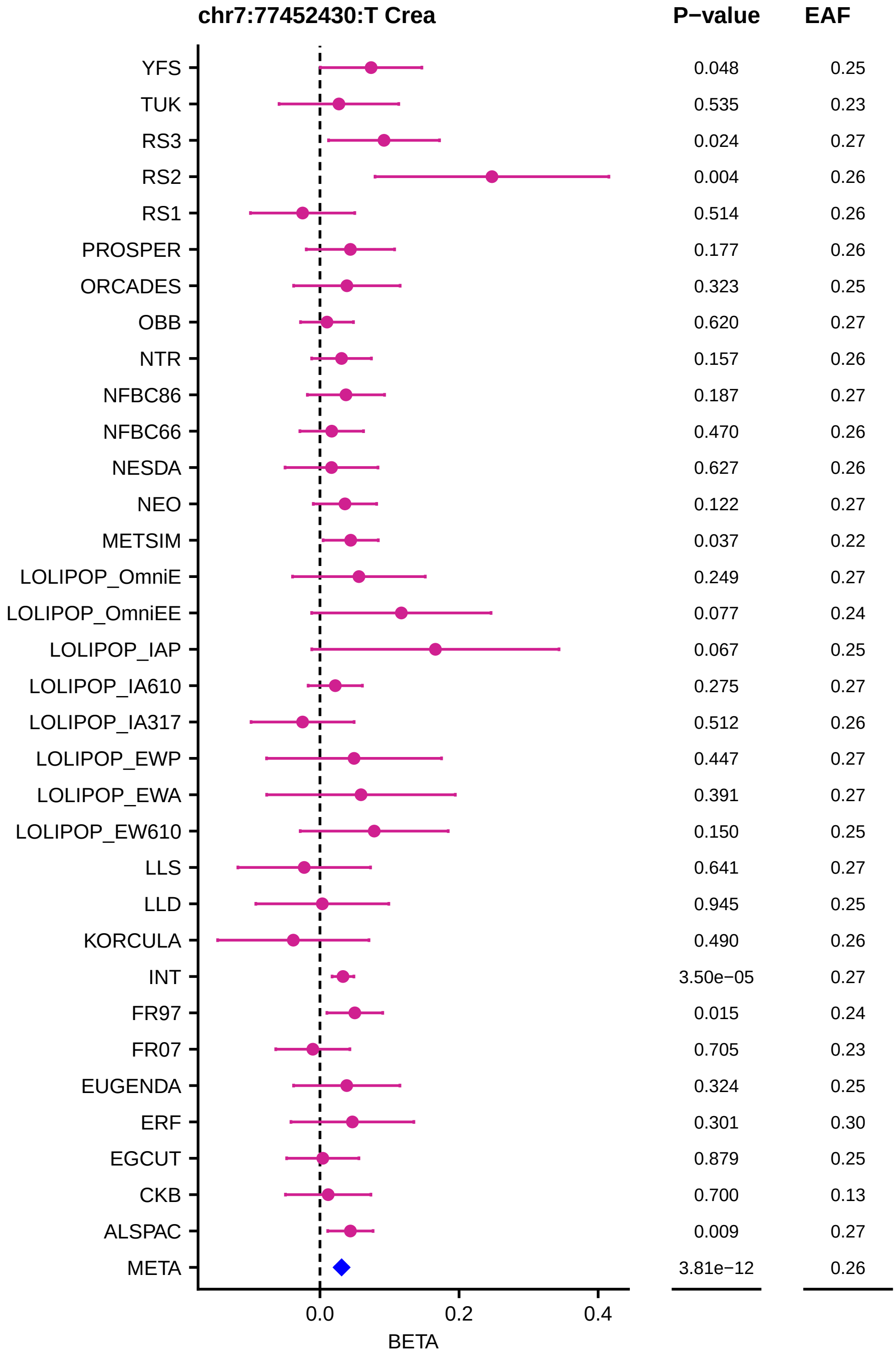
chr7:77452430:T Crea
P−value
EAF
YFS 0.048 0.25
TUK 0.535 0.23
RS3 0.024 0.27
RS2 0.004 0.26
RS1 0.514 0.26
PROSPER 0.177 0.26
ORCADES 0.323 0.25
OBB 0.620 0.27
NTR 0.157 0.26
NFBC86 0.187 0.27
NFBC66 0.470 0.26
NESDA 0.627 0.26
NEO 0.122 0.27
METSIM 0.037 0.22
LOLIPOP_OmniE 0.249 0.27
LOLIPOP_OmniEE 0.077 0.24
LOLIPOP_IAP 0.067 0.25
LOLIPOP_IA610 0.275 0.27
LOLIPOP_IA317 0.512 0.26
LOLIPOP_EWP 0.447 0.27
LOLIPOP_EWA 0.391 0.27
LOLIPOP_EW610 0.150 0.25
LLS 0.641 0.27
LLD 0.945 0.25
KORCULA 0.490 0.26
INT 3.50e−05 0.27
FR97 0.015 0.24
FR07 0.705 0.23
EUGENDA 0.324 0.25
ERF 0.301 0.30
EGCUT 0.879 0.25
CKB 0.700 0.13
ALSPAC 0.009 0.27
META 3.81e−12 0.26
0.0
0.2
0.4
BETA

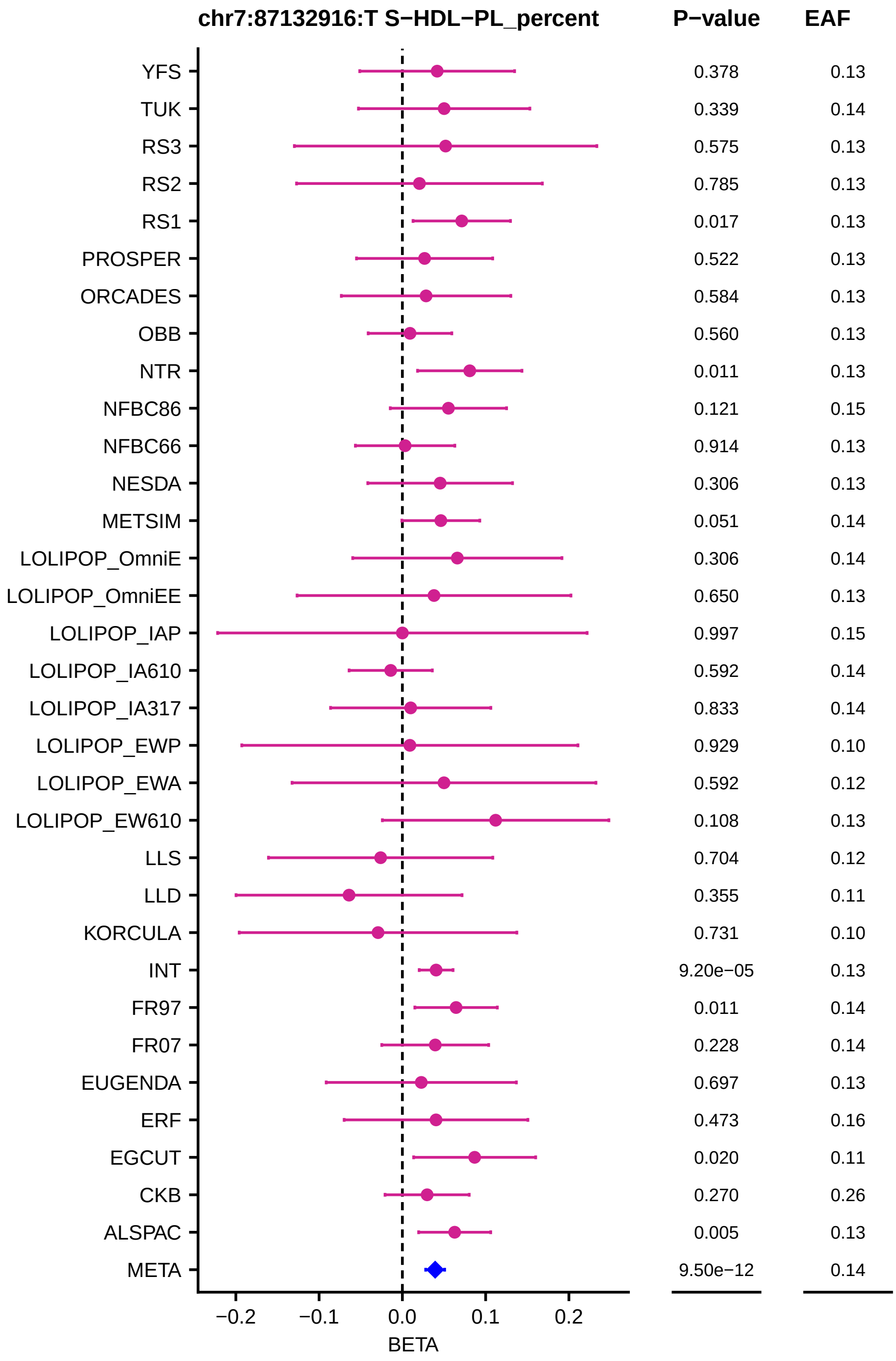
chr7:87132916:T S−HDL−PL_percent
P−value
EAF
YFS 0.378 0.13
TUK 0.339 0.14
RS3 0.575 0.13
RS2 0.785 0.13
RS1 0.017 0.13
PROSPER 0.522 0.13
ORCADES 0.584 0.13
OBB 0.560 0.13
NTR 0.011 0.13
NFBC86 0.121 0.15
NFBC66 0.914 0.13
NESDA 0.306 0.13
METSIM 0.051 0.14
LOLIPOP_OmniE 0.306 0.14
LOLIPOP_OmniEE 0.650 0.13
LOLIPOP_IAP 0.997 0.15
LOLIPOP_IA610 0.592 0.14
LOLIPOP_IA317 0.833 0.14
LOLIPOP_EWP 0.929 0.10
LOLIPOP_EWA 0.592 0.12
LOLIPOP_EW610 0.108 0.13
LLS 0.704 0.12
LLD 0.355 0.11
KORCULA 0.731 0.10
INT 9.20e−05 0.13
FR97 0.011 0.14
FR07 0.228 0.14
EUGENDA 0.697 0.13
ERF 0.473 0.16
EGCUT 0.020 0.11
CKB 0.270 0.26
ALSPAC 0.005 0.13
META 9.50e−12 0.14
−0.2 −0.1 0.0 0.1 0.2
BETA

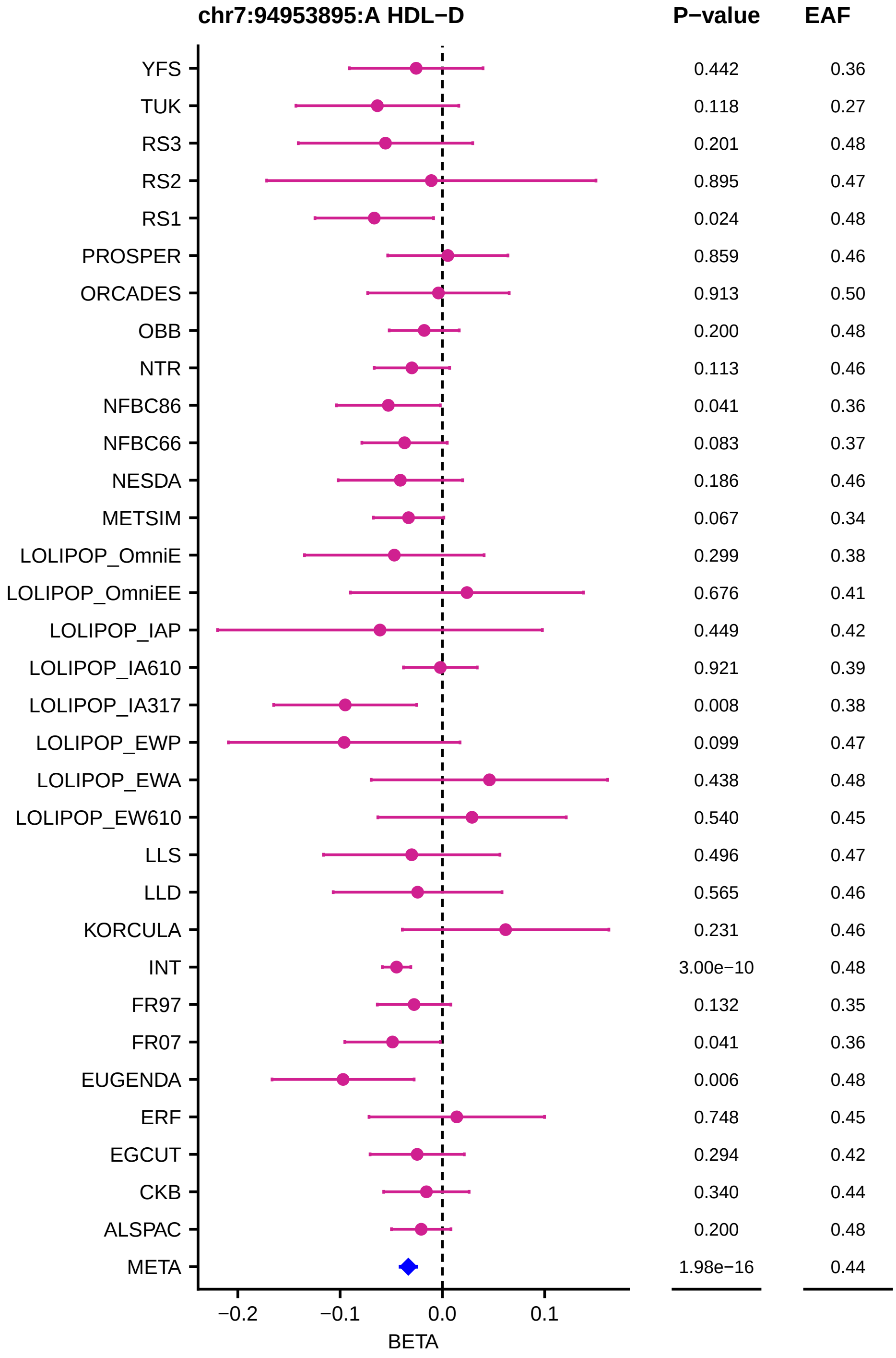
chr7:94953895:A HDL−D
P−value
EAF
YFS 0.442 0.36
TUK 0.118 0.27
RS3 0.201 0.48
RS2 0.895 0.47
RS1 0.024 0.48
PROSPER 0.859 0.46
ORCADES 0.913 0.50
OBB 0.200 0.48
NTR 0.113 0.46
NFBC86 0.041 0.36
NFBC66 0.083 0.37
NESDA 0.186 0.46
METSIM 0.067 0.34
LOLIPOP_OmniE 0.299 0.38
LOLIPOP_OmniEE 0.676 0.41
LOLIPOP_IAP 0.449 0.42
LOLIPOP_IA610 0.921 0.39
LOLIPOP_IA317 0.008 0.38
LOLIPOP_EWP 0.099 0.47
LOLIPOP_EWA 0.438 0.48
LOLIPOP_EW610 0.540 0.45
LLS 0.496 0.47
LLD 0.565 0.46
KORCULA 0.231 0.46
INT 3.00e−10 0.48
FR97 0.132 0.35
FR07 0.041 0.36
EUGENDA 0.006 0.48
ERF 0.748 0.45
EGCUT 0.294 0.42
CKB 0.340 0.44
ALSPAC 0.200 0.48
META 1.98e−16 0.44
−0.2
−0.1
0.0
0.1
BETA

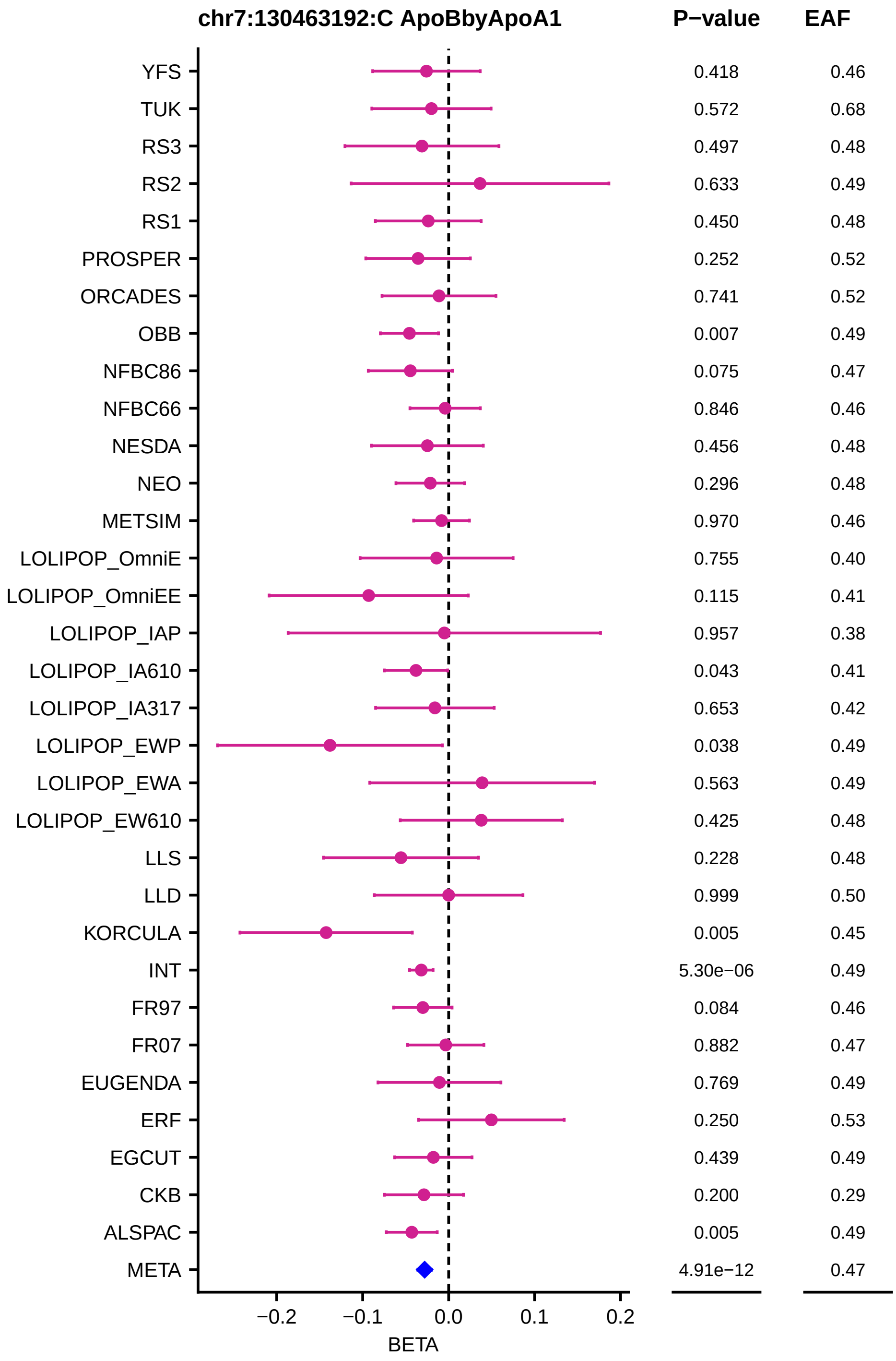
chr7:130463192:C ApoBbyApoA1
P−value
EAF
YFS 0.418 0.46
TUK 0.572 0.68
RS3 0.497 0.48
RS2 0.633 0.49
RS1 0.450 0.48
PROSPER 0.252 0.52
ORCADES 0.741 0.52
OBB 0.007 0.49
NFBC86 0.075 0.47
NFBC66 0.846 0.46
NESDA 0.456 0.48
NEO 0.296 0.48
METSIM 0.970 0.46
LOLIPOP_OmniE 0.755 0.40
LOLIPOP_OmniEE 0.115 0.41
LOLIPOP_IAP 0.957 0.38
LOLIPOP_IA610 0.043 0.41
LOLIPOP_IA317 0.653 0.42
LOLIPOP_EWP 0.038 0.49
LOLIPOP_EWA 0.563 0.49
LOLIPOP_EW610 0.425 0.48
LLS 0.228 0.48
LLD 0.999 0.50
KORCULA 0.005 0.45
INT 5.30e−06 0.49
FR97 0.084 0.46
FR07 0.882 0.47
EUGENDA 0.769 0.49
ERF 0.250 0.53
EGCUT 0.439 0.49
CKB 0.200 0.29
ALSPAC 0.005 0.49
META 4.91e−12 0.47
−0.2
−0.1
0.0
0.1
0.2
BETA

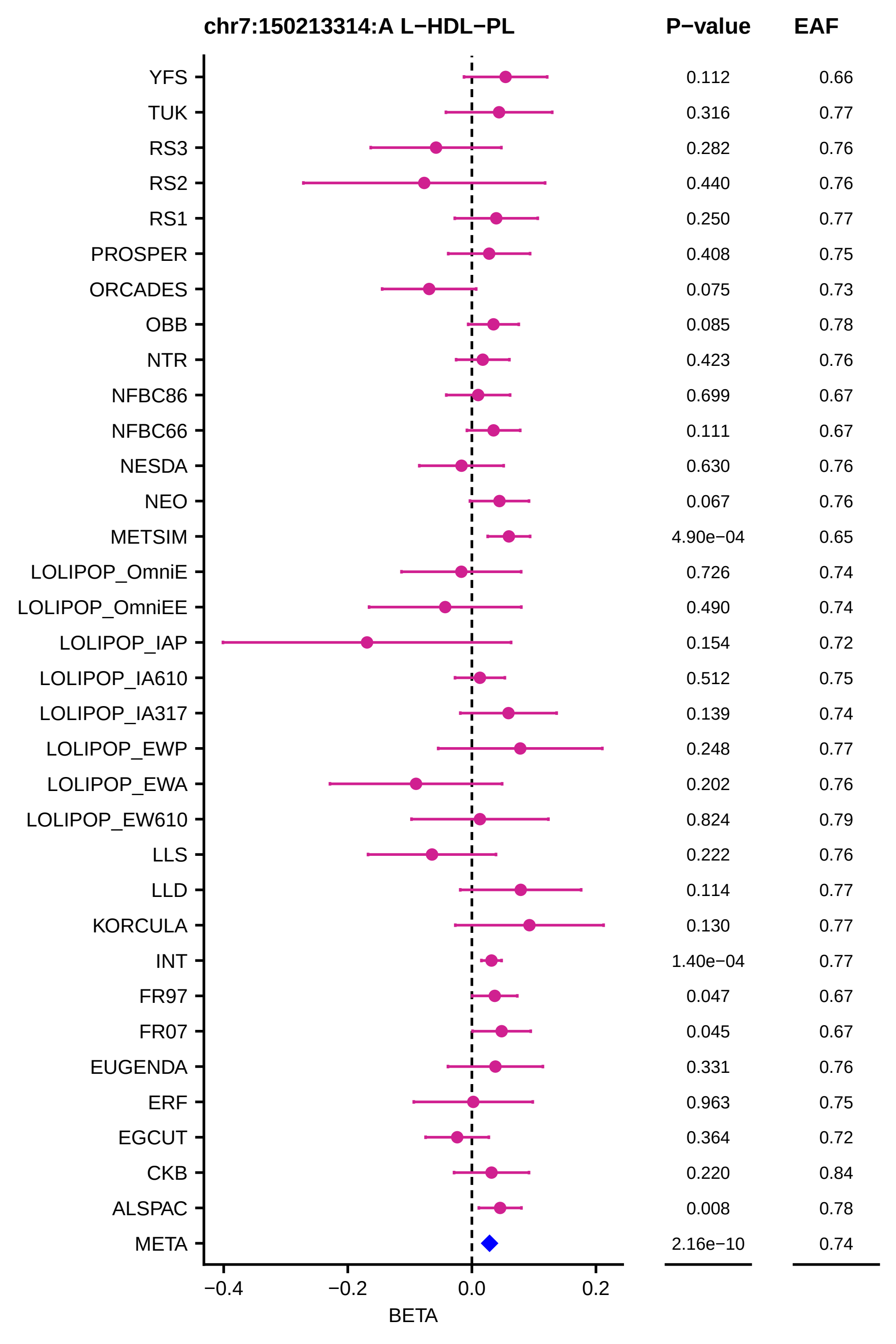
chr7:150213314:A L−HDL−PL
P−value
EAF
YFS
0.112
0.66
TUK
0.316
0.77
RS3
0.282
0.76
RS2
0.440
0.76
RS1
0.250
0.77
PROSPER
0.408
0.75
ORCADES
0.075
0.73
OBB
0.085
0.78
NTR
0.423
0.76
NFBC86
0.699
0.67
NFBC66
0.111
0.67
NESDA
0.630
0.76
NEO
0.067
0.76
METSIM
4.90e−04
0.65
LOLIPOP_OmniE
0.726
0.74
LOLIPOP_OmniEE
0.490
0.74
LOLIPOP_IAP
0.154
0.72
LOLIPOP_IA610
0.512
0.75
LOLIPOP_IA317
0.139
0.74
LOLIPOP_EWP
0.248
0.77
LOLIPOP_EWA
0.202
0.76
LOLIPOP_EW610
0.824
0.79
LLS
0.222
0.76
LLD
0.114
0.77
KORCULA
0.130
0.77
INT
1.40e−04
0.77
FR97
0.047
0.67
FR07
0.045
0.67
EUGENDA
0.331
0.76
ERF
0.963
0.75
EGCUT
0.364
0.72
CKB
0.220
0.84
ALSPAC
0.008
0.78
META
2.16e−10
0.74
−0.4
−0.2
0.0
0.2
BETA

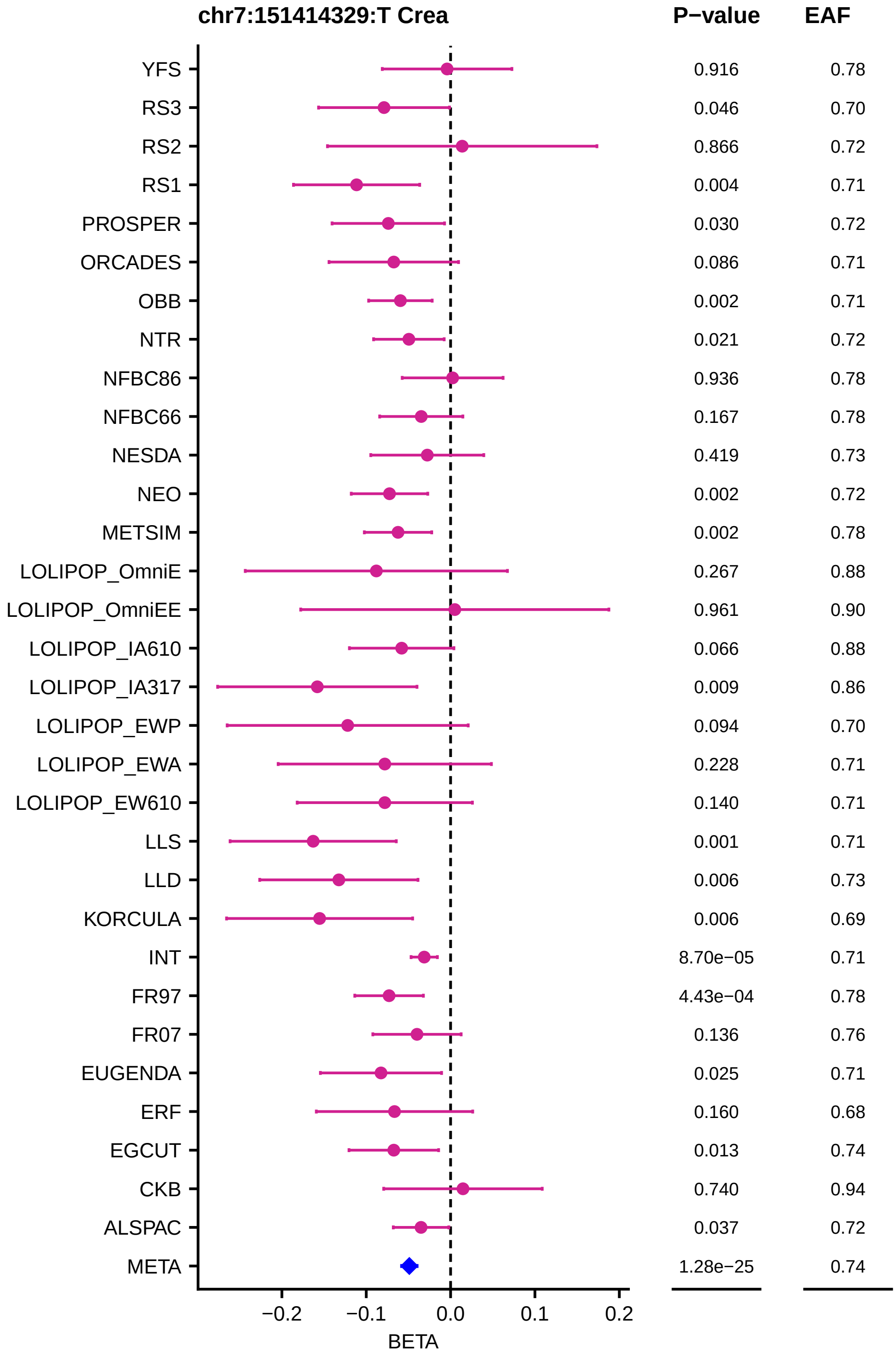

chr7:151414329:T Crea
P−value
EAF
YFS
RS3
RS2
RS1
PROSPER
ORCADES
OBB
NTR
NFBC86
NFBC66
NESDA
NEO
METSIM
LOLIPOP_OmniE
LOLIPOP_OmniEE
LOLIPOP_IA610
LOLIPOP_IA317
LOLIPOP_EWP
LOLIPOP_EWA
LOLIPOP_EW610
LLS
LLD
KORCULA
INT
FR97
FR07
EUGENDA
ERF
EGCUT
CKB
ALSPAC
META
0.916 0.78
0.046 0.70
0.866 0.72
0.004 0.71
0.030 0.72
0.086 0.71
0.002 0.71
0.021 0.72
0.936 0.78
0.167 0.78
0.419 0.73
0.002 0.72
0.002 0.78
0.267 0.88
0.961 0.90
0.066 0.88
0.009 0.86
0.094 0.70
0.228 0.71
0.140 0.71
0.001 0.71
0.006 0.73
0.006 0.69
8.70e−05 0.71
4.43e−04 0.78
0.136 0.76
0.025 0.71
0.160 0.68
0.013 0.74
0.740 0.94
0.037 0.72
1.28e−25 0.74
−0.2
−0.1
0.0
0.1
0.2
BETA

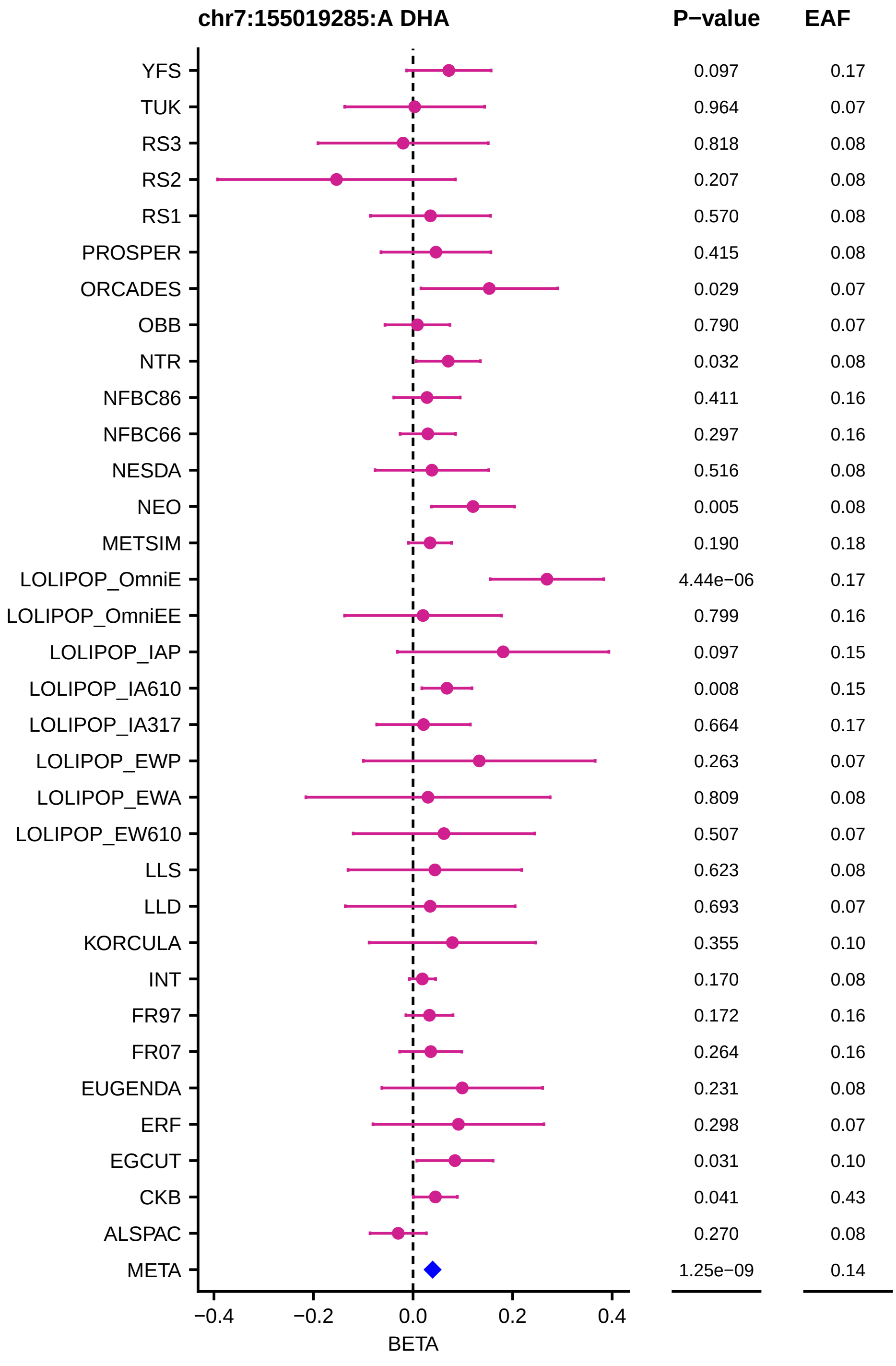
chr7:155019285:A DHA
P−value
EAF
YFS
TUK
RS3
RS2
RS1
PROSPER
ORCADES
OBB
NTR
NFBC86
NFBC66
NESDA
NEO
METSIM
LOLIPOP_OmniE
LOLIPOP_OmniEE
LOLIPOP_IAP
LOLIPOP_IA610
LOLIPOP_IA317
LOLIPOP_EWP
LOLIPOP_EWA
LOLIPOP_EW610
LLS
LLD
KORCULA
INT
FR97
FR07
EUGENDA
ERF
EGCUT
CKB
ALSPAC
META
0.097 0.17
0.964 0.07
0.818 0.08
0.207 0.08
0.570 0.08
0.415 0.08
0.029 0.07
0.790 0.07
0.032 0.08
0.411 0.16
0.297 0.16
0.516 0.08
0.005 0.08
0.190 0.18
4.44e−06 0.17
0.799 0.16
0.097 0.15
0.008 0.15
0.664 0.17
0.263 0.07
0.809 0.08
0.507 0.07
0.623 0.08
0.693 0.07
0.355 0.10
0.170 0.08
0.172 0.16
0.264 0.16
0.231 0.08
0.298 0.07
0.031 0.10
0.041 0.43
0.270 0.08
1.25e−09 0.14
−0.4
−0.2
0.0
0.2
0.4
BETA

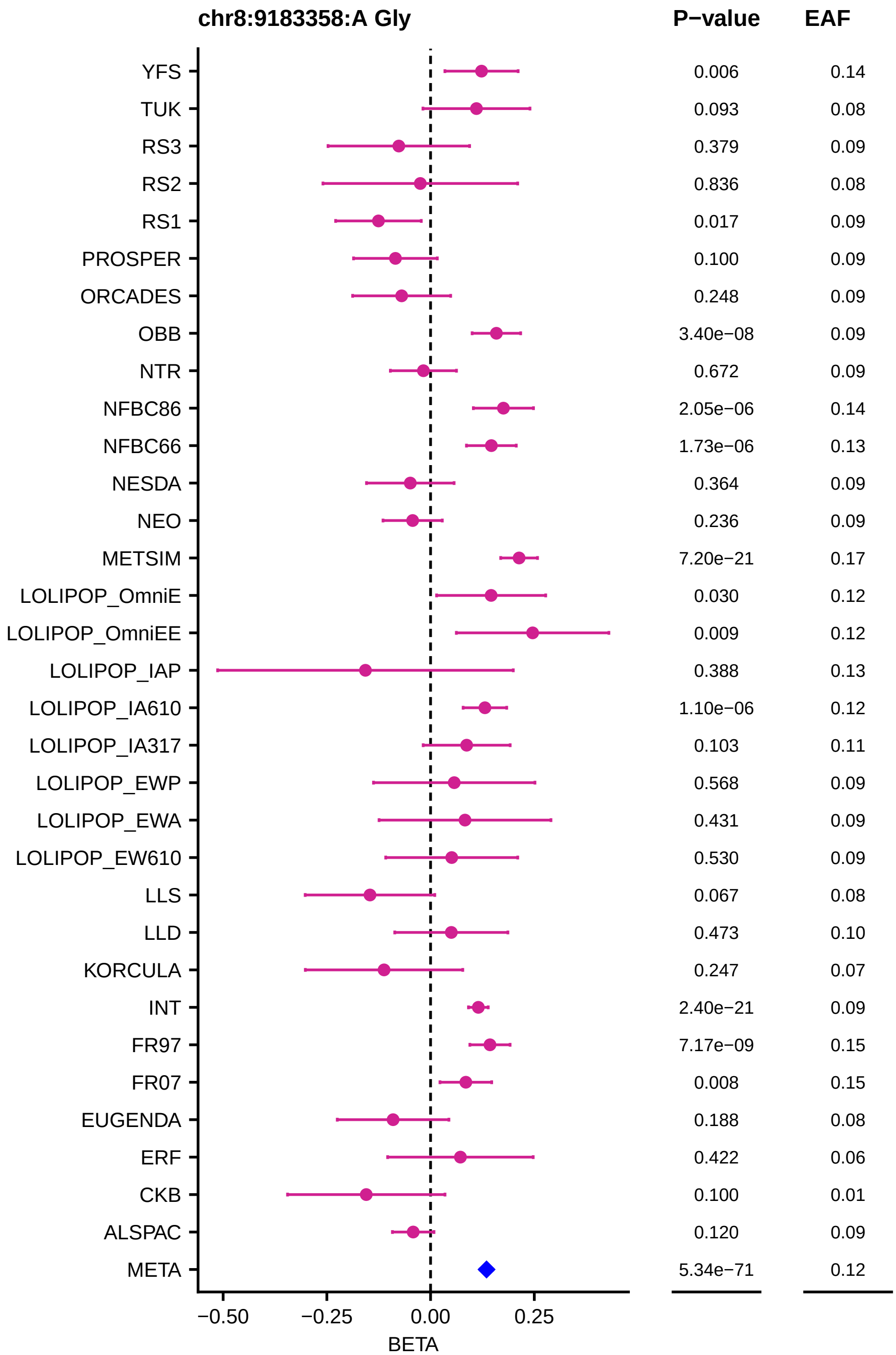
chr8:9183358:A Gly
P−value
EAF
YFS
TUK
RS3
RS2
RS1
PROSPER
ORCADES
OBB
NTR
NFBC86
NFBC66
NESDA
NEO
METSIM
LOLIPOP_OmniE
LOLIPOP_OmniEE
LOLIPOP_IAP
LOLIPOP_IA610
LOLIPOP_IA317
LOLIPOP_EWP
LOLIPOP_EWA
LOLIPOP_EW610
LLS
LLD
KORCULA
INT
FR97
FR07
EUGENDA
ERF
CKB
ALSPAC
META
0.006 0.14
0.093 0.08
0.379 0.09
0.836 0.08
0.017 0.09
0.100 0.09
0.248 0.09
3.40e−08 0.09
0.672 0.09
2.05e−06 0.14
1.73e−06 0.13
0.364 0.09
0.236 0.09
7.20e−21 0.17
0.030 0.12
0.009 0.12
0.388 0.13
1.10e−06 0.12
0.103 0.11
0.568 0.09
0.431 0.09
0.530 0.09
0.067 0.08
0.473 0.10
0.247 0.07
2.40e−21 0.09
7.17e−09 0.15
0.008 0.15
0.188 0.08
0.422 0.06
0.100 0.01
0.120 0.09
5.34e−71 0.12
−0.50
−0.25
0.00
0.25
BETA

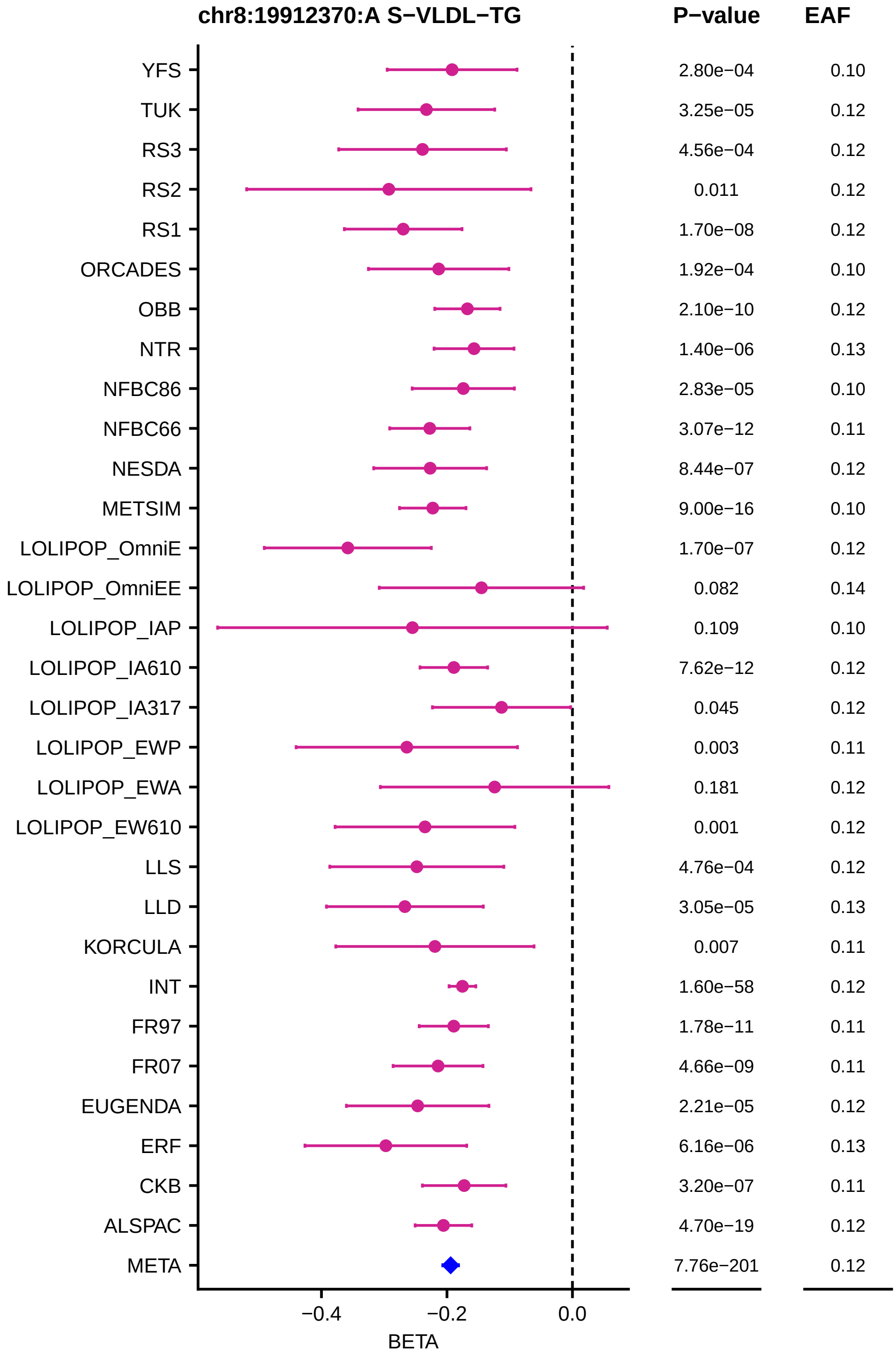
chr8:19912370:A S−VLDL−TG
P−value
EAF
YFS 2.80e−04 0.10
TUK 3.25e−05 0.12
RS3 4.56e−04 0.12
RS2 0.011 0.12
RS1 1.70e−08 0.12
ORCADES 1.92e−04 0.10
OBB 2.10e−10 0.12
NTR 1.40e−06 0.13
NFBC86 2.83e−05 0.10
NFBC66 3.07e−12 0.11
NESDA 8.44e−07 0.12
METSIM 9.00e−16 0.10
LOLIPOP_OmniE 1.70e−07 0.12
LOLIPOP_OmniEE 0.082 0.14
LOLIPOP_IAP 0.109 0.10
LOLIPOP_IA610 7.62e−12 0.12
LOLIPOP_IA317 0.045 0.12
LOLIPOP_EWP 0.003 0.11
LOLIPOP_EWA 0.181 0.12
LOLIPOP_EW610 0.001 0.12
LLS 4.76e−04 0.12
LLD 3.05e−05 0.13
KORCULA 0.007 0.11
INT 1.60e−58 0.12
FR97 1.78e−11 0.11
FR07 4.66e−09 0.11
EUGENDA 2.21e−05 0.12
ERF 6.16e−06 0.13
CKB 3.20e−07 0.11
ALSPAC 4.70e−19 0.12
META 7.76e−201 0.12
−0.4
−0.2
0.0
BETA

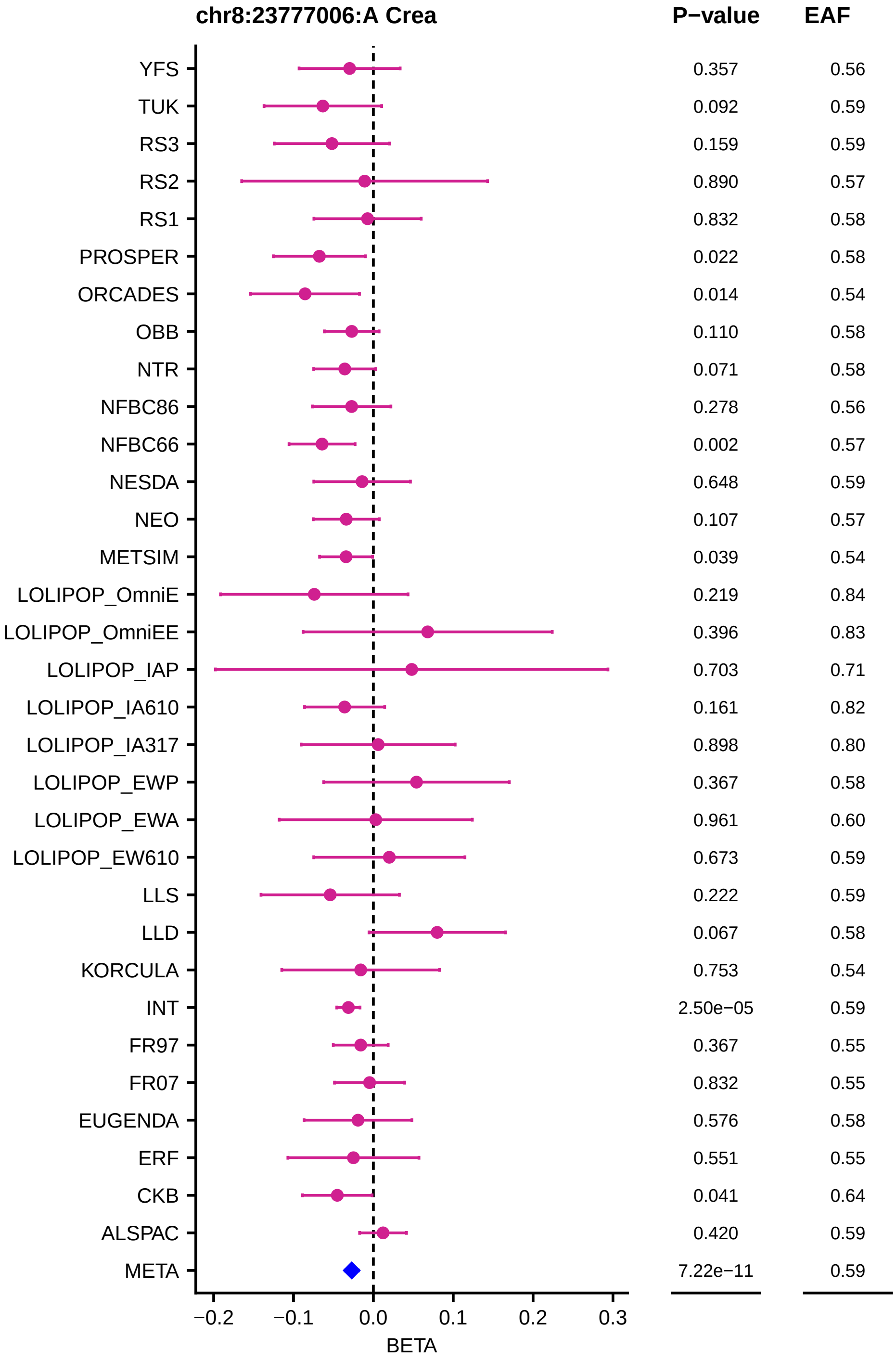

chr8:23777006:A Crea
P−value
EAF
YFS 0.357 0.56
TUK 0.092 0.59
RS3 0.159 0.59
RS2 0.890 0.57
RS1 0.832 0.58
PROSPER 0.022 0.58
ORCADES 0.014 0.54
OBB 0.110 0.58
NTR 0.071 0.58
NFBC86 0.278 0.56
NFBC66 0.002 0.57
NESDA 0.648 0.59
NEO 0.107 0.57
METSIM 0.039 0.54
LOLIPOP_OmniE 0.219 0.84
LOLIPOP_OmniEE 0.396 0.83
LOLIPOP_IAP 0.703 0.71
LOLIPOP_IA610 0.161 0.82
LOLIPOP_IA317 0.898 0.80
LOLIPOP_EWP 0.367 0.58
LOLIPOP_EWA 0.961 0.60
LOLIPOP_EW610 0.673 0.59
LLS 0.222 0.59
LLD 0.067 0.58
KORCULA 0.753 0.54
INT 2.50e−05 0.59
FR97 0.367 0.55
FR07 0.832 0.55
EUGENDA 0.576 0.58
ERF 0.551 0.55
CKB 0.041 0.64
ALSPAC 0.420 0.59
META 7.22e−11 0.59
−0.2 −0.1 0.0 0.1 0.2 0.3
BETA

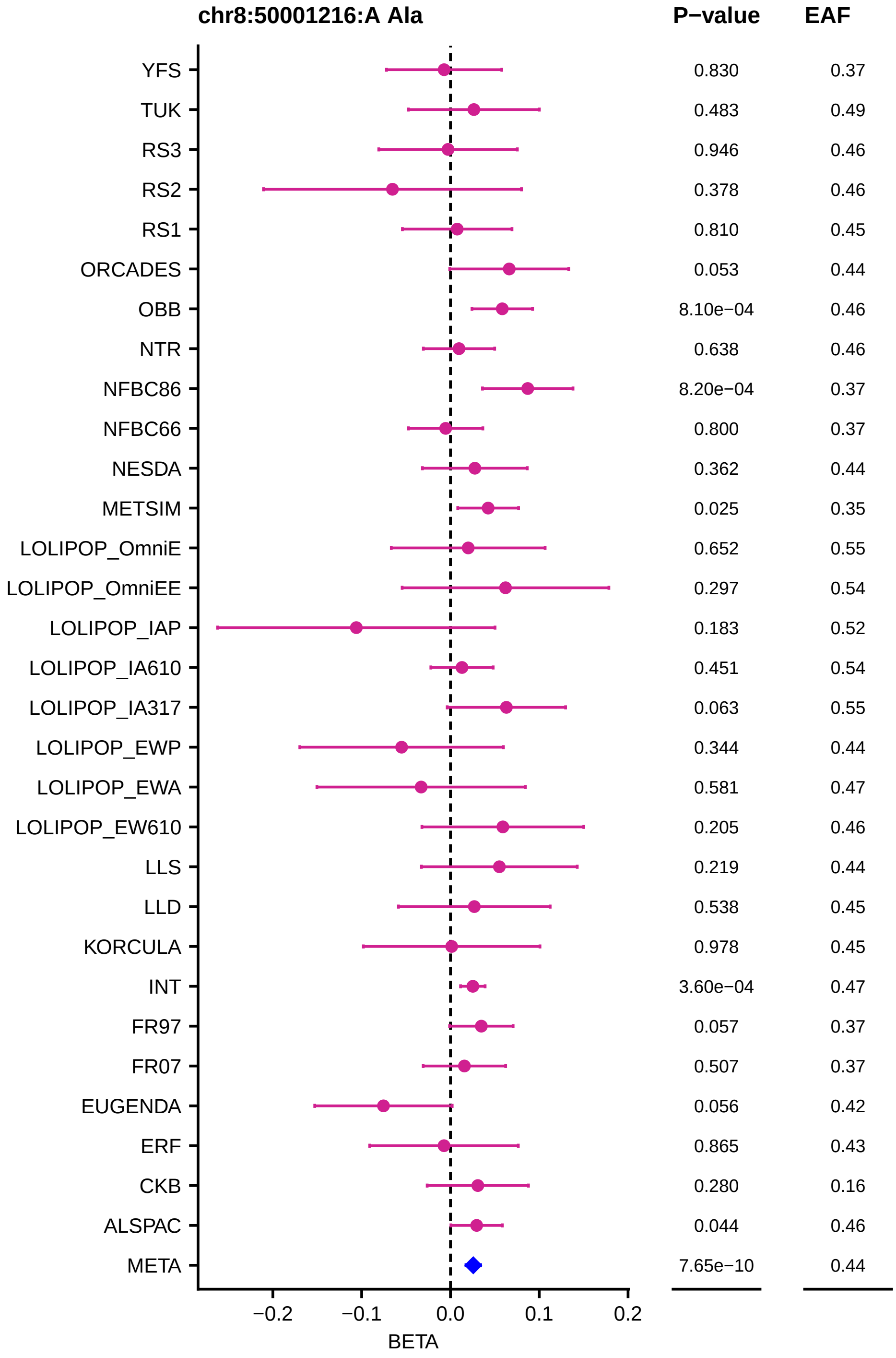

chr8:50001216:A Ala
P−value
EAF
YFS 0.830 0.37
TUK 0.483 0.49
RS3 0.946 0.46
RS2 0.378 0.46
RS1 0.810 0.45
ORCADES 0.053 0.44
OBB 8.10e−04 0.46
NTR 0.638 0.46
NFBC86 8.20e−04 0.37
NFBC66 0.800 0.37
NESDA 0.362 0.44
METSIM 0.025 0.35
LOLIPOP_OmniE 0.652 0.55
LOLIPOP_OmniEE 0.297 0.54
LOLIPOP_IAP 0.183 0.52
LOLIPOP_IA610 0.451 0.54
LOLIPOP_IA317 0.063 0.55
LOLIPOP_EWP 0.344 0.44
LOLIPOP_EWA 0.581 0.47
LOLIPOP_EW610 0.205 0.46
LLS 0.219 0.44
LLD 0.538 0.45
KORCULA 0.978 0.45
INT 3.60e−04 0.47
FR97 0.057 0.37
FR07 0.507 0.37
EUGENDA 0.056 0.42
ERF 0.865 0.43
CKB 0.280 0.16
ALSPAC 0.044 0.46
META 7.65e−10 0.44
−0.2
−0.1
0.0
0.1
0.2
BETA

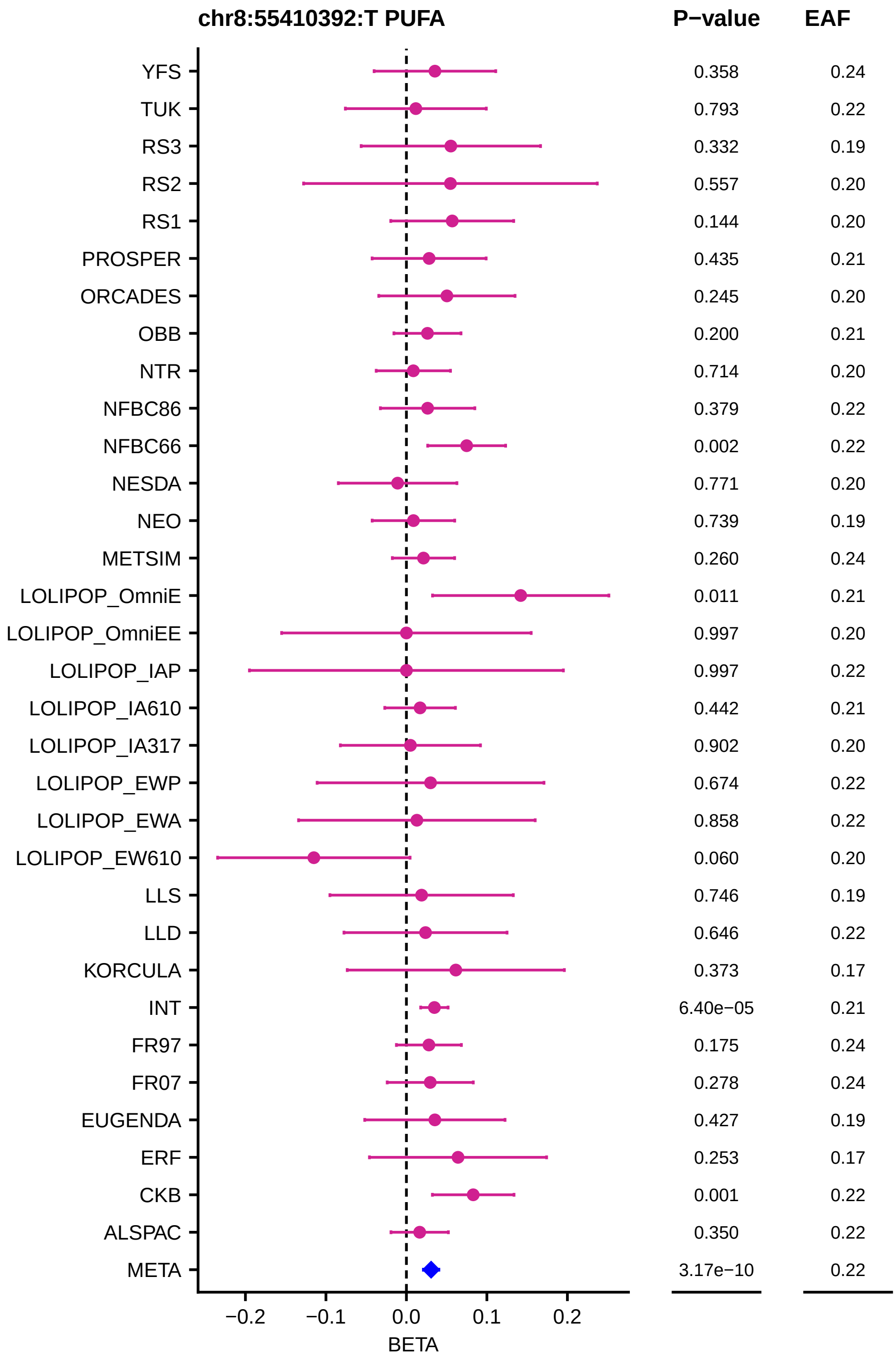
chr8:55410392:T PUFA
P−value
EAF
YFS
TUK
RS3
RS2
RS1
PROSPER
ORCADES
OBB
NTR
NFBC86
NFBC66
NESDA
NEO
METSIM
LOLIPOP_OmniE
LOLIPOP_OmniEE
LOLIPOP_IAP
LOLIPOP_IA610
LOLIPOP_IA317
LOLIPOP_EWP
LOLIPOP_EWA
LOLIPOP_EW610
LLS
LLD
KORCULA
INT
FR97
FR07
EUGENDA
ERF
CKB
ALSPAC
META
0.358 0.24
0.793 0.22
0.332 0.19
0.557 0.20
0.144 0.20
0.435 0.21
0.245 0.20
0.200 0.21
0.714 0.20
0.379 0.22
0.002 0.22
0.771 0.20
0.739 0.19
0.260 0.24
0.011 0.21
0.997 0.20
0.997 0.22
0.442 0.21
0.902 0.20
0.674 0.22
0.858 0.22
0.060 0.20
0.746 0.19
0.646 0.22
0.373 0.17
6.40e−05 0.21
0.175 0.24
0.278 0.24
0.427 0.19
0.253 0.17
0.001 0.22
0.350 0.22
3.17e−10 0.22
−0.2
−0.1
0.0
0.1
0.2
BETA

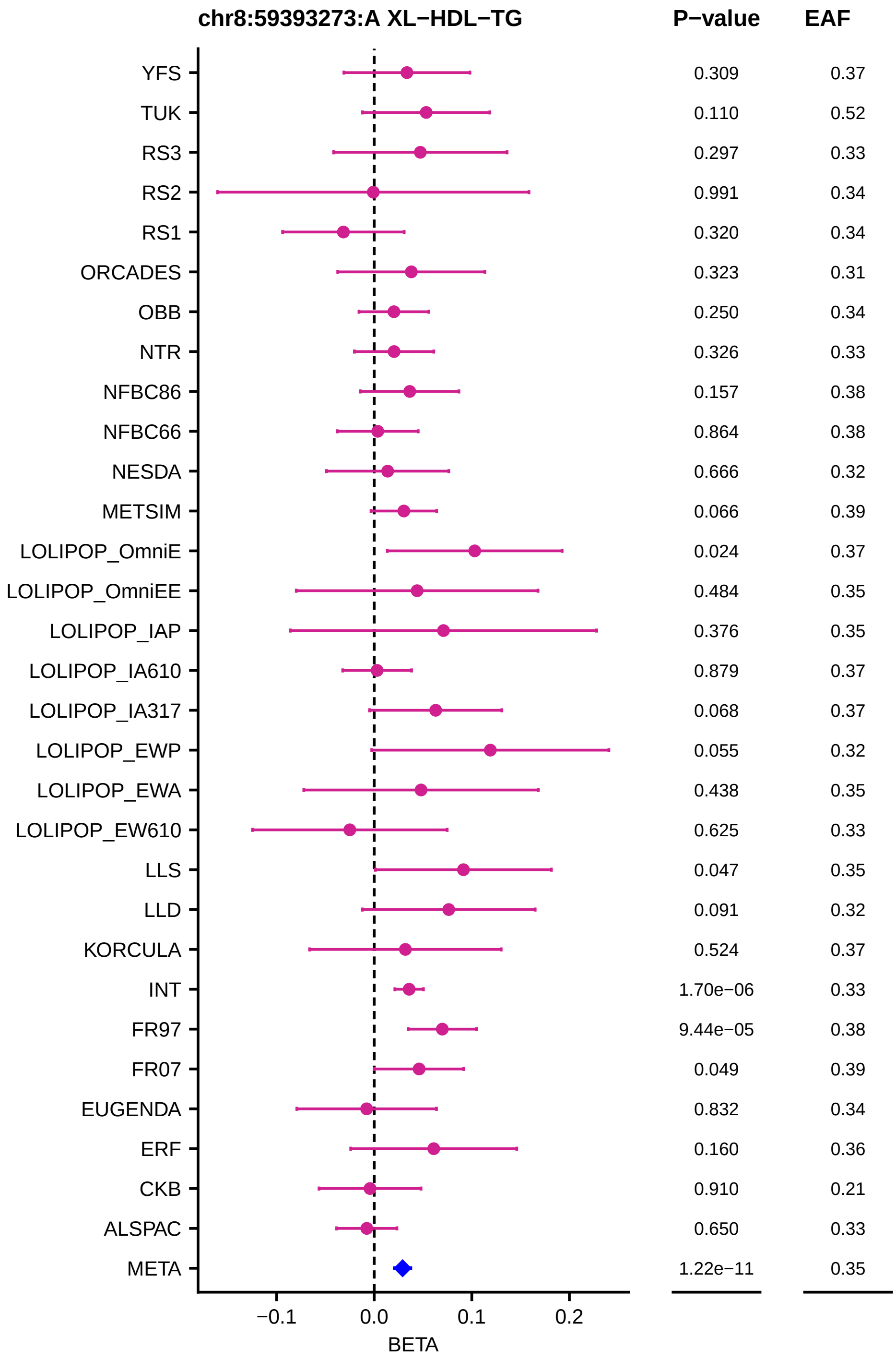
chr8:59393273:A XL−HDL−TG
P−value
EAF
YFS 0.309 0.37
TUK 0.110 0.52
RS3 0.297 0.33
RS2 0.991 0.34
RS1 0.320 0.34
ORCADES 0.323 0.31
OBB 0.250 0.34
NTR 0.326 0.33
NFBC86 0.157 0.38
NFBC66 0.864 0.38
NESDA 0.666 0.32
METSIM 0.066 0.39
LOLIPOP_OmniE 0.024 0.37
LOLIPOP_OmniEE 0.484 0.35
LOLIPOP_IAP 0.376 0.35
LOLIPOP_IA610 0.879 0.37
LOLIPOP_IA317 0.068 0.37
LOLIPOP_EWP 0.055 0.32
LOLIPOP_EWA 0.438 0.35
LOLIPOP_EW610 0.625 0.33
LLS 0.047 0.35
LLD 0.091 0.32
KORCULA 0.524 0.37
INT 1.70e−06 0.33
FR97 9.44e−05 0.38
FR07 0.049 0.39
EUGENDA 0.832 0.34
ERF 0.160 0.36
CKB 0.910 0.21
ALSPAC 0.650 0.33
META 1.22e−11 0.35
−0.1
0.0
0.1
0.2
BETA

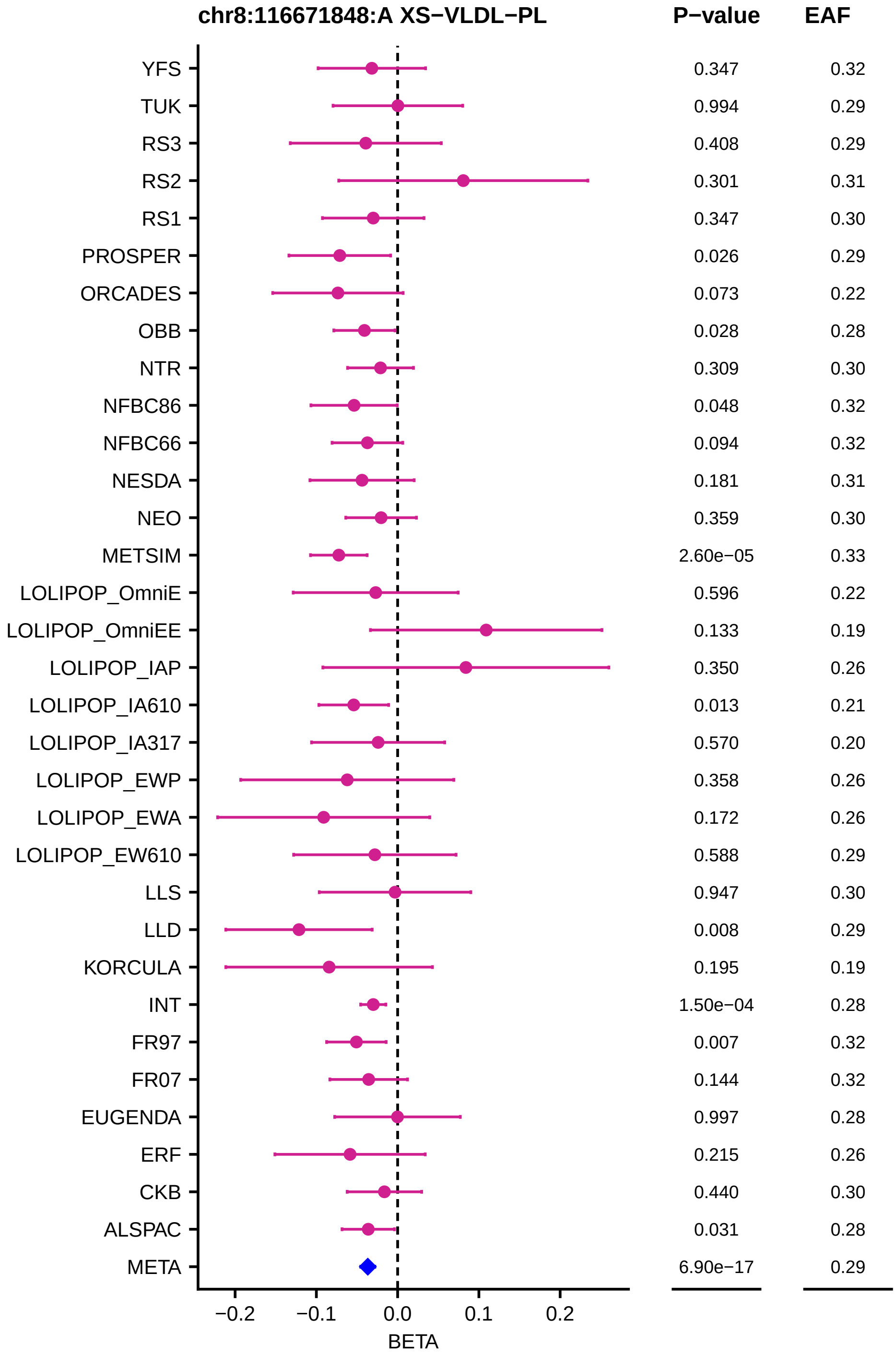
chr8:116671848:A XS−VLDL−PL
P−value
EAF
YFS 0.347 0.32
TUK 0.994 0.29
RS3 0.408 0.29
RS2 0.301 0.31
RS1 0.347 0.30
PROSPER 0.026 0.29
ORCADES 0.073 0.22
OBB 0.028 0.28
NTR 0.309 0.30
NFBC86 0.048 0.32
NFBC66 0.094 0.32
NESDA 0.181 0.31
NEO 0.359 0.30
METSIM 2.60e−05 0.33
LOLIPOP_OmniE 0.596 0.22
LOLIPOP_OmniEE 0.133 0.19
LOLIPOP_IAP 0.350 0.26
LOLIPOP_IA610 0.013 0.21
LOLIPOP_IA317 0.570 0.20
LOLIPOP_EWP 0.358 0.26
LOLIPOP_EWA 0.172 0.26
LOLIPOP_EW610 0.588 0.29
LLS 0.947 0.30
LLD 0.008 0.29
KORCULA 0.195 0.19
INT 1.50e−04 0.28
FR97 0.007 0.32
FR07 0.144 0.32
EUGENDA 0.997 0.28
ERF 0.215 0.26
CKB 0.440 0.30
ALSPAC 0.031 0.28
META 6.90e−17 0.29
−0.2
−0.1
0.0
0.1
0.2
BETA

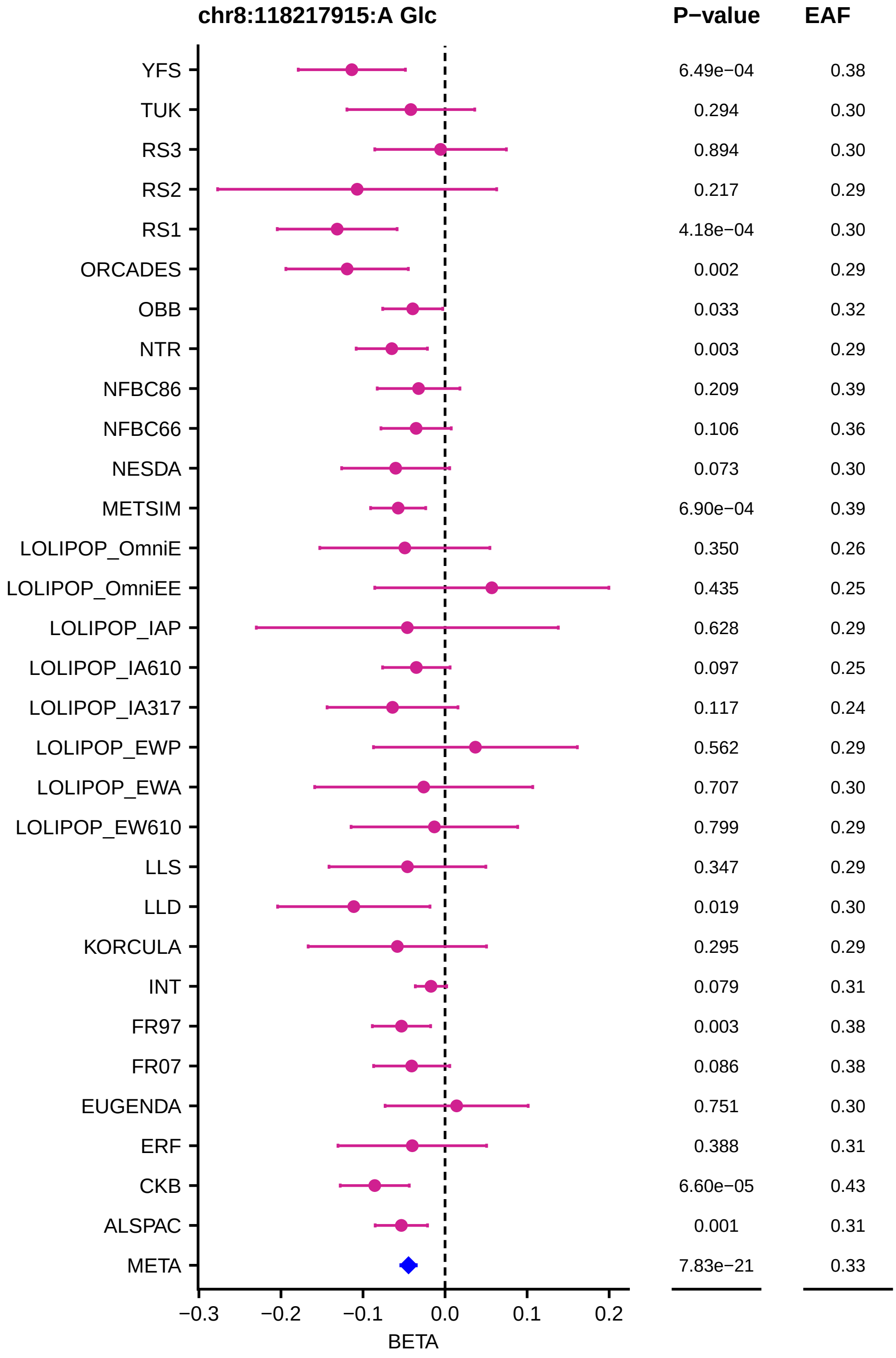

chr8:118217915:A Glc
P−value
EAF
YFS
TUK
RS3
RS2
RS1
ORCADES
OBB
NTR
NFBC86
NFBC66
NESDA
METSIM
LOLIPOP_OmniE
LOLIPOP_OmniEE
LOLIPOP_IAP
LOLIPOP_IA610
LOLIPOP_IA317
LOLIPOP_EWP
LOLIPOP_EWA
LOLIPOP_EW610
LLS
LLD
KORCULA
INT
FR97
FR07
EUGENDA
ERF
CKB
ALSPAC
META
6.49e−04 0.38
0.294 0.30
0.894 0.30
0.217 0.29
4.18e−04 0.30
0.002 0.29
0.033 0.32
0.003 0.29
0.209 0.39
0.106 0.36
0.073 0.30
6.90e−04 0.39
0.350 0.26
0.435 0.25
0.628 0.29
0.097 0.25
0.117 0.24
0.562 0.29
0.707 0.30
0.799 0.29
0.347 0.29
0.019 0.30
0.295 0.29
0.079 0.31
0.003 0.38
0.086 0.38
0.751 0.30
0.388 0.31
6.60e−05 0.43
0.001 0.31
7.83e−21 0.33
−0.3
−0.2
−0.1
0.0
0.1
0.2
BETA

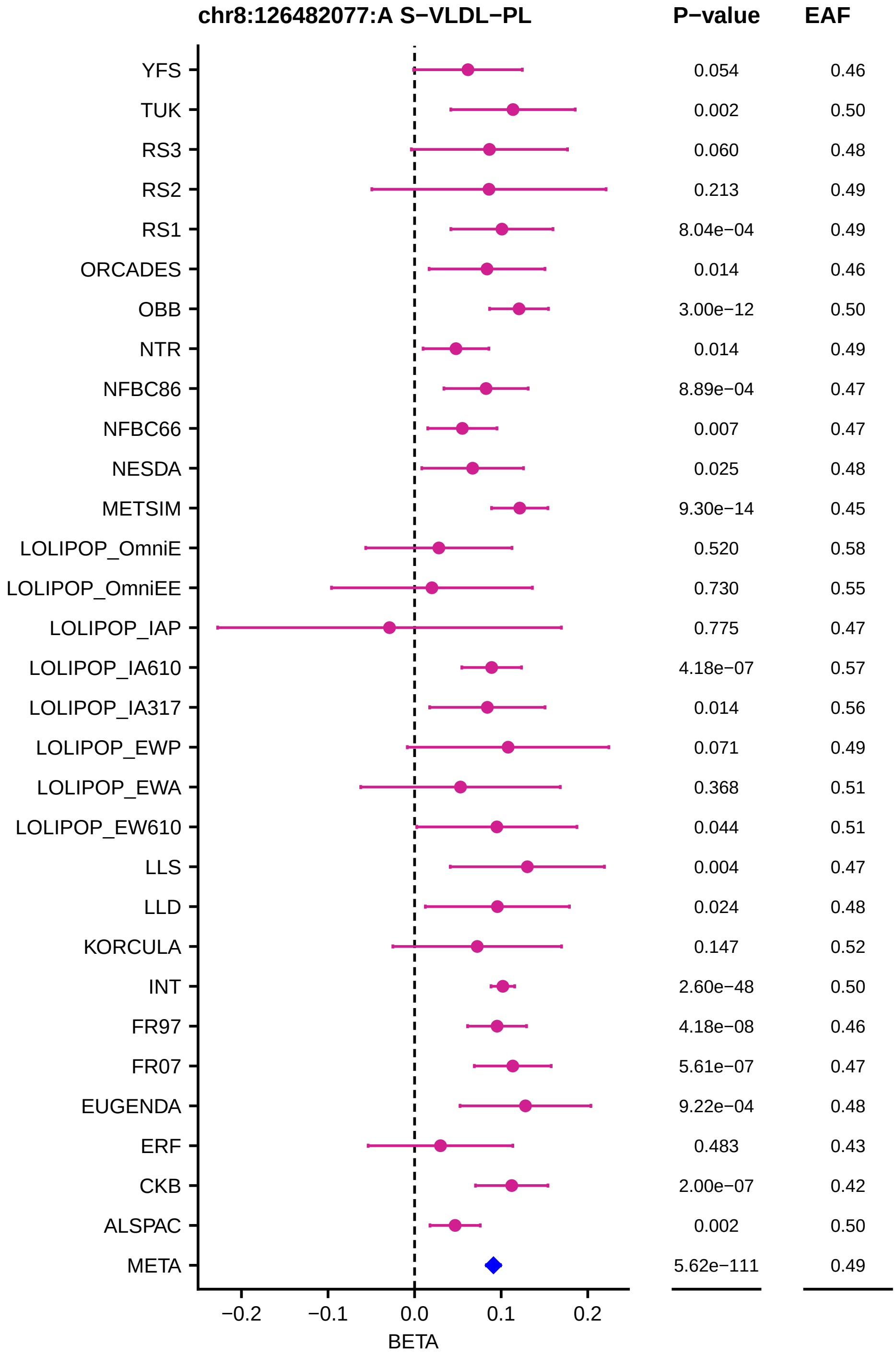
chr8:126482077:A S−VLDL−PL
P−value
EAF
YFS 0.054 0.46
TUK 0.002 0.50
RS3 0.060 0.48
RS2 0.213 0.49
RS1 8.04e−04 0.49
ORCADES 0.014 0.46
OBB 3.00e−12 0.50
NTR 0.014 0.49
NFBC86 8.89e−04 0.47
NFBC66 0.007 0.47
NESDA 0.025 0.48
METSIM 9.30e−14 0.45
LOLIPOP_OmniE 0.520 0.58
LOLIPOP_OmniEE 0.730 0.55
LOLIPOP_IAP 0.775 0.47
LOLIPOP_IA610 4.18e−07 0.57
LOLIPOP_IA317 0.014 0.56
LOLIPOP_EWP 0.071 0.49
LOLIPOP_EWA 0.368 0.51
LOLIPOP_EW610 0.044 0.51
LLS 0.004 0.47
LLD 0.024 0.48
KORCULA 0.147 0.52
INT 2.60e−48 0.50
FR97 4.18e−08 0.46
FR07 5.61e−07 0.47
EUGENDA 9.22e−04 0.48
ERF 0.483 0.43
CKB 2.00e−07 0.42
ALSPAC 0.002 0.50
META 5.62e−111 0.49
−0.2
−0.1
0.0
0.1
0.2
BETA

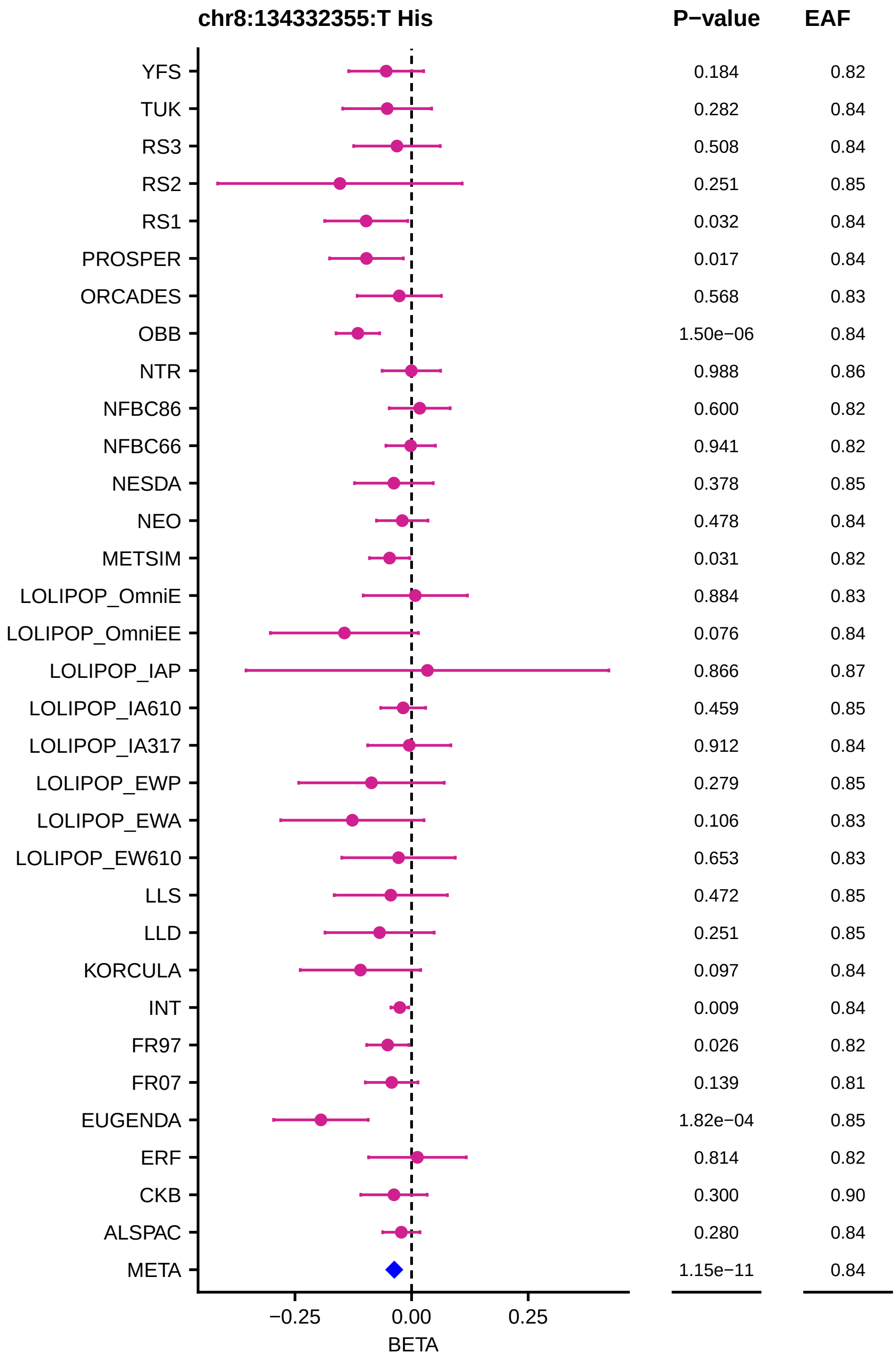
chr8:134332355:T His
P−value
EAF
YFS 0.184 0.82
TUK 0.282 0.84
RS3 0.508 0.84
RS2 0.251 0.85
RS1 0.032 0.84
PROSPER 0.017 0.84
ORCADES 0.568 0.83
OBB 1.50e−06 0.84
NTR 0.988 0.86
NFBC86 0.600 0.82
NFBC66 0.941 0.82
NESDA 0.378 0.85
NEO 0.478 0.84
METSIM 0.031 0.82
LOLIPOP_OmniE 0.884 0.83
LOLIPOP_OmniEE 0.076 0.84
LOLIPOP_IAP 0.866 0.87
LOLIPOP_IA610 0.459 0.85
LOLIPOP_IA317 0.912 0.84
LOLIPOP_EWP 0.279 0.85
LOLIPOP_EWA 0.106 0.83
LOLIPOP_EW610 0.653 0.83
LLS 0.472 0.85
LLD 0.251 0.85
KORCULA 0.097 0.84
INT 0.009 0.84
FR97 0.026 0.82
FR07 0.139 0.81
EUGENDA 1.82e−04 0.85
ERF 0.814 0.82
CKB 0.300 0.90
ALSPAC 0.280 0.84
META 1.15e−11 0.84
−0.25
0.00
0.25
BETA

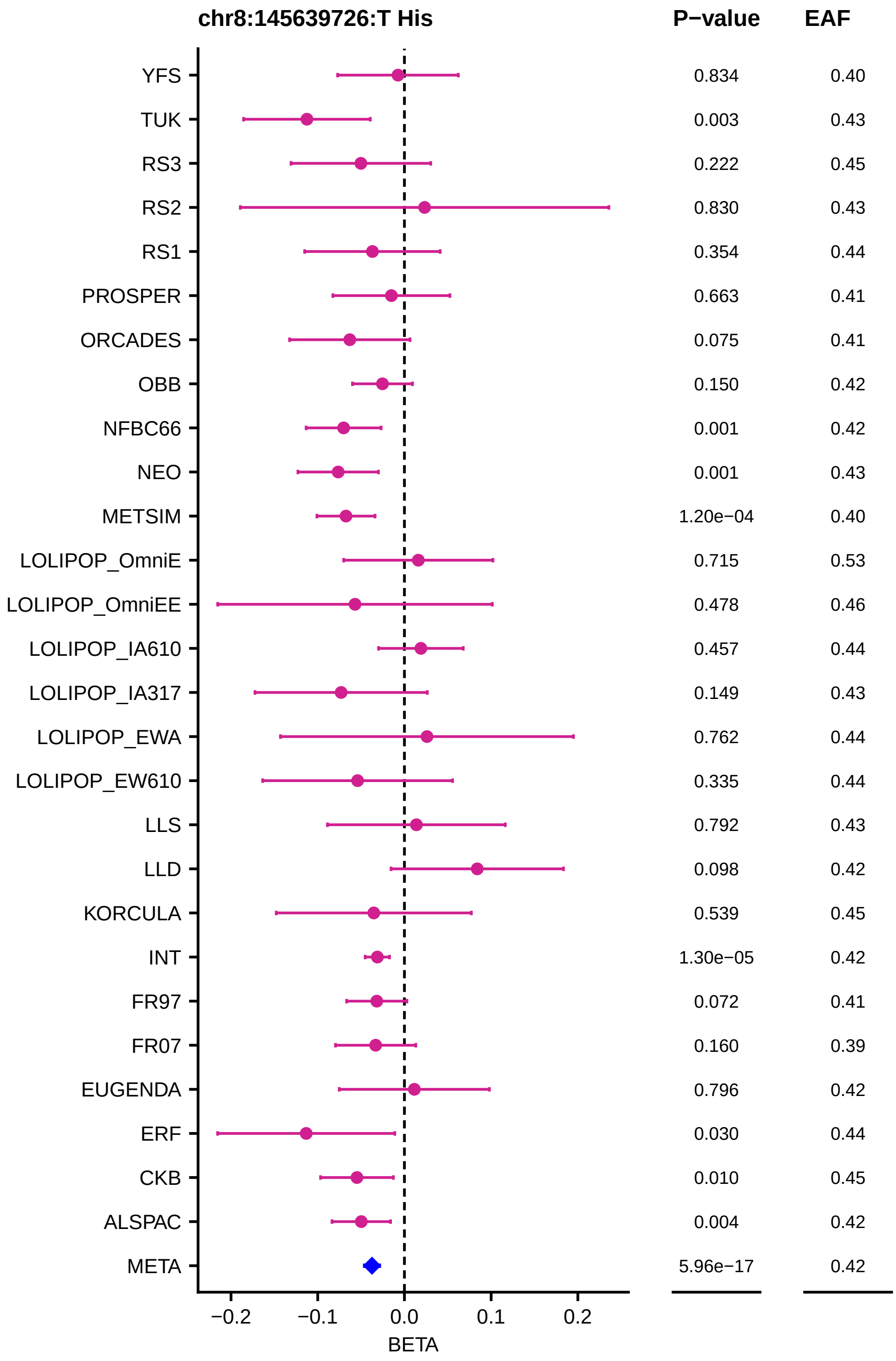
chr8:145639726:T His
P−value
EAF
YFS
TUK
RS3
RS2
RS1
PROSPER
ORCADES
OBB
NFBC66
NEO
METSIM
LOLIPOP_OmniE
LOLIPOP_OmniEE
LOLIPOP_IA610
LOLIPOP_IA317
LOLIPOP_EWA
LOLIPOP_EW610
LLS
LLD
KORCULA
INT
FR97
FR07
EUGENDA
ERF
CKB
ALSPAC
META
0.834 0.40
0.003 0.43
0.222 0.45
0.830 0.43
0.354 0.44
0.663 0.41
0.075 0.41
0.150 0.42
0.001 0.42
0.001 0.43
1.20e−04 0.40
0.715 0.53
0.478 0.46
0.457 0.44
0.149 0.43
0.762 0.44
0.335 0.44
0.792 0.43
0.098 0.42
0.539 0.45
1.30e−05 0.42
0.072 0.41
0.160 0.39
0.796 0.42
0.030 0.44
0.010 0.45
0.004 0.42
5.96e−17 0.42
−0.2
−0.1
0.0
0.1
0.2
BETA

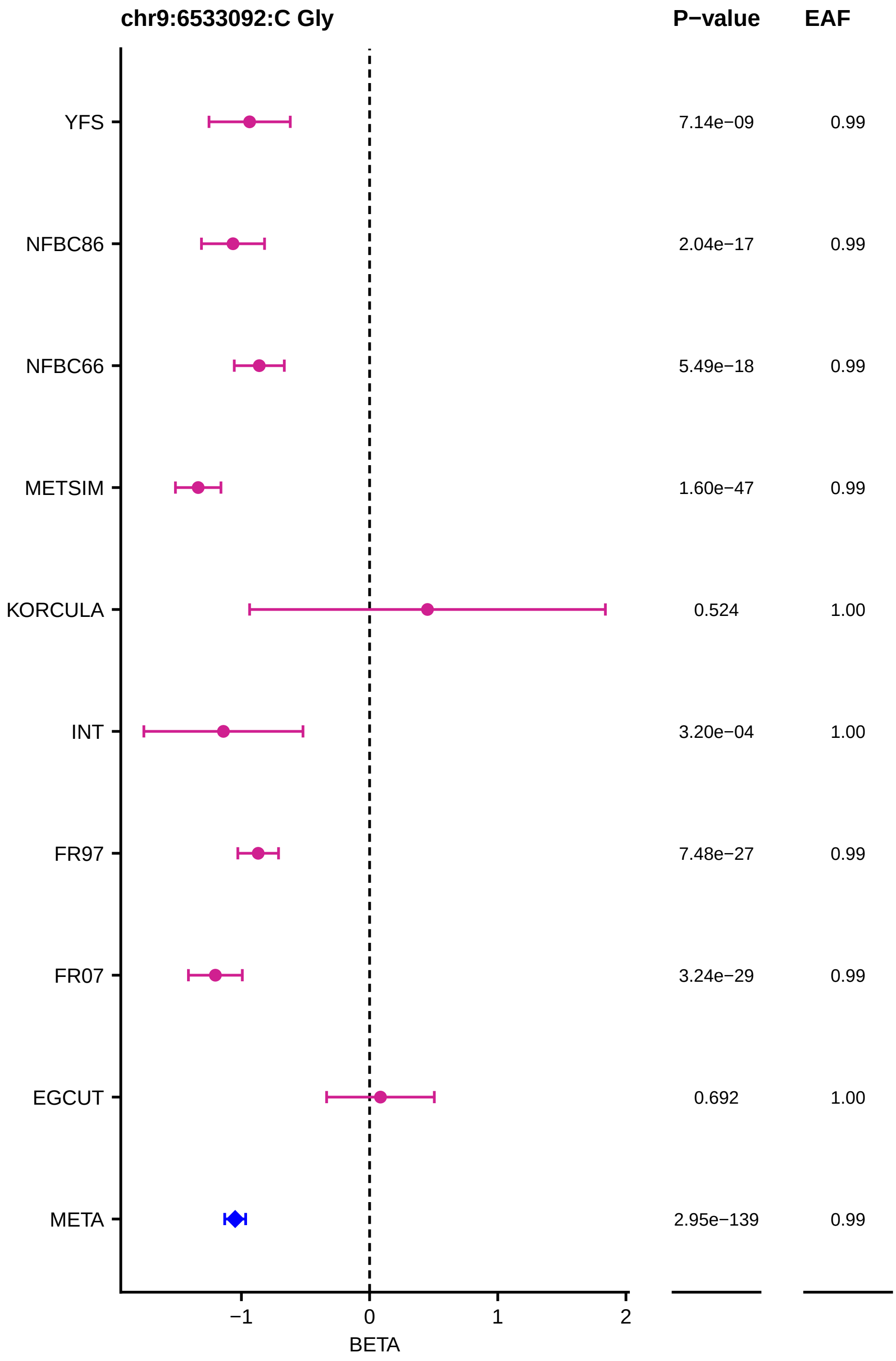
chr9:6533092:C Gly
P−value
EAF
YFS
7.14e−09
0.99
NFBC86
2.04e−17
0.99
NFBC66
5.49e−18
0.99
METSIM
1.60e−47
0.99
KORCULA
0.524
1.00
INT
3.20e−04
1.00
FR97
7.48e−27
0.99
FR07
3.24e−29
0.99
EGCUT
0.692
1.00
META
2.95e−139
0.99
−1
0
1
2
BETA

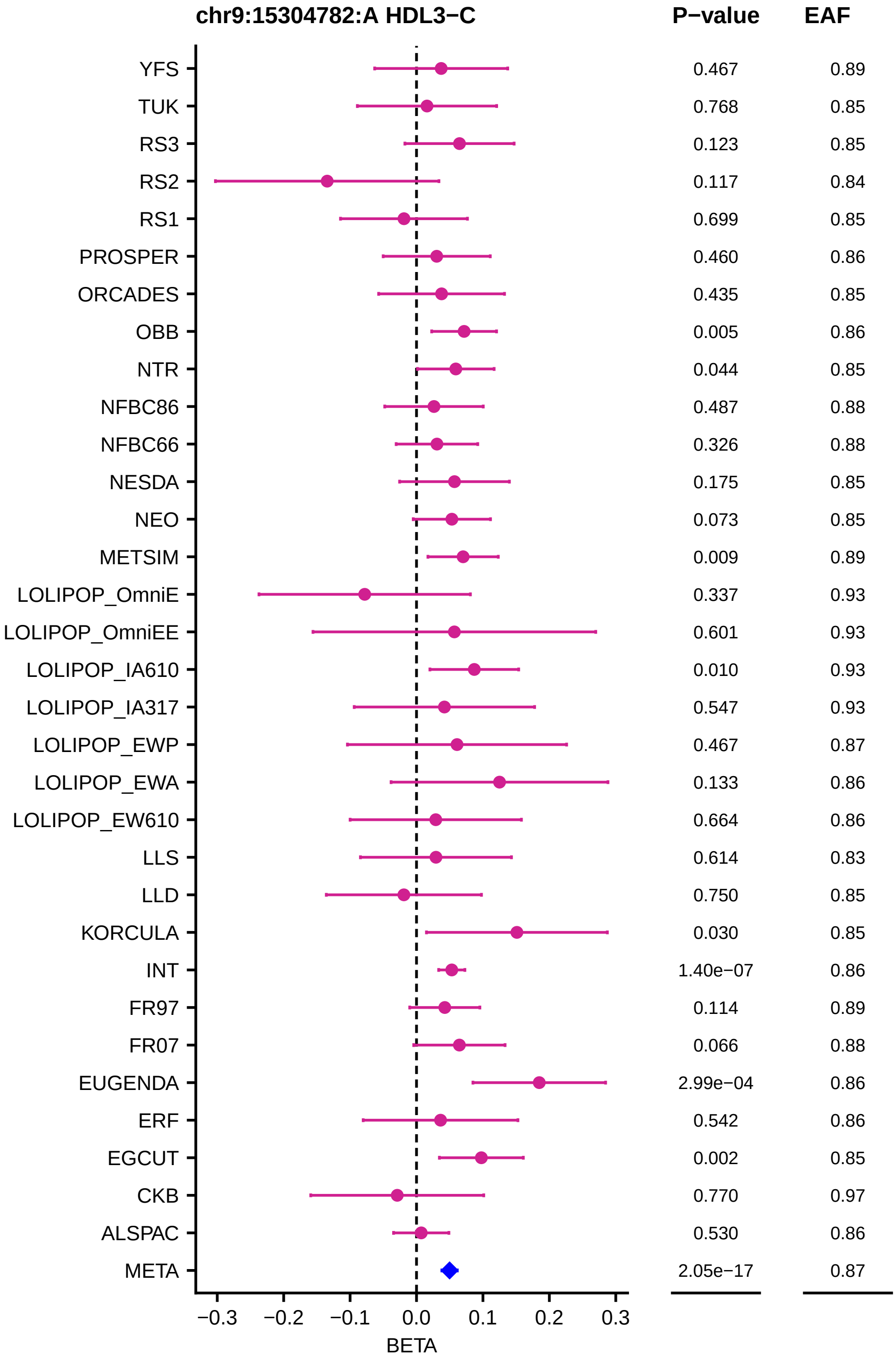
chr9:15304782:A HDL3−C
P−value
EAF
YFS 0.467 0.89
TUK 0.768 0.85
RS3 0.123 0.85
RS2 0.117 0.84
RS1 0.699 0.85
PROSPER 0.460 0.86
ORCADES 0.435 0.85
OBB 0.005 0.86
NTR 0.044 0.85
NFBC86 0.487 0.88
NFBC66 0.326 0.88
NESDA 0.175 0.85
NEO 0.073 0.85
METSIM 0.009 0.89
LOLIPOP_OmniE 0.337 0.93
LOLIPOP_OmniEE 0.601 0.93
LOLIPOP_IA610 0.010 0.93
LOLIPOP_IA317 0.547 0.93
LOLIPOP_EWP 0.467 0.87
LOLIPOP_EWA 0.133 0.86
LOLIPOP_EW610 0.664 0.86
LLS 0.614 0.83
LLD 0.750 0.85
KORCULA 0.030 0.85
INT 1.40e−07 0.86
FR97 0.114 0.89
FR07 0.066 0.88
EUGENDA 2.99e−04 0.86
ERF 0.542 0.86
EGCUT 0.002 0.85
CKB 0.770 0.97
ALSPAC 0.530 0.86
META 2.05e−17 0.87
−0.3 −0.2 −0.1 0.0 0.1 0.2 0.3
BETA

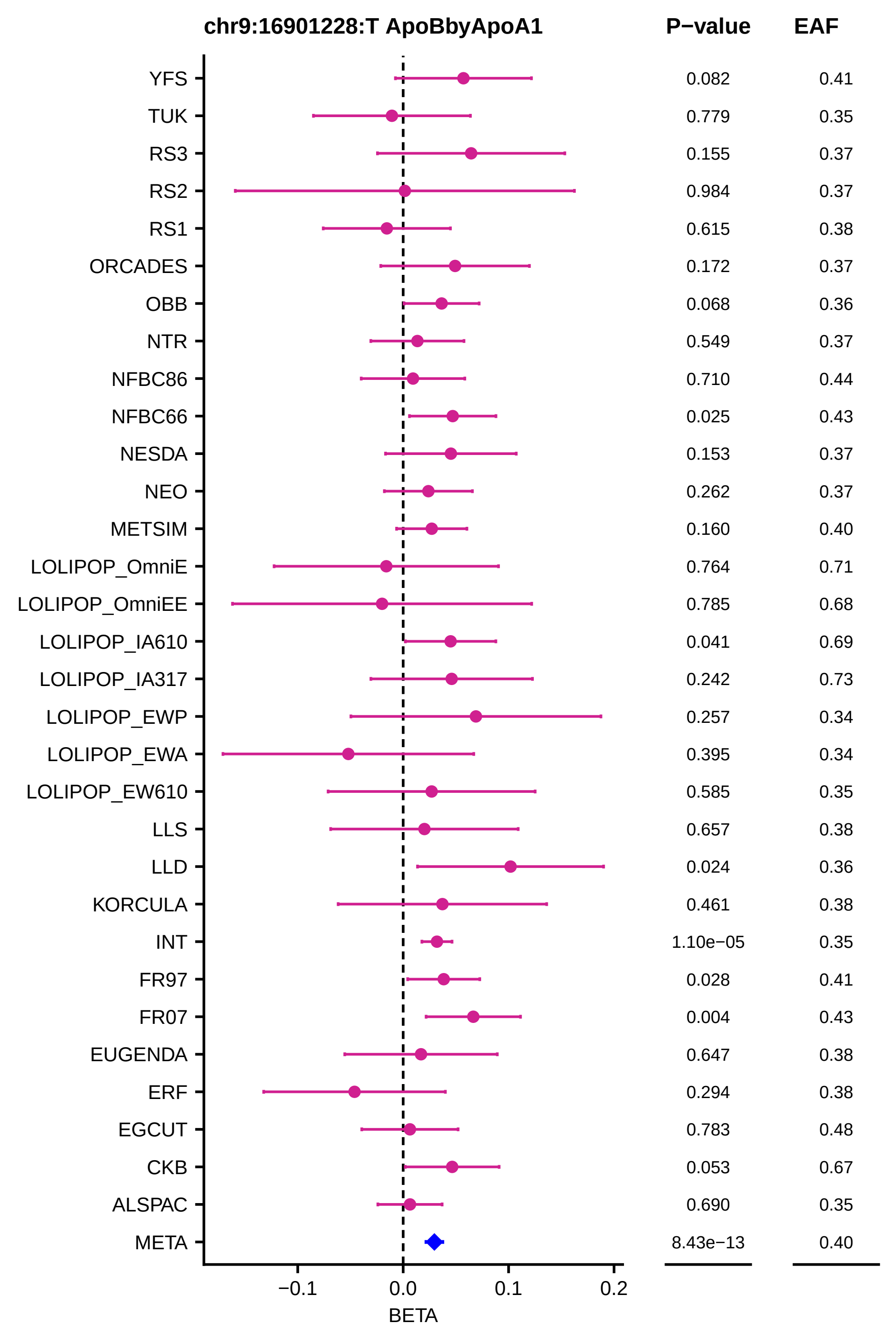
chr9:16901228:T ApoBbyApoA1
P−value
EAF
YFS 0.082 0.41
TUK 0.779 0.35
RS3 0.155 0.37
RS2 0.984 0.37
RS1 0.615 0.38
ORCADES 0.172 0.37
OBB 0.068 0.36
NTR 0.549 0.37
NFBC86 0.710 0.44
NFBC66 0.025 0.43
NESDA 0.153 0.37
NEO 0.262 0.37
METSIM 0.160 0.40
LOLIPOP_OmniE 0.764 0.71
LOLIPOP_OmniEE 0.785 0.68
LOLIPOP_IA610 0.041 0.69
LOLIPOP_IA317 0.242 0.73
LOLIPOP_EWP 0.257 0.34
LOLIPOP_EWA 0.395 0.34
LOLIPOP_EW610 0.585 0.35
LLS 0.657 0.38
LLD 0.024 0.36
KORCULA 0.461 0.38
INT 1.10e−05 0.35
FR97 0.028 0.41
FR07 0.004 0.43
EUGENDA 0.647 0.38
ERF 0.294 0.38
EGCUT 0.783 0.48
CKB 0.053 0.67
ALSPAC 0.690 0.35
META 8.43e−13 0.40
−0.1
0.0
0.1
0.2
BETA

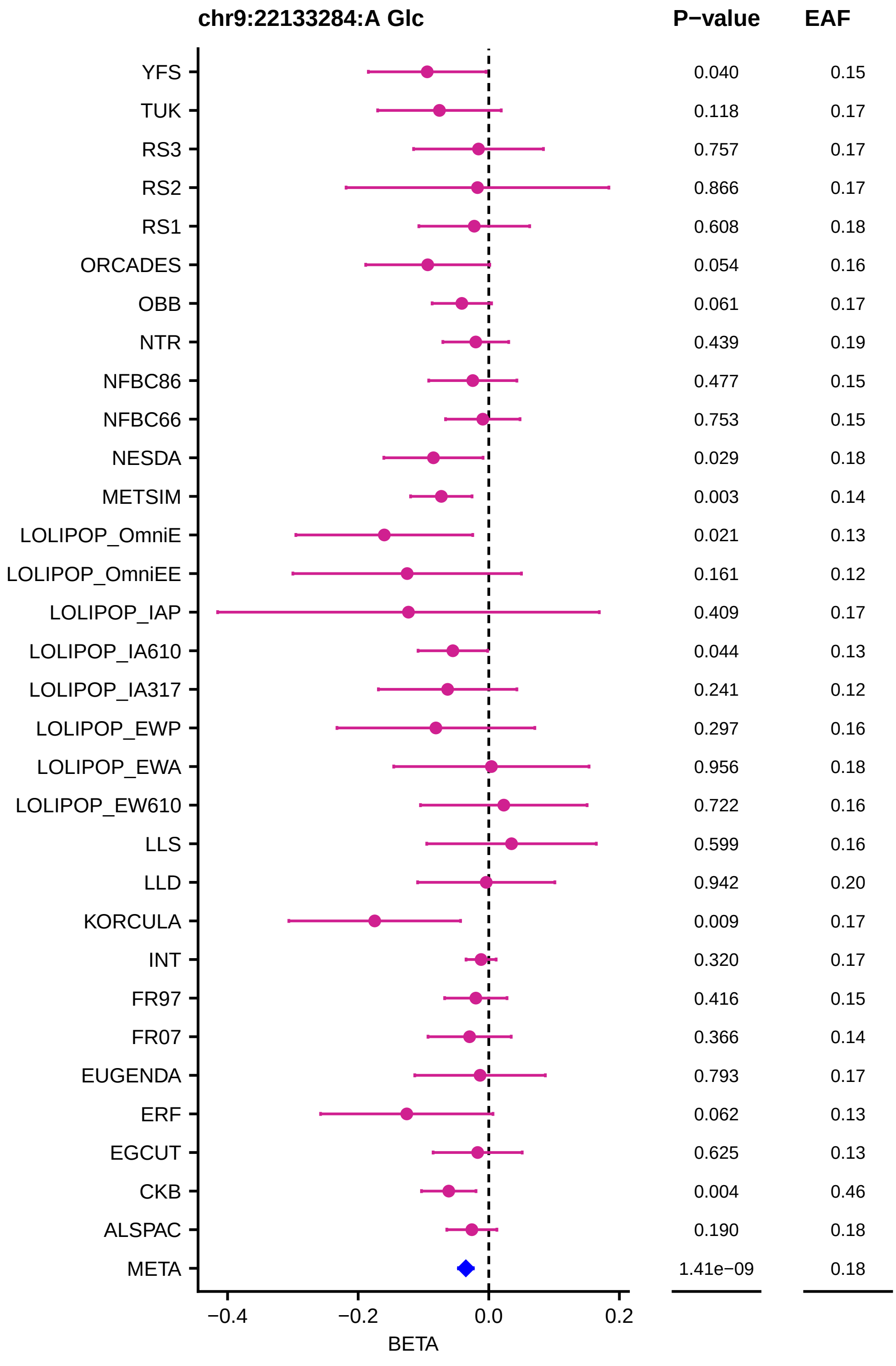
chr9:22133284:A Glc
P−value
EAF
YFS
TUK
RS3
RS2
RS1
ORCADES
OBB
NTR
NFBC86
NFBC66
NESDA
METSIM
LOLIPOP_OmniE
LOLIPOP_OmniEE
LOLIPOP_IAP
LOLIPOP_IA610
LOLIPOP_IA317
LOLIPOP_EWP
LOLIPOP_EWA
LOLIPOP_EW610
LLS
LLD
KORCULA
INT
FR97
FR07
EUGENDA
ERF
EGCUT
CKB
ALSPAC
META
0.040 0.15
0.118 0.17
0.757 0.17
0.866 0.17
0.608 0.18
0.054 0.16
0.061 0.17
0.439 0.19
0.477 0.15
0.753 0.15
0.029 0.18
0.003 0.14
0.021 0.13
0.161 0.12
0.409 0.17
0.044 0.13
0.241 0.12
0.297 0.16
0.956 0.18
0.722 0.16
0.599 0.16
0.942 0.20
0.009 0.17
0.320 0.17
0.416 0.15
0.366 0.14
0.793 0.17
0.062 0.13
0.625 0.13
0.004 0.46
0.190 0.18
1.41e−09 0.18
−0.4
−0.2
0.0
0.2
BETA

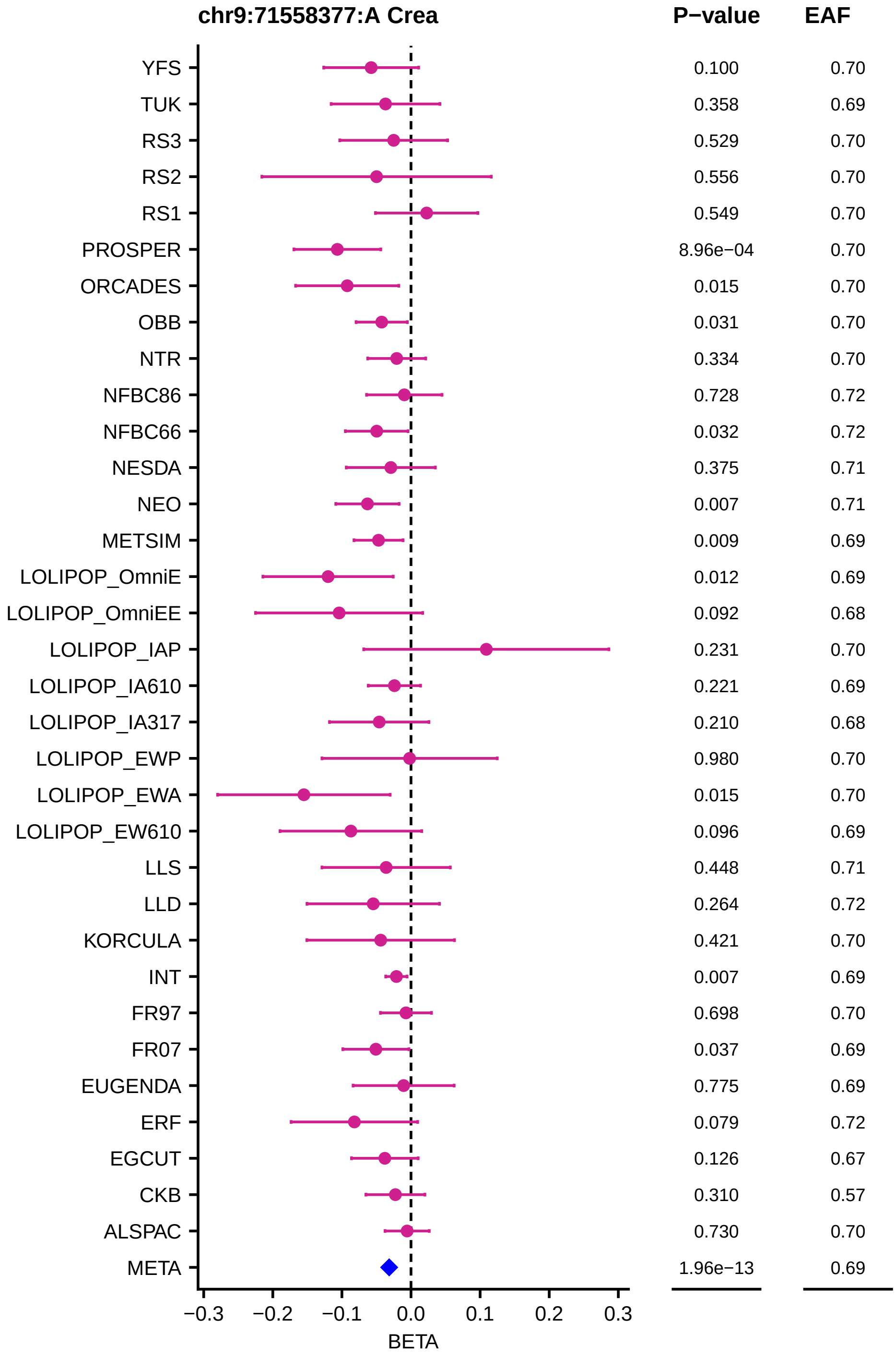

chr9:71558377:A Crea
P−value
EAF
YFS 0.100 0.70
TUK 0.358 0.69
RS3 0.529 0.70
RS2 0.556 0.70
RS1 0.549 0.70
PROSPER 8.96e−04 0.70
ORCADES 0.015 0.70
OBB 0.031 0.70
NTR 0.334 0.70
NFBC86 0.728 0.72
NFBC66 0.032 0.72
NESDA 0.375 0.71
NEO 0.007 0.71
METSIM 0.009 0.69
LOLIPOP_OmniE 0.012 0.69
LOLIPOP_OmniEE 0.092 0.68
LOLIPOP_IAP 0.231 0.70
LOLIPOP_IA610 0.221 0.69
LOLIPOP_IA317 0.210 0.68
LOLIPOP_EWP 0.980 0.70
LOLIPOP_EWA 0.015 0.70
LOLIPOP_EW610 0.096 0.69
LLS 0.448 0.71
LLD 0.264 0.72
KORCULA 0.421 0.70
INT 0.007 0.69
FR97 0.698 0.70
FR07 0.037 0.69
EUGENDA 0.775 0.69
ERF 0.079 0.72
EGCUT 0.126 0.67
CKB 0.310 0.57
ALSPAC 0.730 0.70
META 1.96e−13 0.69
−0.3 −0.2 −0.1 0.0 0.1 0.2 0.3
BETA

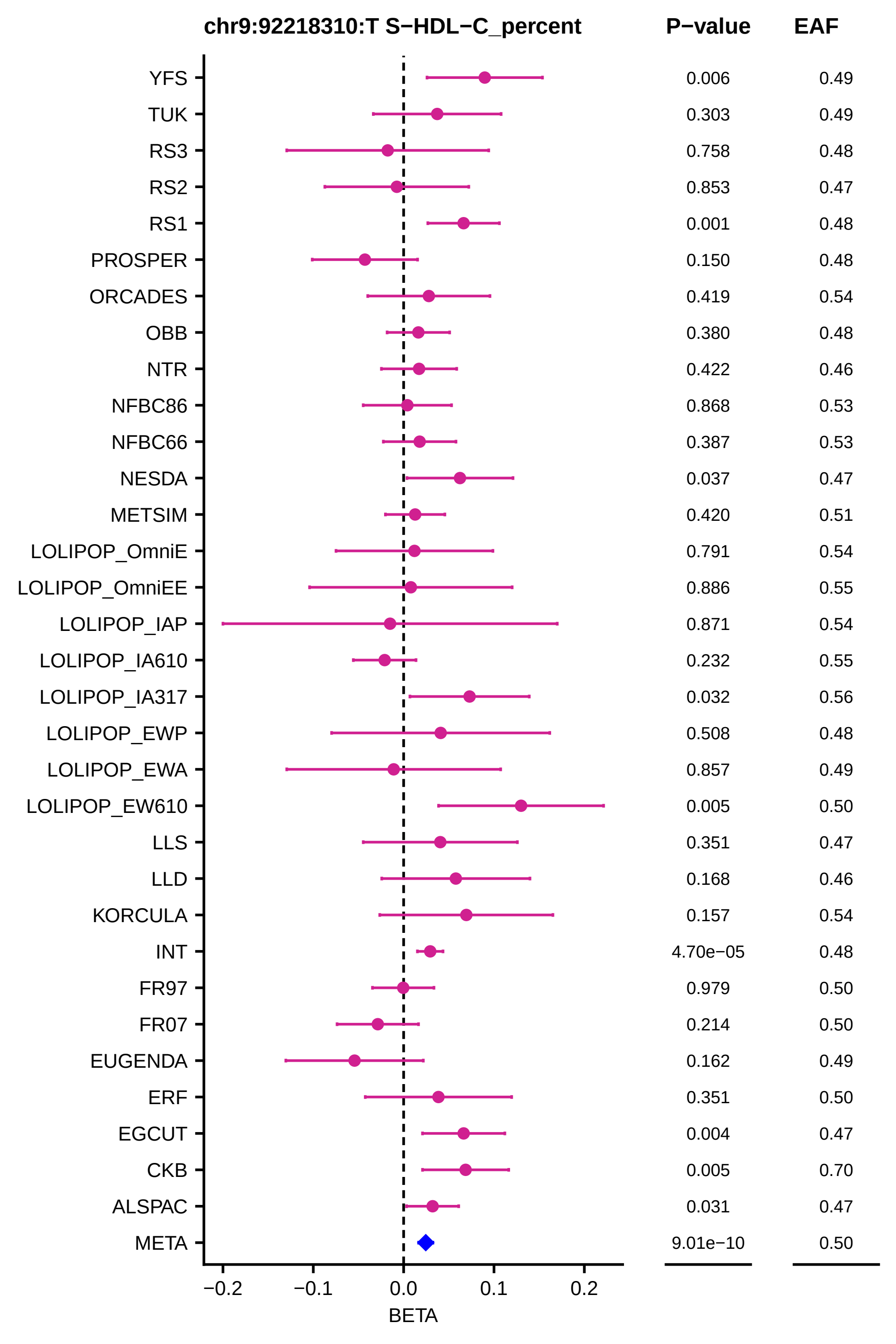
chr9:92218310:T S−HDL−C_percent
P−value
EAF
YFS 0.006 0.49
TUK 0.303 0.49
RS3 0.758 0.48
RS2 0.853 0.47
RS1 0.001 0.48
PROSPER 0.150 0.48
ORCADES 0.419 0.54
OBB 0.380 0.48
NTR 0.422 0.46
NFBC86 0.868 0.53
NFBC66 0.387 0.53
NESDA 0.037 0.47
METSIM 0.420 0.51
LOLIPOP_OmniE 0.791 0.54
LOLIPOP_OmniEE 0.886 0.55
LOLIPOP_IAP 0.871 0.54
LOLIPOP_IA610 0.232 0.55
LOLIPOP_IA317 0.032 0.56
LOLIPOP_EWP 0.508 0.48
LOLIPOP_EWA 0.857 0.49
LOLIPOP_EW610 0.005 0.50
LLS 0.351 0.47
LLD 0.168 0.46
KORCULA 0.157 0.54
INT 4.70e−05 0.48
FR97 0.979 0.50
FR07 0.214 0.50
EUGENDA 0.162 0.49
ERF 0.351 0.50
EGCUT 0.004 0.47
CKB 0.005 0.70
ALSPAC 0.031 0.47
META 9.01e−10 0.50
−0.2
−0.1
0.0
0.1
0.2
BETA

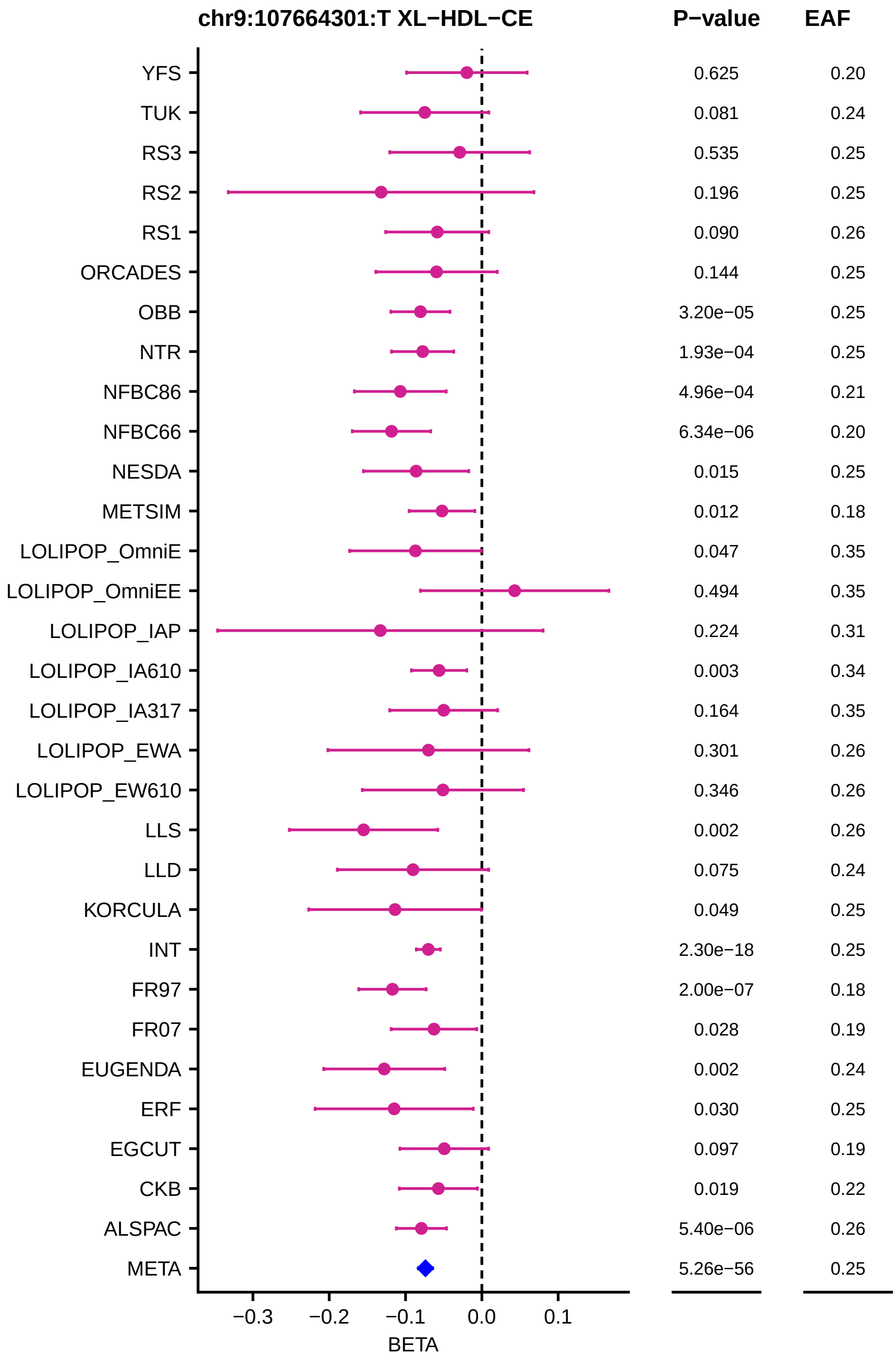
chr9:107664301:T XL−HDL−CE
P−value
EAF
YFS 0.625 0.20
TUK 0.081 0.24
RS3 0.535 0.25
RS2 0.196 0.25
RS1 0.090 0.26
ORCADES 0.144 0.25
OBB 3.20e−05 0.25
NTR 1.93e−04 0.25
NFBC86 4.96e−04 0.21
NFBC66 6.34e−06 0.20
NESDA 0.015 0.25
METSIM 0.012 0.18
LOLIPOP_OmniE 0.047 0.35
LOLIPOP_OmniEE 0.494 0.35
LOLIPOP_IAP 0.224 0.31
LOLIPOP_IA610 0.003 0.34
LOLIPOP_IA317 0.164 0.35
LOLIPOP_EWA 0.301 0.26
LOLIPOP_EW610 0.346 0.26
LLS 0.002 0.26
LLD 0.075 0.24
KORCULA 0.049 0.25
INT 2.30e−18 0.25
FR97 2.00e−07 0.18
FR07 0.028 0.19
EUGENDA 0.002 0.24
ERF 0.030 0.25
EGCUT 0.097 0.19
CKB 0.019 0.22
ALSPAC 5.40e−06 0.26
META 5.26e−56 0.25
−0.3
−0.2
−0.1
0.0
0.1
BETA

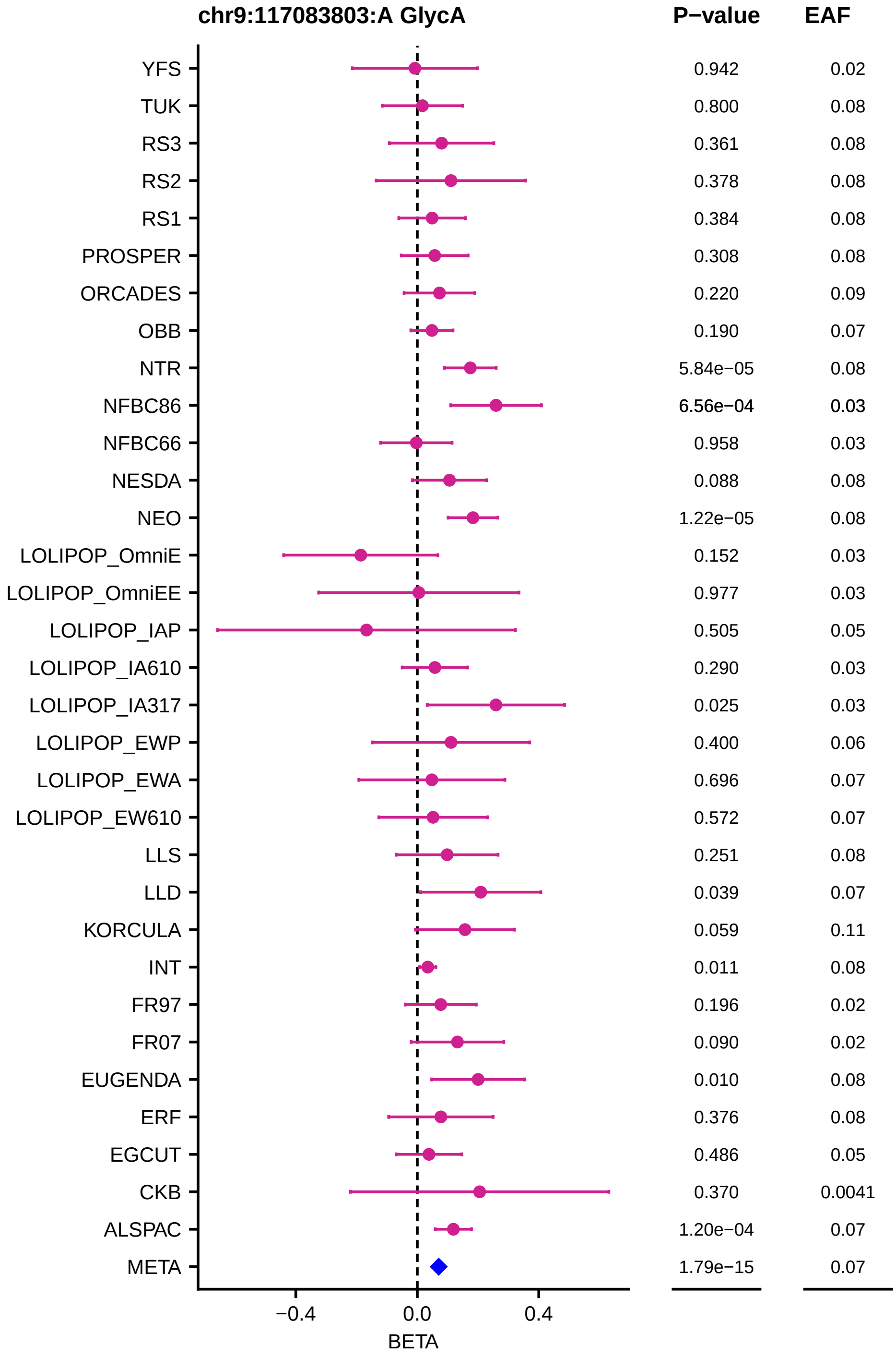

chr9:117083803:A GlycA
P−value
EAF
YFS
TUK
RS3
RS2
RS1
PROSPER
ORCADES
OBB
NTR
NFBC86
NFBC66
NESDA
NEO
LOLIPOP_OmniE
LOLIPOP_OmniEE
LOLIPOP_IAP
LOLIPOP_IA610
LOLIPOP_IA317
LOLIPOP_EWP
LOLIPOP_EWA
LOLIPOP_EW610
LLS
LLD
KORCULA
INT
FR97
FR07
EUGENDA
ERF
EGCUT
CKB
ALSPAC
META
0.942 0.02
0.800 0.08
0.361 0.08
0.378 0.08
0.384 0.08
0.308 0.08
0.220 0.09
0.190 0.07
5.84e−05 0.08
6.56e−04 0.03
0.958 0.03
0.088 0.08
1.22e−05 0.08
0.152 0.03
0.977 0.03
0.505 0.05
0.290 0.03
0.025 0.03
0.400 0.06
0.696 0.07
0.572 0.07
0.251 0.08
0.039 0.07
0.059 0.11
0.011 0.08
0.196 0.02
0.090 0.02
0.010 0.08
0.376 0.08
0.486 0.05
0.370 0.0041
1.20e−04 0.07
1.79e−15 0.07
−0.4
0.0
0.4
BETA

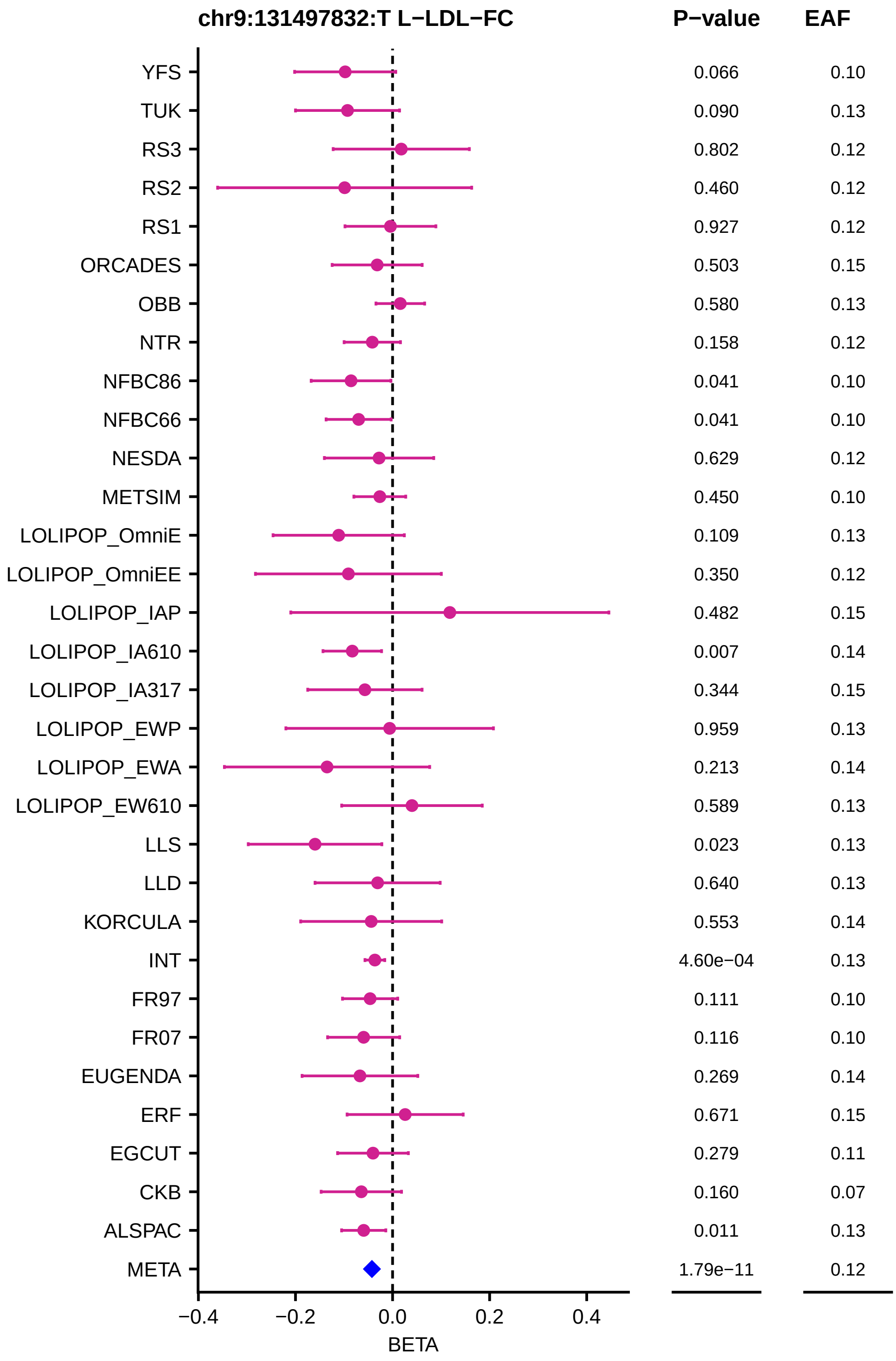
chr9:131497832:T L−LDL−FC
P−value
EAF
YFS 0.066 0.10
TUK 0.090 0.13
RS3 0.802 0.12
RS2 0.460 0.12
RS1 0.927 0.12
ORCADES 0.503 0.15
OBB 0.580 0.13
NTR 0.158 0.12
NFBC86 0.041 0.10
NFBC66 0.041 0.10
NESDA 0.629 0.12
METSIM 0.450 0.10
LOLIPOP_OmniE 0.109 0.13
LOLIPOP_OmniEE 0.350 0.12
LOLIPOP_IAP 0.482 0.15
LOLIPOP_IA610 0.007 0.14
LOLIPOP_IA317 0.344 0.15
LOLIPOP_EWP 0.959 0.13
LOLIPOP_EWA 0.213 0.14
LOLIPOP_EW610 0.589 0.13
LLS 0.023 0.13
LLD 0.640 0.13
KORCULA 0.553 0.14
INT 4.60e−04 0.13
FR97 0.111 0.10
FR07 0.116 0.10
EUGENDA 0.269 0.14
ERF 0.671 0.15
EGCUT 0.279 0.11
CKB 0.160 0.07
ALSPAC 0.011 0.13
META 1.79e−11 0.12
−0.4
−0.2
0.0
0.2
0.4
BETA

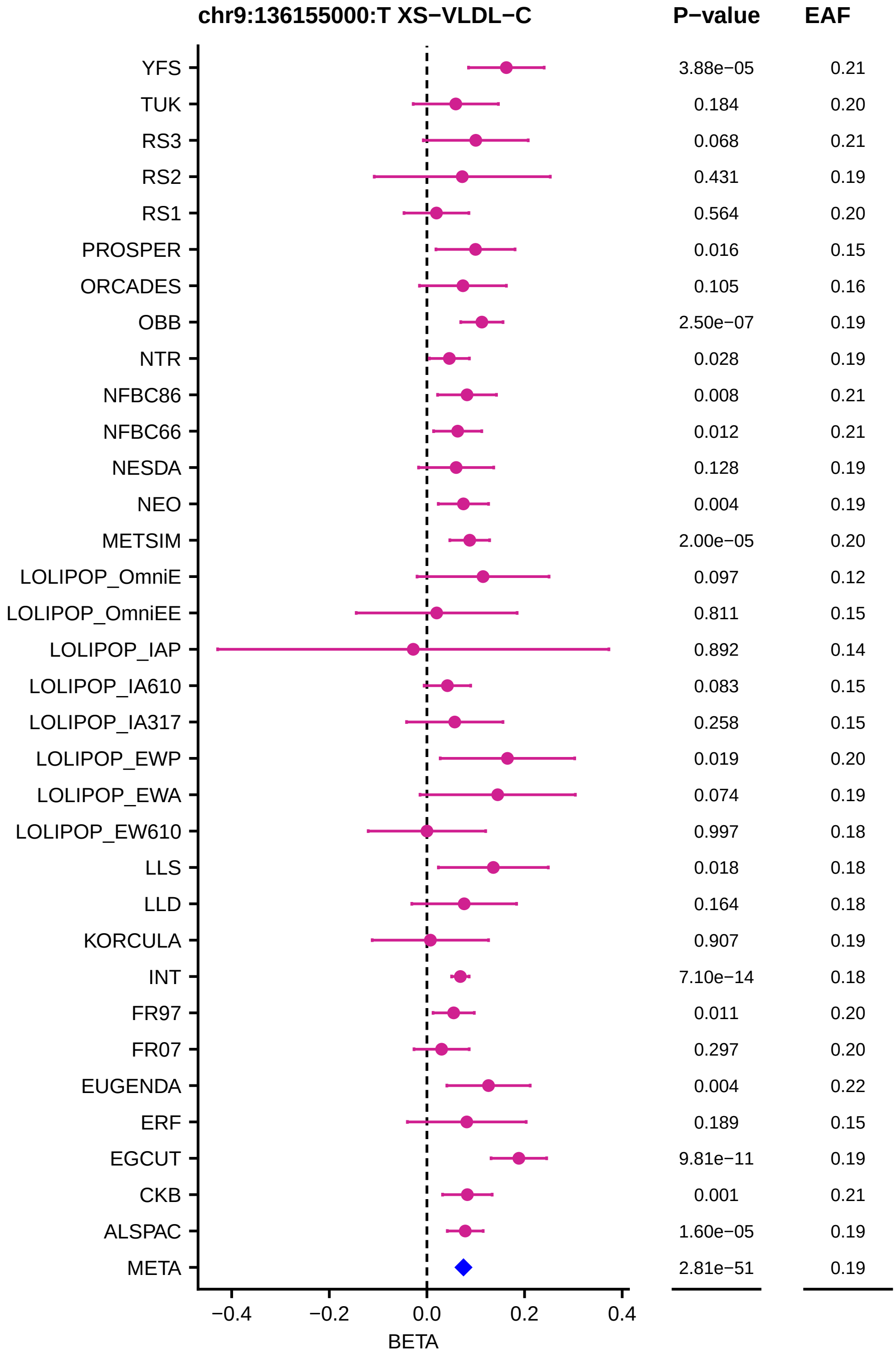

chr9:136155000:T XS−VLDL−C
P−value
EAF
YFS
TUK
RS3
RS2
RS1
PROSPER
ORCADES
OBB
NTR
NFBC86
NFBC66
NESDA
NEO
METSIM
LOLIPOP_OmniE
LOLIPOP_OmniEE
LOLIPOP_IAP
LOLIPOP_IA610
LOLIPOP_IA317
LOLIPOP_EWP
LOLIPOP_EWA
LOLIPOP_EW610
LLS
LLD
KORCULA
INT
FR97
FR07
EUGENDA
ERF
EGCUT
CKB
ALSPAC
META
3.88e−05 0.21
0.184 0.20
0.068 0.21
0.431 0.19
0.564 0.20
0.016 0.15
0.105 0.16
2.50e−07 0.19
0.028 0.19
0.008 0.21
0.012 0.21
0.128 0.19
0.004 0.19
2.00e−05 0.20
0.097 0.12
0.811 0.15
0.892 0.14
0.083 0.15
0.258 0.15
0.019 0.20
0.074 0.19
0.997 0.18
0.018 0.18
0.164 0.18
0.907 0.19
7.10e−14 0.18
0.011 0.20
0.297 0.20
0.004 0.22
0.189 0.15
9.81e−11 0.19
0.001 0.21
1.60e−05 0.19
2.81e−51 0.19
−0.4
−0.2
0.0
0.2
0.4
BETA

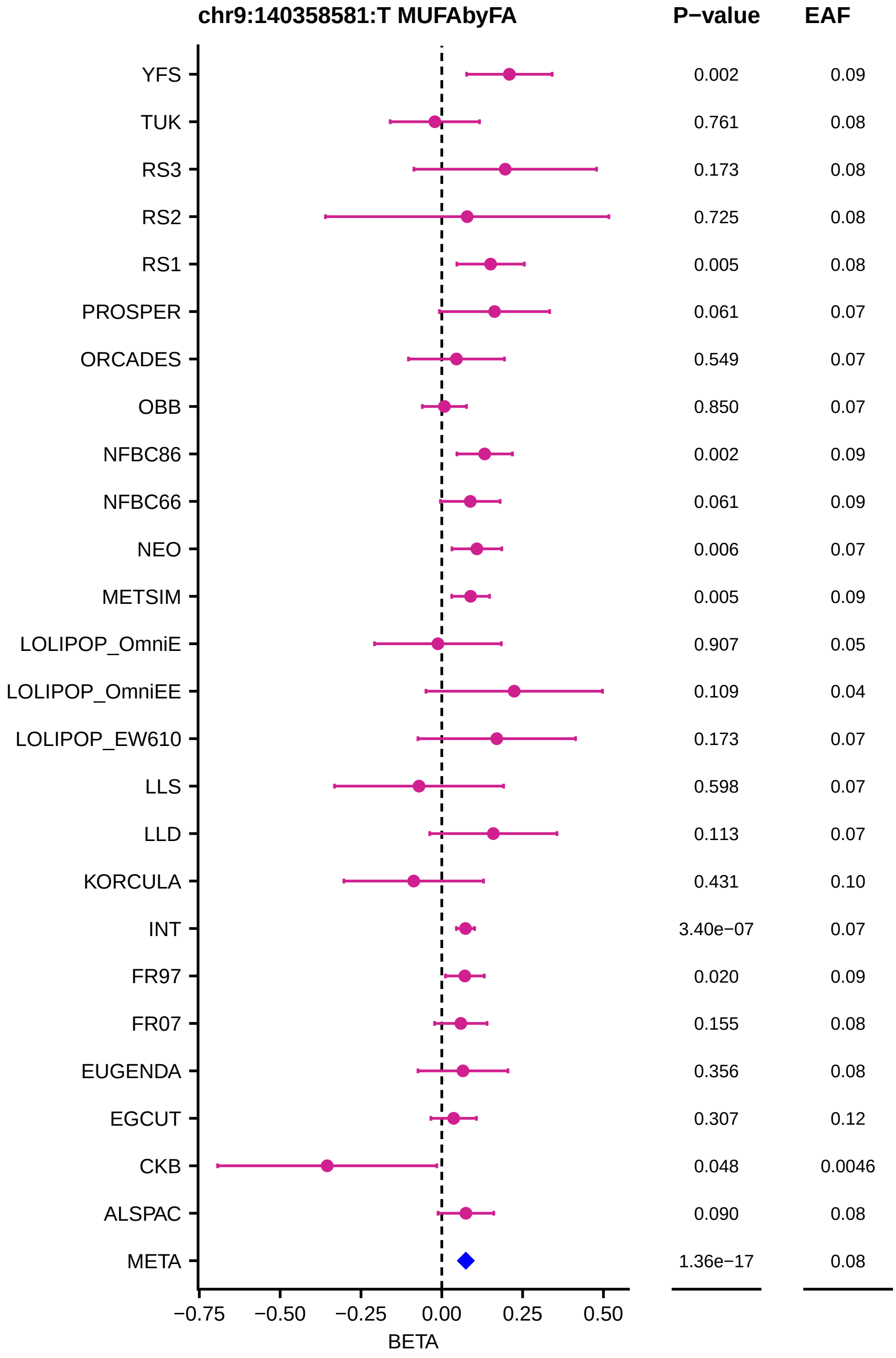
chr9:140358581:T MUFAbyFA
P−value
EAF
YFS 0.002 0.09
TUK 0.761 0.08
RS3 0.173 0.08
RS2 0.725 0.08
RS1 0.005 0.08
PROSPER 0.061 0.07
ORCADES 0.549 0.07
OBB 0.850 0.07
NFBC86 0.002 0.09
NFBC66 0.061 0.09
NEO 0.006 0.07
METSIM 0.005 0.09
LOLIPOP_OmniE 0.907 0.05
LOLIPOP_OmniEE 0.109 0.04
LOLIPOP_EW610 0.173 0.07
LLS 0.598 0.07
LLD 0.113 0.07
KORCULA 0.431 0.10
INT 3.40e−07 0.07
FR97 0.020 0.09
FR07 0.155 0.08
EUGENDA 0.356 0.08
EGCUT 0.307 0.12
CKB 0.048 0.0046
ALSPAC 0.090 0.08
META 1.36e−17 0.08
−0.75 −0.50 −0.25 0.00 0.25 0.50
BETA

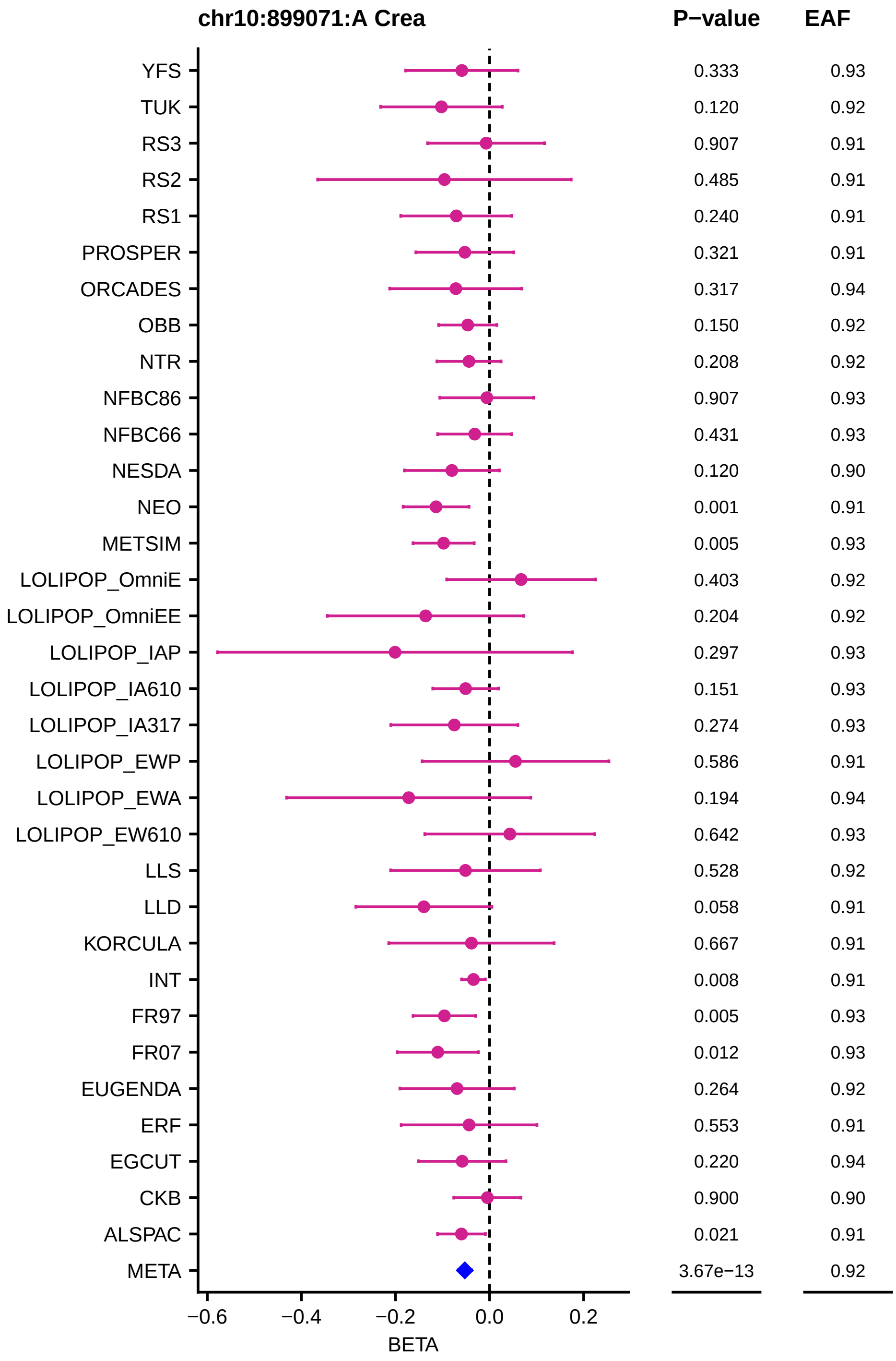
chr10:899071:A Crea
P−value
EAF
YFS 0.333 0.93
TUK 0.120 0.92
RS3 0.907 0.91
RS2 0.485 0.91
RS1 0.240 0.91
PROSPER 0.321 0.91
ORCADES 0.317 0.94
OBB 0.150 0.92
NTR 0.208 0.92
NFBC86 0.907 0.93
NFBC66 0.431 0.93
NESDA 0.120 0.90
NEO 0.001 0.91
METSIM 0.005 0.93
LOLIPOP_OmniE 0.403 0.92
LOLIPOP_OmniEE 0.204 0.92
LOLIPOP_IAP 0.297 0.93
LOLIPOP_IA610 0.151 0.93
LOLIPOP_IA317 0.274 0.93
LOLIPOP_EWP 0.586 0.91
LOLIPOP_EWA 0.194 0.94
LOLIPOP_EW610 0.642 0.93
LLS 0.528 0.92
LLD 0.058 0.91
KORCULA 0.667 0.91
INT 0.008 0.91
FR97 0.005 0.93
FR07 0.012 0.93
EUGENDA 0.264 0.92
ERF 0.553 0.91
EGCUT 0.220 0.94
CKB 0.900 0.90
ALSPAC 0.021 0.91
META 3.67e−13 0.92
−0.6
−0.4
−0.2
0.0
0.2
BETA

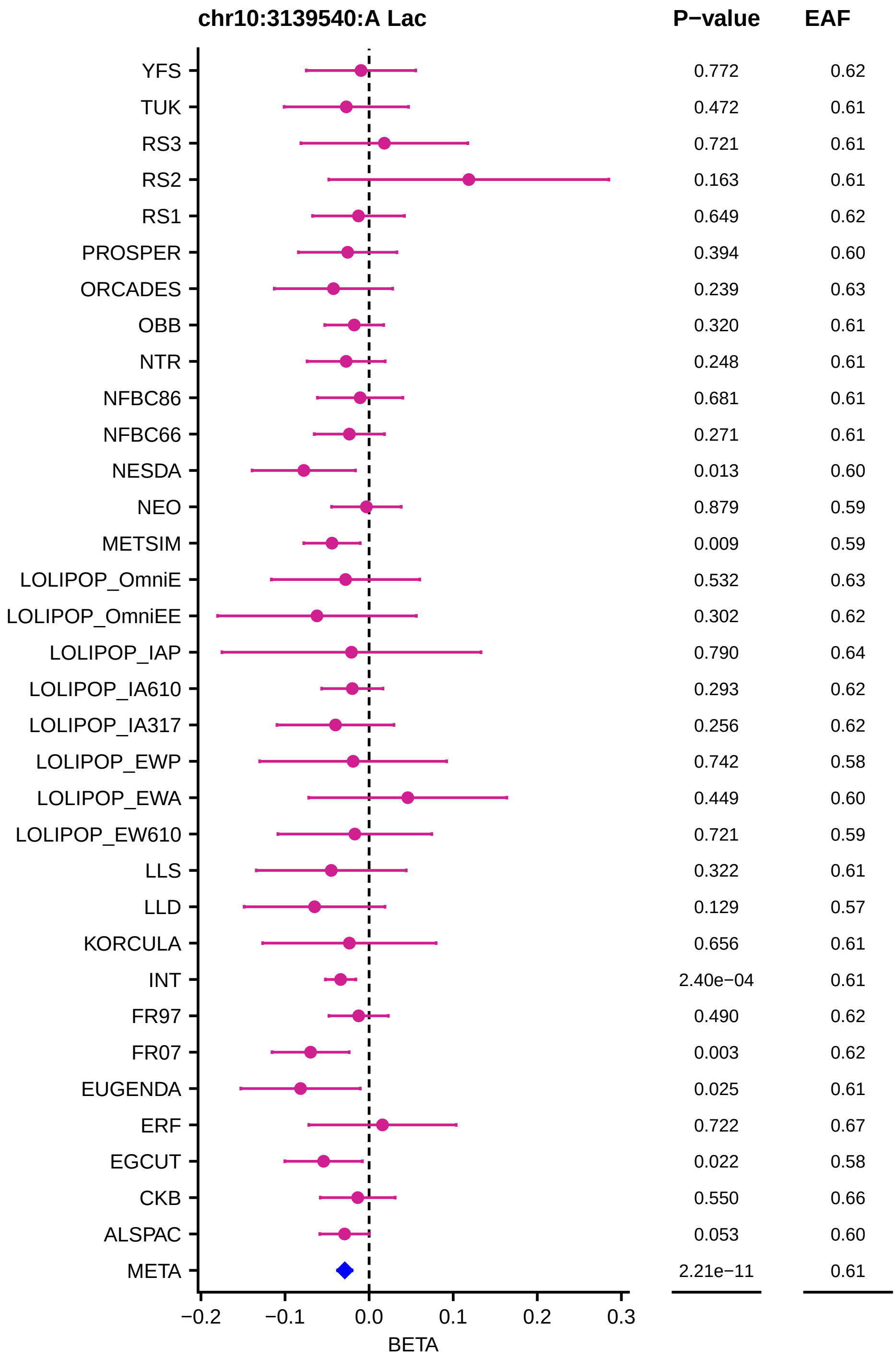
chr10:3139540:A Lac
P−value
EAF
YFS 0.772 0.62
TUK 0.472 0.61
RS3 0.721 0.61
RS2 0.163 0.61
RS1 0.649 0.62
PROSPER 0.394 0.60
ORCADES 0.239 0.63
OBB 0.320 0.61
NTR 0.248 0.61
NFBC86 0.681 0.61
NFBC66 0.271 0.61
NESDA 0.013 0.60
NEO 0.879 0.59
METSIM 0.009 0.59
LOLIPOP_OmniE 0.532 0.63
LOLIPOP_OmniEE 0.302 0.62
LOLIPOP_IAP 0.790 0.64
LOLIPOP_IA610 0.293 0.62
LOLIPOP_IA317 0.256 0.62
LOLIPOP_EWP 0.742 0.58
LOLIPOP_EWA 0.449 0.60
LOLIPOP_EW610 0.721 0.59
LLS 0.322 0.61
LLD 0.129 0.57
KORCULA 0.656 0.61
INT 2.40e−04 0.61
FR97 0.490 0.62
FR07 0.003 0.62
EUGENDA 0.025 0.61
ERF 0.722 0.67
EGCUT 0.022 0.58
CKB 0.550 0.66
ALSPAC 0.053 0.60
META 2.21e−11 0.61
−0.2
−0.1
0.0
0.1
0.2
0.3
BETA

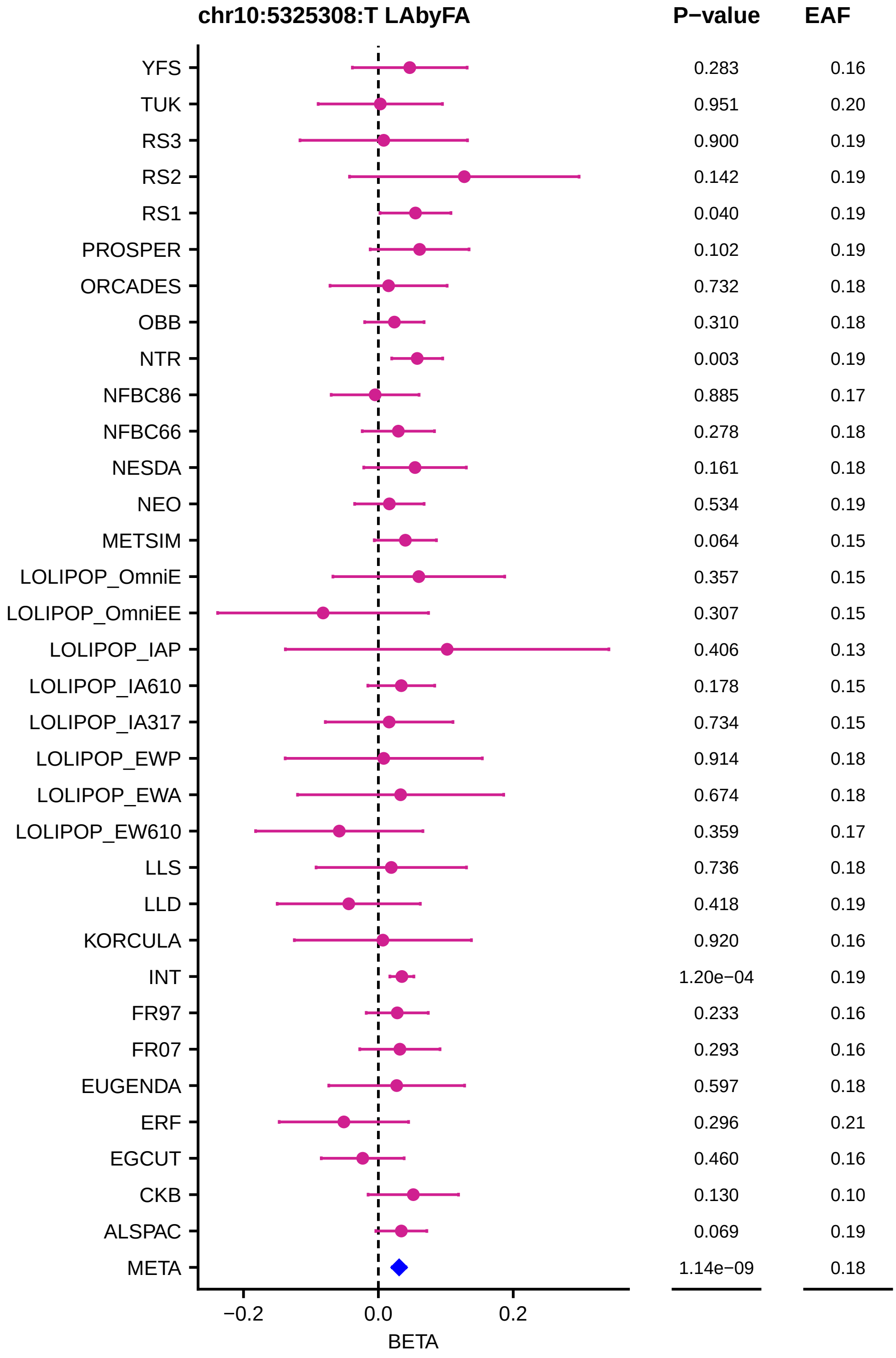
chr10:5325308:T LAbyFA
P−value
EAF
YFS 0.283 0.16
TUK 0.951 0.20
RS3 0.900 0.19
RS2 0.142 0.19
RS1 0.040 0.19
PROSPER 0.102 0.19
ORCADES 0.732 0.18
OBB 0.310 0.18
NTR 0.003 0.19
NFBC86 0.885 0.17
NFBC66 0.278 0.18
NESDA 0.161 0.18
NEO 0.534 0.19
METSIM 0.064 0.15
LOLIPOP_OmniE 0.357 0.15
LOLIPOP_OmniEE 0.307 0.15
LOLIPOP_IAP 0.406 0.13
LOLIPOP_IA610 0.178 0.15
LOLIPOP_IA317 0.734 0.15
LOLIPOP_EWP 0.914 0.18
LOLIPOP_EWA 0.674 0.18
LOLIPOP_EW610 0.359 0.17
LLS 0.736 0.18
LLD 0.418 0.19
KORCULA 0.920 0.16
INT 1.20e−04 0.19
FR97 0.233 0.16
FR07 0.293 0.16
EUGENDA 0.597 0.18
ERF 0.296 0.21
EGCUT 0.460 0.16
CKB 0.130 0.10
ALSPAC 0.069 0.19
META 1.14e−09 0.18
−0.2
0.0
0.2
BETA

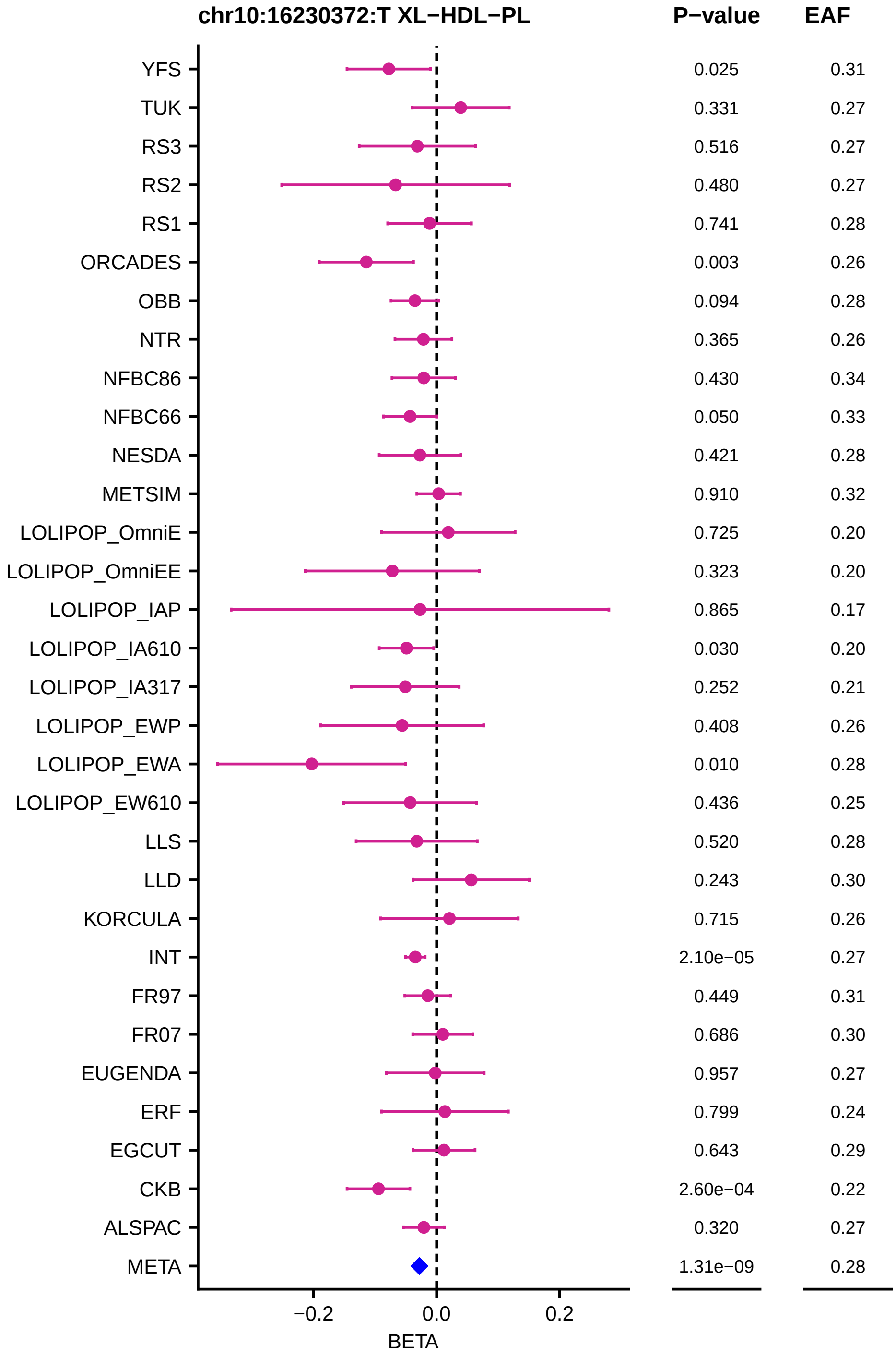

chr10:16230372:T XL−HDL−PL
P−value
EAF
YFS 0.025 0.31
TUK 0.331 0.27
RS3 0.516 0.27
RS2 0.480 0.27
RS1 0.741 0.28
ORCADES 0.003 0.26
OBB 0.094 0.28
NTR 0.365 0.26
NFBC86 0.430 0.34
NFBC66 0.050 0.33
NESDA 0.421 0.28
METSIM 0.910 0.32
LOLIPOP_OmniE 0.725 0.20
LOLIPOP_OmniEE 0.323 0.20
LOLIPOP_IAP 0.865 0.17
LOLIPOP_IA610 0.030 0.20
LOLIPOP_IA317 0.252 0.21
LOLIPOP_EWP 0.408 0.26
LOLIPOP_EWA 0.010 0.28
LOLIPOP_EW610 0.436 0.25
LLS 0.520 0.28
LLD 0.243 0.30
KORCULA 0.715 0.26
INT 2.10e−05 0.27
FR97 0.449 0.31
FR07 0.686 0.30
EUGENDA 0.957 0.27
ERF 0.799 0.24
EGCUT 0.643 0.29
CKB 2.60e−04 0.22
ALSPAC 0.320 0.27
META 1.31e−09 0.28
−0.2
0.0
0.2
BETA

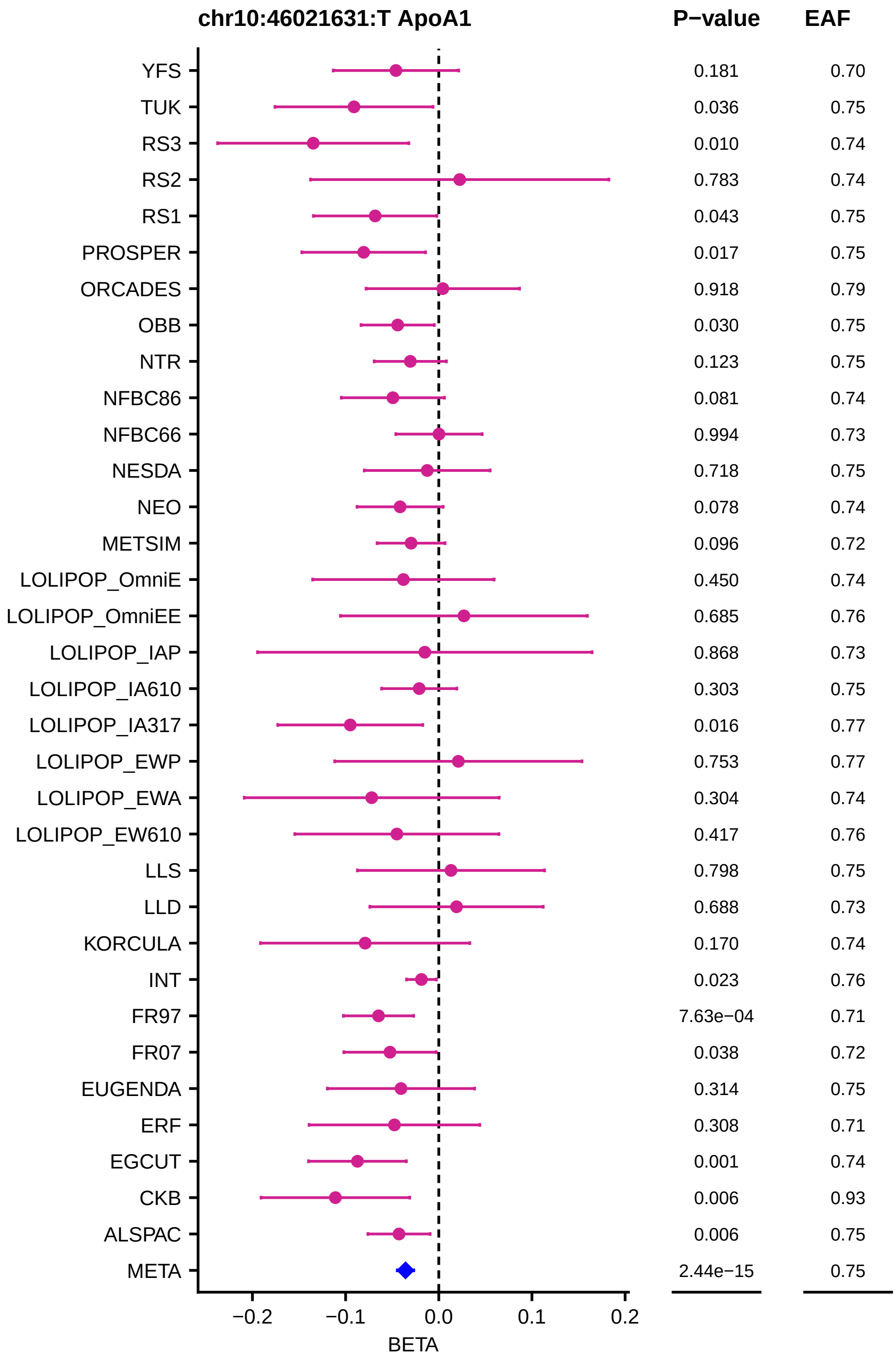
chr10:46021631:T ApoA1
P−value
EAF
YFS 0.181 0.70
TUK 0.036 0.75
RS3 0.010 0.74
RS2 0.783 0.74
RS1 0.043 0.75
PROSPER 0.017 0.75
ORCADES 0.918 0.79
OBB 0.030 0.75
NTR 0.123 0.75
NFBC86 0.081 0.74
NFBC66 0.994 0.73
NESDA 0.718 0.75
NEO 0.078 0.74
METSIM 0.096 0.72
LOLIPOP_OmniE 0.450 0.74
LOLIPOP_OmniEE 0.685 0.76
LOLIPOP_IAP 0.868 0.73
LOLIPOP_IA610 0.303 0.75
LOLIPOP_IA317 0.016 0.77
LOLIPOP_EWP 0.753 0.77
LOLIPOP_EWA 0.304 0.74
LOLIPOP_EW610 0.417 0.76
LLS 0.798 0.75
LLD 0.688 0.73
KORCULA 0.170 0.74
INT 0.023 0.76
FR97 7.63e−04 0.71
FR07 0.038 0.72
EUGENDA 0.314 0.75
ERF 0.308 0.71
EGCUT 0.001 0.74
CKB 0.006 0.93
ALSPAC 0.006 0.75
META 2.44e−15 0.75
−0.2
−0.1
0.0
0.1
0.2
BETA

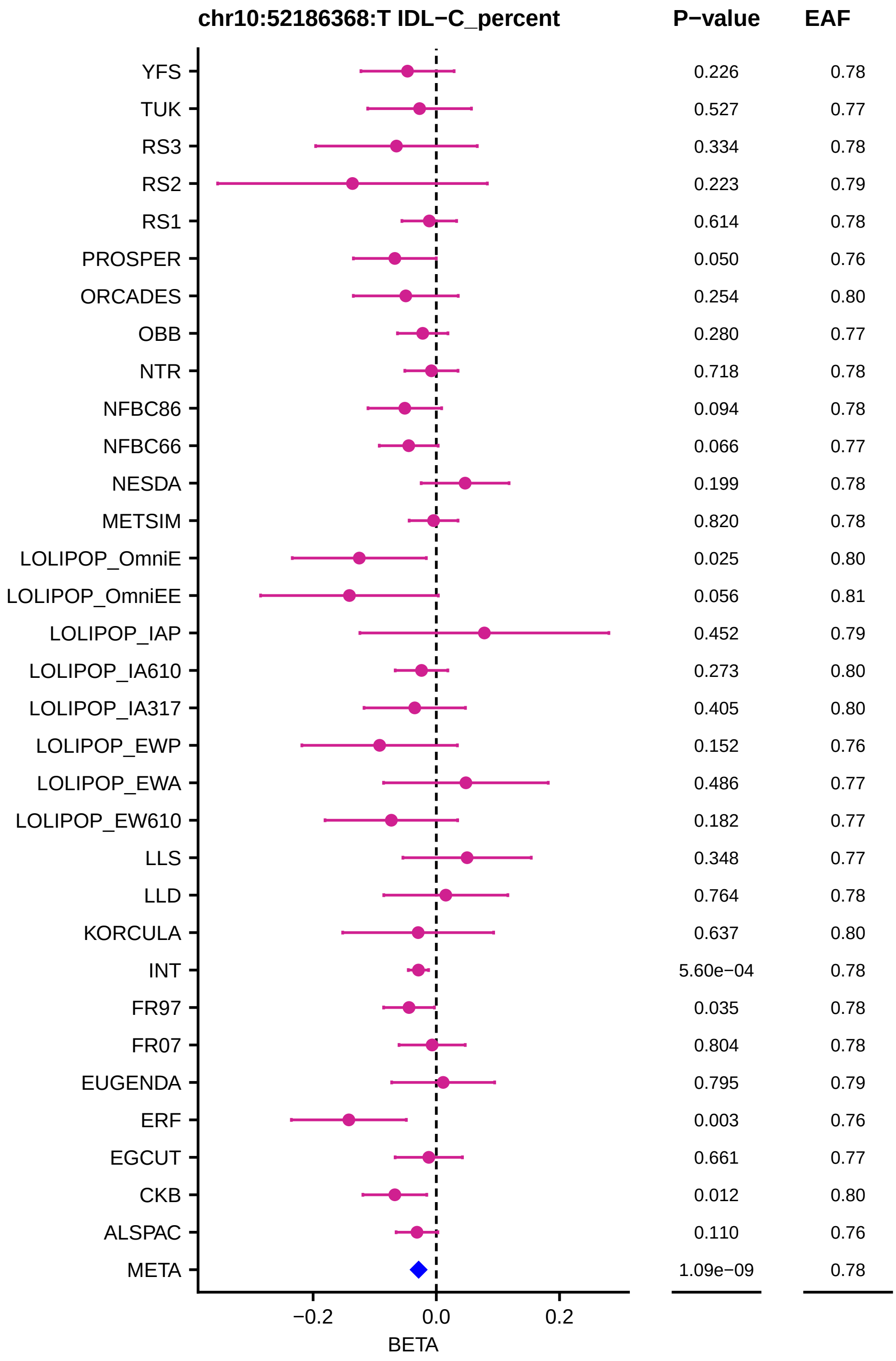
chr10:52186368:T IDL−C_percent
P−value
EAF
YFS 0.226 0.78
TUK 0.527 0.77
RS3 0.334 0.78
RS2 0.223 0.79
RS1 0.614 0.78
PROSPER 0.050 0.76
ORCADES 0.254 0.80
OBB 0.280 0.77
NTR 0.718 0.78
NFBC86 0.094 0.78
NFBC66 0.066 0.77
NESDA 0.199 0.78
METSIM 0.820 0.78
LOLIPOP_OmniE 0.025 0.80
LOLIPOP_OmniEE 0.056 0.81
LOLIPOP_IAP 0.452 0.79
LOLIPOP_IA610 0.273 0.80
LOLIPOP_IA317 0.405 0.80
LOLIPOP_EWP 0.152 0.76
LOLIPOP_EWA 0.486 0.77
LOLIPOP_EW610 0.182 0.77
LLS 0.348 0.77
LLD 0.764 0.78
KORCULA 0.637 0.80
INT 5.60e−04 0.78
FR97 0.035 0.78
FR07 0.804 0.78
EUGENDA 0.795 0.79
ERF 0.003 0.76
EGCUT 0.661 0.77
CKB 0.012 0.80
ALSPAC 0.110 0.76
META 1.09e−09 0.78
−0.2
0.0
0.2
BETA

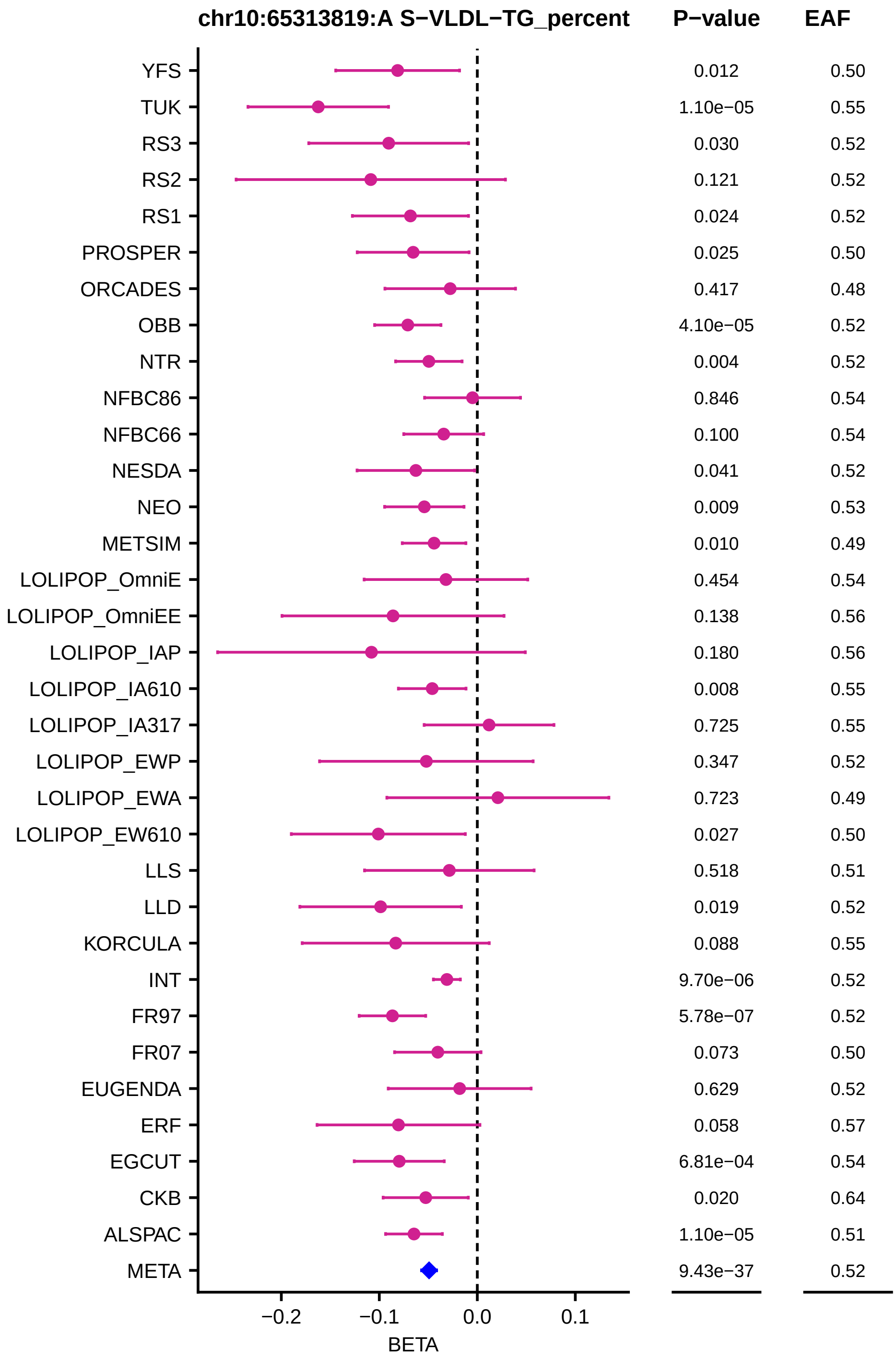
chr10:65313819:A S−VLDL−TG_percent
P−value
EAF
YFS 0.012 0.50
TUK 1.10e−05 0.55
RS3 0.030 0.52
RS2 0.121 0.52
RS1 0.024 0.52
PROSPER 0.025 0.50
ORCADES 0.417 0.48
OBB 4.10e−05 0.52
NTR 0.004 0.52
NFBC86 0.846 0.54
NFBC66 0.100 0.54
NESDA 0.041 0.52
NEO 0.009 0.53
METSIM 0.010 0.49
LOLIPOP_OmniE 0.454 0.54
LOLIPOP_OmniEE 0.138 0.56
LOLIPOP_IAP 0.180 0.56
LOLIPOP_IA610 0.008 0.55
LOLIPOP_IA317 0.725 0.55
LOLIPOP_EWP 0.347 0.52
LOLIPOP_EWA 0.723 0.49
LOLIPOP_EW610 0.027 0.50
LLS 0.518 0.51
LLD 0.019 0.52
KORCULA 0.088 0.55
INT 9.70e−06 0.52
FR97 5.78e−07 0.52
FR07 0.073 0.50
EUGENDA 0.629 0.52
ERF 0.058 0.57
EGCUT 6.81e−04 0.54
CKB 0.020 0.64
ALSPAC 1.10e−05 0.51
META 9.43e−37 0.52
−0.2
−0.1
0.0
0.1
BETA

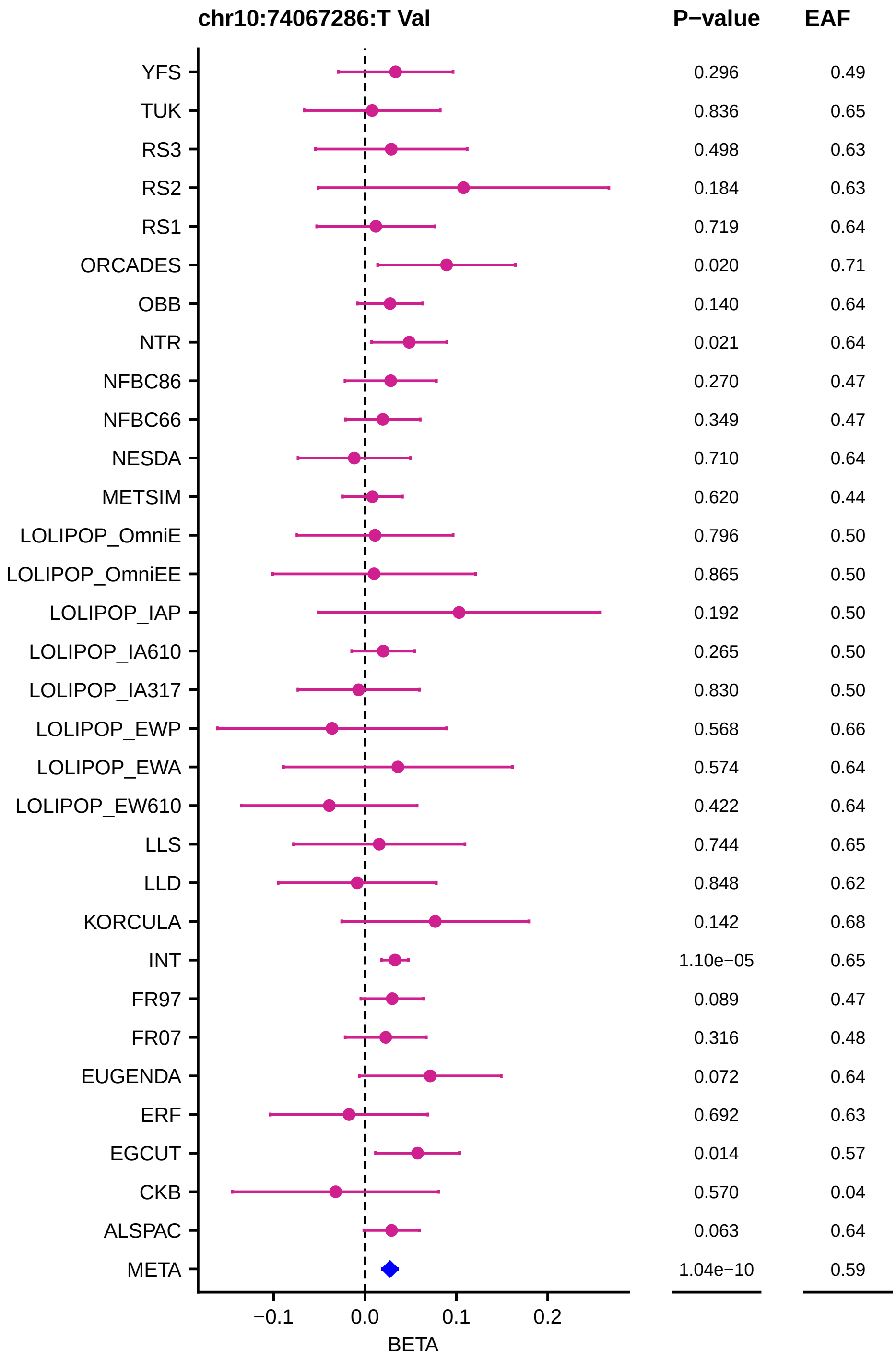
chr10:74067286:T Val
P−value
EAF
YFS 0.296 0.49
TUK 0.836 0.65
RS3 0.498 0.63
RS2 0.184 0.63
RS1 0.719 0.64
ORCADES 0.020 0.71
OBB 0.140 0.64
NTR 0.021 0.64
NFBC86 0.270 0.47
NFBC66 0.349 0.47
NESDA 0.710 0.64
METSIM 0.620 0.44
LOLIPOP_OmniE 0.796 0.50
LOLIPOP_OmniEE 0.865 0.50
LOLIPOP_IAP 0.192 0.50
LOLIPOP_IA610 0.265 0.50
LOLIPOP_IA317 0.830 0.50
LOLIPOP_EWP 0.568 0.66
LOLIPOP_EWA 0.574 0.64
LOLIPOP_EW610 0.422 0.64
LLS 0.744 0.65
LLD 0.848 0.62
KORCULA 0.142 0.68
INT 1.10e−05 0.65
FR97 0.089 0.47
FR07 0.316 0.48
EUGENDA 0.072 0.64
ERF 0.692 0.63
EGCUT 0.014 0.57
CKB 0.570 0.04
ALSPAC 0.063 0.64
META 1.04e−10 0.59
−0.1
0.0
0.1
0.2
BETA

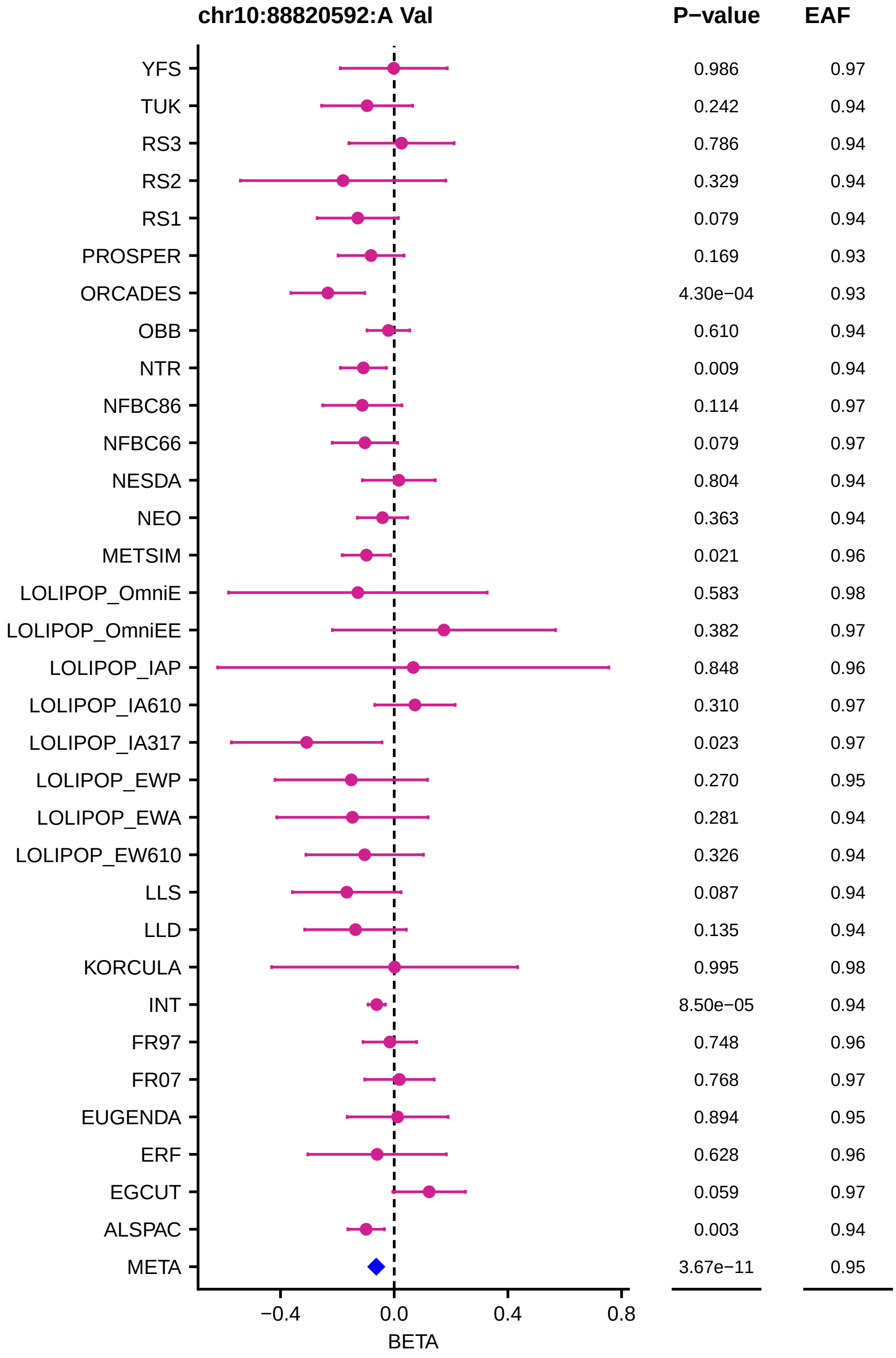

chr10:88820592:A Val
P−value
EAF
YFS 0.986 0.97
TUK 0.242 0.94
RS3 0.786 0.94
RS2 0.329 0.94
RS1 0.079 0.94
PROSPER 0.169 0.93
ORCADES 4.30e−04 0.93
OBB 0.610 0.94
NTR 0.009 0.94
NFBC86 0.114 0.97
NFBC66 0.079 0.97
NESDA 0.804 0.94
NEO 0.363 0.94
METSIM 0.021 0.96
LOLIPOP_OmniE 0.583 0.98
LOLIPOP_OmniEE 0.382 0.97
LOLIPOP_IAP 0.848 0.96
LOLIPOP_IA610 0.310 0.97
LOLIPOP_IA317 0.023 0.97
LOLIPOP_EWP 0.270 0.95
LOLIPOP_EWA 0.281 0.94
LOLIPOP_EW610 0.326 0.94
LLS 0.087 0.94
LLD 0.135 0.94
KORCULA 0.995 0.98
INT 8.50e−05 0.94
FR97 0.748 0.96
FR07 0.768 0.97
EUGENDA 0.894 0.95
ERF 0.628 0.96
EGCUT 0.059 0.97
ALSPAC 0.003 0.94
META 3.67e−11 0.95
−0.4
0.0
0.4
0.8
BETA

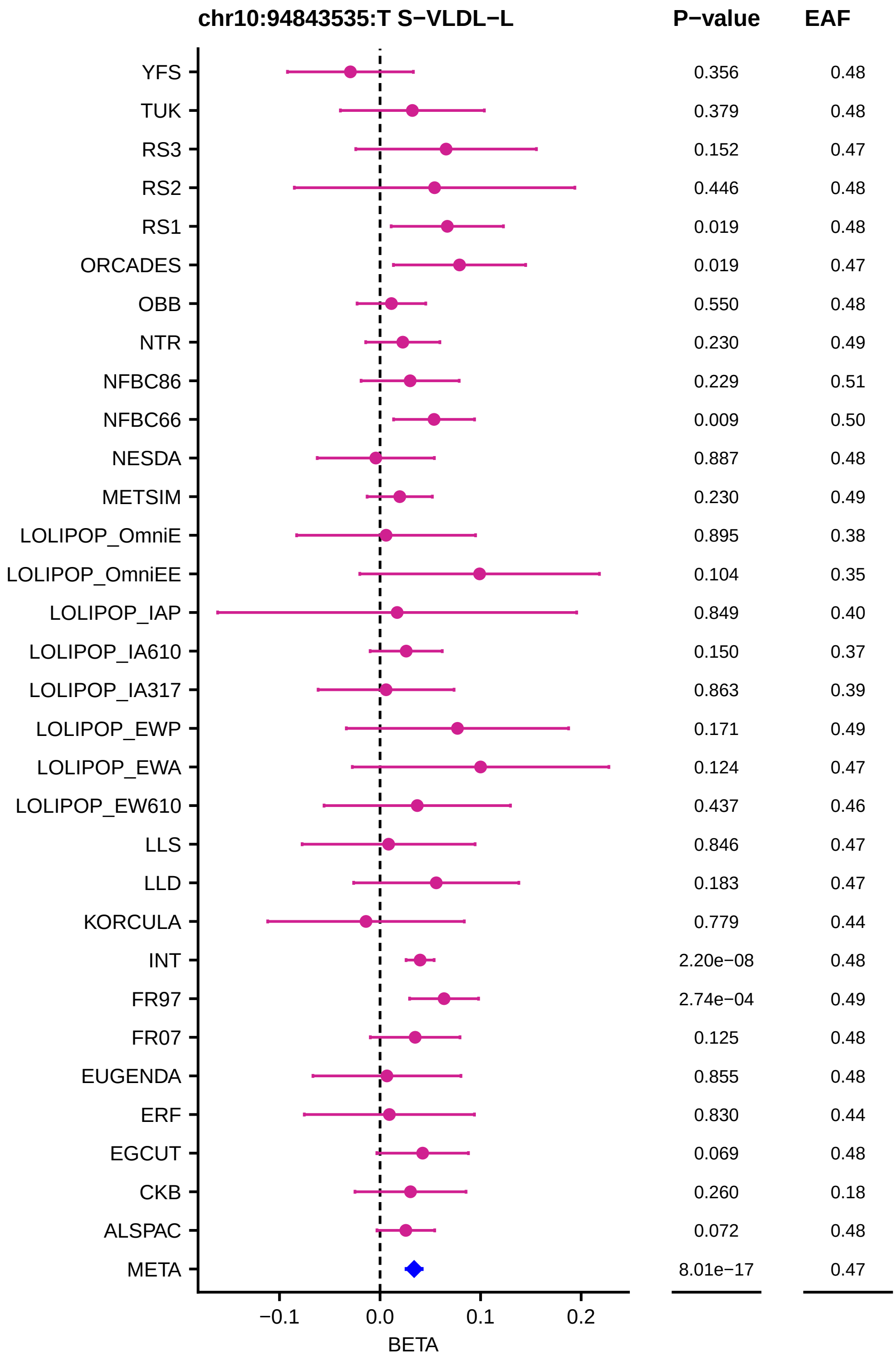
chr10:94843535:T S−VLDL−L
P−value
EAF
YFS
TUK
RS3
RS2
RS1
ORCADES
OBB
NTR
NFBC86
NFBC66
NESDA
METSIM
LOLIPOP_OmniE
LOLIPOP_OmniEE
LOLIPOP_IAP
LOLIPOP_IA610
LOLIPOP_IA317
LOLIPOP_EWP
LOLIPOP_EWA
LOLIPOP_EW610
LLS
LLD
KORCULA
INT
FR97
FR07
EUGENDA
ERF
EGCUT
CKB
ALSPAC
META
0.356 0.48
0.379 0.48
0.152 0.47
0.446 0.48
0.019 0.48
0.019 0.47
0.550 0.48
0.230 0.49
0.229 0.51
0.009 0.50
0.887 0.48
0.230 0.49
0.895 0.38
0.104 0.35
0.849 0.40
0.150 0.37
0.863 0.39
0.171 0.49
0.124 0.47
0.437 0.46
0.846 0.47
0.183 0.47
0.779 0.44
2.20e−08 0.48
2.74e−04 0.49
0.125 0.48
0.855 0.48
0.830 0.44
0.069 0.48
0.260 0.18
0.072 0.48
8.01e−17 0.47
−0.1
0.0
0.1
0.2
BETA

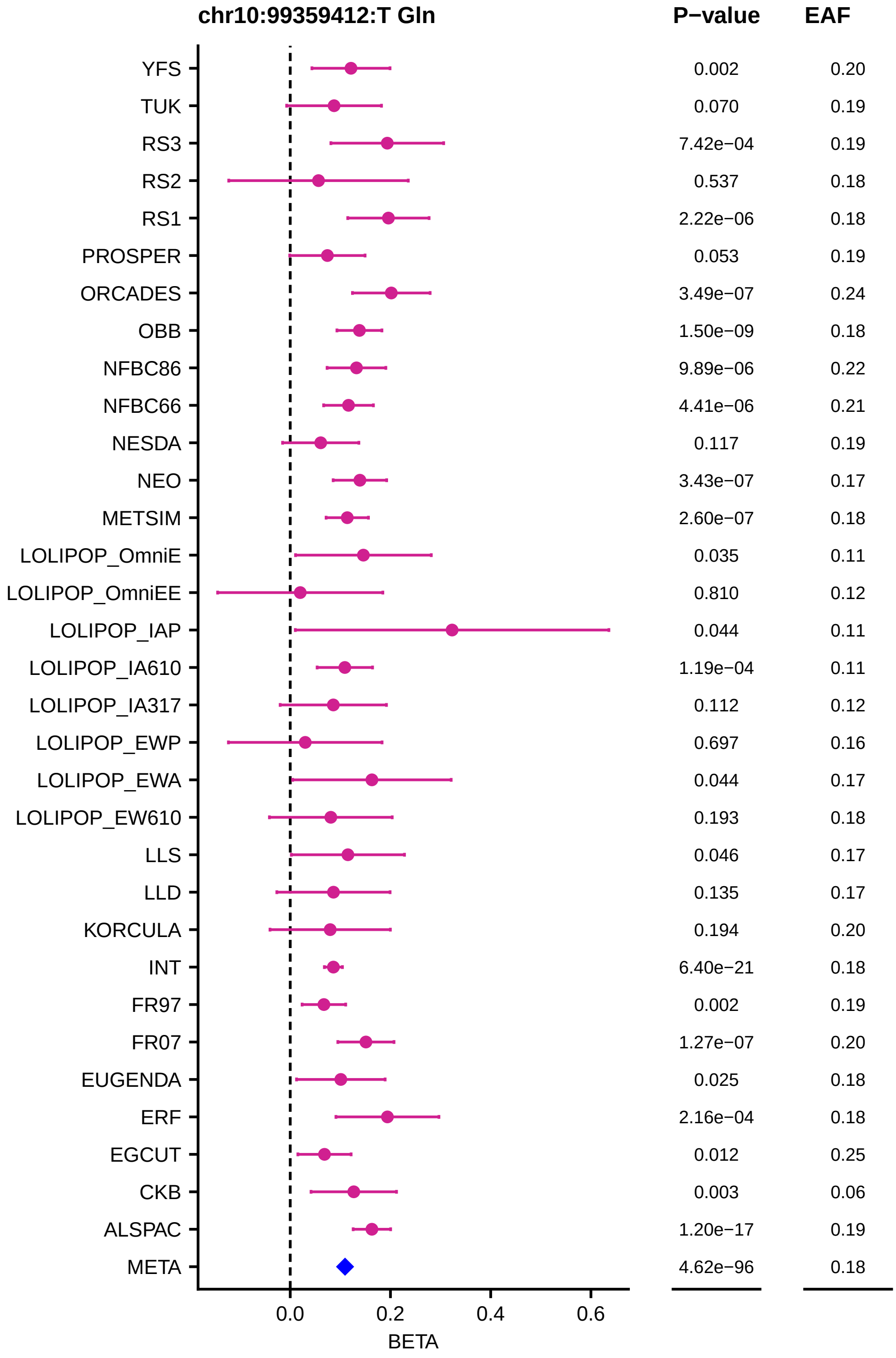

chr10:99359412:T Gln
P−value
EAF
YFS
TUK
RS3
RS2
RS1
PROSPER
ORCADES
OBB
NFBC86
NFBC66
NESDA
NEO
METSIM
LOLIPOP_OmniE
LOLIPOP_OmniEE
LOLIPOP_IAP
LOLIPOP_IA610
LOLIPOP_IA317
LOLIPOP_EWP
LOLIPOP_EWA
LOLIPOP_EW610
LLS
LLD
KORCULA
INT
FR97
FR07
EUGENDA
ERF
EGCUT
CKB
ALSPAC
META
0.002 0.20
0.070 0.19
7.42e−04 0.19
0.537 0.18
2.22e−06 0.18
0.053 0.19
3.49e−07 0.24
1.50e−09 0.18
9.89e−06 0.22
4.41e−06 0.21
0.117 0.19
3.43e−07 0.17
2.60e−07 0.18
0.035 0.11
0.810 0.12
0.044 0.11
1.19e−04 0.11
0.112 0.12
0.697 0.16
0.044 0.17
0.193 0.18
0.046 0.17
0.135 0.17
0.194 0.20
6.40e−21 0.18
0.002 0.19
1.27e−07 0.20
0.025 0.18
2.16e−04 0.18
0.012 0.25
0.003 0.06
1.20e−17 0.19
4.62e−96 0.18
0.0
0.2
0.4
0.6
BETA

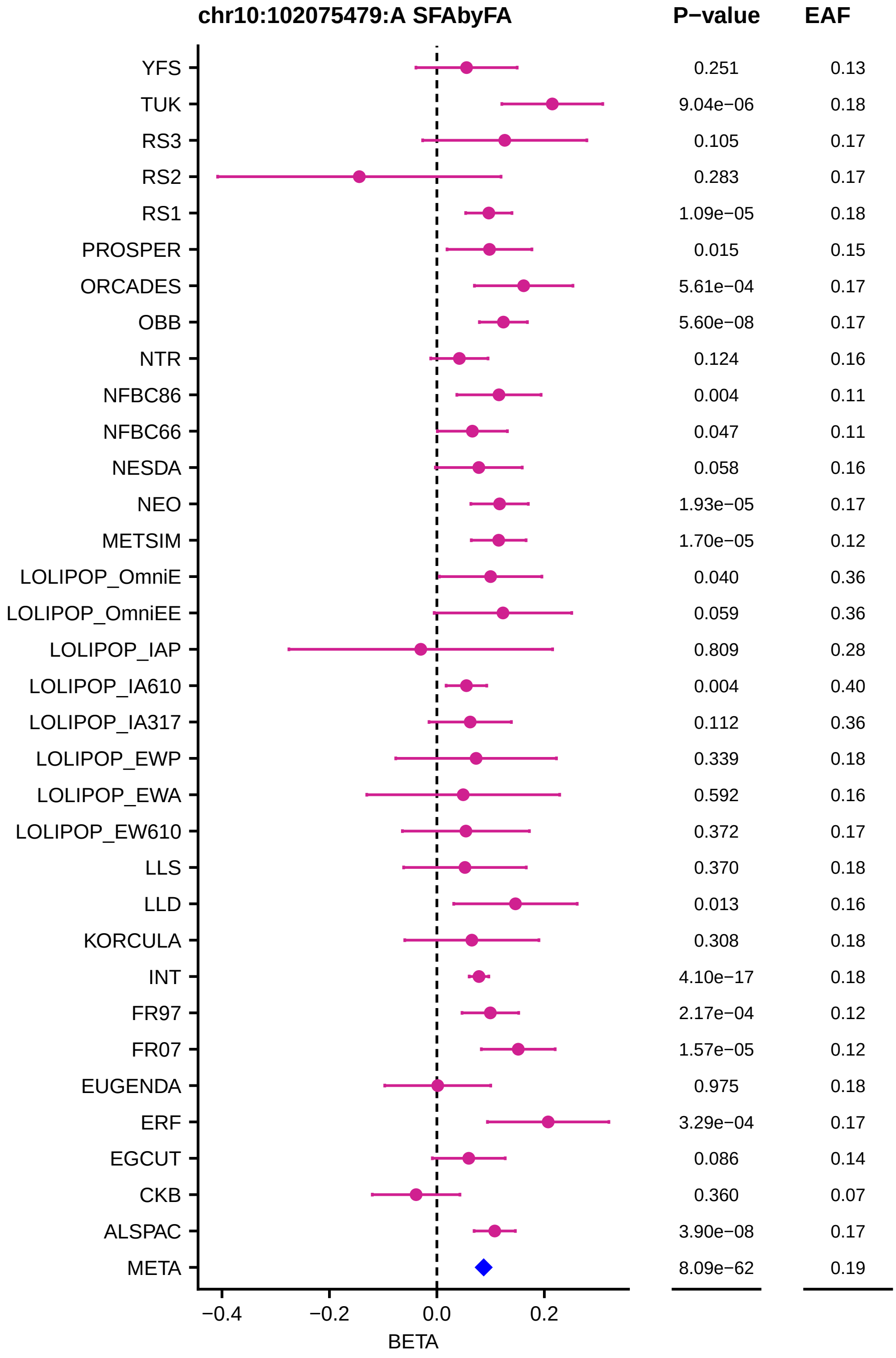
chr10:102075479:A SFAbyFA
P−value
EAF
YFS 0.251 0.13
TUK 9.04e−06 0.18
RS3 0.105 0.17
RS2 0.283 0.17
RS1 1.09e−05 0.18
PROSPER 0.015 0.15
ORCADES 5.61e−04 0.17
OBB 5.60e−08 0.17
NTR 0.124 0.16
NFBC86 0.004 0.11
NFBC66 0.047 0.11
NESDA 0.058 0.16
NEO 1.93e−05 0.17
METSIM 1.70e−05 0.12
LOLIPOP_OmniE 0.040 0.36
LOLIPOP_OmniEE 0.059 0.36
LOLIPOP_IAP 0.809 0.28
LOLIPOP_IA610 0.004 0.40
LOLIPOP_IA317 0.112 0.36
LOLIPOP_EWP 0.339 0.18
LOLIPOP_EWA 0.592 0.16
LOLIPOP_EW610 0.372 0.17
LLS 0.370 0.18
LLD 0.013 0.16
KORCULA 0.308 0.18
INT 4.10e−17 0.18
FR97 2.17e−04 0.12
FR07 1.57e−05 0.12
EUGENDA 0.975 0.18
ERF 3.29e−04 0.17
EGCUT 0.086 0.14
CKB 0.360 0.07
ALSPAC 3.90e−08 0.17
META 8.09e−62 0.19
−0.4
−0.2
0.0
0.2
BETA

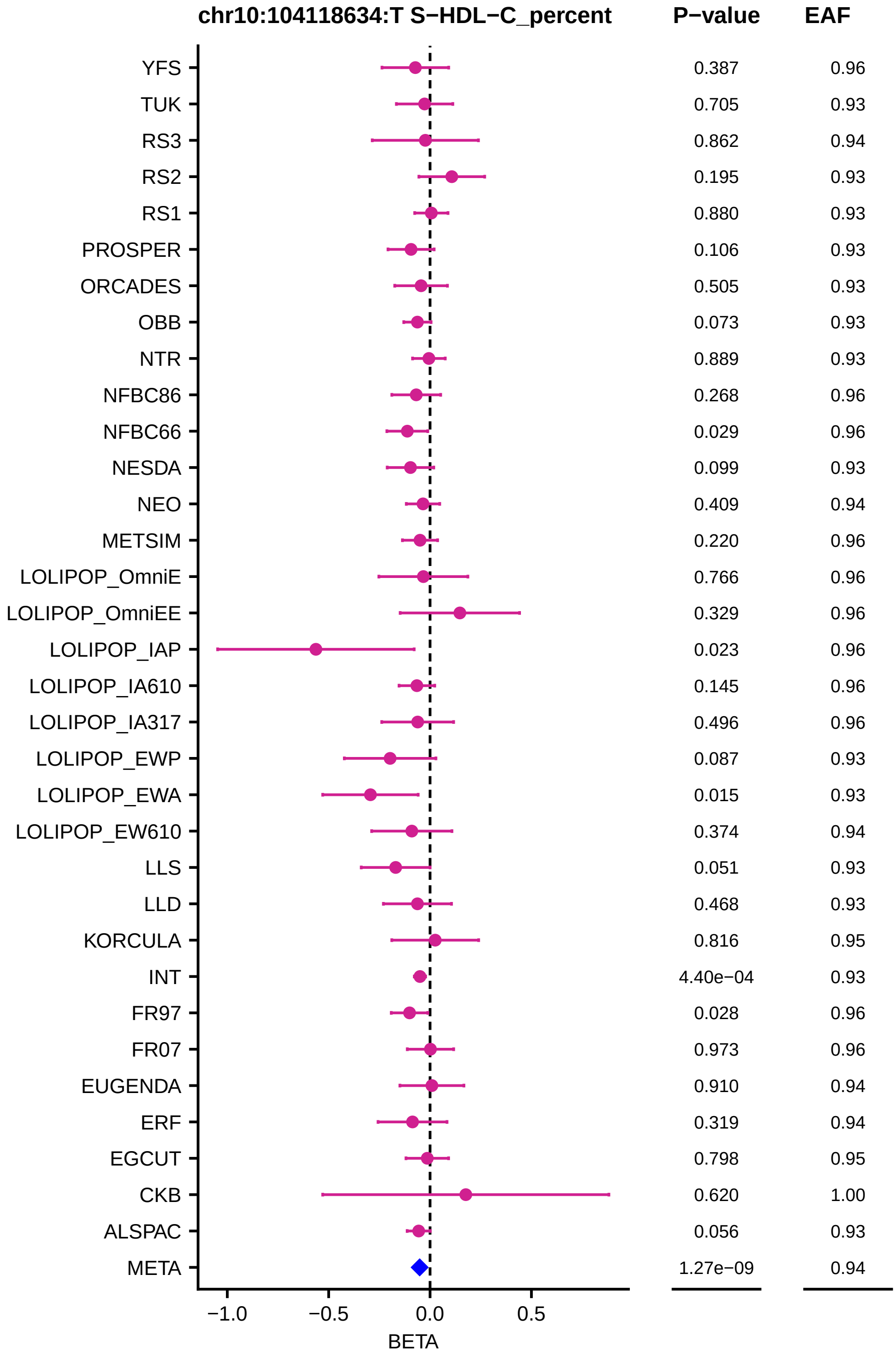
chr10:104118634:T S−HDL−C_percent
P−value
EAF
YFS 0.387 0.96
TUK 0.705 0.93
RS3 0.862 0.94
RS2 0.195 0.93
RS1 0.880 0.93
PROSPER 0.106 0.93
ORCADES 0.505 0.93
OBB 0.073 0.93
NTR 0.889 0.93
NFBC86 0.268 0.96
NFBC66 0.029 0.96
NESDA 0.099 0.93
NEO 0.409 0.94
METSIM 0.220 0.96
LOLIPOP_OmniE 0.766 0.96
LOLIPOP_OmniEE 0.329 0.96
LOLIPOP_IAP 0.023 0.96
LOLIPOP_IA610 0.145 0.96
LOLIPOP_IA317 0.496 0.96
LOLIPOP_EWP 0.087 0.93
LOLIPOP_EWA 0.015 0.93
LOLIPOP_EW610 0.374 0.94
LLS 0.051 0.93
LLD 0.468 0.93
KORCULA 0.816 0.95
INT 4.40e−04 0.93
FR97 0.028 0.96
FR07 0.973 0.96
EUGENDA 0.910 0.94
ERF 0.319 0.94
EGCUT 0.798 0.95
CKB 0.620 1.00
ALSPAC 0.056 0.93
META 1.27e−09 0.94
−1.0
−0.5
0.0
0.5
BETA

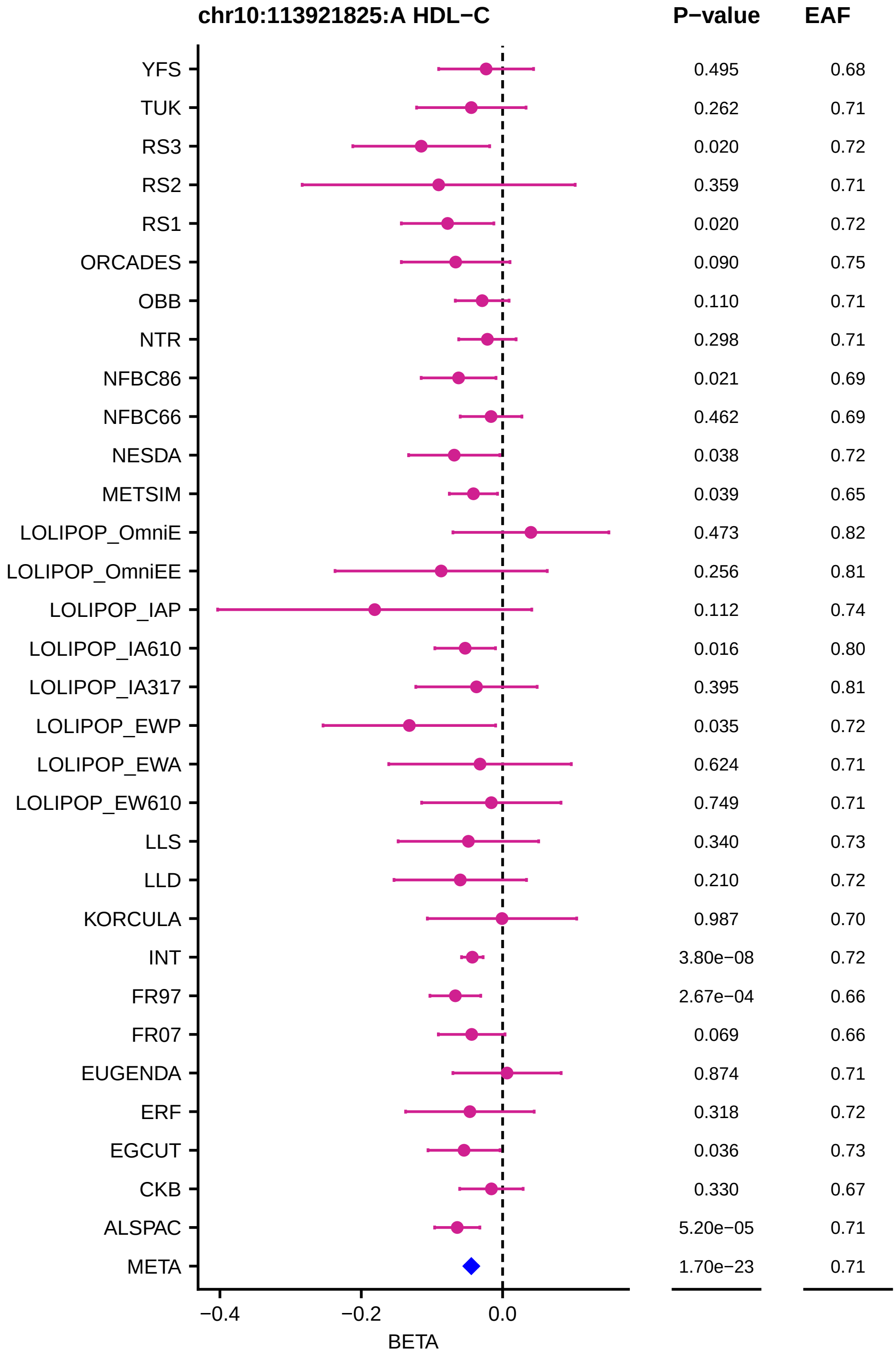
chr10:113921825:A HDL−C
P−value
EAF
YFS 0.495 0.68
TUK 0.262 0.71
RS3 0.020 0.72
RS2 0.359 0.71
RS1 0.020 0.72
ORCADES 0.090 0.75
OBB 0.110 0.71
NTR 0.298 0.71
NFBC86 0.021 0.69
NFBC66 0.462 0.69
NESDA 0.038 0.72
METSIM 0.039 0.65
LOLIPOP_OmniE 0.473 0.82
LOLIPOP_OmniEE 0.256 0.81
LOLIPOP_IAP 0.112 0.74
LOLIPOP_IA610 0.016 0.80
LOLIPOP_IA317 0.395 0.81
LOLIPOP_EWP 0.035 0.72
LOLIPOP_EWA 0.624 0.71
LOLIPOP_EW610 0.749 0.71
LLS 0.340 0.73
LLD 0.210 0.72
KORCULA 0.987 0.70
INT 3.80e−08 0.72
FR97 2.67e−04 0.66
FR07 0.069 0.66
EUGENDA 0.874 0.71
ERF 0.318 0.72
EGCUT 0.036 0.73
CKB 0.330 0.67
ALSPAC 5.20e−05 0.71
META 1.70e−23 0.71
−0.4
−0.2
0.0
BETA

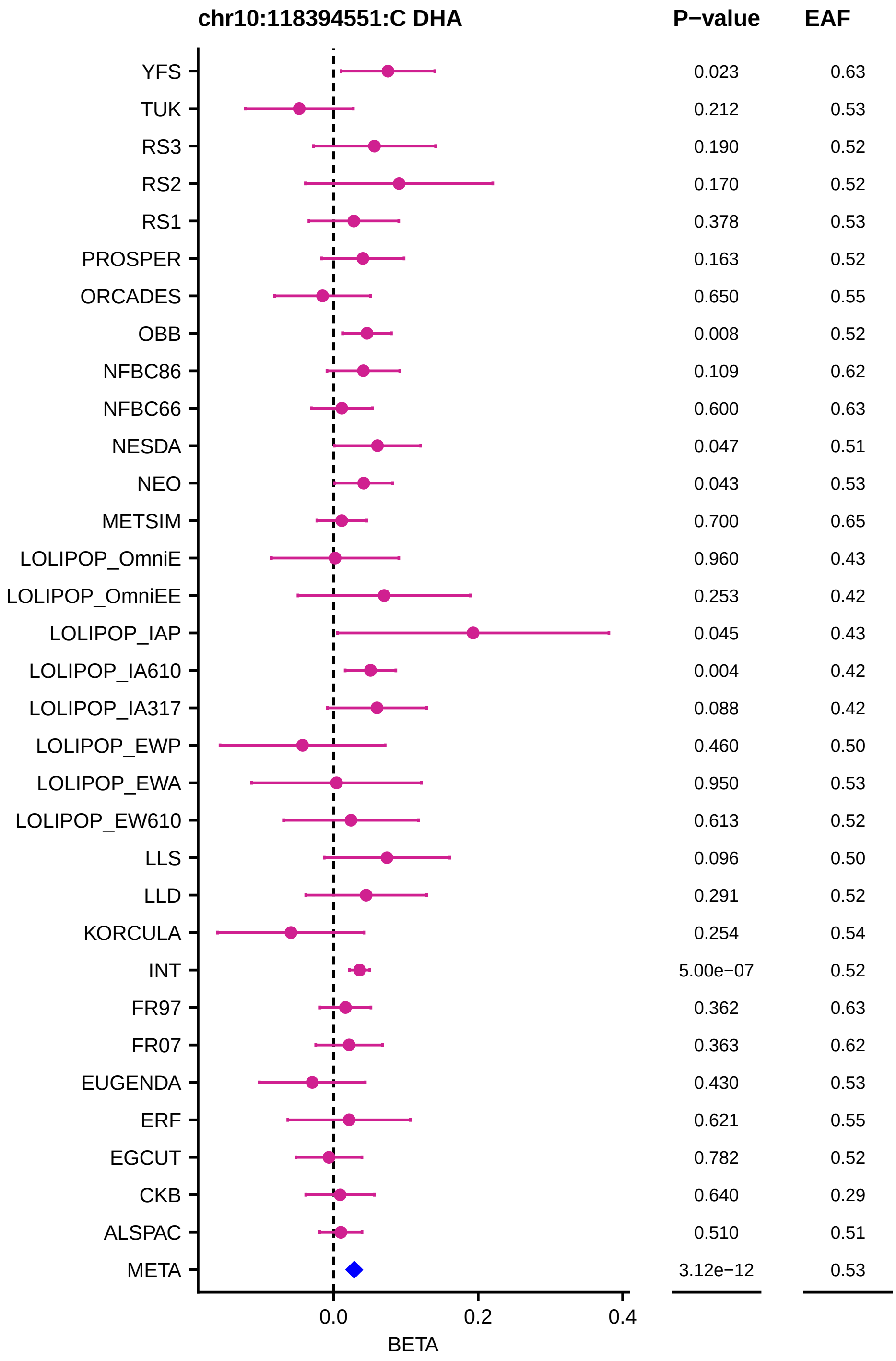
chr10:118394551:C DHA
P−value
EAF
YFS 0.023 0.63
TUK 0.212 0.53
RS3 0.190 0.52
RS2 0.170 0.52
RS1 0.378 0.53
PROSPER 0.163 0.52
ORCADES 0.650 0.55
OBB 0.008 0.52
NFBC86 0.109 0.62
NFBC66 0.600 0.63
NESDA 0.047 0.51
NEO 0.043 0.53
METSIM 0.700 0.65
LOLIPOP_OmniE 0.960 0.43
LOLIPOP_OmniEE 0.253 0.42
LOLIPOP_IAP 0.045 0.43
LOLIPOP_IA610 0.004 0.42
LOLIPOP_IA317 0.088 0.42
LOLIPOP_EWP 0.460 0.50
LOLIPOP_EWA 0.950 0.53
LOLIPOP_EW610 0.613 0.52
LLS 0.096 0.50
LLD 0.291 0.52
KORCULA 0.254 0.54
INT 5.00e−07 0.52
FR97 0.362 0.63
FR07 0.363 0.62
EUGENDA 0.430 0.53
ERF 0.621 0.55
EGCUT 0.782 0.52
CKB 0.640 0.29
ALSPAC 0.510 0.51
META 3.12e−12 0.53
0.0
0.2
0.4
BETA

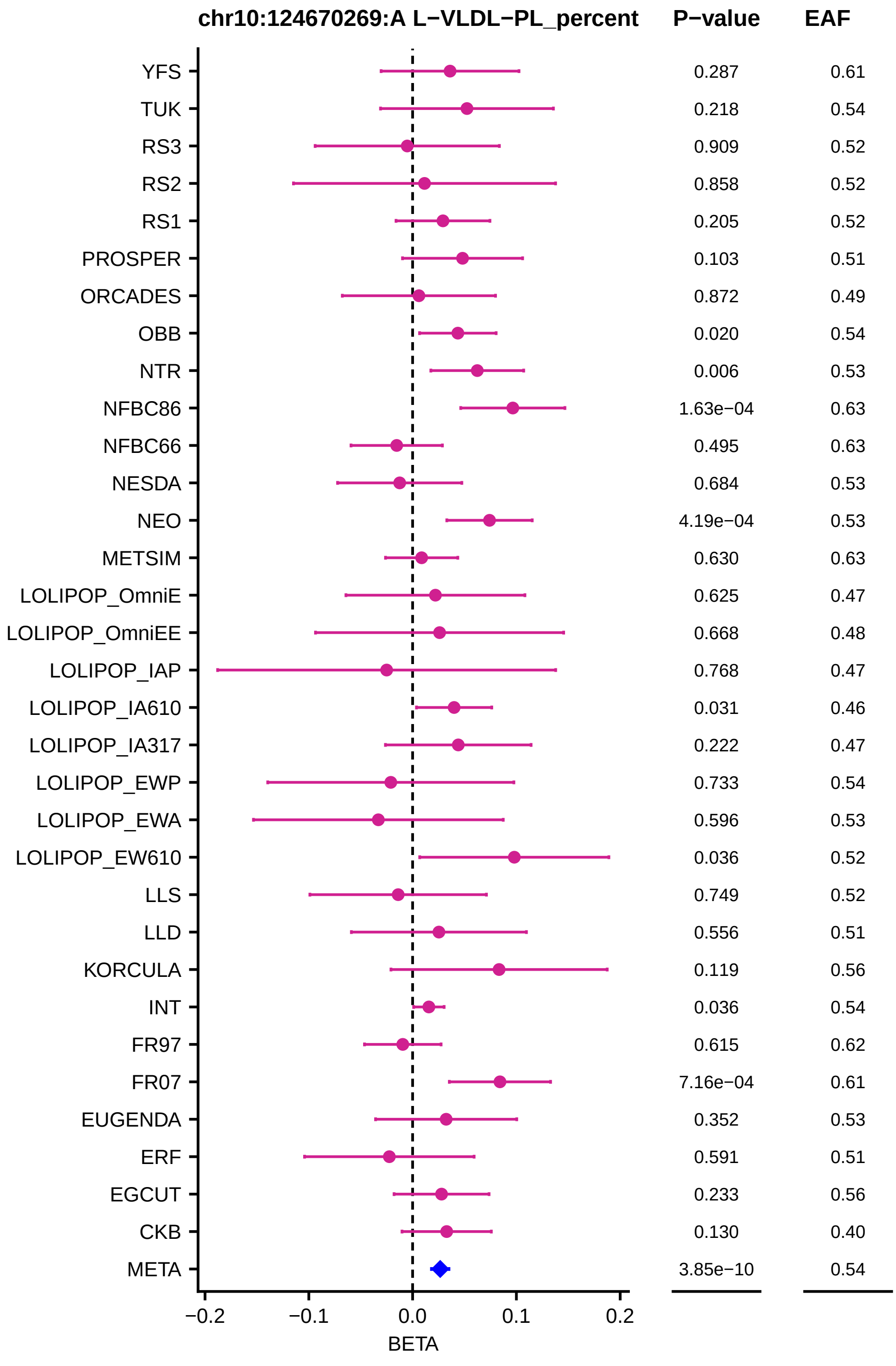
chr10:124670269:A L−VLDL−PL_percent
P−value
EAF
YFS 0.287 0.61
TUK 0.218 0.54
RS3 0.909 0.52
RS2 0.858 0.52
RS1 0.205 0.52
PROSPER 0.103 0.51
ORCADES 0.872 0.49
OBB 0.020 0.54
NTR 0.006 0.53
NFBC86 1.63e−04 0.63
NFBC66 0.495 0.63
NESDA 0.684 0.53
NEO 4.19e−04 0.53
METSIM 0.630 0.63
LOLIPOP_OmniE 0.625 0.47
LOLIPOP_OmniEE 0.668 0.48
LOLIPOP_IAP 0.768 0.47
LOLIPOP_IA610 0.031 0.46
LOLIPOP_IA317 0.222 0.47
LOLIPOP_EWP 0.733 0.54
LOLIPOP_EWA 0.596 0.53
LOLIPOP_EW610 0.036 0.52
LLS 0.749 0.52
LLD 0.556 0.51
KORCULA 0.119 0.56
INT 0.036 0.54
FR97 0.615 0.62
FR07 7.16e−04 0.61
EUGENDA 0.352 0.53
ERF 0.591 0.51
EGCUT 0.233 0.56
CKB 0.130 0.40
META 3.85e−10 0.54
−0.2 −0.1 0.0 0.1 0.2
BETA

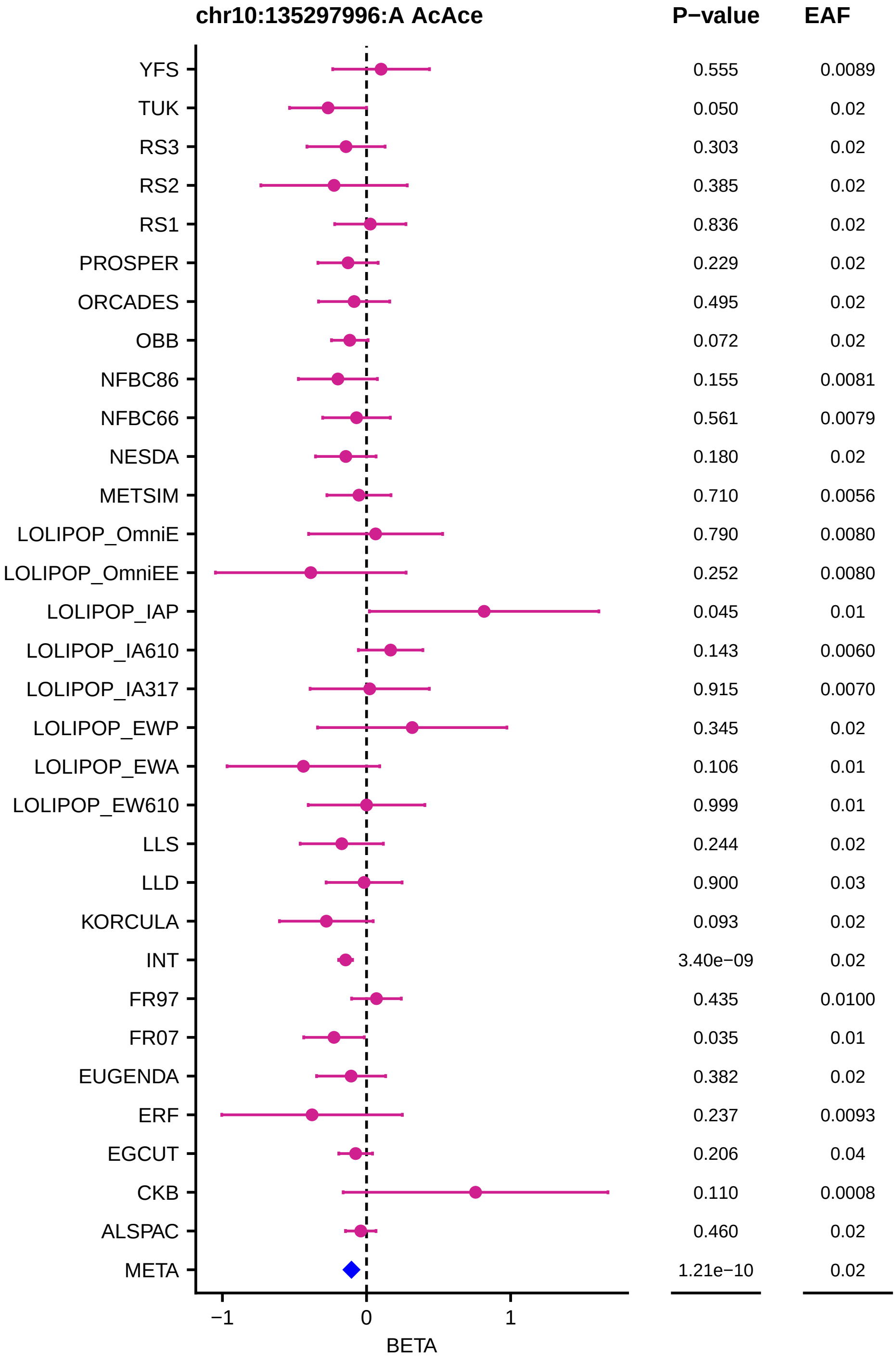
chr10:135297996:A AcAce
P−value
EAF
YFS 0.555 0.0089
TUK 0.050 0.02
RS3 0.303 0.02
RS2 0.385 0.02
RS1 0.836 0.02
PROSPER 0.229 0.02
ORCADES 0.495 0.02
OBB 0.072 0.02
NFBC86 0.155 0.0081
NFBC66 0.561 0.0079
NESDA 0.180 0.02
METSIM 0.710 0.0056
LOLIPOP_OmniE 0.790 0.0080
LOLIPOP_OmniEE 0.252 0.0080
LOLIPOP_IAP 0.045 0.01
LOLIPOP_IA610 0.143 0.0060
LOLIPOP_IA317 0.915 0.0070
LOLIPOP_EWP 0.345 0.02
LOLIPOP_EWA 0.106 0.01
LOLIPOP_EW610 0.999 0.01
LLS 0.244 0.02
LLD 0.900 0.03
KORCULA 0.093 0.02
INT 3.40e−09 0.02
FR97 0.435 0.0100
FR07 0.035 0.01
EUGENDA 0.382 0.02
ERF 0.237 0.0093
EGCUT 0.206 0.04
CKB 0.110 0.0008
ALSPAC 0.460 0.02
META 1.21e−10 0.02
−1
0
1
BETA

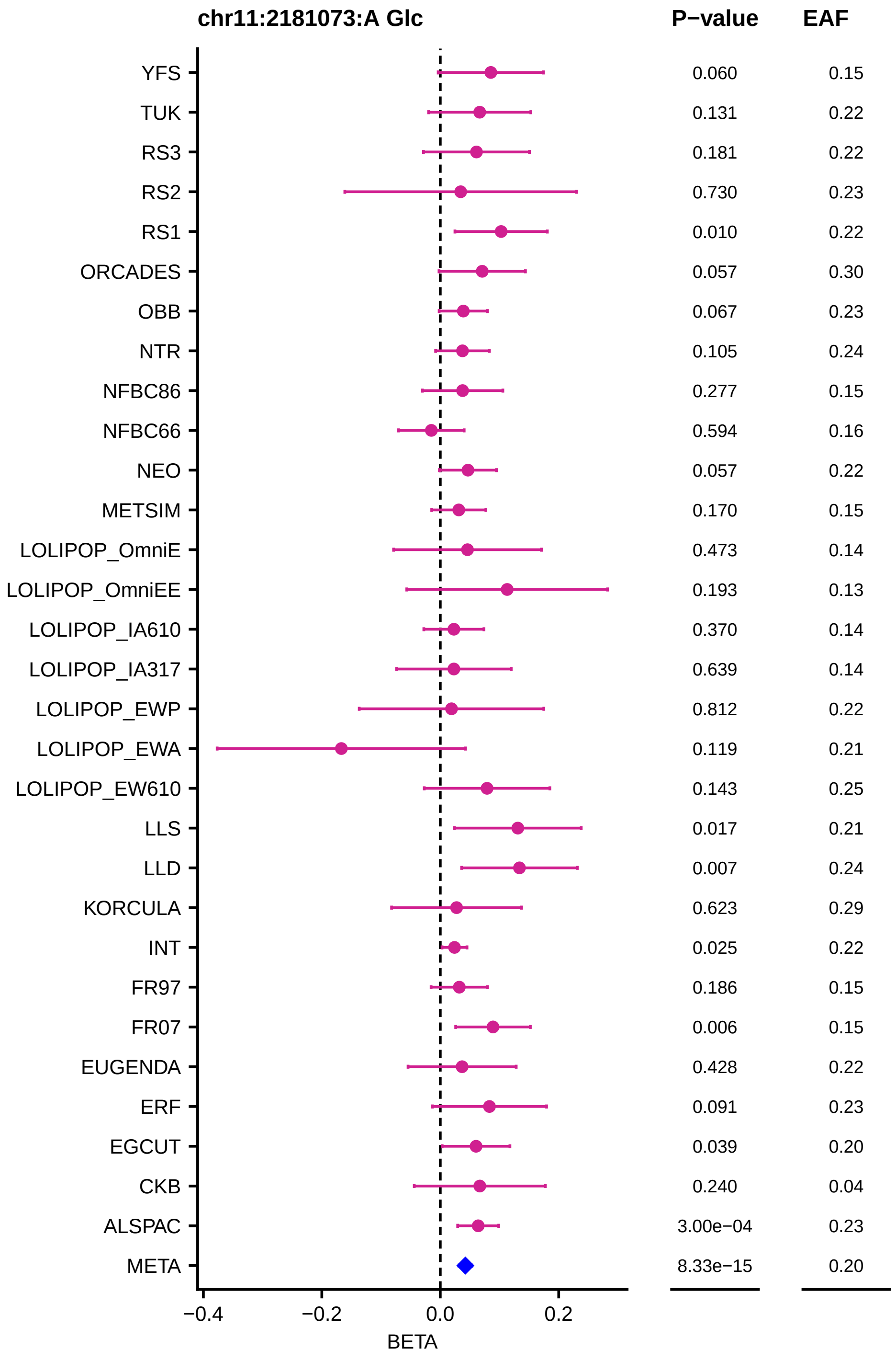
chr11:2181073:A Glc
P−value
EAF
YFS
0.060
0.15
TUK
0.131
0.22
RS3
0.181
0.22
RS2
0.730
0.23
RS1
0.010
0.22
ORCADES
0.057
0.30
OBB
0.067
0.23
NTR
0.105
0.24
NFBC86
0.277
0.15
NFBC66
0.594
0.16
NEO
0.057
0.22
METSIM
0.170
0.15
LOLIPOP_OmniE
0.473
0.14
LOLIPOP_OmniEE
0.193
0.13
LOLIPOP_IA610
0.370
0.14
LOLIPOP_IA317
0.639
0.14
LOLIPOP_EWP
0.812
0.22
LOLIPOP_EWA
0.119
0.21
LOLIPOP_EW610
0.143
0.25
LLS
0.017
0.21
LLD
0.007
0.24
KORCULA
0.623
0.29
INT
0.025
0.22
FR97
0.186
0.15
FR07
0.006
0.15
EUGENDA
0.428
0.22
ERF
0.091
0.23
EGCUT
0.039
0.20
CKB
0.240
0.04
ALSPAC
3.00e−04
0.23
META
8.33e−15
0.20
−0.4
−0.2
0.0
0.2
BETA

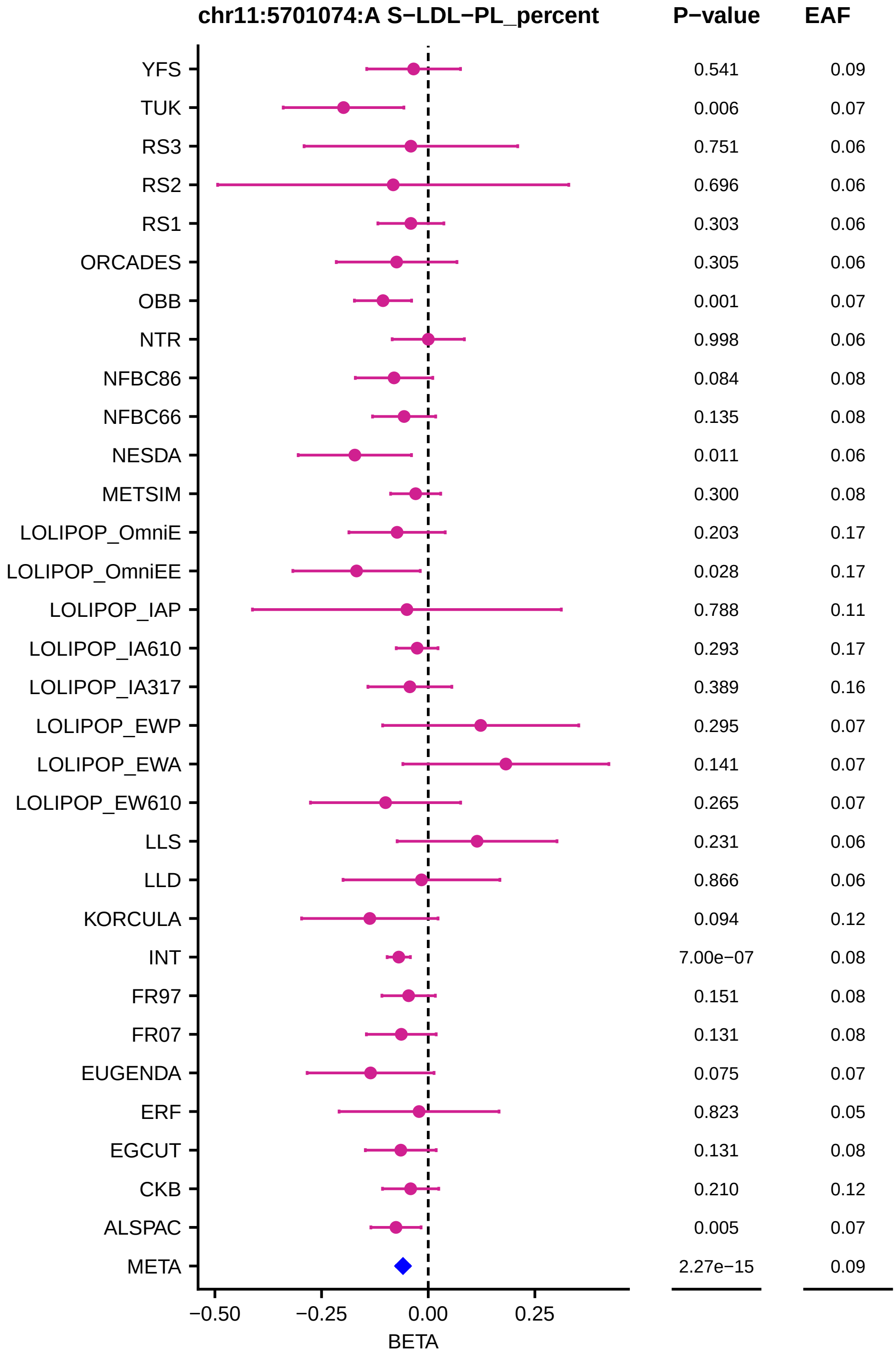

chr11:5701074:A S−LDL−PL_percent
P−value
EAF
YFS 0.541 0.09
TUK 0.006 0.07
RS3 0.751 0.06
RS2 0.696 0.06
RS1 0.303 0.06
ORCADES 0.305 0.06
OBB 0.001 0.07
NTR 0.998 0.06
NFBC86 0.084 0.08
NFBC66 0.135 0.08
NESDA 0.011 0.06
METSIM 0.300 0.08
LOLIPOP_OmniE 0.203 0.17
LOLIPOP_OmniEE 0.028 0.17
LOLIPOP_IAP 0.788 0.11
LOLIPOP_IA610 0.293 0.17
LOLIPOP_IA317 0.389 0.16
LOLIPOP_EWP 0.295 0.07
LOLIPOP_EWA 0.141 0.07
LOLIPOP_EW610 0.265 0.07
LLS 0.231 0.06
LLD 0.866 0.06
KORCULA 0.094 0.12
INT 7.00e−07 0.08
FR97 0.151 0.08
FR07 0.131 0.08
EUGENDA 0.075 0.07
ERF 0.823 0.05
EGCUT 0.131 0.08
CKB 0.210 0.12
ALSPAC 0.005 0.07
META 2.27e−15 0.09
−0.50
−0.25
0.00
0.25
BETA

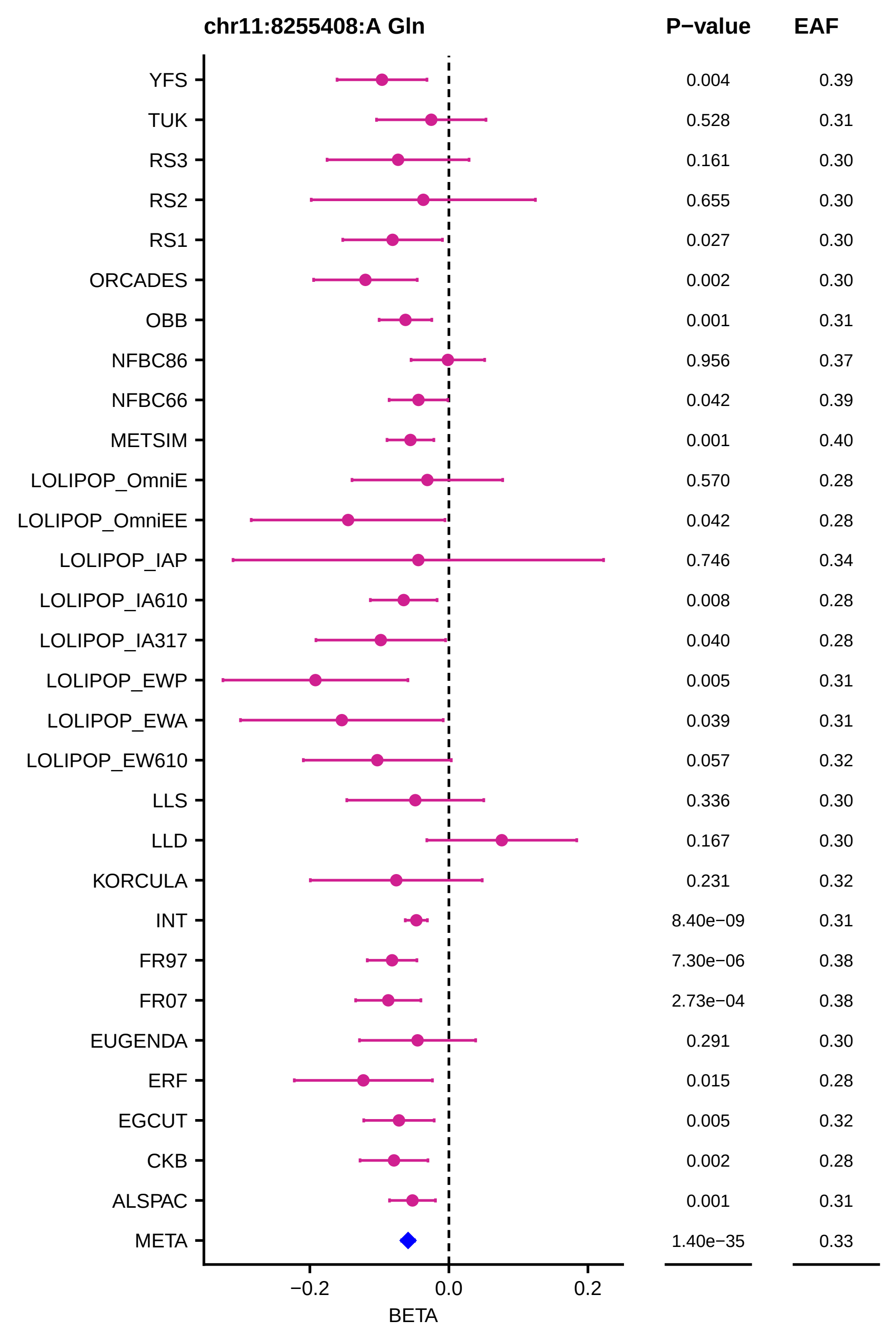
chr11:8255408:A Gln
P−value
EAF
YFS 0.004 0.39
TUK 0.528 0.31
RS3 0.161 0.30
RS2 0.655 0.30
RS1 0.027 0.30
ORCADES 0.002 0.30
OBB 0.001 0.31
NFBC86 0.956 0.37
NFBC66 0.042 0.39
METSIM 0.001 0.40
LOLIPOP_OmniE 0.570 0.28
LOLIPOP_OmniEE 0.042 0.28
LOLIPOP_IAP 0.746 0.34
LOLIPOP_IA610 0.008 0.28
LOLIPOP_IA317 0.040 0.28
LOLIPOP_EWP 0.005 0.31
LOLIPOP_EWA 0.039 0.31
LOLIPOP_EW610 0.057 0.32
LLS 0.336 0.30
LLD 0.167 0.30
KORCULA 0.231 0.32
INT 8.40e−09 0.31
FR97 7.30e−06 0.38
FR07 2.73e−04 0.38
EUGENDA 0.291 0.30
ERF 0.015 0.28
EGCUT 0.005 0.32
CKB 0.002 0.28
ALSPAC 0.001 0.31
META 1.40e−35 0.33
−0.2
0.0
0.2
BETA

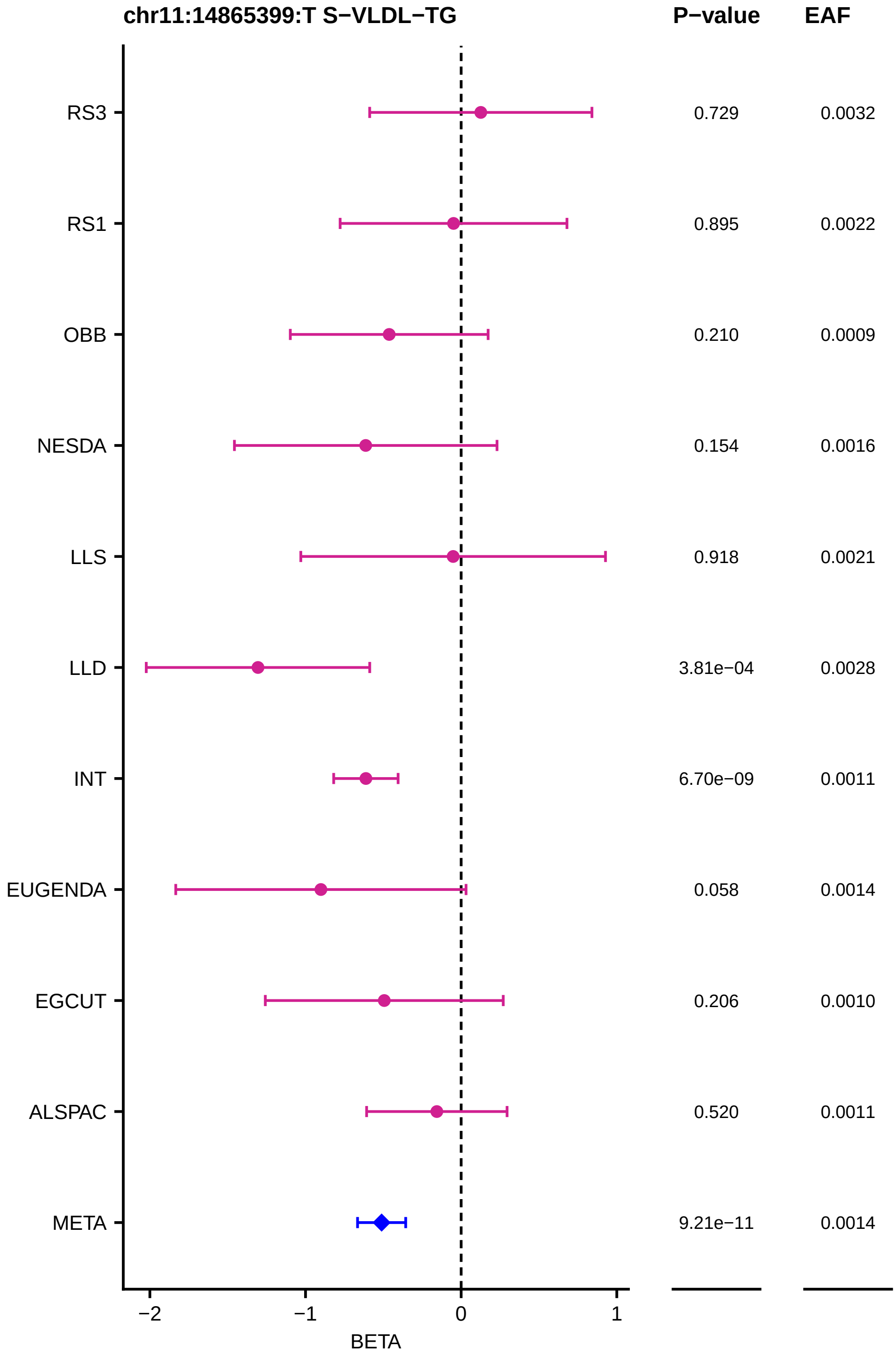
chr11:14865399:T S−VLDL−TG
P−value
EAF
RS3
0.729
0.0032
RS1
0.895
0.0022
OBB
0.210
0.0009
NESDA
0.154
0.0016
LLS
0.918
0.0021
LLD
3.81e−04
0.0028
INT
6.70e−09
0.0011
EUGENDA
0.058
0.0014
EGCUT
0.206
0.0010
ALSPAC
0.520
0.0011
META
9.21e−11
0.0014
−2
−1
0
1
BETA

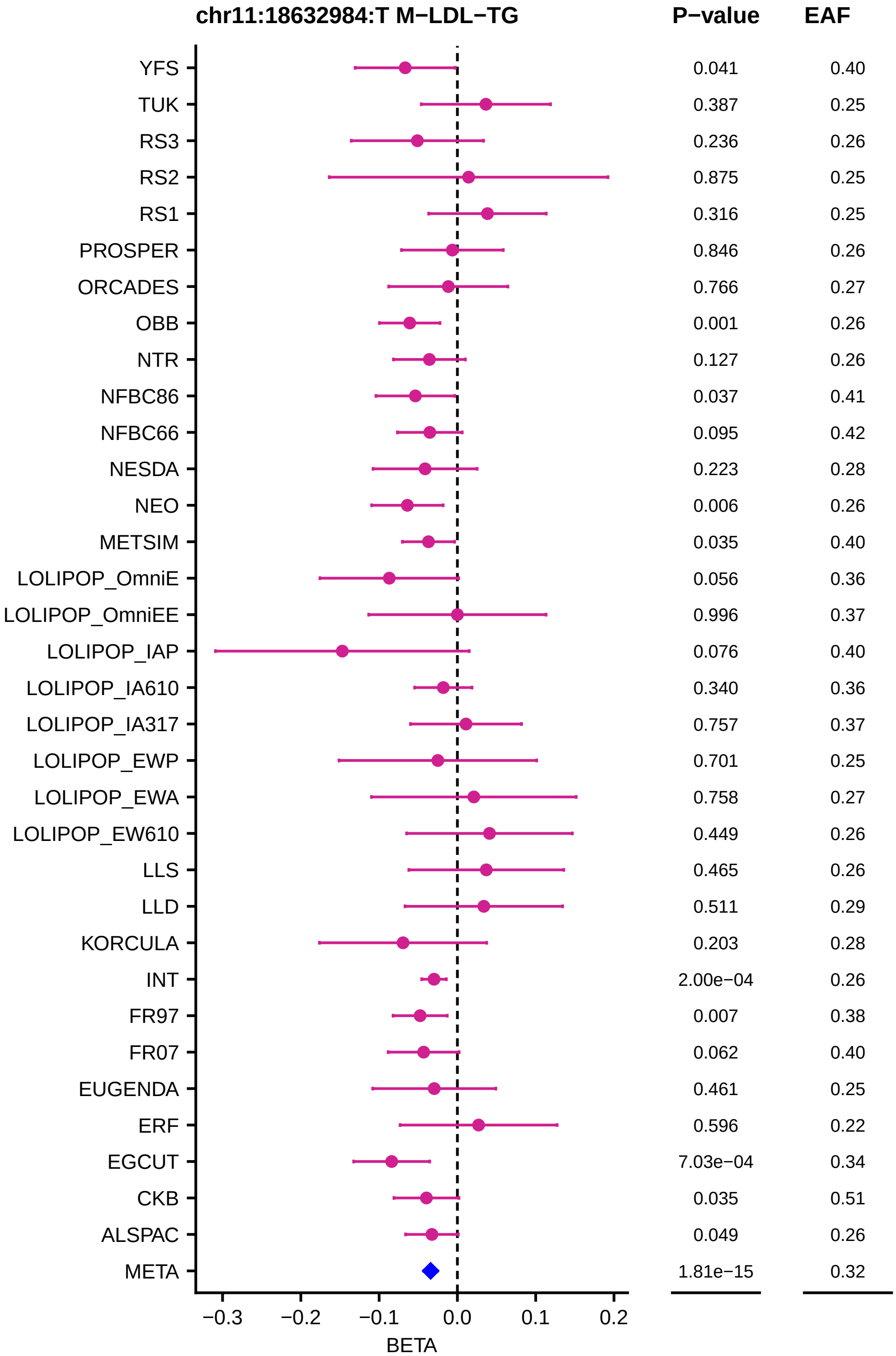

chr11:18632984:T M−LDL−TG
P−value
EAF
YFS 0.041 0.40
TUK 0.387 0.25
RS3 0.236 0.26
RS2 0.875 0.25
RS1 0.316 0.25
PROSPER 0.846 0.26
ORCADES 0.766 0.27
OBB 0.001 0.26
NTR 0.127 0.26
NFBC86 0.037 0.41
NFBC66 0.095 0.42
NESDA 0.223 0.28
NEO 0.006 0.26
METSIM 0.035 0.40
LOLIPOP_OmniE 0.056 0.36
LOLIPOP_OmniEE 0.996 0.37
LOLIPOP_IAP 0.076 0.40
LOLIPOP_IA610 0.340 0.36
LOLIPOP_IA317 0.757 0.37
LOLIPOP_EWP 0.701 0.25
LOLIPOP_EWA 0.758 0.27
LOLIPOP_EW610 0.449 0.26
LLS 0.465 0.26
LLD 0.511 0.29
KORCULA 0.203 0.28
INT 2.00e−04 0.26
FR97 0.007 0.38
FR07 0.062 0.40
EUGENDA 0.461 0.25
ERF 0.596 0.22
EGCUT 7.03e−04 0.34
CKB 0.035 0.51
ALSPAC 0.049 0.26
META 1.81e−15 0.32
−0.3 −0.2 −0.1 0.0 0.1 0.2
BETA

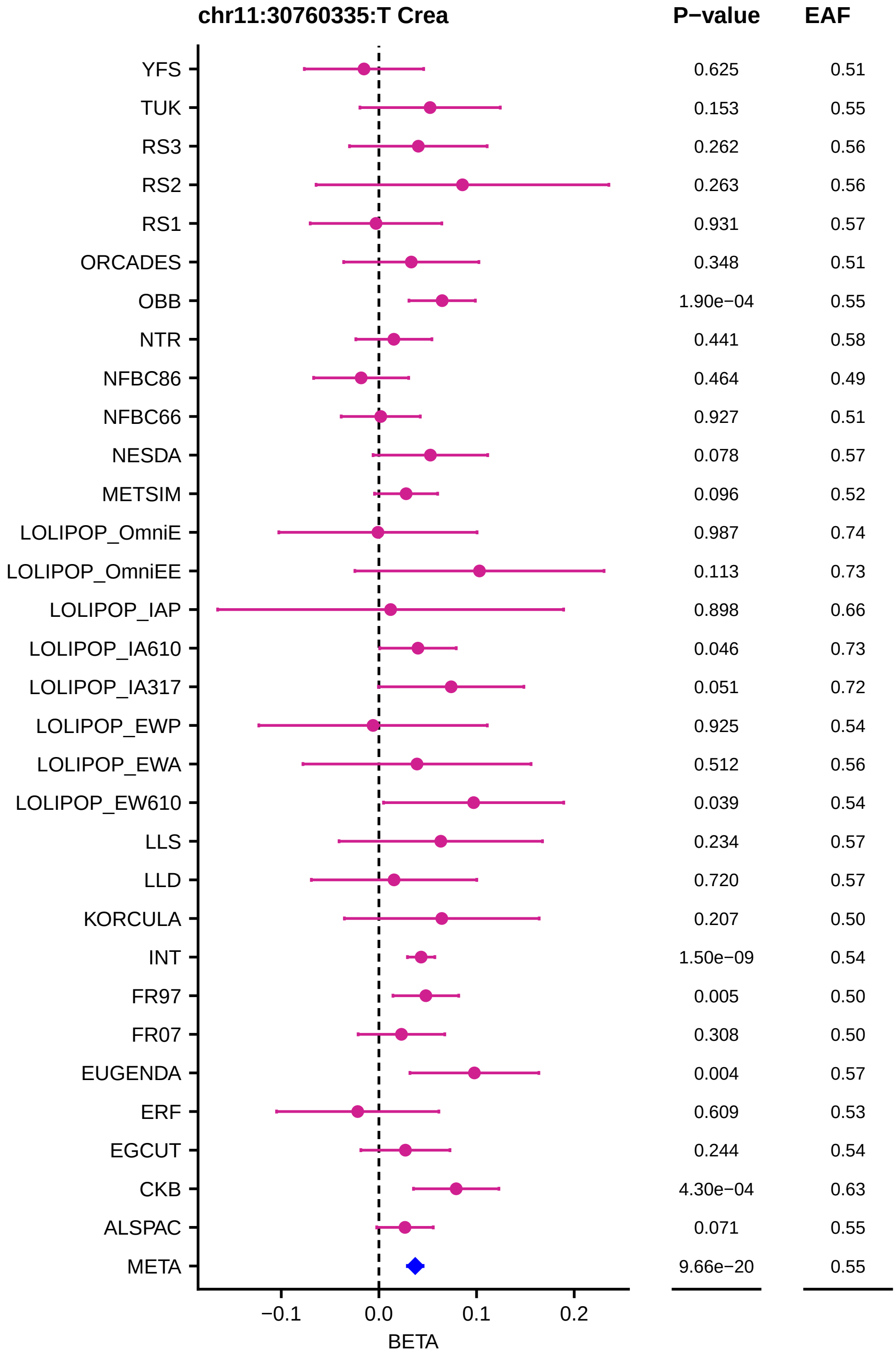

chr11:30760335:T Crea
P−value
EAF
YFS 0.625 0.51
TUK 0.153 0.55
RS3 0.262 0.56
RS2 0.263 0.56
RS1 0.931 0.57
ORCADES 0.348 0.51
OBB 1.90e−04 0.55
NTR 0.441 0.58
NFBC86 0.464 0.49
NFBC66 0.927 0.51
NESDA 0.078 0.57
METSIM 0.096 0.52
LOLIPOP_OmniE 0.987 0.74
LOLIPOP_OmniEE 0.113 0.73
LOLIPOP_IAP 0.898 0.66
LOLIPOP_IA610 0.046 0.73
LOLIPOP_IA317 0.051 0.72
LOLIPOP_EWP 0.925 0.54
LOLIPOP_EWA 0.512 0.56
LOLIPOP_EW610 0.039 0.54
LLS 0.234 0.57
LLD 0.720 0.57
KORCULA 0.207 0.50
INT 1.50e−09 0.54
FR97 0.005 0.50
FR07 0.308 0.50
EUGENDA 0.004 0.57
ERF 0.609 0.53
EGCUT 0.244 0.54
CKB 4.30e−04 0.63
ALSPAC 0.071 0.55
META 9.66e−20 0.55
−0.1
0.0
0.1
0.2
BETA

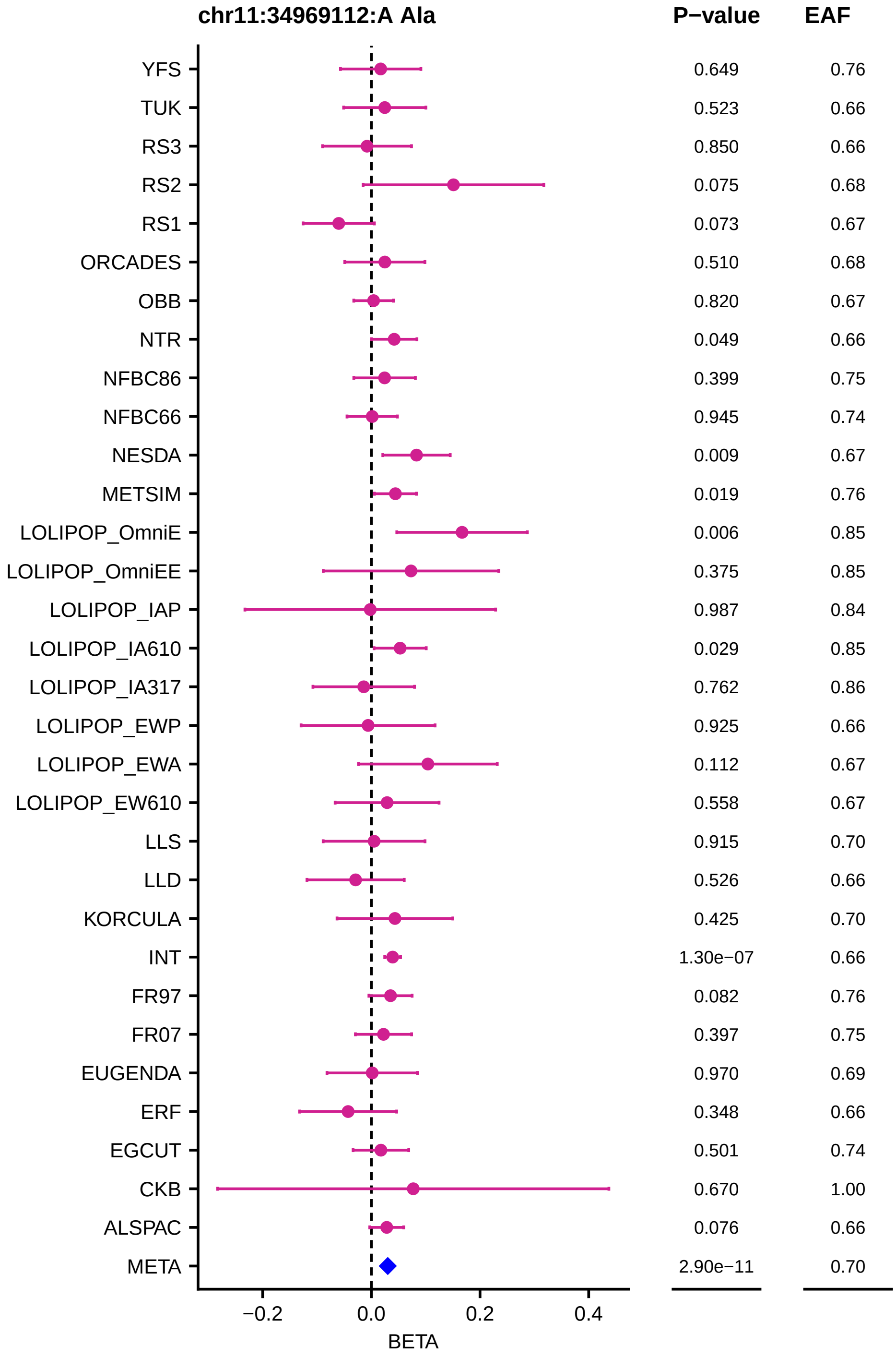
chr11:34969112:A Ala
P−value
EAF
YFS 0.649 0.76
TUK 0.523 0.66
RS3 0.850 0.66
RS2 0.075 0.68
RS1 0.073 0.67
ORCADES 0.510 0.68
OBB 0.820 0.67
NTR 0.049 0.66
NFBC86 0.399 0.75
NFBC66 0.945 0.74
NESDA 0.009 0.67
METSIM 0.019 0.76
LOLIPOP_OmniE 0.006 0.85
LOLIPOP_OmniEE 0.375 0.85
LOLIPOP_IAP 0.987 0.84
LOLIPOP_IA610 0.029 0.85
LOLIPOP_IA317 0.762 0.86
LOLIPOP_EWP 0.925 0.66
LOLIPOP_EWA 0.112 0.67
LOLIPOP_EW610 0.558 0.67
LLS 0.915 0.70
LLD 0.526 0.66
KORCULA 0.425 0.70
INT 1.30e−07 0.66
FR97 0.082 0.76
FR07 0.397 0.75
EUGENDA 0.970 0.69
ERF 0.348 0.66
EGCUT 0.501 0.74
CKB 0.670 1.00
ALSPAC 0.076 0.66
META 2.90e−11 0.70
−0.2
0.0
0.2
0.4
BETA

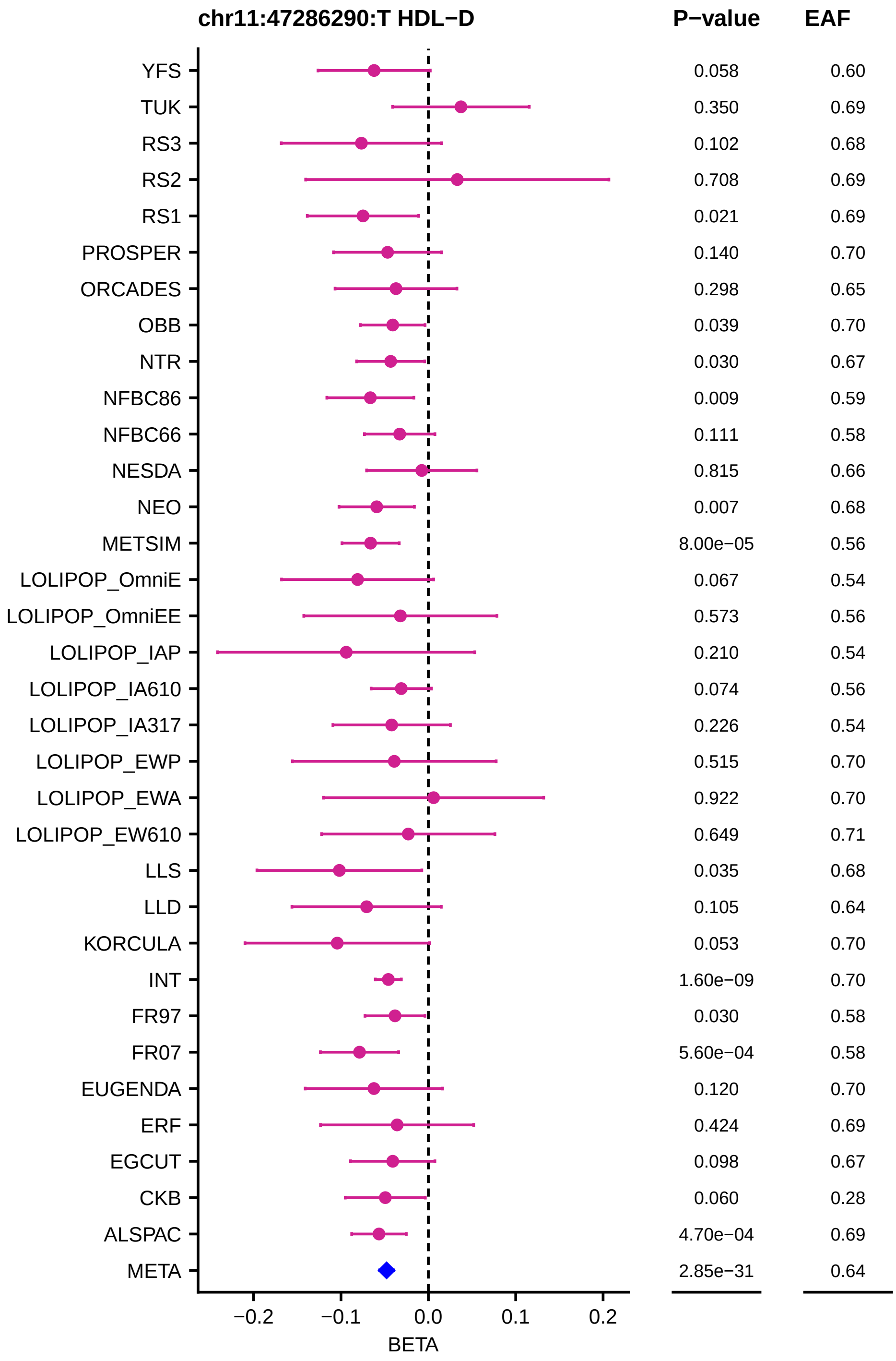

chr11:47286290:T HDL−D
P−value
EAF
YFS 0.058 0.60
TUK 0.350 0.69
RS3 0.102 0.68
RS2 0.708 0.69
RS1 0.021 0.69
PROSPER 0.140 0.70
ORCADES 0.298 0.65
OBB 0.039 0.70
NTR 0.030 0.67
NFBC86 0.009 0.59
NFBC66 0.111 0.58
NESDA 0.815 0.66
NEO 0.007 0.68
METSIM 8.00e−05 0.56
LOLIPOP_OmniE 0.067 0.54
LOLIPOP_OmniEE 0.573 0.56
LOLIPOP_IAP 0.210 0.54
LOLIPOP_IA610 0.074 0.56
LOLIPOP_IA317 0.226 0.54
LOLIPOP_EWP 0.515 0.70
LOLIPOP_EWA 0.922 0.70
LOLIPOP_EW610 0.649 0.71
LLS 0.035 0.68
LLD 0.105 0.64
KORCULA 0.053 0.70
INT 1.60e−09 0.70
FR97 0.030 0.58
FR07 5.60e−04 0.58
EUGENDA 0.120 0.70
ERF 0.424 0.69
EGCUT 0.098 0.67
CKB 0.060 0.28
ALSPAC 4.70e−04 0.69
META 2.85e−31 0.64
−0.2
−0.1
0.0
0.1
0.2
BETA

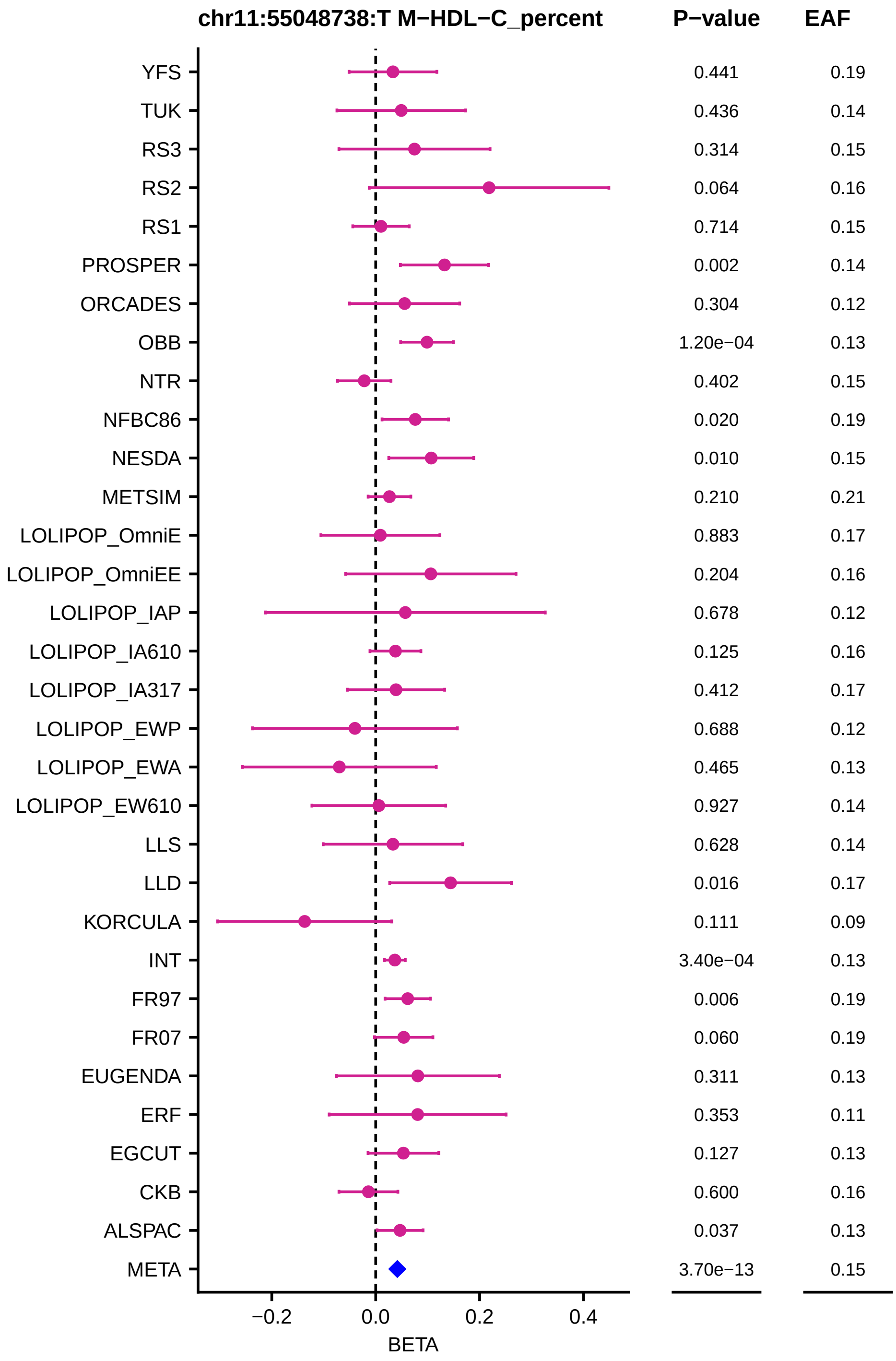
chr11:55048738:T M−HDL−C_percent
P−value
EAF
YFS 0.441 0.19
TUK 0.436 0.14
RS3 0.314 0.15
RS2 0.064 0.16
RS1 0.714 0.15
PROSPER 0.002 0.14
ORCADES 0.304 0.12
OBB 1.20e−04 0.13
NTR 0.402 0.15
NFBC86 0.020 0.19
NESDA 0.010 0.15
METSIM 0.210 0.21
LOLIPOP_OmniE 0.883 0.17
LOLIPOP_OmniEE 0.204 0.16
LOLIPOP_IAP 0.678 0.12
LOLIPOP_IA610 0.125 0.16
LOLIPOP_IA317 0.412 0.17
LOLIPOP_EWP 0.688 0.12
LOLIPOP_EWA 0.465 0.13
LOLIPOP_EW610 0.927 0.14
LLS 0.628 0.14
LLD 0.016 0.17
KORCULA 0.111 0.09
INT 3.40e−04 0.13
FR97 0.006 0.19
FR07 0.060 0.19
EUGENDA 0.311 0.13
ERF 0.353 0.11
EGCUT 0.127 0.13
CKB 0.600 0.16
ALSPAC 0.037 0.13
META 3.70e−13 0.15
−0.2
0.0
0.2
0.4
BETA

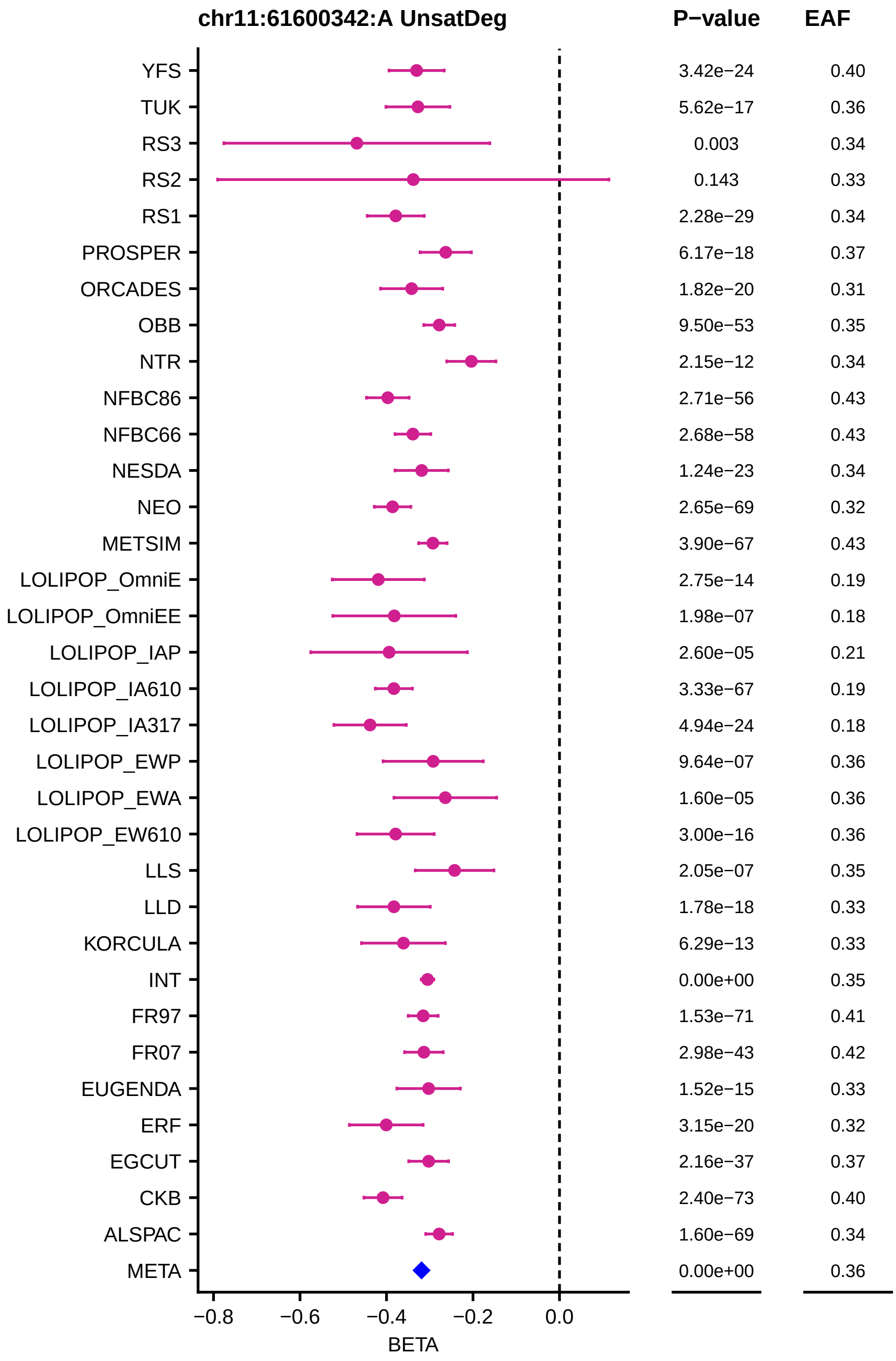
chr11:61600342:A UnsatDeg
P−value
EAF
YFS 3.42e−24 0.40
TUK 5.62e−17 0.36
RS3 0.003 0.34
RS2 0.143 0.33
RS1 2.28e−29 0.34
PROSPER 6.17e−18 0.37
ORCADES 1.82e−20 0.31
OBB 9.50e−53 0.35
NTR 2.15e−12 0.34
NFBC86 2.71e−56 0.43
NFBC66 2.68e−58 0.43
NESDA 1.24e−23 0.34
NEO 2.65e−69 0.32
METSIM 3.90e−67 0.43
LOLIPOP_OmniE 2.75e−14 0.19
LOLIPOP_OmniEE 1.98e−07 0.18
LOLIPOP_IAP 2.60e−05 0.21
LOLIPOP_IA610 3.33e−67 0.19
LOLIPOP_IA317 4.94e−24 0.18
LOLIPOP_EWP 9.64e−07 0.36
LOLIPOP_EWA 1.60e−05 0.36
LOLIPOP_EW610 3.00e−16 0.36
LLS 2.05e−07 0.35
LLD 1.78e−18 0.33
KORCULA 6.29e−13 0.33
INT 0.00e+00 0.35
FR97 1.53e−71 0.41
FR07 2.98e−43 0.42
EUGENDA 1.52e−15 0.33
ERF 3.15e−20 0.32
EGCUT 2.16e−37 0.37
CKB 2.40e−73 0.40
ALSPAC 1.60e−69 0.34
META 0.00e+00 0.36
−0.8
−0.6
−0.4
−0.2
0.0
BETA

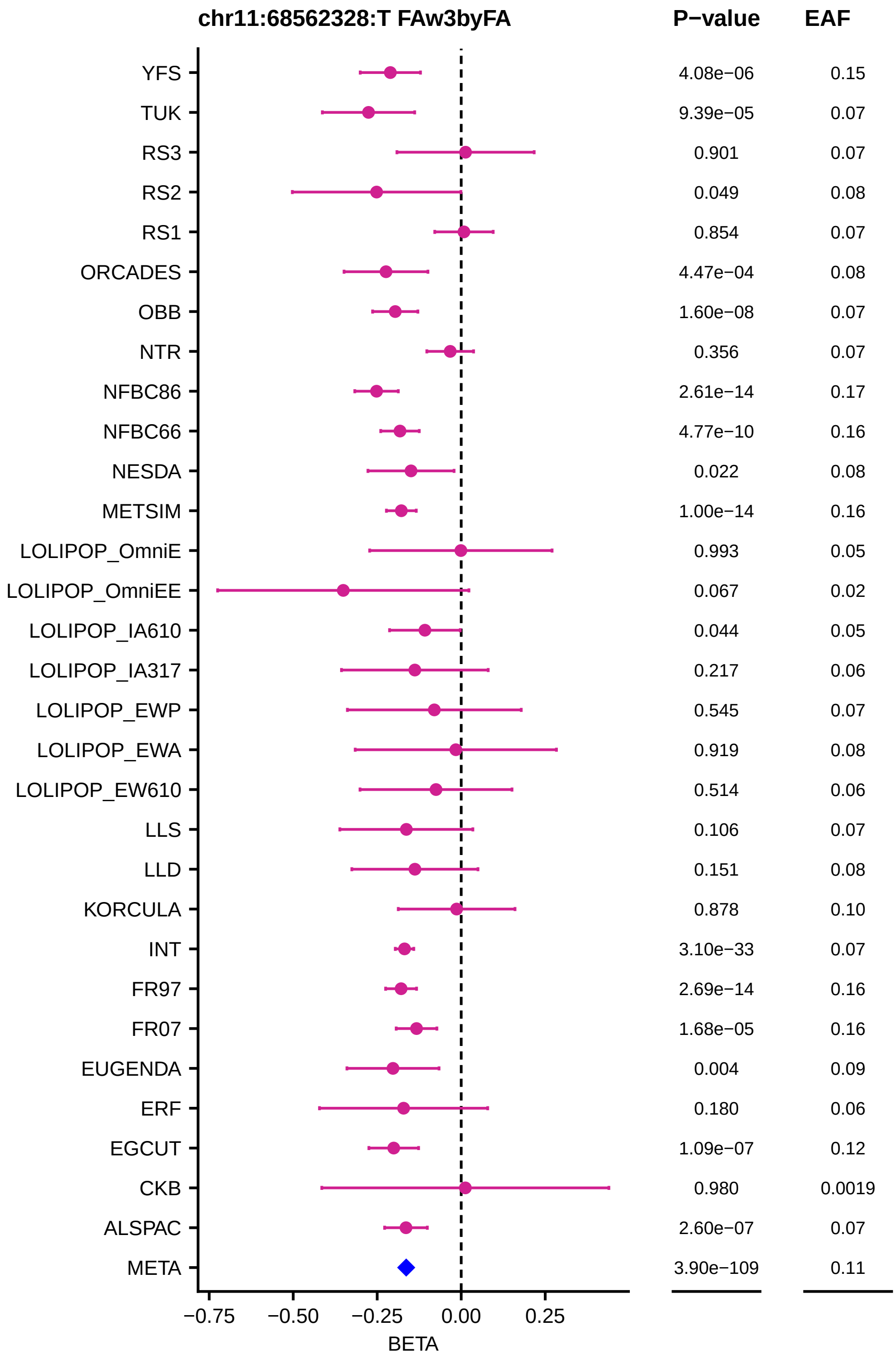

chr11:68562328:T FAw3byFA
P−value
EAF
YFS 4.08e−06 0.15
TUK 9.39e−05 0.07
RS3 0.901 0.07
RS2 0.049 0.08
RS1 0.854 0.07
ORCADES 4.47e−04 0.08
OBB 1.60e−08 0.07
NTR 0.356 0.07
NFBC86 2.61e−14 0.17
NFBC66 4.77e−10 0.16
NESDA 0.022 0.08
METSIM 1.00e−14 0.16
LOLIPOP_OmniE 0.993 0.05
LOLIPOP_OmniEE 0.067 0.02
LOLIPOP_IA610 0.044 0.05
LOLIPOP_IA317 0.217 0.06
LOLIPOP_EWP 0.545 0.07
LOLIPOP_EWA 0.919 0.08
LOLIPOP_EW610 0.514 0.06
LLS 0.106 0.07
LLD 0.151 0.08
KORCULA 0.878 0.10
INT 3.10e−33 0.07
FR97 2.69e−14 0.16
FR07 1.68e−05 0.16
EUGENDA 0.004 0.09
ERF 0.180 0.06
EGCUT 1.09e−07 0.12
CKB 0.980 0.0019
ALSPAC 2.60e−07 0.07
META 3.90e−109 0.11
−0.75
−0.50
−0.25
0.00
0.25
BETA

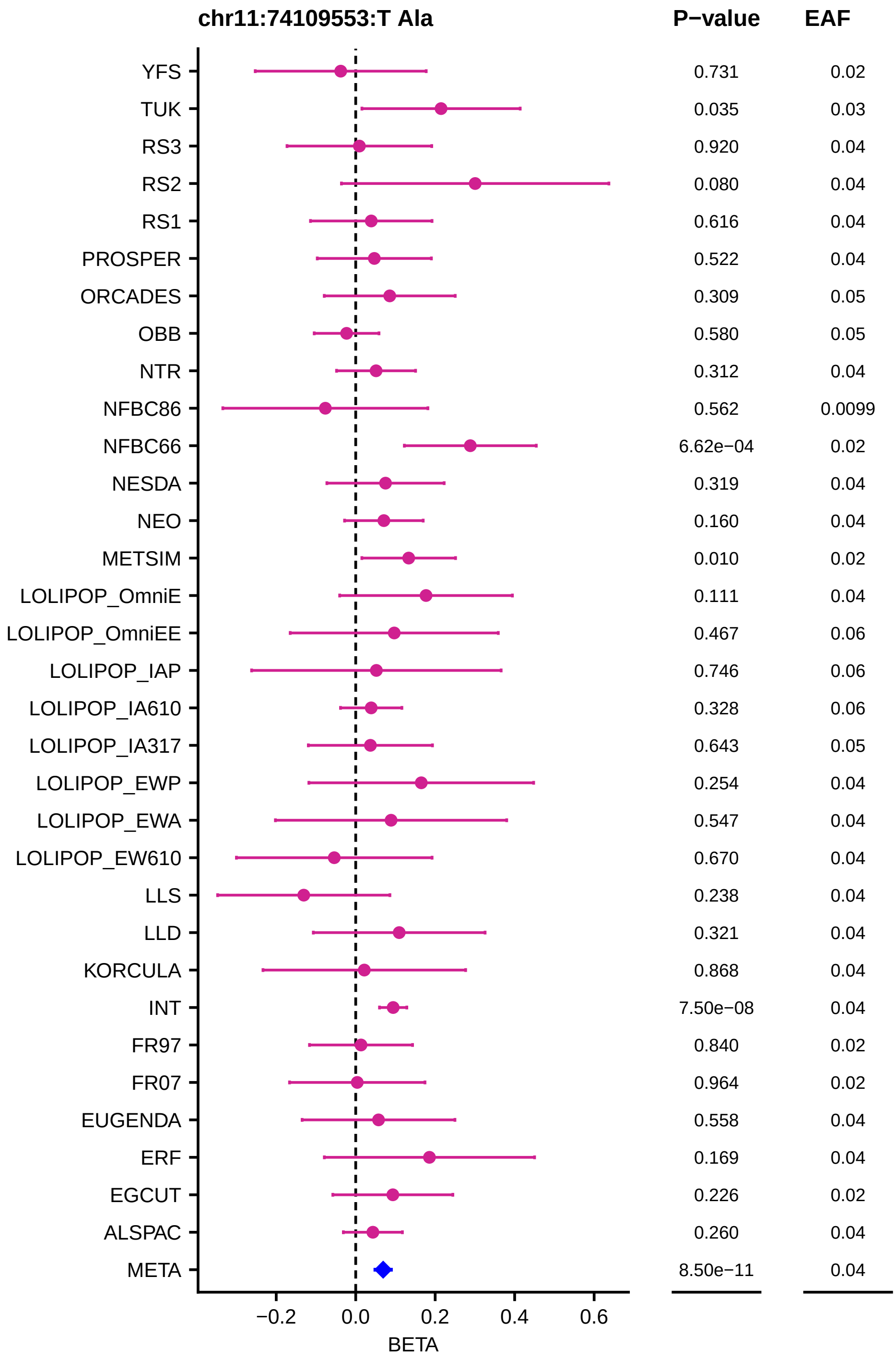
chr11:74109553:T Ala
P−value
EAF
YFS 0.731 0.02
TUK 0.035 0.03
RS3 0.920 0.04
RS2 0.080 0.04
RS1 0.616 0.04
PROSPER 0.522 0.04
ORCADES 0.309 0.05
OBB 0.580 0.05
NTR 0.312 0.04
NFBC86 0.562 0.0099
NFBC66 6.62e−04 0.02
NESDA 0.319 0.04
NEO 0.160 0.04
METSIM 0.010 0.02
LOLIPOP_OmniE 0.111 0.04
LOLIPOP_OmniEE 0.467 0.06
LOLIPOP_IAP 0.746 0.06
LOLIPOP_IA610 0.328 0.06
LOLIPOP_IA317 0.643 0.05
LOLIPOP_EWP 0.254 0.04
LOLIPOP_EWA 0.547 0.04
LOLIPOP_EW610 0.670 0.04
LLS 0.238 0.04
LLD 0.321 0.04
KORCULA 0.868 0.04
INT 7.50e−08 0.04
FR97 0.840 0.02
FR07 0.964 0.02
EUGENDA 0.558 0.04
ERF 0.169 0.04
EGCUT 0.226 0.02
ALSPAC 0.260 0.04
META 8.50e−11 0.04
−0.2
0.0
0.2
0.4
0.6
BETA

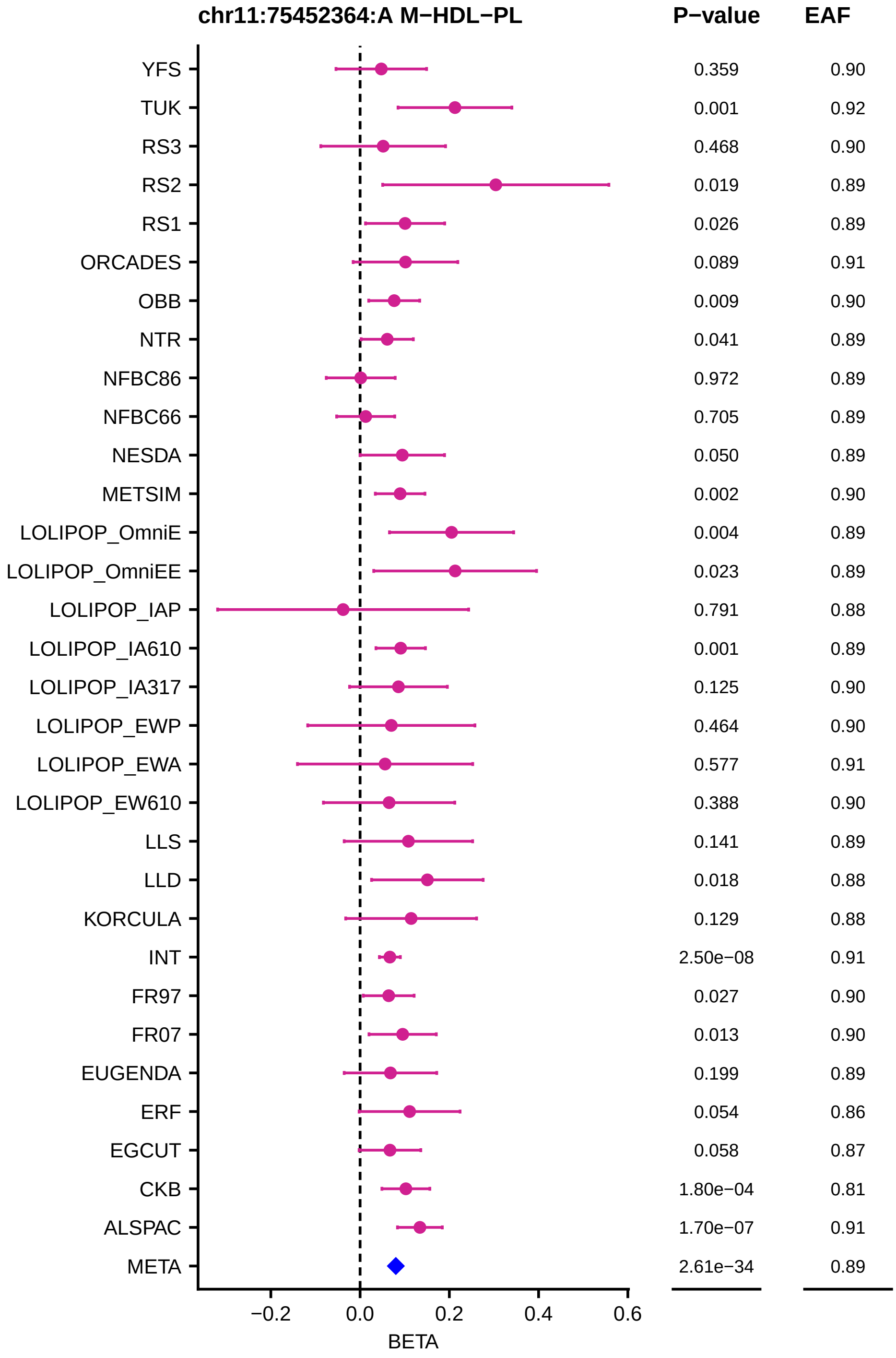
chr11:75452364:A M−HDL−PL
P−value
EAF
YFS 0.359 0.90
TUK 0.001 0.92
RS3 0.468 0.90
RS2 0.019 0.89
RS1 0.026 0.89
ORCADES 0.089 0.91
OBB 0.009 0.90
NTR 0.041 0.89
NFBC86 0.972 0.89
NFBC66 0.705 0.89
NESDA 0.050 0.89
METSIM 0.002 0.90
LOLIPOP_OmniE 0.004 0.89
LOLIPOP_OmniEE 0.023 0.89
LOLIPOP_IAP 0.791 0.88
LOLIPOP_IA610 0.001 0.89
LOLIPOP_IA317 0.125 0.90
LOLIPOP_EWP 0.464 0.90
LOLIPOP_EWA 0.577 0.91
LOLIPOP_EW610 0.388 0.90
LLS 0.141 0.89
LLD 0.018 0.88
KORCULA 0.129 0.88
INT 2.50e−08 0.91
FR97 0.027 0.90
FR07 0.013 0.90
EUGENDA 0.199 0.89
ERF 0.054 0.86
EGCUT 0.058 0.87
CKB 1.80e−04 0.81
ALSPAC 1.70e−07 0.91
META 2.61e−34 0.89
−0.2
0.0
0.2
0.4
0.6
BETA

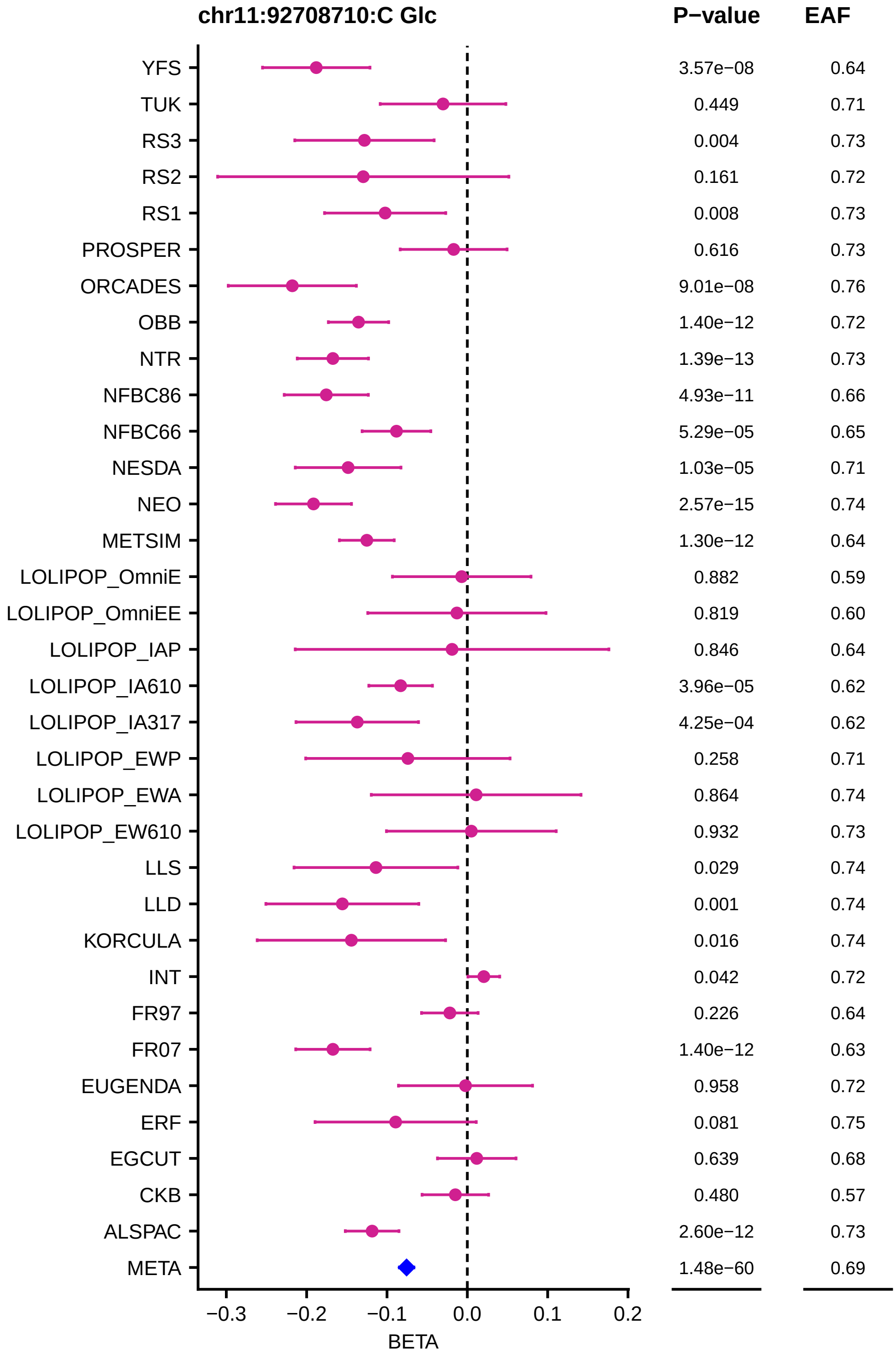
chr11:92708710:C Glc
P−value
EAF
YFS
TUK
RS3
RS2
RS1
PROSPER
ORCADES
OBB
NTR
NFBC86
NFBC66
NESDA
NEO
METSIM
LOLIPOP_OmniE
LOLIPOP_OmniEE
LOLIPOP_IAP
LOLIPOP_IA610
LOLIPOP_IA317
LOLIPOP_EWP
LOLIPOP_EWA
LOLIPOP_EW610
LLS
LLD
KORCULA
INT
FR97
FR07
EUGENDA
ERF
EGCUT
CKB
ALSPAC
META
3.57e−08 0.64
0.449 0.71
0.004 0.73
0.161 0.72
0.008 0.73
0.616 0.73
9.01e−08 0.76
1.40e−12 0.72
1.39e−13 0.73
4.93e−11 0.66
5.29e−05 0.65
1.03e−05 0.71
2.57e−15 0.74
1.30e−12 0.64
0.882 0.59
0.819 0.60
0.846 0.64
3.96e−05 0.62
4.25e−04 0.62
0.258 0.71
0.864 0.74
0.932 0.73
0.029 0.74
0.001 0.74
0.016 0.74
0.042 0.72
0.226 0.64
1.40e−12 0.63
0.958 0.72
0.081 0.75
0.639 0.68
0.480 0.57
2.60e−12 0.73
1.48e−60 0.69
−0.3
−0.2
−0.1
0.0
0.1
0.2
BETA

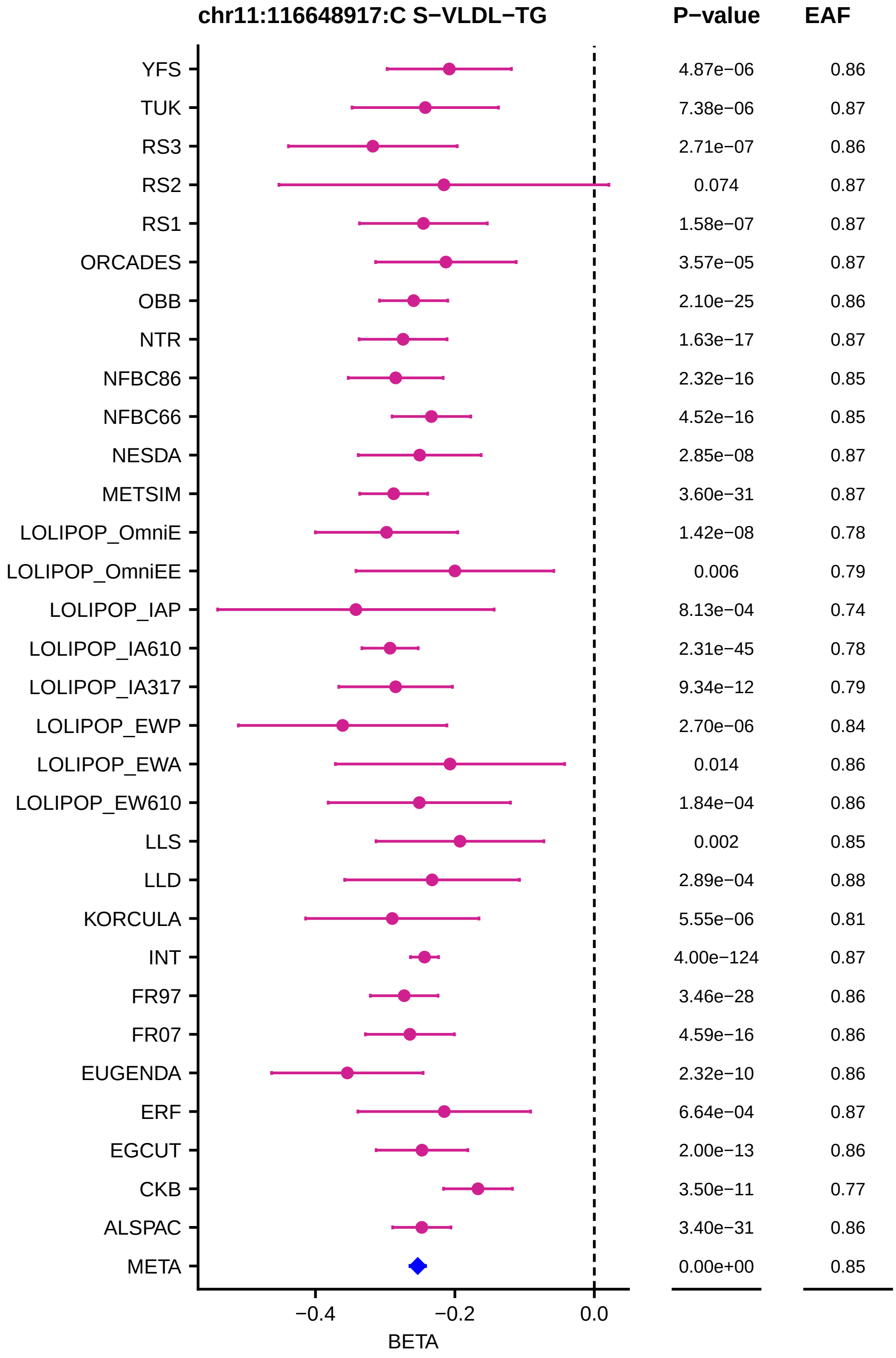
chr11:116648917:C S−VLDL−TG
P−value
EAF
YFS
TUK
RS3
RS2
RS1
ORCADES
OBB
NTR
NFBC86
NFBC66
NESDA
METSIM
LOLIPOP_OmniE
LOLIPOP_OmniEE
LOLIPOP_IAP
LOLIPOP_IA610
LOLIPOP_IA317
LOLIPOP_EWP
LOLIPOP_EWA
LOLIPOP_EW610
LLS
LLD
KORCULA
INT
FR97
FR07
EUGENDA
ERF
EGCUT
CKB
ALSPAC
META
4.87e−06
7.38e−06
2.71e−07
0.074
1.58e−07
3.57e−05
2.10e−25
1.63e−17
2.32e−16
4.52e−16
2.85e−08
3.60e−31
1.42e−08
0.006
8.13e−04
2.31e−45
9.34e−12
2.70e−06
0.014
1.84e−04
0.002
2.89e−04
5.55e−06
4.00e−124
3.46e−28
4.59e−16
2.32e−10
6.64e−04
2.00e−13
3.50e−11
3.40e−31
0.00e+00
0.86
0.87
0.86
0.87
0.87
0.87
0.86
0.87
0.85
0.85
0.87
0.87
0.78
0.79
0.74
0.78
0.79
0.84
0.86
0.86
0.85
0.88
0.81
0.87
0.86
0.86
0.86
0.87
0.86
0.77
0.86
0.85
−0.4
−0.2
0.0
BETA

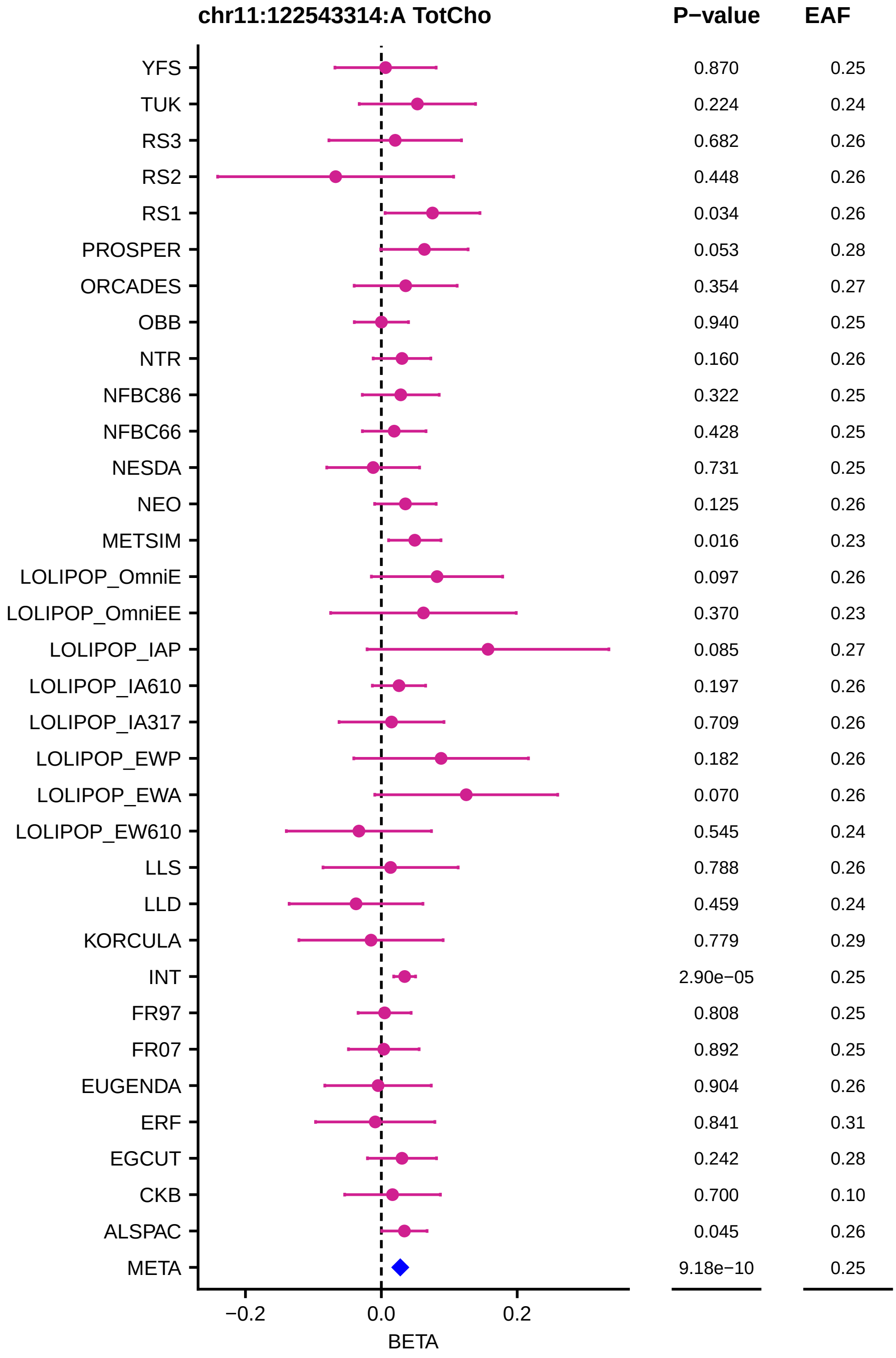
chr11:122543314:A TotCho
P−value
EAF
YFS 0.870 0.25
TUK 0.224 0.24
RS3 0.682 0.26
RS2 0.448 0.26
RS1 0.034 0.26
PROSPER 0.053 0.28
ORCADES 0.354 0.27
OBB 0.940 0.25
NTR 0.160 0.26
NFBC86 0.322 0.25
NFBC66 0.428 0.25
NESDA 0.731 0.25
NEO 0.125 0.26
METSIM 0.016 0.23
LOLIPOP_OmniE 0.097 0.26
LOLIPOP_OmniEE 0.370 0.23
LOLIPOP_IAP 0.085 0.27
LOLIPOP_IA610 0.197 0.26
LOLIPOP_IA317 0.709 0.26
LOLIPOP_EWP 0.182 0.26
LOLIPOP_EWA 0.070 0.26
LOLIPOP_EW610 0.545 0.24
LLS 0.788 0.26
LLD 0.459 0.24
KORCULA 0.779 0.29
INT 2.90e−05 0.25
FR97 0.808 0.25
FR07 0.892 0.25
EUGENDA 0.904 0.26
ERF 0.841 0.31
EGCUT 0.242 0.28
CKB 0.700 0.10
ALSPAC 0.045 0.26
META 9.18e−10 0.25
−0.2
0.0
0.2
BETA

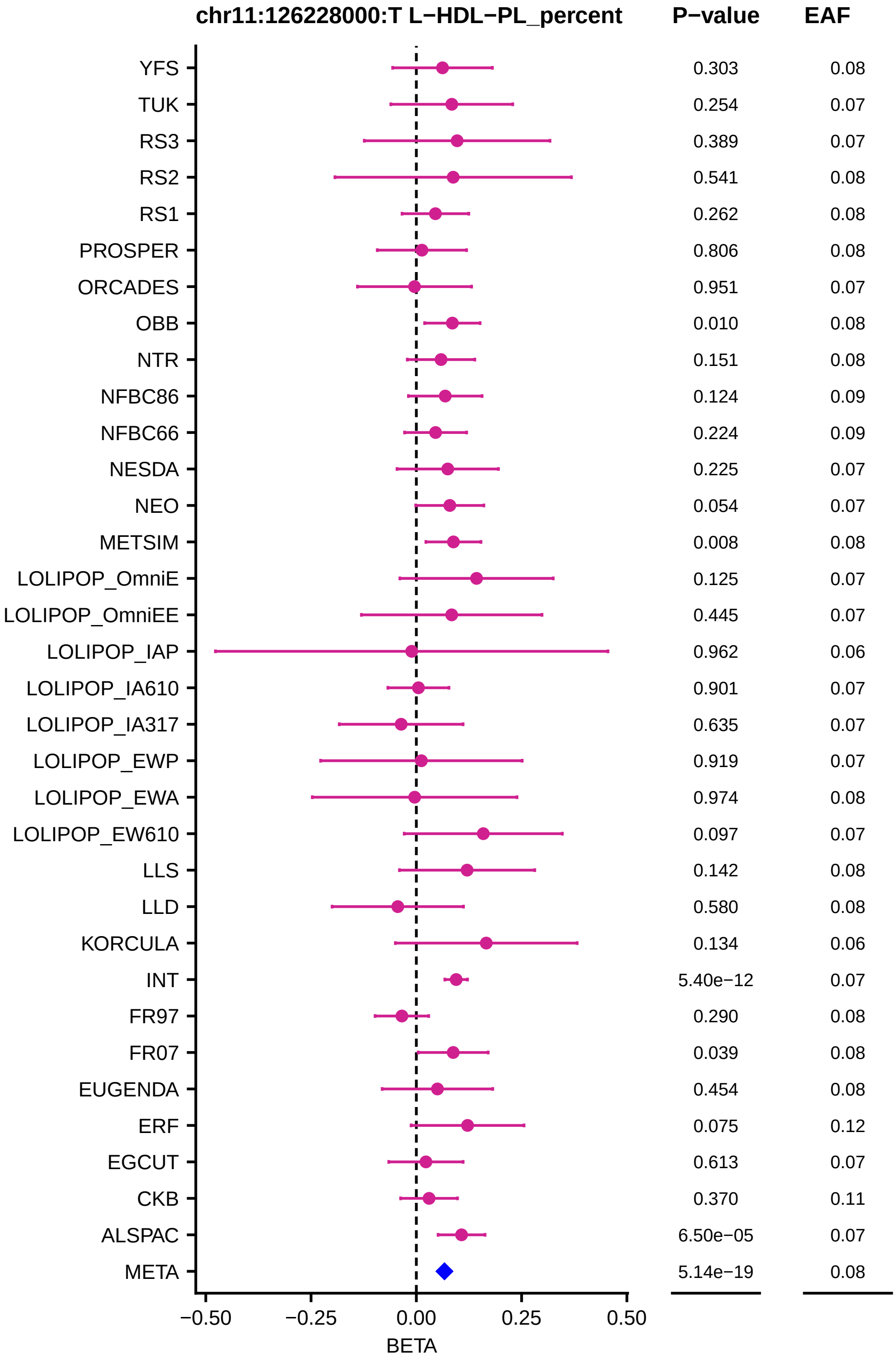
chr11:126228000:T L−HDL−PL_percent
P−value
EAF
YFS 0.303 0.08
TUK 0.254 0.07
RS3 0.389 0.07
RS2 0.541 0.08
RS1 0.262 0.08
PROSPER 0.806 0.08
ORCADES 0.951 0.07
OBB 0.010 0.08
NTR 0.151 0.08
NFBC86 0.124 0.09
NFBC66 0.224 0.09
NESDA 0.225 0.07
NEO 0.054 0.07
METSIM 0.008 0.08
LOLIPOP_OmniE 0.125 0.07
LOLIPOP_OmniEE 0.445 0.07
LOLIPOP_IAP 0.962 0.06
LOLIPOP_IA610 0.901 0.07
LOLIPOP_IA317 0.635 0.07
LOLIPOP_EWP 0.919 0.07
LOLIPOP_EWA 0.974 0.08
LOLIPOP_EW610 0.097 0.07
LLS 0.142 0.08
LLD 0.580 0.08
KORCULA 0.134 0.06
INT 5.40e−12 0.07
FR97 0.290 0.08
FR07 0.039 0.08
EUGENDA 0.454 0.08
ERF 0.075 0.12
EGCUT 0.613 0.07
CKB 0.370 0.11
ALSPAC 6.50e−05 0.07
META 5.14e−19 0.08
−0.50
−0.25
0.00
0.25
0.50
BETA

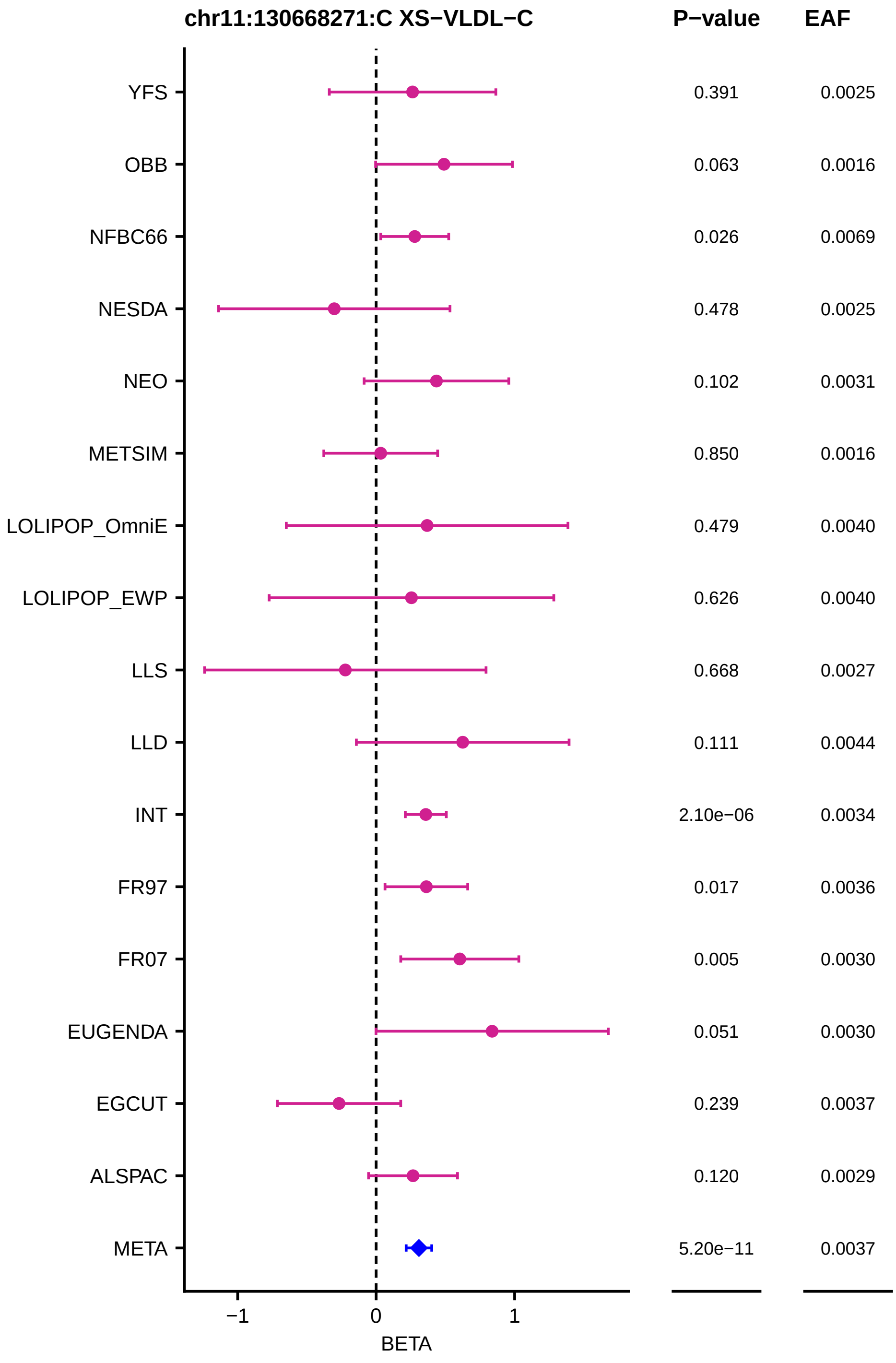
chr11:130668271:C XS−VLDL−C
P−value
EAF
YFS
0.391
0.0025
OBB
0.063
0.0016
NFBC66
0.026
0.0069
NESDA
0.478
0.0025
NEO
0.102
0.0031
METSIM
0.850
0.0016
LOLIPOP_OmniE
0.479
0.0040
LOLIPOP_EWP
0.626
0.0040
LLS
0.668
0.0027
LLD
0.111
0.0044
INT
2.10e−06
0.0034
FR97
0.017
0.0036
FR07
0.005
0.0030
EUGENDA
0.051
0.0030
EGCUT
0.239
0.0037
ALSPAC
0.120
0.0029
META
5.20e−11
0.0037
−1
0
1
BETA

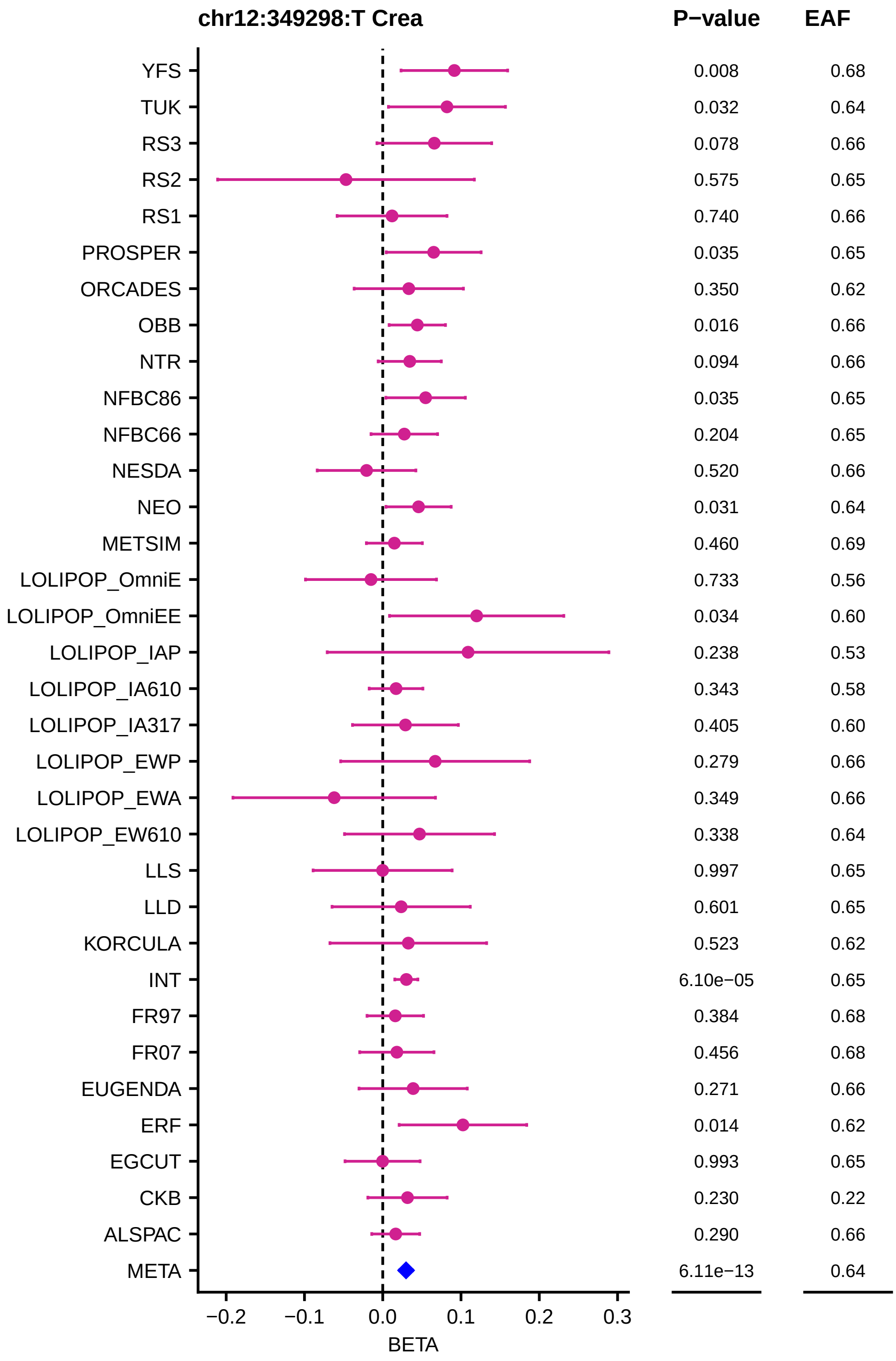
chr12:349298:T Crea
P−value
EAF
YFS 0.008 0.68
TUK 0.032 0.64
RS3 0.078 0.66
RS2 0.575 0.65
RS1 0.740 0.66
PROSPER 0.035 0.65
ORCADES 0.350 0.62
OBB 0.016 0.66
NTR 0.094 0.66
NFBC86 0.035 0.65
NFBC66 0.204 0.65
NESDA 0.520 0.66
NEO 0.031 0.64
METSIM 0.460 0.69
LOLIPOP_OmniE 0.733 0.56
LOLIPOP_OmniEE 0.034 0.60
LOLIPOP_IAP 0.238 0.53
LOLIPOP_IA610 0.343 0.58
LOLIPOP_IA317 0.405 0.60
LOLIPOP_EWP 0.279 0.66
LOLIPOP_EWA 0.349 0.66
LOLIPOP_EW610 0.338 0.64
LLS 0.997 0.65
LLD 0.601 0.65
KORCULA 0.523 0.62
INT 6.10e−05 0.65
FR97 0.384 0.68
FR07 0.456 0.68
EUGENDA 0.271 0.66
ERF 0.014 0.62
EGCUT 0.993 0.65
CKB 0.230 0.22
ALSPAC 0.290 0.66
META 6.11e−13 0.64
−0.2 −0.1 0.0 0.1 0.2 0.3
BETA

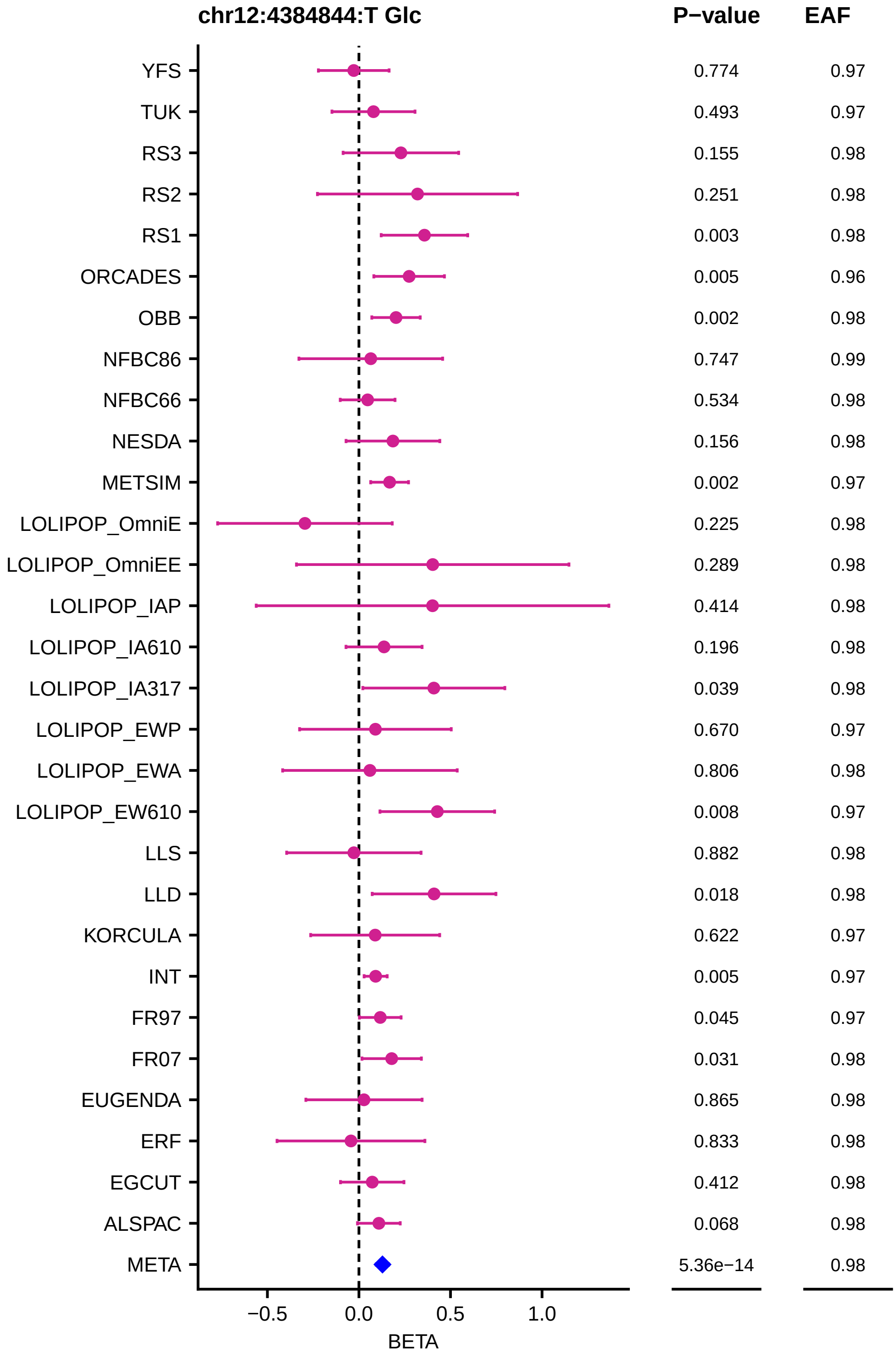
chr12:4384844:T Glc
P−value
EAF
YFS 0.774 0.97
TUK 0.493 0.97
RS3 0.155 0.98
RS2 0.251 0.98
RS1 0.003 0.98
ORCADES 0.005 0.96
OBB 0.002 0.98
NFBC86 0.747 0.99
NFBC66 0.534 0.98
NESDA 0.156 0.98
METSIM 0.002 0.97
LOLIPOP_OmniE 0.225 0.98
LOLIPOP_OmniEE 0.289 0.98
LOLIPOP_IAP 0.414 0.98
LOLIPOP_IA610 0.196 0.98
LOLIPOP_IA317 0.039 0.98
LOLIPOP_EWP 0.670 0.97
LOLIPOP_EWA 0.806 0.98
LOLIPOP_EW610 0.008 0.97
LLS 0.882 0.98
LLD 0.018 0.98
KORCULA 0.622 0.97
INT 0.005 0.97
FR97 0.045 0.97
FR07 0.031 0.98
EUGENDA 0.865 0.98
ERF 0.833 0.98
EGCUT 0.412 0.98
ALSPAC 0.068 0.98
META 5.36e−14 0.98
−0.5
0.0
0.5
1.0
BETA

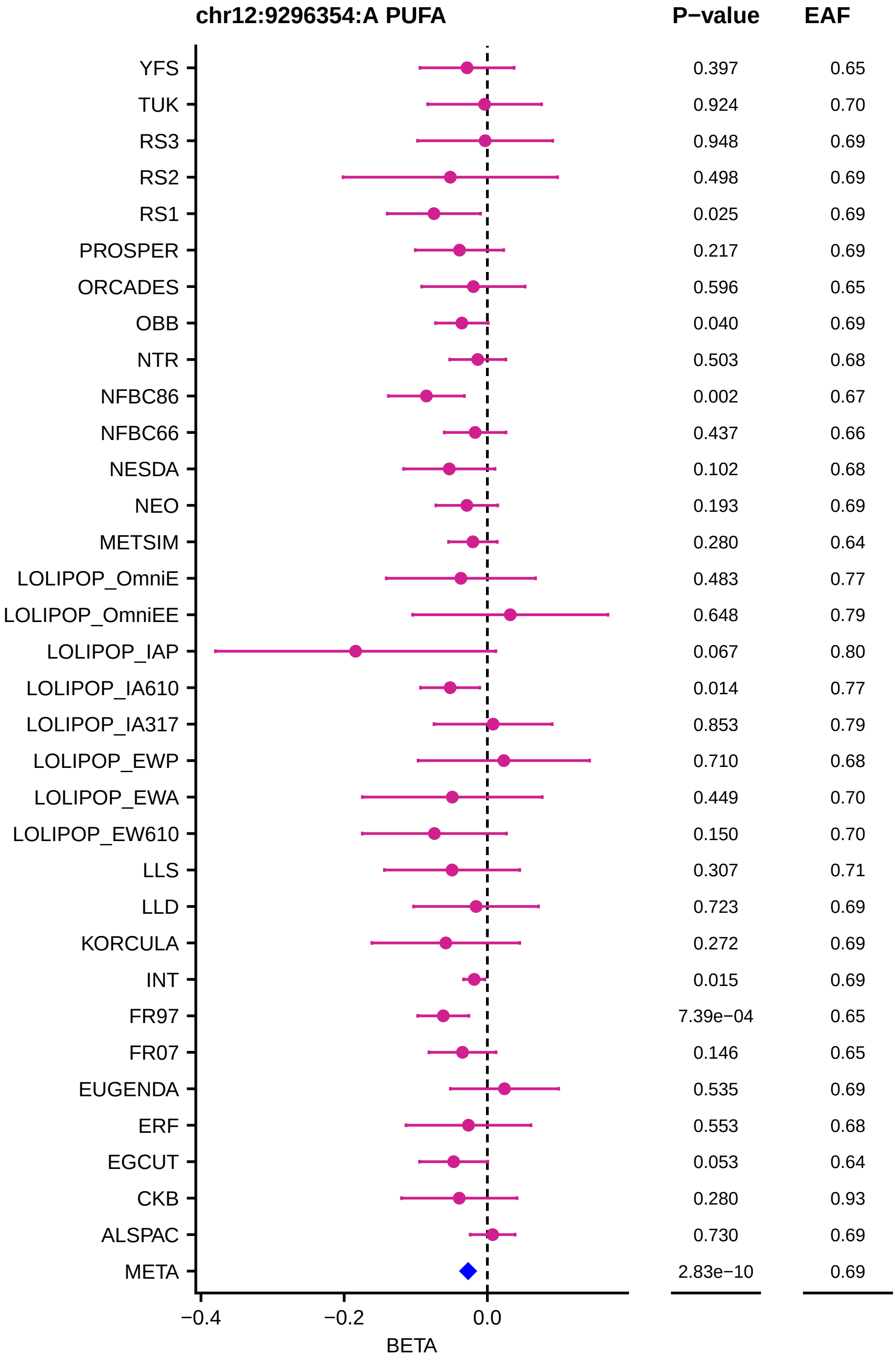
chr12:9296354:A PUFA
P−value
EAF
YFS 0.397 0.65
TUK 0.924 0.70
RS3 0.948 0.69
RS2 0.498 0.69
RS1 0.025 0.69
PROSPER 0.217 0.69
ORCADES 0.596 0.65
OBB 0.040 0.69
NTR 0.503 0.68
NFBC86 0.002 0.67
NFBC66 0.437 0.66
NESDA 0.102 0.68
NEO 0.193 0.69
METSIM 0.280 0.64
LOLIPOP_OmniE 0.483 0.77
LOLIPOP_OmniEE 0.648 0.79
LOLIPOP_IAP 0.067 0.80
LOLIPOP_IA610 0.014 0.77
LOLIPOP_IA317 0.853 0.79
LOLIPOP_EWP 0.710 0.68
LOLIPOP_EWA 0.449 0.70
LOLIPOP_EW610 0.150 0.70
LLS 0.307 0.71
LLD 0.723 0.69
KORCULA 0.272 0.69
INT 0.015 0.69
FR97 7.39e−04 0.65
FR07 0.146 0.65
EUGENDA 0.535 0.69
ERF 0.553 0.68
EGCUT 0.053 0.64
CKB 0.280 0.93
ALSPAC 0.730 0.69
META 2.83e−10 0.69
−0.4
−0.2
0.0
BETA

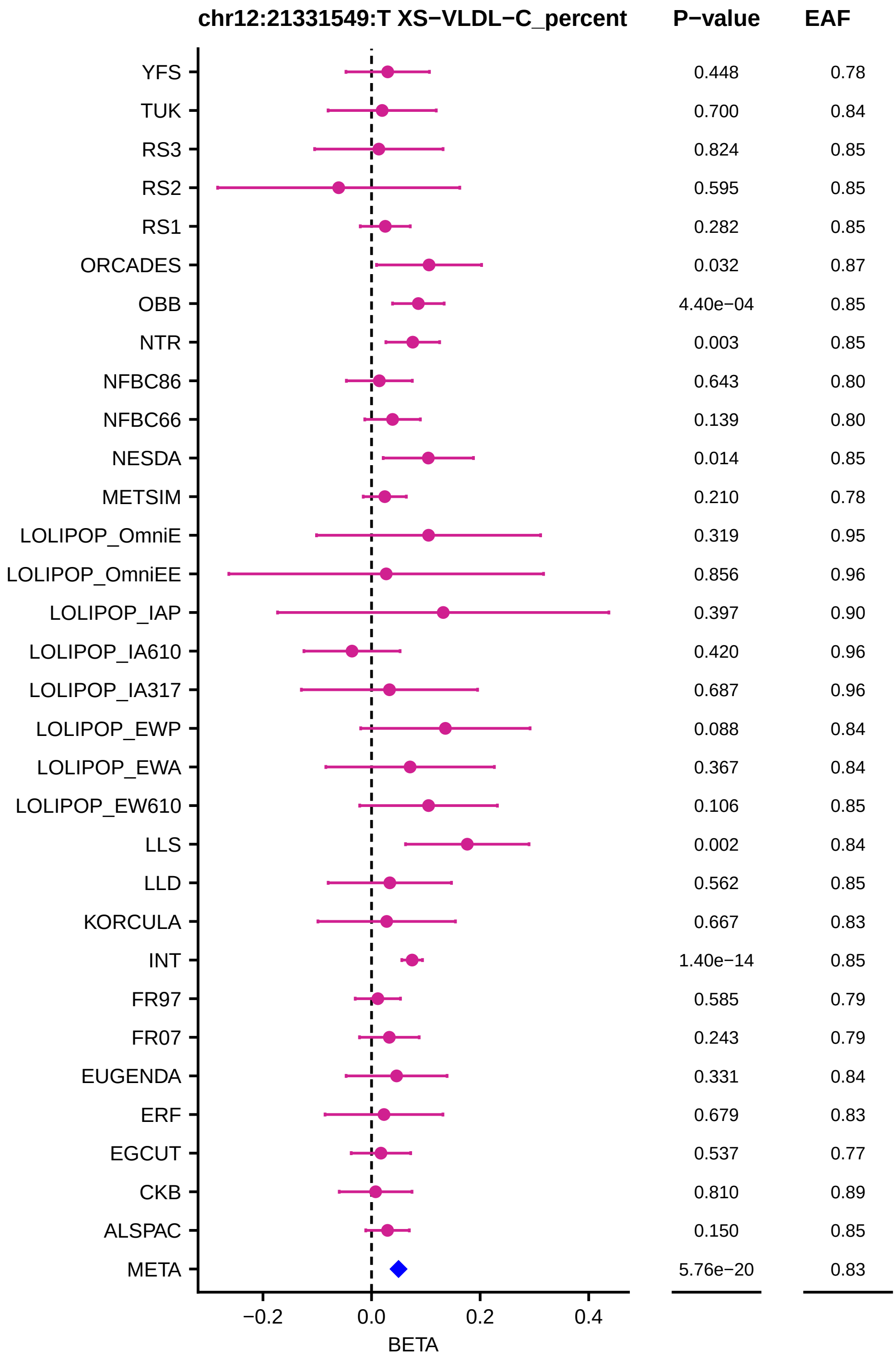
chr12:21331549:T XS−VLDL−C_percent
P−value
EAF
YFS 0.448 0.78
TUK 0.700 0.84
RS3 0.824 0.85
RS2 0.595 0.85
RS1 0.282 0.85
ORCADES 0.032 0.87
OBB 4.40e−04 0.85
NTR 0.003 0.85
NFBC86 0.643 0.80
NFBC66 0.139 0.80
NESDA 0.014 0.85
METSIM 0.210 0.78
LOLIPOP_OmniE 0.319 0.95
LOLIPOP_OmniEE 0.856 0.96
LOLIPOP_IAP 0.397 0.90
LOLIPOP_IA610 0.420 0.96
LOLIPOP_IA317 0.687 0.96
LOLIPOP_EWP 0.088 0.84
LOLIPOP_EWA 0.367 0.84
LOLIPOP_EW610 0.106 0.85
LLS 0.002 0.84
LLD 0.562 0.85
KORCULA 0.667 0.83
INT 1.40e−14 0.85
FR97 0.585 0.79
FR07 0.243 0.79
EUGENDA 0.331 0.84
ERF 0.679 0.83
EGCUT 0.537 0.77
CKB 0.810 0.89
ALSPAC 0.150 0.85
META 5.76e−20 0.83
−0.2
0.0
0.2
0.4
BETA

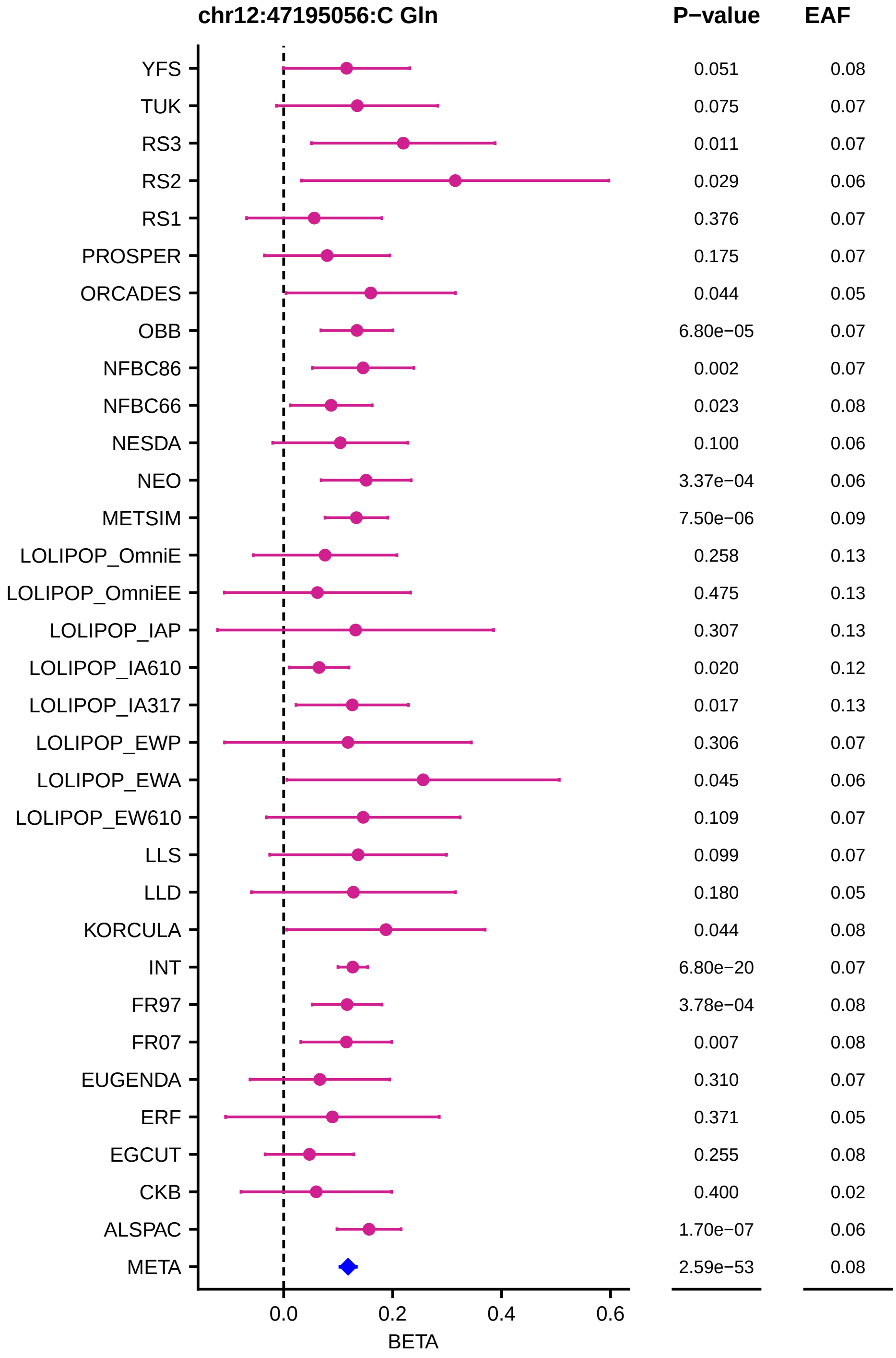

chr12:47195056:C Gln
P−value
EAF
YFS 0.051 0.08
TUK 0.075 0.07
RS3 0.011 0.07
RS2 0.029 0.06
RS1 0.376 0.07
PROSPER 0.175 0.07
ORCADES 0.044 0.05
OBB 6.80e−05 0.07
NFBC86 0.002 0.07
NFBC66 0.023 0.08
NESDA 0.100 0.06
NEO 3.37e−04 0.06
METSIM 7.50e−06 0.09
LOLIPOP_OmniE 0.258 0.13
LOLIPOP_OmniEE 0.475 0.13
LOLIPOP_IAP 0.307 0.13
LOLIPOP_IA610 0.020 0.12
LOLIPOP_IA317 0.017 0.13
LOLIPOP_EWP 0.306 0.07
LOLIPOP_EWA 0.045 0.06
LOLIPOP_EW610 0.109 0.07
LLS 0.099 0.07
LLD 0.180 0.05
KORCULA 0.044 0.08
INT 6.80e−20 0.07
FR97 3.78e−04 0.08
FR07 0.007 0.08
EUGENDA 0.310 0.07
ERF 0.371 0.05
EGCUT 0.255 0.08
CKB 0.400 0.02
ALSPAC 1.70e−07 0.06
META 2.59e−53 0.08
0.0
0.2
0.4
0.6
BETA

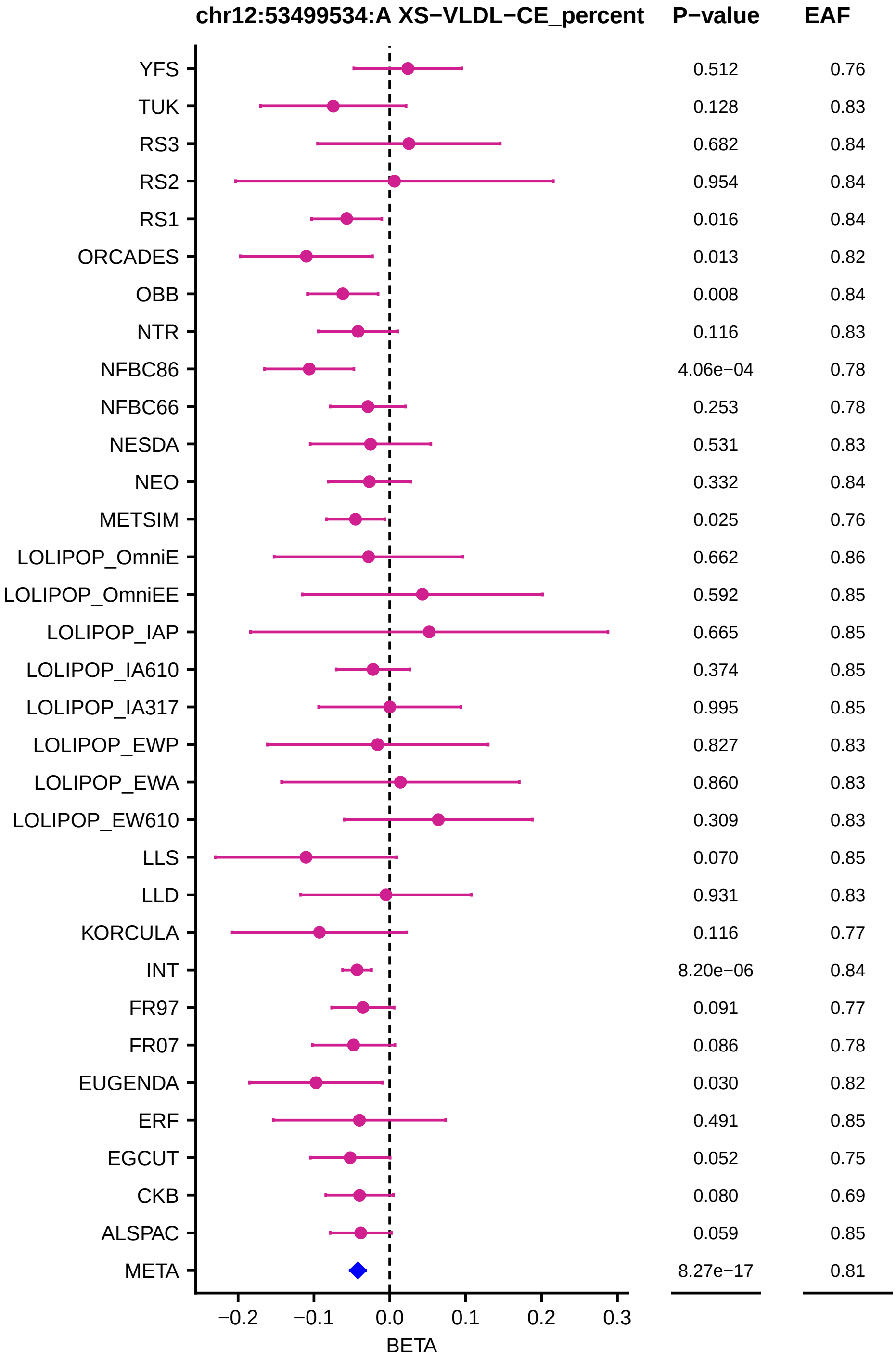
chr12:53499534:A XS−VLDL−CE_percent
P−value
EAF
YFS
TUK
RS3
RS2
RS1
ORCADES
OBB
NTR
NFBC86
NFBC66
NESDA
NEO
METSIM
LOLIPOP_OmniE
LOLIPOP_OmniEE
LOLIPOP_IAP
LOLIPOP_IA610
LOLIPOP_IA317
LOLIPOP_EWP
LOLIPOP_EWA
LOLIPOP_EW610
LLS
LLD
KORCULA
INT
FR97
FR07
EUGENDA
ERF
EGCUT
CKB
ALSPAC
META
0.512 0.76
0.128 0.83
0.682 0.84
0.954 0.84
0.016 0.84
0.013 0.82
0.008 0.84
0.116 0.83
4.06e−04 0.78
0.253 0.78
0.531 0.83
0.332 0.84
0.025 0.76
0.662 0.86
0.592 0.85
0.665 0.85
0.374 0.85
0.995 0.85
0.827 0.83
0.860 0.83
0.309 0.83
0.070 0.85
0.931 0.83
0.116 0.77
8.20e−06 0.84
0.091 0.77
0.086 0.78
0.030 0.82
0.491 0.85
0.052 0.75
0.080 0.69
0.059 0.85
8.27e−17 0.81
−0.2
−0.1
0.0
0.1
0.2
0.3
BETA

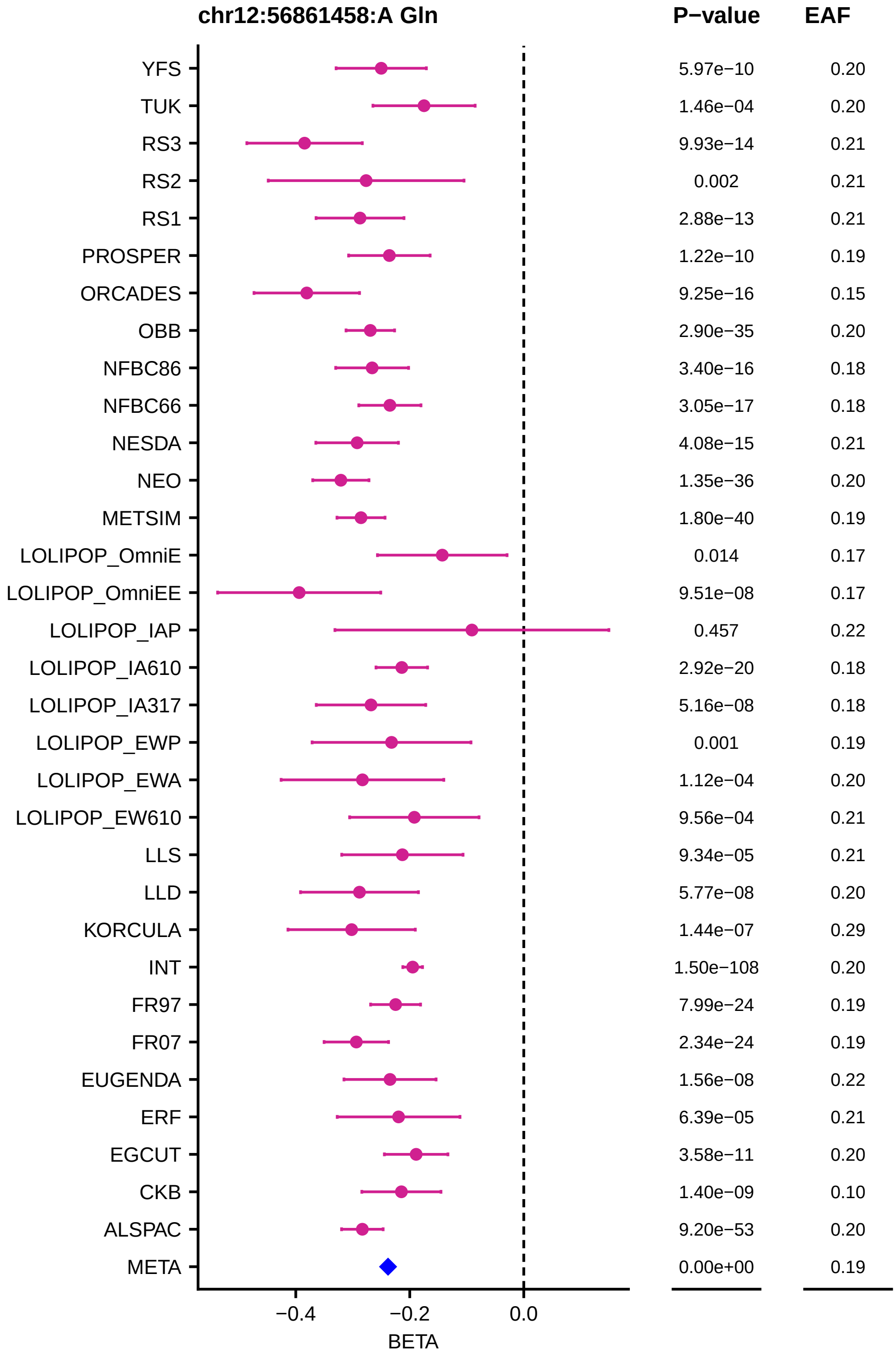
chr12:56861458:A Gln
P−value
EAF
YFS 5.97e−10 0.20
TUK 1.46e−04 0.20
RS3 9.93e−14 0.21
RS2 0.002 0.21
RS1 2.88e−13 0.21
PROSPER 1.22e−10 0.19
ORCADES 9.25e−16 0.15
OBB 2.90e−35 0.20
NFBC86 3.40e−16 0.18
NFBC66 3.05e−17 0.18
NESDA 4.08e−15 0.21
NEO 1.35e−36 0.20
METSIM 1.80e−40 0.19
LOLIPOP_OmniE 0.014 0.17
LOLIPOP_OmniEE 9.51e−08 0.17
LOLIPOP_IAP 0.457 0.22
LOLIPOP_IA610 2.92e−20 0.18
LOLIPOP_IA317 5.16e−08 0.18
LOLIPOP_EWP 0.001 0.19
LOLIPOP_EWA 1.12e−04 0.20
LOLIPOP_EW610 9.56e−04 0.21
LLS 9.34e−05 0.21
LLD 5.77e−08 0.20
KORCULA 1.44e−07 0.29
INT 1.50e−108 0.20
FR97 7.99e−24 0.19
FR07 2.34e−24 0.19
EUGENDA 1.56e−08 0.22
ERF 6.39e−05 0.21
EGCUT 3.58e−11 0.20
CKB 1.40e−09 0.10
ALSPAC 9.20e−53 0.20
META 0.00e+00 0.19
−0.4
−0.2
0.0
BETA

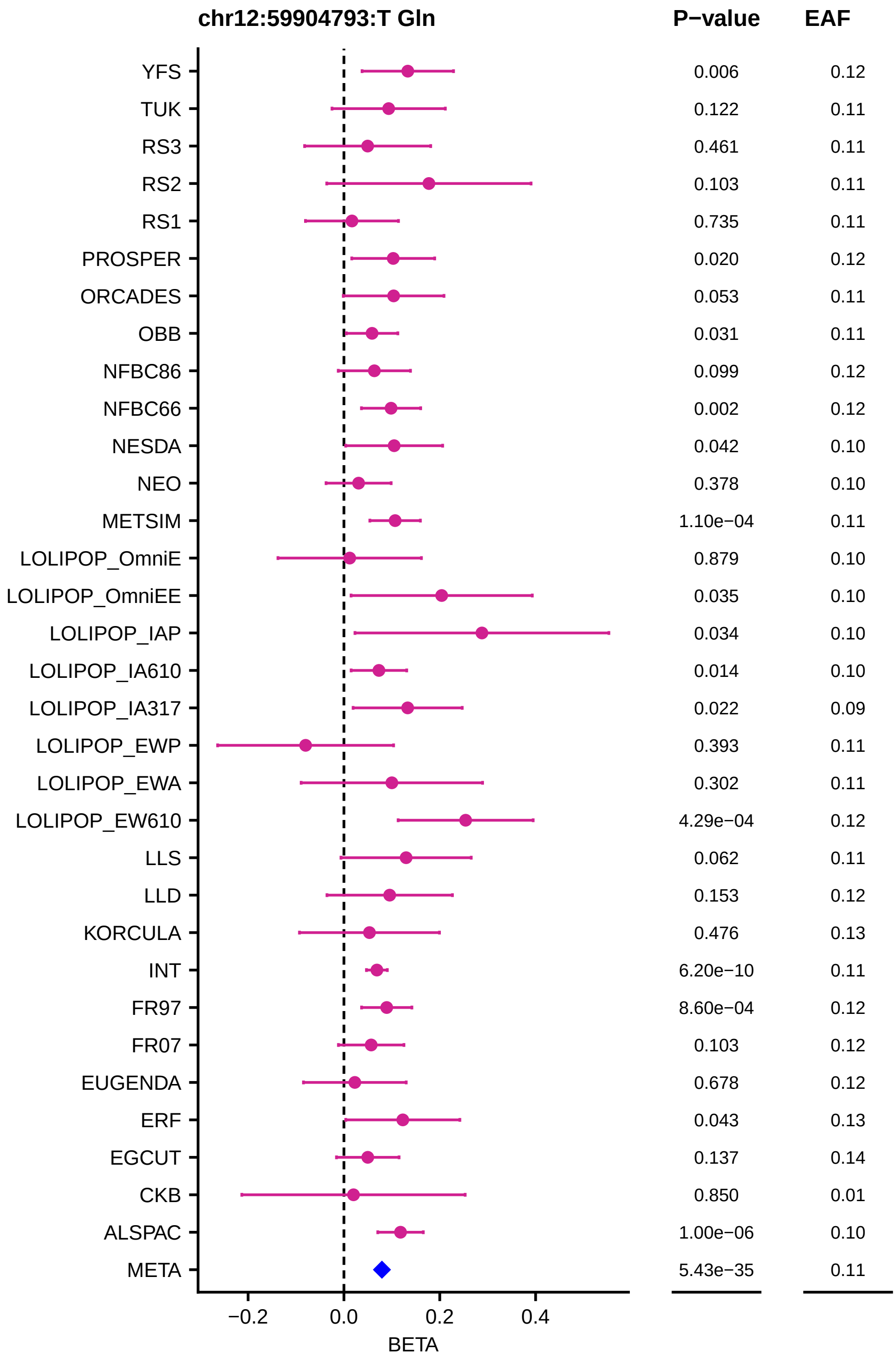
chr12:59904793:T Gln
P−value
EAF
YFS 0.006 0.12
TUK 0.122 0.11
RS3 0.461 0.11
RS2 0.103 0.11
RS1 0.735 0.11
PROSPER 0.020 0.12
ORCADES 0.053 0.11
OBB 0.031 0.11
NFBC86 0.099 0.12
NFBC66 0.002 0.12
NESDA 0.042 0.10
NEO 0.378 0.10
METSIM 1.10e−04 0.11
LOLIPOP_OmniE 0.879 0.10
LOLIPOP_OmniEE 0.035 0.10
LOLIPOP_IAP 0.034 0.10
LOLIPOP_IA610 0.014 0.10
LOLIPOP_IA317 0.022 0.09
LOLIPOP_EWP 0.393 0.11
LOLIPOP_EWA 0.302 0.11
LOLIPOP_EW610 4.29e−04 0.12
LLS 0.062 0.11
LLD 0.153 0.12
KORCULA 0.476 0.13
INT 6.20e−10 0.11
FR97 8.60e−04 0.12
FR07 0.103 0.12
EUGENDA 0.678 0.12
ERF 0.043 0.13
EGCUT 0.137 0.14
CKB 0.850 0.01
ALSPAC 1.00e−06 0.10
META 5.43e−35 0.11
−0.2
0.0
0.2
0.4
BETA

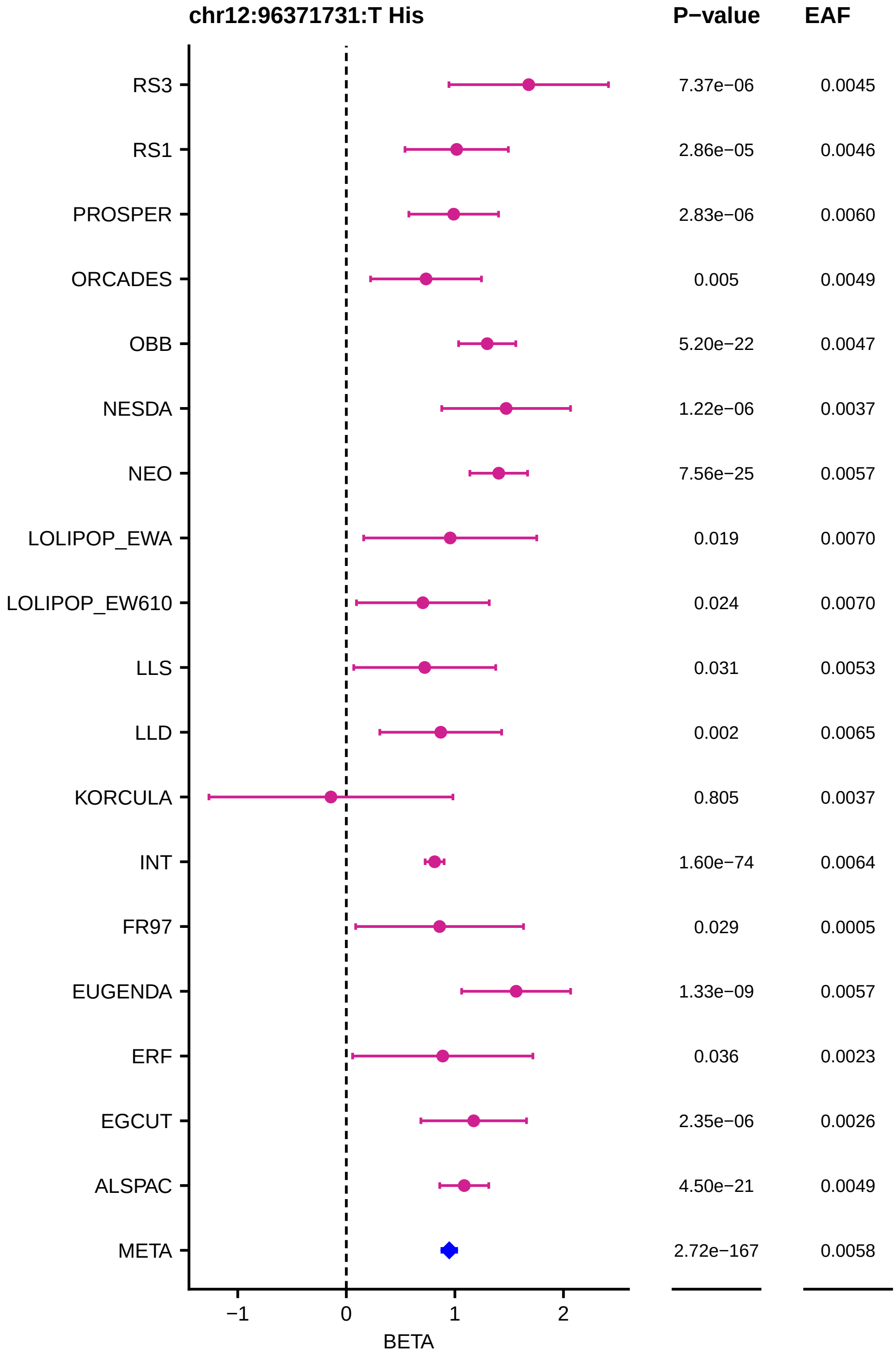
chr12:96371731:T His
P−value
EAF
RS3
7.37e−06
0.0045
RS1
2.86e−05
0.0046
PROSPER
2.83e−06
0.0060
ORCADES
0.005
0.0049
OBB
5.20e−22
0.0047
NESDA
1.22e−06
0.0037
NEO
7.56e−25
0.0057
LOLIPOP_EWA
0.019
0.0070
LOLIPOP_EW610
0.024
0.0070
LLS
0.031
0.0053
LLD
0.002
0.0065
KORCULA
0.805
0.0037
INT
1.60e−74
0.0064
FR97
0.029
0.0005
EUGENDA
1.33e−09
0.0057
ERF
0.036
0.0023
EGCUT
2.35e−06
0.0026
ALSPAC
4.50e−21
0.0049
META
2.72e−167
0.0058
−1
0
1
2
BETA

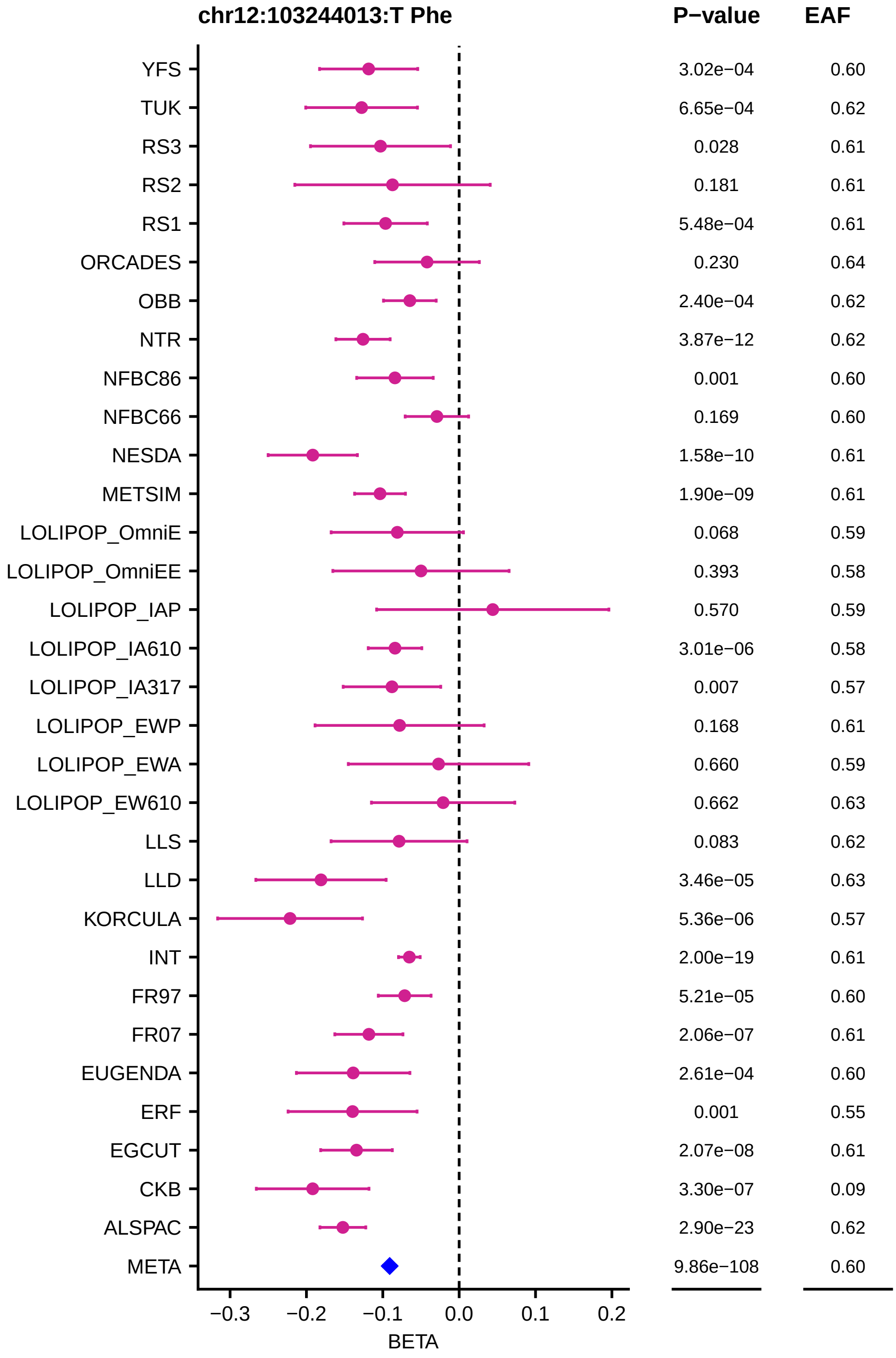
chr12:103244013:T Phe
P−value
EAF
YFS
3.02e−04
0.60
TUK
6.65e−04
0.62
RS3
0.028
0.61
RS2
0.181
0.61
RS1
5.48e−04
0.61
ORCADES
0.230
0.64
OBB
2.40e−04
0.62
NTR
3.87e−12
0.62
NFBC86
0.001
0.60
NFBC66
0.169
0.60
NESDA
1.58e−10
0.61
METSIM
1.90e−09
0.61
LOLIPOP_OmniE
0.068
0.59
LOLIPOP_OmniEE
0.393
0.58
LOLIPOP_IAP
0.570
0.59
LOLIPOP_IA610
3.01e−06
0.58
LOLIPOP_IA317
0.007
0.57
LOLIPOP_EWP
0.168
0.61
LOLIPOP_EWA
0.660
0.59
LOLIPOP_EW610
0.662
0.63
LLS
0.083
0.62
LLD
3.46e−05
0.63
KORCULA
5.36e−06
0.57
INT
2.00e−19
0.61
FR97
5.21e−05
0.60
FR07
2.06e−07
0.61
EUGENDA
2.61e−04
0.60
ERF
0.001
0.55
EGCUT
2.07e−08
0.61
CKB
3.30e−07
0.09
ALSPAC
2.90e−23
0.62
META
9.86e−108
0.60
−0.3
−0.2
−0.1
0.0
0.1
0.2
BETA

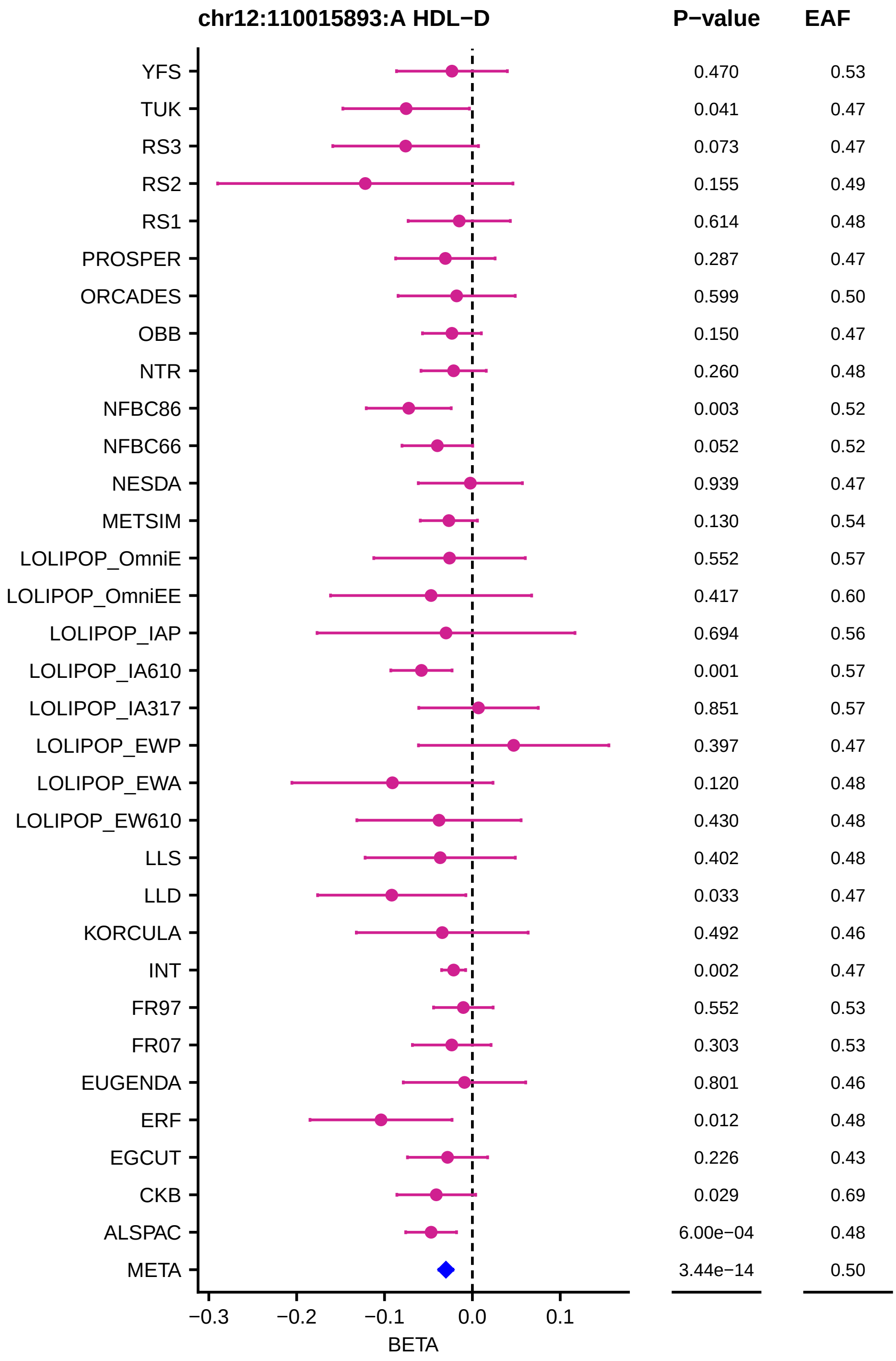
chr12:110015893:A HDL−D
P−value
EAF
YFS 0.470 0.53
TUK 0.041 0.47
RS3 0.073 0.47
RS2 0.155 0.49
RS1 0.614 0.48
PROSPER 0.287 0.47
ORCADES 0.599 0.50
OBB 0.150 0.47
NTR 0.260 0.48
NFBC86 0.003 0.52
NFBC66 0.052 0.52
NESDA 0.939 0.47
METSIM 0.130 0.54
LOLIPOP_OmniE 0.552 0.57
LOLIPOP_OmniEE 0.417 0.60
LOLIPOP_IAP 0.694 0.56
LOLIPOP_IA610 0.001 0.57
LOLIPOP_IA317 0.851 0.57
LOLIPOP_EWP 0.397 0.47
LOLIPOP_EWA 0.120 0.48
LOLIPOP_EW610 0.430 0.48
LLS 0.402 0.48
LLD 0.033 0.47
KORCULA 0.492 0.46
INT 0.002 0.47
FR97 0.552 0.53
FR07 0.303 0.53
EUGENDA 0.801 0.46
ERF 0.012 0.48
EGCUT 0.226 0.43
CKB 0.029 0.69
ALSPAC 6.00e−04 0.48
META 3.44e−14 0.50
−0.3
−0.2
−0.1
0.0
0.1
BETA

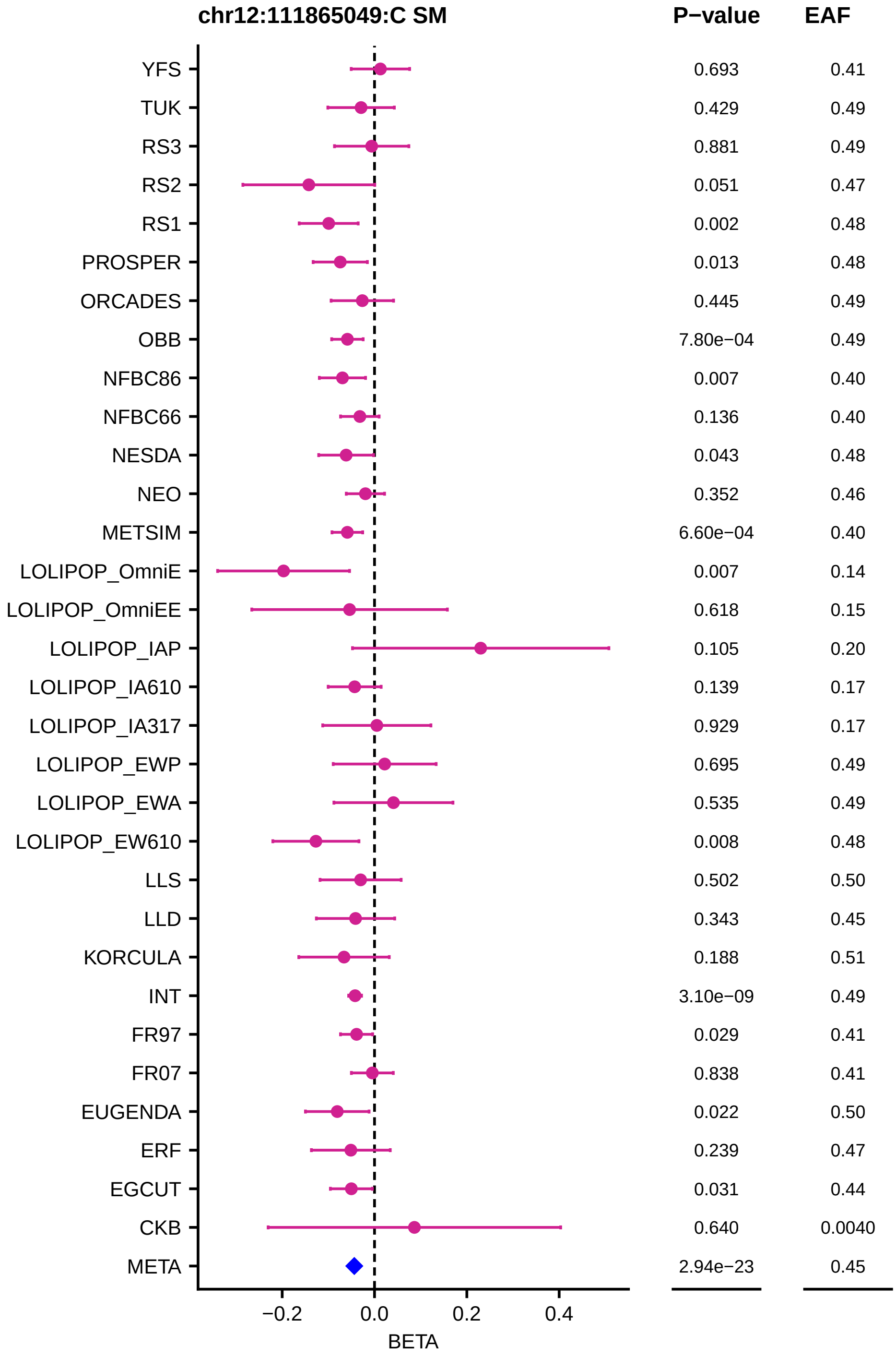
chr12:111865049:C SM
P−value
EAF
YFS
TUK
RS3
RS2
RS1
PROSPER
ORCADES
OBB
NFBC86
NFBC66
NESDA
NEO
METSIM
LOLIPOP_OmniE
LOLIPOP_OmniEE
LOLIPOP_IAP
LOLIPOP_IA610
LOLIPOP_IA317
LOLIPOP_EWP
LOLIPOP_EWA
LOLIPOP_EW610
LLS
LLD
KORCULA
INT
FR97
FR07
EUGENDA
ERF
EGCUT
CKB
META
0.693 0.41
0.429 0.49
0.881 0.49
0.051 0.47
0.002 0.48
0.013 0.48
0.445 0.49
7.80e−04 0.49
0.007 0.40
0.136 0.40
0.043 0.48
0.352 0.46
6.60e−04 0.40
0.007 0.14
0.618 0.15
0.105 0.20
0.139 0.17
0.929 0.17
0.695 0.49
0.535 0.49
0.008 0.48
0.502 0.50
0.343 0.45
0.188 0.51
3.10e−09 0.49
0.029 0.41
0.838 0.41
0.022 0.50
0.239 0.47
0.031 0.44
0.640 0.0040
2.94e−23 0.45
−0.2
0.0
0.2
0.4
BETA

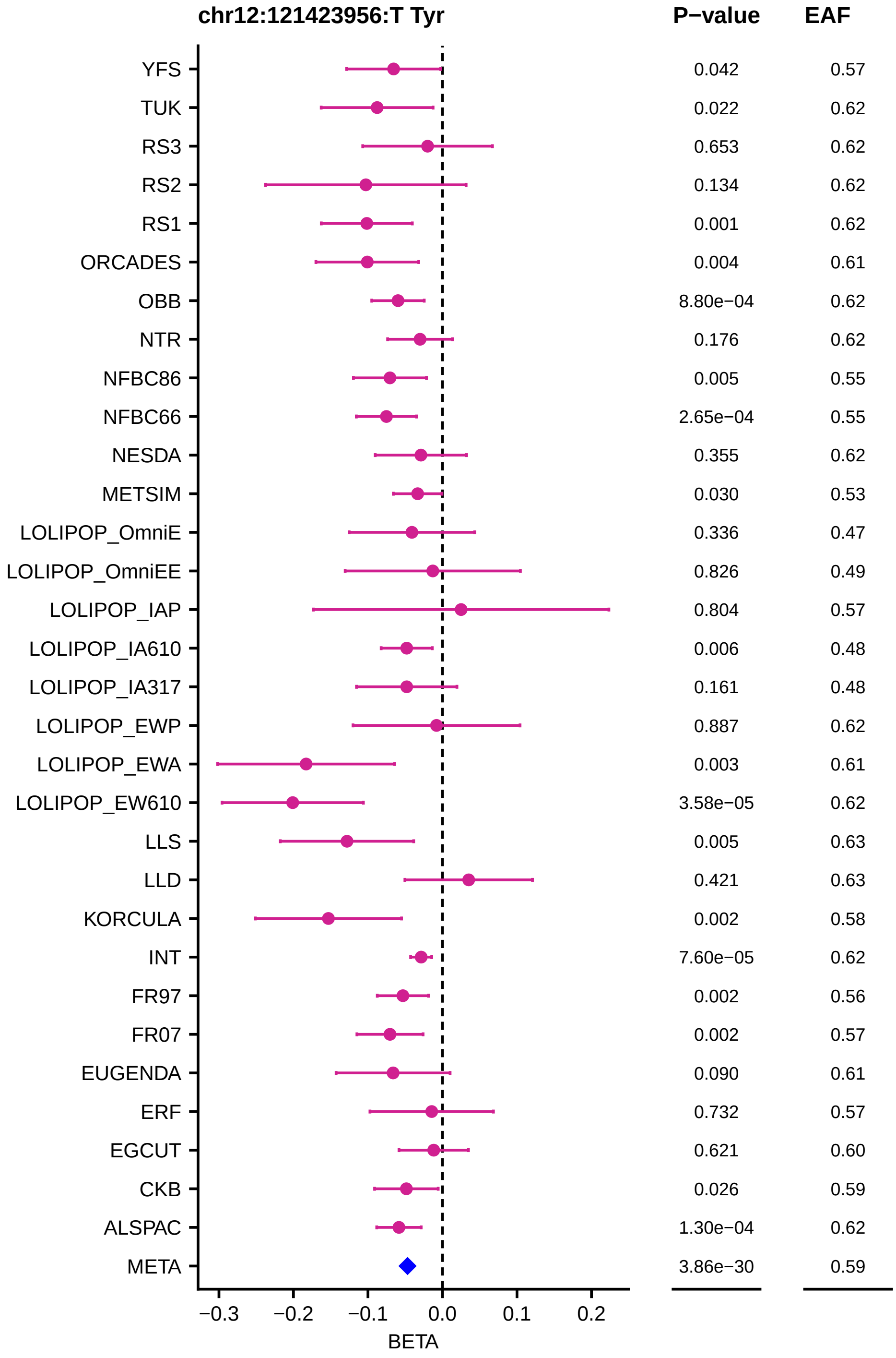
chr12:121423956:T Tyr
P−value
EAF
YFS 0.042 0.57
TUK 0.022 0.62
RS3 0.653 0.62
RS2 0.134 0.62
RS1 0.001 0.62
ORCADES 0.004 0.61
OBB 8.80e−04 0.62
NTR 0.176 0.62
NFBC86 0.005 0.55
NFBC66 2.65e−04 0.55
NESDA 0.355 0.62
METSIM 0.030 0.53
LOLIPOP_OmniE 0.336 0.47
LOLIPOP_OmniEE 0.826 0.49
LOLIPOP_IAP 0.804 0.57
LOLIPOP_IA610 0.006 0.48
LOLIPOP_IA317 0.161 0.48
LOLIPOP_EWP 0.887 0.62
LOLIPOP_EWA 0.003 0.61
LOLIPOP_EW610 3.58e−05 0.62
LLS 0.005 0.63
LLD 0.421 0.63
KORCULA 0.002 0.58
INT 7.60e−05 0.62
FR97 0.002 0.56
FR07 0.002 0.57
EUGENDA 0.090 0.61
ERF 0.732 0.57
EGCUT 0.621 0.60
CKB 0.026 0.59
ALSPAC 1.30e−04 0.62
META 3.86e−30 0.59
−0.3
−0.2
−0.1
0.0
0.1
0.2
BETA

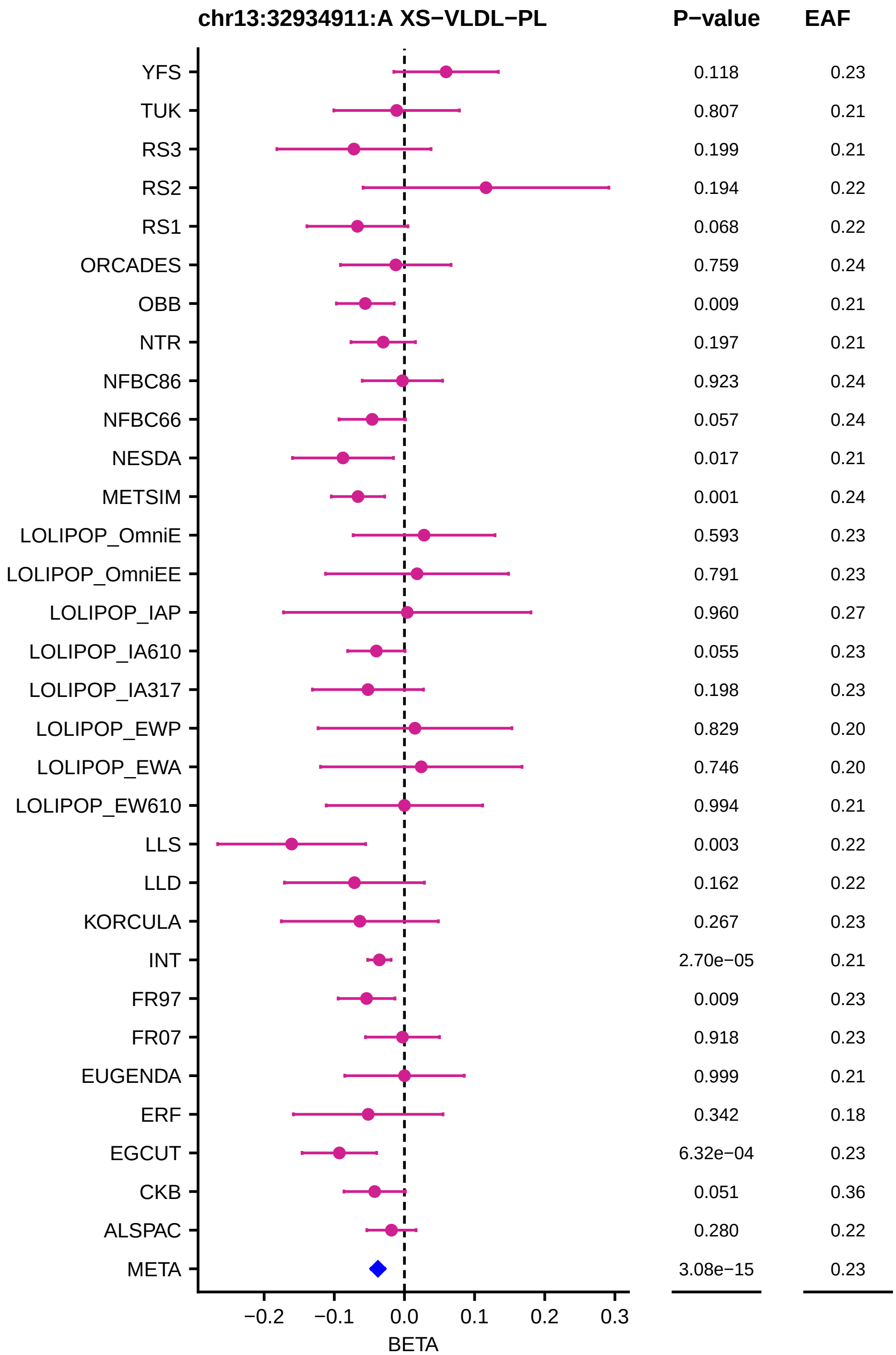
chr13:32934911:A XS−VLDL−PL
P−value
EAF
YFS 0.118 0.23
TUK 0.807 0.21
RS3 0.199 0.21
RS2 0.194 0.22
RS1 0.068 0.22
ORCADES 0.759 0.24
OBB 0.009 0.21
NTR 0.197 0.21
NFBC86 0.923 0.24
NFBC66 0.057 0.24
NESDA 0.017 0.21
METSIM 0.001 0.24
LOLIPOP_OmniE 0.593 0.23
LOLIPOP_OmniEE 0.791 0.23
LOLIPOP_IAP 0.960 0.27
LOLIPOP_IA610 0.055 0.23
LOLIPOP_IA317 0.198 0.23
LOLIPOP_EWP 0.829 0.20
LOLIPOP_EWA 0.746 0.20
LOLIPOP_EW610 0.994 0.21
LLS 0.003 0.22
LLD 0.162 0.22
KORCULA 0.267 0.23
INT 2.70e−05 0.21
FR97 0.009 0.23
FR07 0.918 0.23
EUGENDA 0.999 0.21
ERF 0.342 0.18
EGCUT 6.32e−04 0.23
CKB 0.051 0.36
ALSPAC 0.280 0.22
META 3.08e−15 0.23
−0.2 −0.1 0.0 0.1 0.2 0.3
BETA

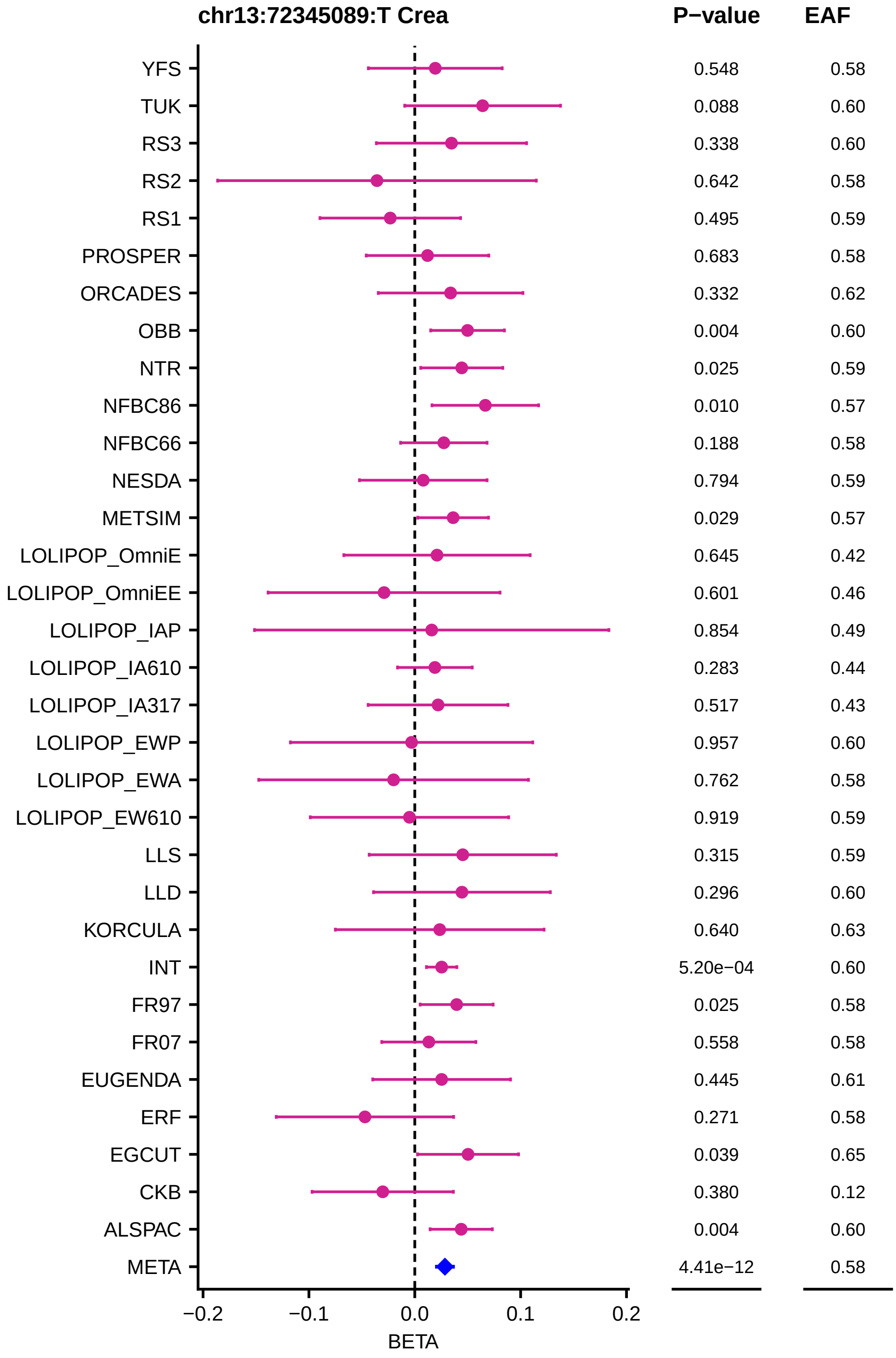

chr13:72345089:T Crea
P−value
EAF
YFS 0.548 0.58
TUK 0.088 0.60
RS3 0.338 0.60
RS2 0.642 0.58
RS1 0.495 0.59
PROSPER 0.683 0.58
ORCADES 0.332 0.62
OBB 0.004 0.60
NTR 0.025 0.59
NFBC86 0.010 0.57
NFBC66 0.188 0.58
NESDA 0.794 0.59
METSIM 0.029 0.57
LOLIPOP_OmniE 0.645 0.42
LOLIPOP_OmniEE 0.601 0.46
LOLIPOP_IAP 0.854 0.49
LOLIPOP_IA610 0.283 0.44
LOLIPOP_IA317 0.517 0.43
LOLIPOP_EWP 0.957 0.60
LOLIPOP_EWA 0.762 0.58
LOLIPOP_EW610 0.919 0.59
LLS 0.315 0.59
LLD 0.296 0.60
KORCULA 0.640 0.63
INT 5.20e−04 0.60
FR97 0.025 0.58
FR07 0.558 0.58
EUGENDA 0.445 0.61
ERF 0.271 0.58
EGCUT 0.039 0.65
CKB 0.380 0.12
ALSPAC 0.004 0.60
META 4.41e−12 0.58
−0.2
−0.1
0.0
0.1
0.2
BETA

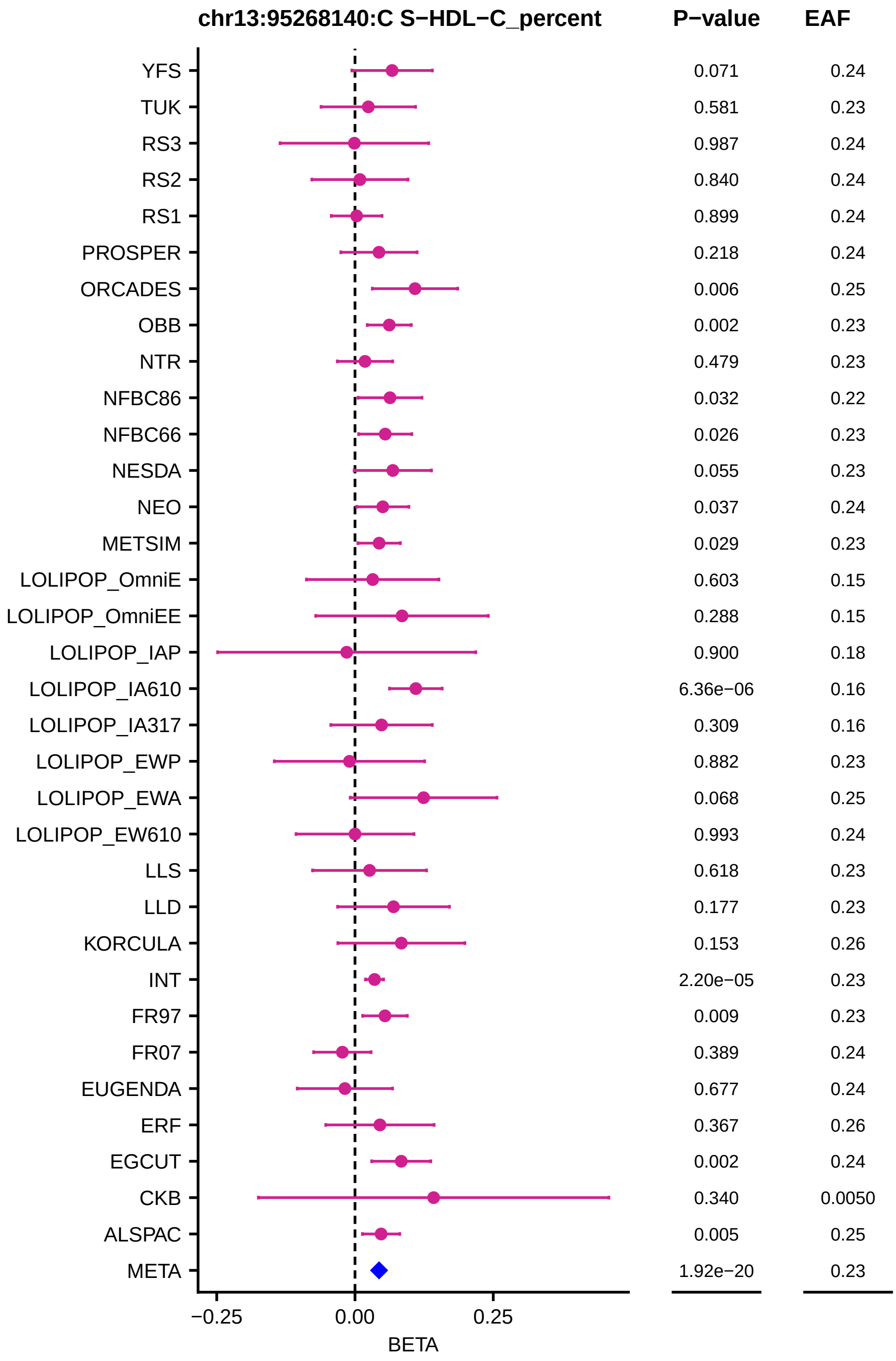

chr13:95268140:C S−HDL−C_percent
P−value
EAF
YFS 0.071 0.24
TUK 0.581 0.23
RS3 0.987 0.24
RS2 0.840 0.24
RS1 0.899 0.24
PROSPER 0.218 0.24
ORCADES 0.006 0.25
OBB 0.002 0.23
NTR 0.479 0.23
NFBC86 0.032 0.22
NFBC66 0.026 0.23
NESDA 0.055 0.23
NEO 0.037 0.24
METSIM 0.029 0.23
LOLIPOP_OmniE 0.603 0.15
LOLIPOP_OmniEE 0.288 0.15
LOLIPOP_IAP 0.900 0.18
LOLIPOP_IA610 6.36e−06 0.16
LOLIPOP_IA317 0.309 0.16
LOLIPOP_EWP 0.882 0.23
LOLIPOP_EWA 0.068 0.25
LOLIPOP_EW610 0.993 0.24
LLS 0.618 0.23
LLD 0.177 0.23
KORCULA 0.153 0.26
INT 2.20e−05 0.23
FR97 0.009 0.23
FR07 0.389 0.24
EUGENDA 0.677 0.24
ERF 0.367 0.26
EGCUT 0.002 0.24
CKB 0.340 0.0050
ALSPAC 0.005 0.25
META 1.92e−20 0.23
−0.25
0.00
0.25
BETA

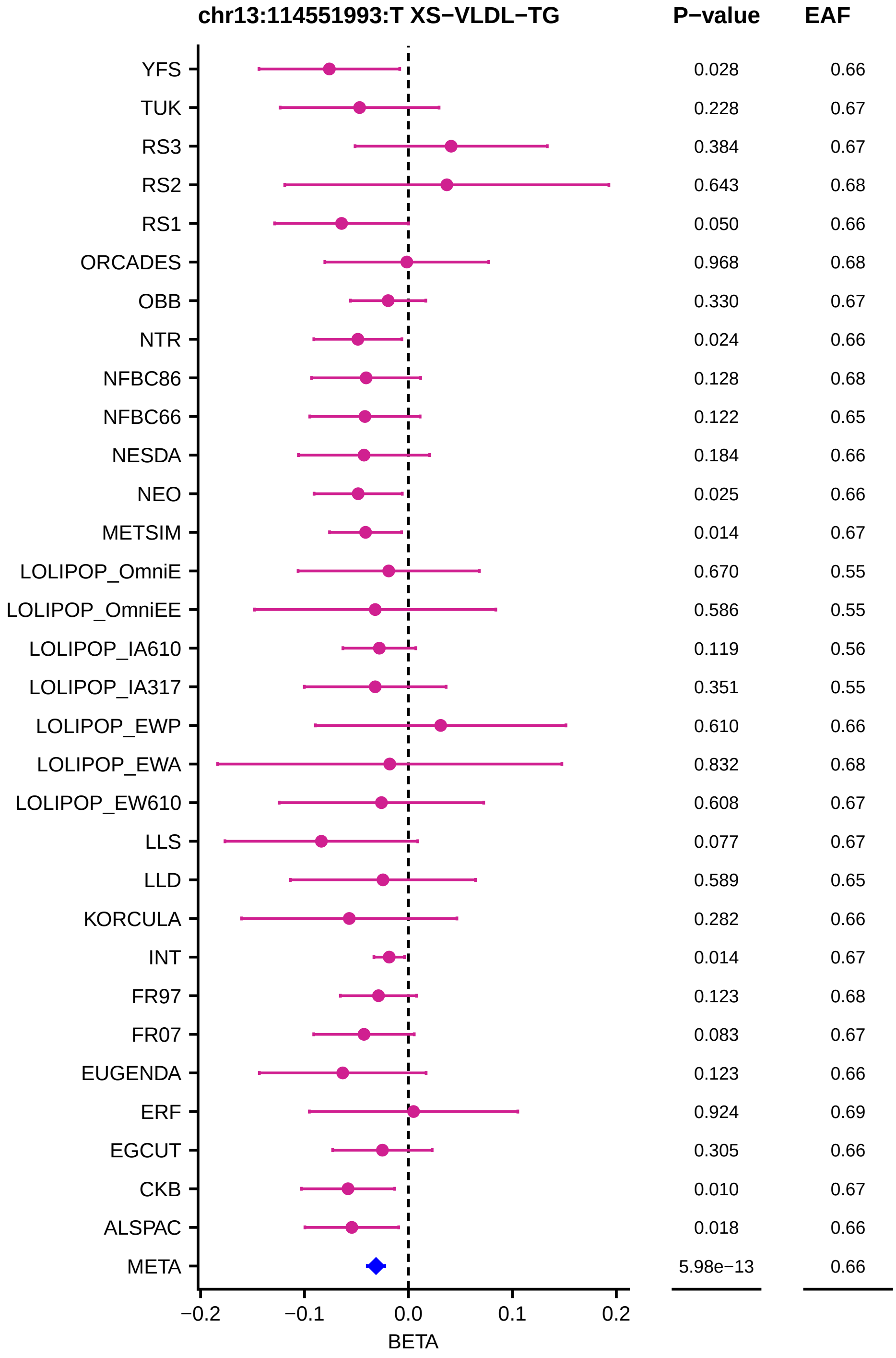
chr13:114551993:T XS−VLDL−TG
P−value
EAF
YFS
0.028
0.66
TUK
0.228
0.67
RS3
0.384
0.67
RS2
0.643
0.68
RS1
0.050
0.66
ORCADES
0.968
0.68
OBB
0.330
0.67
NTR
0.024
0.66
NFBC86
0.128
0.68
NFBC66
0.122
0.65
NESDA
0.184
0.66
NEO
0.025
0.66
METSIM
0.014
0.67
LOLIPOP_OmniE
0.670
0.55
LOLIPOP_OmniEE
0.586
0.55
LOLIPOP_IA610
0.119
0.56
LOLIPOP_IA317
0.351
0.55
LOLIPOP_EWP
0.610
0.66
LOLIPOP_EWA
0.832
0.68
LOLIPOP_EW610
0.608
0.67
LLS
0.077
0.67
LLD
0.589
0.65
KORCULA
0.282
0.66
INT
0.014
0.67
FR97
0.123
0.68
FR07
0.083
0.67
EUGENDA
0.123
0.66
ERF
0.924
0.69
EGCUT
0.305
0.66
CKB
0.010
0.67
ALSPAC
0.018
0.66
META
5.98e−13
0.66
−0.2
−0.1
0.0
0.1
0.2
BETA

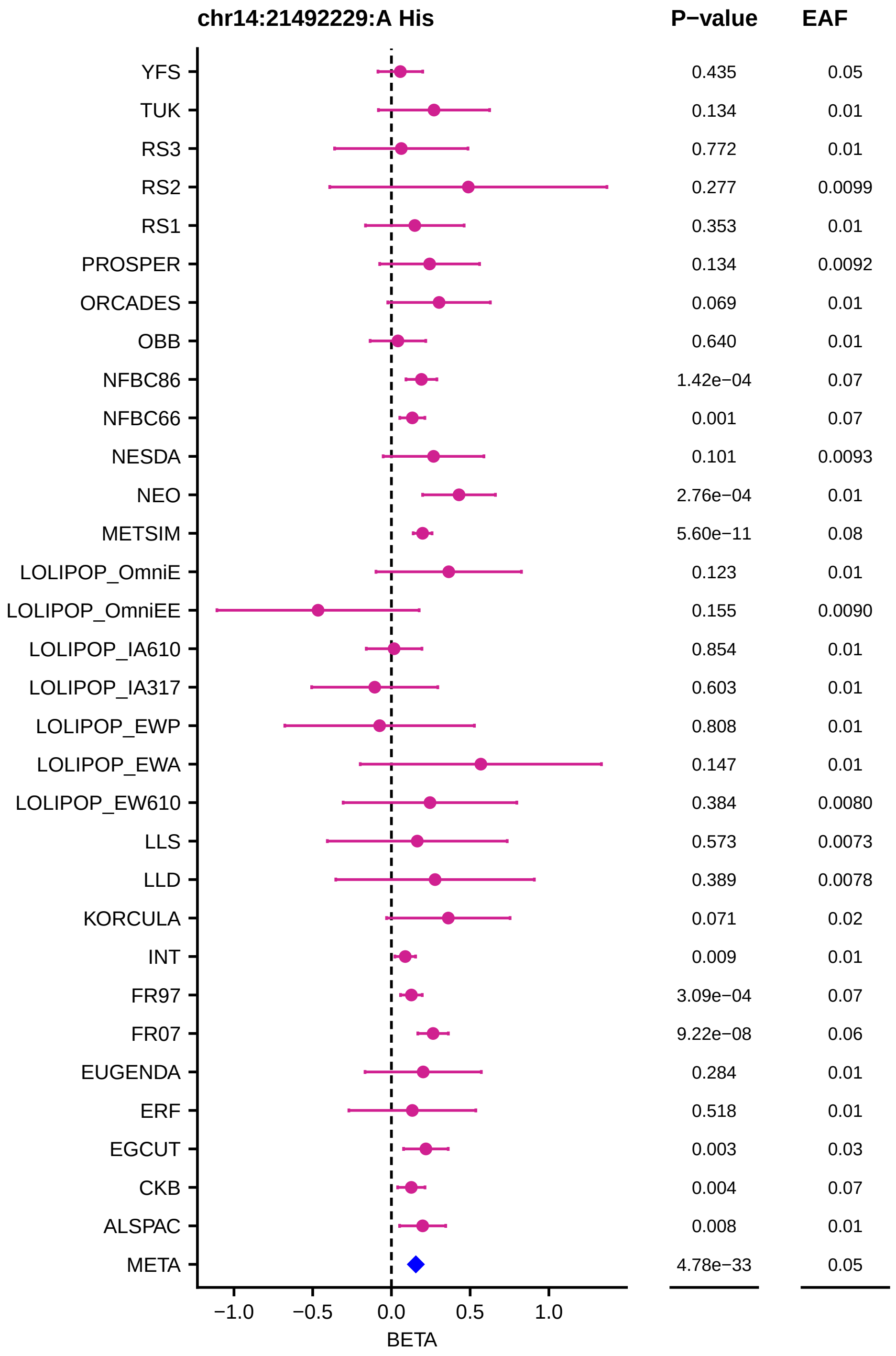
chr14:21492229:A His
P−value
EAF
YFS 0.435 0.05
TUK 0.134 0.01
RS3 0.772 0.01
RS2 0.277 0.0099
RS1 0.353 0.01
PROSPER 0.134 0.0092
ORCADES 0.069 0.01
OBB 0.640 0.01
NFBC86 1.42e−04 0.07
NFBC66 0.001 0.07
NESDA 0.101 0.0093
NEO 2.76e−04 0.01
METSIM 5.60e−11 0.08
LOLIPOP_OmniE 0.123 0.01
LOLIPOP_OmniEE 0.155 0.0090
LOLIPOP_IA610 0.854 0.01
LOLIPOP_IA317 0.603 0.01
LOLIPOP_EWP 0.808 0.01
LOLIPOP_EWA 0.147 0.01
LOLIPOP_EW610 0.384 0.0080
LLS 0.573 0.0073
LLD 0.389 0.0078
KORCULA 0.071 0.02
INT 0.009 0.01
FR97 3.09e−04 0.07
FR07 9.22e−08 0.06
EUGENDA 0.284 0.01
ERF 0.518 0.01
EGCUT 0.003 0.03
CKB 0.004 0.07
ALSPAC 0.008 0.01
META 4.78e−33 0.05
−1.0
−0.5
0.0
0.5
1.0
BETA

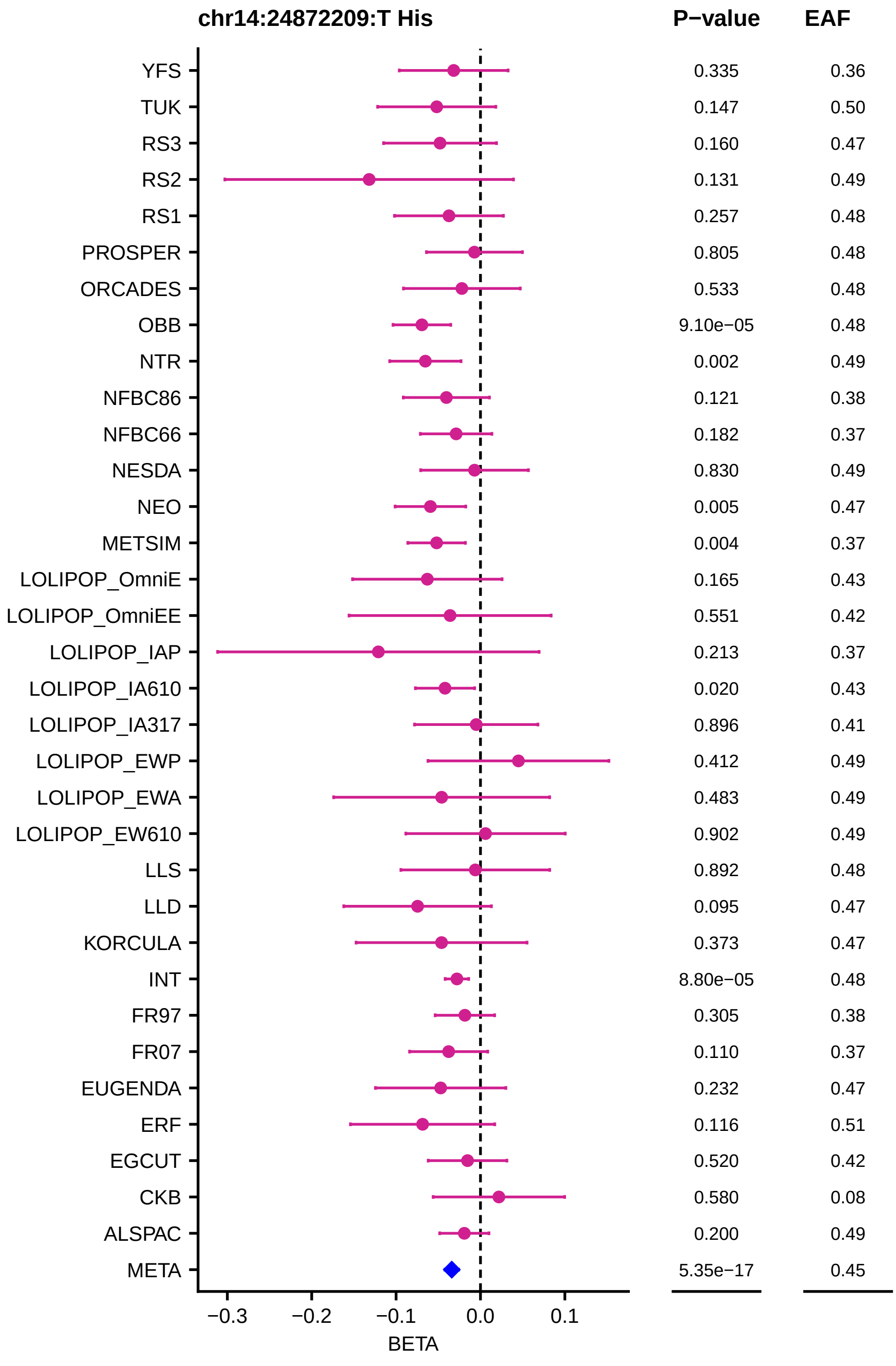
chr14:24872209:T His
P−value
EAF
YFS 0.335 0.36
TUK 0.147 0.50
RS3 0.160 0.47
RS2 0.131 0.49
RS1 0.257 0.48
PROSPER 0.805 0.48
ORCADES 0.533 0.48
OBB 9.10e−05 0.48
NTR 0.002 0.49
NFBC86 0.121 0.38
NFBC66 0.182 0.37
NESDA 0.830 0.49
NEO 0.005 0.47
METSIM 0.004 0.37
LOLIPOP_OmniE 0.165 0.43
LOLIPOP_OmniEE 0.551 0.42
LOLIPOP_IAP 0.213 0.37
LOLIPOP_IA610 0.020 0.43
LOLIPOP_IA317 0.896 0.41
LOLIPOP_EWP 0.412 0.49
LOLIPOP_EWA 0.483 0.49
LOLIPOP_EW610 0.902 0.49
LLS 0.892 0.48
LLD 0.095 0.47
KORCULA 0.373 0.47
INT 8.80e−05 0.48
FR97 0.305 0.38
FR07 0.110 0.37
EUGENDA 0.232 0.47
ERF 0.116 0.51
EGCUT 0.520 0.42
CKB 0.580 0.08
ALSPAC 0.200 0.49
META 5.35e−17 0.45
−0.3
−0.2
−0.1
0.0
0.1
BETA

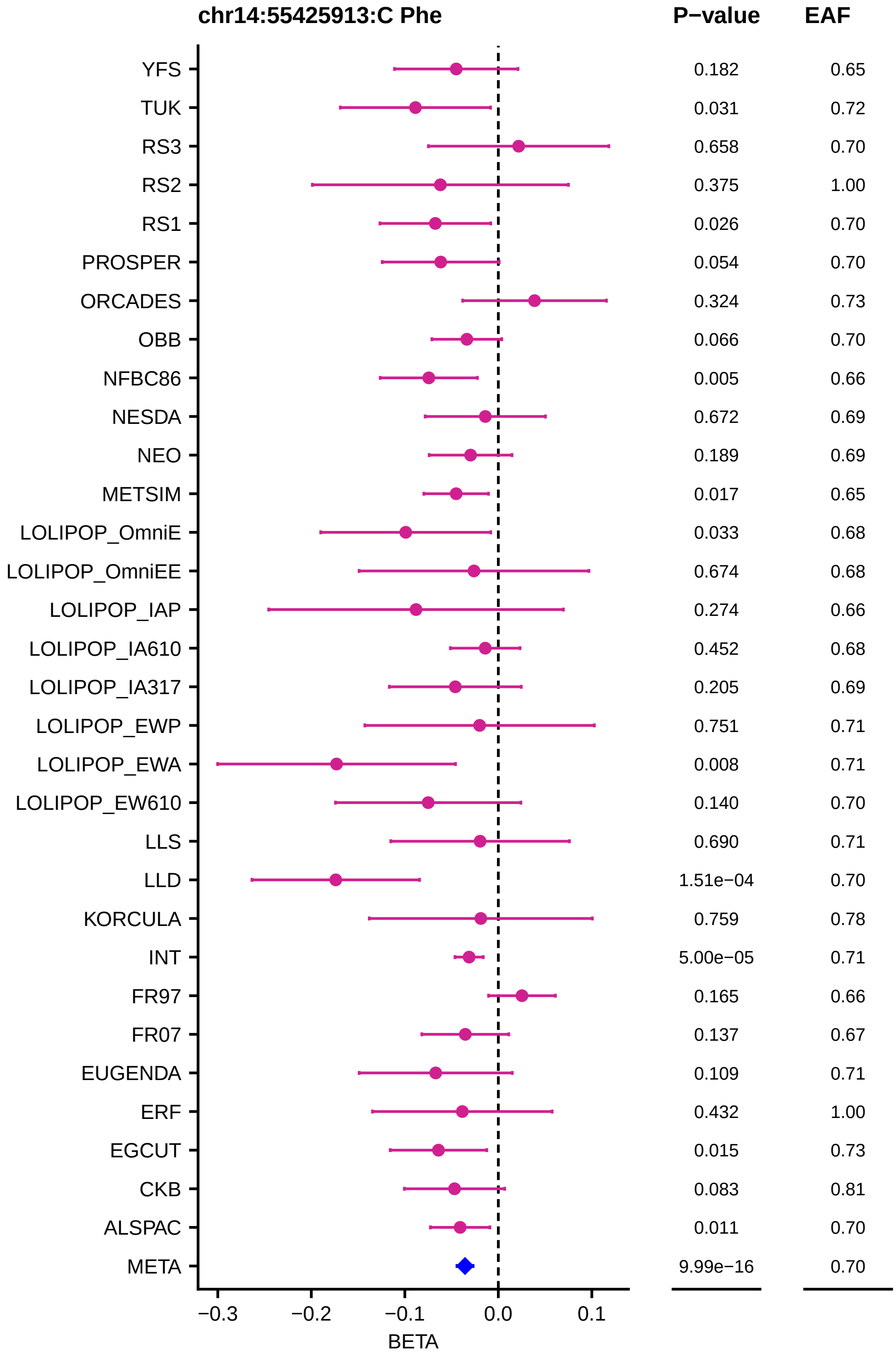
chr14:55425913:C Phe
P−value
EAF
YFS 0.182 0.65
TUK 0.031 0.72
RS3 0.658 0.70
RS2 0.375 1.00
RS1 0.026 0.70
PROSPER 0.054 0.70
ORCADES 0.324 0.73
OBB 0.066 0.70
NFBC86 0.005 0.66
NESDA 0.672 0.69
NEO 0.189 0.69
METSIM 0.017 0.65
LOLIPOP_OmniE 0.033 0.68
LOLIPOP_OmniEE 0.674 0.68
LOLIPOP_IAP 0.274 0.66
LOLIPOP_IA610 0.452 0.68
LOLIPOP_IA317 0.205 0.69
LOLIPOP_EWP 0.751 0.71
LOLIPOP_EWA 0.008 0.71
LOLIPOP_EW610 0.140 0.70
LLS 0.690 0.71
LLD 1.51e−04 0.70
KORCULA 0.759 0.78
INT 5.00e−05 0.71
FR97 0.165 0.66
FR07 0.137 0.67
EUGENDA 0.109 0.71
ERF 0.432 1.00
EGCUT 0.015 0.73
CKB 0.083 0.81
ALSPAC 0.011 0.70
META 9.99e−16 0.70
−0.3
−0.2
−0.1
0.0
0.1
BETA

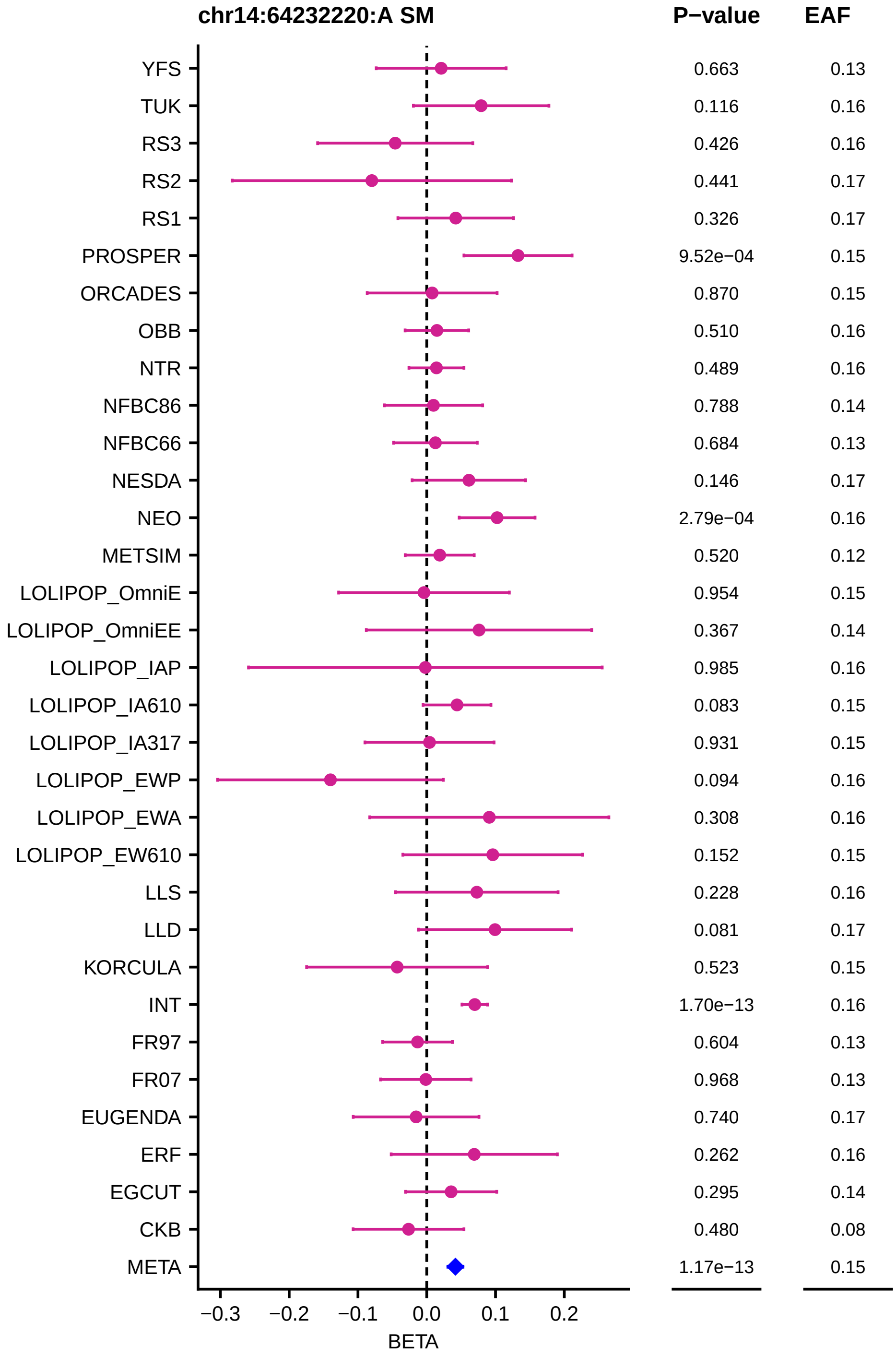

chr14:64232220:A SM
P−value
EAF
YFS 0.663 0.13
TUK 0.116 0.16
RS3 0.426 0.16
RS2 0.441 0.17
RS1 0.326 0.17
PROSPER 9.52e−04 0.15
ORCADES 0.870 0.15
OBB 0.510 0.16
NTR 0.489 0.16
NFBC86 0.788 0.14
NFBC66 0.684 0.13
NESDA 0.146 0.17
NEO 2.79e−04 0.16
METSIM 0.520 0.12
LOLIPOP_OmniE 0.954 0.15
LOLIPOP_OmniEE 0.367 0.14
LOLIPOP_IAP 0.985 0.16
LOLIPOP_IA610 0.083 0.15
LOLIPOP_IA317 0.931 0.15
LOLIPOP_EWP 0.094 0.16
LOLIPOP_EWA 0.308 0.16
LOLIPOP_EW610 0.152 0.15
LLS 0.228 0.16
LLD 0.081 0.17
KORCULA 0.523 0.15
INT 1.70e−13 0.16
FR97 0.604 0.13
FR07 0.968 0.13
EUGENDA 0.740 0.17
ERF 0.262 0.16
EGCUT 0.295 0.14
CKB 0.480 0.08
META 1.17e−13 0.15
−0.3 −0.2 −0.1 0.0 0.1 0.2
BETA

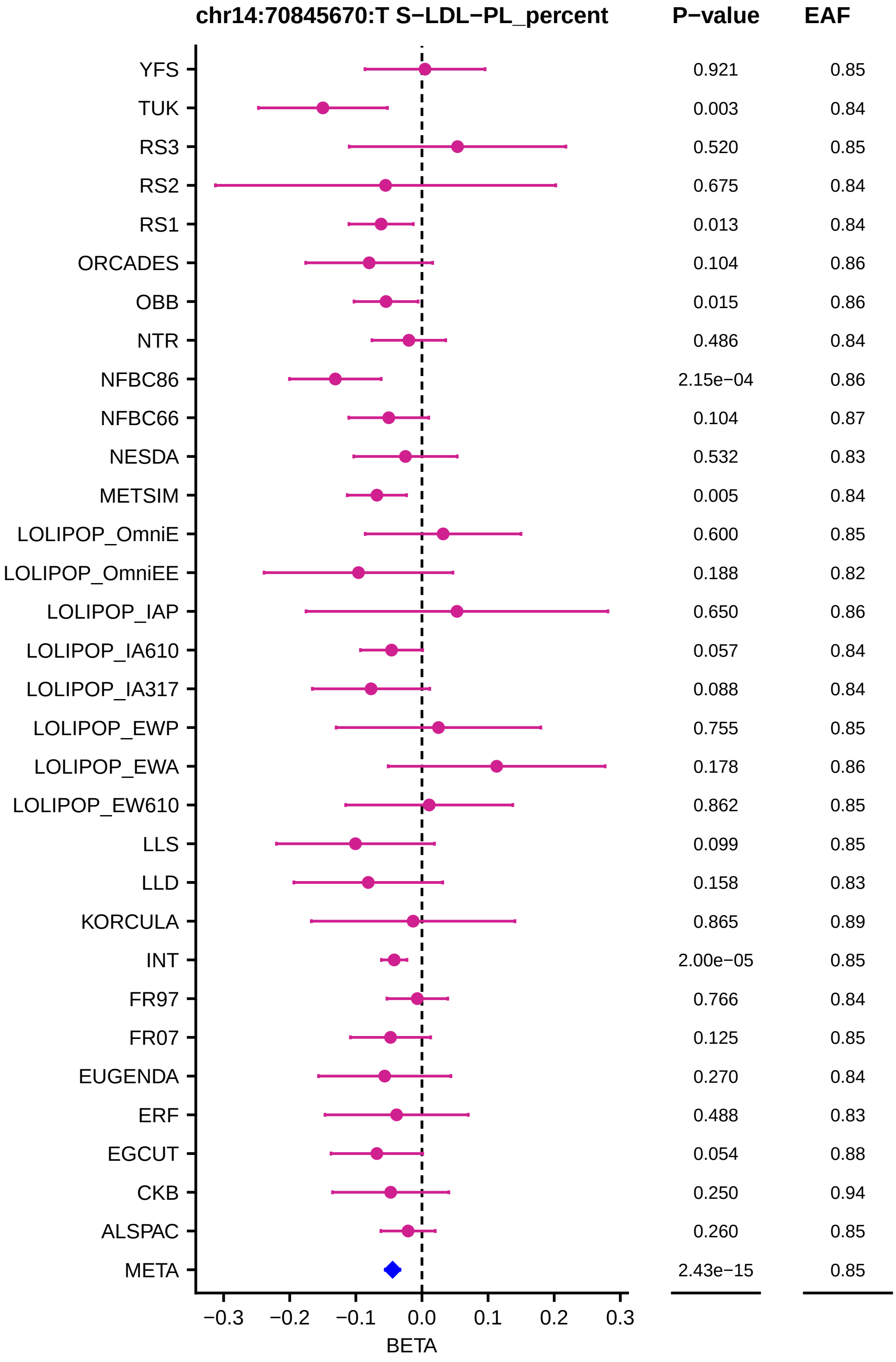
chr14:70845670:T S−LDL−PL_percent
P−value
EAF
YFS 0.921 0.85
TUK 0.003 0.84
RS3 0.520 0.85
RS2 0.675 0.84
RS1 0.013 0.84
ORCADES 0.104 0.86
OBB 0.015 0.86
NTR 0.486 0.84
NFBC86 2.15e−04 0.86
NFBC66 0.104 0.87
NESDA 0.532 0.83
METSIM 0.005 0.84
LOLIPOP_OmniE 0.600 0.85
LOLIPOP_OmniEE 0.188 0.82
LOLIPOP_IAP 0.650 0.86
LOLIPOP_IA610 0.057 0.84
LOLIPOP_IA317 0.088 0.84
LOLIPOP_EWP 0.755 0.85
LOLIPOP_EWA 0.178 0.86
LOLIPOP_EW610 0.862 0.85
LLS 0.099 0.85
LLD 0.158 0.83
KORCULA 0.865 0.89
INT 2.00e−05 0.85
FR97 0.766 0.84
FR07 0.125 0.85
EUGENDA 0.270 0.84
ERF 0.488 0.83
EGCUT 0.054 0.88
CKB 0.250 0.94
ALSPAC 0.260 0.85
META 2.43e−15 0.85
−0.3 −0.2 −0.1 0.0 0.1 0.2 0.3
BETA

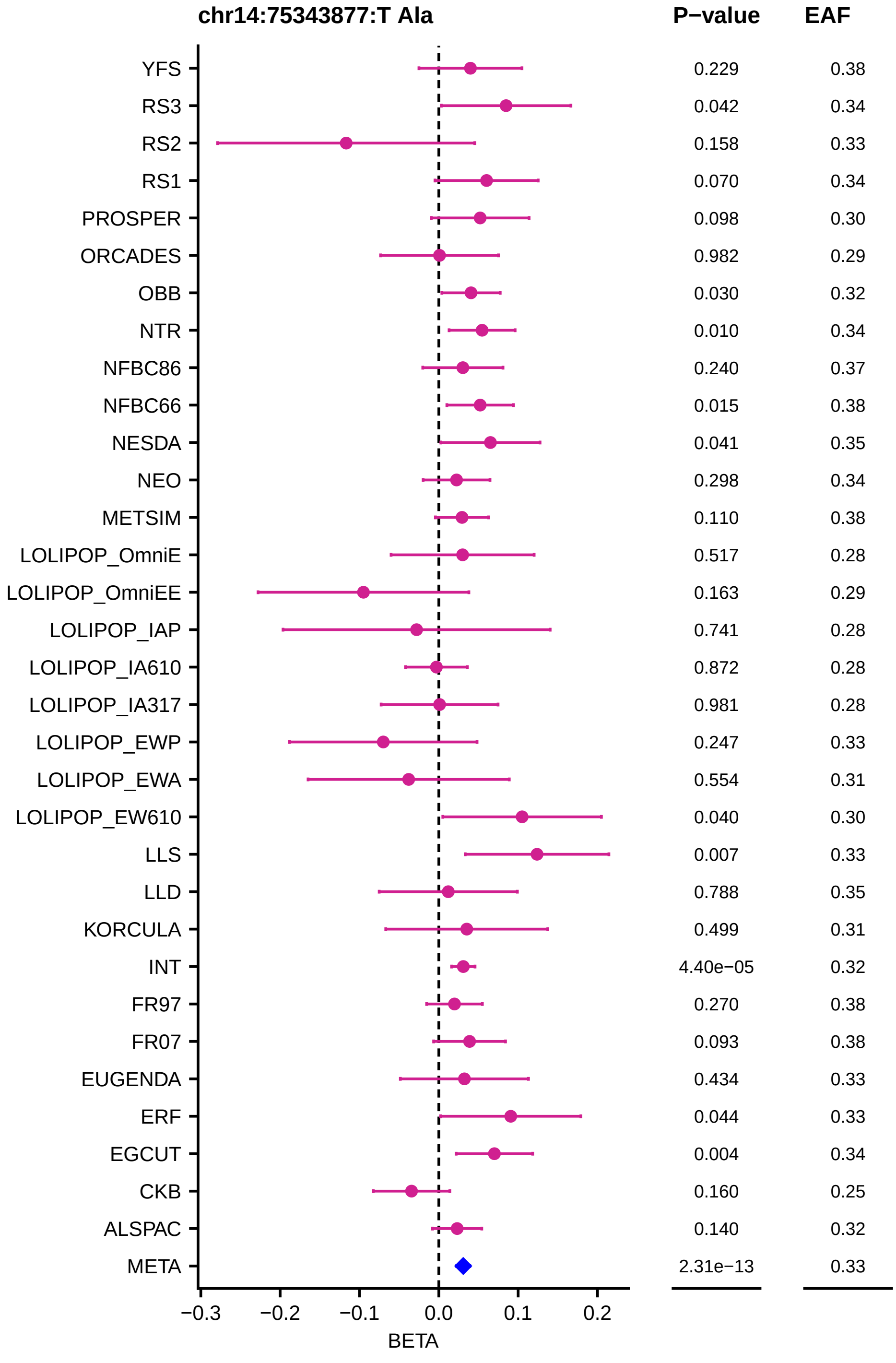
chr14:75343877:T Ala
P−value
EAF
YFS 0.229 0.38
RS3 0.042 0.34
RS2 0.158 0.33
RS1 0.070 0.34
PROSPER 0.098 0.30
ORCADES 0.982 0.29
OBB 0.030 0.32
NTR 0.010 0.34
NFBC86 0.240 0.37
NFBC66 0.015 0.38
NESDA 0.041 0.35
NEO 0.298 0.34
METSIM 0.110 0.38
LOLIPOP_OmniE 0.517 0.28
LOLIPOP_OmniEE 0.163 0.29
LOLIPOP_IAP 0.741 0.28
LOLIPOP_IA610 0.872 0.28
LOLIPOP_IA317 0.981 0.28
LOLIPOP_EWP 0.247 0.33
LOLIPOP_EWA 0.554 0.31
LOLIPOP_EW610 0.040 0.30
LLS 0.007 0.33
LLD 0.788 0.35
KORCULA 0.499 0.31
INT 4.40e−05 0.32
FR97 0.270 0.38
FR07 0.093 0.38
EUGENDA 0.434 0.33
ERF 0.044 0.33
EGCUT 0.004 0.34
CKB 0.160 0.25
ALSPAC 0.140 0.32
META 2.31e−13 0.33
−0.3 −0.2 −0.1 0.0 0.1 0.2
BETA

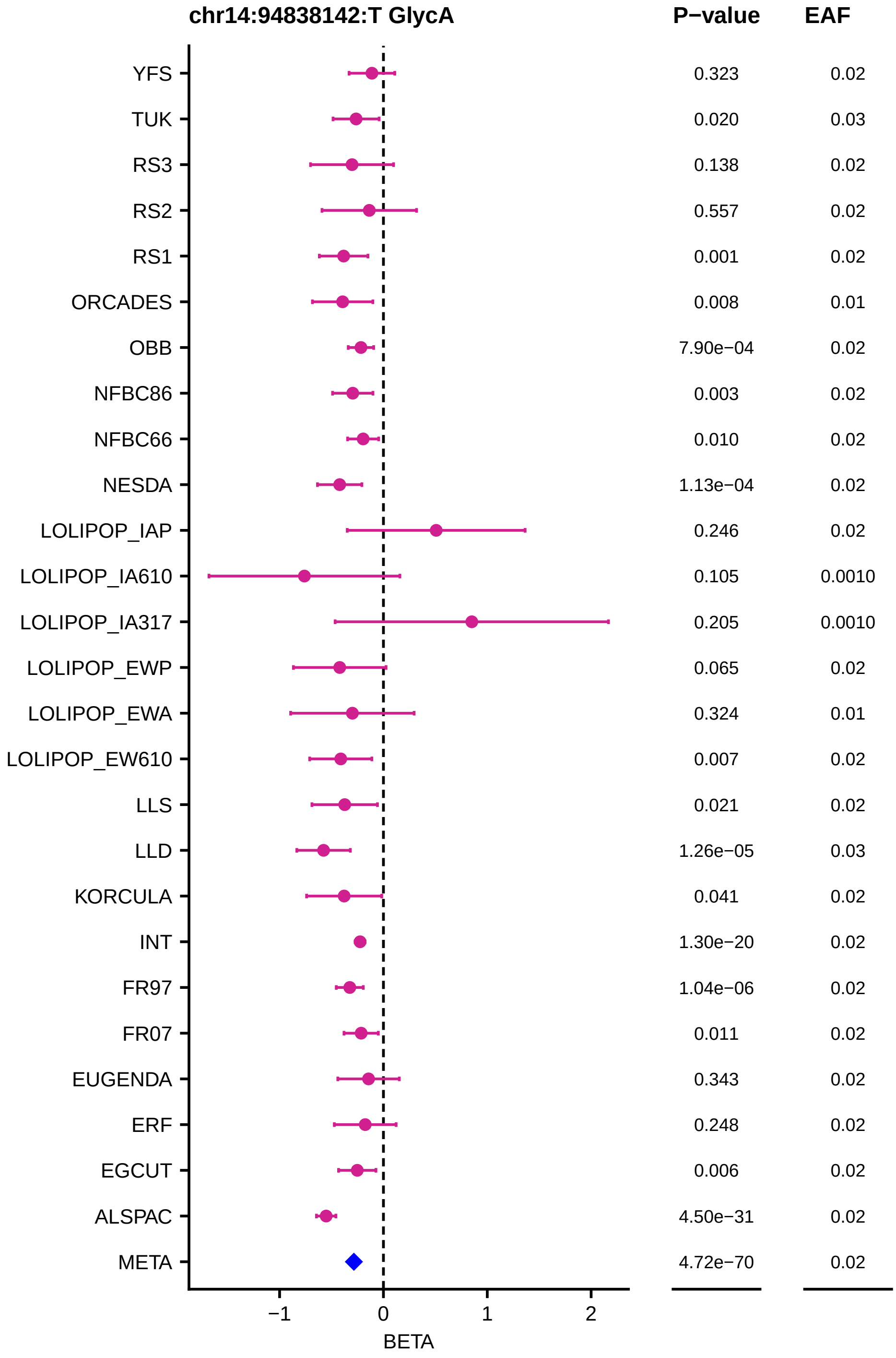
chr14:94838142:T GlycA
P−value
EAF
YFS 0.323 0.02
TUK 0.020 0.03
RS3 0.138 0.02
RS2 0.557 0.02
RS1 0.001 0.02
ORCADES 0.008 0.01
OBB 7.90e−04 0.02
NFBC86 0.003 0.02
NFBC66 0.010 0.02
NESDA 1.13e−04 0.02
LOLIPOP_IAP 0.246 0.02
LOLIPOP_IA610 0.105 0.0010
LOLIPOP_IA317 0.205 0.0010
LOLIPOP_EWP 0.065 0.02
LOLIPOP_EWA 0.324 0.01
LOLIPOP_EW610 0.007 0.02
LLS 0.021 0.02
LLD 1.26e−05 0.03
KORCULA 0.041 0.02
INT 1.30e−20 0.02
FR97 1.04e−06 0.02
FR07 0.011 0.02
EUGENDA 0.343 0.02
ERF 0.248 0.02
EGCUT 0.006 0.02
ALSPAC 4.50e−31 0.02
META 4.72e−70 0.02
−1
0
1
2
BETA

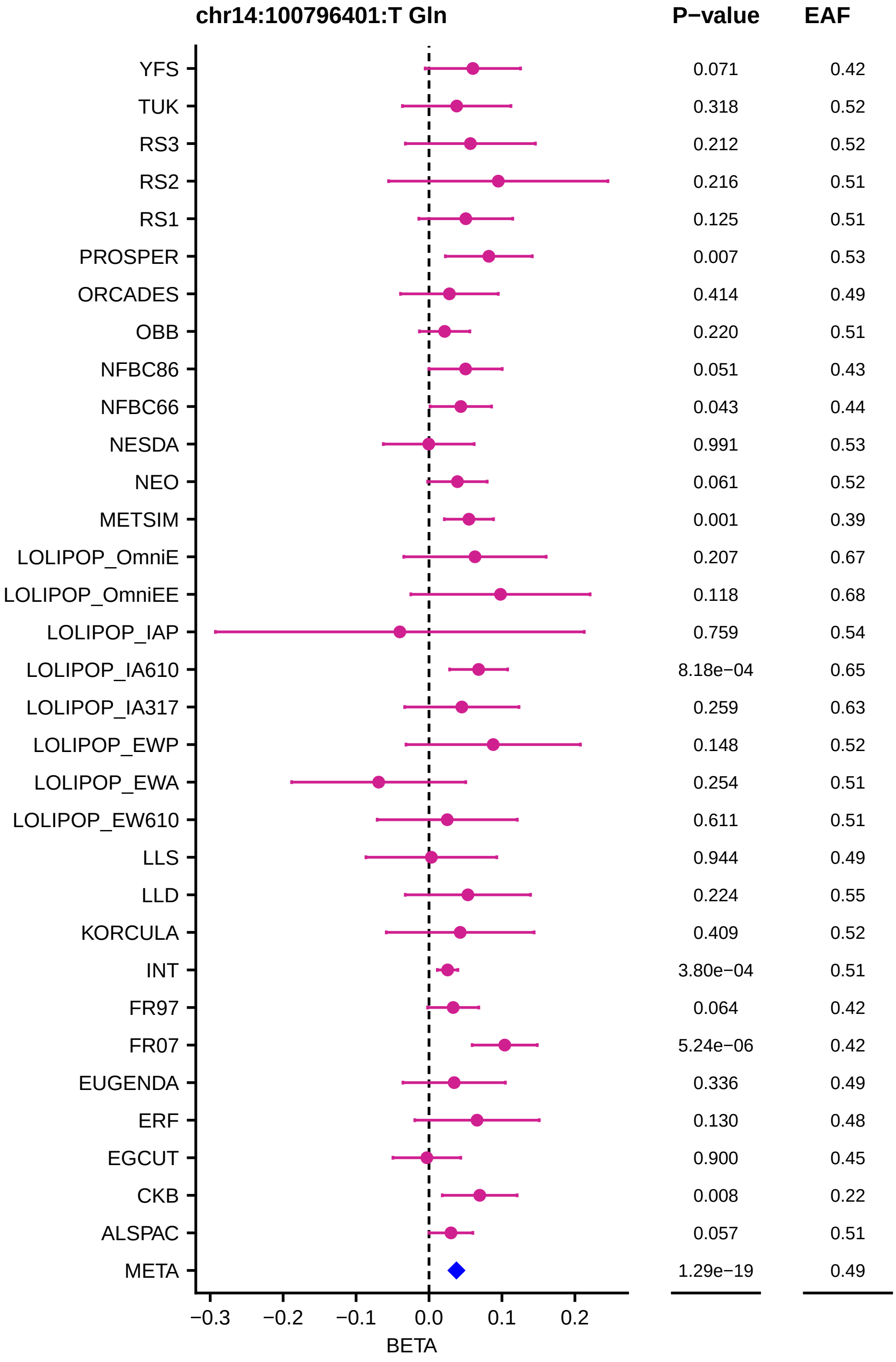

chr14:100796401:T Gln
P−value
EAF
YFS 0.071 0.42
TUK 0.318 0.52
RS3 0.212 0.52
RS2 0.216 0.51
RS1 0.125 0.51
PROSPER 0.007 0.53
ORCADES 0.414 0.49
OBB 0.220 0.51
NFBC86 0.051 0.43
NFBC66 0.043 0.44
NESDA 0.991 0.53
NEO 0.061 0.52
METSIM 0.001 0.39
LOLIPOP_OmniE 0.207 0.67
LOLIPOP_OmniEE 0.118 0.68
LOLIPOP_IAP 0.759 0.54
LOLIPOP_IA610 8.18e−04 0.65
LOLIPOP_IA317 0.259 0.63
LOLIPOP_EWP 0.148 0.52
LOLIPOP_EWA 0.254 0.51
LOLIPOP_EW610 0.611 0.51
LLS 0.944 0.49
LLD 0.224 0.55
KORCULA 0.409 0.52
INT 3.80e−04 0.51
FR97 0.064 0.42
FR07 5.24e−06 0.42
EUGENDA 0.336 0.49
ERF 0.130 0.48
EGCUT 0.900 0.45
CKB 0.008 0.22
ALSPAC 0.057 0.51
META 1.29e−19 0.49
−0.3 −0.2 −0.1 0.0 0.1 0.2
BETA

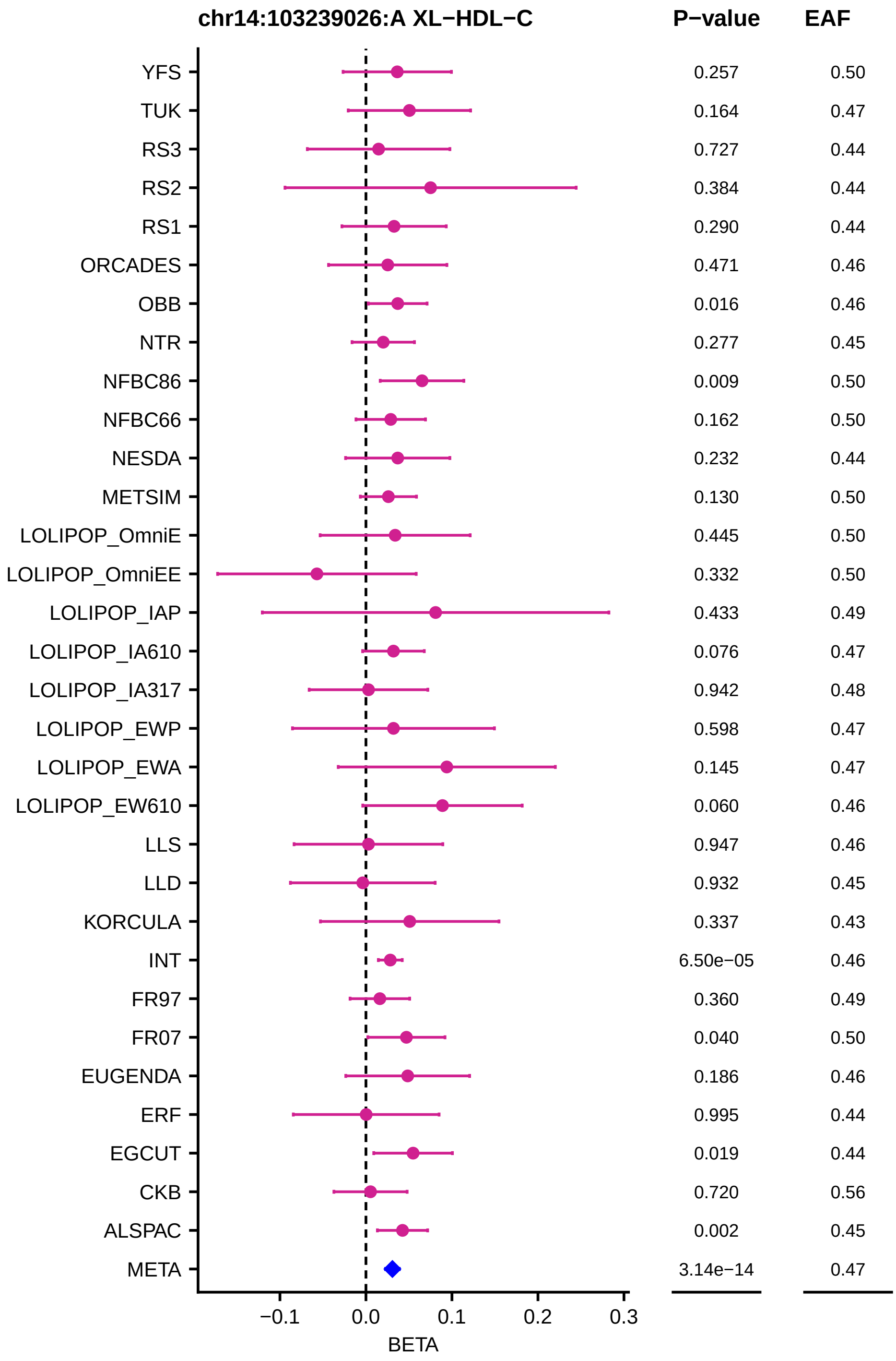
chr14:103239026:A XL−HDL−C
P−value
EAF
YFS 0.257 0.50
TUK 0.164 0.47
RS3 0.727 0.44
RS2 0.384 0.44
RS1 0.290 0.44
ORCADES 0.471 0.46
OBB 0.016 0.46
NTR 0.277 0.45
NFBC86 0.009 0.50
NFBC66 0.162 0.50
NESDA 0.232 0.44
METSIM 0.130 0.50
LOLIPOP_OmniE 0.445 0.50
LOLIPOP_OmniEE 0.332 0.50
LOLIPOP_IAP 0.433 0.49
LOLIPOP_IA610 0.076 0.47
LOLIPOP_IA317 0.942 0.48
LOLIPOP_EWP 0.598 0.47
LOLIPOP_EWA 0.145 0.47
LOLIPOP_EW610 0.060 0.46
LLS 0.947 0.46
LLD 0.932 0.45
KORCULA 0.337 0.43
INT 6.50e−05 0.46
FR97 0.360 0.49
FR07 0.040 0.50
EUGENDA 0.186 0.46
ERF 0.995 0.44
EGCUT 0.019 0.44
CKB 0.720 0.56
ALSPAC 0.002 0.45
META 3.14e−14 0.47
−0.1
0.0
0.1
0.2
0.3
BETA

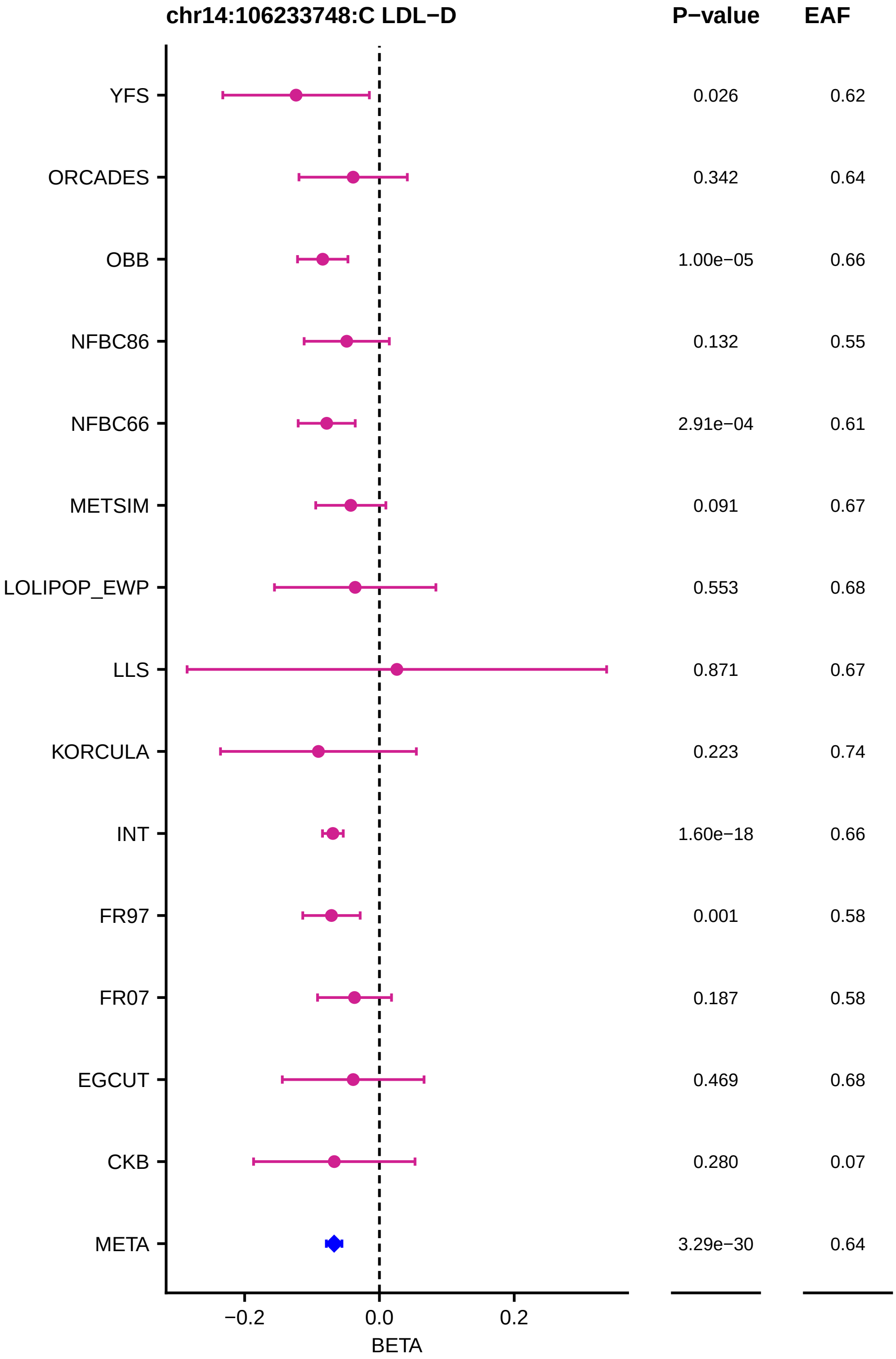
chr14:106233748:C LDL−D
P−value
EAF
YFS 0.026 0.62
ORCADES 0.342 0.64
OBB 1.00e−05 0.66
NFBC86 0.132 0.55
NFBC66 2.91e−04 0.61
METSIM 0.091 0.67
LOLIPOP_EWP 0.553 0.68
LLS 0.871 0.67
KORCULA 0.223 0.74
INT 1.60e−18 0.66
FR97 0.001 0.58
FR07 0.187 0.58
EGCUT 0.469 0.68
CKB 0.280 0.07
META 3.29e−30 0.64
−0.2
0.0
0.2
BETA

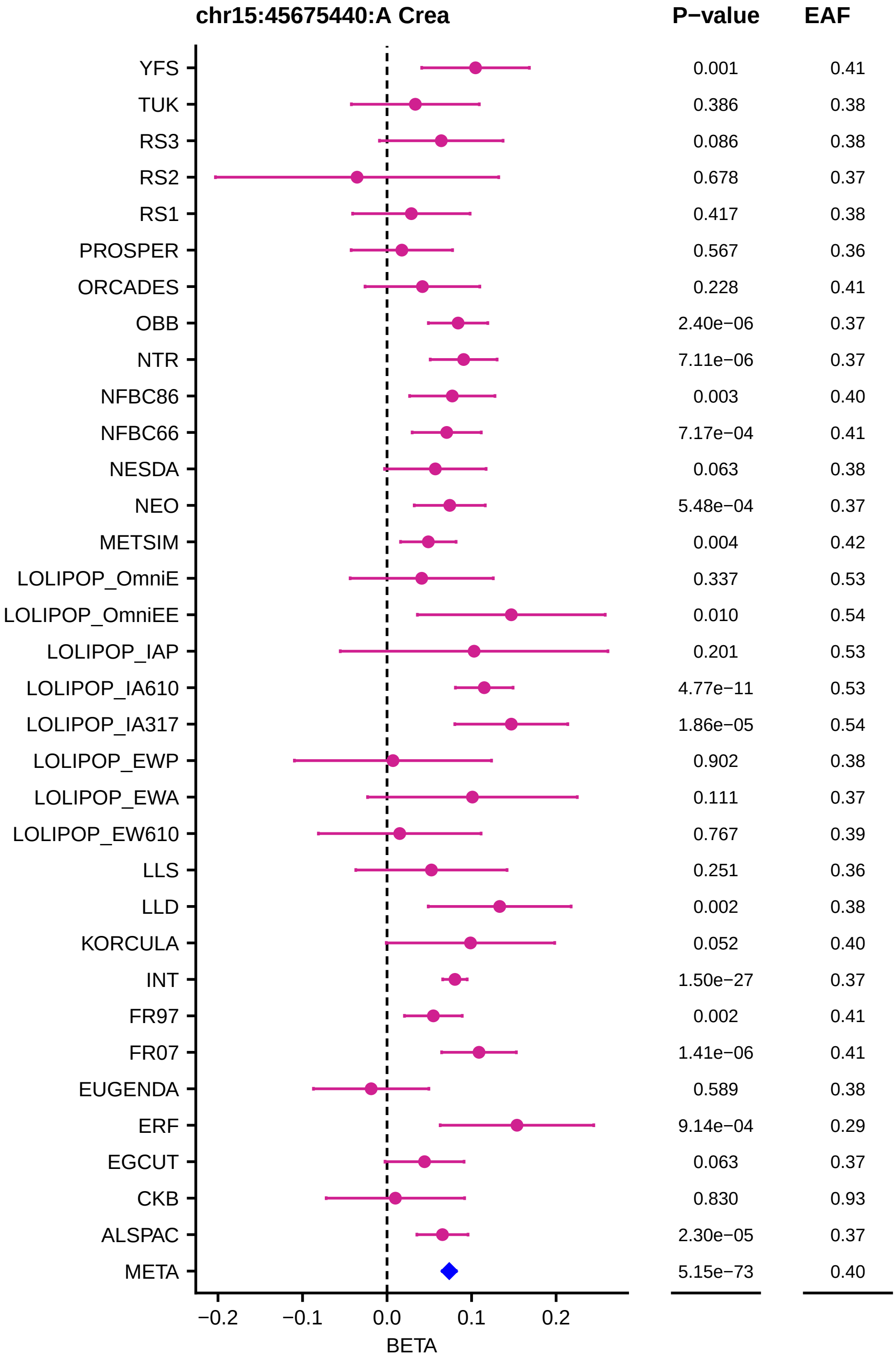
chr15:45675440:A Crea
P−value
EAF
YFS 0.001 0.41
TUK 0.386 0.38
RS3 0.086 0.38
RS2 0.678 0.37
RS1 0.417 0.38
PROSPER 0.567 0.36
ORCADES 0.228 0.41
OBB 2.40e−06 0.37
NTR 7.11e−06 0.37
NFBC86 0.003 0.40
NFBC66 7.17e−04 0.41
NESDA 0.063 0.38
NEO 5.48e−04 0.37
METSIM 0.004 0.42
LOLIPOP_OmniE 0.337 0.53
LOLIPOP_OmniEE 0.010 0.54
LOLIPOP_IAP 0.201 0.53
LOLIPOP_IA610 4.77e−11 0.53
LOLIPOP_IA317 1.86e−05 0.54
LOLIPOP_EWP 0.902 0.38
LOLIPOP_EWA 0.111 0.37
LOLIPOP_EW610 0.767 0.39
LLS 0.251 0.36
LLD 0.002 0.38
KORCULA 0.052 0.40
INT 1.50e−27 0.37
FR97 0.002 0.41
FR07 1.41e−06 0.41
EUGENDA 0.589 0.38
ERF 9.14e−04 0.29
EGCUT 0.063 0.37
CKB 0.830 0.93
ALSPAC 2.30e−05 0.37
META 5.15e−73 0.40
−0.2
−0.1
0.0
0.1
0.2
BETA

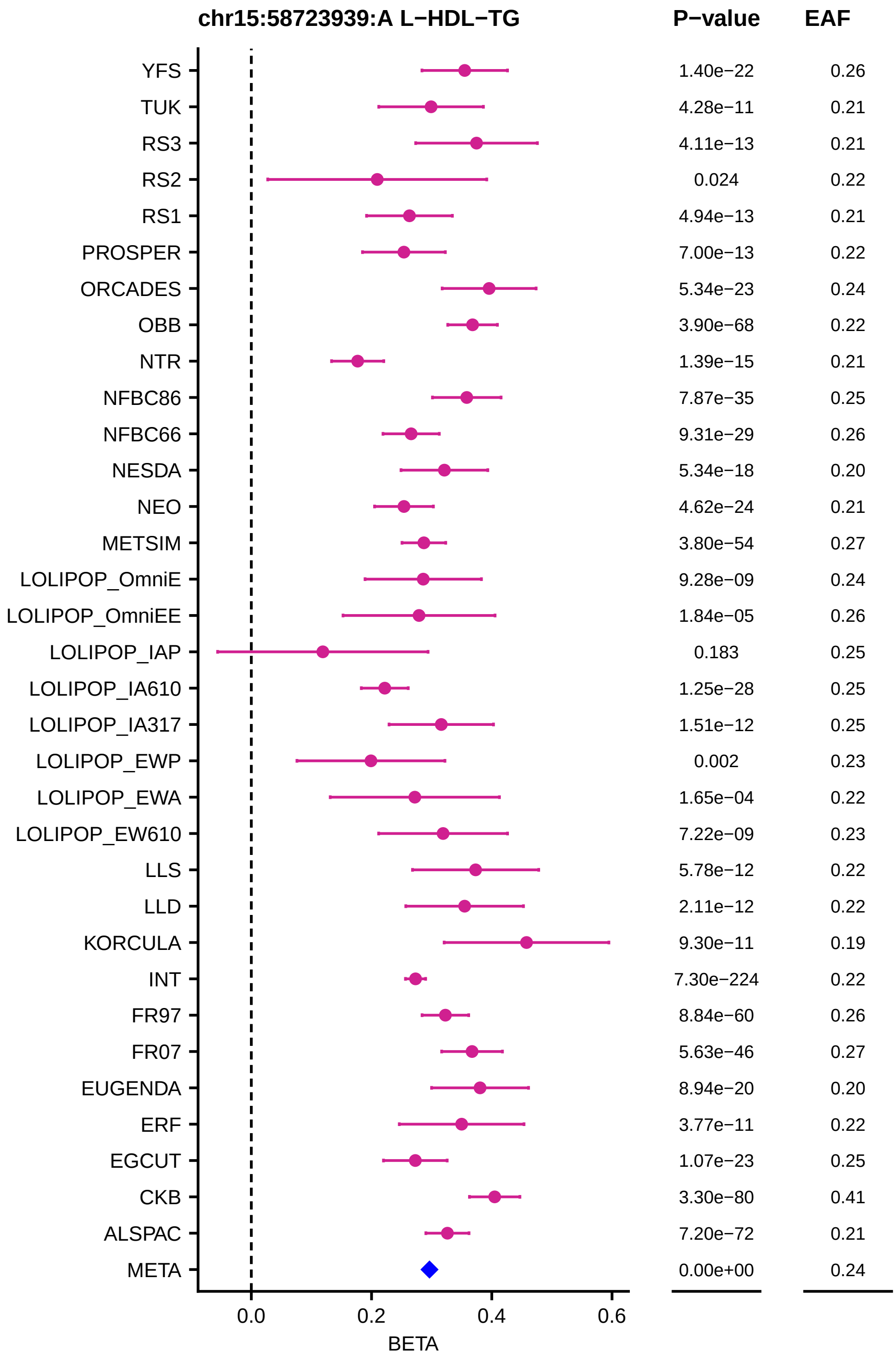
chr15:58723939:A L−HDL−TG
P−value
EAF
YFS 1.40e−22 0.26
TUK 4.28e−11 0.21
RS3 4.11e−13 0.21
RS2 0.024 0.22
RS1 4.94e−13 0.21
PROSPER 7.00e−13 0.22
ORCADES 5.34e−23 0.24
OBB 3.90e−68 0.22
NTR 1.39e−15 0.21
NFBC86 7.87e−35 0.25
NFBC66 9.31e−29 0.26
NESDA 5.34e−18 0.20
NEO 4.62e−24 0.21
METSIM 3.80e−54 0.27
LOLIPOP_OmniE 9.28e−09 0.24
LOLIPOP_OmniEE 1.84e−05 0.26
LOLIPOP_IAP 0.183 0.25
LOLIPOP_IA610 1.25e−28 0.25
LOLIPOP_IA317 1.51e−12 0.25
LOLIPOP_EWP 0.002 0.23
LOLIPOP_EWA 1.65e−04 0.22
LOLIPOP_EW610 7.22e−09 0.23
LLS 5.78e−12 0.22
LLD 2.11e−12 0.22
KORCULA 9.30e−11 0.19
INT 7.30e−224 0.22
FR97 8.84e−60 0.26
FR07 5.63e−46 0.27
EUGENDA 8.94e−20 0.20
ERF 3.77e−11 0.22
EGCUT 1.07e−23 0.25
CKB 3.30e−80 0.41
ALSPAC 7.20e−72 0.21
META 0.00e+00 0.24
0.0
0.2
0.4
0.6
BETA

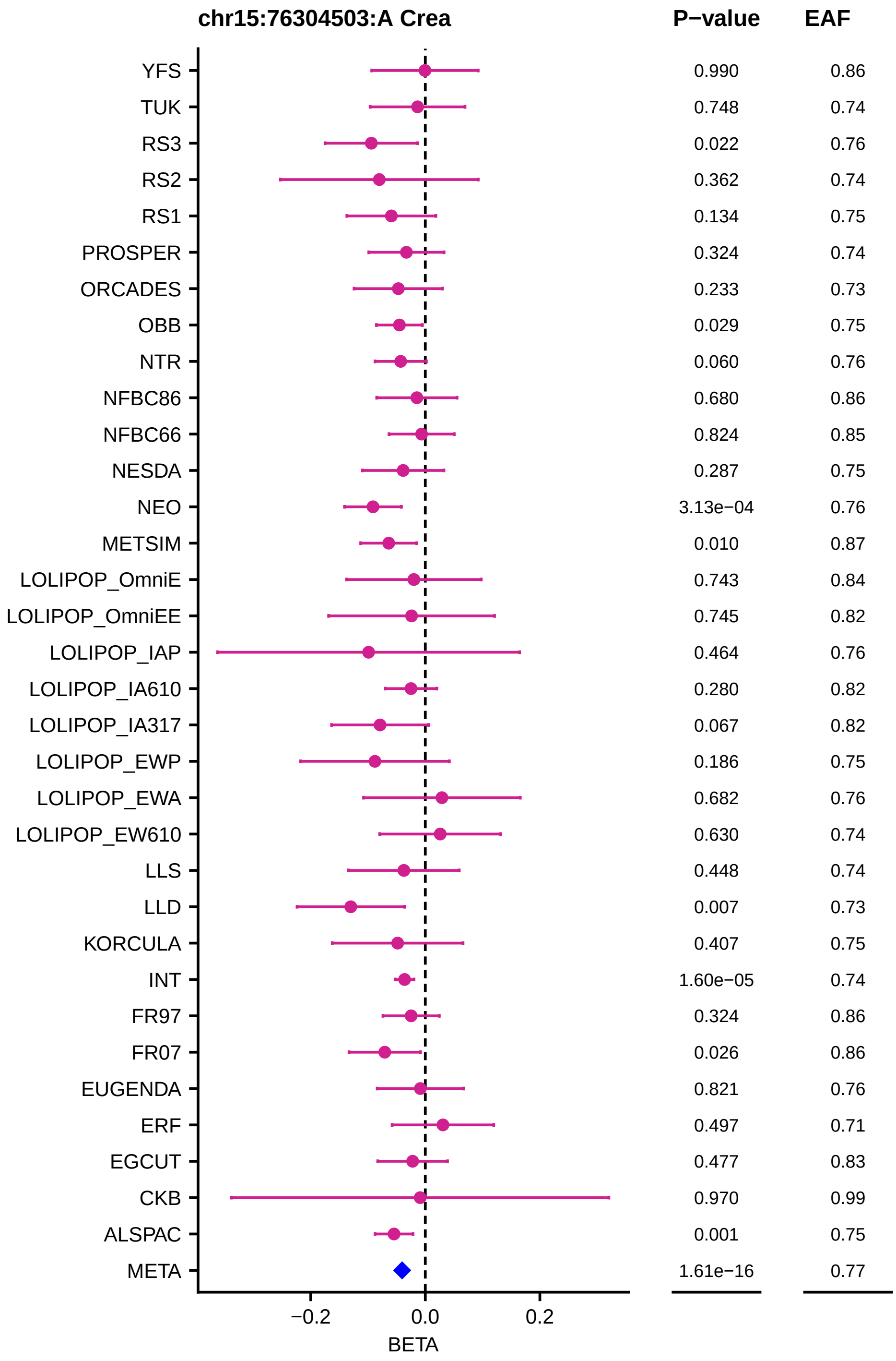
chr15:76304503:A Crea
P−value
EAF
YFS 0.990 0.86
TUK 0.748 0.74
RS3 0.022 0.76
RS2 0.362 0.74
RS1 0.134 0.75
PROSPER 0.324 0.74
ORCADES 0.233 0.73
OBB 0.029 0.75
NTR 0.060 0.76
NFBC86 0.680 0.86
NFBC66 0.824 0.85
NESDA 0.287 0.75
NEO 3.13e−04 0.76
METSIM 0.010 0.87
LOLIPOP_OmniE 0.743 0.84
LOLIPOP_OmniEE 0.745 0.82
LOLIPOP_IAP 0.464 0.76
LOLIPOP_IA610 0.280 0.82
LOLIPOP_IA317 0.067 0.82
LOLIPOP_EWP 0.186 0.75
LOLIPOP_EWA 0.682 0.76
LOLIPOP_EW610 0.630 0.74
LLS 0.448 0.74
LLD 0.007 0.73
KORCULA 0.407 0.75
INT 1.60e−05 0.74
FR97 0.324 0.86
FR07 0.026 0.86
EUGENDA 0.821 0.76
ERF 0.497 0.71
EGCUT 0.477 0.83
CKB 0.970 0.99
ALSPAC 0.001 0.75
META 1.61e−16 0.77
−0.2
0.0
0.2
BETA

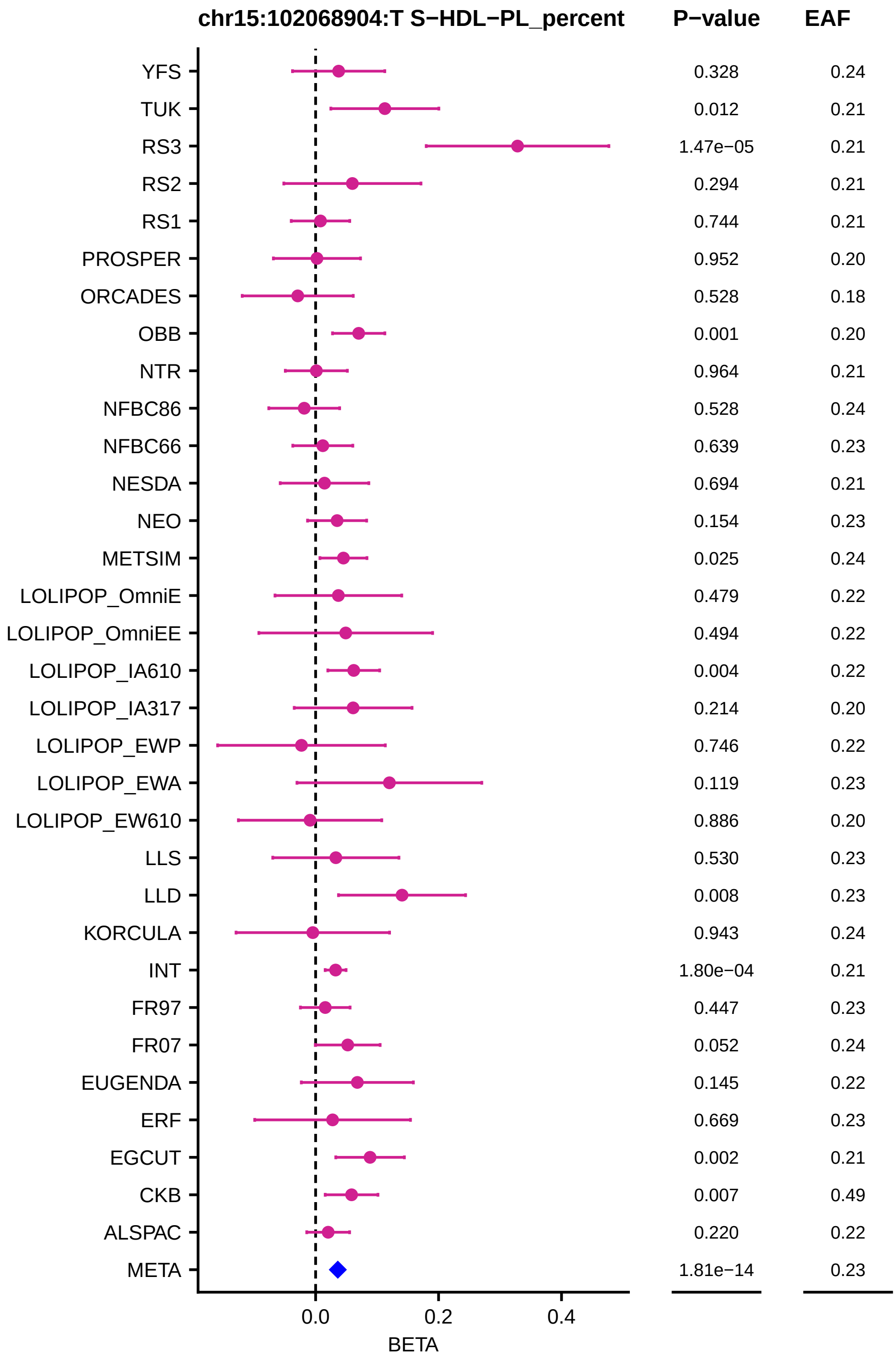

chr15:102068904:T S−HDL−PL_percent
P−value
EAF
YFS
TUK
RS3
RS2
RS1
PROSPER
ORCADES
OBB
NTR
NFBC86
NFBC66
NESDA
NEO
METSIM
LOLIPOP_OmniE
LOLIPOP_OmniEE
LOLIPOP_IA610
LOLIPOP_IA317
LOLIPOP_EWP
LOLIPOP_EWA
LOLIPOP_EW610
LLS
LLD
KORCULA
INT
FR97
FR07
EUGENDA
ERF
EGCUT
CKB
ALSPAC
META
0.328
0.012
1.47e−05
0.294
0.744
0.952
0.528
0.001
0.964
0.528
0.639
0.694
0.154
0.025
0.479
0.494
0.004
0.214
0.746
0.119
0.886
0.530
0.008
0.943
1.80e−04
0.447
0.052
0.145
0.669
0.002
0.007
0.220
1.81e−14
0.24
0.21
0.21
0.21
0.21
0.20
0.18
0.20
0.21
0.24
0.23
0.21
0.23
0.24
0.22
0.22
0.22
0.20
0.22
0.23
0.20
0.23
0.23
0.24
0.21
0.23
0.24
0.22
0.23
0.21
0.49
0.22
0.23
0.0
0.2
0.4
BETA

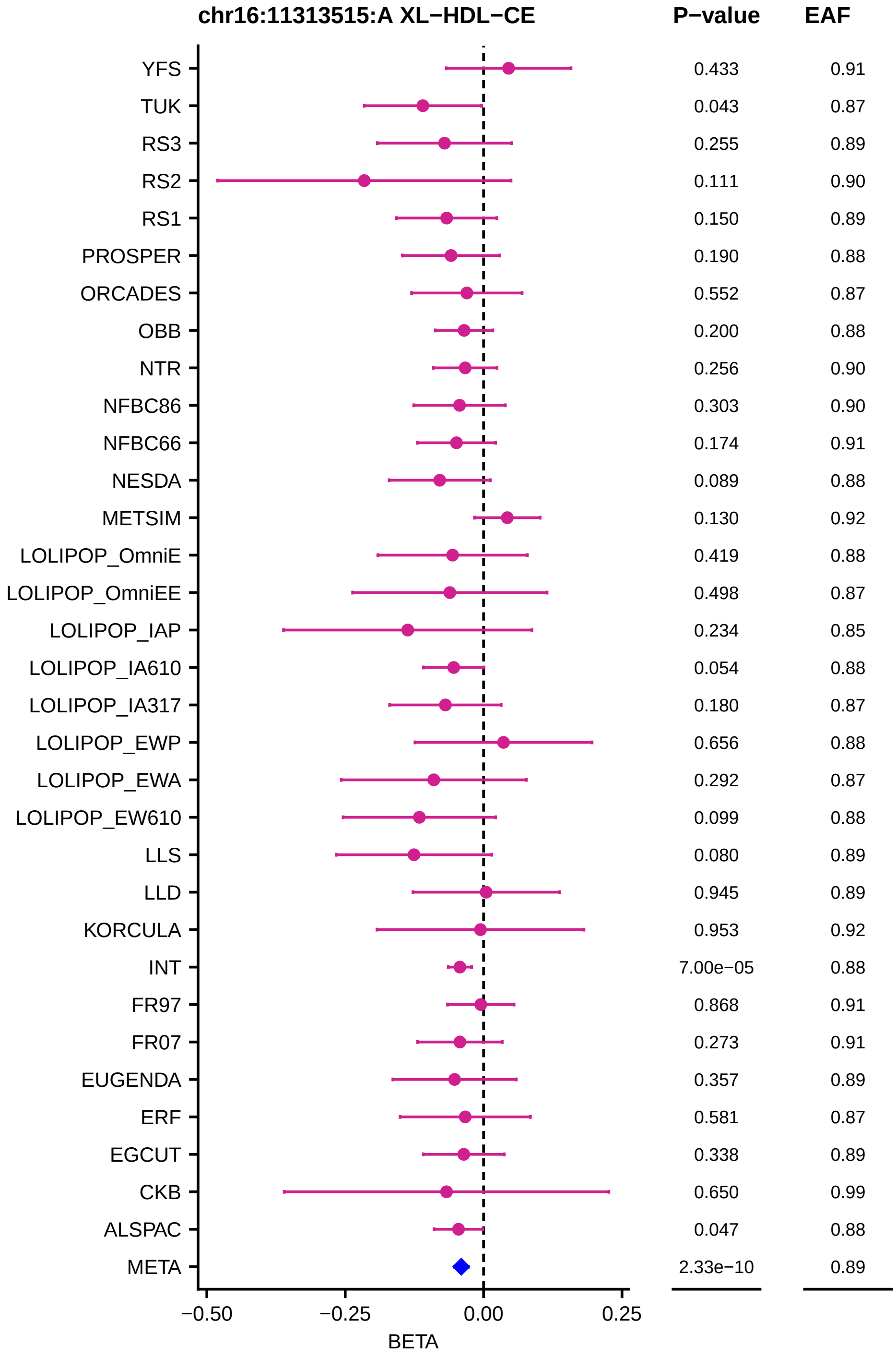
chr16:11313515:A XL−HDL−CE
P−value
EAF
YFS 0.433 0.91
TUK 0.043 0.87
RS3 0.255 0.89
RS2 0.111 0.90
RS1 0.150 0.89
PROSPER 0.190 0.88
ORCADES 0.552 0.87
OBB 0.200 0.88
NTR 0.256 0.90
NFBC86 0.303 0.90
NFBC66 0.174 0.91
NESDA 0.089 0.88
METSIM 0.130 0.92
LOLIPOP_OmniE 0.419 0.88
LOLIPOP_OmniEE 0.498 0.87
LOLIPOP_IAP 0.234 0.85
LOLIPOP_IA610 0.054 0.88
LOLIPOP_IA317 0.180 0.87
LOLIPOP_EWP 0.656 0.88
LOLIPOP_EWA 0.292 0.87
LOLIPOP_EW610 0.099 0.88
LLS 0.080 0.89
LLD 0.945 0.89
KORCULA 0.953 0.92
INT 7.00e−05 0.88
FR97 0.868 0.91
FR07 0.273 0.91
EUGENDA 0.357 0.89
ERF 0.581 0.87
EGCUT 0.338 0.89
CKB 0.650 0.99
ALSPAC 0.047 0.88
META 2.33e−10 0.89
−0.50
−0.25
0.00
0.25
BETA

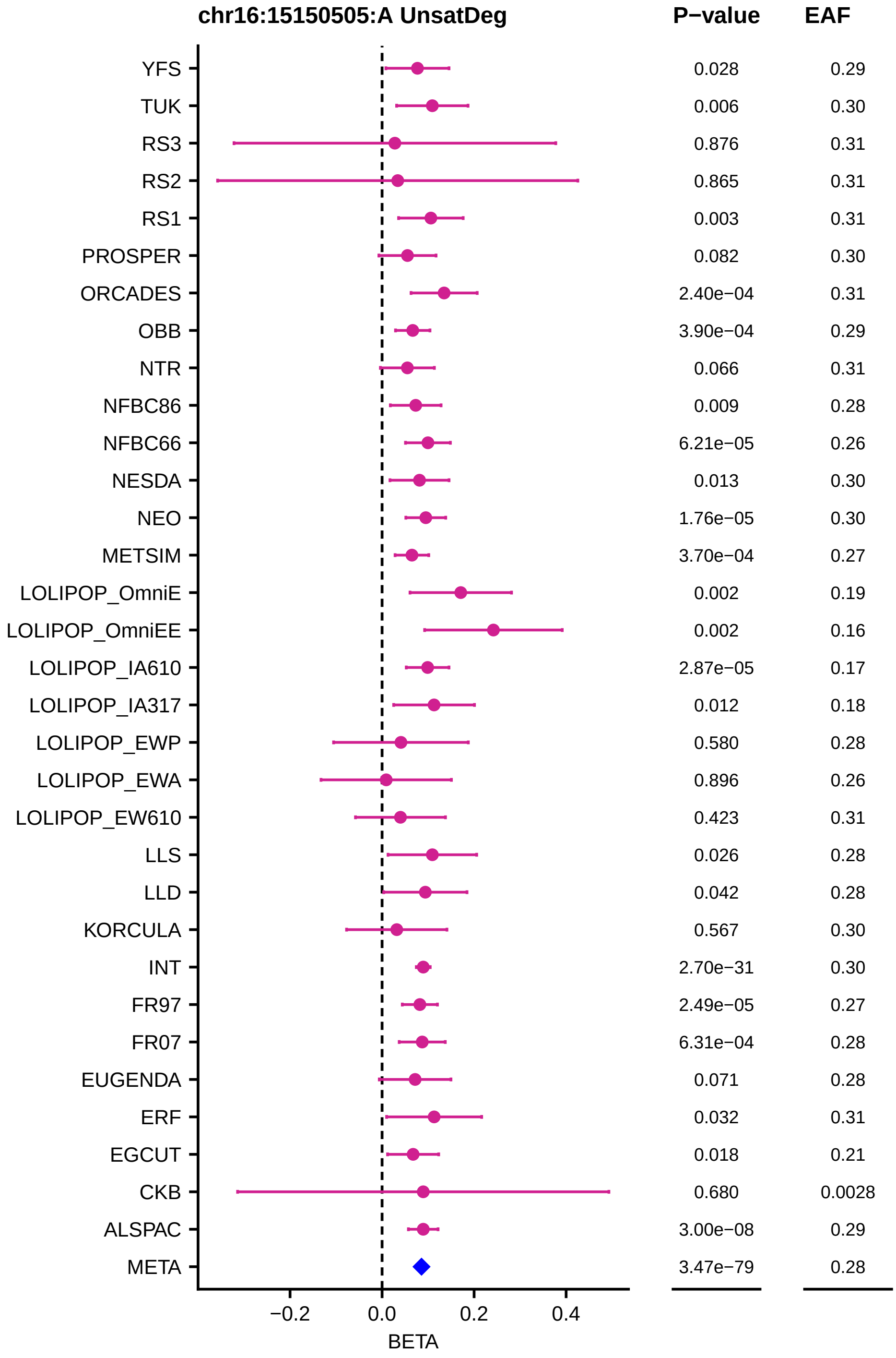
chr16:15150505:A UnsatDeg
P−value
EAF
YFS
TUK
RS3
RS2
RS1
PROSPER
ORCADES
OBB
NTR
NFBC86
NFBC66
NESDA
NEO
METSIM
LOLIPOP_OmniE
LOLIPOP_OmniEE
LOLIPOP_IA610
LOLIPOP_IA317
LOLIPOP_EWP
LOLIPOP_EWA
LOLIPOP_EW610
LLS
LLD
KORCULA
INT
FR97
FR07
EUGENDA
ERF
EGCUT
CKB
ALSPAC
META
0.028
0.006
0.876
0.865
0.003
0.082
2.40e−04
3.90e−04
0.066
0.009
6.21e−05
0.013
1.76e−05
3.70e−04
0.002
0.002
2.87e−05
0.012
0.580
0.896
0.423
0.026
0.042
0.567
2.70e−31
2.49e−05
6.31e−04
0.071
0.032
0.018
0.680
3.00e−08
3.47e−79
0.29
0.30
0.31
0.31
0.31
0.30
0.31
0.29
0.31
0.28
0.26
0.30
0.30
0.27
0.19
0.16
0.17
0.18
0.28
0.26
0.31
0.28
0.28
0.30
0.30
0.27
0.28
0.28
0.31
0.21
0.0028
0.29
0.28
−0.2
0.0
0.2
0.4
BETA

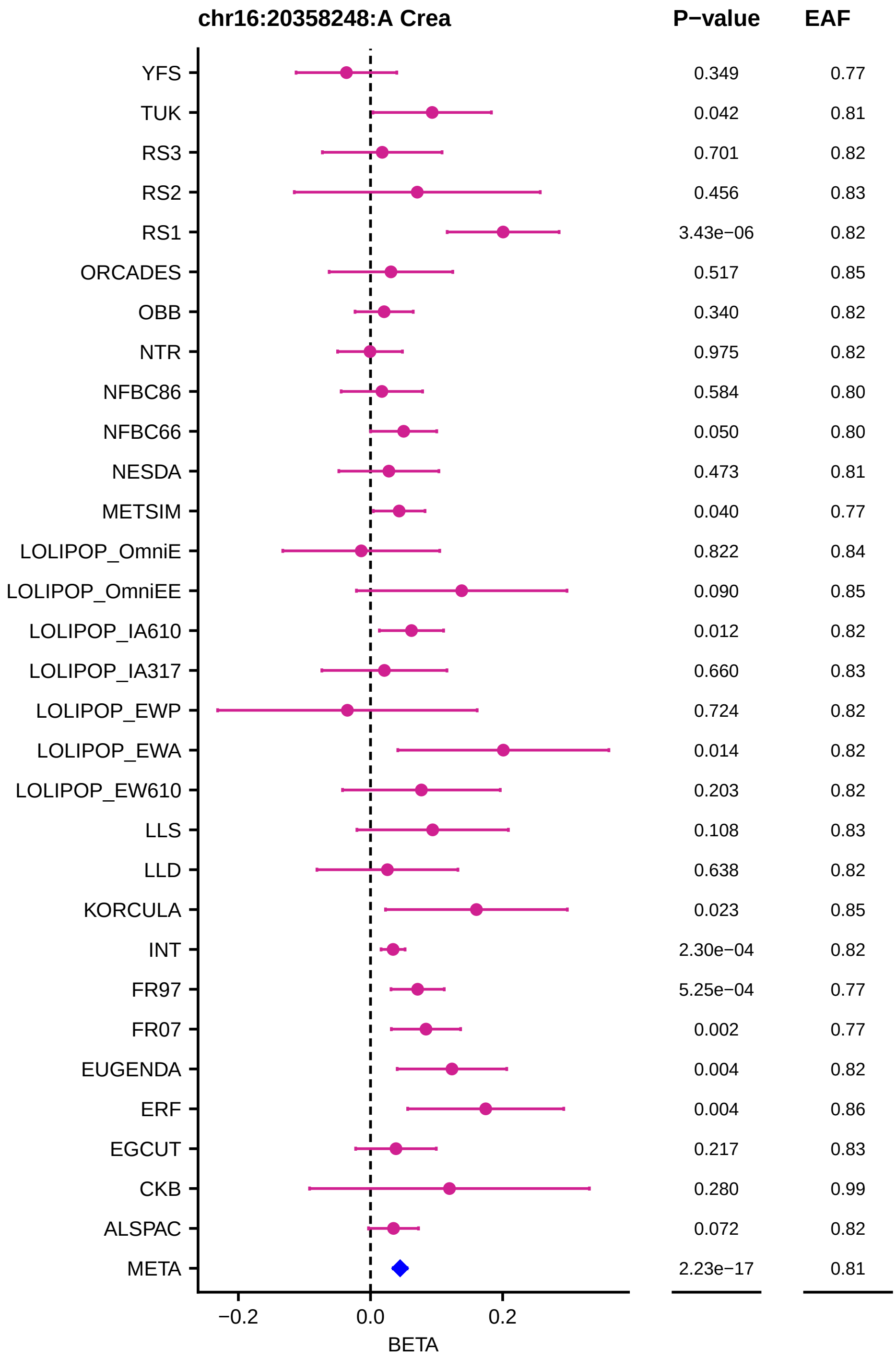
chr16:20358248:A Crea
P−value
EAF
YFS 0.349 0.77
TUK 0.042 0.81
RS3 0.701 0.82
RS2 0.456 0.83
RS1 3.43e−06 0.82
ORCADES 0.517 0.85
OBB 0.340 0.82
NTR 0.975 0.82
NFBC86 0.584 0.80
NFBC66 0.050 0.80
NESDA 0.473 0.81
METSIM 0.040 0.77
LOLIPOP_OmniE 0.822 0.84
LOLIPOP_OmniEE 0.090 0.85
LOLIPOP_IA610 0.012 0.82
LOLIPOP_IA317 0.660 0.83
LOLIPOP_EWP 0.724 0.82
LOLIPOP_EWA 0.014 0.82
LOLIPOP_EW610 0.203 0.82
LLS 0.108 0.83
LLD 0.638 0.82
KORCULA 0.023 0.85
INT 2.30e−04 0.82
FR97 5.25e−04 0.77
FR07 0.002 0.77
EUGENDA 0.004 0.82
ERF 0.004 0.86
EGCUT 0.217 0.83
CKB 0.280 0.99
ALSPAC 0.072 0.82
META 2.23e−17 0.81
−0.2
0.0
0.2
BETA

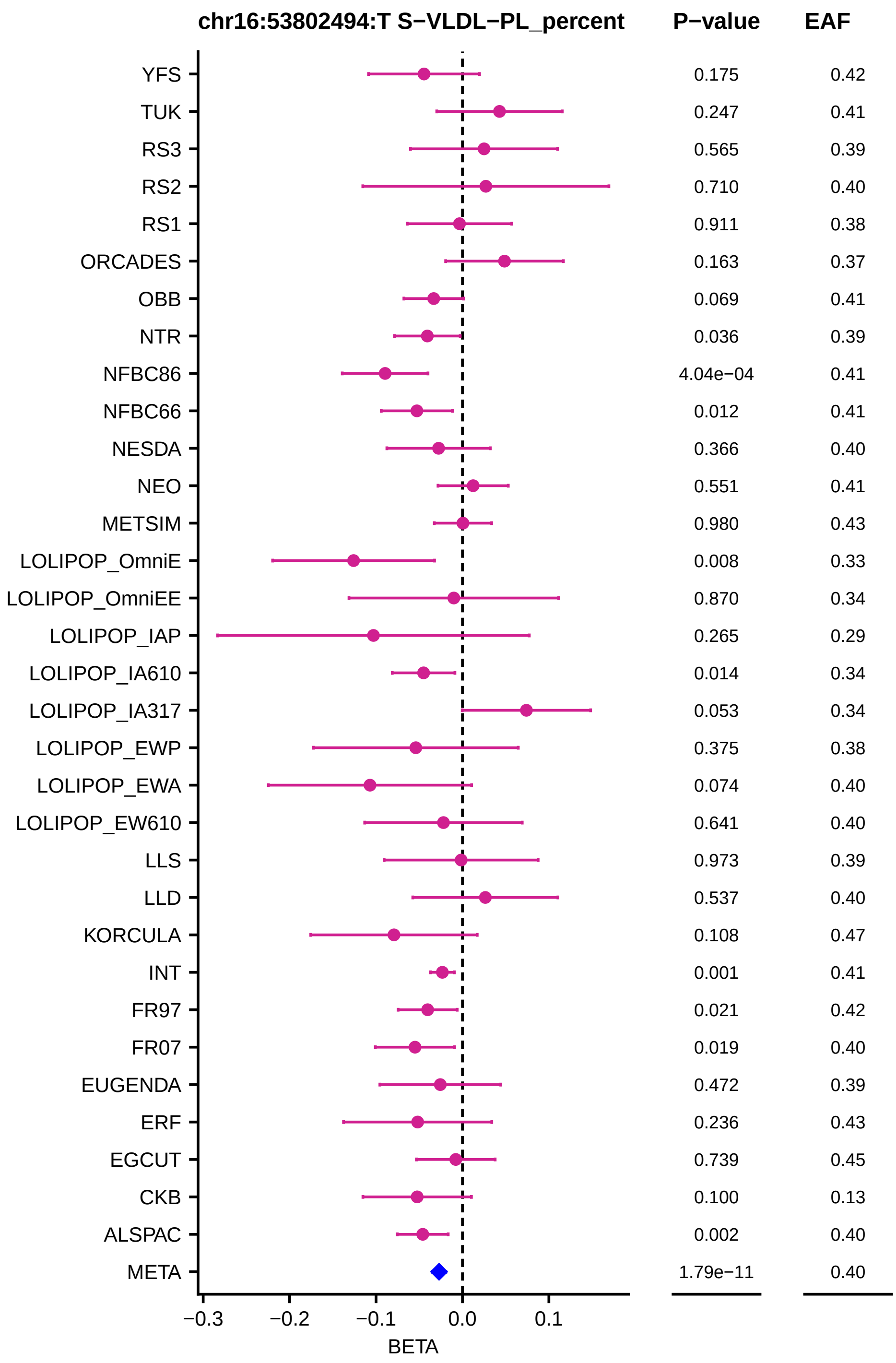
chr16:53802494:T S−VLDL−PL_percent
P−value
EAF
YFS
TUK
RS3
RS2
RS1
ORCADES
OBB
NTR
NFBC86
NFBC66
NESDA
NEO
METSIM
LOLIPOP_OmniE
LOLIPOP_OmniEE
LOLIPOP_IAP
LOLIPOP_IA610
LOLIPOP_IA317
LOLIPOP_EWP
LOLIPOP_EWA
LOLIPOP_EW610
LLS
LLD
KORCULA
INT
FR97
FR07
EUGENDA
ERF
EGCUT
CKB
ALSPAC
META
0.175
0.247
0.565
0.710
0.911
0.163
0.069
0.036
4.04e−04
0.012
0.366
0.551
0.980
0.008
0.870
0.265
0.014
0.053
0.375
0.074
0.641
0.973
0.537
0.108
0.001
0.021
0.019
0.472
0.236
0.739
0.100
0.002
1.79e−11
0.42
0.41
0.39
0.40
0.38
0.37
0.41
0.39
0.41
0.41
0.40
0.41
0.43
0.33
0.34
0.29
0.34
0.34
0.38
0.40
0.40
0.39
0.40
0.47
0.41
0.42
0.40
0.39
0.43
0.45
0.13
0.40
0.40
−0.3
−0.2
−0.1
0.0
0.1
BETA

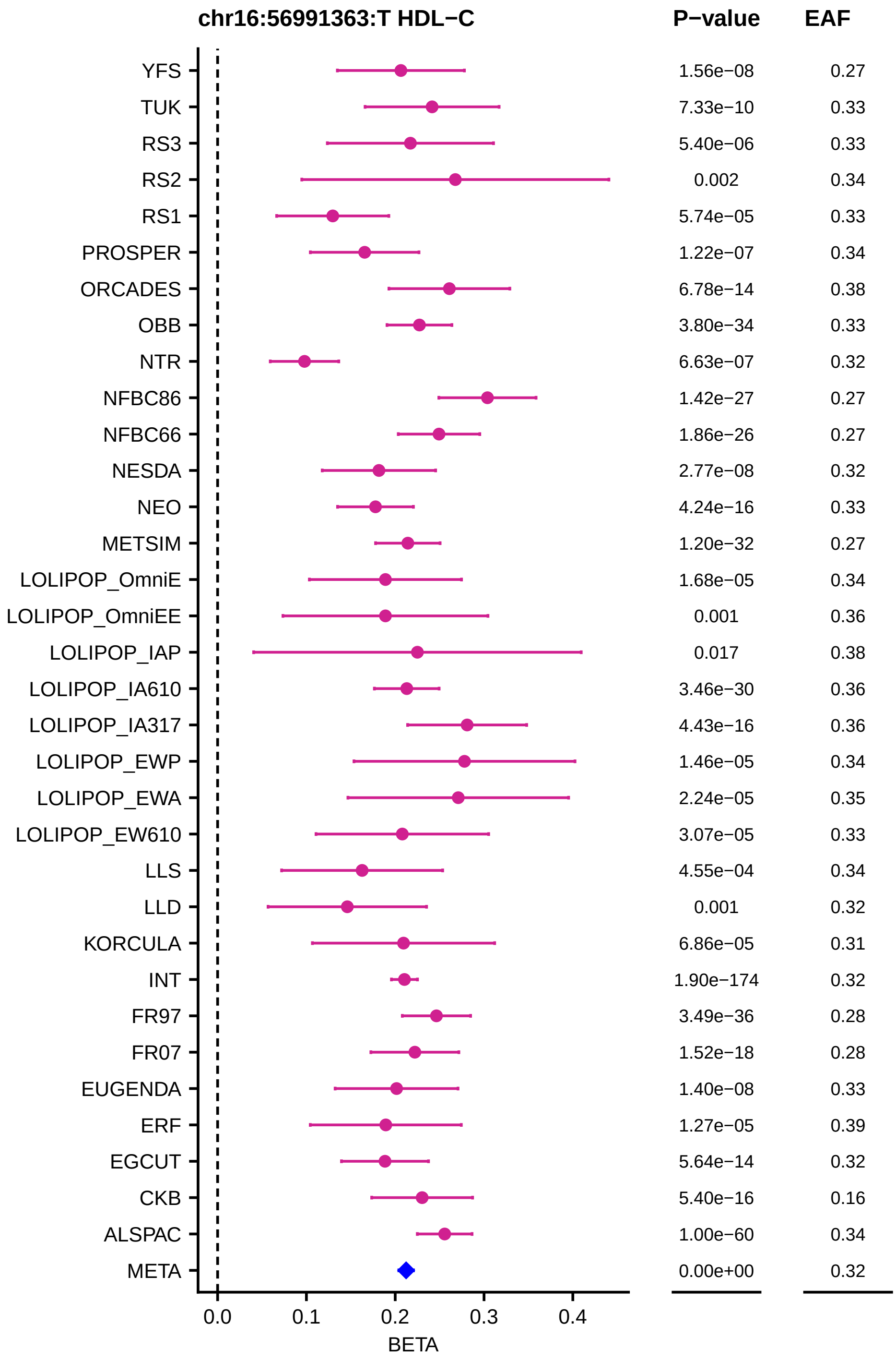
chr16:56991363:T HDL−C
P−value
EAF
YFS 1.56e−08 0.27
TUK 7.33e−10 0.33
RS3 5.40e−06 0.33
RS2 0.002 0.34
RS1 5.74e−05 0.33
PROSPER 1.22e−07 0.34
ORCADES 6.78e−14 0.38
OBB 3.80e−34 0.33
NTR 6.63e−07 0.32
NFBC86 1.42e−27 0.27
NFBC66 1.86e−26 0.27
NESDA 2.77e−08 0.32
NEO 4.24e−16 0.33
METSIM 1.20e−32 0.27
LOLIPOP_OmniE 1.68e−05 0.34
LOLIPOP_OmniEE 0.001 0.36
LOLIPOP_IAP 0.017 0.38
LOLIPOP_IA610 3.46e−30 0.36
LOLIPOP_IA317 4.43e−16 0.36
LOLIPOP_EWP 1.46e−05 0.34
LOLIPOP_EWA 2.24e−05 0.35
LOLIPOP_EW610 3.07e−05 0.33
LLS 4.55e−04 0.34
LLD 0.001 0.32
KORCULA 6.86e−05 0.31
INT 1.90e−174 0.32
FR97 3.49e−36 0.28
FR07 1.52e−18 0.28
EUGENDA 1.40e−08 0.33
ERF 1.27e−05 0.39
EGCUT 5.64e−14 0.32
CKB 5.40e−16 0.16
ALSPAC 1.00e−60 0.34
META 0.00e+00 0.32
0.0
0.1
0.2
0.3
0.4
BETA

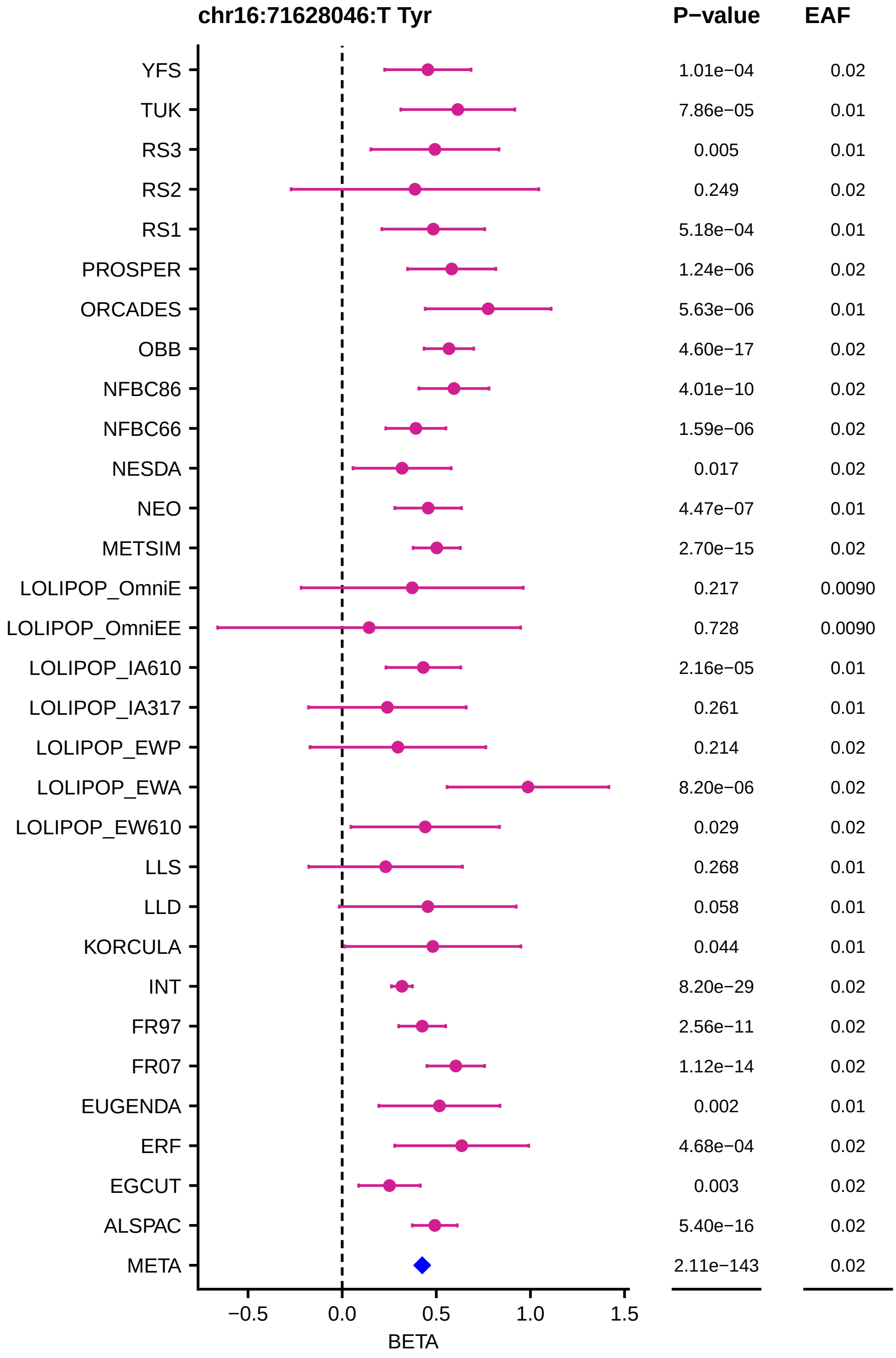
chr16:71628046:T Tyr
P−value
EAF
YFS
TUK
RS3
RS2
RS1
PROSPER
ORCADES
OBB
NFBC86
NFBC66
NESDA
NEO
METSIM
LOLIPOP_OmniE
LOLIPOP_OmniEE
LOLIPOP_IA610
LOLIPOP_IA317
LOLIPOP_EWP
LOLIPOP_EWA
LOLIPOP_EW610
LLS
LLD
KORCULA
INT
FR97
FR07
EUGENDA
ERF
EGCUT
ALSPAC
META
1.01e−04 0.02
7.86e−05 0.01
0.005 0.01
0.249 0.02
5.18e−04 0.01
1.24e−06 0.02
5.63e−06 0.01
4.60e−17 0.02
4.01e−10 0.02
1.59e−06 0.02
0.017 0.02
4.47e−07 0.01
2.70e−15 0.02
0.217 0.0090
0.728 0.0090
2.16e−05 0.01
0.261 0.01
0.214 0.02
8.20e−06 0.02
0.029 0.02
0.268 0.01
0.058 0.01
0.044 0.01
8.20e−29 0.02
2.56e−11 0.02
1.12e−14 0.02
0.002 0.01
4.68e−04 0.02
0.003 0.02
5.40e−16 0.02
2.11e−143 0.02
−0.5
0.0
0.5
1.0
1.5
BETA

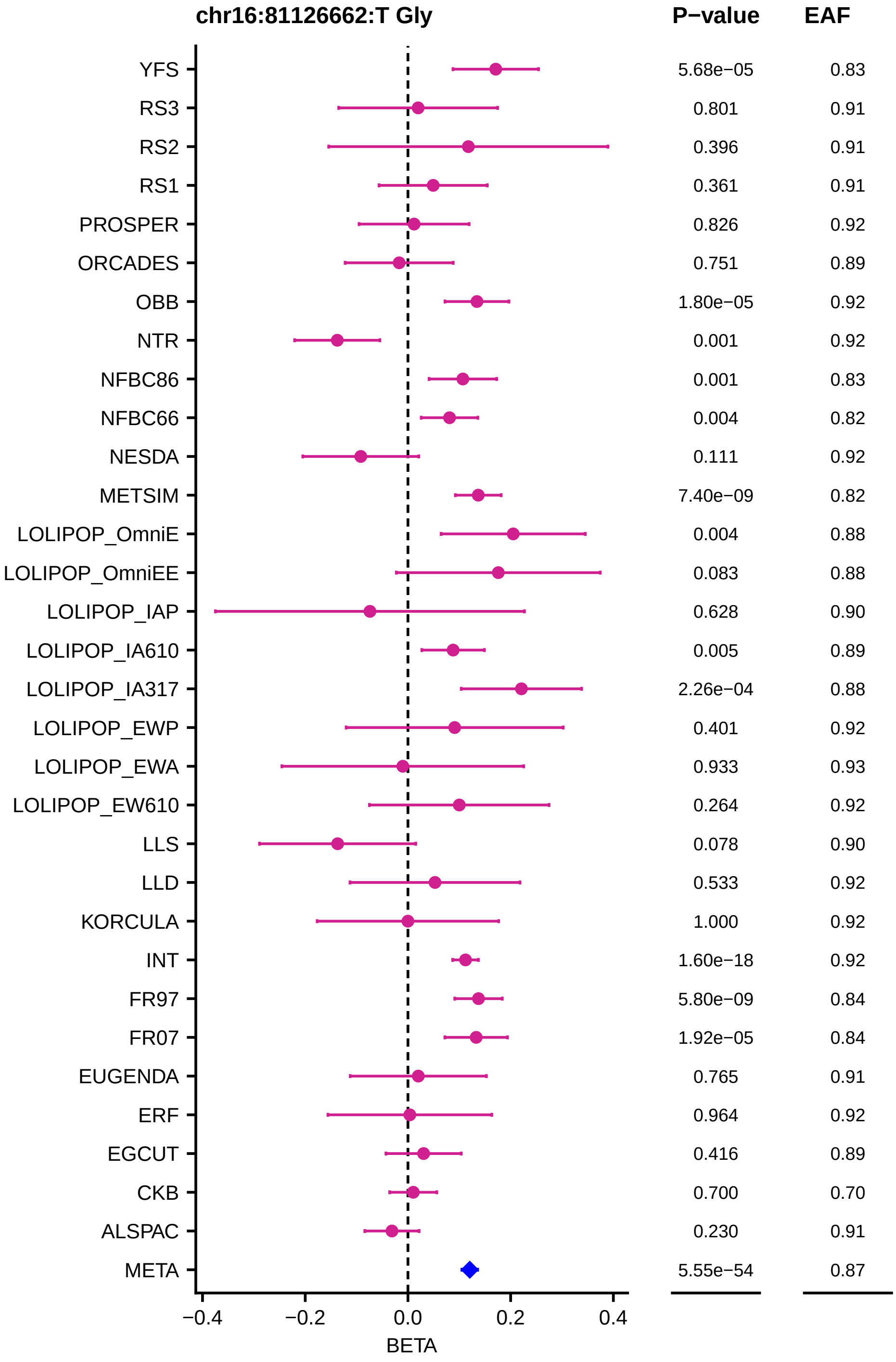
chr16:81126662:T Gly
P−value
EAF
YFS 5.68e−05 0.83
RS3 0.801 0.91
RS2 0.396 0.91
RS1 0.361 0.91
PROSPER 0.826 0.92
ORCADES 0.751 0.89
OBB 1.80e−05 0.92
NTR 0.001 0.92
NFBC86 0.001 0.83
NFBC66 0.004 0.82
NESDA 0.111 0.92
METSIM 7.40e−09 0.82
LOLIPOP_OmniE 0.004 0.88
LOLIPOP_OmniEE 0.083 0.88
LOLIPOP_IAP 0.628 0.90
LOLIPOP_IA610 0.005 0.89
LOLIPOP_IA317 2.26e−04 0.88
LOLIPOP_EWP 0.401 0.92
LOLIPOP_EWA 0.933 0.93
LOLIPOP_EW610 0.264 0.92
LLS 0.078 0.90
LLD 0.533 0.92
KORCULA 1.000 0.92
INT 1.60e−18 0.92
FR97 5.80e−09 0.84
FR07 1.92e−05 0.84
EUGENDA 0.765 0.91
ERF 0.964 0.92
EGCUT 0.416 0.89
CKB 0.700 0.70
ALSPAC 0.230 0.91
META 5.55e−54 0.87
−0.4
−0.2
0.0
0.2
0.4
BETA

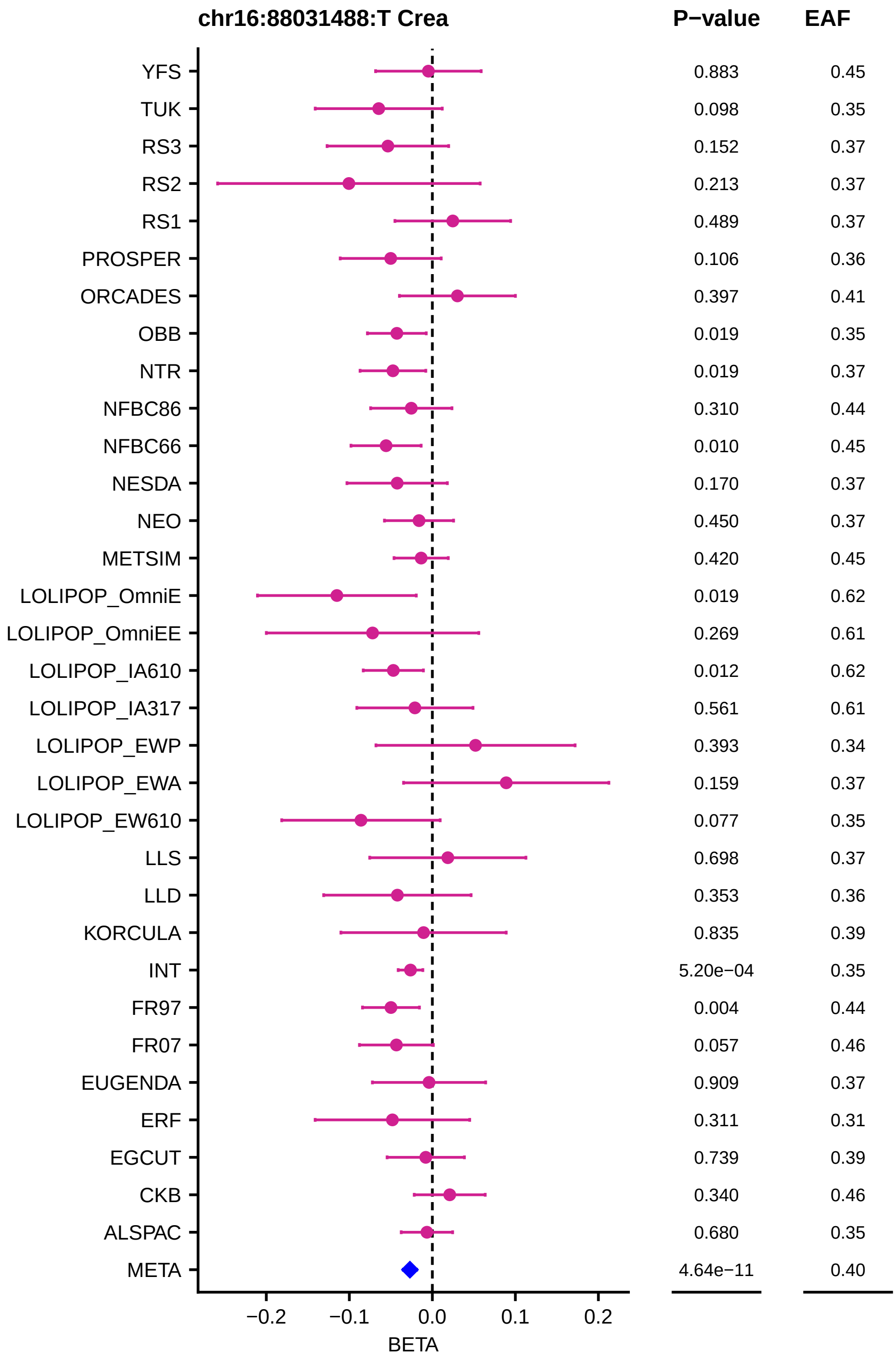
chr16:88031488:T Crea
P−value
EAF
YFS 0.883 0.45
TUK 0.098 0.35
RS3 0.152 0.37
RS2 0.213 0.37
RS1 0.489 0.37
PROSPER 0.106 0.36
ORCADES 0.397 0.41
OBB 0.019 0.35
NTR 0.019 0.37
NFBC86 0.310 0.44
NFBC66 0.010 0.45
NESDA 0.170 0.37
NEO 0.450 0.37
METSIM 0.420 0.45
LOLIPOP_OmniE 0.019 0.62
LOLIPOP_OmniEE 0.269 0.61
LOLIPOP_IA610 0.012 0.62
LOLIPOP_IA317 0.561 0.61
LOLIPOP_EWP 0.393 0.34
LOLIPOP_EWA 0.159 0.37
LOLIPOP_EW610 0.077 0.35
LLS 0.698 0.37
LLD 0.353 0.36
KORCULA 0.835 0.39
INT 5.20e−04 0.35
FR97 0.004 0.44
FR07 0.057 0.46
EUGENDA 0.909 0.37
ERF 0.311 0.31
EGCUT 0.739 0.39
CKB 0.340 0.46
ALSPAC 0.680 0.35
META 4.64e−11 0.40
−0.2 −0.1 0.0 0.1 0.2
BETA

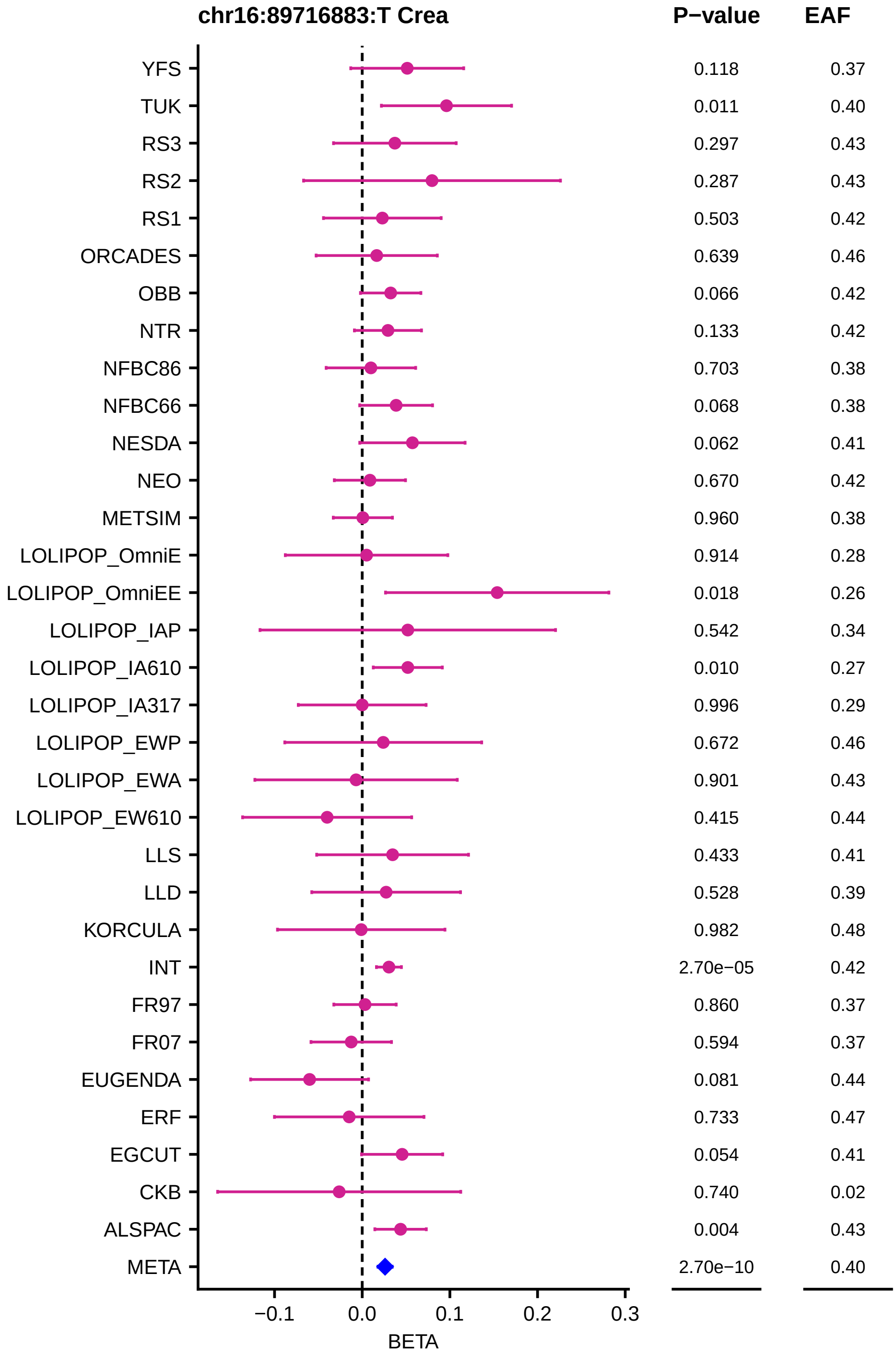
chr16:89716883:T Crea
P−value
EAF
YFS 0.118 0.37
TUK 0.011 0.40
RS3 0.297 0.43
RS2 0.287 0.43
RS1 0.503 0.42
ORCADES 0.639 0.46
OBB 0.066 0.42
NTR 0.133 0.42
NFBC86 0.703 0.38
NFBC66 0.068 0.38
NESDA 0.062 0.41
NEO 0.670 0.42
METSIM 0.960 0.38
LOLIPOP_OmniE 0.914 0.28
LOLIPOP_OmniEE 0.018 0.26
LOLIPOP_IAP 0.542 0.34
LOLIPOP_IA610 0.010 0.27
LOLIPOP_IA317 0.996 0.29
LOLIPOP_EWP 0.672 0.46
LOLIPOP_EWA 0.901 0.43
LOLIPOP_EW610 0.415 0.44
LLS 0.433 0.41
LLD 0.528 0.39
KORCULA 0.982 0.48
INT 2.70e−05 0.42
FR97 0.860 0.37
FR07 0.594 0.37
EUGENDA 0.081 0.44
ERF 0.733 0.47
EGCUT 0.054 0.41
CKB 0.740 0.02
ALSPAC 0.004 0.43
META 2.70e−10 0.40
−0.1
0.0
0.1
0.2
0.3
BETA

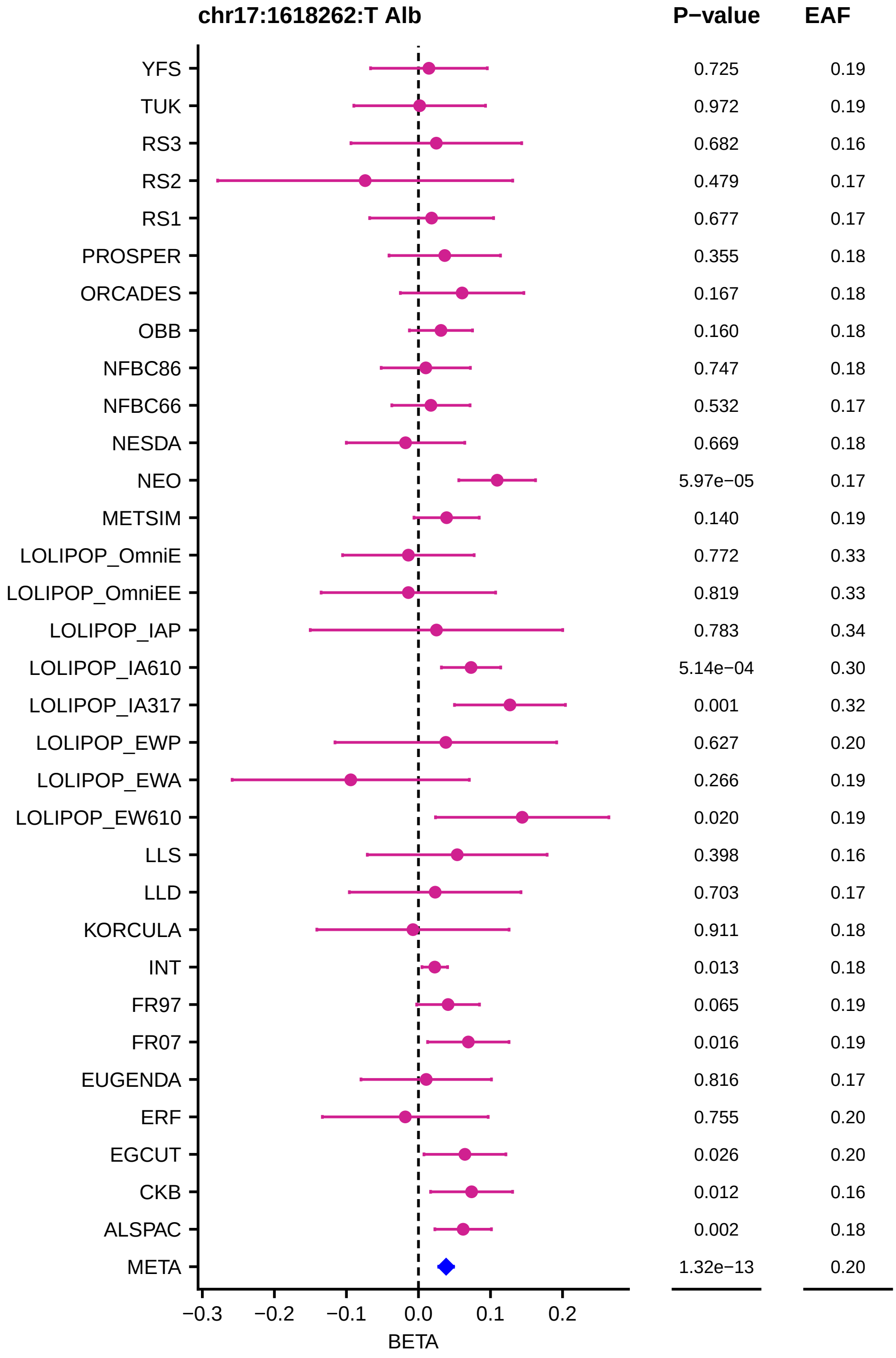
chr17:1618262:T Alb
P−value
EAF
YFS
TUK
RS3
RS2
RS1
PROSPER
ORCADES
OBB
NFBC86
NFBC66
NESDA
NEO
METSIM
LOLIPOP_OmniE
LOLIPOP_OmniEE
LOLIPOP_IAP
LOLIPOP_IA610
LOLIPOP_IA317
LOLIPOP_EWP
LOLIPOP_EWA
LOLIPOP_EW610
LLS
LLD
KORCULA
INT
FR97
FR07
EUGENDA
ERF
EGCUT
CKB
ALSPAC
META
0.725 0.19
0.972 0.19
0.682 0.16
0.479 0.17
0.677 0.17
0.355 0.18
0.167 0.18
0.160 0.18
0.747 0.18
0.532 0.17
0.669 0.18
5.97e−05 0.17
0.140 0.19
0.772 0.33
0.819 0.33
0.783 0.34
5.14e−04 0.30
0.001 0.32
0.627 0.20
0.266 0.19
0.020 0.19
0.398 0.16
0.703 0.17
0.911 0.18
0.013 0.18
0.065 0.19
0.016 0.19
0.816 0.17
0.755 0.20
0.026 0.20
0.012 0.16
0.002 0.18
1.32e−13 0.20
−0.3
−0.2
−0.1
0.0
0.1
0.2
BETA

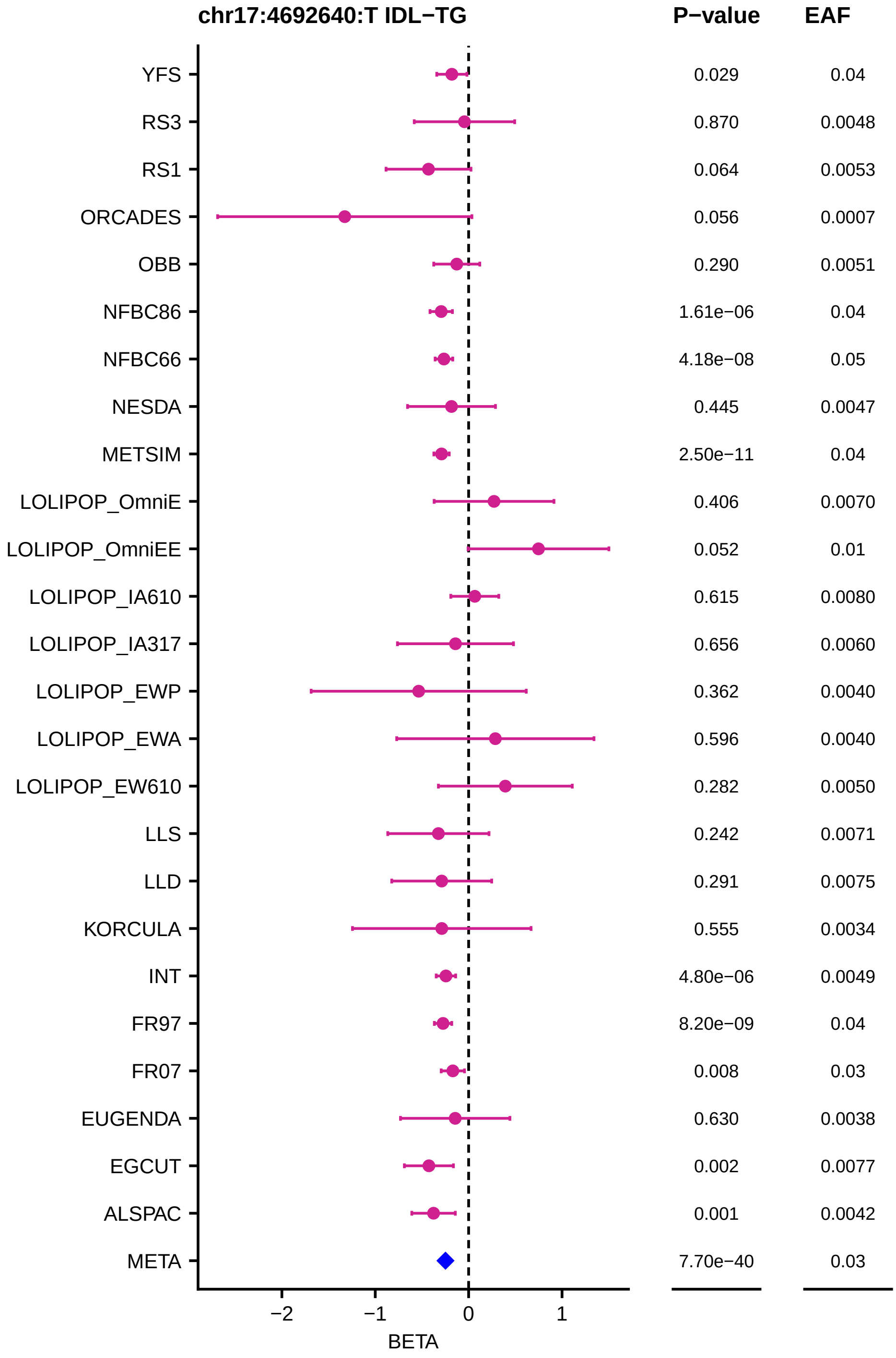
chr17:4692640:T IDL−TG
P−value
EAF
YFS 0.029 0.04
RS3 0.870 0.0048
RS1 0.064 0.0053
ORCADES 0.056 0.0007
OBB 0.290 0.0051
NFBC86 1.61e−06 0.04
NFBC66 4.18e−08 0.05
NESDA 0.445 0.0047
METSIM 2.50e−11 0.04
LOLIPOP_OmniE 0.406 0.0070
LOLIPOP_OmniEE 0.052 0.01
LOLIPOP_IA610 0.615 0.0080
LOLIPOP_IA317 0.656 0.0060
LOLIPOP_EWP 0.362 0.0040
LOLIPOP_EWA 0.596 0.0040
LOLIPOP_EW610 0.282 0.0050
LLS 0.242 0.0071
LLD 0.291 0.0075
KORCULA 0.555 0.0034
INT 4.80e−06 0.0049
FR97 8.20e−09 0.04
FR07 0.008 0.03
EUGENDA 0.630 0.0038
EGCUT 0.002 0.0077
ALSPAC 0.001 0.0042
META 7.70e−40 0.03
−2
−1
0
1
BETA

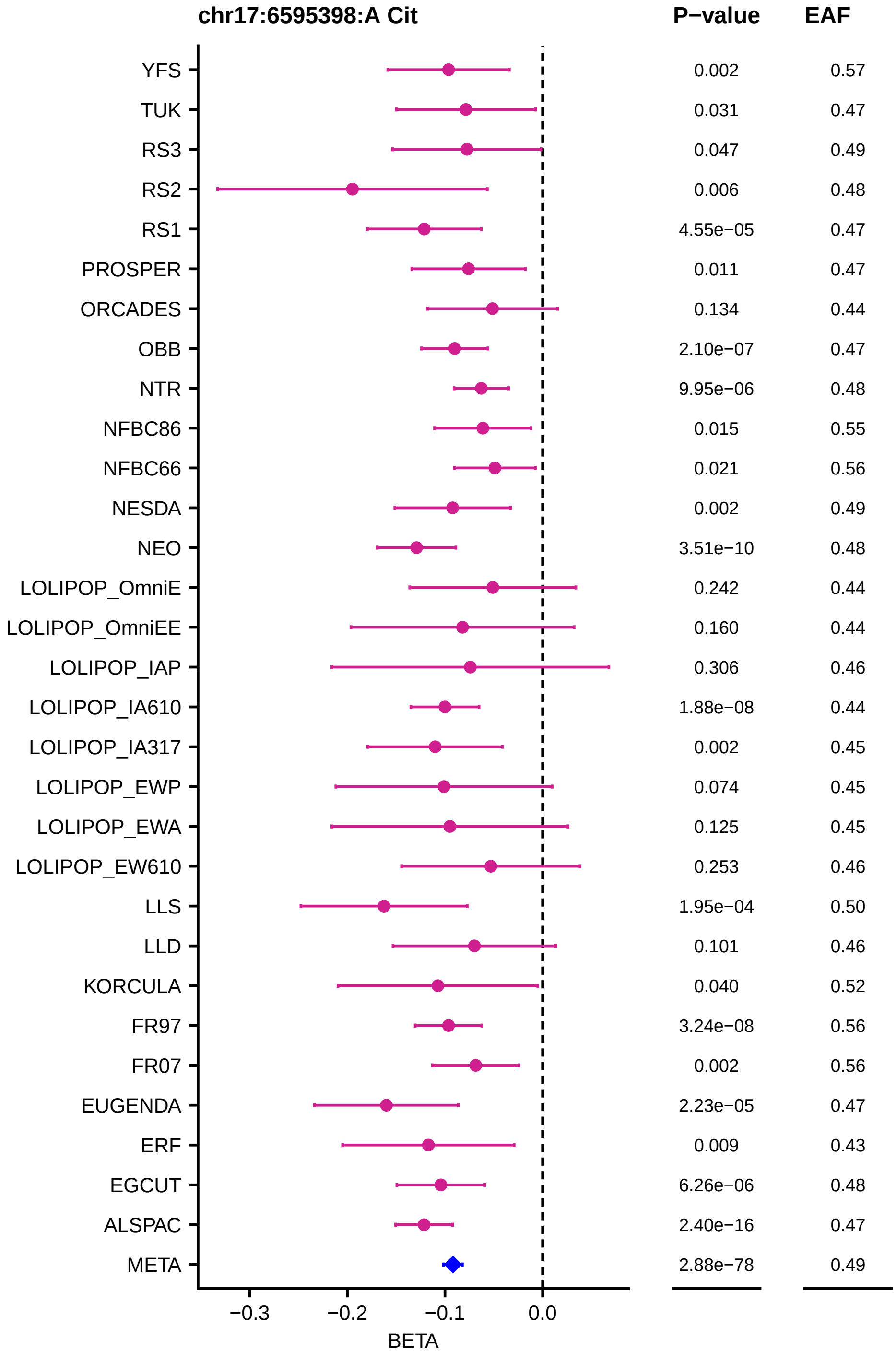
chr17:6595398:A Cit
P−value
EAF
YFS 0.002 0.57
TUK 0.031 0.47
RS3 0.047 0.49
RS2 0.006 0.48
RS1 4.55e−05 0.47
PROSPER 0.011 0.47
ORCADES 0.134 0.44
OBB 2.10e−07 0.47
NTR 9.95e−06 0.48
NFBC86 0.015 0.55
NFBC66 0.021 0.56
NESDA 0.002 0.49
NEO 3.51e−10 0.48
LOLIPOP_OmniE 0.242 0.44
LOLIPOP_OmniEE 0.160 0.44
LOLIPOP_IAP 0.306 0.46
LOLIPOP_IA610 1.88e−08 0.44
LOLIPOP_IA317 0.002 0.45
LOLIPOP_EWP 0.074 0.45
LOLIPOP_EWA 0.125 0.45
LOLIPOP_EW610 0.253 0.46
LLS 1.95e−04 0.50
LLD 0.101 0.46
KORCULA 0.040 0.52
FR97 3.24e−08 0.56
FR07 0.002 0.56
EUGENDA 2.23e−05 0.47
ERF 0.009 0.43
EGCUT 6.26e−06 0.48
ALSPAC 2.40e−16 0.47
META 2.88e−78 0.49
−0.3
−0.2
−0.1
0.0
BETA

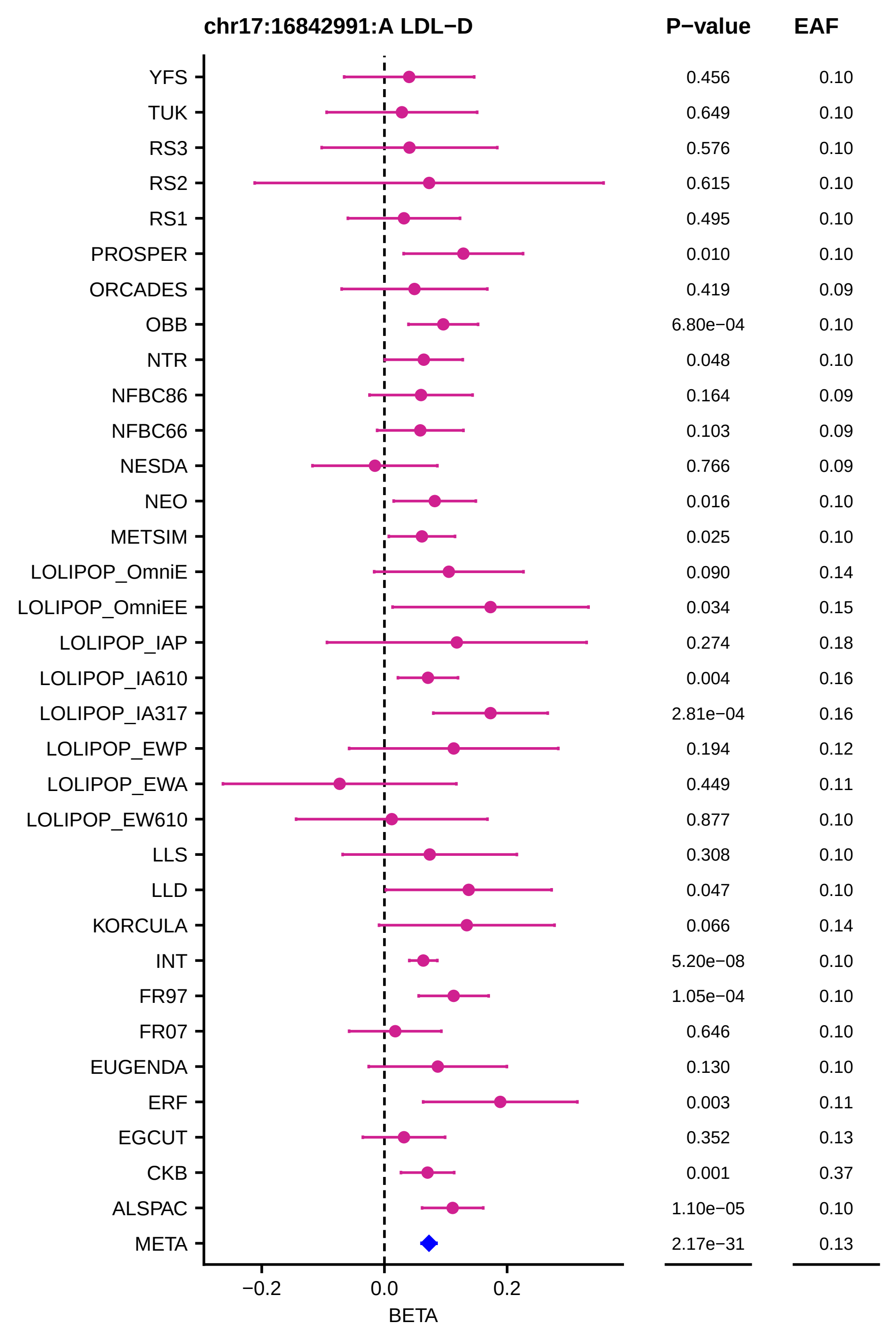
chr17:16842991:A LDL−D
P−value
EAF
YFS
TUK
RS3
RS2
RS1
PROSPER
ORCADES
OBB
NTR
NFBC86
NFBC66
NESDA
NEO
METSIM
LOLIPOP_OmniE
LOLIPOP_OmniEE
LOLIPOP_IAP
LOLIPOP_IA610
LOLIPOP_IA317
LOLIPOP_EWP
LOLIPOP_EWA
LOLIPOP_EW610
LLS
LLD
KORCULA
INT
FR97
FR07
EUGENDA
ERF
EGCUT
CKB
ALSPAC
META
0.456 0.10
0.649 0.10
0.576 0.10
0.615 0.10
0.495 0.10
0.010 0.10
0.419 0.09
6.80e−04 0.10
0.048 0.10
0.164 0.09
0.103 0.09
0.766 0.09
0.016 0.10
0.025 0.10
0.090 0.14
0.034 0.15
0.274 0.18
0.004 0.16
2.81e−04 0.16
0.194 0.12
0.449 0.11
0.877 0.10
0.308 0.10
0.047 0.10
0.066 0.14
5.20e−08 0.10
1.05e−04 0.10
0.646 0.10
0.130 0.10
0.003 0.11
0.352 0.13
0.001 0.37
1.10e−05 0.10
2.17e−31 0.13
−0.2
0.0
0.2
BETA

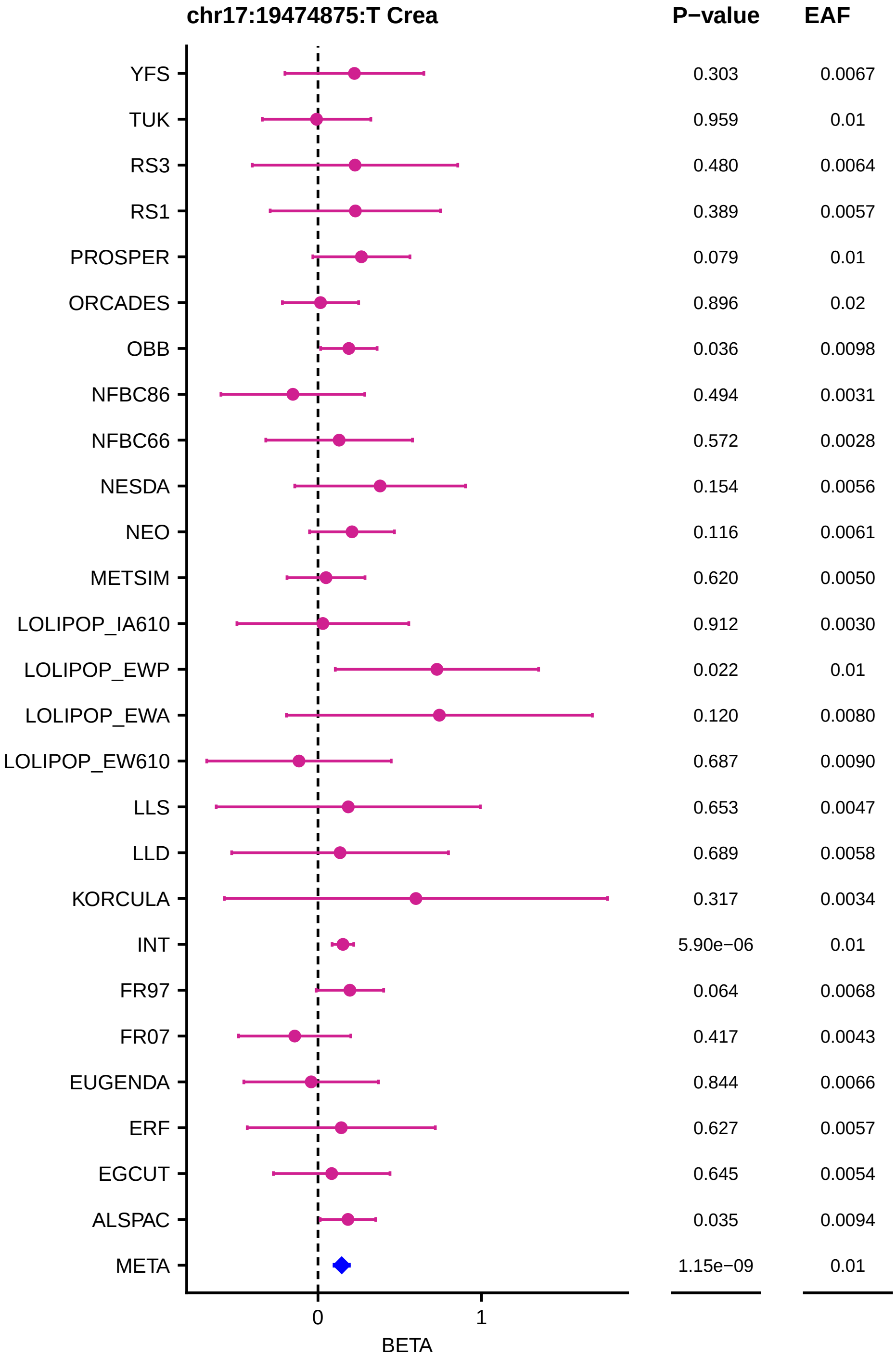

chr17:19474875:T Crea
P−value
EAF
YFS 0.303 0.0067
TUK 0.959 0.01
RS3 0.480 0.0064
RS1 0.389 0.0057
PROSPER 0.079 0.01
ORCADES 0.896 0.02
OBB 0.036 0.0098
NFBC86 0.494 0.0031
NFBC66 0.572 0.0028
NESDA 0.154 0.0056
NEO 0.116 0.0061
METSIM 0.620 0.0050
LOLIPOP_IA610 0.912 0.0030
LOLIPOP_EWP 0.022 0.01
LOLIPOP_EWA 0.120 0.0080
LOLIPOP_EW610 0.687 0.0090
LLS 0.653 0.0047
LLD 0.689 0.0058
KORCULA 0.317 0.0034
INT 5.90e−06 0.01
FR97 0.064 0.0068
FR07 0.417 0.0043
EUGENDA 0.844 0.0066
ERF 0.627 0.0057
EGCUT 0.645 0.0054
ALSPAC 0.035 0.0094
META 1.15e−09 0.01
0
1
BETA

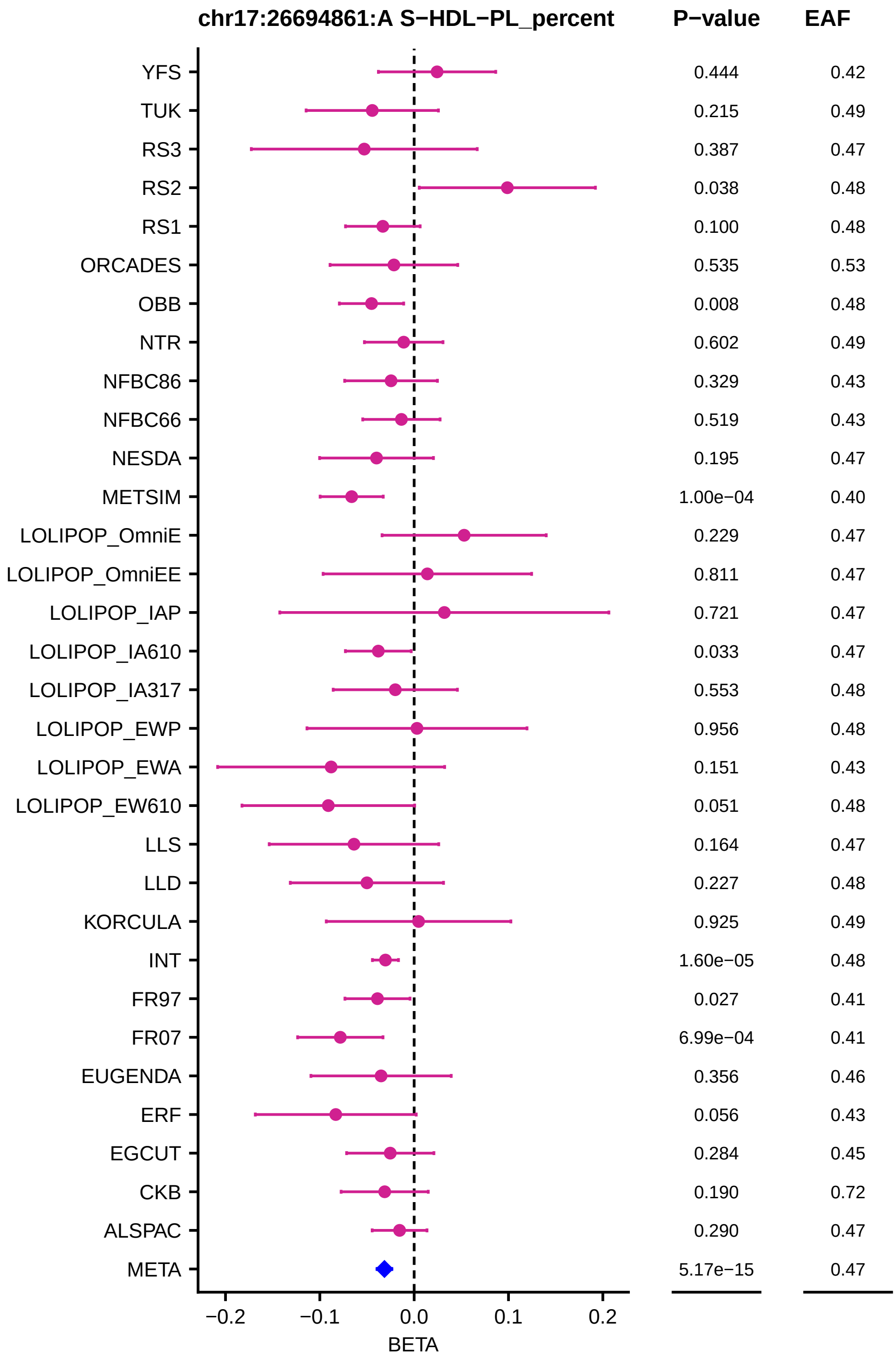
chr17:26694861:A S−HDL−PL_percent
P−value
EAF
YFS 0.444 0.42
TUK 0.215 0.49
RS3 0.387 0.47
RS2 0.038 0.48
RS1 0.100 0.48
ORCADES 0.535 0.53
OBB 0.008 0.48
NTR 0.602 0.49
NFBC86 0.329 0.43
NFBC66 0.519 0.43
NESDA 0.195 0.47
METSIM 1.00e−04 0.40
LOLIPOP_OmniE 0.229 0.47
LOLIPOP_OmniEE 0.811 0.47
LOLIPOP_IAP 0.721 0.47
LOLIPOP_IA610 0.033 0.47
LOLIPOP_IA317 0.553 0.48
LOLIPOP_EWP 0.956 0.48
LOLIPOP_EWA 0.151 0.43
LOLIPOP_EW610 0.051 0.48
LLS 0.164 0.47
LLD 0.227 0.48
KORCULA 0.925 0.49
INT 1.60e−05 0.48
FR97 0.027 0.41
FR07 6.99e−04 0.41
EUGENDA 0.356 0.46
ERF 0.056 0.43
EGCUT 0.284 0.45
CKB 0.190 0.72
ALSPAC 0.290 0.47
META 5.17e−15 0.47
−0.2
−0.1
0.0
0.1
0.2
BETA

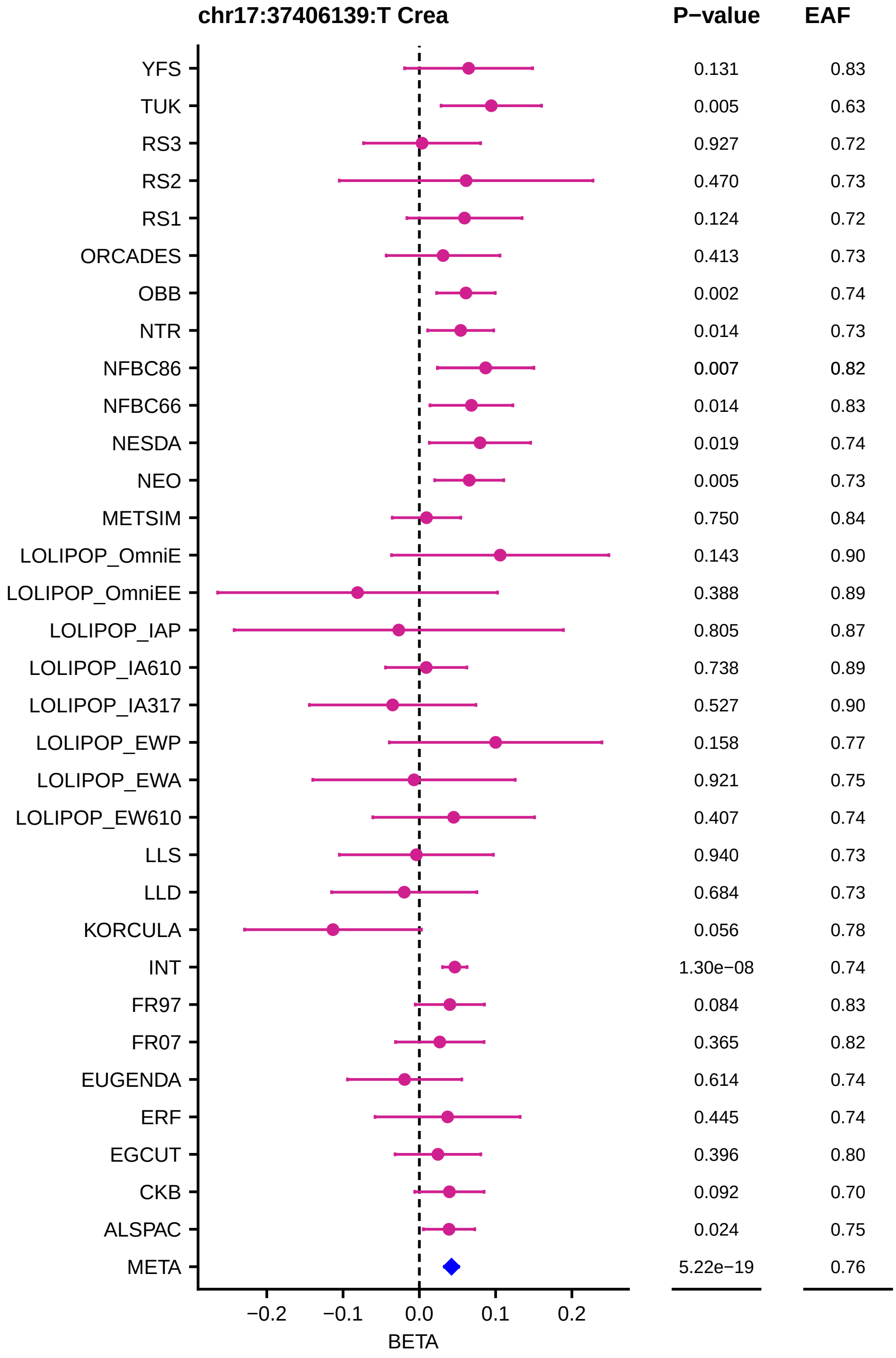
chr17:37406139:T Crea
P−value
EAF
YFS 0.131 0.83
TUK 0.005 0.63
RS3 0.927 0.72
RS2 0.470 0.73
RS1 0.124 0.72
ORCADES 0.413 0.73
OBB 0.002 0.74
NTR 0.014 0.73
NFBC86 0.007 0.82
NFBC66 0.014 0.83
NESDA 0.019 0.74
NEO 0.005 0.73
METSIM 0.750 0.84
LOLIPOP_OmniE 0.143 0.90
LOLIPOP_OmniEE 0.388 0.89
LOLIPOP_IAP 0.805 0.87
LOLIPOP_IA610 0.738 0.89
LOLIPOP_IA317 0.527 0.90
LOLIPOP_EWP 0.158 0.77
LOLIPOP_EWA 0.921 0.75
LOLIPOP_EW610 0.407 0.74
LLS 0.940 0.73
LLD 0.684 0.73
KORCULA 0.056 0.78
INT 1.30e−08 0.74
FR97 0.084 0.83
FR07 0.365 0.82
EUGENDA 0.614 0.74
ERF 0.445 0.74
EGCUT 0.396 0.80
CKB 0.092 0.70
ALSPAC 0.024 0.75
META 5.22e−19 0.76
−0.2
−0.1
0.0
0.1
0.2
BETA

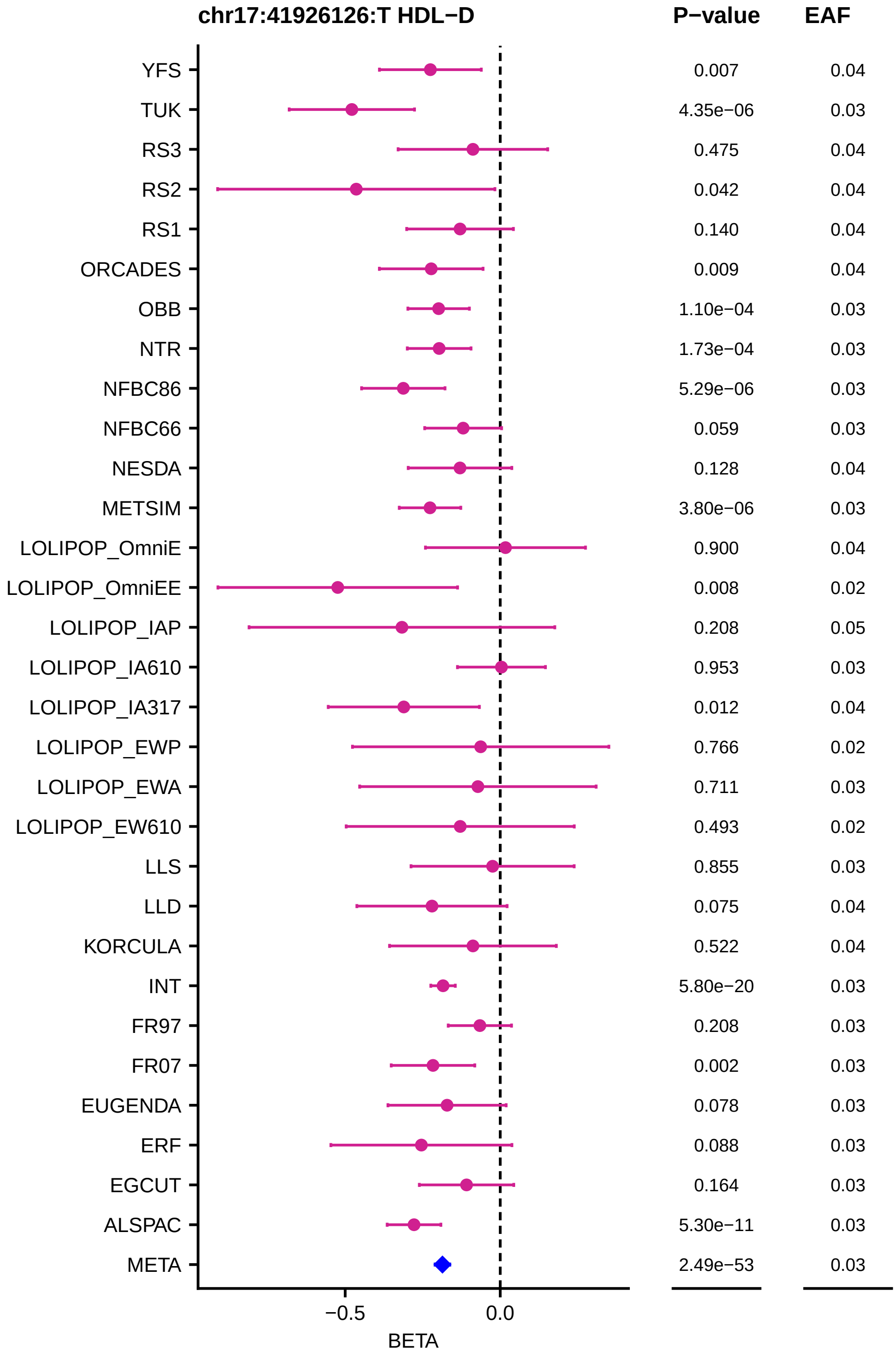
chr17:41926126:T HDL−D
P−value
EAF
YFS 0.007 0.04
TUK 4.35e−06 0.03
RS3 0.475 0.04
RS2 0.042 0.04
RS1 0.140 0.04
ORCADES 0.009 0.04
OBB 1.10e−04 0.03
NTR 1.73e−04 0.03
NFBC86 5.29e−06 0.03
NFBC66 0.059 0.03
NESDA 0.128 0.04
METSIM 3.80e−06 0.03
LOLIPOP_OmniE 0.900 0.04
LOLIPOP_OmniEE 0.008 0.02
LOLIPOP_IAP 0.208 0.05
LOLIPOP_IA610 0.953 0.03
LOLIPOP_IA317 0.012 0.04
LOLIPOP_EWP 0.766 0.02
LOLIPOP_EWA 0.711 0.03
LOLIPOP_EW610 0.493 0.02
LLS 0.855 0.03
LLD 0.075 0.04
KORCULA 0.522 0.04
INT 5.80e−20 0.03
FR97 0.208 0.03
FR07 0.002 0.03
EUGENDA 0.078 0.03
ERF 0.088 0.03
EGCUT 0.164 0.03
ALSPAC 5.30e−11 0.03
META 2.49e−53 0.03
−0.5
0.0
BETA

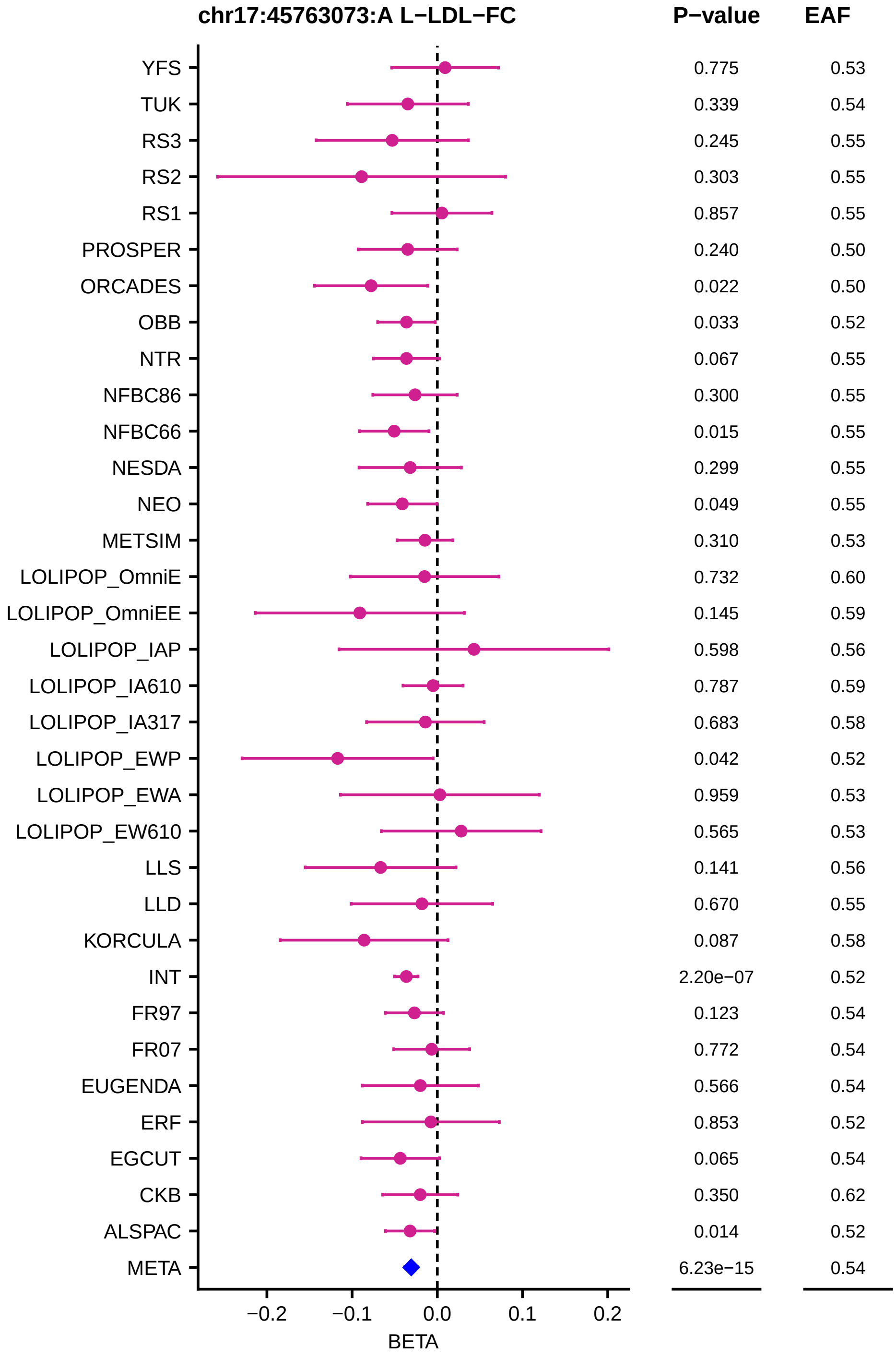
chr17:45763073:A L−LDL−FC
P−value
EAF
YFS 0.775 0.53
TUK 0.339 0.54
RS3 0.245 0.55
RS2 0.303 0.55
RS1 0.857 0.55
PROSPER 0.240 0.50
ORCADES 0.022 0.50
OBB 0.033 0.52
NTR 0.067 0.55
NFBC86 0.300 0.55
NFBC66 0.015 0.55
NESDA 0.299 0.55
NEO 0.049 0.55
METSIM 0.310 0.53
LOLIPOP_OmniE 0.732 0.60
LOLIPOP_OmniEE 0.145 0.59
LOLIPOP_IAP 0.598 0.56
LOLIPOP_IA610 0.787 0.59
LOLIPOP_IA317 0.683 0.58
LOLIPOP_EWP 0.042 0.52
LOLIPOP_EWA 0.959 0.53
LOLIPOP_EW610 0.565 0.53
LLS 0.141 0.56
LLD 0.670 0.55
KORCULA 0.087 0.58
INT 2.20e−07 0.52
FR97 0.123 0.54
FR07 0.772 0.54
EUGENDA 0.566 0.54
ERF 0.853 0.52
EGCUT 0.065 0.54
CKB 0.350 0.62
ALSPAC 0.014 0.52
META 6.23e−15 0.54
−0.2
−0.1
0.0
0.1
0.2
BETA

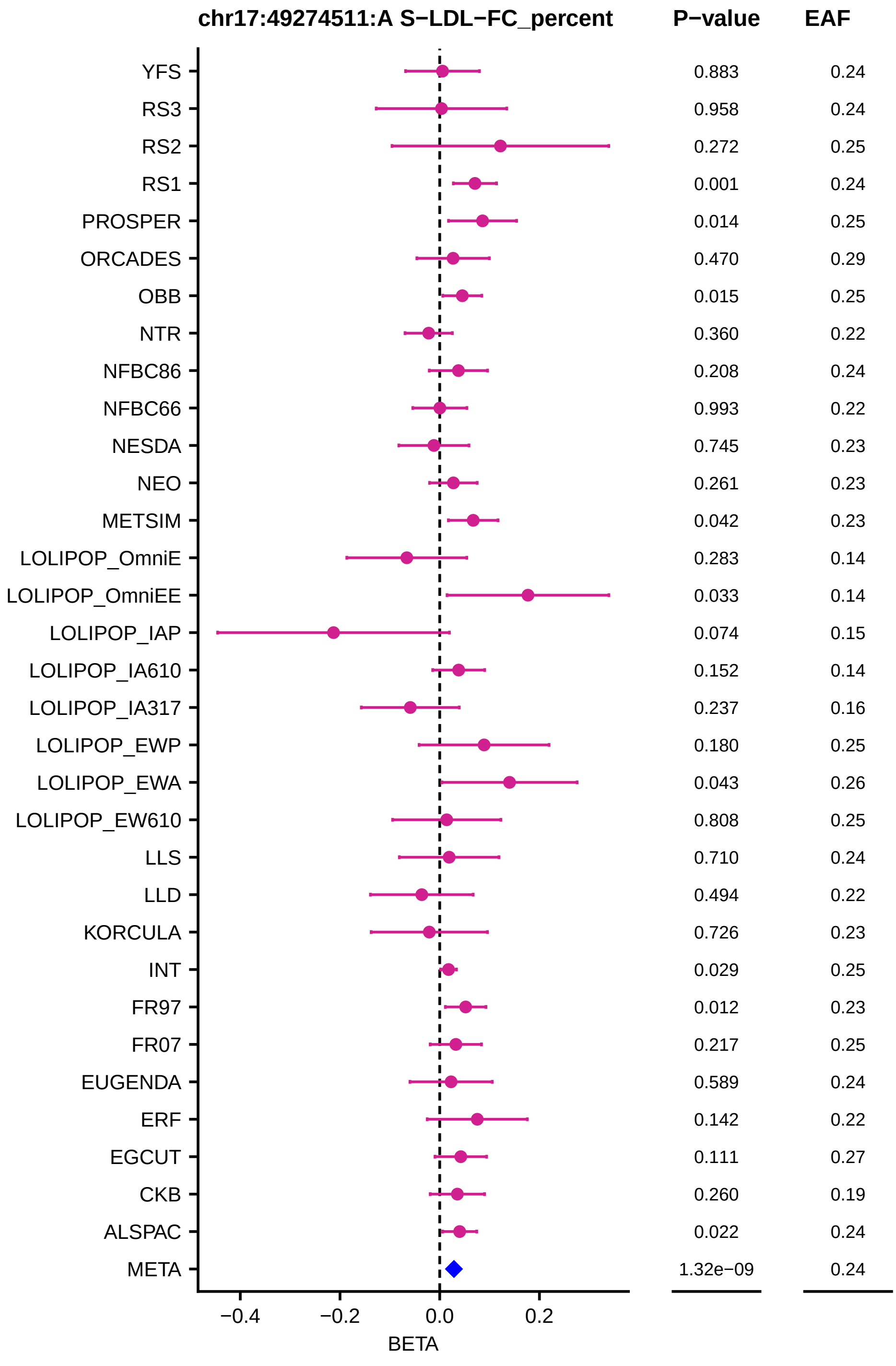
chr17:49274511:A S−LDL−FC_percent
P−value
EAF
YFS 0.883 0.24
RS3 0.958 0.24
RS2 0.272 0.25
RS1 0.001 0.24
PROSPER 0.014 0.25
ORCADES 0.470 0.29
OBB 0.015 0.25
NTR 0.360 0.22
NFBC86 0.208 0.24
NFBC66 0.993 0.22
NESDA 0.745 0.23
NEO 0.261 0.23
METSIM 0.042 0.23
LOLIPOP_OmniE 0.283 0.14
LOLIPOP_OmniEE 0.033 0.14
LOLIPOP_IAP 0.074 0.15
LOLIPOP_IA610 0.152 0.14
LOLIPOP_IA317 0.237 0.16
LOLIPOP_EWP 0.180 0.25
LOLIPOP_EWA 0.043 0.26
LOLIPOP_EW610 0.808 0.25
LLS 0.710 0.24
LLD 0.494 0.22
KORCULA 0.726 0.23
INT 0.029 0.25
FR97 0.012 0.23
FR07 0.217 0.25
EUGENDA 0.589 0.24
ERF 0.142 0.22
EGCUT 0.111 0.27
CKB 0.260 0.19
ALSPAC 0.022 0.24
META 1.32e−09 0.24
−0.4
−0.2
0.0
0.2
BETA

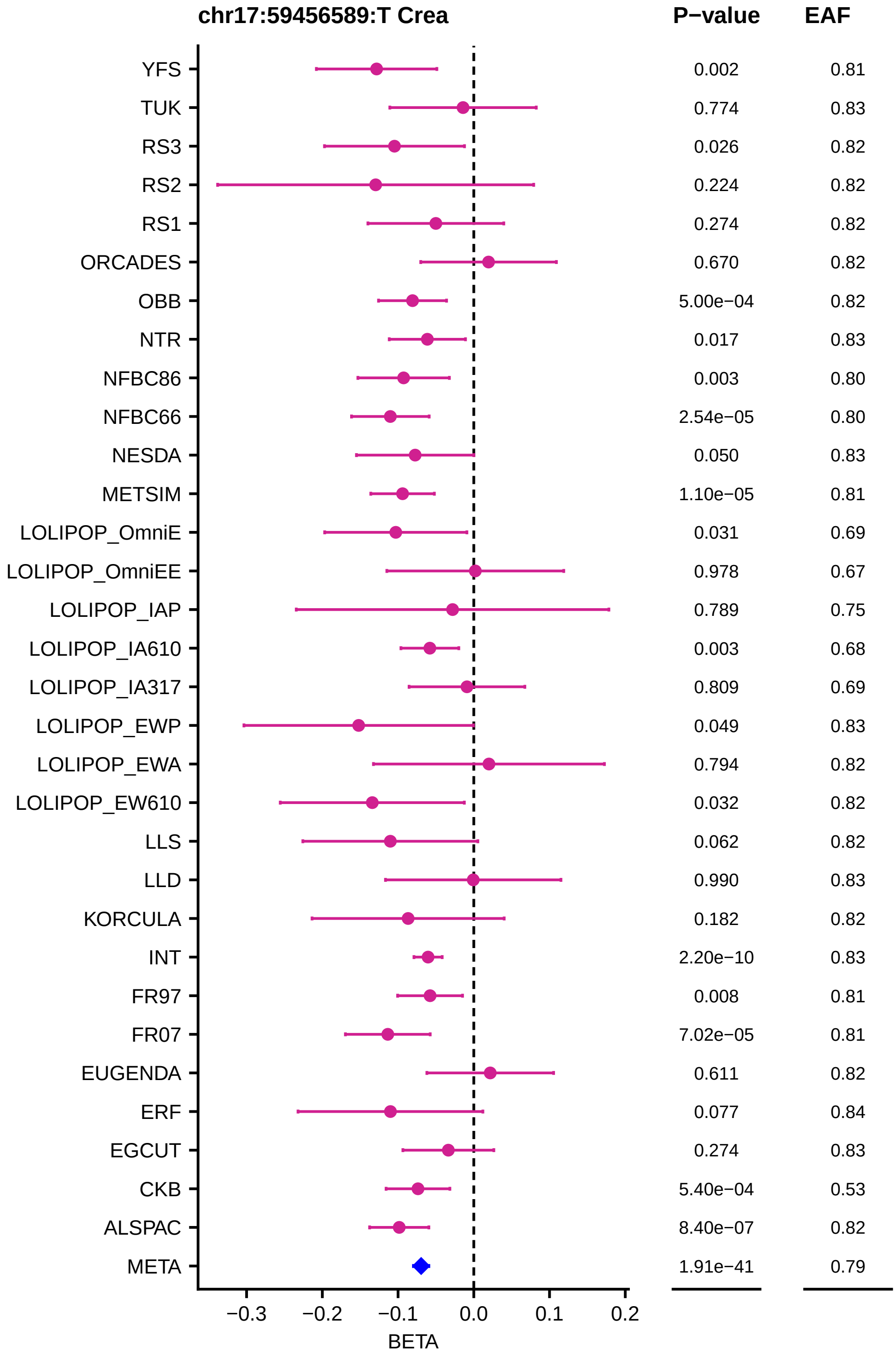
chr17:59456589:T Crea
P−value
EAF
YFS 0.002 0.81
TUK 0.774 0.83
RS3 0.026 0.82
RS2 0.224 0.82
RS1 0.274 0.82
ORCADES 0.670 0.82
OBB 5.00e−04 0.82
NTR 0.017 0.83
NFBC86 0.003 0.80
NFBC66 2.54e−05 0.80
NESDA 0.050 0.83
METSIM 1.10e−05 0.81
LOLIPOP_OmniE 0.031 0.69
LOLIPOP_OmniEE 0.978 0.67
LOLIPOP_IAP 0.789 0.75
LOLIPOP_IA610 0.003 0.68
LOLIPOP_IA317 0.809 0.69
LOLIPOP_EWP 0.049 0.83
LOLIPOP_EWA 0.794 0.82
LOLIPOP_EW610 0.032 0.82
LLS 0.062 0.82
LLD 0.990 0.83
KORCULA 0.182 0.82
INT 2.20e−10 0.83
FR97 0.008 0.81
FR07 7.02e−05 0.81
EUGENDA 0.611 0.82
ERF 0.077 0.84
EGCUT 0.274 0.83
CKB 5.40e−04 0.53
ALSPAC 8.40e−07 0.82
META 1.91e−41 0.79
−0.3
−0.2
−0.1
0.0
0.1
0.2
BETA

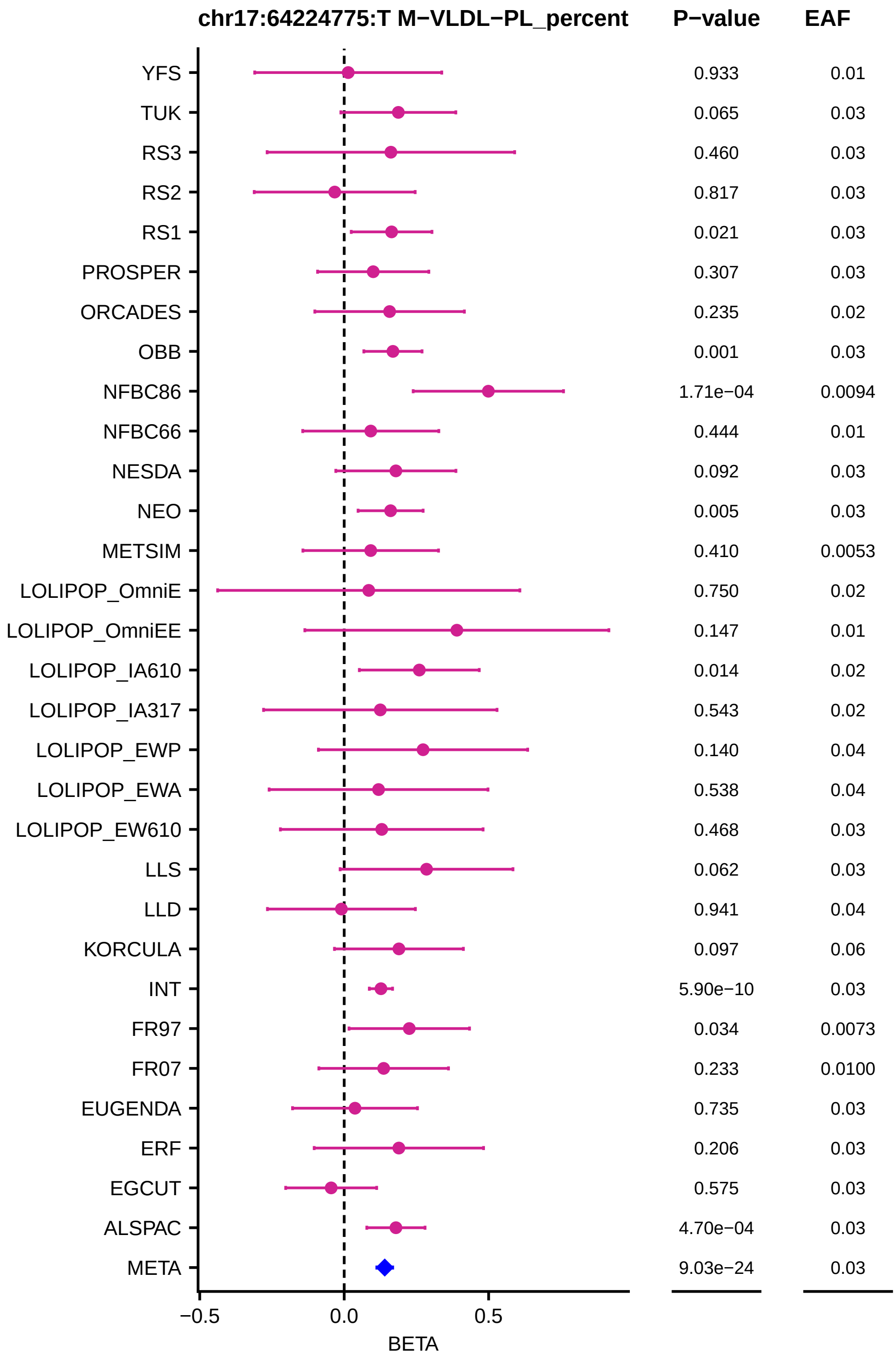
chr17:64224775:T M−VLDL−PL_percent
P−value
EAF
YFS 0.933 0.01
TUK 0.065 0.03
RS3 0.460 0.03
RS2 0.817 0.03
RS1 0.021 0.03
PROSPER 0.307 0.03
ORCADES 0.235 0.02
OBB 0.001 0.03
NFBC86 1.71e−04 0.0094
NFBC66 0.444 0.01
NESDA 0.092 0.03
NEO 0.005 0.03
METSIM 0.410 0.0053
LOLIPOP_OmniE 0.750 0.02
LOLIPOP_OmniEE 0.147 0.01
LOLIPOP_IA610 0.014 0.02
LOLIPOP_IA317 0.543 0.02
LOLIPOP_EWP 0.140 0.04
LOLIPOP_EWA 0.538 0.04
LOLIPOP_EW610 0.468 0.03
LLS 0.062 0.03
LLD 0.941 0.04
KORCULA 0.097 0.06
INT 5.90e−10 0.03
FR97 0.034 0.0073
FR07 0.233 0.0100
EUGENDA 0.735 0.03
ERF 0.206 0.03
EGCUT 0.575 0.03
ALSPAC 4.70e−04 0.03
META 9.03e−24 0.03
−0.5
0.0
0.5
BETA

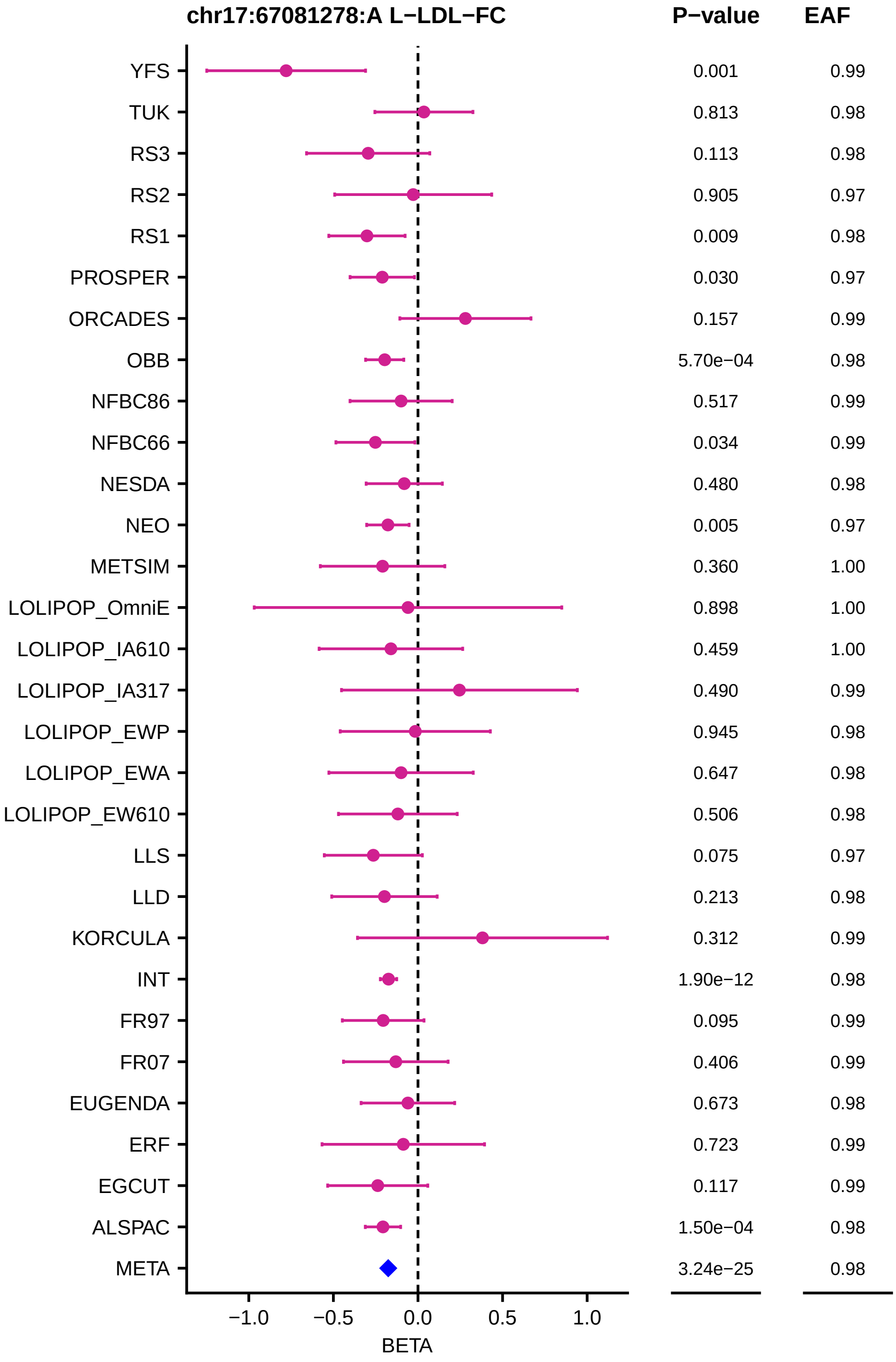
chr17:67081278:A L−LDL−FC
P−value
EAF
YFS 0.001 0.99
TUK 0.813 0.98
RS3 0.113 0.98
RS2 0.905 0.97
RS1 0.009 0.98
PROSPER 0.030 0.97
ORCADES 0.157 0.99
OBB 5.70e−04 0.98
NFBC86 0.517 0.99
NFBC66 0.034 0.99
NESDA 0.480 0.98
NEO 0.005 0.97
METSIM 0.360 1.00
LOLIPOP_OmniE 0.898 1.00
LOLIPOP_IA610 0.459 1.00
LOLIPOP_IA317 0.490 0.99
LOLIPOP_EWP 0.945 0.98
LOLIPOP_EWA 0.647 0.98
LOLIPOP_EW610 0.506 0.98
LLS 0.075 0.97
LLD 0.213 0.98
KORCULA 0.312 0.99
INT 1.90e−12 0.98
FR97 0.095 0.99
FR07 0.406 0.99
EUGENDA 0.673 0.98
ERF 0.723 0.99
EGCUT 0.117 0.99
ALSPAC 1.50e−04 0.98
META 3.24e−25 0.98
−1.0
−0.5
0.0
0.5
1.0
BETA

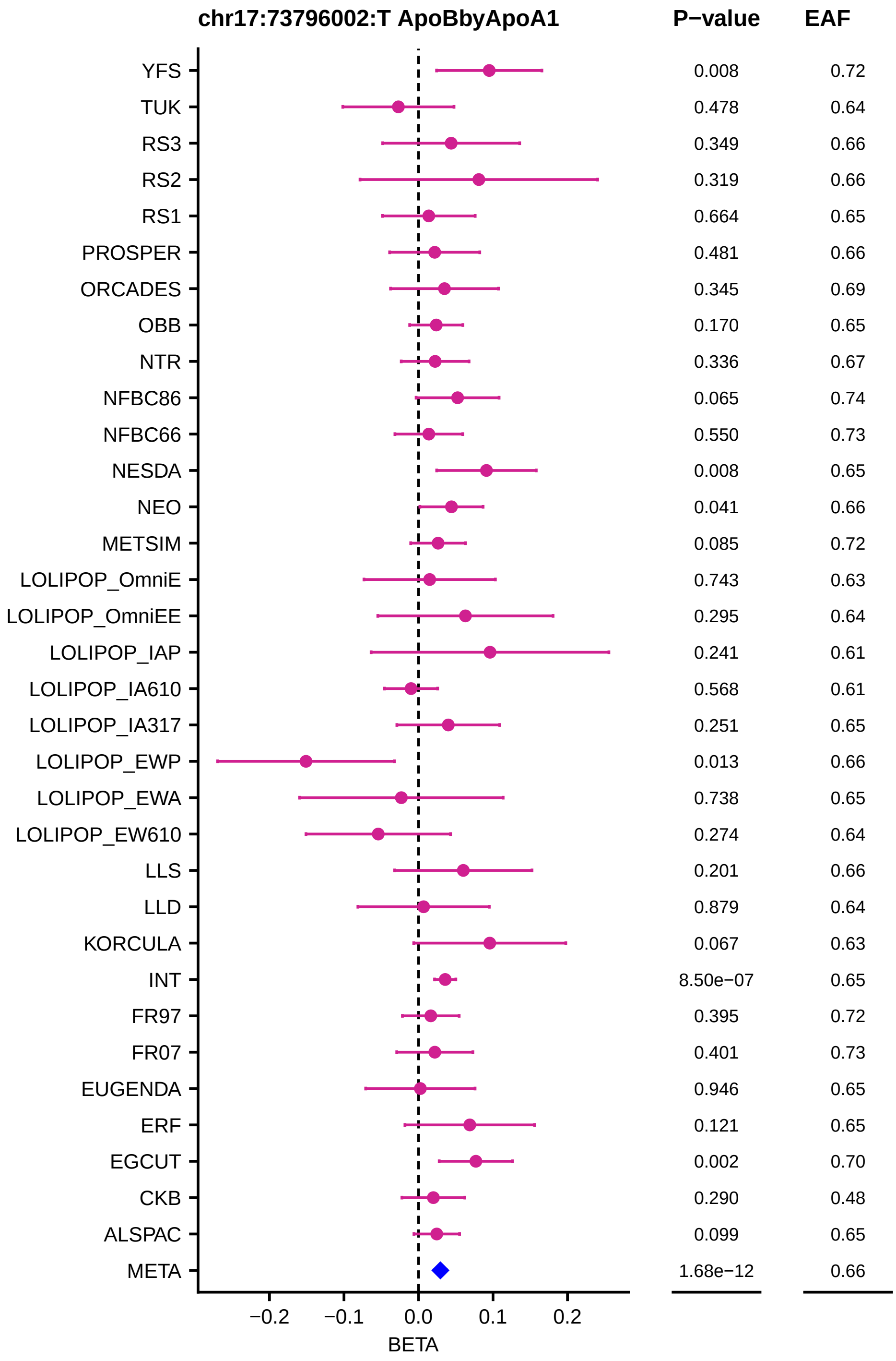
chr17:73796002:T ApoBbyApoA1
P−value
EAF
YFS 0.008 0.72
TUK 0.478 0.64
RS3 0.349 0.66
RS2 0.319 0.66
RS1 0.664 0.65
PROSPER 0.481 0.66
ORCADES 0.345 0.69
OBB 0.170 0.65
NTR 0.336 0.67
NFBC86 0.065 0.74
NFBC66 0.550 0.73
NESDA 0.008 0.65
NEO 0.041 0.66
METSIM 0.085 0.72
LOLIPOP_OmniE 0.743 0.63
LOLIPOP_OmniEE 0.295 0.64
LOLIPOP_IAP 0.241 0.61
LOLIPOP_IA610 0.568 0.61
LOLIPOP_IA317 0.251 0.65
LOLIPOP_EWP 0.013 0.66
LOLIPOP_EWA 0.738 0.65
LOLIPOP_EW610 0.274 0.64
LLS 0.201 0.66
LLD 0.879 0.64
KORCULA 0.067 0.63
INT 8.50e−07 0.65
FR97 0.395 0.72
FR07 0.401 0.73
EUGENDA 0.946 0.65
ERF 0.121 0.65
EGCUT 0.002 0.70
CKB 0.290 0.48
ALSPAC 0.099 0.65
META 1.68e−12 0.66
−0.2 −0.1 0.0 0.1 0.2
BETA

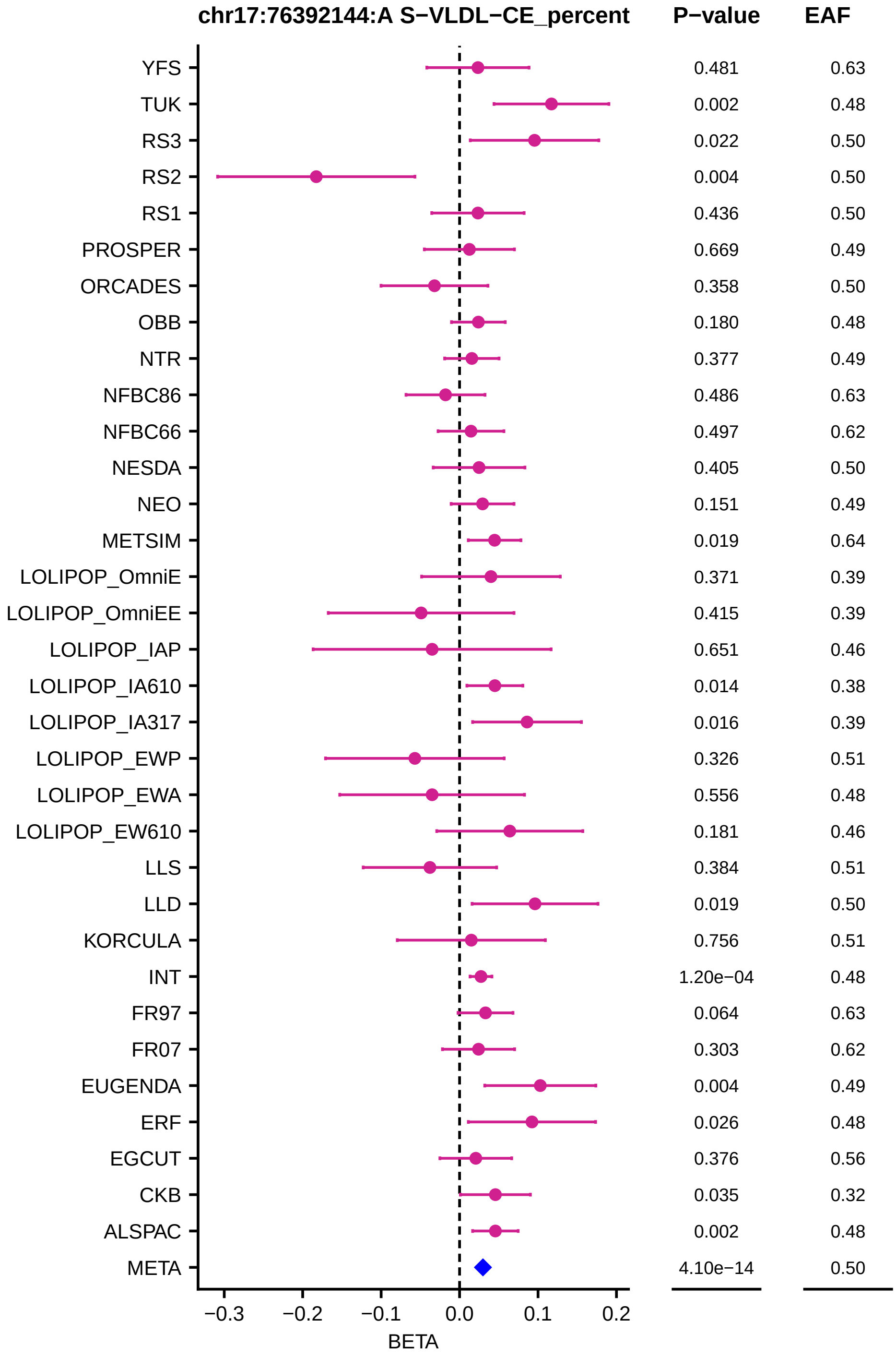
chr17:76392144:A S−VLDL−CE_percent
P−value
EAF
YFS 0.481 0.63
TUK 0.002 0.48
RS3 0.022 0.50
RS2 0.004 0.50
RS1 0.436 0.50
PROSPER 0.669 0.49
ORCADES 0.358 0.50
OBB 0.180 0.48
NTR 0.377 0.49
NFBC86 0.486 0.63
NFBC66 0.497 0.62
NESDA 0.405 0.50
NEO 0.151 0.49
METSIM 0.019 0.64
LOLIPOP_OmniE 0.371 0.39
LOLIPOP_OmniEE 0.415 0.39
LOLIPOP_IAP 0.651 0.46
LOLIPOP_IA610 0.014 0.38
LOLIPOP_IA317 0.016 0.39
LOLIPOP_EWP 0.326 0.51
LOLIPOP_EWA 0.556 0.48
LOLIPOP_EW610 0.181 0.46
LLS 0.384 0.51
LLD 0.019 0.50
KORCULA 0.756 0.51
INT 1.20e−04 0.48
FR97 0.064 0.63
FR07 0.303 0.62
EUGENDA 0.004 0.49
ERF 0.026 0.48
EGCUT 0.376 0.56
CKB 0.035 0.32
ALSPAC 0.002 0.48
META 4.10e−14 0.50
−0.3 −0.2 −0.1 0.0 0.1 0.2
BETA

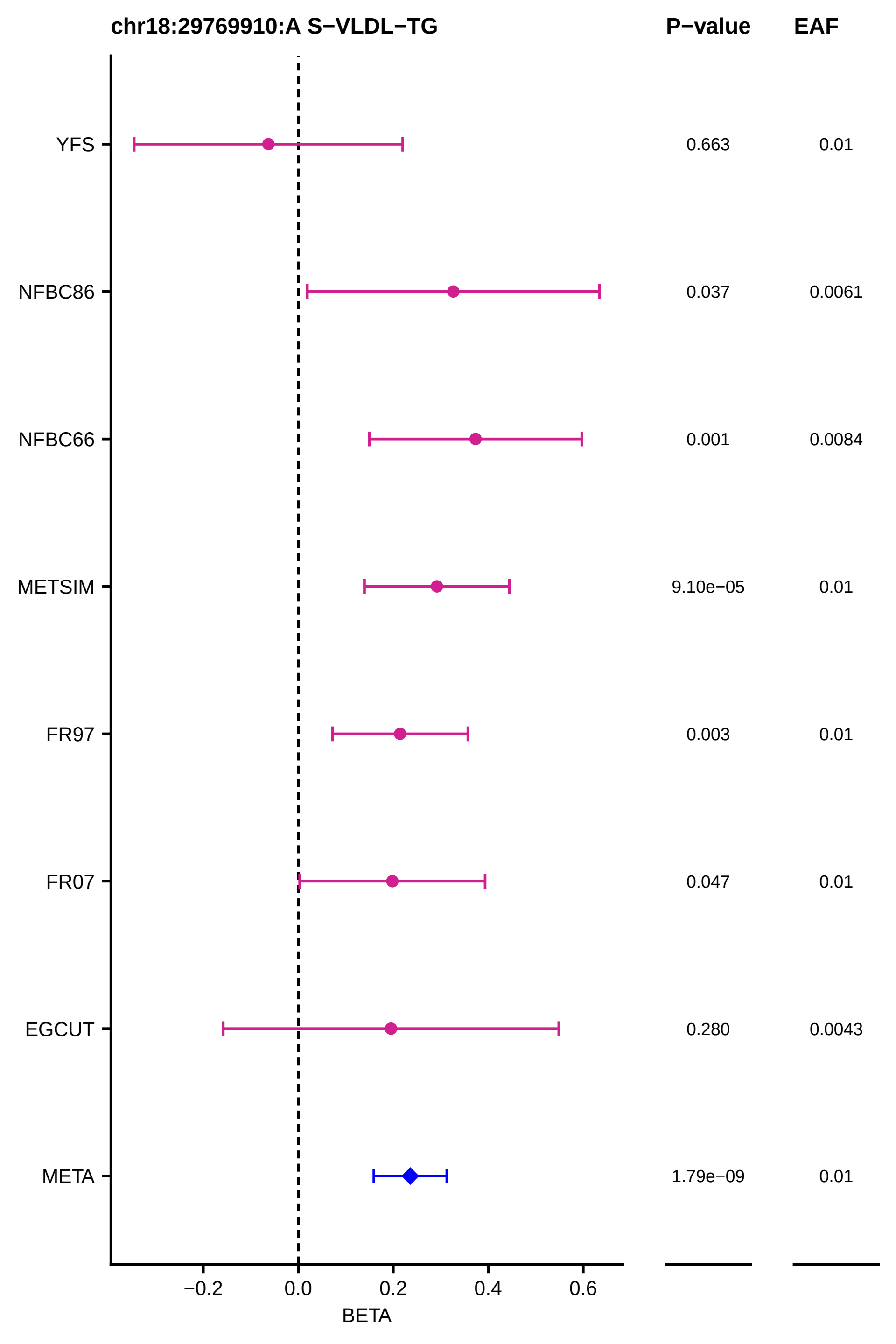

chr18:29769910:A S−VLDL−TG
P−value
EAF
YFS
0.663
0.01
NFBC86
0.037
0.0061
NFBC66
0.001
0.0084
METSIM
9.10e−05
0.01
FR97
0.003
0.01
FR07
0.047
0.01
EGCUT
0.280
0.0043
META
1.79e−09
0.01
−0.2
0.0
0.2
0.4
0.6
BETA

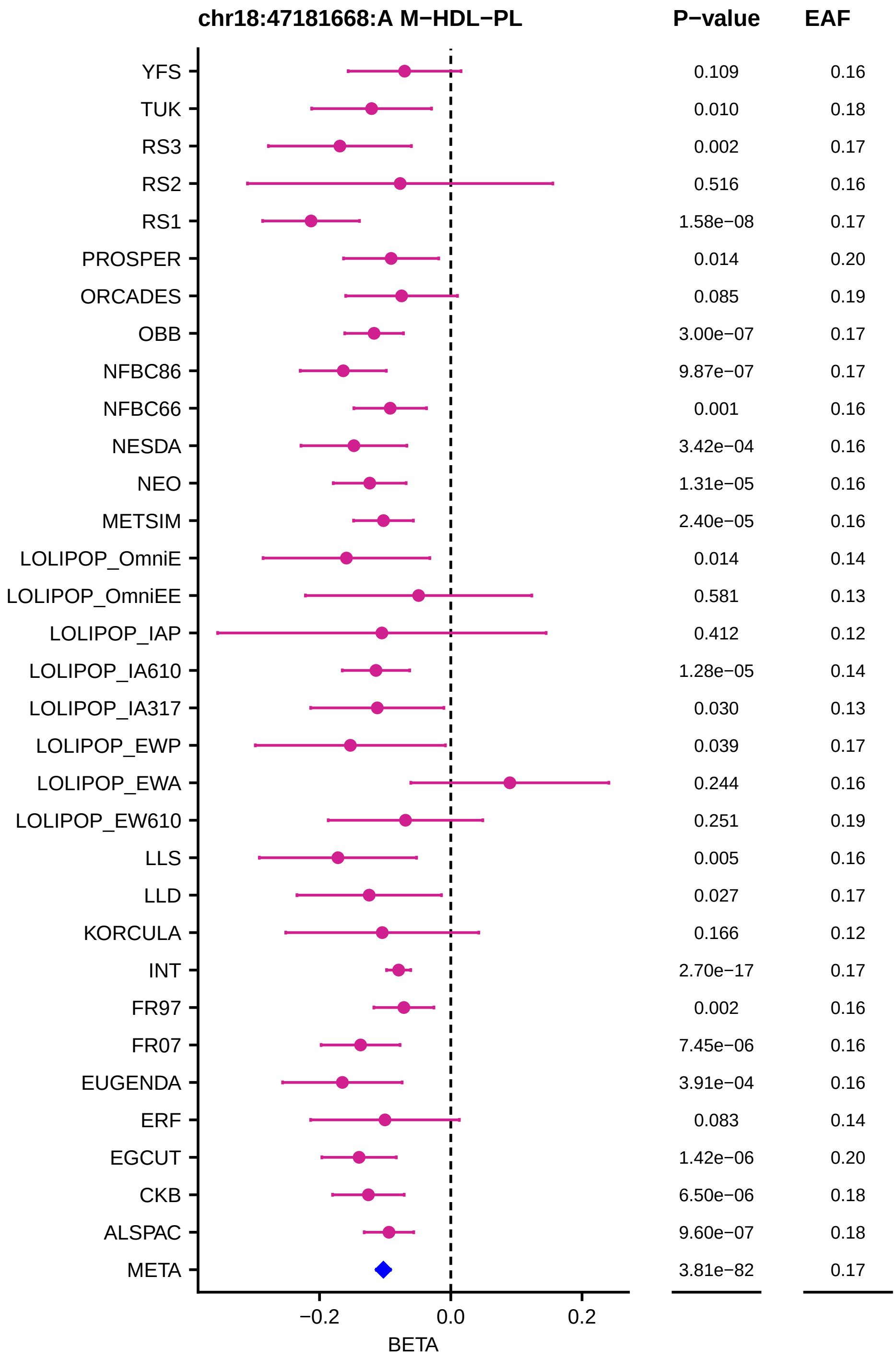
chr18:47181668:A M−HDL−PL
P−value
EAF
YFS 0.109 0.16
TUK 0.010 0.18
RS3 0.002 0.17
RS2 0.516 0.16
RS1 1.58e−08 0.17
PROSPER 0.014 0.20
ORCADES 0.085 0.19
OBB 3.00e−07 0.17
NFBC86 9.87e−07 0.17
NFBC66 0.001 0.16
NESDA 3.42e−04 0.16
NEO 1.31e−05 0.16
METSIM 2.40e−05 0.16
LOLIPOP_OmniE 0.014 0.14
LOLIPOP_OmniEE 0.581 0.13
LOLIPOP_IAP 0.412 0.12
LOLIPOP_IA610 1.28e−05 0.14
LOLIPOP_IA317 0.030 0.13
LOLIPOP_EWP 0.039 0.17
LOLIPOP_EWA 0.244 0.16
LOLIPOP_EW610 0.251 0.19
LLS 0.005 0.16
LLD 0.027 0.17
KORCULA 0.166 0.12
INT 2.70e−17 0.17
FR97 0.002 0.16
FR07 7.45e−06 0.16
EUGENDA 3.91e−04 0.16
ERF 0.083 0.14
EGCUT 1.42e−06 0.20
CKB 6.50e−06 0.18
ALSPAC 9.60e−07 0.18
META 3.81e−82 0.17
−0.2
0.0
0.2
BETA

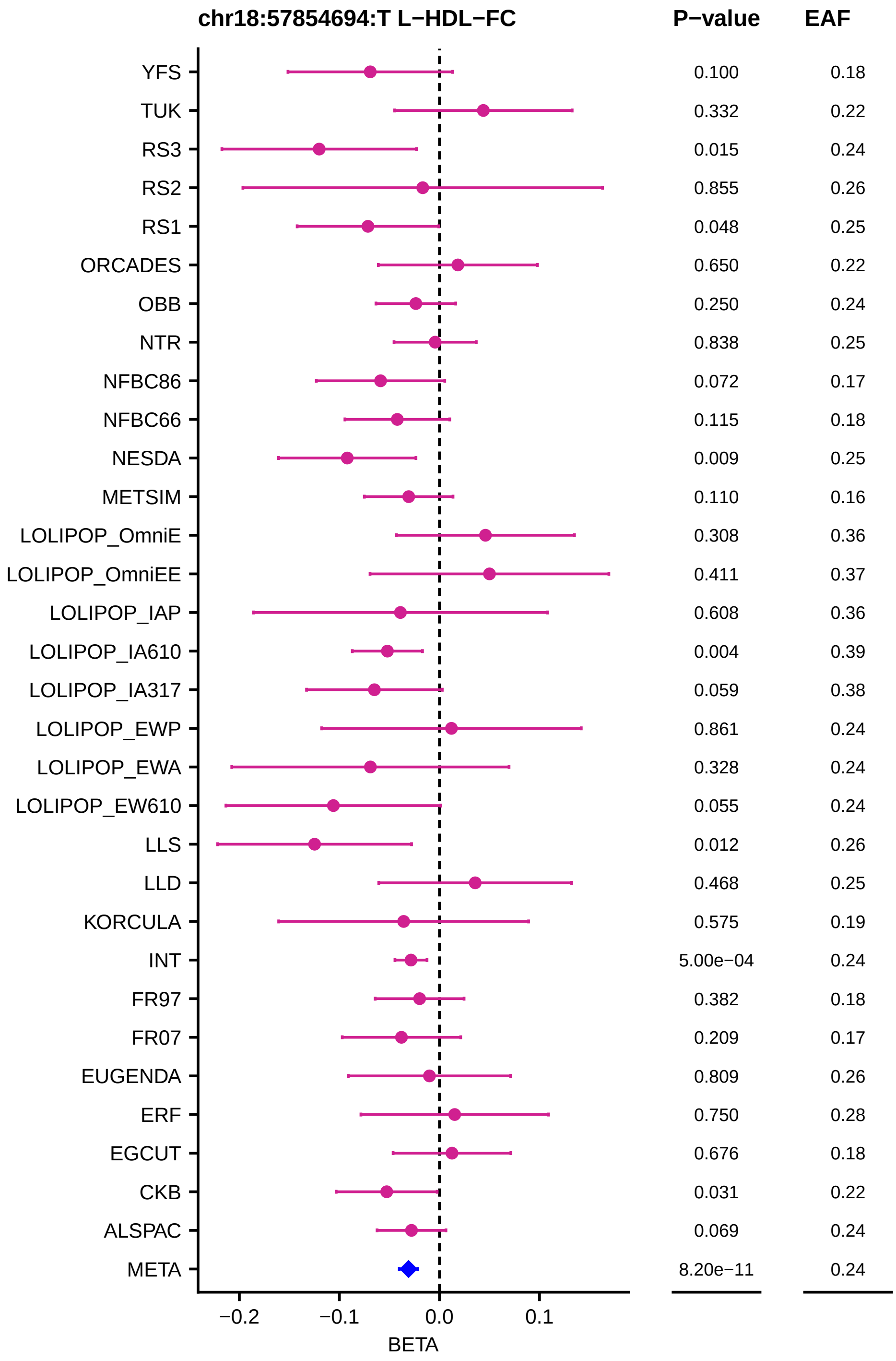
chr18:57854694:T L−HDL−FC
P−value
EAF
YFS 0.100 0.18
TUK 0.332 0.22
RS3 0.015 0.24
RS2 0.855 0.26
RS1 0.048 0.25
ORCADES 0.650 0.22
OBB 0.250 0.24
NTR 0.838 0.25
NFBC86 0.072 0.17
NFBC66 0.115 0.18
NESDA 0.009 0.25
METSIM 0.110 0.16
LOLIPOP_OmniE 0.308 0.36
LOLIPOP_OmniEE 0.411 0.37
LOLIPOP_IAP 0.608 0.36
LOLIPOP_IA610 0.004 0.39
LOLIPOP_IA317 0.059 0.38
LOLIPOP_EWP 0.861 0.24
LOLIPOP_EWA 0.328 0.24
LOLIPOP_EW610 0.055 0.24
LLS 0.012 0.26
LLD 0.468 0.25
KORCULA 0.575 0.19
INT 5.00e−04 0.24
FR97 0.382 0.18
FR07 0.209 0.17
EUGENDA 0.809 0.26
ERF 0.750 0.28
EGCUT 0.676 0.18
CKB 0.031 0.22
ALSPAC 0.069 0.24
META 8.20e−11 0.24
−0.2
−0.1
0.0
0.1
BETA

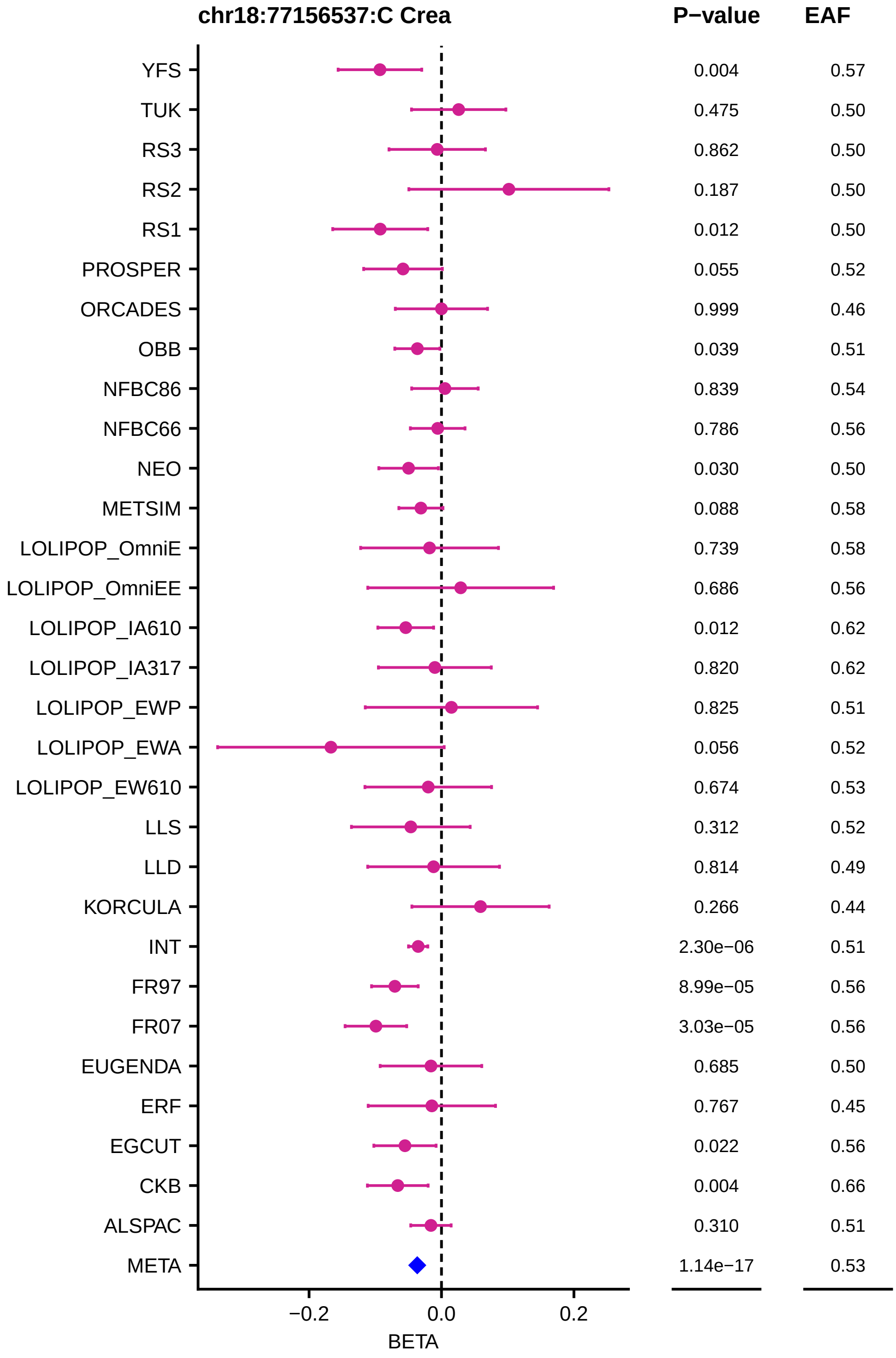
chr18:77156537:C Crea
P−value
EAF
YFS
TUK
RS3
RS2
RS1
PROSPER
ORCADES
OBB
NFBC86
NFBC66
NEO
METSIM
LOLIPOP_OmniE
LOLIPOP_OmniEE
LOLIPOP_IA610
LOLIPOP_IA317
LOLIPOP_EWP
LOLIPOP_EWA
LOLIPOP_EW610
LLS
LLD
KORCULA
INT
FR97
FR07
EUGENDA
ERF
EGCUT
CKB
ALSPAC
META
0.004 0.57
0.475 0.50
0.862 0.50
0.187 0.50
0.012 0.50
0.055 0.52
0.999 0.46
0.039 0.51
0.839 0.54
0.786 0.56
0.030 0.50
0.088 0.58
0.739 0.58
0.686 0.56
0.012 0.62
0.820 0.62
0.825 0.51
0.056 0.52
0.674 0.53
0.312 0.52
0.814 0.49
0.266 0.44
2.30e−06 0.51
8.99e−05 0.56
3.03e−05 0.56
0.685 0.50
0.767 0.45
0.022 0.56
0.004 0.66
0.310 0.51
1.14e−17 0.53
−0.2
0.0
0.2
BETA

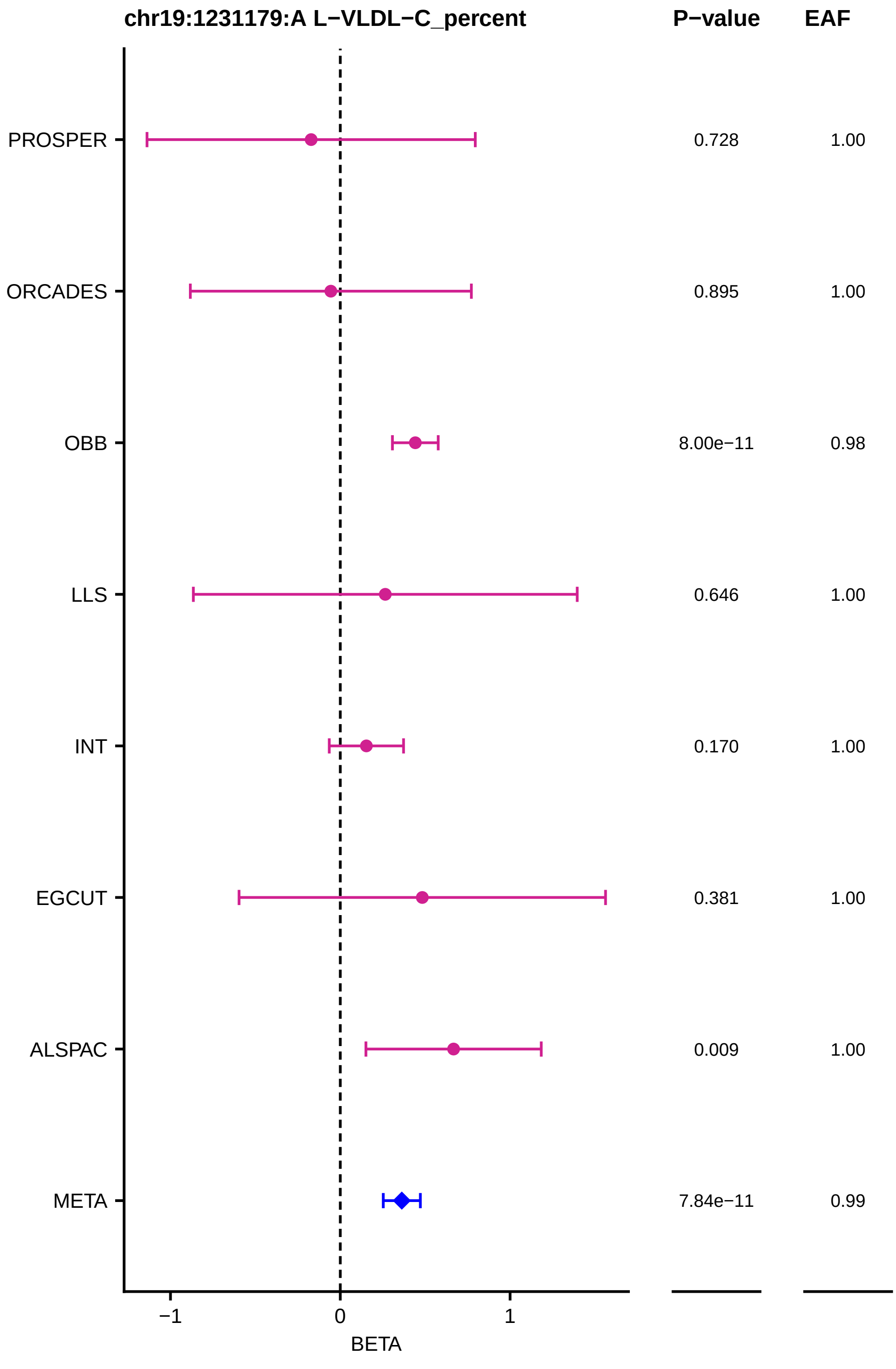
chr19:1231179:A L−VLDL−C_percent
P−value
EAF
PROSPER
0.728
1.00
ORCADES
0.895
1.00
OBB
8.00e−11
0.98
LLS
0.646
1.00
INT
0.170
1.00
EGCUT
0.381
1.00
ALSPAC
0.009
1.00
META
7.84e−11
0.99
−1
0
1
BETA

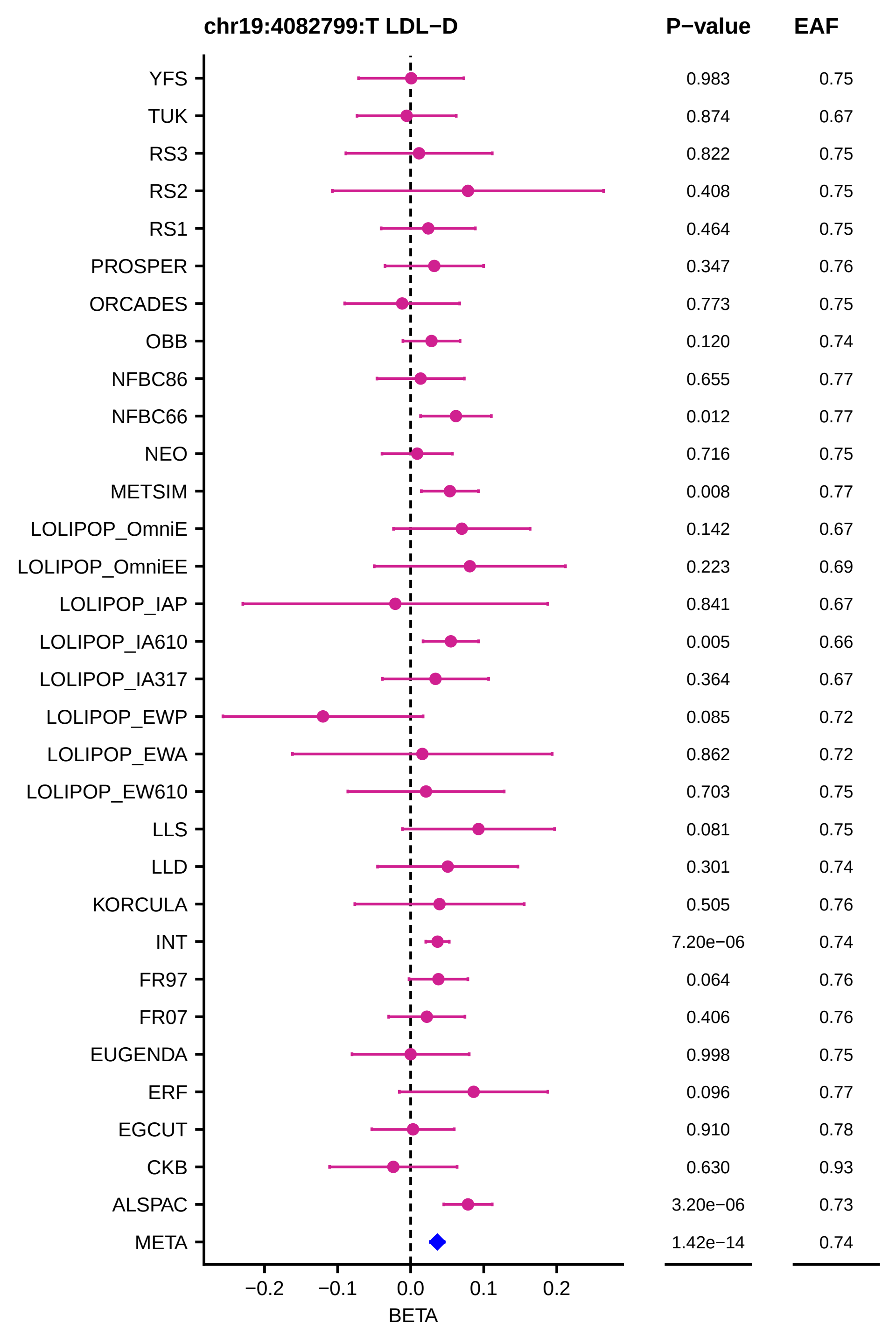
chr19:4082799:T LDL−D
P−value
EAF
YFS 0.983 0.75
TUK 0.874 0.67
RS3 0.822 0.75
RS2 0.408 0.75
RS1 0.464 0.75
PROSPER 0.347 0.76
ORCADES 0.773 0.75
OBB 0.120 0.74
NFBC86 0.655 0.77
NFBC66 0.012 0.77
NEO 0.716 0.75
METSIM 0.008 0.77
LOLIPOP_OmniE 0.142 0.67
LOLIPOP_OmniEE 0.223 0.69
LOLIPOP_IAP 0.841 0.67
LOLIPOP_IA610 0.005 0.66
LOLIPOP_IA317 0.364 0.67
LOLIPOP_EWP 0.085 0.72
LOLIPOP_EWA 0.862 0.72
LOLIPOP_EW610 0.703 0.75
LLS 0.081 0.75
LLD 0.301 0.74
KORCULA 0.505 0.76
INT 7.20e−06 0.74
FR97 0.064 0.76
FR07 0.406 0.76
EUGENDA 0.998 0.75
ERF 0.096 0.77
EGCUT 0.910 0.78
CKB 0.630 0.93
ALSPAC 3.20e−06 0.73
META 1.42e−14 0.74
−0.2
−0.1
0.0
0.1
0.2
BETA

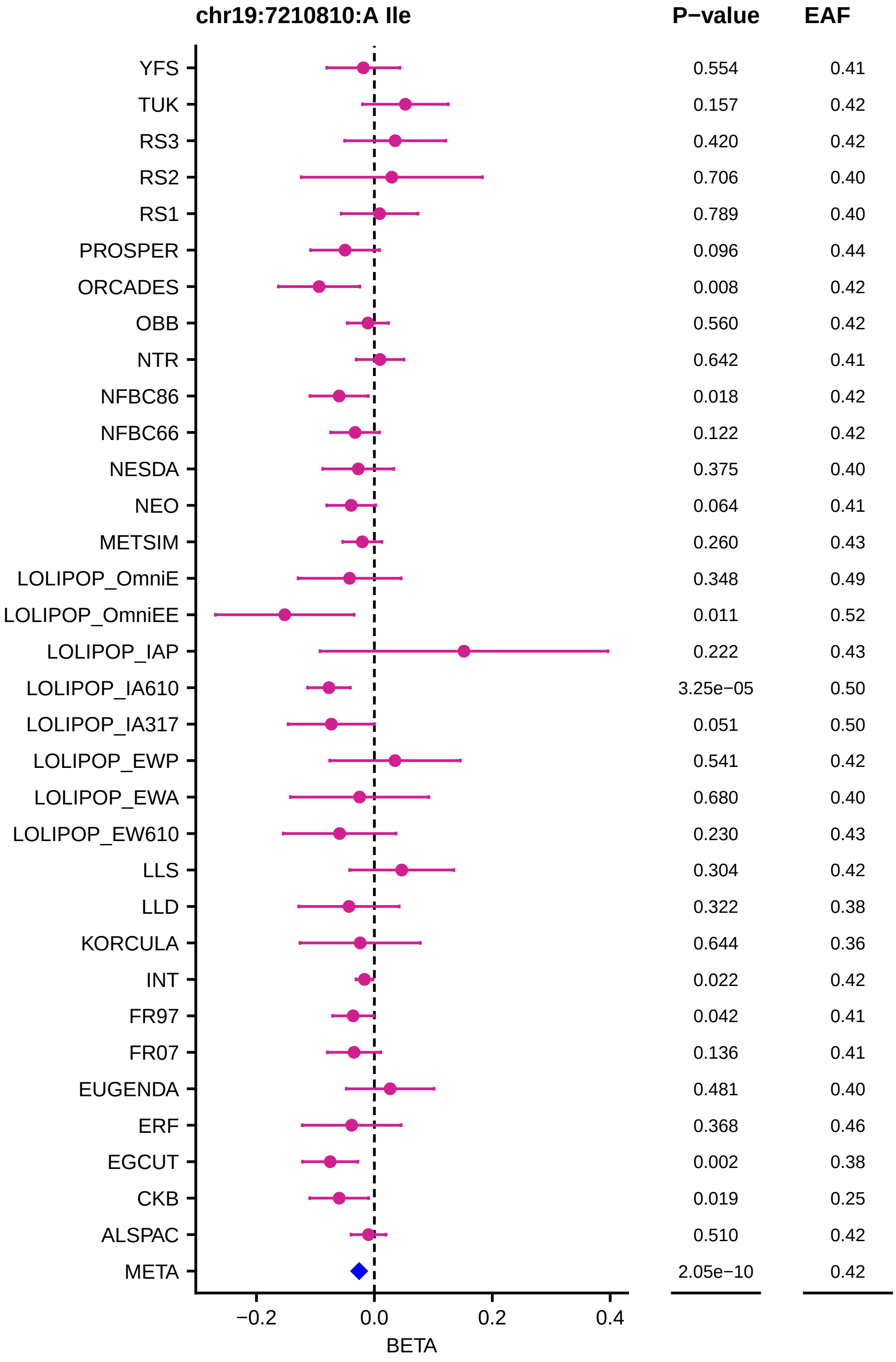
chr19:7210810:A Ile
P−value
EAF
YFS 0.554 0.41
TUK 0.157 0.42
RS3 0.420 0.42
RS2 0.706 0.40
RS1 0.789 0.40
PROSPER 0.096 0.44
ORCADES 0.008 0.42
OBB 0.560 0.42
NTR 0.642 0.41
NFBC86 0.018 0.42
NFBC66 0.122 0.42
NESDA 0.375 0.40
NEO 0.064 0.41
METSIM 0.260 0.43
LOLIPOP_OmniE 0.348 0.49
LOLIPOP_OmniEE 0.011 0.52
LOLIPOP_IAP 0.222 0.43
LOLIPOP_IA610 3.25e−05 0.50
LOLIPOP_IA317 0.051 0.50
LOLIPOP_EWP 0.541 0.42
LOLIPOP_EWA 0.680 0.40
LOLIPOP_EW610 0.230 0.43
LLS 0.304 0.42
LLD 0.322 0.38
KORCULA 0.644 0.36
INT 0.022 0.42
FR97 0.042 0.41
FR07 0.136 0.41
EUGENDA 0.481 0.40
ERF 0.368 0.46
EGCUT 0.002 0.38
CKB 0.019 0.25
ALSPAC 0.510 0.42
META 2.05e−10 0.42
−0.2
0.0
0.2
0.4
BETA

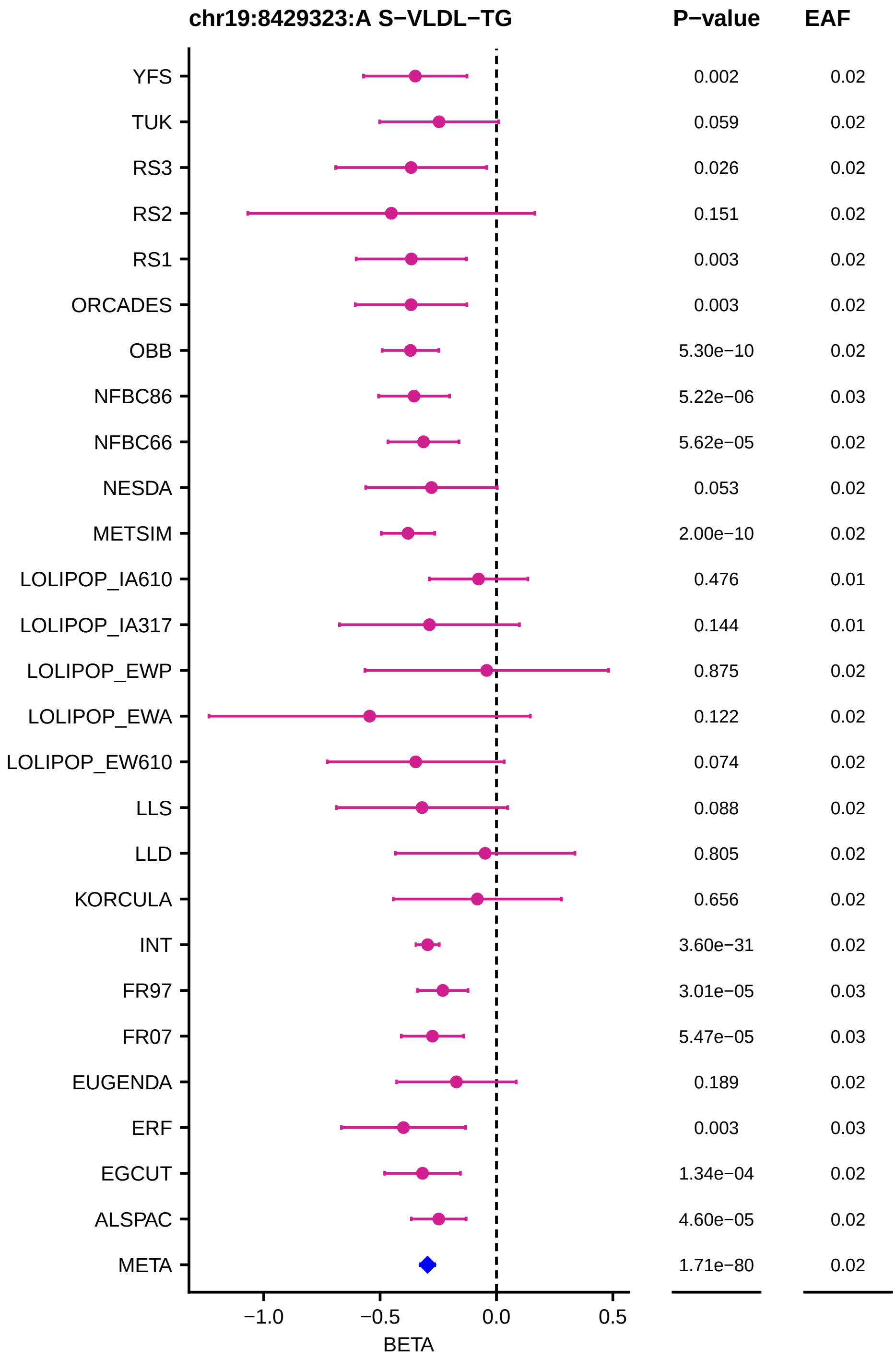
chr19:8429323:A S−VLDL−TG
P−value
EAF
YFS 0.002 0.02
TUK 0.059 0.02
RS3 0.026 0.02
RS2 0.151 0.02
RS1 0.003 0.02
ORCADES 0.003 0.02
OBB 5.30e−10 0.02
NFBC86 5.22e−06 0.03
NFBC66 5.62e−05 0.02
NESDA 0.053 0.02
METSIM 2.00e−10 0.02
LOLIPOP_IA610 0.476 0.01
LOLIPOP_IA317 0.144 0.01
LOLIPOP_EWP 0.875 0.02
LOLIPOP_EWA 0.122 0.02
LOLIPOP_EW610 0.074 0.02
LLS 0.088 0.02
LLD 0.805 0.02
KORCULA 0.656 0.02
INT 3.60e−31 0.02
FR97 3.01e−05 0.03
FR07 5.47e−05 0.03
EUGENDA 0.189 0.02
ERF 0.003 0.03
EGCUT 1.34e−04 0.02
ALSPAC 4.60e−05 0.02
META 1.71e−80 0.02
−1.0
−0.5
0.0
0.5
BETA

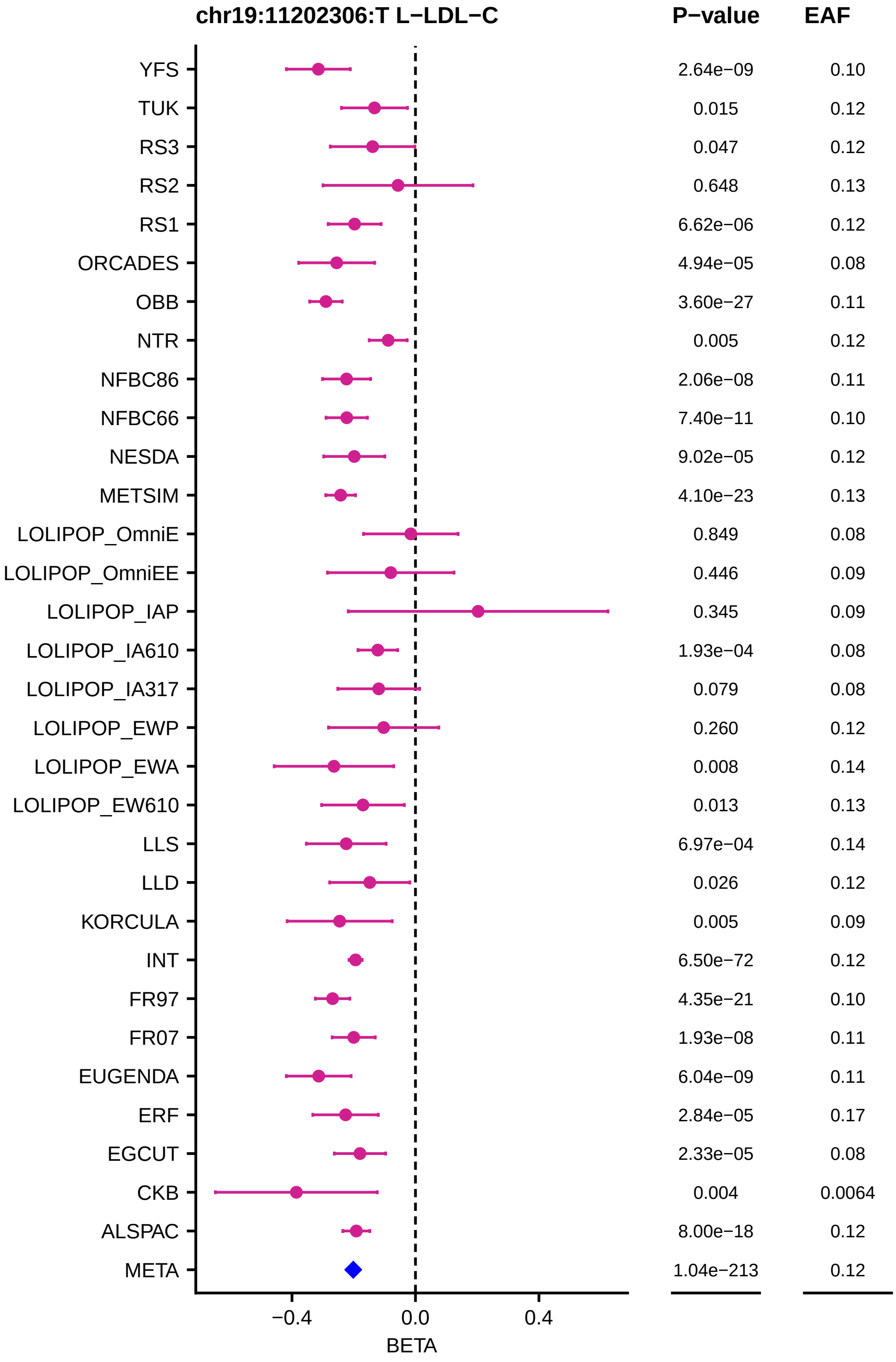
chr19:11202306:T L−LDL−C
P−value
EAF
YFS 2.64e−09 0.10
TUK 0.015 0.12
RS3 0.047 0.12
RS2 0.648 0.13
RS1 6.62e−06 0.12
ORCADES 4.94e−05 0.08
OBB 3.60e−27 0.11
NTR 0.005 0.12
NFBC86 2.06e−08 0.11
NFBC66 7.40e−11 0.10
NESDA 9.02e−05 0.12
METSIM 4.10e−23 0.13
LOLIPOP_OmniE 0.849 0.08
LOLIPOP_OmniEE 0.446 0.09
LOLIPOP_IAP 0.345 0.09
LOLIPOP_IA610 1.93e−04 0.08
LOLIPOP_IA317 0.079 0.08
LOLIPOP_EWP 0.260 0.12
LOLIPOP_EWA 0.008 0.14
LOLIPOP_EW610 0.013 0.13
LLS 6.97e−04 0.14
LLD 0.026 0.12
KORCULA 0.005 0.09
INT 6.50e−72 0.12
FR97 4.35e−21 0.10
FR07 1.93e−08 0.11
EUGENDA 6.04e−09 0.11
ERF 2.84e−05 0.17
EGCUT 2.33e−05 0.08
CKB 0.004 0.0064
ALSPAC 8.00e−18 0.12
META 1.04e−213 0.12
−0.4
0.0
0.4
BETA

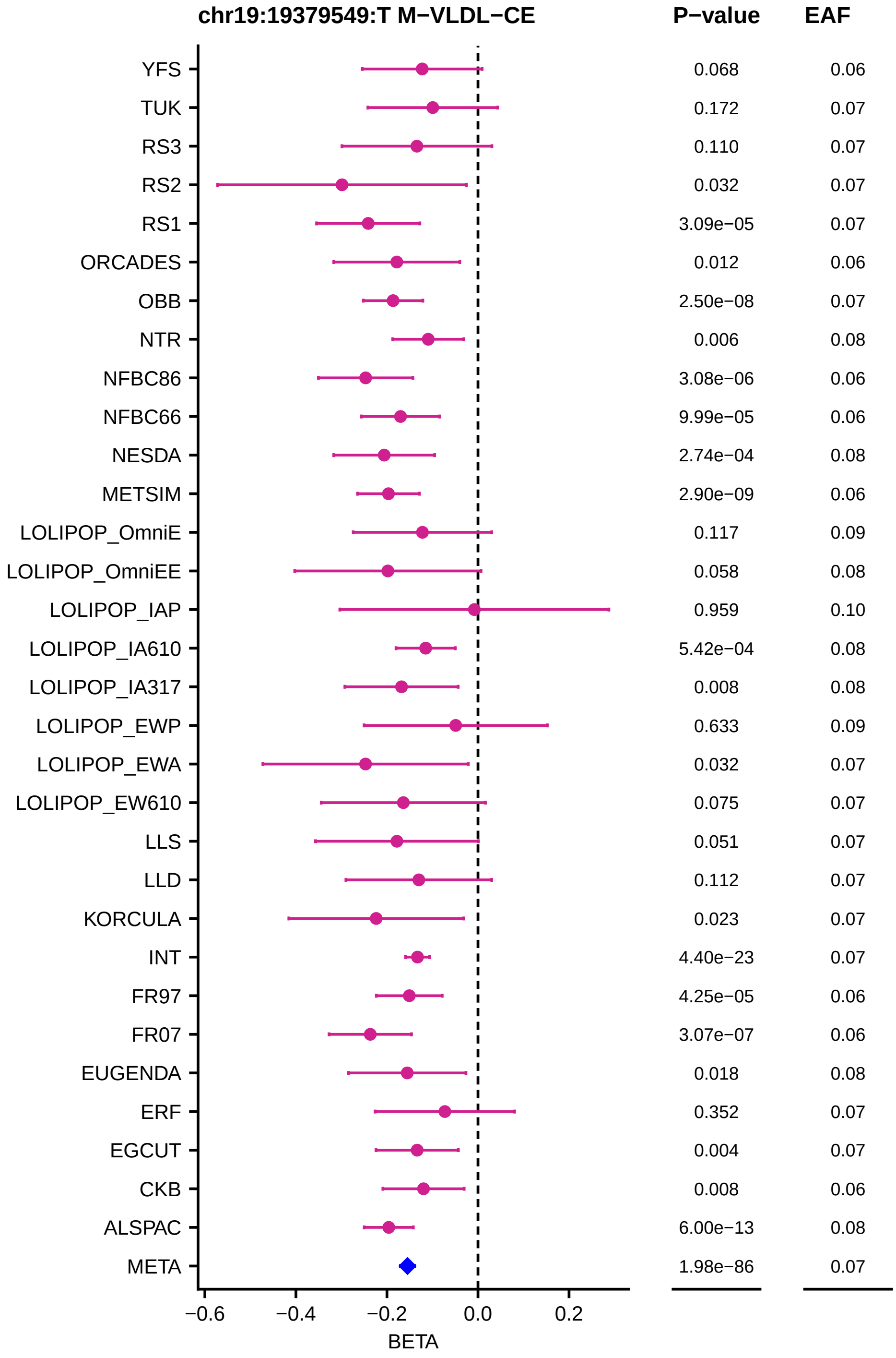
chr19:19379549:T M−VLDL−CE
P−value
EAF
YFS 0.068 0.06
TUK 0.172 0.07
RS3 0.110 0.07
RS2 0.032 0.07
RS1 3.09e−05 0.07
ORCADES 0.012 0.06
OBB 2.50e−08 0.07
NTR 0.006 0.08
NFBC86 3.08e−06 0.06
NFBC66 9.99e−05 0.06
NESDA 2.74e−04 0.08
METSIM 2.90e−09 0.06
LOLIPOP_OmniE 0.117 0.09
LOLIPOP_OmniEE 0.058 0.08
LOLIPOP_IAP 0.959 0.10
LOLIPOP_IA610 5.42e−04 0.08
LOLIPOP_IA317 0.008 0.08
LOLIPOP_EWP 0.633 0.09
LOLIPOP_EWA 0.032 0.07
LOLIPOP_EW610 0.075 0.07
LLS 0.051 0.07
LLD 0.112 0.07
KORCULA 0.023 0.07
INT 4.40e−23 0.07
FR97 4.25e−05 0.06
FR07 3.07e−07 0.06
EUGENDA 0.018 0.08
ERF 0.352 0.07
EGCUT 0.004 0.07
CKB 0.008 0.06
ALSPAC 6.00e−13 0.08
META 1.98e−86 0.07
−0.6 −0.4 −0.2 0.0 0.2
BETA

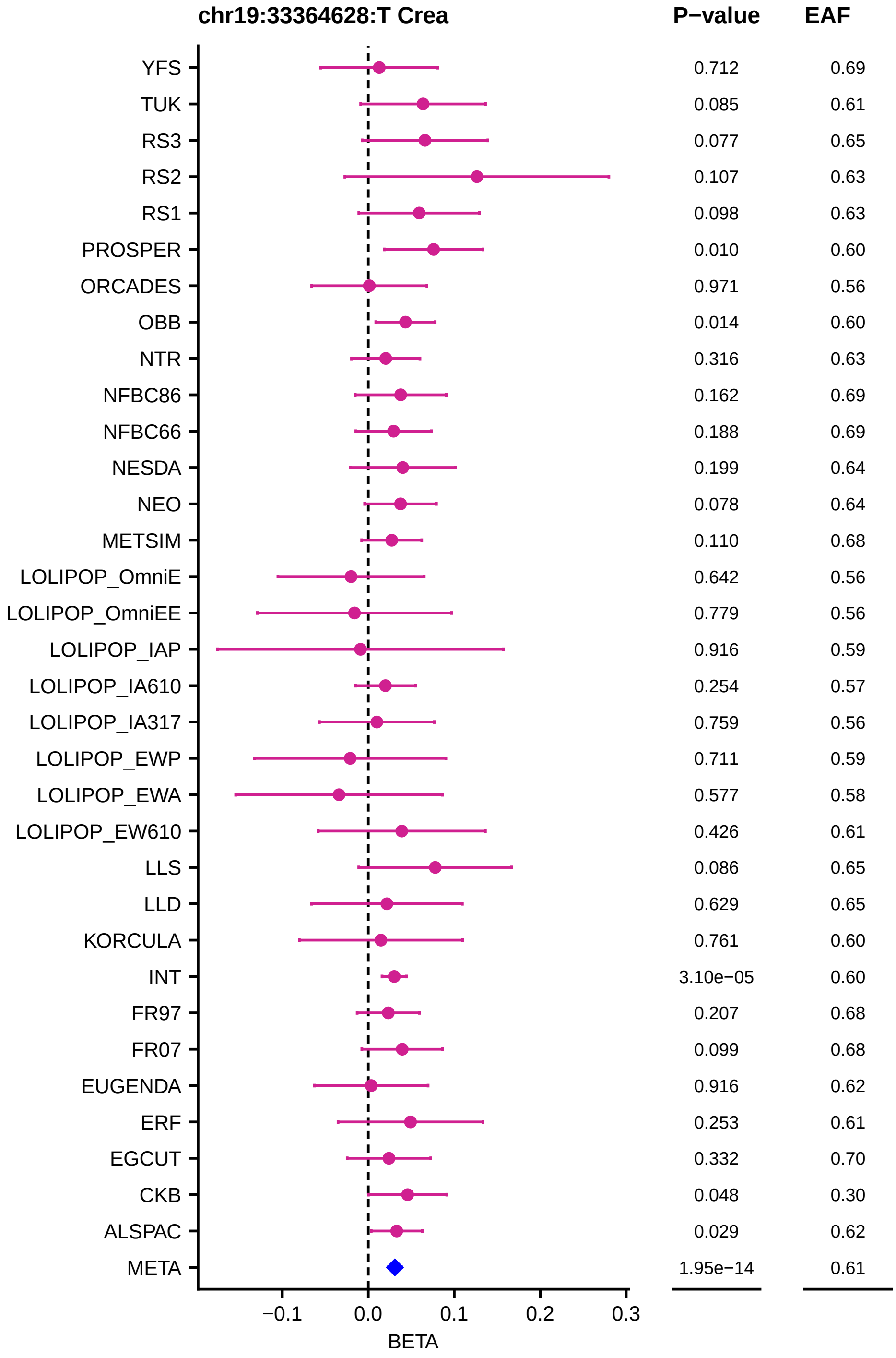

chr19:33364628:T Crea
P−value
EAF
YFS 0.712 0.69
TUK 0.085 0.61
RS3 0.077 0.65
RS2 0.107 0.63
RS1 0.098 0.63
PROSPER 0.010 0.60
ORCADES 0.971 0.56
OBB 0.014 0.60
NTR 0.316 0.63
NFBC86 0.162 0.69
NFBC66 0.188 0.69
NESDA 0.199 0.64
NEO 0.078 0.64
METSIM 0.110 0.68
LOLIPOP_OmniE 0.642 0.56
LOLIPOP_OmniEE 0.779 0.56
LOLIPOP_IAP 0.916 0.59
LOLIPOP_IA610 0.254 0.57
LOLIPOP_IA317 0.759 0.56
LOLIPOP_EWP 0.711 0.59
LOLIPOP_EWA 0.577 0.58
LOLIPOP_EW610 0.426 0.61
LLS 0.086 0.65
LLD 0.629 0.65
KORCULA 0.761 0.60
INT 3.10e−05 0.60
FR97 0.207 0.68
FR07 0.099 0.68
EUGENDA 0.916 0.62
ERF 0.253 0.61
EGCUT 0.332 0.70
CKB 0.048 0.30
ALSPAC 0.029 0.62
META 1.95e−14 0.61
−0.1
0.0
0.1
0.2
0.3
BETA

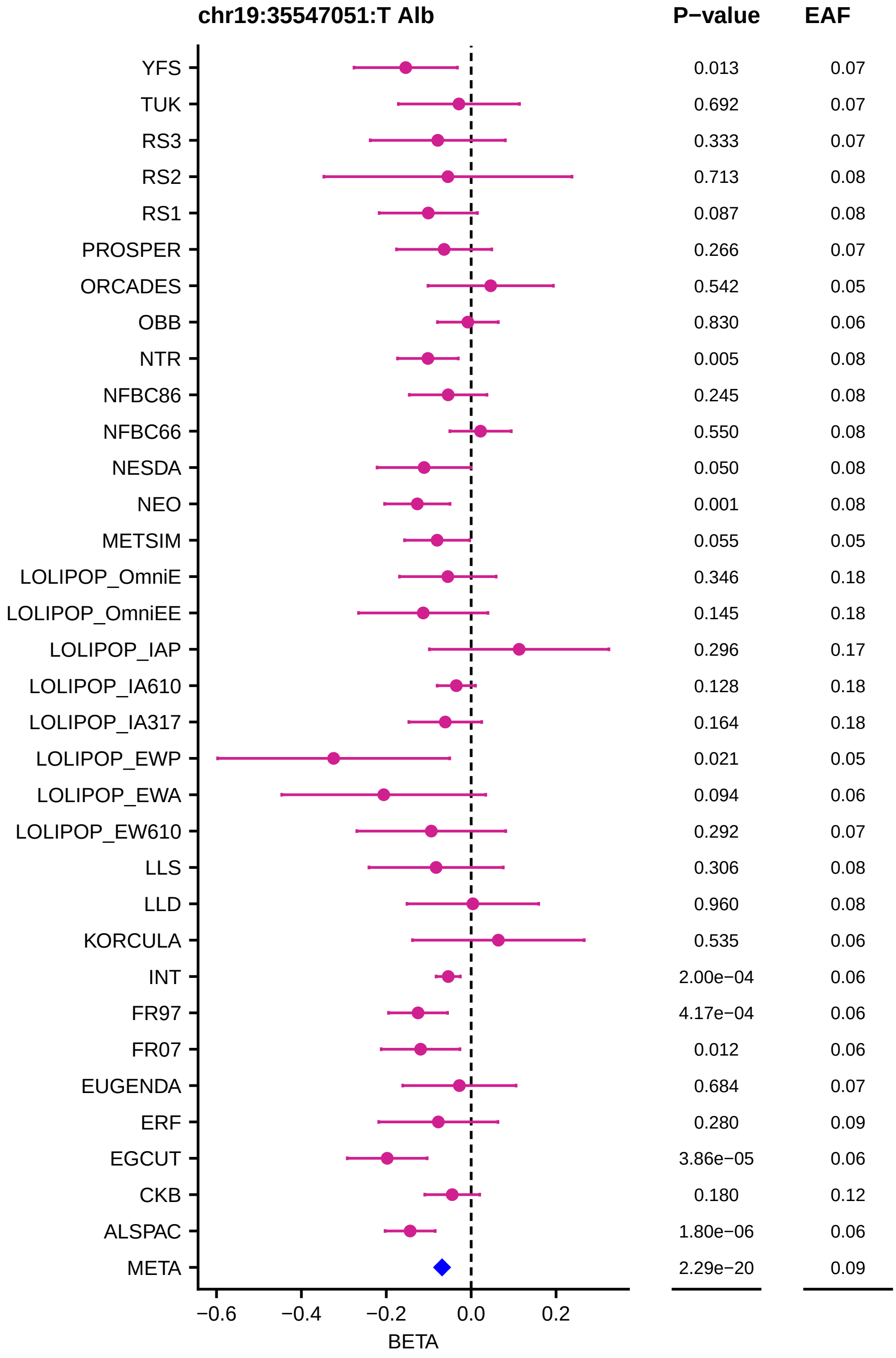
chr19:35547051:T Alb
P−value
EAF
YFS 0.013 0.07
TUK 0.692 0.07
RS3 0.333 0.07
RS2 0.713 0.08
RS1 0.087 0.08
PROSPER 0.266 0.07
ORCADES 0.542 0.05
OBB 0.830 0.06
NTR 0.005 0.08
NFBC86 0.245 0.08
NFBC66 0.550 0.08
NESDA 0.050 0.08
NEO 0.001 0.08
METSIM 0.055 0.05
LOLIPOP_OmniE 0.346 0.18
LOLIPOP_OmniEE 0.145 0.18
LOLIPOP_IAP 0.296 0.17
LOLIPOP_IA610 0.128 0.18
LOLIPOP_IA317 0.164 0.18
LOLIPOP_EWP 0.021 0.05
LOLIPOP_EWA 0.094 0.06
LOLIPOP_EW610 0.292 0.07
LLS 0.306 0.08
LLD 0.960 0.08
KORCULA 0.535 0.06
INT 2.00e−04 0.06
FR97 4.17e−04 0.06
FR07 0.012 0.06
EUGENDA 0.684 0.07
ERF 0.280 0.09
EGCUT 3.86e−05 0.06
CKB 0.180 0.12
ALSPAC 1.80e−06 0.06
META 2.29e−20 0.09
−0.6
−0.4
−0.2
0.0
0.2
BETA

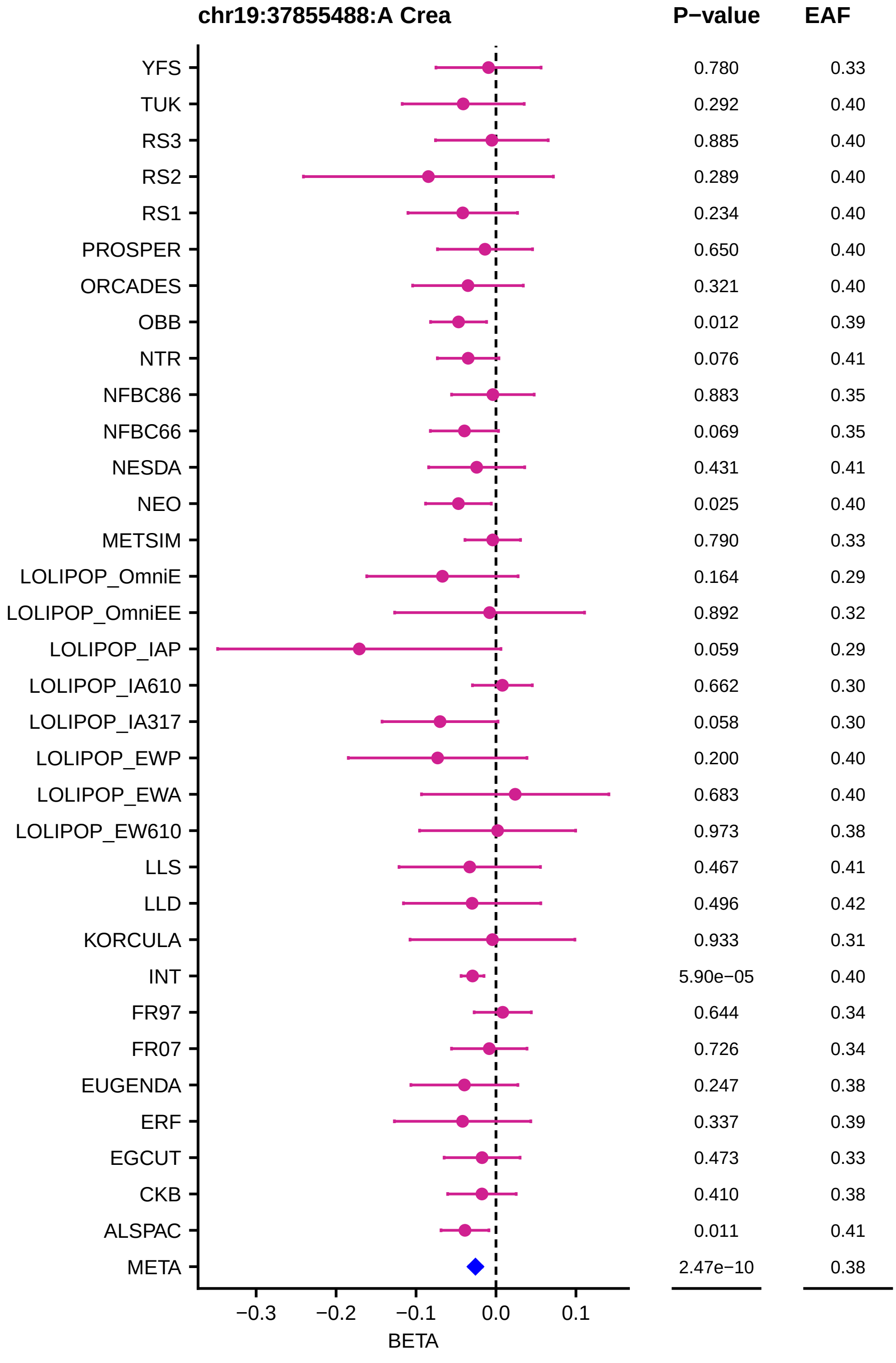
chr19:37855488:A Crea
P−value
EAF
YFS
TUK
RS3
RS2
RS1
PROSPER
ORCADES
OBB
NTR
NFBC86
NFBC66
NESDA
NEO
METSIM
LOLIPOP_OmniE
LOLIPOP_OmniEE
LOLIPOP_IAP
LOLIPOP_IA610
LOLIPOP_IA317
LOLIPOP_EWP
LOLIPOP_EWA
LOLIPOP_EW610
LLS
LLD
KORCULA
INT
FR97
FR07
EUGENDA
ERF
EGCUT
CKB
ALSPAC
META
0.780 0.33
0.292 0.40
0.885 0.40
0.289 0.40
0.234 0.40
0.650 0.40
0.321 0.40
0.012 0.39
0.076 0.41
0.883 0.35
0.069 0.35
0.431 0.41
0.025 0.40
0.790 0.33
0.164 0.29
0.892 0.32
0.059 0.29
0.662 0.30
0.058 0.30
0.200 0.40
0.683 0.40
0.973 0.38
0.467 0.41
0.496 0.42
0.933 0.31
5.90e−05 0.40
0.644 0.34
0.726 0.34
0.247 0.38
0.337 0.39
0.473 0.33
0.410 0.38
0.011 0.41
2.47e−10 0.38
−0.3
−0.2
−0.1
0.0
0.1
BETA

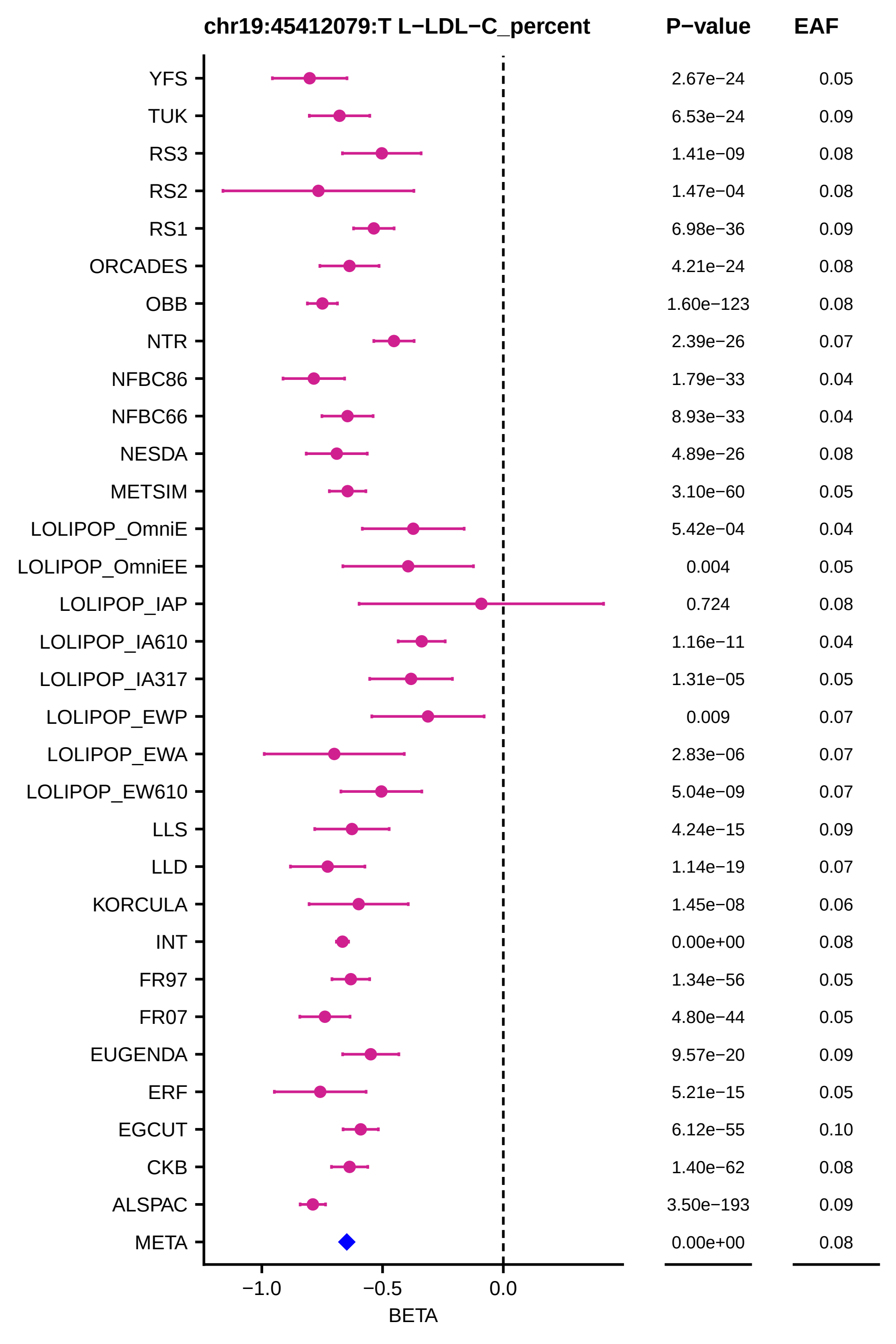
chr19:45412079:T L−LDL−C_percent
P−value
EAF
YFS 2.67e−24 0.05
TUK 6.53e−24 0.09
RS3 1.41e−09 0.08
RS2 1.47e−04 0.08
RS1 6.98e−36 0.09
ORCADES 4.21e−24 0.08
OBB 1.60e−123 0.08
NTR 2.39e−26 0.07
NFBC86 1.79e−33 0.04
NFBC66 8.93e−33 0.04
NESDA 4.89e−26 0.08
METSIM 3.10e−60 0.05
LOLIPOP_OmniE 5.42e−04 0.04
LOLIPOP_OmniEE 0.004 0.05
LOLIPOP_IAP 0.724 0.08
LOLIPOP_IA610 1.16e−11 0.04
LOLIPOP_IA317 1.31e−05 0.05
LOLIPOP_EWP 0.009 0.07
LOLIPOP_EWA 2.83e−06 0.07
LOLIPOP_EW610 5.04e−09 0.07
LLS 4.24e−15 0.09
LLD 1.14e−19 0.07
KORCULA 1.45e−08 0.06
INT 0.00e+00 0.08
FR97 1.34e−56 0.05
FR07 4.80e−44 0.05
EUGENDA 9.57e−20 0.09
ERF 5.21e−15 0.05
EGCUT 6.12e−55 0.10
CKB 1.40e−62 0.08
ALSPAC 3.50e−193 0.09
META 0.00e+00 0.08
−1.0
−0.5
0.0
BETA

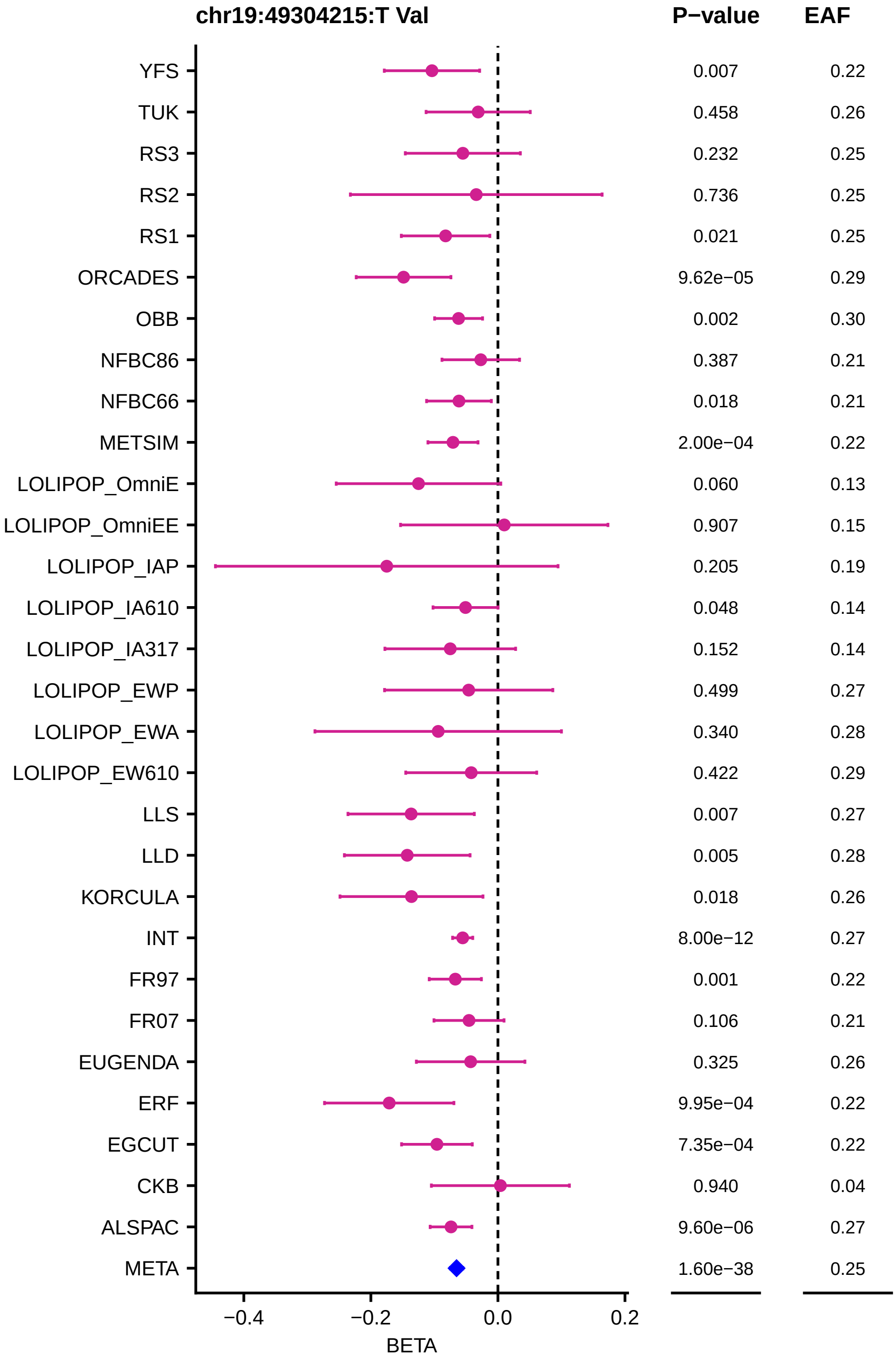
chr19:49304215:T Val
P−value
EAF
YFS 0.007 0.22
TUK 0.458 0.26
RS3 0.232 0.25
RS2 0.736 0.25
RS1 0.021 0.25
ORCADES 9.62e−05 0.29
OBB 0.002 0.30
NFBC86 0.387 0.21
NFBC66 0.018 0.21
METSIM 2.00e−04 0.22
LOLIPOP_OmniE 0.060 0.13
LOLIPOP_OmniEE 0.907 0.15
LOLIPOP_IAP 0.205 0.19
LOLIPOP_IA610 0.048 0.14
LOLIPOP_IA317 0.152 0.14
LOLIPOP_EWP 0.499 0.27
LOLIPOP_EWA 0.340 0.28
LOLIPOP_EW610 0.422 0.29
LLS 0.007 0.27
LLD 0.005 0.28
KORCULA 0.018 0.26
INT 8.00e−12 0.27
FR97 0.001 0.22
FR07 0.106 0.21
EUGENDA 0.325 0.26
ERF 9.95e−04 0.22
EGCUT 7.35e−04 0.22
CKB 0.940 0.04
ALSPAC 9.60e−06 0.27
META 1.60e−38 0.25
−0.4
−0.2
0.0
0.2
BETA

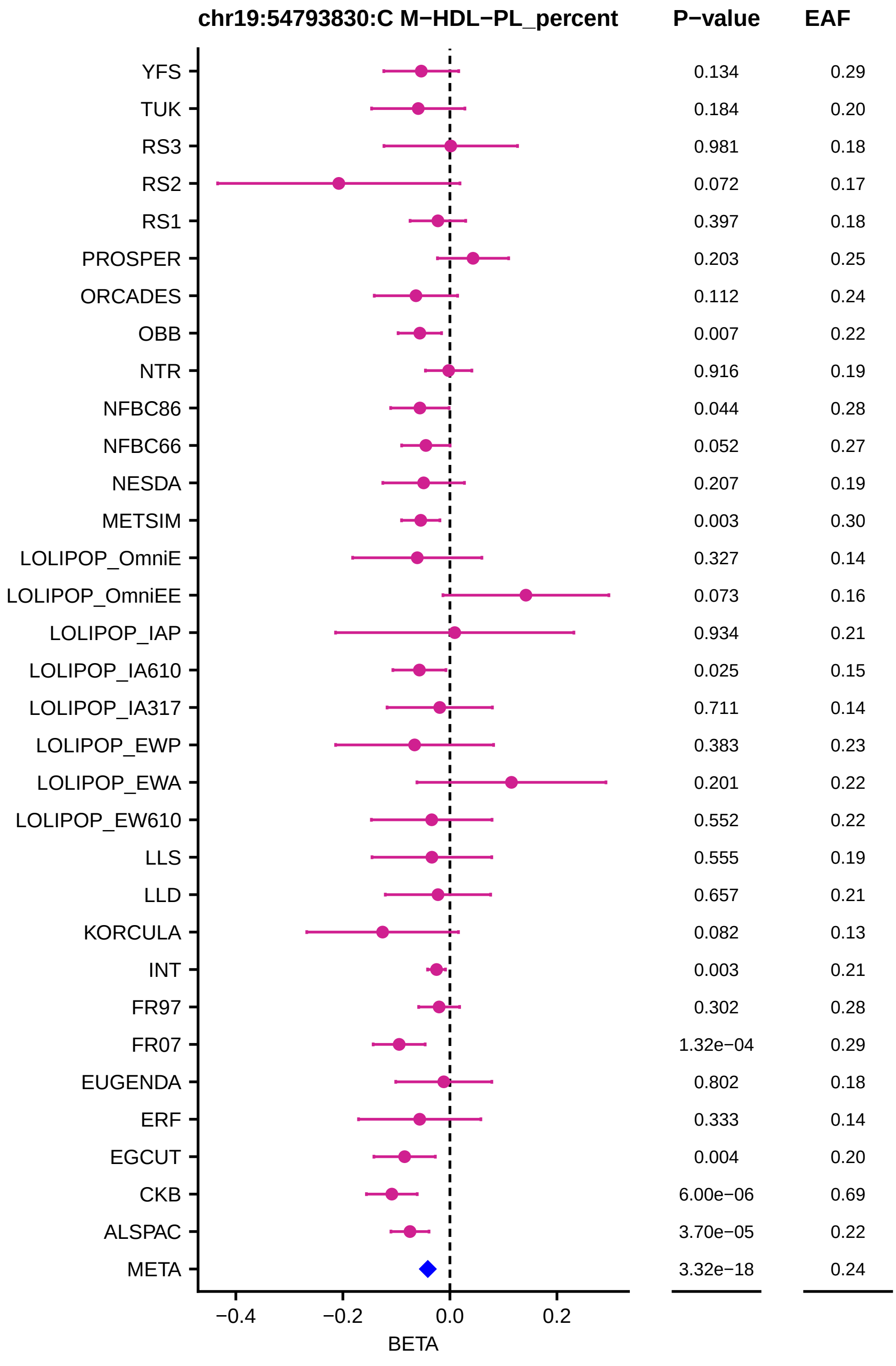
chr19:54793830:C M−HDL−PL_percent
P−value
EAF
YFS 0.134 0.29
TUK 0.184 0.20
RS3 0.981 0.18
RS2 0.072 0.17
RS1 0.397 0.18
PROSPER 0.203 0.25
ORCADES 0.112 0.24
OBB 0.007 0.22
NTR 0.916 0.19
NFBC86 0.044 0.28
NFBC66 0.052 0.27
NESDA 0.207 0.19
METSIM 0.003 0.30
LOLIPOP_OmniE 0.327 0.14
LOLIPOP_OmniEE 0.073 0.16
LOLIPOP_IAP 0.934 0.21
LOLIPOP_IA610 0.025 0.15
LOLIPOP_IA317 0.711 0.14
LOLIPOP_EWP 0.383 0.23
LOLIPOP_EWA 0.201 0.22
LOLIPOP_EW610 0.552 0.22
LLS 0.555 0.19
LLD 0.657 0.21
KORCULA 0.082 0.13
INT 0.003 0.21
FR97 0.302 0.28
FR07 1.32e−04 0.29
EUGENDA 0.802 0.18
ERF 0.333 0.14
EGCUT 0.004 0.20
CKB 6.00e−06 0.69
ALSPAC 3.70e−05 0.22
META 3.32e−18 0.24
−0.4
−0.2
0.0
0.2
BETA

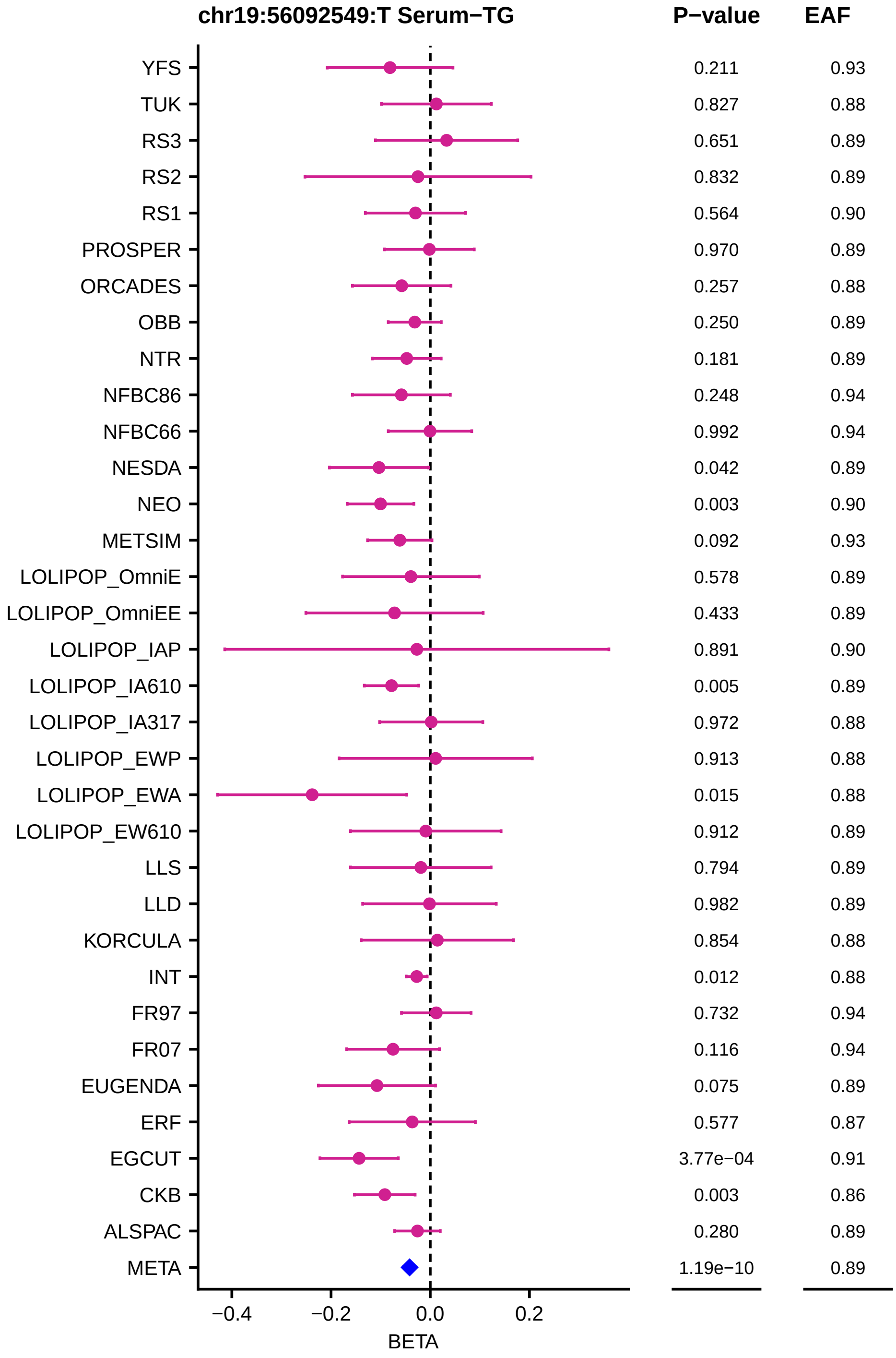
chr19:56092549:T Serum−TG
P−value
EAF
YFS
TUK
RS3
RS2
RS1
PROSPER
ORCADES
OBB
NTR
NFBC86
NFBC66
NESDA
NEO
METSIM
LOLIPOP_OmniE
LOLIPOP_OmniEE
LOLIPOP_IAP
LOLIPOP_IA610
LOLIPOP_IA317
LOLIPOP_EWP
LOLIPOP_EWA
LOLIPOP_EW610
LLS
LLD
KORCULA
INT
FR97
FR07
EUGENDA
ERF
EGCUT
CKB
ALSPAC
META
0.211 0.93
0.827 0.88
0.651 0.89
0.832 0.89
0.564 0.90
0.970 0.89
0.257 0.88
0.250 0.89
0.181 0.89
0.248 0.94
0.992 0.94
0.042 0.89
0.003 0.90
0.092 0.93
0.578 0.89
0.433 0.89
0.891 0.90
0.005 0.89
0.972 0.88
0.913 0.88
0.015 0.88
0.912 0.89
0.794 0.89
0.982 0.89
0.854 0.88
0.012 0.88
0.732 0.94
0.116 0.94
0.075 0.89
0.577 0.87
3.77e−04 0.91
0.003 0.86
0.280 0.89
1.19e−10 0.89
−0.4
−0.2
0.0
0.2
BETA

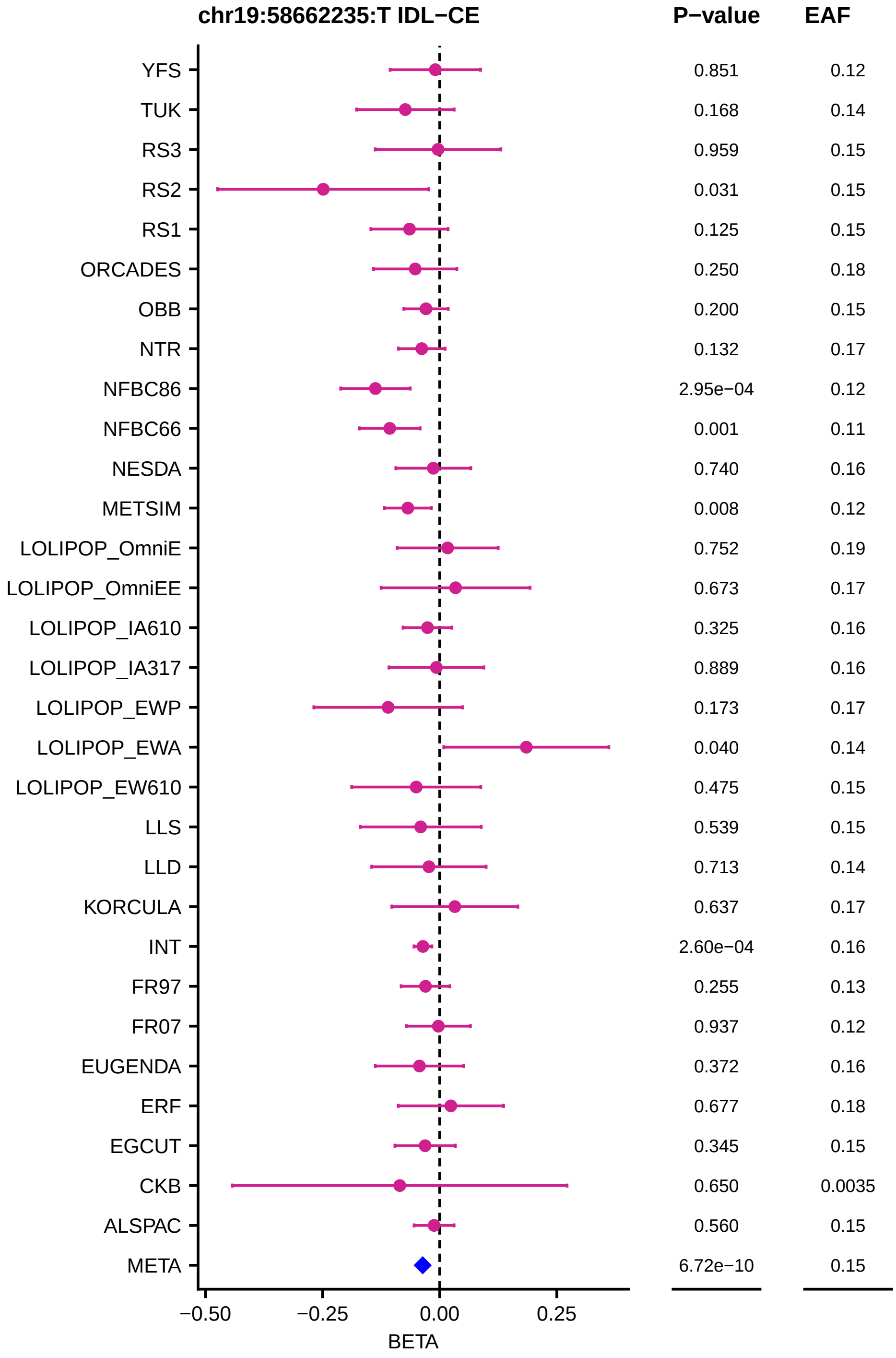

chr19:58662235:T IDL−CE
P−value
EAF
YFS
0.851
0.12
TUK
0.168
0.14
RS3
0.959
0.15
RS2
0.031
0.15
RS1
0.125
0.15
ORCADES
0.250
0.18
OBB
0.200
0.15
NTR
0.132
0.17
NFBC86
2.95e−04
0.12
NFBC66
0.001
0.11
NESDA
0.740
0.16
METSIM
0.008
0.12
LOLIPOP_OmniE
0.752
0.19
LOLIPOP_OmniEE
0.673
0.17
LOLIPOP_IA610
0.325
0.16
LOLIPOP_IA317
0.889
0.16
LOLIPOP_EWP
0.173
0.17
LOLIPOP_EWA
0.040
0.14
LOLIPOP_EW610
0.475
0.15
LLS
0.539
0.15
LLD
0.713
0.14
KORCULA
0.637
0.17
INT
2.60e−04
0.16
FR97
0.255
0.13
FR07
0.937
0.12
EUGENDA
0.372
0.16
ERF
0.677
0.18
EGCUT
0.345
0.15
CKB
0.650
0.0035
ALSPAC
0.560
0.15
META
6.72e−10
0.15
−0.50
−0.25
0.00
0.25
BETA

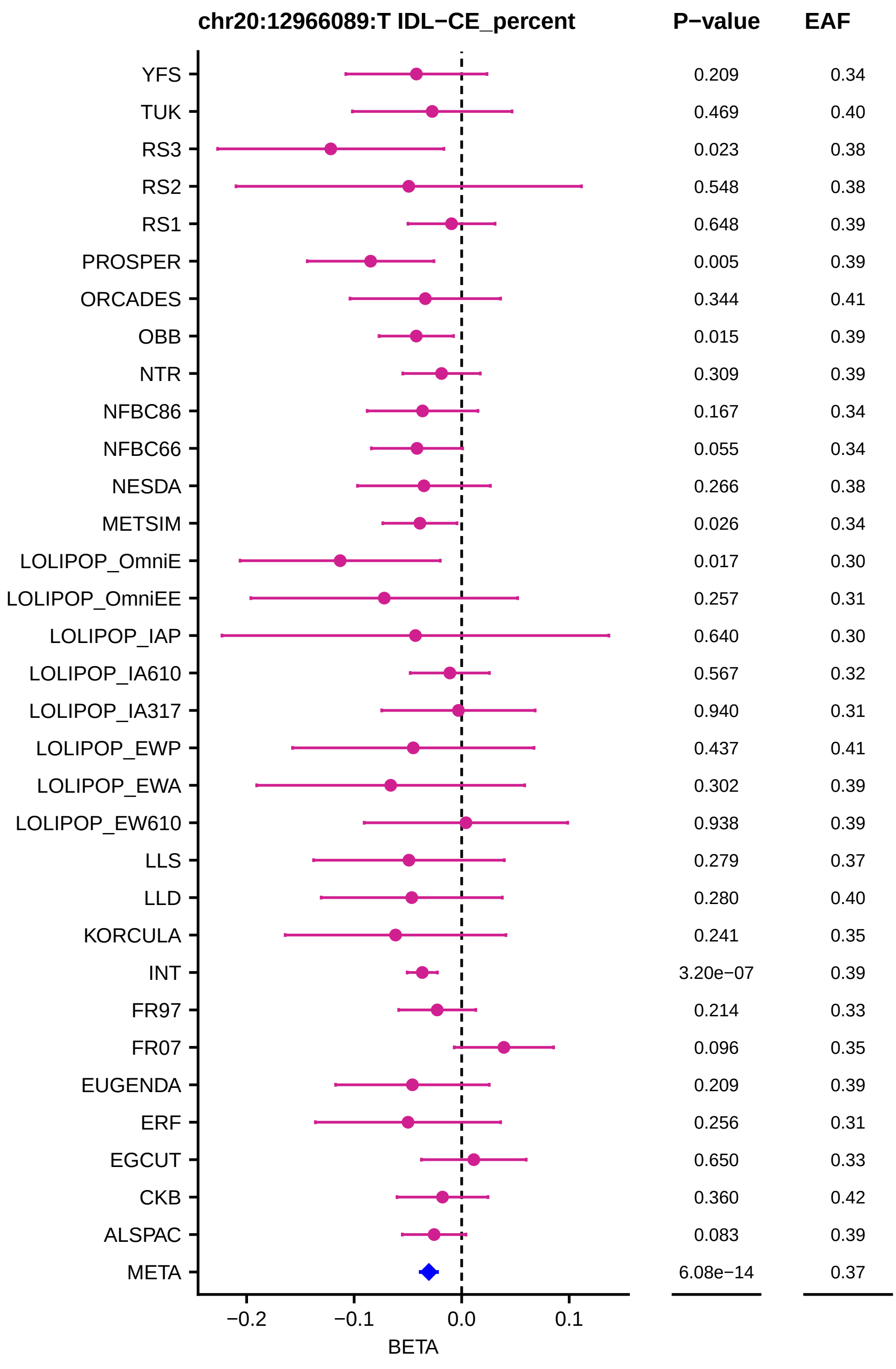
chr20:12966089:T IDL−CE_percent
P−value
EAF
YFS 0.209 0.34
TUK 0.469 0.40
RS3 0.023 0.38
RS2 0.548 0.38
RS1 0.648 0.39
PROSPER 0.005 0.39
ORCADES 0.344 0.41
OBB 0.015 0.39
NTR 0.309 0.39
NFBC86 0.167 0.34
NFBC66 0.055 0.34
NESDA 0.266 0.38
METSIM 0.026 0.34
LOLIPOP_OmniE 0.017 0.30
LOLIPOP_OmniEE 0.257 0.31
LOLIPOP_IAP 0.640 0.30
LOLIPOP_IA610 0.567 0.32
LOLIPOP_IA317 0.940 0.31
LOLIPOP_EWP 0.437 0.41
LOLIPOP_EWA 0.302 0.39
LOLIPOP_EW610 0.938 0.39
LLS 0.279 0.37
LLD 0.280 0.40
KORCULA 0.241 0.35
INT 3.20e−07 0.39
FR97 0.214 0.33
FR07 0.096 0.35
EUGENDA 0.209 0.39
ERF 0.256 0.31
EGCUT 0.650 0.33
CKB 0.360 0.42
ALSPAC 0.083 0.39
META 6.08e−14 0.37
−0.2
−0.1
0.0
0.1
BETA

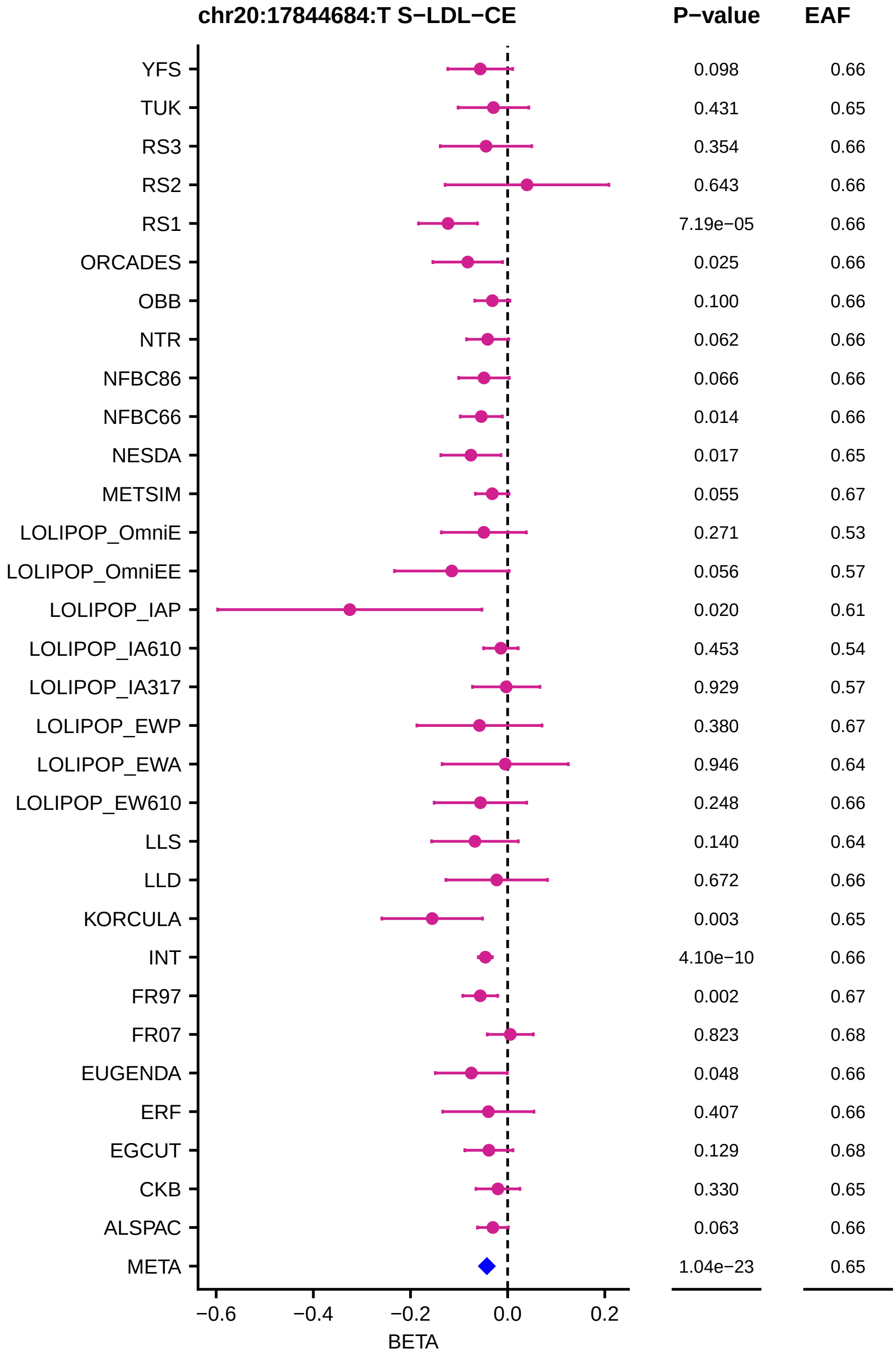

chr20:17844684:T S−LDL−CE
P−value
EAF
YFS 0.098 0.66
TUK 0.431 0.65
RS3 0.354 0.66
RS2 0.643 0.66
RS1 7.19e−05 0.66
ORCADES 0.025 0.66
OBB 0.100 0.66
NTR 0.062 0.66
NFBC86 0.066 0.66
NFBC66 0.014 0.66
NESDA 0.017 0.65
METSIM 0.055 0.67
LOLIPOP_OmniE 0.271 0.53
LOLIPOP_OmniEE 0.056 0.57
LOLIPOP_IAP 0.020 0.61
LOLIPOP_IA610 0.453 0.54
LOLIPOP_IA317 0.929 0.57
LOLIPOP_EWP 0.380 0.67
LOLIPOP_EWA 0.946 0.64
LOLIPOP_EW610 0.248 0.66
LLS 0.140 0.64
LLD 0.672 0.66
KORCULA 0.003 0.65
INT 4.10e−10 0.66
FR97 0.002 0.67
FR07 0.823 0.68
EUGENDA 0.048 0.66
ERF 0.407 0.66
EGCUT 0.129 0.68
CKB 0.330 0.65
ALSPAC 0.063 0.66
META 1.04e−23 0.65
−0.6
−0.4
−0.2
0.0
0.2
BETA

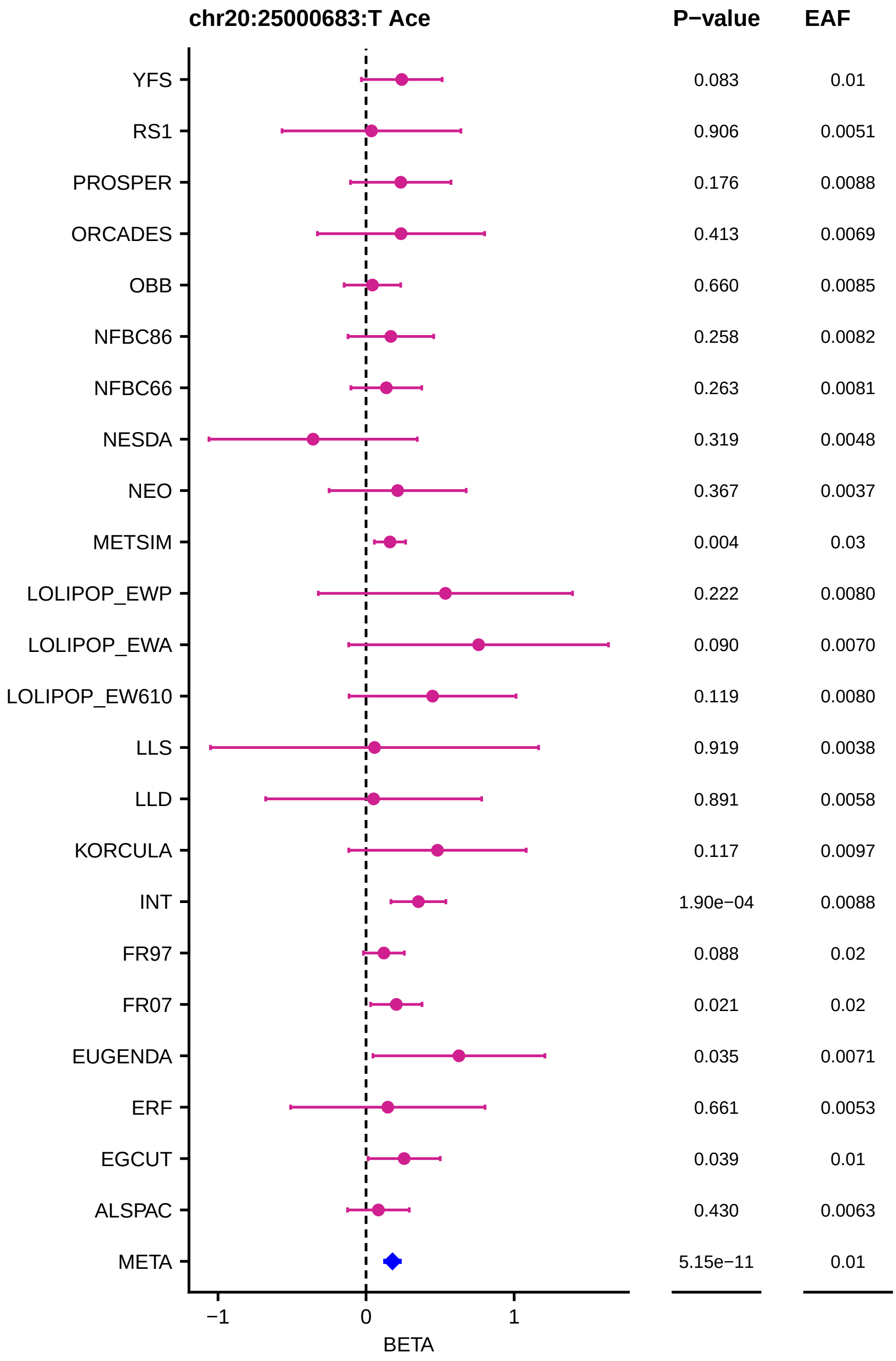
chr20:25000683:T Ace
P−value
EAF
YFS 0.083 0.01
RS1 0.906 0.0051
PROSPER 0.176 0.0088
ORCADES 0.413 0.0069
OBB 0.660 0.0085
NFBC86 0.258 0.0082
NFBC66 0.263 0.0081
NESDA 0.319 0.0048
NEO 0.367 0.0037
METSIM 0.004 0.03
LOLIPOP_EWP 0.222 0.0080
LOLIPOP_EWA 0.090 0.0070
LOLIPOP_EW610 0.119 0.0080
LLS 0.919 0.0038
LLD 0.891 0.0058
KORCULA 0.117 0.0097
INT 1.90e−04 0.0088
FR97 0.088 0.02
FR07 0.021 0.02
EUGENDA 0.035 0.0071
ERF 0.661 0.0053
EGCUT 0.039 0.01
ALSPAC 0.430 0.0063
META 5.15e−11 0.01
−1
0
1
BETA

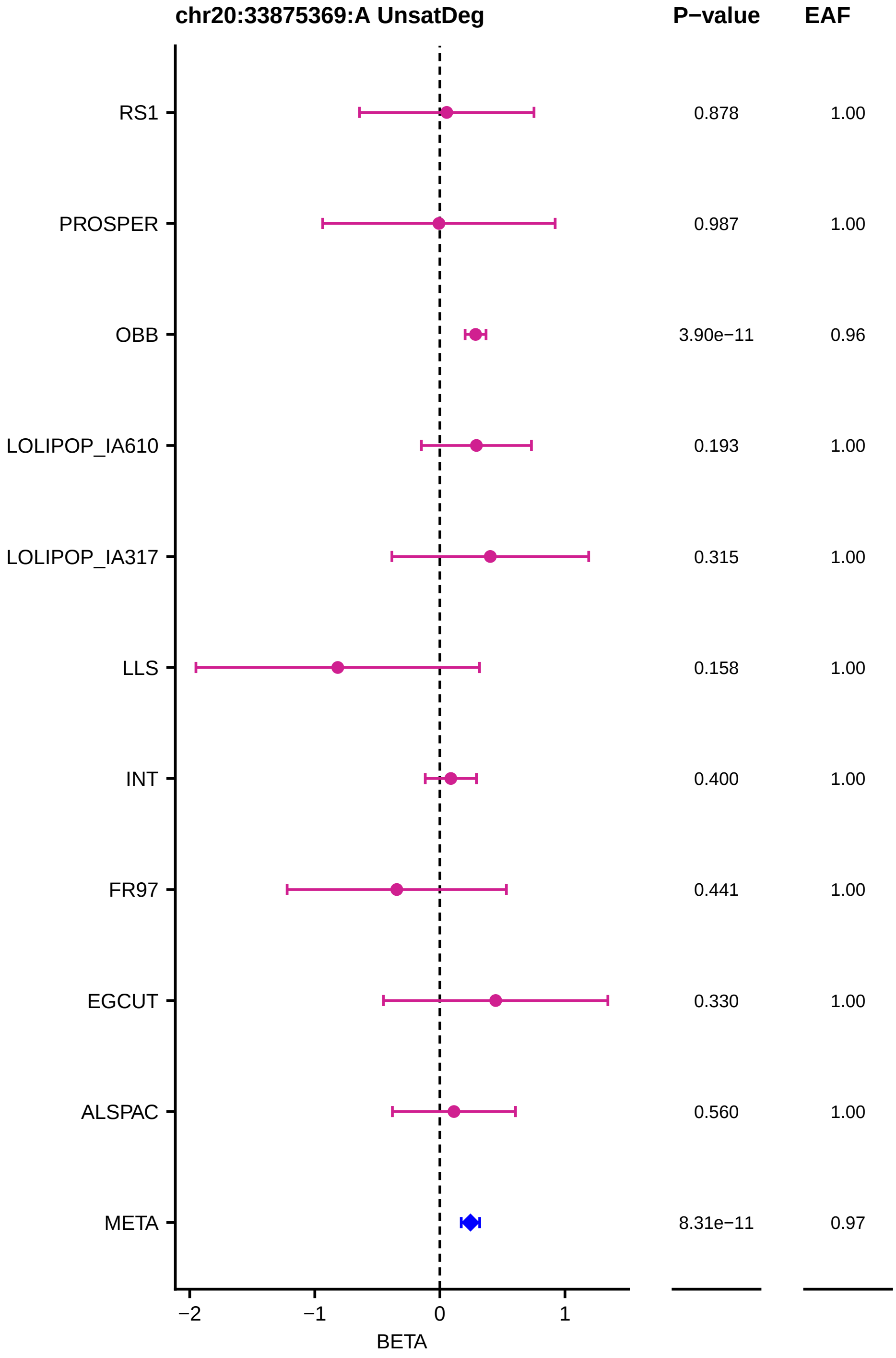
chr20:33875369:A UnsatDeg
P−value
EAF
RS1
PROSPER
OBB
LOLIPOP_IA610
LOLIPOP_IA317
LLS
INT
FR97
EGCUT
ALSPAC
META
0.878
0.987
3.90e−11
0.193
0.315
0.158
0.400
0.441
0.330
0.560
8.31e−11
1.00
1.00
0.96
1.00
1.00
1.00
1.00
1.00
1.00
1.00
0.97
−2
−1
0
1
BETA

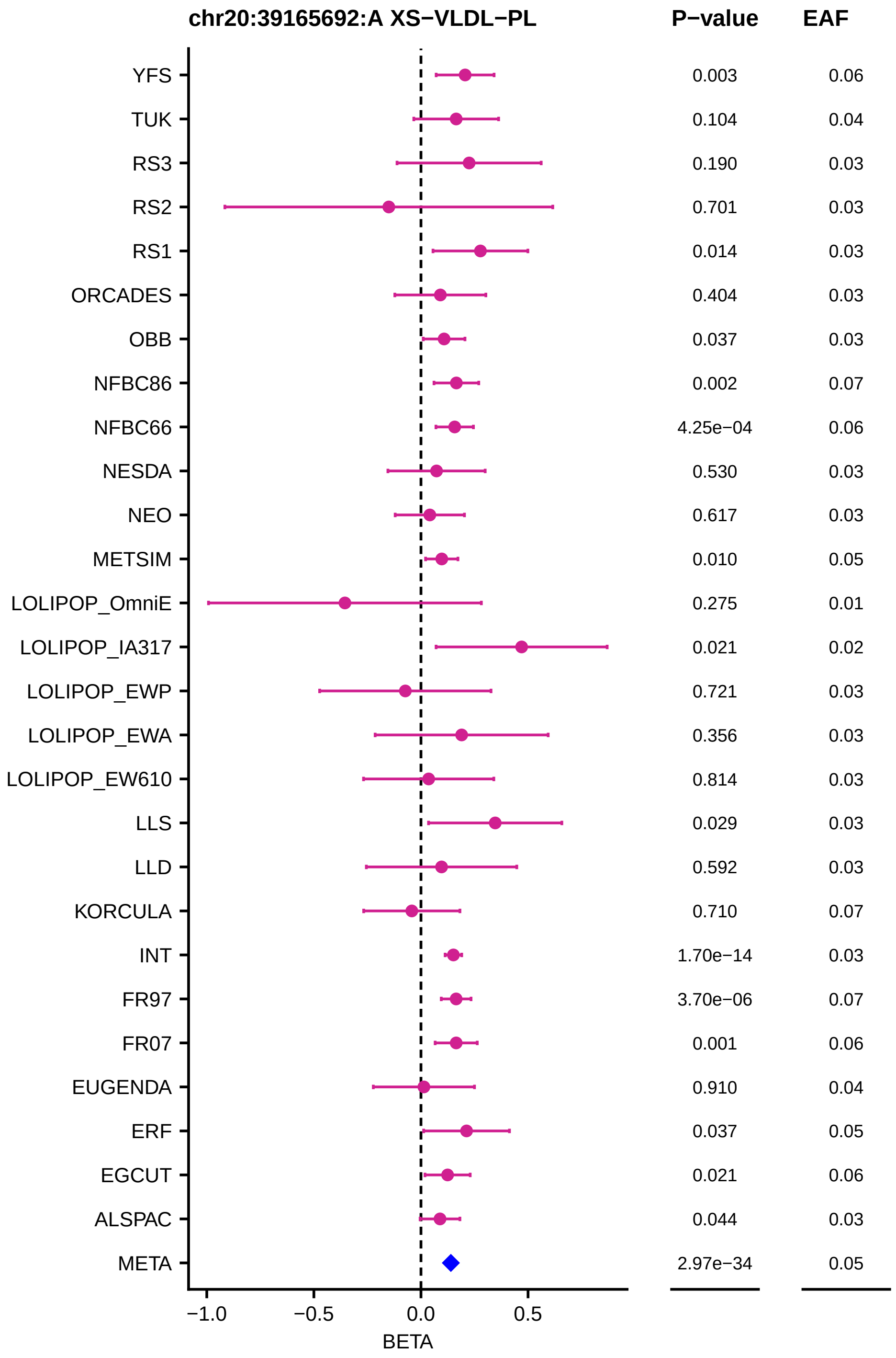
chr20:39165692:A XS−VLDL−PL
P−value
EAF
YFS 0.003 0.06
TUK 0.104 0.04
RS3 0.190 0.03
RS2 0.701 0.03
RS1 0.014 0.03
ORCADES 0.404 0.03
OBB 0.037 0.03
NFBC86 0.002 0.07
NFBC66 4.25e−04 0.06
NESDA 0.530 0.03
NEO 0.617 0.03
METSIM 0.010 0.05
LOLIPOP_OmniE 0.275 0.01
LOLIPOP_IA317 0.021 0.02
LOLIPOP_EWP 0.721 0.03
LOLIPOP_EWA 0.356 0.03
LOLIPOP_EW610 0.814 0.03
LLS 0.029 0.03
LLD 0.592 0.03
KORCULA 0.710 0.07
INT 1.70e−14 0.03
FR97 3.70e−06 0.07
FR07 0.001 0.06
EUGENDA 0.910 0.04
ERF 0.037 0.05
EGCUT 0.021 0.06
ALSPAC 0.044 0.03
META 2.97e−34 0.05
−1.0
−0.5
0.0
0.5
BETA

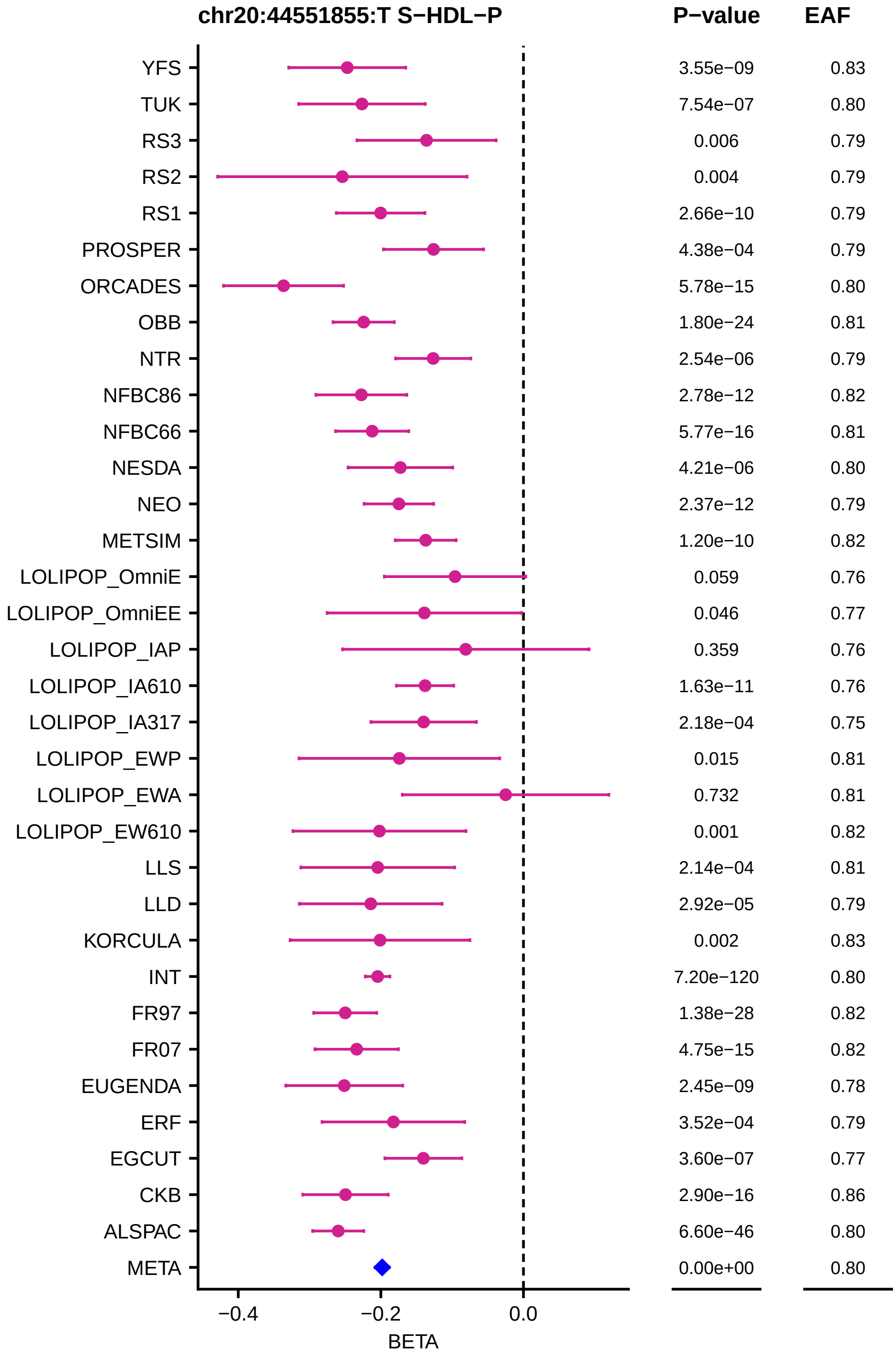

chr20:44551855:T S−HDL−P
P−value
EAF
YFS 3.55e−09 0.83
TUK 7.54e−07 0.80
RS3 0.006 0.79
RS2 0.004 0.79
RS1 2.66e−10 0.79
PROSPER 4.38e−04 0.79
ORCADES 5.78e−15 0.80
OBB 1.80e−24 0.81
NTR 2.54e−06 0.79
NFBC86 2.78e−12 0.82
NFBC66 5.77e−16 0.81
NESDA 4.21e−06 0.80
NEO 2.37e−12 0.79
METSIM 1.20e−10 0.82
LOLIPOP_OmniE 0.059 0.76
LOLIPOP_OmniEE 0.046 0.77
LOLIPOP_IAP 0.359 0.76
LOLIPOP_IA610 1.63e−11 0.76
LOLIPOP_IA317 2.18e−04 0.75
LOLIPOP_EWP 0.015 0.81
LOLIPOP_EWA 0.732 0.81
LOLIPOP_EW610 0.001 0.82
LLS 2.14e−04 0.81
LLD 2.92e−05 0.79
KORCULA 0.002 0.83
INT 7.20e−120 0.80
FR97 1.38e−28 0.82
FR07 4.75e−15 0.82
EUGENDA 2.45e−09 0.78
ERF 3.52e−04 0.79
EGCUT 3.60e−07 0.77
CKB 2.90e−16 0.86
ALSPAC 6.60e−46 0.80
META 0.00e+00 0.80
−0.4
−0.2
0.0
BETA

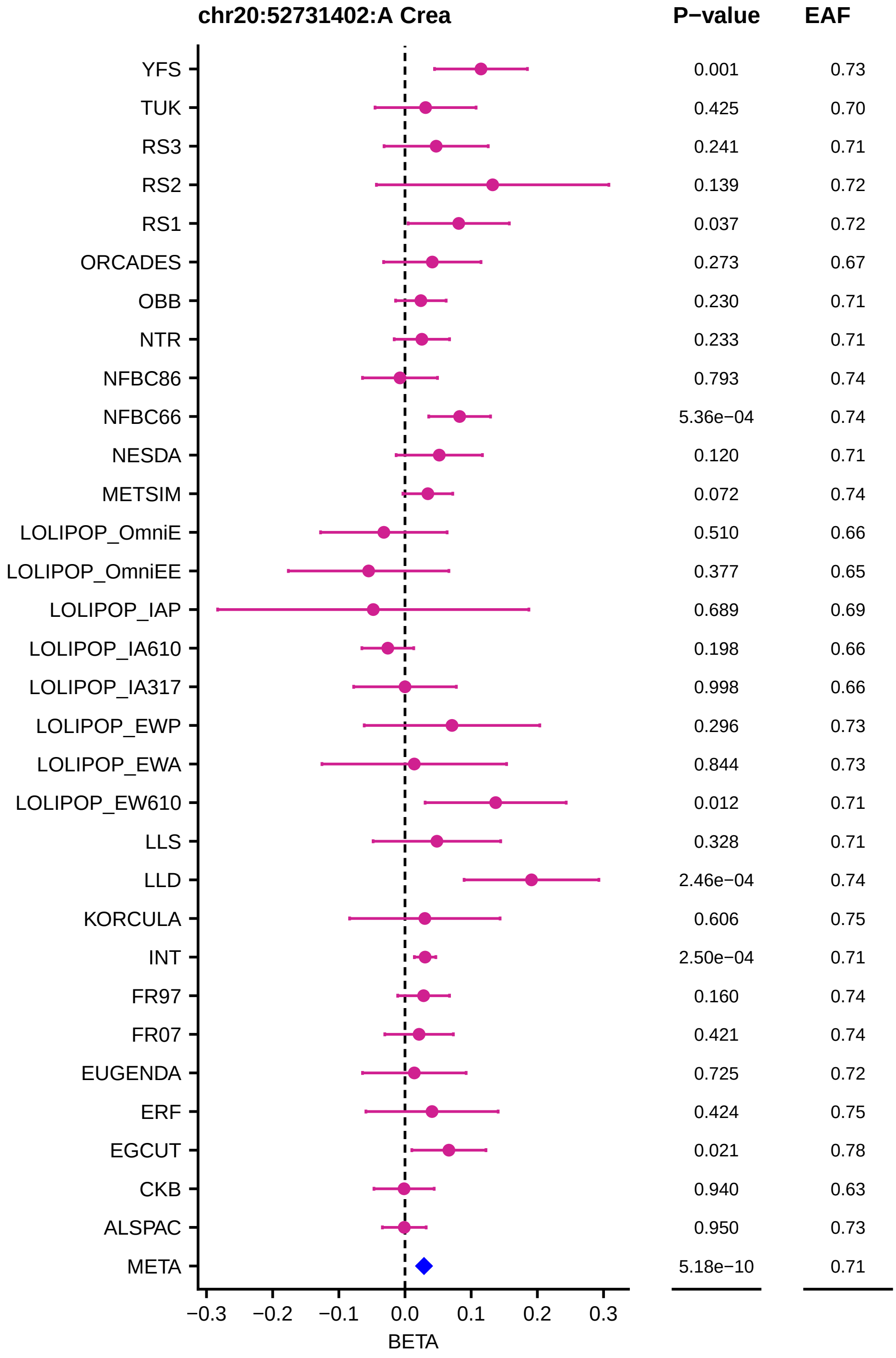
chr20:52731402:A Crea
P−value
EAF
YFS 0.001 0.73
TUK 0.425 0.70
RS3 0.241 0.71
RS2 0.139 0.72
RS1 0.037 0.72
ORCADES 0.273 0.67
OBB 0.230 0.71
NTR 0.233 0.71
NFBC86 0.793 0.74
NFBC66 5.36e−04 0.74
NESDA 0.120 0.71
METSIM 0.072 0.74
LOLIPOP_OmniE 0.510 0.66
LOLIPOP_OmniEE 0.377 0.65
LOLIPOP_IAP 0.689 0.69
LOLIPOP_IA610 0.198 0.66
LOLIPOP_IA317 0.998 0.66
LOLIPOP_EWP 0.296 0.73
LOLIPOP_EWA 0.844 0.73
LOLIPOP_EW610 0.012 0.71
LLS 0.328 0.71
LLD 2.46e−04 0.74
KORCULA 0.606 0.75
INT 2.50e−04 0.71
FR97 0.160 0.74
FR07 0.421 0.74
EUGENDA 0.725 0.72
ERF 0.424 0.75
EGCUT 0.021 0.78
CKB 0.940 0.63
ALSPAC 0.950 0.73
META 5.18e−10 0.71
−0.3 −0.2 −0.1 0.0 0.1 0.2 0.3
BETA

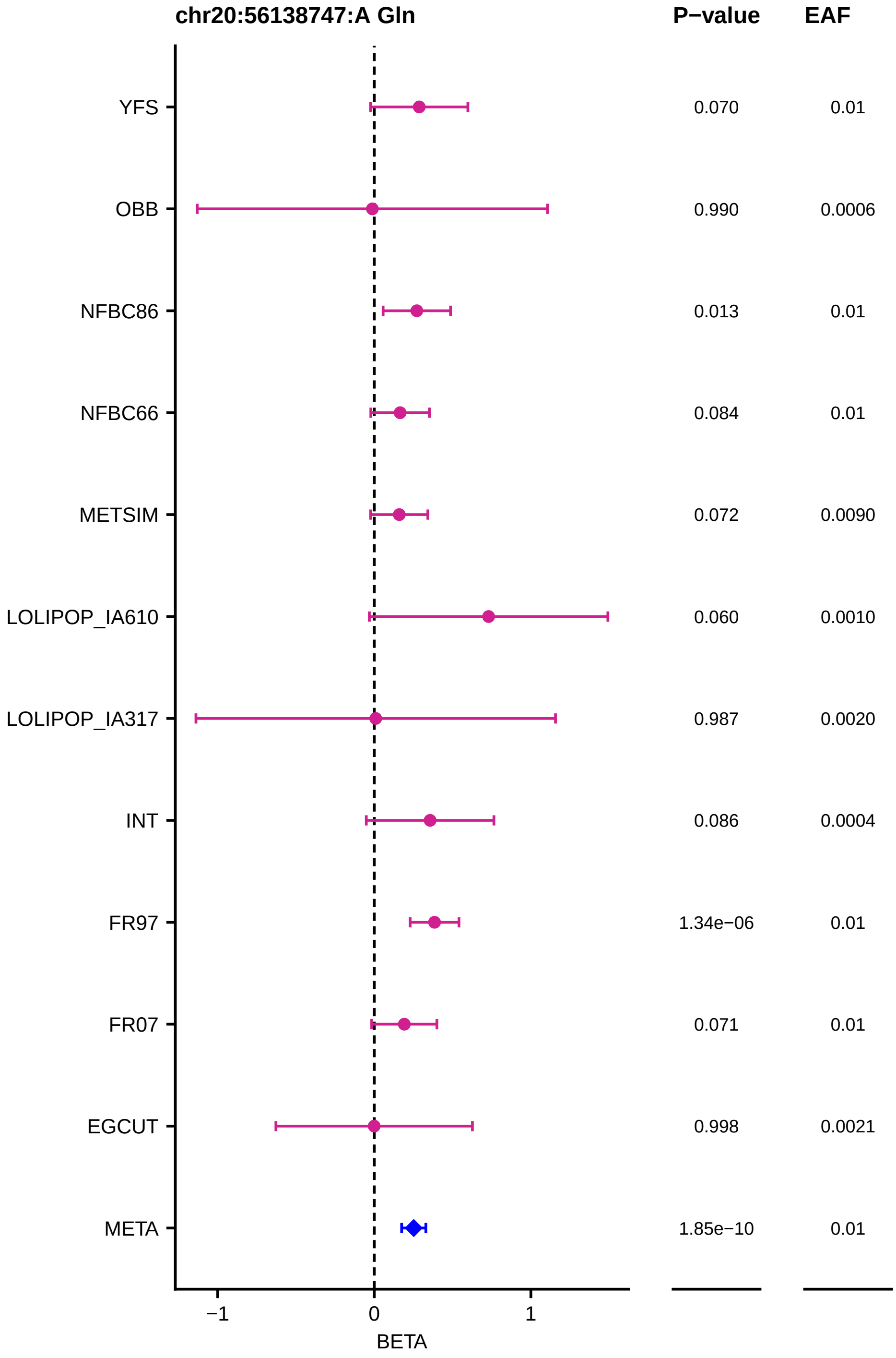
chr20:56138747:A Gln
P−value
EAF
YFS
OBB
NFBC86
NFBC66
METSIM
LOLIPOP_IA610
LOLIPOP_IA317
INT
FR97
FR07
EGCUT
META
0.070
0.990
0.013
0.084
0.072
0.060
0.987
0.086
1.34e−06
0.071
0.998
1.85e−10
0.01
0.0006
0.01
0.01
0.0090
0.0010
0.0020
0.0004
0.01
0.01
0.0021
0.01
−1
0
1
BETA

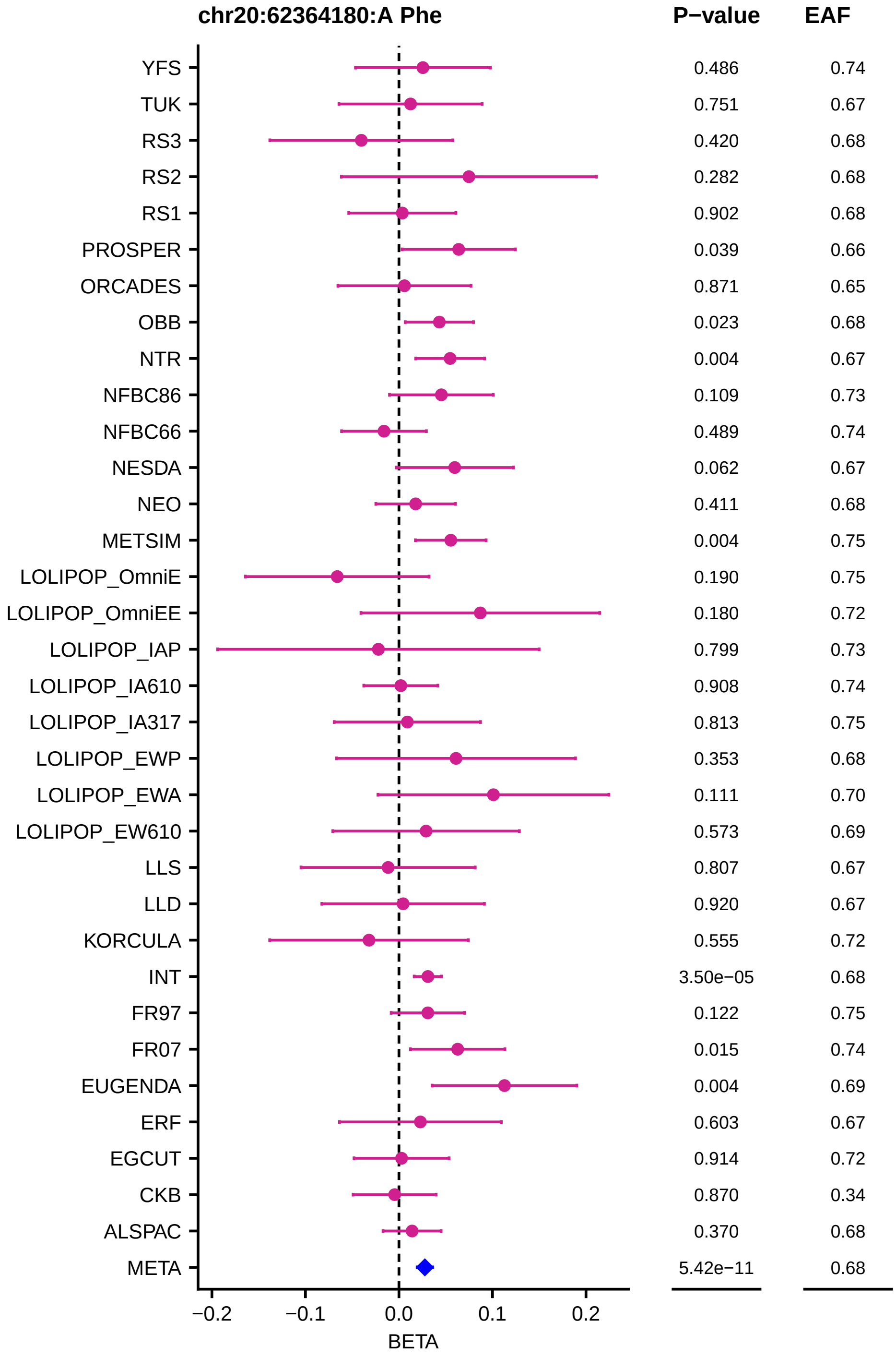
chr20:62364180:A Phe
P−value
EAF
YFS 0.486 0.74
TUK 0.751 0.67
RS3 0.420 0.68
RS2 0.282 0.68
RS1 0.902 0.68
PROSPER 0.039 0.66
ORCADES 0.871 0.65
OBB 0.023 0.68
NTR 0.004 0.67
NFBC86 0.109 0.73
NFBC66 0.489 0.74
NESDA 0.062 0.67
NEO 0.411 0.68
METSIM 0.004 0.75
LOLIPOP_OmniE 0.190 0.75
LOLIPOP_OmniEE 0.180 0.72
LOLIPOP_IAP 0.799 0.73
LOLIPOP_IA610 0.908 0.74
LOLIPOP_IA317 0.813 0.75
LOLIPOP_EWP 0.353 0.68
LOLIPOP_EWA 0.111 0.70
LOLIPOP_EW610 0.573 0.69
LLS 0.807 0.67
LLD 0.920 0.67
KORCULA 0.555 0.72
INT 3.50e−05 0.68
FR97 0.122 0.75
FR07 0.015 0.74
EUGENDA 0.004 0.69
ERF 0.603 0.67
EGCUT 0.914 0.72
CKB 0.870 0.34
ALSPAC 0.370 0.68
META 5.42e−11 0.68
−0.2
−0.1
0.0
0.1
0.2
BETA

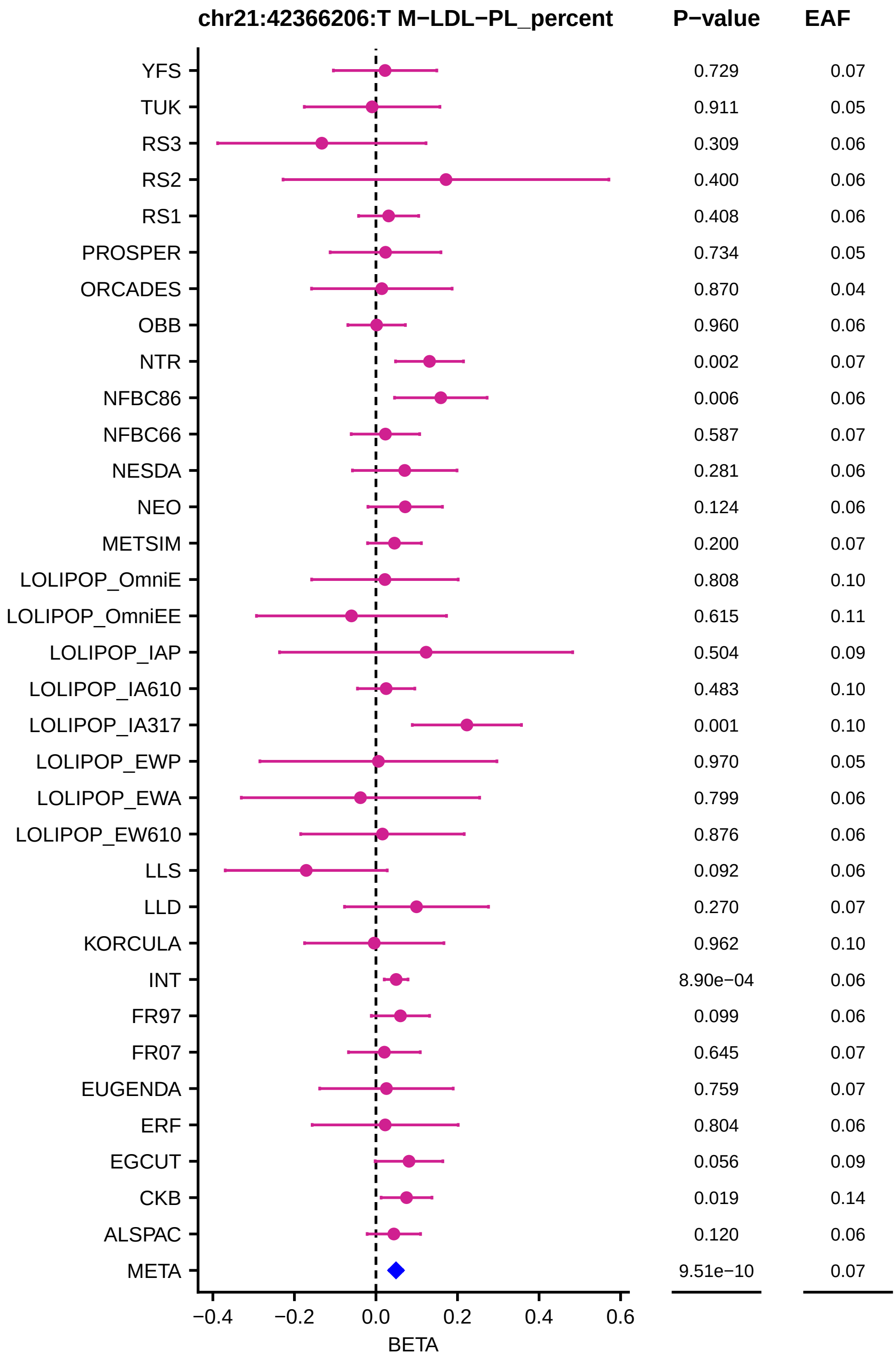

chr21:42366206:T M−LDL−PL_percent
P−value
EAF
YFS
0.729
0.07
TUK
0.911
0.05
RS3
0.309
0.06
RS2
0.400
0.06
RS1
0.408
0.06
PROSPER
0.734
0.05
ORCADES
0.870
0.04
OBB
0.960
0.06
NTR
0.002
0.07
NFBC86
0.006
0.06
NFBC66
0.587
0.07
NESDA
0.281
0.06
NEO
0.124
0.06
METSIM
0.200
0.07
LOLIPOP_OmniE
0.808
0.10
LOLIPOP_OmniEE
0.615
0.11
LOLIPOP_IAP
0.504
0.09
LOLIPOP_IA610
0.483
0.10
LOLIPOP_IA317
0.001
0.10
LOLIPOP_EWP
0.970
0.05
LOLIPOP_EWA
0.799
0.06
LOLIPOP_EW610
0.876
0.06
LLS
0.092
0.06
LLD
0.270
0.07
KORCULA
0.962
0.10
INT
8.90e−04
0.06
FR97
0.099
0.06
FR07
0.645
0.07
EUGENDA
0.759
0.07
ERF
0.804
0.06
EGCUT
0.056
0.09
CKB
0.019
0.14
ALSPAC
0.120
0.06
META
9.51e−10
0.07
−0.4
−0.2
0.0
0.2
0.4
0.6
BETA

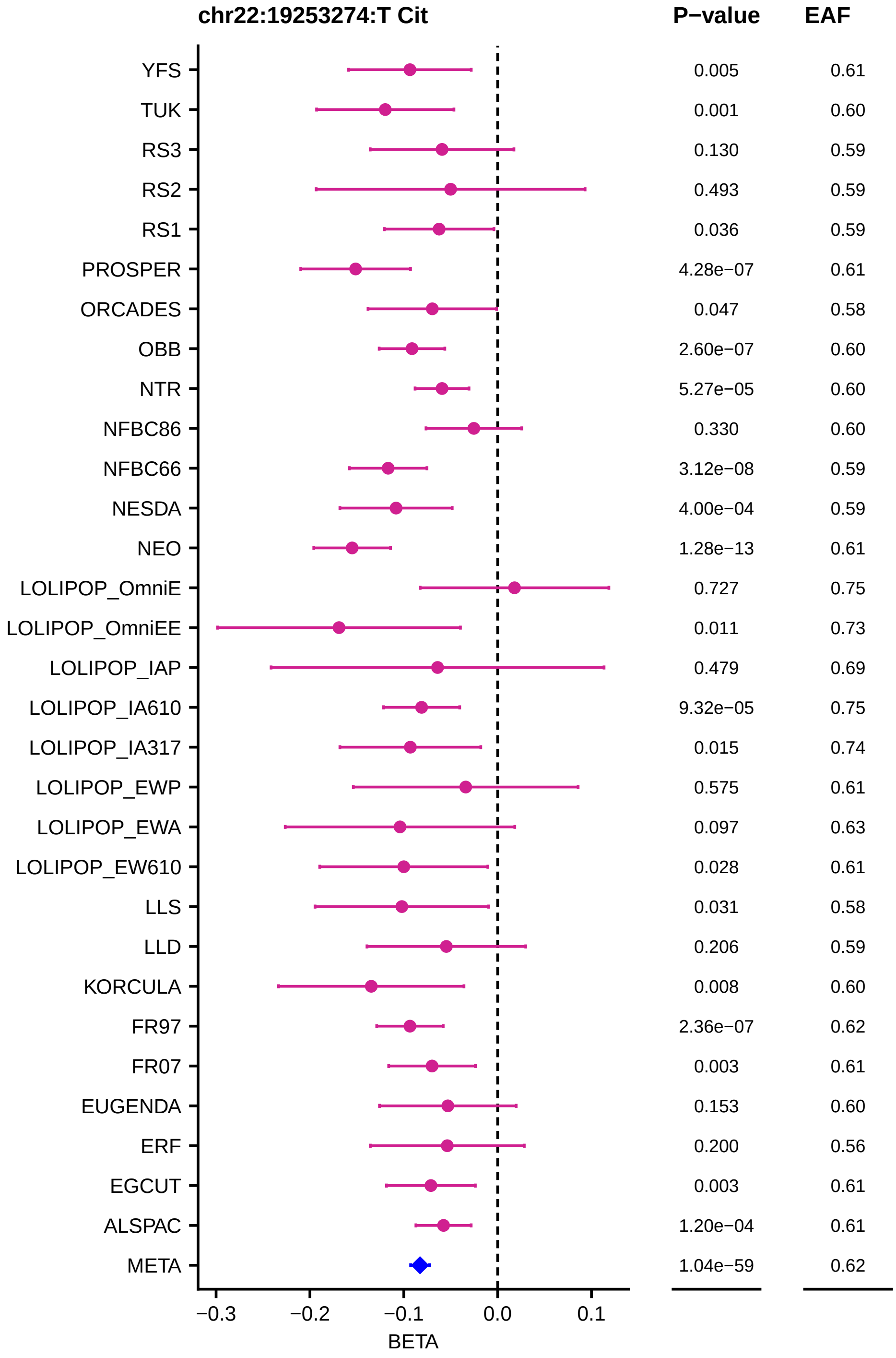
chr22:19253274:T Cit
P−value
EAF
YFS 0.005 0.61
TUK 0.001 0.60
RS3 0.130 0.59
RS2 0.493 0.59
RS1 0.036 0.59
PROSPER 4.28e−07 0.61
ORCADES 0.047 0.58
OBB 2.60e−07 0.60
NTR 5.27e−05 0.60
NFBC86 0.330 0.60
NFBC66 3.12e−08 0.59
NESDA 4.00e−04 0.59
NEO 1.28e−13 0.61
LOLIPOP_OmniE 0.727 0.75
LOLIPOP_OmniEE 0.011 0.73
LOLIPOP_IAP 0.479 0.69
LOLIPOP_IA610 9.32e−05 0.75
LOLIPOP_IA317 0.015 0.74
LOLIPOP_EWP 0.575 0.61
LOLIPOP_EWA 0.097 0.63
LOLIPOP_EW610 0.028 0.61
LLS 0.031 0.58
LLD 0.206 0.59
KORCULA 0.008 0.60
FR97 2.36e−07 0.62
FR07 0.003 0.61
EUGENDA 0.153 0.60
ERF 0.200 0.56
EGCUT 0.003 0.61
ALSPAC 1.20e−04 0.61
META 1.04e−59 0.62
−0.3
−0.2
−0.1
0.0
0.1
BETA

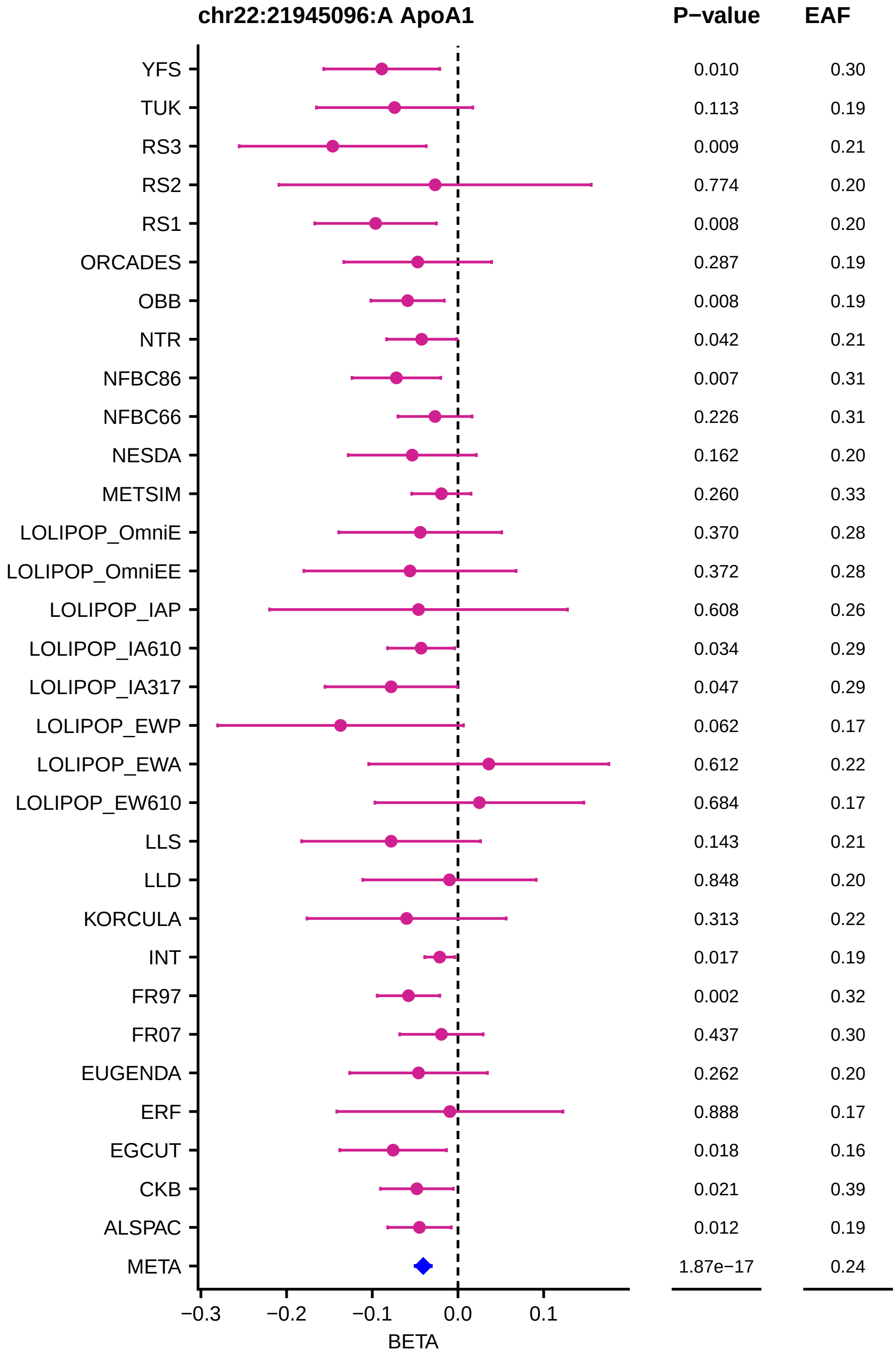
chr22:21945096:A ApoA1
P−value
EAF
YFS 0.010 0.30
TUK 0.113 0.19
RS3 0.009 0.21
RS2 0.774 0.20
RS1 0.008 0.20
ORCADES 0.287 0.19
OBB 0.008 0.19
NTR 0.042 0.21
NFBC86 0.007 0.31
NFBC66 0.226 0.31
NESDA 0.162 0.20
METSIM 0.260 0.33
LOLIPOP_OmniE 0.370 0.28
LOLIPOP_OmniEE 0.372 0.28
LOLIPOP_IAP 0.608 0.26
LOLIPOP_IA610 0.034 0.29
LOLIPOP_IA317 0.047 0.29
LOLIPOP_EWP 0.062 0.17
LOLIPOP_EWA 0.612 0.22
LOLIPOP_EW610 0.684 0.17
LLS 0.143 0.21
LLD 0.848 0.20
KORCULA 0.313 0.22
INT 0.017 0.19
FR97 0.002 0.32
FR07 0.437 0.30
EUGENDA 0.262 0.20
ERF 0.888 0.17
EGCUT 0.018 0.16
CKB 0.021 0.39
ALSPAC 0.012 0.19
META 1.87e−17 0.24
−0.3 −0.2 −0.1 0.0 0.1
BETA

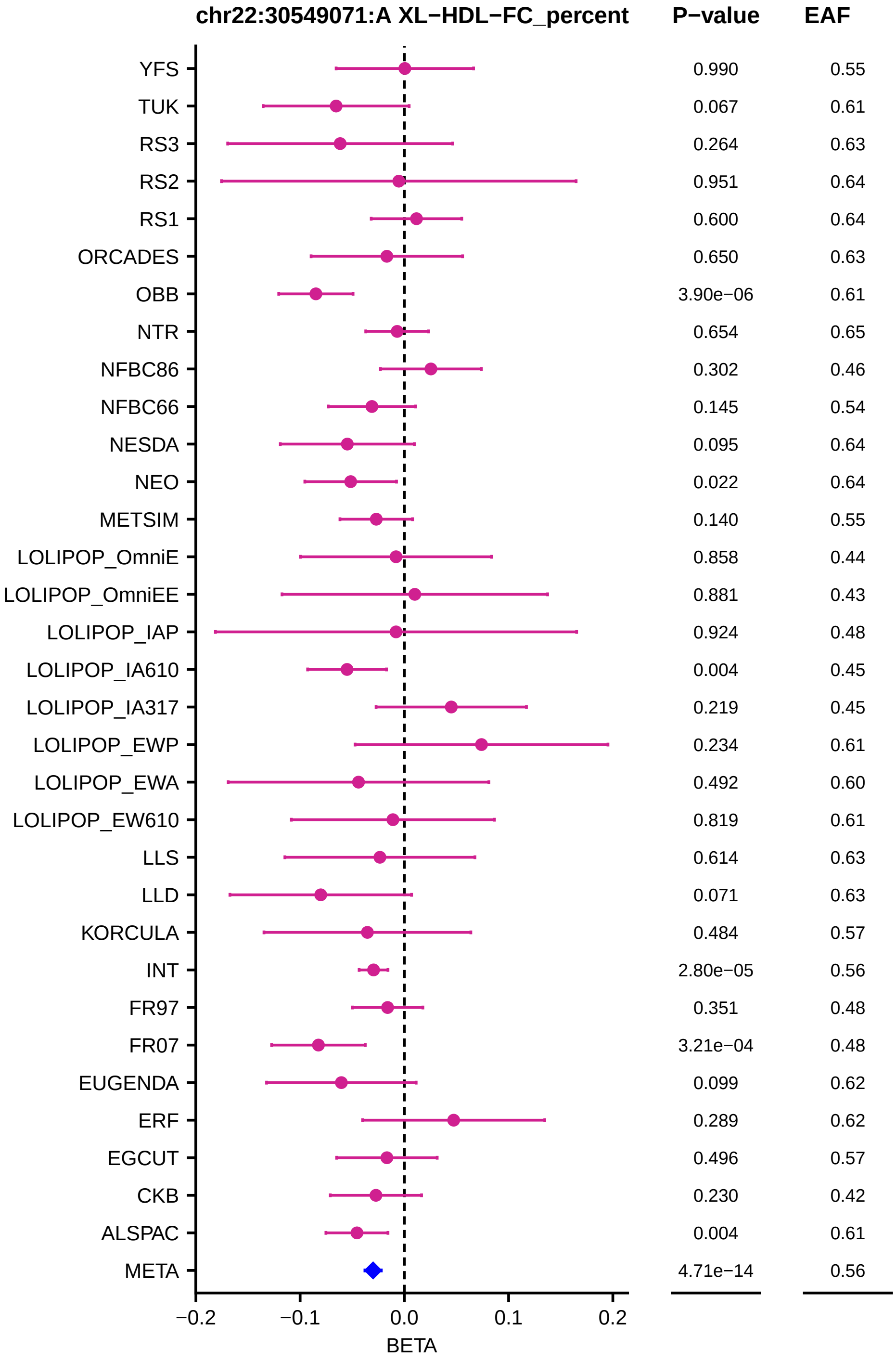
chr22:30549071:A XL−HDL−FC_percent
P−value
EAF
YFS 0.990 0.55
TUK 0.067 0.61
RS3 0.264 0.63
RS2 0.951 0.64
RS1 0.600 0.64
ORCADES 0.650 0.63
OBB 3.90e−06 0.61
NTR 0.654 0.65
NFBC86 0.302 0.46
NFBC66 0.145 0.54
NESDA 0.095 0.64
NEO 0.022 0.64
METSIM 0.140 0.55
LOLIPOP_OmniE 0.858 0.44
LOLIPOP_OmniEE 0.881 0.43
LOLIPOP_IAP 0.924 0.48
LOLIPOP_IA610 0.004 0.45
LOLIPOP_IA317 0.219 0.45
LOLIPOP_EWP 0.234 0.61
LOLIPOP_EWA 0.492 0.60
LOLIPOP_EW610 0.819 0.61
LLS 0.614 0.63
LLD 0.071 0.63
KORCULA 0.484 0.57
INT 2.80e−05 0.56
FR97 0.351 0.48
FR07 3.21e−04 0.48
EUGENDA 0.099 0.62
ERF 0.289 0.62
EGCUT 0.496 0.57
CKB 0.230 0.42
ALSPAC 0.004 0.61
META 4.71e−14 0.56
−0.2
−0.1
0.0
0.1
0.2
BETA

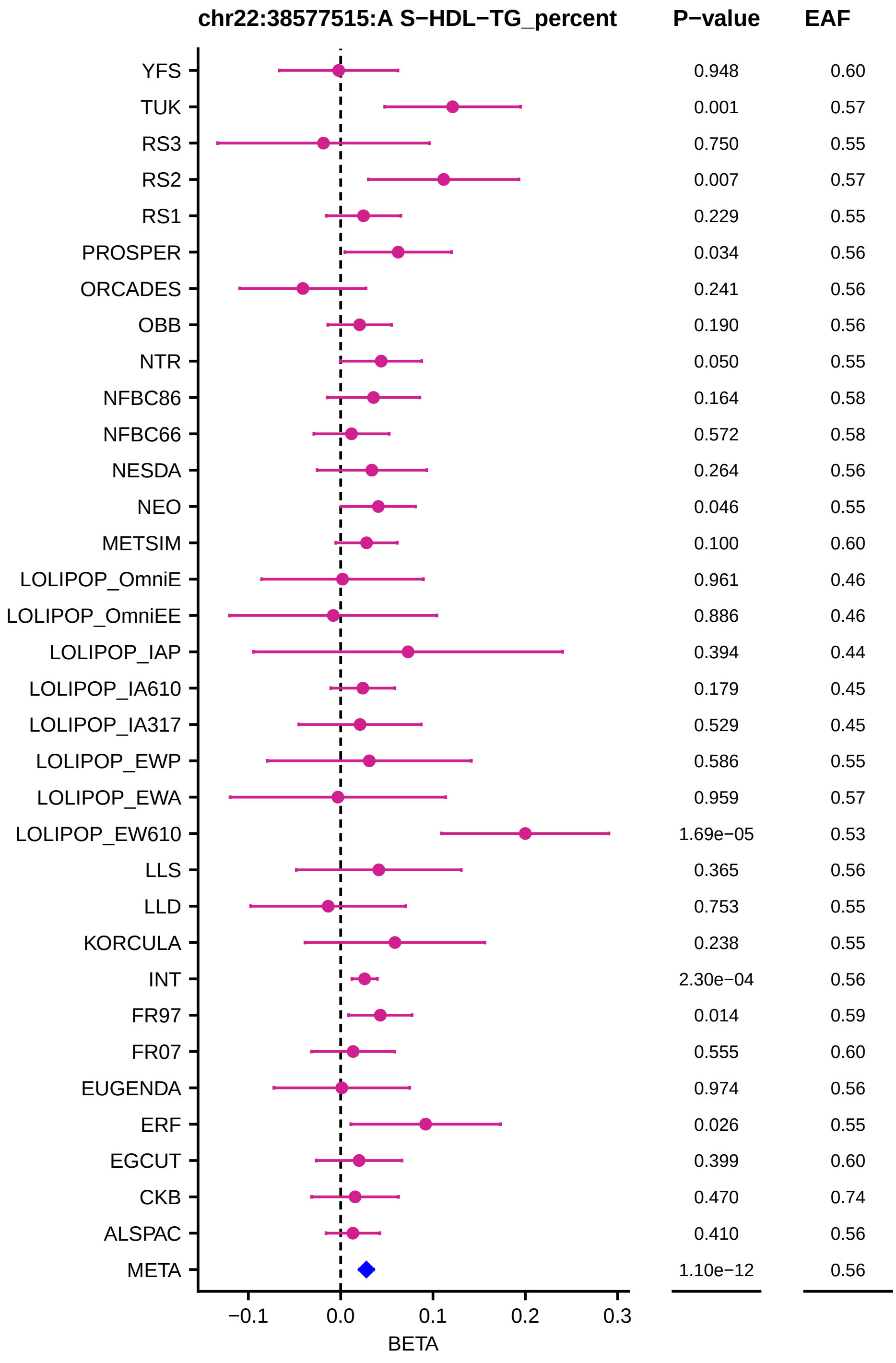
chr22:38577515:A S−HDL−TG_percent
P−value
EAF
YFS 0.948 0.60
TUK 0.001 0.57
RS3 0.750 0.55
RS2 0.007 0.57
RS1 0.229 0.55
PROSPER 0.034 0.56
ORCADES 0.241 0.56
OBB 0.190 0.56
NTR 0.050 0.55
NFBC86 0.164 0.58
NFBC66 0.572 0.58
NESDA 0.264 0.56
NEO 0.046 0.55
METSIM 0.100 0.60
LOLIPOP_OmniE 0.961 0.46
LOLIPOP_OmniEE 0.886 0.46
LOLIPOP_IAP 0.394 0.44
LOLIPOP_IA610 0.179 0.45
LOLIPOP_IA317 0.529 0.45
LOLIPOP_EWP 0.586 0.55
LOLIPOP_EWA 0.959 0.57
LOLIPOP_EW610 1.69e−05 0.53
LLS 0.365 0.56
LLD 0.753 0.55
KORCULA 0.238 0.55
INT 2.30e−04 0.56
FR97 0.014 0.59
FR07 0.555 0.60
EUGENDA 0.974 0.56
ERF 0.026 0.55
EGCUT 0.399 0.60
CKB 0.470 0.74
ALSPAC 0.410 0.56
META 1.10e−12 0.56
−0.1
0.0
0.1
0.2
0.3
BETA

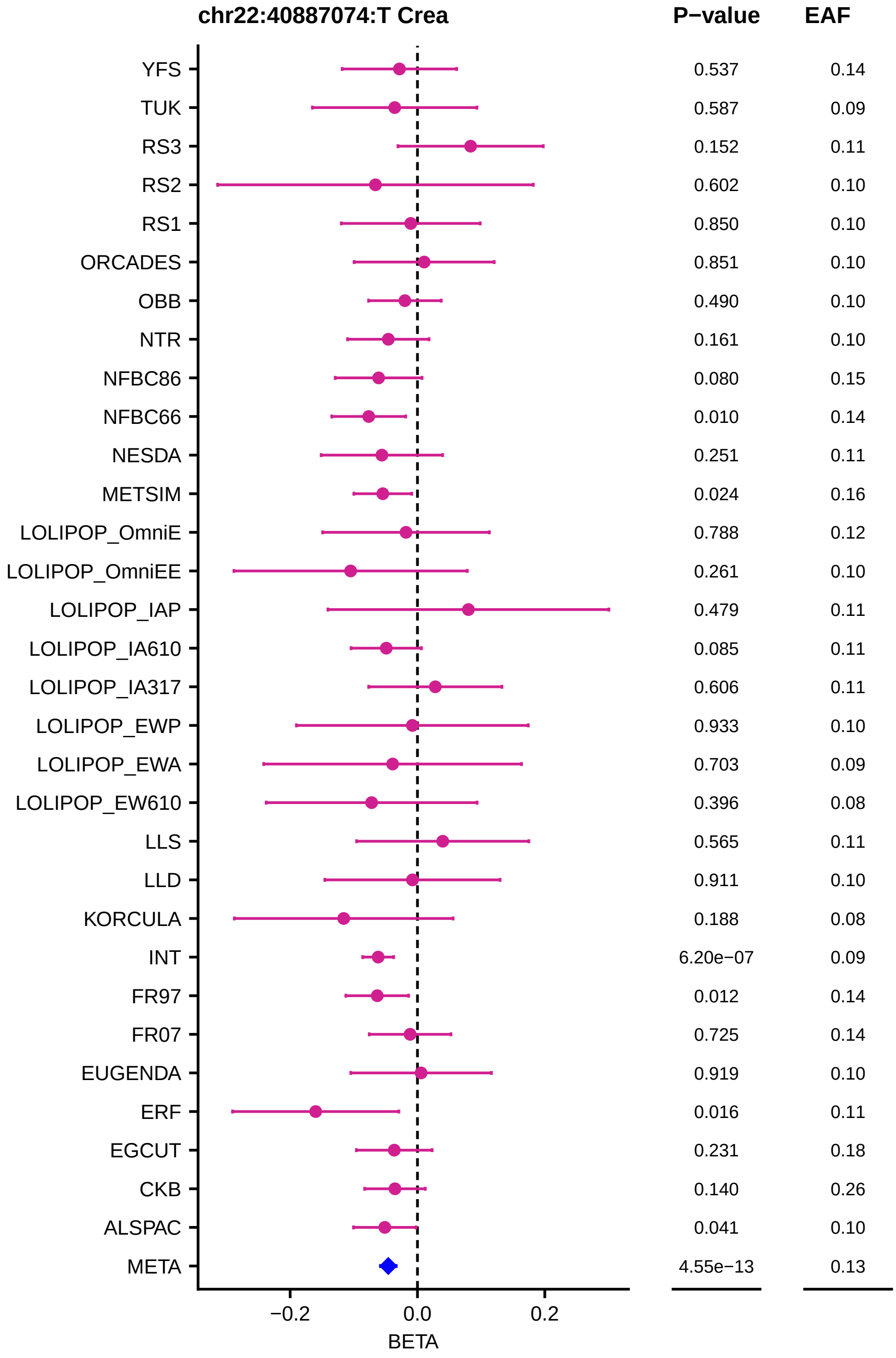
chr22:40887074:T Crea
P−value
EAF
YFS 0.537 0.14
TUK 0.587 0.09
RS3 0.152 0.11
RS2 0.602 0.10
RS1 0.850 0.10
ORCADES 0.851 0.10
OBB 0.490 0.10
NTR 0.161 0.10
NFBC86 0.080 0.15
NFBC66 0.010 0.14
NESDA 0.251 0.11
METSIM 0.024 0.16
LOLIPOP_OmniE 0.788 0.12
LOLIPOP_OmniEE 0.261 0.10
LOLIPOP_IAP 0.479 0.11
LOLIPOP_IA610 0.085 0.11
LOLIPOP_IA317 0.606 0.11
LOLIPOP_EWP 0.933 0.10
LOLIPOP_EWA 0.703 0.09
LOLIPOP_EW610 0.396 0.08
LLS 0.565 0.11
LLD 0.911 0.10
KORCULA 0.188 0.08
INT 6.20e−07 0.09
FR97 0.012 0.14
FR07 0.725 0.14
EUGENDA 0.919 0.10
ERF 0.016 0.11
EGCUT 0.231 0.18
CKB 0.140 0.26
ALSPAC 0.041 0.10
META 4.55e−13 0.13
−0.2
0.0
0.2
BETA

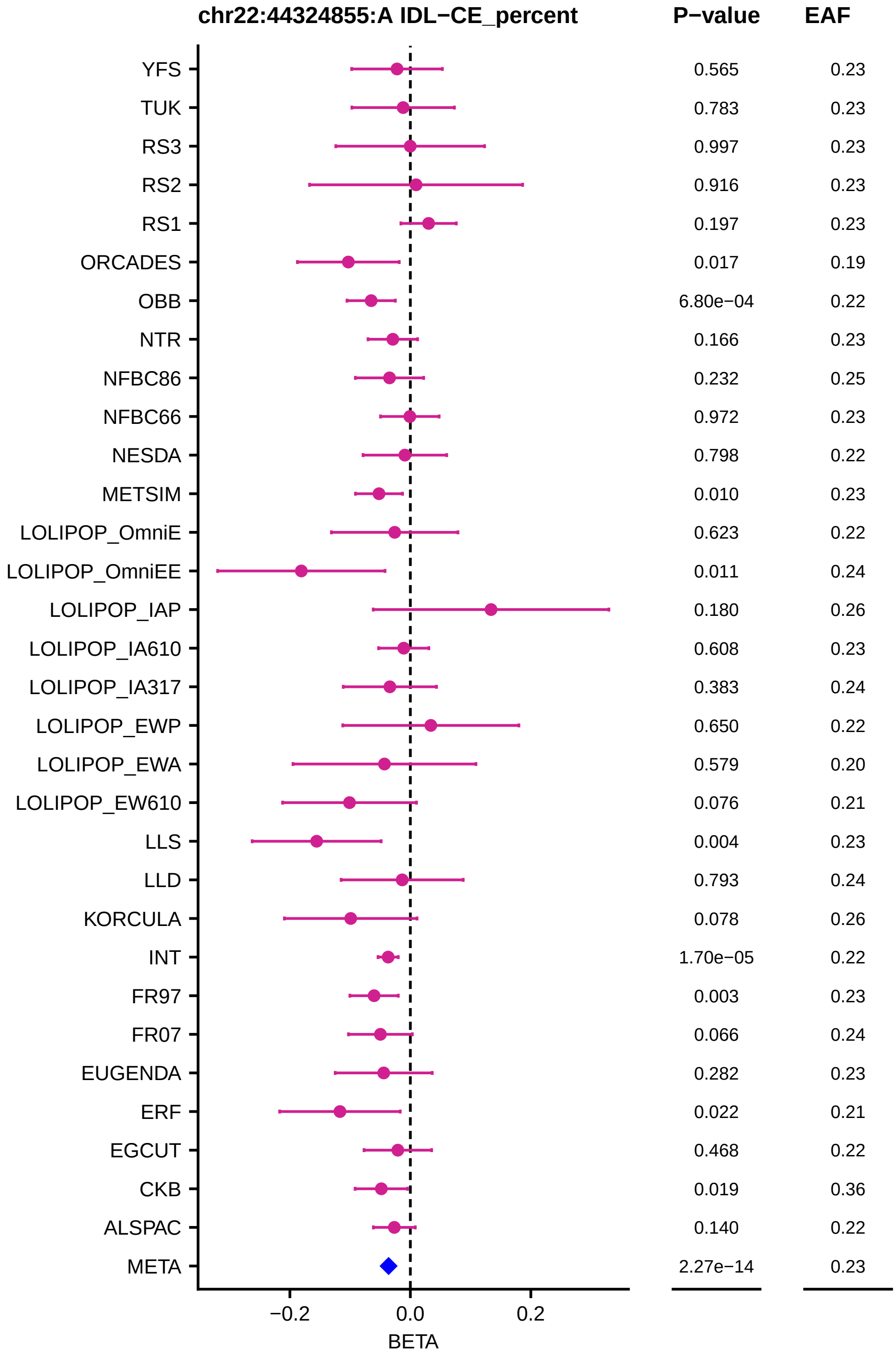
chr22:44324855:A IDL−CE_percent
P−value
EAF
YFS 0.565 0.23
TUK 0.783 0.23
RS3 0.997 0.23
RS2 0.916 0.23
RS1 0.197 0.23
ORCADES 0.017 0.19
OBB 6.80e−04 0.22
NTR 0.166 0.23
NFBC86 0.232 0.25
NFBC66 0.972 0.23
NESDA 0.798 0.22
METSIM 0.010 0.23
LOLIPOP_OmniE 0.623 0.22
LOLIPOP_OmniEE 0.011 0.24
LOLIPOP_IAP 0.180 0.26
LOLIPOP_IA610 0.608 0.23
LOLIPOP_IA317 0.383 0.24
LOLIPOP_EWP 0.650 0.22
LOLIPOP_EWA 0.579 0.20
LOLIPOP_EW610 0.076 0.21
LLS 0.004 0.23
LLD 0.793 0.24
KORCULA 0.078 0.26
INT 1.70e−05 0.22
FR97 0.003 0.23
FR07 0.066 0.24
EUGENDA 0.282 0.23
ERF 0.022 0.21
EGCUT 0.468 0.22
CKB 0.019 0.36
ALSPAC 0.140 0.22
META 2.27e−14 0.23
−0.2
0.0
0.2
BETA
