## Supplementary Notes for "Genome-wide characterization of circulating metabolic biomarkers reveals substantial pleiotropy and novel disease pathways"

### Supplementary notes for the manuscript entitled *Genome-wide characterization of circulating metabolic biomarkers reveals substantial pleiotropy and novel disease pathways*

#### Study descriptions, acknowledgements, and funding

**Datasets:**

We used genotyping and lipoprotein profiling data from 33 studies/subcohorts, totaling up to 136,016 individuals. Descriptions of the studies are provided below, and specific details are shown in Supplementary Table S1. Details of genotyping and GWAS are shown in Supplementary Table S3. All studies were approved by ethical committees of relevant institutions. In addition, we used summary-level data from the FinnGen study[1] (description below).

**Study descriptions, acknowledgements, and funding:**

***Avon Longitudinal Study of Parents and Children (ALSPAC)***

*Study description*: Pregnant women resident in Avon, UK with expected dates of delivery 1st April 1991 to 31st December 1992 were invited to take part in the ALSPAC study[2,3]. The initial number of pregnancies enrolled is 14,541 (for these at least one questionnaire has been returned or a "Children in Focus" clinic had been attended by 19/07/99). Of these initial pregnancies, there was a total of 14,676 foetuses, resulting in 14,062 live births and 13,988 children who were alive at 1 year of age. When the oldest children were approximately 7 years of age, an attempt was made to bolster the initial sample with eligible cases who had failed to join the study originally. As a result, when considering variables collected from the age of seven onwards (and potentially abstracted from obstetric notes) there are data available for more than the 14,541 pregnancies mentioned above. The number of new pregnancies not in the initial sample (known as Phase I enrolment) that are currently represented on the built files and reflecting enrolment status at the age of 24 is 913 (456, 262 and 195 recruited during Phases II, III and IV respectively), resulting in an additional 913 children being enrolled. The phases of enrolment are described in more detail in the cohort profile paper and its update (see footnote 4 below). The total sample size for analyses using any data collected after the age of seven is therefore 15,454 pregnancies, resulting in 15,589 foetuses. Of these 14,901 were alive at 1 year of age. A 10% sample of the ALSPAC cohort, known as the Children in Focus (CiF) group, attended clinics at the University of Bristol at various time intervals between 4 to 61 months of age. The CiF group were chosen at random from the last 6 months of ALSPAC births (1432 families attended at least one clinic). Excluded were those mothers who had moved out of the area or were lost to follow-up, and those partaking in another study of infant development in Avon. Please note that the study website contains details of all the data that is available through a fully searchable data dictionary and variable search tool (http://www.bristol.ac.uk/alspac/researchers/our-data/). Ethical approval for the study was obtained from the ALSPAC Law and Ethics committee and local research ethics committees. Consent for biological samples has been collected in accordance with the Human Tissue Act (2004). Informed consent for the use of data collected via questionnaires and clinics was obtained from participants following the recommendations of the ALSPAC Ethics and Law Committee at the time.

*Acknowledgements/funding*: We are extremely grateful to all the families who took part in this study, the midwives for their help in recruiting them, and the whole ALSPAC team, which includes interviewers, computer and laboratory technicians, clerical workers, research scientists, volunteers, managers, receptionists and nurses. The UK Medical Research Council and Wellcome (Grant ref: 217065/Z/19/Z) and the University of Bristol provide core support for ALSPAC. This publication is the work of the authors and Johannes Kettunen will serve as guarantor for the contents of this paper. A comprehensive list of grants funding is available on the ALSPAC website (http://www.bristol.ac.uk/alspac/external/documents/grant-acknowledgements.pdf). GWAS data was generated by Sample Logistics and Genotyping Facilities at Wellcome Sanger Institute and LabCorp (Laboratory Corporation of America) using support from 23andMe. D.A.H. is funded by N.J.T.'s Wellcome Trust Investigator (202802/Z/16/Z) & University of Bristol and UK Medical Research Council (MC_UU_00011/1 and MC_UU_00011/6). N.J.T. is a Wellcome Trust Investigator (202802/Z/16/Z), is the PI of the Avon Longitudinal Study of Parents and Children (MRC & WT 217065/Z/19/Z), is supported by the University of Bristol NIHR Biomedical Research Centre (BRC-1215-2001), the MRC Integrative Epidemiology Unit (MC_UU_00011/1) and works within the CRUK Integrative Cancer Epidemiology Programme (C18281/A29019).

***China Kadoorie Biobank (CKB)***

*Study description*: China Kadoorie Biobank (CKB)[4] is a large prospective study in which more than 512,000 adults were recruited from ten geographically defined and diverse areas of China between 2004 and 2008. Extensive data (questionnaires, physical measurements and blood samples) was collected at baseline and subsequent periodic resurveys, with follow-up (median 11 years) through linkage to death and disease registries and to in-patient records through the national health insurance system. Genotyping of >100,000 participants used 2 custom-designed Affymetrix Axiom arrays, with imputation to the 1,000 Genomes Phase 3 reference (EAS MAF > 0)[5]. A subset of 4,435 genotyped participants, comprising 3,198 incident cases of cardiovascular disease and 1,237 controls, had Nightingale NMR-metabolomics measurements for 225 parameters.

*Acknowledgements/funding:* The most important acknowledgement is to the participants in the study and the members of the survey teams in each of the 10 regional centres, and to the project development and management teams based at Beijing, Oxford and the 10 regional centres. China's National Health Insurance provides electronic linkage to all hospital treatments. Members of The China Kadoorie Biobank (CKB) Collaborative Group (banner authors) are also acknowledged: International Steering Committee: Junshi Chen, Zhengming Chen (PI), Robert Clarke, Rory Collins, Yu Guo, Liming Li (PI), Chen Wang, Jun Lv, Richard Peto, Robin Walters.; International Co-ordinating Centre, Oxford: Daniel Avery, Derrick Bennett, Ruth Boxall, Ka Hung Chan, Yumei Chang, Yiping Chen, Zhengming Chen, Johnathan Clarke; Robert Clarke, Huaidong Du, Zammy Fairhurst-Hunter, Hannah Fry, Simon Gilbert, Alex Hacker, Mike Hill, Michael Holmes, Pek Kei Im, Andri Iona, Maria Kakkoura, Christiana Kartsonaki, Rene Kerosi, Kuang Lin, Mohsen Mazidi, Iona Millwood, Qunhua Nie, Alfred Pozarickij, Paul Ryder, Saredo Said, Sam Sansome, Dan Schmidt, Paul Sherliker, Rajani Sohoni, Becky Stevens, Iain Turnbull, Robin Walters, Lin Wang, Neil Wright, Ling Yang, Xiaoming Yang, Pang Yao. ; National Co-ordinating Centre, Beijing: Yu Guo, Xiao Han, Can Hou, Chun Li, Chao Liu, Jun Lv, Pei Pei, Canqing Yu.; Regional Co-ordinating Centres: Gansu: Gansu Provincial CDC – Caixia Dong, Pengfei Ge, Xiaolan Ren. Maiji CDC – Zhongxiao Li, Enke Mao, Tao Wang, Hui Zhang, Xi Zhang. Haikou: Hainan Provincial CDC – Jinyan Chen, Ximin Hu, Xiaohuan Wang. Meilan CDC – Zhendong Guo, Huimei Li, Yilei Li, Min Weng, Shukuan Wu. Harbin: Heilongjiang Provincial CDC – Shichun Yan, Mingyuan Zou, Xue Zhou. Nangang CDC – Ziyan Guo, Quan Kang, Yanjie Li, Bo Yu, Qinai Xu. Henan: Henan Provincial CDC – Liang Chang, Lei Fan, Shixian Feng, Ding Zhang, Gang Zhou. Huixian CDC – Yulian Gao, Tianyou He, Pan He, Chen Hu, Huarong Sun, Xukui Zhang. Hunan: Hunan Provincial CDC – Biyun Chen, Zhongxi Fu, Yuelong Huang, Huilin Liu, Qiaohua Xu, Li Yin. Liuyang CDC – Huajun Long, Xin Xu, Hao Zhang, Libo Zhang. Liuzhou:

Guangxi Provincial CDC – Naying Chen, Duo Liu, Zhenzhu Tang. Liuzhou CDC – Ningyu Chen, Qilian Jiang, Jian Lan, Mingqiang Li, Yun Liu, Fanwen Meng, Jinhuai Meng, Rong Pan, Yulu Qin, Ping Wang, Sisi Wang, Liuping Wei, Liyuan Zhou. Qingdao: Qingdao CDC – Liang Cheng, Ranran Du, Ruqin Gao, Feifei Li, Shanpeng Li, Yongmei Liu, Feng Ning, Zengchang Pang, Xiaohui Sun, Xiaocao Tian, Shaojie Wang, Yaoming Zhai, Hua Zhang, Licang CDC – Wei Hou, Silu Lv, Junzheng Wang. Sichuan: Sichuan Provincial CDC – Xiaofang Chen, Xianping Wu, Ningmei Zhang, Weiwei Zhou. Pengzhou CDC – Xiaofang Chen, Jianguo Li, Jiaqiu Liu, Guojin Luo, Qiang Sun, Xunfu Zhong. Suzhou: Jiangsu Provincial CDC – Jian Su, Ran Tao, Ming Wu, Jie Yang, Jinyi Zhou, Yonglin Zhou. Suzhou CDC – Yihe Hu, Yujie Hua, Jianrong Jin Fang Liu, Jingchao Liu, Yan Lu, Liangcai Ma, Aiyu Tang, Jun Zhang. Zhejiang: Zhejiang Provincial CDC – Weiwei Gong, Ruying Hu, Hao Wang, Meng Wang, Min Yu. Tongxiang CDC – Lingli Chen, Qijun Gu, Dongxia Pan, Chunmei Wang, Kaixu Xie, Xiaoyi Zhang. DNA extraction and genotyping was funded by GlaxoSmithKline, and the UK Medical Research Council (MC-PC-13049, MC-PC-14135). The CKB baseline survey and the first re-survey were supported by the Kadoorie Charitable Foundation in Hong Kong. Long-term follow-up was supported by the Wellcome Trust (212946/Z/18/Z, 202922/Z/16/Z, 104085/Z/14/Z, 088158/Z/09/Z), the National Key Research and Development Program of China (2016YFC0900500, 2016YFC0900501, 2016YFC0900504, 2016YFC1303904), and the National Natural Science Foundation of China (91843302). The project is supported by core funding from the UK Medical Research Council (MC_UU_00017/1,MC_UU_12026/2, MC_U137686851), Cancer Research UK (C16077/A29186; C500/A16896), and the British Heart Foundation (CH/1996001/9454) to the Clinical Trial Service Unit and Epidemiological Studies Unit and to the MRC Population Health Research Unit at Oxford University.

***CROATIA_Korcula***

*Study description*: The Croatian Biobank (The 10001 Dalmatians) is an isolated population biobank, with a focus on genetic basis of complex diseases[6]. The current study included a cohort of adult participants from the island of Korcula. Participants were invited by mail, posters, radio, and personal contacts in 2007. A large number of traits were measured, and biological samples were collected as described before[7].

*Acknowledgements/funding*: We would like to acknowledge the staff of several institutions in Croatia that supported the field work, including but not limited, to the University of Split and Zagreb Medical Schools, Institute for Anthropological Research in Zagreb, and the Croatian Institute for Public Health. We also thank all of the participants from the island of Korcula. CROATIA-Korcula was funded by grants from the Medical Research Council (UK), from the Republic of Croatia Ministry of Science, Education and Sports (108-1080315-0302; 216-1080315-0302) and the Croatian Science Foundation (8875); and the CROATIA-Korčula genotyping was funded by the European Union framework program 6 project EUROSPAN (LSHGCT2006018947). Genetic analyses were supported by the MRC HGU "QTL in Health and Disease" (MRC University Unit Programme Grant MC_UU_00007/10). CH was supported by the The Croatian Biobank (10001 Dalmatians) and an MRC Human Genetics Unit program grant 'Quantitative traits in health and disease' (U.MC_UU_00007/10).

***Estonian Genome Center of the University of Tartu (EGCUT)***

*Study description:* The Estonian Genome Center of University of Tartu (EGCUT)[8,9] cohort is a sample of the Estonian resident adult population. All participants were recruited randomly. Questionnaire data, various measurements, and biological samples were collected in 2002-2013.

*Acknowledgments and funding*: Estonian Biobank research was supported by the European Union through the European Regional Development Fund (Project No. 2014-2020.4.01.15-0012), T.E. was supported by the Estonian Research Council grant PUT (PRG1291). We acknowledge the High Performance Computing Centre of the University of Tartu. We also acknowledge the Estonian Biobank Research Team contributors (banner authors) Andres Metspalu, Lili Milani, Reedik Mägi and Mari Nelis.

***The Erasmus Rucphen Family (ERF)***

*Study description*: The Erasmus Rucphen Family (ERF) study is a family-based study that includes inhabitants of a genetically isolated community in the South-West of the Netherlands, ascertained as part of the Genetic Research in Isolated Population program. The ERF cohort includes approximately 3,000 living descendants of 22 founder couples who had at least 6 children baptized in the community church. Individuals who were 18 years or older were invited to participate in the study. Baseline data were collected between 2002 and 2005 and follow-up data between 2015 and 2018. Metabolic biomarkers were successfully quantified in fasting EDTA plasma samples of the selected individuals. The ERF study was approved by the medical ethics committee of the Erasmus Medical Center, Rotterdam, the Netherlands. All participants provided written informed consents and all investigations were carried out in accordance with the Declaration of Helsinki.

*Acknowledgements/funding*: The ERF study was supported by the Consortium for Systems Biology (NCSB), both within the framework of the Netherlands Genomics Initiative (NGI)/Netherlands Organisation for Scientific Research (NWO). ERF study as a part of EUROSPAN (European Special Populations Research Network) was supported by European Commission FP6 STRP grant number 018947 (LSHG-CT-2006-01947) and also received funding from the European Community's Seventh Framework Programme (FP7/2007-

2013)/grant agreement HEALTH-F4-2007-201413 by the European Commission under the programme "Quality of Life and Management of the Living Resources" of 5th Framework Programme (no. QLG2-CT-2002-01254) as well as FP7 project EUROHEADPAIN (nr 602633). High-throughput analysis of the ERF data was supported by joint grant from Netherlands Organisation for Scientific Research and the Russian Foundation for Basic Research (NWO-RFBR 047.017.043). Exome sequencing analysis in ERF was supported by the ZonMw grant (2010/31471/ZONMW). The funders had no role in study design, data collection and analysis, decision to publish, or preparation of the manuscripts. The authors are grateful to all ERF study participants and their relatives, general practitioners and other clinicians for their contributions and to P. Veraart for her help in genealogy, J. Vergeer for the supervision of the laboratory work and assistance with the DNA sample preparation, both S.J. van der Lee and A. van der Spek for collection of the follow-up data and P. Snijders for help with data collection and sample information of both baseline and follow-up data.

***European Genetic Database (EUGENDA)***

*Study description*: The EUGENDA (European Genetic Database) is a case-control study focusing on genetic and nongenetic factors in age-related macular degeneration (AMD)[10]. Subjects were recruited from the clinic in Nijmegen (Netherlands) and Cologne (Germany). Nutrition and lifestyle variables were assessed by questionnaire. The study was approved by the ethics committees in both Cologne and Nijmegen.

*Acknowledgements/funding:* The metabolomics dataset of the EUGENDA cohort was supported by the Dutch Research Council (VICI grant no.: 016.VICL170.024 [A.I.d.H.]); and the European Union's Horizon 2020 research and innovation programme (grant no.: 634479 [A.I.d.H.]).

***FINRISK***

*Study description:* The FINRISK study[11] is a series of population-based cardiovascular risk factor surveys carried out every five years in five (or six in 2002) districts of Finland, including North Karelia, Northern Savo (former Kuopio), Southwestern Finland, Oulu Province, Lapland province (in 2002 only), and the region of Helsinki and Vantaa. A stratified random sample was drawn for each survey from the national population register; the age-range was 25-74 years. All individuals enrolled in the study received a physical examination, a self-administered questionnaire, and a blood sample was drawn. During follow-up, the National Hospital Discharge Register, the National Causes of Death Register and the National Drug Reimbursement Register were used to identify endpoints. In these analyses, the follow-up extends until 31st December 2010. The present study included eligible individuals from the 1997 ($n$=6,643) and 2007 ($n$=3,896) surveys. The Coordinating Ethics Committee of the Helsinki and Uusimaa Hospital District approved the study, which followed the declaration of Helsinki. All subjects gave their informed consent.

*Acknowledgements/funding*: FINRISK cohorts have been mainly funded from budgetary funds of THL. Important additional funding has been obtained from the Finnish Academy and several domestic foundations. V.S. was supported by the Finnish Foundation for Cardiovascular Research and Juho Vainio Foundation.

***INTERVAL***

*Study description*: The INTERVAL Bioresource involves nearly 50,000 blood donors recruited from 25 centres across England between 2012 and 2014. Donors were aged 18-80 with roughly equal proportions of men and women. Blood samples were collected at routine blood donation appointments with serum, plasma, whole blood and buffy coat stored for future analysis. Informed consent was provided at the appointment and phenotypic information was collected later by web questionnaire and via linkage to electronic health records. NMR assays were performed in all participants with an available baseline plasma sample.

*Acknowledgements/funding:* Participants in the INTERVAL randomised controlled trial were recruited with the active collaboration of NHS Blood and Transplant England (www.nhsbt.nhs.uk), which has supported field work and other elements of the trial. DNA extraction and genotyping were co-funded by the National Institute for Health and Care Research (NIHR), the NIHR BioResource (http://bioresource.nihr.ac.uk) and the NIHR Cambridge Biomedical Research Centre (BRC-1215-20014) [*]. The academic coordinating centre for INTERVAL was supported by core funding from the: NIHR Blood and Transplant Research Unit in Donor Health and Genomics (NIHR BTRU-2014-10024), NIHR BTRU in Donor Health and Behaviour (NIHR203337), UK Medical Research Council (MR/L003120/1), British Heart Foundation (SP/09/002; RG/13/13/30194; RG/18/13/33946) and NIHR Cambridge BRC (BRC-1215-20014) [*]. A complete list of the investigators and contributors to the INTERVAL trial is provided in reference[12]. The academic coordinating centre would like to thank blood donor centre staff and blood donors for participating in the INTERVAL trial. NMR assays in INTERVAL were partially funded by a European Commission Framework Programme 7 award (HEALTH-F2-2012-279233). This work was supported by Health Data Research UK, which is funded by the UK Medical Research Council, Engineering and Physical Sciences Research Council, Economic and Social Research Council, Department of Health and Social Care (England), Chief Scientist Office of the Scottish Government Health and Social Care Directorates, Health and Social Care Research and Development Division (Welsh Government), Public Health Agency (Northern Ireland), British Heart Foundation and Wellcome. E.A. is funded by the BHF Programme Grant RG/18/13/33946. P.S. is supported by a Rutherford Fund Fellowship from the Medical Research Council grant (MR/S003746/1). J.D. holds a British Heart Foundation Professorship and a NIHR Senior Investigator Award

[*]. *The views expressed are those of the author(s) and not necessarily those of the NIHR or the Department of Health and Social Care.

***Leiden Longevity Study (LLS)***

*Study description:* The Leiden Longevity Study consists of 1671 members of long-lived families (mean age 60 years) and their 744 partners (mean age 60 years) as population controls[13,14]. For the current study we used a subset of 1,038 unrelated offspring of nonagenarian siblings and the partners thereof, with GWAS data and Nightingale metabolites levels available. In accordance with the Declaration of Helsinki, we obtained informed consent from all participants prior to their entering the study. Good clinical practice guidelines were maintained. The study protocol was approved by the ethical committee of the Leiden University Medical Center before the start of the study (P01.113).

*Acknowledgements/funding*: The Leiden Longevity Study has received funding from the European Union's Seventh Framework Programme (FP7/2007-2011) under grant agreement number 259679. This study was financially supported by the Innovation-Oriented Research Program on Genomics (SenterNovem IGE05007), the Centre for Medical Systems Biology and the Netherlands Consortium for Healthy Ageing (grant 050-060-810), all in the framework of the Netherlands Genomics Initiative, Netherlands Organization for Scientific Research (NWO), by Unilever Colworth and by BBMRI-NL, a Research Infrastructure financed by the Dutch government (NWO 184.021.007 and 184.033.111).

***Lifelines-DEEP (LLD)***

*Study description*: The Lifelines-DEEP cohort[15] is a subset of 1,539 individuals, randomly selected from the prospective, population-based LifeLines cohort from northern provinces of the Netherlands (n=167,000). These participants were examined more thoroughly, specifically with respect to multidimensional omics data that include genetics, epigenetics, transcriptomics, proteomics, metabolomics and the gut microbiome. Moreover, additional biological materials and information on health status are collected. This allows for a more in-depth investigation of molecular pathways underlying the association between genetic and phenotypic variation.

*Acknowledgements/funding*: A.K. is supported by NWO Gravitation grant Exposome-NL (024.004.017). L.F. is supported by grants from the Dutch Research Council (ZonMW-VIDI 917.14.374 and ZonMW-VICI to L.F.), and by an ERC Starting Grant, grant agreement 637640 (ImmRisk) and through a Senior Investigator Grant from the Oncode Institute. JF is supported by the Dutch Heart Foundation IN-CONTROL (CVON2018-27), the ERC Consolidator grant (grant agreement No. 101001678), NWO-VICI grant VI.C.202.022, and the Netherlands Organ-on-Chip Initiative, an NWO Gravitation project (024.003.001) funded by the Ministry of Education, Culture and Science of the government of The Netherlands. A.Z. is supported by the Dutch Heart Foundation IN-CONTROL (CVON2018-27), the ERC Starting Grant 715772, NWO-VIDI grant 016.178.056, and the NWO Gravitation grant Exposome-NL (024.004.017).

***London Life Sciences Prospective Population Study (LOLIPOP)***

*Study description:* The London Life Sciences Prospective Population Study (LOLIPOP)[16,17] is a prospective population study of 28,372 South Asian and European men and women, recruited at age 35–74 years from the lists of 58 General Practitioners in West London, UK, between May 1, 2002, and Sept 12, 2008. Indian Asians had all four grandparents born on the Indian subcontinent (India, Pakistan, Sri Lanka, or Bangladesh); Europeans were of self-reported white ancestry. At enrolment, all participants completed a structured assessment of cardiovascular and metabolic health, including collection of blood samples after an eight-hour fasting time. Aliquots of whole blood were stored at -80C for extraction of genomic DNA. Serum samples were subjected to NMR profiling and genome-wide genotyping and imputation for over 10,000 participants, as described previously[16–19]. The LOLIPOP study is approved by the National Research Ethics Service (07/H0712/150) and all participants gave written informed consent at enrolment. The current study included individuals from eight sub-cohorts of the LOLIPOP study (see Table S1 for details).

*Acknowledgements/funding*: The London Life Sciences Prospective Population (LOLIPOP) Study is supported by the National Institute for Health Research (NIHR) Comprehensive Biomedical Research Centre Imperial College Healthcare NHS Trust, the British Heart Foundation (SP/04/002), the Medical Research Council (G0601966, G0700931), the Wellcome Trust (084723/Z/08/Z, 090532 & 098381) the NIHR (RP-PG-0407-10371), the NIHR Official Development Assistance (ODA, award 16/136/68), the European Union FP7 (EpiMigrant, 279143) and H2020 programs (iHealth-T2D, 643774). We acknowledge support of the MRC-PHE Centre for Environment and Health, and the NIHR Health Protection Research Unit on Health Impact of Environmental Hazards. The work was carried out in part at the NIHR/Wellcome Trust Imperial Clinical Research Facility. The views expressed are those of the author(s) and not necessarily those of the Imperial College Healthcare NHS Trust, the NHS, the NIHR or the Department of Health. We thank the participants and research staff who made the study possible. J.C. is supported by the Singapore Ministry of Health's National Medical Research Council under its Singapore Translational Research Investigator (STaR) Award (NMRC/STaR/0028/2017).

***METabolic Syndrome In Men Study (METSIM)***

*Study description*: The METSIM Study includes 10,197 men, aged from 45 to 73 years at baseline, randomly selected from the population register of the Kuopio town, Eastern Finland, and examined in 2005-2010[20]. The aim of the study is to investigate genetic and non-genetic factors associated with the risk of type 2 diabetes and cardiovascular diseases. Proton NMR data was obtained from all METSIM participants.

*Acknowledgements/funding*: MB was supported by National Institutes of Health (grant no. DK062370). ML was supported by the Academy of Finland (grant no. 321428) and the Sigrid Juselius Foundation. XY was supported by an American Diabetes Association Postdoctoral Fellowship (grant no. 1-19-PDF-061) and a University of Michigan Precision Health Scholarship.

***Northern Finland Birth Cohorts (NFBC) 1966 and 1986***

*Study descriptions*: Northern Finland Birth Cohort 1966 (NFBC1966)[21–23] is a prospective birth cohort started in 1965. At baseline all mothers with expected dates of delivery between the 1st of January and the 31st of December 1966 were recruited from the two northernmost provinces in Finland. NFBC1966 study population comprised 12,231 children and their parents. The follow-ups have been conducted at four different time points i.e. at 1, 14, 31 and 46 years of age. Collected data include prenatal and early life measurements, information on motor, social, psychological and mental development in childhood. In adulthood data has been collected on social background, lifestyle, medication, diagnosed diseases, organ-specific and psychiatric symptoms, workload and occupational health, economy, personal traits, functioning, quality of life, use of health services, and family history of diseases. The current study included eligible individuals from NFBC1966 at the age of 31 years (*n*=4,668). Northern Finland Birth Cohort 1986 (NFBC1986)[24,25] is a prospective cohort, which included mothers in the two northernmost provinces on Finland (Oulu and Lapland) with an expected date of birth between 1.7.1985 and 30.6.1986. Altogether 9,479 children were born into the cohort, 9,432 of them live-born. In addition to the data on delivery, follow-ups have been conducted at four different time points i.e. at 1, 7-8, 15-16 and 33-35 years of age. Collected data include prenatal and early life measurements, information on motor, social, psychological and mental development in childhood. In adulthood data has been collected on social background, lifestyle, medication, diagnosed diseases, organ-specific and psychiatric symptoms, workload and occupational health, economy, personal traits, functioning, quality of life, and family history of diseases. The current study included eligible individuals from NFBC1986 at the age of 15-16 years (*n*=3,215). More detailed description of NFBC can be found at https://www.oulu.fi/nfbc/.

*Acknowledgements/funding*: We thank all cohort members and researchers who participated in the study. We also wish to acknowledge the work of the NFBC project center. NFBC1966 and NFBC1986 have received core support from multiple funders. NFBC1966 31 year follow up received financial support from University of Oulu Grant no. 65354, Oulu University Hospital Grant no. 2/97, 8/97, Ministry of Health and Social Affairs Grant no. 23/251/97, 160/97, 190/97, National Institute for Health and Welfare, Helsinki Grant no. 54121, Regional Institute of Occupational Health, Oulu, Finland Grant no. 50621, 54231. NFBC1966 46 year follow up received financial support from University of Oulu Grant no. 24000692, Oulu University Hospital Grant no. 24301140, and ERDF European Regional Development Fund Grant no. 539/2010 A31592. The data generation, curation and manpower were also supported by the following EU H2020 grants for NFBC1966 and NFBC1986: DynaHEALTH (grant no 633595), LifeCycle (733206), LongITools (873749), EarlyCause, EDCMET (825762) and the Medical Research Council, UK: grant number MRC/BBSRC MR/S03658X/1 (JPI HDHL H2020). E.S. is supported by Academy of Finland (grant no 338229) and Orion Research Foundation. M.R.J. is partly funded by the Medical Research Council (MR/S019669/1). M.A.-K. is supported by by the Sigrid Juselius Foundation. J.K. is supported by the Sigrid Juselius Foundation, Academy of Finland (grant nos. 297338 and 307247) and the Novo Nordisk Foundation (NNF17OC0026062).

***The Netherlands Epidemiology of Obesity (NEO) study***

*Study description*: The Netherlands Epidemiology of Obesity (NEO) study[26] was designed for extensive phenotyping to investigate pathways that lead to obesity-related diseases. The NEO study is a population-based, prospective cohort study that includes 6,671 individuals aged 45–65 years, with an oversampling of individuals with overweight or obesity. At baseline, information on demography, lifestyle, and medical history have been collected by questionnaires. In addition, samples of 24-h urine, fasting and postprandial blood plasma and serum, and DNA were collected. Genotyping was performed using the Illumina HumanCoreExome chip, which was subsequently imputed to the 1000 genome reference panel and the Haplotype Reference Consortium (HRC) reference panel. Participants underwent an extensive physical examination, including anthropometry, electrocardiography, spirometry, and measurement of the carotid artery intima-media thickness by ultrasonography. In random subsamples of participants, magnetic resonance imaging of abdominal fat, pulse wave velocity of the aorta, heart, and brain, magnetic resonance spectroscopy of the liver, indirect calorimetry, dual energy X-ray absorptiometry, or accelerometry measurements were performed. The collection of data started in September 2008 and completed at the end of September 2012. Participants are currently being followed for the incidence of obesity-related diseases and mortality.

*Acknowledgements/funding*: The authors of the NEO study thank all individuals who participated in the Netherlands Epidemiology of Obesity study, all participating general practitioners for inviting eligible participants and all research nurses for collection of the data. We thank the NEO study group, Pat van Beelen, Petra Noordijk and Ingeborg de Jonge for the coordination, lab and data management of the NEO study. The genotyping in the NEO study was supported by the Centre National de Génotypage (Paris, France), headed by Jean-Francois Deleuze. The NEO study is supported by the participating Departments, the Division and the

Board of Directors of the Leiden University Medical Center, and by the Leiden University, Research Profile Area Vascular and Regenerative Medicine. R.L-G. is supported by Stichting Management Apothekers en de Gezondheidszorg (STIMAG). D.M.-K. is supported by Dutch Science Organization (ZonMW-VENI Grant 916.14.023).

***The Netherlands Study of Depression and Anxiety (NESDA)***

*Study description:* The Netherlands Study of Depression and Anxiety (NESDA, www.nesda.nl) is a longitudinal, multi-site, naturalistic, case-control cohort study set up to examine the etiology, course and consequences of depressive and anxiety disorders. Detailed descriptions of the rationale, design and methods for NESDA are given elsewhere[27]. Briefly, in 2004 to 2007, 2981 participants with depressive and/or anxiety disorder and healthy control subjects were recruited from the community (19%), general practice (54%), and secondary mental health care (27%). The research protocol was approved by the ethical committee of participating universities, and all respondents provided written informed consent.

*Acknowledgements/funding:* Funding was obtained from the Netherlands Organization for Scientific Research (Geestkracht program grant 10-000-1002); the Center for Medical Systems Biology (CSMB, NWO Genomics), Biobanking and Biomolecular Resources Research Infrastructure (BBMRI-NL), VU University's Institutes for Health and Care Research (EMGO+) and Neuroscience Campus Amsterdam, University Medical Center Groningen, Leiden University Medical Center, National Institutes of Health (NIH, R01D0042157-01A, MH081802, Grand Opportunity grants 1RC2 MH089951 and 1RC2 MH089995). Part of the genotyping and analyses were funded by the Genetic Association Information Network (GAIN) of the Foundation for the National Institutes of Health. Computing was supported by BiG Grid, the Dutch e-Science Grid, which is financially supported by NWO.

***Netherlands Twin Register (NTR)***

*Study description*: The Netherlands Twin Register (NTR; https://tweelingenregister.vu.nl/) recruits twins and their family members to study the causes of individual differences in health, behavior, and lifestyle. Participants of the NTR are followed longitudinally by surveys[28]. A subsample of unselected twins and their family members took part in the NTR-Biobank project in which biological samples, including DNA and RNA, were collected in a standardized manner after overnight fasting[29]. Fertile women were bled in their pill-free week or on day 2–4 of their menstrual cycle. Of the participants included in the current study, 96% had fasted for more than 12 hours. For metabolomics measurements, blood (plasma) was collected in the morning using a safety lock butterfly needle. Study protocols were approved by the Central Ethics Committee on Research Involving Human Subjects of the VU University Medical Center, Amsterdam, an Institutional Review Board certified by the US Office of Human Research Protections (IRB number IRB-2991 under Federal-wide Assurance-3703; IRB/institute codes, NTR 03-180) and informed consent was obtained from all participants. Metabolomics and genotyping information were available for 4,677 NTR participants. They come from larger group of genotyped subjects (N=23,601; NTR release M8) who were genotyped on the Affymetrix 6.0, Affymetrix Perlegen 5.0, Illumina Human Beadchip 660, Illumina Omni Express 1M, or Illumina GSA genotyping platforms, or had sequence data from the Netherlands reference genome project GONL (BGI full sequence at least 12x)[30]. Genome-wide association analyses included dummy variables to account for genotyping platform. Calls were made with platform specific software (Birdseed, ATP, and Genome studio) following manufacturers protocols. For each genotyping array samples were removed if the genotype call rate was <=0.95 or the heterozygosity (Plink F statistic) fell outside the range of −0.10 to 0.10. SNPs were removed if they were palindromic AT/GC SNPs with a MAF range between 0.4 and 0.5, if the MAF was below 0.01, if Hardy Weinberg Equilibrium (HWE) had $p$<1e-5, or if the number of Mendelian errors was greater than 20. Data of SNPs present on all platforms were first imputed cross platform with respect to the GoNL reference panel, and the result was then used for imputation on the Michigan imputation server to HRC panel[31]. Filtering of the imputed dataset included the exclusion of monomorphic variants, removal of variants with info <0.6, variants with HWE p<1e-5, and variants not in 1kGP3 reference.

*Acknowledgements/funding*: The BBMRI Metabolomics Consortium is funded by BBMRI-NL, a research infrastructure financed by the Dutch government (NWO, no. 184.021.007 and 184.033.111); the Netherlands Organization for Scientific Research: Netherlands Twin Registry Repository (NWO-480-15-001/674); OCW Gravity program, NWO-024.001.003: Why some children thrive; NWO Large Scale infrastructures, X-Omics (184.034.019); European Union FP7/2007-2013: ACTION Consortium: Aggression in Children: Unravelling gene-environment interplay to inform Treatment and InterventiON strategies (grant number 602768); NWO/SPI 56-464-14192, Genetic Association Information Network (GAIN) of the Foundation for the NIH; Rutgers University Cell and DNA Repository (NIMH U24 MH068457-06), the Avera Institute, Sioux Falls (USA); NIH R01 HD042157-01A1, MH081802, Grand Opportunity grants 1RC2 MH089951 and 1RC2 MH089995; European Research Council (ERC-230374). Genotyping support was received from Amsterdam Reproduction and Development Research Institute. D.B. has received the Royal Netherlands Academy of Science Professor Award (PAH/6635).

***The Orkney Complex Disease Study (ORCADES)***

*Study description*: The Orkney Complex Disease Study (ORCADES) is a family-based study that seeks to identify genetic factors influencing cardiovascular and other disease risk in the isolated archipelago of the Orkney Isles in northern Scotland[32]. Genetic diversity in this population is decreased compared to Mainland Scotland, consistent with the high levels of endogamy historically. 2,078 participants aged 16-100 years were recruited between 2005 and 2011, most having three or four grandparents from Orkney, the remainder with two Orcadian grandparents. Fasting blood samples were collected and many health-related phenotypes and environmental exposures were measured in each individual. All participants gave written informed consent and the study was approved by Research Ethics Committees in Orkney, Aberdeen (North of Scotland REC), and South East Scotland REC, NHS Lothian (reference: 12/SS/0151).

*Acknowledgements/funding*: The Orkney Complex Disease Study (ORCADES) was supported by the Chief Scientist Office of the Scottish Government (CZB/4/276, CZB/4/710), a Royal Society URF to J.F.W., the MRC Human Genetics Unit quinquennial programme "QTL in Health and Disease", Arthritis Research UK and the European Union framework program 6 EUROSPAN project (contract no. LSHG-CT-2006-018947).

***Oxford Biobank (OBB)***

*Study description:* The Oxford BioBank (OBB) is a collection of 30-50 year old healthy participants living in Oxfordshire. Recruitment of the participants started in 2000. All participants have undergone a detailed examination and given DNA samples. The participants have given informed consent to be re-approached. The OBB research focuses on common diseases like diabetes, obesity and cardiovascular disease. Details have been described before[33].

*Acknowledgements/funding*: We thank the volunteers from the Oxford Biobank (www.oxfordbiobank.org.uk) for their participation in this recall study. The Oxford BioBank and Oxford Bioresource are funded by the NIHR Oxford Biomedical Research Centre (BRC). The views expressed are those of the author(s) and not necessarily those of the NIHR or the Department of Health and Social care. M.I.M. is a Wellcome Senior Investigator. N.v.Z. was supported by DOLORisk. M.N. and F.K. were supported by the NIHR Oxford Biomedical Research Centre.

***PROspective Study of Pravastatin in the Elderly at Risk (PROSPER)***

*Study description:* All data come from the PROspective Study of Pravastatin in the Elderly at Risk (PROSPER). A detailed description of the study has been published elsewhere[34–36]. PROSPER was a prospective multicenter randomized placebo-controlled trial to assess whether treatment with pravastatin diminishes the risk of major vascular events in elderly. Between December 1997 and May 1999, we screened and enrolled subjects in Scotland (Glasgow), Ireland (Cork), and the Netherlands (Leiden). Men and women aged 70-82 years were recruited if they had pre-existing vascular disease or increased risk of such disease because of smoking, hypertension, or diabetes. A total number of 5,804 subjects were randomly assigned to pravastatin or placebo. A large number of prospective tests were performed including Biobank tests and cognitive function measurements. A whole genome wide screening has been performed in the sequential PHASE project. Of 5,763 subjects DNA was available for genotyping. Genotyping was performed with the Illumina 660K beadchip, after QC (call rate <95%) 5,244 subjects and 557,192 SNPs were left for analysis. These SNPs were imputed to 2.5 million SNPs based on the HAPMAP built 36 with MACH imputation software. The study was approved by the institutional ethics review boards of centers of Cork University (Ireland), Glasgow University (Scotland) and Leiden University Medical Center (the Netherlands) and all participants gave written informed consent.

*Acknowledgements/funding*: The PROSPER study was supported by an investigator initiated grant obtained from Bristol-Myers Squibb. Prof. Dr. J. W. Jukema is an Established Clinical Investigator of the Netherlands Heart Foundation (grant 2001 D 032). Support for genotyping was provided by the seventh framework program of the European commission (grant 223004) and by the Netherlands Genomics Initiative (Netherlands Consortium for Healthy Aging grant 050-060-810).

***The Rotterdam Study (RS)***

*Study description*: The Rotterdam Study (RS)[37] is a prospective population-based study designed to investigate the determinants of disease occurrence and progression in the elderly. The RS cohort was initially defined in 1990 among 7,983 persons living in the well-defined Ommoord district in Rotterdam, the Netherlands. All participants underwent a home interview and extensive physical examination at baseline and during follow-up examinations occurring every 3–4 years (RS-I). The cohort was further extended in 2000 (RS-II) and 2006 (RS-III), establishing a total of 14,926 participants aged 45 years or over. Metabolic biomarkers were successfully quantified in fasting EDTA plasma samples for the selected individuals. The RS has been approved by the Medical Ethics Committee of the Erasmus MC (registration number MEC 02.1015) and by the Dutch Ministry of Health, Welfare and Sport (Population Screening Act WBO, license number 1071272-159521-PG). The RS has been entered into the Netherlands National Trial Register (NTR; www.trialregister.nl) and into the WHO International Clinical Trials Registry Platform (ICTRP; www.who.int/ictrp/network/primary/en/) under shared catalogue number NTR6831. All participants provided written informed consent to participate in the study and to have their information obtained from treating physicians. The current study included individuals from the three sub-cohorts of the Rotterdam Study (see Table S1 for details).

*Acknowledgements/funding*: The Rotterdam Study is supported by the Erasmus MC University Medical Center and Erasmus University Rotterdam; The Netherlands Organisation for Scientific Research (NWO); The Netherlands Organisation for Health Research and Development (ZonMw); the Research Institute for Diseases in the Elderly (RIDE); The

Netherlands Genomics Initiative (NGI); the Ministry of Education, Culture and Science; the Ministry of Health, Welfare and Sports; the European Commission (DG XII); and the Municipality of Rotterdam. The authors are grateful to the Rotterdam Study participants and staff, and in particular, general practitioners and pharmacists. Metabolomics measurements were funded by Biobanking and Biomolecular Resources Research Infrastructure (BBMRI)–NL (184.021.007) and the JNPD under the project PERADES (grant number 733051021, Defining Genetic, Polygenic and Environmental Risk for Alzheimer's Disease using multiple powerful cohorts, focused Epigenetics and Stem cell metabolomics).

***TwinsUK***

*Study description*: TwinsUK is a national register of adult twins recruited as volunteers without selecting for any particular disease or traits[38]. The cohort was initially developed to investigate the heritability and genetics of disease prevalence in women. It now comprises over 14,000 predominantly female twins and is representative of the British general population.

*Acknowledgements/funding*: The Dept of Twin Research receives support from grants from the Wellcome Trust (212904/Z/18/Z) and the Medical Research Council (MRC)/British Heart Foundation Ancestry and Biological Informative Markers for Stratification of Hypertension (AIMHY; MR/M016560/1), European Union, Chronic Disease Research Foundation (CDRF), Zoe Global Ltd, NIH and the National Institute for Health Research (NIHR)-funded BioResource, Clinical Research Facility and Biomedical Research Centre based at Guy's and St Thomas' NHS Foundation Trust in partnership with King's College London. C.M. is funded by the Chronic Disease Research Foundation (CDRF).

***The Cardiovascular Risk in Young Finns Study (YFS)***

*Study description*: The Cardiovascular Risk in Young Finns Study (YFS) is a prospective multi-center follow-up study investigating cardiovascular risk factors from childhood to adulthood[39]. The study was initiated in 1980 with 3596 children and adolescents aged 3–18 years. The participants were randomly selected from the areas of five university hospitals in Finland (Turku, Tampere, Helsinki, Kuopio and Oulu) and have been followed for over 40 years. The serum samples used ($n$=1,948) in this study were collected in 2007.

*Acknowledgements/funding*: The Young Finns Study has been financially supported by the Academy of Finland: grants 322098, 286284, 134309 (Eye), 126925, 121584, 124282, 129378 (Salve), 117787 (Gendi), 41071 (Skidi), 330809, 338395 and 349708 (PPM); the Social Insurance Institution of Finland; Competitive State Research Financing of the Expert Responsibility area of Kuopio, Tampere and Turku University Hospitals (grant X51001); Juho Vainio Foundation; Paavo Nurmi Foundation; Finnish Foundation for Cardiovascular Research; Finnish Cultural Foundation; Sigrid Jusélius Foundation; Tampere Tuberculosis Foundation; Emil Aaltonen Foundation; Yrjö Jahnsson Foundation; Signe and Ane Gyllenberg Foundation; Diabetes Research Foundation of Finnish Diabetes Association; This project has received funding from the European Union's Horizon 2020 research and innovation programme under grant agreements No 848146 (To Aition) and No 755320 (TAXINOMISIS); This project has received funding from the European Research Council (ERC) advanced grants under grant agreement No 742927 (MULTIEPIGEN project); Tampere University Hospital Supporting Foundation and Finnish Society of Clinical Chemistry.

***FinnGen***

*Study description*: In the present study, we used GWAS summary statistics of 3,095 disease endpoints from FinnGen Data Freeze 7. The FinnGen (www.finngen.fi/en) study[1] was launched in 2017 as a public-private collaboration. The project aims to improve human health by combining genetic information with digital health care data: the genotype data from a nationwide network of Finnish biobanks are linked to national hospital discharge, death, cancer, and medication reimbursement registries using the unique national personal identification codes. Once ~200,000 existing samples from Finnish biobanks are combined with ~300,000 samples from the ongoing collections, the data resource will cover roughly 10% of the Finnish population. The present study comprised data from FinnGen Preparatory Phase Data Freeze 7, consisting of 309,154 individuals. A full list of FinnGen contributors (banner authors) is presented in Supplementary Table S11.

FinnGen participants provided informed consent for biobank research, based on the Finnish Biobank Act. Alternatively, separate research cohorts, collected prior the Finnish Biobank Act came into effect (in September 2013) and start of FinnGen (August 2017), were collected based on study-specific consents and later transferred to the Finnish biobanks after approval by Fimea (Finnish Medicines Agency), the National Supervisory Authority for Welfare and Health. Recruitment protocols followed the biobank protocols approved by Fimea. The Coordinating Ethics Committee of the Hospital District of Helsinki and Uusimaa (HUS) statement number for the FinnGen study is Nr HUS/990/2017.

The FinnGen study is approved by Finnish Institute for Health and Welfare (permit numbers: THL/2031/6.02.00/2017, THL/1101/5.05.00/2017, THL/341/6.02.00/2018, THL/2222/6.02.00/2018, THL/283/6.02.00/2019, THL/1721/5.05.00/2019, THL/1524/5.05.00/2020, and THL/2364/14.02/2020), Digital and population data service agency (permit numbers: VRK43431/2017-3, VRK/6909/2018-3, VRK/4415/2019-3), the

Social Insurance Institution (permit numbers: KELA 58/522/2017, KELA 131/522/2018, KELA 70/522/2019, KELA 98/522/2019, KELA 138/522/2019, KELA 2/522/2020, KELA 16/522/2020, Findata THL/2364/14.02/2020 and Statistics Finland (permit numbers: TK-53-1041-17 and TK/143/07.03.00/2020 (earlier TK-53-90-20).

The Biobank Access Decisions for FinnGen samples and data utilized in FinnGen Data Freeze 7 include: THL Biobank BB2017_55, BB2017_111, BB2018_19, BB_2018_34, BB_2018_67, BB2018_71, BB2019_7, BB2019_8, BB2019_26, BB2020_1, Finnish Red Cross Blood Service Biobank 7.12.2017, Helsinki Biobank HUS/359/2017, Auria Biobank AB17-5154 and amendment #1 (August 17 2020), Biobank Borealis of Northern Finland_2017_1013, Biobank of Eastern Finland 1186/2018 and amendment 22 § /2020, Finnish Clinical Biobank Tampere MH0004 and amendments (21.02.2020 & 06.10.2020), Central Finland Biobank 1-2017, and Terveystalo Biobank STB 2018001.

Genotyping of the FinnGen samples was performed using Illumina and Affymetrix arrays (Illumina Inc., San Diego, and Thermo Fisher Scientific, Santa Clara, CA, USA). In sample quality control (QC), individuals with high genotype missingness (>5%), ambiguous gender, excess heterozygosity (±4SD) and non-Finnish ancestry were excluded. In variant QC, variants with low Hardy-Weinberg equilibrium (HWE) p-value (<1e-06), high missingness (>2%) and minor allele count (MAC)<3 were excluded. Chip genotyped samples were pre-phased with Eagle 2.3.5 with the number of conditioning haplotypes set to 20,000. Genotype imputation was carried out by using the Finnish population-specific SISu v3 reference panel with Beagle 4.1 (version 08Jun17.d8b) as described in the following protocol:

dx.doi.org/10.17504/protocols.io.nmndc5e. In post-imputation QC, variants with imputation INFO<0.6 were excluded. In FinnGen, GWASs were completed using the Scalable and Accurate Implementation of Generalized (SAIGE) software[40]. Age, sex, the first 10 genetic principal components, and genotyping batch were included as covariates and only variants with minimum allele count of 5 were included in association testing.

*Acknowledgements/funding*: We want to acknowledge the participants and investigators of FinnGen study. The FinnGen project is funded by two grants from Business Finland (HUS 4685/31/2016 and UH 4386/31/2016) and the following industry partners: AbbVie Inc., AstraZeneca UK Ltd, Biogen MA Inc., Bristol Myers Squibb (and Celgene Corporation & Celgene International II Sàrl), Genentech Inc., Merck Sharp & Dohme Corp, Pfizer Inc., GlaxoSmithKline Intellectual Property Development Ltd., Sanofi US Services Inc., Maze Therapeutics Inc., Janssen Biotech Inc, Novartis AG, and Boehringer Ingelheim. Following biobanks are acknowledged for delivering biobank samples to FinnGen: Auria Biobank (www.auria.fi/biopankki), THL Biobank (www.thl.fi/biobank), Helsinki Biobank (www.helsinginbiopankki.fi), Biobank Borealis of Northern Finland (https://www.ppshp.fi/Tutkimus-ja-opetus/Biopankki/Pages/Biobank-Borealis-briefly-in-English.aspx), Finnish Clinical Biobank Tampere (www.tays.fi/en-US/Research_and_development/Finnish_Clinical_Biobank_Tampere), Biobank of Eastern Finland (www.ita-suomenbiopankki.fi/en), Central Finland Biobank (www.ksshp.fi/fi-FI/Potilaalle/Biopankki), Finnish Red Cross Blood Service Biobank (www.veripalvelu.fi/verenluovutus/biopankkitoiminta) and Terveystalo Biobank (www.terveystalo.com/fi/Yritystietoa/Terveystalo-Biopankki/Biopankki/). All Finnish Biobanks are members of BBMRI.fi infrastructure (www.bbmri.fi). Finnish Biobank Cooperative -FINBB (https://finbb.fi/) is the coordinator of BBMRI-ERIC operations in

Finland. The Finnish biobank data can be accessed through the Fingenious® services (https://site.fingenious.fi/en/) managed by FINBB.

***Other funding***

F.A. is supported by UCL Hospitals NIHR Biomedical Research Centre.
